## Supplementary material for "Comprehensive profiling of the human intestinal DNA virome and prediction of disease-associated bacterial hosts in severe Myalgic Encephalomyelitis/Chronic Fatigue Syndrome (ME/CFS)": S Table 10 WMS viral cluster analysis

| | VC | Size | Internal Weight | External Weight | Quality | P-value | Min Dist | Max Dist | Total Dist | Below Thres | Taxon Prediction Score | Avg Dist | Genera | Families | Orders | Members |
|---|---|---|---|---|---|---|---|---|---|---|---|---|---|---|---|---|
| 0 | VC_0_0 | 3 | 77.25303487 | 0 | 1 | 0 | 1 | 2 | 3 | 3 | 1 | 1.577350269 | 0 | 0 | 0 | AB002632,KC357596,OP297622 |
| 1 | VC_1000_0 | 21 | 1764.69294 | 326.8331113 | 0.843734621 | 2.47E-08 | 0 | 2.236067977 | 210 | 210 | 1 | 1.45145382 | 0 | 0 | 0 | MG945161,MK249131,MK249144,MK249156,MK249159,MK249160,MK249167,MK249179,MK249181,MZ364226,MZ364241,MZ364244,MZ364245,MZ364246,MZ364247,MZ364249,MZ364250,MZ364260,MZ364263,MZ364266,MZ364270 |
| 2 | VC_1002_0 | 2 | 5.394552787 | 24.43350627 | 0.180854972 | 0.279092325 | 5.567764363 | 5.567764363 | 1 | 1 | 1 | 5.567764363 | 0 | 0 | 0 | MT152698,MZ334527 |
| 3 | VC_1003_0 | 4 | 613.6260189 | 90.27156737 | 0.871754685 | 0.00016219 | 5 | 6.782329983 | 6 | 6 | 1 | 6.016528108 | 0 | 0 | 0 | MN698240,MN698245,MN698246,MN698248 |
| 4 | VC_1004_0 | 2 | 212.1262903 | 245.2302544 | 0.463809456 | 0.057744643 | 0 | 0 | 1 | 1 | 1 | 0 | 0 | 0 | 0 | MW202462,MW202544 |
| 5 | VC_1006_0 | 8 | 7918.777591 | 2772.145143 | 0.74070104 | 4.06E-14 | 0 | 8.774964387 | 28 | 28 | 1 | 6.710466857 | 0 | 0 | 0 | KC595511,KT070866,MG967616,MG967617,MG967618,MK288022,MW392801,MW392802 |
| 6 | VC_1007_0 | 19 | 26158.92735 | 4259.906749 | 0.85995825 | 5.31E-42 | 0 | 4 | 171 | 171 | 1 | 2.534708956 | 0 | 0 | 0 | MT074134,MT074135,MT074143,MT074144,MT074145,MT074147,MT074148,MT074149,MT074150,MT074151,MT074152,MT074153,MT074154,MT074155,MT074156,MT074157,MT074158,MT074159,MT074160 |
| 7 | VC_1008_0 | 4 | 615.7103743 | 316.9364352 | 0.660175286 | 6.46E-05 | 1.732050808 | 6 | 6 | 6 | 1 | 4.451677537 | 0 | 0 | 0 | MT635598,MT806185,MT806186,S4-wms_141607 |
| 8 | VC_1009_0 | 3 | 454.8895356 | 3127.647895 | 0.126974119 | 0.001855388 | 1.732050808 | 3 | 3 | 3 | 1 | 2.520159311 | 0 | 0 | 0 | ON721384,ON721385,S7-wms_244866 |
| 9 | VC_100_0 | 15 | 18911.93957 | 10023.80534 | 0.653583989 | 3.42E-16 | 0 | 8.246211251 | 105 | 105 | 1 | 5.36854133 | 0 | 0 | 0 | KC330679,KC330680,KC330681,KC330682,KC330683,KC330684,KF669648,KF669651,KF669659,KU836751,MG983742,MH884513,MN727882,MT422786,MT514532 |
| 10 | VC_1010_0 | 2 | 7.940981737 | 31.06519054 | 0.203582697 | 0.226627352 | 5.477225575 | 5.477225575 | 1 | 1 | 1 | 5.477225575 | 0 | 0 | 0 | MN732896,S11-wms_330479 |
| 11 | VC_1011_0 | 9 | 1542.44428 | 0 | 1 | 0 | 0 | 1.732050808 | 36 | 36 | 1 | 0.962250449 | 0 | 0 | 0 | KC821606,KC821623,KC821628,KC821631,MT732445,MT732446,MT732447,MT732448,MT732449 |
| 12 | VC_1012_0 | 4 | 1231.694827 | 181.6559425 | 0.871471438 | 4.61E-05 | 6.8556546 | 7.615773106 | 6 | 6 | 1 | 7.27476053 | 0 | 0 | 0 | KC821608,KC821620,KC821622,KC821633 |
| 13 | VC_1013_0 | 34 | 94222.53757 | 578.6058074 | 0.993896637 | 1.78E-58 | 0 | 7.141428429 | 561 | 561 | 1 | 5.010295196 | 0 | 0 | 0 | MN812206,MN812207,MN812208,MN812209,MN812210,MN812212,MN812213,MN812214,MN812215,MN812216,MN812217,MN812218,MN812219,MN812220,MN812221,MN812222,MN812223,MN812224,MN812225,MN812226,MN812227,MN812228,MN812229,MN812230,MN812231,MN812232,MN812233,MN812234,MN812235,MN812236,MN812237,MN812238,MN812240,MN812241 |
| 14 | VC_1014_0 | 3 | 50.89806485 | 200.2682651 | 0.202646847 | 0.144087817 | 7.211102551 | 9 | 3 | 2 | 0.666666667 | 8.290452196 | 0 | 0 | 0 | S11-wms-combined_92114,S17-wms-combined_199746,S7-wms-combined_163980 |
| 15 | VC_1015_0 | 62 | 553967.0099 | 2972.02003 | 0.994663653 | 1.16E-278 | 0 | 3.872983346 | 1891 | 1891 | 1 | 1.145404917 | 0 | 0 | 0 | MT497017,MT497065,MT497066,MT497067,MT497068,MT497069,MT497070,MT497071,MT497072,MT497073,MT497074,MT497075,MT497076,MT497077,MT497078,MT497079,MT497080,MT497081,MT497082,MT497083,MT497084,MT497085,MT497086,MT497087,MT497088,MT497089,MT497090,MT497091,MT497092,MT497093,MT497094,MT497095,MT497096,MT497097,MT497098,MT497099,MT497100,MT497101,MT497102,MT497103,MT497104,MT497105,MT497106,MT497107,MT497108,MT497109,MT497110,MT497111,MT497112,MT497113,MT497114,MT497115,MT497116,MT497117,MT497118,MT497119,MT497120,MT497121,MT497122,MT497123,MT497124,MT497125 |
| 16 | VC_1016_0 | 8 | 5433.763075 | 297.5763986 | 0.948079083 | 9.82E-16 | 0 | 0 | 28 | 28 | 1 | 0 | 0 | 0 | 0 | MT732435,MT732436,MT732437,MT732438,MT732439,MT732440,MT732441,MT732442 |
| 17 | VC_1017_0 | 5 | 3000 | 74.05087486 | 0.975910979 | 3.90E-06 | 0 | 4.898979486 | 10 | 10 | 1 | 2.939387691 | 0 | 0 | 0 | KC821609,KC821632,KT962245,KT962246,KT962247 |
| 18 | VC_1018_0 | 2 | 188.1416897 | 2021.96321 | 0.085127946 | 0.055980677 | 5.830951895 | 5.830951895 | 1 | 1 | 1 | 5.830951895 | 0 | 0 | 0 | KJ003983,MT774389 |
| 19 | VC_1018_1 | 10 | 7017.239745 | 2316.291925 | 0.751831139 | 1.94E-11 | 2 | 9.539392014 | 45 | 36 | 0.8 | 8.188178164 | 0 | 0 | 0 | MH675552,MT074136,MT074138,MT774378,MT774385,MT774387,MT774388,MT774390,MT774391,MT774392 |
| 20 | VC_1019_0 | 5 | 3000 | 464.0184448 | 0.866046197 | 1.33E-08 | 0 | 2.449489743 | 10 | 10 | 1 | 1.469693846 | 0 | 0 | 0 | KC821612,KC821614,MT732432,MT732433,MT732434 |
| 21 | VC_101_0 | 3 | 497.2311074 | 876.078347 | 0.362067781 | 0.002351157 | 2.449489743 | 5.385164807 | 3 | 3 | 1 | 4.278218183 | 0 | 0 | 0 | KY565347,MH884514,OP341625 |
| 22 | VC_1020_0 | 5 | 1846.320447 | 386.2957948 | 0.826976178 | 2.96E-07 | 0 | 4.242640687 | 10 | 10 | 1 | 3.328642689 | 0 | 0 | 0 | KC821613,KC821615,KC821617,KC821619,KC821627 |
| 23 | VC_1021_0 | 10 | 4853.66425 | 614.4769338 | 0.887625993 | 1.89E-20 | 0 | 6.8556546 | 45 | 45 | 1 | 4.191349673 | 0 | 0 | 0 | MK764442,MK764445,MK764448,MN812203,MN812204,MN812205,MW421581,MW421582,MW421583,MW421584 |
| 24 | VC_1022_0 | 4 | 1476.028711 | 358.302652 | 0.804668524 | 3.91E-05 | 1 | 3.605551275 | 6 | 6 | 1 | 2.469417422 | 0 | 0 | 0 | KY206887,MK017819,MN417334,OP381444 |
| 25 | VC_1023_0 | 8 | 4865.42966 | 809.4334527 | 0.857365114 | 4.02E-14 | 1 | 5.656854249 | 28 | 28 | 1 | 3.918338992 | 0 | 0 | 0 | KC959568,KU599887,KU599888,KU599889,MK764437,MK764440,MK764450,MW421576 |
| 26 | VC_1024_0 | 2 | 300 | 0 | 1 | 0 | 0 | 0 | 1 | 1 | 1 | 0 | 0 | 0 | 0 | KC960489,KF981876 |
| 27 | VC_1025_0 | 4 | 1800 | 0 | 1 | 1 | 1 | 3.464101615 | 6 | 6 | 1 | 2.665279944 | 0 | 0 | 0 | KF005317,KF005318,KF005319,KF005320 |
| 28 | VC_1026_0 | 2 | 2.699997311 | 6.368472666 | 0.297734603 | 0.185546685 | 7.141428429 | 7.141428429 | 1 | 1 | 1 | 7.141428429 | 0 | 0 | 0 | KX257490,MW805362 |
| 29 | VC_1027_0 | 20 | 17450.45256 | 7080.649275 | 0.711360324 | 9.35E-59 | 0 | 7.211102551 | 190 | 190 | 1 | 5.585094879 | 0 | 0 | 0 | KF322026,KF800937,KJ502657,KY290756,KY626176,KY626177,MF481197,MG018929,MG545917,MH113812,MH113813,MH113814,MH113815,MK455769,MK455770,MK820013,MN807240,MT941682,MW451247,MW865291 |
| 30 | VC_1028_0 | 8 | 8400 | 609.8557848 | 0.93231237 | 7.68E-17 | 1 | 3.741657387 | 28 | 28 | 1 | 2.48352807 | 0 | 0 | 0 | KU245542,OK638201,OK638202,OK638203,OL343755,OL473897,OL622073,ON857939 |
| 31 | VC_1029_0 | 4 | 1800 | 198.6255297 | 0.900618937 | 4.24E-05 | 2.449489743 | 4.582575695 | 6 | 6 | 1 | 3.64296812 | 0 | 0 | 0 | MW677511,MW677527,OP009274,OP009281 |
| 32 | VC_102_0 | 2 | 35.70708253 | 277.4142268 | 0.114035939 | 0.053624467 | 7.348469228 | 7.348469228 | 1 | 1 | 1 | 7.348469228 | 0 | 0 | 0 | KJ159566,KT001912 |
| 33 | VC_1030_0 | 3 | 639.8512308 | 170.5937414 | 0.789506077 | 0.004575844 | 6 | 7.211102551 | 3 | 3 | 1 | 6.760723454 | 0 | 0 | 0 | MG596799,MK473373,MZ605292 |
| 34 | VC_1031_0 | 4 | 1566.723002 | 420.5238217 | 0.788388731 | 2.38E-05 | 5.196152423 | 6.557438524 | 6 | 6 | 1 | 5.843023616 | 0 | 0 | 0 | MN096357,MN096358,MT818426,ON526969 |
| 35 | VC_1032_0 | 7 | 3021.390644 | 34.59405021 | 0.988679901 | 4.96E-06 | 1.414213562 | 4.472135955 | 21 | 21 | 1 | 2.984375755 | 0 | 0 | 0 | KF669655,KF669657,KF669660,KM236247,KM236248,KP411017,KT001911 |
| 36 | VC_1034_0 | 2 | 145.6618651 | 1905.736805 | 0.071006122 | 0.046736356 | 8 | 8 | 1 | 1 | 1 | 8 | 0 | 0 | 0 | ON645346,OP580516 |
| 37 | VC_1034_1 | 1 | 0 | 351.2232494 | 0 | 0 | 0 | 0 | 1 | 1 | 1 | 0 | 0 | 0 | 0 | KF692088 |
| 38 | VC_1035_0 | 6 | 2585.886475 | 2741.774604 | 0.485369928 | 1.23E-10 | 3.464101615 | 5.744562647 | 15 | 15 | 1 | 4.896734247 | 0 | 0 | 0 | MH834603,MH834605,MH834611,MH834614,MH834623,ON970561 |
| 39 | VC_1036_0 | 11 | 9323.246069 | 4889.391776 | 0.655982807 | 4.76E-31 | 2.449489743 | 9.591663047 | 55 | 48 | 0.872727273 | 7.824293519 | 0 | 0 | 0 | MH834598,MH834612,MH834625,MH910036,MH910037,MH910041,MW712719,ON724017,ON970598,OP434442,OP434463 |
| 40 | VC_1037_0 | 6 | 2456.770905 | 1156.974224 | 0.679840669 | 2.17E-10 | 3.605551275 | 6.08276253 | 15 | 15 | 1 | 4.992253228 | 0 | 0 | 0 | MH834621,MH834627,MK392368,MZ150784,MZ274308,ON392166 |
| 41 | VC_1039_0 | 5 | 1699.537165 | 0 | 1 | 0 | 0 | 3.605551275 | 10 | 10 | 1 | 2.682239499 | 0 | 0 | 0 | KF751793,KF751794,KF751795,KF751796,KF751797 |
| 42 | VC_103_0 | 6 | 4294.375821 | 1558.407207 | 0.733732277 | 2.22E-10 | 1 | 3.464101615 | 15 | 15 | 1 | 2.598958212 | 0 | 0 | 0 | MN176222,MN176223,MN176224,MN176225,MN176226,MN176229 |
| 43 | VC_1040_0 | 23 | 60461.10187 | 2488.877802 | 0.960462612 | 5.32E-41 | 0 | 1.732050808 | 253 | 253 | 1 | 0.207235319 | 0 | 0 | 0 | KT876725,KU599880,KU599881,KU599882,KU599883,KU599884,KU599885,MK764436,MK764438,MK764439,MK764443,MK764444,MK764446,MK764447,OK268422,OK268423,OK268424,OK268425,OK268426,OK268427,OK340817,OK340818,OK340819 |
| 44 | VC_1041_0 | 9 | 5541.177777 | 2930.572458 | 0.654077094 | 1.10E-21 | 0 | 5.385164807 | 36 | 36 | 1 | 3.49415694 | 0 | 0 | 0 | MG592391,MG592421,MG592436,MG592443,MG592445,MG592449,MG592661,OX241561,OX241569 |
| 45 | VC_1043_0 | 2 | 300 | 4909.815617 | 0.057583612 | 0.043457527 | 0 | 0 | 1 | 1 | 1 | 0 | 0 | 0 | 0 | MN183282,MT771347 |
| 46 | VC_1044_0 | 3 | 788.466022 | 8563.810208 | 0.084307392 | 0.001476374 | 3.31662479 | 3.872983346 | 3 | 3 | 1 | 3.643755174 | 0 | 0 | 0 | OM913586,ON970564,ON970608 |
| 47 | VC_1045_0 | 2 | 156.6572271 | 206.1842793 | 0.431751121 | 0.05601967 | 5.916079783 | 5.916079783 | 1 | 1 | 1 | 5.916079783 | 0 | 0 | 0 | MN988468,MN988516 |
| 48 | VC_1046_0 | 7 | 4220.673924 | 263.2927084 | 0.941281296 | 2.16E-11 | 0 | 4.123105626 | 21 | 21 | 1 | 2.712907607 | 0 | 0 | 0 | KJ510413,KM591905,MF155936,MG925349,MH001451,MK494089,MN735432 |
| 49 | VC_1047_0 | 5 | 1430.948215 | 734.1050901 | 0.660929785 | 6.85E-08 | 0 | 8.185352772 | 10 | 10 | 1 | 6.112687395 | 0 | 0 | 0 | MW584157,MW584173,MW584189,MW584206,MW584209 |
| 50 | VC_1048_0 | 3 | 280.8724475 | 0 | 1 | 0 | 1.414213562 | 1.732050808 | 3 | 3 | 1 | 1.626105059 | 0 | 0 | 0 | KJ617393,LK392619,Z47794 |
| 51 | VC_1049_0 | 8 | 5407.635344 | 3024.792047 | 0.641290472 | 4.95E-18 | 0 | 8.888194417 | 28 | 28 | 1 | 7.273862469 | 0 | 0 | 0 | KJ802832,KX987158,KY002061,MH586730,MZ726797,OL604152,ON602746,OP617744 |
| 52 | VC_104_0 | 6 | 2530.405176 | 521.4160736 | 0.829145933 | 4.69E-10 | 2.645751311 | 7.810249676 | 15 | 15 | 1 | 6.109451441 | 0 | 0 | 0 | MH752385,MN013088,MN176219,MN176227,MN176228,X97918 |
| 53 | VC_1050_0 | 24 | 80027.74598 | 71723.91298 | 0.527359941 | 1.01E-131 | 0 | 6.164414003 | 276 | 276 | 1 | 1.905224889 | 0 | 0 | 0 | MG592411,MG592538,MG592563,MG592564,MG592565,MG592571,MG592632,MG592633,MG592634,MG592635,MG592636,MG592637,MG592638,MG592639,MG592640,MG592642,MG592645,MG592646,MG592648,MG592649,MG592650,MG592651,MG592652,MG592653 |
| 54 | VC_1050_1 | 29 | 68140.94969 | 72040.74473 | 0.486090213 | 2.39E-176 | 0 | 9.486832981 | 406 | 389 | 0.958128079 | 7.897424148 | 0 | 0 | 0 | MG592397,MG592406,MG592407,MG592408,MG592416,MG592417,MG592428,MG592463,MG592464,MG592471,MG592475,MG592476,MG592492,MG592493,MG592528,MG592601,MG592602,MG592617,MG592618,MG592655,MH925092,MW824418,MW824430,MW824432,MW824433,MW824434,OX241427,OX241556,OX241563 |
| 55 | VC_1051_0 | 2 | 76.66564858 | 0 | 1 | 0 | 4.795831523 | 4.795831523 | 1 | 1 | 1 | 4.795831523 | 0 | 0 | 0 | KM058087,MW900438 |
| 56 | VC_1052_0 | 2 | 300 | 3558.158445 | 0.077757304 | 0.053993533 | 1 | 1 | 1 | 1 | 1 | 1 | 0 | 0 | 0 | MT234342,MT234343 |
| 57 | VC_1052_1 | 1 | 0 | 3801.338718 | 0 | 0 | 0 | 0 | 1 | 1 | 1 | 0 | 0 | 0 | 0 | KP881232 |
| 58 | VC_1052_2 | 1 | 0 | 3340.460153 | 0 | 0 | 0 | 0 | 1 | 1 | 1 | 0 | 0 | 0 | 0 | MF403007 |
| 59 | VC_1052_3 | 5 | 3000 | 13220.40126 | 0.184952268 | 0.013439523 | 4.582575695 | 7.141428429 | 10 | 10 | 1 | 6.239792949 | 0 | 0 | 0 | KM199770,MN549360,MN549361,MT778837,MT778839 |
| 60 | VC_1052_4 | 6 | 4500 | 13966.28309 | 0.243687372 | 0.002040746 | 0 | 8.660254038 | 15 | 15 | 1 | 4.744674703 | 0 | 0 | 0 | MN988486,MN988509,MN988520,MN988532,MN988534,MN988553 |
| 61 | VC_1053_0 | 1 | 0 | 62.55824749 | 0 | 0 | 0 | 0 | 1 | 1 | 1 | 0 | 0 | 0 | 0 | MG945594 |
| 62 | VC_1054_0 | 12 | 366.5388527 | 913.8579835 | 0.286269727 | 0.00253356 | 0 | 2.828427125 | 66 | 66 | 1 | 2.095388801 | 0 | 0 | 0 | KM589498,KT264826,KT264827,MG945473,MG945703,MH931003,MK496723,MN379633,MN379651,OM522999,OM523001,OM869624 |
| 63 | VC_1056_0 | 4 | 72.93892166 | 358.720217 | 0.168973422 | 0.158893734 | 0 | 2.645751311 | 6 | 6 | 1 | 1.794280176 | 0 | 0 | 0 | KT264807,KT264829,KX513869,MG945560 |
| 64 | VC_1058_0 | 2 | 8.282189768 | 8.07565711 | 0.506312954 | 0.270145687 | 2.236067977 | 2.236067977 | 1 | 1 | 1 | 2.236067977 | 0 | 0 | 0 | OM869532,OM869702 |
| 65 | VC_1059_0 | 3 | 18.75727903 | 28.99904547 | 0.392770575 | 0.016005398 | 1.414213562 | 2 | 3 | 3 | 1 | 1.804737854 | 0 | 0 | 0 | MG945835,MH992170,MH992193 |
| 66 | VC_1061_0 | 2 | 20.86797703 | 8.66893268 | 0.706505089 | 0.239750061 | 0 | 0 | 1 | 1 | 1 | 0 | 0 | 0 | 0 | KM589499,KM589507 |
| 67 | VC_1062_0 | 45 | 6527.953136 | 178.3586877 | 0.973404355 | 0.490501528 | 0 | 1.732050808 | 990 | 990 | 1 | 0.997707419 | 0 | 0 | 0 | KM589500,KM589503,KM589504,KM589505,KM589509,KM589510,KM589512,KM589513,KP087948,MG945178,MG945463,MH572367,MH572368,MH572377,MH572378,MH572379,MH572459,MH572460,MH572461,MH572462,MH572463,MH572465,MH992180,MK249133,MK249138,MK249191,MK249199,MK249207,MK249208,MK249228,MK249232,MK765579,MK765582,MK765639,MK765642,MK765643,MK765644,MK765646,MN311488,MN311489,MN311490,MN311491,MZ364242,OL617032,OL617033 |
| 68 | VC_1066_0 | 52 | 16759.48627 | 5058.433614 | 0.768152343 | 1.05E-284 | 0 | 2.645751311 | 1326 | 1326 | 1 | 1.350829078 | 0 | 0 | 0 | KM589508,KT264808,KT264809,KT264810,MH992165,MK249127,MK249128,MK249129,MK249139,MK249149,MK249150,MK249153,MK249154,MK249155,MK249158,MK249162,MK249163,MK249166,MK249168,MK249174,MK249176,MK249178,MK249180,MK249188,MK249192,MK249194,MK249195,MK249196,MK249198,MK249204,MK249212,MK249214,MK249219,MN862338,MZ364222,MZ364223,MZ364232,MZ364248,MZ364254,MZ364255,MZ364261,MZ364262,MZ364264,MZ364265,MZ364268,MZ364269,MZ364271,MZ364272,MZ364273,MZ364275,MZ364276,MZ364278 |
| 69 | VC_1067_0 | 2 | 8.583214966 | 708.0442186 | 0.011977235 | 0.141922278 | 1.414213562 | 1.414213562 | 1 | 1 | 1 | 1.414213562 | 0 | 0 | 0 | MK249132,MK249205 |
| 70 | VC_1068_0 | 5 | 81.24792706 | 0 | 1 | 0 | 0 | 1.414213562 | 10 | 10 | 1 | 0.882842712 | 0 | 0 | 0 | KM589514,MK249222,MZ364231,MZ364280,MZ364284 |
| 71 | VC_1069_0 | 4 | 65.20443085 | 0 | 1 | 0 | 0 | 1.414213562 | 6 | 6 | 1 | 1.178511302 | 0 | 0 | 0 | KM589515,KM589516,MT135405,OM869497 |
| 72 | VC_1070_0 | 2 | 300 | 2008.693666 | 0.129943615 | 0.046247325 | 7.141428429 | 7.141428429 | 1 | 1 | 1 | 7.141428429 | 0 | 0 | 0 | MH153807,MH153812 |
| 73 | VC_1070_1 | 1 | 0 | 1264.24364 | 0 | 0 | 0 | 0 | 1 | 1 | 1 | 0 | 0 | 0 | 0 | KX774321 |
| 74 | VC_1071_0 | 20 | 57000 | 117435.2819 | 0.326768756 | 1.31E-102 | 2 | 9.055385138 | 190 | 187 | 0.984210526 | 6.328649617 | 0 | 0 | 0 | MH576964,MH576965,MH576968,MH590597,MH590599,MH727564,MK359332,MK359351,MK801722,MN369750,MN369757,MN428060,MN484599,MW291014,MW507134,ON260819,ON260828,ON970579,ON970590,OP021680 |
| 75 | VC_1071_1 | 1 | 0 | 12914.8746 | 0 | 0 | 0 | 0 | 1 | 1 | 1 | 0 | 0 | 0 | 0 | OL829978 |
| 76 | VC_1071_2 | 5 | 3000 | 53141.80486 | 0.053436116 | 2.64E-05 | 3.464101615 | 9.38083152 | 10 | 9 | 0.9 | 7.174740102 | 0 | 0 | 0 | MF155946,MT310865,MT684590,MT897905,ON970591 |
| 77 | VC_1071_3 | 16 | 36000 | 116914.0826 | 0.235426322 | 9.11E-57 | 3.31662479 | 9.327379053 | 120 | 116 | 0.966666667 | 7.367608895 | 0 | 0 | 0 | KM652554,MF347636,MF347637,MF347638,MF347639,MF358541,MF358542,MK460248,MK494112,MK977711,MN096371,MN369743,MW507136,MZ958750,OK310502,ON108650 |
| 78 | VC_1071_4 | 1 | 0 | 12034.67914 | 0 | 0 | 0 | 0 | 1 | 1 | 1 | 0 | 0 | 0 | 0 | MN444876 |
| 79 | VC_1071_5 | 1 | 0 | 11767.87188 | 0 | 0 | 0 | 0 | 1 | 1 | 1 | 0 | 0 | 0 | 0 | MT711976 |
| 80 | VC_1072_0 | 2 | 300 | 11048.06758 | 0.026436219 | 0.048825988 | 3.605551275 | 3.605551275 | 1 | 1 | 1 | 3.605551275 | 0 | 0 | 0 | MG757153,MK620896 |
| 81 | VC_1072_1 | 2 | 300 | 11282.93201 | 0.025900178 | 0.048719576 | 4 | 4 | 1 | 1 | 1 | 4 | 0 | 0 | 0 | MT024865,MT024872 |
| 82 | VC_1072_2 | 5 | 3000 | 26912.46432 | 0.100292639 | 6.85E-06 | 2.236067977 | 5.099019514 | 10 | 10 | 1 | 4.103919087 | 0 | 0 | 0 | MH590589,MH651172,MW365952,MZ820095,ON456343 |
| 83 | VC_1072_3 | 6 | 4500 | 30019.34189 | 0.130361697 | 6.48E-08 | 3.605551275 | 9.110433579 | 15 | 13 | 0.866666667 | 6.980707437 | 0 | 0 | 0 | MK061412,MT114162,MW435853,MZ820093,MZ820094,ON208830 |
| 84 | VC_1072_4 | 4 | 1800 | 22659.45077 | 0.073591186 | 0.000434408 | 3.741657387 | 7.937253933 | 6 | 6 | 1 | 6.887275578 | 0 | 0 | 0 | MH536811,MH536814,MN369754,MT952852 |
| 85 | VC_1072_5 | 3 | 900 | 18056.33942 | 0.047477521 | 0.010026794 | 6.92820323 | 8.94427191 | 3 | 3 | 1 | 8.234745336 | 0 | 0 | 0 | MT684598,MW822145,ON645339 |
| 86 | VC_1073_0 | 24 | 56348.25696 | 10828.27477 | 0.838808666 | 3.17E-125 | 1.414213562 | 7.141428429 | 276 | 276 | 1 | 4.357263141 | 0 | 0 | 0 | KT365401,KU160651,KU160656,KU160665,KU160668,KU647629,MF189176,MF189177,MF189178,MF324907,MF377441,MF377442,MH825703,MK919479,MN183279,MN183283,MT024868,MT114161,MT310895,MT776811,MT889388,MW862982,OK999980,ON970601 |
| 87 | VC_1074_0 | 19 | 51212.62536 | 4462.845979 | 0.919841793 | 1.25E-88 | 0 | 5.477225575 | 171 | 171 | 1 | 3.069561112 | 0 | 0 | 0 | KM879463,KU160660,KX522650,KX670786,KX670787,KX855961,KX855962,MF185718,MF185721,MF185723,MF185724,MF185725,MF668274,MH744419,MK494107,MN204499,MN204503,MN234234,MN703411 |
| 88 | VC_1075_0 | 4 | 1166.060237 | 6694.97929 | 0.148334102 | 1.57E-05 | 3.872983346 | 6.633249581 | 6 | 6 | 1 | 5.169198155 | 0 | 0 | 0 | MT498055,MW601223,MZ747522,OL742560 |
| 89 | VC_1076_0 | 2 | 37.25022147 | 70.38665183 | 0.346073054 | 0.361836805 | 7.874007874 | 7.874007874 | 1 | 1 | 1 | 7.874007874 | 0 | 0 | 0 | S12-wms-combined_376326,S8-wms-combined_72911 |
| 90 | VC_1077_0 | 48 | 6785.315608 | 1174.292656 | 0.852468536 | 1.82E-44 | 0 | 2.236067977 | 1128 | 1128 | 1 | 1.407328578 | 0 | 0 | 0 | KP087938,KP087942,MG883726,MG945129,MG945206,MG945670,MG945836,MG945840,MG945846,MH572362,MH572410,MH572411,MH572412,MH572486,MH572521,MH931004,MH992163,MH992164,MH992192,MH992199,MH992202,MH992206,MH992208,MH992214,MH992215,MH992216,MH992217,MH992219,MH992220,MH992222,MK765645,MN862346,MW202445,MW202609,MW202760,MW202819,MZ089752,MZ364224,MZ364225,MZ364227,MZ364277,MZ364279,MZ364283,MZ364285,MZ364286,MZ364291,OL617030,OL617031 |
| 91 | VC_1079_0 | 6 | 136.6462375 | 10.27097468 | 0.930090052 | 0.109034215 | 0 | 2 | 15 | 15 | 1 | 1.013076828 | 0 | 0 | 0 | MH572360,MH572361,MH572409,MH572457,MH572482,MH572483 |
| 92 | VC_107_0 | 5 | 1502.822416 | 1825.786854 | 0.45148658 | 5.28E-08 | 4 | 6.92820323 | 10 | 10 | 1 | 6.061721007 | 0 | 0 | 0 | OP341624,OP341626,OP341627,OP341628,OP341630 |
| 93 | VC_1081_0 | 1 | 0 | 13.04261974 | 0 | 0 | 0 | 0 | 1 | 1 | 1 | 0 | 0 | 0 | 0 | MN379632 |
| 94 | VC_1082_0 | 3 | 24.02393131 | 10.66891169 | 0.69247514 | 0.380453364 | 0 | 1.732050808 | 3 | 3 | 1 | 1.154700538 | 0 | 0 | 0 | MH572366,MW202464,MW202671 |
| 95 | VC_1084_0 | 9 | 557.0840498 | 292.3370352 | 0.655839677 | 1.47E-17 | 0 | 1.414213562 | 36 | 36 | 1 | 0.791257816 | 0 | 0 | 0 | MH572287,MH572288,MH572467,MH572468,MH572496,MH572497,MH572498,MH572499,MH572500 |
| 96 | VC_1085_0 | 2 | 25.35975681 | 17.56982715 | 0.590729154 | 0.066807201 | 0 | 0 | 1 | 1 | 1 | 0 | 0 | 0 | 0 | KT264812,MG945475 |
| 97 | VC_1086_0 | 3 | 14.03030183 | 2.639931621 | 0.841637993 | 0.185546685 | 1 | 1.732050808 | 3 | 3 | 1 | 1.382088123 | 0 | 0 | 0 | KP087943,KP087951,KP087957 |
| 98 | VC_1087_0 | 5 | 24.06238446 | 5.612535498 | 0.810866027 | 0.012404212 | 1 | 2.236067977 | 10 | 10 | 1 | 1.567486109 | 0 | 0 | 0 | KP087953,MH572371,MH572373,MH572380,MK249215 |
| 99 | VC_1088_0 | 3 | 16.40130604 | 4.790390798 | 0.773949635 | 0.172889293 | 0 | 1 | 3 | 3 | 1 | 0.666666667 | 0 | 0 | 0 | KP087955,MH572364,MH572458 |
| 100 | VC_1089_0 | 2 | 3.175008833 | 2.639931621 | 0.54600883 | 0.5 | 1 | 1 | 1 | 1 | 1 | 1 | 0 | 0 | 0 | MH572489,MW202618 |
| 101 | VC_108_0 | 33 | 124113.4748 | 6664.107101 | 0.949042435 | 3.43E-84 | 0 | 7.937253933 | 528 | 528 | 1 | 6.145397794 | 0 | 0 | 0 | AB712291,HE962497,JF731128,KF192053,KJ094032,KJ127303,KT932699,KY303907,KY472224,LC547238,MH618488,MH791415,MK570225,MK721189,MK721194,MK800154,MN103542,MN871443,MN939408,MW677132,MZ333457,OL505085,OL870612,ON086985,ON113171,ON113172,ON113173,ON113174,ON113175,ON113177,ON113178,ON113179,ON113180 |
| 102 | VC_1090_0 | 4 | 183.6191782 | 0 | 1 | 0 | 7.348469228 | 8.602325267 | 6 | 6 | 1 | 7.978251757 | 0 | 0 | 0 | KP202971,KT001914,MF098558,ON323491 |
| 103 | VC_1091_0 | 2 | 150.9423405 | 0 | 1 | 0 | 3.741657387 | 3.741657387 | 1 | 1 | 1 | 3.741657387 | 0 | 0 | 0 | KP836355,KP836356 |
| 104 | VC_1092_0 | 12 | 11676.69435 | 1452.418658 | 0.889374198 | 8.74E-33 | 0 | 5.291502622 | 66 | 66 | 1 | 3.716604004 | 0 | 0 | 0 | MW314851,MW314853,MW314857,MW584149,MW584151,MW584184,MW584193,MW584194,MW584198,MW584200,MW584205,MW584210 |
| 105 | VC_1093_0 | 40 | 165629.108 | 23580.81472 | 0.875372209 | 2.54E-278 | 1 | 7.615773106 | 780 | 780 | 1 | 5.728244738 | 0 | 0 | 0 | KU963250,MF919514,MG812487,MH153809,MH450129,MH479924,MH651166,MH651168,MH976506,MH976507,MH976508,MH976509,MH976510,MH976511,MH976512,MH976513,MH976514,MH976518,MH976519,MK937594,MK937601,MK967396,MK977702,MN329673,MN329677,MT498035,MT639640,MT723934,MT723937,MT723941,MW822146,MW924645,MZ322015,MZ681510,ON081332,ON456336,ON637761,ON724014,OP172877,OP434456 |
| 106 | VC_1095_0 | 16 | 23546.79785 | 27866.13825 | 0.457993642 | 1.40E-70 | 0 | 8.831760866 | 120 | 120 | 1 | 6.188558907 | 0 | 0 | 0 | KU963253,MH669000,MK937607,MK937611,MN586026,MN586028,MN586047,MT498036,MW314847,MZ322009,MZ322020,ON392159,ON456337,ON456349,ON755186,OP068342 |
| 107 | VC_1096_0 | 5 | 2079.239624 | 348.1711688 | 0.856566853 | 0.001300234 | 2.645751311 | 8.246211251 | 10 | 10 | 1 | 6.159672454 | 0 | 0 | 0 | KR063280,MK279853,MN586022,MZ820089,ON755184 |
| 108 | VC_1098_0 | 4 | 1800 | 1493.830995 | 0.546476125 | 3.54E-05 | 1 | 7.810249676 | 6 | 6 | 1 | 4.672372311 | 0 | 0 | 0 | MK814760,MK894436,MN484601,OP297539 |
| 109 | VC_1098_1 | 1 | 0 | 737.4900434 | 0 | 0 | 0 | 0 | 1 | 1 | 1 | 0 | 0 | 0 | 0 | KR063281 |
| 110 | VC_1099_0 | 2 | 300 | 848.430591 | 0.261226061 | 0.053268464 | 2.449489743 | 2.449489743 | 1 | 1 | 1 | 2.449489743 | 0 | 0 | 0 | MN234206,MN234211 |
| 111 | VC_1099_1 | 1 | 0 | 670.1181876 | 0 | 0 | 0 | 0 | 1 | 1 | 1 | 0 | 0 | 0 | 0 | OK040794 |
| 112 | VC_109_0 | 3 | 847.6412643 | 4121.825521 | 0.170569862 | 0.001643767 | 0 | 3.16227766 | 3 | 3 | 1 | 2.108185107 | 0 | 0 | 0 | MZ398135,MZ398136,MZ399596 |
| 113 | VC_10_0 | 25 | 32380.36153 | 61024.34232 | 0.346667354 | 5.46E-178 | 0 | 9.219544457 | 300 | 292 | 0.973333333 | 7.391403197 | 0 | 0 | 0 | AP011955,AP011956,AY508486,CP062430,CP062436,CP062447,DQ530361,GQ398772,KC595278,KF831354,KJ452292,KJ596420,KT809368,KT809369,KT878766,KY653123,MG029510,MG029512,MH384259,MH401416,MW924889,OP038320,OP136135,OP328423,OP493558 |
| 114 | VC_1100_0 | 2 | 300 | 2034.126921 | 0.128527715 | 0.053268464 | 3.464101615 | 3.464101615 | 1 | 1 | 1 | 3.464101615 | 0 | 0 | 0 | MN908685,MZ150783 |
| 115 | VC_1100_1 | 2 | 300 | 2152.360589 | 0.122331113 | 0.053268464 | 6.08276253 | 6.08276253 | 1 | 1 | 1 | 6.08276253 | 0 | 0 | 0 | OK040793,ON637770 |
| 116 | VC_1100_2 | 1 | 0 | 953.1209951 | 0 | 0 | 0 | 0 | 1 | 1 | 1 | 0 | 0 | 0 | 0 | OK040790 |
| 117 | VC_1100_3 | 1 | 0 | 1262.046244 | 0 | 0 | 0 | 0 | 1 | 1 | 1 | 0 | 0 | 0 | 0 | OP068343 |
| 118 | VC_1101_0 | 2 | 255.3834155 | 5.573705406 | 0.978641298 | 0.270145687 | 2.449489743 | 2.449489743 | 1 | 1 | 1 | 2.449489743 | 0 | 0 | 0 | KR136259,KR136260 |
| 119 | VC_1103_0 | 2 | 14.75294763 | 0 | 1 | 0 | 0 | 0 | 1 | 1 | 1 | 0 | 0 | 0 | 0 | KR704913,KR704915 |
| 120 | VC_1104_0 | 154 | 131803.624 | 23550.04811 | 0.848410097 | 0 | 0 | 3.464101615 | 11781 | 11781 | 1 | 1.808109624 | 0 | 0 | 0 | KT264749,KT264750,KT264751,KT264752,KT264753,KT264754,KT264755,KT264756,KT264757,KT264758,KT264759,KT264760,KT264761,KT264762,KT264763,KT264764,KT264765,KT264766,KT264767,KT264768,KT264769,KT264779,KX513865,MG025518,MG641179,MG641180,MG641186,MG883717,MG945153,MG945158,MG945163,MG945190,MG945203,MG945205,MG945207,MG945213,MG945227,MG945237,MG945238,MG945318,MG945325,MG945330,MG945333,MG945346,MG945351,MG945355,MG945361,MG945363,MG945387,MG945393,MG945398,MG945402,MG945407,MG945415,MG945424,MG945429,MG945435,MG945438,MG945444,MG945459,MG945461,MG945467,MG945476,MG945486,MG945499,MG945508,MG945517,MG945526,MG945541,MG945548,MG945559,MG945565,MG945566,MG945570,MG945586,MG945587,MG945589,MG945596,MG945602,MG945646,MG945671,MG945673,MG945674,MG945681,MG945691,MG945699,MG945705,MG945708,MG945710,MG945728,MG945731,MG945744,MG945758,MG945761,MG945765,MG945768,MG945770,MG945786,MG945787,MG945793,MG945796,MG945801,MG945802,MG945820,MG945853,MG945855,MG945862,MG945864,MG945869,MK765620,MT135257,MT135263,MT135264,MT135272,MT135273,MT135278,MT135338,MZ089765,MZ089767,MZ089774,MZ089775,OM869524,OM869525,OM869526,OM869527,OM869528,OM869531,OM869533,OM869534,OM869535,OM869537,OM869538,OM869543,OM869546,OM869547,OM869550,OM869606,OM869635,OM869641,OM869642,OM869643,OM869644,OM869645,OM869664,OM869697,OM869698,OM869700,OM869701,OM869703,OM869704,OP549869,OP549872,OP549875,OP549876 |
| 121 | VC_1105_0 | 3 | 19.55315005 | 239.4936747 | 0.075481142 | 0.005331249 | 1.732050808 | 2 | 3 | 3 | 1 | 1.821367205 | 0 | 0 | 0 | MG945597,MK496743,MK496744 |
| 122 | VC_1108_0 | 4 | 98.49456099 | 1932.484699 | 0.048496094 | 9.79E-06 | 0 | 2.236067977 | 6 | 6 | 1 | 1.558408522 | 0 | 0 | 0 | KT264775,MG883718,MG945386,MG945775 |
| 123 | VC_1109_0 | 2 | 11.84558033 | 6.52357444 | 0.644862569 | 0.144422183 | 7.071067812 | 7.071067812 | 1 | 1 | 1 | 7.071067812 | 0 | 0 | 0 | KR816341,S7-wms-combined_235165 |
| 124 | VC_110_0 | 4 | 786.3198875 | 6093.327532 | 0.114296539 | 1.27E-05 | 0 | 6 | 6 | 6 | 1 | 4.362373124 | 0 | 0 | 0 | FR687252,MZ333135,OM513679,OM513680 |
| 125 | VC_1110_0 | 8 | 356.5398977 | 159.0330362 | 0.691541146 | 1.09E-10 | 0 | 2.449489743 | 28 | 28 | 1 | 1.787604653 | 0 | 0 | 0 | MG945181,MG945211,MG945232,MG945718,MG945783,MT135261,MT135334,MZ089771 |
| 126 | VC_1111_0 | 2 | 15.53109722 | 43.42024723 | 0.2634562 | 0.101520076 | 1 | 1 | 1 | 1 | 1 | 1 | 0 | 0 | 0 | MG945489,MT135282 |
| 127 | VC_1112_0 | 2 | 8.282189768 | 29.15743477 | 0.221214552 | 0.276226452 | 1.732050808 | 1.732050808 | 1 | 1 | 1 | 1.732050808 | 0 | 0 | 0 | MK496719,MK496738 |
| 128 | VC_1113_0 | 1 | 0 | 116.1832688 | 0 | 0 | 0 | 0 | 1 | 1 | 1 | 0 | 0 | 0 | 0 | MG945591 |
| 129 | VC_1114_0 | 2 | 8.583214966 | 155.8955653 | 0.05218433 | 0.05239548 | 1.414213562 | 1.414213562 | 1 | 1 | 1 | 1.414213562 | 0 | 0 | 0 | MG945131,MG945742 |
| 130 | VC_1116_0 | 14 | 989.6359613 | 258.7226044 | 0.792749766 | 5.26E-26 | 0 | 2.236067977 | 91 | 91 | 1 | 1.539755979 | 0 | 0 | 0 | KT264776,MG883719,MG945139,MG945339,MG945394,MG945403,MG945542,MG945550,MG945645,MG945736,MG945740,MG945771,MG945819,OP209174 |
| 131 | VC_1117_0 | 5 | 172.0997629 | 194.9832313 | 0.468830661 | 2.61E-07 | 0 | 1 | 10 | 10 | 1 | 0.6 | 0 | 0 | 0 | KT264781,MG945481,MG945807,MT135256,MT135266 |
| 132 | VC_1119_0 | 20 | 1151.152751 | 553.9510847 | 0.675121788 | 2.11E-05 | 0 | 2.449489743 | 190 | 190 | 1 | 1.492478073 | 0 | 0 | 0 | KT264782,KT264783,KX513866,KX513870,MG945154,MG945472,MG945534,MG945613,MG945826,MK496754,MK496805,MK496812,MK496813,MK765559,MK765603,MK765621,MK765669,MZ089756,MZ089773,OM869503 |
| 133 | VC_111_0 | 91 | 460400.0387 | 19850.42961 | 0.958666507 | 0 | 0 | 7.615773106 | 4095 | 4095 | 1 | 6.033690095 | 0 | 0 | 0 | AB716666,GQ413938,KF765493,KM576124,KP708985,KP708986,KT367886,KT964103,KU666550,KX211991,KX237514,KX587949,KX856662,KY000081,KY385423,KY684082,MF612071,MG972768,MH885472,MK135468,MK562503,MK598851,MK693005,MN101217,MN101219,MN101220,MN116494,MN176572,MN176573,MN380459,MN794000,MN794001,MN794002,MN794003,MT090077,MT197175,MT197176,MT360680,MT559526,MT559527,MT682065,MT701589,MT939252,MW590329,MZ208805,MZ424864,MZ571827,MZ602146,MZ634338,MZ634339,MZ634340,MZ634342,MZ634343,OK562429,OK583892,OK583893,OL362278,OL362279,OL474125,OL976437,OL979478,OL979479,OM864357,ON132389,ON146449,ON165415,ON602723,ON602724,ON602725,ON602726,ON602727,ON602728,ON602732,ON602739,ON602740,ON602743,ON602748,ON602752,ON602754,ON602756,ON602757,ON602761,ON602762,ON602767,ON809559,OP114731,OP413829,OU509533,OU509534,OX335379,OX335428 |

| | | | | | | | | | | | | | | | |
|---|---|---|---|---|---|---|---|---|---|---|---|---|---|---|---|
| 134 | VC_1120_0 | 7 | 139.7627318 | 380.2096518 | 0.268788759 | 0.004813204 | 1 | 2 | 21 | 21 | 1 | 1.558634374 | 0 | 0 | 0 MG945704,MZ089755,MZ089759,MZ089761,MZ089764,OM869627,OM869629 |
| 135 | VC_1122_0 | 10 | 738.4083418 | 27.33738998 | 0.964299651 | 3.03E-06 | 0 | 2.449489743 | 45 | 45 | 1 | 1.837559165 | 0 | 0 | 0 KT264785,KX513873,MG945186,MG945569,MG945689,MG945797,MG945870,MN379630,MN379640,MT135351 |
| 136 | VC_1123_0 | 2 | 25.35975681 | 54.22329692 | 0.31865775 | 0.067297332 | 0 | 0 | 1 | 1 | 1 | 0 | 0 | 0 | 0 KT264784,KX513864 |
| 137 | VC_1124_0 | 10 | 775.5435605 | 461.7117706 | 0.626825798 | 1.01E-21 | 0 | 2.828427125 | 45 | 45 | 1 | 2.013385987 | 0 | 0 | 0 KT264798,KT264799,KT264800,KT264801,MG945135,MG945136,MG945157,MG945391,MG945867,MT135258 |
| 138 | VC_1125_0 | 20 | 1879.265887 | 1477.037791 | 0.559921291 | 2.38E-35 | 0 | 2.645751311 | 190 | 190 | 1 | 1.776025311 | 0 | 0 | 0 KT264796,KT264797,MG883722,MG945286,MG945319,MG945338,MG945347,MG945385,MG945389,MG945410,MG945416,MG945437,MG945538,MG945635,MG945639,MG945746,MG945767,MG945858,OM869558,OM869663 |
| 139 | VC_1126_0 | 8 | 393.5066845 | 980.3011821 | 0.286435021 | 1.56E-12 | 0 | 2.828427125 | 28 | 28 | 1 | 1.829974799 | 0 | 0 | 0 KT264788,KT264789,KT264790,MG945138,MG945381,MG945382,MG945780,MT135259 |
| 140 | VC_1127_0 | 19 | 3585.755056 | 1125.35752 | 0.761127016 | 1.30E-64 | 0 | 2.645751311 | 171 | 171 | 1 | 1.591464515 | 0 | 0 | 0 KT264791,KT264792,KT264793,KX513868,KX513877,MG945162,MG945233,MG945334,MG945383,MG945388,MG945430,MG945447,MG945514,MG945642,MG945694,MG945753,MG945781,MT135277,MZ821031 |
| 141 | VC_1128_0 | 2 | 13.78291419 | 138.669696 | 0.09040786 | 0.048566868 | 1.414213562 | 1.414213562 | 1 | 1 | 1 | 1.414213562 | 0 | 0 | 0 KT264794,MG945231 |
| 142 | VC_112_0 | 8 | 3390.940014 | 9975.342762 | 0.253693571 | 1.16E-19 | 3.31662479 | 6.480740698 | 28 | 28 | 1 | 4.900898397 | 0 | 0 | 0 KF319020,KM819694,MG575418,MG575419,MK387869,MW358930,OL416096,OM234793 |
| 143 | VC_1132_0 | 2 | 4.09143279 | 37.74211098 | 0.097802682 | 0.168052265 | 2 | 2 | 1 | 1 | 1 | 2 | 0 | 0 | 0 KT264795,MG945738 |
| 144 | VC_1133_0 | 4 | 38.92455113 | 271.6296645 | 0.125338988 | 0.024932995 | 1.414213562 | 2.449489743 | 6 | 6 | 1 | 2.002227107 | 0 | 0 | 0 MG641184,MZ089778,OM869517,OM869640 |
| 145 | VC_1134_0 | 5 | 53.56080877 | 361.780039 | 0.128956275 | 0.236291101 | 0 | 2.449489743 | 10 | 10 | 1 | 1.709513191 | 0 | 0 | 0 MK496714,MK496779,MK496792,MZ089777,OM869545 |
| 146 | VC_1136_0 | 2 | 26.62182152 | 19.27661572 | 0.580015859 | 0.117381831 | 2 | 2 | 1 | 1 | 1 | 2 | 0 | 0 | 0 KT264803,MG945142 |
| 147 | VC_1137_0 | 3 | 63.77440347 | 5.72793687 | 0.917586417 | 0.069320317 | 1.414213562 | 1.732050808 | 3 | 3 | 1 | 1.626105059 | 0 | 0 | 0 KT264802,MG945620,MG945741 |
| 148 | VC_1138_0 | 4 | 137.7798897 | 8.212978528 | 0.943743974 | 0.010350137 | 1.414213562 | 2 | 6 | 6 | 1 | 1.670763258 | 0 | 0 | 0 KT264805,MG945519,MG945693,MG945809 |
| 149 | VC_1139_0 | 4 | 115.3245102 | 0 | 1 | 0 | 0 | 2.236067977 | 6 | 6 | 1 | 1.578689326 | 0 | 0 | 0 KT264806,MG945320,MG945561,MG945721 |
| 150 | VC_113_0 | 9 | 3153.164405 | 2922.381764 | 0.518992748 | 1.08E-22 | 2 | 8.717797887 | 36 | 36 | 1 | 6.87188797 | 0 | 0 | 0 AB720063,AP008979,AY299121,DQ777876,EU717894,MW934263,MZ701794,OK258140,OM913894 |
| 151 | VC_1140_0 | 4 | 42.35797099 | 277.3736397 | 0.132479772 | 0.337928011 | 1.414213562 | 2.645751311 | 6 | 6 | 1 | 2.22279108 | 0 | 0 | 0 KT264820,KT264821,MG945644,MK765602 |
| 152 | VC_1143_0 | 9 | 866.3168129 | 325.6739771 | 0.726781465 | 6.92E-19 | 0 | 1.414213562 | 36 | 36 | 1 | 0.712690396 | 0 | 0 | 0 KT264818,KT264819,KX513867,KX513872,KX513874,MG945573,MG945759,MG945760,MG945811 |
| 153 | VC_1144_0 | 7 | 534.3856341 | 220.8613688 | 0.707564058 | 7.01E-13 | 0 | 1.414213562 | 21 | 21 | 1 | 0.824462309 | 0 | 0 | 0 MG883714,MG945364,MG945525,MG945719,MG945805,MT135337,MT135424 |
| 154 | VC_1145_0 | 5 | 159.9798797 | 21.66125994 | 0.88074695 | 0.000789379 | 0 | 0 | 10 | 10 | 1 | 0 | 0 | 0 | 0 OM869513,OM869516,OM869519,OM869634,OM869637 |
| 155 | VC_1146_0 | 3 | 62.6039311 | 109.3607466 | 0.364051106 | 0.001329228 | 0 | 0 | 3 | 3 | 1 | 0 | 0 | 0 | 0 MG945234,MG945405,MG945827 |
| 156 | VC_1147_0 | 3 | 12.51733192 | 16.36573266 | 0.433379633 | 0.346640909 | 1.414213562 | 1.732050808 | 3 | 3 | 1 | 1.626105059 | 0 | 0 | 0 KT264822,KT264830,MK496745 |
| 157 | VC_1148_0 | 16 | 1653.669296 | 221.092342 | 0.882069092 | 1.68E-24 | 0 | 2 | 120 | 120 | 1 | 1.099336914 | 0 | 0 | 0 KT264823,KT264824,KT264825,MG945179,MG945357,MG945413,MG945506,MG945507,MG945609,MG945676,MG945706,MG945764,MG945817,MT135267,MT135271,MT135281 |
| 158 | VC_1149_0 | 5 | 194.4233581 | 213.3694372 | 0.476769968 | 1.96E-07 | 0 | 2 | 10 | 10 | 1 | 1.219615242 | 0 | 0 | 0 MG945441,MG945487,MG945505,MT135255,MT135275 |
| 159 | VC_1151_0 | 9 | 588.4458773 | 99.9913509 | 0.854756038 | 1.41E-11 | 0 | 1.414213562 | 36 | 36 | 1 | 0.909108946 | 0 | 0 | 0 KT264828,MG945228,MG945426,MG945726,MG945754,MG945816,OP549870,OP549873,OP549877 |
| 160 | VC_1152_0 | 4 | 125.2078622 | 48.5485187 | 0.720594326 | 0.000288354 | 0 | 0 | 6 | 6 | 1 | 0 | 0 | 0 | 0 MG945204,MG945727,MG945785,MG945810 |
| 161 | VC_1153_0 | 2 | 3.644286755 | 46.24911685 | 0.073041454 | 0.194373584 | 2.236067977 | 2.236067977 | 1 | 1 | 1 | 2.236067977 | 0 | 0 | 0 KT264831,MN379641 |
| 162 | VC_1154_0 | 5 | 74.23000791 | 0 | 1 | 0 | 0 | 1.414213562 | 10 | 10 | 1 | 0.848528137 | 0 | 0 | 0 KT264833,MG945418,MG945549,MG945747,OM869677 |
| 163 | VC_1155_0 | 2 | 8.951186952 | 0 | 1 | 0 | 1.414213562 | 1.414213562 | 1 | 1 | 1 | 1.414213562 | 0 | 0 | 0 KT264834,MG945678 |
| 164 | VC_1156_0 | 8 | 6486.058456 | 1485.393041 | 0.813660907 | 1.12E-14 | 0 | 6.557438524 | 28 | 28 | 1 | 4.718654796 | 0 | 0 | 0 KT755656,MH431938,MH454076,MH454080,MH460824,MH460825,MH460826,MH460827 |
| 165 | VC_1157_0 | 24 | 79745.203 | 0 | 1 | 0 | 0 | 5 | 276 | 276 | 1 | 2.885243546 | 0 | 0 | 0 KT876724,KT876726,KU599877,KU599878,KU599879,KU599886,MK764441,MK764449,MW421577,MW421578,MW421579,MW421580,MW442825,MW442826,MW442827,MW442828,MW442829,MW442830,MW448539,MW448540,MW448541,OK340820,OK340821,OK340822 |
| 166 | VC_1158_0 | 3 | 900 | 1930.902276 | 0.317919841 | 0.006644424 | 0 | 1.732050808 | 3 | 3 | 1 | 1.154700538 | 0 | 0 | 0 MH576962,MK620894,MW291021 |
| 167 | VC_1158_1 | 2 | 300 | 1885.967613 | 0.137238996 | 0.101796862 | 3.464101615 | 3.464101615 | 1 | 1 | 1 | 3.464101615 | 0 | 0 | 0 MH744418,MT310898 |
| 168 | VC_1159_0 | 3 | 75.95706824 | 153.9104374 | 0.330438476 | 0.01451641 | 6.480740698 | 7.615773106 | 3 | 3 | 1 | 7.193276193 | 0 | 0 | 0 MF668275,MZ955870,OV696619 |
| 169 | VC_115_0 | 2 | 60.73640616 | 86.78846027 | 0.411702838 | 0.092556008 | 5.196152423 | 5.196152423 | 1 | 1 | 1 | 5.196152423 | 0 | 0 | 0 MG592523,OX241425 |
| 170 | VC_1160_0 | 73 | 350016.3117 | 5854.845117 | 0.983547852 | 0 | 0 | 8.366600265 | 2628 | 2628 | 1 | 5.997615649 | 0 | 0 | 0 KU160640,KU160643,KU160645,KU160648,KU160649,KU160652,KU160653,KU160659,KU160661,KU160662,KU160671,KU160672,KX576640,KX576641,KX621004,KX621005,KX688102,KX688103,MF140400,MF140404,MF140407,MF140424,MF140425,MF668285,MG198779,MG198781,MG198782,MG210949,MH450117,MH450118,MH450121,MH450131,MH744421,MK112526,MK112528,MK112529,MK112531,MK112535,MK112538,MK279844,MK279845,MK279847,MK279851,MK279854,MK279856,MK279860,MK279862,MK279863,MK279867,MK279868,MK279869,MK279872,MK279874,MK279875,MK279876,MK279880,MK279884,MK279886,MK279892,MK279896,MK279901,MK279907,MK660714,MK814749,MK919476,MK919484,MN586061,MN735429,MN813680,MT498045,MT498050,MW314848,OP434444 |
| 171 | VC_1161_0 | 2 | 96.52230796 | 3163.60741 | 0.029606892 | 0.042628099 | 5.656854249 | 5.656854249 | 1 | 1 | 1 | 5.656854249 | 0 | 0 | 0 KX557278,MT771339 |
| 172 | VC_1162_0 | 10 | 8202.982884 | 6149.279255 | 0.571546339 | 7.39E-30 | 0 | 5.656854249 | 45 | 45 | 1 | 3.238904209 | 0 | 0 | 0 MK016495,MK016497,MK737942,MN369765,MT310855,MT498057,MZ150782,MZ747515,ON260821,ON970583 |
| 173 | VC_1163_0 | 28 | 45361.45591 | 46782.97634 | 0.492286455 | 1.05E-220 | 0 | 8.306623863 | 378 | 378 | 1 | 6.119889509 | 0 | 0 | 0 MG839027,MG925343,MG925345,MG925347,MG944216,MG944218,MG944226,MH513982,MH590604,MH590606,MK308637,MK620901,MK620904,MK814762,MK894432,MK937602,MN096367,MN096378,MN284901,MN310543,MT553340,MT657343,MT657344,MZ005670,OK318961,ON526972,ON970571,OP068339 |
| 174 | VC_1164_0 | 25 | 70031.14352 | 25741.19561 | 0.731225155 | 2.60E-139 | 0 | 7.141428429 | 300 | 300 | 1 | 5.342705459 | 0 | 0 | 0 KU160641,KU160646,MF038790,MF038791,MF140401,MF140423,MF140432,MH399789,MH450115,MH450125,MH479926,MH825711,MN234168,MN234194,MN234221,MN284902,MN703412,MW055912,MZ209303,MZ274309,OK310503,ON456335,ON456340,ON645338,OP434462 |
| 175 | VC_1166_0 | 20 | 30763.22744 | 311.9133725 | 0.989962608 | 4.71E-40 | 1.732050808 | 7.549834435 | 190 | 190 | 1 | 5.406810937 | 0 | 0 | 0 KU160644,MF140397,MH834595,MH834606,MH834608,MH834609,MH834616,MH834618,MH834620,MH834624,MN234214,MN484600,MN703409,MT889373,MT952853,MW507131,ON260822,ON970604,OP297550,OP297552 |
| 176 | VC_1167_0 | 6 | 2474.88811 | 388.9488032 | 0.86418612 | 5.45E-07 | 0 | 4 | 15 | 15 | 1 | 2.59761825 | 0 | 0 | 0 KU160650,MF668266,MF668273,MF668279,MH450114,ON337195 |
| 177 | VC_1168_0 | 2 | 39.06987812 | 355.3833776 | 0.099048183 | 0.064498006 | 4.69041576 | 4.69041576 | 1 | 1 | 1 | 4.69041576 | 0 | 0 | 0 MW507126,MW590223 |
| 178 | VC_1169_0 | 7 | 6112.725839 | 118.016966 | 0.981058925 | 4.39E-05 | 2.236067977 | 4.472135955 | 21 | 21 | 1 | 3.654633973 | 0 | 0 | 0 KU160654,KU160664,MF140418,MF140427,MF140434,MG099950,MN585972 |
| 179 | VC_116_0 | 13 | 14237.49555 | 0 | 1 | 0 | 0 | 4.795831523 | 78 | 78 | 1 | 3.10338801 | 0 | 0 | 0 AB720064,LR743523,LR743528,LR743529,LR743531,MW802488,MW822535,MW822536,MW822537,MW822538,MW822539,OK913679,OK913680 |
| 180 | VC_1170_0 | 3 | 900 | 1847.977088 | 0.327513648 | 0.001657868 | 2.828427125 | 3.464101615 | 3 | 3 | 1 | 3.040318622 | 0 | 0 | 0 MN586021,MT889394,OP434452 |
| 181 | VC_1170_1 | 3 | 834.9295532 | 1876.061447 | 0.307979463 | 0.001635534 | 2.828427125 | 5 | 3 | 3 | 1 | 4.276142375 | 0 | 0 | 0 KU160669,KU160673,MW712735 |
| 182 | VC_1170_2 | 2 | 144.4804482 | 1734.323269 | 0.076900235 | 0.045810936 | 7.549834435 | 7.549834435 | 1 | 1 | 1 | 7.549834435 | 0 | 0 | 0 MW862992,OM818331 |
| 183 | VC_1171_0 | 21 | 58214.54746 | 23487.41835 | 0.712523216 | 4.44E-95 | 0 | 7.416198487 | 210 | 210 | 1 | 5.406578821 | 0 | 0 | 0 MH153803,MH153813,MN183284,MT114166,MT639641,MT657333,MW365946,MW365949,MW365950,MW712724,MW712727,MW712728,MW924638,MZ322011,MZ322018,OM203157,OP068331,OP068332,OP068336,OP068337,OP068344 |
| 184 | VC_1173_0 | 6 | 4500 | 22.25367723 | 0.995079074 | 7.04E-06 | 0 | 7.071067812 | 15 | 15 | 1 | 4.997587108 | 0 | 0 | 0 KU230356,MK605242,MK605243,MK605244,MK605245,MK605246 |
| 185 | VC_1174_0 | 4 | 1417.86243 | 0 | 1 | 0 | 3 | 4.582575695 | 6 | 6 | 1 | 3.979988005 | 0 | 0 | 0 KU234532,KU234533,MT426122,MZ078141 |
| 186 | VC_1175_0 | 22 | 41914.35759 | 49960.57373 | 0.456211036 | 1.02E-137 | 0 | 8.717797887 | 231 | 231 | 1 | 4.88055141 | 0 | 0 | 0 KU963246,KU963247,MG099941,MH271295,MK279911,MK279912,MK359313,MK801734,MK875797,MN586055,MT771342,MT818424,MW055908,MW291013,MW507130,MW822147,MZ150788,MZ171371,MZ681509,OM203160,ON645343,OP297544 |
| 187 | VC_1176_0 | 41 | 138099.1816 | 6392.659702 | 0.955757642 | 0 | 0 | 9.16515139 | 820 | 818 | 0.997560976 | 6.922079792 | 0 | 0 | 0 KU963251,KU963254,KU963259,KU998236,KU998248,KX557274,KX557275,MF919521,MH020241,MH153808,MH536818,MH576969,MK501729,MK501730,MK801721,MK801724,MK814757,MK864263,MK878896,MK919470,MK967389,MN010762,MN062704,MN096365,MN234161,MN234179,MN369740,MN585963,MN586031,MT553336,MT723935,MT723936,MT723942,MW291016,MW601225,MZ150785,MZ622164,ON724008,ON724011,ON724012,ON970567 |
| 188 | VC_1177_0 | 14 | 10743.127 | 2946.929446 | 0.784739423 | 3.55E-53 | 2.449489743 | 9.273618495 | 91 | 90 | 0.989010989 | 6.915847147 | 0 | 0 | 0 KU963252,KU998235,KX557279,MH479915,MK061410,MK284521,MK878901,MK937599,MK967377,MN813694,MT498059,MT522006,MT771341,MW055901 |
| 189 | VC_1178_0 | 24 | 67757.39262 | 3384.346427 | 0.952428118 | 1.18E-89 | 0 | 7.681145748 | 276 | 276 | 1 | 5.428561403 | 0 | 0 | 0 KU998234,KX557286,MH479910,MH479917,MH669015,MK305889,MK814761,MK864264,MK864265,MK864266,MK864267,MK937603,MK967381,MN010760,MN284907,MT310872,MT521998,MT639651,MW507125,MZ005680,MZ150791,MZ622177,ON970602,ON970619 |
| 190 | VC_1179_0 | 3 | 900 | 286.0342172 | 0.758831395 | 0.031801285 | 1.732050808 | 2.645751311 | 3 | 3 | 1 | 2.275763954 | 0 | 0 | 0 KU647626,KU647627,MN586053 |
| 191 | VC_117_0 | 9 | 2958.977649 | 6.703616968 | 0.997739603 | 0.010113628 | 1 | 6.32455532 | 36 | 36 | 1 | 4.715993154 | 0 | 0 | 0 AB757800,AB757801,JX880072,KX365747,MH059635,MK574011,MW202665,MW202744,OL959999 |
| 192 | VC_1180_0 | 3 | 528.6241114 | 495.6882675 | 0.516077051 | 0.001821403 | 2.449489743 | 4.69041576 | 3 | 3 | 1 | 3.713301834 | 0 | 0 | 0 MK814759,MZ209302,OM818327 |
| 193 | VC_1181_0 | 2 | 300 | 6201.586617 | 0.046142583 | 0.048735676 | 7.141428429 | 7.141428429 | 1 | 1 | 1 | 7.141428429 | 0 | 0 | 0 KU998245,MK524501 |
| 194 | VC_1181_1 | 10 | 13500 | 9664.244735 | 0.582794741 | 6.04E-22 | 1.732050808 | 7.810249676 | 45 | 45 | 1 | 4.747876313 | 0 | 0 | 0 KU998244,KU998246,KU998247,KX557276,MF919520,MF919537,MK376956,MK376959,MT498054,MZ388552 |
| 195 | VC_1182_0 | 4 | 1800 | 4526.359409 | 0.284523829 | 2.32E-05 | 6 | 8.306623863 | 6 | 6 | 1 | 7.574172865 | 0 | 0 | 0 KX557272,MK977705,MN908691,MT657339 |
| 196 | VC_1182_1 | 1 | 0 | 1900.171593 | 0 | 0 | 0 | 0 | 1 | 1 | 1 | 0 | 0 | 0 | 0 MH590603 |
| 197 | VC_1183_0 | 6 | 4384.377814 | 21.97870762 | 0.995012045 | 3.03E-05 | 0 | 6.633249581 | 15 | 15 | 1 | 3.64029784 | 0 | 0 | 0 MH479913,MH479925,MK919478,MN586014,MN813698,MW601220 |
| 198 | VC_1184_0 | 5 | 1488.123041 | 769.8319638 | 0.659057881 | 1.79E-07 | 3.872983346 | 4.795831523 | 10 | 10 | 1 | 4.371613925 | 0 | 0 | 0 KX557282,MK524505,MK814750,MW862986,ON724009 |
| 199 | VC_1186_0 | 4 | 847.3932277 | 1001.954357 | 0.458211985 | 8.06E-05 | 3.464101615 | 5.916079783 | 6 | 6 | 1 | 5.078476877 | 0 | 0 | 0 KX620750,KX620751,KX620753,KX620754 |
| 200 | VC_1187_0 | 3 | 66.0942646 | 5.435273925 | 0.924013575 | 0.074457337 | 3.741657387 | 4.898979486 | 3 | 3 | 1 | 4.213545624 | 0 | 0 | 0 KX077892,OL774866,OL774876 |
| 201 | VC_1188_0 | 3 | 550.5303161 | 61.10675933 | 0.900093108 | 0.005586199 | 0 | 1.414213562 | 3 | 3 | 1 | 0.942809042 | 0 | 0 | 0 MT344103,MT344104,MT344105 |
| 202 | VC_1189_0 | 9 | 7392.870039 | 244.540247 | 0.967981261 | 1.20E-11 | 2.236067977 | 6.244997998 | 36 | 36 | 1 | 4.696770336 | 0 | 0 | 0 KX455876,KY290947,KY290949,KY290950,KY290952,MH179470,MH179471,MH179479,MT276994 |
| 203 | VC_118_0 | 3 | 457.2346591 | 185.0008775 | 0.711942322 | 0.007885065 | 1 | 6.557438524 | 3 | 3 | 1 | 4.679393074 | 0 | 0 | 0 AB746912,LC545443,LC545444 |
| 204 | VC_1190_0 | 2 | 237.0894915 | 227.3824808 | 0.510449512 | 0.052354615 | 0 | 0 | 1 | 1 | 1 | 0 | 0 | 0 | 0 MG592484,MG592485 |
| 205 | VC_1191_0 | 3 | 23.42188031 | 142.9237229 | 0.140802521 | 0.010230257 | 1 | 2.236067977 | 3 | 3 | 1 | 1.745355992 | 0 | 0 | 0 KX513871,MG945160,MG945185 |
| 206 | VC_1192_0 | 3 | 28.52794827 | 41.57361211 | 0.406951687 | 0.005726732 | 1.414213562 | 1.732050808 | 3 | 3 | 1 | 1.626105059 | 0 | 0 | 0 MT135371,MT135422,MT135432 |
| 207 | VC_1193_0 | 3 | 61.07984183 | 50.52249268 | 0.547298962 | 0.004762206 | 1 | 1.414213562 | 3 | 3 | 1 | 1.138071187 | 0 | 0 | 0 MG945352,MG945735,MG945752 |
| 208 | VC_1194_0 | 5 | 153.4685656 | 139.6363789 | 0.523595963 | 1.01E-06 | 0 | 2 | 10 | 10 | 1 | 1.312095586 | 0 | 0 | 0 MG945210,MG945327,MG945623,MT135440,MT135446 |
| 209 | VC_1196_0 | 3 | 60.70502737 | 119.0249068 | 0.337756911 | 0.00256025 | 1 | 1.414213562 | 3 | 3 | 1 | 1.138071187 | 0 | 0 | 0 MG945748,MG945799,MG945800 |
| 210 | VC_1197_0 | 15 | 536.1783476 | 60.1606615 | 0.899116676 | 0.233916058 | 0 | 2 | 105 | 105 | 1 | 1.566607408 | 0 | 0 | 0 KX513875,MG883712,MG945284,MG945309,MG945433,MG945502,MG945616,MG945794,MK496693,MK496774,MZ089789,OM869564,OM869565,OM869570,OM869709 |
| 211 | VC_1198_0 | 3 | 40.5016314 | 104.4554578 | 0.279404282 | 0.048518588 | 6.32455532 | 7.483314774 | 3 | 3 | 1 | 7.052113107 | 0 | 0 | 0 MT006237,S5-wms_94965,S7-wms-combined_189250 |
| 212 | VC_1199_0 | 8 | 4339.436071 | 1133.97735 | 0.792820812 | 2.37E-17 | 0 | 6.708203932 | 28 | 28 | 1 | 4.790919079 | 0 | 0 | 0 MH727560,MH926061,MK937613,MK977706,MK977710,MK977712,MK977714,MK977715 |
| 213 | VC_119_0 | 2 | 219.3274309 | 178.2972606 | 0.551594093 | 0.081593387 | 0 | 0 | 1 | 1 | 1 | 0 | 0 | 0 | 0 LC545445,LC545446 |
| 214 | VC_11_0 | 19 | 25857.86752 | 8338.75262 | 0.756152725 | 1.59E-90 | 0 | 8.366600265 | 171 | 171 | 1 | 5.930331419 | 0 | 0 | 0 AP019560,AP019561,AP019562,KX827368,KX827369,KX827370,MF428478,MF428479,MF428480,MF428481,MK075003,MK075006,MN028536,MT423823,MT423824,OM373551,OM373552,OM373553,OP168678 |
| 215 | VC_1200_0 | 4 | 1251.179227 | 188.376379 | 0.869142687 | 8.04E-07 | 0 | 0 | 6 | 6 | 1 | 0 | 0 | 0 | 0 MF197383,MH271317,MN234233,OP172878 |
| 216 | VC_1201_0 | 2 | 60.63961284 | 7.353353373 | 0.891851264 | 0.185546685 | 2 | 2 | 1 | 1 | 1 | 2 | 0 | 0 | 0 KY817360,OL656105 |
| 217 | VC_1202_0 | 2 | 24.45666682 | 0 | 1 | 0 | 1 | 1 | 1 | 1 | 1 | 1 | 0 | 0 | 0 LC035386,LC210520 |
| 218 | VC_1203_0 | 7 | 6300 | 3370.525347 | 0.65146409 | 4.57E-11 | 1.732050808 | 7 | 21 | 21 | 1 | 4.510617844 | 0 | 0 | 0 MN988521,MN988522,MN988523,MN988524,MN988525,MN988559,MN988561 |
| 219 | VC_1203_1 | 1 | 0 | 2200.623707 | 0 | 0 | 0 | 0 | 1 | 1 | 1 | 0 | 0 | 0 | 0 MN988560 |
| 220 | VC_1204_0 | 2 | 300 | 39.73220869 | 0.883048449 | 0.080290302 | 1.414213562 | 1.414213562 | 1 | 1 | 1 | 1.414213562 | 0 | 0 | 0 LC644975,LC644976 |
| 221 | VC_1205_0 | 3 | 784.0286798 | 67.04974997 | 0.921217895 | 0.004981863 | 0 | 2.828427125 | 3 | 3 | 1 | 1.885618083 | 0 | 0 | 0 MT740726,MT740743,MT740745 |
| 222 | VC_1206_0 | 8 | 2728.653322 | 11532.40349 | 0.191335983 | 2.29E-18 | 1 | 8.888194417 | 28 | 28 | 1 | 7.514897582 | 0 | 0 | 0 MH271298,MH271308,MH271320,MK880124,MT952845,MW862981,OK040779,ON970568 |
| 223 | VC_1207_0 | 29 | 63820.65503 | 13768.46972 | 0.822546397 | 2.00E-116 | 0 | 8.366600265 | 406 | 406 | 1 | 5.719313717 | 0 | 0 | 0 MH834610,MH834619,MH834629,MK061413,MN703413,MN813676,MN908689,MT024869,MT024871,MT639653,MT889366,MT889375,MT889376,MT897906,MW055910,MZ005682,MZ681506,MZ747516,MZ820087,OK040777,OL455896,OL549188,OL549189,OL549191,OL549192,ON208833,ON528933,ON970616,OP434455 |
| 224 | VC_1208_0 | 11 | 16500 | 0 | 1 | 1 | 2 | 6.244997998 | 55 | 55 | 1 | 4.995261868 | 0 | 0 | 0 LR535901,LR535903,LR535904,LR535905,LR535906,LR535907,LR535908,LR535911,LR535912,LR535913,LR535917 |
| 225 | VC_1209_0 | 3 | 135.7682431 | 738.2387754 | 0.155339992 | 0.008479812 | 8 | 8.602325267 | 3 | 3 | 1 | 8.362535547 | 0 | 0 | 0 S1-wms-combined_41369,S1-wms_130214,S8-wms-combined_276211 |
| 226 | VC_1209_1 | 1 | 0 | 356.7893026 | 0 | 0 | 0 | 0 | 1 | 1 | 1 | 0 | 0 | 0 | 0 S12-wms-combined_23517 |
| 227 | VC_1209_2 | 1 | 0 | 387.2390441 | 0 | 0 | 0 | 0 | 1 | 1 | 1 | 0 | 0 | 0 | 0 S11-wms-combined_3507 |
| 228 | VC_1210_0 | 2 | 300 | 7881.824057 | 0.03666664 | 0.318790317 | 7.071067812 | 7.071067812 | 1 | 1 | 1 | 7.071067812 | 0 | 0 | 0 MK250021,MK250023 |
| 229 | VC_1210_1 | 7 | 6300 | 17097.63814 | 0.269257947 | 0.001258589 | 3 | 8.485281374 | 21 | 21 | 1 | 6.216553301 | 0 | 0 | 0 MK250020,MK250022,MK250024,MK250025,MK250026,MK250027,MK250028 |
| 230 | VC_1210_2 | 4 | 1800 | 13409.42458 | 0.118347673 | 0.079377892 | 1.414213562 | 5.099019514 | 6 | 6 | 1 | 4.043296893 | 0 | 0 | 0 MK250015,MK250016,MK250017,MK250018 |
| 231 | VC_1210_3 | 2 | 300 | 7901.575323 | 0.036578339 | 0.318790317 | 8.94427191 | 8.94427191 | 1 | 1 | 1 | 8.94427191 | 0 | 0 | 0 MK250019,MK250029 |
| 232 | VC_1211_0 | 25 | 3603.464777 | 0 | 1 | 0 | 0 | 2.236067977 | 300 | 300 | 1 | 1.576290331 | 0 | 0 | 0 LT986678,MG945141,MG945453,MG945455,MG945456,MG945498,MG945509,MG945564,MG945664,MG945680,MG945683,MG945713,MG945716,MG945730,MG945784,MG945790,MG945813,MG945845,MK765557,MK765655,MK765661,MK765667,MT135342,OM869561,OM869610 |
| 233 | VC_1212_0 | 2 | 16.30924847 | 0 | 1 | 0 | 0 | 0 | 1 | 1 | 1 | 0 | 0 | 0 | 0 MF101922,MH015251 |
| 234 | VC_1213_0 | 2 | 90.65925481 | 0 | 1 | 0 | 0 | 0 | 1 | 1 | 1 | 0 | 0 | 0 | 0 MF443783,MF443784 |
| 235 | VC_1215_0 | 3 | 451.5757549 | 0 | 1 | 0 | 0 | 2.236067977 | 3 | 3 | 1 | 1.490711985 | 0 | 0 | 0 MF580955,MF580957,MF580958 |
| 236 | VC_1216_0 | 3 | 733.9828396 | 168.3287969 | 0.813447162 | 0.007745576 | 0 | 1.732050808 | 3 | 3 | 1 | 1.154700538 | 0 | 0 | 0 MF580956,MF580959,MF580960 |
| 237 | VC_1217_0 | 3 | 670.0375996 | 168.3287969 | 0.799218101 | 0.007837249 | 0 | 2 | 3 | 3 | 1 | 1.333333333 | 0 | 0 | 0 MF580961,MF580962,MF629150 |
| 238 | VC_1218_0 | 4 | 304.1725247 | 37.41721771 | 0.89046153 | 0.004694091 | 0 | 1.732050808 | 6 | 6 | 1 | 0.866025404 | 0 | 0 | 0 MF979562,MK080312,MT074429,MT074440 |
| 239 | VC_1219_0 | 1 | 0 | 25.92395319 | 0 | 0 | 0 | 0 | 1 | 1 | 1 | 0 | 0 | 0 | 0 MG065691 |
| 240 | VC_1219_1 | 1 | 0 | 23.61272473 | 0 | 0 | 0 | 0 | 1 | 1 | 1 | 0 | 0 | 0 | 0 S5-wms-combined_66662 |
| 241 | VC_121_0 | 4 | 1008.728937 | 4553.009423 | 0.181369362 | 1.47E-05 | 0 | 5.744562647 | 6 | 6 | 1 | 4.713584711 | 0 | 0 | 0 JQ177061,JQ177064,KJ572844,MN379460 |
| 242 | VC_1221_0 | 1 | 0 | 275.4167061 | 0 | 0 | 0 | 0 | 1 | 1 | 1 | 0 | 0 | 0 | 0 MW660583 |
| 243 | VC_1221_1 | 1 | 0 | 316.0874499 | 0 | 0 | 0 | 0 | 1 | 1 | 1 | 0 | 0 | 0 | 0 S7-wms-combined_62119 |
| 244 | VC_1222_0 | 3 | 14.8095565 | 0 | 1 | 1 | 0 | 1 | 3 | 3 | 1 | 0.666666667 | 0 | 0 | 0 MG214764,MW202596,OL617037 |
| 245 | VC_1223_0 | 2 | 3.427695194 | 0 | 1 | 0 | 5 | 5 | 1 | 1 | 1 | 5 | 0 | 0 | 0 MG269973,MW202566 |
| 246 | VC_1224_0 | 19 | 29427.15172 | 0 | 1 | 0 | 0 | 6.557438524 | 171 | 171 | 1 | 4.078194401 | 0 | 0 | 0 MG515223,MG518519,MG518520,MG663582,MG663583,MG663584,MG663585,MG663586,MH155868,MK112550,MN119376,MN284894,MN994275,MT684596,MZ622160,OL455897,ON108647,ON970569,OP021678 |
| 247 | VC_1225_0 | 4 | 411.6172006 | 16.3108134 | 0.961884212 | 0.006251147 | 1 | 1.414213562 | 6 | 6 | 1 | 1.138071187 | 0 | 0 | 0 MG592394,MG592402,MG592427,MG592455 |
| 248 | VC_1226_0 | 2 | 138.4797217 | 74.67781679 | 0.64965904 | 0.131776239 | 0 | 0 | 1 | 1 | 1 | 0 | 0 | 0 | 0 MG592533,MG592560 |
| 249 | VC_1227_0 | 6 | 4247.323365 | 318.2488974 | 0.930293755 | 3.15E-07 | 0 | 3.741657387 | 15 | 15 | 1 | 2.405757917 | 0 | 0 | 0 MG592536,MG592554,MG592555,MG592556,MG592591,MG592625 |
| 250 | VC_1228_0 | 2 | 278.3645715 | 2.748292682 | 0.990223526 | 0.270145687 | 2.645751311 | 2.645751311 | 1 | 1 | 1 | 2.645751311 | 0 | 0 | 0 MK373772,OP136151 |
| 251 | VC_1229_0 | 7 | 4516.020158 | 34.42698443 | 0.992434373 | 1.33E-07 | 0 | 7.280109889 | 21 | 21 | 1 | 4.621331987 | 0 | 0 | 0 MG592596,MW824384,MW824385,MW824386,MW824395,MW824396,MW824397 |
| 252 | VC_122_0 | 5 | 1836.144968 | 7022.95615 | 0.207260866 | 4.49E-08 | 3.872983346 | 8.062257748 | 10 | 10 | 1 | 7.187067897 | 0 | 0 | 0 MG592400,MG592422,MG592425,MG592448,MG592486 |
| 253 | VC_122_1 | 1 | 0 | 665.783618 | 0 | 0 | 0 | 0 | 1 | 1 | 1 | 0 | 0 | 0 | 0 MG592434 |
| 254 | VC_1230_0 | 3 | 10.47501181 | 11.34108823 | 0.480150521 | 0.172889293 | 2 | 2 | 3 | 3 | 1 | 2 | 0 | 0 | 0 MG945523,MG945677,OM869552 |
| 255 | VC_1231_0 | 2 | 13.66481654 | 19.28092176 | 0.414767349 | 0.106686854 | 1.414213562 | 1.414213562 | 1 | 1 | 1 | 1.414213562 | 0 | 0 | 0 MZ089785,OM869555 |
| 256 | VC_1232_0 | 2 | 15.53109722 | 17.45384775 | 0.470854119 | 0.144422183 | 0 | 0 | 1 | 1 | 1 | 0 | 0 | 0 | 0 MG641185,MG945667 |
| 257 | VC_1233_0 | 16 | 1239.958404 | 142.2362437 | 0.897093912 | 8.30E-19 | 0 | 2.449489743 | 120 | 120 | 1 | 1.403640257 | 0 | 0 | 0 MG641188,MG945130,MG945256,MG945457,MG945557,MG945725,MG945757,MN379637,MN379647,MT135339,MT135348,MT135361,MT135384,MT135409,MT135435,OL617023 |
| 258 | VC_1234_0 | 44 | 149904.9295 | 0 | 1 | 0 | 0 | 8.602325267 | 946 | 946 | 1 | 5.801147403 | 0 | 0 | 0 MG670586,MH045566,MH045567,MH045568,MH509447,MH834596,MH834599,MH834602,MH834604,MH834626,MK814753,MK937598,MK937610,MN062714,MN062717,MN183281,MN329675,MT024860,MT310853,MT310896,MT639649,MT818427,MT889365,MT889372,MT889389,MT889392,MW601221,MW601224,MW712721,MW924640,MW924647,MZ322012,MZ747513,MZ820083,OK318958,OK999979,OK999981,OL742562,OM818329,ON724018,OP172875,OP434439,OP434443,OP434445 |
| 259 | VC_1235_0 | 12 | 6765.183035 | 1693.982031 | 0.799745954 | 2.14E-20 | 3 | 8.717797887 | 66 | 66 | 1 | 5.748341058 | 0 | 0 | 0 MG711460,S1-wms_4277,S11-wms-combined_283220,S11-wms_3116,S12-wms-combined_299872,S12-wms_193565,S17-wms-combined_175924,S17-wms-combined_193958,S2-wms-combined_165833,S7-wms_113137,S7-wms_200936,S8-wms_30498 |
| 260 | VC_1235_1 | 1 | 0 | 898.6757519 | 0 | 0 | 0 | 0 | 1 | 1 | 1 | 0 | 0 | 0 | 0 S2-wms-combined_157934 |
| 261 | VC_1236_0 | 3 | 66.88739326 | 405.8522353 | 0.141488865 | 0.002252261 | 4.123105626 | 6.557438524 | 3 | 3 | 1 | 5.720428283 | 0 | 0 | 0 S8-wms-combined_134284,S8-wms-combined_211119,S8-wms-combined_49369 |
| 262 | VC_1238_0 | 2 | 21.2201661 | 179.9390334 | 0.105489414 | 0.08545176 | 8.94427191 | 8.94427191 | 1 | 1 | 1 | 8.94427191 | 0 | 0 | 0 S12-wms-combined_147628,S12-wms-combined_41160 |
| 263 | VC_1239_0 | 6 | 307.2945309 | 346.7136276 | 0.469863452 | 2.78E-07 | 5.291502622 | 8.831760866 | 15 | 15 | 1 | 6.821570877 | 0 | 0 | 0 S1-wms_185046,S11-wms_303398,S12-wms-combined_334806,S7-wms-combined_40779,S8-wms-combined_134098,S8-wms-combined_25607 |
| 264 | VC_123_0 | 23 | 37012.61764 | 8289.344677 | 0.817020185 | 1.60E-95 | 4.123105626 | 8.717797887 | 253 | 253 | 1 | 6.763447395 | 0 | 0 | 0 JX866719,KJ021043,KX237516,MF663761,MG197810,MG432137,MK521906,MK685667,MK798142,MN689778,MT559528,MT757392,MT894004,MZ374361,MZ398021,MZ398245,MZ504995,OK499978,ON086804,ON602758,ON807605,ON809560,OP114730 |
| 265 | VC_1240_0 | 82 | 413485.3 | 106963.1009 | 0.794478952 | 0 | 0 | 8.94427191 | 3321 | 3321 | 1 | 6.48111885 | 0 | 0 | 0 MG839015,MG839016,MG839017,MG839018,MG839020,MG839021,MG839022,MG839023,MG839026,MG839028,MG839029,MG920061,MG925352,MG944219,MG944222,MG962360,MG962367,MH045554,MH045558,MH045560,MH045562,MH045563,MH045564,MH045565,MH153802,MH153805,MH153806,MH153811,MH513981,MK620895,MK620902,MK620903,MK620905,MK757447,MK919471,MK967388,MK967391,MK967394,MK967400,MK977697,MN062702,MN062709,MN062713,MN234163,MN234177,MN234181,MN234204,MN234222,MN369737,MN617840,MN735430,MN735435,MN807246,MN892483,MT024864,MT310851,MT310856,MT310861,MT310866,MT310873,MT451985,MT498049,MT553339,MT553341,MT553343,MT771350,MT952846,MT986026,MT986027,MW055911,MW291026,MW435859,MZ005684,MZ958749,OK310506,ON260823,ON337194,ON456354,ON970581,ON970605,ON970606,OP021683 |
| 266 | VC_1243_0 | 52 | 6748.865982 | 645.9660172 | 0.912646289 | 0.072916892 | 0 | 3.16227766 | 1326 | 1326 | 1 | 2.070750549 | 0 | 0 | 0 MG883709,MG945260,MG945282,MG945390,MG945396,MG945452,MG945485,MG945511,MG945515,MG945618,MG945634,MG945641,MG945647,MG945743,MH572525,MH572526,MK496694,MK496775,MK765561,MK765563,MK765564,MK765568,MK765569,MK765594,MK765638,MN311493,MT135347,MT135352,MT135355,MT135366,MT135381,MT135382,MT135387,MT135444,MW202703,MW202824,MZ089786,MZ089791,MZ089795,MZ089806,OL617028,OM869549,OM869559,OM869560,OM869562,OM869569,OM869650,OP549868,OP549871,OP549874,S1-wms-combined_357373,S9-wms_128232 |
| 267 | VC_1245_0 | 2 | 14.75294763 | 0 | 1 | 0 | 0 | 0 | 1 | 1 | 1 | 0 | 0 | 0 | 0 MG883710,MG945464 |
| 268 | VC_1246_0 | 2 | 18.57572439 | 0 | 1 | 0 | 0 | 0 | 1 | 1 | 1 | 0 | 0 | 0 | 0 MG883713,MT135394 |
| 269 | VC_1247_0 | 7 | 328.4875003 | 33.05080297 | 0.908582845 | 1.04E-05 | 0 | 1.414213562 | 21 | 21 | 1 | 0.543533979 | 0 | 0 | 0 MG883720,MG945384,MG945482,MG945692,MG945697,MG945698,MG945734 |
| 270 | VC_124_0 | 2 | 93.60729906 | 144.3398453 | 0.393395346 | 0.046629082 | 6.32455532 | 6.32455532 | 1 | 1 | 1 | 6.32455532 | 0 | 0 | 0 KF977490,MW960031 |
| 271 | VC_1250_0 | 5 | 91.09487203 | 51.60415612 | 0.638370655 | 0.000810551 | 0 | 1.732050808 | 10 | 10 | 1 | 1.258505748 | 0 | 0 | 0 MG883724,MG945625,MG945629,MG945632,MG945633 |
| 272 | VC_1251_0 | 15 | 1494.860866 | 151.1350927 | 0.908180156 | 4.90E-21 | 0 | 2.645751311 | 105 | 105 | 1 | 2.134916383 | 0 | 0 | 0 MG945128,MG945159,MG945188,MG945229,MG945255,MG945323,MG945332,MG945401,MG945685,MG945745,MG945774,MG945803,MG945829,MT135336,MT135441 |
| 273 | VC_1252_0 | 3 | 56.32923145 | 117.6897036 | 0.32369599 | 0.002096069 | 1.414213562 | 1.414213562 | 3 | 3 | 1 | 1.414213562 | 0 | 0 | 0 MG945544,MG945723,MT135439 |
| 274 | VC_1253_0 | 6 | 286.7807044 | 183.5543355 | 0.60973706 | 2.35E-09 | 0 | 2.236067977 | 15 | 15 | 1 | 1.52363122 | 0 | 0 | 0 MG945414,MG945445,MG945578,MG945611,MT135367,MT135433 |
| 275 | VC_1255_0 | 4 | 47.73982594 | 144.7590607 | 0.24800053 | 0.001718468 | 0 | 2.236067977 | 6 | 6 | 1 | 1.578689326 | 0 | 0 | 0 MG945460,MG945496,MG945622,MG945830 |

| | | | | | | | | | | | | | | | | | |
|---|---|---|---|---|---|---|---|---|---|---|---|---|---|---|---|---|---|
| 276 | VC_1256_0 | 4 | 43.53488751 | 31.39820017 | 0.580983499 | 0.052482568 | 1 | 2 | 6 | 6 | | 1 | 1.54875479 | 0 | 0 | 0 | MG945241,MG945276,MG945448,MG945643 |
| 277 | VC_1257_0 | 6 | 161.0906594 | 86.07779142 | 0.651744423 | 3.06E-07 | 0 | 1 | 15 | 15 | | 1 | 0.333333333 | 0 | 0 | 0 | MG945140,MG945257,MG945449,MG945458,MG945662,MG945815 |
| 278 | VC_1258_0 | 3 | 13.11938141 | 41.05708426 | 0.242160157 | 0.140741807 | 1 | 1.414213562 | 3 | 3 | | 1 | 1.138071187 | 0 | 0 | 0 | MG945580,MG945804,MG945909 |
| 279 | VC_1259_0 | 8 | 604.3146253 | 130.4088202 | 0.822506249 | 3.73E-12 | 0 | 1.414213562 | 28 | 28 | | 1 | 0.63672954 | 0 | 0 | 0 | MG945166,MG945208,MG945329,MG945365,MG945512,MG945540,MG945687,MT135329 |
| 280 | VC_125_0 | 17 | 13545.03156 | 1107.121122 | 0.924439695 | 2.11E-65 | 1.732050808 | 8.246211251 | 136 | 136 | | 1 | 6.421128798 | 0 | 0 | 0 | AB775548,GU903191,KM360178,KX231828,KY705409,MH252365,MT012302,MT161381,MT161382,MT161383,MT161384,MT375042,MT613935,MT701586,MT708550,MZ345003,ON853583 |
| 281 | VC_1260_0 | 2 | 9.796278594 | 0 | 1 | 0 | 1 | 1 | 1 | 1 | | 1 | 1 | 0 | 0 | 0 | MG945165,MT135354 |
| 282 | VC_1261_0 | 5 | 152.6286803 | 46.88638112 | 0.764998287 | 7.00E-05 | 0 | 1.414213562 | 10 | 10 | | 1 | 0.965685425 | 0 | 0 | 0 | MG945177,MG945349,MG945411,MG945462,MG945495 |
| 283 | VC_1262_0 | 2 | 16.30924847 | 120.35741 | 0.119335972 | 0.043864358 | 0 | 0 | 1 | 1 | | 1 | 0 | 0 | 0 | 0 | MN311487,MN311492 |
| 284 | VC_1263_0 | 2 | 3.84840074 | 0 | 1 | 0 | 0 | 0 | 1 | 1 | | 1 | 0 | 0 | 0 | 0 | MG945183,MG945314 |
| 285 | VC_1264_0 | 6 | 52.05710841 | 21.66126294 | 0.706161944 | 0.079442752 | 0 | 2 | 15 | 15 | | 1 | 1.214715465 | 0 | 0 | 0 | MG945261,MG945420,MG945470,MG945518,MK249148,MK496685 |
| 286 | VC_1266_0 | 5 | 57.93390719 | 0 | 1 | 0 | 0 | 2 | 10 | 10 | | 1 | 1.292820323 | 0 | 0 | 0 | MG945189,MG945275,MG945408,MG945848,MG945852 |
| 287 | VC_1267_0 | 7 | 233.5670107 | 0 | 1 | 0 | 0 | 1.414213562 | 21 | 21 | | 1 | 0.808122036 | 0 | 0 | 0 | MG945209,MG945263,MG945279,MG945404,MG945750,MG945776,MG945791 |
| 288 | VC_1268_0 | 3 | 14.50934719 | 165.3295597 | 0.08067969 | 0.313746804 | 1.414213562 | 1.414213562 | 3 | 3 | | 1 | 1.414213562 | 0 | 0 | 0 | MG945212,MG945582,MT135345 |
| 289 | VC_126_0 | 2 | 81.74986358 | 410.4355049 | 0.16609568 | 0.048184325 | 5.196152423 | 5.196152423 | 1 | 1 | | 1 | 5.196152423 | 0 | 0 | 0 | EF057797,EU399241 |
| 290 | VC_1270_0 | 3 | 54.79870562 | 25.69470678 | 0.680784973 | 0.013119721 | 1 | 1.414213562 | 3 | 3 | | 1 | 1.138071187 | 0 | 0 | 0 | MG945214,MT135380,MT135423 |
| 291 | VC_1271_0 | 2 | 24.45666682 | 0 | 1 | 0 | 0 | 0 | 1 | 1 | | 1 | 0 | 0 | 0 | 0 | MG945230,MT135357 |
| 292 | VC_1272_0 | 5 | 166.2088324 | 0 | 1 | 0 | 0 | 1 | 10 | 10 | | 1 | 0.6 | 0 | 0 | 0 | MG945235,MG945392,MT135388,MT135443,MT135462 |
| 293 | VC_1273_0 | 6 | 144.1261857 | 51.03733489 | 0.738489372 | 1.96E-05 | 0 | 1.414213562 | 15 | 15 | | 1 | 0.816176046 | 0 | 0 | 0 | MG945236,MG945822,MT135358,MT135377,MT135426,MT135427 |
| 294 | VC_1274_0 | 2 | 13.23885114 | 7.492687495 | 0.638585074 | 0.270145687 | 0 | 0 | 1 | 1 | | 1 | 0 | 0 | 0 | 0 | MG945567,MG945631 |
| 295 | VC_1275_0 | 4 | 83.76514101 | 0 | 1 | 0 | 0 | 1.732050808 | 6 | 6 | | 1 | 1.215421457 | 0 | 0 | 0 | MG945252,MG945584,MN379645,OM869494 |
| 296 | VC_1276_0 | 7 | 240.2427152 | 36.07126754 | 0.869455511 | 0.000175243 | 0 | 2.645751311 | 21 | 21 | | 1 | 1.617774418 | 0 | 0 | 0 | MG945488,MG945572,MZ089802,MZ089808,MZ089809,OP549867,S17-wms_254872 |
| 297 | VC_1277_0 | 14 | 558.2043093 | 121.058678 | 0.821779369 | 2.24E-09 | 0 | 2.236067977 | 91 | 91 | | 1 | 1.532350718 | 0 | 0 | 0 | MG945494,MG945563,MG945585,MK765590,MN379638,MN379648,MT135333,MT135379,MT135391,MT135401,MT135437,MT135445,OM869612,OM869668 |
| 298 | VC_1278_0 | 3 | 56.32923145 | 85.86606972 | 0.396139893 | 0.002947141 | 1 | 1.414213562 | 3 | 3 | | 1 | 1.138071187 | 0 | 0 | 0 | MG945425,MG945789,MT135415 |
| 299 | VC_1279_0 | 4 | 32.68044565 | 21.26440339 | 0.605812162 | 0.230279085 | 1 | 2 | 6 | 6 | | 1 | 1.426746322 | 0 | 0 | 0 | MG945258,MG945434,MT135434,MT135451 |
| 300 | VC_127_0 | 1 | 0 | 152.4074347 | 0 | 0 | 0 | 0 | 1 | 1 | | 1 | 0 | 0 | 0 | 0 | MF663786 |
| 301 | VC_127_1 | 1 | 0 | 131.988923 | 0 | 0 | 0 | 0 | 1 | 1 | | 1 | 0 | 0 | 0 | 0 | OL614104 |
| 302 | VC_127_2 | 1 | 0 | 165.0256777 | 0 | 0 | 0 | 0 | 1 | 1 | | 1 | 0 | 0 | 0 | 0 | MT711888 |
| 303 | VC_1280_0 | 2 | 9.796278594 | 131.1654061 | 0.069496038 | 0.050553389 | 0 | 0 | 1 | 1 | | 1 | 0 | 0 | 0 | 0 | MK765572,OM869521 |
| 304 | VC_1281_0 | 7 | 336.1779181 | 28.75601342 | 0.921202138 | 4.71E-05 | 0 | 1.414213562 | 21 | 21 | | 1 | 0.908146086 | 0 | 0 | 0 | MG945278,MG945285,MG945492,MG945605,MG945751,MG945812,MG945825 |
| 305 | VC_1282_0 | 3 | 14.20750251 | 0 | 1 | 0 | 0 | 0 | 3 | 3 | | 1 | 0 | 0 | 0 | 0 | MG945280,MK765649,OM869500 |
| 306 | VC_1283_0 | 26 | 6186.976235 | 5174.279777 | 0.54456798 | 9.52E-112 | 0 | 3.31662479 | 325 | 325 | | 1 | 2.257867212 | 0 | 0 | 0 | MK765552,MK765553,MK765556,MK765560,MK765562,MK765565,MK765591,MK765592,MK765593,MK765596,MK765606,MK765607,MK765609,MK765611,MK765612,MK765613,MK765615,MK765633,MK765648,MK765651,MK765657,MK765658,MK765660,MK765666,MT135276,MT135425 |
| 307 | VC_1285_0 | 25 | 2092.084258 | 0 | 1 | 0 | 0 | 2.449489743 | 300 | 300 | | 1 | 1.560084368 | 0 | 0 | 0 | MG945316,MG945344,MG945468,MG945524,MG945556,MG945562,MG945579,MG945612,MG945640,MG945668,MG945712,MG945733,MG945806,MG945814,MK765571,MK765574,MK765576,MK765577,MK765580,MT135330,MT135406,MT135431,MT135458,OL617036,OM869623 |
| 308 | VC_1286_0 | 1 | 0 | 4.936518833 | 0 | 0 | 0 | 0 | 1 | 1 | | 1 | 0 | 0 | 0 | 0 | MG945328 |
| 309 | VC_1287_0 | 7 | 245.4147848 | 0 | 1 | 0 | 0 | 1.414213562 | 21 | 21 | | 1 | 0.908146086 | 0 | 0 | 0 | MG945331,MG945343,MG945490,MG945501,MG945665,MG945729,MG945782 |
| 310 | VC_1288_0 | 3 | 54.11289743 | 0 | 1 | 0 | 0 | 2.236067977 | 3 | 3 | | 1 | 1.490711985 | 0 | 0 | 0 | MG945335,MG945778,MT135385 |
| 311 | VC_1289_0 | 2 | 15.53109722 | 17.79379432 | 0.466050946 | 0.144422183 | 1 | 1 | 1 | 1 | | 1 | 1 | 0 | 0 | 0 | MG945336,MT135365 |
| 312 | VC_1290_0 | 2 | 22.90036782 | 0 | 1 | 0 | 1 | 1 | 1 | 1 | | 1 | 1 | 0 | 0 | 0 | MG945337,MT135410 |
| 313 | VC_1291_0 | 2 | 3.84840074 | 0 | 1 | 0 | 1.732050808 | 1.732050808 | 1 | 1 | | 1 | 1.732050808 | 0 | 0 | 0 | MG945345,MG945818 |
| 314 | VC_1292_0 | 3 | 17.37708608 | 0 | 1 | 0 | 0 | 1.414213562 | 3 | 3 | | 1 | 0.942809042 | 0 | 0 | 0 | MG945348,MG945702,MK496732 |
| 315 | VC_1293_0 | 4 | 77.49339696 | 21.26440339 | 0.784681278 | 0.007107019 | 1 | 1.732050808 | 6 | 6 | | 1 | 1.451123717 | 0 | 0 | 0 | MG945350,MG945432,MG945755,MT135447 |
| 316 | VC_1294_0 | 6 | 124.4392727 | 63.36048707 | 0.662616783 | 7.28E-06 | 0 | 2 | 15 | 15 | | 1 | 1.284481349 | 0 | 0 | 0 | MG945354,MG945576,MG945637,MK496765,OM869571,OP209176 |
| 317 | VC_1295_0 | 3 | 76.07927043 | 0 | 1 | 1 | 0 | 0 | 3 | 3 | | 1 | 0 | 0 | 0 | 0 | MG945360,MG945478,MG945614 |
| 318 | VC_1296_0 | 4 | 33.20055058 | 5.033427347 | 0.868351984 | 0.088483245 | 0 | 1 | 6 | 6 | | 1 | 0.5 | 0 | 0 | 0 | MG945362,MK496704,MK496729,MK496782 |
| 319 | VC_1297_0 | 7 | 98.07626717 | 0 | 1 | 0 | 0 | 1.414213562 | 21 | 21 | | 1 | 0.812907991 | 0 | 0 | 0 | MG945395,MG945421,MG945480,MG945539,MG945696,MK496696,MK496711 |
| 320 | VC_1298_0 | 3 | 29.18471806 | 30.51523872 | 0.488856603 | 0.018325274 | 1 | 2 | 3 | 3 | | 1 | 1.577350269 | 0 | 0 | 0 | MG945397,MT135331,MT135461 |
| 321 | VC_1299_0 | 2 | 19.17778267 | 0 | 1 | 0 | 0 | 0 | 1 | 1 | | 1 | 0 | 0 | 0 | 0 | MG945409,OM869563 |
| 322 | VC_129_0 | 43 | 122177.171 | 15725.17723 | 0.885968749 | 1.72E-289 | 0 | 6.633249581 | 903 | 903 | | 1 | 3.916307313 | 0 | 0 | 0 | AF493143,KJ507099,KR091952,KU064779,KY798120,KY798121,KY971611,LC730319,LN889756,LN889995,LN913017,MG250485,MG775258,MN729595,MN729596,MN729597,MN729598,MN729599,MN729600,MT104465,MT104466,MT711889,MT711890,MW812345,MZ501272,MZ598487,MZ826344,MZ826345,MZ826346,MZ826347,MZ826348,MZ826349,MZ826351,MZ826352,MZ826354,OK523998,OL581611,OM256450,OM282085,OM471789,OM630590,ON189046 |
| 323 | VC_12_0 | 21 | 20517.63897 | 69729.17433 | 0.227350288 | 2.58E-127 | 2.828427125 | 9.219544457 | 210 | 201 | 0.957142857 | 1 | 7.894974153 | 0 | 0 | 0 | AP008954,AY954960,AY954961,CP062422,CP062462,CP071049,DQ530362,JQ973847,KJ140076,KJ452291,KP209285,KP893289,KT344895,MK290764,MK618716,MW238767,OM439663,OM439669,OM439672,OM439673,OM439674 |
| 324 | VC_12_1 | 2 | 186.7549706 | 9526.452074 | 0.019226911 | 0.042605046 | 7.141428429 | 7.141428429 | 1 | 1 | | 1 | 7.141428429 | 0 | 0 | 0 | MG543995,MG564297 |
| 325 | VC_12_2 | 19 | 17763.05043 | 70266.55697 | 0.201784956 | 2.99E-104 | 0 | 8.774964387 | 171 | 171 | | 1 | 7.519158643 | 0 | 0 | 0 | AB370268,AP001553,AY954962,AY954963,AY954964,AY954965,AY954966,AY954967,AY954968,DQ908929,JX013863,JX094501,KC595279,KP893290,KR709302,KR709303,MK211557,MN904510,ON571632 |
| 326 | VC_1300_0 | 2 | 13.78291419 | 6.975321055 | 0.663973311 | 0.270145687 | 1 | 1 | 1 | 1 | | 1 | 1 | 0 | 0 | 0 | MG945412,MG945828 |
| 327 | VC_1301_0 | 2 | 29.79354264 | 0 | 1 | 0 | 0 | 0 | 1 | 1 | | 1 | 0 | 0 | 0 | 0 | MG945417,MG945419 |
| 328 | VC_1302_0 | 3 | 23.13612536 | 0 | 1 | 0 | 0 | 1 | 3 | 3 | | 1 | 0.666666667 | 0 | 0 | 0 | MG945427,MG945574,MT135411 |
| 329 | VC_1303_0 | 6 | 86.06224956 | 0 | 1 | 0 | 0 | 1.732050808 | 15 | 15 | | 1 | 1.26994394 | 0 | 0 | 0 | MG945439,MG945628,MK496757,MK765601,MZ089768,OM869696 |
| 330 | VC_1304_0 | 7 | 202.9578452 | 52.93487507 | 0.793136456 | 1.99E-07 | 0 | 1.732050808 | 21 | 21 | | 1 | 1.319333969 | 0 | 0 | 0 | MZ089804,MZ089807,OM869574,OM869576,OM869577,OM869592,OM869653 |
| 331 | VC_1305_0 | 5 | 64.07936426 | 19.12586863 | 0.770136229 | 0.330149589 | 0 | 1.414213562 | 10 | 10 | | 1 | 0.882842712 | 0 | 0 | 0 | MG945469,MG945824,MT135353,MT135400,MT135402 |
| 332 | VC_1306_0 | 2 | 15.53109722 | 0 | 1 | 0 | 0 | 0 | 1 | 1 | | 1 | 0 | 0 | 0 | 0 | MG945483,MT135420 |
| 333 | VC_1307_0 | 2 | 9.796278594 | 8.969958177 | 0.522016146 | 0.270145687 | 1.732050808 | 1.732050808 | 1 | 1 | | 1 | 1.732050808 | 0 | 0 | 0 | MT135397,MT135407 |
| 334 | VC_1308_0 | 12 | 413.9025686 | 91.86867639 | 0.818359234 | 0.018926896 | 0 | 2 | 66 | 66 | | 1 | 1.418512581 | 0 | 0 | 0 | MG945500,MG945621,MH572369,MK765583,MT135326,OM869498,OM869502,OM869505,OM869587,OM869604,OM869633,OM869691 |
| 335 | VC_1309_0 | 2 | 4.635491837 | 10.36788319 | 0.308963272 | 0.270145687 | 0 | 0 | 1 | 1 | | 1 | 0 | 0 | 0 | 0 | MT135328,OM869626 |
| 336 | VC_130_0 | 19 | 21305.78605 | 13807.89769 | 0.606765904 | 7.61E-95 | 0 | 5.830951895 | 171 | 171 | | 1 | 4.244654662 | 0 | 0 | 0 | AB775549,FR823298,GU583987,JX173487,KP025626,KX431888,KX449361,KX449362,KX449363,MH179472,MH179475,MH179478,MH268168,MK033137,MT094431,MT711887,OK094519,OM131411,ON109291 |
| 337 | VC_1310_0 | 2 | 4.09143279 | 0 | 1 | 0 | 2.449489743 | 2.449489743 | 1 | 1 | | 1 | 2.449489743 | 0 | 0 | 0 | MG945504,MG945617 |
| 338 | VC_1311_0 | 5 | 157.3924036 | 31.8757975 | 0.831583978 | 0.001209473 | 0 | 1 | 10 | 10 | | 1 | 0.6 | 0 | 0 | 0 | MG945529,MG945568,MG945717,MG945739,MT135265 |
| 339 | VC_1312_0 | 3 | 71.65227999 | 0 | 1 | 0 | 1 | 2 | 3 | 3 | | 1 | 1.577350269 | 0 | 0 | 0 | MG945533,MT135343,MT135416 |
| 340 | VC_1313_0 | 13 | 534.5643427 | 95.76004215 | 0.848078157 | 0.125435675 | 0 | 2.449489743 | 78 | 78 | | 1 | 1.66468498 | 0 | 0 | 0 | MZ089798,MZ089799,MZ089800,MZ089801,MZ089803,MZ089805,OM869557,OM869648,OM869652,OM869654,OM869655,OM869666,OM869684 |
| 341 | VC_1314_0 | 4 | 79.74794015 | 38.50093659 | 0.674407591 | 0.000970101 | 0 | 1.414213562 | 6 | 6 | | 1 | 0.707106781 | 0 | 0 | 0 | MZ089796,MZ089797,OM869566,OM869568 |
| 342 | VC_1315_0 | 4 | 52.85941249 | 13.17737036 | 0.800454083 | 0.011246719 | 0 | 1.414213562 | 6 | 6 | | 1 | 0.971404521 | 0 | 0 | 0 | MT135349,MT135363,MT135392,MT135408 |
| 343 | VC_1319_0 | 2 | 8.282189768 | 0 | 1 | 0 | 0 | 0 | 1 | 1 | | 1 | 0 | 0 | 0 | 0 | MG945554,MG945575 |
| 344 | VC_131_0 | 17 | 40800 | 111970.7868 | 0.267066766 | 4.43E-68 | 0 | 7.280109889 | 136 | 136 | | 1 | 3.751751135 | 0 | 0 | 0 | HE600015,KJ716335,KM209228,KM209270,KM209320,KY942056,KY942057,MG266157,MH807812,MH807820,MN788369,MN813048,MN813049,MN813050,MN813051,MN813052,MN813053 |
| 345 | VC_131_1 | 6 | 4500 | 74453.41947 | 0.056995631 | 7.64E-09 | 0 | 7.348469228 | 15 | 15 | | 1 | 6.039441195 | 0 | 0 | 0 | MW650886,OL656110,ON184125,ON184126,ON212266,ON862890 |
| 346 | VC_131_2 | 7 | 6300 | 85706.09621 | 0.068473724 | 8.87E-12 | 6.8556546 | 9.16515139 | 21 | 19 | 0.904761905 | 1 | 7.971938225 | 0 | 0 | 0 | FJ373894,JX181829,LC373201,MK509462,MT078988,OL615011,OL615013 |
| 347 | VC_131_3 | 1 | 0 | 14145.51018 | 0 | 0 | 0 | 0 | 1 | 1 | | 1 | 0 | 0 | 0 | 0 | MT857002 |
| 348 | VC_1321_0 | 14 | 920.2479817 | 193.7988885 | 0.826040633 | 7.70E-11 | 0 | 2.645751311 | 91 | 91 | | 1 | 1.663397128 | 0 | 0 | 0 | MZ089770,MZ089772,MZ089780,OM869514,OM869529,OM869540,OM869542,OM869544,OM869548,OM869553,OM869589,OM869590,OM869638,OM869707 |
| 349 | VC_1322_0 | 2 | 5.033427347 | 0 | 1 | 0 | 0 | 0 | 1 | 1 | | 1 | 0 | 0 | 0 | 0 | MG945615,MT135372 |
| 350 | VC_1323_0 | 6 | 119.9922707 | 80.65722269 | 0.598019306 | 0.005396109 | 0 | 1.732050808 | 15 | 15 | | 1 | 1.188513211 | 0 | 0 | 0 | MH572374,MH572375,MH572376,MH572414,MH572485,MH572522 |
| 351 | VC_1324_0 | 4 | 26.18077238 | 17.39585731 | 0.600798469 | 0.455926385 | 0 | 1 | 6 | 6 | | 1 | 0.5 | 0 | 0 | 0 | MG945619,MH572524,MH992173,MZ364234 |
| 352 | VC_1325_0 | 7 | 496.162381 | 45.16902334 | 0.916559389 | 1.89E-07 | 0 | 2.645751311 | 21 | 21 | | 1 | 1.259881577 | 0 | 0 | 0 | MK765555,MK765567,MK765600,MK765610,MK765617,MK765659,MK765664 |
| 353 | VC_1326_0 | 2 | 3.644286755 | 0 | 1 | 0 | 1.414213562 | 1.414213562 | 1 | 1 | | 1 | 1.414213562 | 0 | 0 | 0 | MG945690,MK496692 |
| 354 | VC_1327_0 | 4 | 79.74794015 | 0 | 1 | 0 | 0 | 1 | 6 | 6 | | 1 | 0.5 | 0 | 0 | 0 | MG945724,MT135383,MT135403,MT135456 |
| 355 | VC_1328_0 | 2 | 3.088008248 | 0 | 1 | 0 | 2 | 2 | 1 | 1 | | 1 | 2 | 0 | 0 | 0 | MG945766,MG945823 |
| 356 | VC_1329_0 | 2 | 3.644286755 | 0 | 1 | 0 | 0 | 0 | 1 | 1 | | 1 | 0 | 0 | 0 | 0 | MG945769,MG945792 |
| 357 | VC_132_0 | 4 | 1800 | 48239.29813 | 0.035971728 | 1.22E-05 | 4.795831523 | 7.681145748 | 6 | 6 | | 1 | 6.263796994 | 0 | 0 | 0 | JX878496,KX452697,MF285619,MH929319 |
| 358 | VC_132_1 | 17 | 40800 | 193899.6083 | 0.173839233 | 1.28E-86 | 0 | 9.273618495 | 136 | 134 | 0.985294118 | 1 | 7.342700174 | 0 | 0 | 0 | AB797215,KX147096,KY073123,MG428990,MG428991,MG770379,MK373783,MN045230,MN478483,MZ634341,ON202820,ON323462,ON399185,OX335402,OX335403,OX335405,OX335416 |
| 359 | VC_1330_0 | 2 | 15.53109722 | 0 | 1 | 0 | 0 | 0 | 1 | 1 | | 1 | 0 | 0 | 0 | 0 | MG945788,MG945795 |
| 360 | VC_1331_0 | 4 | 85.81825823 | 31.05885908 | 0.734260565 | 0.001005414 | 0 | 1.414213562 | 6 | 6 | | 1 | 0.707106781 | 0 | 0 | 0 | MG945798,MK765653,MT135350,MT135457 |
| 361 | VC_1332_0 | 2 | 24.45666682 | 31.05885908 | 0.440537425 | 0.080290302 | 0 | 0 | 1 | 1 | | 1 | 0 | 0 | 0 | 0 | OM869639,OM869679 |
| 362 | VC_1333_0 | 2 | 19.17778267 | 0 | 1 | 0 | 0 | 0 | 1 | 1 | | 1 | 0 | 0 | 0 | 0 | MG945808,MG945849 |
| 363 | VC_1334_0 | 1 | 0 | 13.57530437 | 0 | 0 | 0 | 0 | 1 | 1 | | 1 | 0 | 0 | 0 | 0 | MG945821 |
| 364 | VC_1335_0 | 7 | 252.3056609 | 50.00905139 | 0.834579498 | 1.56E-06 | 0 | 1.732050808 | 21 | 21 | | 1 | 1.017314122 | 0 | 0 | 0 | MG945831,MT135370,MT135373,MT135418,MT135430,MT135455,MT135459 |
| 365 | VC_1336_0 | 3 | 48.98470788 | 50.00905139 | 0.494826222 | 0.00471016 | 0 | 1.414213562 | 3 | 3 | | 1 | 0.942809042 | 0 | 0 | 0 | OP549864,OP549865,OP549866 |
| 366 | VC_1337_0 | 2 | 14.98702917 | 0 | 1 | 0 | 1 | 1 | 1 | 1 | | 1 | 1 | 0 | 0 | 0 | MG945832,MT135421 |
| 367 | VC_1338_0 | 4 | 65.83420152 | 0 | 1 | 0 | 0 | 0 | 6 | 6 | | 1 | 0 | 0 | 0 | 0 | MG945839,MH572484,MZ364252,MZ364293 |
| 368 | VC_1339_0 | 7 | 211.2939182 | 18.93990286 | 0.917736227 | 0.002276952 | 0 | 2 | 21 | 21 | | 1 | 1.361355108 | 0 | 0 | 0 | MG945842,MH992159,MH992175,MH992200,MK249218,MK249229,MK249233 |
| 369 | VC_133_0 | 2 | 300 | 47353.71101 | 0.006295417 | 0.059611162 | 6.32455532 | 6.32455532 | 1 | 1 | | 1 | 6.32455532 | 0 | 0 | 0 | KF669653,KF669654 |
| 370 | VC_133_1 | 1 | 0 | 30799.98837 | 0 | 0 | 0 | 0 | 1 | 1 | | 1 | 0 | 0 | 0 | 0 | MG428992 |
| 371 | VC_133_10 | 10 | 13500 | 242843.0135 | 0.052663811 | 1.02E-22 | 1.732050808 | 7.615773106 | 45 | 45 | | 1 | 5.623420291 | 0 | 0 | 0 | MH709121,MN871504,MZ375357,MZ375358,OL964736,OL964737,OL964738,OL964746,OL964747,OL964753 |
| 372 | VC_133_11 | 2 | 300 | 49793.20862 | 0.005988836 | 0.060950463 | 0 | 0 | 1 | 1 | | 1 | 0 | 0 | 0 | 0 | MW006476,MW006480 |
| 373 | VC_133_12 | 2 | 300 | 58813.90421 | 0.005074948 | 0.132791703 | 1.732050808 | 1.732050808 | 1 | 1 | | 1 | 1.732050808 | 0 | 0 | 0 | MN871494,MN871495 |
| 374 | VC_133_13 | 2 | 300 | 58951.9738 | 0.005063122 | 0.132052319 | 7.348469228 | 7.348469228 | 1 | 1 | | 1 | 7.348469228 | 0 | 0 | 0 | JN126049,LR800396 |
| 375 | VC_133_14 | 3 | 900 | 87520.98781 | 0.010178579 | 0.036727444 | 5.099019514 | 6.403124237 | 3 | 3 | | 1 | 5.748902133 | 0 | 0 | 0 | KX438380,MN953776,OM681507 |
| 376 | VC_133_15 | 2 | 300 | 58092.73182 | 0.005137626 | 0.152135634 | 4.472135955 | 4.472135955 | 1 | 1 | | 1 | 4.472135955 | 0 | 0 | 0 | KX765865,MW388005 |
| 377 | VC_133_16 | 1 | 0 | 29932.11568 | 0 | 0 | 0 | 0 | 1 | 1 | | 1 | 0 | 0 | 0 | 0 | KJ174318 |
| 378 | VC_133_17 | 1 | 0 | 30320.54851 | 0 | 0 | 0 | 0 | 1 | 1 | | 1 | 0 | 0 | 0 | 0 | MH709120 |
| 379 | VC_133_18 | 1 | 0 | 29829.57656 | 0 | 0 | 0 | 0 | 1 | 1 | | 1 | 0 | 0 | 0 | 0 | KR296692 |
| 380 | VC_133_19 | 2 | 300 | 58836.74104 | 0.005072988 | 0.130335699 | 6.8556546 | 6.8556546 | 1 | 1 | | 1 | 6.8556546 | 0 | 0 | 0 | MN871500,MT074475 |
| 381 | VC_133_2 | 1 | 0 | 30925.65194 | 0 | 0 | 0 | 0 | 1 | 1 | | 1 | 0 | 0 | 0 | 0 | MT074472 |
| 382 | VC_133_20 | 1 | 0 | 29371.99133 | 0 | 0 | 0 | 0 | 1 | 1 | | 1 | 0 | 0 | 0 | 0 | OM367911 |
| 383 | VC_133_21 | 1 | 0 | 29851.20891 | 0 | 0 | 0 | 0 | 1 | 1 | | 1 | 0 | 0 | 0 | 0 | MK972690 |
| 384 | VC_133_22 | 13 | 23400 | 267560.268 | 0.080423352 | 5.67E-47 | 1.414213562 | 10.81665383 | 78 | 64 | 0.820512821 | 1 | 7.867173998 | 0 | 0 | 0 | FQ312032,JX128257,JX181828,KJ174317,MH427377,MH688040,MH940212,MN066127,MN342150,MW021753,MW021757,OK322699,OP610151 |
| 385 | VC_133_23 | 1 | 0 | 28414.90037 | 0 | 0 | 0 | 0 | 1 | 1 | | 1 | 0 | 0 | 0 | 0 | MT074470 |
| 386 | VC_133_24 | 1 | 0 | 29934.31938 | 0 | 0 | 0 | 0 | 1 | 1 | | 1 | 0 | 0 | 0 | 0 | MF001362 |
| 387 | VC_133_25 | 1 | 0 | 30087.36899 | 0 | 0 | 0 | 0 | 1 | 1 | | 1 | 0 | 0 | 0 | 0 | JN593240 |
| 388 | VC_133_26 | 1 | 0 | 29602.38895 | 0 | 0 | 0 | 0 | 1 | 1 | | 1 | 0 | 0 | 0 | 0 | JN673056 |
| 389 | VC_133_27 | 1 | 0 | 30658.75382 | 0 | 0 | 0 | 0 | 1 | 1 | | 1 | 0 | 0 | 0 | 0 | MG383452 |
| 390 | VC_133_28 | 3 | 900 | 84455.86086 | 0.010544091 | 0.023383628 | 4.795831523 | 9 | 3 | 2 | 0.666666667 | 1 | 7.265277174 | 0 | 0 | 0 | LR811967,LR811969,MW355478 |
| 391 | VC_133_29 | 2 | 300 | 57614.49581 | 0.00518005 | 0.141348401 | 1 | 1 | 1 | 1 | | 1 | 1 | 0 | 0 | 0 | MN871475,MN871501 |
| 392 | VC_133_3 | 3 | 900 | 84849.96131 | 0.010495632 | 0.00880425 | 4.358898944 | 8.124038405 | 3 | 3 | | 1 | 6.677590594 | 0 | 0 | 0 | MH370368,MH370371,MT074469 |
| 393 | VC_133_30 | 2 | 300 | 57389.01091 | 0.005200297 | 0.122611286 | 0 | 0 | 1 | 1 | | 1 | 0 | 0 | 0 | 0 | KX245012,KY000003 |
| 394 | VC_133_31 | 6 | 4500 | 166641.783 | 0.026293988 | 3.06E-06 | 1.414213562 | 8.062257748 | 15 | 15 | | 1 | 6.174372274 | 0 | 0 | 0 | HQ259103,MH370370,MT074476,MW355450,MW355475,MW355477 |
| 395 | VC_133_32 | 1 | 0 | 29753.55514 | 0 | 0 | 0 | 0 | 1 | 1 | | 1 | 0 | 0 | 0 | 0 | OM688977 |
| 396 | VC_133_33 | 1 | 0 | 28471.36416 | 0 | 0 | 0 | 0 | 1 | 1 | | 1 | 0 | 0 | 0 | 0 | MT499896 |
| 397 | VC_133_34 | 3 | 900 | 87040.36724 | 0.010234208 | 0.02630956 | 7.810249676 | 9.16515139 | 3 | 2 | 0.666666667 | 1 | 8.304218333 | 0 | 0 | 0 | KY630163,KY787213,OK283619 |
| 398 | VC_133_35 | 1 | 0 | 29910.96093 | 0 | 0 | 0 | 0 | 1 | 1 | | 1 | 0 | 0 | 0 | 0 | MZ156570 |
| 399 | VC_133_36 | 2 | 300 | 57912.27187 | 0.005153553 | 0.135788523 | 1 | 1 | 1 | 1 | | 1 | 1 | 0 | 0 | 0 | MN871491,MN871492 |
| 400 | VC_133_37 | 1 | 0 | 29721.8579 | 0 | 0 | 0 | 0 | 1 | 1 | | 1 | 0 | 0 | 0 | 0 | MN871493 |
| 401 | VC_133_38 | 1 | 0 | 29185.44119 | 0 | 0 | 0 | 0 | 1 | 1 | | 1 | 0 | 0 | 0 | 0 | MN871502 |
| 402 | VC_133_39 | 1 | 0 | 29958.8431 | 0 | 0 | 0 | 0 | 1 | 1 | | 1 | 0 | 0 | 0 | 0 | MT074478 |
| 403 | VC_133_4 | 1 | 0 | 30039.16754 | 0 | 0 | 0 | 0 | 1 | 1 | | 1 | 0 | 0 | 0 | 0 | LR811968 |
| 404 | VC_133_40 | 1 | 0 | 29943.85904 | 0 | 0 | 0 | 0 | 1 | 1 | | 1 | 0 | 0 | 0 | 0 | MH523359 |
| 405 | VC_133_41 | 1 | 0 | 29493.30729 | 0 | 0 | 0 | 0 | 1 | 1 | | 1 | 0 | 0 | 0 | 0 | MT074474 |
| 406 | VC_133_42 | 1 | 0 | 28165.3901 | 0 | 0 | 0 | 0 | 1 | 1 | | 1 | 0 | 0 | 0 | 0 | MN445184 |
| 407 | VC_133_43 | 4 | 1800 | 118001.9253 | 0.0150248 | 4.32E-05 | 1.732050808 | 2.449489743 | 6 | 6 | | 1 | 2.015855318 | 0 | 0 | 0 | MN871496,MN871497,MN871498,MN871499 |
| 408 | VC_133_44 | 1 | 0 | 29338.26797 | 0 | 0 | 0 | 0 | 1 | 1 | | 1 | 0 | 0 | 0 | 0 | MW021754 |
| 409 | VC_133_45 | 2 | 300 | 57527.04619 | 0.005187884 | 0.051013205 | 7.549834435 | 7.549834435 | 1 | 1 | | 1 | 7.549834435 | 0 | 0 | 0 | MT074471,MT074480 |
| 410 | VC_133_46 | 1 | 0 | 29235.47487 | 0 | 0 | 0 | 0 | 1 | 1 | | 1 | 0 | 0 | 0 | 0 | MW021760 |
| 411 | VC_133_47 | 1 | 0 | 28878.36629 | 0 | 0 | 0 | 0 | 1 | 1 | | 1 | 0 | 0 | 0 | 0 | MT074479 |
| 412 | VC_133_48 | 2 | 300 | 57909.14032 | 0.00515383 | 0.063600477 | 1.732050808 | 1.732050808 | 1 | 1 | | 1 | 1.732050808 | 0 | 0 | 0 | MK972698,MK972699 |
| 413 | VC_133_49 | 1 | 0 | 30125.20818 | 0 | 0 | 0 | 0 | 1 | 1 | | 1 | 0 | 0 | 0 | 0 | ON169963 |
| 414 | VC_133_5 | 1 | 0 | 30484.88589 | 0 | 0 | 0 | 0 | 1 | 1 | | 1 | 0 | 0 | 0 | 0 | MG748547 |
| 415 | VC_133_50 | 1 | 0 | 30634.82056 | 0 | 0 | 0 | 0 | 1 | 1 | | 1 | 0 | 0 | 0 | 0 | ON169961 |
| 416 | VC_133_51 | 1 | 0 | 31280.41455 | 0 | 0 | 0 | 0 | 1 | 1 | | 1 | 0 | 0 | 0 | 0 | ON169962 |
| 417 | VC_133_6 | 1 | 0 | 30595.22212 | 0 | 0 | 0 | 0 | 1 | 1 | | 1 | 0 | 0 | 0 | 0 | MH791405 |
| 418 | VC_133_7 | 1 | 0 | 30113.50112 | 0 | 0 | 0 | 0 | 1 | 1 | | 1 | 0 | 0 | 0 | 0 | MT974436 |
| 419 | VC_133_8 | 1 | 0 | 30068.91172 | 0 | 0 | 0 | 0 | 1 | 1 | | 1 | 0 | 0 | 0 | 0 | MT074477 |
| 420 | VC_133_9 | 3 | 900 | 89951.56384 | 0.009906269 | 0.01542891 | 2.236067977 | 8.544003745 | 3 | 3 | | 1 | 6.421784366 | 0 | 0 | 0 | KP797973,KX171211,MN384979 |
| 421 | VC_1340_0 | 5 | 71.54630972 | 0 | 1 | 0 | 1.414213562 | 2 | 10 | 10 | | 1 | 1.589949494 | 0 | 0 | 0 | MG945850,MT135335,MT135340,MT135376,MT135393 |
| 422 | VC_1341_0 | 2 | 14.98702917 | 4.392456787 | 0.773345031 | 0.5 | 1 | 1 | 1 | 1 | | 1 | 1 | 0 | 0 | 0 | MT135269,MT135429 |
| 423 | VC_1342_0 | 3 | 23.2746182 | 0 | 1 | 0 | 1.414213562 | 2.236067977 | 3 | 3 | | 1 | 1.794110782 | 0 | 0 | 0 | MG945857,MG945859,MG945866 |
| 424 | VC_1343_0 | 21 | 52416.18639 | 836.4594984 | 0.984292621 | 3.56E-52 | 0 | 8.246211251 | 210 | 210 | | 1 | 3.555195494 | 0 | 0 | 0 | MH271292,MH271301,MK737941,MK801731,MK801732,MN497954,MT522001,MT522003,MT639643,MT684591,MT684592,MT818420,MW055903,MW291025,MZ681513,OK040788,OK318957,ON526977,ON637767,ON755183,OP580517 |

| | | | | | | | | | | | | | | | | |
|---|---|---|---|---|---|---|---|---|---|---|---|---|---|---|---|---|
| 425 | VC_1344_0 | 78 | 162099.785 | 4467.495057 | 0.973179036 | 1.99E-73 | 0 | 5.477225575 | 3003 | 3003 | 1 | 3.734033554 | 0 | 0 | 0 | MH045557,MH045561,MH371111,MH371118,MH371124,MH399783,MH576963,MH651181,MH744416,MH779512,MH825700,MK524502,MK620897,MK801730,MK878899,MK894434,MN010755,MN010759,MN284892,MN329679,MN369745,MN428062,MN428064,MN585990,MN586008,MN586010,MT310864,MT316458,MT316459,MT316462,MT498038,MT498040,MT498046,MT498047,MT498051,MT498065,MT498067,MT521990,MT639642,MT639647,MT657332,MT776807,MT952850,MW055907,MW055909,MW291020,MW321655,MW507127,MW924642,MZ005676,MZ005678,MZ028628,MZ209304,MZ274307,MZ322006,MZ322008,MZ322014,MZ322027,MZ622180,MZ648031,OK040781,OK040783,OK310504,ON081334,ON108642,ON108648,ON260812,ON260814,ON392161,ON456350,ON645341,ON645345,ON755182,ON755188,ON970573,ON970589,ON970613,OP580518 |
| 426 | VC_1345_0 | 2 | 300 | 2347.092755 | 0.113331881 | 0.044208267 | 0 | 0 | 1 | 1 | 1 | 0 | 0 | 0 | 0 | MN703407,MN703414 |
| 427 | VC_1345_1 | 2 | 297.2168991 | 2812.090542 | 0.095589421 | 0.0438062 | 6.32455532 | 6.32455532 | 1 | 1 | 1 | 6.32455532 | 0 | 0 | 0 | MH183162,MT639646 |
| 428 | VC_1346_0 | 10 | 8786.703368 | 2501.558935 | 0.778392912 | 1.16E-15 | 0 | 3.741657387 | 45 | 45 | 1 | 2.51186182 | 0 | 0 | 0 | MW021755,MW021763,MW021766,OL539438,OL539439,OL539442,OL539455,OL539456,OL539464,OM135608 |
| 429 | VC_1348_0 | 5 | 2158.980254 | 1793.686937 | 0.546208459 | 1.54E-07 | 1 | 2.828427125 | 10 | 10 | 1 | 2.030600053 | 0 | 0 | 0 | MH153799,MK737944,MZ456037,MZ622170,ON260825 |
| 430 | VC_1349_0 | 7 | 5184.820658 | 15593.64902 | 0.249528514 | 1.22E-14 | 2.645751311 | 8 | 21 | 21 | 1 | 5.142584048 | 0 | 0 | 0 | MH271303,MK937591,MK937606,MK937608,MN813682,MN813684,OK040792 |
| 431 | VC_134_0 | 3 | 900 | 8514.371509 | 0.095598522 | 0.001415056 | 2 | 3.741657387 | 3 | 3 | 1 | 3.068586334 | 0 | 0 | 0 | MH363700,MK795384,OK428602 |
| 432 | VC_1350_0 | 2 | 194.2252171 | 996.3660026 | 0.16313342 | 0.046951483 | 3.16227766 | 3.16227766 | 1 | 1 | 1 | 3.16227766 | 0 | 0 | 0 | MT889371,ON970592 |
| 433 | VC_1351_0 | 2 | 20.32742815 | 0 | 1 | 0 | 6.92820323 | 6.92820323 | 1 | 1 | 1 | 6.92820323 | 0 | 0 | 0 | MH155870,MK240575 |
| 434 | VC_1352_0 | 31 | 118151.4156 | 21014.00908 | 0.848999785 | 2.04E-147 | 0 | 7.211102551 | 465 | 465 | 1 | 4.709667751 | 0 | 0 | 0 | MH155873,MH271304,MH271312,MH271321,MH590600,MK524510,MK875798,MK977698,MN062719,MN175603,MN444877,MN585980,MN585986,MN586007,MN586042,MN586059,MT657336,MT818415,MW578837,MW601217,MZ150792,MZ322026,MZ622179,MZ648035,ON081330,ON260815,ON260820,ON970563,ON970612,OP068338,OP297543 |
| 435 | VC_1353_0 | 10 | 11003.33853 | 0 | 1 | 0 | 0 | 5.099019514 | 45 | 45 | 1 | 3.582205633 | 0 | 0 | 0 | MH229862,MH229863,MH229864,MH229865,MH536815,MH825706,MN224564,MN224565,ON456338,ON456348 |
| 436 | VC_1354_0 | 3 | 509.5064876 | 8.832975739 | 0.982959091 | 0.040427799 | 0 | 4 | 3 | 3 | 1 | 2.666666667 | 0 | 0 | 0 | MH447526,MK064562,MK064563 |
| 437 | VC_1355_0 | 3 | 25.88353688 | 0 | 1 | 0 | 1.414213562 | 1.414213562 | 3 | 3 | 1 | 1.414213562 | 0 | 0 | 0 | MH572274,MH572275,MH572276 |
| 438 | VC_1356_0 | 8 | 268.2858067 | 39.07361524 | 0.872873215 | 1.78E-05 | 0 | 1.414213562 | 28 | 28 | 1 | 0.353553391 | 0 | 0 | 0 | MH572353,MH572354,MH572355,MH572357,MH572358,MH572407,MH572408,MH572516 |
| 439 | VC_1357_0 | 6 | 141.446599 | 0 | 1 | 0 | 0 | 1 | 15 | 15 | 1 | 0.533333333 | 0 | 0 | 0 | MH572490,MH572491,MH572517,MH572518,MH572519,MH572520 |
| 440 | VC_1358_0 | 5 | 2384.922104 | 0 | 1 | 0 | 0 | 1 | 10 | 10 | 1 | 0.4 | 0 | 0 | 0 | MH707430,MW354669,MW354676,MW354677,MW354678 |
| 441 | VC_1359_0 | 11 | 4623.84576 | 0 | 1 | 1 | 0 | 0 | 55 | 55 | 1 | 0 | 0 | 0 | 0 | MH730557,MZ230992,MZ230993,MZ230994,MZ230995,MZ230996,MZ230997,MZ230998,MZ230999,MZ231000,MZ231001 |
| 442 | VC_135_0 | 2 | 300 | 9638.212953 | 0.030186514 | 0.044425325 | 0 | 0 | 1 | 1 | 1 | 0 | 0 | 0 | 0 | MG592609,MG592621 |
| 443 | VC_135_1 | 2 | 300 | 9735.632693 | 0.029893481 | 0.044423634 | 8.602325267 | 8.602325267 | 1 | 1 | 1 | 8.602325267 | 0 | 0 | 0 | MW824370,MW824375 |
| 444 | VC_1360_0 | 31 | 58834.4525 | 7743.30292 | 0.883695344 | 7.52E-89 | 0 | 7.280109889 | 465 | 465 | 1 | 5.170556738 | 0 | 0 | 0 | MK308638,MK757449,MK878904,MK894435,MK894438,MN310546,MN369747,MT310869,MT310875,MT310887,MT310888,MT310889,MT310890,MT310891,MT498064,MT657334,MT657337,MT684587,MT771344,MW055914,MW291018,MW365947,MW365948,MW507124,MW862991,MZ388554,OM913587,ON081328,ON456346,ON970584,OP297538 |
| 445 | VC_1361_0 | 6 | 2405.982619 | 7743.30292 | 0.237059309 | 6.08E-11 | 0 | 4.358898944 | 15 | 15 | 1 | 2.423545924 | 0 | 0 | 0 | MH825698,MN062711,MT024863,ON260817,ON526980,OP068335 |
| 446 | VC_1362_0 | 6 | 663.0981007 | 0 | 1 | 0 | 1.732050808 | 4.69041576 | 15 | 15 | 1 | 3.580623048 | 0 | 0 | 0 | MH834607,MH834622,MK757448,MT024867,MT024870,MT889364 |
| 447 | VC_1363_0 | 2 | 188.7261751 | 37.93253524 | 0.832644705 | 0.270145687 | 0 | 0 | 1 | 1 | 1 | 0 | 0 | 0 | 0 | MH939157,MK071653 |
| 448 | VC_1364_0 | 3 | 34.64653635 | 0 | 1 | 0 | 0 | 1 | 3 | 3 | 1 | 0.666666667 | 0 | 0 | 0 | MH992154,MK249216,MK249217 |
| 449 | VC_1365_0 | 3 | 43.26213071 | 0 | 1 | 0 | 0 | 1.732050808 | 3 | 3 | 1 | 1.154700538 | 0 | 0 | 0 | MH992155,MH992196,MH992205 |
| 450 | VC_1367_0 | 2 | 15.53109722 | 0 | 1 | 0 | 0 | 0 | 1 | 1 | 1 | 0 | 0 | 0 | 0 | MH992167,MK496682 |
| 451 | VC_1368_0 | 2 | 10.97236665 | 0 | 1 | 0 | 0 | 0 | 1 | 1 | 1 | 0 | 0 | 0 | 0 | MH992171,MW202680 |
| 452 | VC_1369_0 | 3 | 377.4785754 | 7.353353373 | 0.980892039 | 0.040427799 | 1 | 1.414213562 | 3 | 3 | 1 | 1.138071187 | 0 | 0 | 0 | MK305890,MK620900,OP434453 |
| 453 | VC_136_0 | 2 | 300 | 6990.138631 | 0.041151481 | 0.042587315 | 3.741657387 | 3.741657387 | 1 | 1 | 1 | 3.741657387 | 0 | 0 | 0 | MH375644,MT366762 |
| 454 | VC_1370_0 | 2 | 137.3136498 | 29.85354423 | 0.821415055 | 0.087622123 | 6.32455532 | 6.32455532 | 1 | 1 | 1 | 6.32455532 | 0 | 0 | 0 | MK411746,MN586027 |
| 455 | VC_1371_0 | 5 | 63.78333871 | 27.2776659 | 0.700446245 | 0.478116184 | 0 | 1.732050808 | 10 | 10 | 1 | 1.17067423 | 0 | 0 | 0 | MK496679,MK496680,MK496776,OM869578,OM869711 |
| 456 | VC_1372_0 | 2 | 4.936518833 | 22.88520911 | 0.177433941 | 0.263544628 | 1.414213562 | 1.414213562 | 1 | 1 | 1 | 1.414213562 | 0 | 0 | 0 | MK496698,MK496772 |
| 457 | VC_1373_0 | 2 | 29.79354264 | 16.15131422 | 0.648463064 | 0.131776239 | 0 | 0 | 1 | 1 | 1 | 0 | 0 | 0 | 0 | MK496681,MK496760 |
| 458 | VC_1374_0 | 2 | 20.86797703 | 0 | 1 | 0 | 0 | 0 | 1 | 1 | 1 | 0 | 0 | 0 | 0 | MK496683,MK496761 |
| 459 | VC_1375_0 | 2 | 4.33446634 | 0 | 1 | 0 | 0 | 0 | 1 | 1 | 1 | 0 | 0 | 0 | 0 | MK496686,MK496764 |
| 460 | VC_1376_0 | 3 | 36.3442126 | 0 | 1 | 0 | 0 | 1.414213562 | 3 | 3 | 1 | 0.942809042 | 0 | 0 | 0 | MK496688,MK496690,MK496766 |
| 461 | VC_1377_0 | 3 | 8.784913575 | 0 | 1 | 1 | 1 | 1.732050808 | 3 | 3 | 1 | 1.382088123 | 0 | 0 | 0 | MK496689,MK496700,MK496784 |
| 462 | VC_1378_0 | 2 | 4.635491837 | 0 | 1 | 0 | 0 | 0 | 1 | 1 | 1 | 0 | 0 | 0 | 0 | MK496697,MK496780 |
| 463 | VC_1379_0 | 2 | 6.334452844 | 0 | 1 | 0 | 0 | 0 | 1 | 1 | 1 | 0 | 0 | 0 | 0 | MK496699,MK496785 |
| 464 | VC_137_0 | 3 | 355.9573481 | 2795.565648 | 0.112947724 | 0.001453003 | 7.549834435 | 8.366600265 | 3 | 3 | 1 | 7.992897483 | 0 | 0 | 0 | AJ609634,MK448910,OL774867 |
| 465 | VC_137_1 | 1 | 0 | 1199.65636 | 0 | 0 | 0 | 0 | 1 | 1 | 1 | 0 | 0 | 0 | 0 | MK448902 |
| 466 | VC_1380_0 | 2 | 10.97236665 | 0 | 1 | 0 | 0 | 0 | 1 | 1 | 1 | 0 | 0 | 0 | 0 | MK496701,MK496702 |
| 467 | VC_1381_0 | 3 | 7.93983353 | 0 | 1 | 0 | 1 | 1.414213562 | 3 | 3 | 1 | 1.138071187 | 0 | 0 | 0 | MK496710,MK496767,MK496768 |
| 468 | VC_1382_0 | 2 | 11.67133666 | 0 | 1 | 0 | 0 | 0 | 1 | 1 | 1 | 0 | 0 | 0 | 0 | MK496712,MK496787 |
| 469 | VC_1383_0 | 2 | 14.20888126 | 7.93983353 | 0.641521704 | 0.270145687 | 0 | 0 | 1 | 1 | 1 | 0 | 0 | 0 | 0 | MK496716,MK496790 |
| 470 | VC_1384_0 | 2 | 9.319160538 | 0 | 1 | 0 | 0 | 0 | 1 | 1 | 1 | 0 | 0 | 0 | 0 | MK496717,MK496804 |
| 471 | VC_1385_0 | 2 | 25.35975681 | 0 | 1 | 0 | 0 | 0 | 1 | 1 | 1 | 0 | 0 | 0 | 0 | MK496720,MK496796 |
| 472 | VC_1386_0 | 3 | 20.80953851 | 0 | 1 | 1 | 0 | 0 | 3 | 3 | 1 | 0 | 0 | 0 | 0 | MK496721,MK496781,MK496786 |
| 473 | VC_1387_0 | 3 | 30.10425588 | 10.72874778 | 0.737253035 | 0.03826125 | 1 | 1.414213562 | 3 | 3 | 1 | 1.138071187 | 0 | 0 | 0 | MK496725,MK496746,MK496811 |
| 474 | VC_1388_0 | 2 | 14.75294763 | 0 | 1 | 0 | 0 | 0 | 1 | 1 | 1 | 0 | 0 | 0 | 0 | MK496748,MK496815 |
| 475 | VC_1389_0 | 2 | 15.53109722 | 8.969958177 | 0.633895029 | 0.270145687 | 0 | 0 | 1 | 1 | 1 | 0 | 0 | 0 | 0 | MK496750,MK496816 |
| 476 | VC_138_0 | 2 | 112.7686339 | 448.4465939 | 0.200936518 | 0.046310926 | 5.196152423 | 5.196152423 | 1 | 1 | 1 | 5.196152423 | 0 | 0 | 0 | DQ340064,DQ840344 |
| 477 | VC_1390_0 | 2 | 16.30924847 | 0 | 1 | 0 | 0 | 0 | 1 | 1 | 1 | 0 | 0 | 0 | 0 | MK496753,MK496818 |
| 478 | VC_1391_0 | 2 | 9.796278594 | 0 | 1 | 0 | 1.414213562 | 1.414213562 | 1 | 1 | 1 | 1.414213562 | 0 | 0 | 0 | MK496756,MK496824 |
| 479 | VC_1392_0 | 2 | 14.75294763 | 0 | 1 | 0 | 0 | 0 | 1 | 1 | 1 | 0 | 0 | 0 | 0 | MK496758,MK496808 |
| 480 | VC_1393_0 | 2 | 16.30924847 | 0 | 1 | 0 | 0 | 0 | 1 | 1 | 1 | 0 | 0 | 0 | 0 | MK496770,MK496771 |
| 481 | VC_1394_0 | 2 | 3.887317306 | 0 | 1 | 0 | 0 | 0 | 1 | 1 | 1 | 0 | 0 | 0 | 0 | MK496820,OM869632 |
| 482 | VC_1395_0 | 3 | 502.6487421 | 144.9856775 | 0.776130371 | 0.036099099 | 0 | 2.236067977 | 3 | 3 | 1 | 1.490711985 | 0 | 0 | 0 | MK570053,MK570054,MK570055 |
| 483 | VC_1396_0 | 110 | 1770844.614 | 0 | 1 | 0 | 0 | 10.24695077 | 5995 | 5843 | 0.974645538 | 4.936735917 | 0 | 0 | 0 | MK719701,MK719702,MK719703,MK719704,MK719705,MK719706,MK719707,MK719708,MK719709,MK719710,MK719711,MK719712,MK719713,MK719714,MK719715,MK719716,MK719717,MK719718,MK719719,MK719720,MK719721,MK719722,MK719723,MK719724,MK719725,MK719726,MK719727,MK719728,MK719729,MK719730,MK719731,MK719732,MK719733,MK719734,MK719735,MK719736,MK719737,MK719738,MK719739,MK719740,MK719741,MK719742,MK719743,MK719744,MK719745,MK719746,MK719747,MK719748,MK719749,MK719750,MK719751,MK719752,MK719753,MK719754,MT497224,MT497225,MT497226,MT497227,MT497228,MT497229,MT497230,MT497231,MT497232,MT497233,MT497234,MT497235,MT497236,MT497237,MT497238,MT497239,MT497240,MT497241,MT497242,MT497243,MT497244,MT497245,MT497246,MT497247,MT497248,MT497249,MT497250,MT497251,MT497252,MT497253,MT497254,MT497255,MT497256,MT497257,MT497258,MT497259,MT497260,MT497261,MT497262,MT497263,MT497264,MT497265,MT497266,MT497267,MT497268,MT497269,MT497270,MT497271,MT497272,MT497273,MT497274,MT497275,MT497276,MT497277,MT497278,MT497279 |
| 484 | VC_1397_0 | 2 | 165.0302466 | 73.96689844 | 0.690511372 | 0.065366293 | 8.124038405 | 8.124038405 | 1 | 1 | 1 | 8.124038405 | 0 | 0 | 0 | S11-wms-combined_73204,S12-wms-combined_320727 |
| 485 | VC_1398_0 | 2 | 20.02287899 | 0 | 1 | 0 | 0 | 0 | 1 | 1 | 1 | 0 | 0 | 0 | 0 | MK765551,MK765650 |
| 486 | VC_1399_0 | 9 | 420.1681197 | 0 | 1 | 1 | 0 | 0 | 36 | 36 | 1 | 0 | 0 | 0 | 0 | MK765558,MK765581,MK765604,MK765622,MK765640,MK765654,MK765656,MK765663,MK765668 |
| 487 | VC_139_0 | 2 | 186.1419215 | 2467.976517 | 0.070133239 | 0.044898152 | 6.480740698 | 6.480740698 | 1 | 1 | 1 | 6.480740698 | 0 | 0 | 0 | AY855346,LK985322 |
| 488 | VC_139_1 | 4 | 1073.274386 | 4973.384526 | 0.177498748 | 1.39E-05 | 3.605551275 | 8.426149773 | 6 | 6 | 1 | 6.979687734 | 0 | 0 | 0 | DQ466086,HG531805,LN681541,LN681542 |
| 489 | VC_1400_0 | 2 | 9.319160538 | 0 | 1 | 0 | 0 | 0 | 1 | 1 | 1 | 0 | 0 | 0 | 0 | MK765573,MK765662 |
| 490 | VC_1401_0 | 2 | 10.97236665 | 0 | 1 | 0 | 0 | 0 | 1 | 1 | 1 | 0 | 0 | 0 | 0 | MK765575,MK765578 |
| 491 | VC_1402_0 | 14 | 412.1442703 | 2051.859504 | 0.167266087 | 0.486096463 | 0 | 2.645751311 | 91 | 91 | 1 | 1.990900852 | 0 | 0 | 0 | MK765587,MK765588,MK765598,MK765599,MK765623,MK765625,MK765628,MK765630,MK765631,MK765634,OL617020,OL617024,OL617026,OM782324 |
| 492 | VC_1403_0 | 28 | 8196.936363 | 2688.272467 | 0.753034369 | 2.07E-168 | 0 | 2.236067977 | 378 | 378 | 1 | 0.666085152 | 0 | 0 | 0 | MK765589,MN988460,MN988467,MN988469,MN988471,MN988473,MN988474,MN988475,MN988478,MN988479,MN988480,MN988481,MN988489,MN988491,MN988494,MN988498,MN988499,MN988503,MN988504,MN988511,MN988518,MW960029,MW960035,MW980067,OL617021,OL617022,OL617025,OL617027 |
| 493 | VC_1404_0 | 1 | 0 | 51.76694088 | 0 | 0 | 0 | 0 | 1 | 1 | 1 | 0 | 0 | 0 | 0 | MK765626 |
| 494 | VC_1406_0 | 2 | 10.27339825 | 0 | 1 | 0 | 0 | 0 | 1 | 1 | 1 | 0 | 0 | 0 | 0 | MK765605,MK765641 |
| 495 | VC_1407_0 | 2 | 20.34523526 | 4.540216016 | 0.817555408 | 0.185546685 | 8.306623863 | 8.306623863 | 1 | 1 | 1 | 8.306623863 | 0 | 0 | 0 | MK929790,MW202545 |
| 496 | VC_1408_0 | 2 | 10.51240781 | 51.50282377 | 0.16951332 | 0.401293674 | 8.94427191 | 8.94427191 | 1 | 1 | 1 | 8.94427191 | 0 | 0 | 0 | S1-wms-combined_197401,S17-wms-combined_162786 |
| 497 | VC_1408_1 | 1 | 0 | 53.04348343 | 0 | 0 | 0 | 0 | 1 | 1 | 1 | 0 | 0 | 0 | 0 | S1-wms-combined_220022 |
| 498 | VC_1409_0 | 2 | 13.23885114 | 0 | 1 | 0 | 0 | 0 | 1 | 1 | 1 | 0 | 0 | 0 | 0 | MN379629,MN379636 |
| 499 | VC_140_0 | 2 | 217.5902156 | 311.6650836 | 0.411125247 | 0.051752086 | 4.472135955 | 4.472135955 | 1 | 1 | 1 | 4.472135955 | 0 | 0 | 0 | DQ003640,MH341453 |
| 500 | VC_1410_0 | 2 | 20.02287899 | 0 | 1 | 0 | 0 | 0 | 1 | 1 | 1 | 0 | 0 | 0 | 0 | MN379639,MN379646 |
| 501 | VC_1411_0 | 2 | 14.75294763 | 0 | 1 | 0 | 0 | 0 | 1 | 1 | 1 | 0 | 0 | 0 | 0 | MN379643,MN379650 |
| 502 | VC_1413_0 | 2 | 171.3104661 | 80.74987012 | 0.679640711 | 0.105649774 | 0 | 0 | 1 | 1 | 1 | 0 | 0 | 0 | 0 | MN882551,MN882552 |
| 503 | VC_1414_0 | 4 | 1800 | 0 | 1 | 1 | 0 | 1.732050808 | 6 | 6 | 1 | 1.093412989 | 0 | 0 | 0 | MN988461,MN988462,MN988463,MN988472 |
| 504 | VC_1415_0 | 2 | 15.53109722 | 0 | 1 | 0 | 0 | 0 | 1 | 1 | 1 | 0 | 0 | 0 | 0 | MN988485,MN988550 |
| 505 | VC_1416_0 | 5 | 3000 | 13.62742601 | 0.995478065 | 0.000116471 | 0 | 6.480740698 | 10 | 10 | 1 | 4.659164571 | 0 | 0 | 0 | MN988515,MN988535,MN988540,MW980063,MW980064 |
| 506 | VC_1417_0 | 15 | 22851.94544 | 40.83131706 | 0.99821641 | 2.09E-10 | 0 | 6.403124237 | 105 | 105 | 1 | 4.041789466 | 0 | 0 | 0 | MT732458,MT732459,MT732460,MT732461,MT732462,MT732463,MT732464,MT732465,MT732466,MT732467,MT732468,MT732469,MT732470,MT732471,MT732472 |
| 507 | VC_1418_0 | 2 | 141.7431816 | 0 | 1 | 0 | 0 | 0 | 1 | 1 | 1 | 0 | 0 | 0 | 0 | MT006234,MT006235 |
| 508 | VC_1419_0 | 6 | 4500 | 215.6728999 | 0.954264661 | 6.96E-06 | 0 | 7.071067812 | 15 | 15 | 1 | 4.628726661 | 0 | 0 | 0 | MT074137,MT074139,MT074140,MT074141,MT074142,MT074146 |
| 509 | VC_141_0 | 6 | 3383.244073 | 2256.676936 | 0.599874372 | 4.94E-11 | 0 | 8 | 15 | 15 | 1 | 5.275870066 | 0 | 0 | 0 | KC595514,KC595515,KC595517,KC595518,KT151956,KT151958 |
| 510 | VC_1420_0 | 5 | 1193.292455 | 0 | 1 | 0 | 2.645751311 | 8 | 10 | 10 | 1 | 6.044250934 | 0 | 0 | 0 | MT121960,MT121964,S12-wms_401372,S2-wms-combined_197577,S2-wms_128831 |
| 511 | VC_1421_0 | 4 | 59.41518686 | 0 | 1 | 0 | 0 | 1.732050808 | 6 | 6 | 1 | 1.093412989 | 0 | 0 | 0 | MT135262,MT135274,MT135389,MT135413 |
| 512 | VC_1422_0 | 2 | 28.13437958 | 0 | 1 | 0 | 1 | 1 | 1 | 1 | 1 | 1 | 0 | 0 | 0 | MT135284,S2-wms-combined_132966 |
| 513 | VC_1423_0 | 8 | 524.5818436 | 60.89151326 | 0.895996099 | 0.000160684 | 0 | 2.828427125 | 28 | 28 | 1 | 1.633823161 | 0 | 0 | 0 | MT135285,MT135286,MT135289,MT135297,MT135302,MT135308,MT135314,S9-wms_104169 |
| 514 | VC_1424_0 | 8 | 514.7654063 | 141.6190616 | 0.784243734 | 5.40E-13 | 0 | 3.605551275 | 28 | 28 | 1 | 2.567446955 | 0 | 0 | 0 | MT135288,MT135290,MT135291,MT135301,MT135303,MT135307,MT135309,MT135311 |
| 515 | VC_1425_0 | 7 | 309.0508475 | 133.9381279 | 0.697649072 | 1.45E-08 | 0 | 4 | 21 | 21 | 1 | 2.664121186 | 0 | 0 | 0 | MT135287,MT135294,MT135298,MT135299,MT135304,MT135306,MT135310 |
| 516 | VC_1426_0 | 4 | 103.7350176 | 0 | 1 | 0 | 0 | 1.414213562 | 6 | 6 | 1 | 0.971404521 | 0 | 0 | 0 | MT135292,MT135293,MT135296,MT135305 |
| 517 | VC_1427_0 | 2 | 5.334455843 | 0 | 1 | 0 | 1 | 1 | 1 | 1 | 1 | 1 | 0 | 0 | 0 | MT135332,MT135436 |
| 518 | VC_1428_0 | 2 | 7.981166171 | 0 | 1 | 0 | 0 | 0 | 1 | 1 | 1 | 0 | 0 | 0 | 0 | MT135346,MT135395 |
| 519 | VC_1429_0 | 8 | 279.6760993 | 8.221783646 | 0.971442014 | 0.05482919 | 1 | 2.828427125 | 28 | 28 | 1 | 1.983155984 | 0 | 0 | 0 | MT135359,MT135368,MT135374,MT135378,MT135399,MT135414,MT135419,MT135463 |
| 520 | VC_142_0 | 2 | 251.0442322 | 295.0561832 | 0.459703427 | 0.057268646 | 0 | 0 | 1 | 1 | 1 | 0 | 0 | 0 | 0 | KM983330,KM983333 |
| 521 | VC_1430_0 | 4 | 36.04712246 | 0 | 1 | 0 | 1.414213562 | 2 | 6 | 6 | 1 | 1.670763258 | 0 | 0 | 0 | MT135369,MT135396,MT135454,MT135460 |
| 522 | VC_1431_0 | 3 | 28.18227414 | 0 | 1 | 0 | 0 | 1.414213562 | 3 | 3 | 1 | 0.942809042 | 0 | 0 | 0 | MT135375,MT135438,MT135453 |
| 523 | VC_1433_0 | 2 | 300 | 0 | 1 | 0 | 5.477225575 | 5.477225575 | 1 | 1 | 1 | 5.477225575 | 0 | 0 | 0 | MT234670,ON014754 |
| 524 | VC_1434_0 | 2 | 273.4640231 | 39.3646605 | 0.874165438 | 0.144422183 | 1.414213562 | 1.414213562 | 1 | 1 | 1 | 1.414213562 | 0 | 0 | 0 | ON529863,ON529867 |
| 525 | VC_1435_0 | 9 | 874.6348922 | 387.3481794 | 0.693063886 | 5.34E-08 | 4.123105626 | 8 | 36 | 36 | 1 | 6.481341919 | 0 | 0 | 0 | S17-wms-combined_16309,S17-wms_124308,S2-wms_156439,S7-wms-combined_185016,S7-wms-combined_90393,S7-wms-combined_90728,S7-wms_123954,S9-wms-combined_33229,S9-wms-combined_33230 |
| 526 | VC_1435_1 | 1 | 0 | 183.8748403 | 0 | 0 | 0 | 0 | 1 | 1 | 1 | 0 | 0 | 0 | 0 | S6-wms-combined_82597 |
| 527 | VC_1436_0 | 4 | 46.74873652 | 35.12833821 | 0.570962466 | 0.03630047 | 3.872983346 | 6.480740698 | 6 | 6 | 1 | 5.708073768 | 0 | 0 | 0 | S12-wms-combined_351847,S12-wms_383851,S2-wms-combined_58121,S7-wms-combined_239784 |
| 528 | VC_1437_0 | 3 | 250.8216948 | 87.23509648 | 0.741951356 | 0.008115024 | 5.291502622 | 6.32455532 | 3 | 3 | 1 | 5.926823982 | 0 | 0 | 0 | S1-wms-combined_263562,S5-wms-combined_207291,S7-wms-combined_104872 |
| 529 | VC_1438_0 | 6 | 4500 | 0 | 1 | 1 | 0 | 3.605551275 | 15 | 15 | 1 | 1.860050056 | 0 | 0 | 0 | MT732450,MT732451,MT732452,MT732453,MT732454,MT732455 |
| 530 | VC_1439_0 | 3 | 38.36357799 | 47.62584872 | 0.446142967 | 0.012635581 | 5.830951895 | 7.141428429 | 3 | 3 | 1 | 6.560194752 | 0 | 0 | 0 | S12-wms-combined_260036,S12-wms-combined_91231,S4-wms_45128 |
| 531 | VC_143_0 | 2 | 39.90191747 | 295.7747569 | 0.118870093 | 0.05004329 | 6.782329983 | 6.782329983 | 1 | 1 | 1 | 6.782329983 | 0 | 0 | 0 | KT626446,ON699007 |
| 532 | VC_1440_0 | 4 | 663.5497664 | 0 | 1 | 0 | 0 | 1 | 6 | 6 | 1 | 0.666666667 | 0 | 0 | 0 | MT952848,MZ150790,OL455892,OP068334 |
| 533 | VC_1441_0 | 2 | 25.35975681 | 0 | 1 | 0 | 0 | 0 | 1 | 1 | 1 | 0 | 0 | 0 | 0 | MT975991,OP433453 |
| 534 | VC_1442_0 | 2 | 256.5191023 | 8.356398473 | 0.968451599 | 0.131776239 | 0 | 0 | 1 | 1 | 1 | 0 | 0 | 0 | 0 | MW202613,MW202748 |
| 535 | VC_1443_0 | 2 | 194.062967 | 31.29142199 | 0.861145718 | 0.239750061 | 0 | 0 | 1 | 1 | 1 | 0 | 0 | 0 | 0 | MW202637,MW202789 |
| 536 | VC_1444_0 | 3 | 89.38062792 | 0 | 1 | 1 | 0 | 0 | 3 | 3 | 1 | 0 | 0 | 0 | 0 | MW202486,MW202591,MW202766 |
| 537 | VC_1445_0 | 4 | 1266.227393 | 14.99889595 | 0.988293328 | 0.003842984 | 1 | 1.414213562 | 6 | 6 | 1 | 1.138071187 | 0 | 0 | 0 | MW202572,MW202631,MW202645,MW202742 |
| 538 | VC_1446_0 | 3 | 721.8807503 | 0 | 1 | 1 | 0 | 0 | 3 | 3 | 1 | 0 | 0 | 0 | 0 | MW202524,MW202718,MW202743 |
| 539 | VC_1447_0 | 2 | 244.0422632 | 0 | 1 | 0 | 0 | 0 | 1 | 1 | 1 | 0 | 0 | 0 | 0 | MW202601,MW202604 |
| 540 | VC_1448_0 | 5 | 2085.645793 | 0 | 1 | 0 | 0 | 1 | 10 | 10 | 1 | 0.6 | 0 | 0 | 0 | MW202608,MW202725,MW202812,MW202826,MW202842 |
| 541 | VC_1449_0 | 2 | 197.6921866 | 0 | 1 | 0 | 0 | 0 | 1 | 1 | 1 | 0 | 0 | 0 | 0 | MW202622,MW202658 |
| 542 | VC_144_0 | 2 | 268.7364357 | 365.5521083 | 0.42368168 | 0.052317582 | 1 | 1 | 1 | 1 | 1 | 1 | 0 | 0 | 0 | MN604230,MN604698 |
| 543 | VC_1450_0 | 3 | 23.13612536 | 0 | 1 | 0 | 0 | 1 | 3 | 3 | 1 | 0.666666667 | 0 | 0 | 0 | MW202642,MW202843,MW202852 |
| 544 | VC_1451_0 | 2 | 265.2030535 | 0 | 1 | 0 | 0 | 0 | 1 | 1 | 1 | 0 | 0 | 0 | 0 | MW202657,MW202864 |
| 545 | VC_1452_0 | 2 | 66.1264962 | 0 | 1 | 0 | 5 | 5 | 1 | 1 | 1 | 5 | 0 | 0 | 0 | MW202753,MW202825 |
| 546 | VC_1453_0 | 7 | 1522.810025 | 147.0001386 | 0.911965958 | 0.000486588 | 0 | 5.196152423 | 21 | 21 | 1 | 3.416689082 | 0 | 0 | 0 | MW353166,MW353167,MW353168,MW353169,MW353170,MW353171,MW353172 |
| 547 | VC_1454_0 | 2 | 33.93969256 | 14.98865778 | 0.693661085 | 0.144422183 | 9 | 9 | 1 | 0 | 0 | 9 | 0 | 0 | 0 | MW462221,MZ047271 |
| 548 | VC_1456_0 | 4 | 551.7060882 | 349.6011421 | 0.612117677 | 0.000122565 | 5.477225575 | 8.717797887 | 6 | 6 | 1 | 7.206631841 | 0 | 0 | 0 | S11-wms-combined_125176,S12-wms_386692,S6-wms-combined_91039,S7-wms-combined_14267 |
| 549 | VC_1457_0 | 2 | 11.67133666 | 0 | 1 | 0 | 0 | 0 | 1 | 1 | 1 | 0 | 0 | 0 | 0 | MZ089747,MZ089748 |
| 550 | VC_1458_0 | 2 | 4.392456787 | 0 | 1 | 0 | 1 | 1 | 1 | 1 | 1 | 1 | 0 | 0 | 0 | MZ089769,OM869588 |
| 551 | VC_1459_0 | 2 | 17.49655 | 0 | 1 | 0 | 0 | 0 | 1 | 1 | 1 | 0 | 0 | 0 | 0 | MZ089783,OM869706 |
| 552 | VC_1460_0 | 3 | 41.32207156 | 11.34108823 | 0.784648542 | 0.03826125 | 0 | 1.732050808 | 3 | 3 | 1 | 1.154700538 | 0 | 0 | 0 | MZ089787,MZ089788,OM869708 |
| 553 | VC_1461_0 | 3 | 47.37144291 | 0 | 1 | 0 | 0 | 0 | 3 | 3 | 1 | 0 | 0 | 0 | 0 | MZ089790,MZ089792,MZ089794 |
| 554 | VC_1462_0 | 4 | 73.07202371 | 0 | 1 | 0 | 0 | 1 | 6 | 6 | 1 | 0.5 | 0 | 0 | 0 | MZ089811,OM869593,OM869669,OM869712 |
| 555 | VC_1463_0 | 6 | 126.1913745 | 0 | 1 | 0 | 0 | 1 | 15 | 15 | 1 | 0.333333333 | 0 | 0 | 0 | MZ364235,MZ364236,MZ364238,MZ364239,MZ364257,MZ364289 |
| 556 | VC_1464_0 | 3 | 23.42188031 | 0 | 1 | 0 | 0 | 2 | 3 | 3 | 1 | 1.333333333 | 0 | 0 | 0 | MZ364251,MZ364282,MZ364294 |
| 557 | VC_1465_0 | 2 | 208.5298712 | 0 | 1 | 0 | 0 | 0 | 1 | 1 | 1 | 0 | 0 | 0 | 0 | MZ447863,MZ570427 |
| 558 | VC_1466_0 | 2 | 2.262782824 | 0 | 1 | 0 | 2.449489743 | 2.449489743 | 1 | 1 | 1 | 2.449489743 | 0 | 0 | 0 | MZ474488,MZ474489 |
| 559 | VC_1467_0 | 3 | 162.2436747 | 0 | 1 | 0 | 2.645751311 | 3.16227766 | 3 | 3 | 1 | 2.817926761 | 0 | 0 | 0 | OL689226,OL689227,OL689230 |
| 560 | VC_1468_0 | 2 | 4.33446634 | 0 | 1 | 0 | 0 | 0 | 1 | 1 | 1 | 0 | 0 | 0 | 0 | OM869510,OM869512 |
| 561 | VC_1469_0 | 2 | 20.86797703 | 0 | 1 | 0 | 0 | 0 | 1 | 1 | 1 | 0 | 0 | 0 | 0 | OM869572,OM869573 |
| 562 | VC_1470_0 | 3 | 5.276002318 | 0 | 1 | 0 | 0 | 0 | 3 | 3 | 1 | 0 | 0 | 0 | 0 | OM869583,OM869672,OM869688 |
| 563 | VC_1471_0 | 2 | 14.20888126 | 0 | 1 | 0 | 0 | 0 | 1 | 1 | 1 | 0 | 0 | 0 | 0 | OM869594,OM869713 |
| 564 | VC_1472_0 | 2 | 2.490056751 | 0 | 1 | 0 | 6.32455532 | 6.32455532 | 1 | 1 | 1 | 6.32455532 | 0 | 0 | 0 | ON014753,ON014757 |
| 565 | VC_1473_0 | 3 | 222.4261317 | 84.92048884 | 0.723697991 | 0.003868342 | 7.810249676 | 9 | 3 | 2 | 0.666666667 | 8.31142936 | 0 | 0 | 0 | S1-wms_244836,S2-wms-combined_229529,S9-wms-combined_37587 |
| 566 | VC_1474_0 | 3 | 900 | 0 | 1 | 1 | 2 | 3 | 3 | 3 | 1 | 2.666666667 | 0 | 0 | 0 | ON464759,ON464763,ON464764 |
| 567 | VC_1475_0 | 2 | 300 | 0 | 1 | 0 | 1.732050808 | 1.732050808 | 1 | 1 | 1 | 1.732050808 | 0 | 0 | 0 | ON529865,ON529866 |

| | | | | | | | | | | | | | | | | |
|---|---|---|---|---|---|---|---|---|---|---|---|---|---|---|---|---|
| 568 | VC_1476_0 | 2 | 16.30924847 | 0 | 1 | 0 | 0 | 0 | 1 | 1 | 1 | 0 | 0 | 0 | 0 | OP549878,OP549879 |
| 569 | VC_1477_0 | 3 | 102.2494938 | 0 | 1 | 0 | 1.732050808 | 3.16227766 | 3 | 3 | 1 | 2.631442823 | 0 | 0 | 0 | OP549880,S4-wms_161443,S7-wms_157264 |
| 570 | VC_1478_0 | 2 | 29.79354264 | 0 | 1 | 0 | 0 | 0 | 1 | 1 | 1 | 0 | 0 | 0 | 0 | PF3COMN,PF3COMNY |
| 571 | VC_1479_0 | 2 | 205.3385226 | 214.8549051 | 0.488676188 | 0.061538965 | 7.549834435 | 7.549834435 | 1 | 1 | 1 | 7.549834435 | 0 | 0 | 0 | S12-wms-combined_51076,S5-wms-combined_78434 |
| 572 | VC_1481_0 | 9 | 152.8826248 | 9.479170006 | 0.941616992 | 0.220826911 | 1.414213562 | 3.464101615 | 36 | 36 | 1 | 2.481661244 | 0 | 0 | 0 | S1-wms-combined_120533,S1-wms-combined_331896,S12-wms-combined_393703,S17-wms-combined_126600,S2-wms-combined_181095,S5-wms_52566,S8-wms-combined_234473,S8-wms-combined_264711,S9-wms-combined_56773 |
| 573 | VC_1482_0 | 2 | 9.152822718 | 16.61800529 | 0.355162151 | 0.144422183 | 1 | 1 | 1 | 1 | 1 | 1 | 0 | 0 | 0 | S17-wms-combined_30308,S4-wms-combined_143879 |
| 574 | VC_1483_0 | 1 | 0 | 59.24370107 | 0 | 0 | 0 | 0 | 1 | 1 | 1 | 0 | 0 | 0 | 0 | S7-wms-combined_169375 |
| 575 | VC_1483_1 | 1 | 0 | 45.35844511 | 0 | 0 | 0 | 0 | 1 | 1 | 1 | 0 | 0 | 0 | 0 | S8-wms-combined_40666 |
| 576 | VC_1484_0 | 10 | 1043.57682 | 464.9838744 | 0.69176986 | 2.08E-08 | 2.236067977 | 10.58300524 | 45 | 40 | 0.888888889 | 7.073707412 | 0 | 0 | 0 | S1-wms-combined_151708,S1-wms-combined_164485,S12-wms-combined_329242,S17-wms-combined_126774,S17-wms-combined_146850,S17-wms-combined_157468,S2-wms-combined_4685,S2-wms-combined_6605,S6-wms-combined_41246,S7-wms-combined_84688 |
| 577 | VC_1485_0 | 1 | 0 | 44.10957618 | 0 | 0 | 0 | 0 | 1 | 1 | 1 | 0 | 0 | 0 | 0 | S2-wms-combined_233107 |
| 578 | VC_1487_0 | 2 | 6.557034017 | 0 | 1 | 0 | 4.472135955 | 4.472135955 | 1 | 1 | 1 | 4.472135955 | 0 | 0 | 0 | S1-wms-combined_156637,S12-wms-combined_210942 |
| 579 | VC_1489_0 | 1 | 0 | 5.515472155 | 0 | 0 | 0 | 0 | 1 | 1 | 1 | 0 | 0 | 0 | 0 | S12-wms-combined_157167 |
| 580 | VC_148_0 | 2 | 170.0183889 | 662.0139995 | 0.20434107 | 0.046334615 | 6.244997998 | 6.244997998 | 1 | 1 | 1 | 6.244997998 | 0 | 0 | 0 | S2-wms-combined_62519,S8-wms-combined_161181 |
| 581 | VC_148_1 | 3 | 417.1453056 | 623.6087881 | 0.400810632 | 0.002477882 | 5.567764363 | 7.745966692 | 3 | 3 | 1 | 6.572285098 | 0 | 0 | 0 | S2-wms-combined_218002,S6-wms-combined_119138,S6-wms-combined_32530 |
| 582 | VC_1491_0 | 2 | 5.635482839 | 0 | 1 | 0 | 1 | 1 | 1 | 1 | 1 | 1 | 0 | 0 | 0 | S1-wms-combined_185336,S1-wms-combined_40115 |
| 583 | VC_1492_0 | 7 | 1518.837228 | 555.5451018 | 0.7321877 | 5.64E-11 | 3.872983346 | 9.539392014 | 21 | 18 | 0.857142857 | 7.746739836 | 0 | 0 | 0 | S1-wms_100010,S1-wms_225995,S12-wms-combined_264425,S12-wms-combined_299293,S12-wms_179388,S12-wms_401366,S12-wms_44817 |
| 584 | VC_1494_0 | 2 | 30.24608585 | 1.463968726 | 0.953832665 | 0.5 | 1.732050808 | 1.732050808 | 1 | 1 | 1 | 1.732050808 | 0 | 0 | 0 | S1-wms-combined_248622,S17-wms-combined_70983 |
| 585 | VC_1495_0 | 3 | 93.82698037 | 38.09746008 | 0.711217573 | 0.006365682 | 5 | 7.810249676 | 3 | 3 | 1 | 6.764521483 | 0 | 0 | 0 | S17-wms-combined_233761,S2-wms-combined_210822,S4-wms-combined_47565 |
| 586 | VC_1496_0 | 5 | 130.1356474 | 10.89296783 | 0.922760584 | 0.011247136 | 2 | 3.605551275 | 10 | 10 | 1 | 2.883076275 | 0 | 0 | 0 | S1-wms-combined_313851,S1-wms_313991,S2-wms_110761,S2-wms_23562,S6-wms_137591 |
| 587 | VC_1497_0 | 3 | 210.1504315 | 0 | 1 | 0 | 3 | 4.358898944 | 3 | 3 | 1 | 3.867179877 | 0 | 0 | 0 | S1-wms-combined_331544,S4-wms-combined_51079,S7-wms-combined_136642 |
| 588 | VC_1498_0 | 2 | 33.56623516 | 3.194404025 | 0.913102598 | 0.5 | 4.472135955 | 4.472135955 | 1 | 1 | 1 | 4.472135955 | 0 | 0 | 0 | S1-wms-combined_354899,S2-wms-combined_248318 |
| 589 | VC_1499_0 | 2 | 1.467479827 | 3.194404025 | 0.31478258 | 0.5 | 6.782329983 | 6.782329983 | 1 | 1 | 1 | 6.782329983 | 0 | 0 | 0 | S5-wms-combined_66017,S5-wms_210460 |
| 590 | VC_149_0 | 5 | 3000 | 103889.1795 | 0.028066452 | 1.03E-06 | 0 | 1.732050808 | 10 | 10 | 1 | 0.897469149 | 0 | 0 | 0 | MT080592,MT080597,MT080598,MT080599,MT080603 |
| 591 | VC_149_1 | 3 | 900 | 69214.93003 | 0.012836068 | 0.006960472 | 1 | 1.414213562 | 3 | 3 | 1 | 1.138071187 | 0 | 0 | 0 | OM850391,OM850392,ON113344 |
| 592 | VC_149_10 | 1 | 0 | 35853.22012 | 0 | 0 | 0 | 0 | 1 | 1 | 1 | 0 | 0 | 0 | 0 | KY794641 |
| 593 | VC_149_11 | 4 | 1800 | 86765.56562 | 0.020323926 | 0.000164304 | 0 | 8.94427191 | 6 | 6 | 1 | 4.906036795 | 0 | 0 | 0 | MT080582,MT080583,MT080586,MT080594 |
| 594 | VC_149_12 | 2 | 300 | 67267.03433 | 0.004440035 | 0.168432916 | 7.874007874 | 7.874007874 | 1 | 1 | 1 | 7.874007874 | 0 | 0 | 0 | MH107769,MN045228 |
| 595 | VC_149_13 | 3 | 900 | 102060.8519 | 0.008741186 | 0.048210974 | 7.348469228 | 8.888194417 | 3 | 3 | 1 | 8.220937806 | 0 | 0 | 0 | AB853330,JX878671,LR215719 |
| 596 | VC_149_14 | 1 | 0 | 35285.51482 | 0 | 0 | 0 | 0 | 1 | 1 | 1 | 0 | 0 | 0 | 0 | AB903967 |
| 597 | VC_149_15 | 16 | 36000 | 421023.6356 | 0.078770543 | 1.24E-67 | 0 | 10.77032961 | 120 | 97 | 0.808333333 | 7.02767147 | 0 | 0 | 0 | JQ686190,KM216423,KY779849,LC574321,LR215718,MF001365,MG557618,MH159197,MK250904,MN304941,MT786458,MW546073,MZ427930,MZ573923,OP235318,OP323059 |
| 598 | VC_149_16 | 1 | 0 | 33802.20141 | 0 | 0 | 0 | 0 | 1 | 1 | 1 | 0 | 0 | 0 | 0 | KJ888149 |
| 599 | VC_149_17 | 4 | 1800 | 136436.969 | 0.013021119 | 0.000481854 | 0 | 2 | 6 | 6 | 1 | 1.333333333 | 0 | 0 | 0 | MT080585,MT080589,MT080591,MT080601 |
| 600 | VC_149_18 | 1 | 0 | 34926.54481 | 0 | 0 | 0 | 0 | 1 | 1 | 1 | 0 | 0 | 0 | 0 | MT554104 |
| 601 | VC_149_19 | 1 | 0 | 34414.99466 | 0 | 0 | 0 | 0 | 1 | 1 | 1 | 0 | 0 | 0 | 0 | OP263968 |
| 602 | VC_149_2 | 1 | 0 | 24781.52512 | 0 | 0 | 0 | 0 | 1 | 1 | 1 | 0 | 0 | 0 | 0 | LR215720 |
| 603 | VC_149_20 | 1 | 0 | 33796.75631 | 0 | 0 | 0 | 0 | 1 | 1 | 1 | 0 | 0 | 0 | 0 | KP027446 |
| 604 | VC_149_21 | 1 | 0 | 38022.37521 | 0 | 0 | 0 | 0 | 1 | 1 | 1 | 0 | 0 | 0 | 0 | MG721208 |
| 605 | VC_149_22 | 2 | 300 | 47850.66213 | 0.006230444 | 0.070834384 | 1 | 1 | 1 | 1 | 1 | 1 | 0 | 0 | 0 | MT080593,MT080596 |
| 606 | VC_149_23 | 1 | 0 | 32659.35798 | 0 | 0 | 0 | 0 | 1 | 1 | 1 | 0 | 0 | 0 | 0 | AB853331 |
| 607 | VC_149_24 | 1 | 0 | 23304.24333 | 0 | 0 | 0 | 0 | 1 | 1 | 1 | 0 | 0 | 0 | 0 | MZ417322 |
| 608 | VC_149_25 | 14 | 27300 | 259934.8427 | 0.09504418 | 5.46E-47 | 2.449489743 | 10.95445115 | 91 | 80 | 0.879120879 | 7.006047118 | 0 | 0 | 0 | MZ417315,MZ417319,MZ417320,MZ417321,MZ417327,MZ417328,MZ417332,MZ417337,MZ417340,MZ417345,MZ417346,MZ417347,MZ417350,MZ417354 |
| 609 | VC_149_26 | 1 | 0 | 23636.88817 | 0 | 0 | 0 | 0 | 1 | 1 | 1 | 0 | 0 | 0 | 0 | MZ417331 |
| 610 | VC_149_27 | 1 | 0 | 17406.87461 | 0 | 0 | 0 | 0 | 1 | 1 | 1 | 0 | 0 | 0 | 0 | KX011028 |
| 611 | VC_149_28 | 1 | 0 | 21591.38333 | 0 | 0 | 0 | 0 | 1 | 1 | 1 | 0 | 0 | 0 | 0 | MN935200 |
| 612 | VC_149_29 | 3 | 900 | 73914.6509 | 0.012029729 | 0.005514229 | 6.403124237 | 8.246211251 | 3 | 3 | 1 | 7.528863141 | 0 | 0 | 0 | KY794642,KY794643,LR215721 |
| 613 | VC_149_3 | 1 | 0 | 23798.4585 | 0 | 0 | 0 | 0 | 1 | 1 | 1 | 0 | 0 | 0 | 0 | MT787017 |
| 614 | VC_149_4 | 1 | 0 | 24237.99408 | 0 | 0 | 0 | 0 | 1 | 1 | 1 | 0 | 0 | 0 | 0 | OL741057 |
| 615 | VC_149_5 | 1 | 0 | 25353.63292 | 0 | 0 | 0 | 0 | 1 | 1 | 1 | 0 | 0 | 0 | 0 | MN091625 |
| 616 | VC_149_6 | 1 | 0 | 26787.2931 | 0 | 0 | 0 | 0 | 1 | 1 | 1 | 0 | 0 | 0 | 0 | OP297178 |
| 617 | VC_149_7 | 1 | 0 | 23354.53964 | 0 | 0 | 0 | 0 | 1 | 1 | 1 | 0 | 0 | 0 | 0 | LC648442 |
| 618 | VC_149_8 | 66 | 643500 | 766163.0263 | 0.456492075 | 0 | 0 | 10.39230485 | 2145 | 1755 | 0.818181818 | 6.463957648 | 0 | 0 | 0 | AY176327,AY954969,EU418428,FR852584,HQ163896,JX080300,JX080301,JX080302,JX080303,JX080304,JX080305,JX875065,KC012913,KF766114,KJ206559,KJ206560,KJ206561,KJ206562,KJ206563,KP687431,KP687432,KR902361,KR908644,KY581279,MF398190,MF405094,MG656408,MH136584,MH844528,MH844529,MK331930,MK331931,MK417514,MK417515,MK417516,MK584893,MK656892,MN047438,MN336261,MN336262,MN336263,MN539736,MN539737,MN539738,MT080600,MT080602,MT226657,MT338525,MW460250,MW528836,MW546064,MW546065,MW546066,MW546067,MW546068,MW546069,MW546070,MW546072,MW546074,OL362280,OL960567,OL960568,OL960569,OL960570,OM030343,OM373557 |
| 619 | VC_149_9 | 3 | 900 | 106201.1063 | 0.008403275 | 0.021454443 | 7.937253933 | 8.888194417 | 3 | 3 | 1 | 8.357219867 | 0 | 0 | 0 | AP018375,MT411892,OP263967 |
| 620 | VC_1500_0 | 2 | 43.47911773 | 1.118016055 | 0.974930764 | 0.5 | 0 | 0 | 1 | 1 | 1 | 0 | 0 | 0 | 0 | S1-wms-combined_394628,S12-wms-combined_401379 |
| 621 | VC_1501_0 | 2 | 4.619287091 | 35.46274095 | 0.115245843 | 0.280637918 | 7.874007874 | 7.874007874 | 1 | 1 | 1 | 7.874007874 | 0 | 0 | 0 | S8-wms-combined_276204,S8-wms_57121 |
| 622 | VC_1502_0 | 2 | 33.367684 | 50.19727374 | 0.39930235 | 0.081867177 | 7.416198487 | 7.416198487 | 1 | 1 | 1 | 7.416198487 | 0 | 0 | 0 | S11-wms-combined_17029,S12-wms-combined_375343 |
| 623 | VC_1503_0 | 3 | 354.7435472 | 34.61511723 | 0.911097093 | 0.009451772 | 5.567764363 | 7 | 3 | 3 | 1 | 6.349501687 | 0 | 0 | 0 | S11-wms-combined_217790,S12-wms-combined_111524,S5-wms-combined_98202 |
| 624 | VC_1504_0 | 2 | 60.7080931 | 2.226001104 | 0.964629647 | 0.5 | 6.32455532 | 6.32455532 | 1 | 1 | 1 | 6.32455532 | 0 | 0 | 0 | S11-wms-combined_221310,S8-wms-combined_215102 |
| 625 | VC_1505_0 | 4 | 365.6148947 | 19.43590799 | 0.949523783 | 0.001207324 | 5 | 7.416198487 | 6 | 6 | 1 | 6.604960038 | 0 | 0 | 0 | S11-wms-combined_41367,S12-wms-combined_256299,S17-wms-combined_158354,S5-wms_85650 |
| 626 | VC_1506_0 | 2 | 10.62980693 | 39.76173325 | 0.210944275 | 0.191366544 | 8.602325267 | 8.602325267 | 1 | 1 | 1 | 8.602325267 | 0 | 0 | 0 | S5-wms-combined_178432,S6-wms-combined_72228 |
| 627 | VC_1507_0 | 2 | 5.776189 | 4.860891598 | 0.543023901 | 0.270145687 | 4.69041576 | 4.69041576 | 1 | 1 | 1 | 4.69041576 | 0 | 0 | 0 | S12-wms-combined_264845,S12-wms_373775 |
| 628 | VC_1508_0 | 3 | 155.4711449 | 17.71885559 | 0.897691232 | 0.040427799 | 8 | 9 | 3 | 2 | 0.666666667 | 8.354085916 | 0 | 0 | 0 | S17-wms-combined_203701,S4-wms-combined_57123,S6-wms-combined_135223 |
| 629 | VC_1509_0 | 2 | 164.7898519 | 111.6200581 | 0.596179247 | 0.087622123 | 3.605551275 | 3.605551275 | 1 | 1 | 1 | 3.605551275 | 0 | 0 | 0 | S17-wms-combined_179771,S7-wms-combined_72070 |
| 630 | VC_150_0 | 13 | 23400 | 73432.19069 | 0.241655175 | 1.34E-51 | 0 | 9 | 78 | 77 | 0.987179487 | 6.230438546 | 0 | 0 | 0 | KF021268,KP027447,MH321490,MH321491,MH542234,MT596501,MT596502,MT596503,MT596504,MT596505,MT596506,MZ417323,MZ417325 |
| 631 | VC_1510_0 | 2 | 10.9800245 | 0 | 1 | 0 | 0 | 0 | 1 | 1 | 1 | 0 | 0 | 0 | 0 | S12-wms-combined_401383,S9-wms-combined_3878 |
| 632 | VC_1511_0 | 2 | 7.44029455 | 0 | 1 | 0 | 5 | 5 | 1 | 1 | 1 | 5 | 0 | 0 | 0 | S12-wms_15700,S8-wms-combined_226076 |
| 633 | VC_1512_0 | 2 | 2.933115285 | 0 | 1 | 0 | 0 | 0 | 1 | 1 | 1 | 0 | 0 | 0 | 0 | S17-wms-combined_223748,S9-wms_64472 |
| 634 | VC_1513_0 | 2 | 15.20842239 | 1.209466374 | 0.926332405 | 0.5 | 4.898979486 | 4.898979486 | 1 | 1 | 1 | 4.898979486 | 0 | 0 | 0 | S17-wms-combined_225356,S4-wms_107182 |
| 635 | VC_1514_0 | 2 | 9.826238618 | 0 | 1 | 0 | 1 | 1 | 1 | 1 | 1 | 1 | 0 | 0 | 0 | S5-wms-combined_39793,S7-wms-combined_143218 |
| 636 | VC_151_0 | 6 | 4500 | 56765.36966 | 0.073450956 | 1.33E-11 | 0 | 8.831760866 | 15 | 15 | 1 | 6.618294809 | 0 | 0 | 0 | KX171212,KX171213,MW430345,MZ417329,MZ417330,MZ417344 |
| 637 | VC_152_0 | 1 | 0 | 5908.481571 | 0 | 0 | 0 | 0 | 1 | 1 | 1 | 0 | 0 | 0 | 0 | LC647030 |
| 638 | VC_152_1 | 1 | 0 | 4169.681694 | 0 | 0 | 0 | 0 | 1 | 1 | 1 | 0 | 0 | 0 | 0 | MZ417326 |
| 639 | VC_153_0 | 25 | 90000 | 26444.95332 | 0.772897386 | 1.12E-142 | 0 | 8.062257748 | 300 | 300 | 1 | 5.573626113 | 0 | 0 | 0 | AP019522,JX194239,JX846612,JX846613,KP881332,KX532239,KY779848,LC492751,LC492752,MT080584,MT080587,MT080588,MT080590,MT080595,MW363798,MW546071,MW546075,MW546076,MW546077,MZ643272,ON814134,ON814135,ON814136,ON866946,OP263969 |
| 640 | VC_153_1 | 1 | 0 | 8761.775594 | 0 | 0 | 0 | 0 | 1 | 1 | 1 | 0 | 0 | 0 | 0 | LC576631 |
| 641 | VC_154_0 | 4 | 300.9148492 | 1402.886472 | 0.176613814 | 4.89E-05 | 4.358898944 | 6.633249581 | 6 | 6 | 1 | 5.845516355 | 0 | 0 | 0 | AB854109,KY117485,MN844877,OL634959 |
| 642 | VC_155_0 | 14 | 11428.51712 | 7323.511316 | 0.60945498 | 4.62E-55 | 0 | 6.782329983 | 91 | 91 | 1 | 4.202627495 | 0 | 0 | 0 | FN594518,MT104468,MT104469,MT104470,MT104471,MT104472,MT104476,MT104477,MT354570,MW627366,MZ868714,MZ868715,MZ868716,MZ868717 |
| 643 | VC_156_0 | 12 | 9950.94663 | 1029.926012 | 0.906207271 | 8.86E-30 | 1 | 5.567764363 | 66 | 66 | 1 | 3.709839687 | 0 | 0 | 0 | FR751545,KM247287,MK629528,MN850568,MN850570,MN850583,MN850592,MN850608,MN850617,MN850624,MN850625,MW250275 |
| 644 | VC_157_0 | 3 | 509.7785864 | 856.0247477 | 0.373244503 | 0.00191903 | 1.414213562 | 2.828427125 | 3 | 3 | 1 | 2.230710143 | 0 | 0 | 0 | MK962628,MK962638,MK962640 |
| 645 | VC_159_0 | 2 | 160.637858 | 1040.78759 | 0.133706056 | 0.04371377 | 2.828427125 | 2.828427125 | 1 | 1 | 1 | 2.828427125 | 0 | 0 | 0 | OM913597,OM913598 |
| 646 | VC_15_0 | 3 | 361.2304471 | 7148.582097 | 0.048101127 | 0.001465399 | 7 | 7.280109889 | 3 | 3 | 1 | 7.16373748 | 0 | 0 | 0 | KX827371,MK075002,OM373558 |
| 647 | VC_15_1 | 9 | 2772.329634 | 12463.93259 | 0.18195602 | 7.69E-23 | 4.472135955 | 9.486832981 | 36 | 32 | 0.888888889 | 7.463468535 | 0 | 0 | 0 | AY954954,MK075001,MK075004,MK450538,OM373550,OM373554,OM373555,OM373556,OM373561 |
| 648 | VC_160_0 | 2 | 148.853439 | 151.4831149 | 0.495622118 | 0.056361467 | 3.605551275 | 3.605551275 | 1 | 1 | 1 | 3.605551275 | 0 | 0 | 0 | MF150911,MN996301 |
| 649 | VC_161_0 | 3 | 487.8368293 | 356.6612016 | 0.577664851 | 0.00195604 | 1.414213562 | 3.741657387 | 3 | 3 | 1 | 2.873324188 | 0 | 0 | 0 | AB863625,MT740737,MT740741 |
| 650 | VC_162_0 | 18 | 17769.63491 | 6234.873331 | 0.740262401 | 2.99E-66 | 1.732050808 | 9.273618495 | 153 | 132 | 0.862745098 | 7.090655067 | 0 | 0 | 0 | AY349011,FJ937737,JN662425,JX104231,MH638294,MT361768,MT740727,MT740730,MT740731,MT740732,MT740733,MT740736,MT740738,MT740739,MT740740,MT740742,OM283127,OV121131 |
| 651 | VC_163_0 | 2 | 36.54933039 | 91.01037225 | 0.286527247 | 0.058939311 | 6.708203932 | 6.708203932 | 1 | 1 | 1 | 6.708203932 | 0 | 0 | 0 | MW202475,MW202867 |
| 652 | VC_164_0 | 3 | 900 | 29296.33813 | 0.029804938 | 0.170162741 | 2 | 4 | 3 | 3 | 1 | 3.154700538 | 0 | 0 | 0 | MK268344,MK773491,MT001829 |
| 653 | VC_164_1 | 1 | 0 | 10350.71338 | 0 | 0 | 0 | 0 | 1 | 1 | 1 | 0 | 0 | 0 | 0 | MT446416 |
| 654 | VC_164_10 | 5 | 3000 | 45799.74405 | 0.061475732 | 0.035164184 | 5 | 8.366600265 | 10 | 10 | 1 | 6.756909001 | 0 | 0 | 0 | AB897757,JQ513383,MT708547,OP617745,OU509535 |
| 655 | VC_164_11 | 10 | 13500 | 76062.51673 | 0.1507327 | 1.58E-09 | 5.744562647 | 9.746794345 | 45 | 41 | 0.911111111 | 7.533132912 | 0 | 0 | 0 | KC295538,KM507819,LC494302,LT603033,MH383160,MH791411,MK327931,MK817115,MW822008,ON568341 |
| 656 | VC_164_2 | 1 | 0 | 9649.720417 | 0 | 0 | 0 | 0 | 1 | 1 | 1 | 0 | 0 | 0 | 0 | MH494197 |
| 657 | VC_164_3 | 1 | 0 | 10334.39817 | 0 | 0 | 0 | 0 | 1 | 1 | 1 | 0 | 0 | 0 | 0 | ON286972 |
| 658 | VC_164_4 | 1 | 0 | 10326.84073 | 0 | 0 | 0 | 0 | 1 | 1 | 1 | 0 | 0 | 0 | 0 | JN882285 |
| 659 | VC_164_5 | 1 | 0 | 10243.70835 | 0 | 0 | 0 | 0 | 1 | 1 | 1 | 0 | 0 | 0 | 0 | KU574722 |
| 660 | VC_164_6 | 1 | 0 | 10341.51957 | 0 | 0 | 0 | 0 | 1 | 1 | 1 | 0 | 0 | 0 | 0 | OP292654 |
| 661 | VC_164_7 | 7 | 6300 | 59614.18713 | 0.095578817 | 2.00E-05 | 3 | 6.08276253 | 21 | 21 | 1 | 4.198196279 | 0 | 0 | 0 | KY630187,MT939486,MT939487,MT939488,MW879342,MW879343,MW879344 |
| 662 | VC_164_8 | 3 | 900 | 29243.38875 | 0.029857293 | 0.150553512 | 0 | 5.656854249 | 3 | 3 | 1 | 3.771236166 | 0 | 0 | 0 | LR596615,LT960551,LT960552 |
| 663 | VC_164_9 | 1 | 0 | 6339.081617 | 0 | 0 | 0 | 0 | 1 | 1 | 1 | 0 | 0 | 0 | 0 | MG592671 |
| 664 | VC_165_0 | 4 | 1800 | 6210.016059 | 0.224718651 | 1.67E-05 | 3 | 4.69041576 | 6 | 6 | 1 | 3.876563835 | 0 | 0 | 0 | LC727697,LC727698,LR588166,MK797984 |
| 665 | VC_166_0 | 26 | 77776.21373 | 20382.99439 | 0.792347608 | 8.76E-165 | 0 | 8.306623863 | 325 | 325 | 1 | 4.660748372 | 0 | 0 | 0 | AM749441,DQ163916,JX131330,KC758116,KT734862,KX198613,KX961385,KY000218,KY000219,KY000220,KY000221,MF490239,MH791403,MH791406,MN871456,MN871463,MN871469,MN871478,MN871482,MN871486,MT118302,MZ089734,MZ089741,OL763419,OM809160,ON165687 |
| 666 | VC_167_0 | 3 | 900 | 394.5491546 | 0.695222732 | 0.00173425 | 1 | 2.828427125 | 3 | 3 | 1 | 2.158059479 | 0 | 0 | 0 | AB910392,MK443266,MK803322 |
| 667 | VC_169_0 | 20 | 37837.01925 | 12415.60697 | 0.752936157 | 1.73E-110 | 1.414213562 | 7.937253933 | 190 | 190 | 1 | 5.556011134 | 0 | 0 | 0 | JQ067089,JQ067093,KU204984,KX241618,LR743532,MG018224,MT664984,OK275492,OK275493,OL674541,ON005621,ON189045,ON189047,ON331942,ON529854,ON932079,ON932080,ON932084,OP361299,OP380269 |
| 668 | VC_16_0 | 12 | 1426.870011 | 730.0879617 | 0.661519617 | 1.66E-22 | 0 | 3.605551275 | 66 | 66 | 1 | 2.414769882 | 0 | 0 | 0 | AB012573,AB012574,AB572858,AF452449,AF453500,AY242528,CP017905,CP017906,D89074,FJ904927,MN536023,MN690600 |
| 669 | VC_170_0 | 1 | 0 | 12.04923717 | 0 | 0 | 0 | 0 | 1 | 1 | 1 | 0 | 0 | 0 | 0 | KT001913 |
| 670 | VC_170_1 | 1 | 0 | 93.05263174 | 0 | 0 | 0 | 0 | 1 | 1 | 1 | 0 | 0 | 0 | 0 | KY006853 |
| 671 | VC_171_0 | 8 | 4962.696141 | 3471.388723 | 0.588409557 | 8.35E-19 | 1 | 7.141428429 | 28 | 28 | 1 | 4.075828958 | 0 | 0 | 0 | KP202969,KP202970,MH746817,MK962627,MK962633,MK962634,MT241605,MT241607 |
| 672 | VC_172_0 | 5 | 1913.78375 | 3433.349007 | 0.357908404 | 3.05E-08 | 2.236067977 | 5.830951895 | 10 | 10 | 1 | 4.580025852 | 0 | 0 | 0 | KF626665,KF626668,LR743524,MZ326855,ON189048 |
| 673 | VC_173_0 | 2 | 169.7809307 | 1953.91116 | 0.079946114 | 0.045109088 | 5.385164807 | 5.385164807 | 1 | 1 | 1 | 5.385164807 | 0 | 0 | 0 | KT887558,MG707188 |
| 674 | VC_174_0 | 3 | 900 | 35.19812395 | 0.962362923 | 0.006664428 | 1 | 1.732050808 | 3 | 3 | 1 | 1.382088123 | 0 | 0 | 0 | AB910393,LC102729,OK094707 |
| 675 | VC_175_0 | 4 | 869.5914639 | 1391.110991 | 0.384655425 | 1.74E-05 | 3.464101615 | 7.348469228 | 6 | 6 | 1 | 5.770075875 | 0 | 0 | 0 | MG592578,MG592660,MW865292,OX241572 |
| 676 | VC_176_0 | 2 | 208.9662403 | 3991.28597 | 0.049750879 | 0.043362593 | 6.92820323 | 6.92820323 | 1 | 1 | 1 | 6.92820323 | 0 | 0 | 0 | AF165214,HQ711985 |
| 677 | VC_176_1 | 2 | 135.9459687 | 3613.836762 | 0.036254359 | 0.043424325 | 8.306623863 | 8.306623863 | 1 | 1 | 1 | 8.306623863 | 0 | 0 | 0 | KT887557,ON857940 |
| 678 | VC_176_2 | 1 | 0 | 1948.343483 | 0 | 0 | 0 | 0 | 1 | 1 | 1 | 0 | 0 | 0 | 0 | OM141125 |
| 679 | VC_176_3 | 1 | 0 | 1502.463013 | 0 | 0 | 0 | 0 | 1 | 1 | 1 | 0 | 0 | 0 | 0 | KT968831 |
| 680 | VC_177_0 | 5 | 145.4409 | 49.29896565 | 0.74684708 | 0.001143542 | 1.414213562 | 3.464101615 | 10 | 10 | 1 | 2.665768372 | 0 | 0 | 0 | AB910602,HG315669,KX181651,KY853667,MN335248 |
| 681 | VC_178_0 | 5 | 227.9381438 | 66.50805902 | 0.774124922 | 1.01E-05 | 1 | 2.828427125 | 10 | 10 | 1 | 2.301171671 | 0 | 0 | 0 | MH206183,MH206184,MH218848,MK512531,MN263053 |
| 682 | VC_179_0 | 53 | 217386.3131 | 20524.01427 | 0.913732142 | 0 | 0 | 6.708203932 | 1378 | 1378 | 1 | 4.275079744 | 0 | 0 | 0 | AY394005,DQ631426,DQ873690,EU272036,EU272037,HE584812,HM624080,JN808773,JN811560,JQ067085,JQ762257,JX434030,JX434031,JX434032,JX434033,KF475786,KJ477077,KM233689,KT968832,KU199707,KU199708,KU199709,LN610590,LN907801,LT608331,LT999987,MH643777,MH643778,MK144667,MK511017,MK511018,MK511019,MK511020,MK511021,MK511022,MK511023,MK511024,MK511025,MK511026,MK511027,MK511028,MK511029,MK511030,MK511031,MK511032,MK511033,MK511034,MK511035,MK511036,MK511037,MK511038,MW390886,OM541307 |
| 683 | VC_17_0 | 9 | 1323.577384 | 575.5641335 | 0.696934574 | 2.73E-18 | 1.414213562 | 2.645751311 | 36 | 36 | 1 | 1.966193006 | 0 | 0 | 0 | AB043678,AB043679,MN719123,MT188662,MT188663,MT188664,MT188665,MT188666,MT193890 |
| 684 | VC_180_0 | 4 | 773.0916907 | 2986.222222 | 0.205647017 | 1.38E-05 | 0 | 3.872983346 | 6 | 6 | 1 | 2.363324182 | 0 | 0 | 0 | AB916497,ON170148,ON170170,ON170354 |
| 685 | VC_181_0 | 5 | 457.526721 | 3459.059229 | 0.11681774 | 5.35E-08 | 4 | 7.071067812 | 10 | 10 | 1 | 5.899422226 | 0 | 0 | 0 | JF832915,KP137440,MK388859,S1-wms-combined_19031,S11-wms-combined_268967 |
| 686 | VC_182_0 | 5 | 627.5162961 | 387.2362285 | 0.618393432 | 3.26E-07 | 0 | 8.366600265 | 10 | 10 | 1 | 6.391654069 | 0 | 0 | 0 | JF974307,JF974308,KT381864,KT381865,KX077179 |
| 687 | VC_183_0 | 2 | 67.83568441 | 949.0846561 | 0.06670698 | 0.045101067 | 4.69041576 | 4.69041576 | 1 | 1 | 1 | 4.69041576 | 0 | 0 | 0 | KF302035,MZ568827 |
| 688 | VC_184_0 | 2 | 202.5113226 | 51.76539008 | 0.796421035 | 0.063315229 | 3.31662479 | 3.31662479 | 1 | 1 | 1 | 3.31662479 | 0 | 0 | 0 | MH727593,MZ547449 |
| 689 | VC_185_0 | 3 | 16.01522219 | 19.67897952 | 0.448678537 | 0.028519937 | 6.708203932 | 7.141428429 | 3 | 3 | 1 | 6.925945197 | 0 | 0 | 0 | LR797314,MF959999,MK088078 |
| 690 | VC_187_0 | 4 | 1800 | 25432.74038 | 0.06609691 | 5.90E-05 | 6.403124237 | 7.681145748 | 6 | 6 | 1 | 7.012485515 | 0 | 0 | 0 | KX961630,MN150686,OK499977,OK499983 |
| 691 | VC_187_1 | 7 | 6300 | 37968.02708 | 0.142314903 | 7.64E-14 | 3.31662479 | 8.366600265 | 21 | 21 | 1 | 6.89654448 | 0 | 0 | 0 | JN797797,JN797798,KT187252,KT224359,KX987999,MH487649,MN013089 |
| 692 | VC_187_2 | 2 | 300 | 14036.894 | 0.020925034 | 0.069424051 | 6.782329983 | 6.782329983 | 1 | 1 | 1 | 6.782329983 | 0 | 0 | 0 | KJ081346,KJ676859 |
| 693 | VC_187_3 | 9 | 10800 | 44006.57942 | 0.197056633 | 2.21E-22 | 0 | 9.746794345 | 36 | 31 | 0.861111111 | 7.931741392 | 0 | 0 | 0 | JX094431,KJ451625,KT995480,KU577463,KX495186,MG602477,MH638309,MH638312,OL964058 |
| 694 | VC_187_4 | 1 | 0 | 7235.550894 | 0 | 0 | 0 | 0 | 1 | 1 | 1 | 0 | 0 | 0 | 0 | MN364664 |
| 695 | VC_188_0 | 13 | 838.2350435 | 2.245014199 | 0.997328891 | 0.001761978 | 0 | 1 | 78 | 78 | 1 | 0.153846154 | 0 | 0 | 0 | AB971354,AF059242,AF059243,EF068134,FJ483840,FJ483841,FJ483842,FJ483843,FJ483844,FJ539132,FJ539133,FJ539134,FJ539135 |
| 696 | VC_189_0 | 2 | 40.28399931 | 3850.248397 | 0.010354367 | 0.060465563 | 4 | 4 | 1 | 1 | 1 | 4 | 0 | 0 | 0 | AJ604530,AJ604531 |
| 697 | VC_18_0 | 4 | 732.3688929 | 72.6237079 | 0.909783385 | 0.000176534 | 1 | 6.633249581 | 6 | 6 | 1 | 4.600777349 | 0 | 0 | 0 | AB016282,HM072038,OM236514,ON528935 |
| 698 | VC_190_0 | 2 | 300 | 34290.63765 | 0.008672867 | 0.042365267 | 5.916079783 | 5.916079783 | 1 | 1 | 1 | 5.916079783 | 0 | 0 | 0 | KP143763,KX130863 |
| 699 | VC_190_1 | 1 | 0 | 19389.39865 | 0 | 0 | 0 | 0 | 1 | 1 | 1 | 0 | 0 | 0 | 0 | MK373792 |
| 700 | VC_190_10 | 1 | 0 | 20350.10251 | 0 | 0 | 0 | 0 | 1 | 1 | 1 | 0 | 0 | 0 | 0 | KR422353 |
| 701 | VC_190_11 | 1 | 0 | 15969.89458 | 0 | 0 | 0 | 0 | 1 | 1 | 1 | 0 | 0 | 0 | 0 | AC171169 |
| 702 | VC_190_12 | 1 | 0 | 20754.3707 | 0 | 0 | 0 | 0 | 1 | 1 | 1 | 0 | 0 | 0 | 0 | LR027384 |
| 703 | VC_190_13 | 4 | 1800 | 81486.62085 | 0.021612115 | 2.96E-05 | 1.414213562 | 9 | 6 | 4 | 0.666666667 | 7.437928288 | 0 | 0 | 0 | MH370377,MH370378,MN970211,MN994502 |
| 704 | VC_190_14 | 3 | 900 | 61781.97448 | 0.014358195 | 0.003031482 | 8.717797887 | 9.16515139 | 3 | 2 | 0.666666667 | 8.866915721 | 0 | 0 | 0 | LR597655,MN022787,MN316588 |
| 705 | VC_190_15 | 3 | 900 | 60919.38448 | 0.01455854 | 0.002405347 | 8.366600265 | 9.219544457 | 3 | 2 | 0.666666667 | 8.710049489 | 0 | 0 | 0 | KJ190157,KY677846,MN994496 |
| 706 | VC_190_16 | 1 | 0 | 21832.32772 | 0 | 0 | 0 | 0 | 1 | 1 | 1 | 0 | 0 | 0 | 0 | OK272480 |
| 707 | VC_190_17 | 1 | 0 | 21043.48228 | 0 | 0 | 0 | 0 | 1 | 1 | 1 | 0 | 0 | 0 | 0 | AP019559 |
| 708 | VC_190_18 | 1 | 0 | 21629.52886 | 0 | 0 | 0 | 0 | 1 | 1 | 1 | 0 | 0 | 0 | 0 | MF402939 |
| 709 | VC_190_19 | 1 | 0 | 20394.03052 | 0 | 0 | 0 | 0 | 1 | 1 | 1 | 0 | 0 | 0 | 0 | MZ501075 |
| 710 | VC_190_2 | 2 | 300 | 40921.62771 | 0.007277733 | 0.044689377 | 1.414213562 | 1.414213562 | 1 | 1 | 1 | 1.414213562 | 0 | 0 | 0 | MK373797,MK373799 |
| 711 | VC_190_20 | 4 | 1800 | 76642.237 | 0.022946821 | 2.11E-05 | 0 | 8.544003745 | 6 | 6 | 1 | 6.794462362 | 0 | 0 | 0 | LN887948,MK972715,MK972716,MT843274 |
| 712 | VC_190_21 | 1 | 0 | 19586.76816 | 0 | 0 | 0 | 0 | 1 | 1 | 1 | 0 | 0 | 0 | 0 | KY114934 |
| 713 | VC_190_22 | 3 | 900 | 47611.59315 | 0.018552266 | 0.001403304 | 4 | 6.92820323 | 3 | 3 | 1 | 5.952135487 | 0 | 0 | 0 | KM979354,KM979355,MH203051 |
| 714 | VC_190_23 | 10 | 13343.52773 | 176202.3531 | 0.07039735 | 1.48E-27 | 5.477225575 | 9.695359715 | 45 | 38 | 0.844444444 | 8.105580876 | 0 | 0 | 0 | HQ406778,KY963424,MH102285,MN026741,MT311645,MW149273,MW149275,OK108607,OK254197,OP394141 |
| 715 | VC_190_24 | 5 | 3000 | 99378.93128 | 0.029302904 | 4.03E-07 | 4 | 9.38083152 | 10 | 7 | 0.7 | 8.224818939 | 0 | 0 | 0 | HQ665011,LR597659,MZ398246,OK143207,ON185585 |
| 716 | VC_190_25 | 1 | 0 | 19730.74874 | 0 | 0 | 0 | 0 | 1 | 1 | 1 | 0 | 0 | 0 | 0 | MN956514 |

| | | | | | | | | | | | | | | | | |
|---|---|---|---|---|---|---|---|---|---|---|---|---|---|---|---|---|
| 717 | VC_190_26 | 1 | 0 | 19443.74958 | 0 | 0 | 0 | 0 | 1 | 1 | 1 | 0 | 0 | 0 | 0 | MG387042 |
| 718 | VC_190_27 | 1 | 0 | 19229.34703 | 0 | 0 | 0 | 0 | 1 | 1 | 1 | 0 | 0 | 0 | 0 | KR233164 |
| 719 | VC_190_28 | 5 | 3000 | 95735.91224 | 0.030384081 | 6.09E-08 | 1.414213562 | 4.242640687 | 10 | 10 | 1 | 3.028504821 | 0 | 0 | 0 | OK665835,ON185581,ON185582,ON185583,ON185584 |
| 720 | VC_190_29 | 1 | 0 | 22183.2875 | 0 | 0 | 0 | 0 | 1 | 1 | 1 | 0 | 0 | 0 | 0 | MT080103 |
| 721 | VC_190_3 | 1 | 0 | 20610.01085 | 0 | 0 | 0 | 0 | 1 | 1 | 1 | 0 | 0 | 0 | 0 | MN994504 |
| 722 | VC_190_30 | 1 | 0 | 10861.73543 | 0 | 0 | 0 | 0 | 1 | 1 | 1 | 0 | 0 | 0 | 0 | MF774688 |
| 723 | VC_190_31 | 1 | 0 | 16233.54405 | 0 | 0 | 0 | 0 | 1 | 1 | 1 | 0 | 0 | 0 | 0 | MG065692 |
| 724 | VC_190_32 | 1 | 0 | 21108.90924 | 0 | 0 | 0 | 0 | 1 | 1 | 1 | 0 | 0 | 0 | 0 | MZ501058 |
| 725 | VC_190_33 | 1 | 0 | 16783.0568 | 0 | 0 | 0 | 0 | 1 | 1 | 1 | 0 | 0 | 0 | 0 | MZ501103 |
| 726 | VC_190_34 | 1 | 0 | 19218.17837 | 0 | 0 | 0 | 0 | 1 | 1 | 1 | 0 | 0 | 0 | 0 | MN445182 |
| 727 | VC_190_35 | 1 | 0 | 18558.21395 | 0 | 0 | 0 | 0 | 1 | 1 | 1 | 0 | 0 | 0 | 0 | OK272477 |
| 728 | VC_190_36 | 1 | 0 | 20376.27901 | 0 | 0 | 0 | 0 | 1 | 1 | 1 | 0 | 0 | 0 | 0 | MT653143 |
| 729 | VC_190_37 | 1 | 0 | 20362.63132 | 0 | 0 | 0 | 0 | 1 | 1 | 1 | 0 | 0 | 0 | 0 | MT653146 |
| 730 | VC_190_38 | 3 | 900 | 50631.32546 | 0.017465105 | 0.001520484 | 1.732050808 | 3.741657387 | 3 | 3 | 1 | 3.026419823 | 0 | 0 | 0 | AY543070,AY587007,AY692264 |
| 731 | VC_190_39 | 1 | 0 | 19812.33004 | 0 | 0 | 0 | 0 | 1 | 1 | 1 | 0 | 0 | 0 | 0 | MT233524 |
| 732 | VC_190_4 | 1 | 0 | 20626.66511 | 0 | 0 | 0 | 0 | 1 | 1 | 1 | 0 | 0 | 0 | 0 | MT577844 |
| 733 | VC_190_40 | 2 | 300 | 35807.64594 | 0.00830849 | 0.044436046 | 8.485281374 | 8.485281374 | 1 | 1 | 1 | 8.485281374 | 0 | 0 | 0 | MG969412,MW006479 |
| 734 | VC_190_5 | 1 | 0 | 20300.36538 | 0 | 0 | 0 | 0 | 1 | 1 | 1 | 0 | 0 | 0 | 0 | OM386657 |
| 735 | VC_190_6 | 1 | 0 | 19253.03664 | 0 | 0 | 0 | 0 | 1 | 1 | 1 | 0 | 0 | 0 | 0 | MZ501074 |
| 736 | VC_190_7 | 7 | 6300 | 126108.2022 | 0.047580134 | 8.72E-15 | 0 | 8.888194417 | 21 | 21 | 1 | 3.176010077 | 0 | 0 | 0 | MF431730,MF431731,MF431732,MF431733,MF431734,MF431735,MT074448 |
| 737 | VC_190_8 | 1 | 0 | 20290.9754 | 0 | 0 | 0 | 0 | 1 | 1 | 1 | 0 | 0 | 0 | 0 | OM386660 |
| 738 | VC_190_9 | 1 | 0 | 19891.84901 | 0 | 0 | 0 | 0 | 1 | 1 | 1 | 0 | 0 | 0 | 0 | MK373796 |
| 739 | VC_191_0 | 2 | 300 | 44720.68094 | 0.006663604 | 0.042390289 | 8.124038405 | 8.124038405 | 1 | 1 | 1 | 8.124038405 | 0 | 0 | 0 | MW006478,MW006481 |
| 740 | VC_191_1 | 1 | 0 | 27471.69922 | 0 | 0 | 0 | 0 | 1 | 1 | 1 | 0 | 0 | 0 | 0 | MK370036 |
| 741 | VC_191_10 | 1 | 0 | 26254.57197 | 0 | 0 | 0 | 0 | 1 | 1 | 1 | 0 | 0 | 0 | 0 | MN197465 |
| 742 | VC_191_11 | 1 | 0 | 24943.49915 | 0 | 0 | 0 | 0 | 1 | 1 | 1 | 0 | 0 | 0 | 0 | MF681663 |
| 743 | VC_191_12 | 1 | 0 | 26917.93676 | 0 | 0 | 0 | 0 | 1 | 1 | 1 | 0 | 0 | 0 | 0 | MT074464 |
| 744 | VC_191_13 | 5 | 3000 | 125656.1973 | 0.02331796 | 7.25E-07 | 0 | 1.732050808 | 10 | 10 | 1 | 0.897469149 | 0 | 0 | 0 | MK907267,MK907268,MK907269,MK907270,MK907271 |
| 745 | VC_191_14 | 1 | 0 | 26048.52431 | 0 | 0 | 0 | 0 | 1 | 1 | 1 | 0 | 0 | 0 | 0 | LR027389 |
| 746 | VC_191_15 | 2 | 300 | 52544.0857 | 0.005677078 | 0.063689909 | 8.602325267 | 8.602325267 | 1 | 1 | 1 | 8.602325267 | 0 | 0 | 0 | OL451946,OL502173 |
| 747 | VC_191_16 | 12 | 19800 | 287909.0451 | 0.0643465 | 9.81E-27 | 1.414213562 | 9.643650761 | 66 | 62 | 0.939393939 | 7.002171715 | 0 | 0 | 0 | KY787212,MF001363,MH370369,MK393882,MK947458,MK972717,MN218190,MN218191,MN379739,MN994501,MT074450,MT074458 |
| 748 | VC_191_17 | 4 | 1800 | 106823.2528 | 0.016571038 | 0.000699373 | 6.782329983 | 9.219544457 | 6 | 5 | 0.833333333 | 8.00216241 | 0 | 0 | 0 | CP000917,MK947459,MZ501105,OL770310 |
| 749 | VC_191_18 | 2 | 300 | 52167.54735 | 0.00571782 | 0.080964108 | 9 | 9 | 1 | 0 | 0 | 9 | 0 | 0 | 0 | MT833283,MZ501072 |
| 750 | VC_191_19 | 12 | 19800 | 282290.8336 | 0.0655432 | 2.30E-31 | 1 | 9.486832981 | 66 | 57 | 0.863636364 | 8.017454343 | 0 | 0 | 0 | KM236244,KU927497,KX017521,LC659915,MG065642,MH631454,MK728824,MK728825,MN187969,MW357609,OK288022,OP045498 |
| 751 | VC_191_2 | 1 | 0 | 25778.49115 | 0 | 0 | 0 | 0 | 1 | 1 | 1 | 0 | 0 | 0 | 0 | MZ501109 |
| 752 | VC_191_20 | 1 | 0 | 27227.36716 | 0 | 0 | 0 | 0 | 1 | 1 | 1 | 0 | 0 | 0 | 0 | MK005300 |
| 753 | VC_191_21 | 1 | 0 | 27051.75957 | 0 | 0 | 0 | 0 | 1 | 1 | 1 | 0 | 0 | 0 | 0 | MZ501076 |
| 754 | VC_191_22 | 1 | 0 | 25737.13538 | 0 | 0 | 0 | 0 | 1 | 1 | 1 | 0 | 0 | 0 | 0 | MW567727 |
| 755 | VC_191_23 | 1 | 0 | 27061.48273 | 0 | 0 | 0 | 0 | 1 | 1 | 1 | 0 | 0 | 0 | 0 | MH370379 |
| 756 | VC_191_24 | 1 | 0 | 26153.08948 | 0 | 0 | 0 | 0 | 1 | 1 | 1 | 0 | 0 | 0 | 0 | MZ570151 |
| 757 | VC_191_25 | 1 | 0 | 25347.00592 | 0 | 0 | 0 | 0 | 1 | 1 | 1 | 0 | 0 | 0 | 0 | MW206381 |
| 758 | VC_191_26 | 1 | 0 | 21567.70159 | 0 | 0 | 0 | 0 | 1 | 1 | 1 | 0 | 0 | 0 | 0 | HE956708 |
| 759 | VC_191_27 | 1 | 0 | 25191.98158 | 0 | 0 | 0 | 0 | 1 | 1 | 1 | 0 | 0 | 0 | 0 | MK867835 |
| 760 | VC_191_28 | 1 | 0 | 23661.39424 | 0 | 0 | 0 | 0 | 1 | 1 | 1 | 0 | 0 | 0 | 0 | MZ501108 |
| 761 | VC_191_29 | 1 | 0 | 22075.88908 | 0 | 0 | 0 | 0 | 1 | 1 | 1 | 0 | 0 | 0 | 0 | MH370375 |
| 762 | VC_191_3 | 5 | 3000 | 126652.1684 | 0.023138834 | 1.51E-06 | 4.358898944 | 9.797958971 | 10 | 8 | 0.8 | 7.829415765 | 0 | 0 | 0 | MK972704,MK972707,MK972709,MT074453,MT074457 |
| 763 | VC_191_30 | 2 | 300 | 49258.17705 | 0.006053491 | 0.044682493 | 8.660254038 | 8.660254038 | 1 | 1 | 1 | 8.660254038 | 0 | 0 | 0 | MT074447,MT074461 |
| 764 | VC_191_31 | 2 | 300 | 52544.96014 | 0.005676984 | 0.075756206 | 2 | 2 | 1 | 1 | 1 | 2 | 0 | 0 | 0 | MK770410,MK972708 |
| 765 | VC_191_32 | 12 | 19800 | 270243.2705 | 0.068265676 | 1.33E-36 | 0 | 9.746794345 | 66 | 50 | 0.757575758 | 6.759904261 | 0 | 0 | 0 | MF431736,MF431737,MF431738,MF431739,MF431740,MF431741,MK050846,MK809530,MT074449,MT074465,MT653145,ON526840 |
| 766 | VC_191_33 | 1 | 0 | 23888.23863 | 0 | 0 | 0 | 0 | 1 | 1 | 1 | 0 | 0 | 0 | 0 | MW423798 |
| 767 | VC_191_4 | 2 | 300 | 54457.66338 | 0.005478685 | 0.110634862 | 8.602325267 | 8.602325267 | 1 | 1 | 1 | 8.602325267 | 0 | 0 | 0 | MH370366,MT074455 |
| 768 | VC_191_5 | 6 | 4500 | 153396.7086 | 0.028499644 | 3.93E-08 | 5.385164807 | 8.94427191 | 15 | 15 | 1 | 7.240986422 | 0 | 0 | 0 | MH370386,MT004791,MT074452,MT074456,MT074460,MT074463 |
| 769 | VC_191_6 | 1 | 0 | 26395.09908 | 0 | 0 | 0 | 0 | 1 | 1 | 1 | 0 | 0 | 0 | 0 | MT074468 |
| 770 | VC_191_7 | 1 | 0 | 26692.48171 | 0 | 0 | 0 | 0 | 1 | 1 | 1 | 0 | 0 | 0 | 0 | MT074467 |
| 771 | VC_191_8 | 2 | 300 | 54163.04677 | 0.005508322 | 0.081686479 | 8.246211251 | 8.246211251 | 1 | 1 | 1 | 8.246211251 | 0 | 0 | 0 | MH370376,MT074454 |
| 772 | VC_191_9 | 5 | 3000 | 128725.7283 | 0.022774594 | 3.38E-06 | 1.414213562 | 9.486832981 | 10 | 7 | 0.7 | 7.691184777 | 0 | 0 | 0 | MH370367,MH370380,MT074459,MT074466,MW411578 |
| 773 | VC_192_0 | 15 | 31500 | 30622.84972 | 0.507059804 | 6.21E-68 | 2.449489743 | 5.830951895 | 105 | 105 | 1 | 4.46625316 | 0 | 0 | 0 | KX845404,MG459987,MK521907,MK630230,MN101228,MN163280,MN434094,MN434096,MT380195,MW713578,MZ571828,MZ571830,MZ634344,ON637170,OP171943 |
| 774 | VC_192_1 | 1 | 0 | 11380.64928 | 0 | 0 | 0 | 0 | 1 | 1 | 1 | 0 | 0 | 0 | 0 | MZ079855 |
| 775 | VC_192_2 | 1 | 0 | 7988.346869 | 0 | 0 | 0 | 0 | 1 | 1 | 1 | 0 | 0 | 0 | 0 | MN478377 |
| 776 | VC_193_0 | 2 | 300 | 9034.215628 | 0.032139819 | 0.04237344 | 4.795831523 | 4.795831523 | 1 | 1 | 1 | 4.795831523 | 0 | 0 | 0 | KY363465,MW057857 |
| 777 | VC_195_0 | 2 | 300 | 2197.117871 | 0.120138502 | 0.043199927 | 6.08276253 | 6.08276253 | 1 | 1 | 1 | 6.08276253 | 0 | 0 | 0 | MG676224,MG676225 |
| 778 | VC_195_1 | 1 | 0 | 1388.333863 | 0 | 0 | 0 | 0 | 1 | 1 | 1 | 0 | 0 | 0 | 0 | MK318083 |
| 779 | VC_196_0 | 2 | 300 | 13432.15991 | 0.021846527 | 0.042373039 | 4.582575695 | 4.582575695 | 1 | 1 | 1 | 4.582575695 | 0 | 0 | 0 | MN095770,ON287373 |
| 780 | VC_197_0 | 5 | 42.9951916 | 281.6890521 | 0.132421552 | 0.153015986 | 0 | 2.645751311 | 10 | 10 | 1 | 1.825951226 | 0 | 0 | 0 | AE002163,CMU41758,MG641183,MN862345,MW202581 |
| 781 | VC_198_0 | 28 | 2059.615789 | 6418.464807 | 0.242934207 | 0.000288251 | 0 | 2.645751311 | 378 | 378 | 1 | 2.059570554 | 0 | 0 | 0 | AJ270057,AJ550635,AY769964,MG883723,MG945324,MG945546,MG945571,MG945669,MH572277,MH572280,MH572282,MH572290,MH572330,MH572334,MH572422,MH572438,MH572444,MH572448,MH572452,MH572454,MH992157,MH992174,MH992182,MH992210,MN862343,MW202732,MW202889,OL617035 |
| 782 | VC_199_0 | 12 | 925.7847476 | 1140.824961 | 0.447972708 | 9.20E-37 | 0 | 1.414213562 | 66 | 66 | 1 | 0.449977043 | 0 | 0 | 0 | KC131021,MH572341,MH572342,MH572343,MH572399,MH572400,MH572401,MH572402,MH572403,MH572453,MH572513,MH572514 |
| 783 | VC_19_0 | 2 | 14.94602377 | 342.7214179 | 0.041787488 | 0.079551936 | 7 | 7 | 1 | 1 | 1 | 7 | 0 | 0 | 0 | CP000315,S5-wms-combined_83412 |
| 784 | VC_1_0 | 54 | 124307.3014 | 21467.29167 | 0.852736398 | 0 | 0 | 8.124038405 | 1431 | 1431 | 1 | 5.336925346 | 0 | 0 | 0 | AB276040,AB981169,GU911303,KP966108,MG747435,MT701591,ON170026,ON170032,ON170053,ON170059,ON170061,ON170067,ON170070,ON170073,ON170076,ON170078,ON170080,ON170081,ON170082,ON170084,ON170128,ON170130,ON170131,ON170135,ON170151,ON170173,ON170210,ON170213,ON170227,ON170230,ON170233,ON170235,ON170237,ON170239,ON170241,ON170243,ON170258,ON170261,ON170263,ON170266,ON170268,ON170270,ON170272,ON170276,ON170304,ON170305,ON170321,ON170325,ON170329,ON170344,ON170347,ON170348,ON170351,ON642070 |
| 785 | VC_200_0 | 1 | 0 | 38.34816644 | 0 | 0 | 0 | 0 | 1 | 1 | 1 | 0 | 0 | 0 | 0 | MH992181 |
| 786 | VC_201_0 | 3 | 18.76623677 | 169.2877527 | 0.09979175 | 0.303092391 | 1.414213562 | 2 | 3 | 3 | 1 | 1.804737854 | 0 | 0 | 0 | MH572434,MH572475,MZ286294 |
| 787 | VC_202_0 | 102 | 232117.7805 | 341121.4321 | 0.404923068 | 0 | 0 | 9.899494937 | 5151 | 4057 | 0.787614056 | 8.407074484 | 0 | 0 | 0 | AF009630,AF011378,DQ054536,DQ227763,DQ227764,EU221285,FJ848881,FJ848882,FJ848883,FJ848884,FJ848885,GQ979703,KC182542,KC182543,KC182544,KC182545,KC182546,KC182547,KC182548,KC182549,KC182550,KC182551,KC182552,KC522412,KF676640,KM091442,KP793104,KP793105,KP793108,KP793109,KP793111,KP793113,KP793115,KP793116,KP793117,KP793118,KP793123,KP793124,KP793129,KP793134,KX346246,KX379667,KX379668,KX379670,KX379671,KY554760,KY554761,MF346933,MF448554,MF448555,MF448556,MF448557,MF448558,MF448559,MF448560,MF448561,MF448562,MF448563,MF448564,MF448565,MF448566,MF448567,MF448568,MF448569,MF448570,MF448571,MF448572,MF775703,MF775704,MF775705,MG253647,MG253648,MG253649,MG253650,MG253651,MG253652,MG253653,MG309717,MG309718,MK301437,MN552148,MN552149,MN552150,MN689514,MN689517,MN689518,MN689519,MN689520,MN689522,MN689523,MN689524,MN689525,MW032477,MW041632,MW041633,MW041634,MW041635,MW041636,MW041637,MW041638,MW041639,MW041640 |
| 788 | VC_209_0 | 4 | 1081.170469 | 8440.042981 | 0.113553853 | 1.22E-05 | 0 | 4.123105626 | 6 | 6 | 1 | 2.061552813 | 0 | 0 | 0 | KY888143,KY926698,KY926699,MZ368140 |
| 789 | VC_20_0 | 5 | 479.8840249 | 613.9025757 | 0.438736427 | 6.57E-07 | 6.32455532 | 7.615773106 | 10 | 10 | 1 | 6.905980583 | 0 | 0 | 0 | KT372714,KT852578,KX190835,KX534340,MN065183 |
| 790 | VC_211_0 | 1 | 0 | 455.3646207 | 0 | 0 | 0 | 0 | 1 | 1 | 1 | 0 | 0 | 0 | 0 | AP008983 |
| 791 | VC_211_1 | 1 | 0 | 998.0831717 | 0 | 0 | 0 | 0 | 1 | 1 | 1 | 0 | 0 | 0 | 0 | KP144388 |
| 792 | VC_211_2 | 1 | 0 | 1373.112111 | 0 | 0 | 0 | 0 | 1 | 1 | 1 | 0 | 0 | 0 | 0 | KT151959 |
| 793 | VC_211_3 | 1 | 0 | 654.209181 | 0 | 0 | 0 | 0 | 1 | 1 | 1 | 0 | 0 | 0 | 0 | MT325768 |
| 794 | VC_212_0 | 2 | 153.6749663 | 357.7748628 | 0.300469289 | 0.053311599 | 5.477225575 | 5.477225575 | 1 | 1 | 1 | 5.477225575 | 0 | 0 | 0 | S12-wms-combined_263377,S6-wms_132588 |
| 795 | VC_212_1 | 2 | 86.47065646 | 440.8725463 | 0.163974156 | 0.048000263 | 7.071067812 | 7.071067812 | 1 | 1 | 1 | 7.071067812 | 0 | 0 | 0 | S2-wms-combined_31084,S7-wms-combined_7205 |
| 796 | VC_213_0 | 2 | 300 | 398.8432816 | 0.429280796 | 0.046997734 | 4.358898944 | 4.358898944 | 1 | 1 | 1 | 4.358898944 | 0 | 0 | 0 | KT429160,MT880872 |
| 797 | VC_214_0 | 2 | 300 | 1076.689234 | 0.217914103 | 0.046411969 | 1.732050808 | 1.732050808 | 1 | 1 | 1 | 1.732050808 | 0 | 0 | 0 | ON210834,ON210835 |
| 798 | VC_214_1 | 1 | 0 | 779.7045849 | 0 | 0 | 0 | 0 | 1 | 1 | 1 | 0 | 0 | 0 | 0 | OM654379 |
| 799 | VC_215_0 | 5 | 2522.172713 | 32301.54977 | 0.072426855 | 2.54E-08 | 3.741657387 | 6.403124237 | 10 | 10 | 1 | 5.340275076 | 0 | 0 | 0 | DQ398043,KF017927,KF416344,MG099947,MN585989 |
| 800 | VC_215_1 | 19 | 42660.29283 | 59198.23532 | 0.418819058 | 3.13E-107 | 0 | 6.8556546 | 171 | 171 | 1 | 3.81392702 | 0 | 0 | 0 | AF022214,AP018476,AP018480,KC787102,KC787105,KC787106,KX574455,KX758538,KX897981,LC730318,MH513985,MK494087,MN234169,MT310876,MT310883,MT310884,MT310885,MW924646,OP297553 |
| 801 | VC_215_2 | 5 | 1992.199069 | 32823.9773 | 0.057220501 | 2.57E-08 | 4.242640687 | 8.062257748 | 10 | 10 | 1 | 7.08658422 | 0 | 0 | 0 | JF946695,KT381276,MF919536,MH020237,Z18946 |
| 802 | VC_216_0 | 4 | 1618.484435 | 32067.70813 | 0.04804593 | 1.22E-05 | 2.236067977 | 4.582575695 | 6 | 6 | 1 | 3.712064485 | 0 | 0 | 0 | MF185727,MF324899,MK801735,MN369748 |
| 803 | VC_216_1 | 1 | 0 | 9396.898193 | 0 | 0 | 0 | 0 | 1 | 1 | 1 | 0 | 0 | 0 | 0 | MK814748 |
| 804 | VC_216_2 | 2 | 247.5387595 | 17613.84865 | 0.013858876 | 0.042550033 | 4.242640687 | 4.242640687 | 1 | 1 | 1 | 4.242640687 | 0 | 0 | 0 | MN585968,MN586002 |
| 805 | VC_216_3 | 8 | 4913.201601 | 65160.611 | 0.070114661 | 7.36E-20 | 3.741657387 | 9.273618495 | 28 | 24 | 0.857142857 | 7.77084591 | 0 | 0 | 0 | GU339467,JN408461,KF024721,KU761558,KU761559,KX641263,MF140403,MH825709 |
| 806 | VC_216_4 | 13 | 19307.44912 | 97238.69853 | 0.165663555 | 5.79E-51 | 0 | 8.306623863 | 78 | 78 | 1 | 4.828184227 | 0 | 0 | 0 | KJ959632,KM197169,KM677210,KU297783,KX683875,MF668269,MF919538,MH825701,MK061408,MK284522,MK967395,MZ322019,OM203159 |
| 807 | VC_216_5 | 2 | 195.4757455 | 20599.53135 | 0.009400129 | 0.042521879 | 7.141428429 | 7.141428429 | 1 | 1 | 1 | 7.141428429 | 0 | 0 | 0 | MW601216,MZ209305 |
| 808 | VC_216_6 | 42 | 163568.6864 | 182270.8701 | 0.472961185 | 0 | 0 | 10 | 861 | 719 | 0.835075494 | 7.215970855 | 0 | 0 | 0 | AP018479,JN408460,JX899358,KF981601,KJ410132,KJ410134,KM463009,KM591906,KT588442,KU985093,KU985095,KX576639,KX619650,KY965065,LC730320,MF472896,MF919490,MF919500,MF919531,MH051252,MH077576,MH338234,MH697591,MH825704,MK524491,MK919472,MN234203,MN585981,MN585983,MN585998,MT310877,MT310886,MT498041,MT498060,MT498066,MT658804,MW924643,MZ355728,MZ622171,MZ622182,MZ820090,OP297537 |
| 809 | VC_216_7 | 1 | 0 | 8856.177242 | 0 | 0 | 0 | 0 | 1 | 1 | 1 | 0 | 0 | 0 | 0 | MT310858 |
| 810 | VC_217_0 | 5 | 2289.501023 | 30890.49691 | 0.069002446 | 2.46E-08 | 3.872983346 | 6.92820323 | 10 | 10 | 1 | 5.476779411 | 0 | 0 | 0 | EU744250,KU568494,KX758539,MK279857,MN703408 |
| 811 | VC_217_1 | 1 | 0 | 7912.560291 | 0 | 0 | 0 | 0 | 1 | 1 | 1 | 0 | 0 | 0 | 0 | MN062708 |
| 812 | VC_218_0 | 3 | 782.2795157 | 15465.16003 | 0.048147864 | 0.001390214 | 3 | 5 | 3 | 3 | 1 | 4 | 0 | 0 | 0 | KF861510,MN096376,MT639645 |
| 813 | VC_219_0 | 5 | 1595.157751 | 18353.03061 | 0.079965044 | 2.48E-08 | 2.828427125 | 8.485281374 | 10 | 10 | 1 | 6.384485748 | 0 | 0 | 0 | JF957060,KC661275,KP273225,MG793454,MT553337 |
| 814 | VC_21_0 | 4 | 111.4714721 | 84.21485042 | 0.569643656 | 0.127375581 | 7.681145748 | 9.110433579 | 6 | 4 | 0.666666667 | 8.587254879 | 0 | 0 | 0 | JF974314,KJ746502,MF614627,MF614628 |
| 815 | VC_220_0 | 4 | 1453.906695 | 22193.69812 | 0.061482197 | 1.20E-05 | 5.567764363 | 6.782329983 | 6 | 6 | 1 | 6.165049233 | 0 | 0 | 0 | KR080193,MN310541,MT522004,OM203161 |
| 816 | VC_221_0 | 22 | 67367.22787 | 23093.52086 | 0.744712252 | 2.11E-137 | 0 | 5.099019514 | 231 | 231 | 1 | 3.019205911 | 0 | 0 | 0 | KU695581,KY471460,MF140410,MH338236,MH338237,MH536828,MK494130,MK524507,MK524511,MK524513,MK524514,MK524519,MN310547,MN586006,MN703405,MN892487,MW055904,MZ747517,MZ747521,ON108640,OP297532,OP297549 |
| 817 | VC_222_0 | 5 | 1517.821101 | 25362.36497 | 0.056466168 | 2.53E-08 | 6.244997998 | 8.426149773 | 10 | 10 | 1 | 7.638507314 | 0 | 0 | 0 | KX657793,MH513984,MK494113,MK524530,OM203158 |
| 818 | VC_223_0 | 38 | 179471.2197 | 17863.58826 | 0.909475736 | 0 | 0 | 7.141428429 | 703 | 703 | 1 | 4.653610138 | 0 | 0 | 0 | JF704097,JF937092,JF937094,JN049605,JN698999,JN699019,KF560332,KF560333,KF560334,KM401838,KU695582,KU985096,KX375815,KX576638,KX641261,MF668288,MG099945,MG099952,MH020240,MH051253,MH155876,MH779517,MK359352,MK359360,MK494090,MK894433,MN586039,MN728683,MT024861,MT657338,MW822143,MZ005685,MZ171376,MZ322007,MZ456034,MZ681516,OM913585,OP434447 |
| 819 | VC_224_0 | 19 | 47555.34642 | 2948.112595 | 0.941625531 | 5.60E-92 | 0 | 7.745966692 | 171 | 171 | 1 | 3.912475107 | 0 | 0 | 0 | KU998249,KU998250,KU998251,KX557280,KX557283,KX557284,MF668282,MH001454,MH316566,MH513974,MK967382,MN284898,MN369756,MT522007,MZ028625,MZ622176,MZ681508,OM818328,OP021679 |
| 820 | VC_225_0 | 32 | 113319.9526 | 20305.77267 | 0.848039944 | 7.66E-251 | 0 | 9.110433579 | 496 | 494 | 0.995967742 | 5.745797411 | 0 | 0 | 0 | JF704110,JN699005,KT285706,KU613353,KU716094,KU716095,KX585252,KX664447,KX685355,MG872835,MG872840,MG872842,MH371122,MK061409,MK468591,MK494108,MK524489,MK559428,MK977713,MN234167,MN234200,MN234209,MN234235,MN310542,MN369741,MN369763,MN369764,MN585974,MN585978,MN585979,MN585994,ON724016 |
| 821 | VC_227_0 | 2 | 1.1225071 | 2.245014199 | 0.333333333 | 1 | 0 | 0 | 1 | 1 | 1 | 0 | 0 | 0 | 0 | AF334111,HM066936 |
| 822 | VC_228_0 | 60 | 155914.9098 | 9598.257091 | 0.942009103 | 0 | 0 | 7 | 1770 | 1770 | 1 | 4.921659751 | 0 | 0 | 0 | AF063097,AY135486,AY135739,AY251033,B1U32222,HQ110084,KC139521,KC618326,KF322032,KR073660,KR073661,KT630644,KX833211,LR595869,LR595870,LR595873,LR595883,LR595885,LR595887,MF276773,MK416018,MK907236,MK907237,MK907238,MK907239,MK972710,MK972711,MN016939,MT239398,MT374855,MT374857,MT374858,MZ420230,MZ520833,OK490408,OK490461,OK490464,OK490467,OK490470,OK490473,OM046626,OM046627,OM046628,OM140653,OM475426,OM475429,OM475431,OM475433,OM475434,ON146439,ON146440,ON239131,ON470601,ON470606,ON470613,ON782130,OU784549,OU784746,OU784750,OV192280 |
| 823 | VC_229_0 | 32 | 46600.00671 | 4499.977688 | 0.911937787 | 7.67E-112 | 0 | 7.483314774 | 496 | 496 | 1 | 4.474620715 | 0 | 0 | 0 | CP025874,CP025901,CP025923,CP052639,KP063541,KY271397,LR595891,MG589387,MK416015,OK490394,OK490396,OK490398,OK490400,OK490402,OK490404,OK490406,OK490412,OK490414,OK490422,OK490424,OK490426,OK490428,OK490432,OK490435,OK490437,OK490440,OK490443,OK490446,OK490449,OK490453,OK490456,OK490459 |
| 824 | VC_22_0 | 11 | 2920.285837 | 2735.728737 | 0.516315119 | 8.71E-27 | 1 | 8.062257748 | 55 | 55 | 1 | 5.497753648 | 0 | 0 | 0 | AB036666,HQ906662,HQ906663,HQ906664,KX522565,MK976036,MN180248,MN180249,MN180250,MN180251,MN180252 |
| 825 | VC_230_0 | 3 | 70.61879616 | 884.6255288 | 0.07392747 | 0.006504896 | 8.185352772 | 9.327379053 | 3 | 2 | 0.666666667 | 8.6660044 | 0 | 0 | 0 | JQ780327,MK524177,OL964755 |
| 826 | VC_230_1 | 1 | 0 | 477.3565143 | 0 | 0 | 0 | 0 | 1 | 1 | 1 | 0 | 0 | 0 | 0 | HM208537 |
| 827 | VC_231_0 | 3 | 157.6153094 | 510.278316 | 0.235988642 | 0.001754965 | 5.656854249 | 6.92820323 | 3 | 3 | 1 | 6.355266059 | 0 | 0 | 0 | MT366580,MZ496293,OP121182 |
| 828 | VC_232_0 | 2 | 125.0498273 | 9.811724398 | 0.927245948 | 0.270145687 | 1.414213562 | 1.414213562 | 1 | 1 | 1 | 1.414213562 | 0 | 0 | 0 | AF065411,AF301375 |
| 829 | VC_233_0 | 2 | 24.9650949 | 9.811724398 | 0.717865964 | 0.270145687 | 4.123105626 | 4.123105626 | 1 | 1 | 1 | 4.123105626 | 0 | 0 | 0 | MH674343,MN737622 |
| 830 | VC_234_0 | 21 | 61521.00418 | 12109.07517 | 0.835541734 | 3.12E-99 | 0 | 5.099019514 | 210 | 210 | 1 | 2.748225472 | 0 | 0 | 0 | AP018469,AP018470,AP018471,JN831653,KJ944841,KT361920,KX621007,KY087992,MF140402,MF140435,MH051258,MH926058,MK224497,MK524485,MK524488,MN369761,MN945904,MT310882,MW601218,ON526978,ON645342 |
| 831 | VC_236_0 | 72 | 583676.4075 | 136578.7826 | 0.810374456 | 0 | 0 | 8.831760866 | 2556 | 2556 | 1 | 6.128947404 | 0 | 0 | 0 | HM152764,HM152767,JF704105,JF704106,JN185608,JN643714,KJ567045,KM677211,KX585253,KX641264,KX657794,KX657795,KX657796,KY380102,MF140412,MF140428,MF140430,MF377440,MF472892,MF668267,MF919532,MG962364,MG962371,MH001453,MH051246,MH077577,MH371113,MH509444,MH513977,MH536821,MH536822,MH697578,MH727544,MK016503,MK112537,MK460247,MN234182,MN234197,MN234198,MN234205,MN234230,MN234231,MN369742,MN369746,MN908692,MW712722,MW712723,MW712730,MW712733,MW712734,MW862979,MW862984,MW862985,MW862990,MZ005674,MZ355721,MZ648037,OK040778,OK040782,OK040784,OK310505,OM818330,OM913584,ON081329,ON081331,ON260816,ON456339,ON456353,ON526984,ON687735,ON687736,ON724013 |
| 832 | VC_236_1 | 5 | 2221.338035 | 59790.03023 | 0.035821465 | 2.74E-08 | 2.236067977 | 8.306623863 | 10 | 10 | 1 | 6.288541918 | 0 | 0 | 0 | MF140416,MN234165,MN234174,MN234201,MN234226 |
| 833 | VC_237_0 | 4 | 1693.871323 | 37774.13405 | 0.042917581 | 1.24E-05 | 1.732050808 | 5.830951895 | 6 | 6 | 1 | 3.978626572 | 0 | 0 | 0 | KF713486,MF185722,MK310138,MK494119 |
| 834 | VC_237_1 | 13 | 15105.64098 | 70088.68106 | 0.177308072 | 1.61E-49 | 0 | 9.38083152 | 78 | 57 | 0.730769231 | 6.99515529 | 0 | 0 | 0 | JF937104,JX042579,KP027199,KR080206,KX683428,KY087993,MF140398,MF140406,MF324915,MF373841,MN234199,MT723940,MW601222 |
| 835 | VC_237_2 | 11 | 12082.33592 | 73644.12421 | 0.140940567 | 2.39E-36 | 2 | 9.055385138 | 55 | 54 | 0.981818182 | 6.364072338 | 0 | 0 | 0 | KY945355,MF324908,MF324909,MF324911,MF324912,MF324913,MF324914,MN234176,MN369762,MN945901,ON456351 |
| 836 | VC_237_3 | 8 | 6807.995757 | 55536.19069 | 0.10920017 | 9.27E-20 | 2.236067977 | 8.888194417 | 28 | 28 | 1 | 5.997649661 | 0 | 0 | 0 | MH910038,MH910039,MH910042,MK376955,MK460246,MN284895,MT310859,MT310860 |
| 837 | VC_238_0 | 17 | 33731.46578 | 21480.73253 | 0.61094227 | 3.98E-82 | 0 | 7.071067812 | 136 | 136 | 1 | 5.01230262 | 0 | 0 | 0 | JN243855,KJ567042,KM363596,KM923971,KX585251,KX636165,MF185717,MF185719,MF185720,MG962374,MH051255,MH576966,MH651171,MH697592,MW435854,MZ322016,MZ958747 |
| 838 | VC_23_0 | 3 | 519.3022692 | 686.9090739 | 0.43052345 | 0.001626222 | 3.31662479 | 4.358898944 | 3 | 3 | 1 | 3.891841245 | 0 | 0 | 0 | AY133112,FN297812,JX556417 |
| 839 | VC_240_0 | 4 | 1290.629844 | 361.7275595 | 0.781083948 | 1.73E-05 | 0 | 6.403124237 | 6 | 6 | 1 | 4.944050781 | 0 | 0 | 0 | MW314859,MW584165,MW584169,MW584207 |
| 840 | VC_241_0 | 2 | 49.04344142 | 632.3689051 | 0.071973221 | 0.045664183 | 7.874007874 | 7.874007874 | 1 | 1 | 1 | 7.874007874 | 0 | 0 | 0 | MF975720,MT121966 |
| 841 | VC_242_0 | 2 | 212.5664611 | 2240.2481 | 0.086662263 | 0.046207465 | 2.449489743 | 2.449489743 | 1 | 1 | 1 | 2.449489743 | 0 | 0 | 0 | KT588074,MK340941 |
| 842 | VC_242_1 | 20 | 11311.49717 | 5225.186862 | 0.684024509 | 1.47E-79 | 0 | 10.04987562 | 190 | 159 | 0.836842105 | 7.840483877 | 0 | 0 | 0 | KT630649,MK416007,MK416019,MK422455,MK433579,MK448230,MN098327,MN166823,MT135176,MT701587,MW057855,OK665842,OL791266,OM284015,OM621816,ON470579,ON470582,ON470588,ON470593,OP114732 |
| 843 | VC_243_0 | 2 | 103.5828484 | 1523.449572 | 0.063663666 | 0.043901085 | 5.830951895 | 5.830951895 | 1 | 1 | 1 | 5.830951895 | 0 | 0 | 0 | MF695815,MK416014 |
| 844 | VC_244_0 | 12 | 10659.5207 | 15734.79964 | 0.403856609 | 1.30E-41 | 0 | 7.211102551 | 66 | 66 | 1 | 1.63039999 | 0 | 0 | 0 | KY271401,MK416017,OK490430,OK490433,OK490438,OK490441,OK490444,OK490447,OK490450,OK490451,OK490454,OK490458 |
| 845 | VC_245_0 | 10 | 2287.093513 | 10384.39387 | 0.180491322 | 1.09E-27 | 0 | 9.055385138 | 45 | 44 | 0.977777778 | 6.408248785 | 0 | 0 | 0 | KY653119,LN997803,LR597642,ON470580,ON470583,ON470586,ON470600,ON470607,ON470612,ON470621 |
| 846 | VC_246_0 | 3 | 161.0427249 | 50.18066681 | 0.762428458 | 0.040427799 | 1.414213562 | 2.645751311 | 3 | 3 | 1 | 2.098677617 | 0 | 0 | 0 | AY583234,AY583235,AY583236 |
| 847 | VC_247_0 | 14 | 20555.10116 | 1985.918371 | 0.91189758 | 9.30E-25 | 0 | 9.486832981 | 91 | 88 | 0.967032967 | 6.977853044 | 0 | 0 | 0 | AF234172,AF234173,AF503408,FO818745,KU760857,MH160767,MH422554,MH445380,MH445381,MZ188976,ON018986,ON470615,ON470617,OP279344 |
| 848 | VC_247_1 | 1 | 0 | 1916.570517 | 0 | 0 | 0 | 0 | 1 | 1 | 1 | 0 | 0 | 0 | 0 | MF356679 |
| 849 | VC_248_0 | 11 | 9681.814553 | 4087.764906 | 0.703130737 | 3.27E-31 | 0 | 4 | 55 | 55 | 1 | 1.28942858 | 0 | 0 | 0 | HQ316581,MG592412,MG592507,MG592513,MG592524,MG592527,MG592588,MG592589,MG592669,MG592670,MZ568828 |
| 850 | VC_249_0 | 5 | 1074.600487 | 873.8413154 | 0.551517877 | 1.01E-07 | 0 | 3.605551275 | 10 | 10 | 1 | 1.44222051 | 0 | 0 | 0 | ON170062,ON170209,ON170231,ON170302,ON170328 |
| 851 | VC_24_0 | 3 | 791.7970641 | 1499.699553 | 0.345537086 | 0.001551749 | 0 | 0 | 3 | 3 | 1 | 0 | 0 | 0 | 0 | JX507079,MH142219,MH142220 |
| 852 | VC_250_0 | 4 | 623.7934919 | 1980.439952 | 0.239530559 | 1.76E-05 | 3.16227766 | 6.403124237 | 6 | 6 | 1 | 5.365907086 | 0 | 0 | 0 | AY539836,CP000622,ON170188,S9-wms-combined_64277 |
| 853 | VC_251_0 | 22 | 29398.77358 | 15776.95091 | 0.65076485 | 2.43E-130 | 0 | 7.615773106 | 231 | 231 | 1 | 5.176158801 | 0 | 0 | 0 | AF232233,JX495041,JX495042,JX495043,LN898172,MH719189,MH719190,MH719191,MH719192,MH719193,MH719194,MH719195,MK511057,MK511058,MK511059,MK511060,MK511061,MK511063,MK511064,MK511065,MK934841,MN536027 |

| | | | | | | | | | | | | | | | | |
|---|---|---|---|---|---|---|---|---|---|---|---|---|---|---|---|---|
| 854 | VC_252_0 | 121 | 300525.8772 | 106891.1821 | 0.737636951 | 0 | 0 | 9 | 7260 | 7259 | 0.999862259 | 7.165646137 | 0 | 0 | 0 | AF085222,AF115102,AF115103,AF145054,FJ236310,KT717083,KT717084,KT717085,KY705251,KY705256,KY705257,KY705259,KY705260,KY705261,KY705262,KY705263,KY705264,KY705265,KY705266,KY705267,KY705270,KY705271,KY705272,KY705273,KY705274,KY705275,KY705276,KY705282,KY705283,KY705284,KY705287,KY705288,KY705289,KY705290,MF161328,MF580759,MF580760,MF580761,MF580762,MF580763,MF580764,MF580765,MF580766,MF580767,MF580768,MF580769,MF580770,MF580771,MF580772,MF580773,MF580774,MF580775,MG708273,MG708274,MG708275,MH000602,MH000603,MH000604,MH375594,MH375595,MH375596,MH375597,MH375598,MH375599,MH375600,MH892351,MH892352,MH892353,MH892354,MH892356,MH892357,MH892358,MH892359,MH892360,MH892361,MH892375,MH892377,MH892378,MH892379,MH892380,MH892381,MH892382,MH937458,MH937459,MH937461,MH937462,MH937465,MH937466,MH937467,MH937468,MH937469,MH937470,MH937471,MH937472,MH937474,MH937477,MH937480,MH937481,MH937482,MH937483,MH937485,MH937486,MH937487,MH937488,MH937489,MH937490,MH937491,MH937492,MH937493,MH937495,MH937496,MH937497,MH937499,MH937500,MH937501,MH937504,MH937506,MH937508,MH973661,MH973662,MZ090946 |
| 855 | VC_253_0 | 1 | 0 | 2509.267235 | 0 | 0 | 0 | 0 | 1 | 1 | 1 | 0 | 0 | 0 | 0 | MK448899 |
| 856 | VC_254_0 | 46 | 82669.64514 | 77566.04759 | 0.515925283 | 0 | 0 | 8.485281374 | 1035 | 1035 | 1 | 6.236329199 | 0 | 0 | 0 | AF158600,AY699705,EF529515,FJ226752,HE861935,HG380752,HG424323,KY705258,KY705268,KY705269,KY705277,KY705278,KY705279,KY705280,KY705281,KY705285,KY705286,MH375593,MH892362,MH892363,MH892364,MH892366,MH892376,MH892383,MH892384,MH892385,MH937457,MH937460,MH937463,MH937464,MH937473,MH937475,MH937476,MH937478,MH937479,MH937484,MH937494,MH937498,MH937502,MH937503,MH937505,MH937507,MH937509,MH937510,MH937511,U88974 |
| 857 | VC_255_0 | 13 | 8178.08362 | 31747.58641 | 0.204832721 | 1.70E-51 | 1.414213562 | 6.782329983 | 78 | 78 | 1 | 5.049151107 | 0 | 0 | 0 | FJ965538,KX879643,KY705252,KY705253,KY705254,KY705255,MH892355,MH892367,MH892373,MH973663,MK202159,MK202160,MK202161 |
| 858 | VC_255_1 | 1 | 0 | 1633.64622 | 0 | 0 | 0 | 0 | 1 | 1 | 1 | 0 | 0 | 0 | 0 | S9-wms-combined_20485 |
| 859 | VC_256_0 | 14 | 11169.0841 | 35992.45271 | 0.236826127 | 2.53E-59 | 1 | 6.633249581 | 91 | 91 | 1 | 3.98042113 | 0 | 0 | 0 | KU678389,KU678390,KU678391,KU678392,KX879641,KX879642,MH892350,MH892365,MH892368,MH892369,MH892370,MH892371,MH892372,MH892374 |
| 860 | VC_259_0 | 3 | 715.2446236 | 5514.662951 | 0.114808224 | 0.001413984 | 0 | 1.414213562 | 3 | 3 | 1 | 0.942809042 | 0 | 0 | 0 | MK448824,MK449002,MK449003 |
| 861 | VC_25_0 | 12 | 3650.572096 | 944.3943397 | 0.794471983 | 1.57E-31 | 0 | 5.385164807 | 66 | 66 | 1 | 1.982165596 | 0 | 0 | 0 | ON170017,ON170042,ON170051,ON170056,ON170060,ON170066,ON170072,ON170166,ON170169,ON170274,ON170278,ON170357 |
| 862 | VC_260_0 | 13 | 5184.965108 | 8807.861649 | 0.370544508 | 5.13E-48 | 0 | 8.366600265 | 78 | 78 | 1 | 6.547127137 | 0 | 0 | 0 | AF176025,AF323669,AF489521,AJ245616,LT985568,LT985571,LT993224,MN534316,MN534317,MN689509,MN689516,MN689521,MN689530 |
| 863 | VC_262_0 | 5 | 631.3449179 | 8090.698635 | 0.072384977 | 1.08E-07 | 5.656854249 | 8.366600265 | 10 | 10 | 1 | 7.291158126 | 0 | 0 | 0 | KX160204,KX160219,KX160220,KX456211,MN534320 |
| 864 | VC_263_0 | 7 | 1631.360157 | 5268.286225 | 0.236441126 | 3.25E-15 | 0 | 7.681145748 | 21 | 21 | 1 | 5.263473119 | 0 | 0 | 0 | AF323668,LT985569,LT985574,LT985578,LT985583,LT993229,LT996082 |
| 865 | VC_265_0 | 12 | 7292.891574 | 9276.60908 | 0.44013949 | 2.46E-41 | 0 | 7.211102551 | 66 | 66 | 1 | 5.336945711 | 0 | 0 | 0 | DQ394807,DQ394808,DQ394809,DQ394810,KX456206,KX456208,KX456209,KX456210,KX456213,LT985575,LT985576,LT996081 |
| 866 | VC_267_0 | 2 | 125.4364865 | 1143.55802 | 0.098847147 | 0.044420663 | 6.164414003 | 6.164414003 | 1 | 1 | 1 | 6.164414003 | 0 | 0 | 0 | GQ452243,KT945994 |
| 867 | VC_268_0 | 7 | 3085.10951 | 5917.14834 | 0.342703971 | 4.14E-15 | 1.732050808 | 6.403124237 | 21 | 21 | 1 | 3.672350891 | 0 | 0 | 0 | JX564242,JX567312,KX160206,KX160209,KX160210,KX160211,MN552144 |
| 868 | VC_269_0 | 3 | 56.88816804 | 235.1701242 | 0.194783608 | 0.006918233 | 5.567764363 | 7.615773106 | 3 | 3 | 1 | 6.866578652 | 0 | 0 | 0 | KJ608189,KU981050,KY744240 |
| 869 | VC_26_0 | 4 | 67.67918338 | 23.37704642 | 0.743268017 | 0.000638472 | 0 | 2.828427125 | 6 | 6 | 1 | 2.265684697 | 0 | 0 | 0 | AY714348,JN848801,MN200777,MN200778 |
| 870 | VC_271_0 | 5 | 529.8493955 | 823.3568471 | 0.391551102 | 1.62E-06 | 6 | 9.219544457 | 10 | 8 | 0.8 | 7.942422439 | 0 | 0 | 0 | S11-wms-combined_58538,S11-wms_14390,S2-wms-combined_93845,S8-wms-combined_146562,S8-wms-combined_68780 |
| 871 | VC_271_1 | 1 | 0 | 523.080296 | 0 | 0 | 0 | 0 | 1 | 1 | 1 | 0 | 0 | 0 | 0 | S8-wms-combined_225114 |
| 872 | VC_271_2 | 1 | 0 | 177.8336898 | 0 | 0 | 0 | 0 | 1 | 1 | 1 | 0 | 0 | 0 | 0 | S7-wms-combined_238231 |
| 873 | VC_272_0 | 11 | 3331.362768 | 3447.040706 | 0.491467169 | 2.32E-30 | 1.732050808 | 9.38083152 | 55 | 48 | 0.872727273 | 7.721002822 | 0 | 0 | 0 | AJ251789,AP018361,AY131267,AY605066,EU246945,KC171646,KC171647,MF999224,MH983004,MN830258,MT956952 |
| 874 | VC_274_0 | 2 | 35.63643261 | 410.4036909 | 0.079895128 | 0.046291003 | 5.656854249 | 5.656854249 | 1 | 1 | 1 | 5.656854249 | 0 | 0 | 0 | AJ421943,MH220877 |
| 875 | VC_276_0 | 2 | 18.89078523 | 0 | 1 | 0 | 1 | 1 | 1 | 1 | 1 | 1 | 0 | 0 | 0 | AF133242,AJ969242 |
| 876 | VC_277_0 | 3 | 129.2226826 | 0 | 1 | 0 | 1 | 3.16227766 | 3 | 3 | 1 | 2.387425887 | 0 | 0 | 0 | AF155037,MG966533,MK424903 |
| 877 | VC_278_0 | 3 | 582.1889009 | 1325.620708 | 0.305160902 | 0.001511229 | 0 | 0 | 3 | 3 | 1 | 0 | 0 | 0 | 0 | AY029185,AY526908,AY526909 |
| 878 | VC_279_0 | 2 | 54.34164394 | 233.4106092 | 0.188848717 | 0.048753352 | 7.071067812 | 7.071067812 | 1 | 1 | 1 | 7.071067812 | 0 | 0 | 0 | AY605181,MH128984 |
| 879 | VC_27_0 | 2 | 99.41867932 | 0 | 1 | 0 | 2.828427125 | 2.828427125 | 1 | 1 | 1 | 2.828427125 | 0 | 0 | 0 | AB063393,GQ403789 |
| 880 | VC_280_0 | 2 | 109.4155329 | 212.4090253 | 0.339985033 | 0.048416719 | 3.16227766 | 3.16227766 | 1 | 1 | 1 | 3.16227766 | 0 | 0 | 0 | GQ421471,MT084350 |
| 881 | VC_281_0 | 3 | 446.7391569 | 807.6483375 | 0.356141271 | 0.001716806 | 0 | 1.414213562 | 3 | 3 | 1 | 0.942809042 | 0 | 0 | 0 | MK733262,MK733263,MK733264 |
| 882 | VC_283_0 | 2 | 300 | 119789.507 | 0.002498137 | 0.050634469 | 2.236067977 | 2.236067977 | 1 | 1 | 1 | 2.236067977 | 0 | 0 | 0 | MN508614,MN508618 |
| 883 | VC_283_1 | 1 | 0 | 57794.13165 | 0 | 0 | 0 | 0 | 1 | 1 | 1 | 0 | 0 | 0 | 0 | MZ502379 |
| 884 | VC_283_10 | 1 | 0 | 59310.5022 | 0 | 0 | 0 | 0 | 1 | 1 | 1 | 0 | 0 | 0 | 0 | AP011113 |
| 885 | VC_283_100 | 2 | 300 | 111981.7135 | 0.002671851 | 0.091082117 | 1.414213562 | 1.414213562 | 1 | 1 | 1 | 1.414213562 | 0 | 0 | 0 | MT515752,MT526905 |
| 886 | VC_283_101 | 1 | 0 | 59790.40236 | 0 | 0 | 0 | 0 | 1 | 1 | 1 | 0 | 0 | 0 | 0 | LR597660 |
| 887 | VC_283_102 | 1 | 0 | 35201.49716 | 0 | 0 | 0 | 0 | 1 | 1 | 1 | 0 | 0 | 0 | 0 | OK570375 |
| 888 | VC_283_103 | 1 | 0 | 57336.87394 | 0 | 0 | 0 | 0 | 1 | 1 | 1 | 0 | 0 | 0 | 0 | ON528735 |
| 889 | VC_283_104 | 1 | 0 | 53313.35613 | 0 | 0 | 0 | 0 | 1 | 1 | 1 | 0 | 0 | 0 | 0 | OL310488 |
| 890 | VC_283_105 | 1 | 0 | 56264.59798 | 0 | 0 | 0 | 0 | 1 | 1 | 1 | 0 | 0 | 0 | 0 | MZ234013 |
| 891 | VC_283_106 | 1 | 0 | 54679.41562 | 0 | 0 | 0 | 0 | 1 | 1 | 1 | 0 | 0 | 0 | 0 | MZ753803 |
| 892 | VC_283_107 | 1 | 0 | 58364.35483 | 0 | 0 | 0 | 0 | 1 | 1 | 1 | 0 | 0 | 0 | 0 | ON528739 |
| 893 | VC_283_108 | 1 | 0 | 56752.13998 | 0 | 0 | 0 | 0 | 1 | 1 | 1 | 0 | 0 | 0 | 0 | OK148441 |
| 894 | VC_283_109 | 2 | 300 | 112659.7316 | 0.002655814 | 0.084642774 | 1.732050808 | 1.732050808 | 1 | 1 | 1 | 1.732050808 | 0 | 0 | 0 | KY608967,OK275722 |
| 895 | VC_283_11 | 1 | 0 | 60377.51078 | 0 | 0 | 0 | 0 | 1 | 1 | 1 | 0 | 0 | 0 | 0 | ON528741 |
| 896 | VC_283_110 | 2 | 300 | 111929.1516 | 0.002673102 | 0.090895844 | 1.732050808 | 1.732050808 | 1 | 1 | 1 | 1.732050808 | 0 | 0 | 0 | MN508616,MN508617 |
| 897 | VC_283_111 | 1 | 0 | 56430.69191 | 0 | 0 | 0 | 0 | 1 | 1 | 1 | 0 | 0 | 0 | 0 | KY290975 |
| 898 | VC_283_112 | 1 | 0 | 58313.26299 | 0 | 0 | 0 | 0 | 1 | 1 | 1 | 0 | 0 | 0 | 0 | OL870317 |
| 899 | VC_283_113 | 1 | 0 | 57554.29511 | 0 | 0 | 0 | 0 | 1 | 1 | 1 | 0 | 0 | 0 | 0 | LR215724 |
| 900 | VC_283_114 | 1 | 0 | 59367.06972 | 0 | 0 | 0 | 0 | 1 | 1 | 1 | 0 | 0 | 0 | 0 | KX130861 |
| 901 | VC_283_115 | 1 | 0 | 58301.33869 | 0 | 0 | 0 | 0 | 1 | 1 | 1 | 0 | 0 | 0 | 0 | JN986846 |
| 902 | VC_283_116 | 1 | 0 | 61944.16504 | 0 | 0 | 0 | 0 | 1 | 1 | 1 | 0 | 0 | 0 | 0 | MZ501089 |
| 903 | VC_283_117 | 3 | 900 | 169892.4395 | 0.005269554 | 0.002479634 | 1 | 2.236067977 | 3 | 3 | 1 | 1.745355992 | 0 | 0 | 0 | MZ234031,MZ234035,MZ234038 |
| 904 | VC_283_118 | 3 | 900 | 166565.7181 | 0.005374234 | 0.008892194 | 3.16227766 | 6.633249581 | 3 | 3 | 1 | 5.425422646 | 0 | 0 | 0 | MZ501113,OL741432,OL870316 |
| 905 | VC_283_119 | 1 | 0 | 62238.8327 | 0 | 0 | 0 | 0 | 1 | 1 | 1 | 0 | 0 | 0 | 0 | MK327939 |
| 906 | VC_283_12 | 1 | 0 | 60183.16132 | 0 | 0 | 0 | 0 | 1 | 1 | 1 | 0 | 0 | 0 | 0 | MK373785 |
| 907 | VC_283_120 | 1 | 0 | 60561.70395 | 0 | 0 | 0 | 0 | 1 | 1 | 1 | 0 | 0 | 0 | 0 | MK962752 |
| 908 | VC_283_121 | 2 | 300 | 115568.7289 | 0.002589137 | 0.092994048 | 1.414213562 | 1.414213562 | 1 | 1 | 1 | 1.414213562 | 0 | 0 | 0 | MK327940,MK327945 |
| 909 | VC_283_122 | 1 | 0 | 57253.87028 | 0 | 0 | 0 | 0 | 1 | 1 | 1 | 0 | 0 | 0 | 0 | OK274152 |
| 910 | VC_283_123 | 1 | 0 | 57651.75063 | 0 | 0 | 0 | 0 | 1 | 1 | 1 | 0 | 0 | 0 | 0 | OL362041 |
| 911 | VC_283_124 | 1 | 0 | 58602.44421 | 0 | 0 | 0 | 0 | 1 | 1 | 1 | 0 | 0 | 0 | 0 | MK373779 |
| 912 | VC_283_125 | 1 | 0 | 59253.52705 | 0 | 0 | 0 | 0 | 1 | 1 | 1 | 0 | 0 | 0 | 0 | KX130862 |
| 913 | VC_283_126 | 1 | 0 | 59527.49259 | 0 | 0 | 0 | 0 | 1 | 1 | 1 | 0 | 0 | 0 | 0 | OM386666 |
| 914 | VC_283_127 | 1 | 0 | 55530.80668 | 0 | 0 | 0 | 0 | 1 | 1 | 1 | 0 | 0 | 0 | 0 | MK327929 |
| 915 | VC_283_128 | 1 | 0 | 57758.85566 | 0 | 0 | 0 | 0 | 1 | 1 | 1 | 0 | 0 | 0 | 0 | OK076929 |
| 916 | VC_283_129 | 1 | 0 | 58587.47647 | 0 | 0 | 0 | 0 | 1 | 1 | 1 | 0 | 0 | 0 | 0 | MH992510 |
| 917 | VC_283_13 | 1 | 0 | 55951.3093 | 0 | 0 | 0 | 0 | 1 | 1 | 1 | 0 | 0 | 0 | 0 | ON528745 |
| 918 | VC_283_130 | 1 | 0 | 60864.94969 | 0 | 0 | 0 | 0 | 1 | 1 | 1 | 0 | 0 | 0 | 0 | MN022785 |
| 919 | VC_283_131 | 1 | 0 | 57161.60568 | 0 | 0 | 0 | 0 | 1 | 1 | 1 | 0 | 0 | 0 | 0 | MN716856 |
| 920 | VC_283_132 | 1 | 0 | 57258.18646 | 0 | 0 | 0 | 0 | 1 | 1 | 1 | 0 | 0 | 0 | 0 | ON528740 |
| 921 | VC_283_133 | 1 | 0 | 56472.12297 | 0 | 0 | 0 | 0 | 1 | 1 | 1 | 0 | 0 | 0 | 0 | MK327936 |
| 922 | VC_283_134 | 1 | 0 | 55392.28672 | 0 | 0 | 0 | 0 | 1 | 1 | 1 | 0 | 0 | 0 | 0 | MK327944 |
| 923 | VC_283_135 | 1 | 0 | 58649.98311 | 0 | 0 | 0 | 0 | 1 | 1 | 1 | 0 | 0 | 0 | 0 | OP168898 |
| 924 | VC_283_136 | 2 | 300 | 115202.4883 | 0.002597347 | 0.058838631 | 0 | 0 | 1 | 1 | 1 | 0 | 0 | 0 | 0 | MT515753,MT515755 |
| 925 | VC_283_137 | 1 | 0 | 58166.83875 | 0 | 0 | 0 | 0 | 1 | 1 | 1 | 0 | 0 | 0 | 0 | LR597657 |
| 926 | VC_283_138 | 1 | 0 | 56006.85911 | 0 | 0 | 0 | 0 | 1 | 1 | 1 | 0 | 0 | 0 | 0 | MG867727 |
| 927 | VC_283_139 | 1 | 0 | 58494.75773 | 0 | 0 | 0 | 0 | 1 | 1 | 1 | 0 | 0 | 0 | 0 | KX009778 |
| 928 | VC_283_14 | 1 | 0 | 59989.80019 | 0 | 0 | 0 | 0 | 1 | 1 | 1 | 0 | 0 | 0 | 0 | MF001360 |
| 929 | VC_283_140 | 2 | 300 | 107088.1139 | 0.002793605 | 0.061880847 | 7.416198487 | 7.416198487 | 1 | 1 | 1 | 7.416198487 | 0 | 0 | 0 | OV876900,OV877085 |
| 930 | VC_283_141 | 1 | 0 | 54265.91385 | 0 | 0 | 0 | 0 | 1 | 1 | 1 | 0 | 0 | 0 | 0 | LR746311 |
| 931 | VC_283_142 | 2 | 300 | 104621.4334 | 0.002859282 | 0.057160884 | 5.916079783 | 5.916079783 | 1 | 1 | 1 | 5.916079783 | 0 | 0 | 0 | ON528726,ON528744 |
| 932 | VC_283_143 | 1 | 0 | 53476.04152 | 0 | 0 | 0 | 0 | 1 | 1 | 1 | 0 | 0 | 0 | 0 | KX452698 |
| 933 | VC_283_144 | 1 | 0 | 53181.19378 | 0 | 0 | 0 | 0 | 1 | 1 | 1 | 0 | 0 | 0 | 0 | MW341595 |
| 934 | VC_283_145 | 1 | 0 | 51542.93427 | 0 | 0 | 0 | 0 | 1 | 1 | 1 | 0 | 0 | 0 | 0 | KM501444 |
| 935 | VC_283_146 | 1 | 0 | 52758.63402 | 0 | 0 | 0 | 0 | 1 | 1 | 1 | 0 | 0 | 0 | 0 | ON528746 |
| 936 | VC_283_147 | 1 | 0 | 55122.49188 | 0 | 0 | 0 | 0 | 1 | 1 | 1 | 0 | 0 | 0 | 0 | MZ501096 |
| 937 | VC_283_148 | 1 | 0 | 52657.83641 | 0 | 0 | 0 | 0 | 1 | 1 | 1 | 0 | 0 | 0 | 0 | JN202312 |
| 938 | VC_283_149 | 1 | 0 | 54913.00302 | 0 | 0 | 0 | 0 | 1 | 1 | 1 | 0 | 0 | 0 | 0 | ON528736 |
| 939 | VC_283_15 | 1 | 0 | 56866.96279 | 0 | 0 | 0 | 0 | 1 | 1 | 1 | 0 | 0 | 0 | 0 | LC465543 |
| 940 | VC_283_150 | 1 | 0 | 55558.58535 | 0 | 0 | 0 | 0 | 1 | 1 | 1 | 0 | 0 | 0 | 0 | MZ065353 |
| 941 | VC_283_151 | 1 | 0 | 54893.21566 | 0 | 0 | 0 | 0 | 1 | 1 | 1 | 0 | 0 | 0 | 0 | MH051915 |
| 942 | VC_283_152 | 1 | 0 | 57278.48967 | 0 | 0 | 0 | 0 | 1 | 1 | 1 | 0 | 0 | 0 | 0 | OL960580 |
| 943 | VC_283_153 | 1 | 0 | 59012.05502 | 0 | 0 | 0 | 0 | 1 | 1 | 1 | 0 | 0 | 0 | 0 | MW749000 |
| 944 | VC_283_154 | 1 | 0 | 60428.93842 | 0 | 0 | 0 | 0 | 1 | 1 | 1 | 0 | 0 | 0 | 0 | HM035025 |

| | | | | | | | | | | | | | | | | |
|---|---|---|---|---|---|---|---|---|---|---|---|---|---|---|---|---|
| 945 | VC_283_155 | 2 | 300 | 110916.1506 | 0.00269745 | 0.047142304 | 1.414213562 | 1.414213562 | 1 | 1 | 1 | 1.414213562 | 0 | 0 | 0 | MT630408,MZ681930 |
| 946 | VC_283_156 | 1 | 0 | 55884.30038 | 0 | 0 | 0 | 0 | 1 | 1 | 1 | 0 | 0 | 0 | 0 | MK327942 |
| 947 | VC_283_157 | 1 | 0 | 55108.60927 | 0 | 0 | 0 | 0 | 1 | 1 | 1 | 0 | 0 | 0 | 0 | OL960581 |
| 948 | VC_283_158 | 1 | 0 | 59233.37871 | 0 | 0 | 0 | 0 | 1 | 1 | 1 | 0 | 0 | 0 | 0 | MF327007 |
| 949 | VC_283_159 | 1 | 0 | 54860.41539 | 0 | 0 | 0 | 0 | 1 | 1 | 1 | 0 | 0 | 0 | 0 | MH837626 |
| 950 | VC_283_16 | 1 | 0 | 60712.32051 | 0 | 0 | 0 | 0 | 1 | 1 | 1 | 0 | 0 | 0 | 0 | MK327937 |
| 951 | VC_283_160 | 1 | 0 | 54251.52797 | 0 | 0 | 0 | 0 | 1 | 1 | 1 | 0 | 0 | 0 | 0 | ON229909 |
| 952 | VC_283_161 | 1 | 0 | 53236.83215 | 0 | 0 | 0 | 0 | 1 | 1 | 1 | 0 | 0 | 0 | 0 | MT179807 |
| 953 | VC_283_162 | 1 | 0 | 48856.56246 | 0 | 0 | 0 | 0 | 1 | 1 | 1 | 0 | 0 | 0 | 0 | ON720975 |
| 954 | VC_283_163 | 2 | 300 | 103200.2982 | 0.002898542 | 0.065792539 | 2.449489743 | 2.449489743 | 1 | 1 | 1 | 2.449489743 | 0 | 0 | 0 | KF925357,KR233165 |
| 955 | VC_283_164 | 2 | 300 | 99767.4428 | 0.002997978 | 0.065242118 | 6 | 6 | 1 | 1 | 1 | 6 | 0 | 0 | 0 | MK886800,MZ502380 |
| 956 | VC_283_165 | 1 | 0 | 53832.30059 | 0 | 0 | 0 | 0 | 1 | 1 | 1 | 0 | 0 | 0 | 0 | OK018184 |
| 957 | VC_283_166 | 1 | 0 | 50780.09139 | 0 | 0 | 0 | 0 | 1 | 1 | 1 | 0 | 0 | 0 | 0 | MZ189262 |
| 958 | VC_283_167 | 1 | 0 | 50764.0544 | 0 | 0 | 0 | 0 | 1 | 1 | 1 | 0 | 0 | 0 | 0 | OM135583 |
| 959 | VC_283_168 | 1 | 0 | 53099.15931 | 0 | 0 | 0 | 0 | 1 | 1 | 1 | 0 | 0 | 0 | 0 | MW822007 |
| 960 | VC_283_169 | 1 | 0 | 61933.86525 | 0 | 0 | 0 | 0 | 1 | 1 | 1 | 0 | 0 | 0 | 0 | MK962750 |
| 961 | VC_283_17 | 1 | 0 | 60859.95597 | 0 | 0 | 0 | 0 | 1 | 1 | 1 | 0 | 0 | 0 | 0 | MK327946 |
| 962 | VC_283_170 | 1 | 0 | 53059.50458 | 0 | 0 | 0 | 0 | 1 | 1 | 1 | 0 | 0 | 0 | 0 | MH051916 |
| 963 | VC_283_171 | 1 | 0 | 53533.24226 | 0 | 0 | 0 | 0 | 1 | 1 | 1 | 0 | 0 | 0 | 0 | MT682709 |
| 964 | VC_283_172 | 1 | 0 | 53032.94284 | 0 | 0 | 0 | 0 | 1 | 1 | 1 | 0 | 0 | 0 | 0 | MW883059 |
| 965 | VC_283_173 | 1 | 0 | 54677.05552 | 0 | 0 | 0 | 0 | 1 | 1 | 1 | 0 | 0 | 0 | 0 | MT968995 |
| 966 | VC_283_174 | 1 | 0 | 56641.60881 | 0 | 0 | 0 | 0 | 1 | 1 | 1 | 0 | 0 | 0 | 0 | MZ417521 |
| 967 | VC_283_175 | 1 | 0 | 53799.65906 | 0 | 0 | 0 | 0 | 1 | 1 | 1 | 0 | 0 | 0 | 0 | MW822011 |
| 968 | VC_283_176 | 1 | 0 | 52917.87372 | 0 | 0 | 0 | 0 | 1 | 1 | 1 | 0 | 0 | 0 | 0 | MZ150757 |
| 969 | VC_283_177 | 1 | 0 | 49687.20753 | 0 | 0 | 0 | 0 | 1 | 1 | 1 | 0 | 0 | 0 | 0 | OL770073 |
| 970 | VC_283_178 | 1 | 0 | 51434.2806 | 0 | 0 | 0 | 0 | 1 | 1 | 1 | 0 | 0 | 0 | 0 | ON645936 |
| 971 | VC_283_179 | 1 | 0 | 54102.54061 | 0 | 0 | 0 | 0 | 1 | 1 | 1 | 0 | 0 | 0 | 0 | MN580668 |
| 972 | VC_283_18 | 1 | 0 | 62863.35367 | 0 | 0 | 0 | 0 | 1 | 1 | 1 | 0 | 0 | 0 | 0 | MK327928 |
| 973 | VC_283_180 | 1 | 0 | 60265.38709 | 0 | 0 | 0 | 0 | 1 | 1 | 1 | 0 | 0 | 0 | 0 | MT515756 |
| 974 | VC_283_181 | 1 | 0 | 57768.90131 | 0 | 0 | 0 | 0 | 1 | 1 | 1 | 0 | 0 | 0 | 0 | MH243439 |
| 975 | VC_283_182 | 1 | 0 | 58640.56435 | 0 | 0 | 0 | 0 | 1 | 1 | 1 | 0 | 0 | 0 | 0 | LC348379 |
| 976 | VC_283_183 | 1 | 0 | 55239.42686 | 0 | 0 | 0 | 0 | 1 | 1 | 1 | 0 | 0 | 0 | 0 | MF001359 |
| 977 | VC_283_184 | 2 | 300 | 119497.3894 | 0.002504228 | 0.064687762 | 2 | 2 | 1 | 1 | 1 | 2 | 0 | 0 | 0 | MT884006,MT884007 |
| 978 | VC_283_185 | 1 | 0 | 59479.61663 | 0 | 0 | 0 | 0 | 1 | 1 | 1 | 0 | 0 | 0 | 0 | MN994497 |
| 979 | VC_283_186 | 1 | 0 | 58649.21804 | 0 | 0 | 0 | 0 | 1 | 1 | 1 | 0 | 0 | 0 | 0 | ON528742 |
| 980 | VC_283_187 | 1 | 0 | 59458.78497 | 0 | 0 | 0 | 0 | 1 | 1 | 1 | 0 | 0 | 0 | 0 | AP018932 |
| 981 | VC_283_188 | 1 | 0 | 62557.4859 | 0 | 0 | 0 | 0 | 1 | 1 | 1 | 0 | 0 | 0 | 0 | MH992121 |
| 982 | VC_283_189 | 1 | 0 | 58897.63014 | 0 | 0 | 0 | 0 | 1 | 1 | 1 | 0 | 0 | 0 | 0 | MF158046 |
| 983 | VC_283_19 | 1 | 0 | 57093.09284 | 0 | 0 | 0 | 0 | 1 | 1 | 1 | 0 | 0 | 0 | 0 | MF327008 |
| 984 | VC_283_190 | 1 | 0 | 58728.9995 | 0 | 0 | 0 | 0 | 1 | 1 | 1 | 0 | 0 | 0 | 0 | MK977694 |
| 985 | VC_283_2 | 2 | 300 | 111377.1494 | 0.002686315 | 0.061573814 | 1.732050808 | 1.732050808 | 1 | 1 | 1 | 1.732050808 | 0 | 0 | 0 | HM997020,KP869105 |
| 986 | VC_283_20 | 1 | 0 | 57187.35527 | 0 | 0 | 0 | 0 | 1 | 1 | 1 | 0 | 0 | 0 | 0 | MF327009 |
| 987 | VC_283_21 | 1 | 0 | 59772.10288 | 0 | 0 | 0 | 0 | 1 | 1 | 1 | 0 | 0 | 0 | 0 | MN781580 |
| 988 | VC_283_22 | 1 | 0 | 55714.97966 | 0 | 0 | 0 | 0 | 1 | 1 | 1 | 0 | 0 | 0 | 0 | OP197928 |
| 989 | VC_283_23 | 1 | 0 | 56296.8144 | 0 | 0 | 0 | 0 | 1 | 1 | 1 | 0 | 0 | 0 | 0 | MT828552 |
| 990 | VC_283_24 | 1 | 0 | 57236.96471 | 0 | 0 | 0 | 0 | 1 | 1 | 1 | 0 | 0 | 0 | 0 | MG775043 |
| 991 | VC_283_25 | 1 | 0 | 60232.55887 | 0 | 0 | 0 | 0 | 1 | 1 | 1 | 0 | 0 | 0 | 0 | KU867876 |
| 992 | VC_283_26 | 1 | 0 | 59263.77784 | 0 | 0 | 0 | 0 | 1 | 1 | 1 | 0 | 0 | 0 | 0 | OL615012 |
| 993 | VC_283_27 | 1 | 0 | 60875.45337 | 0 | 0 | 0 | 0 | 1 | 1 | 1 | 0 | 0 | 0 | 0 | MT176424 |
| 994 | VC_283_28 | 1 | 0 | 60868.82152 | 0 | 0 | 0 | 0 | 1 | 1 | 1 | 0 | 0 | 0 | 0 | MT176426 |
| 995 | VC_283_29 | 1 | 0 | 60523.66594 | 0 | 0 | 0 | 0 | 1 | 1 | 1 | 0 | 0 | 0 | 0 | ON528727 |
| 996 | VC_283_3 | 3 | 900 | 161887.8459 | 0.005528668 | 0.003198296 | 2 | 3.872983346 | 3 | 3 | 1 | 3.248655564 | 0 | 0 | 0 | FJ839693,KM607004,OK040907 |
| 997 | VC_283_30 | 1 | 0 | 59203.79146 | 0 | 0 | 0 | 0 | 1 | 1 | 1 | 0 | 0 | 0 | 0 | ON528728 |
| 998 | VC_283_31 | 1 | 0 | 59299.02445 | 0 | 0 | 0 | 0 | 1 | 1 | 1 | 0 | 0 | 0 | 0 | MN895437 |
| 999 | VC_283_32 | 1 | 0 | 60696.81481 | 0 | 0 | 0 | 0 | 1 | 1 | 1 | 0 | 0 | 0 | 0 | MZ436830 |
| 1000 | VC_283_33 | 4 | 1800 | 236990.7064 | 0.007537982 | 0.000205293 | 0 | 2 | 6 | 6 | 1 | 1.276142375 | 0 | 0 | 0 | LN881726,LN881729,LN881734,LN881737 |
| 1001 | VC_283_34 | 4 | 1800 | 217882.2524 | 0.008193652 | 0.000251904 | 0 | 1.732050808 | 6 | 6 | 1 | 1.093412989 | 0 | 0 | 0 | MH809535,MT515751,MT828551,MT828553 |
| 1002 | VC_283_35 | 1 | 0 | 60693.33042 | 0 | 0 | 0 | 0 | 1 | 1 | 1 | 0 | 0 | 0 | 0 | MK962755 |
| 1003 | VC_283_36 | 1 | 0 | 60056.2788 | 0 | 0 | 0 | 0 | 1 | 1 | 1 | 0 | 0 | 0 | 0 | MZ501065 |
| 1004 | VC_283_37 | 1 | 0 | 60123.85751 | 0 | 0 | 0 | 0 | 1 | 1 | 1 | 0 | 0 | 0 | 0 | LN881732 |
| 1005 | VC_283_38 | 1 | 0 | 58969.3354 | 0 | 0 | 0 | 0 | 1 | 1 | 1 | 0 | 0 | 0 | 0 | LR990704 |
| 1006 | VC_283_39 | 1 | 0 | 60195.08236 | 0 | 0 | 0 | 0 | 1 | 1 | 1 | 0 | 0 | 0 | 0 | MN895434 |
| 1007 | VC_283_4 | 5 | 3000 | 264553.5642 | 0.011212708 | 3.06E-07 | 0 | 1 | 10 | 10 | 1 | 0.4 | 0 | 0 | 0 | MZ291552,OP555980,OP555981,OP555982,OP572230 |
| 1008 | VC_283_40 | 1 | 0 | 59421.36744 | 0 | 0 | 0 | 0 | 1 | 1 | 1 | 0 | 0 | 0 | 0 | MZ234018 |
| 1009 | VC_283_41 | 1 | 0 | 59457.10193 | 0 | 0 | 0 | 0 | 1 | 1 | 1 | 0 | 0 | 0 | 0 | MZ501046 |
| 1010 | VC_283_42 | 1 | 0 | 57504.08644 | 0 | 0 | 0 | 0 | 1 | 1 | 1 | 0 | 0 | 0 | 0 | MZ501050 |
| 1011 | VC_283_43 | 1 | 0 | 60793.60963 | 0 | 0 | 0 | 0 | 1 | 1 | 1 | 0 | 0 | 0 | 0 | MK373786 |
| 1012 | VC_283_44 | 1 | 0 | 60470.60139 | 0 | 0 | 0 | 0 | 1 | 1 | 1 | 0 | 0 | 0 | 0 | MN895438 |
| 1013 | VC_283_45 | 1 | 0 | 58018.74176 | 0 | 0 | 0 | 0 | 1 | 1 | 1 | 0 | 0 | 0 | 0 | MH355584 |
| 1014 | VC_283_46 | 1 | 0 | 60275.03869 | 0 | 0 | 0 | 0 | 1 | 1 | 1 | 0 | 0 | 0 | 0 | MZ501066 |
| 1015 | VC_283_47 | 1 | 0 | 61112.49399 | 0 | 0 | 0 | 0 | 1 | 1 | 1 | 0 | 0 | 0 | 0 | MZ501067 |
| 1016 | VC_283_48 | 1 | 0 | 61085.71627 | 0 | 0 | 0 | 0 | 1 | 1 | 1 | 0 | 0 | 0 | 0 | MN895436 |
| 1017 | VC_283_49 | 1 | 0 | 58118.80707 | 0 | 0 | 0 | 0 | 1 | 1 | 1 | 0 | 0 | 0 | 0 | MT176425 |
| 1018 | VC_283_5 | 1 | 0 | 54896.68993 | 0 | 0 | 0 | 0 | 1 | 1 | 1 | 0 | 0 | 0 | 0 | LC620505 |
| 1019 | VC_283_50 | 1 | 0 | 59876.25626 | 0 | 0 | 0 | 0 | 1 | 1 | 1 | 0 | 0 | 0 | 0 | MZ501106 |
| 1020 | VC_283_51 | 1 | 0 | 58831.11302 | 0 | 0 | 0 | 0 | 1 | 1 | 1 | 0 | 0 | 0 | 0 | MN895435 |
| 1021 | VC_283_52 | 1 | 0 | 60956.36154 | 0 | 0 | 0 | 0 | 1 | 1 | 1 | 0 | 0 | 0 | 0 | MH553563 |
| 1022 | VC_283_53 | 1 | 0 | 61297.26374 | 0 | 0 | 0 | 0 | 1 | 1 | 1 | 0 | 0 | 0 | 0 | MK373781 |
| 1023 | VC_283_54 | 1 | 0 | 61206.44555 | 0 | 0 | 0 | 0 | 1 | 1 | 1 | 0 | 0 | 0 | 0 | MT682713 |
| 1024 | VC_283_55 | 1 | 0 | 60110.03395 | 0 | 0 | 0 | 0 | 1 | 1 | 1 | 0 | 0 | 0 | 0 | ON528743 |
| 1025 | VC_283_56 | 1 | 0 | 60463.32819 | 0 | 0 | 0 | 0 | 1 | 1 | 1 | 0 | 0 | 0 | 0 | MZ726795 |
| 1026 | VC_283_57 | 1 | 0 | 57406.97821 | 0 | 0 | 0 | 0 | 1 | 1 | 1 | 0 | 0 | 0 | 0 | KT184311 |
| 1027 | VC_283_58 | 1 | 0 | 60033.79378 | 0 | 0 | 0 | 0 | 1 | 1 | 1 | 0 | 0 | 0 | 0 | KX130864 |
| 1028 | VC_283_59 | 1 | 0 | 59742.00578 | 0 | 0 | 0 | 0 | 1 | 1 | 1 | 0 | 0 | 0 | 0 | OK272473 |
| 1029 | VC_283_6 | 1 | 0 | 56220.49696 | 0 | 0 | 0 | 0 | 1 | 1 | 1 | 0 | 0 | 0 | 0 | ON782582 |
| 1030 | VC_283_60 | 1 | 0 | 58212.43114 | 0 | 0 | 0 | 0 | 1 | 1 | 1 | 0 | 0 | 0 | 0 | OK272476 |
| 1031 | VC_283_61 | 1 | 0 | 60550.67538 | 0 | 0 | 0 | 0 | 1 | 1 | 1 | 0 | 0 | 0 | 0 | MT446387 |
| 1032 | VC_283_62 | 1 | 0 | 59954.74686 | 0 | 0 | 0 | 0 | 1 | 1 | 1 | 0 | 0 | 0 | 0 | OK272484 |
| 1033 | VC_283_63 | 1 | 0 | 57831.52963 | 0 | 0 | 0 | 0 | 1 | 1 | 1 | 0 | 0 | 0 | 0 | KT184312 |
| 1034 | VC_283_64 | 1 | 0 | 59467.34018 | 0 | 0 | 0 | 0 | 1 | 1 | 1 | 0 | 0 | 0 | 0 | MF158045 |
| 1035 | VC_283_65 | 1 | 0 | 59087.49413 | 0 | 0 | 0 | 0 | 1 | 1 | 1 | 0 | 0 | 0 | 0 | OL956808 |
| 1036 | VC_283_66 | 1 | 0 | 57447.13931 | 0 | 0 | 0 | 0 | 1 | 1 | 1 | 0 | 0 | 0 | 0 | KT184308 |
| 1037 | VC_283_67 | 2 | 300 | 109440.9693 | 0.00273371 | 0.092739974 | 7.745966692 | 7.745966692 | 1 | 1 | 1 | 7.745966692 | 0 | 0 | 0 | LR877332,MT515754 |
| 1038 | VC_283_68 | 1 | 0 | 60660.29529 | 0 | 0 | 0 | 0 | 1 | 1 | 1 | 0 | 0 | 0 | 0 | MW749001 |
| 1039 | VC_283_69 | 1 | 0 | 60308.17943 | 0 | 0 | 0 | 0 | 1 | 1 | 1 | 0 | 0 | 0 | 0 | HE956711 |
| 1040 | VC_283_7 | 1 | 0 | 58231.65418 | 0 | 0 | 0 | 0 | 1 | 1 | 1 | 0 | 0 | 0 | 0 | MW286157 |
| 1041 | VC_283_70 | 1 | 0 | 56516.76489 | 0 | 0 | 0 | 0 | 1 | 1 | 1 | 0 | 0 | 0 | 0 | MZ501098 |
| 1042 | VC_283_71 | 1 | 0 | 60240.29782 | 0 | 0 | 0 | 0 | 1 | 1 | 1 | 0 | 0 | 0 | 0 | LR215723 |
| 1043 | VC_283_72 | 1 | 0 | 61497.94227 | 0 | 0 | 0 | 0 | 1 | 1 | 1 | 0 | 0 | 0 | 0 | MT682710 |
| 1044 | VC_283_73 | 3 | 900 | 173836.8854 | 0.005150601 | 0.002546496 | 0 | 1 | 3 | 3 | 1 | 0.666666667 | 0 | 0 | 0 | LN881728,LN881733,LN881736 |
| 1045 | VC_283_74 | 1 | 0 | 58035.56058 | 0 | 0 | 0 | 0 | 1 | 1 | 1 | 0 | 0 | 0 | 0 | MT682714 |
| 1046 | VC_283_75 | 1 | 0 | 59072.10958 | 0 | 0 | 0 | 0 | 1 | 1 | 1 | 0 | 0 | 0 | 0 | MZ170040 |
| 1047 | VC_283_76 | 1 | 0 | 55588.98912 | 0 | 0 | 0 | 0 | 1 | 1 | 1 | 0 | 0 | 0 | 0 | MK373787 |
| 1048 | VC_283_77 | 2 | 300 | 120759.3369 | 0.002478124 | 0.045925809 | 1.732050808 | 1.732050808 | 1 | 1 | 1 | 1.732050808 | 0 | 0 | 0 | MT682712,MZ170041 |
| 1049 | VC_283_78 | 13 | 23400 | 645726.1312 | 0.034970985 | 7.73E-36 | 0 | 11.13552873 | 78 | 62 | 0.794871795 | 6.932772937 | 0 | 0 | 0 | MZ234027,MZ234030,MZ234034,MZ234036,MZ234037,MZ234039,MZ234040,MZ234041,MZ234042,MZ234043,MZ234051,MZ234052,MZ234053 |
| 1050 | VC_283_79 | 1 | 0 | 55462.94152 | 0 | 0 | 0 | 0 | 1 | 1 | 1 | 0 | 0 | 0 | 0 | MF001354 |
| 1051 | VC_283_8 | 1 | 0 | 58502.63356 | 0 | 0 | 0 | 0 | 1 | 1 | 1 | 0 | 0 | 0 | 0 | KU925172 |
| 1052 | VC_283_80 | 1 | 0 | 54613.17947 | 0 | 0 | 0 | 0 | 1 | 1 | 1 | 0 | 0 | 0 | 0 | MT764206 |
| 1053 | VC_283_81 | 1 | 0 | 55298.89282 | 0 | 0 | 0 | 0 | 1 | 1 | 1 | 0 | 0 | 0 | 0 | KR269718 |
| 1054 | VC_283_82 | 6 | 4500 | 342477.0607 | 0.012969157 | 2.03E-11 | 0 | 2.449489743 | 15 | 15 | 1 | 1.494635803 | 0 | 0 | 0 | KM606994,KM606995,KM606996,KM606997,KM606998,KM606999 |
| 1055 | VC_283_83 | 1 | 0 | 61796.8049 | 0 | 0 | 0 | 0 | 1 | 1 | 1 | 0 | 0 | 0 | 0 | KM607000 |
| 1056 | VC_283_84 | 1 | 0 | 62418.83314 | 0 | 0 | 0 | 0 | 1 | 1 | 1 | 0 | 0 | 0 | 0 | FJ839692 |
| 1057 | VC_283_85 | 1 | 0 | 60248.45679 | 0 | 0 | 0 | 0 | 1 | 1 | 1 | 0 | 0 | 0 | 0 | KX828711 |
| 1058 | VC_283_86 | 1 | 0 | 57985.64552 | 0 | 0 | 0 | 0 | 1 | 1 | 1 | 0 | 0 | 0 | 0 | ON528734 |
| 1059 | VC_283_87 | 1 | 0 | 60161.50195 | 0 | 0 | 0 | 0 | 1 | 1 | 1 | 0 | 0 | 0 | 0 | ON528738 |
| 1060 | VC_283_88 | 2 | 300 | 116873.7375 | 0.002560301 | 0.044034531 | 3.464101615 | 3.464101615 | 1 | 1 | 1 | 3.464101615 | 0 | 0 | 0 | AP018814,MH550421 |

| | | | | | | | | | | | | | | | | |
|---|---|---|---|---|---|---|---|---|---|---|---|---|---|---|---|---|
| 1061 | VC_283_89 | 2 | 300 | 110398.306 | 0.002710069 | 0.087352388 | 2.449489743 | 2.449489743 | 1 | 1 | 1 | 2.449489743 | 0 | 0 | 0 | MG781190,MG781191 |
| 1062 | VC_283_9 | 1 | 0 | 57254.40407 | 0 | 0 | 0 | 0 | 1 | 1 | 1 | 0 | 0 | 0 | 0 | MZ501052 |
| 1063 | VC_283_90 | 1 | 0 | 58872.61378 | 0 | 0 | 0 | 0 | 1 | 1 | 1 | 0 | 0 | 0 | 0 | KF208315 |
| 1064 | VC_283_91 | 11 | 16500 | 562694.5584 | 0.028487837 | 2.04E-35 | 0 | 7.071067812 | 55 | 55 | 1 | 3.669044122 | 0 | 0 | 0 | AF158101,HM137666,KJ477684,KJ477685,KJ477686,KM607002,KM607003,MT984581,MZ375356,OL964735,OL964745 |
| 1065 | VC_283_92 | 4 | 1800 | 225479.4939 | 0.007919764 | 4.18E-05 | 1.732050808 | 3.31662479 | 6 | 6 | 1 | 2.490266304 | 0 | 0 | 0 | AP018813,LC348380,MH751506,OM339528 |
| 1066 | VC_283_93 | 1 | 0 | 59566.85807 | 0 | 0 | 0 | 0 | 1 | 1 | 1 | 0 | 0 | 0 | 0 | MT932213 |
| 1067 | VC_283_94 | 2 | 300 | 113276.3023 | 0.002641396 | 0.047062377 | 0 | 0 | 1 | 1 | 1 | 0 | 0 | 0 | 0 | DQ904452,KM607001 |
| 1068 | VC_283_95 | 1 | 0 | 63444.99548 | 0 | 0 | 0 | 0 | 1 | 1 | 1 | 0 | 0 | 0 | 0 | JX128259 |
| 1069 | VC_283_96 | 1 | 0 | 62775.39109 | 0 | 0 | 0 | 0 | 1 | 1 | 1 | 0 | 0 | 0 | 0 | ON506924 |
| 1070 | VC_283_97 | 1 | 0 | 57778.2252 | 0 | 0 | 0 | 0 | 1 | 1 | 1 | 0 | 0 | 0 | 0 | LR215722 |
| 1071 | VC_283_98 | 1 | 0 | 56986.02756 | 0 | 0 | 0 | 0 | 1 | 1 | 1 | 0 | 0 | 0 | 0 | ON210139 |
| 1072 | VC_283_99 | 1 | 0 | 59553.26724 | 0 | 0 | 0 | 0 | 1 | 1 | 1 | 0 | 0 | 0 | 0 | KJ668714 |
| 1073 | VC_284_0 | 3 | 900 | 17299.91444 | 0.049450782 | 0.001390344 | 6.32455532 | 7.483314774 | 3 | 3 | 1 | 7.097061622 | 0 | 0 | 0 | KJ025957,MN095771,OM457001 |
| 1074 | VC_286_0 | 2 | 300 | 40832.54991 | 0.007293494 | 0.225370074 | 7.280109889 | 7.280109889 | 1 | 1 | 1 | 7.280109889 | 0 | 0 | 0 | MW512264,MZ501047 |
| 1075 | VC_286_1 | 21 | 63000 | 308533.0289 | 0.169567697 | 8.06E-32 | 0 | 9.949874371 | 210 | 190 | 0.904761905 | 7.509750767 | 0 | 0 | 0 | KR422352,LR027383,LR027386,LR027387,LR027390,LR027391,MH243438,MK327934,MK327943,MK373778,MK373782,MK373784,MK962754,MK962756,MK962757,MN850579,MN850590,MZ234025,OL960576,OL960577,OM960734 |
| 1076 | VC_286_10 | 9 | 10800 | 164735.8961 | 0.061525877 | 1.22E-06 | 7.071067812 | 9.433981132 | 36 | 24 | 0.666666667 | 8.603631143 | 0 | 0 | 0 | AY303349,KF562341,KM407600,KY703222,MH992122,MT446390,MZ501056,OM982647,ON528737 |
| 1077 | VC_286_11 | 3 | 900 | 60679.26371 | 0.01461531 | 0.06539716 | 6.8556546 | 8.888194417 | 3 | 3 | 1 | 8.191869961 | 0 | 0 | 0 | MH359124,MK524178,MW882907 |
| 1078 | VC_286_12 | 3 | 900 | 60572.96841 | 0.014640581 | 0.076717347 | 8.185352772 | 8.94427191 | 3 | 3 | 1 | 8.672606366 | 0 | 0 | 0 | MG589383,MZ189261,OL770074 |
| 1079 | VC_286_13 | 10 | 13500 | 179846.1995 | 0.06982294 | 7.09E-07 | 1.414213562 | 9.110433579 | 45 | 43 | 0.955555556 | 7.788539478 | 0 | 0 | 0 | KF582788,MF044457,MK639187,MT782071,MW630116,MZ311865,MZ560701,MZ868638,OV754623,OV877511 |
| 1080 | VC_286_14 | 1 | 0 | 21180.8957 | 0 | 0 | 0 | 0 | 1 | 1 | 1 | 0 | 0 | 0 | 0 | JX536493 |
| 1081 | VC_286_15 | 2 | 300 | 40797.75938 | 0.007299668 | 0.243736918 | 7.615773106 | 7.615773106 | 1 | 1 | 1 | 7.615773106 | 0 | 0 | 0 | MK047717,ON528725 |
| 1082 | VC_286_16 | 1 | 0 | 20764.01845 | 0 | 0 | 0 | 0 | 1 | 1 | 1 | 0 | 0 | 0 | 0 | KX130865 |
| 1083 | VC_286_17 | 8 | 8400 | 148648.1791 | 0.053486771 | 2.17E-05 | 0 | 9.433981132 | 28 | 17 | 0.607142857 | 7.943774458 | 0 | 0 | 0 | KP869101,KP869104,KP869111,KT184309,KT184310,MK047718,MN648445,MN850622 |
| 1084 | VC_286_2 | 1 | 0 | 20696.92053 | 0 | 0 | 0 | 0 | 1 | 1 | 1 | 0 | 0 | 0 | 0 | MN850651 |
| 1085 | VC_286_3 | 2 | 300 | 40730.30271 | 0.007311669 | 0.251141957 | 2 | 2 | 1 | 1 | 1 | 2 | 0 | 0 | 0 | MK373774,MK373775 |
| 1086 | VC_286_4 | 1 | 0 | 20834.35497 | 0 | 0 | 0 | 0 | 1 | 1 | 1 | 0 | 0 | 0 | 0 | MK327933 |
| 1087 | VC_286_5 | 1 | 0 | 20904.49631 | 0 | 0 | 0 | 0 | 1 | 1 | 1 | 0 | 0 | 0 | 0 | MT446423 |
| 1088 | VC_286_6 | 1 | 0 | 21137.69818 | 0 | 0 | 0 | 0 | 1 | 1 | 1 | 0 | 0 | 0 | 0 | MN864145 |
| 1089 | VC_286_7 | 1 | 0 | 25804.89913 | 0 | 0 | 0 | 0 | 1 | 1 | 1 | 0 | 0 | 0 | 0 | OM632671 |
| 1090 | VC_286_8 | 1 | 0 | 20691.83756 | 0 | 0 | 0 | 0 | 1 | 1 | 1 | 0 | 0 | 0 | 0 | KU255730 |
| 1091 | VC_286_9 | 1 | 0 | 22049.33248 | 0 | 0 | 0 | 0 | 1 | 1 | 1 | 0 | 0 | 0 | 0 | OM386662 |
| 1092 | VC_287_0 | 2 | 300 | 10443.76437 | 0.027923174 | 0.042285388 | 5.567764363 | 5.567764363 | 1 | 1 | 1 | 5.567764363 | 0 | 0 | 0 | MT457552,MT457553 |
| 1093 | VC_287_1 | 1 | 0 | 6203.789024 | 0 | 0 | 0 | 0 | 1 | 1 | 1 | 0 | 0 | 0 | 0 | KY971610 |
| 1094 | VC_288_0 | 3 | 900 | 20513.78926 | 0.042028993 | 0.001438912 | 5.656854249 | 6.244997998 | 3 | 3 | 1 | 5.882138298 | 0 | 0 | 0 | MW358927,MW385300,OK499989 |
| 1095 | VC_288_1 | 1 | 0 | 6930.472329 | 0 | 0 | 0 | 0 | 1 | 1 | 1 | 0 | 0 | 0 | 0 | KX078569 |
| 1096 | VC_288_2 | 2 | 300 | 12829.18591 | 0.022849855 | 0.044614412 | 2.828427125 | 2.828427125 | 1 | 1 | 1 | 2.828427125 | 0 | 0 | 0 | MG696114,MK295204 |
| 1097 | VC_288_3 | 3 | 900 | 18997.67191 | 0.045231422 | 0.001559747 | 5.916079783 | 8.717797887 | 3 | 3 | 1 | 7.258435366 | 0 | 0 | 0 | KP890823,MF001355,MW367898 |
| 1098 | VC_288_4 | 1 | 0 | 6606.855676 | 0 | 0 | 0 | 0 | 1 | 1 | 1 | 0 | 0 | 0 | 0 | MW057858 |
| 1099 | VC_28_0 | 2 | 232.4492088 | 7305.086568 | 0.030838886 | 0.042803361 | 1.732050808 | 1.732050808 | 1 | 1 | 1 | 1.732050808 | 0 | 0 | 0 | MW722521,MW790496 |
| 1100 | VC_28_1 | 53 | 133920.7211 | 53018.8005 | 0.716385278 | 0 | 0 | 9.746794345 | 1378 | 1285 | 0.932510885 | 7.38035055 | 0 | 0 | 0 | AB102868,AB362338,AB426868,AF217253,AF527608,AY052766,BK000583,CP051272,CP051278,CP051279,CP051285,CP051288,DQ003260,EU570103,FJ000341,GQ422450,GQ422451,GU573886,JF900176,JQ806763,KR296686,KR296687,KR296688,KU878967,KU927491,KU927492,KU927494,KU927495,KU927496,KU927498,KX776161,LC730322,LT714109,MF188997,MG407615,MG969415,MH370387,MK770414,MK770415,MK972685,MK972686,MK972687,MK972692,MK972693,MK972694,MN161573,MT774487,MT932212,MW013502,MW013503,MW355480,OP265889,OP377721 |
| 1101 | VC_290_0 | 10 | 13500 | 124159.6298 | 0.098067967 | 3.34E-24 | 1 | 8.774964387 | 45 | 45 | 1 | 7.474032872 | 0 | 0 | 0 | MT129652,MT446396,MT446411,MT446412,MT446415,MT446421,MT611523,MW233709,MW250787,MW598459 |
| 1102 | VC_290_1 | 8 | 8400 | 102585.303 | 0.075685697 | 1.77E-15 | 7.071067812 | 9.848857802 | 28 | 24 | 0.857142857 | 8.295226522 | 0 | 0 | 0 | GQ981382,KP007360,MT446420,MT446422,MT533174,MW269952,OK254198,OL451225 |
| 1103 | VC_290_2 | 2 | 300 | 29137.65366 | 0.01019103 | 0.076289934 | 8.124038405 | 8.124038405 | 1 | 1 | 1 | 8.124038405 | 0 | 0 | 0 | HM563683,KP007361 |
| 1104 | VC_290_3 | 2 | 300 | 29036.85924 | 0.010226044 | 0.077261572 | 8.888194417 | 8.888194417 | 1 | 1 | 1 | 8.888194417 | 0 | 0 | 0 | KP007362,MT446392 |
| 1105 | VC_290_4 | 2 | 300 | 32475.59243 | 0.009153153 | 0.096539322 | 8.366600265 | 8.366600265 | 1 | 1 | 1 | 8.366600265 | 0 | 0 | 0 | KP007359,OM386655 |
| 1106 | VC_290_5 | 14 | 27300 | 164639.3613 | 0.142232421 | 4.10E-42 | 5.567764363 | 9.486832981 | 91 | 84 | 0.923076923 | 7.823538534 | 0 | 0 | 0 | HM071924,HQ829472,KT176190,KU878968,KU878969,MH051917,MK234886,MT150133,MT682711,MW149274,MW679410,MZ561532,OK335775,OL780484 |
| 1107 | VC_290_6 | 12 | 19800 | 152479.5125 | 0.11492951 | 2.07E-30 | 1 | 9.899494937 | 66 | 54 | 0.818181818 | 7.862790764 | 0 | 0 | 0 | EF469154,EU863409,MH051913,MH791409,MK310184,MN850609,MN850621,MT129654,OL770107,OP292222,OV876907,OV877765 |
| 1108 | VC_290_7 | 1 | 0 | 15037.03231 | 0 | 0 | 0 | 0 | 1 | 1 | 1 | 0 | 0 | 0 | 0 | MK295203 |
| 1109 | VC_292_0 | 4 | 678.1388485 | 3364.723639 | 0.167737303 | 1.33E-05 | 0 | 8.660254038 | 6 | 6 | 1 | 6.62821126 | 0 | 0 | 0 | MK448704,MK448707,MK448709,MK448710 |
| 1110 | VC_293_0 | 1 | 0 | 1271.232262 | 0 | 0 | 0 | 0 | 1 | 1 | 1 | 0 | 0 | 0 | 0 | MK448750 |
| 1111 | VC_293_1 | 1 | 0 | 1560.18993 | 0 | 0 | 0 | 0 | 1 | 1 | 1 | 0 | 0 | 0 | 0 | MK448995 |
| 1112 | VC_293_2 | 2 | 106.5232503 | 3317.199015 | 0.031113286 | 0.042895506 | 7.211102551 | 7.211102551 | 1 | 1 | 1 | 7.211102551 | 0 | 0 | 0 | MK448740,MK448917 |
| 1113 | VC_293_3 | 19 | 10734.86464 | 30128.26795 | 0.262702929 | 3.00E-92 | 1 | 9.848857802 | 171 | 140 | 0.81871345 | 8.188746653 | 0 | 0 | 0 | MK448675,MK448676,MK448688,MK448690,MK448694,MK448772,MK448778,MK448782,MK448850,MK448859,MK448877,MK448931,MK448955,MK448962,MK448966,MK448973,MK448975,MK449004,OL774870 |
| 1114 | VC_294_0 | 2 | 237.0894915 | 2597.267606 | 0.083648419 | 0.042682242 | 0 | 0 | 1 | 1 | 1 | 0 | 0 | 0 | 0 | MK448832,MK449008 |
| 1115 | VC_295_0 | 2 | 53.03792305 | 1525.75293 | 0.033594015 | 0.055357686 | 8.717797887 | 8.717797887 | 1 | 1 | 1 | 8.717797887 | 0 | 0 | 0 | MK819239,ON857931 |
| 1116 | VC_296_0 | 6 | 1795.665816 | 1583.133157 | 0.531450918 | 3.90E-11 | 0 | 6.08276253 | 15 | 15 | 1 | 4.381177478 | 0 | 0 | 0 | MG592403,MG592404,MG592405,MG592551,MN592897,MW824406 |
| 1117 | VC_297_0 | 5 | 1436.999284 | 12172.88337 | 0.105584987 | 7.48E-08 | 4.898979486 | 9.433981132 | 10 | 9 | 0.9 | 7.68797595 | 0 | 0 | 0 | KM389210,KU199710,MW748993,MZ089728,ON778007 |
| 1118 | VC_297_1 | 20 | 27598.51446 | 21223.32415 | 0.565290355 | 3.33E-93 | 0 | 9.643650761 | 190 | 146 | 0.768421053 | 6.308565224 | 0 | 0 | 0 | HQ711984,JX403939,KU310943,MK511001,MK511002,MK511003,MK511004,MK511005,MK511006,MK511007,MK511008,MK511009,MK511010,MK511011,MK511012,MK511013,MK511014,MK511015,MK511016,MZ773939 |
| 1119 | VC_298_0 | 1 | 0 | 368.7083874 | 0 | 0 | 0 | 0 | 1 | 1 | 1 | 0 | 0 | 0 | 0 | KU708004 |
| 1120 | VC_298_1 | 1 | 0 | 475.2702627 | 0 | 0 | 0 | 0 | 1 | 1 | 1 | 0 | 0 | 0 | 0 | MW862109 |
| 1121 | VC_299_0 | 24 | 78018.47597 | 745.4675623 | 0.990535421 | 1.63E-67 | 0 | 4.472135955 | 276 | 276 | 1 | 1.837628284 | 0 | 0 | 0 | KX581090,KX581091,KX581092,KX581093,KX581094,KX581095,KX581096,KX581097,KX581098,KX581099,KX581100,KY658673,KY658674,KY658675,KY658676,KY658677,KY658678,KY658679,KY658680,MK672799,MK672800,MK672801,MK672803,MK672804 |
| 1122 | VC_29_0 | 10 | 5406.663895 | 4973.811783 | 0.520849339 | 5.96E-30 | 0 | 3 | 45 | 45 | 1 | 2.237413959 | 0 | 0 | 0 | AF157835,EU794049,LR794146,LR794147,LR794148,LR794149,LR794150,LR794151,LR794152,LR797923 |
| 1123 | VC_2_0 | 4 | 765.0153412 | 5071.903898 | 0.131064918 | 1.31E-05 | 1.414213562 | 4.358898944 | 6 | 6 | 1 | 3.489654876 | 0 | 0 | 0 | AB008550,MK034952,MK511066,Y13918 |
| 1124 | VC_300_0 | 3 | 123.2750423 | 888.7675128 | 0.121808161 | 0.00158299 | 6.164414003 | 9.219544457 | 3 | 2 | 0.666666667 | 8.052974283 | 0 | 0 | 0 | HQ141411,MG788324,OP650937 |
| 1125 | VC_301_0 | 2 | 22.37749083 | 150.856979 | 0.129174585 | 0.050697955 | 1.732050808 | 1.732050808 | 1 | 1 | 1 | 1.732050808 | 0 | 0 | 0 | LT985589,LT993247 |
| 1126 | VC_302_0 | 2 | 67.45895315 | 284.5212238 | 0.191655546 | 0.050257969 | 6.8556546 | 6.8556546 | 1 | 1 | 1 | 6.8556546 | 0 | 0 | 0 | MH837542,MH837543 |
| 1127 | VC_303_0 | 155 | 170519.0101 | 357.4815823 | 0.997907953 | 4.34E-56 | 0 | 2.645751311 | 11935 | 11935 | 1 | 1.325213926 | 0 | 0 | 0 | AF176027,AF176028,AF176029,AF176030,AF176031,AF176032,AF176033,AF176034,AF274751,AF299300,AF299301,AF299302,AF299303,AF299304,AF299305,AF299306,AF299307,AF299308,AF299309,AF299310,AF299311,AF299312,AF299313,AF299314,CP004084,DQ079880,DQ079881,DQ079882,DQ079883,DQ079884,DQ079885,DQ079886,DQ079887,DQ079888,DQ079889,DQ079890,DQ079891,DQ079892,DQ079893,DQ079894,DQ079895,EF380009,EF380010,EF380011,EF380012,EF380013,EF380014,EF380015,EF380016,EF380017,EF380018,EF380019,EF380020,EF380021,EF380022,EF380023,EF380024,EF380025,EF380026,EF380027,EF380028,EF380029,EF380030,EF380031,EF380032,FJ849058,GQ153912,GQ153913,GQ153914,GQ153915,GU385905,HM753658,HM753659,HM753660,HM753661,HM753662,HM753663,HM753664,HM753665,HM753666,HM753667,HM753668,HM753669,HM753670,HM753671,HM753672,HM753673,HM753674,HM753675,HM753676,HM753677,HM753678,HM753679,HM753680,HM753681,HM753682,HM753683,HM753684,HM753685,HM753686,HM753687,HM753688,HM753689,HM753690,HM753691,HM753692,HM753693,HM753694,HM753695,HM753696,HM753697,HM753698,HM753699,HM753700,HM753701,HM753702,HM753703,HM753704,HM753705,HM753706,HM753707,HM753708,HM753709,HM753710,HM753711,HM753712,HM753713,HM753714,HM753715,HM753716,HM753717,HM753718,HM775306,HM775307,HM775308,HM775309,HM775310,HM775311,JF719726,JF719727,JF719728,JQ764986,JQ764987,JQ764988,JQ764989,JQ764990,KJ997912,KY653237,MH378441,MH378442,MH378443,MN385565,MT947439,PX1CG,S13CG |
| 1128 | VC_304_0 | 11 | 6277.975218 | 1699.818151 | 0.786931289 | 7.13E-25 | 1 | 7.348469228 | 55 | 55 | 1 | 4.61864606 | 0 | 0 | 0 | AF189021,FJ867910,FJ867911,FJ867912,FJ867913,FJ867914,JQ809650,MF431615,MF431616,MF431617,MK613346 |
| 1129 | VC_305_0 | 12 | 3948.569345 | 2924.606861 | 0.574489759 | 2.86E-33 | 4.358898944 | 9.539392014 | 66 | 56 | 0.848484848 | 8.142157322 | 0 | 0 | 0 | GU557055,MK613343,MK613344,MK613345,MW514246,MW514247,MZ892987,MZ892988,MZ892989,MZ892990,MZ892991,MZ892992 |
| 1130 | VC_306_0 | 2 | 59.68224381 | 0 | 1 | 0 | 0 | 0 | 1 | 1 | 1 | 0 | 0 | 0 | 0 | AF191797,MW557853 |
| 1131 | VC_307_0 | 2 | 146.3359871 | 235.1980221 | 0.383546377 | 0.047485936 | 4.358898944 | 4.358898944 | 1 | 1 | 1 | 4.358898944 | 0 | 0 | 0 | AJ131519,MW476489 |
| 1132 | VC_308_0 | 4 | 602.3819499 | 599.1323936 | 0.501352275 | 2.58E-05 | 4.123105626 | 7.348469228 | 6 | 6 | 1 | 6.003764761 | 0 | 0 | 0 | AF195902,DQ320509,HE956704,KF767351 |
| 1133 | VC_309_0 | 3 | 51.47939396 | 355.5847171 | 0.126465077 | 0.005011038 | 7.681145748 | 8.185352772 | 3 | 3 | 1 | 7.913502131 | 0 | 0 | 0 | MF041990,MN830256,X98106 |
| 1134 | VC_30_0 | 2 | 172.3224131 | 4487.442202 | 0.036980927 | 0.042653646 | 5.477225575 | 5.477225575 | 1 | 1 | 1 | 5.477225575 | 0 | 0 | 0 | JQ182733,JQ182734 |
| 1135 | VC_30_1 | 16 | 7766.339179 | 31596.15724 | 0.197303014 | 9.00E-73 | 1.414213562 | 9.695359715 | 120 | 100 | 0.833333333 | 8.265124476 | 0 | 0 | 0 | AF335538,AF547987,CP000711,JF974339,JN600960,KJ832078,MH717096,MN340231,MT813197,MT833387,MZ043897,OM022251,ON470589,ON470594,ON470605,OV696608 |
| 1136 | VC_310_0 | 33 | 158395.828 | 22193.20244 | 0.877106586 | 9.48E-180 | 0 | 6.32455532 | 528 | 528 | 1 | 4.613391631 | 0 | 0 | 0 | AF222060,AY190604,KC292020,KC292022,MN901520,MN901521,MW450817,MZ334492,MZ334493,MZ334494,MZ334495,MZ334496,MZ334497,MZ334498,MZ334499,MZ334500,MZ334501,MZ334502,MZ334503,MZ334504,MZ334505,MZ334506,MZ334507,MZ334508,MZ334509,MZ334510,MZ334511,MZ334512,MZ334513,MZ334514,MZ334515,MZ334516,OK649958 |
| 1137 | VC_311_0 | 6 | 4477.772934 | 17942.8043 | 0.199717112 | 4.85E-11 | 0 | 4.582575695 | 15 | 15 | 1 | 3.552051462 | 0 | 0 | 0 | KC117376,KC292021,MZ334517,MZ334518,MZ334519,MZ334520 |
| 1138 | VC_312_0 | 2 | 13.80349458 | 435.2224777 | 0.030740971 | 0.045635733 | 7.681145748 | 7.681145748 | 1 | 1 | 1 | 7.681145748 | 0 | 0 | 0 | KC292026,MZ334525 |
| 1139 | VC_313_0 | 2 | 300 | 1443.237422 | 0.172093598 | 0.092556008 | 2.236067977 | 2.236067977 | 1 | 1 | 1 | 2.236067977 | 0 | 0 | 0 | KC292029,MZ334524 |
| 1140 | VC_313_1 | 3 | 900 | 1663.147163 | 0.351130834 | 0.001979817 | 0 | 6.782329983 | 3 | 3 | 1 | 4.521553322 | 0 | 0 | 0 | KC117377,KC292027,MZ334523 |
| 1141 | VC_314_0 | 2 | 6.936512835 | 31.43019879 | 0.180795084 | 1.68E-07 | 0 | 0 | 1 | 1 | 1 | 0 | 0 | 0 | 0 | DQ836063,JX625144 |
| 1142 | VC_315_0 | 12 | 457.8098471 | 53.88034078 | 0.894701243 | 1.12E-26 | 0 | 0 | 66 | 66 | 1 | 0 | 0 | 0 | 0 | AF227250,EF108464,EF108465,EF204939,EF204940,FJ483837,FJ483838,FJ483839,JQ966307,LT898437,MK213795,MT830620 |
| 1143 | VC_316_0 | 3 | 681.7498462 | 199.0749145 | 0.773990329 | 0.001795021 | 1.732050808 | 2 | 3 | 3 | 1 | 1.821367205 | 0 | 0 | 0 | MZ326866,MZ611865,ON711490 |
| 1144 | VC_317_0 | 18 | 16158.71709 | 32751.2685 | 0.330376648 | 3.34E-95 | 0 | 8.306623863 | 153 | 153 | 1 | 6.222691273 | 0 | 0 | 0 | MK448685,MK448763,MK448767,MK448771,MK448773,MK448774,MK448777,MK448786,MK448845,MK448853,MK448855,MK448942,MK448943,MK448963,MK448964,MK448969,MK448972,MK449012 |
| 1145 | VC_317_1 | 2 | 55.07230462 | 2905.006366 | 0.018605014 | 0.043428487 | 7.549834435 | 7.549834435 | 1 | 1 | 1 | 7.549834435 | 0 | 0 | 0 | MK448706,MK448721 |
| 1146 | VC_317_2 | 20 | 8886.168695 | 36092.56177 | 0.197563795 | 2.61E-63 | 1 | 10.44030651 | 190 | 130 | 0.684210526 | 8.081126215 | 0 | 0 | 0 | AY007505,FR671408,FR671409,KC348600,KC348601,KC348602,KT337360,KX077894,KY065485,KY065503,KY065504,MK448814,MK448881,MK448882,MK448896,MK448909,MK448988,MK448989,OL774869,OL774877 |
| 1147 | VC_317_3 | 1 | 0 | 1870.892515 | 0 | 0 | 0 | 0 | 1 | 1 | 1 | 0 | 0 | 0 | 0 | KP208803 |
| 1148 | VC_317_4 | 1 | 0 | 1849.04137 | 0 | 0 | 0 | 0 | 1 | 1 | 1 | 0 | 0 | 0 | 0 | MK448900 |
| 1149 | VC_318_0 | 3 | 188.2095086 | 4360.350714 | 0.041377821 | 0.014201475 | 4.472135955 | 8.888194417 | 3 | 3 | 1 | 7.378431587 | 0 | 0 | 0 | KY065466,KY065468,MK448982 |
| 1150 | VC_31_0 | 9 | 3330.646199 | 6365.977827 | 0.343485134 | 3.91E-24 | 0 | 9.695359715 | 36 | 25 | 0.694444444 | 7.898677711 | 0 | 0 | 0 | KY271395,KY271396,MK448231,MK448233,MK448235,MK714353,OK490410,OK490415,OK490416 |
| 1151 | VC_31_1 | 1 | 0 | 1126.729457 | 0 | 0 | 0 | 0 | 1 | 1 | 1 | 0 | 0 | 0 | 0 | OK499971 |
| 1152 | VC_31_2 | 1 | 0 | 1229.270654 | 0 | 0 | 0 | 0 | 1 | 1 | 1 | 0 | 0 | 0 | 0 | HQ201308 |
| 1153 | VC_323_0 | 2 | 70.92740272 | 2449.006464 | 0.028146533 | 0.043434336 | 7.810249676 | 7.810249676 | 1 | 1 | 1 | 7.810249676 | 0 | 0 | 0 | MK448821,MK448822 |
| 1154 | VC_323_1 | 9 | 3811.405081 | 17391.77352 | 0.179756307 | 5.55E-23 | 0 | 9.38083152 | 36 | 27 | 0.75 | 7.235635865 | 0 | 0 | 0 | KY065498,MK448735,MK448757,MK448758,MK448759,MK448893,MK448915,MK448939,OL774872 |
| 1155 | VC_324_0 | 2 | 131.5118847 | 508.490588 | 0.205486526 | 0.044855408 | 4.242640687 | 4.242640687 | 1 | 1 | 1 | 4.242640687 | 0 | 0 | 0 | S2-wms-combined_166665,S6-wms-combined_49909 |
| 1156 | VC_325_0 | 2 | 117.4771498 | 28496.99365 | 0.004105515 | 0.044255606 | 8.831760866 | 8.831760866 | 1 | 1 | 1 | 8.831760866 | 0 | 0 | 0 | MH513971,ON970586 |
| 1157 | VC_325_1 | 2 | 138.4512245 | 32281.57147 | 0.004270547 | 0.043366516 | 8.602325267 | 8.602325267 | 1 | 1 | 1 | 8.602325267 | 0 | 0 | 0 | JN398369,MN617845 |
| 1158 | VC_325_10 | 3 | 588.7649137 | 48324.42096 | 0.012036937 | 0.001408313 | 4.898979486 | 8.062257748 | 3 | 3 | 1 | 6.653745745 | 0 | 0 | 0 | MH744414,MK359331,MN119379 |
| 1159 | VC_325_11 | 1 | 0 | 14416.98793 | 0 | 0 | 0 | 0 | 1 | 1 | 1 | 0 | 0 | 0 | 0 | MH001458 |
| 1160 | VC_325_12 | 1 | 0 | 17081.72166 | 0 | 0 | 0 | 0 | 1 | 1 | 1 | 0 | 0 | 0 | 0 | MN813700 |
| 1161 | VC_325_13 | 2 | 275.2336694 | 33100.82066 | 0.008246441 | 0.042488001 | 4.242640687 | 4.242640687 | 1 | 1 | 1 | 4.242640687 | 0 | 0 | 0 | EU744249,EU744252 |
| 1162 | VC_325_14 | 3 | 457.6794697 | 48240.59259 | 0.009398269 | 0.001408411 | 8.185352772 | 8.717797887 | 3 | 3 | 1 | 8.521134899 | 0 | 0 | 0 | GU247132,KT818595,MT889384 |
| 1163 | VC_325_15 | 1 | 0 | 14217.71429 | 0 | 0 | 0 | 0 | 1 | 1 | 1 | 0 | 0 | 0 | 0 | KJ174156 |
| 1164 | VC_325_16 | 2 | 300 | 31166.24194 | 0.009534027 | 0.042486071 | 2.449489743 | 2.449489743 | 1 | 1 | 1 | 2.449489743 | 0 | 0 | 0 | MK310141,MK524523 |
| 1165 | VC_325_17 | 1 | 0 | 17582.00979 | 0 | 0 | 0 | 0 | 1 | 1 | 1 | 0 | 0 | 0 | 0 | KC701493 |
| 1166 | VC_325_18 | 1 | 0 | 15417.33981 | 0 | 0 | 0 | 0 | 1 | 1 | 1 | 0 | 0 | 0 | 0 | OL549190 |
| 1167 | VC_325_19 | 3 | 492.8467404 | 45344.75553 | 0.010752018 | 0.001407792 | 6 | 8.602325267 | 3 | 3 | 1 | 7.69586888 | 0 | 0 | 0 | JN020140,MH338238,MK967387 |
| 1168 | VC_325_2 | 1 | 0 | 16589.27191 | 0 | 0 | 0 | 0 | 1 | 1 | 1 | 0 | 0 | 0 | 0 | JF937099 |
| 1169 | VC_325_20 | 1 | 0 | 15090.15862 | 0 | 0 | 0 | 0 | 1 | 1 | 1 | 0 | 0 | 0 | 0 | MN585970 |
| 1170 | VC_325_21 | 5 | 1682.031399 | 78826.66127 | 0.020892544 | 2.44E-07 | 0 | 9.539392014 | 10 | 8 | 0.8 | 7.160281335 | 0 | 0 | 0 | EU744248,KX522649,KX522943,KX574454,MK524497 |
| 1171 | VC_325_22 | 1 | 0 | 14933.38509 | 0 | 0 | 0 | 0 | 1 | 1 | 1 | 0 | 0 | 0 | 0 | MK524499 |
| 1172 | VC_325_23 | 17 | 19144.41287 | 248067.595 | 0.071645032 | 3.39E-75 | 4.358898944 | 9.949874371 | 136 | 87 | 0.639705882 | 8.453323842 | 0 | 0 | 0 | JF937103,KC661273,KT259047,MG920060,MG925351,MH020244,MH450113,MH450130,MH576959,MH576971,MH576974,MK359299,MK820638,MT639654,MW507123,MW712732,ON970627 |
| 1173 | VC_325_24 | 3 | 476.5564668 | 52637.37397 | 0.008972344 | 0.001460512 | 7.681145748 | 8.94427191 | 3 | 3 | 1 | 8.350522477 | 0 | 0 | 0 | JF957057,MH230876,MN585976 |
| 1174 | VC_325_25 | 1 | 0 | 16863.18343 | 0 | 0 | 0 | 0 | 1 | 1 | 1 | 0 | 0 | 0 | 0 | MZ648034 |
| 1175 | VC_325_26 | 1 | 0 | 16396.86985 | 0 | 0 | 0 | 0 | 1 | 1 | 1 | 0 | 0 | 0 | 0 | MG944220 |
| 1176 | VC_325_27 | 2 | 136.0071932 | 30847.581 | 0.004389653 | 0.042490455 | 8.306623863 | 8.306623863 | 1 | 1 | 1 | 8.306623863 | 0 | 0 | 0 | MH536816,MH632118 |
| 1177 | VC_325_28 | 1 | 0 | 16171.82291 | 0 | 0 | 0 | 0 | 1 | 1 | 1 | 0 | 0 | 0 | 0 | KX369584 |
| 1178 | VC_325_29 | 3 | 539.9423373 | 46303.20516 | 0.011526602 | 0.001425585 | 4.795831523 | 8.602325267 | 3 | 3 | 1 | 7.153471513 | 0 | 0 | 0 | KP027196,KP027203,MW862983 |
| 1179 | VC_325_3 | 1 | 0 | 15866.13216 | 0 | 0 | 0 | 0 | 1 | 1 | 1 | 0 | 0 | 0 | 0 | KF493880 |
| 1180 | VC_325_30 | 4 | 1005.360273 | 69044.18897 | 0.014352131 | 1.38E-05 | 6.08276253 | 9.055385138 | 6 | 5 | 0.833333333 | 7.832372876 | 0 | 0 | 0 | KJ409696,KT326768,MK494116,OK040791 |
| 1181 | VC_325_31 | 1 | 0 | 17686.55402 | 0 | 0 | 0 | 0 | 1 | 1 | 1 | 0 | 0 | 0 | 0 | MK814754 |
| 1182 | VC_325_32 | 1 | 0 | 15269.28669 | 0 | 0 | 0 | 0 | 1 | 1 | 1 | 0 | 0 | 0 | 0 | MW435858 |
| 1183 | VC_325_33 | 4 | 814.7044672 | 53996.61883 | 0.014863799 | 1.44E-05 | 7.549834435 | 8.831760866 | 6 | 6 | 1 | 8.344148423 | 0 | 0 | 0 | KF560331,MK112532,MK359316,OM203162 |
| 1184 | VC_325_34 | 1 | 0 | 17246.96612 | 0 | 0 | 0 | 0 | 1 | 1 | 1 | 0 | 0 | 0 | 0 | ON677304 |
| 1185 | VC_325_35 | 17 | 20190.06617 | 255693.2912 | 0.073183342 | 2.04E-71 | 3.16227766 | 10.63014581 | 136 | 105 | 0.772058824 | 8.22596587 | 0 | 0 | 0 | EU826470,JN572689,JQ698665,JQ809703,KF024724,KF279415,KF279416,KM597531,KR080201,MF668283,MK359312,MK524506,MK524531,MN428054,MN586000,MT657342,MZ322004 |
| 1186 | VC_325_36 | 2 | 147.8237021 | 34212.86317 | 0.004302117 | 0.042488849 | 8.544003745 | 8.544003745 | 1 | 1 | 1 | 8.544003745 | 0 | 0 | 0 | MH371110,MN369760 |
| 1187 | VC_325_37 | 3 | 419.9137134 | 51783.24547 | 0.008043837 | 0.003650635 | 7.937253933 | 9.055385138 | 3 | 2 | 0.666666667 | 8.512214272 | 0 | 0 | 0 | MK061415,MN585985,MT776812 |
| 1188 | VC_325_38 | 6 | 2478.598114 | 90495.17242 | 0.026659111 | 2.79E-10 | 2.828427125 | 9.695359715 | 15 | 12 | 0.8 | 7.395816482 | 0 | 0 | 0 | KX683876,KX702320,MG099951,MH000606,MH651180,MT952847 |
| 1189 | VC_325_39 | 2 | 136.6321074 | 30523.98669 | 0.004456274 | 0.042491111 | 8.366600265 | 8.366600265 | 1 | 1 | 1 | 8.366600265 | 0 | 0 | 0 | AF271693,MZ355726 |
| 1190 | VC_325_4 | 1 | 0 | 15740.99088 | 0 | 0 | 0 | 0 | 1 | 1 | 1 | 0 | 0 | 0 | 0 | KX369586 |
| 1191 | VC_325_40 | 2 | 126.3905118 | 31657.62837 | 0.003976543 | 0.044256591 | 8.485281374 | 8.485281374 | 1 | 1 | 1 | 8.485281374 | 0 | 0 | 0 | KP027205,MH697581 |
| 1192 | VC_325_41 | 2 | 117.7833678 | 32923.66532 | 0.003564716 | 0.059288714 | 8.774964387 | 8.774964387 | 1 | 1 | 1 | 8.774964387 | 0 | 0 | 0 | JF937100,MK112530 |
| 1193 | VC_325_42 | 4 | 697.3816868 | 61478.73303 | 0.011216231 | 0.000195584 | 7.416198487 | 9.16515139 | 6 | 4 | 0.666666667 | 8.552617406 | 0 | 0 | 0 | JN699016,JX307705,KT363731,MN119377 |
| 1194 | VC_325_43 | 1 | 0 | 16009.35792 | 0 | 0 | 0 | 0 | 1 | 1 | 1 | 0 | 0 | 0 | 0 | KX712238 |
| 1195 | VC_325_44 | 3 | 721.8511321 | 45143.26067 | 0.015738567 | 0.00140829 | 3.741657387 | 5.916079783 | 3 | 3 | 1 | 5.014300659 | 0 | 0 | 0 | EU744251,JN660814,MH669011 |
| 1196 | VC_325_45 | 1 | 0 | 16839.78449 | 0 | 0 | 0 | 0 | 1 | 1 | 1 | 0 | 0 | 0 | 0 | MK524525 |
| 1197 | VC_325_46 | 2 | 135.5831604 | 33414.17532 | 0.004041256 | 0.042483835 | 8.485281374 | 8.485281374 | 1 | 1 | 1 | 8.485281374 | 0 | 0 | 0 | MH576975,MK967401 |
| 1198 | VC_325_47 | 2 | 127.1045418 | 28153.53666 | 0.004494401 | 0.042490803 | 8.366600265 | 8.366600265 | 1 | 1 | 1 | 8.366600265 | 0 | 0 | 0 | JN699000,MG962370 |
| 1199 | VC_325_48 | 2 | 117.2664193 | 29691.13608 | 0.003934005 | 0.049910593 | 8.426149773 | 8.426149773 | 1 | 1 | 1 | 8.426149773 | 0 | 0 | 0 | KY213952,MK112540 |
| 1200 | VC_325_49 | 4 | 886.5199348 | 62344.19895 | 0.014020399 | 4.04E-05 | 0 | 8.94427191 | 6 | 6 | 1 | 7.263079288 | 0 | 0 | 0 | AP018477,AP018478,KF416340,MH632117 |
| 1201 | VC_325_5 | 11 | 7704.904691 | 156370.1075 | 0.046959647 | 3.05E-34 | 6.403124237 | 9.949874371 | 55 | 42 | 0.763636364 | 8.509909372 | 0 | 0 | 0 | AY500152,JF937108,JF937110,KJ194585,KT626047,MG872833,MG925344,MG962372,MK359354,MN585999,MT310897 |
| 1202 | VC_325_50 | 4 | 1246.485376 | 67358.66735 | 0.018168976 | 1.22E-05 | 0 | 8.544003745 | 6 | 6 | 1 | 5.736322004 | 0 | 0 | 0 | MG812486,MG812489,MG812495,MG872836 |
| 1203 | VC_325_51 | 2 | 129.7220855 | 33340.9662 | 0.003875692 | 0.043361739 | 8.426149773 | 8.426149773 | 1 | 1 | 1 | 8.426149773 | 0 | 0 | 0 | KJ690250,MN585991 |
| 1204 | VC_325_52 | 5 | 1187.163243 | 79335.26088 | 0.014743263 | 9.27E-06 | 7.211102551 | 9.327379053 | 10 | 7 | 0.7 | 8.63969743 | 0 | 0 | 0 | JN153085,MH230878,MH697575,MN062710,MN234212 |
| 1205 | VC_325_53 | 7 | 2915.34069 | 117417.2785 | 0.024227352 | 1.13E-12 | 5.916079783 | 9.539392014 | 21 | 19 | 0.904761905 | 8.248639306 | 0 | 0 | 0 | KT388014,KX550443,MH020236,MH399782,MH590595,MK359300,MK359304 |

| | | | | | | | | | | | | | | | | |
|---|---|---|---|---|---|---|---|---|---|---|---|---|---|---|---|---|
| 1206 | VC_325_54 | 1 | 0 | 17015.92601 | 0 | 0 | 0 | 0 | 1 | 1 | 1 | 0 | 0 | 0 | 0 | KY224001 |
| 1207 | VC_325_55 | 2 | 181.3958628 | 29798.92568 | 0.006050498 | 0.042490047 | 7 | 7 | 1 | 1 | 1 | 7 | 0 | 0 | 0 | MK305893,MZ355722 |
| 1208 | VC_325_56 | 2 | 116.558312 | 32060.34917 | 0.003622421 | 0.053952016 | 9 | 9 | 1 | 0 | 0 | 9 | 0 | 0 | 0 | MF919508,MN585997 |
| 1209 | VC_325_57 | 1 | 0 | 17230.06883 | 0 | 0 | 0 | 0 | 1 | 1 | 1 | 0 | 0 | 0 | 0 | ON208829 |
| 1210 | VC_325_58 | 1 | 0 | 15914.13453 | 0 | 0 | 0 | 0 | 1 | 1 | 1 | 0 | 0 | 0 | 0 | ON970624 |
| 1211 | VC_325_59 | 1 | 0 | 15893.16976 | 0 | 0 | 0 | 0 | 1 | 1 | 1 | 0 | 0 | 0 | 0 | MT771340 |
| 1212 | VC_325_6 | 1 | 0 | 16105.01703 | 0 | 0 | 0 | 0 | 1 | 1 | 1 | 0 | 0 | 0 | 0 | MN444869 |
| 1213 | VC_325_60 | 2 | 125.0356719 | 32679.04259 | 0.003811589 | 0.042486995 | 8.660254038 | 8.660254038 | 1 | 1 | 1 | 8.660254038 | 0 | 0 | 0 | JN687951,MH338239 |
| 1214 | VC_325_61 | 8 | 4997.55715 | 121440.9436 | 0.039525596 | 1.66E-19 | 0 | 9.695359715 | 28 | 22 | 0.785714286 | 7.441945008 | 0 | 0 | 0 | JN153086,JN698998,MG925340,MG925342,MH450133,MH479919,MK310142,MT639644 |
| 1215 | VC_325_62 | 3 | 887.7068008 | 46311.90201 | 0.018807503 | 0.001408061 | 1.732050808 | 3.16227766 | 3 | 3 | 1 | 2.631442823 | 0 | 0 | 0 | OL455898,OM105884,OP021677 |
| 1216 | VC_325_7 | 3 | 411.0483533 | 47825.20079 | 0.008521565 | 0.002602184 | 8 | 9.219544457 | 3 | 2 | 0.666666667 | 8.664836282 | 0 | 0 | 0 | KR997929,KR997934,MG944221 |
| 1217 | VC_325_8 | 2 | 300 | 32088.76977 | 0.00926247 | 0.042487323 | 2 | 2 | 1 | 1 | 1 | 2 | 0 | 0 | 0 | KF024726,KP027204 |
| 1218 | VC_325_9 | 2 | 182.0366701 | 32716.06605 | 0.005533349 | 0.042490663 | 7.348469228 | 7.348469228 | 1 | 1 | 1 | 7.348469228 | 0 | 0 | 0 | AY500153,KT438500 |
| 1219 | VC_326_0 | 1 | 0 | 16.42479477 | 0 | 0 | 0 | 0 | 1 | 1 | 1 | 0 | 0 | 0 | 0 | MW202895 |
| 1220 | VC_327_0 | 12 | 807.2837481 | 97.27810752 | 0.892458313 | 4.46E-12 | 0 | 2 | 66 | 66 | 1 | 1.238268715 | 0 | 0 | 0 | AF306496,MH992158,MH992160,MH992177,MH992178,MH992189,MH992191,MH992194,MH992197,MH992207,MH992218,MW202535 |
| 1221 | VC_328_0 | 11 | 16500 | 18554.53226 | 0.470695198 | 7.57E-37 | 0 | 3 | 55 | 55 | 1 | 2.233767648 | 0 | 0 | 0 | EU710883,FQ482083,HQ728263,MG589384,MH426725,MK759884,MK770412,MN604407,OK483342,OM522317,ON706965 |
| 1222 | VC_329_0 | 2 | 263.4920828 | 51733.14663 | 0.005067483 | 0.04535418 | 8.185352772 | 8.185352772 | 1 | 1 | 1 | 8.185352772 | 0 | 0 | 0 | MG646672,MH586731 |
| 1223 | VC_329_1 | 17 | 34199.01442 | 360377.2376 | 0.086672764 | 1.42E-57 | 1.732050808 | 10.24695077 | 136 | 112 | 0.823529412 | 7.472644634 | 0 | 0 | 0 | AF320576,JF461087,JX181822,KP143762,KY626162,MF370225,MG251391,MG251392,MG969405,MG969406,MK965969,MK965970,MW006474,MW006475,MW006482,MZ327261,MZ327262 |
| 1224 | VC_329_10 | 1 | 0 | 24312.36389 | 0 | 0 | 0 | 0 | 1 | 1 | 1 | 0 | 0 | 0 | 0 | MN994498 |
| 1225 | VC_329_11 | 15 | 26960.41785 | 313734.9721 | 0.079133498 | 1.00E-62 | 0 | 9.273618495 | 105 | 98 | 0.933333333 | 7.981978837 | 0 | 0 | 0 | KP010413,MG646669,MG646670,MG646671,MN227145,MN481367,MN655999,MW311372,MW321605,MW362867,MZ150758,OL474141,OL800606,OL825705,ON239128 |
| 1226 | VC_329_12 | 2 | 186.965864 | 45589.79227 | 0.004084297 | 0.273945336 | 8.888194417 | 8.888194417 | 1 | 1 | 1 | 8.888194417 | 0 | 0 | 0 | MG065664,OK272472 |
| 1227 | VC_329_13 | 1 | 0 | 25806.9484 | 0 | 0 | 0 | 0 | 1 | 1 | 1 | 0 | 0 | 0 | 0 | KM657822 |
| 1228 | VC_329_14 | 1 | 0 | 24594.96613 | 0 | 0 | 0 | 0 | 1 | 1 | 1 | 0 | 0 | 0 | 0 | MG969404 |
| 1229 | VC_329_15 | 1 | 0 | 34625.44868 | 0 | 0 | 0 | 0 | 1 | 1 | 1 | 0 | 0 | 0 | 0 | GU169904 |
| 1230 | VC_329_16 | 4 | 1634.886699 | 102137.8843 | 0.015754486 | 1.93E-05 | 6.782329983 | 8.888194417 | 6 | 6 | 1 | 8.307422475 | 0 | 0 | 0 | KF055347,KX405003,MT409176,MW423797 |
| 1231 | VC_329_17 | 4 | 1610.40083 | 97479.13649 | 0.016251976 | 3.75E-05 | 2 | 8.888194417 | 6 | 6 | 1 | 6.821291528 | 0 | 0 | 0 | KM092515,MT322327,OK040806,OK040807 |
| 1232 | VC_329_18 | 7 | 5635.440122 | 166132.2487 | 0.032808499 | 4.90E-13 | 5.830951895 | 9.486832981 | 21 | 17 | 0.80952381 | 8.100208534 | 0 | 0 | 0 | JX560968,MK524176,MN882542,MT833282,MT887289,MW296865,ON540300 |
| 1233 | VC_329_19 | 1 | 0 | 25560.90188 | 0 | 0 | 0 | 0 | 1 | 1 | 1 | 0 | 0 | 0 | 0 | ON868915 |
| 1234 | VC_329_2 | 2 | 255.117896 | 51899.91047 | 0.00489153 | 0.044399149 | 8.366600265 | 8.366600265 | 1 | 1 | 1 | 8.366600265 | 0 | 0 | 0 | MN850631,MN850637 |
| 1235 | VC_329_20 | 4 | 1737.835029 | 98200.81716 | 0.017389018 | 1.22E-05 | 0 | 7.615773106 | 6 | 6 | 1 | 4.279291074 | 0 | 0 | 0 | LC739530,LC739531,LC739532,ON210138 |
| 1236 | VC_329_21 | 1 | 0 | 23252.27476 | 0 | 0 | 0 | 0 | 1 | 1 | 1 | 0 | 0 | 0 | 0 | MH051914 |
| 1237 | VC_329_22 | 5 | 2764.613608 | 122697.3371 | 0.022035474 | 6.49E-08 | 4.582575695 | 8.94427191 | 10 | 10 | 1 | 6.70791858 | 0 | 0 | 0 | MZ311863,OM386661,OM386663,OM386664,OM386665 |
| 1238 | VC_329_3 | 24 | 69063.49042 | 503433.1308 | 0.12063563 | 3.77E-140 | 6.557438524 | 10.04987562 | 276 | 201 | 0.72826087 | 8.604064902 | 0 | 0 | 0 | KX377933,LR880803,MK962758,MN850564,MN850566,MN850577,MN850580,MN850585,MN850603,MN850605,MN850610,MN850611,MN850614,MN850619,MN850633,MN850635,MN850636,MN850639,MN850644,MN850647,MZ501086,OK272490,OM630585,OX016465 |
| 1239 | VC_329_4 | 2 | 273.6694968 | 52415.17981 | 0.005194069 | 0.042535462 | 8.366600265 | 8.366600265 | 1 | 1 | 1 | 8.366600265 | 0 | 0 | 0 | JN225449,KR014248 |
| 1240 | VC_329_5 | 1 | 0 | 25567.08366 | 0 | 0 | 0 | 0 | 1 | 1 | 1 | 0 | 0 | 0 | 0 | MN994499 |
| 1241 | VC_329_6 | 3 | 838.9145994 | 77680.48507 | 0.01068417 | 0.001665311 | 5 | 8.485281374 | 3 | 3 | 1 | 7.182513041 | 0 | 0 | 0 | LR597658,MZ726791,MZ726792 |
| 1242 | VC_329_7 | 18 | 40984.24225 | 395811.3452 | 0.093829341 | 6.99E-90 | 1 | 10.04987562 | 153 | 119 | 0.777777778 | 7.869979305 | 0 | 0 | 0 | EU877232,KP869100,KP869106,KP869109,KP869110,KP869113,KT184313,KT184314,KT184315,KT184316,LC519489,LR699048,MH571750,MK482688,MK482689,MK482690,MT446388,MW481326 |
| 1243 | VC_329_8 | 2 | 242.1912643 | 49736.22644 | 0.004845917 | 0.043459863 | 8.485281374 | 8.485281374 | 1 | 1 | 1 | 8.485281374 | 0 | 0 | 0 | LC473039,MN994503 |
| 1244 | VC_329_9 | 1 | 0 | 25368.62508 | 0 | 0 | 0 | 0 | 1 | 1 | 1 | 0 | 0 | 0 | 0 | LR597663 |
| 1245 | VC_32_0 | 1 | 0 | 643.5305745 | 0 | 0 | 0 | 0 | 1 | 1 | 1 | 0 | 0 | 0 | 0 | MH443100 |
| 1246 | VC_330_0 | 27 | 102495.595 | 98965.79554 | 0.508760486 | 3.75E-212 | 0 | 9.591663047 | 351 | 304 | 0.866096866 | 6.130080127 | 0 | 0 | 0 | KM236239,KT001916,KT363872,KT454805,KT454806,KY654690,MF158040,MF158041,MF158043,MF158044,MF327003,MF327004,MF327005,MH791400,MH791413,MH920362,MK562502,MK562504,MK562505,MN342151,MN850587,OK539913,OM953433,ON528729,ON528730,ON528731,ON528732 |
| 1247 | VC_331_0 | 4 | 1800 | 18562.65746 | 0.088397107 | 1.23E-05 | 0 | 4.898979486 | 6 | 6 | 1 | 4.030908924 | 0 | 0 | 0 | MK290737,MK290738,MN270891,MN270892 |
| 1248 | VC_332_0 | 3 | 900 | 21002.73684 | 0.041090755 | 0.001412463 | 3.605551275 | 4.795831523 | 3 | 3 | 1 | 4.214674495 | 0 | 0 | 0 | MW749004,MW749010,OK283618 |
| 1249 | VC_333_0 | 25 | 29999.05292 | 34086.20443 | 0.468111609 | 1.52E-169 | 0 | 8.485281374 | 300 | 300 | 1 | 6.221382089 | 0 | 0 | 0 | FN391954,KT337342,KY065453,KY065454,KY065458,KY065459,KY065460,KY065461,KY065462,KY065463,KY065469,KY065470,KY065471,KY065494,KY065495,KY065499,MK448732,MK448739,MK448901,MK448912,MK448914,MK448918,MK448920,MT900487,OL774868 |
| 1250 | VC_334_0 | 1 | 0 | 802.8355074 | 0 | 0 | 0 | 0 | 1 | 1 | 1 | 0 | 0 | 0 | 0 | MK449000 |
| 1251 | VC_335_0 | 6 | 2297.296305 | 3949.529958 | 0.367754154 | 2.58E-11 | 0 | 5.656854249 | 15 | 15 | 1 | 3.141487628 | 0 | 0 | 0 | MK448804,MK448805,MK448807,MK448808,MK448815,MK448991 |
| 1252 | VC_337_0 | 6 | 1712.736173 | 1114.460314 | 0.605807266 | 1.15E-10 | 3.741657387 | 7.280109889 | 15 | 15 | 1 | 6.109691569 | 0 | 0 | 0 | MT980836,MT980837,MT980838,MT980839,MT980840,MT980841 |
| 1253 | VC_338_0 | 5 | 206.4548077 | 417.509107 | 0.330876198 | 1.31E-07 | 4.472135955 | 7.745966692 | 10 | 10 | 1 | 6.590364851 | 0 | 0 | 0 | S12-wms-combined_120133,S17-wms-combined_130215,S2-wms-combined_55495,S6-wms-combined_118484,S9-wms-combined_36459 |
| 1254 | VC_339_0 | 9 | 981.738133 | 62.08083222 | 0.940525288 | 2.29E-06 | 0 | 5 | 36 | 36 | 1 | 3.721984593 | 0 | 0 | 0 | AF323671,LT985572,LT985581,LT985584,LT985586,LT985587,LT985593,LT993227,LT996080 |
| 1255 | VC_33_0 | 4 | 369.5376562 | 5786.734801 | 0.060026202 | 5.17E-05 | 6.633249581 | 7.483314774 | 6 | 6 | 1 | 6.991144846 | 0 | 0 | 0 | HQ201307,JQ182729,MH178096,MN688132 |
| 1256 | VC_340_0 | 31 | 26290.12297 | 1240.327757 | 0.95494706 | 1.21E-54 | 0 | 8.831760866 | 465 | 465 | 1 | 7.151238168 | 0 | 0 | 0 | AF338467,AP013538,AP013539,AY939843,EF372997,GU071093,GU071100,GU071102,GU071104,GU071107,GU943054,HM559717,HQ316584,HQ317388,HQ317389,HQ332137,HQ332140,HQ337022,HQ633062,HQ634152,HQ634188,KC310802,KC310803,KC310804,KC310805,KC310806,MH540083,MK016662,MT424636,MW015080,MZ803112 |
| 1257 | VC_341_0 | 3 | 29.54426776 | 87.58841794 | 0.252229065 | 0.0603454 | 6.164414003 | 8.246211251 | 3 | 3 | 1 | 7.160564355 | 0 | 0 | 0 | AF396866,MW202489,OK085710 |
| 1258 | VC_342_0 | 2 | 54.63205894 | 245.9994971 | 0.1817243 | 0.065885926 | 8 | 8 | 1 | 1 | 1 | 8 | 0 | 0 | 0 | MN988526,MN988556 |
| 1259 | VC_344_0 | 21 | 63000 | 52890.80955 | 0.543615152 | 5.93E-76 | 0 | 7.348469228 | 210 | 210 | 1 | 4.629051515 | 0 | 0 | 0 | AF399011,AP019418,JX233784,KU521356,LC727695,LC727696,LC727699,LC727701,MF805716,MN563784,MN871480,MN871489,MT133560,MT995926,MT995927,MT995928,MW057919,MZ444140,ON631220,OP310983,OP310984 |
| 1260 | VC_344_1 | 3 | 900 | 24157.56909 | 0.035917291 | 0.066511228 | 2.828427125 | 3.464101615 | 3 | 3 | 1 | 3.040318622 | 0 | 0 | 0 | MK599315,MN103543,OU343167 |
| 1261 | VC_344_2 | 1 | 0 | 7196.500975 | 0 | 0 | 0 | 0 | 1 | 1 | 1 | 0 | 0 | 0 | 0 | MF063068 |
| 1262 | VC_344_3 | 1 | 0 | 8153.192185 | 0 | 0 | 0 | 0 | 1 | 1 | 1 | 0 | 0 | 0 | 0 | MK552327 |
| 1263 | VC_344_4 | 1 | 0 | 8072.992379 | 0 | 0 | 0 | 0 | 1 | 1 | 1 | 0 | 0 | 0 | 0 | EU197055 |
| 1264 | VC_344_5 | 1 | 0 | 8311.146891 | 0 | 0 | 0 | 0 | 1 | 1 | 1 | 0 | 0 | 0 | 0 | MF042360 |
| 1265 | VC_344_6 | 1 | 0 | 8637.082971 | 0 | 0 | 0 | 0 | 1 | 1 | 1 | 0 | 0 | 0 | 0 | HQ630627 |
| 1266 | VC_345_0 | 2 | 300 | 4078.759555 | 0.068512554 | 0.043458474 | 3.605551275 | 3.605551275 | 1 | 1 | 1 | 3.605551275 | 0 | 0 | 0 | AP017924,AP017925 |
| 1267 | VC_346_0 | 4 | 1800 | 2107.678745 | 0.46063152 | 2.09E-05 | 0 | 3.464101615 | 6 | 6 | 1 | 1.732050808 | 0 | 0 | 0 | MK308674,MK308675,MK308676,MK308677 |
| 1268 | VC_346_1 | 2 | 300 | 2122.465727 | 0.123840761 | 0.046465516 | 2.828427125 | 2.828427125 | 1 | 1 | 1 | 2.828427125 | 0 | 0 | 0 | MT811961,OM319461 |
| 1269 | VC_346_2 | 1 | 0 | 1337.595263 | 0 | 0 | 0 | 0 | 1 | 1 | 1 | 0 | 0 | 0 | 0 | OV032902 |
| 1270 | VC_346_3 | 1 | 0 | 1409.942802 | 0 | 0 | 0 | 0 | 1 | 1 | 1 | 0 | 0 | 0 | 0 | MH595538 |
| 1271 | VC_347_0 | 3 | 900 | 4924.541563 | 0.154518599 | 0.001555396 | 6.403124237 | 6.782329983 | 3 | 3 | 1 | 6.580964248 | 0 | 0 | 0 | LR881104,MN871444,MN871445 |
| 1272 | VC_348_0 | 22 | 19891.82467 | 825.1353317 | 0.960171023 | 7.82E-21 | 1 | 7.810249676 | 231 | 231 | 1 | 5.56966756 | 0 | 0 | 0 | AJ344259,AJ414696,AJ748296,AJ875026,FM164764,KP282675,KX712143,KY744228,KY744229,KY744230,KY744231,KY744232,KY744233,KY744234,KY744235,MK064565,MN876841,MN876842,MN876843,OM401712,OM401713,OM401714 |
| 1273 | VC_349_0 | 7 | 2570.327721 | 174.7307766 | 0.936347157 | 3.52E-08 | 0 | 8.660254038 | 21 | 21 | 1 | 6.029744064 | 0 | 0 | 0 | AF440571,AM087120,AM087121,AM087122,AM087123,EU545650,KX467643 |
| 1274 | VC_34_0 | 4 | 1015.147178 | 1318.866937 | 0.434936178 | 1.94E-05 | 1.732050808 | 4.358898944 | 6 | 6 | 1 | 3.203663739 | 0 | 0 | 0 | AY374448,CP038007,CP072507,KY271402 |
| 1275 | VC_350_0 | 2 | 78.44058844 | 163.6877283 | 0.323962887 | 0.061345549 | 3.605551275 | 3.605551275 | 1 | 1 | 1 | 3.605551275 | 0 | 0 | 0 | AJ854042,MK064564 |
| 1276 | VC_351_0 | 3 | 122.3463943 | 15.13147339 | 0.889935204 | 0.018444213 | 5.477225575 | 7.141428429 | 3 | 3 | 1 | 6.062139455 | 0 | 0 | 0 | AM419438,HQ332141,MW344765 |
| 1277 | VC_352_0 | 3 | 672.0697793 | 247.8105226 | 0.730605686 | 0.011327247 | 1 | 6.557438524 | 3 | 3 | 1 | 4.679393074 | 0 | 0 | 0 | AF440695,MK002701,MK450543 |
| 1278 | VC_353_0 | 6 | 2137.634998 | 1784.506164 | 0.545017354 | 2.23E-11 | 0 | 7.874007874 | 15 | 15 | 1 | 6.188235649 | 0 | 0 | 0 | AF447491,AY453853,CP000625,OK095358,OM892879,OP293079 |
| 1279 | VC_354_0 | 4 | 647.2701708 | 3053.520662 | 0.174900501 | 1.32E-05 | 7.549834435 | 9.055385138 | 6 | 5 | 0.833333333 | 8.230053652 | 0 | 0 | 0 | MN512538,MN812722,MT227924,OP125534 |
| 1280 | VC_355_0 | 2 | 14.28598213 | 264.1878418 | 0.051300987 | 0.065816124 | 7.280109889 | 7.280109889 | 1 | 1 | 1 | 7.280109889 | 0 | 0 | 0 | MT121967,MW651859 |
| 1281 | VC_356_0 | 2 | 52.50222215 | 211.042249 | 0.199215798 | 0.067889051 | 6.8556546 | 6.8556546 | 1 | 1 | 1 | 6.8556546 | 0 | 0 | 0 | HQ634187,MK105819 |
| 1282 | VC_357_0 | 53 | 125880.8187 | 14320.20516 | 0.897859482 | 0 | 0 | 6.633249581 | 1378 | 1378 | 1 | 2.523892394 | 0 | 0 | 0 | MT740734,ON170024,ON170027,ON170031,ON170052,ON170057,ON170058,ON170068,ON170071,ON170075,ON170077,ON170079,ON170129,ON170133,ON170134,ON170136,ON170150,ON170172,ON170175,ON170179,ON170181,ON170182,ON170185,ON170187,ON170208,ON170214,ON170228,ON170229,ON170232,ON170234,ON170236,ON170238,ON170240,ON170242,ON170254,ON170257,ON170262,ON170264,ON170265,ON170267,ON170275,ON170279,ON170286,ON170287,ON170303,ON170317,ON170323,ON170324,ON170326,ON170327,ON170343,ON170355,ON170359 |
| 1283 | VC_358_0 | 3 | 146.9906332 | 138.6304012 | 0.514635183 | 0.023427089 | 0 | 0 | 3 | 3 | 1 | 0 | 0 | 0 | 0 | ON170149,ON170171,ON170322 |
| 1284 | VC_359_0 | 7 | 2542.15543 | 4323.533868 | 0.370269513 | 4.92E-14 | 0 | 8.774964387 | 21 | 21 | 1 | 6.84867358 | 0 | 0 | 0 | JN585957,MF612073,MH426726,MK618715,MT251347,MT251348,MW722080 |
| 1285 | VC_35_0 | 6 | 1238.457604 | 4255.358782 | 0.225427557 | 6.80E-11 | 5.916079783 | 9.327379053 | 15 | 14 | 0.933333333 | 7.844729837 | 0 | 0 | 0 | KY271399,MK416008,MK416013,MK416022,MK448234,MK448237 |
| 1286 | VC_35_1 | 1 | 0 | 780.8240063 | 0 | 0 | 0 | 0 | 1 | 1 | 1 | 0 | 0 | 0 | 0 | OL617042 |
| 1287 | VC_360_0 | 358 | 933940.8583 | 37349.42269 | 0.961546591 | 0 | 0 | 2.449489743 | 63903 | 63903 | 1 | 1.242094692 | 0 | 0 | 0 | AF454431,AY751298,DQ079896,DQ079897,DQ079899,DQ079900,DQ079901,DQ079902,DQ079903,DQ079904,DQ079906,DQ079908,DQ079909,GQ153920,GQ153922,GQ153923,JF719729,JF719730,JF719731,KX266300,KX266301,KX266302,KX266303,KX266304,KX266305,KX266306,KX266307,KX266308,KX266309,KX266310,KX266311,KX266312,KX266313,KX266314,KX266315,KX266316,KX266317,KX266318,KX266319,KX266320,KX266321,KX266322,KX266323,KX266324,KX266325,KX266326,KX266327,KX266328,KX266329,KX266330,KX266331,KX266332,KX266333,KX266334,KX266335,KX266336,KX266337,KX266338,KX266339,KX266340,KX266341,KX266342,KX266343,KX266344,KX266345,KX266346,KX266347,KX266348,KX266349,KX266350,KX266351,KX266352,KX266353,KX266354,KX266355,KX266356,KX266357,KX266358,KX266359,KX266360,KX266361,KX266362,KX266363,KX266364,KX266365,KX266366,KX266367,KX266368,KX266369,KX266370,KX266371,KX266372,KX266373,KX266374,KX266375,KX266376,KX266377,KX266378,KX266379,KX266380,KX266381,KX266382,KX266383,KX266384,KX266385,KX266386,KX266387,KX266388,KX266389,KX266390,KX266391,KX266392,KX266393,KX266394,KX266395,KX266396,KX266397,KX266398,KX266399,KX266400,KX266401,KX266402,KX266403,KX266404,KX266405,KX266406,KX266407,KX266408,KX266409,KX266410,KX266411,KX266412,KX266413,KX266414,KX266415,KX266416,KX266417,KX266418,KX266419,KX266420,KX266421,KX266422,KX266423,KX266424,KX266425,KX266426,KX266427,KX266428,KX266429,KX266430,KX266431,KX266432,KX266433,KX266434,KX266435,KX266436,KX266437,KX266438,KX266439,KX266440,KX266441,KX266442,KX266443,KX266444,KX266445,KX266446,KX266447,KX266448,KX266449,KX266450,KX266451,KX266452,KX266453,KX266454,KX266455,KX266456,KX266457,KX266458,KX266459,KX266460,KX266461,KX266462,KX266463,KX266464,KX266465,KX266466,KX266467,KX266468,KX266469,KX266470,KX266471,KX266472,KX266473,KX266474,KX266475,KX266476,KX266477,KX266478,KX266479,KX266480,KX266481,KX266482,KX266483,KX266484,KX266485,KX266486,KX266487,KX266488,KX266489,KX266490,KX266491,KX266492,KX266493,KX266494,KX266495,KX266496,KX266497,KX266498,KX266499,KX266500,KX266501,KX266502,KX266503,KX266504,KX266505,KX266506,KX266507,KX266508,KX266509,KX266510,KX266511,KX266512,KX266513,KX266514,KX266515,KX266516,KX266517,KX266518,KX266519,KX266520,KX266521,KX266522,KX266523,KX266524,KX266525,KX266526,KX266527,KX266528,KX266529,KX266530,KX266531,KX266532,KX266533,KX266534,KX266535,KX266536,KX266537,KX266538,KX266539,KX266540,KX266541,KX266542,KX266543,KX266544,KX266545,KX266546,KX266547,KX266548,KX266549,KX266550,KX266551,KX266552,KX266553,KX266554,KX266555,KX266556,KX266557,KX266558,KX266559,KX266560,KX266561,KX266562,KX266563,KX266564,KX266565,KX266566,KX266567,KX266568,KX266569,KX266570,KX266571,KX266572,KX266573,KX266574,KX266575,KX266576,KX266577,KX266578,KX266579,KX266580,KX266581,KX266582,KX266583,KX266584,KX266585,KX266586,KX266587,KX266588,KX266589,KX266590,KX266591,KX266592,KX266593,KX266594,KX266595,KX266596,KX266597,KX266598,KX266599,KX266600,KX266601,KX266602,KX266603,KX266604,KX266605,KX266606,KX266607,KX266608,KX266609,KX266610,KX266611,KX266612,KX266613,KX266614,KX266615,KX266616,KX266617,KX266618,KX266619,KX266620,KX266621,KX266622,KX266623,KX266624,KX266625,KX266626,KX266627,KX266628,KX266629,KX266630,KX266631,KX266632,KX266633,KX266634,KX266635,KX266636,KX266637,MG563770 |
| 1288 | VC_362_0 | 2 | 9.319160538 | 4638.912396 | 0.002004883 | 0.166922652 | 2 | 2 | 1 | 1 | 1 | 2 | 0 | 0 | 0 | GQ153921,KC237308 |
| 1289 | VC_363_0 | 2 | 28.83930013 | 7.11248825 | 0.80216594 | 0.270145687 | 0 | 0 | 1 | 1 | 1 | 0 | 0 | 0 | 0 | OK338687,OM055665 |
| 1290 | VC_364_0 | 16 | 23058.60561 | 12465.34109 | 0.649100332 | 4.60E-59 | 0 | 7.810249676 | 120 | 120 | 1 | 3.926788423 | 0 | 0 | 0 | AP008986,KY210139,MG944227,MG944228,MG944229,MG944230,MG944231,MG944232,MG944233,MG944234,MG944235,MG944236,MN461279,OK275494,ON996340,OP067662 |
| 1291 | VC_369_0 | 6 | 2622.442153 | 1804.123778 | 0.592432643 | 1.77E-10 | 0 | 8.062257748 | 15 | 15 | 1 | 5.46937234 | 0 | 0 | 0 | JQ177063,KY290953,KY290954,MH179474,MK804891,MK804892 |
| 1292 | VC_371_0 | 2 | 178.4560001 | 56108.12328 | 0.003170489 | 0.052680276 | 8.717797887 | 8.717797887 | 1 | 1 | 1 | 8.717797887 | 0 | 0 | 0 | MT897903,ON456342 |
| 1293 | VC_371_1 | 11 | 10733.0163 | 272398.5923 | 0.037908224 | 3.31E-34 | 0 | 10.04987562 | 55 | 47 | 0.854545455 | 7.96321926 | 0 | 0 | 0 | KJ595576,KM347890,KR997967,KY006474,MH926060,MK279910,MK524512,MK524526,MN585967,MN586058,MW534377 |
| 1294 | VC_371_10 | 1 | 0 | 26257.54059 | 0 | 0 | 0 | 0 | 1 | 1 | 1 | 0 | 0 | 0 | 0 | MH590594 |
| 1295 | VC_371_11 | 31 | 82352.98288 | 806460.386 | 0.092654978 | 3.56E-148 | 0 | 10.63014581 | 465 | 326 | 0.701075269 | 8.527191855 | 0 | 0 | 0 | KX592589,MF155947,MF668276,MF919528,MF919539,MG757159,MH230874,MH513980,MH590588,MK112539,MK112546,MK112551,MK279843,MK279865,MK279870,MK279873,MK279878,MK279894,MK279895,MK279897,MN369744,MT310868,MT316460,MT316463,MW534381,MW712720,MW862980,MW862993,MZ648038,ON637769,OP434459 |
| 1296 | VC_371_12 | 4 | 1262.830498 | 99281.29446 | 0.012559963 | 1.51E-05 | 4.242640687 | 9.055385138 | 6 | 5 | 0.833333333 | 7.528073504 | 0 | 0 | 0 | MK112544,MT310867,MT952849,ON456332 |
| 1297 | VC_371_13 | 33 | 106710.7158 | 489555.7132 | 0.178964823 | 0 | 0 | 10.09950494 | 528 | 463 | 0.876893939 | 7.685665692 | 0 | 0 | 0 | JN192463,KM363597,KR029086,KR029087,KX576645,KX576646,KX576647,KX702319,KY385380,KY385382,KY385383,MF919511,MG757164,MG757165,MG944223,MG962363,MH051251,MH155874,MH316568,MH371107,MH479916,MH479918,MH513973,MH576958,MK279850,MK279858,MK279864,MK279883,MK279890,MK279891,MN585965,MN945903,ON456330 |
| 1298 | VC_371_14 | 1 | 0 | 26472.93987 | 0 | 0 | 0 | 0 | 1 | 1 | 1 | 0 | 0 | 0 | 0 | MN369738 |
| 1299 | VC_371_15 | 44 | 166408.6136 | 1030233.935 | 0.139062925 | 0 | 4.123105626 | 10.19803903 | 946 | 707 | 0.747357294 | 8.47829338 | 0 | 0 | 0 | GQ303264,GQ303266,JF704103,JF704109,JF937097,JF957056,JN006063,JN638752,JN638753,JN699008,JN699010,KM408320,KP027208,KT599441,KU867907,KX578071,KX670813,KX683292,KX683293,KY676783,MG770212,MG925346,MG925350,MG944225,MH399775,MH399786,MH576970,MH651184,MH825702,MH825705,MK112527,MK112536,MK112555,MK279855,MK279859,MK279861,MK279866,MK279879,MK279881,MK279882,MK279885,MK279887,MK279888,MW712726 |
| 1300 | VC_371_16 | 42 | 154118.0091 | 994987.1284 | 0.134120024 | 1.58E-278 | 0 | 10.19803903 | 861 | 627 | 0.728222997 | 8.472045595 | 0 | 0 | 0 | AF547430,DQ398046,FJ174694,GU247133,GU247134,JF704102,JN699009,JX649096,JX649097,JX649100,KJ194580,KJ194583,KX369585,KX620786,MG757158,MG925348,MG962365,MH371114,MH371117,MH371125,MH399773,MH399780,MH450116,MH450122,MH479914,MH479921,MH651175,MH651186,MH727555,MH744417,MH779516,MK112542,MK112553,MK279846,MK279852,MK279871,MK494104,MN945899,MT897909,MW534379,MW534380,MZ622173 |
| 1301 | VC_371_17 | 3 | 668.3153086 | 87629.97615 | 0.007568836 | 0.001883237 | 0 | 8.366600265 | 3 | 3 | 1 | 5.57773351 | 0 | 0 | 0 | JF937095,JN698990,JN699017 |
| 1302 | VC_371_18 | 10 | 8923.016739 | 286616.4264 | 0.030192304 | 7.83E-23 | 0 | 9.433981132 | 45 | 36 | 0.8 | 7.681119688 | 0 | 0 | 0 | JF704091,JF937109,JN698989,KJ194579,KP027197,KP027209,KR816508,KX576643,KY965066,MT897901 |
| 1303 | VC_371_19 | 1 | 0 | 28762.5234 | 0 | 0 | 0 | 0 | 1 | 1 | 1 | 0 | 0 | 0 | 0 | MZ622172 |
| 1304 | VC_371_2 | 8 | 5309.103892 | 223490.6376 | 0.023204152 | 8.79E-17 | 6.32455532 | 9.486832981 | 28 | 24 | 0.857142857 | 8.052410028 | 0 | 0 | 0 | KC576784,KJ567044,MG925356,MG962375,MH077582,MK112552,MK279889,MN096366 |
| 1305 | VC_371_20 | 1 | 0 | 29066.00238 | 0 | 0 | 0 | 0 | 1 | 1 | 1 | 0 | 0 | 0 | 0 | MN703415 |
| 1306 | VC_371_21 | 2 | 167.8782629 | 50758.67292 | 0.003296478 | 0.042292629 | 8.888194417 | 8.888194417 | 1 | 1 | 1 | 8.888194417 | 0 | 0 | 0 | JN006064,KR781349 |
| 1307 | VC_371_3 | 22 | 43298.56312 | 507088.4314 | 0.078669306 | 5.02E-136 | 5.477225575 | 9.695359715 | 231 | 209 | 0.904761905 | 8.015235961 | 0 | 0 | 0 | GQ303259,JF704099,JX649098,JX649099,KC661274,KC691258,KF713485,KJ174157,KJ538723,KM236502,MF919503,MF919507,MF919523,MF919530,MG925339,MG962362,MH230875,MH779500,MK279877,MK279908,MN444868,MT897902 |
| 1308 | VC_371_4 | 5 | 1880.778604 | 137693.1926 | 0.013475139 | 2.86E-07 | 0 | 8.888194417 | 10 | 10 | 1 | 7.618358723 | 0 | 0 | 0 | KT364588,MH576954,MH576967,MH651177,MK279904 |
| 1309 | VC_371_5 | 2 | 179.2875557 | 51487.76121 | 0.003470056 | 0.042289286 | 8.602325267 | 8.602325267 | 1 | 1 | 1 | 8.602325267 | 0 | 0 | 0 | MH051264,MK310139 |
| 1310 | VC_371_6 | 1 | 0 | 20520.99468 | 0 | 0 | 0 | 0 | 1 | 1 | 1 | 0 | 0 | 0 | 0 | MF919519 |
| 1311 | VC_371_7 | 1 | 0 | 25721.24865 | 0 | 0 | 0 | 0 | 1 | 1 | 1 | 0 | 0 | 0 | 0 | ON970594 |
| 1312 | VC_371_8 | 1 | 0 | 24054.69344 | 0 | 0 | 0 | 0 | 1 | 1 | 1 | 0 | 0 | 0 | 0 | MZ648041 |
| 1313 | VC_371_9 | 4 | 1095.139234 | 99094.75905 | 0.010930635 | 2.34E-05 | 6.244997998 | 9.38083152 | 6 | 4 | 0.666666667 | 8.31236511 | 0 | 0 | 0 | MT776813,MT889369,MZ681520,ON970577 |
| 1314 | VC_372_0 | 24 | 79908.38324 | 64356.26351 | 0.553901354 | 7.53E-144 | 0 | 6.244997998 | 276 | 276 | 1 | 3.627152774 | 0 | 0 | 0 | DQ398044,EU770221,JF704094,JF704104,JN699011,KF279411,KF279412,KF279413,KF493881,KR080194,KR080200,KT270441,KX589269,MH513970,MK524486,MN234171,MN369759,MN428050,MT310870,MT639648,MT818419,MT897910,MW435857,ON970575 |
| 1315 | VC_372_1 | 9 | 9132.603158 | 37934.01294 | 0.194035686 | 3.62E-24 | 0 | 8.246211251 | 36 | 36 | 1 | 5.080135532 | 0 | 0 | 0 | JN699007,KF024727,KF416341,KR080199,KY224000,MH727561,MK524490,MN284893,MT889381 |
| 1316 | VC_372_2 | 5 | 1779.442795 | 22945.72888 | 0.071968875 | 3.18E-08 | 5.656854249 | 9.055385138 | 10 | 9 | 0.9 | 7.571257399 | 0 | 0 | 0 | MH744423,MK059749,MN617843,MW055906,OL455893 |
| 1317 | VC_372_3 | 5 | 2966.428379 | 23298.26406 | 0.11294358 | 3.30E-08 | 0 | 4.472135955 | 10 | 10 | 1 | 3.779188896 | 0 | 0 | 0 | KF024729,KJ433973,KJ433974,KJ433975,KJ433976 |
| 1318 | VC_372_4 | 3 | 523.2325138 | 15340.53732 | 0.032982861 | 0.001467429 | 6.633249581 | 8.831760866 | 3 | 3 | 1 | 7.983430607 | 0 | 0 | 0 | MN813693,MZ150786,OP297541 |
| 1319 | VC_373_0 | 10 | 5782.863642 | 1957.950674 | 0.747061408 | 1.69E-21 | 2 | 6 | 45 | 45 | 1 | 4.771805838 | 0 | 0 | 0 | AJ006589,AJ550940,JX182372,MK433266,MK450421,MK450426,MK450431,MK450433,MK686068,MK686069 |
| 1320 | VC_374_0 | 2 | 98.89526051 | 687.0879377 | 0.125823632 | 0.044216833 | 4.123105626 | 4.123105626 | 1 | 1 | 1 | 4.123105626 | 0 | 0 | 0 | GQ379227,MK937595 |
| 1321 | VC_375_0 | 3 | 331.4591376 | 1005.620557 | 0.247897817 | 0.001927136 | 0 | 6.708203932 | 3 | 3 | 1 | 4.472135955 | 0 | 0 | 0 | MG593800,MG593803,MH001457 |
| 1322 | VC_376_0 | 3 | 385.8139242 | 1228.980825 | 0.238924436 | 0.002100806 | 4 | 4.358898944 | 3 | 3 | 1 | 4.16066819 | 0 | 0 | 0 | MT701593,MT701595,MT701596 |
| 1323 | VC_377_0 | 3 | 102.0180316 | 159.1354209 | 0.390644009 | 0.002149445 | 6.08276253 | 7.416198487 | 3 | 3 | 1 | 6.856676276 | 0 | 0 | 0 | KR905068,KU848187,LC625742 |
| 1324 | VC_378_0 | 9 | 4816.186316 | 1518.1514 | 0.760329893 | 2.30E-17 | 1.732050808 | 7.937253933 | 36 | 36 | 1 | 6.536314305 | 0 | 0 | 0 | AJ242593,DQ003637,KJ094022,KP399678,MH341452,MZ361825,MZ361826,MZ361827,ON456151 |
| 1325 | VC_379_0 | 5 | 898.198954 | 981.3357567 | 0.477883675 | 7.73E-08 | 4.242640687 | 9 | 10 | 9 | 0.9 | 7.458931357 | 0 | 0 | 0 | AJ312240,DQ003639,JX120799,KJ094023,MT500540 |
| 1326 | VC_37_0 | 5 | 1656.167192 | 1414.830112 | 0.539292949 | 1.31E-07 | 1.414213562 | 7.071067812 | 10 | 10 | 1 | 5.610870988 | 0 | 0 | 0 | OM046621,OM046622,OM046623,OM046625,OM046629 |
| 1327 | VC_37_1 | 3 | 144.2631276 | 1588.359613 | 0.083262862 | 0.051536502 | 8.94427191 | 8.94427191 | 3 | 3 | 1 | 8.94427191 | 0 | 0 | 0 | KU052037,MN013086,OL396571 |
| 1328 | VC_380_0 | 3 | 237.7284585 | 1220.057724 | 0.163074984 | 0.003249027 | 7.071067812 | 7.874007874 | 3 | 3 | 1 | 7.563680793 | 0 | 0 | 0 | DQ003642,KP399677,MH341451 |
| 1329 | VC_381_0 | 2 | 55.31844079 | 612.2201767 | 0.082869274 | 0.046186082 | 6.92820323 | 6.92820323 | 1 | 1 | 1 | 6.92820323 | 0 | 0 | 0 | GU323708,HQ141410 |
| 1330 | VC_385_0 | 51 | 161709.5524 | 23022.52987 | 0.875373408 | 0 | 0 | 6.164414003 | 1275 | 1275 | 1 | 4.163273787 | 0 | 0 | 0 | AJ251805,AJ318471,EU547803,JX421753,KC960671,KM253764,KP313531,KP791805,KP791806,KP791807,KT008108,KU687347,KU687348,KX130726,KX452696,KX778611,LC730324,MF285616,MF285617,MF285620,MH717709,MK608335,MN013074,MN149904,MT419366,MT813142,MW296877,MW600722,MZ359670,MZ428229,OK149215,OL362281,OL362282,OL362283,OL691920,OL744209,OL744212,OL744217,OL800605,ON528720,ON528721,ON528722,ON528723,ON528724,ON950090,OP617747,OX335419,OX335420,OX335431,OX335434,OX335437 |
| 1331 | VC_386_0 | 49 | 120746.5472 | 31298.5354 | 0.794149637 | 0 | 0 | 7 | 1176 | 1176 | 1 | 5.000250071 | 0 | 0 | 0 | AM183667,EU734170,EU734171,FJ194439,GQ468526,HQ333270,JQ965702,KF700371,KM366096,KP090453,KP090454,KY084243,LT960606,LT961836,LT961837,LT961838,LT961840,LT961841,LT961842,LT961843,LT961844,LT961845,LT961846,LT962379,LT962380,LT962475,LT962906,LT962907,MG471392,MH031343,MH809534,MK685668,MK907285,MN080711,MN252582,MT374859,MT515757,MT542512,MT625440,MW147603,MW367418,MW767996,MZ501055,MZ501064,OK663600,OL800603,OL856139,ON754979,ON922920 |
| 1332 | VC_387_0 | 3 | 366.9510029 | 8293.443365 | 0.042371165 | 0.001384107 | 3.16227766 | 4.898979486 | 3 | 3 | 1 | 4.1777977 | 0 | 0 | 0 | HQ728265,MN450150,ON715521 |
| 1333 | VC_388_0 | 4 | 596.9021897 | 16696.71238 | 0.034515757 | 1.19E-05 | 1 | 7 | 6 | 6 | 1 | 5.268252116 | 0 | 0 | 0 | KT321314,ON602733,OX335378,OX335441 |
| 1334 | VC_389_0 | 3 | 539.8748266 | 9940.81592 | 0.051511378 | 0.001384431 | 1 | 1.732050808 | 3 | 3 | 1 | 1.382088123 | 0 | 0 | 0 | MF979561,MW001769,ON995367 |
| 1335 | VC_38_0 | 3 | 900 | 0 | 1 | 1 | 3.741657387 | 4.898979486 | 3 | 3 | 1 | 4.443684211 | 0 | 0 | 0 | AB231700,KF356199,OP394178 |
| 1336 | VC_390_0 | 4 | 693.1883555 | 14133.57017 | 0.046752522 | 1.18E-05 | 3.872983346 | 5.744562647 | 6 | 6 | 1 | 4.973149053 | 0 | 0 | 0 | EU652770,KX078568,MW358931,OK499982 |

| | | | | | | | | | | | | | | | | | |
|---|---|---|---|---|---|---|---|---|---|---|---|---|---|---|---|---|---|
| 1337 | VC_392_0 | 126 | 892534.6732 | 35195.3221 | 0.962062968 | 0 | 0 | 7.280109889 | 7875 | 7875 | | 1 | 4.590539688 | 0 | 0 | 0 | AY247822,AY264774,AY264775,AY264776,AY264777,AY264778,EU734174,FR667955,GU071091,JQ957925,JQ965700,JQ965701,JQ965703,JX000007,JX872508,KM242061,KU927499,LC730325,LR745706,LR745707,LR745708,LR745709,LR745710,MG488277,MG832642,MG833025,MG966531,MH382198,MH717098,MH717099,MH779619,MK310182,MK542821,MN026740,MN098329,MN481365,MN481366,MT350293,MT543041,MT681669,MW074885,MW883060,MZ220766,MZ318361,MZ318362,MZ318363,MZ375233,MZ375234,MZ375235,MZ375236,MZ375237,MZ375238,MZ375239,MZ375240,MZ375241,MZ375242,MZ375243,MZ375244,MZ375245,MZ375246,MZ375247,MZ375248,MZ375249,MZ375250,MZ375251,MZ375252,MZ375253,MZ375254,MZ375255,MZ375256,MZ375257,MZ375258,MZ375259,MZ375260,MZ375261,MZ375262,MZ375263,MZ375264,MZ375265,MZ375266,MZ375267,MZ375268,MZ375269,MZ375270,MZ375271,MZ375272,MZ375273,MZ375274,MZ375275,MZ375276,MZ375277,MZ375278,MZ375279,MZ375280,MZ375281,MZ375282,MZ375283,MZ375284,MZ375285,MZ375286,MZ375287,MZ375288,MZ375289,MZ375290,MZ375291,MZ375292,MZ375293,MZ375294,MZ501078,MZ501081,MZ833439,OK042080,OL964739,OL964740,OL964741,OL964748,OL964749,OL964750,OM630586,ON528715,ON528716,ON528717,ON528719,ON568193,OP331230,V01146 |
| 1338 | VC_393_0 | 7 | 3186.069379 | 14766.61339 | 0.177470377 | 2.99E-15 | 1.414213562 | 4 | 21 | 21 | | 1 | 3.290631022 | 0 | 0 | 0 | MH059639,MH460459,MN013082,MN013085,MN812693,MT822288,ON287378 |
| 1339 | VC_394_0 | 2 | 162.6016698 | 7548.03508 | 0.021087969 | 0.042077996 | 2.236067977 | 2.236067977 | 1 | 1 | | 1 | 2.236067977 | 0 | 0 | 0 | MH370477,MT465573 |
| 1340 | VC_395_0 | 5 | 1818.525602 | 15988.10076 | 0.102126341 | 2.43E-08 | 1 | 2.449489743 | 10 | 10 | | 1 | 1.903600885 | 0 | 0 | 0 | MN184885,MT822284,MT822285,MT822286,MT822287 |
| 1341 | VC_396_0 | 2 | 111.0109439 | 1442.856894 | 0.07144169 | 0.042406414 | 4.123105626 | 4.123105626 | 1 | 1 | | 1 | 4.123105626 | 0 | 0 | 0 | MW057854,MW358928 |
| 1342 | VC_398_0 | 48 | 7635.116631 | 8227.893032 | 0.481315765 | 4.49E-81 | 0 | 2.449489743 | 1128 | 1128 | | 1 | 1.48586544 | 0 | 0 | 0 | KP087939,KP087945,KT264811,MG945164,MG945682,MG945837,MH572269,MH572314,MH572315,MH572316,MH572317,MH572318,MH572319,MH572320,MH572321,MH572325,MH572326,MH572328,MH572329,MH572336,MH572351,MH572394,MH572395,MH572396,MH572397,MH572404,MH572419,MH572431,MH572439,MH572441,MH572443,MH572445,MH572446,MH572451,MH572473,MH572487,MH572492,MH572493,MH572494,MH572503,MH572510,MH572511,MH992195,MH992211,MK249143,MK249151,MK249223,MK249231 |
| 1343 | VC_39_0 | 35 | 145832.3456 | 190456.2075 | 0.433652422 | 0 | 0 | 8.774964387 | 595 | 595 | | 1 | 4.202204325 | 0 | 0 | 0 | AF125520,AP000363,AP000422,AP004402,AP005153,AP005154,AP012529,AP012531,AP012532,AP012535,EU311208,HM208303,KJ909655,KP682372,KP682373,KP682375,KP682376,KP682377,KP682378,KP682379,KP682380,KP682382,KP682383,KP682384,KP682385,KP682386,KP682387,KP682388,KP682389,KP682390,KP682391,KP682392,KU238068,KU238069,KU238070 |
| 1344 | VC_3_0 | 17 | 13843.20867 | 10704.70014 | 0.563926189 | 1.02E-79 | 0 | 6.557438524 | 136 | 136 | | 1 | 4.898300906 | 0 | 0 | 0 | CP000623,CP000624,CP008753,DQ087285,GU911304,HM461982,JX681814,LC528882,LC667452,MW072790,MZ375754,OM304382,OM638608,OM638609,OM638610,OM638611,ON135435 |
| 1345 | VC_400_0 | 1 | 0 | 135.8404689 | 0 | 0 | 0 | 0 | 1 | 1 | | 1 | 0 | 0 | 0 | 0 | MH572340 |
| 1346 | VC_401_0 | 1 | 0 | 23.24090136 | 0 | 0 | 0 | 0 | 1 | 1 | | 1 | 0 | 0 | 0 | 0 | MH572479 |
| 1347 | VC_402_0 | 1 | 0 | 26.21969195 | 0 | 0 | 0 | 0 | 1 | 1 | | 1 | 0 | 0 | 0 | 0 | MH572512 |
| 1348 | VC_403_0 | 20 | 3038.785298 | 1566.935839 | 0.659784908 | 3.21E-84 | 0 | 2.236067977 | 190 | 190 | | 1 | 1.198881836 | 0 | 0 | 0 | KC131023,KC131025,MH572294,MH572295,MH572296,MH572384,MH572385,MH572386,MH572387,MH572388,MH572389,MH572390,MH572391,MH572430,MH572470,MH572471,MH572472,MH572505,MH572506,MH572507 |
| 1349 | VC_404_0 | 25 | 2062.264597 | 1588.44935 | 0.564893505 | 1.16E-22 | 0 | 2.449489743 | 300 | 300 | | 1 | 1.795576552 | 0 | 0 | 0 | MH572279,MH572300,MH572301,MH572302,MH572303,MH572305,MH572306,MH572307,MH572308,MH572309,MH572310,MH572311,MH572312,MH572313,MH572345,MH572392,MH572393,MH572406,MH572435,MH572436,MH572437,MH572469,MH572480,MH572508,MH572509 |
| 1350 | VC_406_0 | 11 | 9660.292554 | 563.1494002 | 0.944915871 | 1.41E-27 | 0 | 6.708203932 | 55 | 55 | | 1 | 5.323437507 | 0 | 0 | 0 | KP790011,KU998237,KU998238,KY471268,KY471269,MH651189,MH697580,MN010758,MT553344,MW132713,ON260826 |
| 1351 | VC_407_0 | 3 | 351.7916377 | 697.822307 | 0.335162885 | 0.001752088 | 3 | 5.477225575 | 3 | 3 | | 1 | 4.620796794 | 0 | 0 | 0 | AJ278322,MG324353,MG324354 |
| 1352 | VC_408_0 | 9 | 6578.977811 | 1070.851939 | 0.860016239 | 1.54E-21 | 0 | 0 | 36 | 36 | | 1 | 0 | 0 | 0 | 0 | KY566218,KY613597,KY624610,KY624611,KY624612,KY624613,KY624614,KY624615,KY624616 |
| 1353 | VC_409_0 | 3 | 71.91390314 | 408.9526625 | 0.149550641 | 0.017058989 | 7 | 8.426149773 | 3 | 3 | | 1 | 7.499072528 | 0 | 0 | 0 | KY203335,KY250482,MW582634 |
| 1354 | VC_40_0 | 5 | 1470.851922 | 15953.28203 | 0.084414636 | 5.31E-08 | 4.123105626 | 5.477225575 | 10 | 10 | | 1 | 4.961042449 | 0 | 0 | 0 | LC567830,LC567838,LC567842,LC616079,ON416862 |
| 1355 | VC_410_0 | 7 | 2464.03842 | 652.9410414 | 0.790521224 | 3.41E-13 | 2.449489743 | 7.416198487 | 21 | 21 | | 1 | 6.268348397 | 0 | 0 | 0 | MG198783,MG198784,MH271305,MH271318,MT521997,MZ681507,ON456352 |
| 1356 | VC_411_0 | 3 | 75.906291 | 536.6301669 | 0.123921262 | 0.010136594 | 7 | 8.717797887 | 3 | 3 | | 1 | 7.755877441 | 0 | 0 | 0 | LC644972,LC644974,MT843275 |
| 1357 | VC_412_0 | 39 | 54043.08926 | 9913.324136 | 0.844998748 | 8.06E-302 | 0 | 8.94427191 | 741 | 741 | | 1 | 7.184510989 | 0 | 0 | 0 | AJ302074,DQ113772,HF563658,JX879087,KC413988,KT337358,KT337364,KT388093,KX550065,KY065455,KY065479,KY065487,KY065490,KY065491,KY065492,KY065493,KY065501,KY065502,KY349816,MK448671,MK448708,MK448714,MK448831,MK448834,MK448861,MK448874,MK448875,MK448886,MK448887,MK448897,MK448898,MK448905,MK448925,MK448927,MK449006,MK449009,MT900488,MW495853,OL774873 |
| 1358 | VC_413_0 | 3 | 189.5358541 | 4373.925467 | 0.041533354 | 0.001582459 | 7.141428429 | 8 | 3 | 3 | | 1 | 7.692894121 | 0 | 0 | 0 | KY065465,MK448733,MK448904 |
| 1359 | VC_414_0 | 37 | 70652.39027 | 50121.5382 | 0.584997037 | 0 | 1.414213562 | 9.591663047 | 666 | 631 | 0.947447447 | 1 | 7.278358705 | 0 | 0 | 0 | FR671405,FR671406,FR671407,FR671410,FR671411,HG799490,HG799496,HG799497,KJ417497,KT337339,KT337340,KT337341,KT337352,KT337354,KT337355,KT337356,KT337359,KT337362,KY065450,KY065451,KY065452,KY065456,KY065472,KY065473,KY065474,KY065475,KY065476,KY065477,KY065478,KY065480,KY065486,KY065488,KY065496,KY065497,KY065500,KY065505,OK173051 |
| 1360 | VC_415_0 | 1 | 0 | 2039.294407 | 0 | 0 | 0 | 0 | 1 | 1 | | 1 | 0 | 0 | 0 | 0 | KY065449 |
| 1361 | VC_416_0 | 14 | 8305.583211 | 23731.79778 | 0.259246635 | 2.19E-58 | 0 | 9 | 91 | 90 | 0.989010989 | 7.181532971 | 0 | 0 | 0 | | MK448686,MK448762,MK448788,MK448791,MK448793,MK448862,MK448949,MK448951,MK448954,MK448958,MK448959,MK448960,MK448965,MK448977 |
| 1362 | VC_417_0 | 5 | 274.4500849 | 139.5623016 | 0.662903077 | 0.002256602 | 5.916079783 | 9.695359715 | 10 | 8 | 0.8 | 7.590082617 | 0 | 0 | 0 | | KC237729,MW478289,MZ503613,S2-wms_130687,S5-wms_49310 |
| 1363 | VC_418_0 | 6 | 403.2804723 | 478.2549863 | 0.457475044 | 4.95E-07 | 6.08276253 | 9.848857802 | 15 | 12 | 0.8 | 8.070324029 | 0 | 0 | 0 | | LR596902,S1-wms-combined_224578,S2-wms-combined_246480,S2-wms_140933,S4-wms-combined_64632,S4-wms_62635 |
| 1364 | VC_419_0 | 5 | 1149.395377 | 3451.249621 | 0.249833529 | 3.43E-08 | 1.414213562 | 6.8556546 | 10 | 10 | | 1 | 5.80724635 | 0 | 0 | 0 | MK448680,MK448684,MK448698,MK448769,MK448895 |
| 1365 | VC_41_0 | 16 | 22058.91815 | 86879.96828 | 0.202488926 | 1.77E-55 | 0 | 11 | 120 | 116 | 0.966666667 | 4.866937692 | 0 | 0 | 0 | | AJ556162,JQ347801,LC727891,LC727892,LC727893,LC727894,LC727895,LC727896,LC727897,LC727898,LC727899,LC727900,LC727901,LC727902,ON470597,ON470625 |
| 1366 | VC_41_1 | 10 | 7272.870373 | 54440.7915 | 0.117848628 | 2.75E-30 | 2.236067977 | 10 | 45 | 37 | 0.822222222 | 7.293530987 | 0 | 0 | 0 | | AP012530,AP012538,AP012539,AP012540,FJ184280,FJ188381,FM180578,LC645408,LC645425,LC645426 |
| 1367 | VC_420_0 | 6 | 92.73700876 | 313.7146824 | 0.228162438 | 0.025379185 | 6.403124237 | 8.717797887 | 15 | 15 | | 1 | 7.456942241 | 0 | 0 | 0 | S1-wms-combined_72167,S2-wms-combined_227800,S7-wms-combined_28138,S7-wms_127801,S8-wms-combined_140059,S9-wms-combined_71647 |
| 1368 | VC_422_0 | 5 | 1509.058092 | 4313.102866 | 0.259192094 | 2.50E-08 | 0 | 3 | 10 | 10 | | 1 | 2.383623342 | 0 | 0 | 0 | MF034659,MF166859,MT902335,MZ995503,MZ995506 |
| 1369 | VC_423_0 | 45 | 172111.3091 | 9361.033478 | 0.948416197 | 2.31E-272 | 0 | 4.69041576 | 990 | 990 | | 1 | 2.82465251 | 0 | 0 | 0 | AJ505558,AM265638,AM910651,EU056923,EU236438,FN263372,JQ307386,JX501340,JX997978,KC969441,KM067278,KR054031,KU743887,KX711710,KY618819,LC727700,LN610574,LN610580,MH107770,MH979674,MN553585,MN553587,MN553588,MN553589,MN602045,MN615698,MN615699,MN692672,MN871455,MN871483,MN901924,MW117144,MW406975,MW406976,MZ553931,MZ687409,OK539824,OL754589,OL802210,ON857926,ON857928,ON857933,ON857935,OP292288,OP556577 |
| 1370 | VC_424_0 | 7 | 1133.815378 | 119.8008654 | 0.904435774 | 0.0001857 | 0 | 5 | 21 | 21 | | 1 | 3.309097027 | 0 | 0 | 0 | AJ536073,AY257527,AY701338,CP013282,EU408779,MZ089978,MZ089979 |
| 1371 | VC_426_0 | 2 | 157.6968698 | 640.3204387 | 0.197610839 | 0.043560592 | 4.358898944 | 4.358898944 | 1 | 1 | | 1 | 4.358898944 | 0 | 0 | 0 | AJ560763,GQ866233 |
| 1372 | VC_426_1 | 3 | 113.8338456 | 874.7302244 | 0.115150701 | 0.001953848 | 7.937253933 | 8.831760866 | 3 | 3 | | 1 | 8.398388191 | 0 | 0 | 0 | CP013974,MG575421,MZ424863 |
| 1373 | VC_427_0 | 3 | 30.36597811 | 453.6216654 | 0.062741226 | 0.031698786 | 7.681145748 | 8 | 3 | 3 | | 1 | 7.872799894 | 0 | 0 | 0 | JF974295,MW202455,S1-wms-combined_324053 |
| 1374 | VC_428_0 | 33 | 123368.9251 | 1055.713699 | 0.991515236 | 3.33E-84 | 0 | 6 | 528 | 528 | | 1 | 3.166296407 | 0 | 0 | 0 | KM366099,KX826077,KY608966,MF740800,MH181876,MK568062,MK673511,MK770411,MK770413,MN379740,MT012730,MT074434,MT074435,MT074436,MT074437,MT074438,MT074439,MT074441,MT074442,MT074443,MT074444,MT074445,MT074446,MT663719,MT764207,MT768059,MW413353,MW508890,MW602648,MZ127825,OM313460,OM313461,OP132241 |
| 1375 | VC_42_0 | 49 | 173362.7203 | 210421.3585 | 0.451719417 | 0 | 0 | 9.643650761 | 1176 | 1011 | 0.859693878 | 6.789528392 | 0 | 0 | 0 | | AP012534,HE664024,HG792102,HG792103,HG792104,HG792105,HG803182,HG803183,HG803184,HG803185,HQ424691,JQ011318,KF971864,KP682371,KP682374,KR781488,KU298437,KY914478,LC645409,LC645410,LC645411,LC645412,LC645413,LC645414,LC645415,LC645416,LC645417,LC645418,LC645419,LC645420,LC645421,LC645422,LC645423,LC645424,LC645427,LC645438,LC645442,LC645443,LC645444,LC645445,LC645446,LC645447,LC645448,LC645449,LC645450,LC645451,LC727903,MG710528,MN067333 |
| 1376 | VC_431_0 | 9 | 2681.014496 | 13346.99325 | 0.167270601 | 9.75E-22 | 0 | 8.717797887 | 36 | 36 | | 1 | 6.926770072 | 0 | 0 | 0 | MK448692,MK448702,MK448711,MK448716,MK448724,MK448730,MK448744,MK448745,MK448923 |
| 1377 | VC_432_0 | 2 | 300 | 7992.37339 | 0.036177821 | 0.043196036 | 0 | 0 | 1 | 1 | | 1 | 0 | 0 | 0 | 0 | MN695334,MN695335 |
| 1378 | VC_432_1 | 1 | 0 | 8278.263398 | 0 | 0 | 0 | 0 | 1 | 1 | | 1 | 0 | 0 | 0 | 0 | MH412654 |
| 1379 | VC_432_10 | 2 | 300 | 13563.39915 | 0.021639715 | 0.042289829 | 1.732050808 | 1.732050808 | 1 | 1 | | 1 | 1.732050808 | 0 | 0 | 0 | GU071098,HQ317391 |
| 1380 | VC_432_11 | 1 | 0 | 9722.949975 | 0 | 0 | 0 | 0 | 1 | 1 | | 1 | 0 | 0 | 0 | 0 | HQ633071 |
| 1381 | VC_432_12 | 1 | 0 | 7041.795082 | 0 | 0 | 0 | 0 | 1 | 1 | | 1 | 0 | 0 | 0 | 0 | MW015081 |
| 1382 | VC_432_13 | 1 | 0 | 8524.066472 | 0 | 0 | 0 | 0 | 1 | 1 | | 1 | 0 | 0 | 0 | 0 | MK799832 |
| 1383 | VC_432_14 | 1 | 0 | 8152.101765 | 0 | 0 | 0 | 0 | 1 | 1 | | 1 | 0 | 0 | 0 | 0 | MN018232 |
| 1384 | VC_432_15 | 3 | 900 | 20632.44506 | 0.04179739 | 0.001443179 | 1.414213562 | 2.645751311 | 3 | 3 | | 1 | 2.098677617 | 0 | 0 | 0 | KU686204,KU686205,KU686206 |
| 1385 | VC_432_16 | 1 | 0 | 6923.837283 | 0 | 0 | 0 | 0 | 1 | 1 | | 1 | 0 | 0 | 0 | 0 | MH629685 |
| 1386 | VC_432_2 | 1 | 0 | 10031.8177 | 0 | 0 | 0 | 0 | 1 | 1 | | 1 | 0 | 0 | 0 | 0 | GU071095 |
| 1387 | VC_432_3 | 1 | 0 | 8850.791003 | 0 | 0 | 0 | 0 | 1 | 1 | | 1 | 0 | 0 | 0 | 0 | MF351863 |
| 1388 | VC_432_4 | 1 | 0 | 8731.803758 | 0 | 0 | 0 | 0 | 1 | 1 | | 1 | 0 | 0 | 0 | 0 | MT408532 |
| 1389 | VC_432_5 | 1 | 0 | 10636.28451 | 0 | 0 | 0 | 0 | 1 | 1 | | 1 | 0 | 0 | 0 | 0 | MK867354 |
| 1390 | VC_432_6 | 1 | 0 | 5226.300768 | 0 | 0 | 0 | 0 | 1 | 1 | | 1 | 0 | 0 | 0 | 0 | MH319749 |
| 1391 | VC_432_7 | 1 | 0 | 7317.645565 | 0 | 0 | 0 | 0 | 1 | 1 | | 1 | 0 | 0 | 0 | 0 | MH319754 |
| 1392 | VC_432_8 | 1 | 0 | 6993.109173 | 0 | 0 | 0 | 0 | 1 | 1 | | 1 | 0 | 0 | 0 | 0 | KM359505 |
| 1393 | VC_432_9 | 3 | 900 | 23316.65955 | 0.037164498 | 0.001486876 | 2.236067977 | 4.123105626 | 3 | 3 | | 1 | 3.366943663 | 0 | 0 | 0 | AY939844,GU071092,HQ632825 |
| 1394 | VC_433_0 | 6 | 4500 | 52678.17455 | 0.078701358 | 2.45E-11 | 1.414213562 | 2.449489743 | 15 | 15 | | 1 | 1.939724305 | 0 | 0 | 0 | HQ634177,KU686192,KU686193,KU686194,KU686195,KU686196 |
| 1395 | VC_433_1 | 9 | 10800 | 73079.42889 | 0.128756241 | 1.37E-24 | 1 | 9.746794345 | 36 | 31 | 0.861111111 | 6.77186817 | 0 | 0 | 0 | | KJ019054,KJ019069,KJ019082,KJ019089,KJ019094,KJ019156,KU686200,KU686201,KU686202 |
| 1396 | VC_433_10 | 1 | 0 | 10400.95313 | 0 | 0 | 0 | 0 | 1 | 1 | | 1 | 0 | 0 | 0 | 0 | KU686211 |
| 1397 | VC_433_2 | 1 | 0 | 10232.49831 | 0 | 0 | 0 | 0 | 1 | 1 | | 1 | 0 | 0 | 0 | 0 | KF156338 |
| 1398 | VC_433_3 | 3 | 900 | 29884.90753 | 0.029235105 | 0.001871896 | 1.414213562 | 2.645751311 | 3 | 3 | | 1 | 2.098677617 | 0 | 0 | 0 | KU686197,KU686198,KU686199 |
| 1399 | VC_433_4 | 9 | 10800 | 73217.09277 | 0.128545271 | 9.86E-24 | 0 | 6.782329983 | 36 | 36 | | 1 | 4.631535894 | 0 | 0 | 0 | GU071105,KU594607,KX349291,KX349292,KX349293,KX349294,KX349295,KX349296,KX349297 |
| 1400 | VC_433_5 | 2 | 300 | 17008.44074 | 0.017332584 | 0.063578424 | 4.69041576 | 4.69041576 | 1 | 1 | | 1 | 4.69041576 | 0 | 0 | 0 | AJ630128,LN828717 |
| 1401 | VC_433_6 | 1 | 0 | 11224.04754 | 0 | 0 | 0 | 0 | 1 | 1 | | 1 | 0 | 0 | 0 | 0 | KU594606 |
| 1402 | VC_433_7 | 1 | 0 | 10852.20659 | 0 | 0 | 0 | 0 | 1 | 1 | | 1 | 0 | 0 | 0 | 0 | MW147367 |
| 1403 | VC_433_8 | 1 | 0 | 10306.34815 | 0 | 0 | 0 | 0 | 1 | 1 | | 1 | 0 | 0 | 0 | 0 | FM207411 |
| 1404 | VC_433_9 | 1 | 0 | 10743.62193 | 0 | 0 | 0 | 0 | 1 | 1 | | 1 | 0 | 0 | 0 | 0 | MW117966 |
| 1405 | VC_435_0 | 2 | 300 | 6789.518525 | 0.042315991 | 0.042722815 | 7.416198487 | 7.416198487 | 1 | 1 | | 1 | 7.416198487 | 0 | 0 | 0 | MT162466,MT162467 |
| 1406 | VC_435_1 | 1 | 0 | 3805.632018 | 0 | 0 | 0 | 0 | 1 | 1 | | 1 | 0 | 0 | 0 | 0 | MK016664 |
| 1407 | VC_436_0 | 4 | 1800 | 3624.513577 | 0.331826988 | 1.28E-05 | 0 | 2.828427125 | 6 | 6 | | 1 | 1.794110782 | 0 | 0 | 0 | MZ577095,MZ577096,MZ577097,MZ577098 |
| 1408 | VC_437_0 | 4 | 1800 | 1460.685381 | 0.55203118 | 1.77E-05 | 2 | 8.185352772 | 6 | 6 | | 1 | 6.341155883 | 0 | 0 | 0 | KF381361,KR052480,KR052481,KR052482 |
| 1409 | VC_438_0 | 6 | 514.4598957 | 5098.47782 | 0.091656085 | 5.73E-05 | 4.123105626 | 8.831760866 | 15 | 15 | | 1 | 7.096854097 | 0 | 0 | 0 | MH319720,MH319733,MH319742,MH319753,MH319758,MH327487 |
| 1410 | VC_439_0 | 2 | 28.66807063 | 956.7655289 | 0.029091834 | 0.09466152 | 8.485281374 | 8.485281374 | 1 | 1 | | 1 | 8.485281374 | 0 | 0 | 0 | MH319750,MH319759 |
| 1411 | VC_43_0 | 38 | 82501.86654 | 57699.31907 | 0.588453416 | 0 | 0 | 9.797958971 | 703 | 679 | 0.965860597 | 6.633677674 | 0 | 0 | 0 | | EU078592,JQ086376,JQ182735,KT232076,LAMCG,LC730321,LR595850,LR595859,LR595860,LR595861,LR595863,LR595864,LR595865,LR595866,LR597635,LR597636,LR597639,LR597643,LR597644,LR597648,LR597650,LR597651,LR597652,LR597653,MF374379,MH547045,OL657226,OL657227,OL657228,OM418625,OM418626,OM418627,OM475427,OM475428,OM475430,OM475432,OM475435,OV696612 |
| 1412 | VC_440_0 | 4 | 181.3255917 | 0 | 1 | 0 | 1.732050808 | 3.872983346 | 6 | 6 | | 1 | 2.974977029 | 0 | 0 | 0 | AJ635161,AY722806,MN876845,MT047590 |
| 1413 | VC_442_0 | 4 | 1800 | 19574.82243 | 0.084211226 | 3.68E-05 | 1.732050808 | 3 | 6 | 6 | | 1 | 2.461117863 | 0 | 0 | 0 | MK838116,MN102098,MN131137,MN159079 |
| 1414 | VC_442_1 | 3 | 900 | 15829.35752 | 0.053797643 | 0.005804482 | 2.645751311 | 5.196152423 | 3 | 3 | | 1 | 4.313641082 | 0 | 0 | 0 | MN642089,ON287370,ON287375 |
| 1415 | VC_442_2 | 4 | 1800 | 19535.6715 | 0.084365753 | 0.000118713 | 7.416198487 | 9.486832981 | 6 | 5 | 0.833333333 | 8.568747127 | 0 | 0 | 0 | | JN627160,MK689364,MZ348422,MZ501267 |
| 1416 | VC_442_3 | 3 | 900 | 16112.08532 | 0.052903567 | 0.003248195 | 3 | 5.656854249 | 3 | 3 | | 1 | 4.617668891 | 0 | 0 | 0 | MN032614,MN453779,OL964756 |
| 1417 | VC_442_4 | 2 | 300 | 11508.22077 | 0.025406029 | 0.081405016 | 6.480740698 | 6.480740698 | 1 | 1 | | 1 | 6.480740698 | 0 | 0 | 0 | MK774614,OM810292 |
| 1418 | VC_443_0 | 2 | 300 | 10048.63301 | 0.028989336 | 0.125903965 | 5.385164807 | 5.385164807 | 1 | 1 | | 1 | 5.385164807 | 0 | 0 | 0 | MH791414,MW272540 |
| 1419 | VC_443_1 | 2 | 300 | 10135.57369 | 0.028747821 | 0.10652622 | 3.605551275 | 3.605551275 | 1 | 1 | | 1 | 3.605551275 | 0 | 0 | 0 | GU459069,KY290955 |
| 1420 | VC_443_2 | 10 | 13500 | 26873.86424 | 0.334374731 | 4.86E-23 | 5 | 9.949874371 | 45 | 42 | 0.933333333 | 7.360303405 | 0 | 0 | 0 | | JX123262,MF448340,MF498773,MF498775,MH791398,MH791404,MN871441,MN871442,MT713136,OM339718 |
| 1421 | VC_444_0 | 2 | 189.2994502 | 43.3529588 | 0.813657813 | 0.270145687 | 1.732050808 | 1.732050808 | 1 | 1 | | 1 | 1.732050808 | 0 | 0 | 0 | AJ783769,JQ287645 |
| 1422 | VC_445_0 | 2 | 300 | 454.0852776 | 0.397832989 | 0.077364462 | 2.236067977 | 2.236067977 | 1 | 1 | | 1 | 2.236067977 | 0 | 0 | 0 | MT732443,MT732444 |
| 1423 | VC_445_1 | 2 | 94.59058088 | 634.4718487 | 0.129742772 | 0.069448331 | 8.717797887 | 8.717797887 | 1 | 1 | | 1 | 8.717797887 | 0 | 0 | 0 | KC821607,KC821618 |
| 1424 | VC_446_0 | 4 | 691.9971506 | 225.7319334 | 0.754032059 | 0.000122324 | 4.472135955 | 6 | 6 | 6 | | 1 | 5.165250435 | 0 | 0 | 0 | HG322870,KP238129,KP282677,KP282678 |
| 1425 | VC_447_0 | 3 | 585.4749419 | 122.6626863 | 0.82678129 | 0.004900713 | 0 | 3.16227766 | 3 | 3 | | 1 | 2.108185107 | 0 | 0 | 0 | AJ888457,KX607101,KX607102 |
| 1426 | VC_448_0 | 4 | 1800 | 4084.747806 | 0.30587547 | 0.000211471 | 0 | 6.32455532 | 6 | 6 | | 1 | 4.094198404 | 0 | 0 | 0 | KU878088,LC680884,MF360957,MW749003 |
| 1427 | VC_448_1 | 1 | 0 | 1907.899768 | 0 | 0 | 0 | 0 | 1 | 1 | | 1 | 0 | 0 | 0 | 0 | KT895374 |
| 1428 | VC_449_0 | 7 | 6300 | 4174.678176 | 0.601450459 | 1.21E-11 | 3 | 8.888194417 | 21 | 21 | | 1 | 7.026236964 | 0 | 0 | 0 | LC680885,MN091626,MW218148,MW248466,MW349128,MW349129,MZ779063 |
| 1429 | VC_450_0 | 16 | 36000 | 1642.758687 | 0.956359238 | 1.75E-34 | 0 | 2.236067977 | 120 | 120 | | 1 | 0.385698076 | 0 | 0 | 0 | AM076770,MN617773,MT366931,MT366932,MT366933,MT366934,MT366935,MT366936,MT366937,MT366938,MT366939,MT366940,MT366941,MT366942,MT366943,MT366944 |
| 1430 | VC_451_0 | 80 | 335903.9773 | 316.3006065 | 0.999059246 | 1.74E-97 | 0 | 7.416198487 | 3160 | 3160 | | 1 | 5.572048164 | 0 | 0 | 0 | AM084414,DQ111067,EU734172,HG813241,HG818823,HG818824,KC109329,KJ748011,KT990215,KU687349,KU687350,KU687351,KX669227,KX689784,KY295894,KY962008,LT594300,MF630921,MG845865,MH400309,MH464253,MH491167,MK033136,MK301532,MK610268,MK903277,MK903279,MK903281,MN067430,MN218775,MN414250,MN510331,MN518893,MN518894,MN715150,MN812686,MT350292,MT446393,MT446394,MT496969,MT682707,MT808983,MT862763,MW258709,MW345254,MW416012,MW748991,MW876471,MZ223858,MZ234009,MZ234011,MZ234021,MZ234022,MZ234023,MZ234024,MZ234044,MZ234045,MZ234046,MZ234047,MZ311861,MZ311862,MZ398247,MZ648215,MZ726794,MZ923531,OK210074,OL362275,OL362276,OL539729,OL773676,OL773677,OM256482,OM355495,OM457002,OM867676,OM908436,OM937123,ON528733,ON548433,ON637250 |
| 1431 | VC_452_0 | 66 | 229009.5116 | 18629.77858 | 0.924770506 | 3.82E-252 | 0 | 8.246211251 | 2145 | 2145 | | 1 | 5.919144799 | 0 | 0 | 0 | AM084415,AY288927,AY370673,AY370674,GU595417,JN986844,KJ837901,KM366097,KM370384,KM819695,KM819696,KP890822,KU946962,KX279892,KY295891,KY295892,KY295893,KY295897,KY295899,KY435490,KY744566,LC730323,MG004687,MG251390,MG775042,MH729818,MK373771,MK373773,MK380014,MN384978,MT496971,MT680615,MW394389,MZ031013,MZ234012,MZ234019,MZ234020,MZ234032,MZ234049,MZ234050,MZ398244,MZ573924,MZ747518,MZ997839,MZ997840,MZ997841,OK625527,OL362272,OL362273,OL744215,OL744218,ON148527,ON185580,ON210144,ON210145,ON461912,ON526839,ON602763,ON602764,ON602765,OP018999,OP038547,OP079918,OP413828,OP413830,OX335382 |
| 1432 | VC_453_0 | 6 | 2385.120897 | 18535.91353 | 0.114005878 | 2.24E-11 | 0 | 4 | 15 | 15 | | 1 | 2.657579136 | 0 | 0 | 0 | EF160123,FQ482084,FQ482086,MG736918,OK570184,OM486945 |
| 1433 | VC_455_0 | 18 | 27764.02161 | 32622.24104 | 0.459773803 | 6.73E-92 | 0 | 7.549834435 | 153 | 153 | | 1 | 5.464752943 | 0 | 0 | 0 | DQ121662,JF770475,JQ182736,KC333879,KC579452,KF771236,KF771237,KF771238,KF771239,KJ668713,KT825490,MF158039,MF158042,MW286156,OL539447,OL539451,OL539453,OL539467 |
| 1434 | VC_456_0 | 62 | 362422.1017 | 39104.94971 | 0.902609427 | 0 | 0 | 8 | 1891 | 1891 | | 1 | 3.83484609 | 0 | 0 | 0 | AM156909,JN986845,KX130960,KY398841,MG050172,MG065688,MK372342,MK373793,MK907226,MK907227,MK907228,MK907229,MK907230,MK907231,MK907232,MK907233,MK907234,MK907235,MK907240,MK907241,MK907242,MK907243,MK907244,MK907245,MK907246,MK907247,MK907248,MK907249,MK907250,MK907251,MK907252,MK907253,MK907254,MK907255,MK907256,MK907257,MK907258,MK907259,MK907260,MK907261,MK907262,MK907263,MK907264,MK907265,MK907266,MK907272,MK907273,MK907274,MK907275,MK907276,MN164484,MN840485,MT511058,MT682706,MW057918,MZ501051,MZ501080,MZ501087,OM258170,ON081052,OP087522,OP251154 |
| 1435 | VC_458_0 | 5 | 1650.186607 | 6863.653045 | 0.193824018 | 3.22E-08 | 0 | 3.31662479 | 10 | 10 | | 1 | 2.520266907 | 0 | 0 | 0 | MH059632,MH807810,MH807813,MH807815,MH807817 |
| 1436 | VC_459_0 | 3 | 230.4568416 | 39.34927899 | 0.854157204 | 0.009451772 | 4.123105626 | 5.567764363 | 3 | 3 | | 1 | 5.056031854 | 0 | 0 | 0 | KC292023,MG550112,MZ334526 |
| 1437 | VC_45_0 | 12 | 5477.354336 | 10170.17797 | 0.350045888 | 6.45E-40 | 0 | 8.774964387 | 66 | 66 | | 1 | 6.994701136 | 0 | 0 | 0 | AJ298298,AY055382,KC736978,KC814930,KU927493,MK733259,MK733260,MK733261,MZ501270,ON470584,ON470603,SFU82619 |
| 1438 | VC_460_0 | 19 | 26606.0772 | 44464.11235 | 0.374363392 | 2.87E-105 | 0 | 8.544003745 | 171 | 171 | | 1 | 7.153407194 | 0 | 0 | 0 | JF974294,JF974312,MK327140,MT682064,MW119258,MW722082,MZ318367,MZ440881,MZ836210,OL362277,OL617041,ON602730,ON602737,ON602738,ON602744,ON602759,ON602760,OP373729,OP481875 |
| 1439 | VC_461_0 | 96 | 663542.7094 | 49388.65482 | 0.930724531 | 0 | 0 | 9.327379053 | 4560 | 4366 | 0.95745614 | 5.931816891 | 0 | 0 | 0 | | AM491472,KX017520,KX523699,KY860935,MF158038,MH023293,MH321493,MH424446,MH717097,MH791402,MH791410,MH791412,MK112901,MK599416,MK759854,MK947460,MN692673,MN737551,MN871490,MN871503,MT768060,MW427215,MW477799,MZ375232,MZ375295,MZ375296,MZ375297,MZ375298,MZ375299,MZ375300,MZ375301,MZ375302,MZ375303,MZ375304,MZ375305,MZ375306,MZ375307,MZ375308,MZ375309,MZ375310,MZ375311,MZ375312,MZ375313,MZ375314,MZ375315,MZ375316,MZ375317,MZ375318,MZ375319,MZ375320,MZ375321,MZ375322,MZ375323,MZ375324,MZ375325,MZ375326,MZ375327,MZ375328,MZ375329,MZ375330,MZ375331,MZ375332,MZ375333,MZ375334,MZ375335,MZ375336,MZ375337,MZ375338,MZ375339,MZ375340,MZ375341,MZ375342,MZ375343,MZ375344,MZ375345,MZ375346,MZ375347,MZ375348,MZ375349,MZ375350,MZ375351,MZ375352,MZ375353,MZ375354,MZ375355,MZ489634,MZ726796,OL362274,OL964734,OL964742,OL964743,OL964744,OL964751,OL964752,ON239132,ON637251 |
| 1440 | VC_463_0 | 4 | 783.1294826 | 207.4362324 | 0.790588116 | 2.84E-05 | 1 | 2.828427125 | 6 | 6 | | 1 | 2.232634022 | 0 | 0 | 0 | AM749121,DQ386162,KR153145,MT430910 |
| 1441 | VC_464_0 | 8 | 5833.561973 | 16351.71835 | 0.262947409 | 1.59E-19 | 1.732050808 | 8.246211251 | 28 | 28 | | 1 | 5.438312341 | 0 | 0 | 0 | JQ067087,KU356689,LN610577,LN610584,LN610585,MT108725,MT108731,MZ926748 |
| 1442 | VC_465_0 | 7 | 5016.018234 | 10084.03633 | 0.332185438 | 2.27E-15 | 2.236067977 | 4.123105626 | 21 | 21 | | 1 | 3.173498152 | 0 | 0 | 0 | JQ067084,KX129925,KX898399,KX898400,MN504636,MW835180,ON911540 |
| 1443 | VC_466_0 | 3 | 83.54543582 | 172.4501267 | 0.326355016 | 0.002526948 | 5.744562647 | 8.366600265 | 3 | 3 | | 1 | 6.78538697 | 0 | 0 | 0 | MW091529,MW202777,MW462191 |
| 1444 | VC_467_0 | 5 | 3000 | 1364.03657 | 0.687436952 | 3.79E-08 | 4 | 6 | 10 | 10 | | 1 | 4.996699561 | 0 | 0 | 0 | MN862068,ON529856,ON529859,ON529862,ON529864 |
| 1445 | VC_468_0 | 4 | 1049.48111 | 6287.364114 | 0.143042558 | 1.52E-05 | 2.449489743 | 6.557438524 | 6 | 6 | | 1 | 5.085990191 | 0 | 0 | 0 | MN988465,MN988541,MN988554,ON014755 |
| 1446 | VC_469_0 | 2 | 212.6706798 | 1212.743534 | 0.149199214 | 0.043418561 | 1.732050808 | 1.732050808 | 1 | 1 | | 1 | 1.732050808 | 0 | 0 | 0 | MT536174,OL960029 |
| 1447 | VC_46_0 | 2 | 202.638716 | 10996.45374 | 0.018094209 | 0.042364634 | 5.477225575 | 5.477225575 | 1 | 1 | | 1 | 5.477225575 | 0 | 0 | 0 | KF766125,KJ603229 |
| 1448 | VC_470_0 | 3 | 286.2659991 | 229.3067432 | 0.555238816 | 0.00176646 | 5.477225575 | 7 | 3 | 3 | | 1 | 6.395143169 | 0 | 0 | 0 | MN698239,MN698241,MW574966 |
| 1449 | VC_471_0 | 2 | 229.9156141 | 2630.600977 | 0.080375557 | 0.043342814 | 3.464101615 | 3.464101615 | 1 | 1 | | 1 | 3.464101615 | 0 | 0 | 0 | MW677525,OP009285 |
| 1450 | VC_472_0 | 47 | 214486.1715 | 23179.59369 | 0.902469783 | 5.18E-148 | 0 | 7.348469228 | 1081 | 1081 | | 1 | 4.367019734 | 0 | 0 | 0 | AM910650,AY078382,HE983844,HF543949,HG518155,JN254801,JX863101,KC294142,KF856712,KP233880,KR054029,LC648443,LN610578,MF768469,MG432151,MG589385,MG589386,MG845684,MH973725,MK837011,MK837012,MN508619,MN553586,MN562749,MN871476,MN871484,MT108723,MT108724,MT118288,MT119362,MT119371,MT119373,MW329986,MW512831,MW526260,MZ089732,MZ089733,MZ089738,MZ089739,MZ089740,MZ089742,OL441337,ON156559,ON376263,ON857927,ON857936,OP342787 |
| 1451 | VC_473_0 | 1 | 0 | 1085.686134 | 0 | 0 | 0 | 0 | 1 | 1 | | 1 | 0 | 0 | 0 | 0 | CP067352 |
| 1452 | VC_473_1 | 1 | 0 | 1226.243411 | 0 | 0 | 0 | 0 | 1 | 1 | | 1 | 0 | 0 | 0 | 0 | CP069348 |
| 1453 | VC_473_2 | 2 | 300 | 2636.857382 | 0.102150006 | 0.053977716 | 0 | 0 | 1 | 1 | | 1 | 0 | 0 | 0 | 0 | CP011970,LN681537 |
| 1454 | VC_473_3 | 2 | 300 | 2522.140781 | 0.10630228 | 0.05796749 | 5.656854249 | 5.656854249 | 1 | 1 | | 1 | 5.656854249 | 0 | 0 | 0 | MF547662,MF547663 |
| 1455 | VC_473_4 | 1 | 0 | 1430.683953 | 0 | 0 | 0 | 0 | 1 | 1 | | 1 | 0 | 0 | 0 | 0 | CP067347 |
| 1456 | VC_474_0 | 14 | 27300 | 4946.05951 | 0.846615072 | 4.87E-49 | 0 | 8.717797887 | 91 | 91 | | 1 | 5.938817616 | 0 | 0 | 0 | HM029250,KF926093,KM677185,KY554768,KY554769,KY554770,KY554771,KY554772,KY554773,KY554774,KY554775,KY554776,KY554777,MN552145 |
| 1457 | VC_475_0 | 3 | 900 | 10670.67508 | 0.077782843 | 0.002016249 | 5.099019514 | 7.549834435 | 3 | 3 | | 1 | 6.642987946 | 0 | 0 | 0 | OM236516,OM634653,ON133270 |
| 1458 | VC_475_1 | 3 | 900 | 14572.562 | 0.058167484 | 0.011339906 | 6.480740698 | 7.745966692 | 3 | 3 | | 1 | 7.003012458 | 0 | 0 | 0 | OK499972,OK499987,OK499994 |
| 1459 | VC_475_10 | 2 | 300 | 10228.17403 | 0.02849497 | 0.122788077 | 8.602325267 | 8.602325267 | 1 | 1 | | 1 | 8.602325267 | 0 | 0 | 0 | OK499992,ON548418 |
| 1460 | VC_475_2 | 3 | 900 | 14497.59297 | 0.058450694 | 0.011169775 | 7.874007874 | 8.366600265 | 3 | 3 | | 1 | 8.080202713 | 0 | 0 | 0 | KC481682,MT254578,ON548420 |
| 1461 | VC_475_3 | 1 | 0 | 4268.345023 | 0 | 0 | 0 | 0 | 1 | 1 | | 1 | 0 | 0 | 0 | 0 | ON881243 |
| 1462 | VC_475_4 | 1 | 0 | 4329.584955 | 0 | 0 | 0 | 0 | 1 | 1 | | 1 | 0 | 0 | 0 | 0 | MH884508 |
| 1463 | VC_475_5 | 2 | 300 | 9917.889658 | 0.02936027 | 0.060126546 | 7.745966692 | 7.745966692 | 1 | 1 | | 1 | 7.745966692 | 0 | 0 | 0 | ON548417,ON548419 |
| 1464 | VC_475_6 | 1 | 0 | 5284.575367 | 0 | 0 | 0 | 0 | 1 | 1 | | 1 | 0 | 0 | 0 | 0 | KT070867 |
| 1465 | VC_475_7 | 1 | 0 | 5343.185285 | 0 | 0 | 0 | 0 | 1 | 1 | | 1 | 0 | 0 | 0 | 0 | MW084976 |
| 1466 | VC_475_8 | 2 | 300 | 10022.13376 | 0.029063758 | 0.098153013 | 8.94427191 | 8.94427191 | 1 | 1 | | 1 | 8.94427191 | 0 | 0 | 0 | MK288021,OK499991 |

| | | | | | | | | | | | | | | | |
|---|---|---|---|---|---|---|---|---|---|---|---|---|---|---|---|
| 1467 | VC_475_9 | 1 | 0 | 5351.539625 | 0 | 0 | 0 | 0 | 1 | 1 | 1 | 0 | 0 | 0 | 0 ON548421 |
| 1468 | VC_476_0 | 19 | 17870.53028 | 4999.411823 | 0.781398143 | 9.35E-88 | 0 | 8.185352772 | 171 | 171 | 1 | 5.573713643 | 0 | 0 | 0 AY150271,DQ126339,JN391180,JQ011317,JQ691610,KC911856,KC911857,KP869107,KP869108,KX231829,MK416010,MK422448,MN187550,MT261384,MT580116,MT580117,MW205203,MW722083,OK490417 |
| 1469 | VC_477_0 | 4 | 1800 | 30.04838867 | 0.98358055 | 0.002510272 | 0 | 3.741657387 | 6 | 6 | 1 | 2.641065979 | 0 | 0 | 0 AP011616,AP011617,DQ997624,MK257744 |
| 1470 | VC_478_0 | 2 | 300 | 556.0086277 | 0.350463757 | 0.052335108 | 5.744562647 | 5.744562647 | 1 | 1 | 1 | 5.744562647 | 0 | 0 | 0 MG592415,OX241562 |
| 1471 | VC_478_1 | 1 | 0 | 352.5184081 | 0 | 0 | 0 | 0 | 1 | 1 | 1 | 0 | 0 | 0 | 0 MW073017 |
| 1472 | VC_479_0 | 17 | 19853.59535 | 1955.583382 | 0.910332094 | 1.98E-55 | 0 | 2 | 136 | 136 | 1 | 1.137372402 | 0 | 0 | 0 OM416763,OM416764,OM416765,OM416766,OM416767,OM416768,OM416769,OM416770,OM416771,OM416772,OM416773,OM416774,OM416775,OM416776,OM416777,OM416778,OM416779 |
| 1473 | VC_47_0 | 48 | 335554.7405 | 95579.23346 | 0.778307349 | 0 | 0 | 5.656854249 | 1128 | 1128 | 1 | 1.061405149 | 0 | 0 | 0 LC616031,LC616032,LC616033,LC616034,LC616035,LC616036,LC616037,LC616038,LC616039,LC616040,LC616041,LC616042,LC616043,LC616044,LC616045,LC616046,LC616047,LC616048,LC616049,LC616050,LC616051,LC616052,LC616053,LC616054,LC616055,LC616056,LC616057,LC616058,LC616059,LC616060,LC616061,LC616062,LC616063,LC616064,LC616065,LC616066,LC616067,LC616068,LC616069,LC616070,LC616071,LC616072,LC616073,LC616074,LC616075,LC616076,LC616077,LC616078 |
| 1474 | VC_480_0 | 19 | 18264.18138 | 15278.7175 | 0.544502174 | 2.23E-94 | 0 | 7.348469228 | 171 | 171 | 1 | 5.684653465 | 0 | 0 | 0 AP013541,AP013542,AP013545,KC465901,MH579717,MH598798,MH598800,MH598802,MH598803,MH598804,MH598805,MH598807,MT375520,MT375521,MT375526,MT375527,MT375528,MT375530,MT375531 |
| 1475 | VC_481_0 | 8 | 1114.725681 | 7153.139226 | 0.134826306 | 3.69E-10 | 2.828427125 | 7 | 28 | 28 | 1 | 6.087486567 | 0 | 0 | 0 AP013544,JX483873,KM199771,KT997847,MT708545,MW023914,MW980070,MZ666938 |
| 1476 | VC_482_0 | 4 | 288.8558882 | 682.1080933 | 0.297493927 | 1.58E-05 | 4 | 5.385164807 | 6 | 6 | 1 | 4.941346208 | 0 | 0 | 0 AP013546,AP013547,AP013548,MW822601 |
| 1477 | VC_484_0 | 3 | 529.0036916 | 1137.761647 | 0.317383425 | 0.001563623 | 1.414213562 | 1.732050808 | 3 | 3 | 1 | 1.626105059 | 0 | 0 | 0 KJ183191,KJ183192,KJ183193 |
| 1478 | VC_485_0 | 3 | 409.0652329 | 1303.516792 | 0.238858768 | 0.001556357 | 2 | 8.246211251 | 3 | 3 | 1 | 6.164140834 | 0 | 0 | 0 MN698242,MN698243,MN698247 |
| 1479 | VC_486_0 | 18 | 19231.23322 | 2450.376477 | 0.886983646 | 1.35E-71 | 0 | 8 | 153 | 153 | 1 | 5.592536978 | 0 | 0 | 0 KC465898,MT375525,MT647605,MW273920,MW273922,MW273923,MW273924,MW273925,MW273926,OP131293,OP131294,OP131295,OP131296,OP131297,OP131299,OP131300,OP131301,OP131302 |
| 1480 | VC_487_0 | 2 | 81.35158442 | 240.279701 | 0.252934301 | 0.050750825 | 5.099019514 | 5.099019514 | 1 | 1 | 1 | 5.099019514 | 0 | 0 | 0 MT375519,MT375529 |
| 1481 | VC_488_0 | 8 | 8400 | 20194.29086 | 0.293764935 | 2.89E-18 | 0 | 7.348469228 | 28 | 28 | 1 | 5.287198181 | 0 | 0 | 0 MN871446,MN871447,MN871448,MN871449,MN871451,MN871452,MN871453,MW394391 |
| 1482 | VC_488_1 | 8 | 8400 | 20302.90266 | 0.292653328 | 3.62E-18 | 2 | 6.92820323 | 28 | 28 | 1 | 4.525775496 | 0 | 0 | 0 MF285618,MK994515,MW021758,OK040170,OL539468,ON287372,ON287376,ON375847 |
| 1483 | VC_488_2 | 2 | 251.6850835 | 7488.692952 | 0.032515865 | 0.044889709 | 8.717797887 | 8.717797887 | 1 | 1 | 1 | 8.717797887 | 0 | 0 | 0 KF623294,MN334766 |
| 1484 | VC_489_0 | 3 | 900 | 12385.30781 | 0.067744008 | 0.017408422 | 0 | 8.94427191 | 3 | 3 | 1 | 5.96284794 | 0 | 0 | 0 MN445183,MW845760,MW879340 |
| 1485 | VC_489_1 | 11 | 16500 | 19207.23294 | 0.462091253 | 1.66E-24 | 1.732050808 | 6.708203932 | 55 | 55 | 1 | 3.302973778 | 0 | 0 | 0 KF806589,KU886222,KU886223,KU886224,KU886225,MG655267,MG655268,MG655269,MG655270,MK514281,MZ501268 |
| 1486 | VC_489_2 | 1 | 0 | 4738.35674 | 0 | 0 | 0 | 0 | 1 | 1 | 1 | 0 | 0 | 0 | 0 MT661596 |
| 1487 | VC_48_0 | 10 | 12285.83385 | 58308.57339 | 0.174034096 | 6.28E-31 | 0 | 2.828427125 | 45 | 45 | 1 | 1.87260851 | 0 | 0 | 0 LC645428,LC645429,LC645430,LC645431,LC645432,LC645433,LC645434,LC645435,LC645436,LC645437 |
| 1488 | VC_490_0 | 2 | 300 | 29137.2416 | 0.010191172 | 0.046551366 | 4.123105626 | 4.123105626 | 1 | 1 | 1 | 4.123105626 | 0 | 0 | 0 AP014714,AP014715 |
| 1489 | VC_490_1 | 5 | 3000 | 64154.24373 | 0.044673275 | 3.84E-08 | 5.567764363 | 8.426149773 | 10 | 10 | 1 | 7.704845158 | 0 | 0 | 0 MN087708,MN882610,MZ348423,OL989991,ON630910 |
| 1490 | VC_490_2 | 1 | 0 | 14602.08382 | 0 | 0 | 0 | 0 | 1 | 1 | 1 | 0 | 0 | 0 | 0 KF835987 |
| 1491 | VC_490_3 | 2 | 300 | 21981.67049 | 0.013463982 | 0.042968507 | 7.937253933 | 7.937253933 | 1 | 1 | 1 | 7.937253933 | 0 | 0 | 0 HE956709,KY593455 |
| 1492 | VC_490_4 | 1 | 0 | 10553.64871 | 0 | 0 | 0 | 0 | 1 | 1 | 1 | 0 | 0 | 0 | 0 KP202158 |
| 1493 | VC_490_5 | 1 | 0 | 10630.23158 | 0 | 0 | 0 | 0 | 1 | 1 | 1 | 0 | 0 | 0 | 0 MH059636 |
| 1494 | VC_490_6 | 1 | 0 | 10869.36129 | 0 | 0 | 0 | 0 | 1 | 1 | 1 | 0 | 0 | 0 | 0 MT920315 |
| 1495 | VC_491_0 | 36 | 189000 | 165177.5171 | 0.53363071 | 0 | 2.236067977 | 11.18033989 | 630 | 612 | 0.971428571 | 5.983972297 | 0 | 0 | 0 CP062992,KR269720,KT239446,KX258185,KY000080,LR877331,MG751100,MH333064,MH729874,MK421971,MN101226,MN101229,MN101230,MN434095,MT580897,MT701588,MW021752,MW239157,MZ560702,MZ602148,MZ890186,OK499980,OK499981,OK499996,OK499997,OK631811,OL539458,OL870318,OL870319,OM337876,OM337877,OM630219,ON550260,OP331213,OP617748,OU509536 |
| 1496 | VC_491_1 | 1 | 0 | 13554.54626 | 0 | 0 | 0 | 0 | 1 | 1 | 1 | 0 | 0 | 0 | 0 MN101225 |
| 1497 | VC_492_0 | 2 | 300 | 18716.53641 | 0.015775743 | 0.3359603 | 3.16227766 | 3.16227766 | 1 | 1 | 1 | 3.16227766 | 0 | 0 | 0 HQ698922,MK240351 |
| 1498 | VC_492_1 | 20 | 57000 | 79480.64906 | 0.417641625 | 3.53E-15 | 0 | 5.099019514 | 190 | 190 | 1 | 4.047077882 | 0 | 0 | 0 CAKLQC020000001,CAKLQD020000001,MH165274,MH460829,MH713599,MN648195,MN662249,MN709128,MN723850,MN732883,MN782535,MT385367,MT409116,MT741943,OL763420,OL770258,OL770259,OL770261,OL770262,OP585103 |
| 1499 | VC_492_2 | 8 | 8400 | 60408.91322 | 0.122077208 | 0.01213426 | 1 | 5 | 28 | 28 | 1 | 3.852043461 | 0 | 0 | 0 MK278860,MN864865,MT949699,MW388001,MW388002,MW388003,MW388004,OL770260 |
| 1500 | VC_492_3 | 1 | 0 | 9804.541038 | 0 | 0 | 0 | 0 | 1 | 1 | 1 | 0 | 0 | 0 | 0 GU911519 |
| 1501 | VC_492_4 | 1 | 0 | 9696.805758 | 0 | 0 | 0 | 0 | 1 | 1 | 1 | 0 | 0 | 0 | 0 HM004124 |
| 1502 | VC_492_5 | 1 | 0 | 7513.711895 | 0 | 0 | 0 | 0 | 1 | 1 | 1 | 0 | 0 | 0 | 0 HM032710 |
| 1503 | VC_492_6 | 1 | 0 | 7211.166009 | 0 | 0 | 0 | 0 | 1 | 1 | 1 | 0 | 0 | 0 | 0 HM114315 |
| 1504 | VC_493_0 | 1 | 0 | 2878.525935 | 0 | 0 | 0 | 0 | 1 | 1 | 1 | 0 | 0 | 0 | 0 KT932418 |
| 1505 | VC_493_1 | 1 | 0 | 5442.420425 | 0 | 0 | 0 | 0 | 1 | 1 | 1 | 0 | 0 | 0 | 0 MG592562 |
| 1506 | VC_493_10 | 1 | 0 | 4101.868649 | 0 | 0 | 0 | 0 | 1 | 1 | 1 | 0 | 0 | 0 | 0 HQ634156 |
| 1507 | VC_493_11 | 1 | 0 | 2871.129988 | 0 | 0 | 0 | 0 | 1 | 1 | 1 | 0 | 0 | 0 | 0 KX507046 |
| 1508 | VC_493_12 | 2 | 300 | 8856.782758 | 0.032762599 | 0.044420764 | 2.645751311 | 2.645751311 | 1 | 1 | 1 | 2.645751311 | 0 | 0 | 0 MW824371,MW824372 |
| 1509 | VC_493_13 | 1 | 0 | 4797.982562 | 0 | 0 | 0 | 0 | 1 | 1 | 1 | 0 | 0 | 0 | 0 MW824376 |
| 1510 | VC_493_14 | 1 | 0 | 4971.14331 | 0 | 0 | 0 | 0 | 1 | 1 | 1 | 0 | 0 | 0 | 0 HQ316579 |
| 1511 | VC_493_15 | 1 | 0 | 4975.972474 | 0 | 0 | 0 | 0 | 1 | 1 | 1 | 0 | 0 | 0 | 0 HQ634195 |
| 1512 | VC_493_16 | 1 | 0 | 4049.83305 | 0 | 0 | 0 | 0 | 1 | 1 | 1 | 0 | 0 | 0 | 0 MG592553 |
| 1513 | VC_493_2 | 1 | 0 | 4362.879406 | 0 | 0 | 0 | 0 | 1 | 1 | 1 | 0 | 0 | 0 | 0 MG592529 |
| 1514 | VC_493_3 | 1 | 0 | 3738.780113 | 0 | 0 | 0 | 0 | 1 | 1 | 1 | 0 | 0 | 0 | 0 OX241437 |
| 1515 | VC_493_4 | 1 | 0 | 4179.119926 | 0 | 0 | 0 | 0 | 1 | 1 | 1 | 0 | 0 | 0 | 0 OX241455 |
| 1516 | VC_493_5 | 1 | 0 | 2716.689211 | 0 | 0 | 0 | 0 | 1 | 1 | 1 | 0 | 0 | 0 | 0 AP014858 |
| 1517 | VC_493_6 | 1 | 0 | 3317.739637 | 0 | 0 | 0 | 0 | 1 | 1 | 1 | 0 | 0 | 0 | 0 MG592495 |
| 1518 | VC_493_7 | 1 | 0 | 3641.006886 | 0 | 0 | 0 | 0 | 1 | 1 | 1 | 0 | 0 | 0 | 0 HQ634157 |
| 1519 | VC_493_8 | 1 | 0 | 3217.344474 | 0 | 0 | 0 | 0 | 1 | 1 | 1 | 0 | 0 | 0 | 0 MG592441 |
| 1520 | VC_493_9 | 1 | 0 | 3210.396277 | 0 | 0 | 0 | 0 | 1 | 1 | 1 | 0 | 0 | 0 | 0 ON922990 |
| 1521 | VC_495_0 | 39 | 222300 | 499350.4637 | 0.308043868 | 2.92E-97 | 0 | 7.681145748 | 741 | 741 | 1 | 3.952205067 | 0 | 0 | 0 KY883638,KY883639,KY883640,MH310933,MH310935,MN419153,MW794150,MW794151,MW794152,MW794153,MW794154,MW794155,MW794156,MW794157,MW794158,MW794159,MW794160,MW794161,MW794162,MW794163,MW794164,MW794165,MW794166,MW794167,MW794168,MW794169,MW794170,MW794171,MW794172,MW794173,MW794176,MW794177,MW794179,MW794180,MW794182,MW794183,MW794188,MW794190,MZ292654 |
| 1522 | VC_495_1 | 41 | 246000 | 504075.9346 | 0.327966795 | 2.55E-115 | 0 | 10.44030651 | 820 | 747 | 0.91097561 | 6.478268626 | 0 | 0 | 0 HQ641347,HQ641348,HQ641349,HQ641350,HQ641351,HQ641352,HQ641353,HQ641354,KY065147,KY883634,KY883635,KY883636,KY883637,MH310934,MH310936,MN402506,MW794140,MW794141,MW794142,MW794143,MW794144,MW794145,MW794146,MW794147,MW794148,MW794149,MW794174,MW794175,MW794178,MW794181,MW794184,MW794185,MW794186,MW794187,MW794189,MW794191,MW794192,MW911612,MW911613,MW911614,MW911615 |
| 1523 | VC_496_0 | 1 | 0 | 1121.016836 | 0 | 0 | 0 | 0 | 1 | 1 | 1 | 0 | 0 | 0 | 0 KF302034 |
| 1524 | VC_496_1 | 1 | 0 | 794.5361574 | 0 | 0 | 0 | 0 | 1 | 1 | 1 | 0 | 0 | 0 | 0 MF370964 |
| 1525 | VC_496_2 | 1 | 0 | 828.2632615 | 0 | 0 | 0 | 0 | 1 | 1 | 1 | 0 | 0 | 0 | 0 KR534323 |
| 1526 | VC_496_3 | 1 | 0 | 1126.219863 | 0 | 0 | 0 | 0 | 1 | 1 | 1 | 0 | 0 | 0 | 0 KX822733 |
| 1527 | VC_497_0 | 1 | 0 | 1562.597451 | 0 | 0 | 0 | 0 | 1 | 1 | 1 | 0 | 0 | 0 | 0 MG592459 |
| 1528 | VC_497_1 | 1 | 0 | 2869.974651 | 0 | 0 | 0 | 0 | 1 | 1 | 1 | 0 | 0 | 0 | 0 MG592473 |
| 1529 | VC_498_0 | 8 | 2780.635543 | 4193.97987 | 0.398679408 | 2.32E-18 | 1 | 7.681145748 | 28 | 28 | 1 | 5.824701134 | 0 | 0 | 0 AP014887,AY328852,AY328853,KF854250,KX196154,KY622015,MG592395,OL658619 |
| 1530 | VC_499_0 | 24 | 30583.14801 | 5109.085961 | 0.85685721 | 1.06E-103 | 0 | 9.38083152 | 276 | 263 | 0.952898551 | 7.681843724 | 0 | 0 | 0 KT160311,MG592392,MG592401,MG592414,MG592451,MG592461,MG592462,MG592467,MG592468,MG592472,MG592499,MG592526,MG592531,MG592541,MG592542,MG592547,MG592561,MG592572,MG592590,MG592592,MG592611,MW824380,MW824381,MW824382 |
| 1531 | VC_49_0 | 36 | 135240.7621 | 86094.16892 | 0.611023129 | 0 | 0 | 6.244997998 | 630 | 630 | 1 | 3.370415202 | 0 | 0 | 0 LC727840,LC727841,LC727842,LC727843,LC727844,LC727845,LC727846,LC727847,LC727848,LC727849,LC727850,LC727851,LC727852,LC727853,LC727854,LC727855,LC727856,LC727857,LC727858,LC727859,LC727860,LC727861,LC727862,LC727863,LC727864,LC727865,LC727866,LC727867,LC727868,LC727869,LC727870,LC727871,LC727872,LC727873,LC727874,LC727875 |
| 1532 | VC_4_0 | 7 | 3658.278678 | 965.6263913 | 0.791166476 | 4.23E-15 | 0 | 3.741657387 | 21 | 21 | 1 | 2.14880429 | 0 | 0 | 0 DQ426904,DQ426905,JN255163,KP137432,KP137434,KP137436,KP137438 |
| 1533 | VC_500_0 | 3 | 548.8343665 | 114.1463582 | 0.827828542 | 0.003082402 | 0 | 2.828427125 | 3 | 3 | 1 | 1.885618083 | 0 | 0 | 0 OM287554,OM732336,OM732337 |
| 1534 | VC_501_0 | 16 | 15471.45467 | 23.82333559 | 0.998462542 | 5.06E-10 | 0 | 4.582575695 | 120 | 120 | 1 | 3.609799117 | 0 | 0 | 0 AP014888,MG592509,MG592546,MG592620,MG592623,OX241426,OX241428,OX241430,OX241431,OX241432,OX241441,OX241448,OX241449,OX241494,OX241567,OX241568 |
| 1535 | VC_502_0 | 2 | 300 | 3768.636081 | 0.073734783 | 0.048286377 | 5 | 5 | 1 | 1 | 1 | 5 | 0 | 0 | 0 S2-wms_128962,S6-wms_138342 |
| 1536 | VC_502_1 | 6 | 1965.585679 | 6436.022048 | 0.233953517 | 1.36E-09 | 4.123105626 | 8.888194417 | 15 | 15 | 1 | 7.288708057 | 0 | 0 | 0 MT774375,MT774376,MT774379,MT774380,MT774382,S4-wms_73990 |
| 1537 | VC_502_2 | 1 | 0 | 1634.059385 | 0 | 0 | 0 | 0 | 1 | 1 | 1 | 0 | 0 | 0 | 0 MT774401 |
| 1538 | VC_502_3 | 2 | 300 | 3843.04071 | 0.072410585 | 0.048286377 | 4.69041576 | 4.69041576 | 1 | 1 | 1 | 4.69041576 | 0 | 0 | 0 MT774399,MT774403 |
| 1539 | VC_502_4 | 3 | 592.2606988 | 5349.540316 | 0.099676966 | 0.002846525 | 7.348469228 | 8.602325267 | 3 | 3 | 1 | 7.94160079 | 0 | 0 | 0 MT774395,MT774397,MT774398 |
| 1540 | VC_502_5 | 2 | 227.3246905 | 3988.923926 | 0.053916339 | 0.064220362 | 8.888194417 | 8.888194417 | 1 | 1 | 1 | 8.888194417 | 0 | 0 | 0 MT774396,MT774405 |
| 1541 | VC_502_6 | 2 | 260.7048296 | 2981.24832 | 0.080415977 | 0.048286377 | 7.280109889 | 7.280109889 | 1 | 1 | 1 | 7.280109889 | 0 | 0 | 0 MT774404,MT774407 |
| 1542 | VC_502_7 | 1 | 0 | 2103.9841 | 0 | 0 | 0 | 0 | 1 | 1 | 1 | 0 | 0 | 0 | 0 MT774400 |
| 1543 | VC_505_0 | 3 | 900 | 122.4445884 | 0.880243301 | 0.031801285 | 0 | 1.732050808 | 3 | 3 | 1 | 1.154700538 | 0 | 0 | 0 MW353175,MW353176,MW353177 |
| 1544 | VC_506_0 | 8 | 8400 | 2988.501572 | 0.73758606 | 3.59E-17 | 1.414213562 | 7.141428429 | 28 | 28 | 1 | 4.73885141 | 0 | 0 | 0 KR560069,MK580972,MN095772,MN098328,MZ042490,MZ326868,MZ398248,OL702939 |
| 1545 | VC_506_1 | 1 | 0 | 1745.127974 | 0 | 0 | 0 | 0 | 1 | 1 | 1 | 0 | 0 | 0 | 0 ON758385 |
| 1546 | VC_507_0 | 8 | 8038.332233 | 9961.642226 | 0.446574647 | 1.48E-19 | 0 | 4 | 28 | 28 | 1 | 2.950966002 | 0 | 0 | 0 AP018462,AP018463,KY130461,MH001446,MH001450,MK494114,MK494131,MK494132 |
| 1547 | VC_508_0 | 32 | 148680.6033 | 15416.97348 | 0.906049963 | 2.98E-235 | 0 | 3.31662479 | 496 | 496 | 1 | 1.053959571 | 0 | 0 | 0 AY129334,DQ398048,JN618996,JN698991,JN699004,KF024722,KM101117,KR997932,KT365402,KT880194,KX443696,MG757162,MG812490,MG812491,MH001452,MH051249,MH590591,MH697584,MH727546,MK061411,MK620898,MK814751,MN234225,MN428049,MN586045,MN586046,MN813678,MT639652,MW712731,MZ456036,ON970587,ON970593 |
| 1548 | VC_509_0 | 37 | 199764.5847 | 6412.154864 | 0.968899717 | 2.62E-283 | 0 | 4.358898944 | 666 | 666 | 1 | 2.494687492 | 0 | 0 | 0 DQ398049,EU816589,FJ641182,JF704095,JN698992,JN699003,JN699006,JN699018,KF493879,KJ194581,KJ194584,KP027207,KR080203,KR080205,MG839014,MG920059,MG925338,MG925353,MH051265,MH316565,MH513976,MH576977,MK061414,MK359357,MN096359,MN096360,MN234223,MN444872,MN444873,MT310871,MT310892,MT952851,MW924644,MZ171375,MZ171381,OK318960,OP434441 |
| 1549 | VC_50_0 | 9 | 6174.451903 | 4988.253745 | 0.553132197 | 4.82E-23 | 1 | 4.69041576 | 36 | 36 | 1 | 3.456672925 | 0 | 0 | 0 AF083977,GQ357916,KP010268,LR595867,MT374852,MT374853,MT374854,MT374856,MT591577 |
| 1550 | VC_510_0 | 12 | 10993.69262 | 16745.87982 | 0.396318027 | 1.33E-43 | 0 | 9 | 66 | 64 | 0.96969697 | 7.028033555 | 0 | 0 | 0 MH153810,MK801725,MN234196,MN369758,MN908693,MT657335,OK216879,OM105886,ON081333,ON108644,ON260813,OP434461 |
| 1551 | VC_511_0 | 3 | 598.7489424 | 4824.004194 | 0.110414199 | 0.001413172 | 6.244997998 | 7.549834435 | 3 | 3 | 1 | 6.758524377 | 0 | 0 | 0 JN698995,OP172873,OP172874 |
| 1552 | VC_513_0 | 32 | 106519.9272 | 48277.93262 | 0.688122739 | 1.86E-283 | 0 | 7.071067812 | 496 | 496 | 1 | 4.711232911 | 0 | 0 | 0 AP018485,JF704111,JF957058,KC172839,KJ510415,KM592966,KT326767,KX640831,KX670828,KX683423,KY464936,LC730317,MF373840,MG812492,MH020248,MH047632,MH047633,MH051259,MH316563,MH976517,MK284519,MK494127,MK524496,MN062706,MN062712,MN586063,MT310878,MT310879,MT310880,MT310881,MW712718,ON970623 |
| 1553 | VC_514_0 | 75 | 616547.0641 | 39213.56701 | 0.940201401 | 0 | 0 | 8.485281374 | 2775 | 2775 | 1 | 5.184703418 | 0 | 0 | 0 AY129332,HM755814,JF704098,JF704101,JF704114,KC661272,KC661281,KF279418,KM101119,KM233454,KM233455,KP017310,KP017311,KP027202,KT359365,KT381277,KU255188,KU578077,KU984914,KU985091,KX458237,KX507362,KX523125,KX664448,KX670788,KX808129,KY965063,MF185729,MF185732,MF185733,MG757161,MG925341,MG925355,MH020238,MH077583,MH155864,MH338240,MH371121,MH399776,MH450119,MH450124,MH450126,MH450128,MH513975,MH536813,MH536825,MH576949,MH651179,MH727554,MK112543,MK279903,MK279913,MK284518,MK310140,MK359341,MK433273,MK494096,MK494128,MK875800,MK967399,MN062718,MN096368,MN284900,MN497953,MN585969,MN585984,MN586048,MN813699,MT657331,MT897907,MW507128,MZ681518,MZ747514,OK318959,OK999978 |
| 1554 | VC_515_0 | 16 | 25202.01822 | 41233.9628 | 0.379342908 | 5.71E-78 | 0 | 6.8556546 | 120 | 120 | 1 | 4.47315232 | 0 | 0 | 0 JQ512844,JX307704,JX411619,KC661279,KJ156985,KM101120,MF773750,MH001459,MK494124,MN586038,MN807247,MT522002,MZ322017,MZ355723,MZ622159,ON337196 |
| 1555 | VC_516_0 | 3 | 800.3460439 | 915.4966441 | 0.466444884 | 0.001432024 | 0 | 4.795831523 | 3 | 3 | 1 | 3.197221016 | 0 | 0 | 0 MW314854,MW584159,MW584197 |
| 1556 | VC_517_0 | 18 | 35131.60624 | 184530.4201 | 0.159934818 | 2.46E-95 | 0 | 8.485281374 | 153 | 153 | 1 | 4.577889885 | 0 | 0 | 0 JF704093,JN243857,KX550442,KX579975,KX641260,KY083059,MF668278,MK279898,MK359310,MK359322,MK494115,MK494117,MK494129,MK875799,MN284905,MN813697,MZ322023,ON970614 |
| 1557 | VC_517_1 | 108 | 1122660.617 | 200717.3553 | 0.848329533 | 0 | 0 | 9.433981132 | 5778 | 5752 | 0.995500173 | 6.241694953 | 0 | 0 | 0 GQ303263,HM152766,JF792674,JN243856,JN561150,JN699015,JQ809701,JQ896627,JX307702,JX307703,KC661271,KF024733,KF841475,KF841476,KF954506,KF959563,KJ510414,KP027198,KP057620,KT365397,KT716493,KU055616,KU867906,KU985092,KU985094,KX458236,KX550441,KX557234,KX619651,KX648373,KX808130,KX817174,KX817175,KY006473,KY083058,KY083060,KY083061,KY083062,KY083063,KY204244,KY204245,KY204246,KY204247,KY204248,KY204249,KY204250,KY204251,KY471266,KY549155,MF919496,MF919509,MG099942,MG944217,MG944224,MG962373,MH020249,MH051247,MH077581,MH155875,MH155879,MH230877,MH399777,MH399785,MH479909,MH479911,MH509443,MH536817,MH651170,MH697588,MH727563,MH825710,MK279902,MK310145,MK359301,MK359323,MK359325,MK359340,MK359356,MK494088,MK494093,MK494094,MK494097,MK494101,MK494125,MK494126,MK878895,MK878905,MN728684,MN813696,MT114158,MT490373,MT498056,MT553338,MT553346,MT723939,MT818417,MT889382,MT889386,MT889390,MW291019,MZ005673,MZ171380,MZ322025,MZ355725,MZ747509,ON637771,ON970565,OP297529 |
| 1558 | VC_518_0 | 6 | 2390.572436 | 16066.30531 | 0.129522039 | 1.99E-11 | 2.236067977 | 7.615773106 | 15 | 15 | 1 | 6.25639766 | 0 | 0 | 0 AP018487,FJ168659,HM152765,JN699001,MN234195,MN234236 |
| 1559 | VC_518_1 | 2 | 144.5496243 | 3695.751541 | 0.037640179 | 0.042812394 | 7.615773106 | 7.615773106 | 1 | 1 | 1 | 7.615773106 | 0 | 0 | 0 AY129333,KP027201 |
| 1560 | VC_519_0 | 39 | 161534.1835 | 36523.81713 | 0.815590297 | 0.00E+00 | 0 | 8.660254038 | 741 | 741 | 1 | 4.793252917 | 0 | 0 | 0 JN412591,JN572061,KC691256,KF841477,KP027200,KR080195,KT347313,KU985090,KX641262,MF133446,MF140399,MF281061,MF373843,MF472894,MF919492,MG198785,MK016499,MK494122,MK524508,MK814758,MK820639,MK919480,MK937604,MN010757,MN096356,MN585977,MN807248,MN807249,MN807250,MN892484,MN892486,MN908683,MT024866,MT498061,MW055902,ON637760,ON970574,OP172867,OP434454 |
| 1561 | VC_51_0 | 10 | 2834.000518 | 13442.39718 | 0.174117183 | 1.18E-25 | 1.414213562 | 8.544003745 | 45 | 45 | 1 | 6.624197138 | 0 | 0 | 0 AB285204,JQ182728,LR595868,LR597641,MF807953,MW057859,ON470578,ON470598,ON470618,OV696610 |
| 1562 | VC_520_0 | 1 | 0 | 11810.96613 | 0 | 0 | 0 | 0 | 1 | 1 | 1 | 0 | 0 | 0 | 0 KT309034 |
| 1563 | VC_520_1 | 1 | 0 | 10197.06664 | 0 | 0 | 0 | 0 | 1 | 1 | 1 | 0 | 0 | 0 | 0 MT889374 |
| 1564 | VC_520_10 | 1 | 0 | 10576.02655 | 0 | 0 | 0 | 0 | 1 | 1 | 1 | 0 | 0 | 0 | 0 MN369752 |
| 1565 | VC_520_11 | 1 | 0 | 11007.27855 | 0 | 0 | 0 | 0 | 1 | 1 | 1 | 0 | 0 | 0 | 0 MN585995 |
| 1566 | VC_520_12 | 1 | 0 | 10563.36717 | 0 | 0 | 0 | 0 | 1 | 1 | 1 | 0 | 0 | 0 | 0 MN735431 |
| 1567 | VC_520_13 | 1 | 0 | 10590.446 | 0 | 0 | 0 | 0 | 1 | 1 | 1 | 0 | 0 | 0 | 0 KT281793 |
| 1568 | VC_520_14 | 1 | 0 | 11296.88435 | 0 | 0 | 0 | 0 | 1 | 1 | 1 | 0 | 0 | 0 | 0 MT684595 |
| 1569 | VC_520_15 | 2 | 173.328997 | 21978.54059 | 0.007824576 | 0.042395997 | 9 | 9 | 1 | 0 | 0 | 9 | 0 | 0 | 0 MF668270,MT684593 |
| 1570 | VC_520_16 | 1 | 0 | 10296.16512 | 0 | 0 | 0 | 0 | 1 | 1 | 1 | 0 | 0 | 0 | 0 KX808131 |
| 1571 | VC_520_17 | 1 | 0 | 8438.314548 | 0 | 0 | 0 | 0 | 1 | 1 | 1 | 0 | 0 | 0 | 0 MN369739 |
| 1572 | VC_520_18 | 1 | 0 | 10684.57847 | 0 | 0 | 0 | 0 | 1 | 1 | 1 | 0 | 0 | 0 | 0 MN586001 |
| 1573 | VC_520_19 | 2 | 220.1261144 | 22286.86624 | 0.009780343 | 0.04238448 | 7.348469228 | 7.348469228 | 1 | 1 | 1 | 7.348469228 | 0 | 0 | 0 KM652553,MN369749 |
| 1574 | VC_520_2 | 1 | 0 | 10487.15943 | 0 | 0 | 0 | 0 | 1 | 1 | 1 | 0 | 0 | 0 | 0 KR824843 |
| 1575 | VC_520_20 | 1 | 0 | 11180.31481 | 0 | 0 | 0 | 0 | 1 | 1 | 1 | 0 | 0 | 0 | 0 JN699012 |
| 1576 | VC_520_21 | 1 | 0 | 10618.41653 | 0 | 0 | 0 | 0 | 1 | 1 | 1 | 0 | 0 | 0 | 0 MH669001 |
| 1577 | VC_520_22 | 1 | 0 | 10108.90708 | 0 | 0 | 0 | 0 | 1 | 1 | 1 | 0 | 0 | 0 | 0 OP068340 |
| 1578 | VC_520_23 | 2 | 178.4560001 | 22956.03437 | 0.007713851 | 0.042365924 | 8.717797887 | 8.717797887 | 1 | 1 | 1 | 8.717797887 | 0 | 0 | 0 JN859129,KF024723 |
| 1579 | VC_520_24 | 1 | 0 | 11039.85058 | 0 | 0 | 0 | 0 | 1 | 1 | 1 | 0 | 0 | 0 | 0 EF536069 |
| 1580 | VC_520_25 | 1 | 0 | 9676.551631 | 0 | 0 | 0 | 0 | 1 | 1 | 1 | 0 | 0 | 0 | 0 MH590598 |
| 1581 | VC_520_26 | 3 | 858.7458015 | 32169.37056 | 0.026000447 | 0.001401023 | 5.099019514 | 6.244997998 | 3 | 3 | 1 | 5.696193386 | 0 | 0 | 0 HQ728524,JN698996,JX121091 |
| 1582 | VC_520_27 | 2 | 300 | 20671.01852 | 0.014305457 | 0.042383652 | 0 | 0 | 1 | 1 | 1 | 0 | 0 | 0 | 0 KT281792,KY471267 |
| 1583 | VC_520_28 | 2 | 300 | 21668.51747 | 0.013655906 | 0.042384694 | 3.605551275 | 3.605551275 | 1 | 1 | 1 | 3.605551275 | 0 | 0 | 0 JN020141,MT114167 |
| 1584 | VC_520_29 | 1 | 0 | 9064.079229 | 0 | 0 | 0 | 0 | 1 | 1 | 1 | 0 | 0 | 0 | 0 KM597530 |
| 1585 | VC_520_3 | 1 | 0 | 9522.278909 | 0 | 0 | 0 | 0 | 1 | 1 | 1 | 0 | 0 | 0 | 0 MF668287 |
| 1586 | VC_520_30 | 2 | 161.7085748 | 22122.93015 | 0.007256504 | 0.045550803 | 8.831760866 | 8.831760866 | 1 | 1 | 1 | 8.831760866 | 0 | 0 | 0 MH825707,ON526976 |
| 1587 | VC_520_31 | 11 | 9796.994206 | 99423.26537 | 0.089699422 | 2.18E-36 | 0 | 9.848857802 | 55 | 33 | 0.6 | 8.51832669 | 0 | 0 | 0 FJ174690,GU060500,JF937093,KT281795,MH020235,MH077580,MK016494,MN585973,MW924649,MZ681519,OP297536 |
| 1588 | VC_520_32 | 1 | 0 | 11923.9228 | 0 | 0 | 0 | 0 | 1 | 1 | 1 | 0 | 0 | 0 | 0 MH651178 |
| 1589 | VC_520_33 | 2 | 191.9853909 | 21888.68256 | 0.008694728 | 0.042384286 | 8.185352772 | 8.185352772 | 1 | 1 | 1 | 8.185352772 | 0 | 0 | 0 MH371123,MN369755 |
| 1590 | VC_520_34 | 1 | 0 | 11816.1091 | 0 | 0 | 0 | 0 | 1 | 1 | 1 | 0 | 0 | 0 | 0 MH077578 |
| 1591 | VC_520_35 | 3 | 848.2095059 | 32936.25638 | 0.025106494 | 0.001401434 | 3 | 6.480740698 | 3 | 3 | 1 | 5.075101115 | 0 | 0 | 0 KR997930,KR997931,KR997968 |
| 1592 | VC_520_36 | 1 | 0 | 9777.770827 | 0 | 0 | 0 | 0 | 1 | 1 | 1 | 0 | 0 | 0 | 0 MH744415 |
| 1593 | VC_520_37 | 1 | 0 | 10379.91041 | 0 | 0 | 0 | 0 | 1 | 1 | 1 | 0 | 0 | 0 | 0 JF937102 |
| 1594 | VC_520_38 | 6 | 3172.802843 | 57905.53997 | 0.051946446 | 1.75E-11 | 3.31662479 | 10.04987562 | 15 | 13 | 0.866666667 | 7.79849748 | 0 | 0 | 0 KJ025956,MH155865,MH155871,MH590602,MK524517,MT310893 |
| 1595 | VC_520_39 | 1 | 0 | 11733.19194 | 0 | 0 | 0 | 0 | 1 | 1 | 1 | 0 | 0 | 0 | 0 MH371120 |
| 1596 | VC_520_4 | 1 | 0 | 10409.715 | 0 | 0 | 0 | 0 | 1 | 1 | 1 | 0 | 0 | 0 | 0 MH155866 |
| 1597 | VC_520_40 | 4 | 1188.670512 | 28795.04166 | 0.039643874 | 1.22E-05 | 5.567764363 | 7.810249676 | 6 | 6 | 1 | 7.136524503 | 0 | 0 | 0 KU865302,KY965064,MH371115,MT553345 |
| 1598 | VC_520_41 | 2 | 300 | 21208.72784 | 0.013947826 | 0.042385428 | 3.741657387 | 3.741657387 | 1 | 1 | 1 | 3.741657387 | 0 | 0 | 0 MH651183,MN586011 |
| 1599 | VC_520_42 | 1 | 0 | 7531.032504 | 0 | 0 | 0 | 0 | 1 | 1 | 1 | 0 | 0 | 0 | 0 MH399771 |
| 1600 | VC_520_43 | 3 | 610.5187355 | 30956.55654 | 0.019340364 | 0.001629566 | 6.92820323 | 8.717797887 | 3 | 3 | 1 | 8.082775461 | 0 | 0 | 0 JN020143,ON456356,ON970562 |
| 1601 | VC_520_44 | 1 | 0 | 11639.30892 | 0 | 0 | 0 | 0 | 1 | 1 | 1 | 0 | 0 | 0 | 0 KY348865 |
| 1602 | VC_520_45 | 7 | 4112.764836 | 60685.73201 | 0.063470065 | 5.05E-15 | 5.744562647 | 9.110433579 | 21 | 17 | 0.80952381 | 7.861683874 | 0 | 0 | 0 KT895281,MG872841,MH316564,MH399784,MH651169,MH834617,MK494110 |
| 1603 | VC_520_46 | 1 | 0 | 9830.676191 | 0 | 0 | 0 | 0 | 1 | 1 | 1 | 0 | 0 | 0 | 0 MZ274306 |
| 1604 | VC_520_47 | 1 | 0 | 10021.24975 | 0 | 0 | 0 | 0 | 1 | 1 | 1 | 0 | 0 | 0 | 0 MT684586 |

| | | | | | | | | | | | | | | | | | |
|---|---|---|---|---|---|---|---|---|---|---|---|---|---|---|---|---|---|
| 1605 | VC_520_48 | 1 | 0 | 9422.671119 | 0 | 0 | 0 | 0 | 1 | 1 | | 1 | 0 | 0 | 0 | 0 | MZ171378 |
| 1606 | VC_520_49 | 1 | 0 | 10598.13752 | 0 | 0 | 0 | 0 | 1 | 1 | | 1 | 0 | 0 | 0 | 0 | KM101122 |
| 1607 | VC_520_5 | 1 | 0 | 10721.87687 | 0 | 0 | 0 | 0 | 1 | 1 | | 1 | 0 | 0 | 0 | 0 | MH727548 |
| 1608 | VC_520_50 | 1 | 0 | 10749.61604 | 0 | 0 | 0 | 0 | 1 | 1 | | 1 | 0 | 0 | 0 | 0 | MT897904 |
| 1609 | VC_520_51 | 1 | 0 | 10465.88992 | 0 | 0 | 0 | 0 | 1 | 1 | | 1 | 0 | 0 | 0 | 0 | MH051257 |
| 1610 | VC_520_52 | 1 | 0 | 8236.15923 | 0 | 0 | 0 | 0 | 1 | 1 | | 1 | 0 | 0 | 0 | 0 | KC736071 |
| 1611 | VC_520_53 | 1 | 0 | 9801.354288 | 0 | 0 | 0 | 0 | 1 | 1 | | 1 | 0 | 0 | 0 | 0 | KY385384 |
| 1612 | VC_520_54 | 1 | 0 | 9383.172625 | 0 | 0 | 0 | 0 | 1 | 1 | | 1 | 0 | 0 | 0 | 0 | OM105885 |
| 1613 | VC_520_55 | 1 | 0 | 10549.09198 | 0 | 0 | 0 | 0 | 1 | 1 | | 1 | 0 | 0 | 0 | 0 | OP297542 |
| 1614 | VC_520_56 | 1 | 0 | 8986.963781 | 0 | 0 | 0 | 0 | 1 | 1 | | 1 | 0 | 0 | 0 | 0 | KM279937 |
| 1615 | VC_520_57 | 1 | 0 | 9563.123029 | 0 | 0 | 0 | 0 | 1 | 1 | | 1 | 0 | 0 | 0 | 0 | MK359343 |
| 1616 | VC_520_58 | 3 | 714.5275273 | 33340.52854 | 0.02098154 | 0.0014007 | 0 | 8.185352772 | 3 | 3 | | 1 | 5.456901848 | 0 | 0 | 0 | DQ398050,EU816590,ON970595 |
| 1617 | VC_520_59 | 5 | 2704.14804 | 56150.51157 | 0.045946201 | 2.56E-08 | 1.732050808 | 7.211102551 | 10 | 10 | | 1 | 5.04516783 | 0 | 0 | 0 | JN542517,MH001448,MN585987,MN586005,MZ681512 |
| 1618 | VC_520_6 | 3 | 900 | 31659.49111 | 0.02764171 | 0.001401594 | 0 | 1.414213562 | 3 | 3 | | 1 | 0.942809042 | 0 | 0 | 0 | MN586004,MT684597,MT684601 |
| 1619 | VC_520_60 | 2 | 227.3489848 | 24667.77269 | 0.00913227 | 0.042387674 | 6.8556546 | 6.8556546 | 1 | 1 | | 1 | 6.8556546 | 0 | 0 | 0 | DQ398045,KT279576 |
| 1620 | VC_520_61 | 1 | 0 | 11146.06478 | 0 | 0 | 0 | 0 | 1 | 1 | | 1 | 0 | 0 | 0 | 0 | MW055915 |
| 1621 | VC_520_62 | 1 | 0 | 10999.16688 | 0 | 0 | 0 | 0 | 1 | 1 | | 1 | 0 | 0 | 0 | 0 | ON970566 |
| 1622 | VC_520_63 | 1 | 0 | 11617.90847 | 0 | 0 | 0 | 0 | 1 | 1 | | 1 | 0 | 0 | 0 | 0 | MF919501 |
| 1623 | VC_520_64 | 1 | 0 | 11087.91545 | 0 | 0 | 0 | 0 | 1 | 1 | | 1 | 0 | 0 | 0 | 0 | MG962361 |
| 1624 | VC_520_65 | 1 | 0 | 11190.68355 | 0 | 0 | 0 | 0 | 1 | 1 | | 1 | 0 | 0 | 0 | 0 | MZ747519 |
| 1625 | VC_520_66 | 1 | 0 | 10669.52564 | 0 | 0 | 0 | 0 | 1 | 1 | | 1 | 0 | 0 | 0 | 0 | MN586012 |
| 1626 | VC_520_67 | 1 | 0 | 10218.28713 | 0 | 0 | 0 | 0 | 1 | 1 | | 1 | 0 | 0 | 0 | 0 | ON970610 |
| 1627 | VC_520_68 | 1 | 0 | 11179.24913 | 0 | 0 | 0 | 0 | 1 | 1 | | 1 | 0 | 0 | 0 | 0 | KM347889 |
| 1628 | VC_520_69 | 1 | 0 | 11375.71701 | 0 | 0 | 0 | 0 | 1 | 1 | | 1 | 0 | 0 | 0 | 0 | MH669014 |
| 1629 | VC_520_7 | 1 | 0 | 11016.66876 | 0 | 0 | 0 | 0 | 1 | 1 | | 1 | 0 | 0 | 0 | 0 | MN310548 |
| 1630 | VC_520_70 | 1 | 0 | 11020.72191 | 0 | 0 | 0 | 0 | 1 | 1 | | 1 | 0 | 0 | 0 | 0 | KC661280 |
| 1631 | VC_520_71 | 1 | 0 | 10917.0638 | 0 | 0 | 0 | 0 | 1 | 1 | | 1 | 0 | 0 | 0 | 0 | FJ174693 |
| 1632 | VC_520_72 | 1 | 0 | 11087.27983 | 0 | 0 | 0 | 0 | 1 | 1 | | 1 | 0 | 0 | 0 | 0 | MK494118 |
| 1633 | VC_520_73 | 2 | 251.1239561 | 23167.32839 | 0.010723337 | 0.042372566 | 6.708203932 | 6.708203932 | 1 | 1 | | 1 | 6.708203932 | 0 | 0 | 0 | KR935214,MH669012 |
| 1634 | VC_520_74 | 2 | 170.2183944 | 22535.57891 | 0.007496693 | 0.04239414 | 8.888194417 | 8.888194417 | 1 | 1 | | 1 | 8.888194417 | 0 | 0 | 0 | JF704117,MK112556 |
| 1635 | VC_520_75 | 1 | 0 | 11705.76837 | 0 | 0 | 0 | 0 | 1 | 1 | | 1 | 0 | 0 | 0 | 0 | MN310544 |
| 1636 | VC_520_76 | 2 | 194.3164302 | 21023.60947 | 0.009158126 | 0.042386242 | 7.745966692 | 7.745966692 | 1 | 1 | | 1 | 7.745966692 | 0 | 0 | 0 | JN398368,MT114164 |
| 1637 | VC_520_77 | 3 | 697.7262384 | 31307.16208 | 0.021800615 | 0.001400887 | 1.414213562 | 8.246211251 | 3 | 3 | | 1 | 5.928154406 | 0 | 0 | 0 | JQ300538,KX576644,MK016496 |
| 1638 | VC_520_78 | 2 | 186.3957452 | 23176.20389 | 0.007978382 | 0.042388467 | 8.062257748 | 8.062257748 | 1 | 1 | | 1 | 8.062257748 | 0 | 0 | 0 | MH590596,OP297528 |
| 1639 | VC_520_79 | 1 | 0 | 10603.49463 | 0 | 0 | 0 | 0 | 1 | 1 | | 1 | 0 | 0 | 0 | 0 | KC661278 |
| 1640 | VC_520_8 | 1 | 0 | 10591.73324 | 0 | 0 | 0 | 0 | 1 | 1 | | 1 | 0 | 0 | 0 | 0 | MZ958744 |
| 1641 | VC_520_80 | 3 | 900 | 29587.9148 | 0.029519894 | 0.001400963 | 0 | 2.645751311 | 3 | 3 | | 1 | 1.763834207 | 0 | 0 | 0 | MH651188,MH669003,MK359315 |
| 1642 | VC_520_81 | 1 | 0 | 11776.88464 | 0 | 0 | 0 | 0 | 1 | 1 | | 1 | 0 | 0 | 0 | 0 | MF919526 |
| 1643 | VC_520_82 | 1 | 0 | 9603.914151 | 0 | 0 | 0 | 0 | 1 | 1 | | 1 | 0 | 0 | 0 | 0 | MN310545 |
| 1644 | VC_520_83 | 1 | 0 | 9485.953804 | 0 | 0 | 0 | 0 | 1 | 1 | | 1 | 0 | 0 | 0 | 0 | KX610764 |
| 1645 | VC_520_84 | 1 | 0 | 11029.18346 | 0 | 0 | 0 | 0 | 1 | 1 | | 1 | 0 | 0 | 0 | 0 | ON970585 |
| 1646 | VC_520_85 | 2 | 300 | 19927.40518 | 0.014831364 | 0.042375953 | 0 | 0 | 1 | 1 | | 1 | 0 | 0 | 0 | 0 | KT895280,MN369751 |
| 1647 | VC_520_86 | 1 | 0 | 8826.798885 | 0 | 0 | 0 | 0 | 1 | 1 | | 1 | 0 | 0 | 0 | 0 | MZ747512 |
| 1648 | VC_520_87 | 1 | 0 | 10766.08325 | 0 | 0 | 0 | 0 | 1 | 1 | | 1 | 0 | 0 | 0 | 0 | MK524503 |
| 1649 | VC_520_88 | 1 | 0 | 9961.080023 | 0 | 0 | 0 | 0 | 1 | 1 | | 1 | 0 | 0 | 0 | 0 | JN020142 |
| 1650 | VC_520_89 | 1 | 0 | 10865.89646 | 0 | 0 | 0 | 0 | 1 | 1 | | 1 | 0 | 0 | 0 | 0 | MN585975 |
| 1651 | VC_520_9 | 1 | 0 | 9166.765787 | 0 | 0 | 0 | 0 | 1 | 1 | | 1 | 0 | 0 | 0 | 0 | KF114874 |
| 1652 | VC_520_90 | 1 | 0 | 10051.24931 | 0 | 0 | 0 | 0 | 1 | 1 | | 1 | 0 | 0 | 0 | 0 | MK016504 |
| 1653 | VC_520_91 | 1 | 0 | 11030.35404 | 0 | 0 | 0 | 0 | 1 | 1 | | 1 | 0 | 0 | 0 | 0 | MZ209299 |
| 1654 | VC_520_92 | 5 | 3000 | 42280.4134 | 0.066253812 | 2.55E-08 | 0 | 3.16227766 | 10 | 10 | | 1 | 1.516227766 | 0 | 0 | 0 | KM402757,MK359307,MN428051,MT889396,MT889397 |
| 1655 | VC_520_93 | 2 | 199.9702461 | 19105.66095 | 0.01035813 | 0.042379207 | 8.366600265 | 8.366600265 | 1 | 1 | | 1 | 8.366600265 | 0 | 0 | 0 | JF937098,MH632119 |
| 1656 | VC_520_94 | 1 | 0 | 9545.867104 | 0 | 0 | 0 | 0 | 1 | 1 | | 1 | 0 | 0 | 0 | 0 | MK967397 |
| 1657 | VC_520_95 | 1 | 0 | 10438.81724 | 0 | 0 | 0 | 0 | 1 | 1 | | 1 | 0 | 0 | 0 | 0 | MK224592 |
| 1658 | VC_520_96 | 1 | 0 | 10489.48078 | 0 | 0 | 0 | 0 | 1 | 1 | | 1 | 0 | 0 | 0 | 0 | AY129330 |
| 1659 | VC_520_97 | 1 | 0 | 9270.754592 | 0 | 0 | 0 | 0 | 1 | 1 | | 1 | 0 | 0 | 0 | 0 | MG925354 |
| 1660 | VC_520_98 | 1 | 0 | 7076.320832 | 0 | 0 | 0 | 0 | 1 | 1 | | 1 | 0 | 0 | 0 | 0 | MG812488 |
| 1661 | VC_521_0 | 37 | 101981.1658 | 9834.697816 | 0.912045594 | 3.31E-267 | 0 | 8.306623863 | 666 | 666 | | 1 | 5.617991731 | 0 | 0 | 0 | JN256079,JN624851,KC576783,KF986246,KM588359,KU935726,KU935727,KU935728,KU935729,KU935730,KU935731,KX756439,MG099936,MG099948,MH316570,MH399787,MH697576,MH697585,MH697593,MH926055,MK392367,MK524500,MK524509,MK524515,MK524518,MK524520,MK524522,MK524528,MK977708,MT498042,MT498044,MT498048,MT684588,MT723932,MZ622162,OL742561,OP297540 |
| 1662 | VC_522_0 | 3 | 395.6454258 | 4335.854974 | 0.083619443 | 0.001473402 | 5.744562647 | 7.416198487 | 3 | 3 | | 1 | 6.790621228 | 0 | 0 | 0 | KU963256,MN175604,ON645344 |
| 1663 | VC_522_1 | 5 | 1047.165441 | 9246.92981 | 0.101724864 | 3.27E-08 | 5.744562647 | 9.38083152 | 10 | 8 | | 0.8 | 7.787670622 | 0 | 0 | 0 | MN428053,MN585966,MT521995,OP297548,OP434457 |
| 1664 | VC_523_0 | 1 | 0 | 5035.835014 | 0 | 0 | 0 | 0 | 1 | 1 | | 1 | 0 | 0 | 0 | 0 | MK977709 |
| 1665 | VC_524_0 | 2 | 300 | 2354.064437 | 0.113034181 | 0.045224332 | 1.414213562 | 1.414213562 | 1 | 1 | | 1 | 1.414213562 | 0 | 0 | 0 | MW584176,MW584199 |
| 1666 | VC_524_1 | 1 | 0 | 1395.526051 | 0 | 0 | 0 | 0 | 1 | 1 | | 1 | 0 | 0 | 0 | 0 | MW584188 |
| 1667 | VC_525_0 | 25 | 55792.22408 | 65572.42871 | 0.45970736 | 3.28E-174 | 0 | 8.602325267 | 300 | 300 | | 1 | 5.879004775 | 0 | 0 | 0 | KU963255,KU963257,KX557273,KX557287,MH479912,MH479927,MK016492,MK112547,MK801727,MK814755,MK820642,MK875793,MK875796,MK878898,MK919483,MT723938,MT889398,MT952844,MW712717,MW924648,MW924650,MZ171374,MZ274304,OP297546,OP434440 |
| 1668 | VC_525_1 | 5 | 1187.724623 | 23290.69672 | 0.048521292 | 5.28E-07 | 7.549834435 | 9 | 10 | 9 | | 0.9 | 8.386121703 | 0 | 0 | 0 | KU963249,KU963263,MK919469,MK967384,MT639650 |
| 1669 | VC_526_0 | 6 | 4500 | 5759.382448 | 0.438622892 | 5.13E-10 | 1.732050808 | 7.549834435 | 15 | 15 | | 1 | 6.453073993 | 0 | 0 | 0 | MG557979,MG765277,MG765279,MH809529,MH809530,MH809531 |
| 1670 | VC_526_1 | 1 | 0 | 2341.779255 | 0 | 0 | 0 | 0 | 1 | 1 | | 1 | 0 | 0 | 0 | 0 | MF787246 |
| 1671 | VC_526_2 | 1 | 0 | 1778.168355 | 0 | 0 | 0 | 0 | 1 | 1 | | 1 | 0 | 0 | 0 | 0 | AY682195 |
| 1672 | VC_526_3 | 1 | 0 | 1359.863636 | 0 | 0 | 0 | 0 | 1 | 1 | | 1 | 0 | 0 | 0 | 0 | MK504444 |
| 1673 | VC_527_0 | 3 | 191.4536294 | 610.73212 | 0.23866496 | 0.001601173 | 3.872983346 | 5.099019514 | 3 | 3 | | 1 | 4.518192852 | 0 | 0 | 0 | MW202473,MW202478,MW202738 |
| 1674 | VC_529_0 | 2 | 191.413696 | 180.8907318 | 0.514132204 | 0.049743124 | 1.414213562 | 1.414213562 | 1 | 1 | | 1 | 1.414213562 | 0 | 0 | 0 | MW824391,MW824401 |
| 1675 | VC_52_0 | 87 | 234431.8252 | 3380.135927 | 0.985786518 | 7.49E-138 | 0 | 6.08276253 | 3741 | 3741 | | 1 | 1.061381729 | 0 | 0 | 0 | AB259123,AB434711,AB937974,AB981170,KX179905,MF716957,ON170020,ON170023,ON170029,ON170030,ON170034,ON170037,ON170041,ON170050,ON170055,ON170087,ON170089,ON170091,ON170093,ON170095,ON170097,ON170101,ON170103,ON170105,ON170107,ON170109,ON170111,ON170113,ON170115,ON170117,ON170119,ON170121,ON170125,ON170127,ON170132,ON170143,ON170145,ON170147,ON170153,ON170155,ON170157,ON170159,ON170161,ON170163,ON170165,ON170168,ON170193,ON170195,ON170197,ON170199,ON170201,ON170203,ON170205,ON170207,ON170212,ON170216,ON170218,ON170220,ON170223,ON170226,ON170245,ON170247,ON170249,ON170251,ON170253,ON170273,ON170277,ON170281,ON170283,ON170285,ON170290,ON170292,ON170294,ON170296,ON170299,ON170301,ON170308,ON170312,ON170314,ON170316,ON170331,ON170333,ON170335,ON170337,ON170339,ON170341,ON170353 |
| 1676 | VC_530_0 | 9 | 6075.930994 | 1493.315585 | 0.802712784 | 7.48E-13 | 2.645751311 | 7.071067812 | 36 | 36 | | 1 | 4.525014673 | 0 | 0 | 0 | AP019417,KJ936628,MF431726,MG592626,MK483107,MN062185,MW217515,MW451248,MW748992 |
| 1677 | VC_531_0 | 9 | 10800 | 30734.32405 | 0.2600259 | 2.12E-09 | 2.645751311 | 6.244997998 | 36 | 36 | | 1 | 4.539354477 | 0 | 0 | 0 | KC131129,KP671755,KT919972,KT919973,MK568540,MT612988,MT612989,ON000910,ON190025 |
| 1678 | VC_531_1 | 6 | 4500 | 25892.57823 | 0.148062463 | 0.000213354 | 2.828427125 | 8.831760866 | 15 | 15 | | 1 | 5.808049561 | 0 | 0 | 0 | AY283928,JN849462,MN794232,MT135026,OK905446,ON190024 |
| 1679 | VC_531_2 | 1 | 0 | 5809.695788 | 0 | 0 | 0 | 0 | 1 | 1 | | 1 | 0 | 0 | 0 | 0 | HQ317393 |
| 1680 | VC_531_3 | 1 | 0 | 5807.809061 | 0 | 0 | 0 | 0 | 1 | 1 | | 1 | 0 | 0 | 0 | 0 | MW824377 |
| 1681 | VC_531_4 | 1 | 0 | 5803.044018 | 0 | 0 | 0 | 0 | 1 | 1 | | 1 | 0 | 0 | 0 | 0 | MG592456 |
| 1682 | VC_531_5 | 1 | 0 | 8136.748311 | 0 | 0 | 0 | 0 | 1 | 1 | | 1 | 0 | 0 | 0 | 0 | MT135025 |
| 1683 | VC_531_6 | 1 | 0 | 5779.187796 | 0 | 0 | 0 | 0 | 1 | 1 | | 1 | 0 | 0 | 0 | 0 | MT135024 |
| 1684 | VC_532_0 | 2 | 300 | 441.4787999 | 0.404596868 | 0.105649774 | 1.414213562 | 1.414213562 | 1 | 1 | | 1 | 1.414213562 | 0 | 0 | 0 | AP019524,AP019525 |
| 1685 | VC_532_1 | 1 | 0 | 368.5904716 | 0 | 0 | 0 | 0 | 1 | 1 | | 1 | 0 | 0 | 0 | 0 | LC168164 |
| 1686 | VC_533_0 | 2 | 300 | 806.0109454 | 0.271245055 | 0.069143071 | 0 | 0 | 1 | 1 | | 1 | 0 | 0 | 0 | 0 | LR756501,LR756503 |
| 1687 | VC_533_1 | 1 | 0 | 877.6125162 | 0 | 0 | 0 | 0 | 1 | 1 | | 1 | 0 | 0 | 0 | 0 | LR745206 |
| 1688 | VC_533_2 | 1 | 0 | 885.9910368 | 0 | 0 | 0 | 0 | 1 | 1 | | 1 | 0 | 0 | 0 | 0 | LR756502 |
| 1689 | VC_533_3 | 1 | 0 | 700.3472986 | 0 | 0 | 0 | 0 | 1 | 1 | | 1 | 0 | 0 | 0 | 0 | LR756504 |
| 1690 | VC_534_0 | 9 | 10146.17645 | 102.1409323 | 0.990033395 | 1.19E-09 | 0 | 4.242640687 | 36 | 36 | | 1 | 2.756800211 | 0 | 0 | 0 | AP019527,FJ429185,KY554762,KY554763,KY554764,KY554765,KY554766,KY554767,MN528768 |
| 1691 | VC_535_0 | 2 | 15.70169036 | 102.1409323 | 0.133242879 | 0.05936654 | 5.744562647 | 5.744562647 | 1 | 1 | | 1 | 5.744562647 | 0 | 0 | 0 | S4-wms_118690,S9-wms_19025 |
| 1692 | VC_536_0 | 3 | 539.7638945 | 5604.981907 | 0.087841534 | 0.001464102 | 5.656854249 | 7.141428429 | 3 | 3 | | 1 | 6.599427559 | 0 | 0 | 0 | EF153632,MT708549,MW460246 |
| 1693 | VC_539_0 | 3 | 168.3768118 | 0 | 1 | 0 | 3 | 3.741657387 | 3 | 3 | | 1 | 3.352760726 | 0 | 0 | 0 | AY048850,BK013337,BK013338 |
| 1694 | VC_53_0 | 5 | 340.1298475 | 343.2957481 | 0.4976838 | 1.92E-06 | 0 | 4.123105626 | 10 | 10 | | 1 | 2.47275816 | 0 | 0 | 0 | HM064452,ON170025,ON170069,ON170139,ON170320 |
| 1695 | VC_540_0 | 15 | 12482.00302 | 31065.41727 | 0.286630136 | 4.22E-68 | 1 | 8.366600265 | 105 | 105 | | 1 | 6.644672412 | 0 | 0 | 0 | KC139518,KT287080,MF158036,MF158037,MH424444,MK214385,MK479295,MN070121,MN393079,MT012729,MW725301,MZ004649,MZ501269,ON720976,S17-wms_266511 |
| 1696 | VC_542_0 | 2 | 130.7598395 | 217.6741788 | 0.375278626 | 0.059077917 | 5.477225575 | 5.477225575 | 1 | 1 | | 1 | 5.477225575 | 0 | 0 | 0 | MW478292,OK247671 |
| 1697 | VC_543_0 | 6 | 3484.410086 | 548.6388398 | 0.863964249 | 5.19E-07 | 0 | 3.16227766 | 15 | 15 | | 1 | 1.913044717 | 0 | 0 | 0 | MG592399,MG592418,MG592419,MG592510,MG592511,MG592579 |
| 1698 | VC_544_0 | 2 | 244.0422632 | 449.8887193 | 0.351680887 | 0.059409431 | 0 | 0 | 1 | 1 | | 1 | 0 | 0 | 0 | 0 | MG592447,MG592453 |
| 1699 | VC_546_0 | 2 | 300 | 21764.79714 | 0.013596318 | 0.362514023 | 8.660254038 | 8.660254038 | 1 | 1 | | 1 | 8.660254038 | 0 | 0 | 0 | JN201525,MH399772 |
| 1700 | VC_546_1 | 2 | 300 | 21828.05743 | 0.013557449 | 0.31594849 | 7.937253933 | 7.937253933 | 1 | 1 | | 1 | 7.937253933 | 0 | 0 | 0 | MN586029,MW314846 |
| 1701 | VC_546_10 | 1 | 0 | 11461.18437 | 0 | 0 | 0 | 0 | 1 | 1 | | 1 | 0 | 0 | 0 | 0 | AY129338 |
| 1702 | VC_546_11 | 1 | 0 | 11323.32455 | 0 | 0 | 0 | 0 | 1 | 1 | | 1 | 0 | 0 | 0 | 0 | JF937101 |
| 1703 | VC_546_2 | 3 | 900 | 31842.1138 | 0.027487535 | 0.147396579 | 4.69041576 | 5.656854249 | 3 | 3 | | 1 | 5.148763174 | 0 | 0 | 0 | JN698997,KM400683,MF919534 |
| 1704 | VC_546_3 | 4 | 1800 | 41277.11567 | 0.041785527 | 0.078050118 | 4.358898944 | 8.485281374 | 6 | 6 | | 1 | 6.592672932 | 0 | 0 | 0 | KM925136,MF072690,MF133445,MF668284 |
| 1705 | VC_546_4 | 16 | 36000 | 107019.4769 | 0.251713968 | 1.97E-07 | 4.472135955 | 9.591663047 | 120 | 113 | | 0.941666667 | 7.461739702 | 0 | 0 | 0 | KF114875,MH077579,MH669017,MH697582,MH697583,MH727551,MK524504,MK524521,MK524527,MK524529,MK967379,MK967402,MN062701,MN183286,MZ622169,MZ958748 |
| 1706 | VC_546_5 | 1 | 0 | 11219.89964 | 0 | 0 | 0 | 0 | 1 | 1 | | 1 | 0 | 0 | 0 | 0 | MF919504 |
| 1707 | VC_546_6 | 2 | 300 | 21959.02769 | 0.013477678 | 0.218907096 | 6.32455532 | 6.32455532 | 1 | 1 | | 1 | 6.32455532 | 0 | 0 | 0 | KF006818,MF919512 |
| 1708 | VC_546_7 | 4 | 1800 | 42115.81183 | 0.040987515 | 0.003674896 | 6.403124237 | 8.94427191 | 6 | 6 | | 1 | 7.722585294 | 0 | 0 | 0 | JF937090,JF957059,KM101123,MK279849 |
| 1709 | VC_546_8 | 1 | 0 | 11189.44157 | 0 | 0 | 0 | 0 | 1 | 1 | | 1 | 0 | 0 | 0 | 0 | MW132714 |
| 1710 | VC_546_9 | 1 | 0 | 11190.99746 | 0 | 0 | 0 | 0 | 1 | 1 | | 1 | 0 | 0 | 0 | 0 | MK524516 |
| 1711 | VC_54_0 | 6 | 489.3313593 | 238.3040791 | 0.672495227 | 1.01E-07 | 0 | 2.236067977 | 15 | 15 | | 1 | 1.454741697 | 0 | 0 | 0 | AB259124,AB828698,AB931172,JQ408219,LC066596,MT495635 |
| 1712 | VC_550_0 | 12 | 18665.12486 | 4608.706307 | 0.801979044 | 2.27E-35 | 0 | 8.306623863 | 66 | 66 | | 1 | 5.696831437 | 0 | 0 | 0 | MW314855,MW314856,MW314858,MW584152,MW584158,MW584161,MW584164,MW584171,MW584179,MW584180,MW584196,MW584204 |
| 1713 | VC_551_0 | 16 | 34817.81906 | 345562.9196 | 0.091534128 | 1.09E-58 | 0 | 10.34408043 | 120 | 80 | | 0.666666667 | 8.305420408 | 0 | 0 | 0 | JF937091,JF937106,MG872831,MG872832,MG872843,MH399774,MH399778,MH536820,MK016502,MK359320,MK801726,MK937593,MN096361,MN586013,MT723943,MW291023 |
| 1714 | VC_551_1 | 4 | 1733.230065 | 107507.4298 | 0.015866163 | 0.000316626 | 6.782329983 | 9.327379053 | 6 | 4 | | 0.666666667 | 8.397948149 | 0 | 0 | 0 | MF919541,MH576956,MN586043,MN586044 |
| 1715 | VC_551_2 | 17 | 40653.29819 | 377299.1555 | 0.097267758 | 2.10E-76 | 0 | 9.273618495 | 136 | 133 | | 0.977941176 | 7.37256991 | 0 | 0 | 0 | EU816591,JF937096,JN006061,JN006062,KF188414,KR080204,KT222941,KT222942,KU865303,KX817173,KY319168,MH513972,MH513978,MN586032,MN586051,MT114165,MZ274305 |
| 1716 | VC_551_3 | 1 | 0 | 27822.30359 | 0 | 0 | 0 | 0 | 1 | 1 | | 1 | 0 | 0 | 0 | 0 | KF306380 |
| 1717 | VC_551_4 | 2 | 300 | 49344.74346 | 0.006042936 | 0.043041426 | 3.741657387 | 3.741657387 | 1 | 1 | | 1 | 3.741657387 | 0 | 0 | 0 | JN412590,KF416343 |
| 1718 | VC_551_5 | 2 | 278.1092754 | 50903.34479 | 0.00543379 | 0.044519377 | 8.888194417 | 8.888194417 | 1 | 1 | | 1 | 8.888194417 | 0 | 0 | 0 | MK016491,MK016498 |
| 1719 | VC_551_6 | 22 | 67760.06557 | 490503.9482 | 0.121376381 | 4.40E-93 | 0 | 10.29563014 | 231 | 187 | | 0.80952381 | 7.78923934 | 0 | 0 | 0 | EU816588,KX611831,KX834009,MG757160,MG872837,MH371112,MH513983,MH536829,MH651174,MH669002,MH727557,MH779503,MK559429,MN428059,MN586034,MN586037,MT952854,MZ622157,MZ681505,OL742559,ON970597,ON970600 |
| 1720 | VC_551_7 | 28 | 111344.4161 | 559083.8721 | 0.166079532 | 7.94E-159 | 0 | 10.34408043 | 378 | 322 | | 0.851851852 | 7.846339488 | 0 | 0 | 0 | AY129331,DQ398041,GQ303265,JN382248,JN391441,KC661277,KC691255,KC748971,KF279417,KT020852,KT373978,KY549152,MF668277,MH000607,MH536827,MH576953,MK359309,MK620893,MK757445,MN586017,MN586019,MN586035,MN586041,MN586050,MZ322024,MZ622158,MZ622163,ON456345 |
| 1721 | VC_551_8 | 19 | 50413.44652 | 434972.1398 | 0.103862677 | 5.04E-79 | 0 | 9.899494937 | 171 | 149 | | 0.871345029 | 7.47701522 | 0 | 0 | 0 | KC748969,KF024732,KF493883,KF562099,KF562100,KF562101,KF562102,MF919506,MF919524,MF919525,MF919529,MF919535,MF919540,MG099953,MH020247,MH590587,MK433262,MN096364,MT522005 |
| 1722 | VC_554_0 | 2 | 77.87004059 | 261.9314456 | 0.229163331 | 0.04806215 | 6.557438524 | 6.557438524 | 1 | 1 | | 1 | 6.557438524 | 0 | 0 | 0 | JN698994,MPU46938 |
| 1723 | VC_555_0 | 13 | 13808.1958 | 4563.869163 | 0.751586489 | 1.09E-45 | 0 | 7.211102551 | 78 | 78 | | 1 | 5.125806796 | 0 | 0 | 0 | MW314852,MW584148,MW584150,MW584154,MW584156,MW584162,MW584163,MW584181,MW584182,MW584192,MW584203,MW584208,MW584212 |
| 1724 | VC_556_0 | 21 | 63000 | 561.4156776 | 0.991167351 | 4.11E-67 | 0 | 5.099019514 | 210 | 210 | | 1 | 3.642219586 | 0 | 0 | 0 | AY129335,JN698993,KF017002,KF024730,KJ829260,KT281796,MG099943,MG872838,MH020246,MH590592,MH926062,MN428052,MN428058,MN585964,MT818418,MT818425,MW314845,MW712737,MZ209306,MZ747511,ON260829 |
| 1725 | VC_557_0 | 3 | 459.9962677 | 5194.234025 | 0.081354357 | 0.00146721 | 8.717797887 | 9.16515139 | 3 | 2 | | 0.666666667 | 8.904903381 | 0 | 0 | 0 | MK937597,MN096370,OP434458 |
| 1726 | VC_557_1 | 11 | 9997.72515 | 10220.21677 | 0.494497669 | 8.09E-35 | 2 | 9.539392014 | 55 | 43 | | 0.781818182 | 8.297248607 | 0 | 0 | 0 | MG099939,MH651176,MH651185,MH779504,MK279906,MK376961,MK814752,MZ388553,MZ747510,ON456334,ON526981 |
| 1727 | VC_557_2 | 1 | 0 | 2254.986348 | 0 | 0 | 0 | 0 | 1 | 1 | | 1 | 0 | 0 | 0 | 0 | MH536812 |
| 1728 | VC_557_3 | 1 | 0 | 2398.146174 | 0 | 0 | 0 | 0 | 1 | 1 | | 1 | 0 | 0 | 0 | 0 | MW534375 |
| 1729 | VC_558_0 | 2 | 300 | 92368.60475 | 0.003237342 | 0.241415006 | 8.426149773 | 8.426149773 | 1 | 1 | | 1 | 8.426149773 | 0 | 0 | 0 | JN204348,JN412592 |
| 1730 | VC_558_1 | 2 | 300 | 92830.90516 | 0.003221272 | 0.215949683 | 9 | 9 | 1 | 0 | | 0 | 9 | 0 | 0 | 0 | GQ303260,MT553335 |
| 1731 | VC_558_10 | 1 | 0 | 47359.44854 | 0 | 0 | 0 | 0 | 1 | 1 | | 1 | 0 | 0 | 0 | 0 | MK112534 |
| 1732 | VC_558_11 | 2 | 300 | 92981.86351 | 0.003216059 | 0.214714127 | 4.358898944 | 4.358898944 | 1 | 1 | | 1 | 4.358898944 | 0 | 0 | 0 | KT281790,MT889395 |
| 1733 | VC_558_12 | 3 | 900 | 138757.9099 | 0.006444318 | 0.121457474 | 7.348469228 | 7.681145748 | 3 | 3 | | 1 | 7.481937821 | 0 | 0 | 0 | KC748970,MH825697,MK359348 |
| 1734 | VC_558_13 | 1 | 0 | 47215.63528 | 0 | 0 | 0 | 0 | 1 | 1 | | 1 | 0 | 0 | 0 | 0 | MK359321 |
| 1735 | VC_558_14 | 8 | 8400 | 360021.3364 | 0.022799982 | 0.000547538 | 2.449489743 | 9.797958971 | 28 | 23 | | 0.821428571 | 7.955531127 | 0 | 0 | 0 | MG872839,MH316561,MK359303,MK359311,MK359358,MK450427,ON526982,ON637768 |
| 1736 | VC_558_15 | 1 | 0 | 47307.97164 | 0 | 0 | 0 | 0 | 1 | 1 | | 1 | 0 | 0 | 0 | 0 | MK359342 |
| 1737 | VC_558_16 | 1 | 0 | 47349.47565 | 0 | 0 | 0 | 0 | 1 | 1 | | 1 | 0 | 0 | 0 | 0 | ON755189 |
| 1738 | VC_558_17 | 1 | 0 | 47419.5358 | 0 | 0 | 0 | 0 | 1 | 1 | | 1 | 0 | 0 | 0 | 0 | MK359328 |
| 1739 | VC_558_18 | 1 | 0 | 47432.14325 | 0 | 0 | 0 | 0 | 1 | 1 | | 1 | 0 | 0 | 0 | 0 | MN183285 |
| 1740 | VC_558_19 | 1 | 0 | 47438.38156 | 0 | 0 | 0 | 0 | 1 | 1 | | 1 | 0 | 0 | 0 | 0 | ON637764 |
| 1741 | VC_558_2 | 2 | 300 | 94325.92395 | 0.003170379 | 0.331400206 | 8.366600265 | 8.366600265 | 1 | 1 | | 1 | 8.366600265 | 0 | 0 | 0 | EU826471,MH727543 |
| 1742 | VC_558_20 | 17 | 40800 | 714683.7422 | 0.054005133 | 1.59E-17 | 0 | 9.219544457 | 136 | 135 | | 0.992647059 | 5.130990877 | 0 | 0 | 0 | JF704096,KR080196,MF919493,MH697586,MH697590,MK359326,MK359335,MK359338,MK359339,MK359347,MK359362,MK450432,MN428066,MZ028626,MZ681511,ON081335,ON260824 |
| 1743 | VC_558_21 | 1 | 0 | 46878.2198 | 0 | 0 | 0 | 0 | 1 | 1 | | 1 | 0 | 0 | 0 | 0 | MT723945 |
| 1744 | VC_558_22 | 4 | 1800 | 178073.2196 | 0.010007048 | 0.00687181 | 7.141428429 | 9.38083152 | 6 | 5 | | 0.833333333 | 8.031610371 | 0 | 0 | 0 | JF704092,MH590593,MK450423,MZ958745 |
| 1745 | VC_558_23 | 1 | 0 | 46933.08472 | 0 | 0 | 0 | 0 | 1 | 1 | | 1 | 0 | 0 | 0 | 0 | KX369583 |
| 1746 | VC_558_24 | 1 | 0 | 47127.02634 | 0 | 0 | 0 | 0 | 1 | 1 | | 1 | 0 | 0 | 0 | 0 | OP021681 |
| 1747 | VC_558_25 | 1 | 0 | 47633.32593 | 0 | 0 | 0 | 0 | 1 | 1 | | 1 | 0 | 0 | 0 | 0 | EU826468 |
| 1748 | VC_558_26 | 1 | 0 | 47564.42932 | 0 | 0 | 0 | 0 | 1 | 1 | | 1 | 0 | 0 | 0 | 0 | EU826469 |
| 1749 | VC_558_27 | 2 | 300 | 94507.21276 | 0.003164316 | 0.358645393 | 6.708203932 | 6.708203932 | 1 | 1 | | 1 | 6.708203932 | 0 | 0 | 0 | MK359361,MW712729 |
| 1750 | VC_558_28 | 1 | 0 | 47332.358 | 0 | 0 | 0 | 0 | 1 | 1 | | 1 | 0 | 0 | 0 | 0 | MK359333 |
| 1751 | VC_558_29 | 1 | 0 | 47620.2144 | 0 | 0 | 0 | 0 | 1 | 1 | | 1 | 0 | 0 | 0 | 0 | MT818416 |

| | | | | | | | | | | | | | | | | |
|---|---|---|---|---|---|---|---|---|---|---|---|---|---|---|---|---|
| 1752 | VC_558_3 | 1 | 0 | 47121.29767 | 0 | 0 | 0 | 0 | 1 | 1 | 1 | 0 | 0 | 0 | 0 | MK359355 |
| 1753 | VC_558_30 | 3 | 900 | 140553.3225 | 0.006362523 | 0.234259467 | 6.782329983 | 8.94427191 | 3 | 3 | 1 | 7.950213433 | 0 | 0 | 0 | EU826467,JN699625,MK359344 |
| 1754 | VC_558_31 | 1 | 0 | 47664.16351 | 0 | 0 | 0 | 0 | 1 | 1 | 1 | 0 | 0 | 0 | 0 | DQ398053 |
| 1755 | VC_558_32 | 1 | 0 | 47617.04415 | 0 | 0 | 0 | 0 | 1 | 1 | 1 | 0 | 0 | 0 | 0 | MK310143 |
| 1756 | VC_558_33 | 1 | 0 | 47421.54567 | 0 | 0 | 0 | 0 | 1 | 1 | 1 | 0 | 0 | 0 | 0 | MH910040 |
| 1757 | VC_558_34 | 1 | 0 | 47642.99373 | 0 | 0 | 0 | 0 | 1 | 1 | 1 | 0 | 0 | 0 | 0 | MK112549 |
| 1758 | VC_558_35 | 1 | 0 | 47493.15754 | 0 | 0 | 0 | 0 | 1 | 1 | 1 | 0 | 0 | 0 | 0 | MT818423 |
| 1759 | VC_558_36 | 4 | 1800 | 186052.3017 | 0.009581996 | 0.14086148 | 0 | 8.602325267 | 6 | 6 | 1 | 6.722553718 | 0 | 0 | 0 | MK359336,MK359363,MK359366,MN234193 |
| 1760 | VC_558_37 | 1 | 0 | 47370.33283 | 0 | 0 | 0 | 0 | 1 | 1 | 1 | 0 | 0 | 0 | 0 | KT365400 |
| 1761 | VC_558_38 | 2 | 300 | 94425.0999 | 0.003167059 | 0.356713713 | 7.615773106 | 7.615773106 | 1 | 1 | 1 | 7.615773106 | 0 | 0 | 0 | MK878897,MN428065 |
| 1762 | VC_558_39 | 3 | 900 | 140956.2253 | 0.006344452 | 0.261504262 | 7.810249676 | 9.327379053 | 3 | 2 | 0.666666667 | 8.420555711 | 0 | 0 | 0 | MK359364,MT723944,OP297534 |
| 1763 | VC_558_4 | 1 | 0 | 47493.56223 | 0 | 0 | 0 | 0 | 1 | 1 | 1 | 0 | 0 | 0 | 0 | MK359353 |
| 1764 | VC_558_40 | 1 | 0 | 47529.22438 | 0 | 0 | 0 | 0 | 1 | 1 | 1 | 0 | 0 | 0 | 0 | MN276192 |
| 1765 | VC_558_41 | 1 | 0 | 47691.65039 | 0 | 0 | 0 | 0 | 1 | 1 | 1 | 0 | 0 | 0 | 0 | MK450424 |
| 1766 | VC_558_42 | 1 | 0 | 47479.50419 | 0 | 0 | 0 | 0 | 1 | 1 | 1 | 0 | 0 | 0 | 0 | MZ856343 |
| 1767 | VC_558_43 | 1 | 0 | 47461.53961 | 0 | 0 | 0 | 0 | 1 | 1 | 1 | 0 | 0 | 0 | 0 | MK359337 |
| 1768 | VC_558_44 | 1 | 0 | 46847.27278 | 0 | 0 | 0 | 0 | 1 | 1 | 1 | 0 | 0 | 0 | 0 | MK359345 |
| 1769 | VC_558_45 | 1 | 0 | 47381.35474 | 0 | 0 | 0 | 0 | 1 | 1 | 1 | 0 | 0 | 0 | 0 | MK359327 |
| 1770 | VC_558_46 | 1 | 0 | 46960.96139 | 0 | 0 | 0 | 0 | 1 | 1 | 1 | 0 | 0 | 0 | 0 | MK359349 |
| 1771 | VC_558_47 | 3 | 900 | 140417.0076 | 0.00636866 | 0.199560568 | 6.92820323 | 8.717797887 | 3 | 3 | 1 | 7.709771964 | 0 | 0 | 0 | MK112541,MK112545,MK112548 |
| 1772 | VC_558_48 | 2 | 300 | 93781.4661 | 0.003188726 | 0.305338744 | 7.810249676 | 7.810249676 | 1 | 1 | 1 | 7.810249676 | 0 | 0 | 0 | KF024734,KX721256 |
| 1773 | VC_558_49 | 7 | 6300 | 318448.6069 | 0.019399621 | 0.004896307 | 6.32455532 | 9.949874371 | 21 | 18 | 0.857142857 | 8.265139531 | 0 | 0 | 0 | JN624850,KC748968,KX721255,MH779498,MK112554,MK359334,OP068341 |
| 1774 | VC_558_5 | 1 | 0 | 47488.13148 | 0 | 0 | 0 | 0 | 1 | 1 | 1 | 0 | 0 | 0 | 0 | KC691257 |
| 1775 | VC_558_50 | 15 | 31500 | 641172.7558 | 0.046828119 | 1.65E-13 | 4.123105626 | 9.949874371 | 105 | 97 | 0.923809524 | 7.416660423 | 0 | 0 | 0 | JQ911768,KJ595575,KM881426,KT321476,MF919499,MH051266,MH316567,MH669008,MH825713,MK359314,MK359365,MK450429,MK450434,OK040789,OP021684 |
| 1776 | VC_558_51 | 1 | 0 | 47204.54291 | 0 | 0 | 0 | 0 | 1 | 1 | 1 | 0 | 0 | 0 | 0 | MK112533 |
| 1777 | VC_558_52 | 1 | 0 | 47219.97891 | 0 | 0 | 0 | 0 | 1 | 1 | 1 | 0 | 0 | 0 | 0 | MH744422 |
| 1778 | VC_558_53 | 1 | 0 | 47570.91209 | 0 | 0 | 0 | 0 | 1 | 1 | 1 | 0 | 0 | 0 | 0 | MK359306 |
| 1779 | VC_558_54 | 1 | 0 | 47180.98025 | 0 | 0 | 0 | 0 | 1 | 1 | 1 | 0 | 0 | 0 | 0 | MH779502 |
| 1780 | VC_558_55 | 1 | 0 | 47112.5107 | 0 | 0 | 0 | 0 | 1 | 1 | 1 | 0 | 0 | 0 | 0 | MF919494 |
| 1781 | VC_558_56 | 1 | 0 | 47180.19033 | 0 | 0 | 0 | 0 | 1 | 1 | 1 | 0 | 0 | 0 | 0 | MH727558 |
| 1782 | VC_558_57 | 1 | 0 | 47038.93777 | 0 | 0 | 0 | 0 | 1 | 1 | 1 | 0 | 0 | 0 | 0 | MK450425 |
| 1783 | VC_558_58 | 1 | 0 | 47669.10236 | 0 | 0 | 0 | 0 | 1 | 1 | 1 | 0 | 0 | 0 | 0 | MZ388551 |
| 1784 | VC_558_59 | 1 | 0 | 47474.74816 | 0 | 0 | 0 | 0 | 1 | 1 | 1 | 0 | 0 | 0 | 0 | MK359317 |
| 1785 | VC_558_6 | 2 | 300 | 93293.9363 | 0.003205336 | 0.268507211 | 8.888194417 | 8.888194417 | 1 | 1 | 1 | 8.888194417 | 0 | 0 | 0 | JF704116,KT365398 |
| 1786 | VC_558_60 | 1 | 0 | 47606.57188 | 0 | 0 | 0 | 0 | 1 | 1 | 1 | 0 | 0 | 0 | 0 | MN183280 |
| 1787 | VC_558_61 | 2 | 300 | 94444.89654 | 0.003166397 | 0.338345214 | 8.888194417 | 8.888194417 | 1 | 1 | 1 | 8.888194417 | 0 | 0 | 0 | MH051260,MK359318 |
| 1788 | VC_558_62 | 1 | 0 | 47432.44668 | 0 | 0 | 0 | 0 | 1 | 1 | 1 | 0 | 0 | 0 | 0 | MK359305 |
| 1789 | VC_558_63 | 1 | 0 | 47645.36359 | 0 | 0 | 0 | 0 | 1 | 1 | 1 | 0 | 0 | 0 | 0 | JN699626 |
| 1790 | VC_558_64 | 1 | 0 | 47371.4805 | 0 | 0 | 0 | 0 | 1 | 1 | 1 | 0 | 0 | 0 | 0 | MK359350 |
| 1791 | VC_558_65 | 2 | 300 | 94787.19385 | 0.003154999 | 0.388258183 | 7.874007874 | 7.874007874 | 1 | 1 | 1 | 7.874007874 | 0 | 0 | 0 | MK450422,MW291012 |
| 1792 | VC_558_66 | 1 | 0 | 47654.53093 | 0 | 0 | 0 | 0 | 1 | 1 | 1 | 0 | 0 | 0 | 0 | MK359330 |
| 1793 | VC_558_67 | 1 | 0 | 47650.65395 | 0 | 0 | 0 | 0 | 1 | 1 | 1 | 0 | 0 | 0 | 0 | OP021682 |
| 1794 | VC_558_68 | 1 | 0 | 47579.9955 | 0 | 0 | 0 | 0 | 1 | 1 | 1 | 0 | 0 | 0 | 0 | MG812494 |
| 1795 | VC_558_69 | 1 | 0 | 47454.45477 | 0 | 0 | 0 | 0 | 1 | 1 | 1 | 0 | 0 | 0 | 0 | MK359329 |
| 1796 | VC_558_7 | 1 | 0 | 47343.74263 | 0 | 0 | 0 | 0 | 1 | 1 | 1 | 0 | 0 | 0 | 0 | MH051261 |
| 1797 | VC_558_70 | 1 | 0 | 47616.04637 | 0 | 0 | 0 | 0 | 1 | 1 | 1 | 0 | 0 | 0 | 0 | ON456344 |
| 1798 | VC_558_71 | 1 | 0 | 46911.88748 | 0 | 0 | 0 | 0 | 1 | 1 | 1 | 0 | 0 | 0 | 0 | MK359346 |
| 1799 | VC_558_72 | 1 | 0 | 47512.46907 | 0 | 0 | 0 | 0 | 1 | 1 | 1 | 0 | 0 | 0 | 0 | MG812493 |
| 1800 | VC_558_73 | 2 | 300 | 91083.29862 | 0.003282876 | 0.156251606 | 6.708203932 | 6.708203932 | 1 | 1 | 1 | 6.708203932 | 0 | 0 | 0 | MH697579,MK359359 |
| 1801 | VC_558_74 | 1 | 0 | 46728.57024 | 0 | 0 | 0 | 0 | 1 | 1 | 1 | 0 | 0 | 0 | 0 | MF919495 |
| 1802 | VC_558_75 | 1 | 0 | 47470.66446 | 0 | 0 | 0 | 0 | 1 | 1 | 1 | 0 | 0 | 0 | 0 | ON551388 |
| 1803 | VC_558_76 | 1 | 0 | 46972.36199 | 0 | 0 | 0 | 0 | 1 | 1 | 1 | 0 | 0 | 0 | 0 | MW822148 |
| 1804 | VC_558_77 | 3 | 900 | 137906.3086 | 0.006483855 | 0.091220611 | 7.745966692 | 9.486832981 | 3 | 2 | 0.666666667 | 8.368935849 | 0 | 0 | 0 | MF919513,MK359308,MN062707 |
| 1805 | VC_558_78 | 1 | 0 | 47242.75258 | 0 | 0 | 0 | 0 | 1 | 1 | 1 | 0 | 0 | 0 | 0 | JN699627 |
| 1806 | VC_558_79 | 1 | 0 | 47305.68387 | 0 | 0 | 0 | 0 | 1 | 1 | 1 | 0 | 0 | 0 | 0 | MN735434 |
| 1807 | VC_558_8 | 2 | 300 | 94033.49784 | 0.003180206 | 0.319694882 | 5.744562647 | 5.744562647 | 1 | 1 | 1 | 5.744562647 | 0 | 0 | 0 | AY129337,JN699013 |
| 1808 | VC_558_80 | 1 | 0 | 47529.16606 | 0 | 0 | 0 | 0 | 1 | 1 | 1 | 0 | 0 | 0 | 0 | JN412588 |
| 1809 | VC_558_81 | 1 | 0 | 46225.18959 | 0 | 0 | 0 | 0 | 1 | 1 | 1 | 0 | 0 | 0 | 0 | KX752698 |
| 1810 | VC_558_82 | 3 | 900 | 138687.6647 | 0.006447561 | 0.115661263 | 8 | 9.433981132 | 3 | 2 | 0.666666667 | 8.580201665 | 0 | 0 | 0 | GQ303262,KX831080,MF919527 |
| 1811 | VC_558_9 | 3 | 900 | 138960.6583 | 0.006434976 | 0.170884714 | 7.211102551 | 9.643650761 | 3 | 2 | 0.666666667 | 8.264002415 | 0 | 0 | 0 | KF006817,KX781992,MK359324 |
| 1812 | VC_559_0 | 2 | 300 | 26550.78442 | 0.011172858 | 0.043487187 | 7.549834435 | 7.549834435 | 1 | 1 | 1 | 7.549834435 | 0 | 0 | 0 | EU826466,MF668280 |
| 1813 | VC_559_1 | 1 | 0 | 14785.88997 | 0 | 0 | 0 | 0 | 1 | 1 | 1 | 0 | 0 | 0 | 0 | MZ820085 |
| 1814 | VC_55_0 | 3 | 412.2044165 | 1050.709626 | 0.281769403 | 0.001498887 | 1.732050808 | 2.828427125 | 3 | 3 | 1 | 2.265515303 | 0 | 0 | 0 | CP006891,CP023680,CP023686 |
| 1815 | VC_563_0 | 2 | 300 | 3827.425423 | 0.072684536 | 0.043574004 | 4.358898944 | 4.358898944 | 1 | 1 | 1 | 4.358898944 | 0 | 0 | 0 | KJ567043,MK016501 |
| 1816 | VC_564_0 | 3 | 799.5228981 | 1537.841199 | 0.342061769 | 0.001500873 | 1 | 2.645751311 | 3 | 3 | 1 | 2.031747018 | 0 | 0 | 0 | MW584155,MW584175,MW584201 |
| 1817 | VC_565_0 | 20 | 50899.66127 | 9706.93229 | 0.839837026 | 1.27E-107 | 0 | 5.916079783 | 190 | 190 | 1 | 2.340816186 | 0 | 0 | 0 | DQ398047,DQ398051,EU676000,FJ168660,FJ168661,FJ168662,JF937107,JN699014,KJ194582,MH025888,MH316562,MH399788,MH834615,MK494102,MK524498,MK559427,MN119378,MN234227,MN284908,MT889385 |
| 1818 | VC_566_0 | 2 | 278.0137445 | 2534.146485 | 0.098861275 | 0.043774736 | 3.16227766 | 3.16227766 | 1 | 1 | 1 | 3.16227766 | 0 | 0 | 0 | AY129339,MK279909 |
| 1819 | VC_567_0 | 8 | 7341.142214 | 7534.251051 | 0.493509118 | 2.33E-19 | 2 | 6.480740698 | 28 | 28 | 1 | 3.692026831 | 0 | 0 | 0 | EU770222,FJ174691,KF986247,KJ567046,MG962376,MH509445,MN010756,MT521996 |
| 1820 | VC_568_0 | 8 | 8286.314136 | 6156.062465 | 0.573750039 | 2.33E-19 | 1 | 3.741657387 | 28 | 28 | 1 | 2.697423168 | 0 | 0 | 0 | JF704112,KF416342,KT591076,KY969628,MH001447,MH926059,MK757446,MT684602 |
| 1821 | VC_569_0 | 3 | 900 | 3397.037133 | 0.209446642 | 0.001501332 | 1.732050808 | 2.449489743 | 3 | 3 | 1 | 2.139202843 | 0 | 0 | 0 | JN412589,KR997933,ON108649 |
| 1822 | VC_570_0 | 9 | 8792.373367 | 8827.992161 | 0.498989272 | 3.61E-24 | 0 | 6.633249581 | 36 | 36 | 1 | 3.7813655 | 0 | 0 | 0 | KX557285,MH025890,MH271313,MH576972,MT639639,MT771345,MT771348,MZ171372,ON970620 |
| 1823 | VC_571_0 | 4 | 1169.929722 | 4919.904907 | 0.192111903 | 1.41E-05 | 2.828427125 | 8.246211251 | 6 | 6 | 1 | 6.399889864 | 0 | 0 | 0 | MG757156,MH834613,MN585993,MT776806 |
| 1824 | VC_572_0 | 16 | 31595.28854 | 11770.00962 | 0.728584603 | 2.12E-71 | 0 | 6.633249581 | 120 | 120 | 1 | 4.221284356 | 0 | 0 | 0 | MN234162,MN234190,MN234207,MN234220,MN234232,MN586003,MZ005671,OL455887,OL455890,OL455895,ON755185,ON755187,ON970611,OP172868,OP172870,OP172876 |
| 1825 | VC_573_0 | 2 | 300 | 2427.425616 | 0.109993834 | 0.043606824 | 0 | 0 | 1 | 1 | 1 | 0 | 0 | 0 | 0 | MN813687,ON108643 |
| 1826 | VC_574_0 | 7 | 3301.506079 | 8774.589514 | 0.273391847 | 8.34E-15 | 1.732050808 | 7.681145748 | 21 | 21 | 1 | 5.269607569 | 0 | 0 | 0 | JX901189,KF024728,MK279840,MW460248,MZ152914,OM793623,ON108645 |
| 1827 | VC_575_0 | 35 | 136272.7823 | 18321.46948 | 0.881486735 | 5.26E-211 | 0 | 6 | 595 | 595 | 1 | 4.089582088 | 0 | 0 | 0 | AY216660,HM035024,JX912252,KP085586,KU194205,KU194206,KX828710,KY985004,LC516895,MF093736,MF468274,MG049919,MH051911,MH051912,MH285980,MH917278,MK213796,MK335533,MK972831,MN153797,MN296515,MN337573,MN342247,MN393473,MT427400,MT682715,MZ501092,OL770108,OL800706,OL960578,OL960579,OL960582,ON185577,OP094641,OP455115 |
| 1828 | VC_576_0 | 4 | 1163.704331 | 18025.63302 | 0.060643279 | 1.23E-05 | 2.645751311 | 8.485281374 | 6 | 6 | 1 | 6.122655419 | 0 | 0 | 0 | MF996376,MH051918,MT023084,MZ501053 |
| 1829 | VC_577_0 | 6 | 1548.022128 | 122.5337887 | 0.926650891 | 7.02E-08 | 1 | 8.246211251 | 15 | 15 | 1 | 5.974610183 | 0 | 0 | 0 | AY236756,JN051154,JX486087,MG765275,MG765278,MH809528 |
| 1830 | VC_578_0 | 2 | 22.31017736 | 108.0381512 | 0.171158139 | 0.060263944 | 7.615773106 | 7.615773106 | 1 | 1 | 1 | 7.615773106 | 0 | 0 | 0 | MG020111,MK504442 |
| 1831 | VC_57_0 | 4 | 552.7163666 | 768.5908318 | 0.418310267 | 1.33E-05 | 0 | 3 | 6 | 6 | 1 | 1.776142375 | 0 | 0 | 0 | ON470596,ON470604,ON470624,ON470627 |
| 1832 | VC_580_0 | 2 | 300 | 4446.40947 | 0.063205672 | 0.042366949 | 2.645751311 | 2.645751311 | 1 | 1 | 1 | 2.645751311 | 0 | 0 | 0 | KC954775,OM638103 |
| 1833 | VC_581_0 | 1 | 0 | 594.9849057 | 0 | 0 | 0 | 0 | 1 | 1 | 1 | 0 | 0 | 0 | 0 | MW805361 |
| 1834 | VC_581_1 | 1 | 0 | 692.8442743 | 0 | 0 | 0 | 0 | 1 | 1 | 1 | 0 | 0 | 0 | 0 | OM982621 |
| 1835 | VC_582_0 | 4 | 1800 | 28784.84824 | 0.058852671 | 3.01E-05 | 2 | 8.185352772 | 6 | 6 | 1 | 5.644110395 | 0 | 0 | 0 | MN184887,MN732867,OP480062,OP480063 |
| 1836 | VC_582_1 | 3 | 900 | 13595.40449 | 0.062088643 | 0.001844336 | 0 | 1 | 3 | 3 | 1 | 0.666666667 | 0 | 0 | 0 | OK136180,OK181858,OK181861 |
| 1837 | VC_582_2 | 1 | 0 | 8711.10173 | 0 | 0 | 0 | 0 | 1 | 1 | 1 | 0 | 0 | 0 | 0 | KM190144 |
| 1838 | VC_582_3 | 12 | 19800 | 64213.85183 | 0.235675422 | 1.08E-32 | 4.795831523 | 9.899494937 | 66 | 57 | 0.863636364 | 7.513816288 | 0 | 0 | 0 | KT934943,KX452695,KY652726,MG250486,MK024806,MT178275,MT438396,MT939253,MZ602147,ON602729,ON602736,OP296941 |
| 1839 | VC_582_4 | 3 | 900 | 24990.57229 | 0.034761688 | 0.01826497 | 7.348469228 | 8.062257748 | 3 | 3 | 1 | 7.740325551 | 0 | 0 | 0 | JN882284,KF550303,MZ311864 |
| 1840 | VC_582_5 | 4 | 1800 | 32301.79282 | 0.052783149 | 0.000643752 | 5.477225575 | 9.486832981 | 6 | 5 | 0.833333333 | 7.52142506 | 0 | 0 | 0 | GU070616,JX181824,MW006477,MZ326168 |
| 1841 | VC_582_6 | 1 | 0 | 7520.844121 | 0 | 0 | 0 | 0 | 1 | 1 | 1 | 0 | 0 | 0 | 0 | MG969411 |
| 1842 | VC_584_0 | 28 | 82831.99977 | 24681.44858 | 0.770433848 | 1.19E-184 | 0 | 8 | 378 | 378 | 1 | 5.084829332 | 0 | 0 | 0 | AY308796,KM236241,KX015770,KX015771,KY657202,KY694971,LR597656,MG241338,MH102284,MH491968,MH729819,MN994500,MT478992,MW021748,MW021750,MW598258,MZ501059,OK499973,OK499984,OL539443,OL539444,OL539461,OL539462,OL539465,OL539466,ON843697,OP292673,OP292674 |
| 1843 | VC_585_0 | 4 | 1279.394267 | 22727.52265 | 0.053292735 | 1.25E-05 | 4.242640687 | 8.124038405 | 6 | 6 | 1 | 6.341460459 | 0 | 0 | 0 | MN019128,MW032452,MZ501077,MZ501085 |
| 1844 | VC_585_1 | 3 | 518.9018679 | 16078.33367 | 0.031264355 | 0.00142332 | 4.795831523 | 7.874007874 | 3 | 3 | 1 | 6.695345961 | 0 | 0 | 0 | MK778457,MN090155,MZ501099 |
| 1845 | VC_585_2 | 1 | 0 | 4166.535327 | 0 | 0 | 0 | 0 | 1 | 1 | 1 | 0 | 0 | 0 | 0 | MW689258 |
| 1846 | VC_585_3 | 1 | 0 | 5417.048994 | 0 | 0 | 0 | 0 | 1 | 1 | 1 | 0 | 0 | 0 | 0 | OM317910 |
| 1847 | VC_586_0 | 3 | 679.2056022 | 13028.51838 | 0.049549116 | 0.001400665 | 4.472135955 | 5.385164807 | 3 | 3 | 1 | 4.884377428 | 0 | 0 | 0 | MN101222,OK019720,ON045001 |
| 1848 | VC_587_0 | 2 | 78.10662623 | 162.1078398 | 0.325153716 | 0.088071125 | 7.681145748 | 7.681145748 | 1 | 1 | 1 | 7.681145748 | 0 | 0 | 0 | AY320035,KT221034 |
| 1849 | VC_588_0 | 2 | 9.923238075 | 109.4041428 | 0.083159774 | 0.056049052 | 7.348469228 | 7.348469228 | 1 | 1 | 1 | 7.348469228 | 0 | 0 | 0 | KU958700,MH744420 |
| 1850 | VC_589_0 | 2 | 36.60716638 | 0 | 1 | 0 | 2.236067977 | 2.236067977 | 1 | 1 | 1 | 2.236067977 | 0 | 0 | 0 | AY324828,MN710383 |
| 1851 | VC_590_0 | 27 | 105300 | 557.7903275 | 0.994730758 | 1.48E-107 | 2.449489743 | 7.211102551 | 351 | 351 | 1 | 5.276747218 | 0 | 0 | 0 | AY343333,EF437941,EU863408,HE978309,KY683735,MF327006,MH560358,MK327932,MK327935,MK327941,MK327947,MK573636,MK962753,MN655998,MN709127,MN850574,MN850615,MN882349,MN894885,MT478991,MT764208,MW748999,MZ504876,MZ504877,MZ504878,ON286974,OX001802 |
| 1852 | VC_591_0 | 14 | 19127.0017 | 17769.9264 | 0.518390085 | 2.94E-57 | 0 | 4.582575695 | 91 | 91 | 1 | 2.11587904 | 0 | 0 | 0 | CP059058,KP063118,MG030346,MN062188,MN604053,MN840486,MW145139,OK499974,OK499975,OK499986,OK499998,OK500000,OL741431,OM962992 |
| 1853 | VC_592_0 | 50 | 261416.1014 | 95983.34738 | 0.731439576 | 0 | 0 | 7.416198487 | 1225 | 1225 | 1 | 4.071210299 | 0 | 0 | 0 | JN871591,JQ340774,JX094499,JX296113,KC139512,KC139514,KC139515,KC139519,KC677662,KM366098,KM458633,KR296689,KR296691,KU927500,KX452699,KX452700,KX765862,KX765863,KX765864,KY014601,KY620117,LC553736,LR026998,MG280946,MH631453,MH791395,MH791396,MK249308,MK286578,MN871450,MT499897,MT499898,MT675124,MT675125,MT675126,MW057860,MW149272,MW355449,MW355460,MW355467,MW355470,MW355474,MW355476,MW355479,OL606627,OL699877,OM025231,OP453108,OP535471,OP535472 |
| 1854 | VC_593_0 | 5 | 2754.527615 | 3222.868399 | 0.460824012 | 5.09E-08 | 0 | 4.358898944 | 10 | 10 | 1 | 1.743559577 | 0 | 0 | 0 | KM581061,MH015249,MH015252,MH015254,MH015258 |
| 1855 | VC_594_0 | 2 | 207.4114343 | 2608.698784 | 0.073651746 | 0.043141407 | 3.872983346 | 3.872983346 | 1 | 1 | 1 | 3.872983346 | 0 | 0 | 0 | KT321316,OK041469 |
| 1856 | VC_597_0 | 6 | 3119.77287 | 14198.54361 | 0.180142964 | 1.89E-11 | 2 | 6.782329983 | 15 | 15 | 1 | 3.924864783 | 0 | 0 | 0 | MK838113,MN317029,MT952005,MW978786,OL840901,OL964754 |
| 1857 | VC_598_0 | 4 | 190.1483101 | 2996.216586 | 0.059675623 | 0.006446423 | 6.403124237 | 8.544003745 | 6 | 6 | 1 | 7.964181499 | 0 | 0 | 0 | HQ632859,KY249644,MF476925,MW269554 |
| 1858 | VC_599_0 | 50 | 49785.4191 | 54.95993934 | 0.998897281 | 2.66E-10 | 0 | 6.92820323 | 1225 | 1225 | 1 | 4.878632408 | 0 | 0 | 0 | AY370762,AY388628,AY423772,EU030938,EU030939,FJ870915,FJ870916,FJ870917,KC618393,KY563228,KY579375,MK054204,MK054205,MK054206,MK054207,MK054208,MK054209,MK054210,MK054211,MK054212,MK054213,MK054214,MK054215,MK054216,MK054217,MK054218,MK054219,MK054220,MK054221,MK054222,MK054223,MK054224,MK054225,MK054226,MK054227,MK054228,MK054229,MK054230,MK054231,MK054232,MK054233,MK054234,MK054235,MK054236,MK054237,MN496305,MN496306,MN496307,MN496308,X07234 |
| 1859 | VC_5_0 | 30 | 23500.21387 | 859.7231277 | 0.964707498 | 3.70E-95 | 0 | 7.141428429 | 435 | 435 | 1 | 5.800118391 | 0 | 0 | 0 | AB374228,AF125163,AY027935,BHU24159,DQ114220,DQ674738,HG803181,HQ110083,KF361475,KT887559,KU504502,KX058879,KX774374,KY045851,LR597637,LR597638,LR597640,LR597649,MG592573,MK972712,MK972713,MK972714,MN563793,MT662115,OM095401,OP484843,S1-wms-combined_159544,S11-wms-combined_154845,S5-wms-combined_18356,S6-wms-combined_119017 |
| 1860 | VC_600_0 | 3 | 900 | 15724.75535 | 0.054136135 | 0.028899307 | 2.449489743 | 5.830951895 | 3 | 3 | 1 | 4.645765296 | 0 | 0 | 0 | MK577502,MN871507,ON042478 |
| 1861 | VC_600_1 | 7 | 6300 | 27987.24762 | 0.183741783 | 3.05E-08 | 0 | 3.16227766 | 21 | 21 | 1 | 2.540839297 | 0 | 0 | 0 | AY375531,AY962392,KY290948,KY290951,KY290956,KY290957,KY290958 |
| 1862 | VC_600_2 | 10 | 13500 | 30900.04694 | 0.304053733 | 2.28E-16 | 3.872983346 | 7.280109889 | 45 | 45 | 1 | 5.629866963 | 0 | 0 | 0 | DQ529280,HM452125,JN377894,JN377895,JX306041,MF479730,MH179476,MH179477,MW584871,OL840900 |
| 1863 | VC_602_0 | 2 | 186.7156519 | 628.600438 | 0.22901014 | 0.044394478 | 1.732050808 | 1.732050808 | 1 | 1 | 1 | 1.732050808 | 0 | 0 | 0 | JN190960,MW677528 |
| 1864 | VC_603_0 | 9 | 3372.761814 | 25.6490689 | 0.992452629 | 5.86E-08 | 0 | 1.414213562 | 36 | 36 | 1 | 0.314269681 | 0 | 0 | 0 | AY441783,AY848684,AY848685,AY848686,AY848687,AY848688,AY848689,OK041468,OM630588 |
| 1865 | VC_604_0 | 22 | 20750.49858 | 5695.694667 | 0.784630831 | 3.31E-82 | 0 | 8.124038405 | 231 | 231 | 1 | 5.775002349 | 0 | 0 | 0 | KR093634,KR093635,KR093636,KR093637,KR093639,KR093640,KR093641,KR093642,KR093643,KR093644,KR093645,KR093646,KR093647,KR093648,KR093649,KR093650,KR093651,KR093652,KR093653,KR093654,KR093655,KR093656 |
| 1866 | VC_606_0 | 4 | 872.5909345 | 1063.605231 | 0.450672793 | 4.85E-05 | 5.830951895 | 8.602325267 | 6 | 6 | 1 | 7.726646001 | 0 | 0 | 0 | MG711465,S11-wms_55610,S2-wms_107329,S8-wms_82880 |
| 1867 | VC_606_1 | 1 | 0 | 882.4291923 | 0 | 0 | 0 | 0 | 1 | 1 | 1 | 0 | 0 | 0 | 0 | S9-wms-combined_29611 |
| 1868 | VC_607_0 | 16 | 18489.69643 | 1654.767957 | 0.917854954 | 8.89E-52 | 0 | 5.477225575 | 120 | 120 | 1 | 2.700670241 | 0 | 0 | 0 | AY505112,AY510084,JF712866,KM612259,KM612260,KM612261,KM612262,KM612263,KM612264,KM612265,KY883642,KY883647,KY883656,MH105773,MW824387,MW824404 |
| 1869 | VC_608_0 | 5 | 1567.62124 | 2051.577509 | 0.433140413 | 4.03E-08 | 1.414213562 | 4.795831523 | 10 | 10 | 1 | 3.76301008 | 0 | 0 | 0 | MG674163,MT992243,MW176032,MW176034,MW590698 |
| 1870 | VC_609_0 | 4 | 169.099382 | 176.6966874 | 0.48901476 | 7.58E-05 | 6.244997998 | 7.483314774 | 6 | 6 | 1 | 6.962169599 | 0 | 0 | 0 | MF074189,MW202496,MW202711,MW202879 |
| 1871 | VC_60_0 | 5 | 2441.575006 | 3321.121314 | 0.423686217 | 3.49E-08 | 0 | 0 | 10 | 10 | 1 | 0 | 0 | 0 | 0 | OK490460,OK490463,OK490466,OK490469,OK490472 |
| 1872 | VC_610_0 | 4 | 523.2076095 | 72.77961896 | 0.877883929 | 0.000146702 | 3.605551275 | 8.774964387 | 6 | 6 | 1 | 7.161281683 | 0 | 0 | 0 | S1-wms-combined_107073,S2-wms-combined_89742,S4-wms-combined_41490,S8-wms-combined_26087 |
| 1873 | VC_611_0 | 2 | 93.91643864 | 0 | 1 | 0 | 1 | 1 | 1 | 1 | 1 | 1 | 0 | 0 | 0 | AY569307,GU080336 |
| 1874 | VC_612_0 | 9 | 6923.000841 | 330.0044463 | 0.954501006 | 1.36E-09 | 0 | 1.414213562 | 36 | 36 | 1 | 0.971404521 | 0 | 0 | 0 | AY575774,DQ163914,KU884561,KU884562,KU884563,KU884564,KU884565,MK514283,ON857941 |
| 1875 | VC_613_0 | 2 | 12.20284227 | 330.0044463 | 0.035659212 | 0.058056818 | 4 | 4 | 1 | 1 | 1 | 4 | 0 | 0 | 0 | JF412296,OM519319 |
| 1876 | VC_614_0 | 5 | 2392.952933 | 7598.207687 | 0.239507003 | 3.05E-08 | 0 | 6.164414003 | 10 | 10 | 1 | 4.582695747 | 0 | 0 | 0 | KX198614,MT795651,MW114771,ON457558,ON457559 |
| 1877 | VC_615_0 | 2 | 68.12788864 | 187.2033649 | 0.266821581 | 0.058219991 | 5.916079783 | 5.916079783 | 1 | 1 | 1 | 5.916079783 | 0 | 0 | 0 | CAKLQA020000003,MZ326864 |
| 1878 | VC_616_0 | 2 | 11.90255483 | 4.352786502 | 0.732224233 | 0.185546685 | 5.196152423 | 5.196152423 | 1 | 1 | 1 | 5.196152423 | 0 | 0 | 0 | S11-wms-combined_193148,S11-wms-combined_204522 |
| 1879 | VC_618_0 | 13 | 21408.53374 | 12834.19385 | 0.625199429 | 1.55E-40 | 2.828427125 | 8.185352772 | 78 | 78 | 1 | 4.432820219 | 0 | 0 | 0 | JX128258,MH051335,MH816966,MK373776,MK373777,MK903282,MN038177,MN176217,MT446405,MT446408,OL856099,ON210137,OP352608 |
| 1880 | VC_61_0 | 12 | 1595.563257 | 494.9021532 | 0.76325743 | 6.80E-23 | 0 | 1.414213562 | 66 | 66 | 1 | 0.54913092 | 0 | 0 | 0 | AB334720,AB334721,F1CCG,GQ153916,GQ153917,GQ153918,GQ153919,JF719732,JF719733,JF719734,JX412914,PFDCG |
| 1881 | VC_620_0 | 27 | 91761.93946 | 12636.70769 | 0.87895717 | 4.28E-154 | 0 | 6.633249581 | 351 | 351 | 1 | 2.174825626 | 0 | 0 | 0 | KT361654,KT361655,KT361656,KT361657,OP494865,OP503543,OP503968,OP503969,OP503970,OP503971,OP503972,OP503973,OP503974,OP503975,OP503976,OP503977,OP503978,OP503979,OP503980,OP503981,OP503982,OP503983,OP503984,OP503985,OP503986,OP503987,OP503988 |
| 1882 | VC_621_0 | 7 | 1324.371047 | 965.9188405 | 0.578254768 | 7.63E-14 | 5.567764363 | 8.485281374 | 21 | 21 | 1 | 6.902947876 | 0 | 0 | 0 | MG711464,S1-wms-combined_23248,S11-wms-combined_140009,S12-wms-combined_345948,S12-wms-combined_71235,S17-wms-combined_90378,S4-wms-combined_42647 |
| 1883 | VC_622_0 | 2 | 15.27436348 | 286.2217024 | 0.0506619 | 0.374060699 | 6.403124237 | 6.403124237 | 1 | 1 | 1 | 6.403124237 | 0 | 0 | 0 | MF176161,MH884512 |
| 1884 | VC_623_0 | 1 | 0 | 723.8746465 | 0 | 0 | 0 | 0 | 1 | 1 | 1 | 0 | 0 | 0 | 0 | MK448818 |
| 1885 | VC_625_0 | 2 | 31.14175232 | 1172.949692 | 0.025863278 | 0.075194384 | 8.246211251 | 8.246211251 | 1 | 1 | 1 | 8.246211251 | 0 | 0 | 0 | MK448890,MK448930 |
| 1886 | VC_626_0 | 23 | 13604.18923 | 7008.47845 | 0.659991683 | 1.21E-89 | 0 | 9.899494937 | 253 | 212 | 0.837944664 | 7.725811803 | 0 | 0 | 0 | MF172979,MK448667,MK448700,MK448720,MK448839,MK448841,MK448843,MK448934,MK448997,MW387018,S1-wms-combined_332236,S11-wms-combined_194205,S12-wms-combined_368462,S12-wms-combined_7174,S12-wms-combined_99704,S17-wms-combined_9864,S2-wms-combined_174241,S2-wms-combined_244564,S5-wms-combined_206050,S5-wms-combined_29272,S8-wms-combined_108917,S8-wms-combined_186054,S9-wms-combined_25007 |
| 1887 | VC_626_1 | 1 | 0 | 1219.139539 | 0 | 0 | 0 | 0 | 1 | 1 | 1 | 0 | 0 | 0 | 0 | S9-wms-combined_9931 |
| 1888 | VC_626_2 | 1 | 0 | 830.7329334 | 0 | 0 | 0 | 0 | 1 | 1 | 1 | 0 | 0 | 0 | 0 | S12-wms-combined_98046 |
| 1889 | VC_626_3 | 1 | 0 | 1161.335998 | 0 | 0 | 0 | 0 | 1 | 1 | 1 | 0 | 0 | 0 | 0 | S5-wms-combined_108794 |
| 1890 | VC_627_0 | 3 | 68.07235785 | 251.9366996 | 0.212720097 | 0.00588632 | 6.480740698 | 7.615773106 | 3 | 3 | 1 | 7.055860539 | 0 | 0 | 0 | S1-wms_170115,S5-wms-combined_73301,S6-wms-combined_113986 |
| 1891 | VC_629_0 | 2 | 3.36577704 | 819.335632 | 0.004091128 | 0.064504847 | 7.810249676 | 7.810249676 | 1 | 1 | 1 | 7.810249676 | 0 | 0 | 0 | S2-wms-combined_196783,S7-wms_80255 |
| 1892 | VC_630_0 | 20 | 37595.95862 | 6569.501769 | 0.851252501 | 1.10E-70 | 0 | 6.164414003 | 190 | 190 | 1 | 3.328174658 | 0 | 0 | 0 | AY625898,KC262634,KC900378,LT603684,MF490237,MK511039,MK511040,MK511041,MK511042,MK511043,MK511044,MK511045,MK511046,MK511047,MK511048,MK511049,MK511050,MK511051,MT118301,MZ285879 |
| 1893 | VC_631_0 | 3 | 583.4978387 | 4225.696033 | 0.121329656 | 0.00180801 | 1.414213562 | 5.477225575 | 3 | 3 | 1 | 4.122888237 | 0 | 0 | 0 | MT152150,MT491207,MT491208 |
| 1894 | VC_632_0 | 37 | 80454.77271 | 58027.82614 | 0.58097388 | 1.44E-299 | 0 | 10.29563014 | 666 | 615 | 0.923423423 | 6.290496877 | 0 | 0 | 0 | FM864213,FN997652,KC348603,KC581799,KT336320,KT336321,KX077896,MK359990,MK448674,MK448741,MK448752,MK448811,MK448819,MK448828,MK448983,MN270258,MN270259,MN270260,MN270261,MN270262,MN270263,MN270264,MN270265,MN270266,MN270267,MN270268,MN270269,MN270270,MN270271,MN270272,MN270273,MN270274,MN270275,MN270276,MN270277,MN270278,MN270279 |
| 1895 | VC_634_0 | 3 | 900 | 3499.194644 | 0.2045829 | 0.001624074 | 4 | 5.385164807 | 3 | 3 | 1 | 4.726998777 | 0 | 0 | 0 | KU052488,MG252693,MG765276 |
| 1896 | VC_634_1 | 6 | 3883.235044 | 5301.34001 | 0.422799642 | 4.42E-11 | 3 | 8 | 15 | 15 | 1 | 6.391047017 | 0 | 0 | 0 | JX486088,KX223815,KY381600,MG744354,MG765274,ON117106 |

| | | | | | | | | | | | | | | | |
|---|---|---|---|---|---|---|---|---|---|---|---|---|---|---|---|
| 1897 | VC_636_0 | 9 | 3588.703747 | 11229.47162 | 0.242182567 | 2.80E-24 | 0 | 9.110433579 | 36 | 35 | 0.972222222 | 7.079862765 | 0 | 0 | 0 | KM289195,MK448681,MK448683,MK448783,MK448790,MK448798,MK448833,MK448967,MK449010 |
| 1898 | VC_637_0 | 115 | 515693.5824 | 189774.6858 | 0.730994725 | 0 | 0 | 9.643650761 | 6555 | 5745 | 0.876430206 | 7.896593563 | 0 | 0 | 0 | AY730274,EF177456,HE775250,HG934469,HG934470,JQ007353,JX202565,KC139511,KR270151,KT881477,KT962832,KU595434,KU951146,KU951147,KX245013,KX405002,KX649889,KY379853,KY626163,LC507823,MF001356,MF001366,MF431252,MG646668,MG969408,MG969409,MG969410,MH358359,MH370358,MH370359,MH370360,MH370361,MH370362,MH370363,MH370365,MH370372,MH370373,MH370374,MH370381,MH370384,MH370385,MH424443,MH424445,MH992509,MK249126,MK761195,MK761196,MK761197,MK761198,MK761199,MK770409,MK972691,MK972700,MK972701,MK972702,MK972703,MK972705,MK972706,MN336264,MN336265,MN336266,MN393078,MN722429,MN850616,MT074431,MT074432,MT074451,MT074462,MT074473,MT074481,MT074482,MT074483,MT074484,MT588083,MT632017,MT632018,MT632019,MT632020,MT653137,MT653144,MT677934,MT740291,MW201959,MW355463,MW355464,MW355465,MW715619,MW790495,MZ156567,MZ156568,MZ156569,MZ274225,MZ955866,OK108608,OK108609,OK143508,OL436139,OL519843,OL652658,OL770276,OM210019,OM291373,OM339548,ON167532,ON381768,ON409568,ON409569,ON409570,ON454038,ON526838,ON540301,ON720977,ON996339,OP484850,OP494211 |
| 1899 | VC_637_1 | 27 | 54781.84372 | 157737.5817 | 0.257773348 | 6.84E-210 | 0 | 9.695359715 | 351 | 346 | 0.985754986 | 5.519142779 | 0 | 0 | 0 | JX297445,KF562864,KF562865,KY176369,MK578530,MK759883,MK931446,MT074430,MT677933,MW355451,MW355452,MW355453,MW355454,MW355455,MW355456,MW355457,MW355458,MW355459,MW355461,MW355462,MW355466,MW355468,MW355469,MW355471,MW355472,MW355473,OK169616 |
| 1900 | VC_637_2 | 2 | 61.84280275 | 16192.21137 | 0.003804762 | 0.071550726 | 8.717797887 | 8.717797887 | 1 | 1 | 1 | 8.717797887 | 0 | 0 | 0 | KF148055,KM272358 |
| 1901 | VC_638_0 | 45 | 134780.0923 | 39334.04894 | 0.774090441 | 0 | 0 | 8.774964387 | 990 | 990 | 1 | 7.066828327 | 0 | 0 | 0 | GU196277,GU196278,GU196279,GU196280,GU196281,KY295895,KY295896,KY295898,MF153391,MG099933,MG252615,MG852086,MH051334,MK542015,MK737937,MK931438,MK931439,MK931440,MK931441,MN927225,MN927226,MT711523,MT711524,MT711525,MT711526,MT711527,MT711528,MW331437,MW722821,MZ234010,MZ234048,MZ576217,MZ576218,MZ832314,MZ997838,OK539620,OL539727,OM272052,OM632667,OM632668,OM810255,ON001686,ON391668,ON548432,ON556632 |
| 1902 | VC_639_0 | 2 | 146.9309448 | 379.5334857 | 0.279089975 | 0.044730829 | 5.744562647 | 5.744562647 | 1 | 1 | 1 | 5.744562647 | 0 | 0 | 0 | MK813940,OM033133 |
| 1903 | VC_63_0 | 2 | 186.1693958 | 1436.314803 | 0.114743426 | 0.043104024 | 0 | 0 | 1 | 1 | 1 | 0 | 0 | 0 | 0 | AP009146,DQ785801 |
| 1904 | VC_640_0 | 7 | 3089.679768 | 333.8623179 | 0.90248044 | 2.24E-11 | 2.449489743 | 3.872983346 | 21 | 21 | 1 | 3.311013351 | 0 | 0 | 0 | AY739900,EU340421,KF188410,KJ564036,KJ564037,KJ564038,MG913376 |
| 1905 | VC_642_0 | 7 | 2417.598764 | 13611.55131 | 0.150825138 | 4.26E-15 | 0 | 9.327379053 | 21 | 16 | 0.761904762 | 7.939462238 | 0 | 0 | 0 | HQ632860,MG592506,MG592545,MG592567,MG592568,MG592600,MG592630 |
| 1906 | VC_642_1 | 1 | 0 | 2702.709744 | 0 | 0 | 0 | 0 | 1 | 1 | 1 | 0 | 0 | 0 | 0 | OX241574 |
| 1907 | VC_642_2 | 6 | 1388.437513 | 6879.206365 | 0.167936299 | 3.78E-11 | 5.744562647 | 8.124038405 | 15 | 15 | 1 | 7.216197275 | 0 | 0 | 0 | AY772740,JF974313,MG676223,MK097141,ON714422,OP375635 |
| 1908 | VC_642_3 | 9 | 4038.243012 | 14915.79995 | 0.21305444 | 8.04E-19 | 0 | 9.327379053 | 36 | 25 | 0.694444444 | 7.723742783 | 0 | 0 | 0 | KX507045,MG592494,MG592501,MG592508,MG592514,MG592583,MG592587,MG592595,MG592644 |
| 1909 | VC_642_4 | 2 | 286.2198086 | 5230.614639 | 0.051881167 | 0.043955187 | 3.605551275 | 3.605551275 | 1 | 1 | 1 | 3.605551275 | 0 | 0 | 0 | MW824412,MW824413 |
| 1910 | VC_642_5 | 4 | 842.4429109 | 8500.710663 | 0.09016687 | 3.34E-05 | 0 | 8.660254038 | 6 | 6 | 1 | 6.832965705 | 0 | 0 | 0 | MG592544,MG592664,MG592665,OX241573 |
| 1911 | VC_643_0 | 2 | 48.5982064 | 396.602511 | 0.109160216 | 0.049146528 | 7.681145748 | 7.681145748 | 1 | 1 | 1 | 7.681145748 | 0 | 0 | 0 | JF974287,MK249872 |
| 1912 | VC_644_0 | 5 | 642.4447683 | 323.798235 | 0.664889439 | 2.49E-07 | 7.211102551 | 8.544003745 | 10 | 10 | 1 | 7.812424806 | 0 | 0 | 0 | KC542353,KP994596,KU747973,MK387309,MN313256 |
| 1913 | VC_645_0 | 8 | 3940.101255 | 8615.808508 | 0.313804521 | 1.07E-18 | 4.472135955 | 7.348469228 | 28 | 28 | 1 | 6.587604139 | 0 | 0 | 0 | MG592390,MG592420,MG592423,MG592439,MG592444,MG592454,MG592465,MG592466 |
| 1914 | VC_646_0 | 29 | 66273.46258 | 6306.494515 | 0.913109696 | 5.45E-161 | 0 | 8.124038405 | 406 | 406 | 1 | 6.928512056 | 0 | 0 | 0 | MG592393,MG592477,MG592478,MG592518,MG592540,MG592552,MG592569,MG592570,MG592608,MG592614,MG592622,MG592627,MG592628,MG592641,MG592647,MG592656,MW824369,MW824373,MW824374,MW824378,MW824379,MW824383,OX241429,OX241438,OX241498,OX241557,OX241558,OX241575,OX241591 |
| 1915 | VC_647_0 | 11 | 7027.924156 | 4724.127414 | 0.59801679 | 3.90E-34 | 1.732050808 | 5.567764363 | 55 | 55 | 1 | 4.204545798 | 0 | 0 | 0 | CP069347,HG796225,HG798901,JX145342,LK985321,LN681538,LN681540,MK473382,MT193276,MW512571,MW512572 |
| 1916 | VC_649_0 | 3 | 439.1384791 | 1011.010578 | 0.302822994 | 0.001613846 | 0 | 7.874007874 | 3 | 3 | 1 | 5.249338583 | 0 | 0 | 0 | KM983328,KM983331,KM983334 |
| 1917 | VC_64_0 | 3 | 758.319256 | 619.7639384 | 0.550271028 | 0.002143148 | 2.828427125 | 3.741657387 | 3 | 3 | 1 | 3.344728709 | 0 | 0 | 0 | JN022534,MK798143,MK798144 |
| 1918 | VC_650_0 | 10 | 1114.899638 | 1341.973229 | 0.453788087 | 2.80E-17 | 3.872983346 | 8.426149773 | 45 | 45 | 1 | 6.864971497 | 0 | 0 | 0 | S1-wms-combined_353757,S11-wms-combined_119215,S11-wms-combined_299954,S11-wms-combined_49632,S17-wms-combined_136622,S17-wms-combined_247494,S17-wms-combined_79709,S2-wms-combined_126134,S5-wms-combined_145067,S7-wms-combined_31068 |
| 1919 | VC_651_0 | 1 | 0 | 311.6378685 | 0 | 0 | 0 | 0 | 1 | 1 | 1 | 0 | 0 | 0 | 0 | S2-wms-combined_24549 |
| 1920 | VC_652_0 | 4 | 1163.472412 | 1289.531447 | 0.47430517 | 1.92E-05 | 1 | 7.141428429 | 6 | 6 | 1 | 5.583143613 | 0 | 0 | 0 | EU874396,GU229986,KX227758,KX227759 |
| 1921 | VC_654_0 | 4 | 816.5700608 | 2001.800487 | 0.289731264 | 1.74E-05 | 1 | 7.280109889 | 6 | 6 | 1 | 5.577400935 | 0 | 0 | 0 | KT725776,KX227757,KX227760,MW302330 |
| 1922 | VC_655_0 | 2 | 300 | 26685.92375 | 0.011116907 | 0.042955046 | 2 | 2 | 1 | 1 | 1 | 2 | 0 | 0 | 0 | GU071099,MH512890 |
| 1923 | VC_655_1 | 1 | 0 | 13662.93816 | 0 | 0 | 0 | 0 | 1 | 1 | 1 | 0 | 0 | 0 | 0 | GU071103 |
| 1924 | VC_655_2 | 3 | 900 | 38009.82916 | 0.023130402 | 0.001392441 | 2 | 7.549834435 | 3 | 3 | 1 | 5.609981442 | 0 | 0 | 0 | AY940168,HQ337021,HQ634176 |
| 1925 | VC_655_3 | 1 | 0 | 14822.53253 | 0 | 0 | 0 | 0 | 1 | 1 | 1 | 0 | 0 | 0 | 0 | HQ634175 |
| 1926 | VC_655_4 | 1 | 0 | 14769.21429 | 0 | 0 | 0 | 0 | 1 | 1 | 1 | 0 | 0 | 0 | 0 | JN371769 |
| 1927 | VC_655_5 | 3 | 900 | 53412.14728 | 0.016570879 | 0.001393701 | 1 | 2.449489743 | 3 | 3 | 1 | 1.895185907 | 0 | 0 | 0 | KU686207,KU686208,KU686209 |
| 1928 | VC_655_6 | 2 | 300 | 38023.22407 | 0.007828151 | 0.042248323 | 2 | 2 | 1 | 1 | 1 | 2 | 0 | 0 | 0 | GU071106,HQ634190 |
| 1929 | VC_655_7 | 3 | 900 | 53384.96291 | 0.016579177 | 0.001430221 | 5.385164807 | 7 | 3 | 3 | 1 | 5.954130127 | 0 | 0 | 0 | HQ634189,MH920639,MH920640 |
| 1930 | VC_655_8 | 1 | 0 | 18916.10699 | 0 | 0 | 0 | 0 | 1 | 1 | 1 | 0 | 0 | 0 | 0 | GU071094 |
| 1931 | VC_655_9 | 1 | 0 | 16998.53129 | 0 | 0 | 0 | 0 | 1 | 1 | 1 | 0 | 0 | 0 | 0 | GU071097 |
| 1932 | VC_656_0 | 2 | 300 | 33440.59173 | 0.008891367 | 0.077434819 | 1.414213562 | 1.414213562 | 1 | 1 | 1 | 1.414213562 | 0 | 0 | 0 | HQ316583,HQ316603 |
| 1933 | VC_656_1 | 46 | 310500 | 54309.80035 | 0.85112845 | 0 | 0 | 6.403124237 | 1035 | 1035 | 1 | 1.322924465 | 0 | 0 | 0 | KJ019028,KJ019029,KJ019031,KJ019032,KJ019034,KJ019035,KJ019036,KJ019046,KJ019047,KJ019048,KJ019050,KJ019056,KJ019057,KJ019062,KJ019070,KJ019072,KJ019073,KJ019074,KJ019075,KJ019077,KJ019078,KJ019079,KJ019080,KJ019083,KJ019112,KJ019113,KJ019115,KJ019117,KJ019118,KJ019119,KJ019120,KJ019121,KJ019124,KJ019125,KJ019126,KJ019127,KJ019129,KJ019130,KJ019131,KJ019136,KJ019139,KJ019140,KJ019160,KJ019162,KJ019164,KJ019165 |
| 1934 | VC_657_0 | 3 | 900 | 40501.8411 | 0.021738164 | 0.074718742 | 3.741657387 | 5.099019514 | 3 | 3 | 1 | 4.646565471 | 0 | 0 | 0 | KU686212,KU686213,MT586120 |
| 1935 | VC_657_1 | 41 | 246000 | 82480.51936 | 0.748902859 | 8.25E-308 | 0 | 7.211102551 | 820 | 820 | 1 | 4.601362915 | 0 | 0 | 0 | KJ019037,KJ019045,KJ019052,KJ019053,KJ019058,KJ019059,KJ019066,KJ019085,KJ019086,KJ019090,KJ019091,KJ019092,KJ019093,KJ019095,KJ019096,KJ019097,KJ019098,KJ019099,KJ019100,KJ019101,KJ019102,KJ019103,KJ019105,KJ019106,KJ019107,KJ019111,KJ019123,KJ019141,KJ019142,KJ019143,KJ019144,KJ019145,KJ019146,KJ019147,KJ019148,KJ019149,KJ019150,KJ019151,KJ019152,KJ019155,KJ019159 |
| 1936 | VC_659_0 | 2 | 5.107437333 | 196.2328999 | 0.025367184 | 0.454010834 | 5.196152423 | 5.196152423 | 1 | 1 | 1 | 5.196152423 | 0 | 0 | 0 | MH319737,MH319746 |
| 1937 | VC_65_0 | 2 | 300 | 1343.601474 | 0.182525998 | 0.044411797 | 6.8556546 | 6.8556546 | 1 | 1 | 1 | 6.8556546 | 0 | 0 | 0 | MT366760,MT366761 |
| 1938 | VC_662_0 | 62 | 567300 | 245957.5768 | 0.697564973 | 0 | 2.236067977 | 7.615773106 | 1891 | 1891 | 1 | 4.887167278 | 0 | 0 | 0 | HQ317290,HQ317291,HQ317292,KX349226,KX349227,KX349228,KX349229,KX349230,KX349231,KX349232,KX349233,KX349234,KX349235,KX349236,KX349237,KX349238,KX349239,KX349240,KX349241,KX349242,KX349243,KX349244,KX349245,KX349246,KX349247,KX349248,KX349249,KX349250,KX349251,KX349252,KX349253,KX349254,KX349255,KX349256,KX349257,KX349258,KX349259,KX349260,KX349261,KX349262,KX349263,KX349264,KX349265,KX349266,KX349267,KX349268,KX349269,KX349270,KX349271,KX349272,KX349273,KX349274,KX349275,KX349276,KX349277,KX349278,KX349279,KX349280,KX349281,KX349282,KX349283,KX349284 |
| 1939 | VC_664_0 | 4 | 1800 | 5044.937411 | 0.262968073 | 1.25E-05 | 1.732050808 | 6.92820323 | 6 | 6 | 1 | 5.173031417 | 0 | 0 | 0 | JQ245707,OP053360,OP053361,OP053362 |
| 1940 | VC_666_0 | 4 | 695.0268285 | 0 | 1 | 0 | 2.449489743 | 3.741657387 | 6 | 6 | 1 | 3.327831774 | 0 | 0 | 0 | AY950802,JN968479,KC252997,KT809302 |
| 1941 | VC_667_0 | 7 | 6297.013839 | 9056.773479 | 0.410127723 | 6.11E-15 | 0 | 7.211102551 | 21 | 21 | 1 | 5.564690762 | 0 | 0 | 0 | KF929199,KJ804259,MT596499,MZ417336,MZ417339,MZ417343,MZ417348 |
| 1942 | VC_667_1 | 1 | 0 | 2006.743693 | 0 | 0 | 0 | 0 | 1 | 1 | 1 | 0 | 0 | 0 | 0 | MH356730 |
| 1943 | VC_667_2 | 1 | 0 | 1615.098383 | 0 | 0 | 0 | 0 | 1 | 1 | 1 | 0 | 0 | 0 | 0 | MK893987 |
| 1944 | VC_668_0 | 25 | 90000 | 72484.09913 | 0.553900354 | 8.51E-141 | 3.31662479 | 9.591663047 | 300 | 288 | 0.96 | 6.259798712 | 0 | 0 | 0 | GU295964,HQ918180,KT001918,KT001919,KT321315,KX130727,LR746310,LT607758,MN101223,MT157285,MW448170,MZ322895,MZ612130,MZ707156,MZ707157,OK631812,OK631813,OM103621,OM971648,OM971649,ON515458,OP045497,OP684128,OX335396,OX335407 |
| 1945 | VC_668_1 | 1 | 0 | 10389.9482 | 0 | 0 | 0 | 0 | 1 | 1 | 1 | 0 | 0 | 0 | 0 | OP617746 |
| 1946 | VC_668_2 | 1 | 0 | 9304.308347 | 0 | 0 | 0 | 0 | 1 | 1 | 1 | 0 | 0 | 0 | 0 | MN101221 |
| 1947 | VC_668_3 | 1 | 0 | 6434.059769 | 0 | 0 | 0 | 0 | 1 | 1 | 1 | 0 | 0 | 0 | 0 | MN101224 |
| 1948 | VC_669_0 | 21 | 63000 | 110875.7817 | 0.362327631 | 2.24E-68 | 3.741657387 | 7.937253933 | 210 | 210 | 1 | 6.051823405 | 0 | 0 | 0 | JN882287,KT381880,KX431559,KX431560,KX443552,LC589952,MH823906,MN508621,MN508622,MN508623,MN508624,MT341500,MW021751,MW021756,MW250785,MW629017,MW767161,OL539470,OL539471,OL551674,ON157416 |
| 1949 | VC_669_1 | 10 | 13500 | 92961.51817 | 0.126806383 | 2.05E-12 | 2.236067977 | 9.055385138 | 45 | 44 | 0.977777778 | 7.066805594 | 0 | 0 | 0 | AY967407,HE858210,HE981739,HM134276,KC801932,KM236237,KR869820,KX245890,MT334653,MW021749 |
| 1950 | VC_669_2 | 1 | 0 | 13644.15417 | 0 | 0 | 0 | 0 | 1 | 1 | 1 | 0 | 0 | 0 | 0 | MN508817 |
| 1951 | VC_669_3 | 1 | 0 | 13041.52098 | 0 | 0 | 0 | 0 | 1 | 1 | 1 | 0 | 0 | 0 | 0 | MN038175 |
| 1952 | VC_669_4 | 1 | 0 | 6577.3216 | 0 | 0 | 0 | 0 | 1 | 1 | 1 | 0 | 0 | 0 | 0 | OK210075 |
| 1953 | VC_669_5 | 1 | 0 | 10457.96563 | 0 | 0 | 0 | 0 | 1 | 1 | 1 | 0 | 0 | 0 | 0 | MT560058 |
| 1954 | VC_66_0 | 2 | 300 | 1203.753494 | 0.199500783 | 0.045125822 | 3.16227766 | 3.16227766 | 1 | 1 | 1 | 3.16227766 | 0 | 0 | 0 | MK552140,MK552141 |
| 1955 | VC_66_1 | 1 | 0 | 886.2692331 | 0 | 0 | 0 | 0 | 1 | 1 | 1 | 0 | 0 | 0 | 0 | MN094788 |
| 1956 | VC_66_2 | 1 | 0 | 1071.180453 | 0 | 0 | 0 | 0 | 1 | 1 | 1 | 0 | 0 | 0 | 0 | MW460245 |
| 1957 | VC_66_3 | 1 | 0 | 1033.657941 | 0 | 0 | 0 | 0 | 1 | 1 | 1 | 0 | 0 | 0 | 0 | AB366653 |
| 1958 | VC_670_0 | 2 | 185.2131867 | 84.17932058 | 0.687521671 | 0.270145687 | 2.645751311 | 2.645751311 | 1 | 1 | 1 | 2.645751311 | 0 | 0 | 0 | CAKLQA020000002,MW176033 |
| 1959 | VC_671_0 | 5 | 2191.328257 | 1952.226306 | 0.528852275 | 4.43E-08 | 1.414213562 | 8.246211251 | 10 | 10 | 1 | 5.836921642 | 0 | 0 | 0 | MN937349,MW393850,MZ326859,MZ398240,MZ398241 |
| 1960 | VC_672_0 | 12 | 1011.273891 | 0 | 1 | 0 | 0 | 1.414213562 | 66 | 66 | 1 | 0.54913092 | 0 | 0 | 0 | BCP1,MG883715,MG883725,MG945215,MG945341,MG945520,MG945551,MG945686,MG945756,MT135344,MT135404,MT135412 |
| 1961 | VC_673_0 | 15 | 16448.71563 | 3527.556499 | 0.823412673 | 1.30E-27 | 0 | 9.848857802 | 105 | 80 | 0.761904762 | 6.771417482 | 0 | 0 | 0 | BK010471,MH552500,MH616963,MK069403,MK238400,MT006214,MT774381,MT774383,MT774384,MT774386,MT774394,MW067000,MW067001,MW067002,MW067003 |
| 1962 | VC_674_0 | 1 | 0 | 1009.741299 | 0 | 0 | 0 | 0 | 1 | 1 | 1 | 0 | 0 | 0 | 0 | S8-wms_111056 |
| 1963 | VC_675_0 | 2 | 70.38024472 | 798.229082 | 0.081026352 | 0.049814183 | 4.582575695 | 4.582575695 | 1 | 1 | 1 | 4.582575695 | 0 | 0 | 0 | MT774393,S7-wms_244425 |
| 1964 | VC_676_0 | 2 | 103.9474573 | 908.9196867 | 0.102626942 | 0.048432853 | 8.185352772 | 8.185352772 | 1 | 1 | 1 | 8.185352772 | 0 | 0 | 0 | S12-wms-combined_2450,S5-wms-combined_29447 |
| 1965 | VC_676_1 | 4 | 813.3028709 | 1350.165691 | 0.37592544 | 3.07E-05 | 5.830951895 | 9.273618495 | 6 | 4 | 0.666666667 | 7.592631815 | 0 | 0 | 0 | S11-wms-combined_83571,S6-wms-combined_50721,S7-wms-combined_107679_0,S9-wms-combined_28805 |
| 1966 | VC_677_0 | 9 | 1998.43589 | 3278.793801 | 0.378690337 | 3.19E-15 | 3 | 10.24695077 | 36 | 30 | 0.833333333 | 6.984753598 | 0 | 0 | 0 | BK010646,S11-wms-combined_195425,S12-wms-combined_250551,S3-wms-combined_73262,S5-wms-combined_59064,S7-wms-combined_43950,S7-wms-combined_5625,S8-wms-combined_97848,S9-wms-combined_31456 |
| 1967 | VC_678_0 | 1 | 0 | 363.4689654 | 0 | 0 | 0 | 0 | 1 | 1 | 1 | 0 | 0 | 0 | 0 | S12-wms-combined_358797 |
| 1968 | VC_679_0 | 4 | 337.9785062 | 889.1482352 | 0.275422656 | 2.26E-05 | 2.645751311 | 6.164414003 | 6 | 6 | 1 | 5.326435648 | 0 | 0 | 0 | S1-wms-combined_246923,S1-wms-combined_340558,S1-wms-combined_52643,S11-wms-combined_172889 |
| 1969 | VC_67_0 | 2 | 300 | 21184.88311 | 0.013963306 | 0.306423895 | 6.782329983 | 6.782329983 | 1 | 1 | 1 | 6.782329983 | 0 | 0 | 0 | KY448244,MW366843 |
| 1970 | VC_67_1 | 8 | 8400 | 70354.72113 | 0.106660272 | 0.001067539 | 0 | 3.741657387 | 28 | 28 | 1 | 2.939946756 | 0 | 0 | 0 | MZ475891,MZ475892,MZ475893,MZ475894,MZ475895,MZ475896,MZ475897,MZ475898 |
| 1971 | VC_67_2 | 2 | 300 | 21194.82003 | 0.013956851 | 0.306423895 | 2.645751311 | 2.645751311 | 1 | 1 | 1 | 2.645751311 | 0 | 0 | 0 | KY984068,MZ443769 |
| 1972 | VC_67_3 | 20 | 57000 | 103922.2986 | 0.354208214 | 2.11E-23 | 0 | 8.485281374 | 190 | 190 | 1 | 6.942078762 | 0 | 0 | 0 | MH248138,MH460463,MZ443770,MZ443771,MZ443772,MZ443773,MZ443774,MZ443776,MZ443777,MZ443778,MZ443779,MZ443780,MZ443782,MZ443783,MZ443784,MZ443785,MZ443786,MZ443787,MZ443788,MZ443789 |
| 1973 | VC_67_4 | 4 | 1800 | 39915.17313 | 0.043149767 | 0.090569651 | 0 | 4.582575695 | 6 | 6 | 1 | 3.159537972 | 0 | 0 | 0 | MH389777,MH460460,MH460461,MH460462 |
| 1974 | VC_67_5 | 1 | 0 | 6188.660477 | 0 | 0 | 0 | 0 | 1 | 1 | 1 | 0 | 0 | 0 | 0 | LT598654 |
| 1975 | VC_680_0 | 2 | 67.76650507 | 515.0306915 | 0.116278022 | 0.050604009 | 5.830951895 | 5.830951895 | 1 | 1 | 1 | 5.830951895 | 0 | 0 | 0 | S5-wms-combined_83370,S9-wms-combined_44402 |
| 1976 | VC_681_0 | 3 | 75.85897246 | 5.549985161 | 0.931825866 | 0.185546685 | 3.741657387 | 4.472135955 | 3 | 3 | 1 | 4.228643099 | 0 | 0 | 0 | BK013345,BK013346,BK013347 |
| 1977 | VC_682_0 | 2 | 91.75309723 | 0 | 1 | 0 | 5.744562647 | 5.744562647 | 1 | 1 | 1 | 5.744562647 | 0 | 0 | 0 | BK059895,OM373202 |
| 1978 | VC_683_0 | 5 | 2481.039374 | 286.985402 | 0.896321231 | 2.57E-07 | 3.31662479 | 6.08276253 | 10 | 10 | 1 | 4.994404776 | 0 | 0 | 0 | BK061308,BK061309,BK061310,BK061311,BK061312 |
| 1979 | VC_684_0 | 5 | 2454.127061 | 3813.759577 | 0.391539797 | 5.15E-08 | 0 | 5.291502622 | 10 | 10 | 1 | 3.353378039 | 0 | 0 | 0 | MW353181,MW584168,MW584172,MW584178,MW584190 |
| 1980 | VC_685_0 | 16 | 15703.29683 | 37129.70216 | 0.297225165 | 3.64E-76 | 0 | 9.433981132 | 120 | 110 | 0.916666667 | 7.675232537 | 0 | 0 | 0 | MG872834,MH513979,MH651167,MK284520,MK433274,MK433275,MK820641,MN062715,MT521999,MW534378,MW601219,MZ322028,MZ622161,MZ622165,ON108651,OP434460 |
| 1981 | VC_686_0 | 2 | 300 | 1097.01388 | 0.21474375 | 0.046080597 | 2.449489743 | 2.449489743 | 1 | 1 | 1 | 2.449489743 | 0 | 0 | 0 | MK494099,ON260818 |
| 1982 | VC_686_1 | 2 | 300 | 1060.930368 | 0.220437435 | 0.086720853 | 2.645751311 | 2.645751311 | 1 | 1 | 1 | 2.645751311 | 0 | 0 | 0 | KU728633,MK977701 |
| 1983 | VC_687_0 | 3 | 900 | 632.737921 | 0.587184533 | 0.007473349 | 0 | 1.414213562 | 3 | 3 | 1 | 0.942809042 | 0 | 0 | 0 | KF114879,KF114880,KX656785 |
| 1984 | VC_687_1 | 2 | 165.531821 | 576.9118011 | 0.222955408 | 0.085815228 | 4 | 4 | 1 | 1 | 1 | 4 | 0 | 0 | 0 | KF114876,KF114877 |
| 1985 | VC_688_0 | 2 | 271.5102337 | 1.476517761 | 0.994591248 | 0.5 | 1.414213562 | 1.414213562 | 1 | 1 | 1 | 1.414213562 | 0 | 0 | 0 | MF974396,MF974397 |
| 1986 | VC_689_0 | 2 | 68.40404897 | 86.97364221 | 0.440243696 | 0.044175637 | 4.242640687 | 4.242640687 | 1 | 1 | 1 | 4.242640687 | 0 | 0 | 0 | DQ490056,MW791312 |
| 1987 | VC_68_0 | 1 | 0 | 870.9884294 | 0 | 0 | 0 | 0 | 1 | 1 | 1 | 0 | 0 | 0 | 0 | AP018399 |
| 1988 | VC_68_1 | 1 | 0 | 891.9527154 | 0 | 0 | 0 | 0 | 1 | 1 | 1 | 0 | 0 | 0 | 0 | MN047793 |
| 1989 | VC_690_0 | 50 | 146258.3169 | 2907.587734 | 0.980507692 | 1.94E-89 | 0 | 3.872983346 | 1225 | 1225 | 1 | 2.68525706 | 0 | 0 | 0 | C2PVCG,KX373685,KX373686,KX373687,KX373688,KX373689,KX373690,KX373691,KX373692,L67CG,MF443119,MF443120,MF443121,MF443122,MF443123,MF443124,MF443125,MF443126,MH779518,MH779519,MH779520,MH779521,MH779522,MH779523,MH779524,MH779525,MH779526,MH779527,MK301442,MK301443,MN337887,MN689503,MN689504,MN689505,MN689506,MN689507,MN689508,MN689510,MN689511,MN689512,MN689513,MN689515,MN689526,MN689527,MN689528,MN689529,MN689531,MN689532,MN689533,MZ955869 |
| 1990 | VC_691_0 | 2 | 300 | 2550.665151 | 0.105238597 | 0.046569618 | 1 | 1 | 1 | 1 | 1 | 1 | 0 | 0 | 0 | MT939241,ON506926 |
| 1991 | VC_691_1 | 5 | 3000 | 3884.292072 | 0.435774655 | 6.53E-08 | 2.828427125 | 4.242640687 | 10 | 10 | 1 | 3.514585849 | 0 | 0 | 0 | CAJCJZ010000002,LT546029,LT615366,MT119360,MT119361 |
| 1992 | VC_692_0 | 2 | 300 | 366.1608314 | 0.450341698 | 0.046592743 | 1 | 1 | 1 | 1 | 1 | 1 | 0 | 0 | 0 | MF001358,MF001361 |
| 1993 | VC_694_0 | 1 | 0 | 3285.002126 | 0 | 0 | 0 | 0 | 1 | 1 | 1 | 0 | 0 | 0 | 0 | MW672041 |
| 1994 | VC_695_0 | 48 | 170278.2921 | 38349.50091 | 0.816182205 | 6.73E-251 | 0 | 7.874007874 | 1128 | 1128 | 1 | 5.826470904 | 0 | 0 | 0 | CAJDJX010000002,CAJDKF010000002,JX193904,KF728385,KF733017,KJ127304,KR131750,KT932701,KX284704,LC623721,MF678788,MF678789,MF678790,MG264739,MG708276,MH193369,MH203383,MH203384,MH355583,MH375074,MK125140,MK360024,MK721186,MK721187,MK721190,MK721191,MK721199,MK721200,MK982307,MT119359,MT520979,MT627482,MT661597,MT661598,MT829326,MT857001,MZ182246,MZ272341,MZ326865,MZ333461,OL505084,OL539449,OL799256,OL799257,ON113176,ON113334,OP254195,OP559177 |
| 1995 | VC_696_0 | 2 | 269.1717675 | 1388.432964 | 0.162385979 | 0.054263206 | 2.645751311 | 2.645751311 | 1 | 1 | 1 | 2.645751311 | 0 | 0 | 0 | KT588075,MZ514874 |
| 1996 | VC_696_1 | 8 | 2948.754805 | 2460.195169 | 0.545162151 | 6.31E-13 | 2.828427125 | 9.38083152 | 28 | 26 | 0.928571429 | 7.533217174 | 0 | 0 | 0 | CAKLQA020000001,JX403940,KP861230,KT588073,MH115576,MT361972,MZ675741,ON391949 |
| 1997 | VC_697_0 | 5 | 2170.252717 | 443.5210452 | 0.830313912 | 2.89E-07 | 2 | 5.567764363 | 10 | 10 | 1 | 4.37314319 | 0 | 0 | 0 | KJ507100,MT104473,MT104475,MZ348425,MZ868718 |
| 1998 | VC_698_0 | 26 | 56402.45878 | 3817.346316 | 0.936609786 | 1.93E-77 | 1 | 8.366600265 | 325 | 325 | 1 | 6.981701229 | 0 | 0 | 0 | HE806280,HM368260,JX976549,KJ817802,KP861229,KP861231,KU510289,KY670595,KY829116,MF346584,MH853786,MH853787,MH853788,MN166083,MN516421,MN516422,MZ593728,MZ618622,MZ712044,OL743187,OM471864,ON036882,ON036883,ON237674,ON480525,OP585104 |
| 1999 | VC_699_0 | 71 | 297286.7845 | 20605.49821 | 0.935180879 | 0 | 0 | 6.480740698 | 2485 | 2485 | 1 | 4.870621324 | 0 | 0 | 0 | CAKLQB020000001,CAKLQG020000001,HQ186308,JX658790,KC311669,KR149290,KT339321,KT388102,KT388103,KT804908,KY082667,KY268295,KY268296,KY888680,MF033347,MF033348,MF033349,MF033350,MF033351,MG459218,MG599035,MH042230,MH763831,MH800200,MK089780,MK257719,MK257720,MK257721,MK257722,MK257723,MK278859,MN061582,MN212906,MN294712,MN395291,MN433707,MN604238,MN604239,MN614471,MN651570,MN807295,MT263719,MT633129,MT741944,MW056501,MW056502,MW056503,MW345241,MW366783,MW366784,MW459163,MW926912,MZ099557,MZ593174,MZ868724,MZ868725,MZ868726,MZ936314,MZ936315,MZ936316,MZ967493,OK546191,OL770263,OL964948,OM925528,ON210142,ON513429,OP019135,OP171942,OP291336,OP508218 |
| 2000 | VC_6_0 | 44 | 106631.7623 | 10800.78664 | 0.908025614 | 0 | 0 | 6.782329983 | 946 | 946 | 1 | 4.1988097 | 0 | 0 | 0 | AY319521,CP019275,CP025900,HM770079,KT630647,KY271398,MK416009,MK416011,MK416012,MK416020,MK422449,MK422453,MK422454,MK433577,MK433578,MK433580,MK433581,MK433582,OK490395,OK490397,OK490399,OK490401,OK490403,OK490405,OK490407,OK490411,OK490413,OK490418,OK490423,OK490425,OK490427,OK490429,OK490431,OK490434,OK490436,OK490439,OK490442,OK490445,OK490448,OK490452,OK490455,OK490457,ON470620,OV696611 |
| 2001 | VC_700_0 | 10 | 13500 | 3216.691654 | 0.807576061 | 7.57E-25 | 4.123105626 | 7.280109889 | 45 | 45 | 1 | 5.907179569 | 0 | 0 | 0 | KJ628499,MH746814,MH800198,MH800199,MK170160,MT188223,MT623546,MW349133,OL960030,OP585105 |
| 2002 | VC_700_1 | 1 | 0 | 2061.708323 | 0 | 0 | 0 | 0 | 1 | 1 | 1 | 0 | 0 | 0 | 0 | KJ473422 |
| 2003 | VC_701_0 | 2 | 300 | 1716.936821 | 0.148740405 | 0.047500846 | 8.602325267 | 8.602325267 | 1 | 1 | 1 | 8.602325267 | 0 | 0 | 0 | MN276049,MZ477002 |
| 2004 | VC_701_1 | 2 | 300 | 2000.902268 | 0.130383634 | 0.048858364 | 8.306623863 | 8.306623863 | 1 | 1 | 1 | 8.306623863 | 0 | 0 | 0 | MH133207,MZ043613 |
| 2005 | VC_701_2 | 1 | 0 | 1229.569888 | 0 | 0 | 0 | 0 | 1 | 1 | 1 | 0 | 0 | 0 | 0 | CAKLQE020000001 |
| 2006 | VC_702_0 | 5 | 3000 | 848.9987429 | 0.779423481 | 1.17E-07 | 5.916079783 | 7.615773106 | 10 | 10 | 1 | 6.736913996 | 0 | 0 | 0 | KM672662,KY000079,MW316731,MW316732,MW316733 |
| 2007 | VC_704_0 | 16 | 16538.32406 | 6196.970899 | 0.727429492 | 7.63E-66 | 0 | 8.246211251 | 120 | 120 | 1 | 4.798018361 | 0 | 0 | 0 | DQ150593,DQ222851,DQ222853,DQ222855,DQ289555,DQ289556,JQ062992,KY963369,KY963370,KY963371,MK085976,MT745954,MT745955,MT745956,OK499979,OK499999 |
| 2008 | VC_705_0 | 11 | 6097.60396 | 784.107456 | 0.886059236 | 1.16E-25 | 2 | 8.062257748 | 55 | 55 | 1 | 6.190584445 | 0 | 0 | 0 | KF591601,MF614100,MH363708,MK105855,MK125141,MK651787,MN850581,MN850588,MN850623,MW258710,MW258711 |
| 2009 | VC_706_0 | 2 | 152.4024416 | 0 | 1 | 0 | 2.449489743 | 2.449489743 | 1 | 1 | 1 | 2.449489743 | 0 | 0 | 0 | CP002495,CP091911 |
| 2010 | VC_707_0 | 6 | 1721.677634 | 4859.569075 | 0.26160357 | 4.64E-11 | 5.477225575 | 9.643650761 | 15 | 13 | 0.866666667 | 7.662822746 | 0 | 0 | 0 | S1-wms-combined_132934,S11-wms-combined_304239,S12-wms-combined_17617,S12-wms-combined_18287,S17-wms-combined_199440,S6-wms-combined_8508 |
| 2011 | VC_707_1 | 9 | 2058.584435 | 5559.310228 | 0.2702301 | 4.30E-14 | 5.099019514 | 9.643650761 | 36 | 32 | 0.888888889 | 7.441568442 | 0 | 0 | 0 | MG711462,S12-wms-combined_129554,S17-wms-combined_127688,S2-wms-combined_180640,S2-wms-combined_85381,S6-wms-combined_117756,S7-wms_159432,S8-wms-combined_200168,S8-wms-combined_217866 |
| 2012 | VC_707_2 | 1 | 0 | 1334.336668 | 0 | 0 | 0 | 0 | 1 | 1 | 1 | 0 | 0 | 0 | 0 | S12-wms-combined_129799 |
| 2013 | VC_707_3 | 1 | 0 | 1057.448786 | 0 | 0 | 0 | 0 | 1 | 1 | 1 | 0 | 0 | 0 | 0 | S11-wms-combined_264050 |
| 2014 | VC_708_0 | 47 | 155269.4686 | 24155.55053 | 0.865372451 | 1.07E-270 | 0 | 7.810249676 | 1081 | 1081 | 1 | 5.727269177 | 0 | 0 | 0 | KF010834,KP202972,KP296791,KP296793,KP296794,KP296795,KP296796,KT361649,KT361650,KT361652,MG727695,MG727696,MG727698,MG727699,MG727700,MG727701,MG727702,MH431931,MH431932,MH431933,MH431934,MH431935,MH431936,MH431937,MH454077,MH454078,MH454079,MH454082,MH454083,MH454084,MW927511,MW927513,MW927514,MW927515,MW927516,MW927517,MW927519,MW927520,MW927521,MW927522,MW927523,MW927524,MW927525,OP503992,OP503993,OP503994,OP503995 |
| 2015 | VC_70_0 | 27 | 19598.84556 | 13168.99846 | 0.598112148 | 7.79E-104 | 0 | 9.110433579 | 351 | 347 | 0.988603989 | 6.738422522 | 0 | 0 | 0 | AB920995,AB983711,GU393987,HG793132,HG796219,HG796220,HG796221,KC853746,KT381879,KX660669,KY316062,KY464836,KY981272,MF621978,MH684921,MN191861,MT740728,MZ326863,OK319016,OK335783,ON170174,ON170178,ON170184,ON170190,ON170318,ON170358,ON529868 |
| 2016 | VC_710_0 | 10 | 2966.817404 | 9161.037562 | 0.244628371 | 2.77E-21 | 6.244997998 | 9.899494937 | 45 | 37 | 0.822222222 | 8.091139012 | 0 | 0 | 0 | MG711466,ON470591,S1-wms_296850,S11-wms-combined_74983,S12-wms-combined_156234,S17-wms-combined_260343,S6-wms-combined_126236,S7-wms-combined_134458,S8-wms_152831,S8-wms_48935 |
| 2017 | VC_710_1 | 1 | 0 | 1507.739953 | 0 | 0 | 0 | 0 | 1 | 1 | 1 | 0 | 0 | 0 | 0 | S2-wms-combined_137119 |
| 2018 | VC_710_2 | 1 | 0 | 1051.567598 | 0 | 0 | 0 | 0 | 1 | 1 | 1 | 0 | 0 | 0 | 0 | S7-wms-combined_157260 |
| 2019 | VC_710_3 | 1 | 0 | 1573.976143 | 0 | 0 | 0 | 0 | 1 | 1 | 1 | 0 | 0 | 0 | 0 | S6-wms-combined_118427 |
| 2020 | VC_710_4 | 1 | 0 | 1459.564153 | 0 | 0 | 0 | 0 | 1 | 1 | 1 | 0 | 0 | 0 | 0 | S2-wms-combined_13246 |
| 2021 | VC_711_0 | 3 | 591.8326688 | 1967.749302 | 0.231222393 | 0.00157505 | 5.567764363 | 7.416198487 | 3 | 3 | 1 | 6.436172723 | 0 | 0 | 0 | MG711467,S1-wms-combined_68998,S7-wms-combined_111004 |
| 2022 | VC_711_1 | 1 | 0 | 941.7882192 | 0 | 0 | 0 | 0 | 1 | 1 | 1 | 0 | 0 | 0 | 0 | S12-wms-combined_323943 |
| 2023 | VC_711_2 | 1 | 0 | 868.7989642 | 0 | 0 | 0 | 0 | 1 | 1 | 1 | 0 | 0 | 0 | 0 | S17-wms-combined_2234 |
| 2024 | VC_711_3 | 1 | 0 | 867.1943258 | 0 | 0 | 0 | 0 | 1 | 1 | 1 | 0 | 0 | 0 | 0 | S2-wms-combined_140996 |
| 2025 | VC_713_0 | 3 | 775.6014264 | 750.4802458 | 0.508230615 | 0.001728838 | 0 | 1.732050808 | 3 | 3 | 1 | 1.154700538 | 0 | 0 | 0 | ON464760,ON464761,ON464762 |
| 2026 | VC_715_0 | 3 | 131.093981 | 307.2034508 | 0.299098218 | 0.001865828 | 7.549834435 | 8.306623863 | 3 | 3 | 1 | 7.910155391 | 0 | 0 | 0 | S1-wms-combined_302166,S11-wms-combined_198512,S12-wms-combined_358010 |
| 2027 | VC_716_0 | 2 | 46.44857974 | 406.7762208 | 0.102484638 | 0.045370232 | 5 | 5 | 1 | 1 | 1 | 5 | 0 | 0 | 0 | S1-wms-combined_360285,S11-wms-combined_84326 |
| 2028 | VC_717_0 | 7 | 2260.479892 | 1628.032166 | 0.581322588 | 6.71E-13 | 0 | 9.695359715 | 21 | 19 | 0.904761905 | 7.328184691 | 0 | 0 | 0 | LR596903,MG711461,S12-wms-combined_259783,S2-wms-combined_207832,S3-wms_22744,S5-wms-combined_134381,S7-wms_118607 |
| 2029 | VC_717_1 | 1 | 0 | 1460.068596 | 0 | 0 | 0 | 0 | 1 | 1 | 1 | 0 | 0 | 0 | 0 | S17-wms-combined_248321 |
| 2030 | VC_718_0 | 4 | 864.1333326 | 487.5788638 | 0.63928796 | 3.32E-05 | 3.741657387 | 9.591663047 | 6 | 5 | 0.833333333 | 6.917246021 | 0 | 0 | 0 | KP137433,KP137435,KP137437,KP137439 |
| 2031 | VC_719_0 | 1 | 0 | 7.187772826 | 0 | 0 | 0 | 0 | 1 | 1 | 1 | 0 | 0 | 0 | 0 | S1-wms_102342 |

| | | | | | | | | | | | | | | | | |
|---|---|---|---|---|---|---|---|---|---|---|---|---|---|---|---|---|
| 2032 | VC_719_1 | 1 | 0 | 70.90669296 | 0 | 0 | 0 | 0 | 1 | 1 | 1 | 0 | 0 | 0 | 0 | S7-wms-combined_20921 |
| 2033 | VC_720_0 | 2 | 300 | 237.7728731 | 0.557856328 | 0.105649774 | 1 | 1 | 1 | 1 | 1 | 1 | 0 | 0 | 0 | KT151955,KT151957 |
| 2034 | VC_720_1 | 1 | 0 | 220.4440851 | 0 | 0 | 0 | 0 | 1 | 1 | 1 | 0 | 0 | 0 | 0 | CP027117 |
| 2035 | VC_721_0 | 9 | 1114.033729 | 0 | 1 | 0 | 0 | 1 | 36 | 36 | 1 | 0.5 | 0 | 0 | 0 | CP045566,MN890136,MN890137,MN890138,MN890139,MN890140,MN890141,MN890142,MN890143 |
| 2036 | VC_722_0 | 13 | 18689.96688 | 17931.75838 | 0.510351895 | 2.49E-47 | 2.236067977 | 5.385164807 | 78 | 78 | 1 | 4.26912753 | 0 | 0 | 0 | KJ419279,KP719132,KP719133,KP719134,KY926791,MK165087,MK373794,MK373795,MT682705,MZ501057,MZ501068,OM386652,ON922918 |
| 2037 | VC_723_0 | 9 | 9387.359787 | 3763.61425 | 0.713814791 | 6.13E-20 | 0 | 8.717797887 | 36 | 36 | 1 | 7.246134483 | 0 | 0 | 0 | CP053388,CP054387,CP058330,CP101890,CP101896,JQ965645,MK422451,MZ779062,OV696614 |
| 2038 | VC_724_0 | 3 | 900 | 4773.545586 | 0.158630963 | 0.001848425 | 4.358898944 | 5.291502622 | 3 | 3 | 1 | 4.81541103 | 0 | 0 | 0 | MT770738,MT778838,MW960030 |
| 2039 | VC_724_1 | 6 | 4500 | 5776.315294 | 0.437900149 | 1.46E-10 | 0 | 8.831760866 | 15 | 15 | 1 | 6.228076692 | 0 | 0 | 0 | JX483881,MN988505,MN988506,MN988543,MN988547,MN988548 |
| 2040 | VC_725_0 | 4 | 1800 | 2417.659507 | 0.426776983 | 2.64E-05 | 2.449489743 | 7.745966692 | 6 | 6 | 1 | 6.044164163 | 0 | 0 | 0 | MG018927,MT354569,OM140839,OM223115 |
| 2041 | VC_726_0 | 3 | 30.64228132 | 773.2673444 | 0.038116575 | 0.110443923 | 8.185352772 | 8.94427191 | 3 | 3 | 1 | 8.634863023 | 0 | 0 | 0 | JN116824,KR053195,MH153804 |
| 2042 | VC_727_0 | 11 | 6153.897136 | 6022.120403 | 0.505411323 | 1.03E-28 | 0 | 7.416198487 | 55 | 55 | 1 | 5.515265293 | 0 | 0 | 0 | JX182371,KY092479,KY092480,KY092481,KY092482,KY092483,KY092484,KY676784,MH825699,MK305888,MT711975 |
| 2043 | VC_728_0 | 3 | 896.8620978 | 1336.013332 | 0.401662397 | 0.001497412 | 2.828427125 | 3.605551275 | 3 | 3 | 1 | 3.346509892 | 0 | 0 | 0 | MK279841,MT316461,MT723933 |
| 2044 | VC_72_0 | 2 | 155.0281624 | 1840.655257 | 0.077681741 | 0.043835387 | 3.16227766 | 3.16227766 | 1 | 1 | 1 | 3.16227766 | 0 | 0 | 0 | JN882298,MG201401 |
| 2045 | VC_730_0 | 9 | 3328.009786 | 0 | 1 | 0 | 0 | 5.291502622 | 36 | 36 | 1 | 3.682573764 | 0 | 0 | 0 | CP060690,HQ377372,HQ377373,JF773396,KX879601,KX879602,KY661963,MT899443,MT899444 |
| 2046 | VC_731_0 | 2 | 108.9578026 | 185.734697 | 0.369733884 | 0.050202073 | 5.830951895 | 5.830951895 | 1 | 1 | 1 | 5.830951895 | 0 | 0 | 0 | KX190833,MK759918 |
| 2047 | VC_732_0 | 2 | 12.56463272 | 227.1135663 | 0.052422927 | 0.092571862 | 7.141428429 | 7.141428429 | 1 | 1 | 1 | 7.141428429 | 0 | 0 | 0 | S12-wms-combined_191570,S2-wms-combined_238929 |
| 2048 | VC_732_1 | 1 | 0 | 22.88917021 | 0 | 0 | 0 | 0 | 1 | 1 | 1 | 0 | 0 | 0 | 0 | S17-wms-combined_95360 |
| 2049 | VC_733_0 | 5 | 1751.560852 | 1675.2474 | 0.511134771 | 6.34E-08 | 0 | 3.605551275 | 10 | 10 | 1 | 2.352846001 | 0 | 0 | 0 | HM568888,KU057941,LN681535,LN681536,LN881738 |
| 2050 | VC_734_0 | 6 | 94.5687546 | 399.4030455 | 0.191445655 | 3.11E-05 | 6.08276253 | 8.774964387 | 15 | 15 | 1 | 7.599088012 | 0 | 0 | 0 | HQ332138,KU517658,MF595878,S2-wms-combined_101826,S2-wms-combined_248329,S5-wms-combined_7292 |
| 2051 | VC_736_0 | 3 | 60.19369253 | 58.85059373 | 0.505641173 | 0.048046165 | 6.164414003 | 8.124038405 | 3 | 3 | 1 | 6.973900663 | 0 | 0 | 0 | S12-wms-combined_168259,S12-wms-combined_282594,S5-wms-combined_52741 |
| 2052 | VC_738_0 | 2 | 80.52440074 | 39.50457993 | 0.670874653 | 0.144422183 | 6.32455532 | 6.32455532 | 1 | 1 | 1 | 6.32455532 | 0 | 0 | 0 | S2-wms-combined_135139,S8-wms-combined_246185 |
| 2053 | VC_739_0 | 26 | 97500 | 4365.885299 | 0.95714085 | 1.15E-120 | 0 | 7.681145748 | 325 | 325 | 1 | 4.322761581 | 0 | 0 | 0 | DQ003638,DQ004855,JQ797329,JX126918,KJ094029,KJ094030,KJ094031,KJ094033,KJ535721,KJ535722,KJ586794,KJ586795,KJ591604,KJ591605,KJ668715,KM373208,MK792752,MN128594,MN172529,MN939539,MN939540,MT178766,MT178767,MT438761,MT668623,MT668624 |
| 2054 | VC_740_0 | 3 | 132.766692 | 524.6203007 | 0.20196124 | 0.001881716 | 3.872983346 | 6 | 3 | 3 | 1 | 5.023045256 | 0 | 0 | 0 | DQ003641,EU855793,GQ403788 |
| 2055 | VC_743_0 | 15 | 2915.121088 | 0 | 1 | 0 | 0 | 2 | 105 | 105 | 1 | 1.057342236 | 0 | 0 | 0 | DQ079870,DQ079871,DQ079872,DQ079873,DQ079874,DQ079875,DQ079876,DQ079878,DQ079879,DQ085810,GQ149088,MG945254,MK629527,X60322,X60323 |
| 2056 | VC_744_0 | 10 | 905.4949876 | 379.1266269 | 0.704872919 | 3.16E-15 | 1.732050808 | 5 | 45 | 45 | 1 | 3.607641204 | 0 | 0 | 0 | DQ123818,MK016490,MK016500,MN988551,MZ333134,MZ333136,ON381767,ON724010,OV696617,S2-wms_79304 |
| 2057 | VC_745_0 | 4 | 491.8710019 | 388.7823469 | 0.558529645 | 4.20E-05 | 0 | 1.732050808 | 6 | 6 | 1 | 1.093412989 | 0 | 0 | 0 | MT006233,MT006238,MT006239,MT006240 |
| 2058 | VC_747_0 | 23 | 41839.43348 | 9169.011548 | 0.820245225 | 3.54E-98 | 0 | 8.185352772 | 253 | 253 | 1 | 4.279196873 | 0 | 0 | 0 | DQ163912,KJ959591,KY707339,MK510962,MK510963,MK510964,MK510965,MK510968,MK510969,MK510970,MK510971,MK510974,MK510975,MK510976,MK510978,MK510984,MK510985,MK510986,MK510987,MK510988,MK510990,MK510992,MK510993 |
| 2059 | VC_748_0 | 12 | 14576.12067 | 6612.811209 | 0.687912008 | 1.44E-38 | 0 | 2.449489743 | 66 | 66 | 1 | 0.527426811 | 0 | 0 | 0 | MK510966,MK510967,MK510972,MK510973,MK510977,MK510979,MK510980,MK510981,MK510982,MK510983,MK510989,MK510991 |
| 2060 | VC_749_0 | 40 | 89068.12443 | 29428.63212 | 0.751650315 | 6.72E-208 | 0 | 8.717797887 | 780 | 780 | 1 | 5.696608124 | 0 | 0 | 0 | DQ163913,HG962376,JF939047,JQ067092,JQ307387,KM434185,KR537871,KR537872,KR935213,KR935215,KU197014,LC552830,LR743530,MG897800,MK165657,MK165658,MK574078,MN062187,MT118304,MT118305,MW512832,MW677514,MW677526,MZ501266,MZ927745,MZ927746,OK094665,OK330455,OK330456,OK539825,OL802211,OM234791,ON189042,ON189043,ON189044,ON911538,ON932082,ON932085,OP009282,OP422637 |
| 2061 | VC_74_0 | 15 | 12583.74351 | 11599.27169 | 0.520354613 | 3.76E-60 | 2 | 7.071067812 | 105 | 105 | 1 | 4.898699302 | 0 | 0 | 0 | KF626667,KU197013,KU595432,KU595433,LR778216,MF375456,MK903278,MN478375,OK258138,OK275495,ON911539,ON932078,ON932081,ON932083,ON960072 |
| 2062 | VC_750_0 | 1 | 0 | 2714.239436 | 0 | 0 | 0 | 0 | 1 | 1 | 1 | 0 | 0 | 0 | 0 | MG366115 |
| 2063 | VC_751_0 | 4 | 1284.182478 | 6000.89537 | 0.176275739 | 1.47E-05 | 1.414213562 | 2.645751311 | 6 | 6 | 1 | 1.9463942 | 0 | 0 | 0 | KP313532,MK962635,MK962636,MK962637 |
| 2064 | VC_752_0 | 3 | 509.8066057 | 5959.402942 | 0.078805085 | 0.001539802 | 2 | 3.872983346 | 3 | 3 | 1 | 3.159511541 | 0 | 0 | 0 | MN734437,MN734438,MN734439 |
| 2065 | VC_754_0 | 8 | 6625.038031 | 2185.22346 | 0.751968377 | 1.29E-18 | 0 | 3.605551275 | 28 | 28 | 1 | 2.356362535 | 0 | 0 | 0 | HQ641345,HQ641346,KM224878,KM224879,KY883657,KY883658,MT066160,MT767883 |
| 2066 | VC_755_0 | 9 | 5135.247874 | 6303.565628 | 0.448931865 | 2.54E-23 | 2.828427125 | 7.483314774 | 36 | 36 | 1 | 5.732572787 | 0 | 0 | 0 | DQ163915,KU535860,LT986460,MF788075,MG676466,MW929175,MZ089731,MZ727333,ON857932 |
| 2067 | VC_756_0 | 2 | 151.4358819 | 773.4472642 | 0.163735151 | 0.045729859 | 2.828427125 | 2.828427125 | 1 | 1 | 1 | 2.828427125 | 0 | 0 | 0 | KC751414,KF302037 |
| 2068 | VC_757_0 | 2 | 219.3274309 | 2797.325369 | 0.07270556 | 0.044726375 | 0 | 0 | 1 | 1 | 1 | 0 | 0 | 0 | 0 | MG592599,MG592619 |
| 2069 | VC_758_0 | 18 | 24677.61951 | 8758.743841 | 0.738047354 | 1.21E-80 | 0 | 6.708203932 | 153 | 153 | 1 | 4.340089816 | 0 | 0 | 0 | MG592629,MW824407,MW824408,MW824409,MW865297,MW865299,MW865302,MW865307,OL676770,OX241435,OX241436,OX241445,OX241446,OX241447,OX241501,OX241504,OX241559,OX241560 |
| 2070 | VC_759_0 | 3 | 874.4873857 | 967.5061026 | 0.47475053 | 0.001866735 | 3 | 5.385164807 | 3 | 3 | 1 | 4.428048098 | 0 | 0 | 0 | MN988507,MN988510,MN988527 |
| 2071 | VC_75_0 | 2 | 163.224133 | 2735.262044 | 0.05631358 | 0.043807214 | 1.732050808 | 1.732050808 | 1 | 1 | 1 | 1.732050808 | 0 | 0 | 0 | MZ666965,MZ666966 |
| 2072 | VC_760_0 | 3 | 43.85797604 | 16.374553 | 0.728144355 | 0.014092901 | 7 | 7.211102551 | 3 | 3 | 1 | 7.117510326 | 0 | 0 | 0 | MW202785,MW202796,OK172329 |
| 2073 | VC_761_0 | 67 | 462930.5845 | 54493.33465 | 0.894683387 | 0 | 0 | 5.477225575 | 2211 | 2211 | 1 | 2.185786347 | 0 | 0 | 0 | DQ398042,EU568876,FJ973624,GQ303261,JN412593,JN699002,KC787103,KC787104,KC787107,KC787108,KC787109,KC787110,KC787111,KC787112,KJ725374,KM923970,KT347314,KT355472,KT355474,KT365399,KX443326,KX534004,KX588251,KX641265,KX641266,KX664455,MF668268,MF668272,MF919497,MG099946,MH001455,MH001456,MH045569,MH077584,MH371116,MH371119,MH450127,MH479920,MH590605,MH779505,MH779507,MH779509,MH779513,MH779514,MH779515,MK305884,MK310146,MK433277,MK433279,MK524493,MK524494,MK919475,MN444875,MT818422,MW119570,MW291028,MZ171377,MZ747508,ON392160,ON392162,ON637759,ON637765,ON637766,ON645337,ON970618,OP172872,OP294982 |
| 2074 | VC_762_0 | 9 | 5276.430779 | 54317.00831 | 0.088540465 | 3.00E-24 | 3 | 6.8556546 | 36 | 36 | 1 | 5.625723684 | 0 | 0 | 0 | KJ410133,KR080197,KR080198,KR080202,MF141540,MK494105,MN234183,MN234185,MN234219 |
| 2075 | VC_763_0 | 2 | 300 | 4620.301278 | 0.060971876 | 0.049047866 | 5.656854249 | 5.656854249 | 1 | 1 | 1 | 5.656854249 | 0 | 0 | 0 | MW584160,MW584202 |
| 2076 | VC_763_1 | 9 | 10800 | 18339.30416 | 0.370633421 | 1.56E-20 | 0 | 6.92820323 | 36 | 36 | 1 | 3.504694552 | 0 | 0 | 0 | JN699628,KC900379,KT591491,MH697587,MK937612,MN586024,MT658803,MW713383,OP297551 |
| 2077 | VC_763_2 | 7 | 6138.695275 | 17365.66776 | 0.261172586 | 1.81E-13 | 2.645751311 | 8.717797887 | 21 | 21 | 1 | 6.239868842 | 0 | 0 | 0 | JF937105,KY223999,KY783914,MF319184,MT657341,MT658802,MW578836 |
| 2078 | VC_764_0 | 4 | 1800 | 3261.795678 | 0.355605029 | 2.53E-05 | 0 | 3.741657387 | 6 | 6 | 1 | 2.687325274 | 0 | 0 | 0 | DQ398052,KP027195,KY385381,MN585992 |
| 2079 | VC_765_0 | 132 | 640830.6623 | 0 | 1 | 0 | 0 | 7.549834435 | 8646 | 8646 | 1 | 5.802307653 | 0 | 0 | 0 | DQ431235,FJ706171,FJ706172,JX262215,JX262216,JX262217,JX262218,JX262219,JX262220,JX262221,JX262222,JX262223,JX262224,JX262225,JX570702,JX570703,JX570704,JX570705,JX570706,JX570707,JX570708,JX570709,JX570710,JX570711,JX570712,JX570713,JX570714,KJ578758,KJ578759,KJ578760,KJ578761,KJ578762,KJ578763,KJ578764,KJ578765,KJ578766,KJ578767,KJ578768,KJ578769,KJ578770,KJ578771,KJ578772,KJ578773,KJ578774,KJ578775,KJ578776,KJ578777,KJ578778,KJ578779,KJ578780,KJ578781,KJ578782,KJ578783,KJ578784,KJ578785,KJ578786,KJ578787,KJ578788,KJ578789,KJ578790,KJ578791,KJ578792,KJ722067,KR337643,KR337644,KR337645,KR337646,KR337647,KR337648,KR337649,KR337650,KR337651,KR337652,KR337653,KR337654,KR347352,KR347353,KR347354,KR347355,KR902978,KR902979,KR902980,KR902981,KR902982,KR902983,KR902984,KR902985,KR902986,KR902987,KT934381,KY926792,KY926793,MF919491,MF919505,MF919515,MF919516,MF919522,MF919533,MG820632,MG820633,MG820634,MG820635,MG820636,MG820637,MG820638,MG820639,MG820640,MG820641,MG820642,MG820643,MG820644,MG820645,MN329678,MN813675,MN813677,MN813679,MN813683,MN813688,MN813689,MN813690,MN813691,MN813692,MT647607,MW161461,MW161462,MW161463,MW161464,MW161465,MW161466,MW161467,MW161468,MW505928 |
| 2080 | VC_766_0 | 2 | 139.3917034 | 381.9073105 | 0.267392993 | 0.050838929 | 3.605551275 | 3.605551275 | 1 | 1 | 1 | 3.605551275 | 0 | 0 | 0 | LN681534,MN718463 |
| 2081 | VC_767_0 | 34 | 116922.9948 | 10538.32175 | 0.917321411 | 3.65E-157 | 0 | 8.306623863 | 561 | 561 | 1 | 6.538373213 | 0 | 0 | 0 | JF923796,JF923797,KR053201,MF919510,MF919542,MG757157,MG757166,MH536819,MH536824,MH576960,MH669007,MH669013,MH727547,MH727552,MH744424,MH779499,MH779501,MH779508,MH779510,MK376954,MK376960,MK376962,MK433272,MK977699,MN369753,MN369766,MN428063,MT889393,MT986025,MW507129,MZ322010,MZ322022,MZ622178,ON456333 |
| 2082 | VC_768_0 | 8 | 7678.085453 | 3272.043043 | 0.70118679 | 9.30E-19 | 0 | 3.741657387 | 28 | 28 | 1 | 2.767178385 | 0 | 0 | 0 | MG198776,MG198777,MG198778,MG198780,MH926056,MH926057,MH976516,MK977700 |
| 2083 | VC_769_0 | 2 | 300 | 5908.36276 | 0.048321919 | 0.044857026 | 0 | 0 | 1 | 1 | 1 | 0 | 0 | 0 | 0 | MW452562,MW582531 |
| 2084 | VC_76_0 | 37 | 199800 | 12675.11806 | 0.940345401 | 1.99E-249 | 0 | 8.366600265 | 666 | 666 | 1 | 4.593315182 | 0 | 0 | 0 | GU988610,HE983845,HQ832595,KC862297,KC862298,KC862300,KR052143,KT736033,KT804923,KU297675,KU497559,KU761955,KY000083,KY073228,LN610572,LN610575,LN610586,LN610587,MF974178,MH370478,MH725810,MH791399,MH791408,MH791416,MK138526,MK224498,MK340761,MK443264,MN013356,MN563785,MN871470,MN871481,MN871488,MW526259,OK539826,ON169972,ON361135 |
| 2085 | VC_76_1 | 1 | 0 | 8764.895999 | 0 | 0 | 0 | 0 | 1 | 1 | 1 | 0 | 0 | 0 | 0 | MK443265 |
| 2086 | VC_770_0 | 5 | 1155.419465 | 220.9835972 | 0.839448485 | 5.95E-06 | 6.633249581 | 8.831760866 | 10 | 10 | 1 | 8.281026192 | 0 | 0 | 0 | DQ500118,MN988496,MW960032,OP484857,OP484858 |
| 2087 | VC_771_0 | 2 | 222.8787542 | 69.62866705 | 0.761959315 | 0.083990929 | 0 | 0 | 1 | 1 | 1 | 0 | 0 | 0 | 0 | MN988490,MN988502 |
| 2088 | VC_772_0 | 6 | 4500 | 143183.2802 | 0.030470612 | 0.189871701 | 1 | 8.485281374 | 15 | 15 | 1 | 5.120721746 | 0 | 0 | 0 | MG065644,MG065662,MG065672,MG065674,MG065677,MG065690 |
| 2089 | VC_772_1 | 2 | 300 | 50058.43737 | 0.005957294 | 0.423176388 | 8.717797887 | 8.717797887 | 1 | 1 | 1 | 8.717797887 | 0 | 0 | 0 | MK962749,ON286975 |
| 2090 | VC_772_10 | 8 | 8400 | 185389.8746 | 0.043345918 | 0.091181698 | 0 | 9.797958971 | 28 | 26 | 0.928571429 | 6.615135979 | 0 | 0 | 0 | MN850630,MT884008,MT884009,MT884010,MT884011,MZ501082,MZ501083,MZ501102 |
| 2091 | VC_772_11 | 6 | 4500 | 142956.7099 | 0.030517431 | 0.157461973 | 1.414213562 | 9.486832981 | 15 | 12 | 0.8 | 7.896730496 | 0 | 0 | 0 | LC739541,LC739542,MH491969,MH752840,MZ501048,MZ501094 |
| 2092 | VC_772_12 | 1 | 0 | 23826.56746 | 0 | 0 | 0 | 0 | 1 | 1 | 1 | 0 | 0 | 0 | 0 | MG065659 |
| 2093 | VC_772_13 | 1 | 0 | 23845.72953 | 0 | 0 | 0 | 0 | 1 | 1 | 1 | 0 | 0 | 0 | 0 | MG065653 |
| 2094 | VC_772_14 | 3 | 900 | 74290.53569 | 0.011969592 | 0.360204934 | 1 | 8.062257748 | 3 | 3 | 1 | 5.687419249 | 0 | 0 | 0 | MG065654,MG065655,MG065666 |
| 2095 | VC_772_15 | 5 | 3000 | 120477.4574 | 0.024295933 | 0.238551388 | 1.414213562 | 9.16515139 | 10 | 9 | 0.9 | 7.618194443 | 0 | 0 | 0 | DQ832317,KC690136,KP869102,KP869103,KP869112 |
| 2096 | VC_772_16 | 10 | 13500 | 226275.5 | 0.056302666 | 0.056507198 | 1.414213562 | 9.899494937 | 45 | 39 | 0.866666667 | 7.991383739 | 0 | 0 | 0 | KJ190158,LC739538,LC739540,MG065646,MG065647,MG065658,MG065661,MG065682,MG963916,MN850642 |
| 2097 | VC_772_17 | 2 | 300 | 50118.27029 | 0.005950224 | 0.429740878 | 7.874007874 | 7.874007874 | 1 | 1 | 1 | 7.874007874 | 0 | 0 | 0 | MW748998,MZ501073 |
| 2098 | VC_772_18 | 1 | 0 | 25204.93072 | 0 | 0 | 0 | 0 | 1 | 1 | 1 | 0 | 0 | 0 | 0 | MZ501061 |
| 2099 | VC_772_19 | 16 | 36000 | 333103.2673 | 0.09753368 | 0.002953397 | 5.477225575 | 9.591663047 | 120 | 97 | 0.808333333 | 8.220898076 | 0 | 0 | 0 | KT001917,LR699804,MG065635,MK327930,MK373780,MN850576,MN850578,MN850595,MN850597,MN850612,MN850626,MN850627,MZ501054,MZ501062,OK272482,OV101294 |
| 2100 | VC_772_2 | 1 | 0 | 25318.4133 | 0 | 0 | 0 | 0 | 1 | 1 | 1 | 0 | 0 | 0 | 0 | MZ501060 |
| 2101 | VC_772_20 | 2 | 300 | 49946.55334 | 0.005970559 | 0.411314129 | 8.831760866 | 8.831760866 | 1 | 1 | 1 | 8.831760866 | 0 | 0 | 0 | MN850593,MT682716 |
| 2102 | VC_772_3 | 1 | 0 | 25289.6848 | 0 | 0 | 0 | 0 | 1 | 1 | 1 | 0 | 0 | 0 | 0 | MZ501093 |
| 2103 | VC_772_4 | 4 | 1800 | 97447.29796 | 0.018136514 | 0.269816967 | 5.196152423 | 8.062257748 | 6 | 6 | 1 | 6.948725067 | 0 | 0 | 0 | JQ031132,MK883717,MK883718,MW883061 |
| 2104 | VC_772_5 | 1 | 0 | 25323.47938 | 0 | 0 | 0 | 0 | 1 | 1 | 1 | 0 | 0 | 0 | 0 | MT740314 |
| 2105 | VC_772_6 | 3 | 900 | 74214.28565 | 0.011981742 | 0.360204934 | 7.874007874 | 8.185352772 | 3 | 3 | 1 | 7.998871526 | 0 | 0 | 0 | MN850646,MN850649,MZ501111 |
| 2106 | VC_772_7 | 2 | 300 | 49969.9172 | 0.005967784 | 0.411312863 | 1.414213562 | 1.414213562 | 1 | 1 | 1 | 1.414213562 | 0 | 0 | 0 | LR694610,LR694611 |
| 2107 | VC_772_8 | 2 | 300 | 50007.49052 | 0.005963327 | 0.417063176 | 8.426149773 | 8.426149773 | 1 | 1 | 1 | 8.426149773 | 0 | 0 | 0 | KR698074,ON286973 |
| 2108 | VC_772_9 | 7 | 6300 | 164419.517 | 0.036902635 | 0.122944969 | 0 | 8.602325267 | 21 | 21 | 1 | 7.15404982 | 0 | 0 | 0 | KY683736,LN881727,LN881735,LR597647,LR694165,MG022439,MG022440 |
| 2109 | VC_773_0 | 23 | 33128.63817 | 26951.18396 | 0.55141039 | 5.19E-147 | 0 | 6.633249581 | 253 | 253 | 1 | 4.681915739 | 0 | 0 | 0 | DQ864624,MK448678,MK448689,MK448693,MK448696,MK448697,MK448699,MK448766,MK448779,MK448784,MK448789,MK448797,MK448852,MK448866,MK448867,MK448868,MK448871,MK448947,MK448952,MK448961,MK448968,MK448974,MK448976 |
| 2110 | VC_774_0 | 5 | 1856.897624 | 3801.168194 | 0.328185936 | 4.45E-08 | 0 | 5 | 10 | 10 | 1 | 3.357615421 | 0 | 0 | 0 | GQ478081,GQ478082,GQ478083,GQ478084,GQ478085 |
| 2111 | VC_775_0 | 3 | 244.7177624 | 6472.95906 | 0.036428928 | 0.00149927 | 8.485281374 | 8.94427191 | 3 | 3 | 1 | 8.67729285 | 0 | 0 | 0 | MK448854,MK448933,MK448950 |
| 2112 | VC_775_1 | 14 | 7306.152807 | 18818.92942 | 0.279660471 | 1.10E-56 | 0 | 10.14889157 | 91 | 69 | 0.758241758 | 8.127152968 | 0 | 0 | 0 | MK448673,MK448765,MK448775,MK448794,MK448795,MK448858,MK448864,MK448941,MK448944,MK448945,MK448946,MK448953,MK448957,MK448970 |
| 2113 | VC_777_0 | 3 | 208.6113942 | 229.3501718 | 0.476323519 | 0.003818798 | 2.236067977 | 5 | 3 | 3 | 1 | 4.045015821 | 0 | 0 | 0 | EF537008,KF598865,MT457475 |
| 2114 | VC_778_0 | 4 | 737.3286073 | 171.2281121 | 0.811538335 | 0.000145652 | 2 | 4.582575695 | 6 | 6 | 1 | 3.686969088 | 0 | 0 | 0 | DQ875742,OM897575,ON677538,OP056300 |
| 2115 | VC_779_0 | 37 | 118325.4986 | 89920.22656 | 0.568201333 | 0 | 0 | 8.544003745 | 666 | 666 | 1 | 6.240049647 | 0 | 0 | 0 | EF056009,HQ259105,JX415535,JX880034,KC206276,KF192075,KF562340,KF620435,KJ135004,LR999871,MG208881,MG770228,MG922974,MH358458,MH669274,MH733496,MH992123,MK931297,MT682708,MW175414,MW630115,MZ358387,MZ394712,MZ501049,OM632664,OM632665,OM632666,OM632669,OM632670,OM632672,OM982646,ON602749,ON602751,ON922919,OP312987,OV049961,OW991345 |
| 2116 | VC_779_1 | 11 | 9792.26289 | 51899.96447 | 0.15872766 | 3.02E-36 | 0 | 8.660254038 | 55 | 55 | 1 | 6.569343817 | 0 | 0 | 0 | KF787094,KF787095,KT321317,MG018928,MK962626,MK962629,MK962630,MK962631,MK962632,MK962639,MK962641 |
| 2117 | VC_779_2 | 5 | 2415.600617 | 25159.05866 | 0.087602193 | 3.02E-08 | 2 | 7 | 10 | 10 | 1 | 4.613887505 | 0 | 0 | 0 | KF806588,KX098389,KX098390,KX098391,MH160392 |
| 2118 | VC_779_3 | 7 | 2525.695849 | 30446.6471 | 0.076600436 | 2.98E-11 | 2.645751311 | 9.695359715 | 21 | 15 | 0.714285714 | 7.440163657 | 0 | 0 | 0 | MG983743,MK863032,MN062186,MZ326856,MZ326857,MZ326861,OM687511 |
| 2119 | VC_77_0 | 20 | 57000 | 2021.61029 | 0.965747965 | 9.64E-94 | 0 | 6.708203932 | 190 | 190 | 1 | 4.229773102 | 0 | 0 | 0 | AB472900,KC862295,KC862296,KC862299,KC862301,KM434186,KY994101,LN610573,LN610576,LN610581,LN610582,LN610583,MK799650,MT108728,MT108730,MT118294,MT118295,MT118300,OM421595,ON653033 |
| 2120 | VC_780_0 | 8 | 6297.213526 | 5796.76405 | 0.520690028 | 1.11E-19 | 3.605551275 | 7.483314774 | 28 | 28 | 1 | 5.888299469 | 0 | 0 | 0 | KC139517,KC139520,KM236242,MH899585,MN163281,MW544066,OM858838,OM908766 |
| 2121 | VC_781_0 | 2 | 265.8677255 | 6825.881525 | 0.037489724 | 0.043087922 | 2.236067977 | 2.236067977 | 1 | 1 | 1 | 2.236067977 | 0 | 0 | 0 | KU356690,KU356691 |
| 2122 | VC_782_0 | 3 | 872.3706223 | 2074.941541 | 0.295988539 | 0.001437147 | 0 | 3.464101615 | 3 | 3 | 1 | 2.309401077 | 0 | 0 | 0 | MG592603,MG592604,MG592610 |
| 2123 | VC_783_0 | 3 | 600.9441881 | 8806.604363 | 0.063878936 | 0.001456927 | 2.236067977 | 7.745966692 | 3 | 3 | 1 | 5.887726806 | 0 | 0 | 0 | MN228696,MW960036,MW960038 |
| 2124 | VC_784_0 | 6 | 3077.842112 | 2501.120836 | 0.55168714 | 6.34E-11 | 2.236067977 | 8.544003745 | 15 | 15 | 1 | 6.155311269 | 0 | 0 | 0 | KC438282,KC438283,KP280062,KY065148,KY612839,OP313112 |
| 2125 | VC_786_0 | 3 | 900 | 10150.08573 | 0.081447332 | 0.001459028 | 0 | 0 | 3 | 3 | 1 | 0 | 0 | 0 | 0 | MT234338,MT234339,MT234340 |
| 2126 | VC_78_0 | 2 | 300 | 9502.196575 | 0.030605385 | 0.043826134 | 6.403124237 | 6.403124237 | 1 | 1 | 1 | 6.403124237 | 0 | 0 | 0 | MG018930,MZ605293 |
| 2127 | VC_78_1 | 6 | 4500 | 28536.24847 | 0.136214014 | 8.37E-11 | 0 | 4.472135955 | 15 | 15 | 1 | 3.313315198 | 0 | 0 | 0 | KU130126,KU130127,KU130128,KU130129,KU130130,KU130131 |
| 2128 | VC_78_2 | 1 | 0 | 7154.663407 | 0 | 0 | 0 | 0 | 1 | 1 | 1 | 0 | 0 | 0 | 0 | OK040171 |
| 2129 | VC_78_3 | 11 | 16500 | 47643.65672 | 0.257235101 | 1.76E-14 | 1.732050808 | 7.483314774 | 55 | 55 | 1 | 5.422404343 | 0 | 0 | 0 | KJ409772,MN905160,MT670417,MT670418,MT670419,MT670420,MT670421,MT670422,MW175491,MZ348426,MZ868713 |
| 2130 | VC_78_4 | 1 | 0 | 7281.899923 | 0 | 0 | 0 | 0 | 1 | 1 | 1 | 0 | 0 | 0 | 0 | MN812694 |
| 2131 | VC_78_5 | 6 | 4500 | 36884.41982 | 0.108736573 | 0.000219177 | 0 | 7.348469228 | 15 | 15 | 1 | 5.279130029 | 0 | 0 | 0 | LN887844,MZ826350,MZ826353,OK258139,OK275491,OM982620 |
| 2132 | VC_78_6 | 1 | 0 | 7501.578133 | 0 | 0 | 0 | 0 | 1 | 1 | 1 | 0 | 0 | 0 | 0 | MF893340 |
| 2133 | VC_791_0 | 3 | 239.1210589 | 0 | 1 | 0 | 2.236067977 | 4.69041576 | 3 | 3 | 1 | 3.683196454 | 0 | 0 | 0 | EF432053,KP282673,KP282674 |
| 2134 | VC_792_0 | 35 | 104343.6366 | 1823.489572 | 0.982824348 | 8.52E-163 | 0 | 8.544003745 | 595 | 595 | 1 | 5.68601442 | 0 | 0 | 0 | EF460875,HQ728264,KF534715,KY087898,MF044458,MH191398,MN045229,MN098326,MN117721,MN122072,MN136198,MN812688,MN850594,MT230534,MT446397,MT446398,MT446399,MT446400,MT446402,MT446403,MT446406,MT446409,MT446419,MW147598,MW147599,MW147600,MW147601,MW147602,MW349138,OK073976,OK129343,OK148439,OL741059,OL960574,ON778463 |
| 2135 | VC_793_0 | 12 | 10587.83541 | 1823.489572 | 0.853078573 | 2.49E-39 | 0 | 7.071067812 | 66 | 66 | 1 | 4.738283606 | 0 | 0 | 0 | KJ002054,KJ858521,KY709296,LR813619,MT623578,MW380983,MW380984,MW380985,MZ336020,OL770365,OM810291,OP056089 |
| 2136 | VC_794_0 | 3 | 60.36276343 | 0 | 1 | 0 | 1.414213562 | 2.236067977 | 3 | 3 | 1 | 1.794110782 | 0 | 0 | 0 | EF506570,SVU28974,X51344 |
| 2137 | VC_795_0 | 19 | 45673.65119 | 21026.96374 | 0.684756074 | 6.72E-94 | 0 | 6.403124237 | 171 | 171 | 1 | 4.524674234 | 0 | 0 | 0 | KM236243,KP064094,KX534337,LN881725,LR025195,LR025196,LR025197,LR025198,LT907986,MH230177,MK672798,MZ501071,MZ501091,MZ501104,MZ501110,MZ501112,OL539454,OL960573,OL960575 |
| 2138 | VC_796_0 | 5 | 2913.103985 | 5777.094967 | 0.33521718 | 3.19E-08 | 0 | 2 | 10 | 10 | 1 | 1.4 | 0 | 0 | 0 | MK816297,MN029011,MN871471,MN871472,MN871474 |
| 2139 | VC_798_0 | 2 | 81.66170869 | 367.4662179 | 0.181822826 | 0.056539872 | 8.124038405 | 8.124038405 | 1 | 1 | 1 | 8.124038405 | 0 | 0 | 0 | S1-wms-combined_307323,S7-wms-combined_147158 |
| 2140 | VC_798_1 | 1 | 0 | 180.9023059 | 0 | 0 | 0 | 0 | 1 | 1 | 1 | 0 | 0 | 0 | 0 | S1-wms-combined_85225 |
| 2141 | VC_798_2 | 1 | 0 | 238.2138971 | 0 | 0 | 0 | 0 | 1 | 1 | 1 | 0 | 0 | 0 | 0 | S17-wms-combined_186314 |
| 2142 | VC_799_0 | 2 | 300 | 7884.073077 | 0.036656564 | 0.044523346 | 2.828427125 | 2.828427125 | 1 | 1 | 1 | 2.828427125 | 0 | 0 | 0 | GU580941,GU580942 |
| 2143 | VC_799_1 | 27 | 80451.01648 | 12972.86533 | 0.861139731 | 6.40E-161 | 0 | 7.681145748 | 351 | 351 | 1 | 5.431412858 | 0 | 0 | 0 | MG962368,MG962369,MK376953,MK376957,MN010761,MN204501,MN284896,MN284904,MN585971,MN585982,MN585988,MN585996,MT310850,MT498039,MT498052,MT521994,MW291022,MW862987,MZ005675,MZ005683,MZ171370,MZ209301,MZ388556,MZ622168,ON526971,ON970572,ON970622 |
| 2144 | VC_79_0 | 64 | 340539.6994 | 473080.0691 | 0.41854895 | 0 | 0 | 10.19803903 | 2016 | 1542 | 0.764880952 | 8.236851446 | 0 | 0 | 0 | AB560486,AP018815,FM201282,JN254800,KP340287,KP340288,KR869157,KX171208,LC102730,LC105987,LC105988,LC105989,LC105990,LC472883,LC472884,LN610579,LN610588,MF490240,MG897799,MN871458,MN871459,MN871461,MN871462,MN871464,MN871465,MN871466,MN871473,MN871479,MN871485,MN871487,MN871508,MN871509,MT118291,MT118297,MT118298,MT118299,MT119363,MT119365,MT119368,MT119376,MT133561,MT133562,MT349888,MT413450,MT933737,MW512833,MW595221,MZ553930,OL362268,OL362269,OL754588,OM953790,ON324181,ON375838,ON375839,ON375840,ON529291,ON857930,ON857934,ON857942,OP310975,OP310977,OP314870,OP314871 |
| 2145 | VC_79_1 | 2 | 169.6194151 | 28098.91702 | 0.00600029 | 0.042512573 | 8.124038405 | 8.124038405 | 1 | 1 | 1 | 8.124038405 | 0 | 0 | 0 | LC472881,MW406974 |
| 2146 | VC_79_10 | 1 | 0 | 14898.99722 | 0 | 0 | 0 | 0 | 1 | 1 | 1 | 0 | 0 | 0 | 0 | AP019535 |
| 2147 | VC_79_11 | 1 | 0 | 14254.58589 | 0 | 0 | 0 | 0 | 1 | 1 | 1 | 0 | 0 | 0 | 0 | KY000082 |
| 2148 | VC_79_2 | 1 | 0 | 13697.49644 | 0 | 0 | 0 | 0 | 1 | 1 | 1 | 0 | 0 | 0 | 0 | FM201281 |
| 2149 | VC_79_3 | 1 | 0 | 14506.83799 | 0 | 0 | 0 | 0 | 1 | 1 | 1 | 0 | 0 | 0 | 0 | OP310979 |
| 2150 | VC_79_4 | 3 | 522.9873019 | 48984.25632 | 0.010563854 | 0.003679085 | 7.615773106 | 9.219544457 | 3 | 2 | 0.666666667 | 8.172154437 | 0 | 0 | 0 | MF623055,MT118289,MT118292 |
| 2151 | VC_79_5 | 2 | 159.6536676 | 34526.90695 | 0.004602753 | 0.07218034 | 8.717797887 | 8.717797887 | 1 | 1 | 1 | 8.717797887 | 0 | 0 | 0 | MK837009,MW595220 |
| 2152 | VC_79_6 | 11 | 9262.439235 | 166082.4306 | 0.052824124 | 4.42E-19 | 2.236067977 | 9.539392014 | 55 | 41 | 0.745454545 | 7.971332033 | 0 | 0 | 0 | MK837010,MN871457,MN871460,MN871467,MT108729,MT491204,MT491205,MT491206,MW512830,MW512834,OP310974 |
| 2153 | VC_79_7 | 33 | 94647.75936 | 343556.7013 | 0.21598995 | 5.13E-286 | 0 | 10.14889157 | 528 | 444 | 0.840909091 | 7.875683632 | 0 | 0 | 0 | FM887021,FM897211,GU815091,KR054033,KU198331,MF490241,MF768470,MK318076,MN131141,MN131142,MN131143,MN615700,MN615701,MN615702,MT108727,MT118290,MT119366,MT119369,MZ285878,ON857925,ON857929,ON857937,ON857938,OP310970,OP310971,OP310972,OP310973,OP310976,OP310978,OP310980,OP310981,OP310982,OP314872 |
| 2154 | VC_79_8 | 1 | 0 | 14710.63843 | 0 | 0 | 0 | 0 | 1 | 1 | 1 | 0 | 0 | 0 | 0 | KM434184 |
| 2155 | VC_79_9 | 2 | 148.7562797 | 24157.58282 | 0.006120061 | 0.051399994 | 7.416198487 | 7.416198487 | 1 | 1 | 1 | 7.416198487 | 0 | 0 | 0 | MK340760,MN871468 |
| 2156 | VC_7_0 | 28 | 35372.94817 | 54946.06413 | 0.391644542 | 9.39E-213 | 0 | 9.273618495 | 378 | 376 | 0.994708995 | 7.330070585 | 0 | 0 | 0 | AB009866,AB044554,AB243556,AF424783,CP062426,CP062435,CP062445,CP062457,EF462197,EF462199,FJ713816,GQ915271,KC342645,KX232515,KY389063,KY389064,KY653125,KY695153,MH384260,MH401414,MH401415,MW147366,MW192778,MW627293,MW650841,OM439664,OP136133,OP493557 |
| 2157 | VC_800_0 | 3 | 718.9121272 | 847.0350659 | 0.459090913 | 0.001526301 | 1.414213562 | 3 | 3 | 3 | 1 | 2.353321624 | 0 | 0 | 0 | EU081845,MK779875,MN552143 |
| 2158 | VC_801_0 | 4 | 1650.228433 | 1767.891007 | 0.482788405 | 1.30E-05 | 1.414213562 | 2.236067977 | 6 | 6 | 1 | 1.710107921 | 0 | 0 | 0 | KJ489010,KJ489011,KJ489012,KJ489013 |
| 2159 | VC_802_0 | 17 | 37435.43938 | 9211.235889 | 0.802531781 | 4.85E-78 | 0 | 5.291502622 | 136 | 136 | 1 | 4.317211283 | 0 | 0 | 0 | KU160642,KU647628,MF189170,MF189171,MF189172,MF189173,MF189174,MF189175,MF324906,MH450120,MK919473,MK967398,MN284897,MZ209300,MZ209307,MZ747520,OP434451 |
| 2160 | VC_803_0 | 3 | 332.9240126 | 6162.528472 | 0.051254938 | 0.001422636 | 3.741657387 | 9.055385138 | 3 | 2 | 0.666666667 | 7.247104812 | 0 | 0 | 0 | MH153800,MH153801,MK814756 |
| 2161 | VC_804_0 | 2 | 134.0553987 | 2264.409768 | 0.05589216 | 0.043092449 | 8.544003745 | 8.544003745 | 1 | 1 | 1 | 8.544003745 | 0 | 0 | 0 | MG711463,S1-wms-combined_304377 |
| 2162 | VC_804_1 | 2 | 91.07192788 | 2113.0944 | 0.041318083 | 0.046477822 | 8.485281374 | 8.485281374 | 1 | 1 | 1 | 8.485281374 | 0 | 0 | 0 | S17-wms-combined_259811,S17-wms-combined_47860 |
| 2163 | VC_804_2 | 6 | 873.586289 | 6651.485243 | 0.116090098 | 7.83E-10 | 7.211102551 | 9.38083152 | 15 | 13 | 0.866666667 | 8.174270944 | 0 | 0 | 0 | S1-wms_212940,S12-wms-combined_107942,S2-wms-combined_62377,S2-wms_248315,S8-wms_127191,S9-wms-combined_26410 |
| 2164 | VC_804_3 | 1 | 0 | 1436.678512 | 0 | 0 | 0 | 0 | 1 | 1 | 1 | 0 | 0 | 0 | 0 | S2-wms-combined_98888 |
| 2165 | VC_804_4 | 1 | 0 | 1261.655568 | 0 | 0 | 0 | 0 | 1 | 1 | 1 | 0 | 0 | 0 | 0 | S5-wms-combined_72410 |
| 2166 | VC_805_0 | 4 | 1434.451062 | 145.2511201 | 0.90805158 | 0.003773117 | 2 | 7.141428429 | 6 | 6 | 1 | 4.764998774 | 0 | 0 | 0 | EU100883,EU100884,KX987127,MH992131 |
| 2167 | VC_806_0 | 3 | 424.7185729 | 373.5634018 | 0.532040791 | 0.00175974 | 0 | 4.123105626 | 3 | 3 | 1 | 2.748737084 | 0 | 0 | 0 | KP792623,MW202557,MW202655 |
| 2168 | VC_807_0 | 17 | 14572.45305 | 1494.023497 | 0.907009885 | 6.93E-58 | 0 | 6.244997998 | 136 | 136 | 1 | 3.702430463 | 0 | 0 | 0 | EU124666,MG592549,MG592550,MG592581,MG592582,MK796797,MW824388,MW824389,MW824392,MW824393,MW824394,MW824398,MW824399,MW824402,MW824403,MW824405,OL762410 |

| | | | | | | | | | | | | | | | | |
|---|---|---|---|---|---|---|---|---|---|---|---|---|---|---|---|---|
| 2169 | VC_808_0 | 2 | 60.17217399 | 96.02497212 | 0.385232224 | 0.05234006 | 5.656854249 | 5.656854249 | 1 | 1 | 1 | 5.656854249 | 0 | 0 | 0 | MW202520,MW202674 |
| 2170 | VC_809_0 | 2 | 279.6474462 | 1274.688656 | 0.179914399 | 0.03700094 | 1.732050808 | 1.732050808 | 1 | 1 | 1 | 1.732050808 | 0 | 0 | 0 | KJ028219,MG770216 |
| 2171 | VC_80_0 | 41 | 203278.4193 | 145002.2244 | 0.583662695 | 0 | 0 | 10.58300524 | 820 | 804 | 0.980487805 | 5.065439501 | 0 | 0 | 0 | DQ163917,EU716414,KF981875,KR054028,KR054030,KT254130,KT254131,KT254132,KT254133,KT372690,KT372691,KT372692,KT372693,KT372694,KT372695,KT372696,KT372697,KT372698,LC472882,LN610589,MH536736,MK317959,MN508620,MN563783,MN564818,MN871454,MT108726,MT118293,MT118303,MT119364,MT119367,MT119370,MT119372,MT119374,MT119375,MT119377,MT119378,MT133563,MW557846,MZ089729,ON361134 |
| 2172 | VC_810_0 | 18 | 44600.26523 | 4295.853672 | 0.912143259 | 4.34E-52 | 0 | 2.449489743 | 153 | 153 | 1 | 0.797589782 | 0 | 0 | 0 | EU203571,KJ538722,KT222940,KT246485,KT454972,MF919517,MH697577,MH727549,MK494095,MK919482,MT114159,MT657330,MT684594,MT818421,MW924639,MZ005672,MZ622181,OP297533 |
| 2173 | VC_812_0 | 3 | 900 | 6767.990741 | 0.117371034 | 0.001576207 | 3.16227766 | 4.69041576 | 3 | 3 | 1 | 4.108276458 | 0 | 0 | 0 | HM997019,MG873442,MW296032 |
| 2174 | VC_812_1 | 4 | 1800 | 6144.275433 | 0.226578247 | 1.57E-05 | 3.16227766 | 4.582575695 | 6 | 6 | 1 | 3.828890753 | 0 | 0 | 0 | LC519601,MT028297,MT664722,MT740244 |
| 2175 | VC_812_2 | 2 | 300 | 4187.734869 | 0.066848869 | 0.044818616 | 1.732050808 | 1.732050808 | 1 | 1 | 1 | 1.732050808 | 0 | 0 | 0 | JN882286,MW343794 |
| 2176 | VC_812_3 | 1 | 0 | 2189.009008 | 0 | 0 | 0 | 0 | 1 | 1 | 1 | 0 | 0 | 0 | 0 | OL799327 |
| 2177 | VC_813_0 | 12 | 19800 | 64562.22281 | 0.234702208 | 4.84E-40 | 0 | 6.32455532 | 66 | 66 | 1 | 4.721547599 | 0 | 0 | 0 | JX415536,LC553734,MK310183,MT446407,MT446413,MT446414,MZ234015,MZ234017,MZ342906,MZ398243,OK148440,OP293233 |
| 2178 | VC_813_1 | 24 | 79583.03124 | 69161.90049 | 0.535030204 | 1.98E-126 | 3.605551275 | 8.246211251 | 276 | 276 | 1 | 6.138831822 | 0 | 0 | 0 | EU330206,JX867715,KF981730,KM044272,KP308307,LC553735,MG673519,MG748548,MG976803,MG983840,MK373770,MN045231,MN148435,MN508615,MN715356,MT127620,MT129653,MT129655,MT446386,MT446410,MW741821,MW929182,MZ234028,MZ648214 |
| 2179 | VC_814_0 | 3 | 228.8215809 | 0 | 1 | 0 | 0 | 4.242640687 | 3 | 3 | 1 | 2.828427125 | 0 | 0 | 0 | EU350361,KY296500,KY296501 |
| 2180 | VC_815_0 | 8 | 5394.352001 | 109.2936615 | 0.980141588 | 4.67E-12 | 0 | 4.898979486 | 28 | 28 | 1 | 3.698123975 | 0 | 0 | 0 | EU588980,GQ443085,JF767208,JF767209,JF767210,MH999280,MZ401006,MZ995505 |
| 2181 | VC_817_0 | 14 | 6440.473296 | 3714.630564 | 0.63421048 | 5.36E-45 | 0 | 4.472135955 | 91 | 91 | 1 | 2.504760607 | 0 | 0 | 0 | EU622808,EU771092,KU831549,MF407276,MT459794,MW354668,MW354670,MW477480,MZ570263,OL744111,OM249958,ON042750,PZACG,X99260 |
| 2182 | VC_818_0 | 20 | 18829.97421 | 782.514746 | 0.960101201 | 4.36E-43 | 0 | 5.656854249 | 190 | 190 | 1 | 4.129435499 | 0 | 0 | 0 | KC685370,KX349899,KX349900,KX349901,KX961631,KY947509,MF156577,MF156578,MF156580,MH817022,MK084630,MK284526,MK284527,MK284528,MN082624,MT777452,MW012634,MW348917,MZ384014,OL744112 |
| 2183 | VC_821_0 | 4 | 37.37090036 | 703.7524964 | 0.050424667 | 0.340348128 | 3.16227766 | 3.872983346 | 6 | 6 | 1 | 3.525304227 | 0 | 0 | 0 | S2-wms_53881,S7-wms_239833,S7-wms_53338,S8-wms_244525 |
| 2184 | VC_822_0 | 2 | 3.698386129 | 93.65553471 | 0.037989082 | 0.105107375 | 4 | 4 | 1 | 1 | 1 | 4 | 0 | 0 | 0 | LT600745,S7-wms-combined_225260 |
| 2185 | VC_823_0 | 124 | 592162.9285 | 31679.87725 | 0.949218173 | 0 | 0 | 8.485281374 | 7626 | 7626 | 1 | 6.663940833 | 0 | 0 | 0 | EU734173,GQ413937,HG994092,HM480846,KR149291,KT367885,KU183006,KX591654,KX712070,KX712071,KY389315,KY389316,KY450753,KY652723,KY652724,KY652725,LC413193,LC413194,LC413195,LN866626,MF285615,MG835568,MH172261,MH172262,MH172263,MH172264,MH587638,MK134560,MK203841,MK380015,MK380016,MK521905,MK618658,MK795385,MK903728,MN052874,MN101216,MN101218,MN101227,MN149903,MN176574,MN434093,MN585130,MT425185,MT446417,MT446418,MT542697,MT813140,MT813141,MT966872,MT966873,MW042787,MW042788,MW042789,MW042790,MW042791,MW042792,MW042793,MW042794,MW042795,MW042796,MW042797,MW042798,MW042799,MW042800,MW042801,MW042803,MW042804,MW042805,MW042806,MW042807,MW042808,MW042809,MW042810,MW042811,MW042812,MZ424865,MZ571832,OK138556,OL409034,OL799328,OM194188,OM194189,OM194190,ON148528,ON148529,ON571554,ON602742,ON602753,ON602755,ON880501,OP013026,OP045496,OP103917,OP103918,OP103919,OP103920,OP103921,OP103922,OP103923,OP256047,OU342754,OX335359,OX335360,OX335375,OX335376,OX335377,OX335389,OX335390,OX335392,OX335394,OX335398,OX335401,OX335408,OX335409,OX335413,OX335414,OX335418,OX335426,OX335427,OX335433,OX335438,OX335440,OX335443 |
| 2186 | VC_825_0 | 7 | 902.3512775 | 687.8379329 | 0.567449 | 1.58E-10 | 4.898979486 | 9.273618495 | 21 | 18 | 0.857142857 | 7.66483343 | 0 | 0 | 0 | S1-wms-combined_118167,S1-wms-combined_169879,S11-wms-combined_200084,S2-wms-combined_176376,S2-wms_79944,S6-wms-combined_140731,S9-wms-combined_105462 |
| 2187 | VC_825_1 | 1 | 0 | 351.9464578 | 0 | 0 | 0 | 0 | 1 | 1 | 1 | 0 | 0 | 0 | 0 | S11-wms-combined_267993 |
| 2188 | VC_825_2 | 1 | 0 | 171.5574266 | 0 | 0 | 0 | 0 | 1 | 1 | 1 | 0 | 0 | 0 | 0 | S5-wms-combined_134743 |
| 2189 | VC_826_0 | 91 | 648813.6754 | 4272.64629 | 0.993457762 | 1.80E-170 | 0 | 4.795831523 | 4095 | 4095 | 1 | 0.906742905 | 0 | 0 | 0 | ON170019,ON170022,ON170028,ON170033,ON170036,ON170038,ON170040,ON170049,ON170054,ON170074,ON170086,ON170088,ON170090,ON170092,ON170094,ON170096,ON170098,ON170100,ON170102,ON170104,ON170106,ON170108,ON170110,ON170112,ON170114,ON170116,ON170118,ON170120,ON170122,ON170124,ON170126,ON170142,ON170144,ON170146,ON170152,ON170154,ON170156,ON170158,ON170160,ON170162,ON170164,ON170167,ON170176,ON170180,ON170186,ON170189,ON170192,ON170194,ON170196,ON170198,ON170200,ON170202,ON170204,ON170206,ON170211,ON170215,ON170217,ON170219,ON170221,ON170222,ON170225,ON170244,ON170246,ON170248,ON170250,ON170252,ON170271,ON170280,ON170282,ON170284,ON170289,ON170291,ON170293,ON170295,ON170297,ON170298,ON170300,ON170306,ON170307,ON170311,ON170313,ON170315,ON170330,ON170332,ON170334,ON170336,ON170338,ON170340,ON170352,ON170356,ON170360 |
| 2190 | VC_827_0 | 6 | 2188.007623 | 732.1101405 | 0.74928746 | 3.97E-10 | 0 | 6.403124237 | 15 | 15 | 1 | 4.839038974 | 0 | 0 | 0 | KJ018213,MG592431,MG592432,MG592435,MG592458,MG592666 |
| 2191 | VC_828_0 | 3 | 491.9267258 | 544.0495258 | 0.474843632 | 0.001564446 | 2 | 3.741657387 | 3 | 3 | 1 | 3.068586334 | 0 | 0 | 0 | JQ619704,MG784342,MH598512 |
| 2192 | VC_829_0 | 5 | 1661.524261 | 221.9730378 | 0.88214847 | 1.68E-06 | 1 | 4.472135955 | 10 | 10 | 1 | 3.302470595 | 0 | 0 | 0 | EU887664,JX887877,KF296717,KX190834,MT136606 |
| 2193 | VC_82_0 | 24 | 47250.58365 | 17027.60803 | 0.735095099 | 1.62E-109 | 0 | 3.605551275 | 276 | 276 | 1 | 1.712322506 | 0 | 0 | 0 | DQ029335,FJ409640,HQ641340,HQ641341,HQ641342,HQ641343,HQ641344,JQ780163,KY883641,KY883643,KY883644,KY883645,KY883646,KY883648,KY883649,KY883650,KY883651,KY883652,KY883653,MF574151,MG957431,MT215167,ON464735,ON464736 |
| 2194 | VC_830_0 | 6 | 2978.428392 | 1578.192926 | 0.653648435 | 3.28E-09 | 2.645751311 | 4.358898944 | 15 | 15 | 1 | 3.653147576 | 0 | 0 | 0 | FJ008913,HE608841,MN078104,MT630433,MW314138,MW916539 |
| 2195 | VC_831_0 | 5 | 1296.59194 | 1671.327294 | 0.436869011 | 4.80E-07 | 4.242640687 | 6.164414003 | 10 | 10 | 1 | 5.220773115 | 0 | 0 | 0 | MN929097,OK632025,S1-wms-combined_394574,S4-wms-combined_14411,S6-wms_54578 |
| 2196 | VC_832_0 | 2 | 88.03814134 | 287.6297805 | 0.234350969 | 0.054738612 | 7.681145748 | 7.681145748 | 1 | 1 | 1 | 7.681145748 | 0 | 0 | 0 | S2-wms-combined_113619,S9-wms-combined_5526 |
| 2197 | VC_832_1 | 1 | 0 | 235.7872272 | 0 | 0 | 0 | 0 | 1 | 1 | 1 | 0 | 0 | 0 | 0 | S7-wms-combined_3697 |
| 2198 | VC_833_0 | 16 | 21402.09654 | 1484.576387 | 0.935133587 | 1.17E-65 | 0 | 7.615773106 | 120 | 120 | 1 | 6.071992627 | 0 | 0 | 0 | FJ591093,FJ591094,FR682616,FR719956,HQ317387,KJ621082,KJ803031,KU885988,KU885989,KU885990,MH015250,MH015253,MH015256,MH015257,MH015259,MZ773648 |
| 2199 | VC_834_0 | 7 | 261.7771128 | 4.120575994 | 0.984503152 | 0.057231067 | 0 | 3.16227766 | 21 | 21 | 1 | 2.381539227 | 0 | 0 | 0 | FJ685651,GU321093,JN882264,JN882266,KF056323,MW344764,MW344766 |
| 2200 | VC_835_0 | 67 | 330702.2629 | 12905.51484 | 0.962441145 | 0 | 0 | 6.403124237 | 2211 | 2211 | 1 | 4.99848368 | 0 | 0 | 0 | FJ750948,GQ502199,JN984867,JQ086375,JX865427,KM233151,KM896878,KX534335,KX534336,KX534338,KX534339,KX534341,LN881730,LN881731,LR597646,LR990702,LR999870,LT960609,MG065648,MG581355,MK138527,MK373789,MK373790,MK373791,MK728541,MN850575,MN850589,MN850628,MN850629,MN850650,MT331608,MT446389,MT446391,MT478993,MT884012,MT884013,MT884014,MT884015,MT951623,MW749005,MW822006,MW862804,MZ234008,MZ234014,MZ234016,MZ234026,MZ234029,MZ501070,MZ501088,MZ501095,MZ501097,MZ501107,OK272474,OK272488,OK310512,OK310513,OK310514,OK499985,OK499988,OK499993,OL615010,OL770278,OM262540,OM262541,ON184124,ON210136,OP046317 |
| 2201 | VC_836_0 | 9 | 5873.392843 | 1721.697178 | 0.773314448 | 3.12E-17 | 0 | 6.164414003 | 36 | 36 | 1 | 4.392069024 | 0 | 0 | 0 | KT367887,MN656993,MT075871,MW419910,OL539440,OM687892,ON602734,ON602747,ON602766 |
| 2202 | VC_837_0 | 29 | 70287.37118 | 317.8494158 | 0.995498217 | 5.79E-49 | 0 | 3.741657387 | 406 | 406 | 1 | 1.765107108 | 0 | 0 | 0 | FJ896200,LR794124,MF754111,MF754112,MK474470,MN974282,MT178448,MT501516,MW250641,MW331544,MZ020222,MZ182247,MZ516827,MZ592921,OK349506,OK585158,OK585159,OK585160,OM362522,OM799543,ON080940,ON080941,ON457554,ON457555,ON457556,ON457557,ON864052,ON872379,OP117450 |
| 2203 | VC_838_0 | 29 | 120231.2952 | 19390.83617 | 0.861119179 | 1.36E-130 | 0 | 5.291502622 | 406 | 406 | 1 | 2.260645968 | 0 | 0 | 0 | FN422399,HG962375,JX194238,KM411958,KM411959,KM411960,KM974184,KP994390,KR052142,KR054032,KU948710,KX171209,KX171210,MF490236,MF490238,MH356729,MN553583,MN553584,MN906996,MT080102,MZ955867,OK665488,OL519844,OM234792,OM937766,ON815901,ON815902,ON815903,ON857943 |
| 2204 | VC_839_0 | 5 | 3000 | 15187.25564 | 0.16495067 | 5.27E-08 | 1.414213562 | 4.123105626 | 10 | 10 | 1 | 3.168442053 | 0 | 0 | 0 | FN422398,LC064302,MT416090,OL519842,OM782452 |
| 2205 | VC_83_0 | 6 | 2461.82057 | 1363.45482 | 0.643566886 | 5.20E-11 | 1 | 4.242640687 | 15 | 15 | 1 | 3.221323242 | 0 | 0 | 0 | AB597179,KP343639,OK274245,OK318991,OP313111,OP617560 |
| 2206 | VC_841_0 | 10 | 12061.73826 | 8051.64278 | 0.599687255 | 1.20E-27 | 0 | 6.08276253 | 45 | 45 | 1 | 4.819756981 | 0 | 0 | 0 | KY083726,KY514264,KY514265,KY549659,KY769270,MH059638,MN812685,MN812687,MN812691,MZ348421 |
| 2207 | VC_842_0 | 2 | 275.6345664 | 4652.857405 | 0.055926756 | 0.044351492 | 3.741657387 | 3.741657387 | 1 | 1 | 1 | 3.741657387 | 0 | 0 | 0 | MG775260,MH179480 |
| 2208 | VC_843_0 | 5 | 3000 | 7553.916253 | 0.284254672 | 5.62E-08 | 0 | 7.211102551 | 10 | 10 | 1 | 5.86727123 | 0 | 0 | 0 | FN667788,FN667789,HE815464,MK408758,MT932329 |
| 2209 | VC_844_0 | 27 | 105300 | 6898.926948 | 0.938511649 | 1.26E-123 | 0 | 5.744562647 | 351 | 351 | 1 | 3.552029463 | 0 | 0 | 0 | FR823450,GU296433,JN132397,JX569801,KF148616,KX229736,KX236333,KX879627,MH107028,MT863714,MT863715,MT863716,MT863717,MT863718,MT863719,MT863720,MT863721,MT863722,MT863723,MT863724,MT863725,MT863726,MT863727,MT863728,MT863729,MT863730,MT863731 |
| 2210 | VC_845_0 | 3 | 900 | 1787.95544 | 0.334826979 | 0.001685027 | 4.123105626 | 6.403124237 | 3 | 3 | 1 | 5.563547955 | 0 | 0 | 0 | MK387337,MW451250,OM249648 |
| 2211 | VC_846_0 | 2 | 300 | 250.0040779 | 0.545450501 | 0.076424876 | 2.645751311 | 2.645751311 | 1 | 1 | 1 | 2.645751311 | 0 | 0 | 0 | MK388689,OK570186 |
| 2212 | VC_847_0 | 5 | 3000 | 45302.50553 | 0.062108579 | 0.000780631 | 0 | 6.32455532 | 10 | 10 | 1 | 4.197077977 | 0 | 0 | 0 | LC739533,LC739534,LC739535,LC739537,LC739539 |
| 2213 | VC_847_1 | 26 | 97500 | 56698.39464 | 0.632302303 | 1.33E-127 | 2.645751311 | 9.16515139 | 325 | 319 | 0.981538462 | 6.137217131 | 0 | 0 | 0 | JX561091,KX552041,KX664695,LR597654,LR746137,MG065650,MH051333,MH160766,MH252123,MK373788,MN850565,MN850571,MN850598,MN850618,MN850648,MT496970,MT944117,MW175890,MZ726793,OL505078,OL739525,OM362897,OM386656,OM386659,OM386667,OP562383 |
| 2214 | VC_848_0 | 28 | 113400 | 29654.77656 | 0.792703346 | 8.02E-191 | 0 | 6.92820323 | 378 | 378 | 1 | 5.016619389 | 0 | 0 | 0 | FR775895,KU522583,LC739536,LR865361,MF805809,MG065639,MG065645,MG065656,MG065689,MH675927,MH816848,MN850573,MN850584,MN850601,MN850632,MN850645,MT932211,MZ501063,MZ501084,MZ501100,OK210077,OK272470,OM386653,OM386654,OM386658,OM401812,OP177727,OX090893 |
| 2215 | VC_848_1 | 1 | 0 | 10018.61617 | 0 | 0 | 0 | 0 | 1 | 1 | 1 | 0 | 0 | 0 | 0 | OP617743 |
| 2216 | VC_849_0 | 3 | 900 | 12137.29383 | 0.069032731 | 0.001520602 | 5.830951895 | 7.681145748 | 3 | 3 | 1 | 7.020644026 | 0 | 0 | 0 | MH533020,ON287369,ON287374 |
| 2217 | VC_849_1 | 1 | 0 | 6544.136617 | 0 | 0 | 0 | 0 | 1 | 1 | 1 | 0 | 0 | 0 | 0 | MZ520832 |
| 2218 | VC_849_2 | 2 | 300 | 11900.64446 | 0.024588865 | 0.134862392 | 8.246211251 | 8.246211251 | 1 | 1 | 1 | 8.246211251 | 0 | 0 | 0 | JQ015307,KY514263 |
| 2219 | VC_849_3 | 2 | 300 | 12024.09481 | 0.024342559 | 0.148117714 | 2 | 2 | 1 | 1 | 1 | 2 | 0 | 0 | 0 | MF979560,MF979563 |
| 2220 | VC_849_4 | 2 | 300 | 12278.21411 | 0.023850763 | 0.135189255 | 4.242640687 | 4.242640687 | 1 | 1 | 1 | 4.242640687 | 0 | 0 | 0 | MT552976,MT560059 |
| 2221 | VC_849_5 | 3 | 900 | 17386.106 | 0.049217696 | 0.024252726 | 4.582575695 | 6.633249581 | 3 | 3 | 1 | 5.594529879 | 0 | 0 | 0 | JQ691612,KC954774,KT353109 |
| 2222 | VC_849_6 | 7 | 6300 | 32104.37033 | 0.16404383 | 1.40E-08 | 0 | 9.110433579 | 21 | 20 | 0.952380952 | 4.916218409 | 0 | 0 | 0 | JQ691611,MN013077,OL744211,OL744214,OL744216,OL744219,OL744220 |
| 2223 | VC_849_7 | 1 | 0 | 6546.979717 | 0 | 0 | 0 | 0 | 1 | 1 | 1 | 0 | 0 | 0 | 0 | ON712643 |
| 2224 | VC_849_8 | 1 | 0 | 6402.655988 | 0 | 0 | 0 | 0 | 1 | 1 | 1 | 0 | 0 | 0 | 0 | MW367417 |
| 2225 | VC_849_9 | 1 | 0 | 6604.110858 | 0 | 0 | 0 | 0 | 1 | 1 | 1 | 0 | 0 | 0 | 0 | MW250786 |
| 2226 | VC_850_0 | 3 | 900 | 9133.805035 | 0.08969678 | 0.001522067 | 0 | 1 | 3 | 3 | 1 | 0.666666667 | 0 | 0 | 0 | OK136181,OK181860,OK181862 |
| 2227 | VC_851_0 | 2 | 233.4908123 | 288.5897858 | 0.447231353 | 0.058957805 | 0 | 0 | 1 | 1 | 1 | 0 | 0 | 0 | 0 | MH444512,MK932884 |
| 2228 | VC_852_0 | 2 | 134.7280456 | 196.7969374 | 0.406388817 | 0.058101215 | 5.567764363 | 5.567764363 | 1 | 1 | 1 | 5.567764363 | 0 | 0 | 0 | GQ141189,S4-wms-combined_17284 |
| 2229 | VC_853_0 | 2 | 2.60778797 | 7.049965784 | 0.270020135 | 0.270145687 | 7.348469228 | 7.348469228 | 1 | 1 | 1 | 7.348469228 | 0 | 0 | 0 | MG941013,MW073100 |
| 2230 | VC_854_0 | 2 | 188.4475972 | 276.2068684 | 0.405565019 | 0.055317198 | 5.567764363 | 5.567764363 | 1 | 1 | 1 | 5.567764363 | 0 | 0 | 0 | S5-wms-combined_117213,S9-wms-combined_76274 |
| 2231 | VC_855_0 | 2 | 16.90845442 | 576.0555946 | 0.028515143 | 0.106210712 | 7.211102551 | 7.211102551 | 1 | 1 | 1 | 7.211102551 | 0 | 0 | 0 | GQ241246,KF669650 |
| 2232 | VC_856_0 | 7 | 6300 | 11892.15584 | 0.346303102 | 9.21E-14 | 2.449489743 | 4.123105626 | 21 | 21 | 1 | 3.448152834 | 0 | 0 | 0 | MH371108,MH371109,MH536823,MZ355724,MZ456035,ON260827,ON970603 |
| 2233 | VC_856_1 | 13 | 17124.3608 | 12155.85791 | 0.584844019 | 6.81E-37 | 0 | 8.94427191 | 78 | 78 | 1 | 6.791594401 | 0 | 0 | 0 | KJ173786,MH271300,MH271307,MH271310,MH271316,MH399779,MK376963,MK919474,MK937596,MN096374,MN908690,MZ622166,OP068333 |
| 2234 | VC_857_0 | 11 | 13551.06864 | 401.8774811 | 0.971197661 | 4.44E-25 | 0 | 4.582575695 | 55 | 55 | 1 | 2.131513 | 0 | 0 | 0 | KU160638,KU160639,KU160647,KU160663,KU160666,MF185731,MN234164,MN234217,MN234224,MT310874,MT986028 |
| 2235 | VC_859_0 | 9 | 9151.351568 | 556.8340387 | 0.942642831 | 3.93E-21 | 1 | 6.480740698 | 36 | 36 | 1 | 4.651904773 | 0 | 0 | 0 | MH019215,MH019216,MK305891,MT498037,MT498053,MT897908,MT936332,MZ648033,MZ648036 |
| 2236 | VC_85_0 | 3 | 900 | 20441.00269 | 0.042172339 | 0.001767581 | 1.414213562 | 7.745966692 | 3 | 3 | 1 | 5.591984454 | 0 | 0 | 0 | KJ010547,KJ010548,KM051843 |
| 2237 | VC_85_1 | 4 | 1800 | 33736.21231 | 0.050652556 | 3.31E-05 | 2 | 6.244997998 | 6 | 6 | 1 | 5.074713749 | 0 | 0 | 0 | MW419084,MW419085,MW419086,MW419087 |
| 2238 | VC_85_10 | 1 | 0 | 8516.537146 | 0 | 0 | 0 | 0 | 1 | 1 | 1 | 0 | 0 | 0 | 0 | MN043729 |
| 2239 | VC_85_11 | 1 | 0 | 8073.157121 | 0 | 0 | 0 | 0 | 1 | 1 | 1 | 0 | 0 | 0 | 0 | MN940411 |
| 2240 | VC_85_12 | 1 | 0 | 8893.936149 | 0 | 0 | 0 | 0 | 1 | 1 | 1 | 0 | 0 | 0 | 0 | MT151604 |
| 2241 | VC_85_13 | 1 | 0 | 6331.402046 | 0 | 0 | 0 | 0 | 1 | 1 | 1 | 0 | 0 | 0 | 0 | KU640380 |
| 2242 | VC_85_2 | 3 | 900 | 26028.53816 | 0.033421792 | 0.002697044 | 6 | 7.348469228 | 3 | 3 | 1 | 6.660572936 | 0 | 0 | 0 | KC699836,MW749002,MW749007 |
| 2243 | VC_85_3 | 19 | 51300 | 88428.33059 | 0.367141007 | 1.86E-72 | 0 | 10.19803903 | 171 | 138 | 0.807017544 | 7.827033601 | 0 | 0 | 0 | AB930182,AP013029,KF669652,LT960608,MF422185,MG000860,MW354667,MZ501260,MZ501261,MZ501262,MZ501263,MZ501265,OM982668,OM982669,OM982670,OM982671,OM982672,OM982673,OM982674 |
| 2244 | VC_85_4 | 3 | 900 | 29739.90445 | 0.02937346 | 0.018747447 | 4.358898944 | 6.633249581 | 3 | 3 | 1 | 5.745715508 | 0 | 0 | 0 | KY368640,MN043730,OK349510 |
| 2245 | VC_85_5 | 1 | 0 | 10667.8394 | 0 | 0 | 0 | 0 | 1 | 1 | 1 | 0 | 0 | 0 | 0 | MH606185 |
| 2246 | VC_85_6 | 1 | 0 | 7281.836584 | 0 | 0 | 0 | 0 | 1 | 1 | 1 | 0 | 0 | 0 | 0 | JX238501 |
| 2247 | VC_85_7 | 1 | 0 | 7209.823838 | 0 | 0 | 0 | 0 | 1 | 1 | 1 | 0 | 0 | 0 | 0 | KM236246 |
| 2248 | VC_85_8 | 1 | 0 | 7158.743652 | 0 | 0 | 0 | 0 | 1 | 1 | 1 | 0 | 0 | 0 | 0 | KU253712 |
| 2249 | VC_85_9 | 1 | 0 | 8029.002578 | 0 | 0 | 0 | 0 | 1 | 1 | 1 | 0 | 0 | 0 | 0 | KM236245 |
| 2250 | VC_860_0 | 16 | 22007.77886 | 78.94550051 | 0.996425658 | 1.56E-34 | 0 | 0 | 120 | 120 | 1 | 0 | 0 | 0 | 0 | MN023176,MN023177,MN023178,MN023179,MN023180,MN023181,MN023182,MN023183,MN023184,MN023185,MN023186,MN023187,MN023188,MN023189,MN023190,MN023191 |
| 2251 | VC_861_0 | 9 | 5111.33448 | 3598.075727 | 0.586874927 | 3.76E-19 | 0 | 2.449489743 | 36 | 36 | 1 | 1.779981331 | 0 | 0 | 0 | JQ659259,KC013021,KC013022,KC013023,KC013024,KC013025,KX555527,MN552146,OV696621 |
| 2252 | VC_862_0 | 11 | 7454.693584 | 3598.075727 | 0.674463872 | 5.04E-25 | 0 | 4.242640687 | 55 | 55 | 1 | 2.862872861 | 0 | 0 | 0 | GQ451696,KC013026,KC013027,KC013028,KC013029,KM262191,KM262192,KX578042,KX578043,KX578044,MN552147 |
| 2253 | VC_864_0 | 2 | 1.001946165 | 0 | 1 | 0 | 8.660254038 | 8.660254038 | 1 | 1 | 1 | 8.660254038 | 0 | 0 | 0 | GQ919031,MN234216 |
| 2254 | VC_865_0 | 2 | 300 | 3616.860453 | 0.076591955 | 0.061155602 | 7.615773106 | 7.615773106 | 1 | 1 | 1 | 7.615773106 | 0 | 0 | 0 | MK804893,MK813939 |
| 2255 | VC_865_1 | 1 | 0 | 2218.581146 | 0 | 0 | 0 | 0 | 1 | 1 | 1 | 0 | 0 | 0 | 0 | MK813938 |
| 2256 | VC_865_2 | 1 | 0 | 2283.114753 | 0 | 0 | 0 | 0 | 1 | 1 | 1 | 0 | 0 | 0 | 0 | MK813941 |
| 2257 | VC_865_3 | 2 | 300 | 3708.914878 | 0.074833218 | 0.065309387 | 2 | 2 | 1 | 1 | 1 | 2 | 0 | 0 | 0 | OM033134,OM033135 |
| 2258 | VC_865_4 | 1 | 0 | 2062.491807 | 0 | 0 | 0 | 0 | 1 | 1 | 1 | 0 | 0 | 0 | 0 | OM033136 |
| 2259 | VC_866_0 | 4 | 1796.549689 | 20807.86997 | 0.079477806 | 1.41E-05 | 2 | 3.16227766 | 6 | 6 | 1 | 2.682841463 | 0 | 0 | 0 | JN035618,KR063278,MW534374,ON526973 |
| 2260 | VC_866_1 | 20 | 56219.05479 | 60569.74447 | 0.481373686 | 1.12E-107 | 0 | 3.741657387 | 190 | 190 | 1 | 2.388197052 | 0 | 0 | 0 | KU998240,MG099935,MH025889,MK450428,MK450430,MK450435,MK450436,MK494106,MK494109,MK494121,MK878903,MK919477,MK919481,ON724015,ON970582,ON970588,ON970596,ON970607,OP297547,OP434450 |
| 2261 | VC_866_2 | 3 | 857.8383138 | 17778.32709 | 0.046030838 | 0.001495893 | 0 | 4.898979486 | 3 | 3 | 1 | 3.265986324 | 0 | 0 | 0 | KR011062,KR011063,KR011064 |
| 2262 | VC_867_0 | 6 | 2391.512762 | 5076.559507 | 0.320231604 | 2.36E-11 | 1.414213562 | 8.544003745 | 15 | 15 | 1 | 6.668070979 | 0 | 0 | 0 | KP790008,KP790009,KP790010,KR063279,KX557281,ON526975 |
| 2263 | VC_868_0 | 25 | 87159.07037 | 63348.60413 | 0.579100505 | 1.26E-159 | 0 | 6.557438524 | 300 | 300 | 1 | 4.585911655 | 0 | 0 | 0 | KU963245,KU998241,MG099940,MG757152,MG757155,MG770211,MG770214,MK359302,MK433261,MK433263,MK433265,MK433267,MK801728,MK801733,MK967390,MN444874,MN945905,MT114160,MT889368,MT889370,MT889379,MT986029,MW507132,MW507133,ON456331 |
| 2264 | VC_869_0 | 9 | 10652.68933 | 29946.36197 | 0.262387642 | 3.38E-24 | 0 | 4.123105626 | 36 | 36 | 1 | 2.674366436 | 0 | 0 | 0 | KU963258,KU963262,KU998242,KU998243,MH576976,MK494098,MN234166,OM203156,OP434448 |
| 2265 | VC_870_0 | 5 | 3000 | 4074.851555 | 0.424037166 | 3.48E-08 | 0 | 3.872983346 | 10 | 10 | 1 | 2.589970548 | 0 | 0 | 0 | KU998252,KU998253,KU998254,MH479923,MN096363 |
| 2266 | VC_872_0 | 31 | 92286.42497 | 20930.86923 | 0.815126572 | 1.06E-180 | 0 | 9.486832981 | 465 | 381 | 0.819354839 | 5.544227595 | 0 | 0 | 0 | KX670789,KX670790,MF467948,MF467949,MG757154,MH155877,MH155880,MH669006,MK433259,MK433260,MK433268,MK433269,MK967386,MN204492,MN204494,MN204495,MN204496,MN204497,MN204500,MN204504,MN586054,MN586056,MT310852,MT310854,MT657340,MT684589,MT684600,MW055905,MW145539,MW712736,ON108646 |
| 2267 | VC_873_0 | 3 | 171.0361318 | 16.77012196 | 0.9107052 | 0.01623666 | 4.898979486 | 6.164414003 | 3 | 3 | 1 | 5.631448461 | 0 | 0 | 0 | GU936715,HM480106,MG271909 |
| 2268 | VC_874_0 | 3 | 510.649276 | 300.807572 | 0.629299361 | 0.001875173 | 0 | 1 | 3 | 3 | 1 | 0.666666667 | 0 | 0 | 0 | KR935216,KR935217,KT253150 |
| 2269 | VC_875_0 | 10 | 874.1649441 | 10.64489206 | 0.987969288 | 0.000515666 | 0 | 3.16227766 | 45 | 45 | 1 | 1.949558002 | 0 | 0 | 0 | GU942563,HQ224500,KJ619459,KP972568,KP972569,KR063267,KR063268,KT728930,KT728931,MF155889 |
| 2270 | VC_876_0 | 3 | 295.8563793 | 1521.595185 | 0.16278639 | 0.002533688 | 5.291502622 | 9.055385138 | 3 | 2 | 0.666666667 | 7.448962587 | 0 | 0 | 0 | S12-wms-combined_245755,S5-wms-combined_26971,S7-wms-combined_139683 |
| 2271 | VC_876_1 | 1 | 0 | 774.6655457 | 0 | 0 | 0 | 0 | 1 | 1 | 1 | 0 | 0 | 0 | 0 | S2-wms-combined_116221 |
| 2272 | VC_877_0 | 2 | 282.570576 | 81.45767269 | 0.776232551 | 0.131776239 | 0 | 0 | 1 | 1 | 1 | 0 | 0 | 0 | 0 | MN830254,MN830255 |
| 2273 | VC_878_0 | 32 | 40879.10866 | 145.3465115 | 0.996457076 | 1.18E-21 | 0 | 7.416198487 | 496 | 496 | 1 | 5.845477352 | 0 | 0 | 0 | HE956707,HE956710,JN651747,KC107834,KT184661,KT949345,KX756572,KY318515,LN878149,MF683623,MG878892,MH179473,MH992513,MK053931,MK330684,MK838107,MK838108,MK838109,MK838110,MK838111,MN509793,MT682386,MT682387,MT682388,MT682389,MT682390,MT682391,MW250276,MZ592920,OL362270,OM654404,ON287377 |
| 2274 | VC_879_0 | 36 | 130335.6355 | 785.5607976 | 0.994008895 | 1.45E-62 | 0 | 3 | 630 | 630 | 1 | 1.282631085 | 0 | 0 | 0 | HF569088,HF569089,HF569090,HF569091,HF569092,HG428758,JN939331,JN939332,KC556893,KC556894,KC556895,KC556896,KC556897,KC556898,KJ133688,KJ133689,KJ133690,KJ133691,KJ133692,KJ133693,KJ133694,KJ133695,KJ133696,KJ133697,KJ133698,KJ133699,KJ133700,KJ133701,KJ133702,KJ133703,KJ133704,KJ133705,KJ133706,KJ133707,KY056619,MT002975 |
| 2275 | VC_87_0 | 8 | 8400 | 6749.295291 | 0.554481237 | 2.37E-15 | 3.605551275 | 6.633249581 | 28 | 28 | 1 | 5.157793624 | 0 | 0 | 0 | FJ230960,KF669649,KY368639,MT601270,MT601271,MW001214,MZ751040,OM236513 |
| 2276 | VC_87_1 | 3 | 900 | 4601.790606 | 0.163583107 | 0.002220758 | 2.645751311 | 4.898979486 | 3 | 3 | 1 | 4.113520773 | 0 | 0 | 0 | KC595512,KC595513,KF554508 |
| 2277 | VC_87_2 | 1 | 0 | 1790.257906 | 0 | 0 | 0 | 0 | 1 | 1 | 1 | 0 | 0 | 0 | 0 | MH884511 |
| 2278 | VC_880_0 | 2 | 6.494897698 | 33.74507264 | 0.161404137 | 0.165449273 | 2.828427125 | 2.828427125 | 1 | 1 | 1 | 2.828427125 | 0 | 0 | 0 | HG007973,HM150760 |
| 2279 | VC_881_0 | 2 | 116.2925148 | 55.20414358 | 0.678103678 | 0.081867177 | 4.795831523 | 4.795831523 | 1 | 1 | 1 | 4.795831523 | 0 | 0 | 0 | KP282679,KP282680 |
| 2280 | VC_882_0 | 20 | 56996.48288 | 477.5613956 | 0.991690834 | 3.68E-42 | 0 | 5 | 190 | 190 | 1 | 3.471682614 | 0 | 0 | 0 | HG992588,JQ692107,JX556418,MF754113,MF754114,MF754115,MF754116,MG602476,MG893203,MH599087,MH925090,MN215888,MW451246,MW451251,MZ592922,OK483201,OK625546,OL314400,OP547477,OP595601 |
| 2281 | VC_883_0 | 10 | 13500 | 53499.9303 | 0.201492747 | 6.62E-30 | 2 | 8.366600265 | 45 | 45 | 1 | 6.93940589 | 0 | 0 | 0 | JF966203,KJ489397,KJ489398,KJ489400,KU737346,MF288921,MF288922,MF498901,MH638310,MK215646 |
| 2282 | VC_883_1 | 12 | 19800 | 61315.56817 | 0.244096176 | 1.58E-29 | 0 | 8.888194417 | 66 | 66 | 1 | 7.231172083 | 0 | 0 | 0 | KF208639,KF669647,KJ489402,KT307976,KU737345,KU737347,MF765814,MN038178,MW281503,OK499976,OK499990,OM350011 |
| 2283 | VC_883_2 | 2 | 300 | 16244.54916 | 0.01813286 | 0.119051476 | 1 | 1 | 1 | 1 | 1 | 1 | 0 | 0 | 0 | JN790865,JN797796 |
| 2284 | VC_883_3 | 2 | 300 | 16178.89877 | 0.0182051 | 0.120997643 | 6.08276253 | 6.08276253 | 1 | 1 | 1 | 6.08276253 | 0 | 0 | 0 | KF669662,KY888882 |
| 2285 | VC_883_4 | 1 | 0 | 8334.787781 | 0 | 0 | 0 | 0 | 1 | 1 | 1 | 0 | 0 | 0 | 0 | MN153503 |
| 2286 | VC_883_5 | 1 | 0 | 8326.763175 | 0 | 0 | 0 | 0 | 1 | 1 | 1 | 0 | 0 | 0 | 0 | MW787012 |
| 2287 | VC_883_6 | 1 | 0 | 8412.098617 | 0 | 0 | 0 | 0 | 1 | 1 | 1 | 0 | 0 | 0 | 0 | OM743306 |
| 2288 | VC_884_0 | 28 | 113400 | 69177.15993 | 0.621107263 | 5.79E-66 | 0 | 9.273618495 | 378 | 372 | 0.984126984 | 5.8583255 | 0 | 0 | 0 | KJ489399,KJ489401,KT207918,KU737344,KU737348,KU737349,KU737350,KU737351,KU737352,KX147229,KX156152,KX349902,KX349903,KX961629,KX961632,MF288917,MF288918,MF288919,MF288920,MG763894,MH538193,MH638311,MN038176,MN038179,MT584805,OK500001,OM350012,OM350013 |
| 2289 | VC_884_1 | 8 | 8400 | 67805.95899 | 0.1102276 | 0.001067699 | 2.236067977 | 9.797958971 | 28 | 21 | 0.75 | 6.416757534 | 0 | 0 | 0 | HM144387,JN654439,KC430106,KU847400,KX011169,MW392803,MW584228,MW596414 |
| 2290 | VC_885_0 | 11 | 12218.33395 | 282.7890493 | 0.977378908 | 4.89E-26 | 1.414213562 | 4.123105626 | 55 | 55 | 1 | 2.609763728 | 0 | 0 | 0 | MW677509,MW677510,MW677512,MW677515,MW677517,MW677520,OP009272,OP009275,OP009278,OP009280,OP009286 |
| 2291 | VC_886_0 | 2 | 254.9815929 | 1599.035974 | 0.137529222 | 0.046476646 | 5.567764363 | 5.567764363 | 1 | 1 | 1 | 5.567764363 | 0 | 0 | 0 | MN988459,MN988558 |
| 2292 | VC_886_1 | 1 | 0 | 947.1266082 | 0 | 0 | 0 | 0 | 1 | 1 | 1 | 0 | 0 | 0 | 0 | MW677529 |
| 2293 | VC_887_0 | 18 | 45806.16967 | 79246.03598 | 0.366296375 | 1.26E-88 | 1.732050808 | 5.830951895 | 153 | 153 | 1 | 4.110901757 | 0 | 0 | 0 | HM152763,JF704108,JF704113,JN600672,MF185726,MH744425,MK279905,MK310144,MK494092,MK494100,MN096382,MN586018,MN586023,MN617841,MT771349,MW435855,MW822149,MZ958746 |
| 2294 | VC_888_0 | 28 | 113369.9368 | 92859.56387 | 0.549727059 | 2.16E-189 | 0 | 6.708203932 | 378 | 378 | 1 | 3.953489101 | 0 | 0 | 0 | JF744988,JN680858,KC661276,KC691254,KF024731,KT591489,KU234099,KU997639,KX507361,KX580961,KX580962,MF140422,MF185728,MF185730,MF324910,MH779511,MH834600,MK937600,MN096380,MN234228,MN586030,MN586052,MN703406,MN703410,MT758688,MW578835,MZ171373,OL455899 |
| 2295 | VC_888_1 | 3 | 841.2723336 | 28264.9645 | 0.028903507 | 0.001515524 | 5.656854249 | 6.92820323 | 3 | 3 | 1 | 6.406102353 | 0 | 0 | 0 | MH576973,MN234187,MZ005679 |
| 2296 | VC_889_0 | 16 | 35864.16096 | 84927.65251 | 0.296908871 | 2.50E-71 | 0 | 6.403124237 | 120 | 120 | 1 | 4.331197159 | 0 | 0 | 0 | KF024725,KT281791,KT281794,KT372003,MF140405,MF140413,MF140414,MF140421,MH727559,MK878900,MN096377,MT889383,MT889391,MW291024,MZ622183,ON755181 |
| 2297 | VC_88_0 | 1 | 0 | 98.74618067 | 0 | 0 | 0 | 0 | 1 | 1 | 1 | 0 | 0 | 0 | 0 | KT624200 |
| 2298 | VC_88_1 | 1 | 0 | 166.3995877 | 0 | 0 | 0 | 0 | 1 | 1 | 1 | 0 | 0 | 0 | 0 | MF418016 |
| 2299 | VC_890_0 | 3 | 891.4359569 | 1608.753319 | 0.356547388 | 0.001690487 | 4.472135955 | 5.744562647 | 3 | 3 | 1 | 5.137617008 | 0 | 0 | 0 | MH651182,MK967378,MT658805 |
| 2300 | VC_890_1 | 4 | 1800 | 2101.850195 | 0.461319607 | 1.54E-05 | 2 | 6.164414003 | 6 | 6 | 1 | 4.342890308 | 0 | 0 | 0 | MH976515,MN444870,MT684599,OK040786 |
| 2301 | VC_891_0 | 2 | 195.93564 | 145.0672348 | 0.574586476 | 0.077364462 | 2.828427125 | 2.828427125 | 1 | 1 | 1 | 2.828427125 | 0 | 0 | 0 | HQ268735,KY629621 |
| 2302 | VC_893_0 | 2 | 34.39247974 | 164.4716374 | 0.172944623 | 0.061538965 | 5.656854249 | 5.656854249 | 1 | 1 | 1 | 5.656854249 | 0 | 0 | 0 | S11-wms-combined_46816,S4-wms-combined_113721 |

| | | | | | | | | | | | | | | | |
|---|---|---|---|---|---|---|---|---|---|---|---|---|---|---|---|
| 2303 | VC_894_0 | 3 | 481.1329333 | 275.5962867 | 0.63580594 | 0.003078484 | 2 | 5.099019514 | 3 | 3 | 1 | 3.999333 | 0 | 0 | 0 HM588722,KF302032,KF302033 |
| 2304 | VC_895_0 | 2 | 94.49741148 | 244.4261373 | 0.278816305 | 0.057487462 | 6.32455532 | 6.32455532 | 1 | 1 | 1 | 6.32455532 | 0 | 0 | 0 KF302036,KX365748 |
| 2305 | VC_896_0 | 5 | 809.90674 | 815.9494359 | 0.498141688 | 2.26E-07 | 4.898979486 | 7 | 10 | 10 | 1 | 6.423050228 | 0 | 0 | 0 KR060090,KX912252,KY379511,MF370965,MT002875 |
| 2306 | VC_897_0 | 6 | 890.1520339 | 1251.605661 | 0.415617526 | 3.89E-10 | 1 | 3.464101615 | 15 | 15 | 1 | 2.258077258 | 0 | 0 | 0 HM640230,JN800508,KY996523,MH999279,MW815121,OP022426 |
| 2307 | VC_898_0 | 10 | 3856.088318 | 1437.859218 | 0.728395643 | 3.32E-22 | 0 | 3.464101615 | 45 | 45 | 1 | 2.214779374 | 0 | 0 | 0 JQ729990,JQ729991,JQ729992,MH248069,MW057920,MZ401005,MZ401007,MZ401008,MZ401009,OL547738 |
| 2308 | VC_899_0 | 4 | 349.1158081 | 965.6244404 | 0.265539759 | 2.90E-05 | 0 | 4.123105626 | 6 | 6 | 1 | 2.697074804 | 0 | 0 | 0 MH393889,MK017820,OM638104,OP256049 |
| 2309 | VC_89_0 | 2 | 300 | 6355.773225 | 0.045073651 | 0.071505289 | 5.656854249 | 5.656854249 | 1 | 1 | 1 | 5.656854249 | 0 | 0 | 0 MT601273,MT601274 |
| 2310 | VC_89_1 | 2 | 300 | 6408.57464 | 0.044718888 | 0.053531097 | 6.633249581 | 6.633249581 | 1 | 1 | 1 | 6.633249581 | 0 | 0 | 0 MT601272,OL580764 |
| 2311 | VC_89_2 | 5 | 3000 | 11756.77214 | 0.203296491 | 3.57E-06 | 1.414213562 | 7 | 10 | 10 | 1 | 4.833658103 | 0 | 0 | 0 KY030782,MT366945,MT366946,MT366947,MT366948 |
| 2312 | VC_89_3 | 1 | 0 | 3619.233123 | 0 | 0 | 0 | 0 | 1 | 1 | 1 | 0 | 0 | 0 | 0 AF020713 |
| 2313 | VC_89_4 | 1 | 0 | 3430.559108 | 0 | 0 | 0 | 0 | 1 | 1 | 1 | 0 | 0 | 0 | 0 ON107264 |
| 2314 | VC_89_5 | 1 | 0 | 3338.672051 | 0 | 0 | 0 | 0 | 1 | 1 | 1 | 0 | 0 | 0 | 0 MZ969646 |
| 2315 | VC_89_6 | 1 | 0 | 2911.32742 | 0 | 0 | 0 | 0 | 1 | 1 | 1 | 0 | 0 | 0 | 0 OM236515 |
| 2316 | VC_8_0 | 58 | 213381.62 | 43508.58969 | 0.830633523 | 0 | 0 | 9.273618495 | 1653 | 1644 | 0.994555354 | 6.674113308 | 0 | 0 | 0 AB045978,AF424782,AP009363,AP012341,AY954955,AY954956,AY954957,CP062434,CP062442,CP062444,CP062461,EF462198,EU861005,JQ779023,JQ957932,JX174275,KF598856,KT186243,KY056620,LC541428,MF580410,MG029509,MG029511,MG029513,MG029514,MG029515,MG029516,MG029517,MG029518,MG770897,MH384261,MK348510,MK801680,MK801681,MK940809,MT366568,MT899504,MW924497,OL690335,OL840290,OM304856,OM439665,OM439667,ON856257,ON856258,ON989481,OP038318,OP038319,OP136131,OP136132,OP136134,OP328420,OP328422,OP374154,OP374155,OP374156,OP584903,OP584904 |
| 2317 | VC_901_0 | 5 | 2264.4853 | 179.5809188 | 0.92652371 | 5.03E-06 | 0 | 6.92820323 | 10 | 10 | 1 | 5.202732593 | 0 | 0 | 0 KF623293,MW824400,MW824417,MW824421,MW824431 |
| 2318 | VC_902_0 | 3 | 900 | 2.027099769 | 0.997752729 | 0.124106539 | 0 | 7 | 3 | 3 | 1 | 4.666666667 | 0 | 0 | 0 HQ316582,MG592662,MG592672 |
| 2319 | VC_903_0 | 7 | 3617.529445 | 1286.438099 | 0.737674019 | 7.07E-13 | 0 | 5.477225575 | 21 | 21 | 1 | 3.175463184 | 0 | 0 | 0 MW824414,MW824420,MW824427,MW824428,MW824429,MW865319,OX241434 |
| 2320 | VC_904_0 | 6 | 3965.497854 | 13054.78931 | 0.232986542 | 1.50E-10 | 0 | 8.124038405 | 15 | 15 | 1 | 6.254827105 | 0 | 0 | 0 JF713456,MG592585,MG592586,MG720309,MK559459,MT735629 |
| 2321 | VC_904_1 | 4 | 1800 | 10316.52946 | 0.148557391 | 2.28E-05 | 0 | 4.582575695 | 6 | 6 | 1 | 3.177976178 | 0 | 0 | 0 HQ316604,KX198615,MT451873,ON015836 |
| 2322 | VC_904_2 | 7 | 5573.038396 | 13932.76392 | 0.285711826 | 3.35E-13 | 0 | 7.280109889 | 21 | 21 | 1 | 3.702864892 | 0 | 0 | 0 KY022433,MH925091,MN871506,MW411580,OK483199,OP547478,OV032860 |
| 2323 | VC_905_0 | 3 | 900 | 1728.865254 | 0.342353036 | 0.001762247 | 3 | 3.605551275 | 3 | 3 | 1 | 3.255942979 | 0 | 0 | 0 MG189906,MT110073,MZ326862 |
| 2324 | VC_907_0 | 3 | 661.5756116 | 334.457848 | 0.664210228 | 0.003668864 | 3 | 5 | 3 | 3 | 1 | 4 | 0 | 0 | 0 HQ317384,HQ332142,KM233261 |
| 2325 | VC_908_0 | 3 | 620.8108253 | 600.4506209 | 0.508335727 | 0.002754703 | 0 | 6.708203932 | 3 | 3 | 1 | 4.472135955 | 0 | 0 | 0 MN988533,MN988538,MW980068 |
| 2326 | VC_909_0 | 5 | 1544.119256 | 540.7655917 | 0.740625679 | 1.39E-05 | 3.872983346 | 4.795831523 | 10 | 10 | 1 | 4.326114384 | 0 | 0 | 0 HQ317386,MG592522,MG640035,MT720689,OM837731 |
| 2327 | VC_90_0 | 1 | 0 | 209.2356595 | 0 | 0 | 0 | 0 | 1 | 1 | 1 | 0 | 0 | 0 | 0 LC597490 |
| 2328 | VC_90_1 | 1 | 0 | 319.5678008 | 0 | 0 | 0 | 0 | 1 | 1 | 1 | 0 | 0 | 0 | 0 OM634661 |
| 2329 | VC_90_2 | 1 | 0 | 115.6778985 | 0 | 0 | 0 | 0 | 1 | 1 | 1 | 0 | 0 | 0 | 0 EF583821 |
| 2330 | VC_910_0 | 2 | 92.94972114 | 1.019761903 | 0.989147946 | 0.5 | 8.306623863 | 8.306623863 | 1 | 1 | 1 | 8.306623863 | 0 | 0 | 0 HQ317390,MZ568826 |
| 2331 | VC_911_0 | 6 | 3701.968809 | 9581.535428 | 0.278689173 | 1.51E-11 | 0 | 4.358898944 | 15 | 15 | 1 | 1.829714663 | 0 | 0 | 0 HQ317392,KC821610,KC821616,KC821629,KC821630,KC821634 |
| 2332 | VC_912_0 | 76 | 708837.691 | 15200.22476 | 0.979006314 | 2.10E-297 | 0 | 6.08276253 | 2850 | 2850 | 1 | 3.006768721 | 0 | 0 | 0 KM873719,KY951963,KY951964,KY979235,KY979236,KY979237,KY979238,KY979239,KY979240,KY979241,KY979242,KY979243,KY979244,KY979245,KY979246,KY979247,KY992519,KY992520,MK756083,MK756084,MK756085,MK756086,MK756087,MK756088,MK756089,MK756090,MK756091,MK756092,MK756093,MK756094,MK756095,MK756096,MT431535,MT585273,MT585274,MT585275,MT585276,MT585277,MT585278,MT585279,MT585280,MT585281,MT585282,MT585283,MT585284,MT585285,MT585286,MT585287,MT585288,MT585289,MT585290,MT585291,MT585292,MT585293,MT585294,MT585295,MT585296,MT585297,MT585298,MT585299,MT585300,MT585301,MT585302,MT585303,MT585304,MT585305,MT585306,MT585307,MT585308,MT585309,MT585310,MT585311,MT585312,MT585313,MT585314,MT585315 |
| 2333 | VC_913_0 | 2 | 3.484199298 | 29.95716172 | 0.104188322 | 0.176932247 | 6.244997998 | 6.244997998 | 1 | 1 | 1 | 6.244997998 | 0 | 0 | 0 HQ332136,MW452941 |
| 2334 | VC_914_0 | 1 | 0 | 29.72877948 | 0 | 0 | 0 | 0 | 1 | 1 | 1 | 0 | 0 | 0 | 0 S5-wms_67169 |
| 2335 | VC_915_0 | 3 | 134.954144 | 473.2973843 | 0.221872265 | 0.001806511 | 6.557438524 | 7.483314774 | 3 | 3 | 1 | 7.106954396 | 0 | 0 | 0 S12-wms-combined_321174,S3-wms_23958,S8-wms-combined_169835 |
| 2336 | VC_916_0 | 4 | 1477.245847 | 266.3523491 | 0.847239834 | 0.00014426 | 0 | 5 | 6 | 6 | 1 | 2.816496581 | 0 | 0 | 0 HQ396194,MZ995504,ON922917,ON933637 |
| 2337 | VC_917_0 | 2 | 199.4292273 | 464.5320311 | 0.300362747 | 0.05145877 | 5.744562647 | 5.744562647 | 1 | 1 | 1 | 5.744562647 | 0 | 0 | 0 S11-wms-combined_48246,S8-wms-combined_270362 |
| 2338 | VC_917_1 | 1 | 0 | 261.8601647 | 0 | 0 | 0 | 0 | 1 | 1 | 1 | 0 | 0 | 0 | 0 S5-wms-combined_90315 |
| 2339 | VC_918_0 | 2 | 63.55645036 | 385.7879135 | 0.141442634 | 0.071233377 | 6.633249581 | 6.633249581 | 1 | 1 | 1 | 6.633249581 | 0 | 0 | 0 S11-wms_320555,S12-wms_12658 |
| 2340 | VC_918_1 | 1 | 0 | 363.2801541 | 0 | 0 | 0 | 0 | 1 | 1 | 1 | 0 | 0 | 0 | 0 S7-wms-combined_26217 |
| 2341 | VC_918_2 | 1 | 0 | 322.874506 | 0 | 0 | 0 | 0 | 1 | 1 | 1 | 0 | 0 | 0 | 0 S12-wms-combined_201125 |
| 2342 | VC_919_0 | 39 | 100299.6999 | 10298.62874 | 0.906882601 | 7.09E-169 | 0 | 8.306623863 | 741 | 741 | 1 | 6.305444185 | 0 | 0 | 0 HQ403646,KU963260,KU998239,MG770215,MH271302,MH450132,MK279900,MK875791,MK875795,MK967383,MK967385,MN062716,MN234215,MN284906,MN310549,MN444871,MN444878,MN586015,MN586036,MN586049,MN586057,MT776808,MT776809,MT776810,MT889367,MT889378,MW507135,MW862988,MZ150793,MZ322013,MZ622167,OK040785,OL455888,OL829977,ON970576,ON970599,ON970609,ON970621,OP297531 |
| 2343 | VC_91_0 | 22 | 69300 | 61019.14218 | 0.531771456 | 3.97E-111 | 0 | 8.426149773 | 231 | 231 | 1 | 6.254413275 | 0 | 0 | 0 AB609718,AP009390,AP018714,AP018715,CAJDJY010000002,CAJDZD010000002,KJ801817,KR049063,LC596377,LC596378,LC596379,LR031359,MK268686,MK693030,MK721192,MN854830,MW633168,MZ420150,ON113167,ON113168,ON113169,ON286976 |
| 2344 | VC_91_1 | 1 | 0 | 7235.329762 | 0 | 0 | 0 | 0 | 1 | 1 | 1 | 0 | 0 | 0 | 0 OP559178 |
| 2345 | VC_920_0 | 2 | 300 | 1495.267577 | 0.167106009 | 0.044437036 | 3 | 3 | 1 | 1 | 1 | 3 | 0 | 0 | 0 MN204493,MW822144 |
| 2346 | VC_921_0 | 2 | 204.0635968 | 11.19622322 | 0.947987399 | 0.270145687 | 0 | 0 | 1 | 1 | 1 | 0 | 0 | 0 | 0 HQ632857,HQ632858 |
| 2347 | VC_922_0 | 2 | 36.72054079 | 50.70014174 | 0.420044087 | 0.095215132 | 6.480740698 | 6.480740698 | 1 | 1 | 1 | 6.480740698 | 0 | 0 | 0 MK291441,MK291443 |
| 2348 | VC_923_0 | 23 | 73209.94788 | 4217.256885 | 0.945532621 | 1.58E-92 | 1 | 6.403124237 | 253 | 253 | 1 | 3.997941658 | 0 | 0 | 0 MW677513,MW677516,MW677518,MW677519,MW677521,MW677522,MW677523,MW677524,OP009265,OP009266,OP009267,OP009268,OP009269,OP009270,OP009271,OP009273,OP009276,OP009277,OP009279,OP009283,OP009284,OP009287,OP009288 |
| 2349 | VC_924_0 | 34 | 53696.4037 | 6355.746279 | 0.894162885 | 5.31E-162 | 0 | 9.16515139 | 561 | 548 | 0.976827094 | 7.592178037 | 0 | 0 | 0 MG592442,MG592450,MG592457,MG592460,MG592482,MG592483,MG592488,MG592489,MG592490,MG592502,MG592503,MG592504,MG592505,MG592516,MG592517,MG592525,MG592530,MG592532,MG592534,MG592535,MG592539,MG592543,MG592557,MG592558,MG592559,MG592576,MG592593,MG592594,MG592597,MG592598,MG592605,MG592606,MG592607,MG592631 |
| 2350 | VC_925_0 | 1 | 0 | 98.13986357 | 0 | 0 | 0 | 0 | 1 | 1 | 1 | 0 | 0 | 0 | 0 MN592896 |
| 2351 | VC_926_0 | 3 | 524.6968328 | 30.84262168 | 0.944481672 | 0.005696139 | 0 | 5.385164807 | 3 | 3 | 1 | 3.590109871 | 0 | 0 | 0 MG592520,OX241566,OX241570 |
| 2352 | VC_927_0 | 2 | 1.929920333 | 20.09416263 | 0.087627727 | 0.280637918 | 8.602325267 | 8.602325267 | 1 | 1 | 1 | 8.602325267 | 0 | 0 | 0 HQ698895,KJ410740 |
| 2353 | VC_928_0 | 3 | 700.6199726 | 192.609007 | 0.784367714 | 0.005586199 | 0 | 1 | 3 | 3 | 1 | 0.666666667 | 0 | 0 | 0 JF974286,JF974321,MT490303 |
| 2354 | VC_929_0 | 4 | 1800 | 327.5743819 | 0.846033876 | 0.002777992 | 0 | 3.16227766 | 6 | 6 | 1 | 1.58113883 | 0 | 0 | 0 HQ634192,KC821604,KC821621,KC821625 |
| 2355 | VC_92_0 | 22 | 69300 | 53583.28562 | 0.563949765 | 7.06E-119 | 2.645751311 | 8.717797887 | 231 | 231 | 1 | 6.80538555 | 0 | 0 | 0 CAJDKA010000002,KP339049,KY549443,LR990835,MH791397,MH880817,MN027503,MN241318,MT856905,MT909815,MW004544,MW004545,MZ147816,MZ333462,OL799258,OL799260,OL870611,OM966901,ON113170,ON125307,ON506927,ON506928 |
| 2356 | VC_930_0 | 7 | 5720.530485 | 5300.767828 | 0.519043249 | 8.33E-15 | 0 | 6.557438524 | 21 | 21 | 1 | 3.647790413 | 0 | 0 | 0 KX889068,MG592409,MG592410,MG592470,MG592519,MG592521,MG592668 |
| 2357 | VC_932_0 | 2 | 17.75718474 | 22.25367723 | 0.443809102 | 0.105649774 | 7.937253933 | 7.937253933 | 1 | 1 | 1 | 7.937253933 | 0 | 0 | 0 HQ683709,MG209611 |
| 2358 | VC_933_0 | 8 | 2536.194403 | 4416.315356 | 0.364788327 | 5.85E-18 | 0 | 7 | 28 | 28 | 1 | 5.73451774 | 0 | 0 | 0 KR093625,KR093626,KR093627,KR093628,KR093629,KR093630,KR093631,KR093633 |
| 2359 | VC_934_0 | 2 | 258.2134949 | 41.57361848 | 0.861322863 | 0.075007761 | 0 | 0 | 1 | 1 | 1 | 0 | 0 | 0 | 0 MT119766,MT210154 |
| 2360 | VC_935_0 | 2 | 16.67520639 | 6.881059614 | 0.707888355 | 0.270145687 | 6.708203932 | 6.708203932 | 1 | 1 | 1 | 6.708203932 | 0 | 0 | 0 MN988536,OP028995 |
| 2361 | VC_936_0 | 2 | 300 | 23.13842396 | 0.928394699 | 0.185546685 | 1.732050808 | 1.732050808 | 1 | 1 | 1 | 1.732050808 | 0 | 0 | 0 MN602266,MN602267 |
| 2362 | VC_937_0 | 40 | 141603.9082 | 66357.10443 | 0.68091565 | 0 | 0 | 10.19803903 | 780 | 730 | 0.935897436 | 6.445293508 | 0 | 0 | 0 JN672684,KR269719,KX237515,KY780482,MN013075,MN013076,MN013078,MN013079,MN013080,MN013083,MN013087,MT701592,MW021762,MW021764,MW021765,MW042802,MW417503,MW722081,MZ428221,MZ428224,MZ598515,MZ634348,MZ634350,MZ634351,OL539450,OL539472,OL865411,OM065837,OM065838,OM065839,OM065841,OM328157,OM735690,OP611406,OV877339,OV877387,OV877766,OX335361,OX335395,OX335410 |
| 2363 | VC_938_0 | 99 | 753703.7546 | 72857.93473 | 0.911854208 | 0 | 0 | 9.746794345 | 4851 | 4816 | 0.992784993 | 6.942443969 | 0 | 0 | 0 JF501022,KP658157,KR262148,KT001920,KX575286,LR757892,MF415412,MF612072,MG552615,MG894052,MH445453,MH633484,MH633485,MH633486,MH633487,MH688453,MH844531,MK170446,MK278861,MK521904,MK618657,MK931442,MK931443,MK931444,MK931445,MN379832,MN395285,MN395286,MN689779,MN783016,MT894005,MW655991,MW672037,MZ172460,MZ221764,MZ398242,MZ428222,MZ428223,MZ428225,MZ428226,MZ428227,MZ428228,MZ571831,MZ571833,MZ571834,MZ634345,OK138555,OK655936,OL539448,OL539460,OL674249,OL702938,OL744210,OL744213,OM032871,OM065840,OM065842,OM065843,OM687893,OM938991,OM938992,OM964875,ON489264,ON602735,ON602741,ON602750,ON881905,OP267563,OP413832,OP617742,OU342755,OW251746,OX335369,OX335370,OX335371,OX335380,OX335381,OX335383,OX335384,OX335385,OX335386,OX335387,OX335388,OX335391,OX335393,OX335400,OX335404,OX335411,OX335412,OX335417,OX335421,OX335422,OX335423,OX335429,OX335430,OX335432,OX335435,OX335439 |
| 2364 | VC_939_0 | 15 | 31500 | 0 | 1 | 1 | 0 | 4 | 105 | 105 | 1 | 2.82037923 | 0 | 0 | 0 JF704100,KJ538721,KX159477,MG757167,MH727553,MH727562,MK061407,MK279842,MK305892,MK494123,MN329674,MN329676,MN428056,MN428061,ON637772 |
| 2365 | VC_93_0 | 2 | 300 | 2522.512373 | 0.106288285 | 0.043343284 | 4.69041576 | 4.69041576 | 1 | 1 | 1 | 4.69041576 | 0 | 0 | 0 FJ822135,MT758689 |
| 2366 | VC_940_0 | 34 | 124426.6099 | 31914.44734 | 0.795866499 | 4.34E-174 | 0 | 6.708203932 | 561 | 561 | 1 | 4.801789614 | 0 | 0 | 0 JF704107,JN083852,JN083853,JN408459,JQ684677,JX042578,KF017001,KF493882,KF560330,KM101118,KT004677,KT246486,KT438501,MG099934,MG099937,MG099938,MG099944,MG099949,MH020239,MH051250,MH051254,MH051256,MH338235,MH338241,MK305887,MK494091,MN096372,MN586016,MN586060,MN586062,MN617844,OL455889,ON970625,ON970626 |
| 2367 | VC_941_0 | 40 | 134790.4737 | 60485.30736 | 0.690256994 | 0 | 0 | 8.366600265 | 780 | 780 | 1 | 6.24999193 | 0 | 0 | 0 KC710998,MG065636,MG065640,MG065641,MG065643,MG065649,MG065657,MG065663,MG065665,MG065667,MG065669,MG065670,MG065671,MG065673,MG065675,MG065679,MG065680,MG065681,MG065687,MK373798,MN055691,MN432485,MN850567,MN850569,MN850572,MN850591,MN850600,MN850602,MN850607,MN850640,MT074433,MT127619,MT360681,MZ501079,MZ501090,OK272479,OK272487,OL539459,ON185589,ON528718 |
| 2368 | VC_942_0 | 36 | 149921.4328 | 41101.33966 | 0.784835394 | 0 | 0 | 7.810249676 | 630 | 630 | 1 | 4.77516521 | 0 | 0 | 0 LR027385,LR027388,LR596614,LT841304,LT961732,MF564201,MK962751,MN781674,MN850582,MN850586,MN850596,MN850604,MN850606,MN850613,MN850620,MN850634,MN850638,MN850641,MN850643,MZ234033,MZ501069,MZ501101,OK272471,OK272478,OK272481,OK272483,OK272485,OK272486,OK272489,OL539445,OL539446,ON185586,ON185587,ON185588,ON711395,OU734268 |
| 2369 | VC_943_0 | 2 | 117.8948035 | 0 | 1 | 0 | 0 | 0 | 1 | 1 | 1 | 0 | 0 | 0 | 0 JF917095,JQ010983 |
| 2370 | VC_944_0 | 6 | 2047.827568 | 450.8353524 | 0.819569359 | 3.24E-10 | 0 | 8.062257748 | 15 | 15 | 1 | 6.453563135 | 0 | 0 | 0 MG592398,MG592496,MG592497,MG592584,MG592612,MG592613 |
| 2371 | VC_946_0 | 11 | 14643.69742 | 5272.956054 | 0.735248893 | 5.87E-32 | 0 | 5.291502622 | 55 | 55 | 1 | 3.218661496 | 0 | 0 | 0 JX483876,JX483877,JX483878,MN988482,MN988500,MN988501,MN988508,MN988530,MN988545,MN988549,MW980073 |
| 2372 | VC_947_0 | 3 | 18.34590779 | 472.5482563 | 0.03737243 | 0.154083248 | 8.062257748 | 8.94427191 | 3 | 3 | 1 | 8.47755981 | 0 | 0 | 0 S2-wms-combined_135154,S9-wms-combined_53991,S9-wms-combined_89249 |
| 2373 | VC_948_0 | 9 | 4605.209468 | 3855.415011 | 0.544310822 | 6.34E-23 | 2.828427125 | 8.246211251 | 36 | 36 | 1 | 6.813621841 | 0 | 0 | 0 MG592429,MG592438,MG592487,MG592566,MG592654,MG592657,MG592658,MG592659,MZ369152 |
| 2374 | VC_949_0 | 10 | 1561.773596 | 1051.267046 | 0.597684388 | 3.67E-26 | 0 | 5.385164807 | 45 | 45 | 1 | 4.103675598 | 0 | 0 | 0 JN116822,KR053197,KR053198,KU998255,KU998256,MG812496,MH271296,MK660712,MK801723,MN234213 |
| 2375 | VC_94_0 | 2 | 300 | 2000.22131 | 0.130422233 | 0.042768664 | 5 | 5 | 1 | 1 | 1 | 5 | 0 | 0 | 0 HM242243,OM135992 |
| 2376 | VC_950_0 | 38 | 61189.70439 | 3610.240361 | 0.944286367 | 3.80E-160 | 0 | 3.31662479 | 703 | 703 | 1 | 0.700224354 | 0 | 0 | 0 KT355471,KT355473,KT355475,KT783672,KU160655,KU160657,KU160658,KU160667,KU160670,KU160674,KX443695,KX576642,KX610765,KY434670,MF140409,MF140415,MF140417,MF140419,MF140420,MF140426,MF140429,MF140431,MF140433,MH576948,MH576950,MH576951,MH576952,MH576955,MH576957,MH576961,MH779506,MK494103,MK494111,MK737943,MK820640,MN617842,MN813695,ON970580 |
| 2377 | VC_951_0 | 3 | 170.828764 | 1457.377717 | 0.104918366 | 0.000781363 | 0 | 2.645751311 | 3 | 3 | 1 | 1.763834207 | 0 | 0 | 0 MH834594,MH834597,MH834628 |
| 2378 | VC_952_0 | 2 | 217.1396412 | 10.08773978 | 0.955605087 | 0.113899997 | 3.31662479 | 3.31662479 | 1 | 1 | 1 | 3.31662479 | 0 | 0 | 0 JN116828,MW654182 |
| 2379 | VC_953_0 | 2 | 37.37261764 | 139.236576 | 0.21161196 | 0.051791431 | 8.246211251 | 8.246211251 | 1 | 1 | 1 | 8.246211251 | 0 | 0 | 0 JN116823,MK967393 |
| 2380 | VC_954_0 | 13 | 19765.2031 | 3137.044472 | 0.863024602 | 5.85E-34 | 0 | 3.16227766 | 78 | 78 | 1 | 2.102148219 | 0 | 0 | 0 KU963261,MH399781,MH509446,MK279893,MK305885,MK359319,MK376958,MK967392,MW291015,MW534376,MZ005677,ON637762,ON637763 |
| 2381 | VC_955_0 | 3 | 900 | 262.0556666 | 0.774489576 | 0.008653781 | 3.31662479 | 3.741657387 | 3 | 3 | 1 | 3.458302322 | 0 | 0 | 0 MZ322021,ON970578,OP297535 |
| 2382 | VC_956_0 | 39 | 161105.8903 | 0 | 1 | 0 | 0 | 4.898979486 | 741 | 741 | 1 | 2.348671665 | 0 | 0 | 0 JN116826,JN116827,KT372002,KT375356,KT959213,KT990218,KX550082,KX611788,KX712236,KX712237,KY549153,KY549154,MF189179,MF324898,MF324900,MF324901,MF324902,MF324903,MF324904,MF324905,MF373842,MF537628,MH271291,MH271293,MH271294,MH271297,MH271299,MH271306,MH271309,MH271311,MH271314,MH271315,MH271319,MH316569,MK524487,MK524495,MN945902,MW291027,MZ402608 |
| 2383 | VC_957_0 | 1 | 0 | 48.27881381 | 0 | 0 | 0 | 0 | 1 | 1 | 1 | 0 | 0 | 0 | 0 JN175269 |
| 2384 | VC_957_1 | 1 | 0 | 239.9820837 | 0 | 0 | 0 | 0 | 1 | 1 | 1 | 0 | 0 | 0 | 0 KU862660 |
| 2385 | VC_958_0 | 3 | 472.9777625 | 2810.343293 | 0.14405468 | 0.001519856 | 4.582575695 | 5.477225575 | 3 | 3 | 1 | 5.085317898 | 0 | 0 | 0 MW570842,MW584170,MW584191 |
| 2386 | VC_959_0 | 5 | 3000 | 57139.48282 | 0.049884034 | 7.69E-06 | 1.732050808 | 6 | 10 | 10 | 1 | 4.673294096 | 0 | 0 | 0 JX094500,KX397367,KX397372,MK514282,MW845759 |
| 2387 | VC_959_1 | 8 | 8400 | 87314.22511 | 0.08776125 | 0.015649005 | 0 | 5.656854249 | 28 | 28 | 1 | 3.902897941 | 0 | 0 | 0 KX397368,KX397370,KX397371,MT710307,MW384823,MW812340,MW812342,MW812343 |
| 2388 | VC_959_2 | 5 | 3000 | 58998.97861 | 0.048387894 | 0.106535744 | 1.414213562 | 7.549834435 | 10 | 10 | 1 | 6.37000768 | 0 | 0 | 0 KX397365,KX397366,MW760841,MW812338,MW812341 |
| 2389 | VC_959_3 | 3 | 900 | 37125.02258 | 0.023668625 | 0.240884522 | 3.31662479 | 5.916079783 | 3 | 3 | 1 | 4.963186274 | 0 | 0 | 0 KX397369,MH426724,MW845758 |
| 2390 | VC_959_4 | 3 | 900 | 37124.50086 | 0.02366895 | 0.228611835 | 3 | 5.830951895 | 3 | 3 | 1 | 4.738705567 | 0 | 0 | 0 JX316028,KX397364,KX397373 |
| 2391 | VC_959_5 | 19 | 51300 | 144057.1605 | 0.262595954 | 9.71E-12 | 0 | 8.185352772 | 171 | 171 | 1 | 4.285256603 | 0 | 0 | 0 JN641803,KU726251,LR535902,LR535909,LR535910,LR535914,LR535915,LR535916,LR535918,LR535919,LR535920,LR535921,MH221129,MN445185,MT679221,MT799840,MW812339,OK428535,OM456492 |
| 2392 | VC_959_6 | 1 | 0 | 12511.73976 | 0 | 0 | 0 | 0 | 1 | 1 | 1 | 0 | 0 | 0 | 0 MH221128 |
| 2393 | VC_95_0 | 3 | 900 | 3402.167684 | 0.209196867 | 0.001474233 | 5 | 7.810249676 | 3 | 3 | 1 | 6.808674261 | 0 | 0 | 0 MK504443,MK504446,OM293948 |
| 2394 | VC_960_0 | 1 | 0 | 10.05535834 | 0 | 0 | 0 | 0 | 1 | 1 | 1 | 0 | 0 | 0 | 0 JN712910 |
| 2395 | VC_960_1 | 1 | 0 | 10.05535834 | 0 | 0 | 0 | 0 | 1 | 1 | 1 | 0 | 0 | 0 | 0 KX925554 |
| 2396 | VC_961_0 | 3 | 106.4086542 | 51.23281036 | 0.675004222 | 0.009451772 | 2.236067977 | 3.31662479 | 3 | 3 | 1 | 2.904990143 | 0 | 0 | 0 KX344510,KY264020,MW344767 |
| 2397 | VC_962_0 | 2 | 23.76754747 | 42.7767567 | 0.357168773 | 0.077364462 | 2 | 2 | 1 | 1 | 1 | 2 | 0 | 0 | 0 JN882267,OM621814 |
| 2398 | VC_963_0 | 4 | 228.149705 | 40.10860974 | 0.85048512 | 0.001207324 | 1 | 3.741657387 | 6 | 6 | 1 | 2.468349637 | 0 | 0 | 0 KY965934,MG550110,MG550111,MG550113 |
| 2399 | VC_965_0 | 2 | 217.0759579 | 684.4014729 | 0.240800214 | 0.043100688 | 2.236067977 | 2.236067977 | 1 | 1 | 1 | 2.236067977 | 0 | 0 | 0 MG596800,OM982619 |
| 2400 | VC_966_0 | 3 | 554.985136 | 1008.96586 | 0.35486095 | 0.001596675 | 2.828427125 | 3.872983346 | 3 | 3 | 1 | 3.339345087 | 0 | 0 | 0 MK608336,OK499995,OL539452 |
| 2401 | VC_967_0 | 16 | 10058.69829 | 1154.346242 | 0.897053272 | 2.90E-29 | 0 | 3.16227766 | 120 | 120 | 1 | 2.324485898 | 0 | 0 | 0 JQ309827,KT932700,LT630001,MK721183,MK721184,MK721185,MK721188,MK721193,MK721195,MK721196,MK721197,MK721198,MT747434,MT757399,MW623430,ON352054 |
| 2402 | VC_968_0 | 4 | 485.128389 | 1154.346242 | 0.295904786 | 2.36E-05 | 0 | 1.414213562 | 6 | 6 | 1 | 0.707106781 | 0 | 0 | 0 KT945995,MK360025,OL799259,ON131181 |
| 2403 | VC_969_0 | 3 | 817.6386765 | 165.7605334 | 0.831441258 | 0.002371493 | 4.69041576 | 6.244997998 | 3 | 3 | 1 | 5.440192855 | 0 | 0 | 0 JQ312117,MK637516,MT234341 |
| 2404 | VC_96_0 | 30 | 27741.43008 | 10893.04648 | 0.718048555 | 1.98E-151 | 0 | 2.645751311 | 435 | 435 | 1 | 1.647346546 | 0 | 0 | 0 AB626962,AB626963,AF513032,AF513033,AY954949,EU136189,HF937074,KJ210330,KM366100,KU992911,KY000084,KY000085,LC709172,MG766218,MG766219,MH107118,MH972260,MK764384,MK903033,MK922546,MK922547,MK922548,MK936475,MK936476,MN098325,MN150710,MT926124,MW286254,MW363799,MZ359091 |
| 2405 | VC_970_0 | 9 | 10690.56724 | 347.8252975 | 0.968489497 | 1.75E-17 | 0 | 8 | 36 | 36 | 1 | 5.505736802 | 0 | 0 | 0 MN988470,MN988484,MN988495,MN988531,MN988537,MN988544,MN988546,MN988557,MW980069 |
| 2406 | VC_971_0 | 3 | 900 | 15520.97344 | 0.054807957 | 0.001610862 | 0 | 4.69041576 | 3 | 3 | 1 | 3.12694384 | 0 | 0 | 0 KP280063,KY883654,KY883655 |
| 2407 | VC_971_1 | 24 | 82800 | 29191.73012 | 0.739340306 | 6.62E-124 | 1 | 9.486832981 | 276 | 246 | 0.891304348 | 6.606302619 | 0 | 0 | 0 JQ340389,LC487410,MG649966,MG649967,MK796244,MK895508,MK907780,MN954399,MN958086,MN966730,MN966731,MN966732,MT448616,MT448617,MT459144,MT460514,MT460515,MT460516,MT460517,MT460518,MW344054,MW344055,MW344056,MW398869 |
| 2408 | VC_971_2 | 1 | 0 | 6336.528846 | 0 | 0 | 0 | 0 | 1 | 1 | 1 | 0 | 0 | 0 | 0 KT345706 |
| 2409 | VC_972_0 | 6 | 4500 | 11062.7367 | 0.289152229 | 3.92E-11 | 1 | 5.291502622 | 15 | 15 | 1 | 3.495571797 | 0 | 0 | 0 MH645904,MH925094,MN871505,MN871510,MN871511,MT442039 |
| 2410 | VC_973_0 | 8 | 8400 | 9394.801359 | 0.472047978 | 9.07E-19 | 0 | 5.196152423 | 28 | 28 | 1 | 2.026288589 | 0 | 0 | 0 MW824415,MW824416,MW824419,MW824422,MW824423,MW824424,MW824425,MW824426 |
| 2411 | VC_974_0 | 2 | 187.0839253 | 0 | 1 | 0 | 2.449489743 | 2.449489743 | 1 | 1 | 1 | 2.449489743 | 0 | 0 | 0 JQ823122,JQ823123 |
| 2412 | VC_975_0 | 11 | 11563.78002 | 373.4405153 | 0.968716293 | 4.21E-12 | 0 | 6.164414003 | 55 | 55 | 1 | 3.295094572 | 0 | 0 | 0 KP876465,KP876466,KT989433,KU189325,KU530220,KY365739,MF975638,MH178381,MH178382,MK686071,ON526979 |
| 2413 | VC_976_0 | 2 | 300 | 9507.483407 | 0.030588887 | 0.072506347 | 4.123105626 | 4.123105626 | 1 | 1 | 1 | 4.123105626 | 0 | 0 | 0 ON529850,ON529857 |
| 2414 | VC_976_1 | 5 | 3000 | 19588.48624 | 0.132811025 | 2.92E-07 | 4.123105626 | 9 | 10 | 9 | 0.9 | 7.135035013 | 0 | 0 | 0 ON529851,ON529852,ON529853,ON529855,ON529860 |
| 2415 | VC_976_2 | 2 | 300 | 10478.7144 | 0.027832633 | 0.045892061 | 5.477225575 | 5.477225575 | 1 | 1 | 1 | 5.477225575 | 0 | 0 | 0 JX100810,MH588545 |
| 2416 | VC_976_3 | 2 | 300 | 11321.42634 | 0.025814387 | 0.115971081 | 5.099019514 | 5.099019514 | 1 | 1 | 1 | 5.099019514 | 0 | 0 | 0 JX100814,MH588544 |
| 2417 | VC_976_4 | 11 | 16500 | 32544.68409 | 0.336427898 | 1.46E-28 | 0 | 6.32455532 | 55 | 55 | 1 | 4.140975138 | 0 | 0 | 0 JX100809,JX100811,JX100812,JX100813,JX163858,KY555142,KY555143,KY555144,KY555145,KY555146,KY555147 |
| 2418 | VC_976_5 | 1 | 0 | 5377.822146 | 0 | 0 | 0 | 0 | 1 | 1 | 1 | 0 | 0 | 0 | 0 ON529858 |
| 2419 | VC_977_0 | 2 | 300 | 3423.122803 | 0.080577519 | 0.060796812 | 2.236067977 | 2.236067977 | 1 | 1 | 1 | 2.236067977 | 0 | 0 | 0 KT870145,MH015255 |
| 2420 | VC_977_1 | 2 | 300 | 3496.813473 | 0.079013626 | 0.060228139 | 5.477225575 | 5.477225575 | 1 | 1 | 1 | 5.477225575 | 0 | 0 | 0 MN602268,MT270409 |
| 2421 | VC_977_2 | 1 | 0 | 2042.548992 | 0 | 0 | 0 | 0 | 1 | 1 | 1 | 0 | 0 | 0 | 0 OP186293 |
| 2422 | VC_978_0 | 2 | 150.5403256 | 2379.365428 | 0.059504322 | 0.045886287 | 8.774964387 | 8.774964387 | 1 | 1 | 1 | 8.774964387 | 0 | 0 | 0 KY629563,OL955261 |
| 2423 | VC_978_1 | 1 | 0 | 1388.167082 | 0 | 0 | 0 | 0 | 1 | 1 | 1 | 0 | 0 | 0 | 0 OL963731 |
| 2424 | VC_97_0 | 5 | 111.5900918 | 1491.210386 | 0.069621948 | 0.018861652 | 2.645751311 | 4.123105626 | 10 | 10 | 1 | 3.445671806 | 0 | 0 | 0 AY212251,KJ528544,LC628082,LC644557,ON637248 |
| 2425 | VC_980_0 | 10 | 13500 | 5.297151533 | 0.999607772 | 1.00E-11 | 0 | 4.242640687 | 45 | 45 | 1 | 3.350118396 | 0 | 0 | 0 JX126919,JX126920,JX442241,KJ094020,KJ094021,MH299806,MN114082,MN114083,MN128593,MN904502 |
| 2426 | VC_981_0 | 3 | 715.2495115 | 12748.06306 | 0.053125819 | 0.001463146 | 1.414213562 | 4.123105626 | 3 | 3 | 1 | 3.136767511 | 0 | 0 | 0 JX182369,ON526970,ON526974 |
| 2427 | VC_981_1 | 26 | 61511.05997 | 26248.4928 | 0.700904438 | 2.42E-181 | 1 | 8.124038405 | 325 | 325 | 1 | 6.076673646 | 0 | 0 | 0 JX182370,JX262376,MF766044,MF766045,MF766046,MF766047,MF766048,MG593802,MH001460,MH590601,MH651190,MH669004,MH669016,MK392363,MK392364,MK392365,MK392366,MK460245,MK524524,MN234192,MN234208,MT310863,MT701594,MT701597,MT708548,OK412919 |
| 2428 | VC_982_0 | 62 | 252374.1182 | 39523.50113 | 0.86459807 | 0 | 0 | 9.949874371 | 1891 | 1405 | 0.742993125 | 7.000905116 | 0 | 0 | 0 JX889246,KC700556,KC700557,KC700558,KT124227,KT124228,KT124229,KT152029,KT184390,KT184391,KT186228,KT186229,KX344445,KX507343,KX507344,KX507345,KX815338,MF541403,MF541404,MF541405,MF541406,MF541407,MF541408,MF541409,MF541410,MG298964,MG757163,MH171093,MH171094,MH171095,MH171096,MH171097,MH171098,MH248947,MK433270,MK433271,MK433276,MK433278,MN096373,MN096375,MN096379,MN096381,MN204498,MN284899,MN586009,MT701598,MT711978,MT711979,MW365951,MW365953,MW435856,MZ648032,MZ648039,MZ648040,MZ820088,MZ820091,MZ820092,OL455891,OL455894,OL742563,ON208832,ON526985 |
| 2429 | VC_984_0 | 38 | 103257.469 | 2561.789282 | 0.975790897 | 1.54E-28 | 0 | 7.348469228 | 703 | 703 | 1 | 4.553077461 | 0 | 0 | 0 JX290549,KR604693,KT240186,KX278418,KX278419,KY953156,LC063634,LC375533,MH807811,MH807814,MH807816,MH807818,MH807819,MK095192,MK095193,MK095194,MK095195,MK095196,MK095197,MK095198,MK095199,MK095200,MK095201,MK095203,MK095204,MK095205,MK095206,MK095207,MK095208,MK095209,MK095210,MK095211,MN270887,MN270888,MN271656,MN812690,MN812692 |
| 2430 | VC_987_0 | 22 | 21989.08812 | 2564.710178 | 0.895547314 | 6.66E-57 | 0 | 8.366600265 | 231 | 231 | 1 | 6.0284453 | 0 | 0 | 0 JX483874,JX483875,JX483879,JX483880,MF403005,MF403006,MN988466,MN988488,MN988492,MN988493,MN988512,MN988513,MN988514,MN988528,MN988529,MT708544,MT708546,MW960034,MW980062,MW980065,MW980074,MZ462995 |
| 2431 | VC_988_0 | 2 | 161.3318315 | 855.7179851 | 0.158627266 | 0.04731066 | 2.645751311 | 2.645751311 | 1 | 1 | 1 | 2.645751311 | 0 | 0 | 0 MF278336,MW043865 |
| 2432 | VC_989_0 | 2 | 106.3185658 | 776.1615822 | 0.120477006 | 0.047487814 | 3.741657387 | 3.741657387 | 1 | 1 | 1 | 3.741657387 | 0 | 0 | 0 KY065149,MT227925 |
| 2433 | VC_990_0 | 3 | 900 | 514.4810762 | 0.636275745 | 0.001804384 | 2 | 6.92820323 | 3 | 3 | 1 | 5.187150937 | 0 | 0 | 0 KF614509,MN988552,MT778840 |
| 2434 | VC_991_0 | 2 | 90.17782025 | 149.8211652 | 0.375742506 | 0.085815228 | 4.358898944 | 4.358898944 | 1 | 1 | 1 | 4.358898944 | 0 | 0 | 0 KC292024,KC292028 |
| 2435 | VC_992_0 | 8 | 302.3992671 | 478.3551084 | 0.387316775 | 6.08E-16 | 0 | 1.732050808 | 28 | 28 | 1 | 0.935429417 | 0 | 0 | 0 KP087954,MH572347,MH572348,MH572349,MH572405,MH572418,MH572455,MH572515 |

| | | | | | | | | | | | | | | | | |
|---|---|---|---|---|---|---|---|---|---|---|---|---|---|---|---|---|
| 2436 | VC_994_0 | 1 | 0 | 27.21090003 | 0 | 0 | 0 | 0 | 1 | 1 | | 1 | 0 | 0 | 0 | 0 MK249135 |
| 2437 | VC_995_0 | 69 | 17880.25327 | 3040.131553 | 0.854680897 | 1.19E-10 | 0 | 2.645751311 | 2346 | 2346 | | 1 | 1.833810674 | 0 | 0 | 0 KC131022,KM589502,KM589511,KT264813,KT264814,KT264815,MG945202,MG945465,MG945477,MG945663,MG945838,MG945843,MH572283,MH572284,MH572285,MH572286,MH572291,MH572292,MH572293,MH572382,MH572383,MH572423,MH572424,MH572427,MH572428,MH572466,MH572495,MH572504,MH992156,MH992161,MH992184,MH992185,MH992213,MK249141,MK249147,MK249165,MK249185,MK249190,MK249197,MK249201,MK249202,MK249203,MK249211,MK249213,MK249221,MK496825,MN862333,MN862335,MN862341,MW202412,MW202414,MW202466,MW202471,MW202549,MW202587,MW202847,MW202853,MW202883,MZ364228,MZ364233,MZ364240,MZ364243,MZ364256,MZ364258,MZ364259,MZ364267,MZ364274,OL617029,OM523002 |
| 2438 | VC_996_0 | 2 | 4.09143279 | 92.95456729 | 0.042159726 | 0.40961847 | 2 | 2 | 1 | 1 | | 1 | 2 | 0 | 0 | 0 MN862339,MN862340 |
| 2439 | VC_997_0 | 1 | 0 | 266.922191 | 0 | 0 | 0 | 0 | 1 | 1 | | 1 | 0 | 0 | 0 | 0 MH992169 |
| 2440 | VC_998_0 | 2 | 9.796278594 | 180.3650543 | 0.051515618 | 0.060981907 | 1.414213562 | 1.414213562 | 1 | 1 | | 1 | 1.414213562 | 0 | 0 | 0 MH572299,MH572425 |
| 2441 | VC_999_0 | 2 | 4.33446634 | 154.352742 | 0.027314529 | 0.260106748 | 2 | 2 | 1 | 1 | | 1 | 2 | 0 | 0 | 0 MH572426,MK249189 |
| 2442 | VC_99_0 | 32 | 45153.49508 | 5995.79033 | 0.88277861 | 5.38E-81 | 0 | 4.472135955 | 496 | 496 | | 1 | 3.000634047 | 0 | 0 | 0 AB647160,AB731695,JF734911,JQ617284,KX119174,KX119175,KX119176,KX119177,KX119182,KX119188,KX119189,KX119190,KX119191,KX119192,KX119193,KX119194,KX119195,KX119196,KX119197,KX119198,KX119199,KX119200,KX119201,KX119202,KX119203,KX119204,KX119205,KX119206,MW247144,MW247145,MW247147,OM515228 |
| 2443 | VC_9_0 | 38 | 72396.55326 | 75510.32145 | 0.48947389 | 0 | 0 | 8.831760866 | 703 | 703 | | 1 | 7.566579362 | 0 | 0 | 0 AB370205,AF424781,AP008953,AY954951,AY954952,AY954953,CP062427,CP062451,CP062454,DQ517338,DQ530359,DQ530360,EU861004,GU477322,HQ127381,JQ779024,JX274646,JX274647,KC677663,KJ716334,MG557619,MH321492,MK658834,MK801682,MK801683,MT013111,MW554488,OM439662,OM439666,OM439668,OM439670,OM439671,OM439675,ON911714,ON911715,ON911716,ON911717,ON911718 |

| | Genome | Order | Family | Genus | VC | VC Status | Size | VC Subcluster | VC Subcluster Size | Quality | Adj P-value | Topology Confidence Score | Genera in VC | Families in VC | Orders in VC | Genus Confidence Score | Reference |
|---|---|---|---|---|---|---|---|---|---|---|---|---|---|---|---|---|---|
| 0 | AB002632 | Unassigned | Unassigned | Unassigned | 0_0 | Clustered | 3 | VC_0_0 | 3 | 1 | 1 | 1 | 0 | 0 | 0 | 1 | Ref |
| 1 | AB008550 | Unassigned | Unassigned | Unassigned | 2_0 | Clustered | 4 | VC_2_0 | 4 | 0.1311 | 0.99998692 | 0.1311 | 0 | 0 | 0 | 1 | Ref |
| 2 | AB009866 | Unassigned | Unassigned | Unassigned | 7_0 | Clustered | 28 | VC_7_0 | 28 | 0.3916 | 1 | 0.3916 | 0 | 0 | 0 | 0.9947 | Ref |
| 3 | AB012573 | Unassigned | Unassigned | Unassigned | 16_0 | Clustered | 12 | VC_16_0 | 12 | 0.6615 | 1 | 0.6615 | 0 | 0 | 0 | 1 | Ref |
| 4 | AB012574 | Unassigned | Unassigned | Unassigned | 16_0 | Clustered | 12 | VC_16_0 | 12 | 0.6615 | 1 | 0.6615 | 0 | 0 | 0 | 1 | Ref |
| 5 | AB016282 | Unassigned | Unassigned | Unassigned | 18_0 | Clustered | 4 | VC_18_0 | 4 | 0.9098 | 0.99982347 | 0.9096 | 0 | 0 | 0 | 1 | Ref |
| 6 | AB036666 | Unassigned | Unassigned | Unassigned | 22_0 | Clustered | 11 | VC_22_0 | 11 | 0.5163 | 1 | 0.5163 | 0 | 0 | 0 | 1 | Ref |
| 7 | AB043678 | Unassigned | Unassigned | Unassigned | 17_0 | Clustered | 9 | VC_17_0 | 9 | 0.6969 | 1 | 0.6969 | 0 | 0 | 0 | 1 | Ref |
| 8 | AB043679 | Unassigned | Unassigned | Unassigned | 17_0 | Clustered | 9 | VC_17_0 | 9 | 0.6969 | 1 | 0.6969 | 0 | 0 | 0 | 1 | Ref |
| 9 | AB044554 | Unassigned | Unassigned | Unassigned | 7_0 | Clustered | 28 | VC_7_0 | 28 | 0.3916 | 1 | 0.3916 | 0 | 0 | 0 | 0.9947 | Ref |
| 10 | AB045978 | Unassigned | Unassigned | Unassigned | 8_0 | Clustered | 58 | VC_8_0 | 58 | 0.8306 | 1 | 0.8306 | 0 | 0 | 0 | 0.9946 | Ref |
| 11 | AB063393 | Unassigned | Unassigned | Unassigned | 27_0 | Clustered | 2 | VC_27_0 | 2 | 1 | 1 | 1 | 0 | 0 | 0 | 1 | Ref |
| 12 | AB102868 | Unassigned | Unassigned | Unassigned | 28_1 | Clustered | 53 | VC_28_1 | 53 | 0.7164 | 1 | 0.7164 | 0 | 0 | 0 | 0.9325 | Ref |
| 13 | AB231700 | Unassigned | Unassigned | Unassigned | 38_0 | Clustered | 3 | VC_38_0 | 3 | 1 | 0 | 0 | 0 | 0 | 0 | 1 | Ref |
| 14 | AB243556 | Unassigned | Unassigned | Unassigned | 7_0 | Clustered | 28 | VC_7_0 | 28 | 0.3916 | 1 | 0.3916 | 0 | 0 | 0 | 0.9947 | Ref |
| 15 | AB259123 | Unassigned | Unassigned | Unassigned | 52_0 | Clustered | 87 | VC_52_0 | 87 | 0.9858 | 1 | 0.9858 | 0 | 0 | 0 | 1 | Ref |
| 16 | AB259124 | Unassigned | Unassigned | Unassigned | 54_0 | Clustered | 6 | VC_54_0 | 6 | 0.6725 | 0.9999999 | 0.6725 | 0 | 0 | 0 | 1 | Ref |
| 17 | AB276040 | Unassigned | Unassigned | Unassigned | 1_0 | Clustered | 54 | VC_1_0 | 54 | 0.8527 | 1 | 0.8527 | 0 | 0 | 0 | 1 | Ref |
| 18 | AB285204 | Unassigned | Unassigned | Unassigned | 51_0 | Clustered | 10 | VC_51_0 | 10 | 0.1741 | 1 | 0.1741 | 0 | 0 | 0 | 1 | Ref |
| 19 | AB334720 | Unassigned | Unassigned | Unassigned | 61_0 | Clustered | 12 | VC_61_0 | 12 | 0.7633 | 1 | 0.7633 | 0 | 0 | 0 | 1 | Ref |
| 20 | AB334721 | Unassigned | Unassigned | Unassigned | 61_0 | Clustered | 12 | VC_61_0 | 12 | 0.7633 | 1 | 0.7633 | 0 | 0 | 0 | 1 | Ref |
| 21 | AB362338 | Unassigned | Unassigned | Unassigned | 28_1 | Clustered | 53 | VC_28_1 | 53 | 0.7164 | 1 | 0.7164 | 0 | 0 | 0 | 0.9325 | Ref |
| 22 | AB366653 | Unassigned | Unassigned | Unassigned | 66_3 | Clustered/Singleton | 1 | VC_66_3 | 1 | 0 | 1 | 0 | 0 | 0 | 0 | 1 | Ref |
| 23 | AB370205 | Unassigned | Unassigned | Unassigned | 9_0 | Clustered | 38 | VC_9_0 | 38 | 0.4895 | 1 | 0.4895 | 0 | 0 | 0 | 1 | Ref |
| 24 | AB370268 | Unassigned | Unassigned | Unassigned | 12_2 | Clustered | 19 | VC_12_2 | 19 | 0.2018 | 1 | 0.2018 | 0 | 0 | 0 | 1 | Ref |
| 25 | AB374228 | Unassigned | Unassigned | Unassigned | 5_0 | Clustered | 30 | VC_5_0 | 30 | 0.9647 | 1 | 0.9647 | 0 | 0 | 0 | 1 | Ref |
| 26 | AB426868 | Unassigned | Unassigned | Unassigned | 28_1 | Clustered | 53 | VC_28_1 | 53 | 0.7164 | 1 | 0.7164 | 0 | 0 | 0 | 0.9325 | Ref |
| 27 | AB434711 | Unassigned | Unassigned | Unassigned | 52_0 | Clustered | 87 | VC_52_0 | 87 | 0.9858 | 1 | 0.9858 | 0 | 0 | 0 | 1 | Ref |
| 28 | AB472900 | Unassigned | Unassigned | Unassigned | 77_0 | Clustered | 20 | VC_77_0 | 20 | 0.9657 | 1 | 0.9657 | 0 | 0 | 0 | 1 | Ref |
| 29 | AB560486 | Unassigned | Unassigned | Unassigned | 79_0 | Clustered | 64 | VC_79_0 | 64 | 0.4185 | 1 | 0.4185 | 0 | 0 | 0 | 0.7649 | Ref |
| 30 | AB572858 | Unassigned | Unassigned | Unassigned | 16_0 | Clustered | 12 | VC_16_0 | 12 | 0.6615 | 1 | 0.6615 | 0 | 0 | 0 | 1 | Ref |
| 31 | AB597179 | Unassigned | Unassigned | Unassigned | 83_0 | Clustered | 6 | VC_83_0 | 6 | 0.6436 | 1 | 0.6436 | 0 | 0 | 0 | 1 | Ref |
| 32 | AB609718 | Unassigned | Unassigned | Unassigned | 91_0 | Clustered | 22 | VC_91_0 | 22 | 0.5318 | 1 | 0.5318 | 0 | 0 | 0 | 1 | Ref |
| 33 | AB626962 | Unassigned | Unassigned | Unassigned | 96_0 | Clustered | 30 | VC_96_0 | 30 | 0.718 | 1 | 0.718 | 0 | 0 | 0 | 1 | Ref |
| 34 | AB626963 | Unassigned | Unassigned | Unassigned | 96_0 | Clustered | 30 | VC_96_0 | 30 | 0.718 | 1 | 0.718 | 0 | 0 | 0 | 1 | Ref |
| 35 | AB647160 | Unassigned | Unassigned | Unassigned | 99_0 | Clustered | 32 | VC_99_0 | 32 | 0.8828 | 1 | 0.8828 | 0 | 0 | 0 | 1 | Ref |
| 36 | AB712291 | Unassigned | Unassigned | Unassigned | 108_0 | Clustered | 33 | VC_108_0 | 33 | 0.949 | 1 | 0.949 | 0 | 0 | 0 | 1 | Ref |
| 37 | AB716666 | Unassigned | Unassigned | Unassigned | 111_0 | Clustered | 91 | VC_111_0 | 91 | 0.9587 | 1 | 0.9587 | 0 | 0 | 0 | 1 | Ref |
| 38 | AB720063 | Unassigned | Unassigned | Unassigned | 113_0 | Clustered | 9 | VC_113_0 | 9 | 0.519 | 1 | 0.519 | 0 | 0 | 0 | 1 | Ref |
| 39 | AB720064 | Unassigned | Unassigned | Unassigned | 116_0 | Clustered | 13 | VC_116_0 | 13 | 1 | 1 | 1 | 0 | 0 | 0 | 1 | Ref |
| 40 | AB731695 | Unassigned | Unassigned | Unassigned | 99_0 | Clustered | 32 | VC_99_0 | 32 | 0.8828 | 1 | 0.8828 | 0 | 0 | 0 | 1 | Ref |
| 41 | AB746912 | Unassigned | Unassigned | Unassigned | 118_0 | Clustered | 3 | VC_118_0 | 3 | 0.7119 | 0.99211494 | 0.7063 | 0 | 0 | 0 | 1 | Ref |
| 42 | AB757800 | Unassigned | Unassigned | Unassigned | 117_0 | Clustered | 9 | VC_117_0 | 9 | 0.9977 | 0.98988637 | 0.9876 | 0 | 0 | 0 | 1 | Ref |
| 43 | AB757801 | Unassigned | Unassigned | Unassigned | 117_0 | Clustered | 9 | VC_117_0 | 9 | 0.9977 | 0.98988637 | 0.9876 | 0 | 0 | 0 | 1 | Ref |
| 44 | AB775548 | Unassigned | Unassigned | Unassigned | 125_0 | Clustered | 17 | VC_125_0 | 17 | 0.9244 | 1 | 0.9244 | 0 | 0 | 0 | 1 | Ref |
| 45 | AB775549 | Unassigned | Unassigned | Unassigned | 130_0 | Clustered | 19 | VC_130_0 | 19 | 0.6068 | 1 | 0.6068 | 0 | 0 | 0 | 1 | Ref |
| 46 | AB797215 | Unassigned | Unassigned | Unassigned | 132_1 | Clustered | 17 | VC_132_1 | 17 | 0.1738 | 1 | 0.1738 | 0 | 0 | 0 | 0.9853 | Ref |
| 47 | AB828698 | Unassigned | Unassigned | Unassigned | 54_0 | Clustered | 6 | VC_54_0 | 6 | 0.6725 | 0.9999999 | 0.6725 | 0 | 0 | 0 | 1 | Ref |
| 48 | AB853330 | Unassigned | Unassigned | Unassigned | 149_13 | Clustered | 3 | VC_149_13 | 3 | 0.0087 | 0.95178903 | 0.0083 | 0 | 0 | 0 | 1 | Ref |
| 49 | AB853331 | Unassigned | Unassigned | Unassigned | 149_23 | Clustered/Singleton | 1 | VC_149_23 | 1 | 0 | 1 | 0 | 0 | 0 | 0 | 1 | Ref |
| 50 | AB854109 | Unassigned | Unassigned | Unassigned | 154_0 | Clustered | 4 | VC_154_0 | 4 | 0.1766 | 0.99995106 | 0.1766 | 0 | 0 | 0 | 1 | Ref |
| 51 | AB863625 | Unassigned | Unassigned | Unassigned | 161_0 | Clustered | 3 | VC_161_0 | 3 | 0.5777 | 0.99804396 | 0.5765 | 0 | 0 | 0 | 1 | Ref |
| 52 | AB897757 | Unassigned | Unassigned | Unassigned | 164_10 | Clustered | 5 | VC_164_10 | 5 | 0.0615 | 0.96483582 | 0.0593 | 0 | 0 | 0 | 1 | Ref |
| 53 | AB903967 | Unassigned | Unassigned | Unassigned | 149_14 | Clustered/Singleton | 1 | VC_149_14 | 1 | 0 | 1 | 0 | 0 | 0 | 0 | 1 | Ref |
| 54 | AB910392 | Unassigned | Unassigned | Unassigned | 167_0 | Clustered | 3 | VC_167_0 | 3 | 0.6952 | 0.99826575 | 0.694 | 0 | 0 | 0 | 1 | Ref |
| 55 | AB910393 | Unassigned | Unassigned | Unassigned | 174_0 | Clustered | 3 | VC_174_0 | 3 | 0.9624 | 0.99333557 | 0.9559 | 0 | 0 | 0 | 1 | Ref |
| 56 | AB910602 | Unassigned | Unassigned | Unassigned | 177_0 | Clustered | 5 | VC_177_0 | 5 | 0.7468 | 0.99885646 | 0.746 | 0 | 0 | 0 | 1 | Ref |
| 57 | AB916497 | Unassigned | Unassigned | Unassigned | 180_0 | Clustered | 4 | VC_180_0 | 4 | 0.2056 | 0.9999862 | 0.2056 | 0 | 0 | 0 | 1 | Ref |
| 58 | AB920995 | Unassigned | Unassigned | Unassigned | 70_0 | Clustered | 27 | VC_70_0 | 27 | 0.5981 | 1 | 0.5981 | 0 | 0 | 0 | 0.9886 | Ref |
| 59 | AB930182 | Unassigned | Unassigned | Unassigned | 85_3 | Clustered | 19 | VC_85_3 | 19 | 0.3671 | 1 | 0.3671 | 0 | 0 | 0 | 0.807 | Ref |
| 60 | AB931172 | Unassigned | Unassigned | Unassigned | 54_0 | Clustered | 6 | VC_54_0 | 6 | 0.6725 | 0.9999999 | 0.6725 | 0 | 0 | 0 | 1 | Ref |
| 61 | AB937974 | Unassigned | Unassigned | Unassigned | 52_0 | Clustered | 87 | VC_52_0 | 87 | 0.9858 | 1 | 0.9858 | 0 | 0 | 0 | 1 | Ref |
| 62 | AB971354 | Unassigned | Unassigned | Unassigned | 188_0 | Clustered | 13 | VC_188_0 | 13 | 0.9973 | 0.99823802 | 0.9956 | 0 | 0 | 0 | 1 | Ref |
| 63 | AB981169 | Unassigned | Unassigned | Unassigned | 1_0 | Clustered | 54 | VC_1_0 | 54 | 0.8527 | 1 | 0.8527 | 0 | 0 | 0 | 1 | Ref |
| 64 | AB981170 | Unassigned | Unassigned | Unassigned | 52_0 | Clustered | 87 | VC_52_0 | 87 | 0.9858 | 1 | 0.9858 | 0 | 0 | 0 | 1 | Ref |
| 65 | AB983711 | Unassigned | Unassigned | Unassigned | 70_0 | Clustered | 27 | VC_70_0 | 27 | 0.5981 | 1 | 0.5981 | 0 | 0 | 0 | 0.9886 | Ref |
| 66 | AC171169 | Unassigned | Unassigned | Unassigned | 190_11 | Clustered/Singleton | 1 | VC_190_11 | 1 | 0 | 1 | 0 | 0 | 0 | 0 | 1 | Ref |
| 67 | AE002163 | Unassigned | Unassigned | Unassigned | 197_0 | Clustered | 5 | VC_197_0 | 5 | 0.1324 | 0.84698401 | 0.1122 | 0 | 0 | 0 | 1 | Ref |
| 68 | AF009630 | Unassigned | Unassigned | Unassigned | 202_0 | Clustered | 102 | VC_202_0 | 102 | 0.4049 | 1 | 0.4049 | 0 | 0 | 0 | 0.7876 | Ref |
| 69 | AF011378 | Unassigned | Unassigned | Unassigned | 202_0 | Clustered | 102 | VC_202_0 | 102 | 0.4049 | 1 | 0.4049 | 0 | 0 | 0 | 0.7876 | Ref |
| 70 | AF020713 | Unassigned | Unassigned | Unassigned | 89_3 | Clustered/Singleton | 1 | VC_89_3 | 1 | 0 | 1 | 0 | 0 | 0 | 0 | 1 | Ref |
| 71 | AF022214 | Unassigned | Unassigned | Unassigned | 215_1 | Clustered | 19 | VC_215_1 | 19 | 0.4188 | 1 | 0.4188 | 0 | 0 | 0 | 1 | Ref |
| 72 | AF059242 | Unassigned | Unassigned | Unassigned | 188_0 | Clustered | 13 | VC_188_0 | 13 | 0.9973 | 0.99823802 | 0.9956 | 0 | 0 | 0 | 1 | Ref |
| 73 | AF059243 | Unassigned | Unassigned | Unassigned | 188_0 | Clustered | 13 | VC_188_0 | 13 | 0.9973 | 0.99823802 | 0.9956 | 0 | 0 | 0 | 1 | Ref |
| 74 | AF063097 | Unassigned | Unassigned | Unassigned | 228_0 | Clustered | 60 | VC_228_0 | 60 | 0.942 | 1 | 0.942 | 0 | 0 | 0 | 1 | Ref |
| 75 | AF065411 | Unassigned | Unassigned | Unassigned | 232_0 | Clustered | 2 | VC_232_0 | 2 | 0.9272 | 0.72985431 | 0.6768 | 0 | 0 | 0 | 1 | Ref |
| 76 | AF083977 | Unassigned | Unassigned | Unassigned | 50_0 | Clustered | 9 | VC_50_0 | 9 | 0.5531 | 1 | 0.5531 | 0 | 0 | 0 | 1 | Ref |
| 77 | AF085222 | Unassigned | Unassigned | Unassigned | 252_0 | Clustered | 121 | VC_252_0 | 121 | 0.7376 | 1 | 0.7376 | 0 | 0 | 0 | 0.9999 | Ref |
| 78 | AF115102 | Unassigned | Unassigned | Unassigned | 252_0 | Clustered | 121 | VC_252_0 | 121 | 0.7376 | 1 | 0.7376 | 0 | 0 | 0 | 0.9999 | Ref |
| 79 | AF115103 | Unassigned | Unassigned | Unassigned | 252_0 | Clustered | 121 | VC_252_0 | 121 | 0.7376 | 1 | 0.7376 | 0 | 0 | 0 | 0.9999 | Ref |
| 80 | AF125163 | Unassigned | Unassigned | Unassigned | 5_0 | Clustered | 30 | VC_5_0 | 30 | 0.9647 | 1 | 0.9647 | 0 | 0 | 0 | 1 | Ref |
| 81 | AF125520 | Unassigned | Unassigned | Unassigned | 39_0 | Clustered | 35 | VC_39_0 | 35 | 0.4337 | 1 | 0.4337 | 0 | 0 | 0 | 1 | Ref |
| 82 | AF133242 | Unassigned | Unassigned | Unassigned | 276_0 | Clustered | 2 | VC_276_0 | 2 | 1 | 1 | 1 | 0 | 0 | 0 | 1 | Ref |
| 83 | AF145054 | Unassigned | Unassigned | Unassigned | 252_0 | Clustered | 121 | VC_252_0 | 121 | 0.7376 | 1 | 0.7376 | 0 | 0 | 0 | 0.9999 | Ref |
| 84 | AF155037 | Unassigned | Unassigned | Unassigned | 277_0 | Clustered | 3 | VC_277_0 | 3 | 1 | 1 | 1 | 0 | 0 | 0 | 1 | Ref |
| 85 | AF157835 | Unassigned | Unassigned | Unassigned | 29_0 | Clustered | 10 | VC_29_0 | 10 | 0.5208 | 1 | 0.5208 | 0 | 0 | 0 | 1 | Ref |
| 86 | AF158101 | Unassigned | Unassigned | Unassigned | 283_91 | Clustered | 11 | VC_283_91 | 11 | 0.0285 | 1 | 0.0285 | 0 | 0 | 0 | 1 | Ref |
| 87 | AF158600 | Unassigned | Unassigned | Unassigned | 254_0 | Clustered | 46 | VC_254_0 | 46 | 0.5159 | 1 | 0.5159 | 0 | 0 | 0 | 1 | Ref |
| 88 | AF165214 | Unassigned | Unassigned | Unassigned | 176_0 | Clustered | 2 | VC_176_0 | 2 | 0.0498 | 0.95663741 | 0.0476 | 0 | 0 | 0 | 1 | Ref |
| 89 | AF176025 | Unassigned | Unassigned | Unassigned | 260_0 | Clustered | 13 | VC_260_0 | 13 | 0.3705 | 1 | 0.3705 | 0 | 0 | 0 | 1 | Ref |
| 90 | AF176027 | Unassigned | Unassigned | Unassigned | 303_0 | Clustered | 155 | VC_303_0 | 155 | 0.9979 | 1 | 0.9979 | 0 | 0 | 0 | 1 | Ref |
| 91 | AF176028 | Unassigned | Unassigned | Unassigned | 303_0 | Clustered | 155 | VC_303_0 | 155 | 0.9979 | 1 | 0.9979 | 0 | 0 | 0 | 1 | Ref |
| 92 | AF176029 | Unassigned | Unassigned | Unassigned | 303_0 | Clustered | 155 | VC_303_0 | 155 | 0.9979 | 1 | 0.9979 | 0 | 0 | 0 | 1 | Ref |
| 93 | AF176030 | Unassigned | Unassigned | Unassigned | 303_0 | Clustered | 155 | VC_303_0 | 155 | 0.9979 | 1 | 0.9979 | 0 | 0 | 0 | 1 | Ref |
| 94 | AF176031 | Unassigned | Unassigned | Unassigned | 303_0 | Clustered | 155 | VC_303_0 | 155 | 0.9979 | 1 | 0.9979 | 0 | 0 | 0 | 1 | Ref |
| 95 | AF176032 | Unassigned | Unassigned | Unassigned | 303_0 | Clustered | 155 | VC_303_0 | 155 | 0.9979 | 1 | 0.9979 | 0 | 0 | 0 | 1 | Ref |
| 96 | AF176033 | Unassigned | Unassigned | Unassigned | 303_0 | Clustered | 155 | VC_303_0 | 155 | 0.9979 | 1 | 0.9979 | 0 | 0 | 0 | 1 | Ref |
| 97 | AF176034 | Unassigned | Unassigned | Unassigned | 303_0 | Clustered | 155 | VC_303_0 | 155 | 0.9979 | 1 | 0.9979 | 0 | 0 | 0 | 1 | Ref |
| 98 | AF189021 | Unassigned | Unassigned | Unassigned | 304_0 | Clustered | 11 | VC_304_0 | 11 | 0.7869 | 1 | 0.7869 | 0 | 0 | 0 | 1 | Ref |
| 99 | AF191797 | Unassigned | Unassigned | Unassigned | 306_0 | Clustered | 2 | VC_306_0 | 2 | 1 | 1 | 1 | 0 | 0 | 0 | 1 | Ref |
| 100 | AF195902 | Unassigned | Unassigned | Unassigned | 308_0 | Clustered | 4 | VC_308_0 | 4 | 0.5014 | 0.99997418 | 0.5013 | 0 | 0 | 0 | 1 | Ref |
| 101 | AF217253 | Unassigned | Unassigned | Unassigned | 28_1 | Clustered | 53 | VC_28_1 | 53 | 0.7164 | 1 | 0.7164 | 0 | 0 | 0 | 0.9325 | Ref |
| 102 | AF222060 | Unassigned | Unassigned | Unassigned | 310_0 | Clustered | 33 | VC_310_0 | 33 | 0.8771 | 1 | 0.8771 | 0 | 0 | 0 | 1 | Ref |
| 103 | AF227250 | Unassigned | Unassigned | Unassigned | 315_0 | Clustered | 12 | VC_315_0 | 12 | 0.8947 | 1 | 0.8947 | 0 | 0 | 0 | 1 | Ref |
| 104 | AF232233 | Unassigned | Unassigned | Unassigned | 251_0 | Clustered | 22 | VC_251_0 | 22 | 0.6508 | 1 | 0.6508 | 0 | 0 | 0 | 1 | Ref |
| 105 | AF234172 | Unassigned | Unassigned | Unassigned | 247_0 | Clustered | 14 | VC_247_0 | 14 | 0.9119 | 1 | 0.9119 | 0 | 0 | 0 | 0.967 | Ref |
| 106 | AF234173 | Unassigned | Unassigned | Unassigned | 247_0 | Clustered | 14 | VC_247_0 | 14 | 0.9119 | 1 | 0.9119 | 0 | 0 | 0 | 0.967 | Ref |
| 107 | AF271693 | Unassigned | Unassigned | Unassigned | 325_39 | Clustered | 2 | VC_325_39 | 2 | 0.0045 | 0.95750889 | 0.0043 | 0 | 0 | 0 | 1 | Ref |
| 108 | AF274751 | Unassigned | Unassigned | Unassigned | 303_0 | Clustered | 155 | VC_303_0 | 155 | 0.9979 | 1 | 0.9979 | 0 | 0 | 0 | 1 | Ref |
| 109 | AF299300 | Unassigned | Unassigned | Unassigned | 303_0 | Clustered | 155 | VC_303_0 | 155 | 0.9979 | 1 | 0.9979 | 0 | 0 | 0 | 1 | Ref |
| 110 | AF299301 | Unassigned | Unassigned | Unassigned | 303_0 | Clustered | 155 | VC_303_0 | 155 | 0.9979 | 1 | 0.9979 | 0 | 0 | 0 | 1 | Ref |
| 111 | AF299302 | Unassigned | Unassigned | Unassigned | 303_0 | Clustered | 155 | VC_303_0 | 155 | 0.9979 | 1 | 0.9979 | 0 | 0 | 0 | 1 | Ref |
| 112 | AF299303 | Unassigned | Unassigned | Unassigned | 303_0 | Clustered | 155 | VC_303_0 | 155 | 0.9979 | 1 | 0.9979 | 0 | 0 | 0 | 1 | Ref |
| 113 | AF299304 | Unassigned | Unassigned | Unassigned | 303_0 | Clustered | 155 | VC_303_0 | 155 | 0.9979 | 1 | 0.9979 | 0 | 0 | 0 | 1 | Ref |
| 114 | AF299305 | Unassigned | Unassigned | Unassigned | 303_0 | Clustered | 155 | VC_303_0 | 155 | 0.9979 | 1 | 0.9979 | 0 | 0 | 0 | 1 | Ref |
| 115 | AF299306 | Unassigned | Unassigned | Unassigned | 303_0 | Clustered | 155 | VC_303_0 | 155 | 0.9979 | 1 | 0.9979 | 0 | 0 | 0 | 1 | Ref |
| 116 | AF299307 | Unassigned | Unassigned | Unassigned | 303_0 | Clustered | 155 | VC_303_0 | 155 | 0.9979 | 1 | 0.9979 | 0 | 0 | 0 | 1 | Ref |
| 117 | AF299308 | Unassigned | Unassigned | Unassigned | 303_0 | Clustered | 155 | VC_303_0 | 155 | 0.9979 | 1 | 0.9979 | 0 | 0 | 0 | 1 | Ref |
| 118 | AF299309 | Unassigned | Unassigned | Unassigned | 303_0 | Clustered | 155 | VC_303_0 | 155 | 0.9979 | 1 | 0.9979 | 0 | 0 | 0 | 1 | Ref |
| 119 | AF299310 | Unassigned | Unassigned | Unassigned | 303_0 | Clustered | 155 | VC_303_0 | 155 | 0.9979 | 1 | 0.9979 | 0 | 0 | 0 | 1 | Ref |
| 120 | AF299311 | Unassigned | Unassigned | Unassigned | 303_0 | Clustered | 155 | VC_303_0 | 155 | 0.9979 | 1 | 0.9979 | 0 | 0 | 0 | 1 | Ref |
| 121 | AF299312 | Unassigned | Unassigned | Unassigned | 303_0 | Clustered | 155 | VC_303_0 | 155 | 0.9979 | 1 | 0.9979 | 0 | 0 | 0 | 1 | Ref |
| 122 | AF299313 | Unassigned | Unassigned | Unassigned | 303_0 | Clustered | 155 | VC_303_0 | 155 | 0.9979 | 1 | 0.9979 | 0 | 0 | 0 | 1 | Ref |
| 123 | AF299314 | Unassigned | Unassigned | Unassigned | 303_0 | Clustered | 155 | VC_303_0 | 155 | 0.9979 | 1 | 0.9979 | 0 | 0 | 0 | 1 | Ref |
| 124 | AF301375 | Unassigned | Unassigned | Unassigned | 232_0 | Clustered | 2 | VC_232_0 | 2 | 0.9272 | 0.72985431 | 0.6768 | 0 | 0 | 0 | 1 | Ref |
| 125 | AF306496 | Unassigned | Unassigned | Unassigned | 327_0 | Clustered | 12 | VC_327_0 | 12 | 0.8925 | 1 | 0.8925 | 0 | 0 | 0 | 1 | Ref |
| 126 | AF320576 | Unassigned | Unassigned | Unassigned | 329_1 | Clustered | 17 | VC_329_1 | 17 | 0.0867 | 1 | 0.0867 | 0 | 0 | 0 | 0.8235 | Ref |
| 127 | AF323668 | Unassigned | Unassigned | Unassigned | 263_0 | Clustered | 7 | VC_263_0 | 7 | 0.2364 | 1 | 0.2364 | 0 | 0 | 0 | 1 | Ref |
| 128 | AF323669 | Unassigned | Unassigned | Unassigned | 260_0 | Clustered | 13 | VC_260_0 | 13 | 0.3705 | 1 | 0.3705 | 0 | 0 | 0 | 1 | Ref |
| 129 | AF323671 | Unassigned | Unassigned | Unassigned | 339_0 | Clustered | 9 | VC_339_0 | 9 | 0.9405 | 0.99999771 | 0.9405 | 0 | 0 | 0 | 1 | Ref |
| 130 | AF334111 | Unassigned | Unassigned | Unassigned | 227_0 | Clustered | 2 | VC_227_0 | 2 | 0.3333 | 0 | 0 | 0 | 0 | 0 | 1 | Ref |
| 131 | AF335538 | Unassigned | Unassigned | Unassigned | 30_1 | Clustered | 16 | VC_30_1 | 16 | 0.1973 | 1 | 0.1973 | 0 | 0 | 0 | 0.8333 | Ref |
| 132 | AF338467 | Unassigned | Unassigned | Unassigned | 340_0 | Clustered | 31 | VC_340_0 | 31 | 0.9549 | 1 | 0.9549 | 0 | 0 | 0 | 1 | Ref |
| 133 | AF396866 | Unassigned | Unassigned | Unassigned | 341_0 | Clustered | 3 | VC_341_0 | 3 | 0.2522 | 0.9396546 | 0.237 | 0 | 0 | 0 | 1 | Ref |
| 134 | AF399011 | Unassigned | Unassigned | Unassigned | 344_0 | Clustered | 21 | VC_344_0 | 21 | 0.5436 | 1 | 0.5436 | 0 | 0 | 0 | 1 | Ref |
| 135 | AF424781 | Unassigned | Unassigned | Unassigned | 9_0 | Clustered | 38 | VC_9_0 | 38 | 0.4895 | 1 | 0.4895 | 0 | 0 | 0 | 1 | Ref |
| 136 | AF424782 | Unassigned | Unassigned | Unassigned | 8_0 | Clustered | 58 | VC_8_0 | 58 | 0.8306 | 1 | 0.8306 | 0 | 0 | 0 | 0.9946 | Ref |
| 137 | AF424783 | Unassigned | Unassigned | Unassigned | 7_0 | Clustered | 28 | VC_7_0 | 28 | 0.3916 | 1 | 0.3916 | 0 | 0 | 0 | 0.9947 | Ref |
| 138 | AF440571 | Unassigned | Unassigned | Unassigned | 349_0 | Clustered | 7 | VC_349_0 | 7 | 0.9363 | 0.99999996 | 0.9363 | 0 | 0 | 0 | 1 | Ref |
| 139 | AF440695 | Unassigned | Unassigned | Unassigned | 352_0 | Clustered | 3 | VC_352_0 | 3 | 0.7306 | 0.98867275 | 0.7223 | 0 | 0 | 0 | 1 | Ref |
| 140 | AF447491 | Unassigned | Unassigned | Unassigned | 353_0 | Clustered | 6 | VC_353_0 | 6 | 0.545 | 1 | 0.545 | 0 | 0 | 0 | 1 | Ref |
| 141 | AF452449 | Unassigned | Unassigned | Unassigned | 16_0 | Clustered | 12 | VC_16_0 | 12 | 0.6615 | 1 | 0.6615 | 0 | 0 | 0 | 1 | Ref |
| 142 | AF453500 | Unassigned | Unassigned | Unassigned | 16_0 | Clustered | 12 | VC_16_0 | 12 | 0.6615 | 1 | 0.6615 | 0 | 0 | 0 | 1 | Ref |
| 143 | AF454431 | Unassigned | Unassigned | Unassigned | 360_0 | Clustered | 358 | VC_360_0 | 358 | 0.9615 | 1 | 0.9615 | 0 | 0 | 0 | 1 | Ref |
| 144 | AF489521 | Unassigned | Unassigned | Unassigned | 260_0 | Clustered | 13 | VC_260_0 | 13 | 0.3705 | 1 | 0.3705 | 0 | 0 | 0 | 1 | Ref |
| 145 | AF493143 | Unassigned | Unassigned | Unassigned | 129_0 | Clustered | 43 | VC_129_0 | 43 | 0.886 | 1 | 0.886 | 0 | 0 | 0 | 1 | Ref |
| 146 | AF503408 | Unassigned | Unassigned | Unassigned | 247_0 | Clustered | 14 | VC_247_0 | 14 | 0.9119 | 1 | 0.9119 | 0 | 0 | 0 | 0.967 | Ref |
| 147 | AF513032 | Unassigned | Unassigned | Unassigned | 96_0 | Clustered | 30 | VC_96_0 | 30 | 0.718 | 1 | 0.718 | 0 | 0 | 0 | 1 | Ref |
| 148 | AF513033 | Unassigned | Unassigned | Unassigned | 96_0 | Clustered | 30 | VC_96_0 | 30 | 0.718 | 1 | 0.718 | 0 | 0 | 0 | 1 | Ref |
| 149 | AF527608 | Unassigned | Unassigned | Unassigned | 28_1 | Clustered | 53 | VC_28_1 | 53 | 0.7164 | 1 | 0.7164 | 0 | 0 | 0 | 0.9325 | Ref |
| 150 | AF547430 | Unassigned | Unassigned | Unassigned | 371_16 | Clustered | 42 | VC_371_16 | 42 | 0.1341 | 1 | 0.1341 | 0 | 0 | 0 | 0.7282 | Ref |
| 151 | AF547987 | Unassigned | Unassigned | Unassigned | 30_1 | Clustered | 16 | VC_30_1 | 16 | 0.1973 | 1 | 0.1973 | 0 | 0 | 0 | 0.8333 | Ref |
| 152 | AJ006589 | Unassigned | Unassigned | Unassigned | 373_0 | Clustered | 10 | VC_373_0 | 10 | 0.7471 | 1 | 0.7471 | 0 | 0 | 0 | 1 | Ref |
| 153 | AJ131519 | Unassigned | Unassigned | Unassigned | 307_0 | Clustered | 2 | VC_307_0 | 2 | 0.3835 | 0.95251406 | 0.3653 | 0 | 0 | 0 | 1 | Ref |
| 154 | AJ242593 | Unassigned | Unassigned | Unassigned | 378_0 | Clustered | 9 | VC_378_0 | 9 | 0.7603 | 1 | 0.7603 | 0 | 0 | 0 | 1 | Ref |
| 155 | AJ245616 | Unassigned | Unassigned | Unassigned | 260_0 | Clustered | 13 | VC_260_0 | 13 | 0.3705 | 1 | 0.3705 | 0 | 0 | 0 | 1 | Ref |
| 156 | AJ251789 | Unassigned | Unassigned | Unassigned | 272_0 | Clustered | 11 | VC_272_0 | 11 | 0.4915 | 1 | 0.4915 | 0 | 0 | 0 | 0.8727 | Ref |
| 157 | AJ251805 | Unassigned | Unassigned | Unassigned | 385_0 | Clustered | 51 | VC_385_0 | 51 | 0.8754 | 1 | 0.8754 | 0 | 0 | 0 | 1 | Ref |
| 158 | AJ270057 | Unassigned | Unassigned | Unassigned | 198_0 | Clustered | 28 | VC_198_0 | 28 | 0.2429 | 0.99971175 | 0.2429 | 0 | 0 | 0 | 1 | Ref |
| 159 | AJ278322 | Unassigned | Unassigned | Unassigned | 407_0 | Clustered | 3 | VC_407_0 | 3 | 0.3352 | 0.99824791 | 0.3346 | 0 | 0 | 0 | 1 | Ref |
| 160 | AJ298298 | Unassigned | Unassigned | Unassigned | 45_0 | Clustered | 12 | VC_45_0 | 12 | 0.35 | 1 | 0.35 | 0 | 0 | 0 | 1 | Ref |

| | | | | | | | | | | | | | | | | | |
|---|---|---|---|---|---|---|---|---|---|---|---|---|---|---|---|---|---|
| 161 | AJ302074 | Unassigned | Unassigned | Unassigned | 412_0 | Clustered | 39 | VC_412_0 | 39 | 0.845 | 1 | 0.845 | 0 | 0 | 0 | 1 | Ref |
| 162 | AJ312240 | Unassigned | Unassigned | Unassigned | 379_0 | Clustered | 5 | VC_379_0 | 5 | 0.4779 | 0.99999992 | 0.4779 | 0 | 0 | 0 | 0.9 | Ref |
| 163 | AJ318471 | Unassigned | Unassigned | Unassigned | 385_0 | Clustered | 51 | VC_385_0 | 51 | 0.8754 | 1 | 0.8754 | 0 | 0 | 0 | 1 | Ref |
| 164 | AJ344259 | Unassigned | Unassigned | Unassigned | 348_0 | Clustered | 22 | VC_348_0 | 22 | 0.9602 | 1 | 0.9602 | 0 | 0 | 0 | 1 | Ref |
| 165 | AJ414696 | Unassigned | Unassigned | Unassigned | 348_0 | Clustered | 22 | VC_348_0 | 22 | 0.9602 | 1 | 0.9602 | 0 | 0 | 0 | 1 | Ref |
| 166 | AJ421943 | Unassigned | Unassigned | Unassigned | 274_0 | Clustered | 2 | VC_274_0 | 2 | 0.0799 | 0.953709 | 0.0762 | 0 | 0 | 0 | 1 | Ref |
| 167 | AJ505558 | Unassigned | Unassigned | Unassigned | 423_0 | Clustered | 45 | VC_423_0 | 45 | 0.9484 | 1 | 0.9484 | 0 | 0 | 0 | 1 | Ref |
| 168 | AJ536073 | Unassigned | Unassigned | Unassigned | 424_0 | Clustered | 7 | VC_424_0 | 7 | 0.9044 | 0.9998143 | 0.9043 | 0 | 0 | 0 | 1 | Ref |
| 169 | AJ550635 | Unassigned | Unassigned | Unassigned | 198_0 | Clustered | 28 | VC_198_0 | 28 | 0.2429 | 0.99971175 | 0.2429 | 0 | 0 | 0 | 1 | Ref |
| 170 | AJ550940 | Unassigned | Unassigned | Unassigned | 373_0 | Clustered | 10 | VC_373_0 | 10 | 0.7471 | 1 | 0.7471 | 0 | 0 | 0 | 1 | Ref |
| 171 | AJ556162 | Unassigned | Unassigned | Unassigned | 41_0 | Clustered | 16 | VC_41_0 | 16 | 0.2025 | 1 | 0.2025 | 0 | 0 | 0 | 0.9667 | Ref |
| 172 | AJ560763 | Unassigned | Unassigned | Unassigned | 426_0 | Clustered | 2 | VC_426_0 | 2 | 0.1976 | 0.95643941 | 0.189 | 0 | 0 | 0 | 1 | Ref |
| 173 | AJ604530 | Unassigned | Unassigned | Unassigned | 189_0 | Clustered | 2 | VC_189_0 | 2 | 0.0104 | 0.93953444 | 0.0097 | 0 | 0 | 0 | 1 | Ref |
| 174 | AJ604531 | Unassigned | Unassigned | Unassigned | 189_0 | Clustered | 2 | VC_189_0 | 2 | 0.0104 | 0.93953444 | 0.0097 | 0 | 0 | 0 | 1 | Ref |
| 175 | AJ609634 | Unassigned | Unassigned | Unassigned | 137_0 | Clustered | 3 | VC_137_0 | 3 | 0.1129 | 0.998547 | 0.1128 | 0 | 0 | 0 | 1 | Ref |
| 176 | AJ630128 | Unassigned | Unassigned | Unassigned | 433_5 | Clustered | 2 | VC_433_5 | 2 | 0.0173 | 0.93642158 | 0.0162 | 0 | 0 | 0 | 1 | Ref |
| 177 | AJ635161 | Unassigned | Unassigned | Unassigned | 440_0 | Clustered | 4 | VC_440_0 | 4 | 1 | 1 | 1 | 0 | 0 | 0 | 1 | Ref |
| 178 | AJ748296 | Unassigned | Unassigned | Unassigned | 348_0 | Clustered | 22 | VC_348_0 | 22 | 0.9602 | 1 | 0.9602 | 0 | 0 | 0 | 1 | Ref |
| 179 | AJ783769 | Unassigned | Unassigned | Unassigned | 444_0 | Clustered | 2 | VC_444_0 | 2 | 0.8137 | 0.72985431 | 0.5939 | 0 | 0 | 0 | 1 | Ref |
| 180 | AJ854042 | Unassigned | Unassigned | Unassigned | 350_0 | Clustered | 2 | VC_350_0 | 2 | 0.324 | 0.93865445 | 0.3041 | 0 | 0 | 0 | 1 | Ref |
| 181 | AJ875026 | Unassigned | Unassigned | Unassigned | 348_0 | Clustered | 22 | VC_348_0 | 22 | 0.9602 | 1 | 0.9602 | 0 | 0 | 0 | 1 | Ref |
| 182 | AJ888457 | Unassigned | Unassigned | Unassigned | 447_0 | Clustered | 3 | VC_447_0 | 3 | 0.8268 | 0.99509929 | 0.8227 | 0 | 0 | 0 | 1 | Ref |
| 183 | AJ969242 | Unassigned | Unassigned | Unassigned | 276_0 | Clustered | 2 | VC_276_0 | 2 | 1 | 1 | 1 | 0 | 0 | 0 | 1 | Ref |
| 184 | AM076770 | Unassigned | Unassigned | Unassigned | 450_0 | Clustered | 16 | VC_450_0 | 16 | 0.9564 | 1 | 0.9564 | 0 | 0 | 0 | 1 | Ref |
| 185 | AM084414 | Unassigned | Unassigned | Unassigned | 451_0 | Clustered | 80 | VC_451_0 | 80 | 0.9991 | 1 | 0.9991 | 0 | 0 | 0 | 1 | Ref |
| 186 | AM084415 | Unassigned | Unassigned | Unassigned | 452_0 | Clustered | 66 | VC_452_0 | 66 | 0.9248 | 1 | 0.9248 | 0 | 0 | 0 | 1 | Ref |
| 187 | AM087120 | Unassigned | Unassigned | Unassigned | 349_0 | Clustered | 7 | VC_349_0 | 7 | 0.9363 | 0.99999996 | 0.9363 | 0 | 0 | 0 | 1 | Ref |
| 188 | AM087121 | Unassigned | Unassigned | Unassigned | 349_0 | Clustered | 7 | VC_349_0 | 7 | 0.9363 | 0.99999996 | 0.9363 | 0 | 0 | 0 | 1 | Ref |
| 189 | AM087122 | Unassigned | Unassigned | Unassigned | 349_0 | Clustered | 7 | VC_349_0 | 7 | 0.9363 | 0.99999996 | 0.9363 | 0 | 0 | 0 | 1 | Ref |
| 190 | AM087123 | Unassigned | Unassigned | Unassigned | 349_0 | Clustered | 7 | VC_349_0 | 7 | 0.9363 | 0.99999996 | 0.9363 | 0 | 0 | 0 | 1 | Ref |
| 191 | AM156909 | Unassigned | Unassigned | Unassigned | 456_0 | Clustered | 62 | VC_456_0 | 62 | 0.9026 | 1 | 0.9026 | 0 | 0 | 0 | 1 | Ref |
| 192 | AM183667 | Unassigned | Unassigned | Unassigned | 386_0 | Clustered | 49 | VC_386_0 | 49 | 0.7941 | 1 | 0.7941 | 0 | 0 | 0 | 1 | Ref |
| 193 | AM265638 | Unassigned | Unassigned | Unassigned | 423_0 | Clustered | 45 | VC_423_0 | 45 | 0.9484 | 1 | 0.9484 | 0 | 0 | 0 | 1 | Ref |
| 194 | AM419438 | Unassigned | Unassigned | Unassigned | 351_0 | Clustered | 3 | VC_351_0 | 3 | 0.8899 | 0.98155579 | 0.8735 | 0 | 0 | 0 | 1 | Ref |
| 195 | AM491472 | Unassigned | Unassigned | Unassigned | 461_0 | Clustered | 96 | VC_461_0 | 96 | 0.9307 | 1 | 0.9307 | 0 | 0 | 0 | 0.9575 | Ref |
| 196 | AM749121 | Unassigned | Unassigned | Unassigned | 463_0 | Clustered | 4 | VC_463_0 | 4 | 0.7906 | 0.99997156 | 0.7906 | 0 | 0 | 0 | 1 | Ref |
| 197 | AM749441 | Unassigned | Unassigned | Unassigned | 166_0 | Clustered | 26 | VC_166_0 | 26 | 0.7923 | 1 | 0.7923 | 0 | 0 | 0 | 1 | Ref |
| 198 | AM910650 | Unassigned | Unassigned | Unassigned | 472_0 | Clustered | 47 | VC_472_0 | 47 | 0.9025 | 1 | 0.9025 | 0 | 0 | 0 | 1 | Ref |
| 199 | AM910651 | Unassigned | Unassigned | Unassigned | 423_0 | Clustered | 45 | VC_423_0 | 45 | 0.9484 | 1 | 0.9484 | 0 | 0 | 0 | 1 | Ref |
| 200 | AP000363 | Unassigned | Unassigned | Unassigned | 39_0 | Clustered | 35 | VC_39_0 | 35 | 0.4337 | 1 | 0.4337 | 0 | 0 | 0 | 1 | Ref |
| 201 | AP000422 | Unassigned | Unassigned | Unassigned | 39_0 | Clustered | 35 | VC_39_0 | 35 | 0.4337 | 1 | 0.4337 | 0 | 0 | 0 | 1 | Ref |
| 202 | AP001553 | Unassigned | Unassigned | Unassigned | 12_2 | Clustered | 19 | VC_12_2 | 19 | 0.2018 | 1 | 0.2018 | 0 | 0 | 0 | 1 | Ref |
| 203 | AP004402 | Unassigned | Unassigned | Unassigned | 39_0 | Clustered | 35 | VC_39_0 | 35 | 0.4337 | 1 | 0.4337 | 0 | 0 | 0 | 1 | Ref |
| 204 | AP005153 | Unassigned | Unassigned | Unassigned | 39_0 | Clustered | 35 | VC_39_0 | 35 | 0.4337 | 1 | 0.4337 | 0 | 0 | 0 | 1 | Ref |
| 205 | AP005154 | Unassigned | Unassigned | Unassigned | 39_0 | Clustered | 35 | VC_39_0 | 35 | 0.4337 | 1 | 0.4337 | 0 | 0 | 0 | 1 | Ref |
| 206 | AP008953 | Unassigned | Unassigned | Unassigned | 9_0 | Clustered | 38 | VC_9_0 | 38 | 0.4895 | 1 | 0.4895 | 0 | 0 | 0 | 1 | Ref |
| 207 | AP008954 | Unassigned | Unassigned | Unassigned | 12_0 | Clustered | 21 | VC_12_0 | 21 | 0.2274 | 1 | 0.2274 | 0 | 0 | 0 | 0.9571 | Ref |
| 208 | AP008979 | Unassigned | Unassigned | Unassigned | 113_0 | Clustered | 9 | VC_113_0 | 9 | 0.519 | 1 | 0.519 | 0 | 0 | 0 | 1 | Ref |
| 209 | AP008983 | Unassigned | Unassigned | Unassigned | 211_0 | Clustered/Singleton | 1 | VC_211_0 | 1 | 0 | 1 | 0 | 0 | 0 | 0 | 1 | Ref |
| 210 | AP008986 | Unassigned | Unassigned | Unassigned | 364_0 | Clustered | 16 | VC_364_0 | 16 | 0.6491 | 1 | 0.6491 | 0 | 0 | 0 | 1 | Ref |
| 211 | AP009146 | Unassigned | Unassigned | Unassigned | 63_0 | Clustered | 2 | VC_63_0 | 2 | 0.1147 | 0.95689598 | 0.1098 | 0 | 0 | 0 | 1 | Ref |
| 212 | AP009363 | Unassigned | Unassigned | Unassigned | 8_0 | Clustered | 58 | VC_8_0 | 58 | 0.8306 | 1 | 0.8306 | 0 | 0 | 0 | 0.9946 | Ref |
| 213 | AP009390 | Unassigned | Unassigned | Unassigned | 91_0 | Clustered | 22 | VC_91_0 | 22 | 0.5318 | 1 | 0.5318 | 0 | 0 | 0 | 1 | Ref |
| 214 | AP011113 | Unassigned | Unassigned | Unassigned | 283_10 | Clustered/Singleton | 1 | VC_283_10 | 1 | 0 | 1 | 0 | 0 | 0 | 0 | 1 | Ref |
| 215 | AP011616 | Unassigned | Unassigned | Unassigned | 477_0 | Clustered | 4 | VC_477_0 | 4 | 0.9836 | 0.99748973 | 0.9811 | 0 | 0 | 0 | 1 | Ref |
| 216 | AP011617 | Unassigned | Unassigned | Unassigned | 477_0 | Clustered | 4 | VC_477_0 | 4 | 0.9836 | 0.99748973 | 0.9811 | 0 | 0 | 0 | 1 | Ref |
| 217 | AP011955 | Unassigned | Unassigned | Unassigned | 10_0 | Clustered | 25 | VC_10_0 | 25 | 0.3467 | 1 | 0.3467 | 0 | 0 | 0 | 0.9733 | Ref |
| 218 | AP011956 | Unassigned | Unassigned | Unassigned | 10_0 | Clustered | 25 | VC_10_0 | 25 | 0.3467 | 1 | 0.3467 | 0 | 0 | 0 | 0.9733 | Ref |
| 219 | AP012341 | Unassigned | Unassigned | Unassigned | 8_0 | Clustered | 58 | VC_8_0 | 58 | 0.8306 | 1 | 0.8306 | 0 | 0 | 0 | 0.9946 | Ref |
| 220 | AP012529 | Unassigned | Unassigned | Unassigned | 39_0 | Clustered | 35 | VC_39_0 | 35 | 0.4337 | 1 | 0.4337 | 0 | 0 | 0 | 1 | Ref |
| 221 | AP012530 | Unassigned | Unassigned | Unassigned | 41_1 | Clustered | 10 | VC_41_1 | 10 | 0.1178 | 1 | 0.1178 | 0 | 0 | 0 | 0.8222 | Ref |
| 222 | AP012531 | Unassigned | Unassigned | Unassigned | 39_0 | Clustered | 35 | VC_39_0 | 35 | 0.4337 | 1 | 0.4337 | 0 | 0 | 0 | 1 | Ref |
| 223 | AP012532 | Unassigned | Unassigned | Unassigned | 39_0 | Clustered | 35 | VC_39_0 | 35 | 0.4337 | 1 | 0.4337 | 0 | 0 | 0 | 1 | Ref |
| 224 | AP012534 | Unassigned | Unassigned | Unassigned | 42_0 | Clustered | 49 | VC_42_0 | 49 | 0.4517 | 1 | 0.4517 | 0 | 0 | 0 | 0.8597 | Ref |
| 225 | AP012535 | Unassigned | Unassigned | Unassigned | 39_0 | Clustered | 35 | VC_39_0 | 35 | 0.4337 | 1 | 0.4337 | 0 | 0 | 0 | 1 | Ref |
| 226 | AP012538 | Unassigned | Unassigned | Unassigned | 41_1 | Clustered | 10 | VC_41_1 | 10 | 0.1178 | 1 | 0.1178 | 0 | 0 | 0 | 0.8222 | Ref |
| 227 | AP012539 | Unassigned | Unassigned | Unassigned | 41_1 | Clustered | 10 | VC_41_1 | 10 | 0.1178 | 1 | 0.1178 | 0 | 0 | 0 | 0.8222 | Ref |
| 228 | AP012540 | Unassigned | Unassigned | Unassigned | 41_1 | Clustered | 10 | VC_41_1 | 10 | 0.1178 | 1 | 0.1178 | 0 | 0 | 0 | 0.8222 | Ref |
| 229 | AP013029 | Unassigned | Unassigned | Unassigned | 85_3 | Clustered | 19 | VC_85_3 | 19 | 0.3671 | 1 | 0.3671 | 0 | 0 | 0 | 0.807 | Ref |
| 230 | AP013538 | Unassigned | Unassigned | Unassigned | 340_0 | Clustered | 31 | VC_340_0 | 31 | 0.9549 | 1 | 0.9549 | 0 | 0 | 0 | 1 | Ref |
| 231 | AP013539 | Unassigned | Unassigned | Unassigned | 340_0 | Clustered | 31 | VC_340_0 | 31 | 0.9549 | 1 | 0.9549 | 0 | 0 | 0 | 1 | Ref |
| 232 | AP013541 | Unassigned | Unassigned | Unassigned | 480_0 | Clustered | 19 | VC_480_0 | 19 | 0.5445 | 1 | 0.5445 | 0 | 0 | 0 | 1 | Ref |
| 233 | AP013542 | Unassigned | Unassigned | Unassigned | 480_0 | Clustered | 19 | VC_480_0 | 19 | 0.5445 | 1 | 0.5445 | 0 | 0 | 0 | 1 | Ref |
| 234 | AP013544 | Unassigned | Unassigned | Unassigned | 481_0 | Clustered | 8 | VC_481_0 | 8 | 0.1348 | 1 | 0.1348 | 0 | 0 | 0 | 1 | Ref |
| 235 | AP013545 | Unassigned | Unassigned | Unassigned | 480_0 | Clustered | 19 | VC_480_0 | 19 | 0.5445 | 1 | 0.5445 | 0 | 0 | 0 | 1 | Ref |
| 236 | AP013546 | Unassigned | Unassigned | Unassigned | 482_0 | Clustered | 4 | VC_482_0 | 4 | 0.2975 | 0.99998416 | 0.2975 | 0 | 0 | 0 | 1 | Ref |
| 237 | AP013547 | Unassigned | Unassigned | Unassigned | 482_0 | Clustered | 4 | VC_482_0 | 4 | 0.2975 | 0.99998416 | 0.2975 | 0 | 0 | 0 | 1 | Ref |
| 238 | AP013548 | Unassigned | Unassigned | Unassigned | 482_0 | Clustered | 4 | VC_482_0 | 4 | 0.2975 | 0.99998416 | 0.2975 | 0 | 0 | 0 | 1 | Ref |
| 239 | AP014714 | Unassigned | Unassigned | Unassigned | 490_0 | Clustered | 2 | VC_490_0 | 2 | 0.0102 | 0.95344863 | 0.0097 | 0 | 0 | 0 | 1 | Ref |
| 240 | AP014715 | Unassigned | Unassigned | Unassigned | 490_0 | Clustered | 2 | VC_490_0 | 2 | 0.0102 | 0.95344863 | 0.0097 | 0 | 0 | 0 | 1 | Ref |
| 241 | AP014858 | Unassigned | Unassigned | Unassigned | 493_5 | Clustered/Singleton | 1 | VC_493_5 | 1 | 0 | 1 | 0 | 0 | 0 | 0 | 1 | Ref |
| 242 | AP014887 | Unassigned | Unassigned | Unassigned | 498_0 | Clustered | 8 | VC_498_0 | 8 | 0.3987 | 1 | 0.3987 | 0 | 0 | 0 | 1 | Ref |
| 243 | AP014888 | Unassigned | Unassigned | Unassigned | 501_0 | Clustered | 16 | VC_501_0 | 16 | 0.9985 | 1 | 0.9985 | 0 | 0 | 0 | 1 | Ref |
| 244 | AP017924 | Unassigned | Unassigned | Unassigned | 345_0 | Clustered | 2 | VC_345_0 | 2 | 0.0685 | 0.95654153 | 0.0655 | 0 | 0 | 0 | 1 | Ref |
| 245 | AP017925 | Unassigned | Unassigned | Unassigned | 345_0 | Clustered | 2 | VC_345_0 | 2 | 0.0685 | 0.95654153 | 0.0655 | 0 | 0 | 0 | 1 | Ref |
| 246 | AP018361 | Unassigned | Unassigned | Unassigned | 272_0 | Clustered | 11 | VC_272_0 | 11 | 0.4915 | 1 | 0.4915 | 0 | 0 | 0 | 0.8727 | Ref |
| 247 | AP018375 | Unassigned | Unassigned | Unassigned | 149_9 | Clustered | 3 | VC_149_9 | 3 | 0.0084 | 0.97854556 | 0.0082 | 0 | 0 | 0 | 1 | Ref |
| 248 | AP018399 | Unassigned | Unassigned | Unassigned | 68_0 | Clustered/Singleton | 1 | VC_68_0 | 1 | 0 | 1 | 0 | 0 | 0 | 0 | 1 | Ref |
| 249 | AP018462 | Unassigned | Unassigned | Unassigned | 507_0 | Clustered | 8 | VC_507_0 | 8 | 0.4466 | 1 | 0.4466 | 0 | 0 | 0 | 1 | Ref |
| 250 | AP018463 | Unassigned | Unassigned | Unassigned | 507_0 | Clustered | 8 | VC_507_0 | 8 | 0.4466 | 1 | 0.4466 | 0 | 0 | 0 | 1 | Ref |
| 251 | AP018469 | Unassigned | Unassigned | Unassigned | 234_0 | Clustered | 21 | VC_234_0 | 21 | 0.8355 | 1 | 0.8355 | 0 | 0 | 0 | 1 | Ref |
| 252 | AP018470 | Unassigned | Unassigned | Unassigned | 234_0 | Clustered | 21 | VC_234_0 | 21 | 0.8355 | 1 | 0.8355 | 0 | 0 | 0 | 1 | Ref |
| 253 | AP018471 | Unassigned | Unassigned | Unassigned | 234_0 | Clustered | 21 | VC_234_0 | 21 | 0.8355 | 1 | 0.8355 | 0 | 0 | 0 | 1 | Ref |
| 254 | AP018476 | Unassigned | Unassigned | Unassigned | 215_1 | Clustered | 19 | VC_215_1 | 19 | 0.4188 | 1 | 0.4188 | 0 | 0 | 0 | 1 | Ref |
| 255 | AP018477 | Unassigned | Unassigned | Unassigned | 325_49 | Clustered | 4 | VC_325_49 | 4 | 0.014 | 0.99995965 | 0.014 | 0 | 0 | 0 | 1 | Ref |
| 256 | AP018478 | Unassigned | Unassigned | Unassigned | 325_49 | Clustered | 4 | VC_325_49 | 4 | 0.014 | 0.99995965 | 0.014 | 0 | 0 | 0 | 1 | Ref |
| 257 | AP018479 | Unassigned | Unassigned | Unassigned | 216_6 | Clustered | 42 | VC_216_6 | 42 | 0.473 | 1 | 0.473 | 0 | 0 | 0 | 0.8351 | Ref |
| 258 | AP018480 | Unassigned | Unassigned | Unassigned | 215_1 | Clustered | 19 | VC_215_1 | 19 | 0.4188 | 1 | 0.4188 | 0 | 0 | 0 | 1 | Ref |
| 259 | AP018485 | Unassigned | Unassigned | Unassigned | 513_0 | Clustered | 32 | VC_513_0 | 32 | 0.6881 | 1 | 0.6881 | 0 | 0 | 0 | 1 | Ref |
| 260 | AP018487 | Unassigned | Unassigned | Unassigned | 518_0 | Clustered | 6 | VC_518_0 | 6 | 0.1295 | 1 | 0.1295 | 0 | 0 | 0 | 1 | Ref |
| 261 | AP018714 | Unassigned | Unassigned | Unassigned | 91_0 | Clustered | 22 | VC_91_0 | 22 | 0.5318 | 1 | 0.5318 | 0 | 0 | 0 | 1 | Ref |
| 262 | AP018715 | Unassigned | Unassigned | Unassigned | 91_0 | Clustered | 22 | VC_91_0 | 22 | 0.5318 | 1 | 0.5318 | 0 | 0 | 0 | 1 | Ref |
| 263 | AP018813 | Unassigned | Unassigned | Unassigned | 283_92 | Clustered | 4 | VC_283_92 | 4 | 0.0079 | 0.9999582 | 0.0079 | 0 | 0 | 0 | 1 | Ref |
| 264 | AP018814 | Unassigned | Unassigned | Unassigned | 283_88 | Clustered | 2 | VC_283_88 | 2 | 0.0026 | 0.95596547 | 0.0024 | 0 | 0 | 0 | 1 | Ref |
| 265 | AP018815 | Unassigned | Unassigned | Unassigned | 79_0 | Clustered | 64 | VC_79_0 | 64 | 0.4185 | 1 | 0.4185 | 0 | 0 | 0 | 0.7649 | Ref |
| 266 | AP018932 | Unassigned | Unassigned | Unassigned | 283_187 | Clustered/Singleton | 1 | VC_283_187 | 1 | 0 | 1 | 0 | 0 | 0 | 0 | 1 | Ref |
| 267 | AP019417 | Unassigned | Unassigned | Unassigned | 530_0 | Clustered | 9 | VC_530_0 | 9 | 0.8027 | 1 | 0.8027 | 0 | 0 | 0 | 1 | Ref |
| 268 | AP019418 | Unassigned | Unassigned | Unassigned | 344_0 | Clustered | 21 | VC_344_0 | 21 | 0.5436 | 1 | 0.5436 | 0 | 0 | 0 | 1 | Ref |
| 269 | AP019522 | Unassigned | Unassigned | Unassigned | 153_0 | Clustered | 25 | VC_153_0 | 25 | 0.7729 | 1 | 0.7729 | 0 | 0 | 0 | 1 | Ref |
| 270 | AP019524 | Unassigned | Unassigned | Unassigned | 532_0 | Clustered | 2 | VC_532_0 | 2 | 0.4046 | 0.89435023 | 0.3619 | 0 | 0 | 0 | 1 | Ref |
| 271 | AP019525 | Unassigned | Unassigned | Unassigned | 532_0 | Clustered | 2 | VC_532_0 | 2 | 0.4046 | 0.89435023 | 0.3619 | 0 | 0 | 0 | 1 | Ref |
| 272 | AP019527 | Unassigned | Unassigned | Unassigned | 534_0 | Clustered | 9 | VC_534_0 | 9 | 0.99 | 1 | 0.99 | 0 | 0 | 0 | 1 | Ref |
| 273 | AP019535 | Unassigned | Unassigned | Unassigned | 79_10 | Clustered/Singleton | 1 | VC_79_10 | 1 | 0 | 1 | 0 | 0 | 0 | 0 | 1 | Ref |
| 274 | AP019559 | Unassigned | Unassigned | Unassigned | 190_17 | Clustered/Singleton | 1 | VC_190_17 | 1 | 0 | 1 | 0 | 0 | 0 | 0 | 1 | Ref |
| 275 | AP019560 | Unassigned | Unassigned | Unassigned | 11_0 | Clustered | 19 | VC_11_0 | 19 | 0.7562 | 1 | 0.7562 | 0 | 0 | 0 | 1 | Ref |
| 276 | AP019561 | Unassigned | Unassigned | Unassigned | 11_0 | Clustered | 19 | VC_11_0 | 19 | 0.7562 | 1 | 0.7562 | 0 | 0 | 0 | 1 | Ref |
| 277 | AP019562 | Unassigned | Unassigned | Unassigned | 11_0 | Clustered | 19 | VC_11_0 | 19 | 0.7562 | 1 | 0.7562 | 0 | 0 | 0 | 1 | Ref |
| 278 | AY007505 | Unassigned | Unassigned | Unassigned | 317_2 | Clustered | 20 | VC_317_2 | 20 | 0.1976 | 1 | 0.1976 | 0 | 0 | 0 | 0.6842 | Ref |
| 279 | AY027935 | Unassigned | Unassigned | Unassigned | 5_0 | Clustered | 30 | VC_5_0 | 30 | 0.9647 | 1 | 0.9647 | 0 | 0 | 0 | 1 | Ref |
| 280 | AY029185 | Unassigned | Unassigned | Unassigned | 278_0 | Clustered | 3 | VC_278_0 | 3 | 0.3052 | 0.99848877 | 0.3047 | 0 | 0 | 0 | 1 | Ref |
| 281 | AY048850 | Unassigned | Unassigned | Unassigned | 539_0 | Clustered | 3 | VC_539_0 | 3 | 1 | 1 | 1 | 0 | 0 | 0 | 1 | Ref |
| 282 | AY052766 | Unassigned | Unassigned | Unassigned | 28_1 | Clustered | 53 | VC_28_1 | 53 | 0.7164 | 1 | 0.7164 | 0 | 0 | 0 | 0.9325 | Ref |
| 283 | AY055382 | Unassigned | Unassigned | Unassigned | 45_0 | Clustered | 12 | VC_45_0 | 12 | 0.35 | 1 | 0.35 | 0 | 0 | 0 | 1 | Ref |
| 284 | AY078382 | Unassigned | Unassigned | Unassigned | 472_0 | Clustered | 47 | VC_472_0 | 47 | 0.9025 | 1 | 0.9025 | 0 | 0 | 0 | 1 | Ref |
| 285 | AY129330 | Unassigned | Unassigned | Unassigned | 520_96 | Clustered/Singleton | 1 | VC_520_96 | 1 | 0 | 1 | 0 | 0 | 0 | 0 | 1 | Ref |
| 286 | AY129331 | Unassigned | Unassigned | Unassigned | 551_7 | Clustered | 28 | VC_551_7 | 28 | 0.1661 | 1 | 0.1661 | 0 | 0 | 0 | 0.8519 | Ref |
| 287 | AY129332 | Unassigned | Unassigned | Unassigned | 514_0 | Clustered | 75 | VC_514_0 | 75 | 0.9402 | 1 | 0.9402 | 0 | 0 | 0 | 1 | Ref |
| 288 | AY129333 | Unassigned | Unassigned | Unassigned | 518_1 | Clustered | 2 | VC_518_1 | 2 | 0.0376 | 0.95718761 | 0.036 | 0 | 0 | 0 | 1 | Ref |
| 289 | AY129334 | Unassigned | Unassigned | Unassigned | 508_0 | Clustered | 32 | VC_508_0 | 32 | 0.906 | 1 | 0.906 | 0 | 0 | 0 | 1 | Ref |
| 290 | AY129335 | Unassigned | Unassigned | Unassigned | 556_0 | Clustered | 21 | VC_556_0 | 21 | 0.9912 | 1 | 0.9912 | 0 | 0 | 0 | 1 | Ref |
| 291 | AY129337 | Unassigned | Unassigned | Unassigned | 558_8 | Clustered | 2 | VC_558_8 | 2 | 0.0032 | 0.68030512 | 0.0022 | 0 | 0 | 0 | 1 | Ref |
| 292 | AY129338 | Unassigned | Unassigned | Unassigned | 546_10 | Clustered/Singleton | 1 | VC_546_10 | 1 | 0 | 1 | 0 | 0 | 0 | 0 | 1 | Ref |
| 293 | AY129339 | Unassigned | Unassigned | Unassigned | 566_0 | Clustered | 2 | VC_566_0 | 2 | 0.0989 | 0.95622526 | 0.0945 | 0 | 0 | 0 | 1 | Ref |
| 294 | AY131267 | Unassigned | Unassigned | Unassigned | 272_0 | Clustered | 11 | VC_272_0 | 11 | 0.4915 | 1 | 0.4915 | 0 | 0 | 0 | 0.8727 | Ref |
| 295 | AY133112 | Unassigned | Unassigned | Unassigned | 23_0 | Clustered | 3 | VC_23_0 | 3 | 0.4305 | 0.99837378 | 0.4298 | 0 | 0 | 0 | 1 | Ref |
| 296 | AY135486 | Unassigned | Unassigned | Unassigned | 228_0 | Clustered | 60 | VC_228_0 | 60 | 0.942 | 1 | 0.942 | 0 | 0 | 0 | 1 | Ref |
| 297 | AY135739 | Unassigned | Unassigned | Unassigned | 228_0 | Clustered | 60 | VC_228_0 | 60 | 0.942 | 1 | 0.942 | 0 | 0 | 0 | 1 | Ref |
| 298 | AY150271 | Unassigned | Unassigned | Unassigned | 476_0 | Clustered | 19 | VC_476_0 | 19 | 0.7814 | 1 | 0.7814 | 0 | 0 | 0 | 1 | Ref |
| 299 | AY176327 | Unassigned | Unassigned | Unassigned | 149_8 | Clustered | 66 | VC_149_8 | 66 | 0.4565 | 1 | 0.4565 | 0 | 0 | 0 | 0.8182 | Ref |
| 300 | AY190604 | Unassigned | Unassigned | Unassigned | 310_0 | Clustered | 33 | VC_310_0 | 33 | 0.8771 | 1 | 0.8771 | 0 | 0 | 0 | 1 | Ref |
| 301 | AY212251 | Unassigned | Unassigned | Unassigned | 97_0 | Clustered | 5 | VC_97_0 | 5 | 0.0696 | 0.98113835 | 0.0683 | 0 | 0 | 0 | 1 | Ref |
| 302 | AY216660 | Unassigned | Unassigned | Unassigned | 575_0 | Clustered | 35 | VC_575_0 | 35 | 0.8815 | 1 | 0.8815 | 0 | 0 | 0 | 1 | Ref |
| 303 | AY236756 | Unassigned | Unassigned | Unassigned | 577_0 | Clustered | 6 | VC_577_0 | 6 | 0.9267 | 0.99999993 | 0.9267 | 0 | 0 | 0 | 1 | Ref |
| 304 | AY242528 | Unassigned | Unassigned | Unassigned | 16_0 | Clustered | 12 | VC_16_0 | 12 | 0.6615 | 1 | 0.6615 | 0 | 0 | 0 | 1 | Ref |
| 305 | AY247822 | Unassigned | Unassigned | Unassigned | 392_0 | Clustered | 126 | VC_392_0 | 126 | 0.9621 | 1 | 0.9621 | 0 | 0 | 0 | 1 | Ref |
| 306 | AY251033 | Unassigned | Unassigned | Unassigned | 228_0 | Clustered | 60 | VC_228_0 | 60 | 0.942 | 1 | 0.942 | 0 | 0 | 0 | 1 | Ref |
| 307 | AY257527 | Unassigned | Unassigned | Unassigned | 424_0 | Clustered | 7 | VC_424_0 | 7 | 0.9044 | 0.9998143 | 0.9043 | 0 | 0 | 0 | 1 | Ref |
| 308 | AY264774 | Unassigned | Unassigned | Unassigned | 392_0 | Clustered | 126 | VC_392_0 | 126 | 0.9621 | 1 | 0.9621 | 0 | 0 | 0 | 1 | Ref |
| 309 | AY264775 | Unassigned | Unassigned | Unassigned | 392_0 | Clustered | 126 | VC_392_0 | 126 | 0.9621 | 1 | 0.9621 | 0 | 0 | 0 | 1 | Ref |
| 310 | AY264776 | Unassigned | Unassigned | Unassigned | 392_0 | Clustered | 126 | VC_392_0 | 126 | 0.9621 | 1 | 0.9621 | 0 | 0 | 0 | 1 | Ref |
| 311 | AY264777 | Unassigned | Unassigned | Unassigned | 392_0 | Clustered | 126 | VC_392_0 | 126 | 0.9621 | 1 | 0.9621 | 0 | 0 | 0 | 1 | Ref |
| 312 | AY264778 | Unassigned | Unassigned | Unassigned | 392_0 | Clustered | 126 | VC_392_0 | 126 | 0.9621 | 1 | 0.9621 | 0 | 0 | 0 | 1 | Ref |
| 313 | AY283928 | Unassigned | Unassigned | Unassigned | 531_1 | Clustered | 6 | VC_531_1 | 6 | 0.1481 | 0.99978665 | 0.148 | 0 | 0 | 0 | 1 | Ref |
| 314 | AY288927 | Unassigned | Unassigned | Unassigned | 452_0 | Clustered | 66 | VC_452_0 | 66 | 0.9248 | 1 | 0.9248 | 0 | 0 | 0 | 1 | Ref |
| 315 | AY299121 | Unassigned | Unassigned | Unassigned | 113_0 | Clustered | 9 | VC_113_0 | 9 | 0.519 | 1 | 0.519 | 0 | 0 | 0 | 1 | Ref |
| 316 | AY303349 | Unassigned | Unassigned | Unassigned | 286_10 | Clustered | 9 | VC_286_10 | 9 | 0.0615 | 0.99999878 | 0.0615 | 0 | 0 | 0 | 0.6667 | Ref |
| 317 | AY308796 | Unassigned | Unassigned | Unassigned | 584_0 | Clustered | 28 | VC_584_0 | 28 | 0.7704 | 1 | 0.7704 | 0 | 0 | 0 | 1 | Ref |
| 318 | AY319521 | Unassigned | Unassigned | Unassigned | 6_0 | Clustered | 44 | VC_6_0 | 44 | 0.908 | 1 | 0.908 | 0 | 0 | 0 | 1 | Ref |
| 319 | AY320035 | Unassigned | Unassigned | Unassigned | 587_0 | Clustered | 2 | VC_587_0 | 2 | 0.3252 | 0.91192888 | 0.2965 | 0 | 0 | 0 | 1 | Ref |
| 320 | AY324828 | Unassigned | Unassigned | Unassigned | 589_0 | Clustered | 2 | VC_589_0 | 2 | 1 | 1 | 1 | 0 | 0 | 0 | 1 | Ref |
| 321 | AY328852 | Unassigned | Unassigned | Unassigned | 498_0 | Clustered | 8 | VC_498_0 | 8 | 0.3987 | 1 | 0.3987 | 0 | 0 | 0 | 1 | Ref |
| 322 | AY328853 | Unassigned | Unassigned | Unassigned | 498_0 | Clustered | 8 | VC_498_0 | 8 | 0.3987 | 1 | 0.3987 | 0 | 0 | 0 | 1 | Ref |

| | | | | | | | | | | | | | | | | | |
|---|---|---|---|---|---|---|---|---|---|---|---|---|---|---|---|---|---|
| 323 | AY343333 | Unassigned | Unassigned | Unassigned | 590_0 | Clustered | 27 | VC_590_0 | 27 | 0.9947 | 1 | 0.9947 | 0 | 0 | 0 | 1 | Ref |
| 324 | AY349011 | Unassigned | Unassigned | Unassigned | 162_0 | Clustered | 18 | VC_162_0 | 18 | 0.7403 | 1 | 0.7403 | 0 | 0 | 0 | 0.8627 | Ref |
| 325 | AY370673 | Unassigned | Unassigned | Unassigned | 452_0 | Clustered | 66 | VC_452_0 | 66 | 0.9248 | 1 | 0.9248 | 0 | 0 | 0 | 1 | Ref |
| 326 | AY370674 | Unassigned | Unassigned | Unassigned | 452_0 | Clustered | 66 | VC_452_0 | 66 | 0.9248 | 1 | 0.9248 | 0 | 0 | 0 | 1 | Ref |
| 327 | AY370762 | Unassigned | Unassigned | Unassigned | 599_0 | Clustered | 50 | VC_599_0 | 50 | 0.9989 | 1 | 0.9989 | 0 | 0 | 0 | 1 | Ref |
| 328 | AY374448 | Unassigned | Unassigned | Unassigned | 34_0 | Clustered | 4 | VC_34_0 | 4 | 0.4349 | 0.99998062 | 0.4349 | 0 | 0 | 0 | 1 | Ref |
| 329 | AY375531 | Unassigned | Unassigned | Unassigned | 600_1 | Clustered | 7 | VC_600_1 | 7 | 0.1837 | 0.99999997 | 0.1837 | 0 | 0 | 0 | 1 | Ref |
| 330 | AY388628 | Unassigned | Unassigned | Unassigned | 599_0 | Clustered | 50 | VC_599_0 | 50 | 0.9989 | 1 | 0.9989 | 0 | 0 | 0 | 1 | Ref |
| 331 | AY394005 | Unassigned | Unassigned | Unassigned | 179_0 | Clustered | 53 | VC_179_0 | 53 | 0.9137 | 1 | 0.9137 | 0 | 0 | 0 | 1 | Ref |
| 332 | AY423772 | Unassigned | Unassigned | Unassigned | 599_0 | Clustered | 50 | VC_599_0 | 50 | 0.9989 | 1 | 0.9989 | 0 | 0 | 0 | 1 | Ref |
| 333 | AY441783 | Unassigned | Unassigned | Unassigned | 603_0 | Clustered | 9 | VC_603_0 | 9 | 0.9925 | 0.99999994 | 0.9925 | 0 | 0 | 0 | 1 | Ref |
| 334 | AY453853 | Unassigned | Unassigned | Unassigned | 353_0 | Clustered | 6 | VC_353_0 | 6 | 0.545 | 1 | 0.545 | 0 | 0 | 0 | 1 | Ref |
| 335 | AY500152 | Unassigned | Unassigned | Unassigned | 325_5 | Clustered | 11 | VC_325_5 | 11 | 0.047 | 1 | 0.047 | 0 | 0 | 0 | 0.7636 | Ref |
| 336 | AY500153 | Unassigned | Unassigned | Unassigned | 325_9 | Clustered | 2 | VC_325_9 | 2 | 0.0055 | 0.95750934 | 0.0053 | 0 | 0 | 0 | 1 | Ref |
| 337 | AY505112 | Unassigned | Unassigned | Unassigned | 607_0 | Clustered | 16 | VC_607_0 | 16 | 0.9179 | 1 | 0.9179 | 0 | 0 | 0 | 1 | Ref |
| 338 | AY508486 | Unassigned | Unassigned | Unassigned | 10_0 | Clustered | 25 | VC_10_0 | 25 | 0.3467 | 1 | 0.3467 | 0 | 0 | 0 | 0.9733 | Ref |
| 339 | AY510084 | Unassigned | Unassigned | Unassigned | 607_0 | Clustered | 16 | VC_607_0 | 16 | 0.9179 | 1 | 0.9179 | 0 | 0 | 0 | 1 | Ref |
| 340 | AY526908 | Unassigned | Unassigned | Unassigned | 278_0 | Clustered | 3 | VC_278_0 | 3 | 0.3052 | 0.99848877 | 0.3047 | 0 | 0 | 0 | 1 | Ref |
| 341 | AY526909 | Unassigned | Unassigned | Unassigned | 278_0 | Clustered | 3 | VC_278_0 | 3 | 0.3052 | 0.99848877 | 0.3047 | 0 | 0 | 0 | 1 | Ref |
| 342 | AY539836 | Unassigned | Unassigned | Unassigned | 250_0 | Clustered | 4 | VC_250_0 | 4 | 0.2395 | 0.99998241 | 0.2395 | 0 | 0 | 0 | 1 | Ref |
| 343 | AY543070 | Unassigned | Unassigned | Unassigned | 190_38 | Clustered | 3 | VC_190_38 | 3 | 0.0175 | 0.99847952 | 0.0174 | 0 | 0 | 0 | 1 | Ref |
| 344 | AY569307 | Unassigned | Unassigned | Unassigned | 611_0 | Clustered | 2 | VC_611_0 | 2 | 1 | 1 | 1 | 0 | 0 | 0 | 1 | Ref |
| 345 | AY575774 | Unassigned | Unassigned | Unassigned | 612_0 | Clustered | 9 | VC_612_0 | 9 | 0.9545 | 1 | 0.9545 | 0 | 0 | 0 | 1 | Ref |
| 346 | AY583234 | Unassigned | Unassigned | Unassigned | 246_0 | Clustered | 3 | VC_246_0 | 3 | 0.7624 | 0.9595722 | 0.7316 | 0 | 0 | 0 | 1 | Ref |
| 347 | AY583235 | Unassigned | Unassigned | Unassigned | 246_0 | Clustered | 3 | VC_246_0 | 3 | 0.7624 | 0.9595722 | 0.7316 | 0 | 0 | 0 | 1 | Ref |
| 348 | AY583236 | Unassigned | Unassigned | Unassigned | 246_0 | Clustered | 3 | VC_246_0 | 3 | 0.7624 | 0.9595722 | 0.7316 | 0 | 0 | 0 | 1 | Ref |
| 349 | AY587007 | Unassigned | Unassigned | Unassigned | 190_38 | Clustered | 3 | VC_190_38 | 3 | 0.0175 | 0.99847952 | 0.0174 | 0 | 0 | 0 | 1 | Ref |
| 350 | AY605066 | Unassigned | Unassigned | Unassigned | 272_0 | Clustered | 11 | VC_272_0 | 11 | 0.4915 | 1 | 0.4915 | 0 | 0 | 0 | 0.8727 | Ref |
| 351 | AY605181 | Unassigned | Unassigned | Unassigned | 279_0 | Clustered | 2 | VC_279_0 | 2 | 0.1888 | 0.95124665 | 0.1796 | 0 | 0 | 0 | 1 | Ref |
| 352 | AY625898 | Unassigned | Unassigned | Unassigned | 630_0 | Clustered | 20 | VC_630_0 | 20 | 0.8513 | 1 | 0.8513 | 0 | 0 | 0 | 1 | Ref |
| 353 | AY682195 | Unassigned | Unassigned | Unassigned | 526_2 | Clustered/Singleton | 1 | VC_526_2 | 1 | 0 | 1 | 0 | 0 | 0 | 0 | 1 | Ref |
| 354 | AY692264 | Unassigned | Unassigned | Unassigned | 190_38 | Clustered | 3 | VC_190_38 | 3 | 0.0175 | 0.99847952 | 0.0174 | 0 | 0 | 0 | 1 | Ref |
| 355 | AY699705 | Unassigned | Unassigned | Unassigned | 254_0 | Clustered | 46 | VC_254_0 | 46 | 0.5159 | 1 | 0.5159 | 0 | 0 | 0 | 1 | Ref |
| 356 | AY701338 | Unassigned | Unassigned | Unassigned | 424_0 | Clustered | 7 | VC_424_0 | 7 | 0.9044 | 0.9998143 | 0.9043 | 0 | 0 | 0 | 1 | Ref |
| 357 | AY714348 | Unassigned | Unassigned | Unassigned | 26_0 | Clustered | 4 | VC_26_0 | 4 | 0.7433 | 0.99936153 | 0.7428 | 0 | 0 | 0 | 1 | Ref |
| 358 | AY722806 | Unassigned | Unassigned | Unassigned | 440_0 | Clustered | 4 | VC_440_0 | 4 | 1 | 1 | 1 | 0 | 0 | 0 | 1 | Ref |
| 359 | AY730274 | Unassigned | Unassigned | Unassigned | 637_0 | Clustered | 115 | VC_637_0 | 115 | 0.731 | 1 | 0.731 | 0 | 0 | 0 | 0.8764 | Ref |
| 360 | AY739900 | Unassigned | Unassigned | Unassigned | 640_0 | Clustered | 7 | VC_640_0 | 7 | 0.9025 | 1 | 0.9025 | 0 | 0 | 0 | 1 | Ref |
| 361 | AY751298 | Unassigned | Unassigned | Unassigned | 360_0 | Clustered | 358 | VC_360_0 | 358 | 0.9615 | 1 | 0.9615 | 0 | 0 | 0 | 1 | Ref |
| 362 | AY769964 | Unassigned | Unassigned | Unassigned | 198_0 | Clustered | 28 | VC_198_0 | 28 | 0.2429 | 0.99971175 | 0.2429 | 0 | 0 | 0 | 1 | Ref |
| 363 | AY772740 | Unassigned | Unassigned | Unassigned | 642_2 | Clustered | 6 | VC_642_2 | 6 | 0.1679 | 1 | 0.1679 | 0 | 0 | 0 | 1 | Ref |
| 364 | AY848684 | Unassigned | Unassigned | Unassigned | 603_0 | Clustered | 9 | VC_603_0 | 9 | 0.9925 | 0.99999994 | 0.9925 | 0 | 0 | 0 | 1 | Ref |
| 365 | AY848685 | Unassigned | Unassigned | Unassigned | 603_0 | Clustered | 9 | VC_603_0 | 9 | 0.9925 | 0.99999994 | 0.9925 | 0 | 0 | 0 | 1 | Ref |
| 366 | AY848686 | Unassigned | Unassigned | Unassigned | 603_0 | Clustered | 9 | VC_603_0 | 9 | 0.9925 | 0.99999994 | 0.9925 | 0 | 0 | 0 | 1 | Ref |
| 367 | AY848687 | Unassigned | Unassigned | Unassigned | 603_0 | Clustered | 9 | VC_603_0 | 9 | 0.9925 | 0.99999994 | 0.9925 | 0 | 0 | 0 | 1 | Ref |
| 368 | AY848688 | Unassigned | Unassigned | Unassigned | 603_0 | Clustered | 9 | VC_603_0 | 9 | 0.9925 | 0.99999994 | 0.9925 | 0 | 0 | 0 | 1 | Ref |
| 369 | AY848689 | Unassigned | Unassigned | Unassigned | 603_0 | Clustered | 9 | VC_603_0 | 9 | 0.9925 | 0.99999994 | 0.9925 | 0 | 0 | 0 | 1 | Ref |
| 370 | AY855346 | Unassigned | Unassigned | Unassigned | 139_0 | Clustered | 2 | VC_139_0 | 2 | 0.0701 | 0.95510185 | 0.067 | 0 | 0 | 0 | 1 | Ref |
| 371 | AY939843 | Unassigned | Unassigned | Unassigned | 340_0 | Clustered | 31 | VC_340_0 | 31 | 0.9549 | 1 | 0.9549 | 0 | 0 | 0 | 1 | Ref |
| 372 | AY939844 | Unassigned | Unassigned | Unassigned | 432_9 | Clustered | 3 | VC_432_9 | 3 | 0.0372 | 0.99851312 | 0.0371 | 0 | 0 | 0 | 1 | Ref |
| 373 | AY940168 | Unassigned | Unassigned | Unassigned | 655_2 | Clustered | 3 | VC_655_2 | 3 | 0.0231 | 0.99860756 | 0.0231 | 0 | 0 | 0 | 1 | Ref |
| 374 | AY950802 | Unassigned | Unassigned | Unassigned | 666_0 | Clustered | 4 | VC_666_0 | 4 | 1 | 1 | 1 | 0 | 0 | 0 | 1 | Ref |
| 375 | AY954949 | Unassigned | Unassigned | Unassigned | 96_0 | Clustered | 30 | VC_96_0 | 30 | 0.718 | 1 | 0.718 | 0 | 0 | 0 | 1 | Ref |
| 376 | AY954951 | Unassigned | Unassigned | Unassigned | 9_0 | Clustered | 38 | VC_9_0 | 38 | 0.4895 | 1 | 0.4895 | 0 | 0 | 0 | 1 | Ref |
| 377 | AY954952 | Unassigned | Unassigned | Unassigned | 9_0 | Clustered | 38 | VC_9_0 | 38 | 0.4895 | 1 | 0.4895 | 0 | 0 | 0 | 1 | Ref |
| 378 | AY954953 | Unassigned | Unassigned | Unassigned | 9_0 | Clustered | 38 | VC_9_0 | 38 | 0.4895 | 1 | 0.4895 | 0 | 0 | 0 | 1 | Ref |
| 379 | AY954954 | Unassigned | Unassigned | Unassigned | 15_1 | Clustered | 9 | VC_15_1 | 9 | 0.182 | 1 | 0.182 | 0 | 0 | 0 | 0.8889 | Ref |
| 380 | AY954955 | Unassigned | Unassigned | Unassigned | 8_0 | Clustered | 58 | VC_8_0 | 58 | 0.8306 | 1 | 0.8306 | 0 | 0 | 0 | 0.9946 | Ref |
| 381 | AY954956 | Unassigned | Unassigned | Unassigned | 8_0 | Clustered | 58 | VC_8_0 | 58 | 0.8306 | 1 | 0.8306 | 0 | 0 | 0 | 0.9946 | Ref |
| 382 | AY954957 | Unassigned | Unassigned | Unassigned | 8_0 | Clustered | 58 | VC_8_0 | 58 | 0.8306 | 1 | 0.8306 | 0 | 0 | 0 | 0.9946 | Ref |
| 383 | AY954960 | Unassigned | Unassigned | Unassigned | 12_0 | Clustered | 21 | VC_12_0 | 21 | 0.2274 | 1 | 0.2274 | 0 | 0 | 0 | 0.9571 | Ref |
| 384 | AY954961 | Unassigned | Unassigned | Unassigned | 12_0 | Clustered | 21 | VC_12_0 | 21 | 0.2274 | 1 | 0.2274 | 0 | 0 | 0 | 0.9571 | Ref |
| 385 | AY954962 | Unassigned | Unassigned | Unassigned | 12_2 | Clustered | 19 | VC_12_2 | 19 | 0.2018 | 1 | 0.2018 | 0 | 0 | 0 | 1 | Ref |
| 386 | AY954963 | Unassigned | Unassigned | Unassigned | 12_2 | Clustered | 19 | VC_12_2 | 19 | 0.2018 | 1 | 0.2018 | 0 | 0 | 0 | 1 | Ref |
| 387 | AY954964 | Unassigned | Unassigned | Unassigned | 12_2 | Clustered | 19 | VC_12_2 | 19 | 0.2018 | 1 | 0.2018 | 0 | 0 | 0 | 1 | Ref |
| 388 | AY954965 | Unassigned | Unassigned | Unassigned | 12_2 | Clustered | 19 | VC_12_2 | 19 | 0.2018 | 1 | 0.2018 | 0 | 0 | 0 | 1 | Ref |
| 389 | AY954966 | Unassigned | Unassigned | Unassigned | 12_2 | Clustered | 19 | VC_12_2 | 19 | 0.2018 | 1 | 0.2018 | 0 | 0 | 0 | 1 | Ref |
| 390 | AY954967 | Unassigned | Unassigned | Unassigned | 12_2 | Clustered | 19 | VC_12_2 | 19 | 0.2018 | 1 | 0.2018 | 0 | 0 | 0 | 1 | Ref |
| 391 | AY954968 | Unassigned | Unassigned | Unassigned | 12_2 | Clustered | 19 | VC_12_2 | 19 | 0.2018 | 1 | 0.2018 | 0 | 0 | 0 | 1 | Ref |
| 392 | AY954969 | Unassigned | Unassigned | Unassigned | 149_8 | Clustered | 66 | VC_149_8 | 66 | 0.4565 | 1 | 0.4565 | 0 | 0 | 0 | 0.8182 | Ref |
| 393 | AY962392 | Unassigned | Unassigned | Unassigned | 600_1 | Clustered | 7 | VC_600_1 | 7 | 0.1837 | 0.99999997 | 0.1837 | 0 | 0 | 0 | 1 | Ref |
| 394 | AY967407 | Unassigned | Unassigned | Unassigned | 669_1 | Clustered | 10 | VC_669_1 | 10 | 0.1268 | 1 | 0.1268 | 0 | 0 | 0 | 0.9778 | Ref |
| 395 | B1U32222 | Unassigned | Unassigned | Unassigned | 228_0 | Clustered | 60 | VC_228_0 | 60 | 0.942 | 1 | 0.942 | 0 | 0 | 0 | 1 | Ref |
| 396 | BCP1 | Unassigned | Unassigned | Unassigned | 672_0 | Clustered | 12 | VC_672_0 | 12 | 1 | 1 | 1 | 0 | 0 | 0 | 1 | Ref |
| 397 | BHU24159 | Unassigned | Unassigned | Unassigned | 5_0 | Clustered | 30 | VC_5_0 | 30 | 0.9647 | 1 | 0.9647 | 0 | 0 | 0 | 1 | Ref |
| 398 | BK000583 | Unassigned | Unassigned | Unassigned | 28_1 | Clustered | 53 | VC_28_1 | 53 | 0.7164 | 1 | 0.7164 | 0 | 0 | 0 | 0.9325 | Ref |
| 399 | BK010471 | Unassigned | Unassigned | Unassigned | 673_0 | Clustered | 15 | VC_673_0 | 15 | 0.8234 | 1 | 0.8234 | 0 | 0 | 0 | 0.7619 | Ref |
| 400 | BK010646 | Unassigned | Unassigned | Unassigned | 677_0 | Clustered | 9 | VC_677_0 | 9 | 0.3787 | 1 | 0.3787 | 0 | 0 | 0 | 0.8333 | Ref |
| 401 | BK013337 | Unassigned | Unassigned | Unassigned | 539_0 | Clustered | 3 | VC_539_0 | 3 | 1 | 1 | 1 | 0 | 0 | 0 | 1 | Ref |
| 402 | BK013338 | Unassigned | Unassigned | Unassigned | 539_0 | Clustered | 3 | VC_539_0 | 3 | 1 | 1 | 1 | 0 | 0 | 0 | 1 | Ref |
| 403 | BK013345 | Unassigned | Unassigned | Unassigned | 681_0 | Clustered | 3 | VC_681_0 | 3 | 0.9318 | 0.81445332 | 0.7589 | 0 | 0 | 0 | 1 | Ref |
| 404 | BK013346 | Unassigned | Unassigned | Unassigned | 681_0 | Clustered | 3 | VC_681_0 | 3 | 0.9318 | 0.81445332 | 0.7589 | 0 | 0 | 0 | 1 | Ref |
| 405 | BK013347 | Unassigned | Unassigned | Unassigned | 681_0 | Clustered | 3 | VC_681_0 | 3 | 0.9318 | 0.81445332 | 0.7589 | 0 | 0 | 0 | 1 | Ref |
| 406 | BK059895 | Unassigned | Unassigned | Unassigned | 682_0 | Clustered | 2 | VC_682_0 | 2 | 1 | 1 | 1 | 0 | 0 | 0 | 1 | Ref |
| 407 | BK061308 | Unassigned | Unassigned | Unassigned | 683_0 | Clustered | 5 | VC_683_0 | 5 | 0.8963 | 0.99999974 | 0.8963 | 0 | 0 | 0 | 1 | Ref |
| 408 | BK061309 | Unassigned | Unassigned | Unassigned | 683_0 | Clustered | 5 | VC_683_0 | 5 | 0.8963 | 0.99999974 | 0.8963 | 0 | 0 | 0 | 1 | Ref |
| 409 | BK061310 | Unassigned | Unassigned | Unassigned | 683_0 | Clustered | 5 | VC_683_0 | 5 | 0.8963 | 0.99999974 | 0.8963 | 0 | 0 | 0 | 1 | Ref |
| 410 | BK061311 | Unassigned | Unassigned | Unassigned | 683_0 | Clustered | 5 | VC_683_0 | 5 | 0.8963 | 0.99999974 | 0.8963 | 0 | 0 | 0 | 1 | Ref |
| 411 | BK061312 | Unassigned | Unassigned | Unassigned | 683_0 | Clustered | 5 | VC_683_0 | 5 | 0.8963 | 0.99999974 | 0.8963 | 0 | 0 | 0 | 1 | Ref |
| 412 | C2PVCG | Unassigned | Unassigned | Unassigned | 690_0 | Clustered | 50 | VC_690_0 | 50 | 0.9805 | 1 | 0.9805 | 0 | 0 | 0 | 1 | Ref |
| 413 | CAJCJZ010000002 | Unassigned | Unassigned | Unassigned | 691_1 | Clustered | 5 | VC_691_1 | 5 | 0.4358 | 0.99999993 | 0.4358 | 0 | 0 | 0 | 1 | Ref |
| 414 | CAJDJX010000002 | Unassigned | Unassigned | Unassigned | 695_0 | Clustered | 48 | VC_695_0 | 48 | 0.8162 | 1 | 0.8162 | 0 | 0 | 0 | 1 | Ref |
| 415 | CAJDJY010000002 | Unassigned | Unassigned | Unassigned | 91_0 | Clustered | 22 | VC_91_0 | 22 | 0.5318 | 1 | 0.5318 | 0 | 0 | 0 | 1 | Ref |
| 416 | CAJDJZ010000002 | Unassigned | Unassigned | Unassigned | 91_0 | Clustered | 22 | VC_91_0 | 22 | 0.5318 | 1 | 0.5318 | 0 | 0 | 0 | 1 | Ref |
| 417 | CAJDKA010000002 | Unassigned | Unassigned | Unassigned | 92_0 | Clustered | 22 | VC_92_0 | 22 | 0.5639 | 1 | 0.5639 | 0 | 0 | 0 | 1 | Ref |
| 418 | CAJDKF010000002 | Unassigned | Unassigned | Unassigned | 695_0 | Clustered | 48 | VC_695_0 | 48 | 0.8162 | 1 | 0.8162 | 0 | 0 | 0 | 1 | Ref |
| 419 | CAKLQA020000001 | Unassigned | Unassigned | Unassigned | 696_1 | Clustered | 8 | VC_696_1 | 8 | 0.5452 | 1 | 0.5452 | 0 | 0 | 0 | 0.9286 | Ref |
| 420 | CAKLQA020000002 | Unassigned | Unassigned | Unassigned | 670_0 | Clustered | 2 | VC_670_0 | 2 | 0.6875 | 0.72985431 | 0.5018 | 0 | 0 | 0 | 1 | Ref |
| 421 | CAKLQA020000003 | Unassigned | Unassigned | Unassigned | 615_0 | Clustered | 2 | VC_615_0 | 2 | 0.2668 | 0.94178001 | 0.2513 | 0 | 0 | 0 | 1 | Ref |
| 422 | CAKLQB020000001 | Unassigned | Unassigned | Unassigned | 699_0 | Clustered | 71 | VC_699_0 | 71 | 0.9352 | 1 | 0.9352 | 0 | 0 | 0 | 1 | Ref |
| 423 | CAKLQC020000001 | Unassigned | Unassigned | Unassigned | 492_1 | Clustered | 20 | VC_492_1 | 20 | 0.4176 | 1 | 0.4176 | 0 | 0 | 0 | 1 | Ref |
| 424 | CAKLQD020000001 | Unassigned | Unassigned | Unassigned | 492_1 | Clustered | 20 | VC_492_1 | 20 | 0.4176 | 1 | 0.4176 | 0 | 0 | 0 | 1 | Ref |
| 425 | CAKLQE020000001 | Unassigned | Unassigned | Unassigned | 701_2 | Clustered/Singleton | 1 | VC_701_2 | 1 | 0 | 1 | 0 | 0 | 0 | 0 | 1 | Ref |
| 426 | CAKLQG020000001 | Unassigned | Unassigned | Unassigned | 699_0 | Clustered | 71 | VC_699_0 | 71 | 0.9352 | 1 | 0.9352 | 0 | 0 | 0 | 1 | Ref |
| 427 | CMU41758 | Unassigned | Unassigned | Unassigned | 197_0 | Clustered | 5 | VC_197_0 | 5 | 0.1324 | 0.84698401 | 0.1122 | 0 | 0 | 0 | 1 | Ref |
| 428 | CP000315 | Unassigned | Unassigned | Unassigned | 19_0 | Clustered | 2 | VC_19_0 | 2 | 0.0418 | 0.92044806 | 0.0385 | 0 | 0 | 0 | 1 | Ref |
| 429 | CP000622 | Unassigned | Unassigned | Unassigned | 250_0 | Clustered | 4 | VC_250_0 | 4 | 0.2395 | 0.99998241 | 0.2395 | 0 | 0 | 0 | 1 | Ref |
| 430 | CP000623 | Unassigned | Unassigned | Unassigned | 3_0 | Clustered | 17 | VC_3_0 | 17 | 0.5639 | 1 | 0.5639 | 0 | 0 | 0 | 1 | Ref |
| 431 | CP000624 | Unassigned | Unassigned | Unassigned | 3_0 | Clustered | 17 | VC_3_0 | 17 | 0.5639 | 1 | 0.5639 | 0 | 0 | 0 | 1 | Ref |
| 432 | CP000625 | Unassigned | Unassigned | Unassigned | 353_0 | Clustered | 6 | VC_353_0 | 6 | 0.545 | 1 | 0.545 | 0 | 0 | 0 | 1 | Ref |
| 433 | CP000711 | Unassigned | Unassigned | Unassigned | 30_1 | Clustered | 16 | VC_30_1 | 16 | 0.1973 | 1 | 0.1973 | 0 | 0 | 0 | 0.8333 | Ref |
| 434 | CP000917 | Unassigned | Unassigned | Unassigned | 191_17 | Clustered | 4 | VC_191_17 | 4 | 0.0166 | 0.99930063 | 0.0166 | 0 | 0 | 0 | 0.8333 | Ref |
| 435 | CP002495 | Unassigned | Unassigned | Unassigned | 706_0 | Clustered | 2 | VC_706_0 | 2 | 1 | 1 | 1 | 0 | 0 | 0 | 1 | Ref |
| 436 | CP004084 | Unassigned | Unassigned | Unassigned | 303_0 | Clustered | 155 | VC_303_0 | 155 | 0.9979 | 1 | 0.9979 | 0 | 0 | 0 | 1 | Ref |
| 437 | CP006891 | Unassigned | Unassigned | Unassigned | 55_0 | Clustered | 3 | VC_55_0 | 3 | 0.2818 | 0.99850111 | 0.2813 | 0 | 0 | 0 | 1 | Ref |
| 438 | CP008753 | Unassigned | Unassigned | Unassigned | 3_0 | Clustered | 17 | VC_3_0 | 17 | 0.5639 | 1 | 0.5639 | 0 | 0 | 0 | 1 | Ref |
| 439 | CP011970 | Unassigned | Unassigned | Unassigned | 473_2 | Clustered | 2 | VC_473_2 | 2 | 0.1022 | 0.94602228 | 0.0966 | 0 | 0 | 0 | 1 | Ref |
| 440 | CP013282 | Unassigned | Unassigned | Unassigned | 424_0 | Clustered | 7 | VC_424_0 | 7 | 0.9044 | 0.9998143 | 0.9043 | 0 | 0 | 0 | 1 | Ref |
| 441 | CP013974 | Unassigned | Unassigned | Unassigned | 426_1 | Clustered | 3 | VC_426_1 | 3 | 0.1152 | 0.99804615 | 0.1149 | 0 | 0 | 0 | 1 | Ref |
| 442 | CP017905 | Unassigned | Unassigned | Unassigned | 16_0 | Clustered | 12 | VC_16_0 | 12 | 0.6615 | 1 | 0.6615 | 0 | 0 | 0 | 1 | Ref |
| 443 | CP017906 | Unassigned | Unassigned | Unassigned | 16_0 | Clustered | 12 | VC_16_0 | 12 | 0.6615 | 1 | 0.6615 | 0 | 0 | 0 | 1 | Ref |
| 444 | CP019275 | Unassigned | Unassigned | Unassigned | 6_0 | Clustered | 44 | VC_6_0 | 44 | 0.908 | 1 | 0.908 | 0 | 0 | 0 | 1 | Ref |
| 445 | CP023680 | Unassigned | Unassigned | Unassigned | 55_0 | Clustered | 3 | VC_55_0 | 3 | 0.2818 | 0.99850111 | 0.2813 | 0 | 0 | 0 | 1 | Ref |
| 446 | CP023686 | Unassigned | Unassigned | Unassigned | 55_0 | Clustered | 3 | VC_55_0 | 3 | 0.2818 | 0.99850111 | 0.2813 | 0 | 0 | 0 | 1 | Ref |
| 447 | CP025874 | Unassigned | Unassigned | Unassigned | 229_0 | Clustered | 32 | VC_229_0 | 32 | 0.9119 | 1 | 0.9119 | 0 | 0 | 0 | 1 | Ref |
| 448 | CP025900 | Unassigned | Unassigned | Unassigned | 6_0 | Clustered | 44 | VC_6_0 | 44 | 0.908 | 1 | 0.908 | 0 | 0 | 0 | 1 | Ref |
| 449 | CP025901 | Unassigned | Unassigned | Unassigned | 229_0 | Clustered | 32 | VC_229_0 | 32 | 0.9119 | 1 | 0.9119 | 0 | 0 | 0 | 1 | Ref |
| 450 | CP025923 | Unassigned | Unassigned | Unassigned | 229_0 | Clustered | 32 | VC_229_0 | 32 | 0.9119 | 1 | 0.9119 | 0 | 0 | 0 | 1 | Ref |
| 451 | CP027117 | Unassigned | Unassigned | Unassigned | 720_1 | Clustered/Singleton | 1 | VC_720_1 | 1 | 0 | 1 | 0 | 0 | 0 | 0 | 1 | Ref |
| 452 | CP038007 | Unassigned | Unassigned | Unassigned | 34_0 | Clustered | 4 | VC_34_0 | 4 | 0.4349 | 0.99998062 | 0.4349 | 0 | 0 | 0 | 1 | Ref |
| 453 | CP045566 | Unassigned | Unassigned | Unassigned | 721_0 | Clustered | 9 | VC_721_0 | 9 | 1 | 1 | 1 | 0 | 0 | 0 | 1 | Ref |
| 454 | CP051272 | Unassigned | Unassigned | Unassigned | 28_1 | Clustered | 53 | VC_28_1 | 53 | 0.7164 | 1 | 0.7164 | 0 | 0 | 0 | 0.9325 | Ref |
| 455 | CP051278 | Unassigned | Unassigned | Unassigned | 28_1 | Clustered | 53 | VC_28_1 | 53 | 0.7164 | 1 | 0.7164 | 0 | 0 | 0 | 0.9325 | Ref |
| 456 | CP051279 | Unassigned | Unassigned | Unassigned | 28_1 | Clustered | 53 | VC_28_1 | 53 | 0.7164 | 1 | 0.7164 | 0 | 0 | 0 | 0.9325 | Ref |
| 457 | CP051285 | Unassigned | Unassigned | Unassigned | 28_1 | Clustered | 53 | VC_28_1 | 53 | 0.7164 | 1 | 0.7164 | 0 | 0 | 0 | 0.9325 | Ref |
| 458 | CP051288 | Unassigned | Unassigned | Unassigned | 28_1 | Clustered | 53 | VC_28_1 | 53 | 0.7164 | 1 | 0.7164 | 0 | 0 | 0 | 0.9325 | Ref |
| 459 | CP052639 | Unassigned | Unassigned | Unassigned | 229_0 | Clustered | 32 | VC_229_0 | 32 | 0.9119 | 1 | 0.9119 | 0 | 0 | 0 | 1 | Ref |
| 460 | CP053388 | Unassigned | Unassigned | Unassigned | 723_0 | Clustered | 9 | VC_723_0 | 9 | 0.7138 | 1 | 0.7138 | 0 | 0 | 0 | 1 | Ref |
| 461 | CP054387 | Unassigned | Unassigned | Unassigned | 723_0 | Clustered | 9 | VC_723_0 | 9 | 0.7138 | 1 | 0.7138 | 0 | 0 | 0 | 1 | Ref |
| 462 | CP058330 | Unassigned | Unassigned | Unassigned | 723_0 | Clustered | 9 | VC_723_0 | 9 | 0.7138 | 1 | 0.7138 | 0 | 0 | 0 | 1 | Ref |
| 463 | CP059058 | Unassigned | Unassigned | Unassigned | 591_0 | Clustered | 14 | VC_591_0 | 14 | 0.5184 | 1 | 0.5184 | 0 | 0 | 0 | 1 | Ref |
| 464 | CP060690 | Unassigned | Unassigned | Unassigned | 730_0 | Clustered | 9 | VC_730_0 | 9 | 1 | 1 | 1 | 0 | 0 | 0 | 1 | Ref |
| 465 | CP062422 | Unassigned | Unassigned | Unassigned | 12_0 | Clustered | 21 | VC_12_0 | 21 | 0.2274 | 1 | 0.2274 | 0 | 0 | 0 | 0.9571 | Ref |
| 466 | CP062426 | Unassigned | Unassigned | Unassigned | 7_0 | Clustered | 28 | VC_7_0 | 28 | 0.3916 | 1 | 0.3916 | 0 | 0 | 0 | 0.9947 | Ref |
| 467 | CP062427 | Unassigned | Unassigned | Unassigned | 9_0 | Clustered | 38 | VC_9_0 | 38 | 0.4895 | 1 | 0.4895 | 0 | 0 | 0 | 1 | Ref |
| 468 | CP062430 | Unassigned | Unassigned | Unassigned | 10_0 | Clustered | 25 | VC_10_0 | 25 | 0.3467 | 1 | 0.3467 | 0 | 0 | 0 | 0.9733 | Ref |
| 469 | CP062434 | Unassigned | Unassigned | Unassigned | 8_0 | Clustered | 58 | VC_8_0 | 58 | 0.8306 | 1 | 0.8306 | 0 | 0 | 0 | 0.9946 | Ref |
| 470 | CP062435 | Unassigned | Unassigned | Unassigned | 7_0 | Clustered | 28 | VC_7_0 | 28 | 0.3916 | 1 | 0.3916 | 0 | 0 | 0 | 0.9947 | Ref |
| 471 | CP062436 | Unassigned | Unassigned | Unassigned | 10_0 | Clustered | 25 | VC_10_0 | 25 | 0.3467 | 1 | 0.3467 | 0 | 0 | 0 | 0.9733 | Ref |
| 472 | CP062442 | Unassigned | Unassigned | Unassigned | 8_0 | Clustered | 58 | VC_8_0 | 58 | 0.8306 | 1 | 0.8306 | 0 | 0 | 0 | 0.9946 | Ref |
| 473 | CP062444 | Unassigned | Unassigned | Unassigned | 8_0 | Clustered | 58 | VC_8_0 | 58 | 0.8306 | 1 | 0.8306 | 0 | 0 | 0 | 0.9946 | Ref |
| 474 | CP062445 | Unassigned | Unassigned | Unassigned | 7_0 | Clustered | 28 | VC_7_0 | 28 | 0.3916 | 1 | 0.3916 | 0 | 0 | 0 | 0.9947 | Ref |
| 475 | CP062447 | Unassigned | Unassigned | Unassigned | 10_0 | Clustered | 25 | VC_10_0 | 25 | 0.3467 | 1 | 0.3467 | 0 | 0 | 0 | 0.9733 | Ref |
| 476 | CP062451 | Unassigned | Unassigned | Unassigned | 9_0 | Clustered | 38 | VC_9_0 | 38 | 0.4895 | 1 | 0.4895 | 0 | 0 | 0 | 1 | Ref |
| 477 | CP062454 | Unassigned | Unassigned | Unassigned | 9_0 | Clustered | 38 | VC_9_0 | 38 | 0.4895 | 1 | 0.4895 | 0 | 0 | 0 | 1 | Ref |
| 478 | CP062457 | Unassigned | Unassigned | Unassigned | 7_0 | Clustered | 28 | VC_7_0 | 28 | 0.3916 | 1 | 0.3916 | 0 | 0 | 0 | 0.9947 | Ref |
| 479 | CP062461 | Unassigned | Unassigned | Unassigned | 8_0 | Clustered | 58 | VC_8_0 | 58 | 0.8306 | 1 | 0.8306 | 0 | 0 | 0 | 0.9946 | Ref |
| 480 | CP062462 | Unassigned | Unassigned | Unassigned | 12_0 | Clustered | 21 | VC_12_0 | 21 | 0.2274 | 1 | 0.2274 | 0 | 0 | 0 | 0.9571 | Ref |
| 481 | CP062992 | Unassigned | Unassigned | Unassigned | 491_0 | Clustered | 36 | VC_491_0 | 36 | 0.5336 | 1 | 0.5336 | 0 | 0 | 0 | 0.9714 | Ref |
| 482 | CP067347 | Unassigned | Unassigned | Unassigned | 473_4 | Clustered/Singleton | 1 | VC_473_4 | 1 | 0 | 1 | 0 | 0 | 0 | 0 | 1 | Ref |
| 483 | CP067352 | Unassigned | Unassigned | Unassigned | 473_0 | Clustered/Singleton | 1 | VC_473_0 | 1 | 0 | 1 | 0 | 0 | 0 | 0 | 1 | Ref |
| 484 | CP069347 | Unassigned | Unassigned | Unassigned | 647_0 | Clustered | 11 | VC_647_0 | 11 | 0.598 | 1 | 0.598 | 0 | 0 | 0 | 1 | Ref |

| | | | | | | | | | | | | | | | | | |
|---|---|---|---|---|---|---|---|---|---|---|---|---|---|---|---|---|---|
| 485 | CP069348 | Unassigned | Unassigned | Unassigned | 473_1 | Clustered/Singleton | 1 | VC_473_1 | 1 | 0 | 1 | 0 | 0 | 0 | 0 | 1 | Ref |
| 486 | CP071049 | Unassigned | Unassigned | Unassigned | 12_0 | Clustered | 21 | VC_12_0 | 21 | 0.2274 | 1 | 0.2274 | 0 | 0 | 0 | 0.9571 | Ref |
| 487 | CP072507 | Unassigned | Unassigned | Unassigned | 34_0 | Clustered | 4 | VC_34_0 | 4 | 0.4349 | 0.99998062 | 0.4349 | 0 | 0 | 0 | 1 | Ref |
| 488 | CP091911 | Unassigned | Unassigned | Unassigned | 706_0 | Clustered | 2 | VC_706_0 | 2 | 1 | 1 | 1 | 0 | 0 | 0 | 1 | Ref |
| 489 | CP101890 | Unassigned | Unassigned | Unassigned | 723_0 | Clustered | 9 | VC_723_0 | 9 | 0.7138 | 1 | 0.7138 | 0 | 0 | 0 | 1 | Ref |
| 490 | CP101896 | Unassigned | Unassigned | Unassigned | 723_0 | Clustered | 9 | VC_723_0 | 9 | 0.7138 | 1 | 0.7138 | 0 | 0 | 0 | 1 | Ref |
| 491 | D89074 | Unassigned | Unassigned | Unassigned | 16_0 | Clustered | 12 | VC_16_0 | 12 | 0.6615 | 1 | 0.6615 | 0 | 0 | 0 | 1 | Ref |
| 492 | DQ003260 | Unassigned | Unassigned | Unassigned | 28_1 | Clustered | 53 | VC_28_1 | 53 | 0.7164 | 1 | 0.7164 | 0 | 0 | 0 | 0.9325 | Ref |
| 493 | DQ003637 | Unassigned | Unassigned | Unassigned | 378_0 | Clustered | 9 | VC_378_0 | 9 | 0.7603 | 1 | 0.7603 | 0 | 0 | 0 | 1 | Ref |
| 494 | DQ003638 | Unassigned | Unassigned | Unassigned | 739_0 | Clustered | 26 | VC_739_0 | 26 | 0.9571 | 1 | 0.9571 | 0 | 0 | 0 | 1 | Ref |
| 495 | DQ003639 | Unassigned | Unassigned | Unassigned | 379_0 | Clustered | 5 | VC_379_0 | 5 | 0.4779 | 0.99999992 | 0.4779 | 0 | 0 | 0 | 0.9 | Ref |
| 496 | DQ003640 | Unassigned | Unassigned | Unassigned | 140_0 | Clustered | 2 | VC_140_0 | 2 | 0.4111 | 0.94824791 | 0.3898 | 0 | 0 | 0 | 1 | Ref |
| 497 | DQ003641 | Unassigned | Unassigned | Unassigned | 740_0 | Clustered | 3 | VC_740_0 | 3 | 0.202 | 0.99811828 | 0.2016 | 0 | 0 | 0 | 1 | Ref |
| 498 | DQ003642 | Unassigned | Unassigned | Unassigned | 380_0 | Clustered | 3 | VC_380_0 | 3 | 0.1631 | 0.99675097 | 0.1625 | 0 | 0 | 0 | 1 | Ref |
| 499 | DQ004855 | Unassigned | Unassigned | Unassigned | 739_0 | Clustered | 26 | VC_739_0 | 26 | 0.9571 | 1 | 0.9571 | 0 | 0 | 0 | 1 | Ref |
| 500 | DQ029335 | Unassigned | Unassigned | Unassigned | 82_0 | Clustered | 24 | VC_82_0 | 24 | 0.7351 | 1 | 0.7351 | 0 | 0 | 0 | 1 | Ref |
| 501 | DQ054536 | Unassigned | Unassigned | Unassigned | 202_0 | Clustered | 102 | VC_202_0 | 102 | 0.4049 | 1 | 0.4049 | 0 | 0 | 0 | 0.7876 | Ref |
| 502 | DQ079870 | Unassigned | Unassigned | Unassigned | 743_0 | Clustered | 15 | VC_743_0 | 15 | 1 | 1 | 1 | 0 | 0 | 0 | 1 | Ref |
| 503 | DQ079871 | Unassigned | Unassigned | Unassigned | 743_0 | Clustered | 15 | VC_743_0 | 15 | 1 | 1 | 1 | 0 | 0 | 0 | 1 | Ref |
| 504 | DQ079872 | Unassigned | Unassigned | Unassigned | 743_0 | Clustered | 15 | VC_743_0 | 15 | 1 | 1 | 1 | 0 | 0 | 0 | 1 | Ref |
| 505 | DQ079873 | Unassigned | Unassigned | Unassigned | 743_0 | Clustered | 15 | VC_743_0 | 15 | 1 | 1 | 1 | 0 | 0 | 0 | 1 | Ref |
| 506 | DQ079874 | Unassigned | Unassigned | Unassigned | 743_0 | Clustered | 15 | VC_743_0 | 15 | 1 | 1 | 1 | 0 | 0 | 0 | 1 | Ref |
| 507 | DQ079875 | Unassigned | Unassigned | Unassigned | 743_0 | Clustered | 15 | VC_743_0 | 15 | 1 | 1 | 1 | 0 | 0 | 0 | 1 | Ref |
| 508 | DQ079876 | Unassigned | Unassigned | Unassigned | 743_0 | Clustered | 15 | VC_743_0 | 15 | 1 | 1 | 1 | 0 | 0 | 0 | 1 | Ref |
| 509 | DQ079878 | Unassigned | Unassigned | Unassigned | 743_0 | Clustered | 15 | VC_743_0 | 15 | 1 | 1 | 1 | 0 | 0 | 0 | 1 | Ref |
| 510 | DQ079879 | Unassigned | Unassigned | Unassigned | 743_0 | Clustered | 15 | VC_743_0 | 15 | 1 | 1 | 1 | 0 | 0 | 0 | 1 | Ref |
| 511 | DQ079880 | Unassigned | Unassigned | Unassigned | 303_0 | Clustered | 155 | VC_303_0 | 155 | 0.9979 | 1 | 0.9979 | 0 | 0 | 0 | 1 | Ref |
| 512 | DQ079881 | Unassigned | Unassigned | Unassigned | 303_0 | Clustered | 155 | VC_303_0 | 155 | 0.9979 | 1 | 0.9979 | 0 | 0 | 0 | 1 | Ref |
| 513 | DQ079882 | Unassigned | Unassigned | Unassigned | 303_0 | Clustered | 155 | VC_303_0 | 155 | 0.9979 | 1 | 0.9979 | 0 | 0 | 0 | 1 | Ref |
| 514 | DQ079883 | Unassigned | Unassigned | Unassigned | 303_0 | Clustered | 155 | VC_303_0 | 155 | 0.9979 | 1 | 0.9979 | 0 | 0 | 0 | 1 | Ref |
| 515 | DQ079884 | Unassigned | Unassigned | Unassigned | 303_0 | Clustered | 155 | VC_303_0 | 155 | 0.9979 | 1 | 0.9979 | 0 | 0 | 0 | 1 | Ref |
| 516 | DQ079885 | Unassigned | Unassigned | Unassigned | 303_0 | Clustered | 155 | VC_303_0 | 155 | 0.9979 | 1 | 0.9979 | 0 | 0 | 0 | 1 | Ref |
| 517 | DQ079886 | Unassigned | Unassigned | Unassigned | 303_0 | Clustered | 155 | VC_303_0 | 155 | 0.9979 | 1 | 0.9979 | 0 | 0 | 0 | 1 | Ref |
| 518 | DQ079887 | Unassigned | Unassigned | Unassigned | 303_0 | Clustered | 155 | VC_303_0 | 155 | 0.9979 | 1 | 0.9979 | 0 | 0 | 0 | 1 | Ref |
| 519 | DQ079888 | Unassigned | Unassigned | Unassigned | 303_0 | Clustered | 155 | VC_303_0 | 155 | 0.9979 | 1 | 0.9979 | 0 | 0 | 0 | 1 | Ref |
| 520 | DQ079889 | Unassigned | Unassigned | Unassigned | 303_0 | Clustered | 155 | VC_303_0 | 155 | 0.9979 | 1 | 0.9979 | 0 | 0 | 0 | 1 | Ref |
| 521 | DQ079890 | Unassigned | Unassigned | Unassigned | 303_0 | Clustered | 155 | VC_303_0 | 155 | 0.9979 | 1 | 0.9979 | 0 | 0 | 0 | 1 | Ref |
| 522 | DQ079891 | Unassigned | Unassigned | Unassigned | 303_0 | Clustered | 155 | VC_303_0 | 155 | 0.9979 | 1 | 0.9979 | 0 | 0 | 0 | 1 | Ref |
| 523 | DQ079892 | Unassigned | Unassigned | Unassigned | 303_0 | Clustered | 155 | VC_303_0 | 155 | 0.9979 | 1 | 0.9979 | 0 | 0 | 0 | 1 | Ref |
| 524 | DQ079893 | Unassigned | Unassigned | Unassigned | 303_0 | Clustered | 155 | VC_303_0 | 155 | 0.9979 | 1 | 0.9979 | 0 | 0 | 0 | 1 | Ref |
| 525 | DQ079894 | Unassigned | Unassigned | Unassigned | 303_0 | Clustered | 155 | VC_303_0 | 155 | 0.9979 | 1 | 0.9979 | 0 | 0 | 0 | 1 | Ref |
| 526 | DQ079895 | Unassigned | Unassigned | Unassigned | 303_0 | Clustered | 155 | VC_303_0 | 155 | 0.9979 | 1 | 0.9979 | 0 | 0 | 0 | 1 | Ref |
| 527 | DQ079896 | Unassigned | Unassigned | Unassigned | 360_0 | Clustered | 358 | VC_360_0 | 358 | 0.9615 | 1 | 0.9615 | 0 | 0 | 0 | 1 | Ref |
| 528 | DQ079897 | Unassigned | Unassigned | Unassigned | 360_0 | Clustered | 358 | VC_360_0 | 358 | 0.9615 | 1 | 0.9615 | 0 | 0 | 0 | 1 | Ref |
| 529 | DQ079899 | Unassigned | Unassigned | Unassigned | 360_0 | Clustered | 358 | VC_360_0 | 358 | 0.9615 | 1 | 0.9615 | 0 | 0 | 0 | 1 | Ref |
| 530 | DQ079900 | Unassigned | Unassigned | Unassigned | 360_0 | Clustered | 358 | VC_360_0 | 358 | 0.9615 | 1 | 0.9615 | 0 | 0 | 0 | 1 | Ref |
| 531 | DQ079901 | Unassigned | Unassigned | Unassigned | 360_0 | Clustered | 358 | VC_360_0 | 358 | 0.9615 | 1 | 0.9615 | 0 | 0 | 0 | 1 | Ref |
| 532 | DQ079902 | Unassigned | Unassigned | Unassigned | 360_0 | Clustered | 358 | VC_360_0 | 358 | 0.9615 | 1 | 0.9615 | 0 | 0 | 0 | 1 | Ref |
| 533 | DQ079903 | Unassigned | Unassigned | Unassigned | 360_0 | Clustered | 358 | VC_360_0 | 358 | 0.9615 | 1 | 0.9615 | 0 | 0 | 0 | 1 | Ref |
| 534 | DQ079904 | Unassigned | Unassigned | Unassigned | 360_0 | Clustered | 358 | VC_360_0 | 358 | 0.9615 | 1 | 0.9615 | 0 | 0 | 0 | 1 | Ref |
| 535 | DQ079906 | Unassigned | Unassigned | Unassigned | 360_0 | Clustered | 358 | VC_360_0 | 358 | 0.9615 | 1 | 0.9615 | 0 | 0 | 0 | 1 | Ref |
| 536 | DQ079908 | Unassigned | Unassigned | Unassigned | 360_0 | Clustered | 358 | VC_360_0 | 358 | 0.9615 | 1 | 0.9615 | 0 | 0 | 0 | 1 | Ref |
| 537 | DQ079909 | Unassigned | Unassigned | Unassigned | 360_0 | Clustered | 358 | VC_360_0 | 358 | 0.9615 | 1 | 0.9615 | 0 | 0 | 0 | 1 | Ref |
| 538 | DQ085810 | Unassigned | Unassigned | Unassigned | 743_0 | Clustered | 15 | VC_743_0 | 15 | 1 | 1 | 1 | 0 | 0 | 0 | 1 | Ref |
| 539 | DQ087285 | Unassigned | Unassigned | Unassigned | 3_0 | Clustered | 17 | VC_3_0 | 17 | 0.5639 | 1 | 0.5639 | 0 | 0 | 0 | 1 | Ref |
| 540 | DQ111067 | Unassigned | Unassigned | Unassigned | 451_0 | Clustered | 80 | VC_451_0 | 80 | 0.9991 | 1 | 0.9991 | 0 | 0 | 0 | 1 | Ref |
| 541 | DQ113772 | Unassigned | Unassigned | Unassigned | 412_0 | Clustered | 39 | VC_412_0 | 39 | 0.845 | 1 | 0.845 | 0 | 0 | 0 | 1 | Ref |
| 542 | DQ114220 | Unassigned | Unassigned | Unassigned | 5_0 | Clustered | 30 | VC_5_0 | 30 | 0.9647 | 1 | 0.9647 | 0 | 0 | 0 | 1 | Ref |
| 543 | DQ121662 | Unassigned | Unassigned | Unassigned | 455_0 | Clustered | 18 | VC_455_0 | 18 | 0.4598 | 1 | 0.4598 | 0 | 0 | 0 | 1 | Ref |
| 544 | DQ123818 | Unassigned | Unassigned | Unassigned | 744_0 | Clustered | 10 | VC_744_0 | 10 | 0.7049 | 1 | 0.7049 | 0 | 0 | 0 | 1 | Ref |
| 545 | DQ126339 | Unassigned | Unassigned | Unassigned | 476_0 | Clustered | 19 | VC_476_0 | 19 | 0.7814 | 1 | 0.7814 | 0 | 0 | 0 | 1 | Ref |
| 546 | DQ150593 | Unassigned | Unassigned | Unassigned | 704_0 | Clustered | 16 | VC_704_0 | 16 | 0.7274 | 1 | 0.7274 | 0 | 0 | 0 | 1 | Ref |
| 547 | DQ163912 | Unassigned | Unassigned | Unassigned | 747_0 | Clustered | 23 | VC_747_0 | 23 | 0.8202 | 1 | 0.8202 | 0 | 0 | 0 | 1 | Ref |
| 548 | DQ163913 | Unassigned | Unassigned | Unassigned | 749_0 | Clustered | 40 | VC_749_0 | 40 | 0.7517 | 1 | 0.7517 | 0 | 0 | 0 | 1 | Ref |
| 549 | DQ163914 | Unassigned | Unassigned | Unassigned | 612_0 | Clustered | 9 | VC_612_0 | 9 | 0.9545 | 1 | 0.9545 | 0 | 0 | 0 | 1 | Ref |
| 550 | DQ163915 | Unassigned | Unassigned | Unassigned | 755_0 | Clustered | 9 | VC_755_0 | 9 | 0.4489 | 1 | 0.4489 | 0 | 0 | 0 | 1 | Ref |
| 551 | DQ163916 | Unassigned | Unassigned | Unassigned | 166_0 | Clustered | 26 | VC_166_0 | 26 | 0.7923 | 1 | 0.7923 | 0 | 0 | 0 | 1 | Ref |
| 552 | DQ163917 | Unassigned | Unassigned | Unassigned | 80_0 | Clustered | 41 | VC_80_0 | 41 | 0.5837 | 1 | 0.5837 | 0 | 0 | 0 | 0.9805 | Ref |
| 553 | DQ222851 | Unassigned | Unassigned | Unassigned | 704_0 | Clustered | 16 | VC_704_0 | 16 | 0.7274 | 1 | 0.7274 | 0 | 0 | 0 | 1 | Ref |
| 554 | DQ222853 | Unassigned | Unassigned | Unassigned | 704_0 | Clustered | 16 | VC_704_0 | 16 | 0.7274 | 1 | 0.7274 | 0 | 0 | 0 | 1 | Ref |
| 555 | DQ222855 | Unassigned | Unassigned | Unassigned | 704_0 | Clustered | 16 | VC_704_0 | 16 | 0.7274 | 1 | 0.7274 | 0 | 0 | 0 | 1 | Ref |
| 556 | DQ227763 | Unassigned | Unassigned | Unassigned | 202_0 | Clustered | 102 | VC_202_0 | 102 | 0.4049 | 1 | 0.4049 | 0 | 0 | 0 | 0.7876 | Ref |
| 557 | DQ227764 | Unassigned | Unassigned | Unassigned | 202_0 | Clustered | 102 | VC_202_0 | 102 | 0.4049 | 1 | 0.4049 | 0 | 0 | 0 | 0.7876 | Ref |
| 558 | DQ289555 | Unassigned | Unassigned | Unassigned | 704_0 | Clustered | 16 | VC_704_0 | 16 | 0.7274 | 1 | 0.7274 | 0 | 0 | 0 | 1 | Ref |
| 559 | DQ289556 | Unassigned | Unassigned | Unassigned | 704_0 | Clustered | 16 | VC_704_0 | 16 | 0.7274 | 1 | 0.7274 | 0 | 0 | 0 | 1 | Ref |
| 560 | DQ320509 | Unassigned | Unassigned | Unassigned | 308_0 | Clustered | 4 | VC_308_0 | 4 | 0.5014 | 0.99997418 | 0.5013 | 0 | 0 | 0 | 1 | Ref |
| 561 | DQ340064 | Unassigned | Unassigned | Unassigned | 138_0 | Clustered | 2 | VC_138_0 | 2 | 0.2009 | 0.95368907 | 0.1916 | 0 | 0 | 0 | 1 | Ref |
| 562 | DQ386162 | Unassigned | Unassigned | Unassigned | 463_0 | Clustered | 4 | VC_463_0 | 4 | 0.7906 | 0.99997156 | 0.7906 | 0 | 0 | 0 | 1 | Ref |
| 563 | DQ394807 | Unassigned | Unassigned | Unassigned | 265_0 | Clustered | 12 | VC_265_0 | 12 | 0.4401 | 1 | 0.4401 | 0 | 0 | 0 | 1 | Ref |
| 564 | DQ394808 | Unassigned | Unassigned | Unassigned | 265_0 | Clustered | 12 | VC_265_0 | 12 | 0.4401 | 1 | 0.4401 | 0 | 0 | 0 | 1 | Ref |
| 565 | DQ394809 | Unassigned | Unassigned | Unassigned | 265_0 | Clustered | 12 | VC_265_0 | 12 | 0.4401 | 1 | 0.4401 | 0 | 0 | 0 | 1 | Ref |
| 566 | DQ394810 | Unassigned | Unassigned | Unassigned | 265_0 | Clustered | 12 | VC_265_0 | 12 | 0.4401 | 1 | 0.4401 | 0 | 0 | 0 | 1 | Ref |
| 567 | DQ398041 | Unassigned | Unassigned | Unassigned | 551_7 | Clustered | 28 | VC_551_7 | 28 | 0.1661 | 1 | 0.1661 | 0 | 0 | 0 | 0.8519 | Ref |
| 568 | DQ398042 | Unassigned | Unassigned | Unassigned | 761_0 | Clustered | 67 | VC_761_0 | 67 | 0.8947 | 1 | 0.8947 | 0 | 0 | 0 | 1 | Ref |
| 569 | DQ398043 | Unassigned | Unassigned | Unassigned | 215_0 | Clustered | 5 | VC_215_0 | 5 | 0.0724 | 0.99999997 | 0.0724 | 0 | 0 | 0 | 1 | Ref |
| 570 | DQ398044 | Unassigned | Unassigned | Unassigned | 372_0 | Clustered | 24 | VC_372_0 | 24 | 0.5539 | 1 | 0.5539 | 0 | 0 | 0 | 1 | Ref |
| 571 | DQ398045 | Unassigned | Unassigned | Unassigned | 520_60 | Clustered | 2 | VC_520_60 | 2 | 0.0091 | 0.95761233 | 0.0087 | 0 | 0 | 0 | 1 | Ref |
| 572 | DQ398046 | Unassigned | Unassigned | Unassigned | 371_16 | Clustered | 42 | VC_371_16 | 42 | 0.1341 | 1 | 0.1341 | 0 | 0 | 0 | 0.7282 | Ref |
| 573 | DQ398047 | Unassigned | Unassigned | Unassigned | 565_0 | Clustered | 20 | VC_565_0 | 20 | 0.8398 | 1 | 0.8398 | 0 | 0 | 0 | 1 | Ref |
| 574 | DQ398048 | Unassigned | Unassigned | Unassigned | 508_0 | Clustered | 32 | VC_508_0 | 32 | 0.906 | 1 | 0.906 | 0 | 0 | 0 | 1 | Ref |
| 575 | DQ398049 | Unassigned | Unassigned | Unassigned | 509_0 | Clustered | 37 | VC_509_0 | 37 | 0.9689 | 1 | 0.9689 | 0 | 0 | 0 | 1 | Ref |
| 576 | DQ398050 | Unassigned | Unassigned | Unassigned | 520_58 | Clustered | 3 | VC_520_58 | 3 | 0.021 | 0.9985993 | 0.021 | 0 | 0 | 0 | 1 | Ref |
| 577 | DQ398051 | Unassigned | Unassigned | Unassigned | 565_0 | Clustered | 20 | VC_565_0 | 20 | 0.8398 | 1 | 0.8398 | 0 | 0 | 0 | 1 | Ref |
| 578 | DQ398052 | Unassigned | Unassigned | Unassigned | 764_0 | Clustered | 4 | VC_764_0 | 4 | 0.3556 | 0.99997469 | 0.3556 | 0 | 0 | 0 | 1 | Ref |
| 579 | DQ398053 | Unassigned | Unassigned | Unassigned | 558_31 | Clustered/Singleton | 1 | VC_558_31 | 1 | 0 | 1 | 0 | 0 | 0 | 0 | 1 | Ref |
| 580 | DQ426904 | Unassigned | Unassigned | Unassigned | 4_0 | Clustered | 7 | VC_4_0 | 7 | 0.7912 | 1 | 0.7912 | 0 | 0 | 0 | 1 | Ref |
| 581 | DQ426905 | Unassigned | Unassigned | Unassigned | 4_0 | Clustered | 7 | VC_4_0 | 7 | 0.7912 | 1 | 0.7912 | 0 | 0 | 0 | 1 | Ref |
| 582 | DQ431235 | Unassigned | Unassigned | Unassigned | 765_0 | Clustered | 132 | VC_765_0 | 132 | 1 | 1 | 1 | 0 | 0 | 0 | 1 | Ref |
| 583 | DQ466086 | Unassigned | Unassigned | Unassigned | 139_1 | Clustered | 4 | VC_139_1 | 4 | 0.1775 | 0.99998609 | 0.1775 | 0 | 0 | 0 | 1 | Ref |
| 584 | DQ490056 | Unassigned | Unassigned | Unassigned | 689_0 | Clustered | 2 | VC_689_0 | 2 | 0.4402 | 0.95582436 | 0.4208 | 0 | 0 | 0 | 1 | Ref |
| 585 | DQ500118 | Unassigned | Unassigned | Unassigned | 770_0 | Clustered | 5 | VC_770_0 | 5 | 0.8394 | 0.99999405 | 0.8394 | 0 | 0 | 0 | 1 | Ref |
| 586 | DQ517338 | Unassigned | Unassigned | Unassigned | 9_0 | Clustered | 38 | VC_9_0 | 38 | 0.4895 | 1 | 0.4895 | 0 | 0 | 0 | 1 | Ref |
| 587 | DQ529280 | Unassigned | Unassigned | Unassigned | 600_2 | Clustered | 10 | VC_600_2 | 10 | 0.3041 | 1 | 0.3041 | 0 | 0 | 0 | 1 | Ref |
| 588 | DQ530359 | Unassigned | Unassigned | Unassigned | 9_0 | Clustered | 38 | VC_9_0 | 38 | 0.4895 | 1 | 0.4895 | 0 | 0 | 0 | 1 | Ref |
| 589 | DQ530360 | Unassigned | Unassigned | Unassigned | 9_0 | Clustered | 38 | VC_9_0 | 38 | 0.4895 | 1 | 0.4895 | 0 | 0 | 0 | 1 | Ref |
| 590 | DQ530361 | Unassigned | Unassigned | Unassigned | 10_0 | Clustered | 25 | VC_10_0 | 25 | 0.3467 | 1 | 0.3467 | 0 | 0 | 0 | 0.9733 | Ref |
| 591 | DQ530362 | Unassigned | Unassigned | Unassigned | 12_0 | Clustered | 21 | VC_12_0 | 21 | 0.2274 | 1 | 0.2274 | 0 | 0 | 0 | 0.9571 | Ref |
| 592 | DQ631426 | Unassigned | Unassigned | Unassigned | 179_0 | Clustered | 53 | VC_179_0 | 53 | 0.9137 | 1 | 0.9137 | 0 | 0 | 0 | 1 | Ref |
| 593 | DQ674738 | Unassigned | Unassigned | Unassigned | 5_0 | Clustered | 30 | VC_5_0 | 30 | 0.9647 | 1 | 0.9647 | 0 | 0 | 0 | 1 | Ref |
| 594 | DQ777876 | Unassigned | Unassigned | Unassigned | 113_0 | Clustered | 9 | VC_113_0 | 9 | 0.519 | 1 | 0.519 | 0 | 0 | 0 | 1 | Ref |
| 595 | DQ785801 | Unassigned | Unassigned | Unassigned | 63_0 | Clustered | 2 | VC_63_0 | 2 | 0.1147 | 0.95689598 | 0.1098 | 0 | 0 | 0 | 1 | Ref |
| 596 | DQ832317 | Unassigned | Unassigned | Unassigned | 772_15 | Clustered | 5 | VC_772_15 | 5 | 0.0243 | 0.76144861 | 0.0185 | 0 | 0 | 0 | 0.9 | Ref |
| 597 | DQ836063 | Unassigned | Unassigned | Unassigned | 314_0 | Clustered | 2 | VC_314_0 | 2 | 0.1808 | 0.99999983 | 0.1808 | 0 | 0 | 0 | 1 | Ref |
| 598 | DQ840344 | Unassigned | Unassigned | Unassigned | 138_0 | Clustered | 2 | VC_138_0 | 2 | 0.2009 | 0.95368907 | 0.1916 | 0 | 0 | 0 | 1 | Ref |
| 599 | DQ864624 | Unassigned | Unassigned | Unassigned | 773_0 | Clustered | 23 | VC_773_0 | 23 | 0.5514 | 1 | 0.5514 | 0 | 0 | 0 | 1 | Ref |
| 600 | DQ873690 | Unassigned | Unassigned | Unassigned | 179_0 | Clustered | 53 | VC_179_0 | 53 | 0.9137 | 1 | 0.9137 | 0 | 0 | 0 | 1 | Ref |
| 601 | DQ875742 | Unassigned | Unassigned | Unassigned | 778_0 | Clustered | 4 | VC_778_0 | 4 | 0.8115 | 0.99985435 | 0.8114 | 0 | 0 | 0 | 1 | Ref |
| 602 | DQ904452 | Unassigned | Unassigned | Unassigned | 283_94 | Clustered | 2 | VC_283_94 | 2 | 0.0026 | 0.95293762 | 0.0025 | 0 | 0 | 0 | 1 | Ref |
| 603 | DQ908929 | Unassigned | Unassigned | Unassigned | 12_2 | Clustered | 19 | VC_12_2 | 19 | 0.2018 | 1 | 0.2018 | 0 | 0 | 0 | 1 | Ref |
| 604 | DQ997624 | Unassigned | Unassigned | Unassigned | 477_0 | Clustered | 4 | VC_477_0 | 4 | 0.9836 | 0.99748973 | 0.9811 | 0 | 0 | 0 | 1 | Ref |
| 605 | EF056009 | Unassigned | Unassigned | Unassigned | 779_0 | Clustered | 37 | VC_779_0 | 37 | 0.5682 | 1 | 0.5682 | 0 | 0 | 0 | 1 | Ref |
| 606 | EF057797 | Unassigned | Unassigned | Unassigned | 126_0 | Clustered | 2 | VC_126_0 | 2 | 0.1661 | 0.95181568 | 0.1581 | 0 | 0 | 0 | 1 | Ref |
| 607 | EF068134 | Unassigned | Unassigned | Unassigned | 188_0 | Clustered | 13 | VC_188_0 | 13 | 0.9973 | 0.99823802 | 0.9956 | 0 | 0 | 0 | 1 | Ref |
| 608 | EF108464 | Unassigned | Unassigned | Unassigned | 315_0 | Clustered | 12 | VC_315_0 | 12 | 0.8947 | 1 | 0.8947 | 0 | 0 | 0 | 1 | Ref |
| 609 | EF108465 | Unassigned | Unassigned | Unassigned | 315_0 | Clustered | 12 | VC_315_0 | 12 | 0.8947 | 1 | 0.8947 | 0 | 0 | 0 | 1 | Ref |
| 610 | EF153632 | Unassigned | Unassigned | Unassigned | 536_0 | Clustered | 3 | VC_536_0 | 3 | 0.0878 | 0.9985359 | 0.0877 | 0 | 0 | 0 | 1 | Ref |
| 611 | EF160123 | Unassigned | Unassigned | Unassigned | 453_0 | Clustered | 6 | VC_453_0 | 6 | 0.114 | 1 | 0.114 | 0 | 0 | 0 | 1 | Ref |
| 612 | EF177456 | Unassigned | Unassigned | Unassigned | 637_0 | Clustered | 115 | VC_637_0 | 115 | 0.731 | 1 | 0.731 | 0 | 0 | 0 | 0.8764 | Ref |
| 613 | EF204939 | Unassigned | Unassigned | Unassigned | 315_0 | Clustered | 12 | VC_315_0 | 12 | 0.8947 | 1 | 0.8947 | 0 | 0 | 0 | 1 | Ref |
| 614 | EF204940 | Unassigned | Unassigned | Unassigned | 315_0 | Clustered | 12 | VC_315_0 | 12 | 0.8947 | 1 | 0.8947 | 0 | 0 | 0 | 1 | Ref |
| 615 | EF372997 | Unassigned | Unassigned | Unassigned | 340_0 | Clustered | 31 | VC_340_0 | 31 | 0.9549 | 1 | 0.9549 | 0 | 0 | 0 | 1 | Ref |
| 616 | EF380009 | Unassigned | Unassigned | Unassigned | 303_0 | Clustered | 155 | VC_303_0 | 155 | 0.9979 | 1 | 0.9979 | 0 | 0 | 0 | 1 | Ref |
| 617 | EF380010 | Unassigned | Unassigned | Unassigned | 303_0 | Clustered | 155 | VC_303_0 | 155 | 0.9979 | 1 | 0.9979 | 0 | 0 | 0 | 1 | Ref |
| 618 | EF380011 | Unassigned | Unassigned | Unassigned | 303_0 | Clustered | 155 | VC_303_0 | 155 | 0.9979 | 1 | 0.9979 | 0 | 0 | 0 | 1 | Ref |
| 619 | EF380012 | Unassigned | Unassigned | Unassigned | 303_0 | Clustered | 155 | VC_303_0 | 155 | 0.9979 | 1 | 0.9979 | 0 | 0 | 0 | 1 | Ref |
| 620 | EF380013 | Unassigned | Unassigned | Unassigned | 303_0 | Clustered | 155 | VC_303_0 | 155 | 0.9979 | 1 | 0.9979 | 0 | 0 | 0 | 1 | Ref |
| 621 | EF380014 | Unassigned | Unassigned | Unassigned | 303_0 | Clustered | 155 | VC_303_0 | 155 | 0.9979 | 1 | 0.9979 | 0 | 0 | 0 | 1 | Ref |
| 622 | EF380015 | Unassigned | Unassigned | Unassigned | 303_0 | Clustered | 155 | VC_303_0 | 155 | 0.9979 | 1 | 0.9979 | 0 | 0 | 0 | 1 | Ref |
| 623 | EF380016 | Unassigned | Unassigned | Unassigned | 303_0 | Clustered | 155 | VC_303_0 | 155 | 0.9979 | 1 | 0.9979 | 0 | 0 | 0 | 1 | Ref |
| 624 | EF380017 | Unassigned | Unassigned | Unassigned | 303_0 | Clustered | 155 | VC_303_0 | 155 | 0.9979 | 1 | 0.9979 | 0 | 0 | 0 | 1 | Ref |
| 625 | EF380018 | Unassigned | Unassigned | Unassigned | 303_0 | Clustered | 155 | VC_303_0 | 155 | 0.9979 | 1 | 0.9979 | 0 | 0 | 0 | 1 | Ref |
| 626 | EF380019 | Unassigned | Unassigned | Unassigned | 303_0 | Clustered | 155 | VC_303_0 | 155 | 0.9979 | 1 | 0.9979 | 0 | 0 | 0 | 1 | Ref |
| 627 | EF380020 | Unassigned | Unassigned | Unassigned | 303_0 | Clustered | 155 | VC_303_0 | 155 | 0.9979 | 1 | 0.9979 | 0 | 0 | 0 | 1 | Ref |
| 628 | EF380021 | Unassigned | Unassigned | Unassigned | 303_0 | Clustered | 155 | VC_303_0 | 155 | 0.9979 | 1 | 0.9979 | 0 | 0 | 0 | 1 | Ref |
| 629 | EF380022 | Unassigned | Unassigned | Unassigned | 303_0 | Clustered | 155 | VC_303_0 | 155 | 0.9979 | 1 | 0.9979 | 0 | 0 | 0 | 1 | Ref |
| 630 | EF380023 | Unassigned | Unassigned | Unassigned | 303_0 | Clustered | 155 | VC_303_0 | 155 | 0.9979 | 1 | 0.9979 | 0 | 0 | 0 | 1 | Ref |
| 631 | EF380024 | Unassigned | Unassigned | Unassigned | 303_0 | Clustered | 155 | VC_303_0 | 155 | 0.9979 | 1 | 0.9979 | 0 | 0 | 0 | 1 | Ref |
| 632 | EF380025 | Unassigned | Unassigned | Unassigned | 303_0 | Clustered | 155 | VC_303_0 | 155 | 0.9979 | 1 | 0.9979 | 0 | 0 | 0 | 1 | Ref |
| 633 | EF380026 | Unassigned | Unassigned | Unassigned | 303_0 | Clustered | 155 | VC_303_0 | 155 | 0.9979 | 1 | 0.9979 | 0 | 0 | 0 | 1 | Ref |
| 634 | EF380027 | Unassigned | Unassigned | Unassigned | 303_0 | Clustered | 155 | VC_303_0 | 155 | 0.9979 | 1 | 0.9979 | 0 | 0 | 0 | 1 | Ref |
| 635 | EF380028 | Unassigned | Unassigned | Unassigned | 303_0 | Clustered | 155 | VC_303_0 | 155 | 0.9979 | 1 | 0.9979 | 0 | 0 | 0 | 1 | Ref |
| 636 | EF380029 | Unassigned | Unassigned | Unassigned | 303_0 | Clustered | 155 | VC_303_0 | 155 | 0.9979 | 1 | 0.9979 | 0 | 0 | 0 | 1 | Ref |
| 637 | EF380030 | Unassigned | Unassigned | Unassigned | 303_0 | Clustered | 155 | VC_303_0 | 155 | 0.9979 | 1 | 0.9979 | 0 | 0 | 0 | 1 | Ref |
| 638 | EF380031 | Unassigned | Unassigned | Unassigned | 303_0 | Clustered | 155 | VC_303_0 | 155 | 0.9979 | 1 | 0.9979 | 0 | 0 | 0 | 1 | Ref |
| 639 | EF380032 | Unassigned | Unassigned | Unassigned | 303_0 | Clustered | 155 | VC_303_0 | 155 | 0.9979 | 1 | 0.9979 | 0 | 0 | 0 | 1 | Ref |
| 640 | EF432053 | Unassigned | Unassigned | Unassigned | 791_0 | Clustered | 3 | VC_791_0 | 3 | 1 | 1 | 1 | 0 | 0 | 0 | 1 | Ref |
| 641 | EF437941 | Unassigned | Unassigned | Unassigned | 590_0 | Clustered | 27 | VC_590_0 | 27 | 0.9947 | 1 | 0.9947 | 0 | 0 | 0 | 1 | Ref |
| 642 | EF460875 | Unassigned | Unassigned | Unassigned | 792_0 | Clustered | 35 | VC_792_0 | 35 | 0.9828 | 1 | 0.9828 | 0 | 0 | 0 | 1 | Ref |
| 643 | EF462197 | Unassigned | Unassigned | Unassigned | 7_0 | Clustered | 28 | VC_7_0 | 28 | 0.3916 | 1 | 0.3916 | 0 | 0 | 0 | 0.9947 | Ref |
| 644 | EF462198 | Unassigned | Unassigned | Unassigned | 8_0 | Clustered | 58 | VC_8_0 | 58 | 0.8306 | 1 | 0.8306 | 0 | 0 | 0 | 0.9946 | Ref |
| 645 | EF462199 | Unassigned | Unassigned | Unassigned | 7_0 | Clustered | 28 | VC_7_0 | 28 | 0.3916 | 1 | 0.3916 | 0 | 0 | 0 | 0.9947 | Ref |
| 646 | EF469154 | Unassigned | Unassigned | Unassigned | 290_6 | Clustered | 12 | VC_290_6 | 12 | 0.1149 | 1 | 0.1149 | 0 | 0 | 0 | 0.8182 | Ref |

| | | | | | | | | | | | | | | | | | |
|---|---|---|---|---|---|---|---|---|---|---|---|---|---|---|---|---|---|
| 647 | EF506570 | Unassigned | Unassigned | Unassigned | 794_0 | Clustered | 3 | VC_794_0 | 3 | 1 | 1 | 1 | 0 | 0 | 0 | 1 | Ref |
| 648 | EF529515 | Unassigned | Unassigned | Unassigned | 254_0 | Clustered | 46 | VC_254_0 | 46 | 0.5159 | 1 | 0.5159 | 0 | 0 | 0 | 1 | Ref |
| 649 | EF536069 | Unassigned | Unassigned | Unassigned | 520_24 | Clustered/Singleton | 1 | VC_520_24 | 1 | 0 | 1 | 0 | 0 | 0 | 0 | 1 | Ref |
| 650 | EF537008 | Unassigned | Unassigned | Unassigned | 777_0 | Clustered | 3 | VC_777_0 | 3 | 0.4763 | 0.9961812 | 0.4745 | 0 | 0 | 0 | 1 | Ref |
| 651 | EF583821 | Unassigned | Unassigned | Unassigned | 90_2 | Clustered/Singleton | 1 | VC_90_2 | 1 | 0 | 1 | 0 | 0 | 0 | 0 | 1 | Ref |
| 652 | EU030938 | Unassigned | Unassigned | Unassigned | 599_0 | Clustered | 50 | VC_599_0 | 50 | 0.9989 | 1 | 0.9989 | 0 | 0 | 0 | 1 | Ref |
| 653 | EU030939 | Unassigned | Unassigned | Unassigned | 599_0 | Clustered | 50 | VC_599_0 | 50 | 0.9989 | 1 | 0.9989 | 0 | 0 | 0 | 1 | Ref |
| 654 | EU056923 | Unassigned | Unassigned | Unassigned | 423_0 | Clustered | 45 | VC_423_0 | 45 | 0.9484 | 1 | 0.9484 | 0 | 0 | 0 | 1 | Ref |
| 655 | EU078592 | Unassigned | Unassigned | Unassigned | 43_0 | Clustered | 38 | VC_43_0 | 38 | 0.5885 | 1 | 0.5885 | 0 | 0 | 0 | 0.9659 | Ref |
| 656 | EU081845 | Unassigned | Unassigned | Unassigned | 800_0 | Clustered | 3 | VC_800_0 | 3 | 0.4591 | 0.9984737 | 0.4584 | 0 | 0 | 0 | 1 | Ref |
| 657 | EU100883 | Unassigned | Unassigned | Unassigned | 805_0 | Clustered | 4 | VC_805_0 | 4 | 0.9081 | 0.99622688 | 0.9046 | 0 | 0 | 0 | 1 | Ref |
| 658 | EU100884 | Unassigned | Unassigned | Unassigned | 805_0 | Clustered | 4 | VC_805_0 | 4 | 0.9081 | 0.99622688 | 0.9046 | 0 | 0 | 0 | 1 | Ref |
| 659 | EU124666 | Unassigned | Unassigned | Unassigned | 807_0 | Clustered | 17 | VC_807_0 | 17 | 0.907 | 1 | 0.907 | 0 | 0 | 0 | 1 | Ref |
| 660 | EU136189 | Unassigned | Unassigned | Unassigned | 96_0 | Clustered | 30 | VC_96_0 | 30 | 0.718 | 1 | 0.718 | 0 | 0 | 0 | 1 | Ref |
| 661 | EU197055 | Unassigned | Unassigned | Unassigned | 344_4 | Clustered/Singleton | 1 | VC_344_4 | 1 | 0 | 1 | 0 | 0 | 0 | 0 | 1 | Ref |
| 662 | EU203571 | Unassigned | Unassigned | Unassigned | 810_0 | Clustered | 18 | VC_810_0 | 18 | 0.9121 | 1 | 0.9121 | 0 | 0 | 0 | 1 | Ref |
| 663 | EU221285 | Unassigned | Unassigned | Unassigned | 202_0 | Clustered | 102 | VC_202_0 | 102 | 0.4049 | 1 | 0.4049 | 0 | 0 | 0 | 0.7876 | Ref |
| 664 | EU236438 | Unassigned | Unassigned | Unassigned | 423_0 | Clustered | 45 | VC_423_0 | 45 | 0.9484 | 1 | 0.9484 | 0 | 0 | 0 | 1 | Ref |
| 665 | EU246945 | Unassigned | Unassigned | Unassigned | 272_0 | Clustered | 11 | VC_272_0 | 11 | 0.4915 | 1 | 0.4915 | 0 | 0 | 0 | 0.8727 | Ref |
| 666 | EU272036 | Unassigned | Unassigned | Unassigned | 179_0 | Clustered | 53 | VC_179_0 | 53 | 0.9137 | 1 | 0.9137 | 0 | 0 | 0 | 1 | Ref |
| 667 | EU272037 | Unassigned | Unassigned | Unassigned | 179_0 | Clustered | 53 | VC_179_0 | 53 | 0.9137 | 1 | 0.9137 | 0 | 0 | 0 | 1 | Ref |
| 668 | EU311208 | Unassigned | Unassigned | Unassigned | 39_0 | Clustered | 35 | VC_39_0 | 35 | 0.4337 | 1 | 0.4337 | 0 | 0 | 0 | 1 | Ref |
| 669 | EU330206 | Unassigned | Unassigned | Unassigned | 813_1 | Clustered | 24 | VC_813_1 | 24 | 0.535 | 1 | 0.535 | 0 | 0 | 0 | 1 | Ref |
| 670 | EU340421 | Unassigned | Unassigned | Unassigned | 640_0 | Clustered | 7 | VC_640_0 | 7 | 0.9025 | 1 | 0.9025 | 0 | 0 | 0 | 1 | Ref |
| 671 | EU350361 | Unassigned | Unassigned | Unassigned | 814_0 | Clustered | 3 | VC_814_0 | 3 | 1 | 1 | 1 | 0 | 0 | 0 | 1 | Ref |
| 672 | EU399241 | Unassigned | Unassigned | Unassigned | 126_0 | Clustered | 2 | VC_126_0 | 2 | 0.1661 | 0.95181568 | 0.1581 | 0 | 0 | 0 | 1 | Ref |
| 673 | EU408779 | Unassigned | Unassigned | Unassigned | 424_0 | Clustered | 7 | VC_424_0 | 7 | 0.9044 | 0.9998143 | 0.9043 | 0 | 0 | 0 | 1 | Ref |
| 674 | EU418428 | Unassigned | Unassigned | Unassigned | 149_8 | Clustered | 66 | VC_149_8 | 66 | 0.4565 | 1 | 0.4565 | 0 | 0 | 0 | 0.8182 | Ref |
| 675 | EU545650 | Unassigned | Unassigned | Unassigned | 349_0 | Clustered | 7 | VC_349_0 | 7 | 0.9363 | 0.99999996 | 0.9363 | 0 | 0 | 0 | 1 | Ref |
| 676 | EU547803 | Unassigned | Unassigned | Unassigned | 385_0 | Clustered | 51 | VC_385_0 | 51 | 0.8754 | 1 | 0.8754 | 0 | 0 | 0 | 1 | Ref |
| 677 | EU568876 | Unassigned | Unassigned | Unassigned | 761_0 | Clustered | 67 | VC_761_0 | 67 | 0.8947 | 1 | 0.8947 | 0 | 0 | 0 | 1 | Ref |
| 678 | EU570103 | Unassigned | Unassigned | Unassigned | 28_1 | Clustered | 53 | VC_28_1 | 53 | 0.7164 | 1 | 0.7164 | 0 | 0 | 0 | 0.9325 | Ref |
| 679 | EU588980 | Unassigned | Unassigned | Unassigned | 815_0 | Clustered | 8 | VC_815_0 | 8 | 0.9801 | 1 | 0.9801 | 0 | 0 | 0 | 1 | Ref |
| 680 | EU622808 | Unassigned | Unassigned | Unassigned | 817_0 | Clustered | 14 | VC_817_0 | 14 | 0.6342 | 1 | 0.6342 | 0 | 0 | 0 | 1 | Ref |
| 681 | EU652770 | Unassigned | Unassigned | Unassigned | 390_0 | Clustered | 4 | VC_390_0 | 4 | 0.0468 | 0.9999882 | 0.0468 | 0 | 0 | 0 | 1 | Ref |
| 682 | EU676000 | Unassigned | Unassigned | Unassigned | 565_0 | Clustered | 20 | VC_565_0 | 20 | 0.8398 | 1 | 0.8398 | 0 | 0 | 0 | 1 | Ref |
| 683 | EU710883 | Unassigned | Unassigned | Unassigned | 328_0 | Clustered | 11 | VC_328_0 | 11 | 0.4707 | 1 | 0.4707 | 0 | 0 | 0 | 1 | Ref |
| 684 | EU716414 | Unassigned | Unassigned | Unassigned | 80_0 | Clustered | 41 | VC_80_0 | 41 | 0.5837 | 1 | 0.5837 | 0 | 0 | 0 | 0.9805 | Ref |
| 685 | EU717894 | Unassigned | Unassigned | Unassigned | 113_0 | Clustered | 9 | VC_113_0 | 9 | 0.519 | 1 | 0.519 | 0 | 0 | 0 | 1 | Ref |
| 686 | EU734170 | Unassigned | Unassigned | Unassigned | 386_0 | Clustered | 49 | VC_386_0 | 49 | 0.7941 | 1 | 0.7941 | 0 | 0 | 0 | 1 | Ref |
| 687 | EU734171 | Unassigned | Unassigned | Unassigned | 386_0 | Clustered | 49 | VC_386_0 | 49 | 0.7941 | 1 | 0.7941 | 0 | 0 | 0 | 1 | Ref |
| 688 | EU734172 | Unassigned | Unassigned | Unassigned | 451_0 | Clustered | 80 | VC_451_0 | 80 | 0.9991 | 1 | 0.9991 | 0 | 0 | 0 | 1 | Ref |
| 689 | EU734173 | Unassigned | Unassigned | Unassigned | 823_0 | Clustered | 124 | VC_823_0 | 124 | 0.9492 | 1 | 0.9492 | 0 | 0 | 0 | 1 | Ref |
| 690 | EU734174 | Unassigned | Unassigned | Unassigned | 392_0 | Clustered | 126 | VC_392_0 | 126 | 0.9621 | 1 | 0.9621 | 0 | 0 | 0 | 1 | Ref |
| 691 | EU744248 | Unassigned | Unassigned | Unassigned | 325_21 | Clustered | 5 | VC_325_21 | 5 | 0.0209 | 0.99999976 | 0.0209 | 0 | 0 | 0 | 0.8 | Ref |
| 692 | EU744249 | Unassigned | Unassigned | Unassigned | 325_13 | Clustered | 2 | VC_325_13 | 2 | 0.0082 | 0.957512 | 0.0079 | 0 | 0 | 0 | 1 | Ref |
| 693 | EU744250 | Unassigned | Unassigned | Unassigned | 217_0 | Clustered | 5 | VC_217_0 | 5 | 0.069 | 0.99999998 | 0.069 | 0 | 0 | 0 | 1 | Ref |
| 694 | EU744251 | Unassigned | Unassigned | Unassigned | 325_44 | Clustered | 3 | VC_325_44 | 3 | 0.0157 | 0.99859171 | 0.0157 | 0 | 0 | 0 | 1 | Ref |
| 695 | EU744252 | Unassigned | Unassigned | Unassigned | 325_13 | Clustered | 2 | VC_325_13 | 2 | 0.0082 | 0.957512 | 0.0079 | 0 | 0 | 0 | 1 | Ref |
| 696 | EU770221 | Unassigned | Unassigned | Unassigned | 372_0 | Clustered | 24 | VC_372_0 | 24 | 0.5539 | 1 | 0.5539 | 0 | 0 | 0 | 1 | Ref |
| 697 | EU770222 | Unassigned | Unassigned | Unassigned | 567_0 | Clustered | 8 | VC_567_0 | 8 | 0.4935 | 1 | 0.4935 | 0 | 0 | 0 | 1 | Ref |
| 698 | EU771092 | Unassigned | Unassigned | Unassigned | 817_0 | Clustered | 14 | VC_817_0 | 14 | 0.6342 | 1 | 0.6342 | 0 | 0 | 0 | 1 | Ref |
| 699 | EU794049 | Unassigned | Unassigned | Unassigned | 29_0 | Clustered | 10 | VC_29_0 | 10 | 0.5208 | 1 | 0.5208 | 0 | 0 | 0 | 1 | Ref |
| 700 | EU816588 | Unassigned | Unassigned | Unassigned | 551_6 | Clustered | 22 | VC_551_6 | 22 | 0.1214 | 1 | 0.1214 | 0 | 0 | 0 | 0.8095 | Ref |
| 701 | EU816589 | Unassigned | Unassigned | Unassigned | 509_0 | Clustered | 37 | VC_509_0 | 37 | 0.9689 | 1 | 0.9689 | 0 | 0 | 0 | 1 | Ref |
| 702 | EU816590 | Unassigned | Unassigned | Unassigned | 520_58 | Clustered | 3 | VC_520_58 | 3 | 0.021 | 0.9985993 | 0.021 | 0 | 0 | 0 | 1 | Ref |
| 703 | EU816591 | Unassigned | Unassigned | Unassigned | 551_2 | Clustered | 17 | VC_551_2 | 17 | 0.0973 | 1 | 0.0973 | 0 | 0 | 0 | 0.9779 | Ref |
| 704 | EU826466 | Unassigned | Unassigned | Unassigned | 559_0 | Clustered | 2 | VC_559_0 | 2 | 0.0112 | 0.95651281 | 0.0107 | 0 | 0 | 0 | 1 | Ref |
| 705 | EU826467 | Unassigned | Unassigned | Unassigned | 558_30 | Clustered | 3 | VC_558_30 | 3 | 0.0064 | 0.76574053 | 0.0049 | 0 | 0 | 0 | 1 | Ref |
| 706 | EU826468 | Unassigned | Unassigned | Unassigned | 558_25 | Clustered/Singleton | 1 | VC_558_25 | 1 | 0 | 1 | 0 | 0 | 0 | 0 | 1 | Ref |
| 707 | EU826469 | Unassigned | Unassigned | Unassigned | 558_26 | Clustered/Singleton | 1 | VC_558_26 | 1 | 0 | 1 | 0 | 0 | 0 | 0 | 1 | Ref |
| 708 | EU826470 | Unassigned | Unassigned | Unassigned | 325_35 | Clustered | 17 | VC_325_35 | 17 | 0.0732 | 1 | 0.0732 | 0 | 0 | 0 | 0.7721 | Ref |
| 709 | EU826471 | Unassigned | Unassigned | Unassigned | 558_2 | Clustered | 2 | VC_558_2 | 2 | 0.0032 | 0.66859979 | 0.0021 | 0 | 0 | 0 | 1 | Ref |
| 710 | EU855793 | Unassigned | Unassigned | Unassigned | 740_0 | Clustered | 3 | VC_740_0 | 3 | 0.202 | 0.99811828 | 0.2016 | 0 | 0 | 0 | 1 | Ref |
| 711 | EU861004 | Unassigned | Unassigned | Unassigned | 9_0 | Clustered | 38 | VC_9_0 | 38 | 0.4895 | 1 | 0.4895 | 0 | 0 | 0 | 1 | Ref |
| 712 | EU861005 | Unassigned | Unassigned | Unassigned | 8_0 | Clustered | 58 | VC_8_0 | 58 | 0.8306 | 1 | 0.8306 | 0 | 0 | 0 | 0.9946 | Ref |
| 713 | EU863408 | Unassigned | Unassigned | Unassigned | 590_0 | Clustered | 27 | VC_590_0 | 27 | 0.9947 | 1 | 0.9947 | 0 | 0 | 0 | 1 | Ref |
| 714 | EU863409 | Unassigned | Unassigned | Unassigned | 290_6 | Clustered | 12 | VC_290_6 | 12 | 0.1149 | 1 | 0.1149 | 0 | 0 | 0 | 0.8182 | Ref |
| 715 | EU874396 | Unassigned | Unassigned | Unassigned | 652_0 | Clustered | 4 | VC_652_0 | 4 | 0.4743 | 0.99998076 | 0.4743 | 0 | 0 | 0 | 1 | Ref |
| 716 | EU877232 | Unassigned | Unassigned | Unassigned | 329_7 | Clustered | 18 | VC_329_7 | 18 | 0.0938 | 1 | 0.0938 | 0 | 0 | 0 | 0.7778 | Ref |
| 717 | EU887664 | Unassigned | Unassigned | Unassigned | 829_0 | Clustered | 5 | VC_829_0 | 5 | 0.8821 | 0.99999832 | 0.8821 | 0 | 0 | 0 | 1 | Ref |
| 718 | F1CCG | Unassigned | Unassigned | Unassigned | 61_0 | Clustered | 12 | VC_61_0 | 12 | 0.7633 | 1 | 0.7633 | 0 | 0 | 0 | 1 | Ref |
| 719 | FJ000341 | Unassigned | Unassigned | Unassigned | 28_1 | Clustered | 53 | VC_28_1 | 53 | 0.7164 | 1 | 0.7164 | 0 | 0 | 0 | 0.9325 | Ref |
| 720 | FJ008913 | Unassigned | Unassigned | Unassigned | 830_0 | Clustered | 6 | VC_830_0 | 6 | 0.6536 | 1 | 0.6536 | 0 | 0 | 0 | 1 | Ref |
| 721 | FJ168659 | Unassigned | Unassigned | Unassigned | 518_0 | Clustered | 6 | VC_518_0 | 6 | 0.1295 | 1 | 0.1295 | 0 | 0 | 0 | 1 | Ref |
| 722 | FJ168660 | Unassigned | Unassigned | Unassigned | 565_0 | Clustered | 20 | VC_565_0 | 20 | 0.8398 | 1 | 0.8398 | 0 | 0 | 0 | 1 | Ref |
| 723 | FJ168661 | Unassigned | Unassigned | Unassigned | 565_0 | Clustered | 20 | VC_565_0 | 20 | 0.8398 | 1 | 0.8398 | 0 | 0 | 0 | 1 | Ref |
| 724 | FJ168662 | Unassigned | Unassigned | Unassigned | 565_0 | Clustered | 20 | VC_565_0 | 20 | 0.8398 | 1 | 0.8398 | 0 | 0 | 0 | 1 | Ref |
| 725 | FJ174690 | Unassigned | Unassigned | Unassigned | 520_31 | Clustered | 11 | VC_520_31 | 11 | 0.0897 | 1 | 0.0897 | 0 | 0 | 0 | 0.6 | Ref |
| 726 | FJ174691 | Unassigned | Unassigned | Unassigned | 567_0 | Clustered | 8 | VC_567_0 | 8 | 0.4935 | 1 | 0.4935 | 0 | 0 | 0 | 1 | Ref |
| 727 | FJ174693 | Unassigned | Unassigned | Unassigned | 520_71 | Clustered/Singleton | 1 | VC_520_71 | 1 | 0 | 1 | 0 | 0 | 0 | 0 | 1 | Ref |
| 728 | FJ174694 | Unassigned | Unassigned | Unassigned | 371_16 | Clustered | 42 | VC_371_16 | 42 | 0.1341 | 1 | 0.1341 | 0 | 0 | 0 | 0.7282 | Ref |
| 729 | FJ184280 | Unassigned | Unassigned | Unassigned | 41_1 | Clustered | 10 | VC_41_1 | 10 | 0.1178 | 1 | 0.1178 | 0 | 0 | 0 | 0.8222 | Ref |
| 730 | FJ188381 | Unassigned | Unassigned | Unassigned | 41_1 | Clustered | 10 | VC_41_1 | 10 | 0.1178 | 1 | 0.1178 | 0 | 0 | 0 | 0.8222 | Ref |
| 731 | FJ194439 | Unassigned | Unassigned | Unassigned | 386_0 | Clustered | 49 | VC_386_0 | 49 | 0.7941 | 1 | 0.7941 | 0 | 0 | 0 | 1 | Ref |
| 732 | FJ226752 | Unassigned | Unassigned | Unassigned | 254_0 | Clustered | 46 | VC_254_0 | 46 | 0.5159 | 1 | 0.5159 | 0 | 0 | 0 | 1 | Ref |
| 733 | FJ230960 | Unassigned | Unassigned | Unassigned | 87_0 | Clustered | 8 | VC_87_0 | 8 | 0.5545 | 1 | 0.5545 | 0 | 0 | 0 | 1 | Ref |
| 734 | FJ236310 | Unassigned | Unassigned | Unassigned | 252_0 | Clustered | 121 | VC_252_0 | 121 | 0.7376 | 1 | 0.7376 | 0 | 0 | 0 | 0.9999 | Ref |
| 735 | FJ373894 | Unassigned | Unassigned | Unassigned | 131_2 | Clustered | 7 | VC_131_2 | 7 | 0.0685 | 1 | 0.0685 | 0 | 0 | 0 | 0.9048 | Ref |
| 736 | FJ409640 | Unassigned | Unassigned | Unassigned | 82_0 | Clustered | 24 | VC_82_0 | 24 | 0.7351 | 1 | 0.7351 | 0 | 0 | 0 | 1 | Ref |
| 737 | FJ429185 | Unassigned | Unassigned | Unassigned | 534_0 | Clustered | 9 | VC_534_0 | 9 | 0.99 | 1 | 0.99 | 0 | 0 | 0 | 1 | Ref |
| 738 | FJ483837 | Unassigned | Unassigned | Unassigned | 315_0 | Clustered | 12 | VC_315_0 | 12 | 0.8947 | 1 | 0.8947 | 0 | 0 | 0 | 1 | Ref |
| 739 | FJ483838 | Unassigned | Unassigned | Unassigned | 315_0 | Clustered | 12 | VC_315_0 | 12 | 0.8947 | 1 | 0.8947 | 0 | 0 | 0 | 1 | Ref |
| 740 | FJ483839 | Unassigned | Unassigned | Unassigned | 315_0 | Clustered | 12 | VC_315_0 | 12 | 0.8947 | 1 | 0.8947 | 0 | 0 | 0 | 1 | Ref |
| 741 | FJ483840 | Unassigned | Unassigned | Unassigned | 188_0 | Clustered | 13 | VC_188_0 | 13 | 0.9973 | 0.99823802 | 0.9956 | 0 | 0 | 0 | 1 | Ref |
| 742 | FJ483841 | Unassigned | Unassigned | Unassigned | 188_0 | Clustered | 13 | VC_188_0 | 13 | 0.9973 | 0.99823802 | 0.9956 | 0 | 0 | 0 | 1 | Ref |
| 743 | FJ483842 | Unassigned | Unassigned | Unassigned | 188_0 | Clustered | 13 | VC_188_0 | 13 | 0.9973 | 0.99823802 | 0.9956 | 0 | 0 | 0 | 1 | Ref |
| 744 | FJ483843 | Unassigned | Unassigned | Unassigned | 188_0 | Clustered | 13 | VC_188_0 | 13 | 0.9973 | 0.99823802 | 0.9956 | 0 | 0 | 0 | 1 | Ref |
| 745 | FJ483844 | Unassigned | Unassigned | Unassigned | 188_0 | Clustered | 13 | VC_188_0 | 13 | 0.9973 | 0.99823802 | 0.9956 | 0 | 0 | 0 | 1 | Ref |
| 746 | FJ539132 | Unassigned | Unassigned | Unassigned | 188_0 | Clustered | 13 | VC_188_0 | 13 | 0.9973 | 0.99823802 | 0.9956 | 0 | 0 | 0 | 1 | Ref |
| 747 | FJ539133 | Unassigned | Unassigned | Unassigned | 188_0 | Clustered | 13 | VC_188_0 | 13 | 0.9973 | 0.99823802 | 0.9956 | 0 | 0 | 0 | 1 | Ref |
| 748 | FJ539134 | Unassigned | Unassigned | Unassigned | 188_0 | Clustered | 13 | VC_188_0 | 13 | 0.9973 | 0.99823802 | 0.9956 | 0 | 0 | 0 | 1 | Ref |
| 749 | FJ539135 | Unassigned | Unassigned | Unassigned | 188_0 | Clustered | 13 | VC_188_0 | 13 | 0.9973 | 0.99823802 | 0.9956 | 0 | 0 | 0 | 1 | Ref |
| 750 | FJ591093 | Unassigned | Unassigned | Unassigned | 833_0 | Clustered | 16 | VC_833_0 | 16 | 0.9351 | 1 | 0.9351 | 0 | 0 | 0 | 1 | Ref |
| 751 | FJ591094 | Unassigned | Unassigned | Unassigned | 833_0 | Clustered | 16 | VC_833_0 | 16 | 0.9351 | 1 | 0.9351 | 0 | 0 | 0 | 1 | Ref |
| 752 | FJ641182 | Unassigned | Unassigned | Unassigned | 509_0 | Clustered | 37 | VC_509_0 | 37 | 0.9689 | 1 | 0.9689 | 0 | 0 | 0 | 1 | Ref |
| 753 | FJ685651 | Unassigned | Unassigned | Unassigned | 834_0 | Clustered | 7 | VC_834_0 | 7 | 0.9845 | 0.94276893 | 0.9282 | 0 | 0 | 0 | 1 | Ref |
| 754 | FJ706171 | Unassigned | Unassigned | Unassigned | 765_0 | Clustered | 132 | VC_765_0 | 132 | 1 | 1 | 1 | 0 | 0 | 0 | 1 | Ref |
| 755 | FJ706172 | Unassigned | Unassigned | Unassigned | 765_0 | Clustered | 132 | VC_765_0 | 132 | 1 | 1 | 1 | 0 | 0 | 0 | 1 | Ref |
| 756 | FJ713816 | Unassigned | Unassigned | Unassigned | 7_0 | Clustered | 28 | VC_7_0 | 28 | 0.3916 | 1 | 0.3916 | 0 | 0 | 0 | 0.9947 | Ref |
| 757 | FJ750948 | Unassigned | Unassigned | Unassigned | 835_0 | Clustered | 67 | VC_835_0 | 67 | 0.9624 | 1 | 0.9624 | 0 | 0 | 0 | 1 | Ref |
| 758 | FJ822135 | Unassigned | Unassigned | Unassigned | 93_0 | Clustered | 2 | VC_93_0 | 2 | 0.1063 | 0.95665672 | 0.1017 | 0 | 0 | 0 | 1 | Ref |
| 759 | FJ839692 | Unassigned | Unassigned | Unassigned | 283_84 | Clustered/Singleton | 1 | VC_283_84 | 1 | 0 | 1 | 0 | 0 | 0 | 0 | 1 | Ref |
| 760 | FJ839693 | Unassigned | Unassigned | Unassigned | 283_3 | Clustered | 3 | VC_283_3 | 3 | 0.0055 | 0.9968017 | 0.0055 | 0 | 0 | 0 | 1 | Ref |
| 761 | FJ848881 | Unassigned | Unassigned | Unassigned | 202_0 | Clustered | 102 | VC_202_0 | 102 | 0.4049 | 1 | 0.4049 | 0 | 0 | 0 | 0.7876 | Ref |
| 762 | FJ848882 | Unassigned | Unassigned | Unassigned | 202_0 | Clustered | 102 | VC_202_0 | 102 | 0.4049 | 1 | 0.4049 | 0 | 0 | 0 | 0.7876 | Ref |
| 763 | FJ848883 | Unassigned | Unassigned | Unassigned | 202_0 | Clustered | 102 | VC_202_0 | 102 | 0.4049 | 1 | 0.4049 | 0 | 0 | 0 | 0.7876 | Ref |
| 764 | FJ848884 | Unassigned | Unassigned | Unassigned | 202_0 | Clustered | 102 | VC_202_0 | 102 | 0.4049 | 1 | 0.4049 | 0 | 0 | 0 | 0.7876 | Ref |
| 765 | FJ848885 | Unassigned | Unassigned | Unassigned | 202_0 | Clustered | 102 | VC_202_0 | 102 | 0.4049 | 1 | 0.4049 | 0 | 0 | 0 | 0.7876 | Ref |
| 766 | FJ849058 | Unassigned | Unassigned | Unassigned | 303_0 | Clustered | 155 | VC_303_0 | 155 | 0.9979 | 1 | 0.9979 | 0 | 0 | 0 | 1 | Ref |
| 767 | FJ867910 | Unassigned | Unassigned | Unassigned | 304_0 | Clustered | 11 | VC_304_0 | 11 | 0.7869 | 1 | 0.7869 | 0 | 0 | 0 | 1 | Ref |
| 768 | FJ867911 | Unassigned | Unassigned | Unassigned | 304_0 | Clustered | 11 | VC_304_0 | 11 | 0.7869 | 1 | 0.7869 | 0 | 0 | 0 | 1 | Ref |
| 769 | FJ867912 | Unassigned | Unassigned | Unassigned | 304_0 | Clustered | 11 | VC_304_0 | 11 | 0.7869 | 1 | 0.7869 | 0 | 0 | 0 | 1 | Ref |
| 770 | FJ867913 | Unassigned | Unassigned | Unassigned | 304_0 | Clustered | 11 | VC_304_0 | 11 | 0.7869 | 1 | 0.7869 | 0 | 0 | 0 | 1 | Ref |
| 771 | FJ867914 | Unassigned | Unassigned | Unassigned | 304_0 | Clustered | 11 | VC_304_0 | 11 | 0.7869 | 1 | 0.7869 | 0 | 0 | 0 | 1 | Ref |
| 772 | FJ870915 | Unassigned | Unassigned | Unassigned | 599_0 | Clustered | 50 | VC_599_0 | 50 | 0.9989 | 1 | 0.9989 | 0 | 0 | 0 | 1 | Ref |
| 773 | FJ870916 | Unassigned | Unassigned | Unassigned | 599_0 | Clustered | 50 | VC_599_0 | 50 | 0.9989 | 1 | 0.9989 | 0 | 0 | 0 | 1 | Ref |
| 774 | FJ870917 | Unassigned | Unassigned | Unassigned | 599_0 | Clustered | 50 | VC_599_0 | 50 | 0.9989 | 1 | 0.9989 | 0 | 0 | 0 | 1 | Ref |
| 775 | FJ896200 | Unassigned | Unassigned | Unassigned | 837_0 | Clustered | 29 | VC_837_0 | 29 | 0.9955 | 1 | 0.9955 | 0 | 0 | 0 | 1 | Ref |
| 776 | FJ904927 | Unassigned | Unassigned | Unassigned | 16_0 | Clustered | 12 | VC_16_0 | 12 | 0.6615 | 1 | 0.6615 | 0 | 0 | 0 | 1 | Ref |
| 777 | FJ937737 | Unassigned | Unassigned | Unassigned | 162_0 | Clustered | 18 | VC_162_0 | 18 | 0.7403 | 1 | 0.7403 | 0 | 0 | 0 | 0.8627 | Ref |
| 778 | FJ965538 | Unassigned | Unassigned | Unassigned | 255_0 | Clustered | 13 | VC_255_0 | 13 | 0.2048 | 1 | 0.2048 | 0 | 0 | 0 | 1 | Ref |
| 779 | FJ973624 | Unassigned | Unassigned | Unassigned | 761_0 | Clustered | 67 | VC_761_0 | 67 | 0.8947 | 1 | 0.8947 | 0 | 0 | 0 | 1 | Ref |
| 780 | FM164764 | Unassigned | Unassigned | Unassigned | 348_0 | Clustered | 22 | VC_348_0 | 22 | 0.9602 | 1 | 0.9602 | 0 | 0 | 0 | 1 | Ref |
| 781 | FM180578 | Unassigned | Unassigned | Unassigned | 41_1 | Clustered | 10 | VC_41_1 | 10 | 0.1178 | 1 | 0.1178 | 0 | 0 | 0 | 0.8222 | Ref |
| 782 | FM201281 | Unassigned | Unassigned | Unassigned | 79_2 | Clustered/Singleton | 1 | VC_79_2 | 1 | 0 | 1 | 0 | 0 | 0 | 0 | 1 | Ref |
| 783 | FM201282 | Unassigned | Unassigned | Unassigned | 79_0 | Clustered | 64 | VC_79_0 | 64 | 0.4185 | 1 | 0.4185 | 0 | 0 | 0 | 0.7649 | Ref |
| 784 | FM207411 | Unassigned | Unassigned | Unassigned | 433_8 | Clustered/Singleton | 1 | VC_433_8 | 1 | 0 | 1 | 0 | 0 | 0 | 0 | 1 | Ref |
| 785 | FM864213 | Unassigned | Unassigned | Unassigned | 632_0 | Clustered | 37 | VC_632_0 | 37 | 0.581 | 1 | 0.581 | 0 | 0 | 0 | 0.9234 | Ref |
| 786 | FM887021 | Unassigned | Unassigned | Unassigned | 79_7 | Clustered | 33 | VC_79_7 | 33 | 0.216 | 1 | 0.216 | 0 | 0 | 0 | 0.8409 | Ref |
| 787 | FM897211 | Unassigned | Unassigned | Unassigned | 79_7 | Clustered | 33 | VC_79_7 | 33 | 0.216 | 1 | 0.216 | 0 | 0 | 0 | 0.8409 | Ref |
| 788 | FN263372 | Unassigned | Unassigned | Unassigned | 423_0 | Clustered | 45 | VC_423_0 | 45 | 0.9484 | 1 | 0.9484 | 0 | 0 | 0 | 1 | Ref |
| 789 | FN297812 | Unassigned | Unassigned | Unassigned | 23_0 | Clustered | 3 | VC_23_0 | 3 | 0.4305 | 0.99837378 | 0.4298 | 0 | 0 | 0 | 1 | Ref |
| 790 | FN391954 | Unassigned | Unassigned | Unassigned | 333_0 | Clustered | 25 | VC_333_0 | 25 | 0.4681 | 1 | 0.4681 | 0 | 0 | 0 | 1 | Ref |
| 791 | FN422398 | Unassigned | Unassigned | Unassigned | 839_0 | Clustered | 5 | VC_839_0 | 5 | 0.165 | 0.99999995 | 0.165 | 0 | 0 | 0 | 1 | Ref |
| 792 | FN422399 | Unassigned | Unassigned | Unassigned | 838_0 | Clustered | 29 | VC_838_0 | 29 | 0.8611 | 1 | 0.8611 | 0 | 0 | 0 | 1 | Ref |
| 793 | FN594518 | Unassigned | Unassigned | Unassigned | 155_0 | Clustered | 14 | VC_155_0 | 14 | 0.6095 | 1 | 0.6095 | 0 | 0 | 0 | 1 | Ref |
| 794 | FN667788 | Unassigned | Unassigned | Unassigned | 843_0 | Clustered | 5 | VC_843_0 | 5 | 0.2843 | 0.99999994 | 0.2843 | 0 | 0 | 0 | 1 | Ref |
| 795 | FN667789 | Unassigned | Unassigned | Unassigned | 843_0 | Clustered | 5 | VC_843_0 | 5 | 0.2843 | 0.99999994 | 0.2843 | 0 | 0 | 0 | 1 | Ref |
| 796 | FN997652 | Unassigned | Unassigned | Unassigned | 632_0 | Clustered | 37 | VC_632_0 | 37 | 0.581 | 1 | 0.581 | 0 | 0 | 0 | 0.9234 | Ref |
| 797 | FO818745 | Unassigned | Unassigned | Unassigned | 247_0 | Clustered | 14 | VC_247_0 | 14 | 0.9119 | 1 | 0.9119 | 0 | 0 | 0 | 0.967 | Ref |
| 798 | FQ312032 | Unassigned | Unassigned | Unassigned | 133_22 | Clustered | 13 | VC_133_22 | 13 | 0.0804 | 1 | 0.0804 | 0 | 0 | 0 | 0.8205 | Ref |
| 799 | FQ482083 | Unassigned | Unassigned | Unassigned | 328_0 | Clustered | 11 | VC_328_0 | 11 | 0.4707 | 1 | 0.4707 | 0 | 0 | 0 | 1 | Ref |
| 800 | FQ482084 | Unassigned | Unassigned | Unassigned | 453_0 | Clustered | 6 | VC_453_0 | 6 | 0.114 | 1 | 0.114 | 0 | 0 | 0 | 1 | Ref |
| 801 | FQ482086 | Unassigned | Unassigned | Unassigned | 453_0 | Clustered | 6 | VC_453_0 | 6 | 0.114 | 1 | 0.114 | 0 | 0 | 0 | 1 | Ref |
| 802 | FR667955 | Unassigned | Unassigned | Unassigned | 392_0 | Clustered | 126 | VC_392_0 | 126 | 0.9621 | 1 | 0.9621 | 0 | 0 | 0 | 1 | Ref |
| 803 | FR671405 | Unassigned | Unassigned | Unassigned | 414_0 | Clustered | 37 | VC_414_0 | 37 | 0.585 | 1 | 0.585 | 0 | 0 | 0 | 0.9474 | Ref |
| 804 | FR671406 | Unassigned | Unassigned | Unassigned | 414_0 | Clustered | 37 | VC_414_0 | 37 | 0.585 | 1 | 0.585 | 0 | 0 | 0 | 0.9474 | Ref |
| 805 | FR671407 | Unassigned | Unassigned | Unassigned | 414_0 | Clustered | 37 | VC_414_0 | 37 | 0.585 | 1 | 0.585 | 0 | 0 | 0 | 0.9474 | Ref |
| 806 | FR671408 | Unassigned | Unassigned | Unassigned | 317_2 | Clustered | 20 | VC_317_2 | 20 | 0.1976 | 1 | 0.1976 | 0 | 0 | 0 | 0.6842 | Ref |
| 807 | FR671409 | Unassigned | Unassigned | Unassigned | 317_2 | Clustered | 20 | VC_317_2 | 20 | 0.1976 | 1 | 0.1976 | 0 | 0 | 0 | 0.6842 | Ref |
| 808 | FR671410 | Unassigned | Unassigned | Unassigned | 414_0 | Clustered | 37 | VC_414_0 | 37 | 0.585 | 1 | 0.585 | 0 | 0 | 0 | 0.9474 | Ref |

| | | | | | | | | | | | | | | | | | |
|---|---|---|---|---|---|---|---|---|---|---|---|---|---|---|---|---|---|
| 809 | FR671411 | Unassigned | Unassigned | Unassigned | 414_0 | Clustered | 37 | VC_414_0 | 37 | 0.585 | 1 | 0.585 | 0 | 0 | 0 | 0.9474 | Ref |
| 810 | FR682616 | Unassigned | Unassigned | Unassigned | 833_0 | Clustered | 16 | VC_833_0 | 16 | 0.9351 | 1 | 0.9351 | 0 | 0 | 0 | 1 | Ref |
| 811 | FR687252 | Unassigned | Unassigned | Unassigned | 110_0 | Clustered | 4 | VC_110_0 | 4 | 0.1143 | 0.99998732 | 0.1143 | 0 | 0 | 0 | 1 | Ref |
| 812 | FR719956 | Unassigned | Unassigned | Unassigned | 833_0 | Clustered | 16 | VC_833_0 | 16 | 0.9351 | 1 | 0.9351 | 0 | 0 | 0 | 1 | Ref |
| 813 | FR751545 | Unassigned | Unassigned | Unassigned | 156_0 | Clustered | 12 | VC_156_0 | 12 | 0.9062 | 1 | 0.9062 | 0 | 0 | 0 | 1 | Ref |
| 814 | FR775895 | Unassigned | Unassigned | Unassigned | 848_0 | Clustered | 28 | VC_848_0 | 28 | 0.7927 | 1 | 0.7927 | 0 | 0 | 0 | 1 | Ref |
| 815 | FR823298 | Unassigned | Unassigned | Unassigned | 130_0 | Clustered | 19 | VC_130_0 | 19 | 0.6068 | 1 | 0.6068 | 0 | 0 | 0 | 1 | Ref |
| 816 | FR823450 | Unassigned | Unassigned | Unassigned | 844_0 | Clustered | 27 | VC_844_0 | 27 | 0.9385 | 1 | 0.9385 | 0 | 0 | 0 | 1 | Ref |
| 817 | FR852584 | Unassigned | Unassigned | Unassigned | 149_8 | Clustered | 66 | VC_149_8 | 66 | 0.4565 | 1 | 0.4565 | 0 | 0 | 0 | 0.8182 | Ref |
| 818 | GQ141189 | Unassigned | Unassigned | Unassigned | 852_0 | Clustered | 2 | VC_852_0 | 2 | 0.4064 | 0.94189878 | 0.3828 | 0 | 0 | 0 | 1 | Ref |
| 819 | GQ149088 | Unassigned | Unassigned | Unassigned | 743_0 | Clustered | 15 | VC_743_0 | 15 | 1 | 1 | 1 | 0 | 0 | 0 | 1 | Ref |
| 820 | GQ153912 | Unassigned | Unassigned | Unassigned | 303_0 | Clustered | 155 | VC_303_0 | 155 | 0.9979 | 1 | 0.9979 | 0 | 0 | 0 | 1 | Ref |
| 821 | GQ153913 | Unassigned | Unassigned | Unassigned | 303_0 | Clustered | 155 | VC_303_0 | 155 | 0.9979 | 1 | 0.9979 | 0 | 0 | 0 | 1 | Ref |
| 822 | GQ153914 | Unassigned | Unassigned | Unassigned | 303_0 | Clustered | 155 | VC_303_0 | 155 | 0.9979 | 1 | 0.9979 | 0 | 0 | 0 | 1 | Ref |
| 823 | GQ153915 | Unassigned | Unassigned | Unassigned | 303_0 | Clustered | 155 | VC_303_0 | 155 | 0.9979 | 1 | 0.9979 | 0 | 0 | 0 | 1 | Ref |
| 824 | GQ153916 | Unassigned | Unassigned | Unassigned | 61_0 | Clustered | 12 | VC_61_0 | 12 | 0.7633 | 1 | 0.7633 | 0 | 0 | 0 | 1 | Ref |
| 825 | GQ153917 | Unassigned | Unassigned | Unassigned | 61_0 | Clustered | 12 | VC_61_0 | 12 | 0.7633 | 1 | 0.7633 | 0 | 0 | 0 | 1 | Ref |
| 826 | GQ153918 | Unassigned | Unassigned | Unassigned | 61_0 | Clustered | 12 | VC_61_0 | 12 | 0.7633 | 1 | 0.7633 | 0 | 0 | 0 | 1 | Ref |
| 827 | GQ153919 | Unassigned | Unassigned | Unassigned | 61_0 | Clustered | 12 | VC_61_0 | 12 | 0.7633 | 1 | 0.7633 | 0 | 0 | 0 | 1 | Ref |
| 828 | GQ153920 | Unassigned | Unassigned | Unassigned | 360_0 | Clustered | 358 | VC_360_0 | 358 | 0.9615 | 1 | 0.9615 | 0 | 0 | 0 | 1 | Ref |
| 829 | GQ153921 | Unassigned | Unassigned | Unassigned | 362_0 | Clustered | 2 | VC_362_0 | 2 | 0.002 | 0.83307735 | 0.0017 | 0 | 0 | 0 | 1 | Ref |
| 830 | GQ153922 | Unassigned | Unassigned | Unassigned | 360_0 | Clustered | 358 | VC_360_0 | 358 | 0.9615 | 1 | 0.9615 | 0 | 0 | 0 | 1 | Ref |
| 831 | GQ153923 | Unassigned | Unassigned | Unassigned | 360_0 | Clustered | 358 | VC_360_0 | 358 | 0.9615 | 1 | 0.9615 | 0 | 0 | 0 | 1 | Ref |
| 832 | GQ241246 | Unassigned | Unassigned | Unassigned | 855_0 | Clustered | 2 | VC_855_0 | 2 | 0.0285 | 0.89378929 | 0.0255 | 0 | 0 | 0 | 1 | Ref |
| 833 | GQ303259 | Unassigned | Unassigned | Unassigned | 371_3 | Clustered | 22 | VC_371_3 | 22 | 0.0787 | 1 | 0.0787 | 0 | 0 | 0 | 0.9048 | Ref |
| 834 | GQ303260 | Unassigned | Unassigned | Unassigned | 558_1 | Clustered | 2 | VC_558_1 | 2 | 0.0032 | 0.78405032 | 0.0025 | 0 | 0 | 0 | 0 | Ref |
| 835 | GQ303261 | Unassigned | Unassigned | Unassigned | 761_0 | Clustered | 67 | VC_761_0 | 67 | 0.8947 | 1 | 0.8947 | 0 | 0 | 0 | 1 | Ref |
| 836 | GQ303262 | Unassigned | Unassigned | Unassigned | 558_82 | Clustered | 3 | VC_558_82 | 3 | 0.0064 | 0.88433874 | 0.0057 | 0 | 0 | 0 | 0.6667 | Ref |
| 837 | GQ303263 | Unassigned | Unassigned | Unassigned | 517_1 | Clustered | 108 | VC_517_1 | 108 | 0.8483 | 1 | 0.8483 | 0 | 0 | 0 | 0.9955 | Ref |
| 838 | GQ303264 | Unassigned | Unassigned | Unassigned | 371_15 | Clustered | 44 | VC_371_15 | 44 | 0.1391 | 1 | 0.1391 | 0 | 0 | 0 | 0.7474 | Ref |
| 839 | GQ303265 | Unassigned | Unassigned | Unassigned | 551_7 | Clustered | 28 | VC_551_7 | 28 | 0.1661 | 1 | 0.1661 | 0 | 0 | 0 | 0.8519 | Ref |
| 840 | GQ303266 | Unassigned | Unassigned | Unassigned | 371_15 | Clustered | 44 | VC_371_15 | 44 | 0.1391 | 1 | 0.1391 | 0 | 0 | 0 | 0.7474 | Ref |
| 841 | GQ357916 | Unassigned | Unassigned | Unassigned | 50_0 | Clustered | 9 | VC_50_0 | 9 | 0.5531 | 1 | 0.5531 | 0 | 0 | 0 | 1 | Ref |
| 842 | GQ379227 | Unassigned | Unassigned | Unassigned | 374_0 | Clustered | 2 | VC_374_0 | 2 | 0.1258 | 0.95578317 | 0.1203 | 0 | 0 | 0 | 1 | Ref |
| 843 | GQ398772 | Unassigned | Unassigned | Unassigned | 10_0 | Clustered | 25 | VC_10_0 | 25 | 0.3467 | 1 | 0.3467 | 0 | 0 | 0 | 0.9733 | Ref |
| 844 | GQ403788 | Unassigned | Unassigned | Unassigned | 740_0 | Clustered | 3 | VC_740_0 | 3 | 0.202 | 0.99811828 | 0.2016 | 0 | 0 | 0 | 1 | Ref |
| 845 | GQ403789 | Unassigned | Unassigned | Unassigned | 27_0 | Clustered | 2 | VC_27_0 | 2 | 1 | 1 | 1 | 0 | 0 | 0 | 1 | Ref |
| 846 | GQ413937 | Unassigned | Unassigned | Unassigned | 823_0 | Clustered | 124 | VC_823_0 | 124 | 0.9492 | 1 | 0.9492 | 0 | 0 | 0 | 1 | Ref |
| 847 | GQ413938 | Unassigned | Unassigned | Unassigned | 111_0 | Clustered | 91 | VC_111_0 | 91 | 0.9587 | 1 | 0.9587 | 0 | 0 | 0 | 1 | Ref |
| 848 | GQ421471 | Unassigned | Unassigned | Unassigned | 280_0 | Clustered | 2 | VC_280_0 | 2 | 0.34 | 0.95158328 | 0.3235 | 0 | 0 | 0 | 1 | Ref |
| 849 | GQ422450 | Unassigned | Unassigned | Unassigned | 28_1 | Clustered | 53 | VC_28_1 | 53 | 0.7164 | 1 | 0.7164 | 0 | 0 | 0 | 0.9325 | Ref |
| 850 | GQ422451 | Unassigned | Unassigned | Unassigned | 28_1 | Clustered | 53 | VC_28_1 | 53 | 0.7164 | 1 | 0.7164 | 0 | 0 | 0 | 0.9325 | Ref |
| 851 | GQ443085 | Unassigned | Unassigned | Unassigned | 815_0 | Clustered | 8 | VC_815_0 | 8 | 0.9801 | 1 | 0.9801 | 0 | 0 | 0 | 1 | Ref |
| 852 | GQ451696 | Unassigned | Unassigned | Unassigned | 862_0 | Clustered | 11 | VC_862_0 | 11 | 0.6745 | 1 | 0.6745 | 0 | 0 | 0 | 1 | Ref |
| 853 | GQ452243 | Unassigned | Unassigned | Unassigned | 267_0 | Clustered | 2 | VC_267_0 | 2 | 0.0988 | 0.95557934 | 0.0945 | 0 | 0 | 0 | 1 | Ref |
| 854 | GQ468526 | Unassigned | Unassigned | Unassigned | 386_0 | Clustered | 49 | VC_386_0 | 49 | 0.7941 | 1 | 0.7941 | 0 | 0 | 0 | 1 | Ref |
| 855 | GQ478081 | Unassigned | Unassigned | Unassigned | 774_0 | Clustered | 5 | VC_774_0 | 5 | 0.3282 | 0.99999996 | 0.3282 | 0 | 0 | 0 | 1 | Ref |
| 856 | GQ478082 | Unassigned | Unassigned | Unassigned | 774_0 | Clustered | 5 | VC_774_0 | 5 | 0.3282 | 0.99999996 | 0.3282 | 0 | 0 | 0 | 1 | Ref |
| 857 | GQ478083 | Unassigned | Unassigned | Unassigned | 774_0 | Clustered | 5 | VC_774_0 | 5 | 0.3282 | 0.99999996 | 0.3282 | 0 | 0 | 0 | 1 | Ref |
| 858 | GQ478084 | Unassigned | Unassigned | Unassigned | 774_0 | Clustered | 5 | VC_774_0 | 5 | 0.3282 | 0.99999996 | 0.3282 | 0 | 0 | 0 | 1 | Ref |
| 859 | GQ478085 | Unassigned | Unassigned | Unassigned | 774_0 | Clustered | 5 | VC_774_0 | 5 | 0.3282 | 0.99999996 | 0.3282 | 0 | 0 | 0 | 1 | Ref |
| 860 | GQ502199 | Unassigned | Unassigned | Unassigned | 835_0 | Clustered | 67 | VC_835_0 | 67 | 0.9624 | 1 | 0.9624 | 0 | 0 | 0 | 1 | Ref |
| 861 | GQ866233 | Unassigned | Unassigned | Unassigned | 426_0 | Clustered | 2 | VC_426_0 | 2 | 0.1976 | 0.95643941 | 0.189 | 0 | 0 | 0 | 1 | Ref |
| 862 | GQ915271 | Unassigned | Unassigned | Unassigned | 7_0 | Clustered | 28 | VC_7_0 | 28 | 0.3916 | 1 | 0.3916 | 0 | 0 | 0 | 0.9947 | Ref |
| 863 | GQ919031 | Unassigned | Unassigned | Unassigned | 864_0 | Clustered | 2 | VC_864_0 | 2 | 1 | 1 | 1 | 0 | 0 | 0 | 1 | Ref |
| 864 | GQ979703 | Unassigned | Unassigned | Unassigned | 202_0 | Clustered | 102 | VC_202_0 | 102 | 0.4049 | 1 | 0.4049 | 0 | 0 | 0 | 0.7876 | Ref |
| 865 | GQ981382 | Unassigned | Unassigned | Unassigned | 290_1 | Clustered | 8 | VC_290_1 | 8 | 0.0757 | 1 | 0.0757 | 0 | 0 | 0 | 0.8571 | Ref |
| 866 | GU060500 | Unassigned | Unassigned | Unassigned | 520_31 | Clustered | 11 | VC_520_31 | 11 | 0.0897 | 1 | 0.0897 | 0 | 0 | 0 | 0.6 | Ref |
| 867 | GU070616 | Unassigned | Unassigned | Unassigned | 582_5 | Clustered | 4 | VC_582_5 | 4 | 0.0528 | 0.99935625 | 0.0527 | 0 | 0 | 0 | 0.8333 | Ref |
| 868 | GU071091 | Unassigned | Unassigned | Unassigned | 392_0 | Clustered | 126 | VC_392_0 | 126 | 0.9621 | 1 | 0.9621 | 0 | 0 | 0 | 1 | Ref |
| 869 | GU071092 | Unassigned | Unassigned | Unassigned | 432_9 | Clustered | 3 | VC_432_9 | 3 | 0.0372 | 0.99851312 | 0.0371 | 0 | 0 | 0 | 1 | Ref |
| 870 | GU071093 | Unassigned | Unassigned | Unassigned | 340_0 | Clustered | 31 | VC_340_0 | 31 | 0.9549 | 1 | 0.9549 | 0 | 0 | 0 | 1 | Ref |
| 871 | GU071094 | Unassigned | Unassigned | Unassigned | 655_8 | Clustered/Singleton | 1 | VC_655_8 | 1 | 0 | 1 | 0 | 0 | 0 | 0 | 1 | Ref |
| 872 | GU071095 | Unassigned | Unassigned | Unassigned | 432_2 | Clustered/Singleton | 1 | VC_432_2 | 1 | 0 | 1 | 0 | 0 | 0 | 0 | 1 | Ref |
| 873 | GU071097 | Unassigned | Unassigned | Unassigned | 655_9 | Clustered/Singleton | 1 | VC_655_9 | 1 | 0 | 1 | 0 | 0 | 0 | 0 | 1 | Ref |
| 874 | GU071098 | Unassigned | Unassigned | Unassigned | 432_10 | Clustered | 2 | VC_432_10 | 2 | 0.0216 | 0.95771017 | 0.0207 | 0 | 0 | 0 | 1 | Ref |
| 875 | GU071099 | Unassigned | Unassigned | Unassigned | 655_0 | Clustered | 2 | VC_655_0 | 2 | 0.0111 | 0.95704495 | 0.0106 | 0 | 0 | 0 | 1 | Ref |
| 876 | GU071100 | Unassigned | Unassigned | Unassigned | 340_0 | Clustered | 31 | VC_340_0 | 31 | 0.9549 | 1 | 0.9549 | 0 | 0 | 0 | 1 | Ref |
| 877 | GU071102 | Unassigned | Unassigned | Unassigned | 340_0 | Clustered | 31 | VC_340_0 | 31 | 0.9549 | 1 | 0.9549 | 0 | 0 | 0 | 1 | Ref |
| 878 | GU071103 | Unassigned | Unassigned | Unassigned | 655_1 | Clustered/Singleton | 1 | VC_655_1 | 1 | 0 | 1 | 0 | 0 | 0 | 0 | 1 | Ref |
| 879 | GU071104 | Unassigned | Unassigned | Unassigned | 340_0 | Clustered | 31 | VC_340_0 | 31 | 0.9549 | 1 | 0.9549 | 0 | 0 | 0 | 1 | Ref |
| 880 | GU071105 | Unassigned | Unassigned | Unassigned | 433_4 | Clustered | 9 | VC_433_4 | 9 | 0.1285 | 1 | 0.1285 | 0 | 0 | 0 | 1 | Ref |
| 881 | GU071106 | Unassigned | Unassigned | Unassigned | 655_6 | Clustered | 2 | VC_655_6 | 2 | 0.0078 | 0.95775168 | 0.0075 | 0 | 0 | 0 | 1 | Ref |
| 882 | GU071107 | Unassigned | Unassigned | Unassigned | 340_0 | Clustered | 31 | VC_340_0 | 31 | 0.9549 | 1 | 0.9549 | 0 | 0 | 0 | 1 | Ref |
| 883 | GU080336 | Unassigned | Unassigned | Unassigned | 611_0 | Clustered | 2 | VC_611_0 | 2 | 1 | 1 | 1 | 0 | 0 | 0 | 1 | Ref |
| 884 | GU169904 | Unassigned | Unassigned | Unassigned | 329_15 | Clustered/Singleton | 1 | VC_329_15 | 1 | 0 | 1 | 0 | 0 | 0 | 0 | 1 | Ref |
| 885 | GU196277 | Unassigned | Unassigned | Unassigned | 638_0 | Clustered | 45 | VC_638_0 | 45 | 0.7741 | 1 | 0.7741 | 0 | 0 | 0 | 1 | Ref |
| 886 | GU196278 | Unassigned | Unassigned | Unassigned | 638_0 | Clustered | 45 | VC_638_0 | 45 | 0.7741 | 1 | 0.7741 | 0 | 0 | 0 | 1 | Ref |
| 887 | GU196279 | Unassigned | Unassigned | Unassigned | 638_0 | Clustered | 45 | VC_638_0 | 45 | 0.7741 | 1 | 0.7741 | 0 | 0 | 0 | 1 | Ref |
| 888 | GU196280 | Unassigned | Unassigned | Unassigned | 638_0 | Clustered | 45 | VC_638_0 | 45 | 0.7741 | 1 | 0.7741 | 0 | 0 | 0 | 1 | Ref |
| 889 | GU196281 | Unassigned | Unassigned | Unassigned | 638_0 | Clustered | 45 | VC_638_0 | 45 | 0.7741 | 1 | 0.7741 | 0 | 0 | 0 | 1 | Ref |
| 890 | GU229986 | Unassigned | Unassigned | Unassigned | 652_0 | Clustered | 4 | VC_652_0 | 4 | 0.4743 | 0.99998076 | 0.4743 | 0 | 0 | 0 | 1 | Ref |
| 891 | GU247132 | Unassigned | Unassigned | Unassigned | 325_14 | Clustered | 3 | VC_325_14 | 3 | 0.0094 | 0.99859159 | 0.0094 | 0 | 0 | 0 | 1 | Ref |
| 892 | GU247133 | Unassigned | Unassigned | Unassigned | 371_16 | Clustered | 42 | VC_371_16 | 42 | 0.1341 | 1 | 0.1341 | 0 | 0 | 0 | 0.7282 | Ref |
| 893 | GU247134 | Unassigned | Unassigned | Unassigned | 371_16 | Clustered | 42 | VC_371_16 | 42 | 0.1341 | 1 | 0.1341 | 0 | 0 | 0 | 0.7282 | Ref |
| 894 | GU295964 | Unassigned | Unassigned | Unassigned | 668_0 | Clustered | 25 | VC_668_0 | 25 | 0.5539 | 1 | 0.5539 | 0 | 0 | 0 | 0.96 | Ref |
| 895 | GU296433 | Unassigned | Unassigned | Unassigned | 844_0 | Clustered | 27 | VC_844_0 | 27 | 0.9385 | 1 | 0.9385 | 0 | 0 | 0 | 1 | Ref |
| 896 | GU321093 | Unassigned | Unassigned | Unassigned | 834_0 | Clustered | 7 | VC_834_0 | 7 | 0.9845 | 0.94276893 | 0.9282 | 0 | 0 | 0 | 1 | Ref |
| 897 | GU323708 | Unassigned | Unassigned | Unassigned | 381_0 | Clustered | 2 | VC_381_0 | 2 | 0.0829 | 0.95381392 | 0.079 | 0 | 0 | 0 | 1 | Ref |
| 898 | GU339467 | Unassigned | Unassigned | Unassigned | 216_3 | Clustered | 8 | VC_216_3 | 8 | 0.0701 | 1 | 0.0701 | 0 | 0 | 0 | 0.8571 | Ref |
| 899 | GU385905 | Unassigned | Unassigned | Unassigned | 303_0 | Clustered | 155 | VC_303_0 | 155 | 0.9979 | 1 | 0.9979 | 0 | 0 | 0 | 1 | Ref |
| 900 | GU393987 | Unassigned | Unassigned | Unassigned | 70_0 | Clustered | 27 | VC_70_0 | 27 | 0.5981 | 1 | 0.5981 | 0 | 0 | 0 | 0.9886 | Ref |
| 901 | GU459069 | Unassigned | Unassigned | Unassigned | 443_1 | Clustered | 2 | VC_443_1 | 2 | 0.0287 | 0.89347378 | 0.0257 | 0 | 0 | 0 | 1 | Ref |
| 902 | GU477322 | Unassigned | Unassigned | Unassigned | 9_0 | Clustered | 38 | VC_9_0 | 38 | 0.4895 | 1 | 0.4895 | 0 | 0 | 0 | 1 | Ref |
| 903 | GU557055 | Unassigned | Unassigned | Unassigned | 305_0 | Clustered | 12 | VC_305_0 | 12 | 0.5745 | 1 | 0.5745 | 0 | 0 | 0 | 0.8485 | Ref |
| 904 | GU573886 | Unassigned | Unassigned | Unassigned | 28_1 | Clustered | 53 | VC_28_1 | 53 | 0.7164 | 1 | 0.7164 | 0 | 0 | 0 | 0.9325 | Ref |
| 905 | GU580941 | Unassigned | Unassigned | Unassigned | 799_0 | Clustered | 2 | VC_799_0 | 2 | 0.0367 | 0.95547665 | 0.035 | 0 | 0 | 0 | 1 | Ref |
| 906 | GU580942 | Unassigned | Unassigned | Unassigned | 799_0 | Clustered | 2 | VC_799_0 | 2 | 0.0367 | 0.95547665 | 0.035 | 0 | 0 | 0 | 1 | Ref |
| 907 | GU583987 | Unassigned | Unassigned | Unassigned | 130_0 | Clustered | 19 | VC_130_0 | 19 | 0.6068 | 1 | 0.6068 | 0 | 0 | 0 | 1 | Ref |
| 908 | GU595417 | Unassigned | Unassigned | Unassigned | 452_0 | Clustered | 66 | VC_452_0 | 66 | 0.9248 | 1 | 0.9248 | 0 | 0 | 0 | 1 | Ref |
| 909 | GU815091 | Unassigned | Unassigned | Unassigned | 79_7 | Clustered | 33 | VC_79_7 | 33 | 0.216 | 1 | 0.216 | 0 | 0 | 0 | 0.8409 | Ref |
| 910 | GU903191 | Unassigned | Unassigned | Unassigned | 125_0 | Clustered | 17 | VC_125_0 | 17 | 0.9244 | 1 | 0.9244 | 0 | 0 | 0 | 1 | Ref |
| 911 | GU911303 | Unassigned | Unassigned | Unassigned | 1_0 | Clustered | 54 | VC_1_0 | 54 | 0.8527 | 1 | 0.8527 | 0 | 0 | 0 | 1 | Ref |
| 912 | GU911304 | Unassigned | Unassigned | Unassigned | 3_0 | Clustered | 17 | VC_3_0 | 17 | 0.5639 | 1 | 0.5639 | 0 | 0 | 0 | 1 | Ref |
| 913 | GU911519 | Unassigned | Unassigned | Unassigned | 492_3 | Clustered/Singleton | 1 | VC_492_3 | 1 | 0 | 1 | 0 | 0 | 0 | 0 | 1 | Ref |
| 914 | GU936715 | Unassigned | Unassigned | Unassigned | 873_0 | Clustered | 3 | VC_873_0 | 3 | 0.9107 | 0.98376334 | 0.8959 | 0 | 0 | 0 | 1 | Ref |
| 915 | GU942563 | Unassigned | Unassigned | Unassigned | 875_0 | Clustered | 10 | VC_875_0 | 10 | 0.988 | 0.99948433 | 0.9875 | 0 | 0 | 0 | 1 | Ref |
| 916 | GU943054 | Unassigned | Unassigned | Unassigned | 340_0 | Clustered | 31 | VC_340_0 | 31 | 0.9549 | 1 | 0.9549 | 0 | 0 | 0 | 1 | Ref |
| 917 | GU988610 | Unassigned | Unassigned | Unassigned | 76_0 | Clustered | 37 | VC_76_0 | 37 | 0.9403 | 1 | 0.9403 | 0 | 0 | 0 | 1 | Ref |
| 918 | HE584812 | Unassigned | Unassigned | Unassigned | 179_0 | Clustered | 53 | VC_179_0 | 53 | 0.9137 | 1 | 0.9137 | 0 | 0 | 0 | 1 | Ref |
| 919 | HE600015 | Unassigned | Unassigned | Unassigned | 131_0 | Clustered | 17 | VC_131_0 | 17 | 0.2671 | 1 | 0.2671 | 0 | 0 | 0 | 1 | Ref |
| 920 | HE608841 | Unassigned | Unassigned | Unassigned | 830_0 | Clustered | 6 | VC_830_0 | 6 | 0.6536 | 1 | 0.6536 | 0 | 0 | 0 | 1 | Ref |
| 921 | HE664024 | Unassigned | Unassigned | Unassigned | 42_0 | Clustered | 49 | VC_42_0 | 49 | 0.4517 | 1 | 0.4517 | 0 | 0 | 0 | 0.8597 | Ref |
| 922 | HE775250 | Unassigned | Unassigned | Unassigned | 637_0 | Clustered | 115 | VC_637_0 | 115 | 0.731 | 1 | 0.731 | 0 | 0 | 0 | 0.8764 | Ref |
| 923 | HE806280 | Unassigned | Unassigned | Unassigned | 698_0 | Clustered | 26 | VC_698_0 | 26 | 0.9366 | 1 | 0.9366 | 0 | 0 | 0 | 1 | Ref |
| 924 | HE815464 | Unassigned | Unassigned | Unassigned | 843_0 | Clustered | 5 | VC_843_0 | 5 | 0.2843 | 0.99999994 | 0.2843 | 0 | 0 | 0 | 1 | Ref |
| 925 | HE858210 | Unassigned | Unassigned | Unassigned | 669_1 | Clustered | 10 | VC_669_1 | 10 | 0.1268 | 1 | 0.1268 | 0 | 0 | 0 | 0.9778 | Ref |
| 926 | HE861935 | Unassigned | Unassigned | Unassigned | 254_0 | Clustered | 46 | VC_254_0 | 46 | 0.5159 | 1 | 0.5159 | 0 | 0 | 0 | 1 | Ref |
| 927 | HE956704 | Unassigned | Unassigned | Unassigned | 308_0 | Clustered | 4 | VC_308_0 | 4 | 0.5014 | 0.99997418 | 0.5013 | 0 | 0 | 0 | 1 | Ref |
| 928 | HE956707 | Unassigned | Unassigned | Unassigned | 878_0 | Clustered | 32 | VC_878_0 | 32 | 0.9965 | 1 | 0.9965 | 0 | 0 | 0 | 1 | Ref |
| 929 | HE956708 | Unassigned | Unassigned | Unassigned | 191_26 | Clustered/Singleton | 1 | VC_191_26 | 1 | 0 | 1 | 0 | 0 | 0 | 0 | 1 | Ref |
| 930 | HE956709 | Unassigned | Unassigned | Unassigned | 490_3 | Clustered | 2 | VC_490_3 | 2 | 0.0135 | 0.95703149 | 0.0129 | 0 | 0 | 0 | 1 | Ref |
| 931 | HE956710 | Unassigned | Unassigned | Unassigned | 878_0 | Clustered | 32 | VC_878_0 | 32 | 0.9965 | 1 | 0.9965 | 0 | 0 | 0 | 1 | Ref |
| 932 | HE956711 | Unassigned | Unassigned | Unassigned | 283_69 | Clustered/Singleton | 1 | VC_283_69 | 1 | 0 | 1 | 0 | 0 | 0 | 0 | 1 | Ref |
| 933 | HE962497 | Unassigned | Unassigned | Unassigned | 108_0 | Clustered | 33 | VC_108_0 | 33 | 0.949 | 1 | 0.949 | 0 | 0 | 0 | 1 | Ref |
| 934 | HE978309 | Unassigned | Unassigned | Unassigned | 590_0 | Clustered | 27 | VC_590_0 | 27 | 0.9947 | 1 | 0.9947 | 0 | 0 | 0 | 1 | Ref |
| 935 | HE981739 | Unassigned | Unassigned | Unassigned | 669_1 | Clustered | 10 | VC_669_1 | 10 | 0.1268 | 1 | 0.1268 | 0 | 0 | 0 | 0.9778 | Ref |
| 936 | HE983844 | Unassigned | Unassigned | Unassigned | 472_0 | Clustered | 47 | VC_472_0 | 47 | 0.9025 | 1 | 0.9025 | 0 | 0 | 0 | 1 | Ref |
| 937 | HE983845 | Unassigned | Unassigned | Unassigned | 76_0 | Clustered | 37 | VC_76_0 | 37 | 0.9403 | 1 | 0.9403 | 0 | 0 | 0 | 1 | Ref |
| 938 | HF543949 | Unassigned | Unassigned | Unassigned | 472_0 | Clustered | 47 | VC_472_0 | 47 | 0.9025 | 1 | 0.9025 | 0 | 0 | 0 | 1 | Ref |
| 939 | HF563658 | Unassigned | Unassigned | Unassigned | 412_0 | Clustered | 39 | VC_412_0 | 39 | 0.845 | 1 | 0.845 | 0 | 0 | 0 | 1 | Ref |
| 940 | HF569088 | Unassigned | Unassigned | Unassigned | 879_0 | Clustered | 36 | VC_879_0 | 36 | 0.994 | 1 | 0.994 | 0 | 0 | 0 | 1 | Ref |
| 941 | HF569089 | Unassigned | Unassigned | Unassigned | 879_0 | Clustered | 36 | VC_879_0 | 36 | 0.994 | 1 | 0.994 | 0 | 0 | 0 | 1 | Ref |
| 942 | HF569090 | Unassigned | Unassigned | Unassigned | 879_0 | Clustered | 36 | VC_879_0 | 36 | 0.994 | 1 | 0.994 | 0 | 0 | 0 | 1 | Ref |
| 943 | HF569091 | Unassigned | Unassigned | Unassigned | 879_0 | Clustered | 36 | VC_879_0 | 36 | 0.994 | 1 | 0.994 | 0 | 0 | 0 | 1 | Ref |
| 944 | HF569092 | Unassigned | Unassigned | Unassigned | 879_0 | Clustered | 36 | VC_879_0 | 36 | 0.994 | 1 | 0.994 | 0 | 0 | 0 | 1 | Ref |
| 945 | HF937074 | Unassigned | Unassigned | Unassigned | 96_0 | Clustered | 30 | VC_96_0 | 30 | 0.718 | 1 | 0.718 | 0 | 0 | 0 | 1 | Ref |
| 946 | HG007973 | Unassigned | Unassigned | Unassigned | 880_0 | Clustered | 2 | VC_880_0 | 2 | 0.1614 | 0.83455073 | 0.1347 | 0 | 0 | 0 | 1 | Ref |
| 947 | HG315669 | Unassigned | Unassigned | Unassigned | 177_0 | Clustered | 5 | VC_177_0 | 5 | 0.7468 | 0.99885646 | 0.746 | 0 | 0 | 0 | 1 | Ref |
| 948 | HG322870 | Unassigned | Unassigned | Unassigned | 446_0 | Clustered | 4 | VC_446_0 | 4 | 0.754 | 0.99987768 | 0.7539 | 0 | 0 | 0 | 1 | Ref |
| 949 | HG380752 | Unassigned | Unassigned | Unassigned | 254_0 | Clustered | 46 | VC_254_0 | 46 | 0.5159 | 1 | 0.5159 | 0 | 0 | 0 | 1 | Ref |
| 950 | HG424323 | Unassigned | Unassigned | Unassigned | 254_0 | Clustered | 46 | VC_254_0 | 46 | 0.5159 | 1 | 0.5159 | 0 | 0 | 0 | 1 | Ref |
| 951 | HG428758 | Unassigned | Unassigned | Unassigned | 879_0 | Clustered | 36 | VC_879_0 | 36 | 0.994 | 1 | 0.994 | 0 | 0 | 0 | 1 | Ref |
| 952 | HG518155 | Unassigned | Unassigned | Unassigned | 472_0 | Clustered | 47 | VC_472_0 | 47 | 0.9025 | 1 | 0.9025 | 0 | 0 | 0 | 1 | Ref |
| 953 | HG531805 | Unassigned | Unassigned | Unassigned | 139_1 | Clustered | 4 | VC_139_1 | 4 | 0.1775 | 0.99998609 | 0.1775 | 0 | 0 | 0 | 1 | Ref |
| 954 | HG792102 | Unassigned | Unassigned | Unassigned | 42_0 | Clustered | 49 | VC_42_0 | 49 | 0.4517 | 1 | 0.4517 | 0 | 0 | 0 | 0.8597 | Ref |
| 955 | HG792103 | Unassigned | Unassigned | Unassigned | 42_0 | Clustered | 49 | VC_42_0 | 49 | 0.4517 | 1 | 0.4517 | 0 | 0 | 0 | 0.8597 | Ref |
| 956 | HG792104 | Unassigned | Unassigned | Unassigned | 42_0 | Clustered | 49 | VC_42_0 | 49 | 0.4517 | 1 | 0.4517 | 0 | 0 | 0 | 0.8597 | Ref |
| 957 | HG792105 | Unassigned | Unassigned | Unassigned | 42_0 | Clustered | 49 | VC_42_0 | 49 | 0.4517 | 1 | 0.4517 | 0 | 0 | 0 | 0.8597 | Ref |
| 958 | HG793132 | Unassigned | Unassigned | Unassigned | 70_0 | Clustered | 27 | VC_70_0 | 27 | 0.5981 | 1 | 0.5981 | 0 | 0 | 0 | 0.9886 | Ref |
| 959 | HG796219 | Unassigned | Unassigned | Unassigned | 70_0 | Clustered | 27 | VC_70_0 | 27 | 0.5981 | 1 | 0.5981 | 0 | 0 | 0 | 0.9886 | Ref |
| 960 | HG796220 | Unassigned | Unassigned | Unassigned | 70_0 | Clustered | 27 | VC_70_0 | 27 | 0.5981 | 1 | 0.5981 | 0 | 0 | 0 | 0.9886 | Ref |
| 961 | HG796221 | Unassigned | Unassigned | Unassigned | 70_0 | Clustered | 27 | VC_70_0 | 27 | 0.5981 | 1 | 0.5981 | 0 | 0 | 0 | 0.9886 | Ref |
| 962 | HG796225 | Unassigned | Unassigned | Unassigned | 647_0 | Clustered | 11 | VC_647_0 | 11 | 0.598 | 1 | 0.598 | 0 | 0 | 0 | 1 | Ref |
| 963 | HG798901 | Unassigned | Unassigned | Unassigned | 647_0 | Clustered | 11 | VC_647_0 | 11 | 0.598 | 1 | 0.598 | 0 | 0 | 0 | 1 | Ref |
| 964 | HG799490 | Unassigned | Unassigned | Unassigned | 414_0 | Clustered | 37 | VC_414_0 | 37 | 0.585 | 1 | 0.585 | 0 | 0 | 0 | 0.9474 | Ref |
| 965 | HG799496 | Unassigned | Unassigned | Unassigned | 414_0 | Clustered | 37 | VC_414_0 | 37 | 0.585 | 1 | 0.585 | 0 | 0 | 0 | 0.9474 | Ref |
| 966 | HG799497 | Unassigned | Unassigned | Unassigned | 414_0 | Clustered | 37 | VC_414_0 | 37 | 0.585 | 1 | 0.585 | 0 | 0 | 0 | 0.9474 | Ref |
| 967 | HG803181 | Unassigned | Unassigned | Unassigned | 5_0 | Clustered | 30 | VC_5_0 | 30 | 0.9647 | 1 | 0.9647 | 0 | 0 | 0 | 1 | Ref |
| 968 | HG803182 | Unassigned | Unassigned | Unassigned | 42_0 | Clustered | 49 | VC_42_0 | 49 | 0.4517 | 1 | 0.4517 | 0 | 0 | 0 | 0.8597 | Ref |
| 969 | HG803183 | Unassigned | Unassigned | Unassigned | 42_0 | Clustered | 49 | VC_42_0 | 49 | 0.4517 | 1 | 0.4517 | 0 | 0 | 0 | 0.8597 | Ref |
| 970 | HG803184 | Unassigned | Unassigned | Unassigned | 42_0 | Clustered | 49 | VC_42_0 | 49 | 0.4517 | 1 | 0.4517 | 0 | 0 | 0 | 0.8597 | Ref |

| | | | | | | | | | | | | | | | | | |
|---|---|---|---|---|---|---|---|---|---|---|---|---|---|---|---|---|---|
| 971 | HG803185 | Unassigned | Unassigned | Unassigned | 42_0 | Clustered | 49 | VC_42_0 | 49 | 0.4517 | 1 | 0.4517 | 0 | 0 | 0 | 0.8597 | Ref |
| 972 | HG813241 | Unassigned | Unassigned | Unassigned | 451_0 | Clustered | 80 | VC_451_0 | 80 | 0.9991 | 1 | 0.9991 | 0 | 0 | 0 | 1 | Ref |
| 973 | HG818823 | Unassigned | Unassigned | Unassigned | 451_0 | Clustered | 80 | VC_451_0 | 80 | 0.9991 | 1 | 0.9991 | 0 | 0 | 0 | 1 | Ref |
| 974 | HG818824 | Unassigned | Unassigned | Unassigned | 451_0 | Clustered | 80 | VC_451_0 | 80 | 0.9991 | 1 | 0.9991 | 0 | 0 | 0 | 1 | Ref |
| 975 | HG934469 | Unassigned | Unassigned | Unassigned | 637_0 | Clustered | 115 | VC_637_0 | 115 | 0.731 | 1 | 0.731 | 0 | 0 | 0 | 0.8764 | Ref |
| 976 | HG934470 | Unassigned | Unassigned | Unassigned | 637_0 | Clustered | 115 | VC_637_0 | 115 | 0.731 | 1 | 0.731 | 0 | 0 | 0 | 0.8764 | Ref |
| 977 | HG962375 | Unassigned | Unassigned | Unassigned | 838_0 | Clustered | 29 | VC_838_0 | 29 | 0.8611 | 1 | 0.8611 | 0 | 0 | 0 | 1 | Ref |
| 978 | HG962376 | Unassigned | Unassigned | Unassigned | 749_0 | Clustered | 40 | VC_749_0 | 40 | 0.7517 | 1 | 0.7517 | 0 | 0 | 0 | 1 | Ref |
| 979 | HG992588 | Unassigned | Unassigned | Unassigned | 882_0 | Clustered | 20 | VC_882_0 | 20 | 0.9917 | 1 | 0.9917 | 0 | 0 | 0 | 1 | Ref |
| 980 | HG994092 | Unassigned | Unassigned | Unassigned | 823_0 | Clustered | 124 | VC_823_0 | 124 | 0.9492 | 1 | 0.9492 | 0 | 0 | 0 | 1 | Ref |
| 981 | HM004124 | Unassigned | Unassigned | Unassigned | 492_4 | Clustered/Singleton | 1 | VC_492_4 | 1 | 0 | 1 | 0 | 0 | 0 | 0 | 1 | Ref |
| 982 | HM029250 | Unassigned | Unassigned | Unassigned | 474_0 | Clustered | 14 | VC_474_0 | 14 | 0.8466 | 1 | 0.8466 | 0 | 0 | 0 | 1 | Ref |
| 983 | HM032710 | Unassigned | Unassigned | Unassigned | 492_5 | Clustered/Singleton | 1 | VC_492_5 | 1 | 0 | 1 | 0 | 0 | 0 | 0 | 1 | Ref |
| 984 | HM035024 | Unassigned | Unassigned | Unassigned | 575_0 | Clustered | 35 | VC_575_0 | 35 | 0.8815 | 1 | 0.8815 | 0 | 0 | 0 | 1 | Ref |
| 985 | HM035025 | Unassigned | Unassigned | Unassigned | 283_154 | Clustered/Singleton | 1 | VC_283_154 | 1 | 0 | 1 | 0 | 0 | 0 | 0 | 1 | Ref |
| 986 | HM064452 | Unassigned | Unassigned | Unassigned | 53_0 | Clustered | 5 | VC_53_0 | 5 | 0.4977 | 0.99999808 | 0.4977 | 0 | 0 | 0 | 1 | Ref |
| 987 | HM066936 | Unassigned | Unassigned | Unassigned | 227_0 | Clustered | 2 | VC_227_0 | 2 | 0.3333 | 0 | 0 | 0 | 0 | 0 | 1 | Ref |
| 988 | HM071924 | Unassigned | Unassigned | Unassigned | 290_5 | Clustered | 14 | VC_290_5 | 14 | 0.1422 | 1 | 0.1422 | 0 | 0 | 0 | 0.9231 | Ref |
| 989 | HM072038 | Unassigned | Unassigned | Unassigned | 18_0 | Clustered | 4 | VC_18_0 | 4 | 0.9098 | 0.99982347 | 0.9096 | 0 | 0 | 0 | 1 | Ref |
| 990 | HM114315 | Unassigned | Unassigned | Unassigned | 492_6 | Clustered/Singleton | 1 | VC_492_6 | 1 | 0 | 1 | 0 | 0 | 0 | 0 | 1 | Ref |
| 991 | HM134276 | Unassigned | Unassigned | Unassigned | 669_1 | Clustered | 10 | VC_669_1 | 10 | 0.1268 | 1 | 0.1268 | 0 | 0 | 0 | 0.9778 | Ref |
| 992 | HM137666 | Unassigned | Unassigned | Unassigned | 283_91 | Clustered | 11 | VC_283_91 | 11 | 0.0285 | 1 | 0.0285 | 0 | 0 | 0 | 1 | Ref |
| 993 | HM144387 | Unassigned | Unassigned | Unassigned | 884_1 | Clustered | 8 | VC_884_1 | 8 | 0.1102 | 0.9989323 | 0.1101 | 0 | 0 | 0 | 0.75 | Ref |
| 994 | HM150760 | Unassigned | Unassigned | Unassigned | 880_0 | Clustered | 2 | VC_880_0 | 2 | 0.1614 | 0.83455073 | 0.1347 | 0 | 0 | 0 | 1 | Ref |
| 995 | HM152763 | Unassigned | Unassigned | Unassigned | 887_0 | Clustered | 18 | VC_887_0 | 18 | 0.3663 | 1 | 0.3663 | 0 | 0 | 0 | 1 | Ref |
| 996 | HM152764 | Unassigned | Unassigned | Unassigned | 236_0 | Clustered | 72 | VC_236_0 | 72 | 0.8104 | 1 | 0.8104 | 0 | 0 | 0 | 1 | Ref |
| 997 | HM152765 | Unassigned | Unassigned | Unassigned | 518_0 | Clustered | 6 | VC_518_0 | 6 | 0.1295 | 1 | 0.1295 | 0 | 0 | 0 | 1 | Ref |
| 998 | HM152766 | Unassigned | Unassigned | Unassigned | 517_1 | Clustered | 108 | VC_517_1 | 108 | 0.8483 | 1 | 0.8483 | 0 | 0 | 0 | 0.9955 | Ref |
| 999 | HM152767 | Unassigned | Unassigned | Unassigned | 236_0 | Clustered | 72 | VC_236_0 | 72 | 0.8104 | 1 | 0.8104 | 0 | 0 | 0 | 1 | Ref |
| 1000 | HM208303 | Unassigned | Unassigned | Unassigned | 39_0 | Clustered | 35 | VC_39_0 | 35 | 0.4337 | 1 | 0.4337 | 0 | 0 | 0 | 1 | Ref |
| 1001 | HM208537 | Unassigned | Unassigned | Unassigned | 230_1 | Clustered/Singleton | 1 | VC_230_1 | 1 | 0 | 1 | 0 | 0 | 0 | 0 | 1 | Ref |
| 1002 | HM242243 | Unassigned | Unassigned | Unassigned | 94_0 | Clustered | 2 | VC_94_0 | 2 | 0.1304 | 0.95723134 | 0.1248 | 0 | 0 | 0 | 1 | Ref |
| 1003 | HM368260 | Unassigned | Unassigned | Unassigned | 698_0 | Clustered | 26 | VC_698_0 | 26 | 0.9366 | 1 | 0.9366 | 0 | 0 | 0 | 1 | Ref |
| 1004 | HM452125 | Unassigned | Unassigned | Unassigned | 600_2 | Clustered | 10 | VC_600_2 | 10 | 0.3041 | 1 | 0.3041 | 0 | 0 | 0 | 1 | Ref |
| 1005 | HM461982 | Unassigned | Unassigned | Unassigned | 3_0 | Clustered | 17 | VC_3_0 | 17 | 0.5639 | 1 | 0.5639 | 0 | 0 | 0 | 1 | Ref |
| 1006 | HM480106 | Unassigned | Unassigned | Unassigned | 873_0 | Clustered | 3 | VC_873_0 | 3 | 0.9107 | 0.98376334 | 0.8959 | 0 | 0 | 0 | 1 | Ref |
| 1007 | HM480846 | Unassigned | Unassigned | Unassigned | 823_0 | Clustered | 124 | VC_823_0 | 124 | 0.9492 | 1 | 0.9492 | 0 | 0 | 0 | 1 | Ref |
| 1008 | HM559717 | Unassigned | Unassigned | Unassigned | 340_0 | Clustered | 31 | VC_340_0 | 31 | 0.9549 | 1 | 0.9549 | 0 | 0 | 0 | 1 | Ref |
| 1009 | HM563683 | Unassigned | Unassigned | Unassigned | 290_2 | Clustered | 2 | VC_290_2 | 2 | 0.0102 | 0.92371007 | 0.0094 | 0 | 0 | 0 | 1 | Ref |
| 1010 | HM568888 | Unassigned | Unassigned | Unassigned | 733_0 | Clustered | 5 | VC_733_0 | 5 | 0.5111 | 0.99999994 | 0.5111 | 0 | 0 | 0 | 1 | Ref |
| 1011 | HM588722 | Unassigned | Unassigned | Unassigned | 894_0 | Clustered | 3 | VC_894_0 | 3 | 0.6358 | 0.99692152 | 0.6338 | 0 | 0 | 0 | 1 | Ref |
| 1012 | HM624080 | Unassigned | Unassigned | Unassigned | 179_0 | Clustered | 53 | VC_179_0 | 53 | 0.9137 | 1 | 0.9137 | 0 | 0 | 0 | 1 | Ref |
| 1013 | HM640230 | Unassigned | Unassigned | Unassigned | 897_0 | Clustered | 6 | VC_897_0 | 6 | 0.4156 | 1 | 0.4156 | 0 | 0 | 0 | 1 | Ref |
| 1014 | HM753658 | Unassigned | Unassigned | Unassigned | 303_0 | Clustered | 155 | VC_303_0 | 155 | 0.9979 | 1 | 0.9979 | 0 | 0 | 0 | 1 | Ref |
| 1015 | HM753659 | Unassigned | Unassigned | Unassigned | 303_0 | Clustered | 155 | VC_303_0 | 155 | 0.9979 | 1 | 0.9979 | 0 | 0 | 0 | 1 | Ref |
| 1016 | HM753660 | Unassigned | Unassigned | Unassigned | 303_0 | Clustered | 155 | VC_303_0 | 155 | 0.9979 | 1 | 0.9979 | 0 | 0 | 0 | 1 | Ref |
| 1017 | HM753661 | Unassigned | Unassigned | Unassigned | 303_0 | Clustered | 155 | VC_303_0 | 155 | 0.9979 | 1 | 0.9979 | 0 | 0 | 0 | 1 | Ref |
| 1018 | HM753662 | Unassigned | Unassigned | Unassigned | 303_0 | Clustered | 155 | VC_303_0 | 155 | 0.9979 | 1 | 0.9979 | 0 | 0 | 0 | 1 | Ref |
| 1019 | HM753663 | Unassigned | Unassigned | Unassigned | 303_0 | Clustered | 155 | VC_303_0 | 155 | 0.9979 | 1 | 0.9979 | 0 | 0 | 0 | 1 | Ref |
| 1020 | HM753664 | Unassigned | Unassigned | Unassigned | 303_0 | Clustered | 155 | VC_303_0 | 155 | 0.9979 | 1 | 0.9979 | 0 | 0 | 0 | 1 | Ref |
| 1021 | HM753665 | Unassigned | Unassigned | Unassigned | 303_0 | Clustered | 155 | VC_303_0 | 155 | 0.9979 | 1 | 0.9979 | 0 | 0 | 0 | 1 | Ref |
| 1022 | HM753666 | Unassigned | Unassigned | Unassigned | 303_0 | Clustered | 155 | VC_303_0 | 155 | 0.9979 | 1 | 0.9979 | 0 | 0 | 0 | 1 | Ref |
| 1023 | HM753667 | Unassigned | Unassigned | Unassigned | 303_0 | Clustered | 155 | VC_303_0 | 155 | 0.9979 | 1 | 0.9979 | 0 | 0 | 0 | 1 | Ref |
| 1024 | HM753668 | Unassigned | Unassigned | Unassigned | 303_0 | Clustered | 155 | VC_303_0 | 155 | 0.9979 | 1 | 0.9979 | 0 | 0 | 0 | 1 | Ref |
| 1025 | HM753669 | Unassigned | Unassigned | Unassigned | 303_0 | Clustered | 155 | VC_303_0 | 155 | 0.9979 | 1 | 0.9979 | 0 | 0 | 0 | 1 | Ref |
| 1026 | HM753670 | Unassigned | Unassigned | Unassigned | 303_0 | Clustered | 155 | VC_303_0 | 155 | 0.9979 | 1 | 0.9979 | 0 | 0 | 0 | 1 | Ref |
| 1027 | HM753671 | Unassigned | Unassigned | Unassigned | 303_0 | Clustered | 155 | VC_303_0 | 155 | 0.9979 | 1 | 0.9979 | 0 | 0 | 0 | 1 | Ref |
| 1028 | HM753672 | Unassigned | Unassigned | Unassigned | 303_0 | Clustered | 155 | VC_303_0 | 155 | 0.9979 | 1 | 0.9979 | 0 | 0 | 0 | 1 | Ref |
| 1029 | HM753673 | Unassigned | Unassigned | Unassigned | 303_0 | Clustered | 155 | VC_303_0 | 155 | 0.9979 | 1 | 0.9979 | 0 | 0 | 0 | 1 | Ref |
| 1030 | HM753674 | Unassigned | Unassigned | Unassigned | 303_0 | Clustered | 155 | VC_303_0 | 155 | 0.9979 | 1 | 0.9979 | 0 | 0 | 0 | 1 | Ref |
| 1031 | HM753675 | Unassigned | Unassigned | Unassigned | 303_0 | Clustered | 155 | VC_303_0 | 155 | 0.9979 | 1 | 0.9979 | 0 | 0 | 0 | 1 | Ref |
| 1032 | HM753676 | Unassigned | Unassigned | Unassigned | 303_0 | Clustered | 155 | VC_303_0 | 155 | 0.9979 | 1 | 0.9979 | 0 | 0 | 0 | 1 | Ref |
| 1033 | HM753677 | Unassigned | Unassigned | Unassigned | 303_0 | Clustered | 155 | VC_303_0 | 155 | 0.9979 | 1 | 0.9979 | 0 | 0 | 0 | 1 | Ref |
| 1034 | HM753678 | Unassigned | Unassigned | Unassigned | 303_0 | Clustered | 155 | VC_303_0 | 155 | 0.9979 | 1 | 0.9979 | 0 | 0 | 0 | 1 | Ref |
| 1035 | HM753679 | Unassigned | Unassigned | Unassigned | 303_0 | Clustered | 155 | VC_303_0 | 155 | 0.9979 | 1 | 0.9979 | 0 | 0 | 0 | 1 | Ref |
| 1036 | HM753680 | Unassigned | Unassigned | Unassigned | 303_0 | Clustered | 155 | VC_303_0 | 155 | 0.9979 | 1 | 0.9979 | 0 | 0 | 0 | 1 | Ref |
| 1037 | HM753681 | Unassigned | Unassigned | Unassigned | 303_0 | Clustered | 155 | VC_303_0 | 155 | 0.9979 | 1 | 0.9979 | 0 | 0 | 0 | 1 | Ref |
| 1038 | HM753682 | Unassigned | Unassigned | Unassigned | 303_0 | Clustered | 155 | VC_303_0 | 155 | 0.9979 | 1 | 0.9979 | 0 | 0 | 0 | 1 | Ref |
| 1039 | HM753683 | Unassigned | Unassigned | Unassigned | 303_0 | Clustered | 155 | VC_303_0 | 155 | 0.9979 | 1 | 0.9979 | 0 | 0 | 0 | 1 | Ref |
| 1040 | HM753684 | Unassigned | Unassigned | Unassigned | 303_0 | Clustered | 155 | VC_303_0 | 155 | 0.9979 | 1 | 0.9979 | 0 | 0 | 0 | 1 | Ref |
| 1041 | HM753685 | Unassigned | Unassigned | Unassigned | 303_0 | Clustered | 155 | VC_303_0 | 155 | 0.9979 | 1 | 0.9979 | 0 | 0 | 0 | 1 | Ref |
| 1042 | HM753686 | Unassigned | Unassigned | Unassigned | 303_0 | Clustered | 155 | VC_303_0 | 155 | 0.9979 | 1 | 0.9979 | 0 | 0 | 0 | 1 | Ref |
| 1043 | HM753687 | Unassigned | Unassigned | Unassigned | 303_0 | Clustered | 155 | VC_303_0 | 155 | 0.9979 | 1 | 0.9979 | 0 | 0 | 0 | 1 | Ref |
| 1044 | HM753688 | Unassigned | Unassigned | Unassigned | 303_0 | Clustered | 155 | VC_303_0 | 155 | 0.9979 | 1 | 0.9979 | 0 | 0 | 0 | 1 | Ref |
| 1045 | HM753689 | Unassigned | Unassigned | Unassigned | 303_0 | Clustered | 155 | VC_303_0 | 155 | 0.9979 | 1 | 0.9979 | 0 | 0 | 0 | 1 | Ref |
| 1046 | HM753690 | Unassigned | Unassigned | Unassigned | 303_0 | Clustered | 155 | VC_303_0 | 155 | 0.9979 | 1 | 0.9979 | 0 | 0 | 0 | 1 | Ref |
| 1047 | HM753691 | Unassigned | Unassigned | Unassigned | 303_0 | Clustered | 155 | VC_303_0 | 155 | 0.9979 | 1 | 0.9979 | 0 | 0 | 0 | 1 | Ref |
| 1048 | HM753692 | Unassigned | Unassigned | Unassigned | 303_0 | Clustered | 155 | VC_303_0 | 155 | 0.9979 | 1 | 0.9979 | 0 | 0 | 0 | 1 | Ref |
| 1049 | HM753693 | Unassigned | Unassigned | Unassigned | 303_0 | Clustered | 155 | VC_303_0 | 155 | 0.9979 | 1 | 0.9979 | 0 | 0 | 0 | 1 | Ref |
| 1050 | HM753694 | Unassigned | Unassigned | Unassigned | 303_0 | Clustered | 155 | VC_303_0 | 155 | 0.9979 | 1 | 0.9979 | 0 | 0 | 0 | 1 | Ref |
| 1051 | HM753695 | Unassigned | Unassigned | Unassigned | 303_0 | Clustered | 155 | VC_303_0 | 155 | 0.9979 | 1 | 0.9979 | 0 | 0 | 0 | 1 | Ref |
| 1052 | HM753696 | Unassigned | Unassigned | Unassigned | 303_0 | Clustered | 155 | VC_303_0 | 155 | 0.9979 | 1 | 0.9979 | 0 | 0 | 0 | 1 | Ref |
| 1053 | HM753697 | Unassigned | Unassigned | Unassigned | 303_0 | Clustered | 155 | VC_303_0 | 155 | 0.9979 | 1 | 0.9979 | 0 | 0 | 0 | 1 | Ref |
| 1054 | HM753698 | Unassigned | Unassigned | Unassigned | 303_0 | Clustered | 155 | VC_303_0 | 155 | 0.9979 | 1 | 0.9979 | 0 | 0 | 0 | 1 | Ref |
| 1055 | HM753699 | Unassigned | Unassigned | Unassigned | 303_0 | Clustered | 155 | VC_303_0 | 155 | 0.9979 | 1 | 0.9979 | 0 | 0 | 0 | 1 | Ref |
| 1056 | HM753700 | Unassigned | Unassigned | Unassigned | 303_0 | Clustered | 155 | VC_303_0 | 155 | 0.9979 | 1 | 0.9979 | 0 | 0 | 0 | 1 | Ref |
| 1057 | HM753701 | Unassigned | Unassigned | Unassigned | 303_0 | Clustered | 155 | VC_303_0 | 155 | 0.9979 | 1 | 0.9979 | 0 | 0 | 0 | 1 | Ref |
| 1058 | HM753702 | Unassigned | Unassigned | Unassigned | 303_0 | Clustered | 155 | VC_303_0 | 155 | 0.9979 | 1 | 0.9979 | 0 | 0 | 0 | 1 | Ref |
| 1059 | HM753703 | Unassigned | Unassigned | Unassigned | 303_0 | Clustered | 155 | VC_303_0 | 155 | 0.9979 | 1 | 0.9979 | 0 | 0 | 0 | 1 | Ref |
| 1060 | HM753704 | Unassigned | Unassigned | Unassigned | 303_0 | Clustered | 155 | VC_303_0 | 155 | 0.9979 | 1 | 0.9979 | 0 | 0 | 0 | 1 | Ref |
| 1061 | HM753705 | Unassigned | Unassigned | Unassigned | 303_0 | Clustered | 155 | VC_303_0 | 155 | 0.9979 | 1 | 0.9979 | 0 | 0 | 0 | 1 | Ref |
| 1062 | HM753706 | Unassigned | Unassigned | Unassigned | 303_0 | Clustered | 155 | VC_303_0 | 155 | 0.9979 | 1 | 0.9979 | 0 | 0 | 0 | 1 | Ref |
| 1063 | HM753707 | Unassigned | Unassigned | Unassigned | 303_0 | Clustered | 155 | VC_303_0 | 155 | 0.9979 | 1 | 0.9979 | 0 | 0 | 0 | 1 | Ref |
| 1064 | HM753708 | Unassigned | Unassigned | Unassigned | 303_0 | Clustered | 155 | VC_303_0 | 155 | 0.9979 | 1 | 0.9979 | 0 | 0 | 0 | 1 | Ref |
| 1065 | HM753709 | Unassigned | Unassigned | Unassigned | 303_0 | Clustered | 155 | VC_303_0 | 155 | 0.9979 | 1 | 0.9979 | 0 | 0 | 0 | 1 | Ref |
| 1066 | HM753710 | Unassigned | Unassigned | Unassigned | 303_0 | Clustered | 155 | VC_303_0 | 155 | 0.9979 | 1 | 0.9979 | 0 | 0 | 0 | 1 | Ref |
| 1067 | HM753711 | Unassigned | Unassigned | Unassigned | 303_0 | Clustered | 155 | VC_303_0 | 155 | 0.9979 | 1 | 0.9979 | 0 | 0 | 0 | 1 | Ref |
| 1068 | HM753712 | Unassigned | Unassigned | Unassigned | 303_0 | Clustered | 155 | VC_303_0 | 155 | 0.9979 | 1 | 0.9979 | 0 | 0 | 0 | 1 | Ref |
| 1069 | HM753713 | Unassigned | Unassigned | Unassigned | 303_0 | Clustered | 155 | VC_303_0 | 155 | 0.9979 | 1 | 0.9979 | 0 | 0 | 0 | 1 | Ref |
| 1070 | HM753714 | Unassigned | Unassigned | Unassigned | 303_0 | Clustered | 155 | VC_303_0 | 155 | 0.9979 | 1 | 0.9979 | 0 | 0 | 0 | 1 | Ref |
| 1071 | HM753715 | Unassigned | Unassigned | Unassigned | 303_0 | Clustered | 155 | VC_303_0 | 155 | 0.9979 | 1 | 0.9979 | 0 | 0 | 0 | 1 | Ref |
| 1072 | HM753716 | Unassigned | Unassigned | Unassigned | 303_0 | Clustered | 155 | VC_303_0 | 155 | 0.9979 | 1 | 0.9979 | 0 | 0 | 0 | 1 | Ref |
| 1073 | HM753717 | Unassigned | Unassigned | Unassigned | 303_0 | Clustered | 155 | VC_303_0 | 155 | 0.9979 | 1 | 0.9979 | 0 | 0 | 0 | 1 | Ref |
| 1074 | HM753718 | Unassigned | Unassigned | Unassigned | 303_0 | Clustered | 155 | VC_303_0 | 155 | 0.9979 | 1 | 0.9979 | 0 | 0 | 0 | 1 | Ref |
| 1075 | HM755814 | Unassigned | Unassigned | Unassigned | 514_0 | Clustered | 75 | VC_514_0 | 75 | 0.9402 | 1 | 0.9402 | 0 | 0 | 0 | 1 | Ref |
| 1076 | HM770079 | Unassigned | Unassigned | Unassigned | 6_0 | Clustered | 44 | VC_6_0 | 44 | 0.908 | 1 | 0.908 | 0 | 0 | 0 | 1 | Ref |
| 1077 | HM775306 | Unassigned | Unassigned | Unassigned | 303_0 | Clustered | 155 | VC_303_0 | 155 | 0.9979 | 1 | 0.9979 | 0 | 0 | 0 | 1 | Ref |
| 1078 | HM775307 | Unassigned | Unassigned | Unassigned | 303_0 | Clustered | 155 | VC_303_0 | 155 | 0.9979 | 1 | 0.9979 | 0 | 0 | 0 | 1 | Ref |
| 1079 | HM775308 | Unassigned | Unassigned | Unassigned | 303_0 | Clustered | 155 | VC_303_0 | 155 | 0.9979 | 1 | 0.9979 | 0 | 0 | 0 | 1 | Ref |
| 1080 | HM775309 | Unassigned | Unassigned | Unassigned | 303_0 | Clustered | 155 | VC_303_0 | 155 | 0.9979 | 1 | 0.9979 | 0 | 0 | 0 | 1 | Ref |
| 1081 | HM775310 | Unassigned | Unassigned | Unassigned | 303_0 | Clustered | 155 | VC_303_0 | 155 | 0.9979 | 1 | 0.9979 | 0 | 0 | 0 | 1 | Ref |
| 1082 | HM775311 | Unassigned | Unassigned | Unassigned | 303_0 | Clustered | 155 | VC_303_0 | 155 | 0.9979 | 1 | 0.9979 | 0 | 0 | 0 | 1 | Ref |
| 1083 | HM997019 | Unassigned | Unassigned | Unassigned | 812_0 | Clustered | 3 | VC_812_0 | 3 | 0.1174 | 0.99842379 | 0.1172 | 0 | 0 | 0 | 1 | Ref |
| 1084 | HM997020 | Unassigned | Unassigned | Unassigned | 283_2 | Clustered | 2 | VC_283_2 | 2 | 0.0027 | 0.93842619 | 0.0025 | 0 | 0 | 0 | 1 | Ref |
| 1085 | HQ110083 | Unassigned | Unassigned | Unassigned | 5_0 | Clustered | 30 | VC_5_0 | 30 | 0.9647 | 1 | 0.9647 | 0 | 0 | 0 | 1 | Ref |
| 1086 | HQ110084 | Unassigned | Unassigned | Unassigned | 228_0 | Clustered | 60 | VC_228_0 | 60 | 0.942 | 1 | 0.942 | 0 | 0 | 0 | 1 | Ref |
| 1087 | HQ127381 | Unassigned | Unassigned | Unassigned | 9_0 | Clustered | 38 | VC_9_0 | 38 | 0.4895 | 1 | 0.4895 | 0 | 0 | 0 | 1 | Ref |
| 1088 | HQ141410 | Unassigned | Unassigned | Unassigned | 381_0 | Clustered | 2 | VC_381_0 | 2 | 0.0829 | 0.95381392 | 0.079 | 0 | 0 | 0 | 1 | Ref |
| 1089 | HQ141411 | Unassigned | Unassigned | Unassigned | 300_0 | Clustered | 3 | VC_300_0 | 3 | 0.1218 | 0.99841701 | 0.1216 | 0 | 0 | 0 | 0.6667 | Ref |
| 1090 | HQ163896 | Unassigned | Unassigned | Unassigned | 149_8 | Clustered | 66 | VC_149_8 | 66 | 0.4565 | 1 | 0.4565 | 0 | 0 | 0 | 0.8182 | Ref |
| 1091 | HQ186308 | Unassigned | Unassigned | Unassigned | 699_0 | Clustered | 71 | VC_699_0 | 71 | 0.9352 | 1 | 0.9352 | 0 | 0 | 0 | 1 | Ref |
| 1092 | HQ201307 | Unassigned | Unassigned | Unassigned | 33_0 | Clustered | 4 | VC_33_0 | 4 | 0.06 | 0.99994826 | 0.06 | 0 | 0 | 0 | 1 | Ref |
| 1093 | HQ201308 | Unassigned | Unassigned | Unassigned | 31_2 | Clustered/Singleton | 1 | VC_31_2 | 1 | 0 | 1 | 0 | 0 | 0 | 0 | 1 | Ref |
| 1094 | HQ224500 | Unassigned | Unassigned | Unassigned | 875_0 | Clustered | 10 | VC_875_0 | 10 | 0.988 | 0.99948433 | 0.9875 | 0 | 0 | 0 | 1 | Ref |
| 1095 | HQ259103 | Unassigned | Unassigned | Unassigned | 133_31 | Clustered | 6 | VC_133_31 | 6 | 0.0263 | 0.99999694 | 0.0263 | 0 | 0 | 0 | 1 | Ref |
| 1096 | HQ259105 | Unassigned | Unassigned | Unassigned | 779_0 | Clustered | 37 | VC_779_0 | 37 | 0.5682 | 1 | 0.5682 | 0 | 0 | 0 | 1 | Ref |
| 1097 | HQ268735 | Unassigned | Unassigned | Unassigned | 891_0 | Clustered | 2 | VC_891_0 | 2 | 0.5746 | 0.92263554 | 0.5301 | 0 | 0 | 0 | 1 | Ref |
| 1098 | HQ316579 | Unassigned | Unassigned | Unassigned | 493_14 | Clustered/Singleton | 1 | VC_493_14 | 1 | 0 | 1 | 0 | 0 | 0 | 0 | 1 | Ref |
| 1099 | HQ316581 | Unassigned | Unassigned | Unassigned | 248_0 | Clustered | 11 | VC_248_0 | 11 | 0.7031 | 1 | 0.7031 | 0 | 0 | 0 | 1 | Ref |
| 1100 | HQ316582 | Unassigned | Unassigned | Unassigned | 902_0 | Clustered | 3 | VC_902_0 | 3 | 0.9978 | 0.87589346 | 0.8739 | 0 | 0 | 0 | 1 | Ref |
| 1101 | HQ316583 | Unassigned | Unassigned | Unassigned | 656_0 | Clustered | 2 | VC_656_0 | 2 | 0.0089 | 0.92256518 | 0.0082 | 0 | 0 | 0 | 1 | Ref |
| 1102 | HQ316584 | Unassigned | Unassigned | Unassigned | 340_0 | Clustered | 31 | VC_340_0 | 31 | 0.9549 | 1 | 0.9549 | 0 | 0 | 0 | 1 | Ref |
| 1103 | HQ316603 | Unassigned | Unassigned | Unassigned | 656_0 | Clustered | 2 | VC_656_0 | 2 | 0.0089 | 0.92256518 | 0.0082 | 0 | 0 | 0 | 1 | Ref |
| 1104 | HQ316604 | Unassigned | Unassigned | Unassigned | 904_1 | Clustered | 4 | VC_904_1 | 4 | 0.1486 | 0.99997723 | 0.1486 | 0 | 0 | 0 | 1 | Ref |
| 1105 | HQ317290 | Unassigned | Unassigned | Unassigned | 662_0 | Clustered | 62 | VC_662_0 | 62 | 0.6976 | 1 | 0.6976 | 0 | 0 | 0 | 1 | Ref |
| 1106 | HQ317291 | Unassigned | Unassigned | Unassigned | 662_0 | Clustered | 62 | VC_662_0 | 62 | 0.6976 | 1 | 0.6976 | 0 | 0 | 0 | 1 | Ref |
| 1107 | HQ317292 | Unassigned | Unassigned | Unassigned | 662_0 | Clustered | 62 | VC_662_0 | 62 | 0.6976 | 1 | 0.6976 | 0 | 0 | 0 | 1 | Ref |
| 1108 | HQ317384 | Unassigned | Unassigned | Unassigned | 907_0 | Clustered | 3 | VC_907_0 | 3 | 0.6642 | 0.99633114 | 0.6618 | 0 | 0 | 0 | 1 | Ref |
| 1109 | HQ317386 | Unassigned | Unassigned | Unassigned | 909_0 | Clustered | 5 | VC_909_0 | 5 | 0.7406 | 0.9999861 | 0.7406 | 0 | 0 | 0 | 1 | Ref |
| 1110 | HQ317387 | Unassigned | Unassigned | Unassigned | 833_0 | Clustered | 16 | VC_833_0 | 16 | 0.9351 | 1 | 0.9351 | 0 | 0 | 0 | 1 | Ref |
| 1111 | HQ317388 | Unassigned | Unassigned | Unassigned | 340_0 | Clustered | 31 | VC_340_0 | 31 | 0.9549 | 1 | 0.9549 | 0 | 0 | 0 | 1 | Ref |
| 1112 | HQ317389 | Unassigned | Unassigned | Unassigned | 340_0 | Clustered | 31 | VC_340_0 | 31 | 0.9549 | 1 | 0.9549 | 0 | 0 | 0 | 1 | Ref |
| 1113 | HQ317390 | Unassigned | Unassigned | Unassigned | 910_0 | Clustered | 2 | VC_910_0 | 2 | 0.9891 | 0.5 | 0.4946 | 0 | 0 | 0 | 1 | Ref |
| 1114 | HQ317391 | Unassigned | Unassigned | Unassigned | 432_10 | Clustered | 2 | VC_432_10 | 2 | 0.0216 | 0.95771017 | 0.0207 | 0 | 0 | 0 | 1 | Ref |
| 1115 | HQ317392 | Unassigned | Unassigned | Unassigned | 911_0 | Clustered | 6 | VC_911_0 | 6 | 0.2787 | 1 | 0.2787 | 0 | 0 | 0 | 1 | Ref |
| 1116 | HQ317393 | Unassigned | Unassigned | Unassigned | 531_2 | Clustered/Singleton | 1 | VC_531_2 | 1 | 0 | 1 | 0 | 0 | 0 | 0 | 1 | Ref |
| 1117 | HQ332136 | Unassigned | Unassigned | Unassigned | 913_0 | Clustered | 2 | VC_913_0 | 2 | 0.1042 | 0.82306775 | 0.0858 | 0 | 0 | 0 | 1 | Ref |
| 1118 | HQ332137 | Unassigned | Unassigned | Unassigned | 340_0 | Clustered | 31 | VC_340_0 | 31 | 0.9549 | 1 | 0.9549 | 0 | 0 | 0 | 1 | Ref |
| 1119 | HQ332138 | Unassigned | Unassigned | Unassigned | 734_0 | Clustered | 6 | VC_734_0 | 6 | 0.1914 | 0.99996888 | 0.1914 | 0 | 0 | 0 | 1 | Ref |
| 1120 | HQ332140 | Unassigned | Unassigned | Unassigned | 340_0 | Clustered | 31 | VC_340_0 | 31 | 0.9549 | 1 | 0.9549 | 0 | 0 | 0 | 1 | Ref |
| 1121 | HQ332141 | Unassigned | Unassigned | Unassigned | 351_0 | Clustered | 3 | VC_351_0 | 3 | 0.8899 | 0.98155579 | 0.8735 | 0 | 0 | 0 | 1 | Ref |
| 1122 | HQ332142 | Unassigned | Unassigned | Unassigned | 907_0 | Clustered | 3 | VC_907_0 | 3 | 0.6642 | 0.99633114 | 0.6618 | 0 | 0 | 0 | 1 | Ref |
| 1123 | HQ333270 | Unassigned | Unassigned | Unassigned | 386_0 | Clustered | 49 | VC_386_0 | 49 | 0.7941 | 1 | 0.7941 | 0 | 0 | 0 | 1 | Ref |
| 1124 | HQ337021 | Unassigned | Unassigned | Unassigned | 655_2 | Clustered | 3 | VC_655_2 | 3 | 0.0231 | 0.99860756 | 0.0231 | 0 | 0 | 0 | 1 | Ref |
| 1125 | HQ337022 | Unassigned | Unassigned | Unassigned | 340_0 | Clustered | 31 | VC_340_0 | 31 | 0.9549 | 1 | 0.9549 | 0 | 0 | 0 | 1 | Ref |
| 1126 | HQ377372 | Unassigned | Unassigned | Unassigned | 730_0 | Clustered | 9 | VC_730_0 | 9 | 1 | 1 | 1 | 0 | 0 | 0 | 1 | Ref |
| 1127 | HQ377373 | Unassigned | Unassigned | Unassigned | 730_0 | Clustered | 9 | VC_730_0 | 9 | 1 | 1 | 1 | 0 | 0 | 0 | 1 | Ref |
| 1128 | HQ396194 | Unassigned | Unassigned | Unassigned | 916_0 | Clustered | 4 | VC_916_0 | 4 | 0.8472 | 0.99985574 | 0.8471 | 0 | 0 | 0 | 1 | Ref |
| 1129 | HQ403646 | Unassigned | Unassigned | Unassigned | 919_0 | Clustered | 39 | VC_919_0 | 39 | 0.9069 | 1 | 0.9069 | 0 | 0 | 0 | 1 | Ref |
| 1130 | HQ406778 | Unassigned | Unassigned | Unassigned | 190_23 | Clustered | 10 | VC_190_23 | 10 | 0.0704 | 1 | 0.0704 | 0 | 0 | 0 | 0.8444 | Ref |
| 1131 | HQ424691 | Unassigned | Unassigned | Unassigned | 42_0 | Clustered | 49 | VC_42_0 | 49 | 0.4517 | 1 | 0.4517 | 0 | 0 | 0 | 0.8597 | Ref |
| 1132 | HQ630627 | Unassigned | Unassigned | Unassigned | 344_6 | Clustered/Singleton | 1 | VC_344_6 | 1 | 0 | 1 | 0 | 0 | 0 | 0 | 1 | Ref |

| | | | | | | | | | | | | | | | | | |
|---|---|---|---|---|---|---|---|---|---|---|---|---|---|---|---|---|---|
| 1133 | HQ632825 | Unassigned | Unassigned | Unassigned | 432_9 | Clustered | 3 | VC_432_9 | 3 | 0.0372 | 0.99851312 | 0.0371 | 0 | 0 | 0 | 1 | Ref |
| 1134 | HQ632857 | Unassigned | Unassigned | Unassigned | 921_0 | Clustered | 2 | VC_921_0 | 2 | 0.948 | 0.72985431 | 0.6919 | 0 | 0 | 0 | 1 | Ref |
| 1135 | HQ632858 | Unassigned | Unassigned | Unassigned | 921_0 | Clustered | 2 | VC_921_0 | 2 | 0.948 | 0.72985431 | 0.6919 | 0 | 0 | 0 | 1 | Ref |
| 1136 | HQ632859 | Unassigned | Unassigned | Unassigned | 598_0 | Clustered | 4 | VC_598_0 | 4 | 0.0597 | 0.99355358 | 0.0593 | 0 | 0 | 0 | 1 | Ref |
| 1137 | HQ632860 | Unassigned | Unassigned | Unassigned | 642_0 | Clustered | 7 | VC_642_0 | 7 | 0.1508 | 1 | 0.1508 | 0 | 0 | 0 | 0.7619 | Ref |
| 1138 | HQ633062 | Unassigned | Unassigned | Unassigned | 340_0 | Clustered | 31 | VC_340_0 | 31 | 0.9549 | 1 | 0.9549 | 0 | 0 | 0 | 1 | Ref |
| 1139 | HQ633071 | Unassigned | Unassigned | Unassigned | 432_11 | Clustered/Singleton | 1 | VC_432_11 | 1 | 0 | 1 | 0 | 0 | 0 | 0 | 1 | Ref |
| 1140 | HQ634152 | Unassigned | Unassigned | Unassigned | 340_0 | Clustered | 31 | VC_340_0 | 31 | 0.9549 | 1 | 0.9549 | 0 | 0 | 0 | 1 | Ref |
| 1141 | HQ634156 | Unassigned | Unassigned | Unassigned | 493_10 | Clustered/Singleton | 1 | VC_493_10 | 1 | 0 | 1 | 0 | 0 | 0 | 0 | 1 | Ref |
| 1142 | HQ634157 | Unassigned | Unassigned | Unassigned | 493_7 | Clustered/Singleton | 1 | VC_493_7 | 1 | 0 | 1 | 0 | 0 | 0 | 0 | 1 | Ref |
| 1143 | HQ634175 | Unassigned | Unassigned | Unassigned | 655_3 | Clustered/Singleton | 1 | VC_655_3 | 1 | 0 | 1 | 0 | 0 | 0 | 0 | 1 | Ref |
| 1144 | HQ634176 | Unassigned | Unassigned | Unassigned | 655_2 | Clustered | 3 | VC_655_2 | 3 | 0.0231 | 0.99860756 | 0.0231 | 0 | 0 | 0 | 1 | Ref |
| 1145 | HQ634177 | Unassigned | Unassigned | Unassigned | 433_0 | Clustered | 6 | VC_433_0 | 6 | 0.0787 | 1 | 0.0787 | 0 | 0 | 0 | 1 | Ref |
| 1146 | HQ634187 | Unassigned | Unassigned | Unassigned | 356_0 | Clustered | 2 | VC_356_0 | 2 | 0.1992 | 0.93211095 | 0.1857 | 0 | 0 | 0 | 1 | Ref |
| 1147 | HQ634188 | Unassigned | Unassigned | Unassigned | 340_0 | Clustered | 31 | VC_340_0 | 31 | 0.9549 | 1 | 0.9549 | 0 | 0 | 0 | 1 | Ref |
| 1148 | HQ634189 | Unassigned | Unassigned | Unassigned | 655_7 | Clustered | 3 | VC_655_7 | 3 | 0.0166 | 0.99856978 | 0.0166 | 0 | 0 | 0 | 1 | Ref |
| 1149 | HQ634190 | Unassigned | Unassigned | Unassigned | 655_6 | Clustered | 2 | VC_655_6 | 2 | 0.0078 | 0.95775168 | 0.0075 | 0 | 0 | 0 | 1 | Ref |
| 1150 | HQ634192 | Unassigned | Unassigned | Unassigned | 929_0 | Clustered | 4 | VC_929_0 | 4 | 0.846 | 0.99722201 | 0.8437 | 0 | 0 | 0 | 1 | Ref |
| 1151 | HQ634195 | Unassigned | Unassigned | Unassigned | 493_15 | Clustered/Singleton | 1 | VC_493_15 | 1 | 0 | 1 | 0 | 0 | 0 | 0 | 1 | Ref |
| 1152 | HQ641340 | Unassigned | Unassigned | Unassigned | 82_0 | Clustered | 24 | VC_82_0 | 24 | 0.7351 | 1 | 0.7351 | 0 | 0 | 0 | 1 | Ref |
| 1153 | HQ641341 | Unassigned | Unassigned | Unassigned | 82_0 | Clustered | 24 | VC_82_0 | 24 | 0.7351 | 1 | 0.7351 | 0 | 0 | 0 | 1 | Ref |
| 1154 | HQ641342 | Unassigned | Unassigned | Unassigned | 82_0 | Clustered | 24 | VC_82_0 | 24 | 0.7351 | 1 | 0.7351 | 0 | 0 | 0 | 1 | Ref |
| 1155 | HQ641343 | Unassigned | Unassigned | Unassigned | 82_0 | Clustered | 24 | VC_82_0 | 24 | 0.7351 | 1 | 0.7351 | 0 | 0 | 0 | 1 | Ref |
| 1156 | HQ641344 | Unassigned | Unassigned | Unassigned | 82_0 | Clustered | 24 | VC_82_0 | 24 | 0.7351 | 1 | 0.7351 | 0 | 0 | 0 | 1 | Ref |
| 1157 | HQ641345 | Unassigned | Unassigned | Unassigned | 754_0 | Clustered | 8 | VC_754_0 | 8 | 0.752 | 1 | 0.752 | 0 | 0 | 0 | 1 | Ref |
| 1158 | HQ641346 | Unassigned | Unassigned | Unassigned | 754_0 | Clustered | 8 | VC_754_0 | 8 | 0.752 | 1 | 0.752 | 0 | 0 | 0 | 1 | Ref |
| 1159 | HQ641347 | Unassigned | Unassigned | Unassigned | 495_1 | Clustered | 41 | VC_495_1 | 41 | 0.328 | 1 | 0.328 | 0 | 0 | 0 | 0.911 | Ref |
| 1160 | HQ641348 | Unassigned | Unassigned | Unassigned | 495_1 | Clustered | 41 | VC_495_1 | 41 | 0.328 | 1 | 0.328 | 0 | 0 | 0 | 0.911 | Ref |
| 1161 | HQ641349 | Unassigned | Unassigned | Unassigned | 495_1 | Clustered | 41 | VC_495_1 | 41 | 0.328 | 1 | 0.328 | 0 | 0 | 0 | 0.911 | Ref |
| 1162 | HQ641350 | Unassigned | Unassigned | Unassigned | 495_1 | Clustered | 41 | VC_495_1 | 41 | 0.328 | 1 | 0.328 | 0 | 0 | 0 | 0.911 | Ref |
| 1163 | HQ641351 | Unassigned | Unassigned | Unassigned | 495_1 | Clustered | 41 | VC_495_1 | 41 | 0.328 | 1 | 0.328 | 0 | 0 | 0 | 0.911 | Ref |
| 1164 | HQ641352 | Unassigned | Unassigned | Unassigned | 495_1 | Clustered | 41 | VC_495_1 | 41 | 0.328 | 1 | 0.328 | 0 | 0 | 0 | 0.911 | Ref |
| 1165 | HQ641353 | Unassigned | Unassigned | Unassigned | 495_1 | Clustered | 41 | VC_495_1 | 41 | 0.328 | 1 | 0.328 | 0 | 0 | 0 | 0.911 | Ref |
| 1166 | HQ641354 | Unassigned | Unassigned | Unassigned | 495_1 | Clustered | 41 | VC_495_1 | 41 | 0.328 | 1 | 0.328 | 0 | 0 | 0 | 0.911 | Ref |
| 1167 | HQ665011 | Unassigned | Unassigned | Unassigned | 190_24 | Clustered | 5 | VC_190_24 | 5 | 0.0293 | 0.9999996 | 0.0293 | 0 | 0 | 0 | 0.7 | Ref |
| 1168 | HQ683709 | Unassigned | Unassigned | Unassigned | 932_0 | Clustered | 2 | VC_932_0 | 2 | 0.4438 | 0.89435023 | 0.3969 | 0 | 0 | 0 | 1 | Ref |
| 1169 | HQ698895 | Unassigned | Unassigned | Unassigned | 927_0 | Clustered | 2 | VC_927_0 | 2 | 0.0876 | 0.71936208 | 0.063 | 0 | 0 | 0 | 1 | Ref |
| 1170 | HQ698922 | Unassigned | Unassigned | Unassigned | 492_0 | Clustered | 2 | VC_492_0 | 2 | 0.0158 | 0.6640397 | 0.0105 | 0 | 0 | 0 | 1 | Ref |
| 1171 | HQ711984 | Unassigned | Unassigned | Unassigned | 297_1 | Clustered | 20 | VC_297_1 | 20 | 0.5653 | 1 | 0.5653 | 0 | 0 | 0 | 0.7684 | Ref |
| 1172 | HQ711985 | Unassigned | Unassigned | Unassigned | 176_0 | Clustered | 2 | VC_176_0 | 2 | 0.0498 | 0.95663741 | 0.0476 | 0 | 0 | 0 | 1 | Ref |
| 1173 | HQ728263 | Unassigned | Unassigned | Unassigned | 328_0 | Clustered | 11 | VC_328_0 | 11 | 0.4707 | 1 | 0.4707 | 0 | 0 | 0 | 1 | Ref |
| 1174 | HQ728264 | Unassigned | Unassigned | Unassigned | 792_0 | Clustered | 35 | VC_792_0 | 35 | 0.9828 | 1 | 0.9828 | 0 | 0 | 0 | 1 | Ref |
| 1175 | HQ728265 | Unassigned | Unassigned | Unassigned | 387_0 | Clustered | 3 | VC_387_0 | 3 | 0.0424 | 0.99861589 | 0.0423 | 0 | 0 | 0 | 1 | Ref |
| 1176 | HQ728524 | Unassigned | Unassigned | Unassigned | 520_26 | Clustered | 3 | VC_520_26 | 3 | 0.026 | 0.99859898 | 0.026 | 0 | 0 | 0 | 1 | Ref |
| 1177 | HQ829472 | Unassigned | Unassigned | Unassigned | 290_5 | Clustered | 14 | VC_290_5 | 14 | 0.1422 | 1 | 0.1422 | 0 | 0 | 0 | 0.9231 | Ref |
| 1178 | HQ832595 | Unassigned | Unassigned | Unassigned | 76_0 | Clustered | 37 | VC_76_0 | 37 | 0.9403 | 1 | 0.9403 | 0 | 0 | 0 | 1 | Ref |
| 1179 | HQ906662 | Unassigned | Unassigned | Unassigned | 22_0 | Clustered | 11 | VC_22_0 | 11 | 0.5163 | 1 | 0.5163 | 0 | 0 | 0 | 1 | Ref |
| 1180 | HQ906663 | Unassigned | Unassigned | Unassigned | 22_0 | Clustered | 11 | VC_22_0 | 11 | 0.5163 | 1 | 0.5163 | 0 | 0 | 0 | 1 | Ref |
| 1181 | HQ906664 | Unassigned | Unassigned | Unassigned | 22_0 | Clustered | 11 | VC_22_0 | 11 | 0.5163 | 1 | 0.5163 | 0 | 0 | 0 | 1 | Ref |
| 1182 | HQ918180 | Unassigned | Unassigned | Unassigned | 668_0 | Clustered | 25 | VC_668_0 | 25 | 0.5539 | 1 | 0.5539 | 0 | 0 | 0 | 0.96 | Ref |
| 1183 | JF412296 | Unassigned | Unassigned | Unassigned | 613_0 | Clustered | 2 | VC_613_0 | 2 | 0.0357 | 0.94194318 | 0.0336 | 0 | 0 | 0 | 1 | Ref |
| 1184 | JF461087 | Unassigned | Unassigned | Unassigned | 329_1 | Clustered | 17 | VC_329_1 | 17 | 0.0867 | 1 | 0.0867 | 0 | 0 | 0 | 0.8235 | Ref |
| 1185 | JF501022 | Unassigned | Unassigned | Unassigned | 938_0 | Clustered | 99 | VC_938_0 | 99 | 0.9119 | 1 | 0.9119 | 0 | 0 | 0 | 0.9928 | Ref |
| 1186 | JF704091 | Unassigned | Unassigned | Unassigned | 371_18 | Clustered | 10 | VC_371_18 | 10 | 0.0302 | 1 | 0.0302 | 0 | 0 | 0 | 0.8 | Ref |
| 1187 | JF704092 | Unassigned | Unassigned | Unassigned | 558_22 | Clustered | 4 | VC_558_22 | 4 | 0.01 | 0.99312819 | 0.0099 | 0 | 0 | 0 | 0.8333 | Ref |
| 1188 | JF704093 | Unassigned | Unassigned | Unassigned | 517_0 | Clustered | 18 | VC_517_0 | 18 | 0.1599 | 1 | 0.1599 | 0 | 0 | 0 | 1 | Ref |
| 1189 | JF704094 | Unassigned | Unassigned | Unassigned | 372_0 | Clustered | 24 | VC_372_0 | 24 | 0.5539 | 1 | 0.5539 | 0 | 0 | 0 | 1 | Ref |
| 1190 | JF704095 | Unassigned | Unassigned | Unassigned | 509_0 | Clustered | 37 | VC_509_0 | 37 | 0.9689 | 1 | 0.9689 | 0 | 0 | 0 | 1 | Ref |
| 1191 | JF704096 | Unassigned | Unassigned | Unassigned | 558_20 | Clustered | 17 | VC_558_20 | 17 | 0.054 | 1 | 0.054 | 0 | 0 | 0 | 0.9926 | Ref |
| 1192 | JF704097 | Unassigned | Unassigned | Unassigned | 223_0 | Clustered | 38 | VC_223_0 | 38 | 0.9095 | 1 | 0.9095 | 0 | 0 | 0 | 1 | Ref |
| 1193 | JF704098 | Unassigned | Unassigned | Unassigned | 514_0 | Clustered | 75 | VC_514_0 | 75 | 0.9402 | 1 | 0.9402 | 0 | 0 | 0 | 1 | Ref |
| 1194 | JF704099 | Unassigned | Unassigned | Unassigned | 371_3 | Clustered | 22 | VC_371_3 | 22 | 0.0787 | 1 | 0.0787 | 0 | 0 | 0 | 0.9048 | Ref |
| 1195 | JF704100 | Unassigned | Unassigned | Unassigned | 939_0 | Clustered | 15 | VC_939_0 | 15 | 1 | 0 | 0 | 0 | 0 | 0 | 1 | Ref |
| 1196 | JF704101 | Unassigned | Unassigned | Unassigned | 514_0 | Clustered | 75 | VC_514_0 | 75 | 0.9402 | 1 | 0.9402 | 0 | 0 | 0 | 1 | Ref |
| 1197 | JF704102 | Unassigned | Unassigned | Unassigned | 371_16 | Clustered | 42 | VC_371_16 | 42 | 0.1341 | 1 | 0.1341 | 0 | 0 | 0 | 0.7282 | Ref |
| 1198 | JF704103 | Unassigned | Unassigned | Unassigned | 371_15 | Clustered | 44 | VC_371_15 | 44 | 0.1391 | 1 | 0.1391 | 0 | 0 | 0 | 0.7474 | Ref |
| 1199 | JF704104 | Unassigned | Unassigned | Unassigned | 372_0 | Clustered | 24 | VC_372_0 | 24 | 0.5539 | 1 | 0.5539 | 0 | 0 | 0 | 1 | Ref |
| 1200 | JF704105 | Unassigned | Unassigned | Unassigned | 236_0 | Clustered | 72 | VC_236_0 | 72 | 0.8104 | 1 | 0.8104 | 0 | 0 | 0 | 1 | Ref |
| 1201 | JF704106 | Unassigned | Unassigned | Unassigned | 236_0 | Clustered | 72 | VC_236_0 | 72 | 0.8104 | 1 | 0.8104 | 0 | 0 | 0 | 1 | Ref |
| 1202 | JF704107 | Unassigned | Unassigned | Unassigned | 940_0 | Clustered | 34 | VC_940_0 | 34 | 0.7959 | 1 | 0.7959 | 0 | 0 | 0 | 1 | Ref |
| 1203 | JF704108 | Unassigned | Unassigned | Unassigned | 887_0 | Clustered | 18 | VC_887_0 | 18 | 0.3663 | 1 | 0.3663 | 0 | 0 | 0 | 1 | Ref |
| 1204 | JF704109 | Unassigned | Unassigned | Unassigned | 371_15 | Clustered | 44 | VC_371_15 | 44 | 0.1391 | 1 | 0.1391 | 0 | 0 | 0 | 0.7474 | Ref |
| 1205 | JF704110 | Unassigned | Unassigned | Unassigned | 225_0 | Clustered | 32 | VC_225_0 | 32 | 0.848 | 1 | 0.848 | 0 | 0 | 0 | 0.996 | Ref |
| 1206 | JF704111 | Unassigned | Unassigned | Unassigned | 513_0 | Clustered | 32 | VC_513_0 | 32 | 0.6881 | 1 | 0.6881 | 0 | 0 | 0 | 1 | Ref |
| 1207 | JF704112 | Unassigned | Unassigned | Unassigned | 568_0 | Clustered | 8 | VC_568_0 | 8 | 0.5738 | 1 | 0.5738 | 0 | 0 | 0 | 1 | Ref |
| 1208 | JF704113 | Unassigned | Unassigned | Unassigned | 887_0 | Clustered | 18 | VC_887_0 | 18 | 0.3663 | 1 | 0.3663 | 0 | 0 | 0 | 1 | Ref |
| 1209 | JF704114 | Unassigned | Unassigned | Unassigned | 514_0 | Clustered | 75 | VC_514_0 | 75 | 0.9402 | 1 | 0.9402 | 0 | 0 | 0 | 1 | Ref |
| 1210 | JF704116 | Unassigned | Unassigned | Unassigned | 558_6 | Clustered | 2 | VC_558_6 | 2 | 0.0032 | 0.73149279 | 0.0023 | 0 | 0 | 0 | 1 | Ref |
| 1211 | JF704117 | Unassigned | Unassigned | Unassigned | 520_74 | Clustered | 2 | VC_520_74 | 2 | 0.0075 | 0.95760586 | 0.0072 | 0 | 0 | 0 | 1 | Ref |
| 1212 | JF712866 | Unassigned | Unassigned | Unassigned | 607_0 | Clustered | 16 | VC_607_0 | 16 | 0.9179 | 1 | 0.9179 | 0 | 0 | 0 | 1 | Ref |
| 1213 | JF713456 | Unassigned | Unassigned | Unassigned | 904_0 | Clustered | 6 | VC_904_0 | 6 | 0.233 | 1 | 0.233 | 0 | 0 | 0 | 1 | Ref |
| 1214 | JF719726 | Unassigned | Unassigned | Unassigned | 303_0 | Clustered | 155 | VC_303_0 | 155 | 0.9979 | 1 | 0.9979 | 0 | 0 | 0 | 1 | Ref |
| 1215 | JF719727 | Unassigned | Unassigned | Unassigned | 303_0 | Clustered | 155 | VC_303_0 | 155 | 0.9979 | 1 | 0.9979 | 0 | 0 | 0 | 1 | Ref |
| 1216 | JF719728 | Unassigned | Unassigned | Unassigned | 303_0 | Clustered | 155 | VC_303_0 | 155 | 0.9979 | 1 | 0.9979 | 0 | 0 | 0 | 1 | Ref |
| 1217 | JF719729 | Unassigned | Unassigned | Unassigned | 360_0 | Clustered | 358 | VC_360_0 | 358 | 0.9615 | 1 | 0.9615 | 0 | 0 | 0 | 1 | Ref |
| 1218 | JF719730 | Unassigned | Unassigned | Unassigned | 360_0 | Clustered | 358 | VC_360_0 | 358 | 0.9615 | 1 | 0.9615 | 0 | 0 | 0 | 1 | Ref |
| 1219 | JF719731 | Unassigned | Unassigned | Unassigned | 360_0 | Clustered | 358 | VC_360_0 | 358 | 0.9615 | 1 | 0.9615 | 0 | 0 | 0 | 1 | Ref |
| 1220 | JF719732 | Unassigned | Unassigned | Unassigned | 61_0 | Clustered | 12 | VC_61_0 | 12 | 0.7633 | 1 | 0.7633 | 0 | 0 | 0 | 1 | Ref |
| 1221 | JF719733 | Unassigned | Unassigned | Unassigned | 61_0 | Clustered | 12 | VC_61_0 | 12 | 0.7633 | 1 | 0.7633 | 0 | 0 | 0 | 1 | Ref |
| 1222 | JF719734 | Unassigned | Unassigned | Unassigned | 61_0 | Clustered | 12 | VC_61_0 | 12 | 0.7633 | 1 | 0.7633 | 0 | 0 | 0 | 1 | Ref |
| 1223 | JF731128 | Unassigned | Unassigned | Unassigned | 108_0 | Clustered | 33 | VC_108_0 | 33 | 0.949 | 1 | 0.949 | 0 | 0 | 0 | 1 | Ref |
| 1224 | JF734911 | Unassigned | Unassigned | Unassigned | 99_0 | Clustered | 32 | VC_99_0 | 32 | 0.8828 | 1 | 0.8828 | 0 | 0 | 0 | 1 | Ref |
| 1225 | JF744988 | Unassigned | Unassigned | Unassigned | 888_0 | Clustered | 28 | VC_888_0 | 28 | 0.5497 | 1 | 0.5497 | 0 | 0 | 0 | 1 | Ref |
| 1226 | JF767208 | Unassigned | Unassigned | Unassigned | 815_0 | Clustered | 8 | VC_815_0 | 8 | 0.9801 | 1 | 0.9801 | 0 | 0 | 0 | 1 | Ref |
| 1227 | JF767209 | Unassigned | Unassigned | Unassigned | 815_0 | Clustered | 8 | VC_815_0 | 8 | 0.9801 | 1 | 0.9801 | 0 | 0 | 0 | 1 | Ref |
| 1228 | JF767210 | Unassigned | Unassigned | Unassigned | 815_0 | Clustered | 8 | VC_815_0 | 8 | 0.9801 | 1 | 0.9801 | 0 | 0 | 0 | 1 | Ref |
| 1229 | JF770475 | Unassigned | Unassigned | Unassigned | 455_0 | Clustered | 18 | VC_455_0 | 18 | 0.4598 | 1 | 0.4598 | 0 | 0 | 0 | 1 | Ref |
| 1230 | JF773396 | Unassigned | Unassigned | Unassigned | 730_0 | Clustered | 9 | VC_730_0 | 9 | 1 | 1 | 1 | 0 | 0 | 0 | 1 | Ref |
| 1231 | JF792674 | Unassigned | Unassigned | Unassigned | 517_1 | Clustered | 108 | VC_517_1 | 108 | 0.8483 | 1 | 0.8483 | 0 | 0 | 0 | 0.9955 | Ref |
| 1232 | JF832915 | Unassigned | Unassigned | Unassigned | 181_0 | Clustered | 5 | VC_181_0 | 5 | 0.1168 | 0.99999995 | 0.1168 | 0 | 0 | 0 | 1 | Ref |
| 1233 | JF900176 | Unassigned | Unassigned | Unassigned | 28_1 | Clustered | 53 | VC_28_1 | 53 | 0.7164 | 1 | 0.7164 | 0 | 0 | 0 | 0.9325 | Ref |
| 1234 | JF917095 | Unassigned | Unassigned | Unassigned | 943_0 | Clustered | 2 | VC_943_0 | 2 | 1 | 1 | 1 | 0 | 0 | 0 | 1 | Ref |
| 1235 | JF923796 | Unassigned | Unassigned | Unassigned | 767_0 | Clustered | 34 | VC_767_0 | 34 | 0.9173 | 1 | 0.9173 | 0 | 0 | 0 | 1 | Ref |
| 1236 | JF923797 | Unassigned | Unassigned | Unassigned | 767_0 | Clustered | 34 | VC_767_0 | 34 | 0.9173 | 1 | 0.9173 | 0 | 0 | 0 | 1 | Ref |
| 1237 | JF937090 | Unassigned | Unassigned | Unassigned | 546_7 | Clustered | 4 | VC_546_7 | 4 | 0.041 | 0.9963251 | 0.0408 | 0 | 0 | 0 | 1 | Ref |
| 1238 | JF937091 | Unassigned | Unassigned | Unassigned | 551_0 | Clustered | 16 | VC_551_0 | 16 | 0.0915 | 1 | 0.0915 | 0 | 0 | 0 | 0.6667 | Ref |
| 1239 | JF937092 | Unassigned | Unassigned | Unassigned | 223_0 | Clustered | 38 | VC_223_0 | 38 | 0.9095 | 1 | 0.9095 | 0 | 0 | 0 | 1 | Ref |
| 1240 | JF937093 | Unassigned | Unassigned | Unassigned | 520_31 | Clustered | 11 | VC_520_31 | 11 | 0.0897 | 1 | 0.0897 | 0 | 0 | 0 | 0.6 | Ref |
| 1241 | JF937094 | Unassigned | Unassigned | Unassigned | 223_0 | Clustered | 38 | VC_223_0 | 38 | 0.9095 | 1 | 0.9095 | 0 | 0 | 0 | 1 | Ref |
| 1242 | JF937095 | Unassigned | Unassigned | Unassigned | 371_17 | Clustered | 3 | VC_371_17 | 3 | 0.0076 | 0.99811676 | 0.0076 | 0 | 0 | 0 | 1 | Ref |
| 1243 | JF937096 | Unassigned | Unassigned | Unassigned | 551_2 | Clustered | 17 | VC_551_2 | 17 | 0.0973 | 1 | 0.0973 | 0 | 0 | 0 | 0.9779 | Ref |
| 1244 | JF937097 | Unassigned | Unassigned | Unassigned | 371_15 | Clustered | 44 | VC_371_15 | 44 | 0.1391 | 1 | 0.1391 | 0 | 0 | 0 | 0.7474 | Ref |
| 1245 | JF937098 | Unassigned | Unassigned | Unassigned | 520_93 | Clustered | 2 | VC_520_93 | 2 | 0.0104 | 0.95762079 | 0.0099 | 0 | 0 | 0 | 1 | Ref |
| 1246 | JF937099 | Unassigned | Unassigned | Unassigned | 325_2 | Clustered/Singleton | 1 | VC_325_2 | 1 | 0 | 1 | 0 | 0 | 0 | 0 | 1 | Ref |
| 1247 | JF937100 | Unassigned | Unassigned | Unassigned | 325_41 | Clustered | 2 | VC_325_41 | 2 | 0.0036 | 0.94071129 | 0.0034 | 0 | 0 | 0 | 1 | Ref |
| 1248 | JF937101 | Unassigned | Unassigned | Unassigned | 546_11 | Clustered/Singleton | 1 | VC_546_11 | 1 | 0 | 1 | 0 | 0 | 0 | 0 | 1 | Ref |
| 1249 | JF937102 | Unassigned | Unassigned | Unassigned | 520_37 | Clustered/Singleton | 1 | VC_520_37 | 1 | 0 | 1 | 0 | 0 | 0 | 0 | 1 | Ref |
| 1250 | JF937103 | Unassigned | Unassigned | Unassigned | 325_23 | Clustered | 17 | VC_325_23 | 17 | 0.0716 | 1 | 0.0716 | 0 | 0 | 0 | 0.6397 | Ref |
| 1251 | JF937104 | Unassigned | Unassigned | Unassigned | 237_1 | Clustered | 13 | VC_237_1 | 13 | 0.1773 | 1 | 0.1773 | 0 | 0 | 0 | 0.7308 | Ref |
| 1252 | JF937105 | Unassigned | Unassigned | Unassigned | 763_2 | Clustered | 7 | VC_763_2 | 7 | 0.2612 | 1 | 0.2612 | 0 | 0 | 0 | 1 | Ref |
| 1253 | JF937106 | Unassigned | Unassigned | Unassigned | 551_0 | Clustered | 16 | VC_551_0 | 16 | 0.0915 | 1 | 0.0915 | 0 | 0 | 0 | 0.6667 | Ref |
| 1254 | JF937107 | Unassigned | Unassigned | Unassigned | 565_0 | Clustered | 20 | VC_565_0 | 20 | 0.8398 | 1 | 0.8398 | 0 | 0 | 0 | 1 | Ref |
| 1255 | JF937108 | Unassigned | Unassigned | Unassigned | 325_5 | Clustered | 11 | VC_325_5 | 11 | 0.047 | 1 | 0.047 | 0 | 0 | 0 | 0.7636 | Ref |
| 1256 | JF937109 | Unassigned | Unassigned | Unassigned | 371_18 | Clustered | 10 | VC_371_18 | 10 | 0.0302 | 1 | 0.0302 | 0 | 0 | 0 | 0.8 | Ref |
| 1257 | JF937110 | Unassigned | Unassigned | Unassigned | 325_5 | Clustered | 11 | VC_325_5 | 11 | 0.047 | 1 | 0.047 | 0 | 0 | 0 | 0.7636 | Ref |
| 1258 | JF939047 | Unassigned | Unassigned | Unassigned | 749_0 | Clustered | 40 | VC_749_0 | 40 | 0.7517 | 1 | 0.7517 | 0 | 0 | 0 | 1 | Ref |
| 1259 | JF946695 | Unassigned | Unassigned | Unassigned | 215_2 | Clustered | 5 | VC_215_2 | 5 | 0.0572 | 0.99999997 | 0.0572 | 0 | 0 | 0 | 1 | Ref |
| 1260 | JF957056 | Unassigned | Unassigned | Unassigned | 371_15 | Clustered | 44 | VC_371_15 | 44 | 0.1391 | 1 | 0.1391 | 0 | 0 | 0 | 0.7474 | Ref |
| 1261 | JF957057 | Unassigned | Unassigned | Unassigned | 325_24 | Clustered | 3 | VC_325_24 | 3 | 0.009 | 0.99853949 | 0.009 | 0 | 0 | 0 | 1 | Ref |
| 1262 | JF957058 | Unassigned | Unassigned | Unassigned | 513_0 | Clustered | 32 | VC_513_0 | 32 | 0.6881 | 1 | 0.6881 | 0 | 0 | 0 | 1 | Ref |
| 1263 | JF957059 | Unassigned | Unassigned | Unassigned | 546_7 | Clustered | 4 | VC_546_7 | 4 | 0.041 | 0.9963251 | 0.0408 | 0 | 0 | 0 | 1 | Ref |
| 1264 | JF957060 | Unassigned | Unassigned | Unassigned | 219_0 | Clustered | 5 | VC_219_0 | 5 | 0.08 | 0.99999998 | 0.08 | 0 | 0 | 0 | 1 | Ref |
| 1265 | JF966203 | Unassigned | Unassigned | Unassigned | 883_0 | Clustered | 10 | VC_883_0 | 10 | 0.2015 | 1 | 0.2015 | 0 | 0 | 0 | 1 | Ref |
| 1266 | JF974286 | Unassigned | Unassigned | Unassigned | 928_0 | Clustered | 3 | VC_928_0 | 3 | 0.7844 | 0.9944138 | 0.78 | 0 | 0 | 0 | 1 | Ref |
| 1267 | JF974287 | Unassigned | Unassigned | Unassigned | 643_0 | Clustered | 2 | VC_643_0 | 2 | 0.1092 | 0.95085347 | 0.1038 | 0 | 0 | 0 | 1 | Ref |
| 1268 | JF974294 | Unassigned | Unassigned | Unassigned | 460_0 | Clustered | 19 | VC_460_0 | 19 | 0.3744 | 1 | 0.3744 | 0 | 0 | 0 | 1 | Ref |
| 1269 | JF974295 | Unassigned | Unassigned | Unassigned | 427_0 | Clustered | 3 | VC_427_0 | 3 | 0.0627 | 0.96830121 | 0.0608 | 0 | 0 | 0 | 1 | Ref |
| 1270 | JF974307 | Unassigned | Unassigned | Unassigned | 182_0 | Clustered | 5 | VC_182_0 | 5 | 0.6184 | 0.99999967 | 0.6184 | 0 | 0 | 0 | 1 | Ref |
| 1271 | JF974308 | Unassigned | Unassigned | Unassigned | 182_0 | Clustered | 5 | VC_182_0 | 5 | 0.6184 | 0.99999967 | 0.6184 | 0 | 0 | 0 | 1 | Ref |
| 1272 | JF974312 | Unassigned | Unassigned | Unassigned | 460_0 | Clustered | 19 | VC_460_0 | 19 | 0.3744 | 1 | 0.3744 | 0 | 0 | 0 | 1 | Ref |
| 1273 | JF974313 | Unassigned | Unassigned | Unassigned | 642_2 | Clustered | 6 | VC_642_2 | 6 | 0.1679 | 1 | 0.1679 | 0 | 0 | 0 | 1 | Ref |
| 1274 | JF974314 | Unassigned | Unassigned | Unassigned | 21_0 | Clustered | 4 | VC_21_0 | 4 | 0.5696 | 0.87262442 | 0.4971 | 0 | 0 | 0 | 0.6667 | Ref |
| 1275 | JF974321 | Unassigned | Unassigned | Unassigned | 928_0 | Clustered | 3 | VC_928_0 | 3 | 0.7844 | 0.9944138 | 0.78 | 0 | 0 | 0 | 1 | Ref |
| 1276 | JF974339 | Unassigned | Unassigned | Unassigned | 30_1 | Clustered | 16 | VC_30_1 | 16 | 0.1973 | 1 | 0.1973 | 0 | 0 | 0 | 0.8333 | Ref |
| 1277 | JN006061 | Unassigned | Unassigned | Unassigned | 551_2 | Clustered | 17 | VC_551_2 | 17 | 0.0973 | 1 | 0.0973 | 0 | 0 | 0 | 0.9779 | Ref |
| 1278 | JN006062 | Unassigned | Unassigned | Unassigned | 551_2 | Clustered | 17 | VC_551_2 | 17 | 0.0973 | 1 | 0.0973 | 0 | 0 | 0 | 0.9779 | Ref |
| 1279 | JN006063 | Unassigned | Unassigned | Unassigned | 371_15 | Clustered | 44 | VC_371_15 | 44 | 0.1391 | 1 | 0.1391 | 0 | 0 | 0 | 0.7474 | Ref |
| 1280 | JN006064 | Unassigned | Unassigned | Unassigned | 371_21 | Clustered | 2 | VC_371_21 | 2 | 0.0033 | 0.95770737 | 0.0032 | 0 | 0 | 0 | 1 | Ref |
| 1281 | JN020140 | Unassigned | Unassigned | Unassigned | 325_19 | Clustered | 3 | VC_325_19 | 3 | 0.0108 | 0.99859221 | 0.0107 | 0 | 0 | 0 | 1 | Ref |
| 1282 | JN020141 | Unassigned | Unassigned | Unassigned | 520_28 | Clustered | 2 | VC_520_28 | 2 | 0.0137 | 0.95761531 | 0.0131 | 0 | 0 | 0 | 1 | Ref |
| 1283 | JN020142 | Unassigned | Unassigned | Unassigned | 520_88 | Clustered/Singleton | 1 | VC_520_88 | 1 | 0 | 1 | 0 | 0 | 0 | 0 | 1 | Ref |
| 1284 | JN020143 | Unassigned | Unassigned | Unassigned | 520_43 | Clustered | 3 | VC_520_43 | 3 | 0.0193 | 0.99837043 | 0.0193 | 0 | 0 | 0 | 1 | Ref |
| 1285 | JN022534 | Unassigned | Unassigned | Unassigned | 64_0 | Clustered | 3 | VC_64_0 | 3 | 0.5503 | 0.99785685 | 0.5491 | 0 | 0 | 0 | 1 | Ref |
| 1286 | JN035618 | Unassigned | Unassigned | Unassigned | 866_0 | Clustered | 4 | VC_866_0 | 4 | 0.0795 | 0.99998595 | 0.0795 | 0 | 0 | 0 | 1 | Ref |
| 1287 | JN049605 | Unassigned | Unassigned | Unassigned | 223_0 | Clustered | 38 | VC_223_0 | 38 | 0.9095 | 1 | 0.9095 | 0 | 0 | 0 | 1 | Ref |
| 1288 | JN051154 | Unassigned | Unassigned | Unassigned | 577_0 | Clustered | 6 | VC_577_0 | 6 | 0.9267 | 0.99999993 | 0.9267 | 0 | 0 | 0 | 1 | Ref |
| 1289 | JN083852 | Unassigned | Unassigned | Unassigned | 940_0 | Clustered | 34 | VC_940_0 | 34 | 0.7959 | 1 | 0.7959 | 0 | 0 | 0 | 1 | Ref |
| 1290 | JN083853 | Unassigned | Unassigned | Unassigned | 940_0 | Clustered | 34 | VC_940_0 | 34 | 0.7959 | 1 | 0.7959 | 0 | 0 | 0 | 1 | Ref |
| 1291 | JN116822 | Unassigned | Unassigned | Unassigned | 949_0 | Clustered | 10 | VC_949_0 | 10 | 0.5977 | 1 | 0.5977 | 0 | 0 | 0 | 1 | Ref |
| 1292 | JN116823 | Unassigned | Unassigned | Unassigned | 953_0 | Clustered | 2 | VC_953_0 | 2 | 0.2116 | 0.94820857 | 0.2007 | 0 | 0 | 0 | 1 | Ref |
| 1293 | JN116824 | Unassigned | Unassigned | Unassigned | 726_0 | Clustered | 3 | VC_726_0 | 3 | 0.0381 | 0.88955608 | 0.0339 | 0 | 0 | 0 | 1 | Ref |
| 1294 | JN116826 | Unassigned | Unassigned | Unassigned | 956_0 | Clustered | 39 | VC_956_0 | 39 | 1 | 1 | 1 | 0 | 0 | 0 | 1 | Ref |

| | | | | | | | | | | | | | | | | | |
|---|---|---|---|---|---|---|---|---|---|---|---|---|---|---|---|---|---|
| 1295 | JN116827 | Unassigned | Unassigned | Unassigned | 956_0 | Clustered | 39 | VC_956_0 | 39 | 1 | 1 | 1 | 0 | 0 | 0 | 1 | Ref |
| 1296 | JN116828 | Unassigned | Unassigned | Unassigned | 952_0 | Clustered | 2 | VC_952_0 | 2 | 0.9556 | 0.8861 | 0.8468 | 0 | 0 | 0 | 1 | Ref |
| 1297 | JN126049 | Unassigned | Unassigned | Unassigned | 133_13 | Clustered | 2 | VC_133_13 | 2 | 0.0051 | 0.86794768 | 0.0044 | 0 | 0 | 0 | 1 | Ref |
| 1298 | JN132397 | Unassigned | Unassigned | Unassigned | 844_0 | Clustered | 27 | VC_844_0 | 27 | 0.9385 | 1 | 0.9385 | 0 | 0 | 0 | 1 | Ref |
| 1299 | JN153085 | Unassigned | Unassigned | Unassigned | 325_52 | Clustered | 5 | VC_325_52 | 5 | 0.0147 | 0.99999073 | 0.0147 | 0 | 0 | 0 | 0.7 | Ref |
| 1300 | JN153086 | Unassigned | Unassigned | Unassigned | 325_61 | Clustered | 8 | VC_325_61 | 8 | 0.0395 | 1 | 0.0395 | 0 | 0 | 0 | 0.7857 | Ref |
| 1301 | JN175269 | Unassigned | Unassigned | Unassigned | 957_0 | Clustered/Singleton | 1 | VC_957_0 | 1 | 0 | 1 | 0 | 0 | 0 | 0 | 1 | Ref |
| 1302 | JN185608 | Unassigned | Unassigned | Unassigned | 236_0 | Clustered | 72 | VC_236_0 | 72 | 0.8104 | 1 | 0.8104 | 0 | 0 | 0 | 1 | Ref |
| 1303 | JN190960 | Unassigned | Unassigned | Unassigned | 602_0 | Clustered | 2 | VC_602_0 | 2 | 0.229 | 0.95560552 | 0.2188 | 0 | 0 | 0 | 1 | Ref |
| 1304 | JN192463 | Unassigned | Unassigned | Unassigned | 371_13 | Clustered | 33 | VC_371_13 | 33 | 0.179 | 1 | 0.179 | 0 | 0 | 0 | 0.8769 | Ref |
| 1305 | JN201525 | Unassigned | Unassigned | Unassigned | 546_0 | Clustered | 2 | VC_546_0 | 2 | 0.0136 | 0.63748598 | 0.0087 | 0 | 0 | 0 | 1 | Ref |
| 1306 | JN202312 | Unassigned | Unassigned | Unassigned | 283_148 | Clustered/Singleton | 1 | VC_283_148 | 1 | 0 | 1 | 0 | 0 | 0 | 0 | 1 | Ref |
| 1307 | JN204348 | Unassigned | Unassigned | Unassigned | 558_0 | Clustered | 2 | VC_558_0 | 2 | 0.0032 | 0.75858499 | 0.0025 | 0 | 0 | 0 | 1 | Ref |
| 1308 | JN225449 | Unassigned | Unassigned | Unassigned | 329_4 | Clustered | 2 | VC_329_4 | 2 | 0.0052 | 0.95746454 | 0.005 | 0 | 0 | 0 | 1 | Ref |
| 1309 | JN243855 | Unassigned | Unassigned | Unassigned | 238_0 | Clustered | 17 | VC_238_0 | 17 | 0.6109 | 1 | 0.6109 | 0 | 0 | 0 | 1 | Ref |
| 1310 | JN243856 | Unassigned | Unassigned | Unassigned | 517_1 | Clustered | 108 | VC_517_1 | 108 | 0.8483 | 1 | 0.8483 | 0 | 0 | 0 | 0.9955 | Ref |
| 1311 | JN243857 | Unassigned | Unassigned | Unassigned | 517_0 | Clustered | 18 | VC_517_0 | 18 | 0.1599 | 1 | 0.1599 | 0 | 0 | 0 | 1 | Ref |
| 1312 | JN254800 | Unassigned | Unassigned | Unassigned | 79_0 | Clustered | 64 | VC_79_0 | 64 | 0.4185 | 1 | 0.4185 | 0 | 0 | 0 | 0.7649 | Ref |
| 1313 | JN254801 | Unassigned | Unassigned | Unassigned | 472_0 | Clustered | 47 | VC_472_0 | 47 | 0.9025 | 1 | 0.9025 | 0 | 0 | 0 | 1 | Ref |
| 1314 | JN255163 | Unassigned | Unassigned | Unassigned | 4_0 | Clustered | 7 | VC_4_0 | 7 | 0.7912 | 1 | 0.7912 | 0 | 0 | 0 | 1 | Ref |
| 1315 | JN256079 | Unassigned | Unassigned | Unassigned | 521_0 | Clustered | 37 | VC_521_0 | 37 | 0.912 | 1 | 0.912 | 0 | 0 | 0 | 1 | Ref |
| 1316 | JN371769 | Unassigned | Unassigned | Unassigned | 655_4 | Clustered/Singleton | 1 | VC_655_4 | 1 | 0 | 1 | 0 | 0 | 0 | 0 | 1 | Ref |
| 1317 | JN377894 | Unassigned | Unassigned | Unassigned | 600_2 | Clustered | 10 | VC_600_2 | 10 | 0.3041 | 1 | 0.3041 | 0 | 0 | 0 | 1 | Ref |
| 1318 | JN377895 | Unassigned | Unassigned | Unassigned | 600_2 | Clustered | 10 | VC_600_2 | 10 | 0.3041 | 1 | 0.3041 | 0 | 0 | 0 | 1 | Ref |
| 1319 | JN382248 | Unassigned | Unassigned | Unassigned | 551_7 | Clustered | 28 | VC_551_7 | 28 | 0.1661 | 1 | 0.1661 | 0 | 0 | 0 | 0.8519 | Ref |
| 1320 | JN391180 | Unassigned | Unassigned | Unassigned | 476_0 | Clustered | 19 | VC_476_0 | 19 | 0.7814 | 1 | 0.7814 | 0 | 0 | 0 | 1 | Ref |
| 1321 | JN391441 | Unassigned | Unassigned | Unassigned | 551_7 | Clustered | 28 | VC_551_7 | 28 | 0.1661 | 1 | 0.1661 | 0 | 0 | 0 | 0.8519 | Ref |
| 1322 | JN398368 | Unassigned | Unassigned | Unassigned | 520_76 | Clustered | 2 | VC_520_76 | 2 | 0.0092 | 0.95761376 | 0.0088 | 0 | 0 | 0 | 1 | Ref |
| 1323 | JN398369 | Unassigned | Unassigned | Unassigned | 325_1 | Clustered | 2 | VC_325_1 | 2 | 0.0043 | 0.95663348 | 0.0041 | 0 | 0 | 0 | 1 | Ref |
| 1324 | JN408459 | Unassigned | Unassigned | Unassigned | 940_0 | Clustered | 34 | VC_940_0 | 34 | 0.7959 | 1 | 0.7959 | 0 | 0 | 0 | 1 | Ref |
| 1325 | JN408460 | Unassigned | Unassigned | Unassigned | 216_6 | Clustered | 42 | VC_216_6 | 42 | 0.473 | 1 | 0.473 | 0 | 0 | 0 | 0.8351 | Ref |
| 1326 | JN408461 | Unassigned | Unassigned | Unassigned | 216_3 | Clustered | 8 | VC_216_3 | 8 | 0.0701 | 1 | 0.0701 | 0 | 0 | 0 | 0.8571 | Ref |
| 1327 | JN412588 | Unassigned | Unassigned | Unassigned | 558_80 | Clustered/Singleton | 1 | VC_558_80 | 1 | 0 | 1 | 0 | 0 | 0 | 0 | 1 | Ref |
| 1328 | JN412589 | Unassigned | Unassigned | Unassigned | 569_0 | Clustered | 3 | VC_569_0 | 3 | 0.2094 | 0.99849867 | 0.2091 | 0 | 0 | 0 | 1 | Ref |
| 1329 | JN412590 | Unassigned | Unassigned | Unassigned | 551_4 | Clustered | 2 | VC_551_4 | 2 | 0.006 | 0.95695857 | 0.0058 | 0 | 0 | 0 | 1 | Ref |
| 1330 | JN412591 | Unassigned | Unassigned | Unassigned | 519_0 | Clustered | 39 | VC_519_0 | 39 | 0.8156 | 1 | 0.8156 | 0 | 0 | 0 | 1 | Ref |
| 1331 | JN412592 | Unassigned | Unassigned | Unassigned | 558_0 | Clustered | 2 | VC_558_0 | 2 | 0.0032 | 0.75858499 | 0.0025 | 0 | 0 | 0 | 1 | Ref |
| 1332 | JN412593 | Unassigned | Unassigned | Unassigned | 761_0 | Clustered | 67 | VC_761_0 | 67 | 0.8947 | 1 | 0.8947 | 0 | 0 | 0 | 1 | Ref |
| 1333 | JN542517 | Unassigned | Unassigned | Unassigned | 520_59 | Clustered | 5 | VC_520_59 | 5 | 0.0459 | 0.99999997 | 0.0459 | 0 | 0 | 0 | 1 | Ref |
| 1334 | JN561150 | Unassigned | Unassigned | Unassigned | 517_1 | Clustered | 108 | VC_517_1 | 108 | 0.8483 | 1 | 0.8483 | 0 | 0 | 0 | 0.9955 | Ref |
| 1335 | JN572061 | Unassigned | Unassigned | Unassigned | 519_0 | Clustered | 39 | VC_519_0 | 39 | 0.8156 | 1 | 0.8156 | 0 | 0 | 0 | 1 | Ref |
| 1336 | JN572689 | Unassigned | Unassigned | Unassigned | 325_35 | Clustered | 17 | VC_325_35 | 17 | 0.0732 | 1 | 0.0732 | 0 | 0 | 0 | 0.7721 | Ref |
| 1337 | JN585957 | Unassigned | Unassigned | Unassigned | 359_0 | Clustered | 7 | VC_359_0 | 7 | 0.3703 | 1 | 0.3703 | 0 | 0 | 0 | 1 | Ref |
| 1338 | JN593240 | Unassigned | Unassigned | Unassigned | 133_25 | Clustered/Singleton | 1 | VC_133_25 | 1 | 0 | 1 | 0 | 0 | 0 | 0 | 1 | Ref |
| 1339 | JN600672 | Unassigned | Unassigned | Unassigned | 887_0 | Clustered | 18 | VC_887_0 | 18 | 0.3663 | 1 | 0.3663 | 0 | 0 | 0 | 1 | Ref |
| 1340 | JN600960 | Unassigned | Unassigned | Unassigned | 30_1 | Clustered | 16 | VC_30_1 | 16 | 0.1973 | 1 | 0.1973 | 0 | 0 | 0 | 0.8333 | Ref |
| 1341 | JN618996 | Unassigned | Unassigned | Unassigned | 508_0 | Clustered | 32 | VC_508_0 | 32 | 0.906 | 1 | 0.906 | 0 | 0 | 0 | 1 | Ref |
| 1342 | JN624850 | Unassigned | Unassigned | Unassigned | 558_49 | Clustered | 7 | VC_558_49 | 7 | 0.0194 | 0.99510369 | 0.0193 | 0 | 0 | 0 | 0.8571 | Ref |
| 1343 | JN624851 | Unassigned | Unassigned | Unassigned | 521_0 | Clustered | 37 | VC_521_0 | 37 | 0.912 | 1 | 0.912 | 0 | 0 | 0 | 1 | Ref |
| 1344 | JN627160 | Unassigned | Unassigned | Unassigned | 442_2 | Clustered | 4 | VC_442_2 | 4 | 0.0844 | 0.99988129 | 0.0844 | 0 | 0 | 0 | 0.8333 | Ref |
| 1345 | JN638752 | Unassigned | Unassigned | Unassigned | 371_15 | Clustered | 44 | VC_371_15 | 44 | 0.1391 | 1 | 0.1391 | 0 | 0 | 0 | 0.7474 | Ref |
| 1346 | JN638753 | Unassigned | Unassigned | Unassigned | 371_15 | Clustered | 44 | VC_371_15 | 44 | 0.1391 | 1 | 0.1391 | 0 | 0 | 0 | 0.7474 | Ref |
| 1347 | JN641803 | Unassigned | Unassigned | Unassigned | 959_5 | Clustered | 19 | VC_959_5 | 19 | 0.2626 | 1 | 0.2626 | 0 | 0 | 0 | 1 | Ref |
| 1348 | JN643714 | Unassigned | Unassigned | Unassigned | 236_0 | Clustered | 72 | VC_236_0 | 72 | 0.8104 | 1 | 0.8104 | 0 | 0 | 0 | 1 | Ref |
| 1349 | JN651747 | Unassigned | Unassigned | Unassigned | 878_0 | Clustered | 32 | VC_878_0 | 32 | 0.9965 | 1 | 0.9965 | 0 | 0 | 0 | 1 | Ref |
| 1350 | JN654439 | Unassigned | Unassigned | Unassigned | 884_1 | Clustered | 8 | VC_884_1 | 8 | 0.1102 | 0.9989323 | 0.1101 | 0 | 0 | 0 | 0.75 | Ref |
| 1351 | JN660814 | Unassigned | Unassigned | Unassigned | 325_44 | Clustered | 3 | VC_325_44 | 3 | 0.0157 | 0.99859171 | 0.0157 | 0 | 0 | 0 | 1 | Ref |
| 1352 | JN662425 | Unassigned | Unassigned | Unassigned | 162_0 | Clustered | 18 | VC_162_0 | 18 | 0.7403 | 1 | 0.7403 | 0 | 0 | 0 | 0.8627 | Ref |
| 1353 | JN672684 | Unassigned | Unassigned | Unassigned | 937_0 | Clustered | 40 | VC_937_0 | 40 | 0.6809 | 1 | 0.6809 | 0 | 0 | 0 | 0.9359 | Ref |
| 1354 | JN673056 | Unassigned | Unassigned | Unassigned | 133_26 | Clustered/Singleton | 1 | VC_133_26 | 1 | 0 | 1 | 0 | 0 | 0 | 0 | 1 | Ref |
| 1355 | JN680858 | Unassigned | Unassigned | Unassigned | 888_0 | Clustered | 28 | VC_888_0 | 28 | 0.5497 | 1 | 0.5497 | 0 | 0 | 0 | 1 | Ref |
| 1356 | JN687951 | Unassigned | Unassigned | Unassigned | 325_60 | Clustered | 2 | VC_325_60 | 2 | 0.0038 | 0.957513 | 0.0036 | 0 | 0 | 0 | 1 | Ref |
| 1357 | JN698989 | Unassigned | Unassigned | Unassigned | 371_18 | Clustered | 10 | VC_371_18 | 10 | 0.0302 | 1 | 0.0302 | 0 | 0 | 0 | 0.8 | Ref |
| 1358 | JN698990 | Unassigned | Unassigned | Unassigned | 371_17 | Clustered | 3 | VC_371_17 | 3 | 0.0076 | 0.99811676 | 0.0076 | 0 | 0 | 0 | 1 | Ref |
| 1359 | JN698991 | Unassigned | Unassigned | Unassigned | 508_0 | Clustered | 32 | VC_508_0 | 32 | 0.906 | 1 | 0.906 | 0 | 0 | 0 | 1 | Ref |
| 1360 | JN698992 | Unassigned | Unassigned | Unassigned | 509_0 | Clustered | 37 | VC_509_0 | 37 | 0.9689 | 1 | 0.9689 | 0 | 0 | 0 | 1 | Ref |
| 1361 | JN698993 | Unassigned | Unassigned | Unassigned | 556_0 | Clustered | 21 | VC_556_0 | 21 | 0.9912 | 1 | 0.9912 | 0 | 0 | 0 | 1 | Ref |
| 1362 | JN698994 | Unassigned | Unassigned | Unassigned | 554_0 | Clustered | 2 | VC_554_0 | 2 | 0.2292 | 0.95193785 | 0.2181 | 0 | 0 | 0 | 1 | Ref |
| 1363 | JN698995 | Unassigned | Unassigned | Unassigned | 511_0 | Clustered | 3 | VC_511_0 | 3 | 0.1104 | 0.99858683 | 0.1103 | 0 | 0 | 0 | 1 | Ref |
| 1364 | JN698996 | Unassigned | Unassigned | Unassigned | 520_26 | Clustered | 3 | VC_520_26 | 3 | 0.026 | 0.99859898 | 0.026 | 0 | 0 | 0 | 1 | Ref |
| 1365 | JN698997 | Unassigned | Unassigned | Unassigned | 546_2 | Clustered | 3 | VC_546_2 | 3 | 0.0275 | 0.85260342 | 0.0234 | 0 | 0 | 0 | 1 | Ref |
| 1366 | JN698998 | Unassigned | Unassigned | Unassigned | 325_61 | Clustered | 8 | VC_325_61 | 8 | 0.0395 | 1 | 0.0395 | 0 | 0 | 0 | 0.7857 | Ref |
| 1367 | JN698999 | Unassigned | Unassigned | Unassigned | 223_0 | Clustered | 38 | VC_223_0 | 38 | 0.9095 | 1 | 0.9095 | 0 | 0 | 0 | 1 | Ref |
| 1368 | JN699000 | Unassigned | Unassigned | Unassigned | 325_47 | Clustered | 2 | VC_325_47 | 2 | 0.0045 | 0.9575092 | 0.0043 | 0 | 0 | 0 | 1 | Ref |
| 1369 | JN699001 | Unassigned | Unassigned | Unassigned | 518_0 | Clustered | 6 | VC_518_0 | 6 | 0.1295 | 1 | 0.1295 | 0 | 0 | 0 | 1 | Ref |
| 1370 | JN699002 | Unassigned | Unassigned | Unassigned | 761_0 | Clustered | 67 | VC_761_0 | 67 | 0.8947 | 1 | 0.8947 | 0 | 0 | 0 | 1 | Ref |
| 1371 | JN699003 | Unassigned | Unassigned | Unassigned | 509_0 | Clustered | 37 | VC_509_0 | 37 | 0.9689 | 1 | 0.9689 | 0 | 0 | 0 | 1 | Ref |
| 1372 | JN699004 | Unassigned | Unassigned | Unassigned | 508_0 | Clustered | 32 | VC_508_0 | 32 | 0.906 | 1 | 0.906 | 0 | 0 | 0 | 1 | Ref |
| 1373 | JN699005 | Unassigned | Unassigned | Unassigned | 225_0 | Clustered | 32 | VC_225_0 | 32 | 0.848 | 1 | 0.848 | 0 | 0 | 0 | 0.996 | Ref |
| 1374 | JN699006 | Unassigned | Unassigned | Unassigned | 509_0 | Clustered | 37 | VC_509_0 | 37 | 0.9689 | 1 | 0.9689 | 0 | 0 | 0 | 1 | Ref |
| 1375 | JN699007 | Unassigned | Unassigned | Unassigned | 372_1 | Clustered | 9 | VC_372_1 | 9 | 0.194 | 1 | 0.194 | 0 | 0 | 0 | 1 | Ref |
| 1376 | JN699008 | Unassigned | Unassigned | Unassigned | 371_15 | Clustered | 44 | VC_371_15 | 44 | 0.1391 | 1 | 0.1391 | 0 | 0 | 0 | 0.7474 | Ref |
| 1377 | JN699009 | Unassigned | Unassigned | Unassigned | 371_16 | Clustered | 42 | VC_371_16 | 42 | 0.1341 | 1 | 0.1341 | 0 | 0 | 0 | 0.7282 | Ref |
| 1378 | JN699010 | Unassigned | Unassigned | Unassigned | 371_15 | Clustered | 44 | VC_371_15 | 44 | 0.1391 | 1 | 0.1391 | 0 | 0 | 0 | 0.7474 | Ref |
| 1379 | JN699011 | Unassigned | Unassigned | Unassigned | 372_0 | Clustered | 24 | VC_372_0 | 24 | 0.5539 | 1 | 0.5539 | 0 | 0 | 0 | 1 | Ref |
| 1380 | JN699012 | Unassigned | Unassigned | Unassigned | 520_20 | Clustered/Singleton | 1 | VC_520_20 | 1 | 0 | 1 | 0 | 0 | 0 | 0 | 1 | Ref |
| 1381 | JN699013 | Unassigned | Unassigned | Unassigned | 558_8 | Clustered | 2 | VC_558_8 | 2 | 0.0032 | 0.68030512 | 0.0022 | 0 | 0 | 0 | 1 | Ref |
| 1382 | JN699014 | Unassigned | Unassigned | Unassigned | 565_0 | Clustered | 20 | VC_565_0 | 20 | 0.8398 | 1 | 0.8398 | 0 | 0 | 0 | 1 | Ref |
| 1383 | JN699015 | Unassigned | Unassigned | Unassigned | 517_1 | Clustered | 108 | VC_517_1 | 108 | 0.8483 | 1 | 0.8483 | 0 | 0 | 0 | 0.9955 | Ref |
| 1384 | JN699016 | Unassigned | Unassigned | Unassigned | 325_42 | Clustered | 4 | VC_325_42 | 4 | 0.0112 | 0.99980442 | 0.0112 | 0 | 0 | 0 | 0.6667 | Ref |
| 1385 | JN699017 | Unassigned | Unassigned | Unassigned | 371_17 | Clustered | 3 | VC_371_17 | 3 | 0.0076 | 0.99811676 | 0.0076 | 0 | 0 | 0 | 1 | Ref |
| 1386 | JN699018 | Unassigned | Unassigned | Unassigned | 509_0 | Clustered | 37 | VC_509_0 | 37 | 0.9689 | 1 | 0.9689 | 0 | 0 | 0 | 1 | Ref |
| 1387 | JN699019 | Unassigned | Unassigned | Unassigned | 223_0 | Clustered | 38 | VC_223_0 | 38 | 0.9095 | 1 | 0.9095 | 0 | 0 | 0 | 1 | Ref |
| 1388 | JN699625 | Unassigned | Unassigned | Unassigned | 558_30 | Clustered | 3 | VC_558_30 | 3 | 0.0064 | 0.76574053 | 0.0049 | 0 | 0 | 0 | 1 | Ref |
| 1389 | JN699626 | Unassigned | Unassigned | Unassigned | 558_63 | Clustered/Singleton | 1 | VC_558_63 | 1 | 0 | 1 | 0 | 0 | 0 | 0 | 1 | Ref |
| 1390 | JN699627 | Unassigned | Unassigned | Unassigned | 558_78 | Clustered/Singleton | 1 | VC_558_78 | 1 | 0 | 1 | 0 | 0 | 0 | 0 | 1 | Ref |
| 1391 | JN699628 | Unassigned | Unassigned | Unassigned | 763_1 | Clustered | 9 | VC_763_1 | 9 | 0.3706 | 1 | 0.3706 | 0 | 0 | 0 | 1 | Ref |
| 1392 | JN712910 | Unassigned | Unassigned | Unassigned | 960_0 | Clustered/Singleton | 1 | VC_960_0 | 1 | 0 | 1 | 0 | 0 | 0 | 0 | 1 | Ref |
| 1393 | JN790865 | Unassigned | Unassigned | Unassigned | 883_2 | Clustered | 2 | VC_883_2 | 2 | 0.0181 | 0.88094852 | 0.016 | 0 | 0 | 0 | 1 | Ref |
| 1394 | JN797796 | Unassigned | Unassigned | Unassigned | 883_2 | Clustered | 2 | VC_883_2 | 2 | 0.0181 | 0.88094852 | 0.016 | 0 | 0 | 0 | 1 | Ref |
| 1395 | JN797797 | Unassigned | Unassigned | Unassigned | 187_1 | Clustered | 7 | VC_187_1 | 7 | 0.1423 | 1 | 0.1423 | 0 | 0 | 0 | 1 | Ref |
| 1396 | JN797798 | Unassigned | Unassigned | Unassigned | 187_1 | Clustered | 7 | VC_187_1 | 7 | 0.1423 | 1 | 0.1423 | 0 | 0 | 0 | 1 | Ref |
| 1397 | JN800508 | Unassigned | Unassigned | Unassigned | 897_0 | Clustered | 6 | VC_897_0 | 6 | 0.4156 | 1 | 0.4156 | 0 | 0 | 0 | 1 | Ref |
| 1398 | JN808773 | Unassigned | Unassigned | Unassigned | 179_0 | Clustered | 53 | VC_179_0 | 53 | 0.9137 | 1 | 0.9137 | 0 | 0 | 0 | 1 | Ref |
| 1399 | JN811560 | Unassigned | Unassigned | Unassigned | 179_0 | Clustered | 53 | VC_179_0 | 53 | 0.9137 | 1 | 0.9137 | 0 | 0 | 0 | 1 | Ref |
| 1400 | JN831653 | Unassigned | Unassigned | Unassigned | 234_0 | Clustered | 21 | VC_234_0 | 21 | 0.8355 | 1 | 0.8355 | 0 | 0 | 0 | 1 | Ref |
| 1401 | JN848801 | Unassigned | Unassigned | Unassigned | 26_0 | Clustered | 4 | VC_26_0 | 4 | 0.7433 | 0.99936153 | 0.7428 | 0 | 0 | 0 | 1 | Ref |
| 1402 | JN849462 | Unassigned | Unassigned | Unassigned | 531_1 | Clustered | 6 | VC_531_1 | 6 | 0.1481 | 0.99978665 | 0.148 | 0 | 0 | 0 | 1 | Ref |
| 1403 | JN859129 | Unassigned | Unassigned | Unassigned | 520_23 | Clustered | 2 | VC_520_23 | 2 | 0.0077 | 0.95763408 | 0.0074 | 0 | 0 | 0 | 1 | Ref |
| 1404 | JN871591 | Unassigned | Unassigned | Unassigned | 592_0 | Clustered | 50 | VC_592_0 | 50 | 0.7314 | 1 | 0.7314 | 0 | 0 | 0 | 1 | Ref |
| 1405 | JN882264 | Unassigned | Unassigned | Unassigned | 834_0 | Clustered | 7 | VC_834_0 | 7 | 0.9845 | 0.94276893 | 0.9282 | 0 | 0 | 0 | 1 | Ref |
| 1406 | JN882266 | Unassigned | Unassigned | Unassigned | 834_0 | Clustered | 7 | VC_834_0 | 7 | 0.9845 | 0.94276893 | 0.9282 | 0 | 0 | 0 | 1 | Ref |
| 1407 | JN882267 | Unassigned | Unassigned | Unassigned | 962_0 | Clustered | 2 | VC_962_0 | 2 | 0.3572 | 0.92263554 | 0.3295 | 0 | 0 | 0 | 1 | Ref |
| 1408 | JN882284 | Unassigned | Unassigned | Unassigned | 582_4 | Clustered | 3 | VC_582_4 | 3 | 0.0348 | 0.98173503 | 0.0341 | 0 | 0 | 0 | 1 | Ref |
| 1409 | JN882285 | Unassigned | Unassigned | Unassigned | 164_4 | Clustered/Singleton | 1 | VC_164_4 | 1 | 0 | 1 | 0 | 0 | 0 | 0 | 1 | Ref |
| 1410 | JN882286 | Unassigned | Unassigned | Unassigned | 812_2 | Clustered | 2 | VC_812_2 | 2 | 0.0668 | 0.95518138 | 0.0639 | 0 | 0 | 0 | 1 | Ref |
| 1411 | JN882287 | Unassigned | Unassigned | Unassigned | 669_0 | Clustered | 21 | VC_669_0 | 21 | 0.3623 | 1 | 0.3623 | 0 | 0 | 0 | 1 | Ref |
| 1412 | JN882298 | Unassigned | Unassigned | Unassigned | 72_0 | Clustered | 2 | VC_72_0 | 2 | 0.0777 | 0.95616461 | 0.0743 | 0 | 0 | 0 | 1 | Ref |
| 1413 | JN939331 | Unassigned | Unassigned | Unassigned | 879_0 | Clustered | 36 | VC_879_0 | 36 | 0.994 | 1 | 0.994 | 0 | 0 | 0 | 1 | Ref |
| 1414 | JN939332 | Unassigned | Unassigned | Unassigned | 879_0 | Clustered | 36 | VC_879_0 | 36 | 0.994 | 1 | 0.994 | 0 | 0 | 0 | 1 | Ref |
| 1415 | JN968479 | Unassigned | Unassigned | Unassigned | 666_0 | Clustered | 4 | VC_666_0 | 4 | 1 | 1 | 1 | 0 | 0 | 0 | 1 | Ref |
| 1416 | JN984867 | Unassigned | Unassigned | Unassigned | 835_0 | Clustered | 67 | VC_835_0 | 67 | 0.9624 | 1 | 0.9624 | 0 | 0 | 0 | 1 | Ref |
| 1417 | JN986844 | Unassigned | Unassigned | Unassigned | 452_0 | Clustered | 66 | VC_452_0 | 66 | 0.9248 | 1 | 0.9248 | 0 | 0 | 0 | 1 | Ref |
| 1418 | JN986845 | Unassigned | Unassigned | Unassigned | 456_0 | Clustered | 62 | VC_456_0 | 62 | 0.9026 | 1 | 0.9026 | 0 | 0 | 0 | 1 | Ref |
| 1419 | JN986846 | Unassigned | Unassigned | Unassigned | 283_115 | Clustered/Singleton | 1 | VC_283_115 | 1 | 0 | 1 | 0 | 0 | 0 | 0 | 1 | Ref |
| 1420 | JQ007353 | Unassigned | Unassigned | Unassigned | 637_0 | Clustered | 115 | VC_637_0 | 115 | 0.731 | 1 | 0.731 | 0 | 0 | 0 | 0.8764 | Ref |
| 1421 | JQ010983 | Unassigned | Unassigned | Unassigned | 943_0 | Clustered | 2 | VC_943_0 | 2 | 1 | 1 | 1 | 0 | 0 | 0 | 1 | Ref |
| 1422 | JQ011317 | Unassigned | Unassigned | Unassigned | 476_0 | Clustered | 19 | VC_476_0 | 19 | 0.7814 | 1 | 0.7814 | 0 | 0 | 0 | 1 | Ref |
| 1423 | JQ011318 | Unassigned | Unassigned | Unassigned | 42_0 | Clustered | 49 | VC_42_0 | 49 | 0.4517 | 1 | 0.4517 | 0 | 0 | 0 | 0.8597 | Ref |
| 1424 | JQ015307 | Unassigned | Unassigned | Unassigned | 849_2 | Clustered | 2 | VC_849_2 | 2 | 0.0246 | 0.86513761 | 0.0213 | 0 | 0 | 0 | 1 | Ref |
| 1425 | JQ031132 | Unassigned | Unassigned | Unassigned | 772_4 | Clustered | 4 | VC_772_4 | 4 | 0.0181 | 0.73018303 | 0.0132 | 0 | 0 | 0 | 1 | Ref |
| 1426 | JQ062992 | Unassigned | Unassigned | Unassigned | 704_0 | Clustered | 16 | VC_704_0 | 16 | 0.7274 | 1 | 0.7274 | 0 | 0 | 0 | 1 | Ref |
| 1427 | JQ067084 | Unassigned | Unassigned | Unassigned | 465_0 | Clustered | 7 | VC_465_0 | 7 | 0.3322 | 1 | 0.3322 | 0 | 0 | 0 | 1 | Ref |
| 1428 | JQ067085 | Unassigned | Unassigned | Unassigned | 179_0 | Clustered | 53 | VC_179_0 | 53 | 0.9137 | 1 | 0.9137 | 0 | 0 | 0 | 1 | Ref |
| 1429 | JQ067087 | Unassigned | Unassigned | Unassigned | 464_0 | Clustered | 8 | VC_464_0 | 8 | 0.2629 | 1 | 0.2629 | 0 | 0 | 0 | 1 | Ref |
| 1430 | JQ067089 | Unassigned | Unassigned | Unassigned | 169_0 | Clustered | 20 | VC_169_0 | 20 | 0.7529 | 1 | 0.7529 | 0 | 0 | 0 | 1 | Ref |
| 1431 | JQ067092 | Unassigned | Unassigned | Unassigned | 749_0 | Clustered | 40 | VC_749_0 | 40 | 0.7517 | 1 | 0.7517 | 0 | 0 | 0 | 1 | Ref |
| 1432 | JQ067093 | Unassigned | Unassigned | Unassigned | 169_0 | Clustered | 20 | VC_169_0 | 20 | 0.7529 | 1 | 0.7529 | 0 | 0 | 0 | 1 | Ref |
| 1433 | JQ086375 | Unassigned | Unassigned | Unassigned | 835_0 | Clustered | 67 | VC_835_0 | 67 | 0.9624 | 1 | 0.9624 | 0 | 0 | 0 | 1 | Ref |
| 1434 | JQ086376 | Unassigned | Unassigned | Unassigned | 43_0 | Clustered | 38 | VC_43_0 | 38 | 0.5885 | 1 | 0.5885 | 0 | 0 | 0 | 0.9659 | Ref |
| 1435 | JQ177061 | Unassigned | Unassigned | Unassigned | 121_0 | Clustered | 4 | VC_121_0 | 4 | 0.1814 | 0.99998531 | 0.1814 | 0 | 0 | 0 | 1 | Ref |
| 1436 | JQ177063 | Unassigned | Unassigned | Unassigned | 369_0 | Clustered | 6 | VC_369_0 | 6 | 0.5924 | 1 | 0.5924 | 0 | 0 | 0 | 1 | Ref |
| 1437 | JQ177064 | Unassigned | Unassigned | Unassigned | 121_0 | Clustered | 4 | VC_121_0 | 4 | 0.1814 | 0.99998531 | 0.1814 | 0 | 0 | 0 | 1 | Ref |
| 1438 | JQ182728 | Unassigned | Unassigned | Unassigned | 51_0 | Clustered | 10 | VC_51_0 | 10 | 0.1741 | 1 | 0.1741 | 0 | 0 | 0 | 1 | Ref |
| 1439 | JQ182729 | Unassigned | Unassigned | Unassigned | 33_0 | Clustered | 4 | VC_33_0 | 4 | 0.06 | 0.99994826 | 0.06 | 0 | 0 | 0 | 1 | Ref |
| 1440 | JQ182733 | Unassigned | Unassigned | Unassigned | 30_0 | Clustered | 2 | VC_30_0 | 2 | 0.037 | 0.95734635 | 0.0354 | 0 | 0 | 0 | 1 | Ref |
| 1441 | JQ182734 | Unassigned | Unassigned | Unassigned | 30_0 | Clustered | 2 | VC_30_0 | 2 | 0.037 | 0.95734635 | 0.0354 | 0 | 0 | 0 | 1 | Ref |
| 1442 | JQ182735 | Unassigned | Unassigned | Unassigned | 43_0 | Clustered | 38 | VC_43_0 | 38 | 0.5885 | 1 | 0.5885 | 0 | 0 | 0 | 0.9659 | Ref |
| 1443 | JQ182736 | Unassigned | Unassigned | Unassigned | 455_0 | Clustered | 18 | VC_455_0 | 18 | 0.4598 | 1 | 0.4598 | 0 | 0 | 0 | 1 | Ref |
| 1444 | JQ245707 | Unassigned | Unassigned | Unassigned | 664_0 | Clustered | 4 | VC_664_0 | 4 | 0.263 | 0.99998747 | 0.263 | 0 | 0 | 0 | 1 | Ref |
| 1445 | JQ287645 | Unassigned | Unassigned | Unassigned | 444_0 | Clustered | 2 | VC_444_0 | 2 | 0.8137 | 0.72985431 | 0.5939 | 0 | 0 | 0 | 1 | Ref |
| 1446 | JQ300538 | Unassigned | Unassigned | Unassigned | 520_77 | Clustered | 3 | VC_520_77 | 3 | 0.0218 | 0.99859911 | 0.0218 | 0 | 0 | 0 | 1 | Ref |
| 1447 | JQ307386 | Unassigned | Unassigned | Unassigned | 423_0 | Clustered | 45 | VC_423_0 | 45 | 0.9484 | 1 | 0.9484 | 0 | 0 | 0 | 1 | Ref |
| 1448 | JQ307387 | Unassigned | Unassigned | Unassigned | 749_0 | Clustered | 40 | VC_749_0 | 40 | 0.7517 | 1 | 0.7517 | 0 | 0 | 0 | 1 | Ref |
| 1449 | JQ309827 | Unassigned | Unassigned | Unassigned | 967_0 | Clustered | 16 | VC_967_0 | 16 | 0.8971 | 1 | 0.8971 | 0 | 0 | 0 | 1 | Ref |
| 1450 | JQ312117 | Unassigned | Unassigned | Unassigned | 969_0 | Clustered | 3 | VC_969_0 | 3 | 0.8314 | 0.99762851 | 0.8295 | 0 | 0 | 0 | 1 | Ref |
| 1451 | JQ340389 | Unassigned | Unassigned | Unassigned | 971_1 | Clustered | 24 | VC_971_1 | 24 | 0.7393 | 1 | 0.7393 | 0 | 0 | 0 | 0.8913 | Ref |
| 1452 | JQ340774 | Unassigned | Unassigned | Unassigned | 592_0 | Clustered | 50 | VC_592_0 | 50 | 0.7314 | 1 | 0.7314 | 0 | 0 | 0 | 1 | Ref |
| 1453 | JQ347801 | Unassigned | Unassigned | Unassigned | 41_0 | Clustered | 16 | VC_41_0 | 16 | 0.2025 | 1 | 0.2025 | 0 | 0 | 0 | 0.9667 | Ref |
| 1454 | JQ408219 | Unassigned | Unassigned | Unassigned | 54_0 | Clustered | 6 | VC_54_0 | 6 | 0.6725 | 0.9999999 | 0.6725 | 0 | 0 | 0 | 1 | Ref |
| 1455 | JQ512844 | Unassigned | Unassigned | Unassigned | 515_0 | Clustered | 16 | VC_515_0 | 16 | 0.3793 | 1 | 0.3793 | 0 | 0 | 0 | 1 | Ref |
| 1456 | JQ513383 | Unassigned | Unassigned | Unassigned | 164_10 | Clustered | 5 | VC_164_10 | 5 | 0.0615 | 0.96483582 | 0.0593 | 0 | 0 | 0 | 1 | Ref |

| | | | | | | | | | | | | | | | | | |
|---|---|---|---|---|---|---|---|---|---|---|---|---|---|---|---|---|---|
| 1457 | JQ617284 | Unassigned | Unassigned | Unassigned | 99_0 | Clustered | 32 | VC_99_0 | 32 | 0.8828 | 1 | 0.8828 | 0 | 0 | 0 | 1 | Ref |
| 1458 | JQ619704 | Unassigned | Unassigned | Unassigned | 828_0 | Clustered | 3 | VC_828_0 | 3 | 0.4748 | 0.99843555 | 0.4741 | 0 | 0 | 0 | 1 | Ref |
| 1459 | JQ659259 | Unassigned | Unassigned | Unassigned | 861_0 | Clustered | 9 | VC_861_0 | 9 | 0.5869 | 1 | 0.5869 | 0 | 0 | 0 | 1 | Ref |
| 1460 | JQ684677 | Unassigned | Unassigned | Unassigned | 940_0 | Clustered | 34 | VC_940_0 | 34 | 0.7959 | 1 | 0.7959 | 0 | 0 | 0 | 1 | Ref |
| 1461 | JQ686190 | Unassigned | Unassigned | Unassigned | 149_15 | Clustered | 16 | VC_149_15 | 16 | 0.0788 | 1 | 0.0788 | 0 | 0 | 0 | 0.8083 | Ref |
| 1462 | JQ691610 | Unassigned | Unassigned | Unassigned | 476_0 | Clustered | 19 | VC_476_0 | 19 | 0.7814 | 1 | 0.7814 | 0 | 0 | 0 | 1 | Ref |
| 1463 | JQ691611 | Unassigned | Unassigned | Unassigned | 849_6 | Clustered | 7 | VC_849_6 | 7 | 0.164 | 0.99999999 | 0.164 | 0 | 0 | 0 | 0.9524 | Ref |
| 1464 | JQ691612 | Unassigned | Unassigned | Unassigned | 849_5 | Clustered | 3 | VC_849_5 | 3 | 0.0492 | 0.97574727 | 0.048 | 0 | 0 | 0 | 1 | Ref |
| 1465 | JQ692107 | Unassigned | Unassigned | Unassigned | 882_0 | Clustered | 20 | VC_882_0 | 20 | 0.9917 | 1 | 0.9917 | 0 | 0 | 0 | 1 | Ref |
| 1466 | JQ698665 | Unassigned | Unassigned | Unassigned | 325_35 | Clustered | 17 | VC_325_35 | 17 | 0.0732 | 1 | 0.0732 | 0 | 0 | 0 | 0.7721 | Ref |
| 1467 | JQ729990 | Unassigned | Unassigned | Unassigned | 898_0 | Clustered | 10 | VC_898_0 | 10 | 0.7284 | 1 | 0.7284 | 0 | 0 | 0 | 1 | Ref |
| 1468 | JQ729991 | Unassigned | Unassigned | Unassigned | 898_0 | Clustered | 10 | VC_898_0 | 10 | 0.7284 | 1 | 0.7284 | 0 | 0 | 0 | 1 | Ref |
| 1469 | JQ729992 | Unassigned | Unassigned | Unassigned | 898_0 | Clustered | 10 | VC_898_0 | 10 | 0.7284 | 1 | 0.7284 | 0 | 0 | 0 | 1 | Ref |
| 1470 | JQ762257 | Unassigned | Unassigned | Unassigned | 179_0 | Clustered | 53 | VC_179_0 | 53 | 0.9137 | 1 | 0.9137 | 0 | 0 | 0 | 1 | Ref |
| 1471 | JQ764986 | Unassigned | Unassigned | Unassigned | 303_0 | Clustered | 155 | VC_303_0 | 155 | 0.9979 | 1 | 0.9979 | 0 | 0 | 0 | 1 | Ref |
| 1472 | JQ764987 | Unassigned | Unassigned | Unassigned | 303_0 | Clustered | 155 | VC_303_0 | 155 | 0.9979 | 1 | 0.9979 | 0 | 0 | 0 | 1 | Ref |
| 1473 | JQ764988 | Unassigned | Unassigned | Unassigned | 303_0 | Clustered | 155 | VC_303_0 | 155 | 0.9979 | 1 | 0.9979 | 0 | 0 | 0 | 1 | Ref |
| 1474 | JQ764989 | Unassigned | Unassigned | Unassigned | 303_0 | Clustered | 155 | VC_303_0 | 155 | 0.9979 | 1 | 0.9979 | 0 | 0 | 0 | 1 | Ref |
| 1475 | JQ764990 | Unassigned | Unassigned | Unassigned | 303_0 | Clustered | 155 | VC_303_0 | 155 | 0.9979 | 1 | 0.9979 | 0 | 0 | 0 | 1 | Ref |
| 1476 | JQ779023 | Unassigned | Unassigned | Unassigned | 8_0 | Clustered | 58 | VC_8_0 | 58 | 0.8306 | 1 | 0.8306 | 0 | 0 | 0 | 0.9946 | Ref |
| 1477 | JQ779024 | Unassigned | Unassigned | Unassigned | 9_0 | Clustered | 38 | VC_9_0 | 38 | 0.4895 | 1 | 0.4895 | 0 | 0 | 0 | 1 | Ref |
| 1478 | JQ780163 | Unassigned | Unassigned | Unassigned | 82_0 | Clustered | 24 | VC_82_0 | 24 | 0.7351 | 1 | 0.7351 | 0 | 0 | 0 | 1 | Ref |
| 1479 | JQ780327 | Unassigned | Unassigned | Unassigned | 230_0 | Clustered | 3 | VC_230_0 | 3 | 0.0739 | 0.9934951 | 0.0734 | 0 | 0 | 0 | 0.6667 | Ref |
| 1480 | JQ797329 | Unassigned | Unassigned | Unassigned | 739_0 | Clustered | 26 | VC_739_0 | 26 | 0.9571 | 1 | 0.9571 | 0 | 0 | 0 | 1 | Ref |
| 1481 | JQ806763 | Unassigned | Unassigned | Unassigned | 28_1 | Clustered | 53 | VC_28_1 | 53 | 0.7164 | 1 | 0.7164 | 0 | 0 | 0 | 0.9325 | Ref |
| 1482 | JQ809650 | Unassigned | Unassigned | Unassigned | 304_0 | Clustered | 11 | VC_304_0 | 11 | 0.7869 | 1 | 0.7869 | 0 | 0 | 0 | 1 | Ref |
| 1483 | JQ809701 | Unassigned | Unassigned | Unassigned | 517_1 | Clustered | 108 | VC_517_1 | 108 | 0.8483 | 1 | 0.8483 | 0 | 0 | 0 | 0.9955 | Ref |
| 1484 | JQ809703 | Unassigned | Unassigned | Unassigned | 325_35 | Clustered | 17 | VC_325_35 | 17 | 0.0732 | 1 | 0.0732 | 0 | 0 | 0 | 0.7721 | Ref |
| 1485 | JQ823122 | Unassigned | Unassigned | Unassigned | 974_0 | Clustered | 2 | VC_974_0 | 2 | 1 | 1 | 1 | 0 | 0 | 0 | 1 | Ref |
| 1486 | JQ823123 | Unassigned | Unassigned | Unassigned | 974_0 | Clustered | 2 | VC_974_0 | 2 | 1 | 1 | 1 | 0 | 0 | 0 | 1 | Ref |
| 1487 | JQ837901 | Unassigned | Unassigned | Unassigned | 452_0 | Clustered | 66 | VC_452_0 | 66 | 0.9248 | 1 | 0.9248 | 0 | 0 | 0 | 1 | Ref |
| 1488 | JQ896627 | Unassigned | Unassigned | Unassigned | 517_1 | Clustered | 108 | VC_517_1 | 108 | 0.8483 | 1 | 0.8483 | 0 | 0 | 0 | 0.9955 | Ref |
| 1489 | JQ911768 | Unassigned | Unassigned | Unassigned | 558_50 | Clustered | 15 | VC_558_50 | 15 | 0.0468 | 1 | 0.0468 | 0 | 0 | 0 | 0.9238 | Ref |
| 1490 | JQ957925 | Unassigned | Unassigned | Unassigned | 392_0 | Clustered | 126 | VC_392_0 | 126 | 0.9621 | 1 | 0.9621 | 0 | 0 | 0 | 1 | Ref |
| 1491 | JQ957932 | Unassigned | Unassigned | Unassigned | 8_0 | Clustered | 58 | VC_8_0 | 58 | 0.8306 | 1 | 0.8306 | 0 | 0 | 0 | 0.9946 | Ref |
| 1492 | JQ965645 | Unassigned | Unassigned | Unassigned | 723_0 | Clustered | 9 | VC_723_0 | 9 | 0.7138 | 1 | 0.7138 | 0 | 0 | 0 | 1 | Ref |
| 1493 | JQ965700 | Unassigned | Unassigned | Unassigned | 392_0 | Clustered | 126 | VC_392_0 | 126 | 0.9621 | 1 | 0.9621 | 0 | 0 | 0 | 1 | Ref |
| 1494 | JQ965701 | Unassigned | Unassigned | Unassigned | 392_0 | Clustered | 126 | VC_392_0 | 126 | 0.9621 | 1 | 0.9621 | 0 | 0 | 0 | 1 | Ref |
| 1495 | JQ965702 | Unassigned | Unassigned | Unassigned | 386_0 | Clustered | 49 | VC_386_0 | 49 | 0.7941 | 1 | 0.7941 | 0 | 0 | 0 | 1 | Ref |
| 1496 | JQ965703 | Unassigned | Unassigned | Unassigned | 392_0 | Clustered | 126 | VC_392_0 | 126 | 0.9621 | 1 | 0.9621 | 0 | 0 | 0 | 1 | Ref |
| 1497 | JQ966307 | Unassigned | Unassigned | Unassigned | 315_0 | Clustered | 12 | VC_315_0 | 12 | 0.8947 | 1 | 0.8947 | 0 | 0 | 0 | 1 | Ref |
| 1498 | JQ973847 | Unassigned | Unassigned | Unassigned | 12_0 | Clustered | 21 | VC_12_0 | 21 | 0.2274 | 1 | 0.2274 | 0 | 0 | 0 | 0.9571 | Ref |
| 1499 | JX000007 | Unassigned | Unassigned | Unassigned | 392_0 | Clustered | 126 | VC_392_0 | 126 | 0.9621 | 1 | 0.9621 | 0 | 0 | 0 | 1 | Ref |
| 1500 | JX013863 | Unassigned | Unassigned | Unassigned | 12_2 | Clustered | 19 | VC_12_2 | 19 | 0.2018 | 1 | 0.2018 | 0 | 0 | 0 | 1 | Ref |
| 1501 | JX042578 | Unassigned | Unassigned | Unassigned | 940_0 | Clustered | 34 | VC_940_0 | 34 | 0.7959 | 1 | 0.7959 | 0 | 0 | 0 | 1 | Ref |
| 1502 | JX042579 | Unassigned | Unassigned | Unassigned | 237_1 | Clustered | 13 | VC_237_1 | 13 | 0.1773 | 1 | 0.1773 | 0 | 0 | 0 | 0.7308 | Ref |
| 1503 | JX080300 | Unassigned | Unassigned | Unassigned | 149_8 | Clustered | 66 | VC_149_8 | 66 | 0.4565 | 1 | 0.4565 | 0 | 0 | 0 | 0.8182 | Ref |
| 1504 | JX080301 | Unassigned | Unassigned | Unassigned | 149_8 | Clustered | 66 | VC_149_8 | 66 | 0.4565 | 1 | 0.4565 | 0 | 0 | 0 | 0.8182 | Ref |
| 1505 | JX080302 | Unassigned | Unassigned | Unassigned | 149_8 | Clustered | 66 | VC_149_8 | 66 | 0.4565 | 1 | 0.4565 | 0 | 0 | 0 | 0.8182 | Ref |
| 1506 | JX080303 | Unassigned | Unassigned | Unassigned | 149_8 | Clustered | 66 | VC_149_8 | 66 | 0.4565 | 1 | 0.4565 | 0 | 0 | 0 | 0.8182 | Ref |
| 1507 | JX080304 | Unassigned | Unassigned | Unassigned | 149_8 | Clustered | 66 | VC_149_8 | 66 | 0.4565 | 1 | 0.4565 | 0 | 0 | 0 | 0.8182 | Ref |
| 1508 | JX080305 | Unassigned | Unassigned | Unassigned | 149_8 | Clustered | 66 | VC_149_8 | 66 | 0.4565 | 1 | 0.4565 | 0 | 0 | 0 | 0.8182 | Ref |
| 1509 | JX094431 | Unassigned | Unassigned | Unassigned | 187_3 | Clustered | 9 | VC_187_3 | 9 | 0.1971 | 1 | 0.1971 | 0 | 0 | 0 | 0.8611 | Ref |
| 1510 | JX094499 | Unassigned | Unassigned | Unassigned | 592_0 | Clustered | 50 | VC_592_0 | 50 | 0.7314 | 1 | 0.7314 | 0 | 0 | 0 | 1 | Ref |
| 1511 | JX094500 | Unassigned | Unassigned | Unassigned | 959_0 | Clustered | 5 | VC_959_0 | 5 | 0.0499 | 0.99999231 | 0.0499 | 0 | 0 | 0 | 1 | Ref |
| 1512 | JX094501 | Unassigned | Unassigned | Unassigned | 12_2 | Clustered | 19 | VC_12_2 | 19 | 0.2018 | 1 | 0.2018 | 0 | 0 | 0 | 1 | Ref |
| 1513 | JX100809 | Unassigned | Unassigned | Unassigned | 976_4 | Clustered | 11 | VC_976_4 | 11 | 0.3364 | 1 | 0.3364 | 0 | 0 | 0 | 1 | Ref |
| 1514 | JX100810 | Unassigned | Unassigned | Unassigned | 976_2 | Clustered | 2 | VC_976_2 | 2 | 0.0278 | 0.95410794 | 0.0266 | 0 | 0 | 0 | 1 | Ref |
| 1515 | JX100811 | Unassigned | Unassigned | Unassigned | 976_4 | Clustered | 11 | VC_976_4 | 11 | 0.3364 | 1 | 0.3364 | 0 | 0 | 0 | 1 | Ref |
| 1516 | JX100812 | Unassigned | Unassigned | Unassigned | 976_4 | Clustered | 11 | VC_976_4 | 11 | 0.3364 | 1 | 0.3364 | 0 | 0 | 0 | 1 | Ref |
| 1517 | JX100813 | Unassigned | Unassigned | Unassigned | 976_4 | Clustered | 11 | VC_976_4 | 11 | 0.3364 | 1 | 0.3364 | 0 | 0 | 0 | 1 | Ref |
| 1518 | JX100814 | Unassigned | Unassigned | Unassigned | 976_3 | Clustered | 2 | VC_976_3 | 2 | 0.0258 | 0.88402892 | 0.0228 | 0 | 0 | 0 | 1 | Ref |
| 1519 | JX104231 | Unassigned | Unassigned | Unassigned | 162_0 | Clustered | 18 | VC_162_0 | 18 | 0.7403 | 1 | 0.7403 | 0 | 0 | 0 | 0.8627 | Ref |
| 1520 | JX120799 | Unassigned | Unassigned | Unassigned | 379_0 | Clustered | 5 | VC_379_0 | 5 | 0.4779 | 0.99999992 | 0.4779 | 0 | 0 | 0 | 0.9 | Ref |
| 1521 | JX121091 | Unassigned | Unassigned | Unassigned | 520_26 | Clustered | 3 | VC_520_26 | 3 | 0.026 | 0.99859898 | 0.026 | 0 | 0 | 0 | 1 | Ref |
| 1522 | JX123262 | Unassigned | Unassigned | Unassigned | 443_2 | Clustered | 10 | VC_443_2 | 10 | 0.3344 | 1 | 0.3344 | 0 | 0 | 0 | 0.9333 | Ref |
| 1523 | JX126918 | Unassigned | Unassigned | Unassigned | 739_0 | Clustered | 26 | VC_739_0 | 26 | 0.9571 | 1 | 0.9571 | 0 | 0 | 0 | 1 | Ref |
| 1524 | JX126919 | Unassigned | Unassigned | Unassigned | 980_0 | Clustered | 10 | VC_980_0 | 10 | 0.9996 | 1 | 0.9996 | 0 | 0 | 0 | 1 | Ref |
| 1525 | JX126920 | Unassigned | Unassigned | Unassigned | 980_0 | Clustered | 10 | VC_980_0 | 10 | 0.9996 | 1 | 0.9996 | 0 | 0 | 0 | 1 | Ref |
| 1526 | JX128257 | Unassigned | Unassigned | Unassigned | 133_22 | Clustered | 13 | VC_133_22 | 13 | 0.0804 | 1 | 0.0804 | 0 | 0 | 0 | 0.8205 | Ref |
| 1527 | JX128258 | Unassigned | Unassigned | Unassigned | 618_0 | Clustered | 13 | VC_618_0 | 13 | 0.6252 | 1 | 0.6252 | 0 | 0 | 0 | 1 | Ref |
| 1528 | JX128259 | Unassigned | Unassigned | Unassigned | 283_95 | Clustered/Singleton | 1 | VC_283_95 | 1 | 0 | 1 | 0 | 0 | 0 | 0 | 1 | Ref |
| 1529 | JX131330 | Unassigned | Unassigned | Unassigned | 166_0 | Clustered | 26 | VC_166_0 | 26 | 0.7923 | 1 | 0.7923 | 0 | 0 | 0 | 1 | Ref |
| 1530 | JX145342 | Unassigned | Unassigned | Unassigned | 647_0 | Clustered | 11 | VC_647_0 | 11 | 0.598 | 1 | 0.598 | 0 | 0 | 0 | 1 | Ref |
| 1531 | JX163858 | Unassigned | Unassigned | Unassigned | 976_4 | Clustered | 11 | VC_976_4 | 11 | 0.3364 | 1 | 0.3364 | 0 | 0 | 0 | 1 | Ref |
| 1532 | JX173487 | Unassigned | Unassigned | Unassigned | 130_0 | Clustered | 19 | VC_130_0 | 19 | 0.6068 | 1 | 0.6068 | 0 | 0 | 0 | 1 | Ref |
| 1533 | JX174275 | Unassigned | Unassigned | Unassigned | 8_0 | Clustered | 58 | VC_8_0 | 58 | 0.8306 | 1 | 0.8306 | 0 | 0 | 0 | 0.9946 | Ref |
| 1534 | JX181822 | Unassigned | Unassigned | Unassigned | 329_1 | Clustered | 17 | VC_329_1 | 17 | 0.0867 | 1 | 0.0867 | 0 | 0 | 0 | 0.8235 | Ref |
| 1535 | JX181824 | Unassigned | Unassigned | Unassigned | 582_5 | Clustered | 4 | VC_582_5 | 4 | 0.0528 | 0.99935625 | 0.0527 | 0 | 0 | 0 | 0.8333 | Ref |
| 1536 | JX181828 | Unassigned | Unassigned | Unassigned | 133_22 | Clustered | 13 | VC_133_22 | 13 | 0.0804 | 1 | 0.0804 | 0 | 0 | 0 | 0.8205 | Ref |
| 1537 | JX181829 | Unassigned | Unassigned | Unassigned | 131_2 | Clustered | 7 | VC_131_2 | 7 | 0.0685 | 1 | 0.0685 | 0 | 0 | 0 | 0.9048 | Ref |
| 1538 | JX182369 | Unassigned | Unassigned | Unassigned | 981_0 | Clustered | 3 | VC_981_0 | 3 | 0.0531 | 0.99853685 | 0.053 | 0 | 0 | 0 | 1 | Ref |
| 1539 | JX182370 | Unassigned | Unassigned | Unassigned | 981_1 | Clustered | 26 | VC_981_1 | 26 | 0.7009 | 1 | 0.7009 | 0 | 0 | 0 | 1 | Ref |
| 1540 | JX182371 | Unassigned | Unassigned | Unassigned | 727_0 | Clustered | 11 | VC_727_0 | 11 | 0.5054 | 1 | 0.5054 | 0 | 0 | 0 | 1 | Ref |
| 1541 | JX182372 | Unassigned | Unassigned | Unassigned | 373_0 | Clustered | 10 | VC_373_0 | 10 | 0.7471 | 1 | 0.7471 | 0 | 0 | 0 | 1 | Ref |
| 1542 | JX193904 | Unassigned | Unassigned | Unassigned | 695_0 | Clustered | 48 | VC_695_0 | 48 | 0.8162 | 1 | 0.8162 | 0 | 0 | 0 | 1 | Ref |
| 1543 | JX194238 | Unassigned | Unassigned | Unassigned | 838_0 | Clustered | 29 | VC_838_0 | 29 | 0.8611 | 1 | 0.8611 | 0 | 0 | 0 | 1 | Ref |
| 1544 | JX194239 | Unassigned | Unassigned | Unassigned | 153_0 | Clustered | 25 | VC_153_0 | 25 | 0.7729 | 1 | 0.7729 | 0 | 0 | 0 | 1 | Ref |
| 1545 | JX202565 | Unassigned | Unassigned | Unassigned | 637_0 | Clustered | 115 | VC_637_0 | 115 | 0.731 | 1 | 0.731 | 0 | 0 | 0 | 0.8764 | Ref |
| 1546 | JX233784 | Unassigned | Unassigned | Unassigned | 344_0 | Clustered | 21 | VC_344_0 | 21 | 0.5436 | 1 | 0.5436 | 0 | 0 | 0 | 1 | Ref |
| 1547 | JX238501 | Unassigned | Unassigned | Unassigned | 85_6 | Clustered/Singleton | 1 | VC_85_6 | 1 | 0 | 1 | 0 | 0 | 0 | 0 | 1 | Ref |
| 1548 | JX262215 | Unassigned | Unassigned | Unassigned | 765_0 | Clustered | 132 | VC_765_0 | 132 | 1 | 1 | 1 | 0 | 0 | 0 | 1 | Ref |
| 1549 | JX262216 | Unassigned | Unassigned | Unassigned | 765_0 | Clustered | 132 | VC_765_0 | 132 | 1 | 1 | 1 | 0 | 0 | 0 | 1 | Ref |
| 1550 | JX262217 | Unassigned | Unassigned | Unassigned | 765_0 | Clustered | 132 | VC_765_0 | 132 | 1 | 1 | 1 | 0 | 0 | 0 | 1 | Ref |
| 1551 | JX262218 | Unassigned | Unassigned | Unassigned | 765_0 | Clustered | 132 | VC_765_0 | 132 | 1 | 1 | 1 | 0 | 0 | 0 | 1 | Ref |
| 1552 | JX262219 | Unassigned | Unassigned | Unassigned | 765_0 | Clustered | 132 | VC_765_0 | 132 | 1 | 1 | 1 | 0 | 0 | 0 | 1 | Ref |
| 1553 | JX262220 | Unassigned | Unassigned | Unassigned | 765_0 | Clustered | 132 | VC_765_0 | 132 | 1 | 1 | 1 | 0 | 0 | 0 | 1 | Ref |
| 1554 | JX262221 | Unassigned | Unassigned | Unassigned | 765_0 | Clustered | 132 | VC_765_0 | 132 | 1 | 1 | 1 | 0 | 0 | 0 | 1 | Ref |
| 1555 | JX262222 | Unassigned | Unassigned | Unassigned | 765_0 | Clustered | 132 | VC_765_0 | 132 | 1 | 1 | 1 | 0 | 0 | 0 | 1 | Ref |
| 1556 | JX262223 | Unassigned | Unassigned | Unassigned | 765_0 | Clustered | 132 | VC_765_0 | 132 | 1 | 1 | 1 | 0 | 0 | 0 | 1 | Ref |
| 1557 | JX262224 | Unassigned | Unassigned | Unassigned | 765_0 | Clustered | 132 | VC_765_0 | 132 | 1 | 1 | 1 | 0 | 0 | 0 | 1 | Ref |
| 1558 | JX262225 | Unassigned | Unassigned | Unassigned | 765_0 | Clustered | 132 | VC_765_0 | 132 | 1 | 1 | 1 | 0 | 0 | 0 | 1 | Ref |
| 1559 | JX262376 | Unassigned | Unassigned | Unassigned | 981_1 | Clustered | 26 | VC_981_1 | 26 | 0.7009 | 1 | 0.7009 | 0 | 0 | 0 | 1 | Ref |
| 1560 | JX274646 | Unassigned | Unassigned | Unassigned | 9_0 | Clustered | 38 | VC_9_0 | 38 | 0.4895 | 1 | 0.4895 | 0 | 0 | 0 | 1 | Ref |
| 1561 | JX274647 | Unassigned | Unassigned | Unassigned | 9_0 | Clustered | 38 | VC_9_0 | 38 | 0.4895 | 1 | 0.4895 | 0 | 0 | 0 | 1 | Ref |
| 1562 | JX290549 | Unassigned | Unassigned | Unassigned | 984_0 | Clustered | 38 | VC_984_0 | 38 | 0.9758 | 1 | 0.9758 | 0 | 0 | 0 | 1 | Ref |
| 1563 | JX296113 | Unassigned | Unassigned | Unassigned | 592_0 | Clustered | 50 | VC_592_0 | 50 | 0.7314 | 1 | 0.7314 | 0 | 0 | 0 | 1 | Ref |
| 1564 | JX297445 | Unassigned | Unassigned | Unassigned | 637_1 | Clustered | 27 | VC_637_1 | 27 | 0.2578 | 1 | 0.2578 | 0 | 0 | 0 | 0.9858 | Ref |
| 1565 | JX306041 | Unassigned | Unassigned | Unassigned | 600_2 | Clustered | 10 | VC_600_2 | 10 | 0.3041 | 1 | 0.3041 | 0 | 0 | 0 | 1 | Ref |
| 1566 | JX307702 | Unassigned | Unassigned | Unassigned | 517_1 | Clustered | 108 | VC_517_1 | 108 | 0.8483 | 1 | 0.8483 | 0 | 0 | 0 | 0.9955 | Ref |
| 1567 | JX307703 | Unassigned | Unassigned | Unassigned | 517_1 | Clustered | 108 | VC_517_1 | 108 | 0.8483 | 1 | 0.8483 | 0 | 0 | 0 | 0.9955 | Ref |
| 1568 | JX307704 | Unassigned | Unassigned | Unassigned | 515_0 | Clustered | 16 | VC_515_0 | 16 | 0.3793 | 1 | 0.3793 | 0 | 0 | 0 | 1 | Ref |
| 1569 | JX307705 | Unassigned | Unassigned | Unassigned | 325_42 | Clustered | 4 | VC_325_42 | 4 | 0.0112 | 0.99980442 | 0.0112 | 0 | 0 | 0 | 0.6667 | Ref |
| 1570 | JX316028 | Unassigned | Unassigned | Unassigned | 959_4 | Clustered | 3 | VC_959_4 | 3 | 0.0237 | 0.77138817 | 0.0183 | 0 | 0 | 0 | 1 | Ref |
| 1571 | JX403939 | Unassigned | Unassigned | Unassigned | 297_1 | Clustered | 20 | VC_297_1 | 20 | 0.5653 | 1 | 0.5653 | 0 | 0 | 0 | 0.7684 | Ref |
| 1572 | JX403940 | Unassigned | Unassigned | Unassigned | 696_1 | Clustered | 8 | VC_696_1 | 8 | 0.5452 | 1 | 0.5452 | 0 | 0 | 0 | 0.9286 | Ref |
| 1573 | JX411619 | Unassigned | Unassigned | Unassigned | 515_0 | Clustered | 16 | VC_515_0 | 16 | 0.3793 | 1 | 0.3793 | 0 | 0 | 0 | 1 | Ref |
| 1574 | JX412914 | Unassigned | Unassigned | Unassigned | 61_0 | Clustered | 12 | VC_61_0 | 12 | 0.7633 | 1 | 0.7633 | 0 | 0 | 0 | 1 | Ref |
| 1575 | JX415535 | Unassigned | Unassigned | Unassigned | 779_0 | Clustered | 37 | VC_779_0 | 37 | 0.5682 | 1 | 0.5682 | 0 | 0 | 0 | 1 | Ref |
| 1576 | JX415536 | Unassigned | Unassigned | Unassigned | 813_0 | Clustered | 12 | VC_813_0 | 12 | 0.2347 | 1 | 0.2347 | 0 | 0 | 0 | 1 | Ref |
| 1577 | JX421753 | Unassigned | Unassigned | Unassigned | 385_0 | Clustered | 51 | VC_385_0 | 51 | 0.8754 | 1 | 0.8754 | 0 | 0 | 0 | 1 | Ref |
| 1578 | JX434030 | Unassigned | Unassigned | Unassigned | 179_0 | Clustered | 53 | VC_179_0 | 53 | 0.9137 | 1 | 0.9137 | 0 | 0 | 0 | 1 | Ref |
| 1579 | JX434031 | Unassigned | Unassigned | Unassigned | 179_0 | Clustered | 53 | VC_179_0 | 53 | 0.9137 | 1 | 0.9137 | 0 | 0 | 0 | 1 | Ref |
| 1580 | JX434032 | Unassigned | Unassigned | Unassigned | 179_0 | Clustered | 53 | VC_179_0 | 53 | 0.9137 | 1 | 0.9137 | 0 | 0 | 0 | 1 | Ref |
| 1581 | JX434033 | Unassigned | Unassigned | Unassigned | 179_0 | Clustered | 53 | VC_179_0 | 53 | 0.9137 | 1 | 0.9137 | 0 | 0 | 0 | 1 | Ref |
| 1582 | JX442241 | Unassigned | Unassigned | Unassigned | 980_0 | Clustered | 10 | VC_980_0 | 10 | 0.9996 | 1 | 0.9996 | 0 | 0 | 0 | 1 | Ref |
| 1583 | JX483873 | Unassigned | Unassigned | Unassigned | 481_0 | Clustered | 8 | VC_481_0 | 8 | 0.1348 | 1 | 0.1348 | 0 | 0 | 0 | 1 | Ref |
| 1584 | JX483874 | Unassigned | Unassigned | Unassigned | 987_0 | Clustered | 22 | VC_987_0 | 22 | 0.8955 | 1 | 0.8955 | 0 | 0 | 0 | 1 | Ref |
| 1585 | JX483875 | Unassigned | Unassigned | Unassigned | 987_0 | Clustered | 22 | VC_987_0 | 22 | 0.8955 | 1 | 0.8955 | 0 | 0 | 0 | 1 | Ref |
| 1586 | JX483876 | Unassigned | Unassigned | Unassigned | 946_0 | Clustered | 11 | VC_946_0 | 11 | 0.7352 | 1 | 0.7352 | 0 | 0 | 0 | 1 | Ref |
| 1587 | JX483877 | Unassigned | Unassigned | Unassigned | 946_0 | Clustered | 11 | VC_946_0 | 11 | 0.7352 | 1 | 0.7352 | 0 | 0 | 0 | 1 | Ref |
| 1588 | JX483878 | Unassigned | Unassigned | Unassigned | 946_0 | Clustered | 11 | VC_946_0 | 11 | 0.7352 | 1 | 0.7352 | 0 | 0 | 0 | 1 | Ref |
| 1589 | JX483879 | Unassigned | Unassigned | Unassigned | 987_0 | Clustered | 22 | VC_987_0 | 22 | 0.8955 | 1 | 0.8955 | 0 | 0 | 0 | 1 | Ref |
| 1590 | JX483880 | Unassigned | Unassigned | Unassigned | 987_0 | Clustered | 22 | VC_987_0 | 22 | 0.8955 | 1 | 0.8955 | 0 | 0 | 0 | 1 | Ref |
| 1591 | JX483881 | Unassigned | Unassigned | Unassigned | 724_1 | Clustered | 6 | VC_724_1 | 6 | 0.4379 | 1 | 0.4379 | 0 | 0 | 0 | 1 | Ref |
| 1592 | JX486087 | Unassigned | Unassigned | Unassigned | 577_0 | Clustered | 6 | VC_577_0 | 6 | 0.9267 | 0.99999993 | 0.9267 | 0 | 0 | 0 | 1 | Ref |
| 1593 | JX486088 | Unassigned | Unassigned | Unassigned | 634_1 | Clustered | 6 | VC_634_1 | 6 | 0.4228 | 1 | 0.4228 | 0 | 0 | 0 | 1 | Ref |
| 1594 | JX495041 | Unassigned | Unassigned | Unassigned | 251_0 | Clustered | 22 | VC_251_0 | 22 | 0.6508 | 1 | 0.6508 | 0 | 0 | 0 | 1 | Ref |
| 1595 | JX495042 | Unassigned | Unassigned | Unassigned | 251_0 | Clustered | 22 | VC_251_0 | 22 | 0.6508 | 1 | 0.6508 | 0 | 0 | 0 | 1 | Ref |
| 1596 | JX495043 | Unassigned | Unassigned | Unassigned | 251_0 | Clustered | 22 | VC_251_0 | 22 | 0.6508 | 1 | 0.6508 | 0 | 0 | 0 | 1 | Ref |
| 1597 | JX501340 | Unassigned | Unassigned | Unassigned | 423_0 | Clustered | 45 | VC_423_0 | 45 | 0.9484 | 1 | 0.9484 | 0 | 0 | 0 | 1 | Ref |
| 1598 | JX507079 | Unassigned | Unassigned | Unassigned | 24_0 | Clustered | 3 | VC_24_0 | 3 | 0.3455 | 0.99844825 | 0.345 | 0 | 0 | 0 | 1 | Ref |
| 1599 | JX536493 | Unassigned | Unassigned | Unassigned | 286_14 | Clustered/Singleton | 1 | VC_286_14 | 1 | 0 | 1 | 0 | 0 | 0 | 0 | 1 | Ref |
| 1600 | JX556417 | Unassigned | Unassigned | Unassigned | 23_0 | Clustered | 3 | VC_23_0 | 3 | 0.4305 | 0.99837378 | 0.4298 | 0 | 0 | 0 | 1 | Ref |
| 1601 | JX556418 | Unassigned | Unassigned | Unassigned | 882_0 | Clustered | 20 | VC_882_0 | 20 | 0.9917 | 1 | 0.9917 | 0 | 0 | 0 | 1 | Ref |
| 1602 | JX560968 | Unassigned | Unassigned | Unassigned | 329_18 | Clustered | 7 | VC_329_18 | 7 | 0.0328 | 1 | 0.0328 | 0 | 0 | 0 | 0.8095 | Ref |
| 1603 | JX561091 | Unassigned | Unassigned | Unassigned | 847_1 | Clustered | 26 | VC_847_1 | 26 | 0.6323 | 1 | 0.6323 | 0 | 0 | 0 | 0.9815 | Ref |
| 1604 | JX564242 | Unassigned | Unassigned | Unassigned | 268_0 | Clustered | 7 | VC_268_0 | 7 | 0.3427 | 1 | 0.3427 | 0 | 0 | 0 | 1 | Ref |
| 1605 | JX567312 | Unassigned | Unassigned | Unassigned | 268_0 | Clustered | 7 | VC_268_0 | 7 | 0.3427 | 1 | 0.3427 | 0 | 0 | 0 | 1 | Ref |
| 1606 | JX569801 | Unassigned | Unassigned | Unassigned | 844_0 | Clustered | 27 | VC_844_0 | 27 | 0.9385 | 1 | 0.9385 | 0 | 0 | 0 | 1 | Ref |
| 1607 | JX570702 | Unassigned | Unassigned | Unassigned | 765_0 | Clustered | 132 | VC_765_0 | 132 | 1 | 1 | 1 | 0 | 0 | 0 | 1 | Ref |
| 1608 | JX570703 | Unassigned | Unassigned | Unassigned | 765_0 | Clustered | 132 | VC_765_0 | 132 | 1 | 1 | 1 | 0 | 0 | 0 | 1 | Ref |
| 1609 | JX570704 | Unassigned | Unassigned | Unassigned | 765_0 | Clustered | 132 | VC_765_0 | 132 | 1 | 1 | 1 | 0 | 0 | 0 | 1 | Ref |
| 1610 | JX570705 | Unassigned | Unassigned | Unassigned | 765_0 | Clustered | 132 | VC_765_0 | 132 | 1 | 1 | 1 | 0 | 0 | 0 | 1 | Ref |
| 1611 | JX570706 | Unassigned | Unassigned | Unassigned | 765_0 | Clustered | 132 | VC_765_0 | 132 | 1 | 1 | 1 | 0 | 0 | 0 | 1 | Ref |
| 1612 | JX570707 | Unassigned | Unassigned | Unassigned | 765_0 | Clustered | 132 | VC_765_0 | 132 | 1 | 1 | 1 | 0 | 0 | 0 | 1 | Ref |
| 1613 | JX570708 | Unassigned | Unassigned | Unassigned | 765_0 | Clustered | 132 | VC_765_0 | 132 | 1 | 1 | 1 | 0 | 0 | 0 | 1 | Ref |
| 1614 | JX570709 | Unassigned | Unassigned | Unassigned | 765_0 | Clustered | 132 | VC_765_0 | 132 | 1 | 1 | 1 | 0 | 0 | 0 | 1 | Ref |
| 1615 | JX570710 | Unassigned | Unassigned | Unassigned | 765_0 | Clustered | 132 | VC_765_0 | 132 | 1 | 1 | 1 | 0 | 0 | 0 | 1 | Ref |
| 1616 | JX570711 | Unassigned | Unassigned | Unassigned | 765_0 | Clustered | 132 | VC_765_0 | 132 | 1 | 1 | 1 | 0 | 0 | 0 | 1 | Ref |
| 1617 | JX570712 | Unassigned | Unassigned | Unassigned | 765_0 | Clustered | 132 | VC_765_0 | 132 | 1 | 1 | 1 | 0 | 0 | 0 | 1 | Ref |
| 1618 | JX570713 | Unassigned | Unassigned | Unassigned | 765_0 | Clustered | 132 | VC_765_0 | 132 | 1 | 1 | 1 | 0 | 0 | 0 | 1 | Ref |

| | | | | | | | | | | | | | | | | | |
|---|---|---|---|---|---|---|---|---|---|---|---|---|---|---|---|---|---|
| 1619 | JX570714 | Unassigned | Unassigned | Unassigned | 765_0 | Clustered | 132 | VC_765_0 | 132 | 1 | 1 | 1 | 0 | 0 | 0 | 1 | Ref |
| 1620 | JX625144 | Unassigned | Unassigned | Unassigned | 314_0 | Clustered | 2 | VC_314_0 | 2 | 0.1808 | 0.99999983 | 0.1808 | 0 | 0 | 0 | 1 | Ref |
| 1621 | JX649096 | Unassigned | Unassigned | Unassigned | 371_16 | Clustered | 42 | VC_371_16 | 42 | 0.1341 | 1 | 0.1341 | 0 | 0 | 0 | 0.7282 | Ref |
| 1622 | JX649097 | Unassigned | Unassigned | Unassigned | 371_16 | Clustered | 42 | VC_371_16 | 42 | 0.1341 | 1 | 0.1341 | 0 | 0 | 0 | 0.7282 | Ref |
| 1623 | JX649098 | Unassigned | Unassigned | Unassigned | 371_3 | Clustered | 22 | VC_371_3 | 22 | 0.0787 | 1 | 0.0787 | 0 | 0 | 0 | 0.9048 | Ref |
| 1624 | JX649099 | Unassigned | Unassigned | Unassigned | 371_3 | Clustered | 22 | VC_371_3 | 22 | 0.0787 | 1 | 0.0787 | 0 | 0 | 0 | 0.9048 | Ref |
| 1625 | JX649100 | Unassigned | Unassigned | Unassigned | 371_16 | Clustered | 42 | VC_371_16 | 42 | 0.1341 | 1 | 0.1341 | 0 | 0 | 0 | 0.7282 | Ref |
| 1626 | JX658790 | Unassigned | Unassigned | Unassigned | 699_0 | Clustered | 71 | VC_699_0 | 71 | 0.9352 | 1 | 0.9352 | 0 | 0 | 0 | 1 | Ref |
| 1627 | JX681814 | Unassigned | Unassigned | Unassigned | 3_0 | Clustered | 17 | VC_3_0 | 17 | 0.5639 | 1 | 0.5639 | 0 | 0 | 0 | 1 | Ref |
| 1628 | JX846612 | Unassigned | Unassigned | Unassigned | 153_0 | Clustered | 25 | VC_153_0 | 25 | 0.7729 | 1 | 0.7729 | 0 | 0 | 0 | 1 | Ref |
| 1629 | JX846613 | Unassigned | Unassigned | Unassigned | 153_0 | Clustered | 25 | VC_153_0 | 25 | 0.7729 | 1 | 0.7729 | 0 | 0 | 0 | 1 | Ref |
| 1630 | JX863101 | Unassigned | Unassigned | Unassigned | 472_0 | Clustered | 47 | VC_472_0 | 47 | 0.9025 | 1 | 0.9025 | 0 | 0 | 0 | 1 | Ref |
| 1631 | JX865427 | Unassigned | Unassigned | Unassigned | 835_0 | Clustered | 67 | VC_835_0 | 67 | 0.9624 | 1 | 0.9624 | 0 | 0 | 0 | 1 | Ref |
| 1632 | JX866719 | Unassigned | Unassigned | Unassigned | 123_0 | Clustered | 23 | VC_123_0 | 23 | 0.817 | 1 | 0.817 | 0 | 0 | 0 | 1 | Ref |
| 1633 | JX867715 | Unassigned | Unassigned | Unassigned | 813_1 | Clustered | 24 | VC_813_1 | 24 | 0.535 | 1 | 0.535 | 0 | 0 | 0 | 1 | Ref |
| 1634 | JX872508 | Unassigned | Unassigned | Unassigned | 392_0 | Clustered | 126 | VC_392_0 | 126 | 0.9621 | 1 | 0.9621 | 0 | 0 | 0 | 1 | Ref |
| 1635 | JX875065 | Unassigned | Unassigned | Unassigned | 149_8 | Clustered | 66 | VC_149_8 | 66 | 0.4565 | 1 | 0.4565 | 0 | 0 | 0 | 0.8182 | Ref |
| 1636 | JX878496 | Unassigned | Unassigned | Unassigned | 132_0 | Clustered | 4 | VC_132_0 | 4 | 0.036 | 0.99998777 | 0.036 | 0 | 0 | 0 | 1 | Ref |
| 1637 | JX878671 | Unassigned | Unassigned | Unassigned | 149_13 | Clustered | 3 | VC_149_13 | 3 | 0.0087 | 0.95178903 | 0.0083 | 0 | 0 | 0 | 1 | Ref |
| 1638 | JX879087 | Unassigned | Unassigned | Unassigned | 412_0 | Clustered | 39 | VC_412_0 | 39 | 0.845 | 1 | 0.845 | 0 | 0 | 0 | 1 | Ref |
| 1639 | JX880034 | Unassigned | Unassigned | Unassigned | 779_0 | Clustered | 37 | VC_779_0 | 37 | 0.5682 | 1 | 0.5682 | 0 | 0 | 0 | 1 | Ref |
| 1640 | JX880072 | Unassigned | Unassigned | Unassigned | 117_0 | Clustered | 9 | VC_117_0 | 9 | 0.9977 | 0.98988637 | 0.9876 | 0 | 0 | 0 | 1 | Ref |
| 1641 | JX887877 | Unassigned | Unassigned | Unassigned | 829_0 | Clustered | 5 | VC_829_0 | 5 | 0.8821 | 0.99999832 | 0.8821 | 0 | 0 | 0 | 1 | Ref |
| 1642 | JX889246 | Unassigned | Unassigned | Unassigned | 982_0 | Clustered | 62 | VC_982_0 | 62 | 0.8646 | 1 | 0.8646 | 0 | 0 | 0 | 0.743 | Ref |
| 1643 | JX899358 | Unassigned | Unassigned | Unassigned | 216_6 | Clustered | 42 | VC_216_6 | 42 | 0.473 | 1 | 0.473 | 0 | 0 | 0 | 0.8351 | Ref |
| 1644 | JX901189 | Unassigned | Unassigned | Unassigned | 574_0 | Clustered | 7 | VC_574_0 | 7 | 0.2734 | 1 | 0.2734 | 0 | 0 | 0 | 1 | Ref |
| 1645 | JX912252 | Unassigned | Unassigned | Unassigned | 575_0 | Clustered | 35 | VC_575_0 | 35 | 0.8815 | 1 | 0.8815 | 0 | 0 | 0 | 1 | Ref |
| 1646 | JX976549 | Unassigned | Unassigned | Unassigned | 698_0 | Clustered | 26 | VC_698_0 | 26 | 0.9366 | 1 | 0.9366 | 0 | 0 | 0 | 1 | Ref |
| 1647 | JX997978 | Unassigned | Unassigned | Unassigned | 423_0 | Clustered | 45 | VC_423_0 | 45 | 0.9484 | 1 | 0.9484 | 0 | 0 | 0 | 1 | Ref |
| 1648 | KC012913 | Unassigned | Unassigned | Unassigned | 149_8 | Clustered | 66 | VC_149_8 | 66 | 0.4565 | 1 | 0.4565 | 0 | 0 | 0 | 0.8182 | Ref |
| 1649 | KC013021 | Unassigned | Unassigned | Unassigned | 861_0 | Clustered | 9 | VC_861_0 | 9 | 0.5869 | 1 | 0.5869 | 0 | 0 | 0 | 1 | Ref |
| 1650 | KC013022 | Unassigned | Unassigned | Unassigned | 861_0 | Clustered | 9 | VC_861_0 | 9 | 0.5869 | 1 | 0.5869 | 0 | 0 | 0 | 1 | Ref |
| 1651 | KC013023 | Unassigned | Unassigned | Unassigned | 861_0 | Clustered | 9 | VC_861_0 | 9 | 0.5869 | 1 | 0.5869 | 0 | 0 | 0 | 1 | Ref |
| 1652 | KC013024 | Unassigned | Unassigned | Unassigned | 861_0 | Clustered | 9 | VC_861_0 | 9 | 0.5869 | 1 | 0.5869 | 0 | 0 | 0 | 1 | Ref |
| 1653 | KC013025 | Unassigned | Unassigned | Unassigned | 861_0 | Clustered | 9 | VC_861_0 | 9 | 0.5869 | 1 | 0.5869 | 0 | 0 | 0 | 1 | Ref |
| 1654 | KC013026 | Unassigned | Unassigned | Unassigned | 862_0 | Clustered | 11 | VC_862_0 | 11 | 0.6745 | 1 | 0.6745 | 0 | 0 | 0 | 1 | Ref |
| 1655 | KC013027 | Unassigned | Unassigned | Unassigned | 862_0 | Clustered | 11 | VC_862_0 | 11 | 0.6745 | 1 | 0.6745 | 0 | 0 | 0 | 1 | Ref |
| 1656 | KC013028 | Unassigned | Unassigned | Unassigned | 862_0 | Clustered | 11 | VC_862_0 | 11 | 0.6745 | 1 | 0.6745 | 0 | 0 | 0 | 1 | Ref |
| 1657 | KC013029 | Unassigned | Unassigned | Unassigned | 862_0 | Clustered | 11 | VC_862_0 | 11 | 0.6745 | 1 | 0.6745 | 0 | 0 | 0 | 1 | Ref |
| 1658 | KC107834 | Unassigned | Unassigned | Unassigned | 878_0 | Clustered | 32 | VC_878_0 | 32 | 0.9965 | 1 | 0.9965 | 0 | 0 | 0 | 1 | Ref |
| 1659 | KC109329 | Unassigned | Unassigned | Unassigned | 451_0 | Clustered | 80 | VC_451_0 | 80 | 0.9991 | 1 | 0.9991 | 0 | 0 | 0 | 1 | Ref |
| 1660 | KC117376 | Unassigned | Unassigned | Unassigned | 311_0 | Clustered | 6 | VC_311_0 | 6 | 0.1997 | 1 | 0.1997 | 0 | 0 | 0 | 1 | Ref |
| 1661 | KC117377 | Unassigned | Unassigned | Unassigned | 313_1 | Clustered | 3 | VC_313_1 | 3 | 0.3511 | 0.99802018 | 0.3504 | 0 | 0 | 0 | 1 | Ref |
| 1662 | KC131021 | Unassigned | Unassigned | Unassigned | 199_0 | Clustered | 12 | VC_199_0 | 12 | 0.448 | 1 | 0.448 | 0 | 0 | 0 | 1 | Ref |
| 1663 | KC131022 | Unassigned | Unassigned | Unassigned | 995_0 | Clustered | 69 | VC_995_0 | 69 | 0.8547 | 1 | 0.8547 | 0 | 0 | 0 | 1 | Ref |
| 1664 | KC131023 | Unassigned | Unassigned | Unassigned | 403_0 | Clustered | 20 | VC_403_0 | 20 | 0.6598 | 1 | 0.6598 | 0 | 0 | 0 | 1 | Ref |
| 1665 | KC131025 | Unassigned | Unassigned | Unassigned | 403_0 | Clustered | 20 | VC_403_0 | 20 | 0.6598 | 1 | 0.6598 | 0 | 0 | 0 | 1 | Ref |
| 1666 | KC131129 | Unassigned | Unassigned | Unassigned | 531_0 | Clustered | 9 | VC_531_0 | 9 | 0.26 | 1 | 0.26 | 0 | 0 | 0 | 1 | Ref |
| 1667 | KC139511 | Unassigned | Unassigned | Unassigned | 637_0 | Clustered | 115 | VC_637_0 | 115 | 0.731 | 1 | 0.731 | 0 | 0 | 0 | 0.8764 | Ref |
| 1668 | KC139512 | Unassigned | Unassigned | Unassigned | 592_0 | Clustered | 50 | VC_592_0 | 50 | 0.7314 | 1 | 0.7314 | 0 | 0 | 0 | 1 | Ref |
| 1669 | KC139514 | Unassigned | Unassigned | Unassigned | 592_0 | Clustered | 50 | VC_592_0 | 50 | 0.7314 | 1 | 0.7314 | 0 | 0 | 0 | 1 | Ref |
| 1670 | KC139515 | Unassigned | Unassigned | Unassigned | 592_0 | Clustered | 50 | VC_592_0 | 50 | 0.7314 | 1 | 0.7314 | 0 | 0 | 0 | 1 | Ref |
| 1671 | KC139517 | Unassigned | Unassigned | Unassigned | 780_0 | Clustered | 8 | VC_780_0 | 8 | 0.5207 | 1 | 0.5207 | 0 | 0 | 0 | 1 | Ref |
| 1672 | KC139518 | Unassigned | Unassigned | Unassigned | 540_0 | Clustered | 15 | VC_540_0 | 15 | 0.2866 | 1 | 0.2866 | 0 | 0 | 0 | 1 | Ref |
| 1673 | KC139519 | Unassigned | Unassigned | Unassigned | 592_0 | Clustered | 50 | VC_592_0 | 50 | 0.7314 | 1 | 0.7314 | 0 | 0 | 0 | 1 | Ref |
| 1674 | KC139520 | Unassigned | Unassigned | Unassigned | 780_0 | Clustered | 8 | VC_780_0 | 8 | 0.5207 | 1 | 0.5207 | 0 | 0 | 0 | 1 | Ref |
| 1675 | KC139521 | Unassigned | Unassigned | Unassigned | 228_0 | Clustered | 60 | VC_228_0 | 60 | 0.942 | 1 | 0.942 | 0 | 0 | 0 | 1 | Ref |
| 1676 | KC171646 | Unassigned | Unassigned | Unassigned | 272_0 | Clustered | 11 | VC_272_0 | 11 | 0.4915 | 1 | 0.4915 | 0 | 0 | 0 | 0.8727 | Ref |
| 1677 | KC171647 | Unassigned | Unassigned | Unassigned | 272_0 | Clustered | 11 | VC_272_0 | 11 | 0.4915 | 1 | 0.4915 | 0 | 0 | 0 | 0.8727 | Ref |
| 1678 | KC172839 | Unassigned | Unassigned | Unassigned | 513_0 | Clustered | 32 | VC_513_0 | 32 | 0.6881 | 1 | 0.6881 | 0 | 0 | 0 | 1 | Ref |
| 1679 | KC182542 | Unassigned | Unassigned | Unassigned | 202_0 | Clustered | 102 | VC_202_0 | 102 | 0.4049 | 1 | 0.4049 | 0 | 0 | 0 | 0.7876 | Ref |
| 1680 | KC182543 | Unassigned | Unassigned | Unassigned | 202_0 | Clustered | 102 | VC_202_0 | 102 | 0.4049 | 1 | 0.4049 | 0 | 0 | 0 | 0.7876 | Ref |
| 1681 | KC182544 | Unassigned | Unassigned | Unassigned | 202_0 | Clustered | 102 | VC_202_0 | 102 | 0.4049 | 1 | 0.4049 | 0 | 0 | 0 | 0.7876 | Ref |
| 1682 | KC182545 | Unassigned | Unassigned | Unassigned | 202_0 | Clustered | 102 | VC_202_0 | 102 | 0.4049 | 1 | 0.4049 | 0 | 0 | 0 | 0.7876 | Ref |
| 1683 | KC182546 | Unassigned | Unassigned | Unassigned | 202_0 | Clustered | 102 | VC_202_0 | 102 | 0.4049 | 1 | 0.4049 | 0 | 0 | 0 | 0.7876 | Ref |
| 1684 | KC182547 | Unassigned | Unassigned | Unassigned | 202_0 | Clustered | 102 | VC_202_0 | 102 | 0.4049 | 1 | 0.4049 | 0 | 0 | 0 | 0.7876 | Ref |
| 1685 | KC182548 | Unassigned | Unassigned | Unassigned | 202_0 | Clustered | 102 | VC_202_0 | 102 | 0.4049 | 1 | 0.4049 | 0 | 0 | 0 | 0.7876 | Ref |
| 1686 | KC182549 | Unassigned | Unassigned | Unassigned | 202_0 | Clustered | 102 | VC_202_0 | 102 | 0.4049 | 1 | 0.4049 | 0 | 0 | 0 | 0.7876 | Ref |
| 1687 | KC182550 | Unassigned | Unassigned | Unassigned | 202_0 | Clustered | 102 | VC_202_0 | 102 | 0.4049 | 1 | 0.4049 | 0 | 0 | 0 | 0.7876 | Ref |
| 1688 | KC182551 | Unassigned | Unassigned | Unassigned | 202_0 | Clustered | 102 | VC_202_0 | 102 | 0.4049 | 1 | 0.4049 | 0 | 0 | 0 | 0.7876 | Ref |
| 1689 | KC182552 | Unassigned | Unassigned | Unassigned | 202_0 | Clustered | 102 | VC_202_0 | 102 | 0.4049 | 1 | 0.4049 | 0 | 0 | 0 | 0.7876 | Ref |
| 1690 | KC206276 | Unassigned | Unassigned | Unassigned | 779_0 | Clustered | 37 | VC_779_0 | 37 | 0.5682 | 1 | 0.5682 | 0 | 0 | 0 | 1 | Ref |
| 1691 | KC237308 | Unassigned | Unassigned | Unassigned | 362_0 | Clustered | 2 | VC_362_0 | 2 | 0.002 | 0.83307735 | 0.0017 | 0 | 0 | 0 | 1 | Ref |
| 1692 | KC237729 | Unassigned | Unassigned | Unassigned | 417_0 | Clustered | 5 | VC_417_0 | 5 | 0.6629 | 0.9977434 | 0.6614 | 0 | 0 | 0 | 0.8 | Ref |
| 1693 | KC252997 | Unassigned | Unassigned | Unassigned | 666_0 | Clustered | 4 | VC_666_0 | 4 | 1 | 1 | 1 | 0 | 0 | 0 | 1 | Ref |
| 1694 | KC262634 | Unassigned | Unassigned | Unassigned | 630_0 | Clustered | 20 | VC_630_0 | 20 | 0.8513 | 1 | 0.8513 | 0 | 0 | 0 | 1 | Ref |
| 1695 | KC292020 | Unassigned | Unassigned | Unassigned | 310_0 | Clustered | 33 | VC_310_0 | 33 | 0.8771 | 1 | 0.8771 | 0 | 0 | 0 | 1 | Ref |
| 1696 | KC292021 | Unassigned | Unassigned | Unassigned | 311_0 | Clustered | 6 | VC_311_0 | 6 | 0.1997 | 1 | 0.1997 | 0 | 0 | 0 | 1 | Ref |
| 1697 | KC292022 | Unassigned | Unassigned | Unassigned | 310_0 | Clustered | 33 | VC_310_0 | 33 | 0.8771 | 1 | 0.8771 | 0 | 0 | 0 | 1 | Ref |
| 1698 | KC292023 | Unassigned | Unassigned | Unassigned | 459_0 | Clustered | 3 | VC_459_0 | 3 | 0.8542 | 0.99054823 | 0.8461 | 0 | 0 | 0 | 1 | Ref |
| 1699 | KC292024 | Unassigned | Unassigned | Unassigned | 991_0 | Clustered | 2 | VC_991_0 | 2 | 0.3757 | 0.91418477 | 0.3435 | 0 | 0 | 0 | 1 | Ref |
| 1700 | KC292026 | Unassigned | Unassigned | Unassigned | 312_0 | Clustered | 2 | VC_312_0 | 2 | 0.0307 | 0.95436427 | 0.0293 | 0 | 0 | 0 | 1 | Ref |
| 1701 | KC292027 | Unassigned | Unassigned | Unassigned | 313_1 | Clustered | 3 | VC_313_1 | 3 | 0.3511 | 0.99802018 | 0.3504 | 0 | 0 | 0 | 1 | Ref |
| 1702 | KC292028 | Unassigned | Unassigned | Unassigned | 991_0 | Clustered | 2 | VC_991_0 | 2 | 0.3757 | 0.91418477 | 0.3435 | 0 | 0 | 0 | 1 | Ref |
| 1703 | KC292029 | Unassigned | Unassigned | Unassigned | 313_0 | Clustered | 2 | VC_313_0 | 2 | 0.1721 | 0.90744399 | 0.1562 | 0 | 0 | 0 | 1 | Ref |
| 1704 | KC294142 | Unassigned | Unassigned | Unassigned | 472_0 | Clustered | 47 | VC_472_0 | 47 | 0.9025 | 1 | 0.9025 | 0 | 0 | 0 | 1 | Ref |
| 1705 | KC295538 | Unassigned | Unassigned | Unassigned | 164_11 | Clustered | 10 | VC_164_11 | 10 | 0.1507 | 1 | 0.1507 | 0 | 0 | 0 | 0.9111 | Ref |
| 1706 | KC310802 | Unassigned | Unassigned | Unassigned | 340_0 | Clustered | 31 | VC_340_0 | 31 | 0.9549 | 1 | 0.9549 | 0 | 0 | 0 | 1 | Ref |
| 1707 | KC310803 | Unassigned | Unassigned | Unassigned | 340_0 | Clustered | 31 | VC_340_0 | 31 | 0.9549 | 1 | 0.9549 | 0 | 0 | 0 | 1 | Ref |
| 1708 | KC310804 | Unassigned | Unassigned | Unassigned | 340_0 | Clustered | 31 | VC_340_0 | 31 | 0.9549 | 1 | 0.9549 | 0 | 0 | 0 | 1 | Ref |
| 1709 | KC310805 | Unassigned | Unassigned | Unassigned | 340_0 | Clustered | 31 | VC_340_0 | 31 | 0.9549 | 1 | 0.9549 | 0 | 0 | 0 | 1 | Ref |
| 1710 | KC310806 | Unassigned | Unassigned | Unassigned | 340_0 | Clustered | 31 | VC_340_0 | 31 | 0.9549 | 1 | 0.9549 | 0 | 0 | 0 | 1 | Ref |
| 1711 | KC311669 | Unassigned | Unassigned | Unassigned | 699_0 | Clustered | 71 | VC_699_0 | 71 | 0.9352 | 1 | 0.9352 | 0 | 0 | 0 | 1 | Ref |
| 1712 | KC330679 | Unassigned | Unassigned | Unassigned | 100_0 | Clustered | 15 | VC_100_0 | 15 | 0.6536 | 1 | 0.6536 | 0 | 0 | 0 | 1 | Ref |
| 1713 | KC330680 | Unassigned | Unassigned | Unassigned | 100_0 | Clustered | 15 | VC_100_0 | 15 | 0.6536 | 1 | 0.6536 | 0 | 0 | 0 | 1 | Ref |
| 1714 | KC330681 | Unassigned | Unassigned | Unassigned | 100_0 | Clustered | 15 | VC_100_0 | 15 | 0.6536 | 1 | 0.6536 | 0 | 0 | 0 | 1 | Ref |
| 1715 | KC330682 | Unassigned | Unassigned | Unassigned | 100_0 | Clustered | 15 | VC_100_0 | 15 | 0.6536 | 1 | 0.6536 | 0 | 0 | 0 | 1 | Ref |
| 1716 | KC330683 | Unassigned | Unassigned | Unassigned | 100_0 | Clustered | 15 | VC_100_0 | 15 | 0.6536 | 1 | 0.6536 | 0 | 0 | 0 | 1 | Ref |
| 1717 | KC330684 | Unassigned | Unassigned | Unassigned | 100_0 | Clustered | 15 | VC_100_0 | 15 | 0.6536 | 1 | 0.6536 | 0 | 0 | 0 | 1 | Ref |
| 1718 | KC333879 | Unassigned | Unassigned | Unassigned | 455_0 | Clustered | 18 | VC_455_0 | 18 | 0.4598 | 1 | 0.4598 | 0 | 0 | 0 | 1 | Ref |
| 1719 | KC342645 | Unassigned | Unassigned | Unassigned | 7_0 | Clustered | 28 | VC_7_0 | 28 | 0.3916 | 1 | 0.3916 | 0 | 0 | 0 | 0.9947 | Ref |
| 1720 | KC348600 | Unassigned | Unassigned | Unassigned | 317_2 | Clustered | 20 | VC_317_2 | 20 | 0.1976 | 1 | 0.1976 | 0 | 0 | 0 | 0.6842 | Ref |
| 1721 | KC348601 | Unassigned | Unassigned | Unassigned | 317_2 | Clustered | 20 | VC_317_2 | 20 | 0.1976 | 1 | 0.1976 | 0 | 0 | 0 | 0.6842 | Ref |
| 1722 | KC348602 | Unassigned | Unassigned | Unassigned | 317_2 | Clustered | 20 | VC_317_2 | 20 | 0.1976 | 1 | 0.1976 | 0 | 0 | 0 | 0.6842 | Ref |
| 1723 | KC348603 | Unassigned | Unassigned | Unassigned | 632_0 | Clustered | 37 | VC_632_0 | 37 | 0.581 | 1 | 0.581 | 0 | 0 | 0 | 0.9234 | Ref |
| 1724 | KC357596 | Unassigned | Unassigned | Unassigned | 0_0 | Clustered | 3 | VC_0_0 | 3 | 1 | 1 | 1 | 0 | 0 | 0 | 1 | Ref |
| 1725 | KC413988 | Unassigned | Unassigned | Unassigned | 412_0 | Clustered | 39 | VC_412_0 | 39 | 0.845 | 1 | 0.845 | 0 | 0 | 0 | 1 | Ref |
| 1726 | KC430106 | Unassigned | Unassigned | Unassigned | 884_1 | Clustered | 8 | VC_884_1 | 8 | 0.1102 | 0.9989323 | 0.1101 | 0 | 0 | 0 | 0.75 | Ref |
| 1727 | KC438282 | Unassigned | Unassigned | Unassigned | 784_0 | Clustered | 6 | VC_784_0 | 6 | 0.5517 | 1 | 0.5517 | 0 | 0 | 0 | 1 | Ref |
| 1728 | KC438283 | Unassigned | Unassigned | Unassigned | 784_0 | Clustered | 6 | VC_784_0 | 6 | 0.5517 | 1 | 0.5517 | 0 | 0 | 0 | 1 | Ref |
| 1729 | KC465898 | Unassigned | Unassigned | Unassigned | 486_0 | Clustered | 18 | VC_486_0 | 18 | 0.887 | 1 | 0.887 | 0 | 0 | 0 | 1 | Ref |
| 1730 | KC465901 | Unassigned | Unassigned | Unassigned | 480_0 | Clustered | 19 | VC_480_0 | 19 | 0.5445 | 1 | 0.5445 | 0 | 0 | 0 | 1 | Ref |
| 1731 | KC481682 | Unassigned | Unassigned | Unassigned | 475_2 | Clustered | 3 | VC_475_2 | 3 | 0.0585 | 0.98883022 | 0.0578 | 0 | 0 | 0 | 1 | Ref |
| 1732 | KC522412 | Unassigned | Unassigned | Unassigned | 202_0 | Clustered | 102 | VC_202_0 | 102 | 0.4049 | 1 | 0.4049 | 0 | 0 | 0 | 0.7876 | Ref |
| 1733 | KC542353 | Unassigned | Unassigned | Unassigned | 644_0 | Clustered | 5 | VC_644_0 | 5 | 0.6649 | 0.99999975 | 0.6649 | 0 | 0 | 0 | 1 | Ref |
| 1734 | KC556893 | Unassigned | Unassigned | Unassigned | 879_0 | Clustered | 36 | VC_879_0 | 36 | 0.994 | 1 | 0.994 | 0 | 0 | 0 | 1 | Ref |
| 1735 | KC556894 | Unassigned | Unassigned | Unassigned | 879_0 | Clustered | 36 | VC_879_0 | 36 | 0.994 | 1 | 0.994 | 0 | 0 | 0 | 1 | Ref |
| 1736 | KC556895 | Unassigned | Unassigned | Unassigned | 879_0 | Clustered | 36 | VC_879_0 | 36 | 0.994 | 1 | 0.994 | 0 | 0 | 0 | 1 | Ref |
| 1737 | KC556896 | Unassigned | Unassigned | Unassigned | 879_0 | Clustered | 36 | VC_879_0 | 36 | 0.994 | 1 | 0.994 | 0 | 0 | 0 | 1 | Ref |
| 1738 | KC556897 | Unassigned | Unassigned | Unassigned | 879_0 | Clustered | 36 | VC_879_0 | 36 | 0.994 | 1 | 0.994 | 0 | 0 | 0 | 1 | Ref |
| 1739 | KC556898 | Unassigned | Unassigned | Unassigned | 879_0 | Clustered | 36 | VC_879_0 | 36 | 0.994 | 1 | 0.994 | 0 | 0 | 0 | 1 | Ref |
| 1740 | KC576783 | Unassigned | Unassigned | Unassigned | 521_0 | Clustered | 37 | VC_521_0 | 37 | 0.912 | 1 | 0.912 | 0 | 0 | 0 | 1 | Ref |
| 1741 | KC576784 | Unassigned | Unassigned | Unassigned | 371_2 | Clustered | 8 | VC_371_2 | 8 | 0.0232 | 1 | 0.0232 | 0 | 0 | 0 | 0.8571 | Ref |
| 1742 | KC579452 | Unassigned | Unassigned | Unassigned | 455_0 | Clustered | 18 | VC_455_0 | 18 | 0.4598 | 1 | 0.4598 | 0 | 0 | 0 | 1 | Ref |
| 1743 | KC581799 | Unassigned | Unassigned | Unassigned | 632_0 | Clustered | 37 | VC_632_0 | 37 | 0.581 | 1 | 0.581 | 0 | 0 | 0 | 0.9234 | Ref |
| 1744 | KC595278 | Unassigned | Unassigned | Unassigned | 10_0 | Clustered | 25 | VC_10_0 | 25 | 0.3467 | 1 | 0.3467 | 0 | 0 | 0 | 0.9733 | Ref |
| 1745 | KC595279 | Unassigned | Unassigned | Unassigned | 12_2 | Clustered | 19 | VC_12_2 | 19 | 0.2018 | 1 | 0.2018 | 0 | 0 | 0 | 1 | Ref |
| 1746 | KC595511 | Unassigned | Unassigned | Unassigned | 1006_0 | Clustered | 8 | VC_1006_0 | 8 | 0.7407 | 1 | 0.7407 | 0 | 0 | 0 | 1 | Ref |
| 1747 | KC595512 | Unassigned | Unassigned | Unassigned | 87_1 | Clustered | 3 | VC_87_1 | 3 | 0.1636 | 0.99777924 | 0.1632 | 0 | 0 | 0 | 1 | Ref |
| 1748 | KC595513 | Unassigned | Unassigned | Unassigned | 87_1 | Clustered | 3 | VC_87_1 | 3 | 0.1636 | 0.99777924 | 0.1632 | 0 | 0 | 0 | 1 | Ref |
| 1749 | KC595514 | Unassigned | Unassigned | Unassigned | 141_0 | Clustered | 6 | VC_141_0 | 6 | 0.5999 | 1 | 0.5999 | 0 | 0 | 0 | 1 | Ref |
| 1750 | KC595515 | Unassigned | Unassigned | Unassigned | 141_0 | Clustered | 6 | VC_141_0 | 6 | 0.5999 | 1 | 0.5999 | 0 | 0 | 0 | 1 | Ref |
| 1751 | KC595517 | Unassigned | Unassigned | Unassigned | 141_0 | Clustered | 6 | VC_141_0 | 6 | 0.5999 | 1 | 0.5999 | 0 | 0 | 0 | 1 | Ref |
| 1752 | KC595518 | Unassigned | Unassigned | Unassigned | 141_0 | Clustered | 6 | VC_141_0 | 6 | 0.5999 | 1 | 0.5999 | 0 | 0 | 0 | 1 | Ref |
| 1753 | KC618326 | Unassigned | Unassigned | Unassigned | 228_0 | Clustered | 60 | VC_228_0 | 60 | 0.942 | 1 | 0.942 | 0 | 0 | 0 | 1 | Ref |
| 1754 | KC618393 | Unassigned | Unassigned | Unassigned | 599_0 | Clustered | 50 | VC_599_0 | 50 | 0.9989 | 1 | 0.9989 | 0 | 0 | 0 | 1 | Ref |
| 1755 | KC661271 | Unassigned | Unassigned | Unassigned | 517_1 | Clustered | 108 | VC_517_1 | 108 | 0.8483 | 1 | 0.8483 | 0 | 0 | 0 | 0.9955 | Ref |
| 1756 | KC661272 | Unassigned | Unassigned | Unassigned | 514_0 | Clustered | 75 | VC_514_0 | 75 | 0.9402 | 1 | 0.9402 | 0 | 0 | 0 | 1 | Ref |
| 1757 | KC661273 | Unassigned | Unassigned | Unassigned | 325_23 | Clustered | 17 | VC_325_23 | 17 | 0.0716 | 1 | 0.0716 | 0 | 0 | 0 | 0.6397 | Ref |
| 1758 | KC661274 | Unassigned | Unassigned | Unassigned | 371_3 | Clustered | 22 | VC_371_3 | 22 | 0.0787 | 1 | 0.0787 | 0 | 0 | 0 | 0.9048 | Ref |
| 1759 | KC661275 | Unassigned | Unassigned | Unassigned | 219_0 | Clustered | 5 | VC_219_0 | 5 | 0.08 | 0.99999998 | 0.08 | 0 | 0 | 0 | 1 | Ref |
| 1760 | KC661276 | Unassigned | Unassigned | Unassigned | 888_0 | Clustered | 28 | VC_888_0 | 28 | 0.5497 | 1 | 0.5497 | 0 | 0 | 0 | 1 | Ref |
| 1761 | KC661277 | Unassigned | Unassigned | Unassigned | 551_7 | Clustered | 28 | VC_551_7 | 28 | 0.1661 | 1 | 0.1661 | 0 | 0 | 0 | 0.8519 | Ref |
| 1762 | KC661278 | Unassigned | Unassigned | Unassigned | 520_79 | Clustered/Singleton | 1 | VC_520_79 | 1 | 0 | 1 | 0 | 0 | 0 | 0 | 1 | Ref |
| 1763 | KC661279 | Unassigned | Unassigned | Unassigned | 515_0 | Clustered | 16 | VC_515_0 | 16 | 0.3793 | 1 | 0.3793 | 0 | 0 | 0 | 1 | Ref |
| 1764 | KC661280 | Unassigned | Unassigned | Unassigned | 520_70 | Clustered/Singleton | 1 | VC_520_70 | 1 | 0 | 1 | 0 | 0 | 0 | 0 | 1 | Ref |
| 1765 | KC661281 | Unassigned | Unassigned | Unassigned | 514_0 | Clustered | 75 | VC_514_0 | 75 | 0.9402 | 1 | 0.9402 | 0 | 0 | 0 | 1 | Ref |
| 1766 | KC677662 | Unassigned | Unassigned | Unassigned | 592_0 | Clustered | 50 | VC_592_0 | 50 | 0.7314 | 1 | 0.7314 | 0 | 0 | 0 | 1 | Ref |
| 1767 | KC677663 | Unassigned | Unassigned | Unassigned | 9_0 | Clustered | 38 | VC_9_0 | 38 | 0.4895 | 1 | 0.4895 | 0 | 0 | 0 | 1 | Ref |
| 1768 | KC685370 | Unassigned | Unassigned | Unassigned | 818_0 | Clustered | 20 | VC_818_0 | 20 | 0.9601 | 1 | 0.9601 | 0 | 0 | 0 | 1 | Ref |
| 1769 | KC690136 | Unassigned | Unassigned | Unassigned | 772_15 | Clustered | 5 | VC_772_15 | 5 | 0.0243 | 0.76144861 | 0.0185 | 0 | 0 | 0 | 0.9 | Ref |
| 1770 | KC691254 | Unassigned | Unassigned | Unassigned | 888_0 | Clustered | 28 | VC_888_0 | 28 | 0.5497 | 1 | 0.5497 | 0 | 0 | 0 | 1 | Ref |
| 1771 | KC691255 | Unassigned | Unassigned | Unassigned | 551_7 | Clustered | 28 | VC_551_7 | 28 | 0.1661 | 1 | 0.1661 | 0 | 0 | 0 | 0.8519 | Ref |
| 1772 | KC691256 | Unassigned | Unassigned | Unassigned | 519_0 | Clustered | 39 | VC_519_0 | 39 | 0.8156 | 1 | 0.8156 | 0 | 0 | 0 | 1 | Ref |
| 1773 | KC691257 | Unassigned | Unassigned | Unassigned | 558_5 | Clustered/Singleton | 1 | VC_558_5 | 1 | 0 | 1 | 0 | 0 | 0 | 0 | 1 | Ref |
| 1774 | KC691258 | Unassigned | Unassigned | Unassigned | 371_3 | Clustered | 22 | VC_371_3 | 22 | 0.0787 | 1 | 0.0787 | 0 | 0 | 0 | 0.9048 | Ref |
| 1775 | KC699836 | Unassigned | Unassigned | Unassigned | 85_2 | Clustered | 3 | VC_85_2 | 3 | 0.0334 | 0.99730296 | 0.0333 | 0 | 0 | 0 | 1 | Ref |
| 1776 | KC700556 | Unassigned | Unassigned | Unassigned | 982_0 | Clustered | 62 | VC_982_0 | 62 | 0.8646 | 1 | 0.8646 | 0 | 0 | 0 | 0.743 | Ref |
| 1777 | KC700557 | Unassigned | Unassigned | Unassigned | 982_0 | Clustered | 62 | VC_982_0 | 62 | 0.8646 | 1 | 0.8646 | 0 | 0 | 0 | 0.743 | Ref |
| 1778 | KC700558 | Unassigned | Unassigned | Unassigned | 982_0 | Clustered | 62 | VC_982_0 | 62 | 0.8646 | 1 | 0.8646 | 0 | 0 | 0 | 0.743 | Ref |
| 1779 | KC701493 | Unassigned | Unassigned | Unassigned | 325_17 | Clustered/Singleton | 1 | VC_325_17 | 1 | 0 | 1 | 0 | 0 | 0 | 0 | 1 | Ref |
| 1780 | KC710998 | Unassigned | Unassigned | Unassigned | 941_0 | Clustered | 40 | VC_941_0 | 40 | 0.6903 | 1 | 0.6903 | 0 | 0 | 0 | 1 | Ref |

| | | | | | | | | | | | | | | | | | |
|---|---|---|---|---|---|---|---|---|---|---|---|---|---|---|---|---|---|
| 1781 | KC736071 | Unassigned | Unassigned | Unassigned | 520_52 | Clustered/Singleton | 1 | VC_520_52 | 1 | 0 | 1 | 0 | 0 | 0 | 0 | 1 | Ref |
| 1782 | KC736978 | Unassigned | Unassigned | Unassigned | 45_0 | Clustered | 12 | VC_45_0 | 12 | 0.35 | 1 | 0.35 | 0 | 0 | 0 | 1 | Ref |
| 1783 | KC748968 | Unassigned | Unassigned | Unassigned | 558_49 | Clustered | 7 | VC_558_49 | 7 | 0.0194 | 0.99510369 | 0.0193 | 0 | 0 | 0 | 0.8571 | Ref |
| 1784 | KC748969 | Unassigned | Unassigned | Unassigned | 551_8 | Clustered | 19 | VC_551_8 | 19 | 0.1039 | 1 | 0.1039 | 0 | 0 | 0 | 0.8713 | Ref |
| 1785 | KC748970 | Unassigned | Unassigned | Unassigned | 558_12 | Clustered | 3 | VC_558_12 | 3 | 0.0064 | 0.87854253 | 0.0057 | 0 | 0 | 0 | 1 | Ref |
| 1786 | KC748971 | Unassigned | Unassigned | Unassigned | 551_7 | Clustered | 28 | VC_551_7 | 28 | 0.1661 | 1 | 0.1661 | 0 | 0 | 0 | 0.8519 | Ref |
| 1787 | KC751414 | Unassigned | Unassigned | Unassigned | 756_0 | Clustered | 2 | VC_756_0 | 2 | 0.1637 | 0.95427014 | 0.1562 | 0 | 0 | 0 | 1 | Ref |
| 1788 | KC758116 | Unassigned | Unassigned | Unassigned | 166_0 | Clustered | 26 | VC_166_0 | 26 | 0.7923 | 1 | 0.7923 | 0 | 0 | 0 | 1 | Ref |
| 1789 | KC787102 | Unassigned | Unassigned | Unassigned | 215_1 | Clustered | 19 | VC_215_1 | 19 | 0.4188 | 1 | 0.4188 | 0 | 0 | 0 | 1 | Ref |
| 1790 | KC787103 | Unassigned | Unassigned | Unassigned | 761_0 | Clustered | 67 | VC_761_0 | 67 | 0.8947 | 1 | 0.8947 | 0 | 0 | 0 | 1 | Ref |
| 1791 | KC787104 | Unassigned | Unassigned | Unassigned | 761_0 | Clustered | 67 | VC_761_0 | 67 | 0.8947 | 1 | 0.8947 | 0 | 0 | 0 | 1 | Ref |
| 1792 | KC787105 | Unassigned | Unassigned | Unassigned | 215_1 | Clustered | 19 | VC_215_1 | 19 | 0.4188 | 1 | 0.4188 | 0 | 0 | 0 | 1 | Ref |
| 1793 | KC787106 | Unassigned | Unassigned | Unassigned | 215_1 | Clustered | 19 | VC_215_1 | 19 | 0.4188 | 1 | 0.4188 | 0 | 0 | 0 | 1 | Ref |
| 1794 | KC787107 | Unassigned | Unassigned | Unassigned | 761_0 | Clustered | 67 | VC_761_0 | 67 | 0.8947 | 1 | 0.8947 | 0 | 0 | 0 | 1 | Ref |
| 1795 | KC787108 | Unassigned | Unassigned | Unassigned | 761_0 | Clustered | 67 | VC_761_0 | 67 | 0.8947 | 1 | 0.8947 | 0 | 0 | 0 | 1 | Ref |
| 1796 | KC787109 | Unassigned | Unassigned | Unassigned | 761_0 | Clustered | 67 | VC_761_0 | 67 | 0.8947 | 1 | 0.8947 | 0 | 0 | 0 | 1 | Ref |
| 1797 | KC787110 | Unassigned | Unassigned | Unassigned | 761_0 | Clustered | 67 | VC_761_0 | 67 | 0.8947 | 1 | 0.8947 | 0 | 0 | 0 | 1 | Ref |
| 1798 | KC787111 | Unassigned | Unassigned | Unassigned | 761_0 | Clustered | 67 | VC_761_0 | 67 | 0.8947 | 1 | 0.8947 | 0 | 0 | 0 | 1 | Ref |
| 1799 | KC787112 | Unassigned | Unassigned | Unassigned | 761_0 | Clustered | 67 | VC_761_0 | 67 | 0.8947 | 1 | 0.8947 | 0 | 0 | 0 | 1 | Ref |
| 1800 | KC801932 | Unassigned | Unassigned | Unassigned | 669_1 | Clustered | 10 | VC_669_1 | 10 | 0.1268 | 1 | 0.1268 | 0 | 0 | 0 | 0.9778 | Ref |
| 1801 | KC814930 | Unassigned | Unassigned | Unassigned | 45_0 | Clustered | 12 | VC_45_0 | 12 | 0.35 | 1 | 0.35 | 0 | 0 | 0 | 1 | Ref |
| 1802 | KC821604 | Unassigned | Unassigned | Unassigned | 929_0 | Clustered | 4 | VC_929_0 | 4 | 0.846 | 0.99722201 | 0.8437 | 0 | 0 | 0 | 1 | Ref |
| 1803 | KC821606 | Unassigned | Unassigned | Unassigned | 1011_0 | Clustered | 9 | VC_1011_0 | 9 | 1 | 1 | 1 | 0 | 0 | 0 | 1 | Ref |
| 1804 | KC821607 | Unassigned | Unassigned | Unassigned | 445_1 | Clustered | 2 | VC_445_1 | 2 | 0.1297 | 0.93055167 | 0.1207 | 0 | 0 | 0 | 1 | Ref |
| 1805 | KC821608 | Unassigned | Unassigned | Unassigned | 1012_0 | Clustered | 4 | VC_1012_0 | 4 | 0.8715 | 0.99995392 | 0.8714 | 0 | 0 | 0 | 1 | Ref |
| 1806 | KC821609 | Unassigned | Unassigned | Unassigned | 1017_0 | Clustered | 5 | VC_1017_0 | 5 | 0.9759 | 0.9999961 | 0.9759 | 0 | 0 | 0 | 1 | Ref |
| 1807 | KC821610 | Unassigned | Unassigned | Unassigned | 911_0 | Clustered | 6 | VC_911_0 | 6 | 0.2787 | 1 | 0.2787 | 0 | 0 | 0 | 1 | Ref |
| 1808 | KC821612 | Unassigned | Unassigned | Unassigned | 1019_0 | Clustered | 5 | VC_1019_0 | 5 | 0.866 | 0.99999999 | 0.866 | 0 | 0 | 0 | 1 | Ref |
| 1809 | KC821613 | Unassigned | Unassigned | Unassigned | 1020_0 | Clustered | 5 | VC_1020_0 | 5 | 0.827 | 0.9999997 | 0.827 | 0 | 0 | 0 | 1 | Ref |
| 1810 | KC821614 | Unassigned | Unassigned | Unassigned | 1019_0 | Clustered | 5 | VC_1019_0 | 5 | 0.866 | 0.99999999 | 0.866 | 0 | 0 | 0 | 1 | Ref |
| 1811 | KC821615 | Unassigned | Unassigned | Unassigned | 1020_0 | Clustered | 5 | VC_1020_0 | 5 | 0.827 | 0.9999997 | 0.827 | 0 | 0 | 0 | 1 | Ref |
| 1812 | KC821616 | Unassigned | Unassigned | Unassigned | 911_0 | Clustered | 6 | VC_911_0 | 6 | 0.2787 | 1 | 0.2787 | 0 | 0 | 0 | 1 | Ref |
| 1813 | KC821617 | Unassigned | Unassigned | Unassigned | 1020_0 | Clustered | 5 | VC_1020_0 | 5 | 0.827 | 0.9999997 | 0.827 | 0 | 0 | 0 | 1 | Ref |
| 1814 | KC821618 | Unassigned | Unassigned | Unassigned | 445_1 | Clustered | 2 | VC_445_1 | 2 | 0.1297 | 0.93055167 | 0.1207 | 0 | 0 | 0 | 1 | Ref |
| 1815 | KC821619 | Unassigned | Unassigned | Unassigned | 1020_0 | Clustered | 5 | VC_1020_0 | 5 | 0.827 | 0.9999997 | 0.827 | 0 | 0 | 0 | 1 | Ref |
| 1816 | KC821620 | Unassigned | Unassigned | Unassigned | 1012_0 | Clustered | 4 | VC_1012_0 | 4 | 0.8715 | 0.99995392 | 0.8714 | 0 | 0 | 0 | 1 | Ref |
| 1817 | KC821621 | Unassigned | Unassigned | Unassigned | 929_0 | Clustered | 4 | VC_929_0 | 4 | 0.846 | 0.99722201 | 0.8437 | 0 | 0 | 0 | 1 | Ref |
| 1818 | KC821622 | Unassigned | Unassigned | Unassigned | 1012_0 | Clustered | 4 | VC_1012_0 | 4 | 0.8715 | 0.99995392 | 0.8714 | 0 | 0 | 0 | 1 | Ref |
| 1819 | KC821623 | Unassigned | Unassigned | Unassigned | 1011_0 | Clustered | 9 | VC_1011_0 | 9 | 1 | 1 | 1 | 0 | 0 | 0 | 1 | Ref |
| 1820 | KC821625 | Unassigned | Unassigned | Unassigned | 929_0 | Clustered | 4 | VC_929_0 | 4 | 0.846 | 0.99722201 | 0.8437 | 0 | 0 | 0 | 1 | Ref |
| 1821 | KC821627 | Unassigned | Unassigned | Unassigned | 1020_0 | Clustered | 5 | VC_1020_0 | 5 | 0.827 | 0.9999997 | 0.827 | 0 | 0 | 0 | 1 | Ref |
| 1822 | KC821628 | Unassigned | Unassigned | Unassigned | 1011_0 | Clustered | 9 | VC_1011_0 | 9 | 1 | 1 | 1 | 0 | 0 | 0 | 1 | Ref |
| 1823 | KC821629 | Unassigned | Unassigned | Unassigned | 911_0 | Clustered | 6 | VC_911_0 | 6 | 0.2787 | 1 | 0.2787 | 0 | 0 | 0 | 1 | Ref |
| 1824 | KC821630 | Unassigned | Unassigned | Unassigned | 911_0 | Clustered | 6 | VC_911_0 | 6 | 0.2787 | 1 | 0.2787 | 0 | 0 | 0 | 1 | Ref |
| 1825 | KC821631 | Unassigned | Unassigned | Unassigned | 1011_0 | Clustered | 9 | VC_1011_0 | 9 | 1 | 1 | 1 | 0 | 0 | 0 | 1 | Ref |
| 1826 | KC821632 | Unassigned | Unassigned | Unassigned | 1017_0 | Clustered | 5 | VC_1017_0 | 5 | 0.9759 | 0.9999961 | 0.9759 | 0 | 0 | 0 | 1 | Ref |
| 1827 | KC821633 | Unassigned | Unassigned | Unassigned | 1012_0 | Clustered | 4 | VC_1012_0 | 4 | 0.8715 | 0.99995392 | 0.8714 | 0 | 0 | 0 | 1 | Ref |
| 1828 | KC821634 | Unassigned | Unassigned | Unassigned | 911_0 | Clustered | 6 | VC_911_0 | 6 | 0.2787 | 1 | 0.2787 | 0 | 0 | 0 | 1 | Ref |
| 1829 | KC853746 | Unassigned | Unassigned | Unassigned | 70_0 | Clustered | 27 | VC_70_0 | 27 | 0.5981 | 1 | 0.5981 | 0 | 0 | 0 | 0.9886 | Ref |
| 1830 | KC862295 | Unassigned | Unassigned | Unassigned | 77_0 | Clustered | 20 | VC_77_0 | 20 | 0.9657 | 1 | 0.9657 | 0 | 0 | 0 | 1 | Ref |
| 1831 | KC862296 | Unassigned | Unassigned | Unassigned | 77_0 | Clustered | 20 | VC_77_0 | 20 | 0.9657 | 1 | 0.9657 | 0 | 0 | 0 | 1 | Ref |
| 1832 | KC862297 | Unassigned | Unassigned | Unassigned | 76_0 | Clustered | 37 | VC_76_0 | 37 | 0.9403 | 1 | 0.9403 | 0 | 0 | 0 | 1 | Ref |
| 1833 | KC862298 | Unassigned | Unassigned | Unassigned | 76_0 | Clustered | 37 | VC_76_0 | 37 | 0.9403 | 1 | 0.9403 | 0 | 0 | 0 | 1 | Ref |
| 1834 | KC862299 | Unassigned | Unassigned | Unassigned | 77_0 | Clustered | 20 | VC_77_0 | 20 | 0.9657 | 1 | 0.9657 | 0 | 0 | 0 | 1 | Ref |
| 1835 | KC862300 | Unassigned | Unassigned | Unassigned | 76_0 | Clustered | 37 | VC_76_0 | 37 | 0.9403 | 1 | 0.9403 | 0 | 0 | 0 | 1 | Ref |
| 1836 | KC862301 | Unassigned | Unassigned | Unassigned | 77_0 | Clustered | 20 | VC_77_0 | 20 | 0.9657 | 1 | 0.9657 | 0 | 0 | 0 | 1 | Ref |
| 1837 | KC900378 | Unassigned | Unassigned | Unassigned | 630_0 | Clustered | 20 | VC_630_0 | 20 | 0.8513 | 1 | 0.8513 | 0 | 0 | 0 | 1 | Ref |
| 1838 | KC900379 | Unassigned | Unassigned | Unassigned | 763_1 | Clustered | 9 | VC_763_1 | 9 | 0.3706 | 1 | 0.3706 | 0 | 0 | 0 | 1 | Ref |
| 1839 | KC911856 | Unassigned | Unassigned | Unassigned | 476_0 | Clustered | 19 | VC_476_0 | 19 | 0.7814 | 1 | 0.7814 | 0 | 0 | 0 | 1 | Ref |
| 1840 | KC911857 | Unassigned | Unassigned | Unassigned | 476_0 | Clustered | 19 | VC_476_0 | 19 | 0.7814 | 1 | 0.7814 | 0 | 0 | 0 | 1 | Ref |
| 1841 | KC954774 | Unassigned | Unassigned | Unassigned | 849_5 | Clustered | 3 | VC_849_5 | 3 | 0.0492 | 0.97574727 | 0.048 | 0 | 0 | 0 | 1 | Ref |
| 1842 | KC954775 | Unassigned | Unassigned | Unassigned | 580_0 | Clustered | 2 | VC_580_0 | 2 | 0.0632 | 0.95763305 | 0.0605 | 0 | 0 | 0 | 1 | Ref |
| 1843 | KC959568 | Unassigned | Unassigned | Unassigned | 1023_0 | Clustered | 8 | VC_1023_0 | 8 | 0.8574 | 1 | 0.8574 | 0 | 0 | 0 | 1 | Ref |
| 1844 | KC960489 | Unassigned | Unassigned | Unassigned | 1024_0 | Clustered | 2 | VC_1024_0 | 2 | 1 | 1 | 1 | 0 | 0 | 0 | 1 | Ref |
| 1845 | KC960671 | Unassigned | Unassigned | Unassigned | 385_0 | Clustered | 51 | VC_385_0 | 51 | 0.8754 | 1 | 0.8754 | 0 | 0 | 0 | 1 | Ref |
| 1846 | KC969441 | Unassigned | Unassigned | Unassigned | 423_0 | Clustered | 45 | VC_423_0 | 45 | 0.9484 | 1 | 0.9484 | 0 | 0 | 0 | 1 | Ref |
| 1847 | KF005317 | Unassigned | Unassigned | Unassigned | 1025_0 | Clustered | 4 | VC_1025_0 | 4 | 1 | 0 | 0 | 0 | 0 | 0 | 1 | Ref |
| 1848 | KF005318 | Unassigned | Unassigned | Unassigned | 1025_0 | Clustered | 4 | VC_1025_0 | 4 | 1 | 0 | 0 | 0 | 0 | 0 | 1 | Ref |
| 1849 | KF005319 | Unassigned | Unassigned | Unassigned | 1025_0 | Clustered | 4 | VC_1025_0 | 4 | 1 | 0 | 0 | 0 | 0 | 0 | 1 | Ref |
| 1850 | KF005320 | Unassigned | Unassigned | Unassigned | 1025_0 | Clustered | 4 | VC_1025_0 | 4 | 1 | 0 | 0 | 0 | 0 | 0 | 1 | Ref |
| 1851 | KF006817 | Unassigned | Unassigned | Unassigned | 558_9 | Clustered | 3 | VC_558_9 | 3 | 0.0064 | 0.82911529 | 0.0053 | 0 | 0 | 0 | 0.6667 | Ref |
| 1852 | KF006818 | Unassigned | Unassigned | Unassigned | 546_6 | Clustered | 2 | VC_546_6 | 2 | 0.0135 | 0.7810929 | 0.0105 | 0 | 0 | 0 | 1 | Ref |
| 1853 | KF010834 | Unassigned | Unassigned | Unassigned | 708_0 | Clustered | 47 | VC_708_0 | 47 | 0.8654 | 1 | 0.8654 | 0 | 0 | 0 | 1 | Ref |
| 1854 | KF017001 | Unassigned | Unassigned | Unassigned | 940_0 | Clustered | 34 | VC_940_0 | 34 | 0.7959 | 1 | 0.7959 | 0 | 0 | 0 | 1 | Ref |
| 1855 | KF017002 | Unassigned | Unassigned | Unassigned | 556_0 | Clustered | 21 | VC_556_0 | 21 | 0.9912 | 1 | 0.9912 | 0 | 0 | 0 | 1 | Ref |
| 1856 | KF017927 | Unassigned | Unassigned | Unassigned | 215_0 | Clustered | 5 | VC_215_0 | 5 | 0.0724 | 0.99999997 | 0.0724 | 0 | 0 | 0 | 1 | Ref |
| 1857 | KF021268 | Unassigned | Unassigned | Unassigned | 150_0 | Clustered | 13 | VC_150_0 | 13 | 0.2417 | 1 | 0.2417 | 0 | 0 | 0 | 0.9872 | Ref |
| 1858 | KF024721 | Unassigned | Unassigned | Unassigned | 216_3 | Clustered | 8 | VC_216_3 | 8 | 0.0701 | 1 | 0.0701 | 0 | 0 | 0 | 0.8571 | Ref |
| 1859 | KF024722 | Unassigned | Unassigned | Unassigned | 508_0 | Clustered | 32 | VC_508_0 | 32 | 0.906 | 1 | 0.906 | 0 | 0 | 0 | 1 | Ref |
| 1860 | KF024723 | Unassigned | Unassigned | Unassigned | 520_23 | Clustered | 2 | VC_520_23 | 2 | 0.0077 | 0.95763408 | 0.0074 | 0 | 0 | 0 | 1 | Ref |
| 1861 | KF024724 | Unassigned | Unassigned | Unassigned | 325_35 | Clustered | 17 | VC_325_35 | 17 | 0.0732 | 1 | 0.0732 | 0 | 0 | 0 | 0.7721 | Ref |
| 1862 | KF024725 | Unassigned | Unassigned | Unassigned | 889_0 | Clustered | 16 | VC_889_0 | 16 | 0.2969 | 1 | 0.2969 | 0 | 0 | 0 | 1 | Ref |
| 1863 | KF024726 | Unassigned | Unassigned | Unassigned | 325_8 | Clustered | 2 | VC_325_8 | 2 | 0.0093 | 0.95751268 | 0.0089 | 0 | 0 | 0 | 1 | Ref |
| 1864 | KF024727 | Unassigned | Unassigned | Unassigned | 372_1 | Clustered | 9 | VC_372_1 | 9 | 0.194 | 1 | 0.194 | 0 | 0 | 0 | 1 | Ref |
| 1865 | KF024728 | Unassigned | Unassigned | Unassigned | 574_0 | Clustered | 7 | VC_574_0 | 7 | 0.2734 | 1 | 0.2734 | 0 | 0 | 0 | 1 | Ref |
| 1866 | KF024729 | Unassigned | Unassigned | Unassigned | 372_3 | Clustered | 5 | VC_372_3 | 5 | 0.1129 | 0.99999997 | 0.1129 | 0 | 0 | 0 | 1 | Ref |
| 1867 | KF024730 | Unassigned | Unassigned | Unassigned | 556_0 | Clustered | 21 | VC_556_0 | 21 | 0.9912 | 1 | 0.9912 | 0 | 0 | 0 | 1 | Ref |
| 1868 | KF024731 | Unassigned | Unassigned | Unassigned | 888_0 | Clustered | 28 | VC_888_0 | 28 | 0.5497 | 1 | 0.5497 | 0 | 0 | 0 | 1 | Ref |
| 1869 | KF024732 | Unassigned | Unassigned | Unassigned | 551_8 | Clustered | 19 | VC_551_8 | 19 | 0.1039 | 1 | 0.1039 | 0 | 0 | 0 | 0.8713 | Ref |
| 1870 | KF024733 | Unassigned | Unassigned | Unassigned | 517_1 | Clustered | 108 | VC_517_1 | 108 | 0.8483 | 1 | 0.8483 | 0 | 0 | 0 | 0.9955 | Ref |
| 1871 | KF024734 | Unassigned | Unassigned | Unassigned | 558_48 | Clustered | 2 | VC_558_48 | 2 | 0.0032 | 0.69466126 | 0.0022 | 0 | 0 | 0 | 1 | Ref |
| 1872 | KF055347 | Unassigned | Unassigned | Unassigned | 329_16 | Clustered | 4 | VC_329_16 | 4 | 0.0158 | 0.9999807 | 0.0158 | 0 | 0 | 0 | 1 | Ref |
| 1873 | KF056323 | Unassigned | Unassigned | Unassigned | 834_0 | Clustered | 7 | VC_834_0 | 7 | 0.9845 | 0.94276893 | 0.9282 | 0 | 0 | 0 | 1 | Ref |
| 1874 | KF114874 | Unassigned | Unassigned | Unassigned | 520_9 | Clustered/Singleton | 1 | VC_520_9 | 1 | 0 | 1 | 0 | 0 | 0 | 0 | 1 | Ref |
| 1875 | KF114875 | Unassigned | Unassigned | Unassigned | 546_4 | Clustered | 16 | VC_546_4 | 16 | 0.2517 | 0.9999998 | 0.2517 | 0 | 0 | 0 | 0.9417 | Ref |
| 1876 | KF114876 | Unassigned | Unassigned | Unassigned | 687_1 | Clustered | 2 | VC_687_1 | 2 | 0.223 | 0.91418477 | 0.2038 | 0 | 0 | 0 | 1 | Ref |
| 1877 | KF114877 | Unassigned | Unassigned | Unassigned | 687_1 | Clustered | 2 | VC_687_1 | 2 | 0.223 | 0.91418477 | 0.2038 | 0 | 0 | 0 | 1 | Ref |
| 1878 | KF114879 | Unassigned | Unassigned | Unassigned | 687_0 | Clustered | 3 | VC_687_0 | 3 | 0.5872 | 0.99252665 | 0.5828 | 0 | 0 | 0 | 1 | Ref |
| 1879 | KF114880 | Unassigned | Unassigned | Unassigned | 687_0 | Clustered | 3 | VC_687_0 | 3 | 0.5872 | 0.99252665 | 0.5828 | 0 | 0 | 0 | 1 | Ref |
| 1880 | KF148055 | Unassigned | Unassigned | Unassigned | 637_2 | Clustered | 2 | VC_637_2 | 2 | 0.0038 | 0.92844927 | 0.0035 | 0 | 0 | 0 | 1 | Ref |
| 1881 | KF148616 | Unassigned | Unassigned | Unassigned | 844_0 | Clustered | 27 | VC_844_0 | 27 | 0.9385 | 1 | 0.9385 | 0 | 0 | 0 | 1 | Ref |
| 1882 | KF156338 | Unassigned | Unassigned | Unassigned | 433_2 | Clustered/Singleton | 1 | VC_433_2 | 1 | 0 | 1 | 0 | 0 | 0 | 0 | 1 | Ref |
| 1883 | KF188410 | Unassigned | Unassigned | Unassigned | 640_0 | Clustered | 7 | VC_640_0 | 7 | 0.9025 | 1 | 0.9025 | 0 | 0 | 0 | 1 | Ref |
| 1884 | KF188414 | Unassigned | Unassigned | Unassigned | 551_2 | Clustered | 17 | VC_551_2 | 17 | 0.0973 | 1 | 0.0973 | 0 | 0 | 0 | 0.9779 | Ref |
| 1885 | KF192053 | Unassigned | Unassigned | Unassigned | 108_0 | Clustered | 33 | VC_108_0 | 33 | 0.949 | 1 | 0.949 | 0 | 0 | 0 | 1 | Ref |
| 1886 | KF192075 | Unassigned | Unassigned | Unassigned | 779_0 | Clustered | 37 | VC_779_0 | 37 | 0.5682 | 1 | 0.5682 | 0 | 0 | 0 | 1 | Ref |
| 1887 | KF208315 | Unassigned | Unassigned | Unassigned | 283_90 | Clustered/Singleton | 1 | VC_283_90 | 1 | 0 | 1 | 0 | 0 | 0 | 0 | 1 | Ref |
| 1888 | KF208639 | Unassigned | Unassigned | Unassigned | 883_1 | Clustered | 12 | VC_883_1 | 12 | 0.2441 | 1 | 0.2441 | 0 | 0 | 0 | 1 | Ref |
| 1889 | KF279411 | Unassigned | Unassigned | Unassigned | 372_0 | Clustered | 24 | VC_372_0 | 24 | 0.5539 | 1 | 0.5539 | 0 | 0 | 0 | 1 | Ref |
| 1890 | KF279412 | Unassigned | Unassigned | Unassigned | 372_0 | Clustered | 24 | VC_372_0 | 24 | 0.5539 | 1 | 0.5539 | 0 | 0 | 0 | 1 | Ref |
| 1891 | KF279413 | Unassigned | Unassigned | Unassigned | 372_0 | Clustered | 24 | VC_372_0 | 24 | 0.5539 | 1 | 0.5539 | 0 | 0 | 0 | 1 | Ref |
| 1892 | KF279415 | Unassigned | Unassigned | Unassigned | 325_35 | Clustered | 17 | VC_325_35 | 17 | 0.0732 | 1 | 0.0732 | 0 | 0 | 0 | 0.7721 | Ref |
| 1893 | KF279416 | Unassigned | Unassigned | Unassigned | 325_35 | Clustered | 17 | VC_325_35 | 17 | 0.0732 | 1 | 0.0732 | 0 | 0 | 0 | 0.7721 | Ref |
| 1894 | KF279417 | Unassigned | Unassigned | Unassigned | 551_7 | Clustered | 28 | VC_551_7 | 28 | 0.1661 | 1 | 0.1661 | 0 | 0 | 0 | 0.8519 | Ref |
| 1895 | KF279418 | Unassigned | Unassigned | Unassigned | 514_0 | Clustered | 75 | VC_514_0 | 75 | 0.9402 | 1 | 0.9402 | 0 | 0 | 0 | 1 | Ref |
| 1896 | KF296717 | Unassigned | Unassigned | Unassigned | 829_0 | Clustered | 5 | VC_829_0 | 5 | 0.8821 | 0.99999832 | 0.8821 | 0 | 0 | 0 | 1 | Ref |
| 1897 | KF302032 | Unassigned | Unassigned | Unassigned | 894_0 | Clustered | 3 | VC_894_0 | 3 | 0.6358 | 0.99692152 | 0.6338 | 0 | 0 | 0 | 1 | Ref |
| 1898 | KF302033 | Unassigned | Unassigned | Unassigned | 894_0 | Clustered | 3 | VC_894_0 | 3 | 0.6358 | 0.99692152 | 0.6338 | 0 | 0 | 0 | 1 | Ref |
| 1899 | KF302034 | Unassigned | Unassigned | Unassigned | 496_0 | Clustered/Singleton | 1 | VC_496_0 | 1 | 0 | 1 | 0 | 0 | 0 | 0 | 1 | Ref |
| 1900 | KF302035 | Unassigned | Unassigned | Unassigned | 183_0 | Clustered | 2 | VC_183_0 | 2 | 0.0667 | 0.95489893 | 0.0637 | 0 | 0 | 0 | 1 | Ref |
| 1901 | KF302036 | Unassigned | Unassigned | Unassigned | 895_0 | Clustered | 2 | VC_895_0 | 2 | 0.2788 | 0.94251254 | 0.2628 | 0 | 0 | 0 | 1 | Ref |
| 1902 | KF302037 | Unassigned | Unassigned | Unassigned | 756_0 | Clustered | 2 | VC_756_0 | 2 | 0.1637 | 0.95427014 | 0.1562 | 0 | 0 | 0 | 1 | Ref |
| 1903 | KF306380 | Unassigned | Unassigned | Unassigned | 551_3 | Clustered/Singleton | 1 | VC_551_3 | 1 | 0 | 1 | 0 | 0 | 0 | 0 | 1 | Ref |
| 1904 | KF319020 | Unassigned | Unassigned | Unassigned | 112_0 | Clustered | 8 | VC_112_0 | 8 | 0.2537 | 1 | 0.2537 | 0 | 0 | 0 | 1 | Ref |
| 1905 | KF322026 | Unassigned | Unassigned | Unassigned | 1027_0 | Clustered | 20 | VC_1027_0 | 20 | 0.7114 | 1 | 0.7114 | 0 | 0 | 0 | 1 | Ref |
| 1906 | KF322032 | Unassigned | Unassigned | Unassigned | 228_0 | Clustered | 60 | VC_228_0 | 60 | 0.942 | 1 | 0.942 | 0 | 0 | 0 | 1 | Ref |
| 1907 | KF356199 | Unassigned | Unassigned | Unassigned | 38_0 | Clustered | 3 | VC_38_0 | 3 | 1 | 0 | 0 | 0 | 0 | 0 | 1 | Ref |
| 1908 | KF361475 | Unassigned | Unassigned | Unassigned | 5_0 | Clustered | 30 | VC_5_0 | 30 | 0.9647 | 1 | 0.9647 | 0 | 0 | 0 | 1 | Ref |
| 1909 | KF381361 | Unassigned | Unassigned | Unassigned | 437_0 | Clustered | 4 | VC_437_0 | 4 | 0.552 | 0.99998233 | 0.552 | 0 | 0 | 0 | 1 | Ref |
| 1910 | KF416340 | Unassigned | Unassigned | Unassigned | 325_49 | Clustered | 4 | VC_325_49 | 4 | 0.014 | 0.99995965 | 0.014 | 0 | 0 | 0 | 1 | Ref |
| 1911 | KF416341 | Unassigned | Unassigned | Unassigned | 372_1 | Clustered | 9 | VC_372_1 | 9 | 0.194 | 1 | 0.194 | 0 | 0 | 0 | 1 | Ref |
| 1912 | KF416342 | Unassigned | Unassigned | Unassigned | 568_0 | Clustered | 8 | VC_568_0 | 8 | 0.5738 | 1 | 0.5738 | 0 | 0 | 0 | 1 | Ref |
| 1913 | KF416343 | Unassigned | Unassigned | Unassigned | 551_4 | Clustered | 2 | VC_551_4 | 2 | 0.006 | 0.95695857 | 0.0058 | 0 | 0 | 0 | 1 | Ref |
| 1914 | KF416344 | Unassigned | Unassigned | Unassigned | 215_0 | Clustered | 5 | VC_215_0 | 5 | 0.0724 | 0.99999997 | 0.0724 | 0 | 0 | 0 | 1 | Ref |
| 1915 | KF475786 | Unassigned | Unassigned | Unassigned | 179_0 | Clustered | 53 | VC_179_0 | 53 | 0.9137 | 1 | 0.9137 | 0 | 0 | 0 | 1 | Ref |
| 1916 | KF493879 | Unassigned | Unassigned | Unassigned | 509_0 | Clustered | 37 | VC_509_0 | 37 | 0.9689 | 1 | 0.9689 | 0 | 0 | 0 | 1 | Ref |
| 1917 | KF493880 | Unassigned | Unassigned | Unassigned | 325_3 | Clustered/Singleton | 1 | VC_325_3 | 1 | 0 | 1 | 0 | 0 | 0 | 0 | 1 | Ref |
| 1918 | KF493881 | Unassigned | Unassigned | Unassigned | 372_0 | Clustered | 24 | VC_372_0 | 24 | 0.5539 | 1 | 0.5539 | 0 | 0 | 0 | 1 | Ref |
| 1919 | KF493882 | Unassigned | Unassigned | Unassigned | 940_0 | Clustered | 34 | VC_940_0 | 34 | 0.7959 | 1 | 0.7959 | 0 | 0 | 0 | 1 | Ref |
| 1920 | KF493883 | Unassigned | Unassigned | Unassigned | 551_8 | Clustered | 19 | VC_551_8 | 19 | 0.1039 | 1 | 0.1039 | 0 | 0 | 0 | 0.8713 | Ref |
| 1921 | KF534715 | Unassigned | Unassigned | Unassigned | 792_0 | Clustered | 35 | VC_792_0 | 35 | 0.9828 | 1 | 0.9828 | 0 | 0 | 0 | 1 | Ref |
| 1922 | KF550303 | Unassigned | Unassigned | Unassigned | 582_4 | Clustered | 3 | VC_582_4 | 3 | 0.0348 | 0.98173503 | 0.0341 | 0 | 0 | 0 | 1 | Ref |
| 1923 | KF554508 | Unassigned | Unassigned | Unassigned | 87_1 | Clustered | 3 | VC_87_1 | 3 | 0.1636 | 0.99777924 | 0.1632 | 0 | 0 | 0 | 1 | Ref |
| 1924 | KF560330 | Unassigned | Unassigned | Unassigned | 940_0 | Clustered | 34 | VC_940_0 | 34 | 0.7959 | 1 | 0.7959 | 0 | 0 | 0 | 1 | Ref |
| 1925 | KF560331 | Unassigned | Unassigned | Unassigned | 325_33 | Clustered | 4 | VC_325_33 | 4 | 0.0149 | 0.99998565 | 0.0149 | 0 | 0 | 0 | 1 | Ref |
| 1926 | KF560332 | Unassigned | Unassigned | Unassigned | 223_0 | Clustered | 38 | VC_223_0 | 38 | 0.9095 | 1 | 0.9095 | 0 | 0 | 0 | 1 | Ref |
| 1927 | KF560333 | Unassigned | Unassigned | Unassigned | 223_0 | Clustered | 38 | VC_223_0 | 38 | 0.9095 | 1 | 0.9095 | 0 | 0 | 0 | 1 | Ref |
| 1928 | KF560334 | Unassigned | Unassigned | Unassigned | 223_0 | Clustered | 38 | VC_223_0 | 38 | 0.9095 | 1 | 0.9095 | 0 | 0 | 0 | 1 | Ref |
| 1929 | KF562099 | Unassigned | Unassigned | Unassigned | 551_8 | Clustered | 19 | VC_551_8 | 19 | 0.1039 | 1 | 0.1039 | 0 | 0 | 0 | 0.8713 | Ref |
| 1930 | KF562100 | Unassigned | Unassigned | Unassigned | 551_8 | Clustered | 19 | VC_551_8 | 19 | 0.1039 | 1 | 0.1039 | 0 | 0 | 0 | 0.8713 | Ref |
| 1931 | KF562101 | Unassigned | Unassigned | Unassigned | 551_8 | Clustered | 19 | VC_551_8 | 19 | 0.1039 | 1 | 0.1039 | 0 | 0 | 0 | 0.8713 | Ref |
| 1932 | KF562102 | Unassigned | Unassigned | Unassigned | 551_8 | Clustered | 19 | VC_551_8 | 19 | 0.1039 | 1 | 0.1039 | 0 | 0 | 0 | 0.8713 | Ref |
| 1933 | KF562340 | Unassigned | Unassigned | Unassigned | 779_0 | Clustered | 37 | VC_779_0 | 37 | 0.5682 | 1 | 0.5682 | 0 | 0 | 0 | 1 | Ref |
| 1934 | KF562341 | Unassigned | Unassigned | Unassigned | 286_10 | Clustered | 9 | VC_286_10 | 9 | 0.0615 | 0.99999878 | 0.0615 | 0 | 0 | 0 | 0.6667 | Ref |
| 1935 | KF562864 | Unassigned | Unassigned | Unassigned | 637_1 | Clustered | 27 | VC_637_1 | 27 | 0.2578 | 1 | 0.2578 | 0 | 0 | 0 | 0.9858 | Ref |
| 1936 | KF562865 | Unassigned | Unassigned | Unassigned | 637_1 | Clustered | 27 | VC_637_1 | 27 | 0.2578 | 1 | 0.2578 | 0 | 0 | 0 | 0.9858 | Ref |
| 1937 | KF582788 | Unassigned | Unassigned | Unassigned | 286_13 | Clustered | 10 | VC_286_13 | 10 | 0.0698 | 0.99999929 | 0.0698 | 0 | 0 | 0 | 0.9556 | Ref |
| 1938 | KF591601 | Unassigned | Unassigned | Unassigned | 705_0 | Clustered | 11 | VC_705_0 | 11 | 0.8861 | 1 | 0.8861 | 0 | 0 | 0 | 1 | Ref |
| 1939 | KF598856 | Unassigned | Unassigned | Unassigned | 8_0 | Clustered | 58 | VC_8_0 | 58 | 0.8306 | 1 | 0.8306 | 0 | 0 | 0 | 0.9946 | Ref |
| 1940 | KF598865 | Unassigned | Unassigned | Unassigned | 777_0 | Clustered | 3 | VC_777_0 | 3 | 0.4763 | 0.9961812 | 0.4745 | 0 | 0 | 0 | 1 | Ref |
| 1941 | KF614509 | Unassigned | Unassigned | Unassigned | 990_0 | Clustered | 3 | VC_990_0 | 3 | 0.6363 | 0.99819562 | 0.6351 | 0 | 0 | 0 | 1 | Ref |
| 1942 | KF620435 | Unassigned | Unassigned | Unassigned | 779_0 | Clustered | 37 | VC_779_0 | 37 | 0.5682 | 1 | 0.5682 | 0 | 0 | 0 | 1 | Ref |

| | | | | | | | | | | | | | | | | | |
|---|---|---|---|---|---|---|---|---|---|---|---|---|---|---|---|---|---|
| 1943 | KF623293 | Unassigned | Unassigned | Unassigned | 901_0 | Clustered | 5 | VC_901_0 | 5 | 0.9265 | 0.99999497 | 0.9265 | 0 | 0 | 0 | 1 | Ref |
| 1944 | KF623294 | Unassigned | Unassigned | Unassigned | 488_2 | Clustered | 2 | VC_488_2 | 2 | 0.0325 | 0.95511029 | 0.0311 | 0 | 0 | 0 | 1 | Ref |
| 1945 | KF626665 | Unassigned | Unassigned | Unassigned | 172_0 | Clustered | 5 | VC_172_0 | 5 | 0.3579 | 0.99999997 | 0.3579 | 0 | 0 | 0 | 1 | Ref |
| 1946 | KF626667 | Unassigned | Unassigned | Unassigned | 74_0 | Clustered | 15 | VC_74_0 | 15 | 0.5204 | 1 | 0.5204 | 0 | 0 | 0 | 1 | Ref |
| 1947 | KF626668 | Unassigned | Unassigned | Unassigned | 172_0 | Clustered | 5 | VC_172_0 | 5 | 0.3579 | 0.99999997 | 0.3579 | 0 | 0 | 0 | 1 | Ref |
| 1948 | KF669647 | Unassigned | Unassigned | Unassigned | 883_1 | Clustered | 12 | VC_883_1 | 12 | 0.2441 | 1 | 0.2441 | 0 | 0 | 0 | 1 | Ref |
| 1949 | KF669648 | Unassigned | Unassigned | Unassigned | 100_0 | Clustered | 15 | VC_100_0 | 15 | 0.6536 | 1 | 0.6536 | 0 | 0 | 0 | 1 | Ref |
| 1950 | KF669649 | Unassigned | Unassigned | Unassigned | 87_0 | Clustered | 8 | VC_87_0 | 8 | 0.5545 | 1 | 0.5545 | 0 | 0 | 0 | 1 | Ref |
| 1951 | KF669650 | Unassigned | Unassigned | Unassigned | 855_0 | Clustered | 2 | VC_855_0 | 2 | 0.0285 | 0.89378929 | 0.0255 | 0 | 0 | 0 | 1 | Ref |
| 1952 | KF669651 | Unassigned | Unassigned | Unassigned | 100_0 | Clustered | 15 | VC_100_0 | 15 | 0.6536 | 1 | 0.6536 | 0 | 0 | 0 | 1 | Ref |
| 1953 | KF669652 | Unassigned | Unassigned | Unassigned | 85_3 | Clustered | 19 | VC_85_3 | 19 | 0.3671 | 1 | 0.3671 | 0 | 0 | 0 | 0.807 | Ref |
| 1954 | KF669653 | Unassigned | Unassigned | Unassigned | 133_0 | Clustered | 2 | VC_133_0 | 2 | 0.0063 | 0.94038884 | 0.0059 | 0 | 0 | 0 | 1 | Ref |
| 1955 | KF669654 | Unassigned | Unassigned | Unassigned | 133_0 | Clustered | 2 | VC_133_0 | 2 | 0.0063 | 0.94038884 | 0.0059 | 0 | 0 | 0 | 1 | Ref |
| 1956 | KF669655 | Unassigned | Unassigned | Unassigned | 1032_0 | Clustered | 7 | VC_1032_0 | 7 | 0.9887 | 0.99999504 | 0.9887 | 0 | 0 | 0 | 1 | Ref |
| 1957 | KF669657 | Unassigned | Unassigned | Unassigned | 1032_0 | Clustered | 7 | VC_1032_0 | 7 | 0.9887 | 0.99999504 | 0.9887 | 0 | 0 | 0 | 1 | Ref |
| 1958 | KF669659 | Unassigned | Unassigned | Unassigned | 100_0 | Clustered | 15 | VC_100_0 | 15 | 0.6536 | 1 | 0.6536 | 0 | 0 | 0 | 1 | Ref |
| 1959 | KF669660 | Unassigned | Unassigned | Unassigned | 1032_0 | Clustered | 7 | VC_1032_0 | 7 | 0.9887 | 0.99999504 | 0.9887 | 0 | 0 | 0 | 1 | Ref |
| 1960 | KF669662 | Unassigned | Unassigned | Unassigned | 883_3 | Clustered | 2 | VC_883_3 | 2 | 0.0182 | 0.87900236 | 0.016 | 0 | 0 | 0 | 1 | Ref |
| 1961 | KF676640 | Unassigned | Unassigned | Unassigned | 202_0 | Clustered | 102 | VC_202_0 | 102 | 0.4049 | 1 | 0.4049 | 0 | 0 | 0 | 0.7876 | Ref |
| 1962 | KF692088 | Unassigned | Unassigned | Unassigned | 1034_1 | Clustered/Singleton | 1 | VC_1034_1 | 1 | 0 | 1 | 0 | 0 | 0 | 0 | 1 | Ref |
| 1963 | KF700371 | Unassigned | Unassigned | Unassigned | 386_0 | Clustered | 49 | VC_386_0 | 49 | 0.7941 | 1 | 0.7941 | 0 | 0 | 0 | 1 | Ref |
| 1964 | KF713485 | Unassigned | Unassigned | Unassigned | 371_3 | Clustered | 22 | VC_371_3 | 22 | 0.0787 | 1 | 0.0787 | 0 | 0 | 0 | 0.9048 | Ref |
| 1965 | KF713486 | Unassigned | Unassigned | Unassigned | 237_0 | Clustered | 4 | VC_237_0 | 4 | 0.0429 | 0.99998759 | 0.0429 | 0 | 0 | 0 | 1 | Ref |
| 1966 | KF728385 | Unassigned | Unassigned | Unassigned | 695_0 | Clustered | 48 | VC_695_0 | 48 | 0.8162 | 1 | 0.8162 | 0 | 0 | 0 | 1 | Ref |
| 1967 | KF733017 | Unassigned | Unassigned | Unassigned | 695_0 | Clustered | 48 | VC_695_0 | 48 | 0.8162 | 1 | 0.8162 | 0 | 0 | 0 | 1 | Ref |
| 1968 | KF751793 | Unassigned | Unassigned | Unassigned | 1039_0 | Clustered | 5 | VC_1039_0 | 5 | 1 | 1 | 1 | 0 | 0 | 0 | 1 | Ref |
| 1969 | KF751794 | Unassigned | Unassigned | Unassigned | 1039_0 | Clustered | 5 | VC_1039_0 | 5 | 1 | 1 | 1 | 0 | 0 | 0 | 1 | Ref |
| 1970 | KF751795 | Unassigned | Unassigned | Unassigned | 1039_0 | Clustered | 5 | VC_1039_0 | 5 | 1 | 1 | 1 | 0 | 0 | 0 | 1 | Ref |
| 1971 | KF751796 | Unassigned | Unassigned | Unassigned | 1039_0 | Clustered | 5 | VC_1039_0 | 5 | 1 | 1 | 1 | 0 | 0 | 0 | 1 | Ref |
| 1972 | KF751797 | Unassigned | Unassigned | Unassigned | 1039_0 | Clustered | 5 | VC_1039_0 | 5 | 1 | 1 | 1 | 0 | 0 | 0 | 1 | Ref |
| 1973 | KF765493 | Unassigned | Unassigned | Unassigned | 111_0 | Clustered | 91 | VC_111_0 | 91 | 0.9587 | 1 | 0.9587 | 0 | 0 | 0 | 1 | Ref |
| 1974 | KF766114 | Unassigned | Unassigned | Unassigned | 149_8 | Clustered | 66 | VC_149_8 | 66 | 0.4565 | 1 | 0.4565 | 0 | 0 | 0 | 0.8182 | Ref |
| 1975 | KF766125 | Unassigned | Unassigned | Unassigned | 46_0 | Clustered | 2 | VC_46_0 | 2 | 0.0181 | 0.95763537 | 0.0173 | 0 | 0 | 0 | 1 | Ref |
| 1976 | KF767351 | Unassigned | Unassigned | Unassigned | 308_0 | Clustered | 4 | VC_308_0 | 4 | 0.5014 | 0.99997418 | 0.5013 | 0 | 0 | 0 | 1 | Ref |
| 1977 | KF771236 | Unassigned | Unassigned | Unassigned | 455_0 | Clustered | 18 | VC_455_0 | 18 | 0.4598 | 1 | 0.4598 | 0 | 0 | 0 | 1 | Ref |
| 1978 | KF771237 | Unassigned | Unassigned | Unassigned | 455_0 | Clustered | 18 | VC_455_0 | 18 | 0.4598 | 1 | 0.4598 | 0 | 0 | 0 | 1 | Ref |
| 1979 | KF771238 | Unassigned | Unassigned | Unassigned | 455_0 | Clustered | 18 | VC_455_0 | 18 | 0.4598 | 1 | 0.4598 | 0 | 0 | 0 | 1 | Ref |
| 1980 | KF771239 | Unassigned | Unassigned | Unassigned | 455_0 | Clustered | 18 | VC_455_0 | 18 | 0.4598 | 1 | 0.4598 | 0 | 0 | 0 | 1 | Ref |
| 1981 | KF787094 | Unassigned | Unassigned | Unassigned | 779_1 | Clustered | 11 | VC_779_1 | 11 | 0.1587 | 1 | 0.1587 | 0 | 0 | 0 | 1 | Ref |
| 1982 | KF787095 | Unassigned | Unassigned | Unassigned | 779_1 | Clustered | 11 | VC_779_1 | 11 | 0.1587 | 1 | 0.1587 | 0 | 0 | 0 | 1 | Ref |
| 1983 | KF800937 | Unassigned | Unassigned | Unassigned | 1027_0 | Clustered | 20 | VC_1027_0 | 20 | 0.7114 | 1 | 0.7114 | 0 | 0 | 0 | 1 | Ref |
| 1984 | KF806588 | Unassigned | Unassigned | Unassigned | 779_2 | Clustered | 5 | VC_779_2 | 5 | 0.0876 | 0.99999997 | 0.0876 | 0 | 0 | 0 | 1 | Ref |
| 1985 | KF806589 | Unassigned | Unassigned | Unassigned | 489_1 | Clustered | 11 | VC_489_1 | 11 | 0.4621 | 1 | 0.4621 | 0 | 0 | 0 | 1 | Ref |
| 1986 | KF831354 | Unassigned | Unassigned | Unassigned | 10_0 | Clustered | 25 | VC_10_0 | 25 | 0.3467 | 1 | 0.3467 | 0 | 0 | 0 | 0.9733 | Ref |
| 1987 | KF835987 | Unassigned | Unassigned | Unassigned | 490_2 | Clustered/Singleton | 1 | VC_490_2 | 1 | 0 | 1 | 0 | 0 | 0 | 0 | 1 | Ref |
| 1988 | KF841475 | Unassigned | Unassigned | Unassigned | 517_1 | Clustered | 108 | VC_517_1 | 108 | 0.8483 | 1 | 0.8483 | 0 | 0 | 0 | 0.9955 | Ref |
| 1989 | KF841476 | Unassigned | Unassigned | Unassigned | 517_1 | Clustered | 108 | VC_517_1 | 108 | 0.8483 | 1 | 0.8483 | 0 | 0 | 0 | 0.9955 | Ref |
| 1990 | KF841477 | Unassigned | Unassigned | Unassigned | 519_0 | Clustered | 39 | VC_519_0 | 39 | 0.8156 | 1 | 0.8156 | 0 | 0 | 0 | 1 | Ref |
| 1991 | KF854250 | Unassigned | Unassigned | Unassigned | 498_0 | Clustered | 8 | VC_498_0 | 8 | 0.3987 | 1 | 0.3987 | 0 | 0 | 0 | 1 | Ref |
| 1992 | KF856712 | Unassigned | Unassigned | Unassigned | 472_0 | Clustered | 47 | VC_472_0 | 47 | 0.9025 | 1 | 0.9025 | 0 | 0 | 0 | 1 | Ref |
| 1993 | KF861510 | Unassigned | Unassigned | Unassigned | 218_0 | Clustered | 3 | VC_218_0 | 3 | 0.0481 | 0.99860979 | 0.0481 | 0 | 0 | 0 | 1 | Ref |
| 1994 | KF925357 | Unassigned | Unassigned | Unassigned | 283_163 | Clustered | 2 | VC_283_163 | 2 | 0.0029 | 0.93420746 | 0.0027 | 0 | 0 | 0 | 1 | Ref |
| 1995 | KF926093 | Unassigned | Unassigned | Unassigned | 474_0 | Clustered | 14 | VC_474_0 | 14 | 0.8466 | 1 | 0.8466 | 0 | 0 | 0 | 1 | Ref |
| 1996 | KF929199 | Unassigned | Unassigned | Unassigned | 667_0 | Clustered | 7 | VC_667_0 | 7 | 0.4101 | 1 | 0.4101 | 0 | 0 | 0 | 1 | Ref |
| 1997 | KF954506 | Unassigned | Unassigned | Unassigned | 517_1 | Clustered | 108 | VC_517_1 | 108 | 0.8483 | 1 | 0.8483 | 0 | 0 | 0 | 0.9955 | Ref |
| 1998 | KF959563 | Unassigned | Unassigned | Unassigned | 517_1 | Clustered | 108 | VC_517_1 | 108 | 0.8483 | 1 | 0.8483 | 0 | 0 | 0 | 0.9955 | Ref |
| 1999 | KF971864 | Unassigned | Unassigned | Unassigned | 42_0 | Clustered | 49 | VC_42_0 | 49 | 0.4517 | 1 | 0.4517 | 0 | 0 | 0 | 0.8597 | Ref |
| 2000 | KF977490 | Unassigned | Unassigned | Unassigned | 124_0 | Clustered | 2 | VC_124_0 | 2 | 0.3934 | 0.95337092 | 0.3751 | 0 | 0 | 0 | 1 | Ref |
| 2001 | KF981601 | Unassigned | Unassigned | Unassigned | 216_6 | Clustered | 42 | VC_216_6 | 42 | 0.473 | 1 | 0.473 | 0 | 0 | 0 | 0.8351 | Ref |
| 2002 | KF981730 | Unassigned | Unassigned | Unassigned | 813_1 | Clustered | 24 | VC_813_1 | 24 | 0.535 | 1 | 0.535 | 0 | 0 | 0 | 1 | Ref |
| 2003 | KF981875 | Unassigned | Unassigned | Unassigned | 80_0 | Clustered | 41 | VC_80_0 | 41 | 0.5837 | 1 | 0.5837 | 0 | 0 | 0 | 0.9805 | Ref |
| 2004 | KF981876 | Unassigned | Unassigned | Unassigned | 1024_0 | Clustered | 2 | VC_1024_0 | 2 | 1 | 1 | 1 | 0 | 0 | 0 | 1 | Ref |
| 2005 | KF986246 | Unassigned | Unassigned | Unassigned | 521_0 | Clustered | 37 | VC_521_0 | 37 | 0.912 | 1 | 0.912 | 0 | 0 | 0 | 1 | Ref |
| 2006 | KF986247 | Unassigned | Unassigned | Unassigned | 567_0 | Clustered | 8 | VC_567_0 | 8 | 0.4935 | 1 | 0.4935 | 0 | 0 | 0 | 1 | Ref |
| 2007 | KJ002054 | Unassigned | Unassigned | Unassigned | 793_0 | Clustered | 12 | VC_793_0 | 12 | 0.8531 | 1 | 0.8531 | 0 | 0 | 0 | 1 | Ref |
| 2008 | KJ003983 | Unassigned | Unassigned | Unassigned | 1018_0 | Clustered | 2 | VC_1018_0 | 2 | 0.0851 | 0.94401932 | 0.0804 | 0 | 0 | 0 | 1 | Ref |
| 2009 | KJ010547 | Unassigned | Unassigned | Unassigned | 85_0 | Clustered | 3 | VC_85_0 | 3 | 0.0422 | 0.99823242 | 0.0421 | 0 | 0 | 0 | 1 | Ref |
| 2010 | KJ010548 | Unassigned | Unassigned | Unassigned | 85_0 | Clustered | 3 | VC_85_0 | 3 | 0.0422 | 0.99823242 | 0.0421 | 0 | 0 | 0 | 1 | Ref |
| 2011 | KJ018213 | Unassigned | Unassigned | Unassigned | 827_0 | Clustered | 6 | VC_827_0 | 6 | 0.7493 | 1 | 0.7493 | 0 | 0 | 0 | 1 | Ref |
| 2012 | KJ019028 | Unassigned | Unassigned | Unassigned | 656_1 | Clustered | 46 | VC_656_1 | 46 | 0.8511 | 1 | 0.8511 | 0 | 0 | 0 | 1 | Ref |
| 2013 | KJ019029 | Unassigned | Unassigned | Unassigned | 656_1 | Clustered | 46 | VC_656_1 | 46 | 0.8511 | 1 | 0.8511 | 0 | 0 | 0 | 1 | Ref |
| 2014 | KJ019031 | Unassigned | Unassigned | Unassigned | 656_1 | Clustered | 46 | VC_656_1 | 46 | 0.8511 | 1 | 0.8511 | 0 | 0 | 0 | 1 | Ref |
| 2015 | KJ019032 | Unassigned | Unassigned | Unassigned | 656_1 | Clustered | 46 | VC_656_1 | 46 | 0.8511 | 1 | 0.8511 | 0 | 0 | 0 | 1 | Ref |
| 2016 | KJ019034 | Unassigned | Unassigned | Unassigned | 656_1 | Clustered | 46 | VC_656_1 | 46 | 0.8511 | 1 | 0.8511 | 0 | 0 | 0 | 1 | Ref |
| 2017 | KJ019035 | Unassigned | Unassigned | Unassigned | 656_1 | Clustered | 46 | VC_656_1 | 46 | 0.8511 | 1 | 0.8511 | 0 | 0 | 0 | 1 | Ref |
| 2018 | KJ019036 | Unassigned | Unassigned | Unassigned | 656_1 | Clustered | 46 | VC_656_1 | 46 | 0.8511 | 1 | 0.8511 | 0 | 0 | 0 | 1 | Ref |
| 2019 | KJ019037 | Unassigned | Unassigned | Unassigned | 657_1 | Clustered | 41 | VC_657_1 | 41 | 0.7489 | 1 | 0.7489 | 0 | 0 | 0 | 1 | Ref |
| 2020 | KJ019045 | Unassigned | Unassigned | Unassigned | 657_1 | Clustered | 41 | VC_657_1 | 41 | 0.7489 | 1 | 0.7489 | 0 | 0 | 0 | 1 | Ref |
| 2021 | KJ019046 | Unassigned | Unassigned | Unassigned | 656_1 | Clustered | 46 | VC_656_1 | 46 | 0.8511 | 1 | 0.8511 | 0 | 0 | 0 | 1 | Ref |
| 2022 | KJ019047 | Unassigned | Unassigned | Unassigned | 656_1 | Clustered | 46 | VC_656_1 | 46 | 0.8511 | 1 | 0.8511 | 0 | 0 | 0 | 1 | Ref |
| 2023 | KJ019048 | Unassigned | Unassigned | Unassigned | 656_1 | Clustered | 46 | VC_656_1 | 46 | 0.8511 | 1 | 0.8511 | 0 | 0 | 0 | 1 | Ref |
| 2024 | KJ019050 | Unassigned | Unassigned | Unassigned | 656_1 | Clustered | 46 | VC_656_1 | 46 | 0.8511 | 1 | 0.8511 | 0 | 0 | 0 | 1 | Ref |
| 2025 | KJ019052 | Unassigned | Unassigned | Unassigned | 657_1 | Clustered | 41 | VC_657_1 | 41 | 0.7489 | 1 | 0.7489 | 0 | 0 | 0 | 1 | Ref |
| 2026 | KJ019053 | Unassigned | Unassigned | Unassigned | 657_1 | Clustered | 41 | VC_657_1 | 41 | 0.7489 | 1 | 0.7489 | 0 | 0 | 0 | 1 | Ref |
| 2027 | KJ019054 | Unassigned | Unassigned | Unassigned | 433_1 | Clustered | 9 | VC_433_1 | 9 | 0.1288 | 1 | 0.1288 | 0 | 0 | 0 | 0.8611 | Ref |
| 2028 | KJ019056 | Unassigned | Unassigned | Unassigned | 656_1 | Clustered | 46 | VC_656_1 | 46 | 0.8511 | 1 | 0.8511 | 0 | 0 | 0 | 1 | Ref |
| 2029 | KJ019057 | Unassigned | Unassigned | Unassigned | 656_1 | Clustered | 46 | VC_656_1 | 46 | 0.8511 | 1 | 0.8511 | 0 | 0 | 0 | 1 | Ref |
| 2030 | KJ019058 | Unassigned | Unassigned | Unassigned | 657_1 | Clustered | 41 | VC_657_1 | 41 | 0.7489 | 1 | 0.7489 | 0 | 0 | 0 | 1 | Ref |
| 2031 | KJ019059 | Unassigned | Unassigned | Unassigned | 657_1 | Clustered | 41 | VC_657_1 | 41 | 0.7489 | 1 | 0.7489 | 0 | 0 | 0 | 1 | Ref |
| 2032 | KJ019062 | Unassigned | Unassigned | Unassigned | 656_1 | Clustered | 46 | VC_656_1 | 46 | 0.8511 | 1 | 0.8511 | 0 | 0 | 0 | 1 | Ref |
| 2033 | KJ019066 | Unassigned | Unassigned | Unassigned | 657_1 | Clustered | 41 | VC_657_1 | 41 | 0.7489 | 1 | 0.7489 | 0 | 0 | 0 | 1 | Ref |
| 2034 | KJ019069 | Unassigned | Unassigned | Unassigned | 433_1 | Clustered | 9 | VC_433_1 | 9 | 0.1288 | 1 | 0.1288 | 0 | 0 | 0 | 0.8611 | Ref |
| 2035 | KJ019070 | Unassigned | Unassigned | Unassigned | 656_1 | Clustered | 46 | VC_656_1 | 46 | 0.8511 | 1 | 0.8511 | 0 | 0 | 0 | 1 | Ref |
| 2036 | KJ019072 | Unassigned | Unassigned | Unassigned | 656_1 | Clustered | 46 | VC_656_1 | 46 | 0.8511 | 1 | 0.8511 | 0 | 0 | 0 | 1 | Ref |
| 2037 | KJ019073 | Unassigned | Unassigned | Unassigned | 656_1 | Clustered | 46 | VC_656_1 | 46 | 0.8511 | 1 | 0.8511 | 0 | 0 | 0 | 1 | Ref |
| 2038 | KJ019074 | Unassigned | Unassigned | Unassigned | 656_1 | Clustered | 46 | VC_656_1 | 46 | 0.8511 | 1 | 0.8511 | 0 | 0 | 0 | 1 | Ref |
| 2039 | KJ019075 | Unassigned | Unassigned | Unassigned | 656_1 | Clustered | 46 | VC_656_1 | 46 | 0.8511 | 1 | 0.8511 | 0 | 0 | 0 | 1 | Ref |
| 2040 | KJ019077 | Unassigned | Unassigned | Unassigned | 656_1 | Clustered | 46 | VC_656_1 | 46 | 0.8511 | 1 | 0.8511 | 0 | 0 | 0 | 1 | Ref |
| 2041 | KJ019078 | Unassigned | Unassigned | Unassigned | 656_1 | Clustered | 46 | VC_656_1 | 46 | 0.8511 | 1 | 0.8511 | 0 | 0 | 0 | 1 | Ref |
| 2042 | KJ019079 | Unassigned | Unassigned | Unassigned | 656_1 | Clustered | 46 | VC_656_1 | 46 | 0.8511 | 1 | 0.8511 | 0 | 0 | 0 | 1 | Ref |
| 2043 | KJ019080 | Unassigned | Unassigned | Unassigned | 656_1 | Clustered | 46 | VC_656_1 | 46 | 0.8511 | 1 | 0.8511 | 0 | 0 | 0 | 1 | Ref |
| 2044 | KJ019082 | Unassigned | Unassigned | Unassigned | 433_1 | Clustered | 9 | VC_433_1 | 9 | 0.1288 | 1 | 0.1288 | 0 | 0 | 0 | 0.8611 | Ref |
| 2045 | KJ019083 | Unassigned | Unassigned | Unassigned | 656_1 | Clustered | 46 | VC_656_1 | 46 | 0.8511 | 1 | 0.8511 | 0 | 0 | 0 | 1 | Ref |
| 2046 | KJ019085 | Unassigned | Unassigned | Unassigned | 657_1 | Clustered | 41 | VC_657_1 | 41 | 0.7489 | 1 | 0.7489 | 0 | 0 | 0 | 1 | Ref |
| 2047 | KJ019086 | Unassigned | Unassigned | Unassigned | 657_1 | Clustered | 41 | VC_657_1 | 41 | 0.7489 | 1 | 0.7489 | 0 | 0 | 0 | 1 | Ref |
| 2048 | KJ019089 | Unassigned | Unassigned | Unassigned | 433_1 | Clustered | 9 | VC_433_1 | 9 | 0.1288 | 1 | 0.1288 | 0 | 0 | 0 | 0.8611 | Ref |
| 2049 | KJ019090 | Unassigned | Unassigned | Unassigned | 657_1 | Clustered | 41 | VC_657_1 | 41 | 0.7489 | 1 | 0.7489 | 0 | 0 | 0 | 1 | Ref |
| 2050 | KJ019091 | Unassigned | Unassigned | Unassigned | 657_1 | Clustered | 41 | VC_657_1 | 41 | 0.7489 | 1 | 0.7489 | 0 | 0 | 0 | 1 | Ref |
| 2051 | KJ019092 | Unassigned | Unassigned | Unassigned | 657_1 | Clustered | 41 | VC_657_1 | 41 | 0.7489 | 1 | 0.7489 | 0 | 0 | 0 | 1 | Ref |
| 2052 | KJ019093 | Unassigned | Unassigned | Unassigned | 657_1 | Clustered | 41 | VC_657_1 | 41 | 0.7489 | 1 | 0.7489 | 0 | 0 | 0 | 1 | Ref |
| 2053 | KJ019094 | Unassigned | Unassigned | Unassigned | 433_1 | Clustered | 9 | VC_433_1 | 9 | 0.1288 | 1 | 0.1288 | 0 | 0 | 0 | 0.8611 | Ref |
| 2054 | KJ019095 | Unassigned | Unassigned | Unassigned | 657_1 | Clustered | 41 | VC_657_1 | 41 | 0.7489 | 1 | 0.7489 | 0 | 0 | 0 | 1 | Ref |
| 2055 | KJ019096 | Unassigned | Unassigned | Unassigned | 657_1 | Clustered | 41 | VC_657_1 | 41 | 0.7489 | 1 | 0.7489 | 0 | 0 | 0 | 1 | Ref |
| 2056 | KJ019097 | Unassigned | Unassigned | Unassigned | 657_1 | Clustered | 41 | VC_657_1 | 41 | 0.7489 | 1 | 0.7489 | 0 | 0 | 0 | 1 | Ref |
| 2057 | KJ019098 | Unassigned | Unassigned | Unassigned | 657_1 | Clustered | 41 | VC_657_1 | 41 | 0.7489 | 1 | 0.7489 | 0 | 0 | 0 | 1 | Ref |
| 2058 | KJ019099 | Unassigned | Unassigned | Unassigned | 657_1 | Clustered | 41 | VC_657_1 | 41 | 0.7489 | 1 | 0.7489 | 0 | 0 | 0 | 1 | Ref |
| 2059 | KJ019100 | Unassigned | Unassigned | Unassigned | 657_1 | Clustered | 41 | VC_657_1 | 41 | 0.7489 | 1 | 0.7489 | 0 | 0 | 0 | 1 | Ref |
| 2060 | KJ019101 | Unassigned | Unassigned | Unassigned | 657_1 | Clustered | 41 | VC_657_1 | 41 | 0.7489 | 1 | 0.7489 | 0 | 0 | 0 | 1 | Ref |
| 2061 | KJ019102 | Unassigned | Unassigned | Unassigned | 657_1 | Clustered | 41 | VC_657_1 | 41 | 0.7489 | 1 | 0.7489 | 0 | 0 | 0 | 1 | Ref |
| 2062 | KJ019103 | Unassigned | Unassigned | Unassigned | 657_1 | Clustered | 41 | VC_657_1 | 41 | 0.7489 | 1 | 0.7489 | 0 | 0 | 0 | 1 | Ref |
| 2063 | KJ019105 | Unassigned | Unassigned | Unassigned | 657_1 | Clustered | 41 | VC_657_1 | 41 | 0.7489 | 1 | 0.7489 | 0 | 0 | 0 | 1 | Ref |
| 2064 | KJ019106 | Unassigned | Unassigned | Unassigned | 657_1 | Clustered | 41 | VC_657_1 | 41 | 0.7489 | 1 | 0.7489 | 0 | 0 | 0 | 1 | Ref |
| 2065 | KJ019107 | Unassigned | Unassigned | Unassigned | 657_1 | Clustered | 41 | VC_657_1 | 41 | 0.7489 | 1 | 0.7489 | 0 | 0 | 0 | 1 | Ref |
| 2066 | KJ019111 | Unassigned | Unassigned | Unassigned | 657_1 | Clustered | 41 | VC_657_1 | 41 | 0.7489 | 1 | 0.7489 | 0 | 0 | 0 | 1 | Ref |
| 2067 | KJ019112 | Unassigned | Unassigned | Unassigned | 656_1 | Clustered | 46 | VC_656_1 | 46 | 0.8511 | 1 | 0.8511 | 0 | 0 | 0 | 1 | Ref |
| 2068 | KJ019113 | Unassigned | Unassigned | Unassigned | 656_1 | Clustered | 46 | VC_656_1 | 46 | 0.8511 | 1 | 0.8511 | 0 | 0 | 0 | 1 | Ref |
| 2069 | KJ019115 | Unassigned | Unassigned | Unassigned | 656_1 | Clustered | 46 | VC_656_1 | 46 | 0.8511 | 1 | 0.8511 | 0 | 0 | 0 | 1 | Ref |
| 2070 | KJ019117 | Unassigned | Unassigned | Unassigned | 656_1 | Clustered | 46 | VC_656_1 | 46 | 0.8511 | 1 | 0.8511 | 0 | 0 | 0 | 1 | Ref |
| 2071 | KJ019118 | Unassigned | Unassigned | Unassigned | 656_1 | Clustered | 46 | VC_656_1 | 46 | 0.8511 | 1 | 0.8511 | 0 | 0 | 0 | 1 | Ref |
| 2072 | KJ019119 | Unassigned | Unassigned | Unassigned | 656_1 | Clustered | 46 | VC_656_1 | 46 | 0.8511 | 1 | 0.8511 | 0 | 0 | 0 | 1 | Ref |
| 2073 | KJ019120 | Unassigned | Unassigned | Unassigned | 656_1 | Clustered | 46 | VC_656_1 | 46 | 0.8511 | 1 | 0.8511 | 0 | 0 | 0 | 1 | Ref |
| 2074 | KJ019121 | Unassigned | Unassigned | Unassigned | 656_1 | Clustered | 46 | VC_656_1 | 46 | 0.8511 | 1 | 0.8511 | 0 | 0 | 0 | 1 | Ref |
| 2075 | KJ019123 | Unassigned | Unassigned | Unassigned | 657_1 | Clustered | 41 | VC_657_1 | 41 | 0.7489 | 1 | 0.7489 | 0 | 0 | 0 | 1 | Ref |
| 2076 | KJ019124 | Unassigned | Unassigned | Unassigned | 656_1 | Clustered | 46 | VC_656_1 | 46 | 0.8511 | 1 | 0.8511 | 0 | 0 | 0 | 1 | Ref |
| 2077 | KJ019125 | Unassigned | Unassigned | Unassigned | 656_1 | Clustered | 46 | VC_656_1 | 46 | 0.8511 | 1 | 0.8511 | 0 | 0 | 0 | 1 | Ref |
| 2078 | KJ019126 | Unassigned | Unassigned | Unassigned | 656_1 | Clustered | 46 | VC_656_1 | 46 | 0.8511 | 1 | 0.8511 | 0 | 0 | 0 | 1 | Ref |
| 2079 | KJ019127 | Unassigned | Unassigned | Unassigned | 656_1 | Clustered | 46 | VC_656_1 | 46 | 0.8511 | 1 | 0.8511 | 0 | 0 | 0 | 1 | Ref |
| 2080 | KJ019129 | Unassigned | Unassigned | Unassigned | 656_1 | Clustered | 46 | VC_656_1 | 46 | 0.8511 | 1 | 0.8511 | 0 | 0 | 0 | 1 | Ref |
| 2081 | KJ019130 | Unassigned | Unassigned | Unassigned | 656_1 | Clustered | 46 | VC_656_1 | 46 | 0.8511 | 1 | 0.8511 | 0 | 0 | 0 | 1 | Ref |
| 2082 | KJ019131 | Unassigned | Unassigned | Unassigned | 656_1 | Clustered | 46 | VC_656_1 | 46 | 0.8511 | 1 | 0.8511 | 0 | 0 | 0 | 1 | Ref |
| 2083 | KJ019136 | Unassigned | Unassigned | Unassigned | 656_1 | Clustered | 46 | VC_656_1 | 46 | 0.8511 | 1 | 0.8511 | 0 | 0 | 0 | 1 | Ref |
| 2084 | KJ019139 | Unassigned | Unassigned | Unassigned | 656_1 | Clustered | 46 | VC_656_1 | 46 | 0.8511 | 1 | 0.8511 | 0 | 0 | 0 | 1 | Ref |
| 2085 | KJ019140 | Unassigned | Unassigned | Unassigned | 656_1 | Clustered | 46 | VC_656_1 | 46 | 0.8511 | 1 | 0.8511 | 0 | 0 | 0 | 1 | Ref |
| 2086 | KJ019141 | Unassigned | Unassigned | Unassigned | 657_1 | Clustered | 41 | VC_657_1 | 41 | 0.7489 | 1 | 0.7489 | 0 | 0 | 0 | 1 | Ref |
| 2087 | KJ019142 | Unassigned | Unassigned | Unassigned | 657_1 | Clustered | 41 | VC_657_1 | 41 | 0.7489 | 1 | 0.7489 | 0 | 0 | 0 | 1 | Ref |
| 2088 | KJ019143 | Unassigned | Unassigned | Unassigned | 657_1 | Clustered | 41 | VC_657_1 | 41 | 0.7489 | 1 | 0.7489 | 0 | 0 | 0 | 1 | Ref |
| 2089 | KJ019144 | Unassigned | Unassigned | Unassigned | 657_1 | Clustered | 41 | VC_657_1 | 41 | 0.7489 | 1 | 0.7489 | 0 | 0 | 0 | 1 | Ref |
| 2090 | KJ019145 | Unassigned | Unassigned | Unassigned | 657_1 | Clustered | 41 | VC_657_1 | 41 | 0.7489 | 1 | 0.7489 | 0 | 0 | 0 | 1 | Ref |
| 2091 | KJ019146 | Unassigned | Unassigned | Unassigned | 657_1 | Clustered | 41 | VC_657_1 | 41 | 0.7489 | 1 | 0.7489 | 0 | 0 | 0 | 1 | Ref |
| 2092 | KJ019147 | Unassigned | Unassigned | Unassigned | 657_1 | Clustered | 41 | VC_657_1 | 41 | 0.7489 | 1 | 0.7489 | 0 | 0 | 0 | 1 | Ref |
| 2093 | KJ019148 | Unassigned | Unassigned | Unassigned | 657_1 | Clustered | 41 | VC_657_1 | 41 | 0.7489 | 1 | 0.7489 | 0 | 0 | 0 | 1 | Ref |
| 2094 | KJ019149 | Unassigned | Unassigned | Unassigned | 657_1 | Clustered | 41 | VC_657_1 | 41 | 0.7489 | 1 | 0.7489 | 0 | 0 | 0 | 1 | Ref |
| 2095 | KJ019150 | Unassigned | Unassigned | Unassigned | 657_1 | Clustered | 41 | VC_657_1 | 41 | 0.7489 | 1 | 0.7489 | 0 | 0 | 0 | 1 | Ref |
| 2096 | KJ019151 | Unassigned | Unassigned | Unassigned | 657_1 | Clustered | 41 | VC_657_1 | 41 | 0.7489 | 1 | 0.7489 | 0 | 0 | 0 | 1 | Ref |
| 2097 | KJ019152 | Unassigned | Unassigned | Unassigned | 657_1 | Clustered | 41 | VC_657_1 | 41 | 0.7489 | 1 | 0.7489 | 0 | 0 | 0 | 1 | Ref |
| 2098 | KJ019155 | Unassigned | Unassigned | Unassigned | 657_1 | Clustered | 41 | VC_657_1 | 41 | 0.7489 | 1 | 0.7489 | 0 | 0 | 0 | 1 | Ref |
| 2099 | KJ019156 | Unassigned | Unassigned | Unassigned | 433_1 | Clustered | 9 | VC_433_1 | 9 | 0.1288 | 1 | 0.1288 | 0 | 0 | 0 | 0.8611 | Ref |
| 2100 | KJ019159 | Unassigned | Unassigned | Unassigned | 657_1 | Clustered | 41 | VC_657_1 | 41 | 0.7489 | 1 | 0.7489 | 0 | 0 | 0 | 1 | Ref |
| 2101 | KJ019160 | Unassigned | Unassigned | Unassigned | 656_1 | Clustered | 46 | VC_656_1 | 46 | 0.8511 | 1 | 0.8511 | 0 | 0 | 0 | 1 | Ref |
| 2102 | KJ019162 | Unassigned | Unassigned | Unassigned | 656_1 | Clustered | 46 | VC_656_1 | 46 | 0.8511 | 1 | 0.8511 | 0 | 0 | 0 | 1 | Ref |
| 2103 | KJ019164 | Unassigned | Unassigned | Unassigned | 656_1 | Clustered | 46 | VC_656_1 | 46 | 0.8511 | 1 | 0.8511 | 0 | 0 | 0 | 1 | Ref |
| 2104 | KJ019165 | Unassigned | Unassigned | Unassigned | 656_1 | Clustered | 46 | VC_656_1 | 46 | 0.8511 | 1 | 0.8511 | 0 | 0 | 0 | 1 | Ref |

| | | | | | | | | | | | | | | | | | |
|---|---|---|---|---|---|---|---|---|---|---|---|---|---|---|---|---|---|
| 2105 | KJ021043 | Unassigned | Unassigned | Unassigned | 123_0 | Clustered | 23 | VC_123_0 | 23 | 0.817 | 1 | 0.817 | 0 | 0 | 0 | 1 | Ref |
| 2106 | KJ025956 | Unassigned | Unassigned | Unassigned | 520_38 | Clustered | 6 | VC_520_38 | 6 | 0.0519 | 1 | 0.0519 | 0 | 0 | 0 | 0.8667 | Ref |
| 2107 | KJ025957 | Unassigned | Unassigned | Unassigned | 284_0 | Clustered | 3 | VC_284_0 | 3 | 0.0495 | 0.99860966 | 0.0494 | 0 | 0 | 0 | 1 | Ref |
| 2108 | KJ028219 | Unassigned | Unassigned | Unassigned | 809_0 | Clustered | 2 | VC_809_0 | 2 | 0.1799 | 0.96299906 | 0.1733 | 0 | 0 | 0 | 1 | Ref |
| 2109 | KJ081346 | Unassigned | Unassigned | Unassigned | 187_2 | Clustered | 2 | VC_187_2 | 2 | 0.0209 | 0.93057595 | 0.0195 | 0 | 0 | 0 | 1 | Ref |
| 2110 | KJ094020 | Unassigned | Unassigned | Unassigned | 980_0 | Clustered | 10 | VC_980_0 | 10 | 0.9996 | 1 | 0.9996 | 0 | 0 | 0 | 1 | Ref |
| 2111 | KJ094021 | Unassigned | Unassigned | Unassigned | 980_0 | Clustered | 10 | VC_980_0 | 10 | 0.9996 | 1 | 0.9996 | 0 | 0 | 0 | 1 | Ref |
| 2112 | KJ094022 | Unassigned | Unassigned | Unassigned | 378_0 | Clustered | 9 | VC_378_0 | 9 | 0.7603 | 1 | 0.7603 | 0 | 0 | 0 | 1 | Ref |
| 2113 | KJ094023 | Unassigned | Unassigned | Unassigned | 379_0 | Clustered | 5 | VC_379_0 | 5 | 0.4779 | 0.99999992 | 0.4779 | 0 | 0 | 0 | 0.9 | Ref |
| 2114 | KJ094029 | Unassigned | Unassigned | Unassigned | 739_0 | Clustered | 26 | VC_739_0 | 26 | 0.9571 | 1 | 0.9571 | 0 | 0 | 0 | 1 | Ref |
| 2115 | KJ094030 | Unassigned | Unassigned | Unassigned | 739_0 | Clustered | 26 | VC_739_0 | 26 | 0.9571 | 1 | 0.9571 | 0 | 0 | 0 | 1 | Ref |
| 2116 | KJ094031 | Unassigned | Unassigned | Unassigned | 739_0 | Clustered | 26 | VC_739_0 | 26 | 0.9571 | 1 | 0.9571 | 0 | 0 | 0 | 1 | Ref |
| 2117 | KJ094032 | Unassigned | Unassigned | Unassigned | 108_0 | Clustered | 33 | VC_108_0 | 33 | 0.949 | 1 | 0.949 | 0 | 0 | 0 | 1 | Ref |
| 2118 | KJ094033 | Unassigned | Unassigned | Unassigned | 739_0 | Clustered | 26 | VC_739_0 | 26 | 0.9571 | 1 | 0.9571 | 0 | 0 | 0 | 1 | Ref |
| 2119 | KJ127303 | Unassigned | Unassigned | Unassigned | 108_0 | Clustered | 33 | VC_108_0 | 33 | 0.949 | 1 | 0.949 | 0 | 0 | 0 | 1 | Ref |
| 2120 | KJ127304 | Unassigned | Unassigned | Unassigned | 695_0 | Clustered | 48 | VC_695_0 | 48 | 0.8162 | 1 | 0.8162 | 0 | 0 | 0 | 1 | Ref |
| 2121 | KJ133688 | Unassigned | Unassigned | Unassigned | 879_0 | Clustered | 36 | VC_879_0 | 36 | 0.994 | 1 | 0.994 | 0 | 0 | 0 | 1 | Ref |
| 2122 | KJ133689 | Unassigned | Unassigned | Unassigned | 879_0 | Clustered | 36 | VC_879_0 | 36 | 0.994 | 1 | 0.994 | 0 | 0 | 0 | 1 | Ref |
| 2123 | KJ133690 | Unassigned | Unassigned | Unassigned | 879_0 | Clustered | 36 | VC_879_0 | 36 | 0.994 | 1 | 0.994 | 0 | 0 | 0 | 1 | Ref |
| 2124 | KJ133691 | Unassigned | Unassigned | Unassigned | 879_0 | Clustered | 36 | VC_879_0 | 36 | 0.994 | 1 | 0.994 | 0 | 0 | 0 | 1 | Ref |
| 2125 | KJ133692 | Unassigned | Unassigned | Unassigned | 879_0 | Clustered | 36 | VC_879_0 | 36 | 0.994 | 1 | 0.994 | 0 | 0 | 0 | 1 | Ref |
| 2126 | KJ133693 | Unassigned | Unassigned | Unassigned | 879_0 | Clustered | 36 | VC_879_0 | 36 | 0.994 | 1 | 0.994 | 0 | 0 | 0 | 1 | Ref |
| 2127 | KJ133694 | Unassigned | Unassigned | Unassigned | 879_0 | Clustered | 36 | VC_879_0 | 36 | 0.994 | 1 | 0.994 | 0 | 0 | 0 | 1 | Ref |
| 2128 | KJ133695 | Unassigned | Unassigned | Unassigned | 879_0 | Clustered | 36 | VC_879_0 | 36 | 0.994 | 1 | 0.994 | 0 | 0 | 0 | 1 | Ref |
| 2129 | KJ133696 | Unassigned | Unassigned | Unassigned | 879_0 | Clustered | 36 | VC_879_0 | 36 | 0.994 | 1 | 0.994 | 0 | 0 | 0 | 1 | Ref |
| 2130 | KJ133697 | Unassigned | Unassigned | Unassigned | 879_0 | Clustered | 36 | VC_879_0 | 36 | 0.994 | 1 | 0.994 | 0 | 0 | 0 | 1 | Ref |
| 2131 | KJ133698 | Unassigned | Unassigned | Unassigned | 879_0 | Clustered | 36 | VC_879_0 | 36 | 0.994 | 1 | 0.994 | 0 | 0 | 0 | 1 | Ref |
| 2132 | KJ133699 | Unassigned | Unassigned | Unassigned | 879_0 | Clustered | 36 | VC_879_0 | 36 | 0.994 | 1 | 0.994 | 0 | 0 | 0 | 1 | Ref |
| 2133 | KJ133700 | Unassigned | Unassigned | Unassigned | 879_0 | Clustered | 36 | VC_879_0 | 36 | 0.994 | 1 | 0.994 | 0 | 0 | 0 | 1 | Ref |
| 2134 | KJ133701 | Unassigned | Unassigned | Unassigned | 879_0 | Clustered | 36 | VC_879_0 | 36 | 0.994 | 1 | 0.994 | 0 | 0 | 0 | 1 | Ref |
| 2135 | KJ133702 | Unassigned | Unassigned | Unassigned | 879_0 | Clustered | 36 | VC_879_0 | 36 | 0.994 | 1 | 0.994 | 0 | 0 | 0 | 1 | Ref |
| 2136 | KJ133703 | Unassigned | Unassigned | Unassigned | 879_0 | Clustered | 36 | VC_879_0 | 36 | 0.994 | 1 | 0.994 | 0 | 0 | 0 | 1 | Ref |
| 2137 | KJ133704 | Unassigned | Unassigned | Unassigned | 879_0 | Clustered | 36 | VC_879_0 | 36 | 0.994 | 1 | 0.994 | 0 | 0 | 0 | 1 | Ref |
| 2138 | KJ133705 | Unassigned | Unassigned | Unassigned | 879_0 | Clustered | 36 | VC_879_0 | 36 | 0.994 | 1 | 0.994 | 0 | 0 | 0 | 1 | Ref |
| 2139 | KJ133706 | Unassigned | Unassigned | Unassigned | 879_0 | Clustered | 36 | VC_879_0 | 36 | 0.994 | 1 | 0.994 | 0 | 0 | 0 | 1 | Ref |
| 2140 | KJ133707 | Unassigned | Unassigned | Unassigned | 879_0 | Clustered | 36 | VC_879_0 | 36 | 0.994 | 1 | 0.994 | 0 | 0 | 0 | 1 | Ref |
| 2141 | KJ135004 | Unassigned | Unassigned | Unassigned | 779_0 | Clustered | 37 | VC_779_0 | 37 | 0.5682 | 1 | 0.5682 | 0 | 0 | 0 | 1 | Ref |
| 2142 | KJ140076 | Unassigned | Unassigned | Unassigned | 12_0 | Clustered | 21 | VC_12_0 | 21 | 0.2274 | 1 | 0.2274 | 0 | 0 | 0 | 0.9571 | Ref |
| 2143 | KJ156985 | Unassigned | Unassigned | Unassigned | 515_0 | Clustered | 16 | VC_515_0 | 16 | 0.3793 | 1 | 0.3793 | 0 | 0 | 0 | 1 | Ref |
| 2144 | KJ159566 | Unassigned | Unassigned | Unassigned | 102_0 | Clustered | 2 | VC_102_0 | 2 | 0.114 | 0.94637553 | 0.1079 | 0 | 0 | 0 | 1 | Ref |
| 2145 | KJ173786 | Unassigned | Unassigned | Unassigned | 856_1 | Clustered | 13 | VC_856_1 | 13 | 0.5848 | 1 | 0.5848 | 0 | 0 | 0 | 1 | Ref |
| 2146 | KJ174156 | Unassigned | Unassigned | Unassigned | 325_15 | Clustered/Singleton | 1 | VC_325_15 | 1 | 0 | 1 | 0 | 0 | 0 | 0 | 1 | Ref |
| 2147 | KJ174157 | Unassigned | Unassigned | Unassigned | 371_3 | Clustered | 22 | VC_371_3 | 22 | 0.0787 | 1 | 0.0787 | 0 | 0 | 0 | 0.9048 | Ref |
| 2148 | KJ174317 | Unassigned | Unassigned | Unassigned | 133_22 | Clustered | 13 | VC_133_22 | 13 | 0.0804 | 1 | 0.0804 | 0 | 0 | 0 | 0.8205 | Ref |
| 2149 | KJ174318 | Unassigned | Unassigned | Unassigned | 133_16 | Clustered/Singleton | 1 | VC_133_16 | 1 | 0 | 1 | 0 | 0 | 0 | 0 | 1 | Ref |
| 2150 | KJ183191 | Unassigned | Unassigned | Unassigned | 484_0 | Clustered | 3 | VC_484_0 | 3 | 0.3174 | 0.99843638 | 0.3169 | 0 | 0 | 0 | 1 | Ref |
| 2151 | KJ183192 | Unassigned | Unassigned | Unassigned | 484_0 | Clustered | 3 | VC_484_0 | 3 | 0.3174 | 0.99843638 | 0.3169 | 0 | 0 | 0 | 1 | Ref |
| 2152 | KJ183193 | Unassigned | Unassigned | Unassigned | 484_0 | Clustered | 3 | VC_484_0 | 3 | 0.3174 | 0.99843638 | 0.3169 | 0 | 0 | 0 | 1 | Ref |
| 2153 | KJ190157 | Unassigned | Unassigned | Unassigned | 190_15 | Clustered | 3 | VC_190_15 | 3 | 0.0146 | 0.99759465 | 0.0145 | 0 | 0 | 0 | 0.6667 | Ref |
| 2154 | KJ190158 | Unassigned | Unassigned | Unassigned | 772_16 | Clustered | 10 | VC_772_16 | 10 | 0.0563 | 0.9434928 | 0.0531 | 0 | 0 | 0 | 0.8667 | Ref |
| 2155 | KJ194579 | Unassigned | Unassigned | Unassigned | 371_18 | Clustered | 10 | VC_371_18 | 10 | 0.0302 | 1 | 0.0302 | 0 | 0 | 0 | 0.8 | Ref |
| 2156 | KJ194580 | Unassigned | Unassigned | Unassigned | 371_16 | Clustered | 42 | VC_371_16 | 42 | 0.1341 | 1 | 0.1341 | 0 | 0 | 0 | 0.7282 | Ref |
| 2157 | KJ194581 | Unassigned | Unassigned | Unassigned | 509_0 | Clustered | 37 | VC_509_0 | 37 | 0.9689 | 1 | 0.9689 | 0 | 0 | 0 | 1 | Ref |
| 2158 | KJ194582 | Unassigned | Unassigned | Unassigned | 565_0 | Clustered | 20 | VC_565_0 | 20 | 0.8398 | 1 | 0.8398 | 0 | 0 | 0 | 1 | Ref |
| 2159 | KJ194583 | Unassigned | Unassigned | Unassigned | 371_16 | Clustered | 42 | VC_371_16 | 42 | 0.1341 | 1 | 0.1341 | 0 | 0 | 0 | 0.7282 | Ref |
| 2160 | KJ194584 | Unassigned | Unassigned | Unassigned | 509_0 | Clustered | 37 | VC_509_0 | 37 | 0.9689 | 1 | 0.9689 | 0 | 0 | 0 | 1 | Ref |
| 2161 | KJ194585 | Unassigned | Unassigned | Unassigned | 325_5 | Clustered | 11 | VC_325_5 | 11 | 0.047 | 1 | 0.047 | 0 | 0 | 0 | 0.7636 | Ref |
| 2162 | KJ206559 | Unassigned | Unassigned | Unassigned | 149_8 | Clustered | 66 | VC_149_8 | 66 | 0.4565 | 1 | 0.4565 | 0 | 0 | 0 | 0.8182 | Ref |
| 2163 | KJ206560 | Unassigned | Unassigned | Unassigned | 149_8 | Clustered | 66 | VC_149_8 | 66 | 0.4565 | 1 | 0.4565 | 0 | 0 | 0 | 0.8182 | Ref |
| 2164 | KJ206561 | Unassigned | Unassigned | Unassigned | 149_8 | Clustered | 66 | VC_149_8 | 66 | 0.4565 | 1 | 0.4565 | 0 | 0 | 0 | 0.8182 | Ref |
| 2165 | KJ206562 | Unassigned | Unassigned | Unassigned | 149_8 | Clustered | 66 | VC_149_8 | 66 | 0.4565 | 1 | 0.4565 | 0 | 0 | 0 | 0.8182 | Ref |
| 2166 | KJ206563 | Unassigned | Unassigned | Unassigned | 149_8 | Clustered | 66 | VC_149_8 | 66 | 0.4565 | 1 | 0.4565 | 0 | 0 | 0 | 0.8182 | Ref |
| 2167 | KJ210330 | Unassigned | Unassigned | Unassigned | 96_0 | Clustered | 30 | VC_96_0 | 30 | 0.718 | 1 | 0.718 | 0 | 0 | 0 | 1 | Ref |
| 2168 | KJ409696 | Unassigned | Unassigned | Unassigned | 325_30 | Clustered | 4 | VC_325_30 | 4 | 0.0144 | 0.99998616 | 0.0144 | 0 | 0 | 0 | 0.8333 | Ref |
| 2169 | KJ409772 | Unassigned | Unassigned | Unassigned | 78_3 | Clustered | 11 | VC_78_3 | 11 | 0.2572 | 1 | 0.2572 | 0 | 0 | 0 | 1 | Ref |
| 2170 | KJ410132 | Unassigned | Unassigned | Unassigned | 216_6 | Clustered | 42 | VC_216_6 | 42 | 0.473 | 1 | 0.473 | 0 | 0 | 0 | 0.8351 | Ref |
| 2171 | KJ410133 | Unassigned | Unassigned | Unassigned | 762_0 | Clustered | 9 | VC_762_0 | 9 | 0.0885 | 1 | 0.0885 | 0 | 0 | 0 | 1 | Ref |
| 2172 | KJ410134 | Unassigned | Unassigned | Unassigned | 216_6 | Clustered | 42 | VC_216_6 | 42 | 0.473 | 1 | 0.473 | 0 | 0 | 0 | 0.8351 | Ref |
| 2173 | KJ410740 | Unassigned | Unassigned | Unassigned | 927_0 | Clustered | 2 | VC_927_0 | 2 | 0.0876 | 0.71936208 | 0.063 | 0 | 0 | 0 | 1 | Ref |
| 2174 | KJ417497 | Unassigned | Unassigned | Unassigned | 414_0 | Clustered | 37 | VC_414_0 | 37 | 0.585 | 1 | 0.585 | 0 | 0 | 0 | 0.9474 | Ref |
| 2175 | KJ419279 | Unassigned | Unassigned | Unassigned | 722_0 | Clustered | 13 | VC_722_0 | 13 | 0.5104 | 1 | 0.5104 | 0 | 0 | 0 | 1 | Ref |
| 2176 | KJ433973 | Unassigned | Unassigned | Unassigned | 372_3 | Clustered | 5 | VC_372_3 | 5 | 0.1129 | 0.99999997 | 0.1129 | 0 | 0 | 0 | 1 | Ref |
| 2177 | KJ433974 | Unassigned | Unassigned | Unassigned | 372_3 | Clustered | 5 | VC_372_3 | 5 | 0.1129 | 0.99999997 | 0.1129 | 0 | 0 | 0 | 1 | Ref |
| 2178 | KJ433975 | Unassigned | Unassigned | Unassigned | 372_3 | Clustered | 5 | VC_372_3 | 5 | 0.1129 | 0.99999997 | 0.1129 | 0 | 0 | 0 | 1 | Ref |
| 2179 | KJ433976 | Unassigned | Unassigned | Unassigned | 372_3 | Clustered | 5 | VC_372_3 | 5 | 0.1129 | 0.99999997 | 0.1129 | 0 | 0 | 0 | 1 | Ref |
| 2180 | KJ451625 | Unassigned | Unassigned | Unassigned | 187_3 | Clustered | 9 | VC_187_3 | 9 | 0.1971 | 1 | 0.1971 | 0 | 0 | 0 | 0.8611 | Ref |
| 2181 | KJ452291 | Unassigned | Unassigned | Unassigned | 12_0 | Clustered | 21 | VC_12_0 | 21 | 0.2274 | 1 | 0.2274 | 0 | 0 | 0 | 0.9571 | Ref |
| 2182 | KJ452292 | Unassigned | Unassigned | Unassigned | 10_0 | Clustered | 25 | VC_10_0 | 25 | 0.3467 | 1 | 0.3467 | 0 | 0 | 0 | 0.9733 | Ref |
| 2183 | KJ473422 | Unassigned | Unassigned | Unassigned | 700_1 | Clustered/Singleton | 1 | VC_700_1 | 1 | 0 | 1 | 0 | 0 | 0 | 0 | 1 | Ref |
| 2184 | KJ477077 | Unassigned | Unassigned | Unassigned | 179_0 | Clustered | 53 | VC_179_0 | 53 | 0.9137 | 1 | 0.9137 | 0 | 0 | 0 | 1 | Ref |
| 2185 | KJ477684 | Unassigned | Unassigned | Unassigned | 283_91 | Clustered | 11 | VC_283_91 | 11 | 0.0285 | 1 | 0.0285 | 0 | 0 | 0 | 1 | Ref |
| 2186 | KJ477685 | Unassigned | Unassigned | Unassigned | 283_91 | Clustered | 11 | VC_283_91 | 11 | 0.0285 | 1 | 0.0285 | 0 | 0 | 0 | 1 | Ref |
| 2187 | KJ477686 | Unassigned | Unassigned | Unassigned | 283_91 | Clustered | 11 | VC_283_91 | 11 | 0.0285 | 1 | 0.0285 | 0 | 0 | 0 | 1 | Ref |
| 2188 | KJ489010 | Unassigned | Unassigned | Unassigned | 801_0 | Clustered | 4 | VC_801_0 | 4 | 0.4828 | 0.99998705 | 0.4828 | 0 | 0 | 0 | 1 | Ref |
| 2189 | KJ489011 | Unassigned | Unassigned | Unassigned | 801_0 | Clustered | 4 | VC_801_0 | 4 | 0.4828 | 0.99998705 | 0.4828 | 0 | 0 | 0 | 1 | Ref |
| 2190 | KJ489012 | Unassigned | Unassigned | Unassigned | 801_0 | Clustered | 4 | VC_801_0 | 4 | 0.4828 | 0.99998705 | 0.4828 | 0 | 0 | 0 | 1 | Ref |
| 2191 | KJ489013 | Unassigned | Unassigned | Unassigned | 801_0 | Clustered | 4 | VC_801_0 | 4 | 0.4828 | 0.99998705 | 0.4828 | 0 | 0 | 0 | 1 | Ref |
| 2192 | KJ489397 | Unassigned | Unassigned | Unassigned | 883_0 | Clustered | 10 | VC_883_0 | 10 | 0.2015 | 1 | 0.2015 | 0 | 0 | 0 | 1 | Ref |
| 2193 | KJ489398 | Unassigned | Unassigned | Unassigned | 883_0 | Clustered | 10 | VC_883_0 | 10 | 0.2015 | 1 | 0.2015 | 0 | 0 | 0 | 1 | Ref |
| 2194 | KJ489399 | Unassigned | Unassigned | Unassigned | 884_0 | Clustered | 28 | VC_884_0 | 28 | 0.6211 | 1 | 0.6211 | 0 | 0 | 0 | 0.9841 | Ref |
| 2195 | KJ489400 | Unassigned | Unassigned | Unassigned | 883_0 | Clustered | 10 | VC_883_0 | 10 | 0.2015 | 1 | 0.2015 | 0 | 0 | 0 | 1 | Ref |
| 2196 | KJ489401 | Unassigned | Unassigned | Unassigned | 884_0 | Clustered | 28 | VC_884_0 | 28 | 0.6211 | 1 | 0.6211 | 0 | 0 | 0 | 0.9841 | Ref |
| 2197 | KJ489402 | Unassigned | Unassigned | Unassigned | 883_1 | Clustered | 12 | VC_883_1 | 12 | 0.2441 | 1 | 0.2441 | 0 | 0 | 0 | 1 | Ref |
| 2198 | KJ502657 | Unassigned | Unassigned | Unassigned | 1027_0 | Clustered | 20 | VC_1027_0 | 20 | 0.7114 | 1 | 0.7114 | 0 | 0 | 0 | 1 | Ref |
| 2199 | KJ507099 | Unassigned | Unassigned | Unassigned | 129_0 | Clustered | 43 | VC_129_0 | 43 | 0.886 | 1 | 0.886 | 0 | 0 | 0 | 1 | Ref |
| 2200 | KJ507100 | Unassigned | Unassigned | Unassigned | 697_0 | Clustered | 5 | VC_697_0 | 5 | 0.8303 | 0.99999971 | 0.8303 | 0 | 0 | 0 | 1 | Ref |
| 2201 | KJ510413 | Unassigned | Unassigned | Unassigned | 1046_0 | Clustered | 7 | VC_1046_0 | 7 | 0.9413 | 1 | 0.9413 | 0 | 0 | 0 | 1 | Ref |
| 2202 | KJ510414 | Unassigned | Unassigned | Unassigned | 517_1 | Clustered | 108 | VC_517_1 | 108 | 0.8483 | 1 | 0.8483 | 0 | 0 | 0 | 0.9955 | Ref |
| 2203 | KJ510415 | Unassigned | Unassigned | Unassigned | 513_0 | Clustered | 32 | VC_513_0 | 32 | 0.6881 | 1 | 0.6881 | 0 | 0 | 0 | 1 | Ref |
| 2204 | KJ528544 | Unassigned | Unassigned | Unassigned | 97_0 | Clustered | 5 | VC_97_0 | 5 | 0.0696 | 0.98113835 | 0.0683 | 0 | 0 | 0 | 1 | Ref |
| 2205 | KJ535721 | Unassigned | Unassigned | Unassigned | 739_0 | Clustered | 26 | VC_739_0 | 26 | 0.9571 | 1 | 0.9571 | 0 | 0 | 0 | 1 | Ref |
| 2206 | KJ535722 | Unassigned | Unassigned | Unassigned | 739_0 | Clustered | 26 | VC_739_0 | 26 | 0.9571 | 1 | 0.9571 | 0 | 0 | 0 | 1 | Ref |
| 2207 | KJ538721 | Unassigned | Unassigned | Unassigned | 939_0 | Clustered | 15 | VC_939_0 | 15 | 1 | 0 | 0 | 0 | 0 | 0 | 1 | Ref |
| 2208 | KJ538722 | Unassigned | Unassigned | Unassigned | 810_0 | Clustered | 18 | VC_810_0 | 18 | 0.9121 | 1 | 0.9121 | 0 | 0 | 0 | 1 | Ref |
| 2209 | KJ538723 | Unassigned | Unassigned | Unassigned | 371_3 | Clustered | 22 | VC_371_3 | 22 | 0.0787 | 1 | 0.0787 | 0 | 0 | 0 | 0.9048 | Ref |
| 2210 | KJ564036 | Unassigned | Unassigned | Unassigned | 640_0 | Clustered | 7 | VC_640_0 | 7 | 0.9025 | 1 | 0.9025 | 0 | 0 | 0 | 1 | Ref |
| 2211 | KJ564037 | Unassigned | Unassigned | Unassigned | 640_0 | Clustered | 7 | VC_640_0 | 7 | 0.9025 | 1 | 0.9025 | 0 | 0 | 0 | 1 | Ref |
| 2212 | KJ564038 | Unassigned | Unassigned | Unassigned | 640_0 | Clustered | 7 | VC_640_0 | 7 | 0.9025 | 1 | 0.9025 | 0 | 0 | 0 | 1 | Ref |
| 2213 | KJ567042 | Unassigned | Unassigned | Unassigned | 238_0 | Clustered | 17 | VC_238_0 | 17 | 0.6109 | 1 | 0.6109 | 0 | 0 | 0 | 1 | Ref |
| 2214 | KJ567043 | Unassigned | Unassigned | Unassigned | 563_0 | Clustered | 2 | VC_563_0 | 2 | 0.0727 | 0.956426 | 0.0695 | 0 | 0 | 0 | 1 | Ref |
| 2215 | KJ567044 | Unassigned | Unassigned | Unassigned | 371_2 | Clustered | 8 | VC_371_2 | 8 | 0.0232 | 1 | 0.0232 | 0 | 0 | 0 | 0.8571 | Ref |
| 2216 | KJ567045 | Unassigned | Unassigned | Unassigned | 236_0 | Clustered | 72 | VC_236_0 | 72 | 0.8104 | 1 | 0.8104 | 0 | 0 | 0 | 1 | Ref |
| 2217 | KJ567046 | Unassigned | Unassigned | Unassigned | 567_0 | Clustered | 8 | VC_567_0 | 8 | 0.4935 | 1 | 0.4935 | 0 | 0 | 0 | 1 | Ref |
| 2218 | KJ572844 | Unassigned | Unassigned | Unassigned | 121_0 | Clustered | 4 | VC_121_0 | 4 | 0.1814 | 0.99998531 | 0.1814 | 0 | 0 | 0 | 1 | Ref |
| 2219 | KJ578758 | Unassigned | Unassigned | Unassigned | 765_0 | Clustered | 132 | VC_765_0 | 132 | 1 | 1 | 1 | 0 | 0 | 0 | 1 | Ref |
| 2220 | KJ578759 | Unassigned | Unassigned | Unassigned | 765_0 | Clustered | 132 | VC_765_0 | 132 | 1 | 1 | 1 | 0 | 0 | 0 | 1 | Ref |
| 2221 | KJ578760 | Unassigned | Unassigned | Unassigned | 765_0 | Clustered | 132 | VC_765_0 | 132 | 1 | 1 | 1 | 0 | 0 | 0 | 1 | Ref |
| 2222 | KJ578761 | Unassigned | Unassigned | Unassigned | 765_0 | Clustered | 132 | VC_765_0 | 132 | 1 | 1 | 1 | 0 | 0 | 0 | 1 | Ref |
| 2223 | KJ578762 | Unassigned | Unassigned | Unassigned | 765_0 | Clustered | 132 | VC_765_0 | 132 | 1 | 1 | 1 | 0 | 0 | 0 | 1 | Ref |
| 2224 | KJ578763 | Unassigned | Unassigned | Unassigned | 765_0 | Clustered | 132 | VC_765_0 | 132 | 1 | 1 | 1 | 0 | 0 | 0 | 1 | Ref |
| 2225 | KJ578764 | Unassigned | Unassigned | Unassigned | 765_0 | Clustered | 132 | VC_765_0 | 132 | 1 | 1 | 1 | 0 | 0 | 0 | 1 | Ref |
| 2226 | KJ578765 | Unassigned | Unassigned | Unassigned | 765_0 | Clustered | 132 | VC_765_0 | 132 | 1 | 1 | 1 | 0 | 0 | 0 | 1 | Ref |
| 2227 | KJ578766 | Unassigned | Unassigned | Unassigned | 765_0 | Clustered | 132 | VC_765_0 | 132 | 1 | 1 | 1 | 0 | 0 | 0 | 1 | Ref |
| 2228 | KJ578767 | Unassigned | Unassigned | Unassigned | 765_0 | Clustered | 132 | VC_765_0 | 132 | 1 | 1 | 1 | 0 | 0 | 0 | 1 | Ref |
| 2229 | KJ578768 | Unassigned | Unassigned | Unassigned | 765_0 | Clustered | 132 | VC_765_0 | 132 | 1 | 1 | 1 | 0 | 0 | 0 | 1 | Ref |
| 2230 | KJ578769 | Unassigned | Unassigned | Unassigned | 765_0 | Clustered | 132 | VC_765_0 | 132 | 1 | 1 | 1 | 0 | 0 | 0 | 1 | Ref |
| 2231 | KJ578770 | Unassigned | Unassigned | Unassigned | 765_0 | Clustered | 132 | VC_765_0 | 132 | 1 | 1 | 1 | 0 | 0 | 0 | 1 | Ref |
| 2232 | KJ578771 | Unassigned | Unassigned | Unassigned | 765_0 | Clustered | 132 | VC_765_0 | 132 | 1 | 1 | 1 | 0 | 0 | 0 | 1 | Ref |
| 2233 | KJ578772 | Unassigned | Unassigned | Unassigned | 765_0 | Clustered | 132 | VC_765_0 | 132 | 1 | 1 | 1 | 0 | 0 | 0 | 1 | Ref |
| 2234 | KJ578773 | Unassigned | Unassigned | Unassigned | 765_0 | Clustered | 132 | VC_765_0 | 132 | 1 | 1 | 1 | 0 | 0 | 0 | 1 | Ref |
| 2235 | KJ578774 | Unassigned | Unassigned | Unassigned | 765_0 | Clustered | 132 | VC_765_0 | 132 | 1 | 1 | 1 | 0 | 0 | 0 | 1 | Ref |
| 2236 | KJ578775 | Unassigned | Unassigned | Unassigned | 765_0 | Clustered | 132 | VC_765_0 | 132 | 1 | 1 | 1 | 0 | 0 | 0 | 1 | Ref |
| 2237 | KJ578776 | Unassigned | Unassigned | Unassigned | 765_0 | Clustered | 132 | VC_765_0 | 132 | 1 | 1 | 1 | 0 | 0 | 0 | 1 | Ref |
| 2238 | KJ578777 | Unassigned | Unassigned | Unassigned | 765_0 | Clustered | 132 | VC_765_0 | 132 | 1 | 1 | 1 | 0 | 0 | 0 | 1 | Ref |
| 2239 | KJ578778 | Unassigned | Unassigned | Unassigned | 765_0 | Clustered | 132 | VC_765_0 | 132 | 1 | 1 | 1 | 0 | 0 | 0 | 1 | Ref |
| 2240 | KJ578779 | Unassigned | Unassigned | Unassigned | 765_0 | Clustered | 132 | VC_765_0 | 132 | 1 | 1 | 1 | 0 | 0 | 0 | 1 | Ref |
| 2241 | KJ578780 | Unassigned | Unassigned | Unassigned | 765_0 | Clustered | 132 | VC_765_0 | 132 | 1 | 1 | 1 | 0 | 0 | 0 | 1 | Ref |
| 2242 | KJ578781 | Unassigned | Unassigned | Unassigned | 765_0 | Clustered | 132 | VC_765_0 | 132 | 1 | 1 | 1 | 0 | 0 | 0 | 1 | Ref |
| 2243 | KJ578782 | Unassigned | Unassigned | Unassigned | 765_0 | Clustered | 132 | VC_765_0 | 132 | 1 | 1 | 1 | 0 | 0 | 0 | 1 | Ref |
| 2244 | KJ578783 | Unassigned | Unassigned | Unassigned | 765_0 | Clustered | 132 | VC_765_0 | 132 | 1 | 1 | 1 | 0 | 0 | 0 | 1 | Ref |
| 2245 | KJ578784 | Unassigned | Unassigned | Unassigned | 765_0 | Clustered | 132 | VC_765_0 | 132 | 1 | 1 | 1 | 0 | 0 | 0 | 1 | Ref |
| 2246 | KJ578785 | Unassigned | Unassigned | Unassigned | 765_0 | Clustered | 132 | VC_765_0 | 132 | 1 | 1 | 1 | 0 | 0 | 0 | 1 | Ref |
| 2247 | KJ578786 | Unassigned | Unassigned | Unassigned | 765_0 | Clustered | 132 | VC_765_0 | 132 | 1 | 1 | 1 | 0 | 0 | 0 | 1 | Ref |
| 2248 | KJ578787 | Unassigned | Unassigned | Unassigned | 765_0 | Clustered | 132 | VC_765_0 | 132 | 1 | 1 | 1 | 0 | 0 | 0 | 1 | Ref |
| 2249 | KJ578788 | Unassigned | Unassigned | Unassigned | 765_0 | Clustered | 132 | VC_765_0 | 132 | 1 | 1 | 1 | 0 | 0 | 0 | 1 | Ref |
| 2250 | KJ578789 | Unassigned | Unassigned | Unassigned | 765_0 | Clustered | 132 | VC_765_0 | 132 | 1 | 1 | 1 | 0 | 0 | 0 | 1 | Ref |
| 2251 | KJ578790 | Unassigned | Unassigned | Unassigned | 765_0 | Clustered | 132 | VC_765_0 | 132 | 1 | 1 | 1 | 0 | 0 | 0 | 1 | Ref |
| 2252 | KJ578791 | Unassigned | Unassigned | Unassigned | 765_0 | Clustered | 132 | VC_765_0 | 132 | 1 | 1 | 1 | 0 | 0 | 0 | 1 | Ref |
| 2253 | KJ578792 | Unassigned | Unassigned | Unassigned | 765_0 | Clustered | 132 | VC_765_0 | 132 | 1 | 1 | 1 | 0 | 0 | 0 | 1 | Ref |
| 2254 | KJ586794 | Unassigned | Unassigned | Unassigned | 739_0 | Clustered | 26 | VC_739_0 | 26 | 0.9571 | 1 | 0.9571 | 0 | 0 | 0 | 1 | Ref |
| 2255 | KJ586795 | Unassigned | Unassigned | Unassigned | 739_0 | Clustered | 26 | VC_739_0 | 26 | 0.9571 | 1 | 0.9571 | 0 | 0 | 0 | 1 | Ref |
| 2256 | KJ591604 | Unassigned | Unassigned | Unassigned | 739_0 | Clustered | 26 | VC_739_0 | 26 | 0.9571 | 1 | 0.9571 | 0 | 0 | 0 | 1 | Ref |
| 2257 | KJ591605 | Unassigned | Unassigned | Unassigned | 739_0 | Clustered | 26 | VC_739_0 | 26 | 0.9571 | 1 | 0.9571 | 0 | 0 | 0 | 1 | Ref |
| 2258 | KJ595575 | Unassigned | Unassigned | Unassigned | 558_50 | Clustered | 15 | VC_558_50 | 15 | 0.0468 | 1 | 0.0468 | 0 | 0 | 0 | 0.9238 | Ref |
| 2259 | KJ595576 | Unassigned | Unassigned | Unassigned | 371_1 | Clustered | 11 | VC_371_1 | 11 | 0.0379 | 1 | 0.0379 | 0 | 0 | 0 | 0.8545 | Ref |
| 2260 | KJ596420 | Unassigned | Unassigned | Unassigned | 10_0 | Clustered | 25 | VC_10_0 | 25 | 0.3467 | 1 | 0.3467 | 0 | 0 | 0 | 0.9733 | Ref |
| 2261 | KJ603229 | Unassigned | Unassigned | Unassigned | 46_0 | Clustered | 2 | VC_46_0 | 2 | 0.0181 | 0.95763537 | 0.0173 | 0 | 0 | 0 | 1 | Ref |
| 2262 | KJ608189 | Unassigned | Unassigned | Unassigned | 269_0 | Clustered | 3 | VC_269_0 | 3 | 0.1948 | 0.99308177 | 0.1934 | 0 | 0 | 0 | 1 | Ref |
| 2263 | KJ617393 | Unassigned | Unassigned | Unassigned | 1048_0 | Clustered | 3 | VC_1048_0 | 3 | 1 | 1 | 1 | 0 | 0 | 0 | 1 | Ref |
| 2264 | KJ619459 | Unassigned | Unassigned | Unassigned | 875_0 | Clustered | 10 | VC_875_0 | 10 | 0.988 | 0.99948433 | 0.9875 | 0 | 0 | 0 | 1 | Ref |
| 2265 | KJ621082 | Unassigned | Unassigned | Unassigned | 833_0 | Clustered | 16 | VC_833_0 | 16 | 0.9351 | 1 | 0.9351 | 0 | 0 | 0 | 1 | Ref |
| 2266 | KJ628499 | Unassigned | Unassigned | Unassigned | 700_0 | Clustered | 10 | VC_700_0 | 10 | 0.8076 | 1 | 0.8076 | 0 | 0 | 0 | 1 | Ref |

| | | | | | | | | | | | | | | | | |
|---|---|---|---|---|---|---|---|---|---|---|---|---|---|---|---|---|
| 2267 | KJ668713 | Unassigned | Unassigned | Unassigned | 455_0 | Clustered | 18 | VC_455_0 | 18 | 0.4598 | 1 | 0.4598 | 0 | 0 | 0 | 1 Ref |
| 2268 | KJ668714 | Unassigned | Unassigned | Unassigned | 283_99 | Clustered/Singleton | 1 | VC_283_99 | 1 | 0 | 1 | 0 | 0 | 0 | 0 | 1 Ref |
| 2269 | KJ668715 | Unassigned | Unassigned | Unassigned | 739_0 | Clustered | 26 | VC_739_0 | 26 | 0.9571 | 1 | 0.9571 | 0 | 0 | 0 | 1 Ref |
| 2270 | KJ676859 | Unassigned | Unassigned | Unassigned | 187_2 | Clustered | 2 | VC_187_2 | 2 | 0.0209 | 0.93057595 | 0.0195 | 0 | 0 | 0 | 1 Ref |
| 2271 | KJ690250 | Unassigned | Unassigned | Unassigned | 325_51 | Clustered | 2 | VC_325_51 | 2 | 0.0039 | 0.95663826 | 0.0037 | 0 | 0 | 0 | 1 Ref |
| 2272 | KJ716334 | Unassigned | Unassigned | Unassigned | 9_0 | Clustered | 38 | VC_9_0 | 38 | 0.4895 | 1 | 0.4895 | 0 | 0 | 0 | 1 Ref |
| 2273 | KJ716335 | Unassigned | Unassigned | Unassigned | 131_0 | Clustered | 17 | VC_131_0 | 17 | 0.2671 | 1 | 0.2671 | 0 | 0 | 0 | 1 Ref |
| 2274 | KJ722067 | Unassigned | Unassigned | Unassigned | 765_0 | Clustered | 132 | VC_765_0 | 132 | 1 | 1 | 1 | 0 | 0 | 0 | 1 Ref |
| 2275 | KJ725374 | Unassigned | Unassigned | Unassigned | 761_0 | Clustered | 67 | VC_761_0 | 67 | 0.8947 | 1 | 0.8947 | 0 | 0 | 0 | 1 Ref |
| 2276 | KJ746502 | Unassigned | Unassigned | Unassigned | 21_0 | Clustered | 4 | VC_21_0 | 4 | 0.5696 | 0.87262442 | 0.4971 | 0 | 0 | 0 | 0.6667 Ref |
| 2277 | KJ748011 | Unassigned | Unassigned | Unassigned | 451_0 | Clustered | 80 | VC_451_0 | 80 | 0.9991 | 1 | 0.9991 | 0 | 0 | 0 | 1 Ref |
| 2278 | KJ801817 | Unassigned | Unassigned | Unassigned | 91_0 | Clustered | 22 | VC_91_0 | 22 | 0.5318 | 1 | 0.5318 | 0 | 0 | 0 | 1 Ref |
| 2279 | KJ802832 | Unassigned | Unassigned | Unassigned | 1049_0 | Clustered | 8 | VC_1049_0 | 8 | 0.6413 | 1 | 0.6413 | 0 | 0 | 0 | 1 Ref |
| 2280 | KJ803031 | Unassigned | Unassigned | Unassigned | 833_0 | Clustered | 16 | VC_833_0 | 16 | 0.9351 | 1 | 0.9351 | 0 | 0 | 0 | 1 Ref |
| 2281 | KJ804259 | Unassigned | Unassigned | Unassigned | 667_0 | Clustered | 7 | VC_667_0 | 7 | 0.4101 | 1 | 0.4101 | 0 | 0 | 0 | 1 Ref |
| 2282 | KJ817802 | Unassigned | Unassigned | Unassigned | 698_0 | Clustered | 26 | VC_698_0 | 26 | 0.9366 | 1 | 0.9366 | 0 | 0 | 0 | 1 Ref |
| 2283 | KJ829260 | Unassigned | Unassigned | Unassigned | 556_0 | Clustered | 21 | VC_556_0 | 21 | 0.9912 | 1 | 0.9912 | 0 | 0 | 0 | 1 Ref |
| 2284 | KJ832078 | Unassigned | Unassigned | Unassigned | 30_1 | Clustered | 16 | VC_30_1 | 16 | 0.1973 | 1 | 0.1973 | 0 | 0 | 0 | 0.8333 Ref |
| 2285 | KJ858521 | Unassigned | Unassigned | Unassigned | 793_0 | Clustered | 12 | VC_793_0 | 12 | 0.8531 | 1 | 0.8531 | 0 | 0 | 0 | 1 Ref |
| 2286 | KJ888149 | Unassigned | Unassigned | Unassigned | 149_16 | Clustered/Singleton | 1 | VC_149_16 | 1 | 0 | 1 | 0 | 0 | 0 | 0 | 1 Ref |
| 2287 | KJ909655 | Unassigned | Unassigned | Unassigned | 39_0 | Clustered | 35 | VC_39_0 | 35 | 0.4337 | 1 | 0.4337 | 0 | 0 | 0 | 1 Ref |
| 2288 | KJ936628 | Unassigned | Unassigned | Unassigned | 530_0 | Clustered | 9 | VC_530_0 | 9 | 0.8027 | 1 | 0.8027 | 0 | 0 | 0 | 1 Ref |
| 2289 | KJ944841 | Unassigned | Unassigned | Unassigned | 234_0 | Clustered | 21 | VC_234_0 | 21 | 0.8355 | 1 | 0.8355 | 0 | 0 | 0 | 1 Ref |
| 2290 | KJ959591 | Unassigned | Unassigned | Unassigned | 747_0 | Clustered | 23 | VC_747_0 | 23 | 0.8202 | 1 | 0.8202 | 0 | 0 | 0 | 1 Ref |
| 2291 | KJ959632 | Unassigned | Unassigned | Unassigned | 216_4 | Clustered | 13 | VC_216_4 | 13 | 0.1657 | 1 | 0.1657 | 0 | 0 | 0 | 1 Ref |
| 2292 | KJ997912 | Unassigned | Unassigned | Unassigned | 303_0 | Clustered | 155 | VC_303_0 | 155 | 0.9979 | 1 | 0.9979 | 0 | 0 | 0 | 1 Ref |
| 2293 | KM044272 | Unassigned | Unassigned | Unassigned | 813_1 | Clustered | 24 | VC_813_1 | 24 | 0.535 | 1 | 0.535 | 0 | 0 | 0 | 1 Ref |
| 2294 | KM051843 | Unassigned | Unassigned | Unassigned | 85_0 | Clustered | 3 | VC_85_0 | 3 | 0.0422 | 0.99823242 | 0.0421 | 0 | 0 | 0 | 1 Ref |
| 2295 | KM058087 | Unassigned | Unassigned | Unassigned | 1051_0 | Clustered | 2 | VC_1051_0 | 2 | 1 | 1 | 1 | 0 | 0 | 0 | 1 Ref |
| 2296 | KM067278 | Unassigned | Unassigned | Unassigned | 423_0 | Clustered | 45 | VC_423_0 | 45 | 0.9484 | 1 | 0.9484 | 0 | 0 | 0 | 1 Ref |
| 2297 | KM091442 | Unassigned | Unassigned | Unassigned | 202_0 | Clustered | 102 | VC_202_0 | 102 | 0.4049 | 1 | 0.4049 | 0 | 0 | 0 | 0.7876 Ref |
| 2298 | KM092515 | Unassigned | Unassigned | Unassigned | 329_17 | Clustered | 4 | VC_329_17 | 4 | 0.0163 | 0.99996247 | 0.0163 | 0 | 0 | 0 | 1 Ref |
| 2299 | KM101117 | Unassigned | Unassigned | Unassigned | 508_0 | Clustered | 32 | VC_508_0 | 32 | 0.906 | 1 | 0.906 | 0 | 0 | 0 | 1 Ref |
| 2300 | KM101118 | Unassigned | Unassigned | Unassigned | 940_0 | Clustered | 34 | VC_940_0 | 34 | 0.7959 | 1 | 0.7959 | 0 | 0 | 0 | 1 Ref |
| 2301 | KM101119 | Unassigned | Unassigned | Unassigned | 514_0 | Clustered | 75 | VC_514_0 | 75 | 0.9402 | 1 | 0.9402 | 0 | 0 | 0 | 1 Ref |
| 2302 | KM101120 | Unassigned | Unassigned | Unassigned | 515_0 | Clustered | 16 | VC_515_0 | 16 | 0.3793 | 1 | 0.3793 | 0 | 0 | 0 | 1 Ref |
| 2303 | KM101122 | Unassigned | Unassigned | Unassigned | 520_49 | Clustered/Singleton | 1 | VC_520_49 | 1 | 0 | 1 | 0 | 0 | 0 | 0 | 1 Ref |
| 2304 | KM101123 | Unassigned | Unassigned | Unassigned | 546_7 | Clustered | 4 | VC_546_7 | 4 | 0.041 | 0.9963251 | 0.0408 | 0 | 0 | 0 | 1 Ref |
| 2305 | KM190144 | Unassigned | Unassigned | Unassigned | 582_2 | Clustered/Singleton | 1 | VC_582_2 | 1 | 0 | 1 | 0 | 0 | 0 | 0 | 1 Ref |
| 2306 | KM197169 | Unassigned | Unassigned | Unassigned | 216_4 | Clustered | 13 | VC_216_4 | 13 | 0.1657 | 1 | 0.1657 | 0 | 0 | 0 | 1 Ref |
| 2307 | KM199770 | Unassigned | Unassigned | Unassigned | 1052_3 | Clustered | 5 | VC_1052_3 | 5 | 0.185 | 0.98656048 | 0.1825 | 0 | 0 | 0 | 1 Ref |
| 2308 | KM199771 | Unassigned | Unassigned | Unassigned | 481_0 | Clustered | 8 | VC_481_0 | 8 | 0.1348 | 1 | 0.1348 | 0 | 0 | 0 | 1 Ref |
| 2309 | KM209228 | Unassigned | Unassigned | Unassigned | 131_0 | Clustered | 17 | VC_131_0 | 17 | 0.2671 | 1 | 0.2671 | 0 | 0 | 0 | 1 Ref |
| 2310 | KM209270 | Unassigned | Unassigned | Unassigned | 131_0 | Clustered | 17 | VC_131_0 | 17 | 0.2671 | 1 | 0.2671 | 0 | 0 | 0 | 1 Ref |
| 2311 | KM209320 | Unassigned | Unassigned | Unassigned | 131_0 | Clustered | 17 | VC_131_0 | 17 | 0.2671 | 1 | 0.2671 | 0 | 0 | 0 | 1 Ref |
| 2312 | KM216423 | Unassigned | Unassigned | Unassigned | 149_15 | Clustered | 16 | VC_149_15 | 16 | 0.0788 | 1 | 0.0788 | 0 | 0 | 0 | 0.8083 Ref |
| 2313 | KM224878 | Unassigned | Unassigned | Unassigned | 754_0 | Clustered | 8 | VC_754_0 | 8 | 0.752 | 1 | 0.752 | 0 | 0 | 0 | 1 Ref |
| 2314 | KM224879 | Unassigned | Unassigned | Unassigned | 754_0 | Clustered | 8 | VC_754_0 | 8 | 0.752 | 1 | 0.752 | 0 | 0 | 0 | 1 Ref |
| 2315 | KM233151 | Unassigned | Unassigned | Unassigned | 835_0 | Clustered | 67 | VC_835_0 | 67 | 0.9624 | 1 | 0.9624 | 0 | 0 | 0 | 1 Ref |
| 2316 | KM233261 | Unassigned | Unassigned | Unassigned | 907_0 | Clustered | 3 | VC_907_0 | 3 | 0.6642 | 0.99633114 | 0.6618 | 0 | 0 | 0 | 1 Ref |
| 2317 | KM233454 | Unassigned | Unassigned | Unassigned | 514_0 | Clustered | 75 | VC_514_0 | 75 | 0.9402 | 1 | 0.9402 | 0 | 0 | 0 | 1 Ref |
| 2318 | KM233455 | Unassigned | Unassigned | Unassigned | 514_0 | Clustered | 75 | VC_514_0 | 75 | 0.9402 | 1 | 0.9402 | 0 | 0 | 0 | 1 Ref |
| 2319 | KM233689 | Unassigned | Unassigned | Unassigned | 179_0 | Clustered | 53 | VC_179_0 | 53 | 0.9137 | 1 | 0.9137 | 0 | 0 | 0 | 1 Ref |
| 2320 | KM236237 | Unassigned | Unassigned | Unassigned | 669_1 | Clustered | 10 | VC_669_1 | 10 | 0.1268 | 1 | 0.1268 | 0 | 0 | 0 | 0.9778 Ref |
| 2321 | KM236239 | Unassigned | Unassigned | Unassigned | 330_0 | Clustered | 27 | VC_330_0 | 27 | 0.5088 | 1 | 0.5088 | 0 | 0 | 0 | 0.8661 Ref |
| 2322 | KM236241 | Unassigned | Unassigned | Unassigned | 584_0 | Clustered | 28 | VC_584_0 | 28 | 0.7704 | 1 | 0.7704 | 0 | 0 | 0 | 1 Ref |
| 2323 | KM236242 | Unassigned | Unassigned | Unassigned | 780_0 | Clustered | 8 | VC_780_0 | 8 | 0.5207 | 1 | 0.5207 | 0 | 0 | 0 | 1 Ref |
| 2324 | KM236243 | Unassigned | Unassigned | Unassigned | 795_0 | Clustered | 19 | VC_795_0 | 19 | 0.6848 | 1 | 0.6848 | 0 | 0 | 0 | 1 Ref |
| 2325 | KM236244 | Unassigned | Unassigned | Unassigned | 191_19 | Clustered | 12 | VC_191_19 | 12 | 0.0655 | 1 | 0.0655 | 0 | 0 | 0 | 0.8636 Ref |
| 2326 | KM236245 | Unassigned | Unassigned | Unassigned | 85_9 | Clustered/Singleton | 1 | VC_85_9 | 1 | 0 | 1 | 0 | 0 | 0 | 0 | 1 Ref |
| 2327 | KM236246 | Unassigned | Unassigned | Unassigned | 85_7 | Clustered/Singleton | 1 | VC_85_7 | 1 | 0 | 1 | 0 | 0 | 0 | 0 | 1 Ref |
| 2328 | KM236247 | Unassigned | Unassigned | Unassigned | 1032_0 | Clustered | 7 | VC_1032_0 | 7 | 0.9887 | 0.99999504 | 0.9887 | 0 | 0 | 0 | 1 Ref |
| 2329 | KM236248 | Unassigned | Unassigned | Unassigned | 1032_0 | Clustered | 7 | VC_1032_0 | 7 | 0.9887 | 0.99999504 | 0.9887 | 0 | 0 | 0 | 1 Ref |
| 2330 | KM236502 | Unassigned | Unassigned | Unassigned | 371_3 | Clustered | 22 | VC_371_3 | 22 | 0.0787 | 1 | 0.0787 | 0 | 0 | 0 | 0.9048 Ref |
| 2331 | KM242061 | Unassigned | Unassigned | Unassigned | 392_0 | Clustered | 126 | VC_392_0 | 126 | 0.9621 | 1 | 0.9621 | 0 | 0 | 0 | 1 Ref |
| 2332 | KM247287 | Unassigned | Unassigned | Unassigned | 156_0 | Clustered | 12 | VC_156_0 | 12 | 0.9062 | 1 | 0.9062 | 0 | 0 | 0 | 1 Ref |
| 2333 | KM253764 | Unassigned | Unassigned | Unassigned | 385_0 | Clustered | 51 | VC_385_0 | 51 | 0.8754 | 1 | 0.8754 | 0 | 0 | 0 | 1 Ref |
| 2334 | KM262191 | Unassigned | Unassigned | Unassigned | 862_0 | Clustered | 11 | VC_862_0 | 11 | 0.6745 | 1 | 0.6745 | 0 | 0 | 0 | 1 Ref |
| 2335 | KM262192 | Unassigned | Unassigned | Unassigned | 862_0 | Clustered | 11 | VC_862_0 | 11 | 0.6745 | 1 | 0.6745 | 0 | 0 | 0 | 1 Ref |
| 2336 | KM272358 | Unassigned | Unassigned | Unassigned | 637_2 | Clustered | 2 | VC_637_2 | 2 | 0.0038 | 0.92844927 | 0.0035 | 0 | 0 | 0 | 1 Ref |
| 2337 | KM279937 | Unassigned | Unassigned | Unassigned | 520_56 | Clustered/Singleton | 1 | VC_520_56 | 1 | 0 | 1 | 0 | 0 | 0 | 0 | 1 Ref |
| 2338 | KM289195 | Unassigned | Unassigned | Unassigned | 636_0 | Clustered | 9 | VC_636_0 | 9 | 0.2422 | 1 | 0.2422 | 0 | 0 | 0 | 0.9722 Ref |
| 2339 | KM347889 | Unassigned | Unassigned | Unassigned | 520_68 | Clustered/Singleton | 1 | VC_520_68 | 1 | 0 | 1 | 0 | 0 | 0 | 0 | 1 Ref |
| 2340 | KM347890 | Unassigned | Unassigned | Unassigned | 371_1 | Clustered | 11 | VC_371_1 | 11 | 0.0379 | 1 | 0.0379 | 0 | 0 | 0 | 0.8545 Ref |
| 2341 | KM359505 | Unassigned | Unassigned | Unassigned | 432_8 | Clustered/Singleton | 1 | VC_432_8 | 1 | 0 | 1 | 0 | 0 | 0 | 0 | 1 Ref |
| 2342 | KM360178 | Unassigned | Unassigned | Unassigned | 125_0 | Clustered | 17 | VC_125_0 | 17 | 0.9244 | 1 | 0.9244 | 0 | 0 | 0 | 1 Ref |
| 2343 | KM363596 | Unassigned | Unassigned | Unassigned | 238_0 | Clustered | 17 | VC_238_0 | 17 | 0.6109 | 1 | 0.6109 | 0 | 0 | 0 | 1 Ref |
| 2344 | KM363597 | Unassigned | Unassigned | Unassigned | 371_13 | Clustered | 33 | VC_371_13 | 33 | 0.179 | 1 | 0.179 | 0 | 0 | 0 | 0.8769 Ref |
| 2345 | KM366096 | Unassigned | Unassigned | Unassigned | 386_0 | Clustered | 49 | VC_386_0 | 49 | 0.7941 | 1 | 0.7941 | 0 | 0 | 0 | 1 Ref |
| 2346 | KM366097 | Unassigned | Unassigned | Unassigned | 452_0 | Clustered | 66 | VC_452_0 | 66 | 0.9248 | 1 | 0.9248 | 0 | 0 | 0 | 1 Ref |
| 2347 | KM366098 | Unassigned | Unassigned | Unassigned | 592_0 | Clustered | 50 | VC_592_0 | 50 | 0.7314 | 1 | 0.7314 | 0 | 0 | 0 | 1 Ref |
| 2348 | KM366099 | Unassigned | Unassigned | Unassigned | 428_0 | Clustered | 33 | VC_428_0 | 33 | 0.9915 | 1 | 0.9915 | 0 | 0 | 0 | 1 Ref |
| 2349 | KM366100 | Unassigned | Unassigned | Unassigned | 96_0 | Clustered | 30 | VC_96_0 | 30 | 0.718 | 1 | 0.718 | 0 | 0 | 0 | 1 Ref |
| 2350 | KM370384 | Unassigned | Unassigned | Unassigned | 452_0 | Clustered | 66 | VC_452_0 | 66 | 0.9248 | 1 | 0.9248 | 0 | 0 | 0 | 1 Ref |
| 2351 | KM373208 | Unassigned | Unassigned | Unassigned | 739_0 | Clustered | 26 | VC_739_0 | 26 | 0.9571 | 1 | 0.9571 | 0 | 0 | 0 | 1 Ref |
| 2352 | KM389210 | Unassigned | Unassigned | Unassigned | 297_0 | Clustered | 5 | VC_297_0 | 5 | 0.1056 | 0.99999993 | 0.1056 | 0 | 0 | 0 | 0.9 Ref |
| 2353 | KM400683 | Unassigned | Unassigned | Unassigned | 546_2 | Clustered | 3 | VC_546_2 | 3 | 0.0275 | 0.85260342 | 0.0234 | 0 | 0 | 0 | 1 Ref |
| 2354 | KM401838 | Unassigned | Unassigned | Unassigned | 223_0 | Clustered | 38 | VC_223_0 | 38 | 0.9095 | 1 | 0.9095 | 0 | 0 | 0 | 1 Ref |
| 2355 | KM402757 | Unassigned | Unassigned | Unassigned | 520_92 | Clustered | 5 | VC_520_92 | 5 | 0.0663 | 0.99999997 | 0.0663 | 0 | 0 | 0 | 1 Ref |
| 2356 | KM407600 | Unassigned | Unassigned | Unassigned | 286_10 | Clustered | 9 | VC_286_10 | 9 | 0.0615 | 0.99999878 | 0.0615 | 0 | 0 | 0 | 0.6667 Ref |
| 2357 | KM408320 | Unassigned | Unassigned | Unassigned | 371_15 | Clustered | 44 | VC_371_15 | 44 | 0.1391 | 1 | 0.1391 | 0 | 0 | 0 | 0.7474 Ref |
| 2358 | KM411958 | Unassigned | Unassigned | Unassigned | 838_0 | Clustered | 29 | VC_838_0 | 29 | 0.8611 | 1 | 0.8611 | 0 | 0 | 0 | 1 Ref |
| 2359 | KM411959 | Unassigned | Unassigned | Unassigned | 838_0 | Clustered | 29 | VC_838_0 | 29 | 0.8611 | 1 | 0.8611 | 0 | 0 | 0 | 1 Ref |
| 2360 | KM411960 | Unassigned | Unassigned | Unassigned | 838_0 | Clustered | 29 | VC_838_0 | 29 | 0.8611 | 1 | 0.8611 | 0 | 0 | 0 | 1 Ref |
| 2361 | KM434184 | Unassigned | Unassigned | Unassigned | 79_8 | Clustered/Singleton | 1 | VC_79_8 | 1 | 0 | 1 | 0 | 0 | 0 | 0 | 1 Ref |
| 2362 | KM434185 | Unassigned | Unassigned | Unassigned | 749_0 | Clustered | 40 | VC_749_0 | 40 | 0.7517 | 1 | 0.7517 | 0 | 0 | 0 | 1 Ref |
| 2363 | KM434186 | Unassigned | Unassigned | Unassigned | 77_0 | Clustered | 20 | VC_77_0 | 20 | 0.9657 | 1 | 0.9657 | 0 | 0 | 0 | 1 Ref |
| 2364 | KM458633 | Unassigned | Unassigned | Unassigned | 592_0 | Clustered | 50 | VC_592_0 | 50 | 0.7314 | 1 | 0.7314 | 0 | 0 | 0 | 1 Ref |
| 2365 | KM463009 | Unassigned | Unassigned | Unassigned | 216_6 | Clustered | 42 | VC_216_6 | 42 | 0.473 | 1 | 0.473 | 0 | 0 | 0 | 0.8351 Ref |
| 2366 | KM501444 | Unassigned | Unassigned | Unassigned | 283_145 | Clustered/Singleton | 1 | VC_283_145 | 1 | 0 | 1 | 0 | 0 | 0 | 0 | 1 Ref |
| 2367 | KM507819 | Unassigned | Unassigned | Unassigned | 164_11 | Clustered | 10 | VC_164_11 | 10 | 0.1507 | 1 | 0.1507 | 0 | 0 | 0 | 0.9111 Ref |
| 2368 | KM576124 | Unassigned | Unassigned | Unassigned | 111_0 | Clustered | 91 | VC_111_0 | 91 | 0.9587 | 1 | 0.9587 | 0 | 0 | 0 | 1 Ref |
| 2369 | KM581061 | Unassigned | Unassigned | Unassigned | 593_0 | Clustered | 5 | VC_593_0 | 5 | 0.4608 | 0.99999995 | 0.4608 | 0 | 0 | 0 | 1 Ref |
| 2370 | KM588359 | Unassigned | Unassigned | Unassigned | 521_0 | Clustered | 37 | VC_521_0 | 37 | 0.912 | 1 | 0.912 | 0 | 0 | 0 | 1 Ref |
| 2371 | KM589498 | Unassigned | Unassigned | Unassigned | 1054_0 | Clustered | 12 | VC_1054_0 | 12 | 0.2863 | 0.99746644 | 0.2855 | 0 | 0 | 0 | 1 Ref |
| 2372 | KM589499 | Unassigned | Unassigned | Unassigned | 1061_0 | Clustered | 2 | VC_1061_0 | 2 | 0.7065 | 0.76024994 | 0.5371 | 0 | 0 | 0 | 1 Ref |
| 2373 | KM589500 | Unassigned | Unassigned | Unassigned | 1062_0 | Clustered | 45 | VC_1062_0 | 45 | 0.9734 | 0.50949847 | 0.4959 | 0 | 0 | 0 | 1 Ref |
| 2374 | KM589502 | Unassigned | Unassigned | Unassigned | 995_0 | Clustered | 69 | VC_995_0 | 69 | 0.8547 | 1 | 0.8547 | 0 | 0 | 0 | 1 Ref |
| 2375 | KM589503 | Unassigned | Unassigned | Unassigned | 1062_0 | Clustered | 45 | VC_1062_0 | 45 | 0.9734 | 0.50949847 | 0.4959 | 0 | 0 | 0 | 1 Ref |
| 2376 | KM589504 | Unassigned | Unassigned | Unassigned | 1062_0 | Clustered | 45 | VC_1062_0 | 45 | 0.9734 | 0.50949847 | 0.4959 | 0 | 0 | 0 | 1 Ref |
| 2377 | KM589505 | Unassigned | Unassigned | Unassigned | 1062_0 | Clustered | 45 | VC_1062_0 | 45 | 0.9734 | 0.50949847 | 0.4959 | 0 | 0 | 0 | 1 Ref |
| 2378 | KM589507 | Unassigned | Unassigned | Unassigned | 1061_0 | Clustered | 2 | VC_1061_0 | 2 | 0.7065 | 0.76024994 | 0.5371 | 0 | 0 | 0 | 1 Ref |
| 2379 | KM589508 | Unassigned | Unassigned | Unassigned | 1066_0 | Clustered | 52 | VC_1066_0 | 52 | 0.7682 | 1 | 0.7682 | 0 | 0 | 0 | 1 Ref |
| 2380 | KM589509 | Unassigned | Unassigned | Unassigned | 1062_0 | Clustered | 45 | VC_1062_0 | 45 | 0.9734 | 0.50949847 | 0.4959 | 0 | 0 | 0 | 1 Ref |
| 2381 | KM589510 | Unassigned | Unassigned | Unassigned | 1062_0 | Clustered | 45 | VC_1062_0 | 45 | 0.9734 | 0.50949847 | 0.4959 | 0 | 0 | 0 | 1 Ref |
| 2382 | KM589511 | Unassigned | Unassigned | Unassigned | 995_0 | Clustered | 69 | VC_995_0 | 69 | 0.8547 | 1 | 0.8547 | 0 | 0 | 0 | 1 Ref |
| 2383 | KM589512 | Unassigned | Unassigned | Unassigned | 1062_0 | Clustered | 45 | VC_1062_0 | 45 | 0.9734 | 0.50949847 | 0.4959 | 0 | 0 | 0 | 1 Ref |
| 2384 | KM589513 | Unassigned | Unassigned | Unassigned | 1062_0 | Clustered | 45 | VC_1062_0 | 45 | 0.9734 | 0.50949847 | 0.4959 | 0 | 0 | 0 | 1 Ref |
| 2385 | KM589514 | Unassigned | Unassigned | Unassigned | 1068_0 | Clustered | 5 | VC_1068_0 | 5 | 1 | 1 | 1 | 0 | 0 | 0 | 1 Ref |
| 2386 | KM589515 | Unassigned | Unassigned | Unassigned | 1069_0 | Clustered | 4 | VC_1069_0 | 4 | 1 | 1 | 1 | 0 | 0 | 0 | 1 Ref |
| 2387 | KM589516 | Unassigned | Unassigned | Unassigned | 1069_0 | Clustered | 4 | VC_1069_0 | 4 | 1 | 1 | 1 | 0 | 0 | 0 | 1 Ref |
| 2388 | KM591905 | Unassigned | Unassigned | Unassigned | 1046_0 | Clustered | 7 | VC_1046_0 | 7 | 0.9413 | 1 | 0.9413 | 0 | 0 | 0 | 1 Ref |
| 2389 | KM591906 | Unassigned | Unassigned | Unassigned | 216_6 | Clustered | 42 | VC_216_6 | 42 | 0.473 | 1 | 0.473 | 0 | 0 | 0 | 0.8351 Ref |
| 2390 | KM592966 | Unassigned | Unassigned | Unassigned | 513_0 | Clustered | 32 | VC_513_0 | 32 | 0.6881 | 1 | 0.6881 | 0 | 0 | 0 | 1 Ref |
| 2391 | KM597530 | Unassigned | Unassigned | Unassigned | 520_29 | Clustered/Singleton | 1 | VC_520_29 | 1 | 0 | 1 | 0 | 0 | 0 | 0 | 1 Ref |
| 2392 | KM597531 | Unassigned | Unassigned | Unassigned | 325_35 | Clustered | 17 | VC_325_35 | 17 | 0.0732 | 1 | 0.0732 | 0 | 0 | 0 | 0.7721 Ref |
| 2393 | KM606994 | Unassigned | Unassigned | Unassigned | 283_82 | Clustered | 6 | VC_283_82 | 6 | 0.013 | 1 | 0.013 | 0 | 0 | 0 | 1 Ref |
| 2394 | KM606995 | Unassigned | Unassigned | Unassigned | 283_82 | Clustered | 6 | VC_283_82 | 6 | 0.013 | 1 | 0.013 | 0 | 0 | 0 | 1 Ref |
| 2395 | KM606996 | Unassigned | Unassigned | Unassigned | 283_82 | Clustered | 6 | VC_283_82 | 6 | 0.013 | 1 | 0.013 | 0 | 0 | 0 | 1 Ref |
| 2396 | KM606997 | Unassigned | Unassigned | Unassigned | 283_82 | Clustered | 6 | VC_283_82 | 6 | 0.013 | 1 | 0.013 | 0 | 0 | 0 | 1 Ref |
| 2397 | KM606998 | Unassigned | Unassigned | Unassigned | 283_82 | Clustered | 6 | VC_283_82 | 6 | 0.013 | 1 | 0.013 | 0 | 0 | 0 | 1 Ref |
| 2398 | KM606999 | Unassigned | Unassigned | Unassigned | 283_82 | Clustered | 6 | VC_283_82 | 6 | 0.013 | 1 | 0.013 | 0 | 0 | 0 | 1 Ref |
| 2399 | KM607000 | Unassigned | Unassigned | Unassigned | 283_83 | Clustered/Singleton | 1 | VC_283_83 | 1 | 0 | 1 | 0 | 0 | 0 | 0 | 1 Ref |
| 2400 | KM607001 | Unassigned | Unassigned | Unassigned | 283_94 | Clustered | 2 | VC_283_94 | 2 | 0.0026 | 0.95293762 | 0.0025 | 0 | 0 | 0 | 1 Ref |
| 2401 | KM607002 | Unassigned | Unassigned | Unassigned | 283_91 | Clustered | 11 | VC_283_91 | 11 | 0.0285 | 1 | 0.0285 | 0 | 0 | 0 | 1 Ref |
| 2402 | KM607003 | Unassigned | Unassigned | Unassigned | 283_91 | Clustered | 11 | VC_283_91 | 11 | 0.0285 | 1 | 0.0285 | 0 | 0 | 0 | 1 Ref |
| 2403 | KM607004 | Unassigned | Unassigned | Unassigned | 283_3 | Clustered | 3 | VC_283_3 | 3 | 0.0055 | 0.9968017 | 0.0055 | 0 | 0 | 0 | 1 Ref |
| 2404 | KM612259 | Unassigned | Unassigned | Unassigned | 607_0 | Clustered | 16 | VC_607_0 | 16 | 0.9179 | 1 | 0.9179 | 0 | 0 | 0 | 1 Ref |
| 2405 | KM612260 | Unassigned | Unassigned | Unassigned | 607_0 | Clustered | 16 | VC_607_0 | 16 | 0.9179 | 1 | 0.9179 | 0 | 0 | 0 | 1 Ref |
| 2406 | KM612261 | Unassigned | Unassigned | Unassigned | 607_0 | Clustered | 16 | VC_607_0 | 16 | 0.9179 | 1 | 0.9179 | 0 | 0 | 0 | 1 Ref |
| 2407 | KM612262 | Unassigned | Unassigned | Unassigned | 607_0 | Clustered | 16 | VC_607_0 | 16 | 0.9179 | 1 | 0.9179 | 0 | 0 | 0 | 1 Ref |
| 2408 | KM612263 | Unassigned | Unassigned | Unassigned | 607_0 | Clustered | 16 | VC_607_0 | 16 | 0.9179 | 1 | 0.9179 | 0 | 0 | 0 | 1 Ref |
| 2409 | KM612264 | Unassigned | Unassigned | Unassigned | 607_0 | Clustered | 16 | VC_607_0 | 16 | 0.9179 | 1 | 0.9179 | 0 | 0 | 0 | 1 Ref |
| 2410 | KM612265 | Unassigned | Unassigned | Unassigned | 607_0 | Clustered | 16 | VC_607_0 | 16 | 0.9179 | 1 | 0.9179 | 0 | 0 | 0 | 1 Ref |
| 2411 | KM652553 | Unassigned | Unassigned | Unassigned | 520_19 | Clustered | 2 | VC_520_19 | 2 | 0.0098 | 0.95761552 | 0.0094 | 0 | 0 | 0 | 1 Ref |
| 2412 | KM652554 | Unassigned | Unassigned | Unassigned | 1071_3 | Clustered | 16 | VC_1071_3 | 16 | 0.2354 | 1 | 0.2354 | 0 | 0 | 0 | 0.9667 Ref |
| 2413 | KM657822 | Unassigned | Unassigned | Unassigned | 329_13 | Clustered/Singleton | 1 | VC_329_13 | 1 | 0 | 1 | 0 | 0 | 0 | 0 | 1 Ref |
| 2414 | KM672662 | Unassigned | Unassigned | Unassigned | 702_0 | Clustered | 5 | VC_702_0 | 5 | 0.7794 | 0.99999988 | 0.7794 | 0 | 0 | 0 | 1 Ref |
| 2415 | KM677185 | Unassigned | Unassigned | Unassigned | 474_0 | Clustered | 14 | VC_474_0 | 14 | 0.8466 | 1 | 0.8466 | 0 | 0 | 0 | 1 Ref |
| 2416 | KM677210 | Unassigned | Unassigned | Unassigned | 216_4 | Clustered | 13 | VC_216_4 | 13 | 0.1657 | 1 | 0.1657 | 0 | 0 | 0 | 1 Ref |
| 2417 | KM677211 | Unassigned | Unassigned | Unassigned | 236_0 | Clustered | 72 | VC_236_0 | 72 | 0.8104 | 1 | 0.8104 | 0 | 0 | 0 | 1 Ref |
| 2418 | KM819694 | Unassigned | Unassigned | Unassigned | 112_0 | Clustered | 8 | VC_112_0 | 8 | 0.2537 | 1 | 0.2537 | 0 | 0 | 0 | 1 Ref |
| 2419 | KM819695 | Unassigned | Unassigned | Unassigned | 452_0 | Clustered | 66 | VC_452_0 | 66 | 0.9248 | 1 | 0.9248 | 0 | 0 | 0 | 1 Ref |
| 2420 | KM819696 | Unassigned | Unassigned | Unassigned | 452_0 | Clustered | 66 | VC_452_0 | 66 | 0.9248 | 1 | 0.9248 | 0 | 0 | 0 | 1 Ref |
| 2421 | KM873719 | Unassigned | Unassigned | Unassigned | 912_0 | Clustered | 76 | VC_912_0 | 76 | 0.979 | 1 | 0.979 | 0 | 0 | 0 | 1 Ref |
| 2422 | KM879463 | Unassigned | Unassigned | Unassigned | 1074_0 | Clustered | 19 | VC_1074_0 | 19 | 0.9198 | 1 | 0.9198 | 0 | 0 | 0 | 1 Ref |
| 2423 | KM881426 | Unassigned | Unassigned | Unassigned | 558_50 | Clustered | 15 | VC_558_50 | 15 | 0.0468 | 1 | 0.0468 | 0 | 0 | 0 | 0.9238 Ref |
| 2424 | KM896878 | Unassigned | Unassigned | Unassigned | 835_0 | Clustered | 67 | VC_835_0 | 67 | 0.9624 | 1 | 0.9624 | 0 | 0 | 0 | 1 Ref |
| 2425 | KM923970 | Unassigned | Unassigned | Unassigned | 761_0 | Clustered | 67 | VC_761_0 | 67 | 0.8947 | 1 | 0.8947 | 0 | 0 | 0 | 1 Ref |
| 2426 | KM923971 | Unassigned | Unassigned | Unassigned | 238_0 | Clustered | 17 | VC_238_0 | 17 | 0.6109 | 1 | 0.6109 | 0 | 0 | 0 | 1 Ref |
| 2427 | KM925136 | Unassigned | Unassigned | Unassigned | 546_3 | Clustered | 4 | VC_546_3 | 4 | 0.0418 | 0.92194988 | 0.0385 | 0 | 0 | 0 | 1 Ref |
| 2428 | KM974184 | Unassigned | Unassigned | Unassigned | 838_0 | Clustered | 29 | VC_838_0 | 29 | 0.8611 | 1 | 0.8611 | 0 | 0 | 0 | 1 Ref |

| | | | | | | | | | | | | | | | | | |
|---|---|---|---|---|---|---|---|---|---|---|---|---|---|---|---|---|---|
| 2429 | KM979354 | Unassigned | Unassigned | Unassigned | 190_22 | Clustered | 3 | VC_190_22 | 3 | 0.0186 | 0.9985967 | 0.0185 | 0 | 0 | 0 | 1 | Ref |
| 2430 | KM979355 | Unassigned | Unassigned | Unassigned | 190_22 | Clustered | 3 | VC_190_22 | 3 | 0.0186 | 0.9985967 | 0.0185 | 0 | 0 | 0 | 1 | Ref |
| 2431 | KM983328 | Unassigned | Unassigned | Unassigned | 649_0 | Clustered | 3 | VC_649_0 | 3 | 0.3028 | 0.99838615 | 0.3023 | 0 | 0 | 0 | 1 | Ref |
| 2432 | KM983330 | Unassigned | Unassigned | Unassigned | 142_0 | Clustered | 2 | VC_142_0 | 2 | 0.4597 | 0.94273135 | 0.4334 | 0 | 0 | 0 | 1 | Ref |
| 2433 | KM983331 | Unassigned | Unassigned | Unassigned | 649_0 | Clustered | 3 | VC_649_0 | 3 | 0.3028 | 0.99838615 | 0.3023 | 0 | 0 | 0 | 1 | Ref |
| 2434 | KM983333 | Unassigned | Unassigned | Unassigned | 142_0 | Clustered | 2 | VC_142_0 | 2 | 0.4597 | 0.94273135 | 0.4334 | 0 | 0 | 0 | 1 | Ref |
| 2435 | KM983334 | Unassigned | Unassigned | Unassigned | 649_0 | Clustered | 3 | VC_649_0 | 3 | 0.3028 | 0.99838615 | 0.3023 | 0 | 0 | 0 | 1 | Ref |
| 2436 | KP007359 | Unassigned | Unassigned | Unassigned | 290_4 | Clustered | 2 | VC_290_4 | 2 | 0.0092 | 0.90346068 | 0.0083 | 0 | 0 | 0 | 1 | Ref |
| 2437 | KP007360 | Unassigned | Unassigned | Unassigned | 290_1 | Clustered | 8 | VC_290_1 | 8 | 0.0757 | 1 | 0.0757 | 0 | 0 | 0 | 0.8571 | Ref |
| 2438 | KP007361 | Unassigned | Unassigned | Unassigned | 290_2 | Clustered | 2 | VC_290_2 | 2 | 0.0102 | 0.92371007 | 0.0094 | 0 | 0 | 0 | 1 | Ref |
| 2439 | KP007362 | Unassigned | Unassigned | Unassigned | 290_3 | Clustered | 2 | VC_290_3 | 2 | 0.0102 | 0.92273843 | 0.0094 | 0 | 0 | 0 | 1 | Ref |
| 2440 | KP010268 | Unassigned | Unassigned | Unassigned | 50_0 | Clustered | 9 | VC_50_0 | 9 | 0.5531 | 1 | 0.5531 | 0 | 0 | 0 | 1 | Ref |
| 2441 | KP010413 | Unassigned | Unassigned | Unassigned | 329_11 | Clustered | 15 | VC_329_11 | 15 | 0.0791 | 1 | 0.0791 | 0 | 0 | 0 | 0.9333 | Ref |
| 2442 | KP017310 | Unassigned | Unassigned | Unassigned | 514_0 | Clustered | 75 | VC_514_0 | 75 | 0.9402 | 1 | 0.9402 | 0 | 0 | 0 | 1 | Ref |
| 2443 | KP017311 | Unassigned | Unassigned | Unassigned | 514_0 | Clustered | 75 | VC_514_0 | 75 | 0.9402 | 1 | 0.9402 | 0 | 0 | 0 | 1 | Ref |
| 2444 | KP025626 | Unassigned | Unassigned | Unassigned | 130_0 | Clustered | 19 | VC_130_0 | 19 | 0.6068 | 1 | 0.6068 | 0 | 0 | 0 | 1 | Ref |
| 2445 | KP027195 | Unassigned | Unassigned | Unassigned | 764_0 | Clustered | 4 | VC_764_0 | 4 | 0.3556 | 0.99997469 | 0.3556 | 0 | 0 | 0 | 1 | Ref |
| 2446 | KP027196 | Unassigned | Unassigned | Unassigned | 325_29 | Clustered | 3 | VC_325_29 | 3 | 0.0115 | 0.99857442 | 0.0115 | 0 | 0 | 0 | 1 | Ref |
| 2447 | KP027197 | Unassigned | Unassigned | Unassigned | 371_18 | Clustered | 10 | VC_371_18 | 10 | 0.0302 | 1 | 0.0302 | 0 | 0 | 0 | 0.8 | Ref |
| 2448 | KP027198 | Unassigned | Unassigned | Unassigned | 517_1 | Clustered | 108 | VC_517_1 | 108 | 0.8483 | 1 | 0.8483 | 0 | 0 | 0 | 0.9955 | Ref |
| 2449 | KP027199 | Unassigned | Unassigned | Unassigned | 237_1 | Clustered | 13 | VC_237_1 | 13 | 0.1773 | 1 | 0.1773 | 0 | 0 | 0 | 0.7308 | Ref |
| 2450 | KP027200 | Unassigned | Unassigned | Unassigned | 519_0 | Clustered | 39 | VC_519_0 | 39 | 0.8156 | 1 | 0.8156 | 0 | 0 | 0 | 1 | Ref |
| 2451 | KP027201 | Unassigned | Unassigned | Unassigned | 518_1 | Clustered | 2 | VC_518_1 | 2 | 0.0376 | 0.95718761 | 0.036 | 0 | 0 | 0 | 1 | Ref |
| 2452 | KP027202 | Unassigned | Unassigned | Unassigned | 514_0 | Clustered | 75 | VC_514_0 | 75 | 0.9402 | 1 | 0.9402 | 0 | 0 | 0 | 1 | Ref |
| 2453 | KP027203 | Unassigned | Unassigned | Unassigned | 325_29 | Clustered | 3 | VC_325_29 | 3 | 0.0115 | 0.99857442 | 0.0115 | 0 | 0 | 0 | 1 | Ref |
| 2454 | KP027204 | Unassigned | Unassigned | Unassigned | 325_8 | Clustered | 2 | VC_325_8 | 2 | 0.0093 | 0.95751268 | 0.0089 | 0 | 0 | 0 | 1 | Ref |
| 2455 | KP027205 | Unassigned | Unassigned | Unassigned | 325_40 | Clustered | 2 | VC_325_40 | 2 | 0.004 | 0.95574341 | 0.0038 | 0 | 0 | 0 | 1 | Ref |
| 2456 | KP027207 | Unassigned | Unassigned | Unassigned | 509_0 | Clustered | 37 | VC_509_0 | 37 | 0.9689 | 1 | 0.9689 | 0 | 0 | 0 | 1 | Ref |
| 2457 | KP027208 | Unassigned | Unassigned | Unassigned | 371_15 | Clustered | 44 | VC_371_15 | 44 | 0.1391 | 1 | 0.1391 | 0 | 0 | 0 | 0.7474 | Ref |
| 2458 | KP027209 | Unassigned | Unassigned | Unassigned | 371_18 | Clustered | 10 | VC_371_18 | 10 | 0.0302 | 1 | 0.0302 | 0 | 0 | 0 | 0.8 | Ref |
| 2459 | KP027446 | Unassigned | Unassigned | Unassigned | 149_20 | Clustered/Singleton | 1 | VC_149_20 | 1 | 0 | 1 | 0 | 0 | 0 | 0 | 1 | Ref |
| 2460 | KP027447 | Unassigned | Unassigned | Unassigned | 150_0 | Clustered | 13 | VC_150_0 | 13 | 0.2417 | 1 | 0.2417 | 0 | 0 | 0 | 0.9872 | Ref |
| 2461 | KP057620 | Unassigned | Unassigned | Unassigned | 517_1 | Clustered | 108 | VC_517_1 | 108 | 0.8483 | 1 | 0.8483 | 0 | 0 | 0 | 0.9955 | Ref |
| 2462 | KP063118 | Unassigned | Unassigned | Unassigned | 591_0 | Clustered | 14 | VC_591_0 | 14 | 0.5184 | 1 | 0.5184 | 0 | 0 | 0 | 1 | Ref |
| 2463 | KP063541 | Unassigned | Unassigned | Unassigned | 229_0 | Clustered | 32 | VC_229_0 | 32 | 0.9119 | 1 | 0.9119 | 0 | 0 | 0 | 1 | Ref |
| 2464 | KP064094 | Unassigned | Unassigned | Unassigned | 795_0 | Clustered | 19 | VC_795_0 | 19 | 0.6848 | 1 | 0.6848 | 0 | 0 | 0 | 1 | Ref |
| 2465 | KP085586 | Unassigned | Unassigned | Unassigned | 575_0 | Clustered | 35 | VC_575_0 | 35 | 0.8815 | 1 | 0.8815 | 0 | 0 | 0 | 1 | Ref |
| 2466 | KP087938 | Unassigned | Unassigned | Unassigned | 1077_0 | Clustered | 48 | VC_1077_0 | 48 | 0.8525 | 1 | 0.8525 | 0 | 0 | 0 | 1 | Ref |
| 2467 | KP087939 | Unassigned | Unassigned | Unassigned | 398_0 | Clustered | 48 | VC_398_0 | 48 | 0.4813 | 1 | 0.4813 | 0 | 0 | 0 | 1 | Ref |
| 2468 | KP087942 | Unassigned | Unassigned | Unassigned | 1077_0 | Clustered | 48 | VC_1077_0 | 48 | 0.8525 | 1 | 0.8525 | 0 | 0 | 0 | 1 | Ref |
| 2469 | KP087943 | Unassigned | Unassigned | Unassigned | 1086_0 | Clustered | 3 | VC_1086_0 | 3 | 0.8416 | 0.81445332 | 0.6855 | 0 | 0 | 0 | 1 | Ref |
| 2470 | KP087945 | Unassigned | Unassigned | Unassigned | 398_0 | Clustered | 48 | VC_398_0 | 48 | 0.4813 | 1 | 0.4813 | 0 | 0 | 0 | 1 | Ref |
| 2471 | KP087948 | Unassigned | Unassigned | Unassigned | 1062_0 | Clustered | 45 | VC_1062_0 | 45 | 0.9734 | 0.50949847 | 0.4959 | 0 | 0 | 0 | 1 | Ref |
| 2472 | KP087951 | Unassigned | Unassigned | Unassigned | 1086_0 | Clustered | 3 | VC_1086_0 | 3 | 0.8416 | 0.81445332 | 0.6855 | 0 | 0 | 0 | 1 | Ref |
| 2473 | KP087953 | Unassigned | Unassigned | Unassigned | 1087_0 | Clustered | 5 | VC_1087_0 | 5 | 0.8109 | 0.98759579 | 0.8008 | 0 | 0 | 0 | 1 | Ref |
| 2474 | KP087954 | Unassigned | Unassigned | Unassigned | 992_0 | Clustered | 8 | VC_992_0 | 8 | 0.3873 | 1 | 0.3873 | 0 | 0 | 0 | 1 | Ref |
| 2475 | KP087955 | Unassigned | Unassigned | Unassigned | 1088_0 | Clustered | 3 | VC_1088_0 | 3 | 0.7739 | 0.82711071 | 0.6401 | 0 | 0 | 0 | 1 | Ref |
| 2476 | KP087957 | Unassigned | Unassigned | Unassigned | 1086_0 | Clustered | 3 | VC_1086_0 | 3 | 0.8416 | 0.81445332 | 0.6855 | 0 | 0 | 0 | 1 | Ref |
| 2477 | KP090453 | Unassigned | Unassigned | Unassigned | 386_0 | Clustered | 49 | VC_386_0 | 49 | 0.7941 | 1 | 0.7941 | 0 | 0 | 0 | 1 | Ref |
| 2478 | KP090454 | Unassigned | Unassigned | Unassigned | 386_0 | Clustered | 49 | VC_386_0 | 49 | 0.7941 | 1 | 0.7941 | 0 | 0 | 0 | 1 | Ref |
| 2479 | KP137432 | Unassigned | Unassigned | Unassigned | 4_0 | Clustered | 7 | VC_4_0 | 7 | 0.7912 | 1 | 0.7912 | 0 | 0 | 0 | 1 | Ref |
| 2480 | KP137433 | Unassigned | Unassigned | Unassigned | 718_0 | Clustered | 4 | VC_718_0 | 4 | 0.6393 | 0.99996676 | 0.6393 | 0 | 0 | 0 | 0.8333 | Ref |
| 2481 | KP137434 | Unassigned | Unassigned | Unassigned | 4_0 | Clustered | 7 | VC_4_0 | 7 | 0.7912 | 1 | 0.7912 | 0 | 0 | 0 | 1 | Ref |
| 2482 | KP137435 | Unassigned | Unassigned | Unassigned | 718_0 | Clustered | 4 | VC_718_0 | 4 | 0.6393 | 0.99996676 | 0.6393 | 0 | 0 | 0 | 0.8333 | Ref |
| 2483 | KP137436 | Unassigned | Unassigned | Unassigned | 4_0 | Clustered | 7 | VC_4_0 | 7 | 0.7912 | 1 | 0.7912 | 0 | 0 | 0 | 1 | Ref |
| 2484 | KP137437 | Unassigned | Unassigned | Unassigned | 718_0 | Clustered | 4 | VC_718_0 | 4 | 0.6393 | 0.99996676 | 0.6393 | 0 | 0 | 0 | 0.8333 | Ref |
| 2485 | KP137438 | Unassigned | Unassigned | Unassigned | 4_0 | Clustered | 7 | VC_4_0 | 7 | 0.7912 | 1 | 0.7912 | 0 | 0 | 0 | 1 | Ref |
| 2486 | KP137439 | Unassigned | Unassigned | Unassigned | 718_0 | Clustered | 4 | VC_718_0 | 4 | 0.6393 | 0.99996676 | 0.6393 | 0 | 0 | 0 | 0.8333 | Ref |
| 2487 | KP137440 | Unassigned | Unassigned | Unassigned | 181_0 | Clustered | 5 | VC_181_0 | 5 | 0.1168 | 0.99999995 | 0.1168 | 0 | 0 | 0 | 1 | Ref |
| 2488 | KP143762 | Unassigned | Unassigned | Unassigned | 329_1 | Clustered | 17 | VC_329_1 | 17 | 0.0867 | 1 | 0.0867 | 0 | 0 | 0 | 0.8235 | Ref |
| 2489 | KP143763 | Unassigned | Unassigned | Unassigned | 190_0 | Clustered | 2 | VC_190_0 | 2 | 0.0087 | 0.95763473 | 0.0083 | 0 | 0 | 0 | 1 | Ref |
| 2490 | KP144388 | Unassigned | Unassigned | Unassigned | 211_1 | Clustered/Singleton | 1 | VC_211_1 | 1 | 0 | 1 | 0 | 0 | 0 | 0 | 1 | Ref |
| 2491 | KP202158 | Unassigned | Unassigned | Unassigned | 490_4 | Clustered/Singleton | 1 | VC_490_4 | 1 | 0 | 1 | 0 | 0 | 0 | 0 | 1 | Ref |
| 2492 | KP202969 | Unassigned | Unassigned | Unassigned | 171_0 | Clustered | 8 | VC_171_0 | 8 | 0.5884 | 1 | 0.5884 | 0 | 0 | 0 | 1 | Ref |
| 2493 | KP202970 | Unassigned | Unassigned | Unassigned | 171_0 | Clustered | 8 | VC_171_0 | 8 | 0.5884 | 1 | 0.5884 | 0 | 0 | 0 | 1 | Ref |
| 2494 | KP202971 | Unassigned | Unassigned | Unassigned | 1090_0 | Clustered | 4 | VC_1090_0 | 4 | 1 | 1 | 1 | 0 | 0 | 0 | 1 | Ref |
| 2495 | KP202972 | Unassigned | Unassigned | Unassigned | 708_0 | Clustered | 47 | VC_708_0 | 47 | 0.8654 | 1 | 0.8654 | 0 | 0 | 0 | 1 | Ref |
| 2496 | KP208803 | Unassigned | Unassigned | Unassigned | 317_3 | Clustered/Singleton | 1 | VC_317_3 | 1 | 0 | 1 | 0 | 0 | 0 | 0 | 1 | Ref |
| 2497 | KP209285 | Unassigned | Unassigned | Unassigned | 12_0 | Clustered | 21 | VC_12_0 | 21 | 0.2274 | 1 | 0.2274 | 0 | 0 | 0 | 0.9571 | Ref |
| 2498 | KP233880 | Unassigned | Unassigned | Unassigned | 472_0 | Clustered | 47 | VC_472_0 | 47 | 0.9025 | 1 | 0.9025 | 0 | 0 | 0 | 1 | Ref |
| 2499 | KP238129 | Unassigned | Unassigned | Unassigned | 446_0 | Clustered | 4 | VC_446_0 | 4 | 0.754 | 0.99987768 | 0.7539 | 0 | 0 | 0 | 1 | Ref |
| 2500 | KP273225 | Unassigned | Unassigned | Unassigned | 219_0 | Clustered | 5 | VC_219_0 | 5 | 0.08 | 0.99999998 | 0.08 | 0 | 0 | 0 | 1 | Ref |
| 2501 | KP280062 | Unassigned | Unassigned | Unassigned | 784_0 | Clustered | 6 | VC_784_0 | 6 | 0.5517 | 1 | 0.5517 | 0 | 0 | 0 | 1 | Ref |
| 2502 | KP280063 | Unassigned | Unassigned | Unassigned | 971_0 | Clustered | 3 | VC_971_0 | 3 | 0.0548 | 0.99838914 | 0.0547 | 0 | 0 | 0 | 1 | Ref |
| 2503 | KP282673 | Unassigned | Unassigned | Unassigned | 791_0 | Clustered | 3 | VC_791_0 | 3 | 1 | 1 | 1 | 0 | 0 | 0 | 1 | Ref |
| 2504 | KP282674 | Unassigned | Unassigned | Unassigned | 791_0 | Clustered | 3 | VC_791_0 | 3 | 1 | 1 | 1 | 0 | 0 | 0 | 1 | Ref |
| 2505 | KP282675 | Unassigned | Unassigned | Unassigned | 348_0 | Clustered | 22 | VC_348_0 | 22 | 0.9602 | 1 | 0.9602 | 0 | 0 | 0 | 1 | Ref |
| 2506 | KP282677 | Unassigned | Unassigned | Unassigned | 446_0 | Clustered | 4 | VC_446_0 | 4 | 0.754 | 0.99987768 | 0.7539 | 0 | 0 | 0 | 1 | Ref |
| 2507 | KP282678 | Unassigned | Unassigned | Unassigned | 446_0 | Clustered | 4 | VC_446_0 | 4 | 0.754 | 0.99987768 | 0.7539 | 0 | 0 | 0 | 1 | Ref |
| 2508 | KP282679 | Unassigned | Unassigned | Unassigned | 881_0 | Clustered | 2 | VC_881_0 | 2 | 0.6781 | 0.91813282 | 0.6226 | 0 | 0 | 0 | 1 | Ref |
| 2509 | KP282680 | Unassigned | Unassigned | Unassigned | 881_0 | Clustered | 2 | VC_881_0 | 2 | 0.6781 | 0.91813282 | 0.6226 | 0 | 0 | 0 | 1 | Ref |
| 2510 | KP296791 | Unassigned | Unassigned | Unassigned | 708_0 | Clustered | 47 | VC_708_0 | 47 | 0.8654 | 1 | 0.8654 | 0 | 0 | 0 | 1 | Ref |
| 2511 | KP296793 | Unassigned | Unassigned | Unassigned | 708_0 | Clustered | 47 | VC_708_0 | 47 | 0.8654 | 1 | 0.8654 | 0 | 0 | 0 | 1 | Ref |
| 2512 | KP296794 | Unassigned | Unassigned | Unassigned | 708_0 | Clustered | 47 | VC_708_0 | 47 | 0.8654 | 1 | 0.8654 | 0 | 0 | 0 | 1 | Ref |
| 2513 | KP296795 | Unassigned | Unassigned | Unassigned | 708_0 | Clustered | 47 | VC_708_0 | 47 | 0.8654 | 1 | 0.8654 | 0 | 0 | 0 | 1 | Ref |
| 2514 | KP296796 | Unassigned | Unassigned | Unassigned | 708_0 | Clustered | 47 | VC_708_0 | 47 | 0.8654 | 1 | 0.8654 | 0 | 0 | 0 | 1 | Ref |
| 2515 | KP308307 | Unassigned | Unassigned | Unassigned | 813_1 | Clustered | 24 | VC_813_1 | 24 | 0.535 | 1 | 0.535 | 0 | 0 | 0 | 1 | Ref |
| 2516 | KP313531 | Unassigned | Unassigned | Unassigned | 385_0 | Clustered | 51 | VC_385_0 | 51 | 0.8754 | 1 | 0.8754 | 0 | 0 | 0 | 1 | Ref |
| 2517 | KP313532 | Unassigned | Unassigned | Unassigned | 751_0 | Clustered | 4 | VC_751_0 | 4 | 0.1763 | 0.99998526 | 0.1763 | 0 | 0 | 0 | 1 | Ref |
| 2518 | KP339049 | Unassigned | Unassigned | Unassigned | 92_0 | Clustered | 22 | VC_92_0 | 22 | 0.5639 | 1 | 0.5639 | 0 | 0 | 0 | 1 | Ref |
| 2519 | KP340287 | Unassigned | Unassigned | Unassigned | 79_0 | Clustered | 64 | VC_79_0 | 64 | 0.4185 | 1 | 0.4185 | 0 | 0 | 0 | 0.7649 | Ref |
| 2520 | KP340288 | Unassigned | Unassigned | Unassigned | 79_0 | Clustered | 64 | VC_79_0 | 64 | 0.4185 | 1 | 0.4185 | 0 | 0 | 0 | 0.7649 | Ref |
| 2521 | KP343639 | Unassigned | Unassigned | Unassigned | 83_0 | Clustered | 6 | VC_83_0 | 6 | 0.6436 | 1 | 0.6436 | 0 | 0 | 0 | 1 | Ref |
| 2522 | KP399677 | Unassigned | Unassigned | Unassigned | 380_0 | Clustered | 3 | VC_380_0 | 3 | 0.1631 | 0.99675097 | 0.1625 | 0 | 0 | 0 | 1 | Ref |
| 2523 | KP399678 | Unassigned | Unassigned | Unassigned | 378_0 | Clustered | 9 | VC_378_0 | 9 | 0.7603 | 1 | 0.7603 | 0 | 0 | 0 | 1 | Ref |
| 2524 | KP411017 | Unassigned | Unassigned | Unassigned | 1032_0 | Clustered | 7 | VC_1032_0 | 7 | 0.9887 | 0.99999504 | 0.9887 | 0 | 0 | 0 | 1 | Ref |
| 2525 | KP658157 | Unassigned | Unassigned | Unassigned | 938_0 | Clustered | 99 | VC_938_0 | 99 | 0.9119 | 1 | 0.9119 | 0 | 0 | 0 | 0.9928 | Ref |
| 2526 | KP671755 | Unassigned | Unassigned | Unassigned | 531_0 | Clustered | 9 | VC_531_0 | 9 | 0.26 | 1 | 0.26 | 0 | 0 | 0 | 1 | Ref |
| 2527 | KP682371 | Unassigned | Unassigned | Unassigned | 42_0 | Clustered | 49 | VC_42_0 | 49 | 0.4517 | 1 | 0.4517 | 0 | 0 | 0 | 0.8597 | Ref |
| 2528 | KP682372 | Unassigned | Unassigned | Unassigned | 39_0 | Clustered | 35 | VC_39_0 | 35 | 0.4337 | 1 | 0.4337 | 0 | 0 | 0 | 1 | Ref |
| 2529 | KP682373 | Unassigned | Unassigned | Unassigned | 39_0 | Clustered | 35 | VC_39_0 | 35 | 0.4337 | 1 | 0.4337 | 0 | 0 | 0 | 1 | Ref |
| 2530 | KP682374 | Unassigned | Unassigned | Unassigned | 42_0 | Clustered | 49 | VC_42_0 | 49 | 0.4517 | 1 | 0.4517 | 0 | 0 | 0 | 0.8597 | Ref |
| 2531 | KP682375 | Unassigned | Unassigned | Unassigned | 39_0 | Clustered | 35 | VC_39_0 | 35 | 0.4337 | 1 | 0.4337 | 0 | 0 | 0 | 1 | Ref |
| 2532 | KP682376 | Unassigned | Unassigned | Unassigned | 39_0 | Clustered | 35 | VC_39_0 | 35 | 0.4337 | 1 | 0.4337 | 0 | 0 | 0 | 1 | Ref |
| 2533 | KP682377 | Unassigned | Unassigned | Unassigned | 39_0 | Clustered | 35 | VC_39_0 | 35 | 0.4337 | 1 | 0.4337 | 0 | 0 | 0 | 1 | Ref |
| 2534 | KP682378 | Unassigned | Unassigned | Unassigned | 39_0 | Clustered | 35 | VC_39_0 | 35 | 0.4337 | 1 | 0.4337 | 0 | 0 | 0 | 1 | Ref |
| 2535 | KP682379 | Unassigned | Unassigned | Unassigned | 39_0 | Clustered | 35 | VC_39_0 | 35 | 0.4337 | 1 | 0.4337 | 0 | 0 | 0 | 1 | Ref |
| 2536 | KP682380 | Unassigned | Unassigned | Unassigned | 39_0 | Clustered | 35 | VC_39_0 | 35 | 0.4337 | 1 | 0.4337 | 0 | 0 | 0 | 1 | Ref |
| 2537 | KP682382 | Unassigned | Unassigned | Unassigned | 39_0 | Clustered | 35 | VC_39_0 | 35 | 0.4337 | 1 | 0.4337 | 0 | 0 | 0 | 1 | Ref |
| 2538 | KP682383 | Unassigned | Unassigned | Unassigned | 39_0 | Clustered | 35 | VC_39_0 | 35 | 0.4337 | 1 | 0.4337 | 0 | 0 | 0 | 1 | Ref |
| 2539 | KP682384 | Unassigned | Unassigned | Unassigned | 39_0 | Clustered | 35 | VC_39_0 | 35 | 0.4337 | 1 | 0.4337 | 0 | 0 | 0 | 1 | Ref |
| 2540 | KP682385 | Unassigned | Unassigned | Unassigned | 39_0 | Clustered | 35 | VC_39_0 | 35 | 0.4337 | 1 | 0.4337 | 0 | 0 | 0 | 1 | Ref |
| 2541 | KP682386 | Unassigned | Unassigned | Unassigned | 39_0 | Clustered | 35 | VC_39_0 | 35 | 0.4337 | 1 | 0.4337 | 0 | 0 | 0 | 1 | Ref |
| 2542 | KP682387 | Unassigned | Unassigned | Unassigned | 39_0 | Clustered | 35 | VC_39_0 | 35 | 0.4337 | 1 | 0.4337 | 0 | 0 | 0 | 1 | Ref |
| 2543 | KP682388 | Unassigned | Unassigned | Unassigned | 39_0 | Clustered | 35 | VC_39_0 | 35 | 0.4337 | 1 | 0.4337 | 0 | 0 | 0 | 1 | Ref |
| 2544 | KP682389 | Unassigned | Unassigned | Unassigned | 39_0 | Clustered | 35 | VC_39_0 | 35 | 0.4337 | 1 | 0.4337 | 0 | 0 | 0 | 1 | Ref |
| 2545 | KP682390 | Unassigned | Unassigned | Unassigned | 39_0 | Clustered | 35 | VC_39_0 | 35 | 0.4337 | 1 | 0.4337 | 0 | 0 | 0 | 1 | Ref |
| 2546 | KP682391 | Unassigned | Unassigned | Unassigned | 39_0 | Clustered | 35 | VC_39_0 | 35 | 0.4337 | 1 | 0.4337 | 0 | 0 | 0 | 1 | Ref |
| 2547 | KP682392 | Unassigned | Unassigned | Unassigned | 39_0 | Clustered | 35 | VC_39_0 | 35 | 0.4337 | 1 | 0.4337 | 0 | 0 | 0 | 1 | Ref |
| 2548 | KP687431 | Unassigned | Unassigned | Unassigned | 149_8 | Clustered | 66 | VC_149_8 | 66 | 0.4565 | 1 | 0.4565 | 0 | 0 | 0 | 0.8182 | Ref |
| 2549 | KP687432 | Unassigned | Unassigned | Unassigned | 149_8 | Clustered | 66 | VC_149_8 | 66 | 0.4565 | 1 | 0.4565 | 0 | 0 | 0 | 0.8182 | Ref |
| 2550 | KP708985 | Unassigned | Unassigned | Unassigned | 111_0 | Clustered | 91 | VC_111_0 | 91 | 0.9587 | 1 | 0.9587 | 0 | 0 | 0 | 1 | Ref |
| 2551 | KP708986 | Unassigned | Unassigned | Unassigned | 111_0 | Clustered | 91 | VC_111_0 | 91 | 0.9587 | 1 | 0.9587 | 0 | 0 | 0 | 1 | Ref |
| 2552 | KP719132 | Unassigned | Unassigned | Unassigned | 722_0 | Clustered | 13 | VC_722_0 | 13 | 0.5104 | 1 | 0.5104 | 0 | 0 | 0 | 1 | Ref |
| 2553 | KP719133 | Unassigned | Unassigned | Unassigned | 722_0 | Clustered | 13 | VC_722_0 | 13 | 0.5104 | 1 | 0.5104 | 0 | 0 | 0 | 1 | Ref |
| 2554 | KP719134 | Unassigned | Unassigned | Unassigned | 722_0 | Clustered | 13 | VC_722_0 | 13 | 0.5104 | 1 | 0.5104 | 0 | 0 | 0 | 1 | Ref |
| 2555 | KP790008 | Unassigned | Unassigned | Unassigned | 867_0 | Clustered | 6 | VC_867_0 | 6 | 0.3202 | 1 | 0.3202 | 0 | 0 | 0 | 1 | Ref |
| 2556 | KP790009 | Unassigned | Unassigned | Unassigned | 867_0 | Clustered | 6 | VC_867_0 | 6 | 0.3202 | 1 | 0.3202 | 0 | 0 | 0 | 1 | Ref |
| 2557 | KP790010 | Unassigned | Unassigned | Unassigned | 867_0 | Clustered | 6 | VC_867_0 | 6 | 0.3202 | 1 | 0.3202 | 0 | 0 | 0 | 1 | Ref |
| 2558 | KP790011 | Unassigned | Unassigned | Unassigned | 406_0 | Clustered | 11 | VC_406_0 | 11 | 0.9449 | 1 | 0.9449 | 0 | 0 | 0 | 1 | Ref |
| 2559 | KP791805 | Unassigned | Unassigned | Unassigned | 385_0 | Clustered | 51 | VC_385_0 | 51 | 0.8754 | 1 | 0.8754 | 0 | 0 | 0 | 1 | Ref |
| 2560 | KP791806 | Unassigned | Unassigned | Unassigned | 385_0 | Clustered | 51 | VC_385_0 | 51 | 0.8754 | 1 | 0.8754 | 0 | 0 | 0 | 1 | Ref |
| 2561 | KP791807 | Unassigned | Unassigned | Unassigned | 385_0 | Clustered | 51 | VC_385_0 | 51 | 0.8754 | 1 | 0.8754 | 0 | 0 | 0 | 1 | Ref |
| 2562 | KP792623 | Unassigned | Unassigned | Unassigned | 806_0 | Clustered | 3 | VC_806_0 | 3 | 0.532 | 0.99824026 | 0.5311 | 0 | 0 | 0 | 1 | Ref |
| 2563 | KP793104 | Unassigned | Unassigned | Unassigned | 202_0 | Clustered | 102 | VC_202_0 | 102 | 0.4049 | 1 | 0.4049 | 0 | 0 | 0 | 0.7876 | Ref |
| 2564 | KP793105 | Unassigned | Unassigned | Unassigned | 202_0 | Clustered | 102 | VC_202_0 | 102 | 0.4049 | 1 | 0.4049 | 0 | 0 | 0 | 0.7876 | Ref |
| 2565 | KP793108 | Unassigned | Unassigned | Unassigned | 202_0 | Clustered | 102 | VC_202_0 | 102 | 0.4049 | 1 | 0.4049 | 0 | 0 | 0 | 0.7876 | Ref |
| 2566 | KP793109 | Unassigned | Unassigned | Unassigned | 202_0 | Clustered | 102 | VC_202_0 | 102 | 0.4049 | 1 | 0.4049 | 0 | 0 | 0 | 0.7876 | Ref |
| 2567 | KP793111 | Unassigned | Unassigned | Unassigned | 202_0 | Clustered | 102 | VC_202_0 | 102 | 0.4049 | 1 | 0.4049 | 0 | 0 | 0 | 0.7876 | Ref |
| 2568 | KP793113 | Unassigned | Unassigned | Unassigned | 202_0 | Clustered | 102 | VC_202_0 | 102 | 0.4049 | 1 | 0.4049 | 0 | 0 | 0 | 0.7876 | Ref |
| 2569 | KP793115 | Unassigned | Unassigned | Unassigned | 202_0 | Clustered | 102 | VC_202_0 | 102 | 0.4049 | 1 | 0.4049 | 0 | 0 | 0 | 0.7876 | Ref |
| 2570 | KP793116 | Unassigned | Unassigned | Unassigned | 202_0 | Clustered | 102 | VC_202_0 | 102 | 0.4049 | 1 | 0.4049 | 0 | 0 | 0 | 0.7876 | Ref |
| 2571 | KP793117 | Unassigned | Unassigned | Unassigned | 202_0 | Clustered | 102 | VC_202_0 | 102 | 0.4049 | 1 | 0.4049 | 0 | 0 | 0 | 0.7876 | Ref |
| 2572 | KP793118 | Unassigned | Unassigned | Unassigned | 202_0 | Clustered | 102 | VC_202_0 | 102 | 0.4049 | 1 | 0.4049 | 0 | 0 | 0 | 0.7876 | Ref |
| 2573 | KP793123 | Unassigned | Unassigned | Unassigned | 202_0 | Clustered | 102 | VC_202_0 | 102 | 0.4049 | 1 | 0.4049 | 0 | 0 | 0 | 0.7876 | Ref |
| 2574 | KP793124 | Unassigned | Unassigned | Unassigned | 202_0 | Clustered | 102 | VC_202_0 | 102 | 0.4049 | 1 | 0.4049 | 0 | 0 | 0 | 0.7876 | Ref |
| 2575 | KP793129 | Unassigned | Unassigned | Unassigned | 202_0 | Clustered | 102 | VC_202_0 | 102 | 0.4049 | 1 | 0.4049 | 0 | 0 | 0 | 0.7876 | Ref |
| 2576 | KP793134 | Unassigned | Unassigned | Unassigned | 202_0 | Clustered | 102 | VC_202_0 | 102 | 0.4049 | 1 | 0.4049 | 0 | 0 | 0 | 0.7876 | Ref |
| 2577 | KP797973 | Unassigned | Unassigned | Unassigned | 133_9 | Clustered | 3 | VC_133_9 | 3 | 0.0099 | 0.98457109 | 0.0098 | 0 | 0 | 0 | 1 | Ref |
| 2578 | KP836355 | Unassigned | Unassigned | Unassigned | 1091_0 | Clustered | 2 | VC_1091_0 | 2 | 1 | 1 | 1 | 0 | 0 | 0 | 1 | Ref |
| 2579 | KP836356 | Unassigned | Unassigned | Unassigned | 1091_0 | Clustered | 2 | VC_1091_0 | 2 | 1 | 1 | 1 | 0 | 0 | 0 | 1 | Ref |
| 2580 | KP861229 | Unassigned | Unassigned | Unassigned | 698_0 | Clustered | 26 | VC_698_0 | 26 | 0.9366 | 1 | 0.9366 | 0 | 0 | 0 | 1 | Ref |
| 2581 | KP861230 | Unassigned | Unassigned | Unassigned | 696_1 | Clustered | 8 | VC_696_1 | 8 | 0.5452 | 1 | 0.5452 | 0 | 0 | 0 | 0.9286 | Ref |
| 2582 | KP861231 | Unassigned | Unassigned | Unassigned | 698_0 | Clustered | 26 | VC_698_0 | 26 | 0.9366 | 1 | 0.9366 | 0 | 0 | 0 | 1 | Ref |
| 2583 | KP869100 | Unassigned | Unassigned | Unassigned | 329_7 | Clustered | 18 | VC_329_7 | 18 | 0.0938 | 1 | 0.0938 | 0 | 0 | 0 | 0.7778 | Ref |
| 2584 | KP869101 | Unassigned | Unassigned | Unassigned | 286_17 | Clustered | 8 | VC_286_17 | 8 | 0.0535 | 0.99997829 | 0.0535 | 0 | 0 | 0 | 0.6071 | Ref |
| 2585 | KP869102 | Unassigned | Unassigned | Unassigned | 772_15 | Clustered | 5 | VC_772_15 | 5 | 0.0243 | 0.76144861 | 0.0185 | 0 | 0 | 0 | 0.9 | Ref |
| 2586 | KP869103 | Unassigned | Unassigned | Unassigned | 772_15 | Clustered | 5 | VC_772_15 | 5 | 0.0243 | 0.76144861 | 0.0185 | 0 | 0 | 0 | 0.9 | Ref |
| 2587 | KP869104 | Unassigned | Unassigned | Unassigned | 286_17 | Clustered | 8 | VC_286_17 | 8 | 0.0535 | 0.99997829 | 0.0535 | 0 | 0 | 0 | 0.6071 | Ref |
| 2588 | KP869105 | Unassigned | Unassigned | Unassigned | 283_2 | Clustered | 2 | VC_283_2 | 2 | 0.0027 | 0.93842619 | 0.0025 | 0 | 0 | 0 | 1 | Ref |
| 2589 | KP869106 | Unassigned | Unassigned | Unassigned | 329_7 | Clustered | 18 | VC_329_7 | 18 | 0.0938 | 1 | 0.0938 | 0 | 0 | 0 | 0.7778 | Ref |
| 2590 | KP869107 | Unassigned | Unassigned | Unassigned | 476_0 | Clustered | 19 | VC_476_0 | 19 | 0.7814 | 1 | 0.7814 | 0 | 0 | 0 | 1 | Ref |

| | | | | | | | | | | | | | | | | | |
|---|---|---|---|---|---|---|---|---|---|---|---|---|---|---|---|---|---|
| 2591 | KP869108 | Unassigned | Unassigned | Unassigned | 476_0 | Clustered | 19 | VC_476_0 | 19 | 0.7814 | 1 | 0.7814 | 0 | 0 | 0 | 1 | Ref |
| 2592 | KP869109 | Unassigned | Unassigned | Unassigned | 329_7 | Clustered | 18 | VC_329_7 | 18 | 0.0938 | 1 | 0.0938 | 0 | 0 | 0 | 0.7778 | Ref |
| 2593 | KP869110 | Unassigned | Unassigned | Unassigned | 329_7 | Clustered | 18 | VC_329_7 | 18 | 0.0938 | 1 | 0.0938 | 0 | 0 | 0 | 0.7778 | Ref |
| 2594 | KP869111 | Unassigned | Unassigned | Unassigned | 286_17 | Clustered | 8 | VC_286_17 | 8 | 0.0535 | 0.99997829 | 0.0535 | 0 | 0 | 0 | 0.6071 | Ref |
| 2595 | KP869112 | Unassigned | Unassigned | Unassigned | 772_15 | Clustered | 5 | VC_772_15 | 5 | 0.0243 | 0.76144861 | 0.0185 | 0 | 0 | 0 | 0.9 | Ref |
| 2596 | KP869113 | Unassigned | Unassigned | Unassigned | 329_7 | Clustered | 18 | VC_329_7 | 18 | 0.0938 | 1 | 0.0938 | 0 | 0 | 0 | 0.7778 | Ref |
| 2597 | KP876465 | Unassigned | Unassigned | Unassigned | 975_0 | Clustered | 11 | VC_975_0 | 11 | 0.9687 | 1 | 0.9687 | 0 | 0 | 0 | 1 | Ref |
| 2598 | KP876466 | Unassigned | Unassigned | Unassigned | 975_0 | Clustered | 11 | VC_975_0 | 11 | 0.9687 | 1 | 0.9687 | 0 | 0 | 0 | 1 | Ref |
| 2599 | KP881232 | Unassigned | Unassigned | Unassigned | 1052_1 | Clustered/Singleton | 1 | VC_1052_1 | 1 | 0 | 1 | 0 | 0 | 0 | 0 | 1 | Ref |
| 2600 | KP881332 | Unassigned | Unassigned | Unassigned | 153_0 | Clustered | 25 | VC_153_0 | 25 | 0.7729 | 1 | 0.7729 | 0 | 0 | 0 | 1 | Ref |
| 2601 | KP890822 | Unassigned | Unassigned | Unassigned | 452_0 | Clustered | 66 | VC_452_0 | 66 | 0.9248 | 1 | 0.9248 | 0 | 0 | 0 | 1 | Ref |
| 2602 | KP890823 | Unassigned | Unassigned | Unassigned | 288_3 | Clustered | 3 | VC_288_3 | 3 | 0.0452 | 0.99844025 | 0.0452 | 0 | 0 | 0 | 1 | Ref |
| 2603 | KP893289 | Unassigned | Unassigned | Unassigned | 12_0 | Clustered | 21 | VC_12_0 | 21 | 0.2274 | 1 | 0.2274 | 0 | 0 | 0 | 0.9571 | Ref |
| 2604 | KP893290 | Unassigned | Unassigned | Unassigned | 12_2 | Clustered | 19 | VC_12_2 | 19 | 0.2018 | 1 | 0.2018 | 0 | 0 | 0 | 1 | Ref |
| 2605 | KP966108 | Unassigned | Unassigned | Unassigned | 1_0 | Clustered | 54 | VC_1_0 | 54 | 0.8527 | 1 | 0.8527 | 0 | 0 | 0 | 1 | Ref |
| 2606 | KP972568 | Unassigned | Unassigned | Unassigned | 875_0 | Clustered | 10 | VC_875_0 | 10 | 0.988 | 0.99948433 | 0.9875 | 0 | 0 | 0 | 1 | Ref |
| 2607 | KP972569 | Unassigned | Unassigned | Unassigned | 875_0 | Clustered | 10 | VC_875_0 | 10 | 0.988 | 0.99948433 | 0.9875 | 0 | 0 | 0 | 1 | Ref |
| 2608 | KP994390 | Unassigned | Unassigned | Unassigned | 838_0 | Clustered | 29 | VC_838_0 | 29 | 0.8611 | 1 | 0.8611 | 0 | 0 | 0 | 1 | Ref |
| 2609 | KP994596 | Unassigned | Unassigned | Unassigned | 644_0 | Clustered | 5 | VC_644_0 | 5 | 0.6649 | 0.99999975 | 0.6649 | 0 | 0 | 0 | 1 | Ref |
| 2610 | KR011062 | Unassigned | Unassigned | Unassigned | 866_2 | Clustered | 3 | VC_866_2 | 3 | 0.046 | 0.99850411 | 0.046 | 0 | 0 | 0 | 1 | Ref |
| 2611 | KR011063 | Unassigned | Unassigned | Unassigned | 866_2 | Clustered | 3 | VC_866_2 | 3 | 0.046 | 0.99850411 | 0.046 | 0 | 0 | 0 | 1 | Ref |
| 2612 | KR011064 | Unassigned | Unassigned | Unassigned | 866_2 | Clustered | 3 | VC_866_2 | 3 | 0.046 | 0.99850411 | 0.046 | 0 | 0 | 0 | 1 | Ref |
| 2613 | KR014248 | Unassigned | Unassigned | Unassigned | 329_4 | Clustered | 2 | VC_329_4 | 2 | 0.0052 | 0.95746454 | 0.005 | 0 | 0 | 0 | 1 | Ref |
| 2614 | KR029086 | Unassigned | Unassigned | Unassigned | 371_13 | Clustered | 33 | VC_371_13 | 33 | 0.179 | 1 | 0.179 | 0 | 0 | 0 | 0.8769 | Ref |
| 2615 | KR029087 | Unassigned | Unassigned | Unassigned | 371_13 | Clustered | 33 | VC_371_13 | 33 | 0.179 | 1 | 0.179 | 0 | 0 | 0 | 0.8769 | Ref |
| 2616 | KR049063 | Unassigned | Unassigned | Unassigned | 91_0 | Clustered | 22 | VC_91_0 | 22 | 0.5318 | 1 | 0.5318 | 0 | 0 | 0 | 1 | Ref |
| 2617 | KR052142 | Unassigned | Unassigned | Unassigned | 838_0 | Clustered | 29 | VC_838_0 | 29 | 0.8611 | 1 | 0.8611 | 0 | 0 | 0 | 1 | Ref |
| 2618 | KR052143 | Unassigned | Unassigned | Unassigned | 76_0 | Clustered | 37 | VC_76_0 | 37 | 0.9403 | 1 | 0.9403 | 0 | 0 | 0 | 1 | Ref |
| 2619 | KR052480 | Unassigned | Unassigned | Unassigned | 437_0 | Clustered | 4 | VC_437_0 | 4 | 0.552 | 0.99998233 | 0.552 | 0 | 0 | 0 | 1 | Ref |
| 2620 | KR052481 | Unassigned | Unassigned | Unassigned | 437_0 | Clustered | 4 | VC_437_0 | 4 | 0.552 | 0.99998233 | 0.552 | 0 | 0 | 0 | 1 | Ref |
| 2621 | KR052482 | Unassigned | Unassigned | Unassigned | 437_0 | Clustered | 4 | VC_437_0 | 4 | 0.552 | 0.99998233 | 0.552 | 0 | 0 | 0 | 1 | Ref |
| 2622 | KR053195 | Unassigned | Unassigned | Unassigned | 726_0 | Clustered | 3 | VC_726_0 | 3 | 0.0381 | 0.88955608 | 0.0339 | 0 | 0 | 0 | 1 | Ref |
| 2623 | KR053197 | Unassigned | Unassigned | Unassigned | 949_0 | Clustered | 10 | VC_949_0 | 10 | 0.5977 | 1 | 0.5977 | 0 | 0 | 0 | 1 | Ref |
| 2624 | KR053198 | Unassigned | Unassigned | Unassigned | 949_0 | Clustered | 10 | VC_949_0 | 10 | 0.5977 | 1 | 0.5977 | 0 | 0 | 0 | 1 | Ref |
| 2625 | KR053201 | Unassigned | Unassigned | Unassigned | 767_0 | Clustered | 34 | VC_767_0 | 34 | 0.9173 | 1 | 0.9173 | 0 | 0 | 0 | 1 | Ref |
| 2626 | KR054028 | Unassigned | Unassigned | Unassigned | 80_0 | Clustered | 41 | VC_80_0 | 41 | 0.5837 | 1 | 0.5837 | 0 | 0 | 0 | 0.9805 | Ref |
| 2627 | KR054029 | Unassigned | Unassigned | Unassigned | 472_0 | Clustered | 47 | VC_472_0 | 47 | 0.9025 | 1 | 0.9025 | 0 | 0 | 0 | 1 | Ref |
| 2628 | KR054030 | Unassigned | Unassigned | Unassigned | 80_0 | Clustered | 41 | VC_80_0 | 41 | 0.5837 | 1 | 0.5837 | 0 | 0 | 0 | 0.9805 | Ref |
| 2629 | KR054031 | Unassigned | Unassigned | Unassigned | 423_0 | Clustered | 45 | VC_423_0 | 45 | 0.9484 | 1 | 0.9484 | 0 | 0 | 0 | 1 | Ref |
| 2630 | KR054032 | Unassigned | Unassigned | Unassigned | 838_0 | Clustered | 29 | VC_838_0 | 29 | 0.8611 | 1 | 0.8611 | 0 | 0 | 0 | 1 | Ref |
| 2631 | KR054033 | Unassigned | Unassigned | Unassigned | 79_7 | Clustered | 33 | VC_79_7 | 33 | 0.216 | 1 | 0.216 | 0 | 0 | 0 | 0.8409 | Ref |
| 2632 | KR060090 | Unassigned | Unassigned | Unassigned | 896_0 | Clustered | 5 | VC_896_0 | 5 | 0.4981 | 0.99999977 | 0.4981 | 0 | 0 | 0 | 1 | Ref |
| 2633 | KR063267 | Unassigned | Unassigned | Unassigned | 875_0 | Clustered | 10 | VC_875_0 | 10 | 0.988 | 0.99948433 | 0.9875 | 0 | 0 | 0 | 1 | Ref |
| 2634 | KR063268 | Unassigned | Unassigned | Unassigned | 875_0 | Clustered | 10 | VC_875_0 | 10 | 0.988 | 0.99948433 | 0.9875 | 0 | 0 | 0 | 1 | Ref |
| 2635 | KR063278 | Unassigned | Unassigned | Unassigned | 866_0 | Clustered | 4 | VC_866_0 | 4 | 0.0795 | 0.99998595 | 0.0795 | 0 | 0 | 0 | 1 | Ref |
| 2636 | KR063279 | Unassigned | Unassigned | Unassigned | 867_0 | Clustered | 6 | VC_867_0 | 6 | 0.3202 | 1 | 0.3202 | 0 | 0 | 0 | 1 | Ref |
| 2637 | KR063280 | Unassigned | Unassigned | Unassigned | 1096_0 | Clustered | 5 | VC_1096_0 | 5 | 0.8566 | 0.99869977 | 0.8555 | 0 | 0 | 0 | 1 | Ref |
| 2638 | KR063281 | Unassigned | Unassigned | Unassigned | 1098_1 | Clustered/Singleton | 1 | VC_1098_1 | 1 | 0 | 1 | 0 | 0 | 0 | 0 | 1 | Ref |
| 2639 | KR073660 | Unassigned | Unassigned | Unassigned | 228_0 | Clustered | 60 | VC_228_0 | 60 | 0.942 | 1 | 0.942 | 0 | 0 | 0 | 1 | Ref |
| 2640 | KR073661 | Unassigned | Unassigned | Unassigned | 228_0 | Clustered | 60 | VC_228_0 | 60 | 0.942 | 1 | 0.942 | 0 | 0 | 0 | 1 | Ref |
| 2641 | KR080193 | Unassigned | Unassigned | Unassigned | 220_0 | Clustered | 4 | VC_220_0 | 4 | 0.0615 | 0.99998802 | 0.0615 | 0 | 0 | 0 | 1 | Ref |
| 2642 | KR080194 | Unassigned | Unassigned | Unassigned | 372_0 | Clustered | 24 | VC_372_0 | 24 | 0.5539 | 1 | 0.5539 | 0 | 0 | 0 | 1 | Ref |
| 2643 | KR080195 | Unassigned | Unassigned | Unassigned | 519_0 | Clustered | 39 | VC_519_0 | 39 | 0.8156 | 1 | 0.8156 | 0 | 0 | 0 | 1 | Ref |
| 2644 | KR080196 | Unassigned | Unassigned | Unassigned | 558_20 | Clustered | 17 | VC_558_20 | 17 | 0.054 | 1 | 0.054 | 0 | 0 | 0 | 0.9926 | Ref |
| 2645 | KR080197 | Unassigned | Unassigned | Unassigned | 762_0 | Clustered | 9 | VC_762_0 | 9 | 0.0885 | 1 | 0.0885 | 0 | 0 | 0 | 1 | Ref |
| 2646 | KR080198 | Unassigned | Unassigned | Unassigned | 762_0 | Clustered | 9 | VC_762_0 | 9 | 0.0885 | 1 | 0.0885 | 0 | 0 | 0 | 1 | Ref |
| 2647 | KR080199 | Unassigned | Unassigned | Unassigned | 372_1 | Clustered | 9 | VC_372_1 | 9 | 0.194 | 1 | 0.194 | 0 | 0 | 0 | 1 | Ref |
| 2648 | KR080200 | Unassigned | Unassigned | Unassigned | 372_0 | Clustered | 24 | VC_372_0 | 24 | 0.5539 | 1 | 0.5539 | 0 | 0 | 0 | 1 | Ref |
| 2649 | KR080201 | Unassigned | Unassigned | Unassigned | 325_35 | Clustered | 17 | VC_325_35 | 17 | 0.0732 | 1 | 0.0732 | 0 | 0 | 0 | 0.7721 | Ref |
| 2650 | KR080202 | Unassigned | Unassigned | Unassigned | 762_0 | Clustered | 9 | VC_762_0 | 9 | 0.0885 | 1 | 0.0885 | 0 | 0 | 0 | 1 | Ref |
| 2651 | KR080203 | Unassigned | Unassigned | Unassigned | 509_0 | Clustered | 37 | VC_509_0 | 37 | 0.9689 | 1 | 0.9689 | 0 | 0 | 0 | 1 | Ref |
| 2652 | KR080204 | Unassigned | Unassigned | Unassigned | 551_2 | Clustered | 17 | VC_551_2 | 17 | 0.0973 | 1 | 0.0973 | 0 | 0 | 0 | 0.9779 | Ref |
| 2653 | KR080205 | Unassigned | Unassigned | Unassigned | 509_0 | Clustered | 37 | VC_509_0 | 37 | 0.9689 | 1 | 0.9689 | 0 | 0 | 0 | 1 | Ref |
| 2654 | KR080206 | Unassigned | Unassigned | Unassigned | 237_1 | Clustered | 13 | VC_237_1 | 13 | 0.1773 | 1 | 0.1773 | 0 | 0 | 0 | 0.7308 | Ref |
| 2655 | KR091952 | Unassigned | Unassigned | Unassigned | 129_0 | Clustered | 43 | VC_129_0 | 43 | 0.886 | 1 | 0.886 | 0 | 0 | 0 | 1 | Ref |
| 2656 | KR093625 | Unassigned | Unassigned | Unassigned | 933_0 | Clustered | 8 | VC_933_0 | 8 | 0.3648 | 1 | 0.3648 | 0 | 0 | 0 | 1 | Ref |
| 2657 | KR093626 | Unassigned | Unassigned | Unassigned | 933_0 | Clustered | 8 | VC_933_0 | 8 | 0.3648 | 1 | 0.3648 | 0 | 0 | 0 | 1 | Ref |
| 2658 | KR093627 | Unassigned | Unassigned | Unassigned | 933_0 | Clustered | 8 | VC_933_0 | 8 | 0.3648 | 1 | 0.3648 | 0 | 0 | 0 | 1 | Ref |
| 2659 | KR093628 | Unassigned | Unassigned | Unassigned | 933_0 | Clustered | 8 | VC_933_0 | 8 | 0.3648 | 1 | 0.3648 | 0 | 0 | 0 | 1 | Ref |
| 2660 | KR093629 | Unassigned | Unassigned | Unassigned | 933_0 | Clustered | 8 | VC_933_0 | 8 | 0.3648 | 1 | 0.3648 | 0 | 0 | 0 | 1 | Ref |
| 2661 | KR093630 | Unassigned | Unassigned | Unassigned | 933_0 | Clustered | 8 | VC_933_0 | 8 | 0.3648 | 1 | 0.3648 | 0 | 0 | 0 | 1 | Ref |
| 2662 | KR093631 | Unassigned | Unassigned | Unassigned | 933_0 | Clustered | 8 | VC_933_0 | 8 | 0.3648 | 1 | 0.3648 | 0 | 0 | 0 | 1 | Ref |
| 2663 | KR093633 | Unassigned | Unassigned | Unassigned | 933_0 | Clustered | 8 | VC_933_0 | 8 | 0.3648 | 1 | 0.3648 | 0 | 0 | 0 | 1 | Ref |
| 2664 | KR093634 | Unassigned | Unassigned | Unassigned | 604_0 | Clustered | 22 | VC_604_0 | 22 | 0.7846 | 1 | 0.7846 | 0 | 0 | 0 | 1 | Ref |
| 2665 | KR093635 | Unassigned | Unassigned | Unassigned | 604_0 | Clustered | 22 | VC_604_0 | 22 | 0.7846 | 1 | 0.7846 | 0 | 0 | 0 | 1 | Ref |
| 2666 | KR093636 | Unassigned | Unassigned | Unassigned | 604_0 | Clustered | 22 | VC_604_0 | 22 | 0.7846 | 1 | 0.7846 | 0 | 0 | 0 | 1 | Ref |
| 2667 | KR093637 | Unassigned | Unassigned | Unassigned | 604_0 | Clustered | 22 | VC_604_0 | 22 | 0.7846 | 1 | 0.7846 | 0 | 0 | 0 | 1 | Ref |
| 2668 | KR093639 | Unassigned | Unassigned | Unassigned | 604_0 | Clustered | 22 | VC_604_0 | 22 | 0.7846 | 1 | 0.7846 | 0 | 0 | 0 | 1 | Ref |
| 2669 | KR093640 | Unassigned | Unassigned | Unassigned | 604_0 | Clustered | 22 | VC_604_0 | 22 | 0.7846 | 1 | 0.7846 | 0 | 0 | 0 | 1 | Ref |
| 2670 | KR093641 | Unassigned | Unassigned | Unassigned | 604_0 | Clustered | 22 | VC_604_0 | 22 | 0.7846 | 1 | 0.7846 | 0 | 0 | 0 | 1 | Ref |
| 2671 | KR093642 | Unassigned | Unassigned | Unassigned | 604_0 | Clustered | 22 | VC_604_0 | 22 | 0.7846 | 1 | 0.7846 | 0 | 0 | 0 | 1 | Ref |
| 2672 | KR093643 | Unassigned | Unassigned | Unassigned | 604_0 | Clustered | 22 | VC_604_0 | 22 | 0.7846 | 1 | 0.7846 | 0 | 0 | 0 | 1 | Ref |
| 2673 | KR093644 | Unassigned | Unassigned | Unassigned | 604_0 | Clustered | 22 | VC_604_0 | 22 | 0.7846 | 1 | 0.7846 | 0 | 0 | 0 | 1 | Ref |
| 2674 | KR093645 | Unassigned | Unassigned | Unassigned | 604_0 | Clustered | 22 | VC_604_0 | 22 | 0.7846 | 1 | 0.7846 | 0 | 0 | 0 | 1 | Ref |
| 2675 | KR093646 | Unassigned | Unassigned | Unassigned | 604_0 | Clustered | 22 | VC_604_0 | 22 | 0.7846 | 1 | 0.7846 | 0 | 0 | 0 | 1 | Ref |
| 2676 | KR093647 | Unassigned | Unassigned | Unassigned | 604_0 | Clustered | 22 | VC_604_0 | 22 | 0.7846 | 1 | 0.7846 | 0 | 0 | 0 | 1 | Ref |
| 2677 | KR093648 | Unassigned | Unassigned | Unassigned | 604_0 | Clustered | 22 | VC_604_0 | 22 | 0.7846 | 1 | 0.7846 | 0 | 0 | 0 | 1 | Ref |
| 2678 | KR093649 | Unassigned | Unassigned | Unassigned | 604_0 | Clustered | 22 | VC_604_0 | 22 | 0.7846 | 1 | 0.7846 | 0 | 0 | 0 | 1 | Ref |
| 2679 | KR093650 | Unassigned | Unassigned | Unassigned | 604_0 | Clustered | 22 | VC_604_0 | 22 | 0.7846 | 1 | 0.7846 | 0 | 0 | 0 | 1 | Ref |
| 2680 | KR093651 | Unassigned | Unassigned | Unassigned | 604_0 | Clustered | 22 | VC_604_0 | 22 | 0.7846 | 1 | 0.7846 | 0 | 0 | 0 | 1 | Ref |
| 2681 | KR093652 | Unassigned | Unassigned | Unassigned | 604_0 | Clustered | 22 | VC_604_0 | 22 | 0.7846 | 1 | 0.7846 | 0 | 0 | 0 | 1 | Ref |
| 2682 | KR093653 | Unassigned | Unassigned | Unassigned | 604_0 | Clustered | 22 | VC_604_0 | 22 | 0.7846 | 1 | 0.7846 | 0 | 0 | 0 | 1 | Ref |
| 2683 | KR093654 | Unassigned | Unassigned | Unassigned | 604_0 | Clustered | 22 | VC_604_0 | 22 | 0.7846 | 1 | 0.7846 | 0 | 0 | 0 | 1 | Ref |
| 2684 | KR093655 | Unassigned | Unassigned | Unassigned | 604_0 | Clustered | 22 | VC_604_0 | 22 | 0.7846 | 1 | 0.7846 | 0 | 0 | 0 | 1 | Ref |
| 2685 | KR093656 | Unassigned | Unassigned | Unassigned | 604_0 | Clustered | 22 | VC_604_0 | 22 | 0.7846 | 1 | 0.7846 | 0 | 0 | 0 | 1 | Ref |
| 2686 | KR131750 | Unassigned | Unassigned | Unassigned | 695_0 | Clustered | 48 | VC_695_0 | 48 | 0.8162 | 1 | 0.8162 | 0 | 0 | 0 | 1 | Ref |
| 2687 | KR136259 | Unassigned | Unassigned | Unassigned | 1101_0 | Clustered | 2 | VC_1101_0 | 2 | 0.9786 | 0.72985431 | 0.7143 | 0 | 0 | 0 | 1 | Ref |
| 2688 | KR136260 | Unassigned | Unassigned | Unassigned | 1101_0 | Clustered | 2 | VC_1101_0 | 2 | 0.9786 | 0.72985431 | 0.7143 | 0 | 0 | 0 | 1 | Ref |
| 2689 | KR149290 | Unassigned | Unassigned | Unassigned | 699_0 | Clustered | 71 | VC_699_0 | 71 | 0.9352 | 1 | 0.9352 | 0 | 0 | 0 | 1 | Ref |
| 2690 | KR149291 | Unassigned | Unassigned | Unassigned | 823_0 | Clustered | 124 | VC_823_0 | 124 | 0.9492 | 1 | 0.9492 | 0 | 0 | 0 | 1 | Ref |
| 2691 | KR153145 | Unassigned | Unassigned | Unassigned | 463_0 | Clustered | 4 | VC_463_0 | 4 | 0.7906 | 0.99997156 | 0.7906 | 0 | 0 | 0 | 1 | Ref |
| 2692 | KR233164 | Unassigned | Unassigned | Unassigned | 190_27 | Clustered/Singleton | 1 | VC_190_27 | 1 | 0 | 1 | 0 | 0 | 0 | 0 | 1 | Ref |
| 2693 | KR233165 | Unassigned | Unassigned | Unassigned | 283_163 | Clustered | 2 | VC_283_163 | 2 | 0.0029 | 0.93420746 | 0.0027 | 0 | 0 | 0 | 1 | Ref |
| 2694 | KR262148 | Unassigned | Unassigned | Unassigned | 938_0 | Clustered | 99 | VC_938_0 | 99 | 0.9119 | 1 | 0.9119 | 0 | 0 | 0 | 0.9928 | Ref |
| 2695 | KR269718 | Unassigned | Unassigned | Unassigned | 283_81 | Clustered/Singleton | 1 | VC_283_81 | 1 | 0 | 1 | 0 | 0 | 0 | 0 | 1 | Ref |
| 2696 | KR269719 | Unassigned | Unassigned | Unassigned | 937_0 | Clustered | 40 | VC_937_0 | 40 | 0.6809 | 1 | 0.6809 | 0 | 0 | 0 | 0.9359 | Ref |
| 2697 | KR269720 | Unassigned | Unassigned | Unassigned | 491_0 | Clustered | 36 | VC_491_0 | 36 | 0.5336 | 1 | 0.5336 | 0 | 0 | 0 | 0.9714 | Ref |
| 2698 | KR270151 | Unassigned | Unassigned | Unassigned | 637_0 | Clustered | 115 | VC_637_0 | 115 | 0.731 | 1 | 0.731 | 0 | 0 | 0 | 0.8764 | Ref |
| 2699 | KR296686 | Unassigned | Unassigned | Unassigned | 28_1 | Clustered | 53 | VC_28_1 | 53 | 0.7164 | 1 | 0.7164 | 0 | 0 | 0 | 0.9325 | Ref |
| 2700 | KR296687 | Unassigned | Unassigned | Unassigned | 28_1 | Clustered | 53 | VC_28_1 | 53 | 0.7164 | 1 | 0.7164 | 0 | 0 | 0 | 0.9325 | Ref |
| 2701 | KR296688 | Unassigned | Unassigned | Unassigned | 28_1 | Clustered | 53 | VC_28_1 | 53 | 0.7164 | 1 | 0.7164 | 0 | 0 | 0 | 0.9325 | Ref |
| 2702 | KR296689 | Unassigned | Unassigned | Unassigned | 592_0 | Clustered | 50 | VC_592_0 | 50 | 0.7314 | 1 | 0.7314 | 0 | 0 | 0 | 1 | Ref |
| 2703 | KR296691 | Unassigned | Unassigned | Unassigned | 592_0 | Clustered | 50 | VC_592_0 | 50 | 0.7314 | 1 | 0.7314 | 0 | 0 | 0 | 1 | Ref |
| 2704 | KR296692 | Unassigned | Unassigned | Unassigned | 133_18 | Clustered/Singleton | 1 | VC_133_18 | 1 | 0 | 1 | 0 | 0 | 0 | 0 | 1 | Ref |
| 2705 | KR337643 | Unassigned | Unassigned | Unassigned | 765_0 | Clustered | 132 | VC_765_0 | 132 | 1 | 1 | 1 | 0 | 0 | 0 | 1 | Ref |
| 2706 | KR337644 | Unassigned | Unassigned | Unassigned | 765_0 | Clustered | 132 | VC_765_0 | 132 | 1 | 1 | 1 | 0 | 0 | 0 | 1 | Ref |
| 2707 | KR337645 | Unassigned | Unassigned | Unassigned | 765_0 | Clustered | 132 | VC_765_0 | 132 | 1 | 1 | 1 | 0 | 0 | 0 | 1 | Ref |
| 2708 | KR337646 | Unassigned | Unassigned | Unassigned | 765_0 | Clustered | 132 | VC_765_0 | 132 | 1 | 1 | 1 | 0 | 0 | 0 | 1 | Ref |
| 2709 | KR337647 | Unassigned | Unassigned | Unassigned | 765_0 | Clustered | 132 | VC_765_0 | 132 | 1 | 1 | 1 | 0 | 0 | 0 | 1 | Ref |
| 2710 | KR337648 | Unassigned | Unassigned | Unassigned | 765_0 | Clustered | 132 | VC_765_0 | 132 | 1 | 1 | 1 | 0 | 0 | 0 | 1 | Ref |
| 2711 | KR337649 | Unassigned | Unassigned | Unassigned | 765_0 | Clustered | 132 | VC_765_0 | 132 | 1 | 1 | 1 | 0 | 0 | 0 | 1 | Ref |
| 2712 | KR337650 | Unassigned | Unassigned | Unassigned | 765_0 | Clustered | 132 | VC_765_0 | 132 | 1 | 1 | 1 | 0 | 0 | 0 | 1 | Ref |
| 2713 | KR337651 | Unassigned | Unassigned | Unassigned | 765_0 | Clustered | 132 | VC_765_0 | 132 | 1 | 1 | 1 | 0 | 0 | 0 | 1 | Ref |
| 2714 | KR337652 | Unassigned | Unassigned | Unassigned | 765_0 | Clustered | 132 | VC_765_0 | 132 | 1 | 1 | 1 | 0 | 0 | 0 | 1 | Ref |
| 2715 | KR337653 | Unassigned | Unassigned | Unassigned | 765_0 | Clustered | 132 | VC_765_0 | 132 | 1 | 1 | 1 | 0 | 0 | 0 | 1 | Ref |
| 2716 | KR337654 | Unassigned | Unassigned | Unassigned | 765_0 | Clustered | 132 | VC_765_0 | 132 | 1 | 1 | 1 | 0 | 0 | 0 | 1 | Ref |
| 2717 | KR347352 | Unassigned | Unassigned | Unassigned | 765_0 | Clustered | 132 | VC_765_0 | 132 | 1 | 1 | 1 | 0 | 0 | 0 | 1 | Ref |
| 2718 | KR347353 | Unassigned | Unassigned | Unassigned | 765_0 | Clustered | 132 | VC_765_0 | 132 | 1 | 1 | 1 | 0 | 0 | 0 | 1 | Ref |
| 2719 | KR347354 | Unassigned | Unassigned | Unassigned | 765_0 | Clustered | 132 | VC_765_0 | 132 | 1 | 1 | 1 | 0 | 0 | 0 | 1 | Ref |
| 2720 | KR347355 | Unassigned | Unassigned | Unassigned | 765_0 | Clustered | 132 | VC_765_0 | 132 | 1 | 1 | 1 | 0 | 0 | 0 | 1 | Ref |
| 2721 | KR422352 | Unassigned | Unassigned | Unassigned | 286_1 | Clustered | 21 | VC_286_1 | 21 | 0.1696 | 1 | 0.1696 | 0 | 0 | 0 | 0.9048 | Ref |
| 2722 | KR422353 | Unassigned | Unassigned | Unassigned | 190_10 | Clustered/Singleton | 1 | VC_190_10 | 1 | 0 | 1 | 0 | 0 | 0 | 0 | 1 | Ref |
| 2723 | KR534323 | Unassigned | Unassigned | Unassigned | 496_2 | Clustered/Singleton | 1 | VC_496_2 | 1 | 0 | 1 | 0 | 0 | 0 | 0 | 1 | Ref |
| 2724 | KR537871 | Unassigned | Unassigned | Unassigned | 749_0 | Clustered | 40 | VC_749_0 | 40 | 0.7517 | 1 | 0.7517 | 0 | 0 | 0 | 1 | Ref |
| 2725 | KR537872 | Unassigned | Unassigned | Unassigned | 749_0 | Clustered | 40 | VC_749_0 | 40 | 0.7517 | 1 | 0.7517 | 0 | 0 | 0 | 1 | Ref |
| 2726 | KR560069 | Unassigned | Unassigned | Unassigned | 506_0 | Clustered | 8 | VC_506_0 | 8 | 0.7376 | 1 | 0.7376 | 0 | 0 | 0 | 1 | Ref |
| 2727 | KR604693 | Unassigned | Unassigned | Unassigned | 984_0 | Clustered | 38 | VC_984_0 | 38 | 0.9758 | 1 | 0.9758 | 0 | 0 | 0 | 1 | Ref |
| 2728 | KR698074 | Unassigned | Unassigned | Unassigned | 772_8 | Clustered | 2 | VC_772_8 | 2 | 0.006 | 0.58293682 | 0.0035 | 0 | 0 | 0 | 1 | Ref |
| 2729 | KR704913 | Unassigned | Unassigned | Unassigned | 1103_0 | Clustered | 2 | VC_1103_0 | 2 | 1 | 1 | 1 | 0 | 0 | 0 | 1 | Ref |
| 2730 | KR704915 | Unassigned | Unassigned | Unassigned | 1103_0 | Clustered | 2 | VC_1103_0 | 2 | 1 | 1 | 1 | 0 | 0 | 0 | 1 | Ref |
| 2731 | KR709302 | Unassigned | Unassigned | Unassigned | 12_2 | Clustered | 19 | VC_12_2 | 19 | 0.2018 | 1 | 0.2018 | 0 | 0 | 0 | 1 | Ref |
| 2732 | KR709303 | Unassigned | Unassigned | Unassigned | 12_2 | Clustered | 19 | VC_12_2 | 19 | 0.2018 | 1 | 0.2018 | 0 | 0 | 0 | 1 | Ref |
| 2733 | KR781349 | Unassigned | Unassigned | Unassigned | 371_21 | Clustered | 2 | VC_371_21 | 2 | 0.0033 | 0.95770737 | 0.0032 | 0 | 0 | 0 | 1 | Ref |
| 2734 | KR781488 | Unassigned | Unassigned | Unassigned | 42_0 | Clustered | 49 | VC_42_0 | 49 | 0.4517 | 1 | 0.4517 | 0 | 0 | 0 | 0.8597 | Ref |
| 2735 | KR816341 | Unassigned | Unassigned | Unassigned | 1109_0 | Clustered | 2 | VC_1109_0 | 2 | 0.6449 | 0.85557782 | 0.5517 | 0 | 0 | 0 | 1 | Ref |
| 2736 | KR816508 | Unassigned | Unassigned | Unassigned | 371_18 | Clustered | 10 | VC_371_18 | 10 | 0.0302 | 1 | 0.0302 | 0 | 0 | 0 | 0.8 | Ref |
| 2737 | KR824843 | Unassigned | Unassigned | Unassigned | 520_2 | Clustered/Singleton | 1 | VC_520_2 | 1 | 0 | 1 | 0 | 0 | 0 | 0 | 1 | Ref |
| 2738 | KR869157 | Unassigned | Unassigned | Unassigned | 79_0 | Clustered | 64 | VC_79_0 | 64 | 0.4185 | 1 | 0.4185 | 0 | 0 | 0 | 0.7649 | Ref |
| 2739 | KR869820 | Unassigned | Unassigned | Unassigned | 669_1 | Clustered | 10 | VC_669_1 | 10 | 0.1268 | 1 | 0.1268 | 0 | 0 | 0 | 0.9778 | Ref |
| 2740 | KR902361 | Unassigned | Unassigned | Unassigned | 149_8 | Clustered | 66 | VC_149_8 | 66 | 0.4565 | 1 | 0.4565 | 0 | 0 | 0 | 0.8182 | Ref |
| 2741 | KR902978 | Unassigned | Unassigned | Unassigned | 765_0 | Clustered | 132 | VC_765_0 | 132 | 1 | 1 | 1 | 0 | 0 | 0 | 1 | Ref |
| 2742 | KR902979 | Unassigned | Unassigned | Unassigned | 765_0 | Clustered | 132 | VC_765_0 | 132 | 1 | 1 | 1 | 0 | 0 | 0 | 1 | Ref |
| 2743 | KR902980 | Unassigned | Unassigned | Unassigned | 765_0 | Clustered | 132 | VC_765_0 | 132 | 1 | 1 | 1 | 0 | 0 | 0 | 1 | Ref |
| 2744 | KR902981 | Unassigned | Unassigned | Unassigned | 765_0 | Clustered | 132 | VC_765_0 | 132 | 1 | 1 | 1 | 0 | 0 | 0 | 1 | Ref |
| 2745 | KR902982 | Unassigned | Unassigned | Unassigned | 765_0 | Clustered | 132 | VC_765_0 | 132 | 1 | 1 | 1 | 0 | 0 | 0 | 1 | Ref |
| 2746 | KR902983 | Unassigned | Unassigned | Unassigned | 765_0 | Clustered | 132 | VC_765_0 | 132 | 1 | 1 | 1 | 0 | 0 | 0 | 1 | Ref |
| 2747 | KR902984 | Unassigned | Unassigned | Unassigned | 765_0 | Clustered | 132 | VC_765_0 | 132 | 1 | 1 | 1 | 0 | 0 | 0 | 1 | Ref |
| 2748 | KR902985 | Unassigned | Unassigned | Unassigned | 765_0 | Clustered | 132 | VC_765_0 | 132 | 1 | 1 | 1 | 0 | 0 | 0 | 1 | Ref |
| 2749 | KR902986 | Unassigned | Unassigned | Unassigned | 765_0 | Clustered | 132 | VC_765_0 | 132 | 1 | 1 | 1 | 0 | 0 | 0 | 1 | Ref |
| 2750 | KR902987 | Unassigned | Unassigned | Unassigned | 765_0 | Clustered | 132 | VC_765_0 | 132 | 1 | 1 | 1 | 0 | 0 | 0 | 1 | Ref |
| 2751 | KR905068 | Unassigned | Unassigned | Unassigned | 377_0 | Clustered | 3 | VC_377_0 | 3 | 0.3906 | 0.99785056 | 0.3898 | 0 | 0 | 0 | 1 | Ref |
| 2752 | KR908644 | Unassigned | Unassigned | Unassigned | 149_8 | Clustered | 66 | VC_149_8 | 66 | 0.4565 | 1 | 0.4565 | 0 | 0 | 0 | 0.8182 | Ref |

| | | | | | | | | | | | | | | | | | |
|---|---|---|---|---|---|---|---|---|---|---|---|---|---|---|---|---|---|
| 2753 | KR935213 | Unassigned | Unassigned | Unassigned | 749_0 | Clustered | 40 | VC_749_0 | 40 | 0.7517 | 1 | 0.7517 | 0 | 0 | 0 | 1 | Ref |
| 2754 | KR935214 | Unassigned | Unassigned | Unassigned | 520_73 | Clustered | 2 | VC_520_73 | 2 | 0.0107 | 0.95762743 | 0.0103 | 0 | 0 | 0 | 1 | Ref |
| 2755 | KR935215 | Unassigned | Unassigned | Unassigned | 749_0 | Clustered | 40 | VC_749_0 | 40 | 0.7517 | 1 | 0.7517 | 0 | 0 | 0 | 1 | Ref |
| 2756 | KR935216 | Unassigned | Unassigned | Unassigned | 874_0 | Clustered | 3 | VC_874_0 | 3 | 0.6293 | 0.99812483 | 0.6281 | 0 | 0 | 0 | 1 | Ref |
| 2757 | KR935217 | Unassigned | Unassigned | Unassigned | 874_0 | Clustered | 3 | VC_874_0 | 3 | 0.6293 | 0.99812483 | 0.6281 | 0 | 0 | 0 | 1 | Ref |
| 2758 | KR997929 | Unassigned | Unassigned | Unassigned | 325_7 | Clustered | 3 | VC_325_7 | 3 | 0.0085 | 0.99739782 | 0.0085 | 0 | 0 | 0 | 0.6667 | Ref |
| 2759 | KR997930 | Unassigned | Unassigned | Unassigned | 520_35 | Clustered | 3 | VC_520_35 | 3 | 0.0251 | 0.99859857 | 0.0251 | 0 | 0 | 0 | 1 | Ref |
| 2760 | KR997931 | Unassigned | Unassigned | Unassigned | 520_35 | Clustered | 3 | VC_520_35 | 3 | 0.0251 | 0.99859857 | 0.0251 | 0 | 0 | 0 | 1 | Ref |
| 2761 | KR997932 | Unassigned | Unassigned | Unassigned | 508_0 | Clustered | 32 | VC_508_0 | 32 | 0.906 | 1 | 0.906 | 0 | 0 | 0 | 1 | Ref |
| 2762 | KR997933 | Unassigned | Unassigned | Unassigned | 569_0 | Clustered | 3 | VC_569_0 | 3 | 0.2094 | 0.99849867 | 0.2091 | 0 | 0 | 0 | 1 | Ref |
| 2763 | KR997934 | Unassigned | Unassigned | Unassigned | 325_7 | Clustered | 3 | VC_325_7 | 3 | 0.0085 | 0.99739782 | 0.0085 | 0 | 0 | 0 | 0.6667 | Ref |
| 2764 | KR997967 | Unassigned | Unassigned | Unassigned | 371_1 | Clustered | 11 | VC_371_1 | 11 | 0.0379 | 1 | 0.0379 | 0 | 0 | 0 | 0.8545 | Ref |
| 2765 | KR997968 | Unassigned | Unassigned | Unassigned | 520_35 | Clustered | 3 | VC_520_35 | 3 | 0.0251 | 0.99859857 | 0.0251 | 0 | 0 | 0 | 1 | Ref |
| 2766 | KT001911 | Unassigned | Unassigned | Unassigned | 1032_0 | Clustered | 7 | VC_1032_0 | 7 | 0.9887 | 0.99999504 | 0.9887 | 0 | 0 | 0 | 1 | Ref |
| 2767 | KT001912 | Unassigned | Unassigned | Unassigned | 102_0 | Clustered | 2 | VC_102_0 | 2 | 0.114 | 0.94637553 | 0.1079 | 0 | 0 | 0 | 1 | Ref |
| 2768 | KT001913 | Unassigned | Unassigned | Unassigned | 170_0 | Clustered/Singleton | 1 | VC_170_0 | 1 | 0 | 1 | 0 | 0 | 0 | 0 | 1 | Ref |
| 2769 | KT001914 | Unassigned | Unassigned | Unassigned | 1090_0 | Clustered | 4 | VC_1090_0 | 4 | 1 | 1 | 1 | 0 | 0 | 0 | 1 | Ref |
| 2770 | KT001916 | Unassigned | Unassigned | Unassigned | 330_0 | Clustered | 27 | VC_330_0 | 27 | 0.5088 | 1 | 0.5088 | 0 | 0 | 0 | 0.8661 | Ref |
| 2771 | KT001917 | Unassigned | Unassigned | Unassigned | 772_19 | Clustered | 16 | VC_772_19 | 16 | 0.0975 | 0.9970466 | 0.0972 | 0 | 0 | 0 | 0.8083 | Ref |
| 2772 | KT001918 | Unassigned | Unassigned | Unassigned | 668_0 | Clustered | 25 | VC_668_0 | 25 | 0.5539 | 1 | 0.5539 | 0 | 0 | 0 | 0.96 | Ref |
| 2773 | KT001919 | Unassigned | Unassigned | Unassigned | 668_0 | Clustered | 25 | VC_668_0 | 25 | 0.5539 | 1 | 0.5539 | 0 | 0 | 0 | 0.96 | Ref |
| 2774 | KT001920 | Unassigned | Unassigned | Unassigned | 938_0 | Clustered | 99 | VC_938_0 | 99 | 0.9119 | 1 | 0.9119 | 0 | 0 | 0 | 0.9928 | Ref |
| 2775 | KT004677 | Unassigned | Unassigned | Unassigned | 940_0 | Clustered | 34 | VC_940_0 | 34 | 0.7959 | 1 | 0.7959 | 0 | 0 | 0 | 1 | Ref |
| 2776 | KT008108 | Unassigned | Unassigned | Unassigned | 385_0 | Clustered | 51 | VC_385_0 | 51 | 0.8754 | 1 | 0.8754 | 0 | 0 | 0 | 1 | Ref |
| 2777 | KT020852 | Unassigned | Unassigned | Unassigned | 551_7 | Clustered | 28 | VC_551_7 | 28 | 0.1661 | 1 | 0.1661 | 0 | 0 | 0 | 0.8519 | Ref |
| 2778 | KT070866 | Unassigned | Unassigned | Unassigned | 1006_0 | Clustered | 8 | VC_1006_0 | 8 | 0.7407 | 1 | 0.7407 | 0 | 0 | 0 | 1 | Ref |
| 2779 | KT070867 | Unassigned | Unassigned | Unassigned | 475_6 | Clustered/Singleton | 1 | VC_475_6 | 1 | 0 | 1 | 0 | 0 | 0 | 0 | 1 | Ref |
| 2780 | KT124227 | Unassigned | Unassigned | Unassigned | 982_0 | Clustered | 62 | VC_982_0 | 62 | 0.8646 | 1 | 0.8646 | 0 | 0 | 0 | 0.743 | Ref |
| 2781 | KT124228 | Unassigned | Unassigned | Unassigned | 982_0 | Clustered | 62 | VC_982_0 | 62 | 0.8646 | 1 | 0.8646 | 0 | 0 | 0 | 0.743 | Ref |
| 2782 | KT124229 | Unassigned | Unassigned | Unassigned | 982_0 | Clustered | 62 | VC_982_0 | 62 | 0.8646 | 1 | 0.8646 | 0 | 0 | 0 | 0.743 | Ref |
| 2783 | KT151955 | Unassigned | Unassigned | Unassigned | 720_0 | Clustered | 2 | VC_720_0 | 2 | 0.5579 | 0.89435023 | 0.4989 | 0 | 0 | 0 | 1 | Ref |
| 2784 | KT151956 | Unassigned | Unassigned | Unassigned | 141_0 | Clustered | 6 | VC_141_0 | 6 | 0.5999 | 1 | 0.5999 | 0 | 0 | 0 | 1 | Ref |
| 2785 | KT151957 | Unassigned | Unassigned | Unassigned | 720_0 | Clustered | 2 | VC_720_0 | 2 | 0.5579 | 0.89435023 | 0.4989 | 0 | 0 | 0 | 1 | Ref |
| 2786 | KT151958 | Unassigned | Unassigned | Unassigned | 141_0 | Clustered | 6 | VC_141_0 | 6 | 0.5999 | 1 | 0.5999 | 0 | 0 | 0 | 1 | Ref |
| 2787 | KT151959 | Unassigned | Unassigned | Unassigned | 211_2 | Clustered/Singleton | 1 | VC_211_2 | 1 | 0 | 1 | 0 | 0 | 0 | 0 | 1 | Ref |
| 2788 | KT152029 | Unassigned | Unassigned | Unassigned | 982_0 | Clustered | 62 | VC_982_0 | 62 | 0.8646 | 1 | 0.8646 | 0 | 0 | 0 | 0.743 | Ref |
| 2789 | KT160311 | Unassigned | Unassigned | Unassigned | 499_0 | Clustered | 24 | VC_499_0 | 24 | 0.8569 | 1 | 0.8569 | 0 | 0 | 0 | 0.9529 | Ref |
| 2790 | KT176190 | Unassigned | Unassigned | Unassigned | 290_5 | Clustered | 14 | VC_290_5 | 14 | 0.1422 | 1 | 0.1422 | 0 | 0 | 0 | 0.9231 | Ref |
| 2791 | KT184308 | Unassigned | Unassigned | Unassigned | 283_66 | Clustered/Singleton | 1 | VC_283_66 | 1 | 0 | 1 | 0 | 0 | 0 | 0 | 1 | Ref |
| 2792 | KT184309 | Unassigned | Unassigned | Unassigned | 286_17 | Clustered | 8 | VC_286_17 | 8 | 0.0535 | 0.99997829 | 0.0535 | 0 | 0 | 0 | 0.6071 | Ref |
| 2793 | KT184310 | Unassigned | Unassigned | Unassigned | 286_17 | Clustered | 8 | VC_286_17 | 8 | 0.0535 | 0.99997829 | 0.0535 | 0 | 0 | 0 | 0.6071 | Ref |
| 2794 | KT184311 | Unassigned | Unassigned | Unassigned | 283_57 | Clustered/Singleton | 1 | VC_283_57 | 1 | 0 | 1 | 0 | 0 | 0 | 0 | 1 | Ref |
| 2795 | KT184312 | Unassigned | Unassigned | Unassigned | 283_63 | Clustered/Singleton | 1 | VC_283_63 | 1 | 0 | 1 | 0 | 0 | 0 | 0 | 1 | Ref |
| 2796 | KT184313 | Unassigned | Unassigned | Unassigned | 329_7 | Clustered | 18 | VC_329_7 | 18 | 0.0938 | 1 | 0.0938 | 0 | 0 | 0 | 0.7778 | Ref |
| 2797 | KT184314 | Unassigned | Unassigned | Unassigned | 329_7 | Clustered | 18 | VC_329_7 | 18 | 0.0938 | 1 | 0.0938 | 0 | 0 | 0 | 0.7778 | Ref |
| 2798 | KT184315 | Unassigned | Unassigned | Unassigned | 329_7 | Clustered | 18 | VC_329_7 | 18 | 0.0938 | 1 | 0.0938 | 0 | 0 | 0 | 0.7778 | Ref |
| 2799 | KT184316 | Unassigned | Unassigned | Unassigned | 329_7 | Clustered | 18 | VC_329_7 | 18 | 0.0938 | 1 | 0.0938 | 0 | 0 | 0 | 0.7778 | Ref |
| 2800 | KT184390 | Unassigned | Unassigned | Unassigned | 982_0 | Clustered | 62 | VC_982_0 | 62 | 0.8646 | 1 | 0.8646 | 0 | 0 | 0 | 0.743 | Ref |
| 2801 | KT184391 | Unassigned | Unassigned | Unassigned | 982_0 | Clustered | 62 | VC_982_0 | 62 | 0.8646 | 1 | 0.8646 | 0 | 0 | 0 | 0.743 | Ref |
| 2802 | KT184661 | Unassigned | Unassigned | Unassigned | 878_0 | Clustered | 32 | VC_878_0 | 32 | 0.9965 | 1 | 0.9965 | 0 | 0 | 0 | 1 | Ref |
| 2803 | KT186228 | Unassigned | Unassigned | Unassigned | 982_0 | Clustered | 62 | VC_982_0 | 62 | 0.8646 | 1 | 0.8646 | 0 | 0 | 0 | 0.743 | Ref |
| 2804 | KT186229 | Unassigned | Unassigned | Unassigned | 982_0 | Clustered | 62 | VC_982_0 | 62 | 0.8646 | 1 | 0.8646 | 0 | 0 | 0 | 0.743 | Ref |
| 2805 | KT186243 | Unassigned | Unassigned | Unassigned | 8_0 | Clustered | 58 | VC_8_0 | 58 | 0.8306 | 1 | 0.8306 | 0 | 0 | 0 | 0.9946 | Ref |
| 2806 | KT187252 | Unassigned | Unassigned | Unassigned | 187_1 | Clustered | 7 | VC_187_1 | 7 | 0.1423 | 1 | 0.1423 | 0 | 0 | 0 | 1 | Ref |
| 2807 | KT207918 | Unassigned | Unassigned | Unassigned | 884_0 | Clustered | 28 | VC_884_0 | 28 | 0.6211 | 1 | 0.6211 | 0 | 0 | 0 | 0.9841 | Ref |
| 2808 | KT221034 | Unassigned | Unassigned | Unassigned | 587_0 | Clustered | 2 | VC_587_0 | 2 | 0.3252 | 0.91192888 | 0.2965 | 0 | 0 | 0 | 1 | Ref |
| 2809 | KT222940 | Unassigned | Unassigned | Unassigned | 810_0 | Clustered | 18 | VC_810_0 | 18 | 0.9121 | 1 | 0.9121 | 0 | 0 | 0 | 1 | Ref |
| 2810 | KT222941 | Unassigned | Unassigned | Unassigned | 551_2 | Clustered | 17 | VC_551_2 | 17 | 0.0973 | 1 | 0.0973 | 0 | 0 | 0 | 0.9779 | Ref |
| 2811 | KT222942 | Unassigned | Unassigned | Unassigned | 551_2 | Clustered | 17 | VC_551_2 | 17 | 0.0973 | 1 | 0.0973 | 0 | 0 | 0 | 0.9779 | Ref |
| 2812 | KT224359 | Unassigned | Unassigned | Unassigned | 187_1 | Clustered | 7 | VC_187_1 | 7 | 0.1423 | 1 | 0.1423 | 0 | 0 | 0 | 1 | Ref |
| 2813 | KT232076 | Unassigned | Unassigned | Unassigned | 43_0 | Clustered | 38 | VC_43_0 | 38 | 0.5885 | 1 | 0.5885 | 0 | 0 | 0 | 0.9659 | Ref |
| 2814 | KT239446 | Unassigned | Unassigned | Unassigned | 491_0 | Clustered | 36 | VC_491_0 | 36 | 0.5336 | 1 | 0.5336 | 0 | 0 | 0 | 0.9714 | Ref |
| 2815 | KT240186 | Unassigned | Unassigned | Unassigned | 984_0 | Clustered | 38 | VC_984_0 | 38 | 0.9758 | 1 | 0.9758 | 0 | 0 | 0 | 1 | Ref |
| 2816 | KT246485 | Unassigned | Unassigned | Unassigned | 810_0 | Clustered | 18 | VC_810_0 | 18 | 0.9121 | 1 | 0.9121 | 0 | 0 | 0 | 1 | Ref |
| 2817 | KT246486 | Unassigned | Unassigned | Unassigned | 940_0 | Clustered | 34 | VC_940_0 | 34 | 0.7959 | 1 | 0.7959 | 0 | 0 | 0 | 1 | Ref |
| 2818 | KT253150 | Unassigned | Unassigned | Unassigned | 874_0 | Clustered | 3 | VC_874_0 | 3 | 0.6293 | 0.99812483 | 0.6281 | 0 | 0 | 0 | 1 | Ref |
| 2819 | KT254130 | Unassigned | Unassigned | Unassigned | 80_0 | Clustered | 41 | VC_80_0 | 41 | 0.5837 | 1 | 0.5837 | 0 | 0 | 0 | 0.9805 | Ref |
| 2820 | KT254131 | Unassigned | Unassigned | Unassigned | 80_0 | Clustered | 41 | VC_80_0 | 41 | 0.5837 | 1 | 0.5837 | 0 | 0 | 0 | 0.9805 | Ref |
| 2821 | KT254132 | Unassigned | Unassigned | Unassigned | 80_0 | Clustered | 41 | VC_80_0 | 41 | 0.5837 | 1 | 0.5837 | 0 | 0 | 0 | 0.9805 | Ref |
| 2822 | KT254133 | Unassigned | Unassigned | Unassigned | 80_0 | Clustered | 41 | VC_80_0 | 41 | 0.5837 | 1 | 0.5837 | 0 | 0 | 0 | 0.9805 | Ref |
| 2823 | KT259047 | Unassigned | Unassigned | Unassigned | 325_23 | Clustered | 17 | VC_325_23 | 17 | 0.0716 | 1 | 0.0716 | 0 | 0 | 0 | 0.6397 | Ref |
| 2824 | KT264749 | Unassigned | Unassigned | Unassigned | 1104_0 | Clustered | 154 | VC_1104_0 | 154 | 0.8484 | 1 | 0.8484 | 0 | 0 | 0 | 1 | Ref |
| 2825 | KT264750 | Unassigned | Unassigned | Unassigned | 1104_0 | Clustered | 154 | VC_1104_0 | 154 | 0.8484 | 1 | 0.8484 | 0 | 0 | 0 | 1 | Ref |
| 2826 | KT264751 | Unassigned | Unassigned | Unassigned | 1104_0 | Clustered | 154 | VC_1104_0 | 154 | 0.8484 | 1 | 0.8484 | 0 | 0 | 0 | 1 | Ref |
| 2827 | KT264752 | Unassigned | Unassigned | Unassigned | 1104_0 | Clustered | 154 | VC_1104_0 | 154 | 0.8484 | 1 | 0.8484 | 0 | 0 | 0 | 1 | Ref |
| 2828 | KT264753 | Unassigned | Unassigned | Unassigned | 1104_0 | Clustered | 154 | VC_1104_0 | 154 | 0.8484 | 1 | 0.8484 | 0 | 0 | 0 | 1 | Ref |
| 2829 | KT264754 | Unassigned | Unassigned | Unassigned | 1104_0 | Clustered | 154 | VC_1104_0 | 154 | 0.8484 | 1 | 0.8484 | 0 | 0 | 0 | 1 | Ref |
| 2830 | KT264755 | Unassigned | Unassigned | Unassigned | 1104_0 | Clustered | 154 | VC_1104_0 | 154 | 0.8484 | 1 | 0.8484 | 0 | 0 | 0 | 1 | Ref |
| 2831 | KT264756 | Unassigned | Unassigned | Unassigned | 1104_0 | Clustered | 154 | VC_1104_0 | 154 | 0.8484 | 1 | 0.8484 | 0 | 0 | 0 | 1 | Ref |
| 2832 | KT264757 | Unassigned | Unassigned | Unassigned | 1104_0 | Clustered | 154 | VC_1104_0 | 154 | 0.8484 | 1 | 0.8484 | 0 | 0 | 0 | 1 | Ref |
| 2833 | KT264758 | Unassigned | Unassigned | Unassigned | 1104_0 | Clustered | 154 | VC_1104_0 | 154 | 0.8484 | 1 | 0.8484 | 0 | 0 | 0 | 1 | Ref |
| 2834 | KT264759 | Unassigned | Unassigned | Unassigned | 1104_0 | Clustered | 154 | VC_1104_0 | 154 | 0.8484 | 1 | 0.8484 | 0 | 0 | 0 | 1 | Ref |
| 2835 | KT264760 | Unassigned | Unassigned | Unassigned | 1104_0 | Clustered | 154 | VC_1104_0 | 154 | 0.8484 | 1 | 0.8484 | 0 | 0 | 0 | 1 | Ref |
| 2836 | KT264761 | Unassigned | Unassigned | Unassigned | 1104_0 | Clustered | 154 | VC_1104_0 | 154 | 0.8484 | 1 | 0.8484 | 0 | 0 | 0 | 1 | Ref |
| 2837 | KT264762 | Unassigned | Unassigned | Unassigned | 1104_0 | Clustered | 154 | VC_1104_0 | 154 | 0.8484 | 1 | 0.8484 | 0 | 0 | 0 | 1 | Ref |
| 2838 | KT264763 | Unassigned | Unassigned | Unassigned | 1104_0 | Clustered | 154 | VC_1104_0 | 154 | 0.8484 | 1 | 0.8484 | 0 | 0 | 0 | 1 | Ref |
| 2839 | KT264764 | Unassigned | Unassigned | Unassigned | 1104_0 | Clustered | 154 | VC_1104_0 | 154 | 0.8484 | 1 | 0.8484 | 0 | 0 | 0 | 1 | Ref |
| 2840 | KT264765 | Unassigned | Unassigned | Unassigned | 1104_0 | Clustered | 154 | VC_1104_0 | 154 | 0.8484 | 1 | 0.8484 | 0 | 0 | 0 | 1 | Ref |
| 2841 | KT264766 | Unassigned | Unassigned | Unassigned | 1104_0 | Clustered | 154 | VC_1104_0 | 154 | 0.8484 | 1 | 0.8484 | 0 | 0 | 0 | 1 | Ref |
| 2842 | KT264767 | Unassigned | Unassigned | Unassigned | 1104_0 | Clustered | 154 | VC_1104_0 | 154 | 0.8484 | 1 | 0.8484 | 0 | 0 | 0 | 1 | Ref |
| 2843 | KT264768 | Unassigned | Unassigned | Unassigned | 1104_0 | Clustered | 154 | VC_1104_0 | 154 | 0.8484 | 1 | 0.8484 | 0 | 0 | 0 | 1 | Ref |
| 2844 | KT264769 | Unassigned | Unassigned | Unassigned | 1104_0 | Clustered | 154 | VC_1104_0 | 154 | 0.8484 | 1 | 0.8484 | 0 | 0 | 0 | 1 | Ref |
| 2845 | KT264775 | Unassigned | Unassigned | Unassigned | 1108_0 | Clustered | 4 | VC_1108_0 | 4 | 0.0485 | 0.99999021 | 0.0485 | 0 | 0 | 0 | 1 | Ref |
| 2846 | KT264776 | Unassigned | Unassigned | Unassigned | 1116_0 | Clustered | 14 | VC_1116_0 | 14 | 0.7927 | 1 | 0.7927 | 0 | 0 | 0 | 1 | Ref |
| 2847 | KT264779 | Unassigned | Unassigned | Unassigned | 1104_0 | Clustered | 154 | VC_1104_0 | 154 | 0.8484 | 1 | 0.8484 | 0 | 0 | 0 | 1 | Ref |
| 2848 | KT264781 | Unassigned | Unassigned | Unassigned | 1117_0 | Clustered | 5 | VC_1117_0 | 5 | 0.4688 | 0.99999974 | 0.4688 | 0 | 0 | 0 | 1 | Ref |
| 2849 | KT264782 | Unassigned | Unassigned | Unassigned | 1119_0 | Clustered | 20 | VC_1119_0 | 20 | 0.6751 | 0.99997892 | 0.6751 | 0 | 0 | 0 | 1 | Ref |
| 2850 | KT264783 | Unassigned | Unassigned | Unassigned | 1119_0 | Clustered | 20 | VC_1119_0 | 20 | 0.6751 | 0.99997892 | 0.6751 | 0 | 0 | 0 | 1 | Ref |
| 2851 | KT264784 | Unassigned | Unassigned | Unassigned | 1123_0 | Clustered | 2 | VC_1123_0 | 2 | 0.3187 | 0.93270267 | 0.2972 | 0 | 0 | 0 | 1 | Ref |
| 2852 | KT264785 | Unassigned | Unassigned | Unassigned | 1122_0 | Clustered | 10 | VC_1122_0 | 10 | 0.9643 | 0.99999697 | 0.9643 | 0 | 0 | 0 | 1 | Ref |
| 2853 | KT264788 | Unassigned | Unassigned | Unassigned | 1126_0 | Clustered | 8 | VC_1126_0 | 8 | 0.2864 | 1 | 0.2864 | 0 | 0 | 0 | 1 | Ref |
| 2854 | KT264789 | Unassigned | Unassigned | Unassigned | 1126_0 | Clustered | 8 | VC_1126_0 | 8 | 0.2864 | 1 | 0.2864 | 0 | 0 | 0 | 1 | Ref |
| 2855 | KT264790 | Unassigned | Unassigned | Unassigned | 1126_0 | Clustered | 8 | VC_1126_0 | 8 | 0.2864 | 1 | 0.2864 | 0 | 0 | 0 | 1 | Ref |
| 2856 | KT264791 | Unassigned | Unassigned | Unassigned | 1127_0 | Clustered | 19 | VC_1127_0 | 19 | 0.7611 | 1 | 0.7611 | 0 | 0 | 0 | 1 | Ref |
| 2857 | KT264792 | Unassigned | Unassigned | Unassigned | 1127_0 | Clustered | 19 | VC_1127_0 | 19 | 0.7611 | 1 | 0.7611 | 0 | 0 | 0 | 1 | Ref |
| 2858 | KT264793 | Unassigned | Unassigned | Unassigned | 1127_0 | Clustered | 19 | VC_1127_0 | 19 | 0.7611 | 1 | 0.7611 | 0 | 0 | 0 | 1 | Ref |
| 2859 | KT264794 | Unassigned | Unassigned | Unassigned | 1128_0 | Clustered | 2 | VC_1128_0 | 2 | 0.0904 | 0.95143313 | 0.086 | 0 | 0 | 0 | 1 | Ref |
| 2860 | KT264795 | Unassigned | Unassigned | Unassigned | 1132_0 | Clustered | 2 | VC_1132_0 | 2 | 0.0978 | 0.83194774 | 0.0814 | 0 | 0 | 0 | 1 | Ref |
| 2861 | KT264796 | Unassigned | Unassigned | Unassigned | 1125_0 | Clustered | 20 | VC_1125_0 | 20 | 0.5599 | 1 | 0.5599 | 0 | 0 | 0 | 1 | Ref |
| 2862 | KT264797 | Unassigned | Unassigned | Unassigned | 1125_0 | Clustered | 20 | VC_1125_0 | 20 | 0.5599 | 1 | 0.5599 | 0 | 0 | 0 | 1 | Ref |
| 2863 | KT264798 | Unassigned | Unassigned | Unassigned | 1124_0 | Clustered | 10 | VC_1124_0 | 10 | 0.6268 | 1 | 0.6268 | 0 | 0 | 0 | 1 | Ref |
| 2864 | KT264799 | Unassigned | Unassigned | Unassigned | 1124_0 | Clustered | 10 | VC_1124_0 | 10 | 0.6268 | 1 | 0.6268 | 0 | 0 | 0 | 1 | Ref |
| 2865 | KT264800 | Unassigned | Unassigned | Unassigned | 1124_0 | Clustered | 10 | VC_1124_0 | 10 | 0.6268 | 1 | 0.6268 | 0 | 0 | 0 | 1 | Ref |
| 2866 | KT264801 | Unassigned | Unassigned | Unassigned | 1124_0 | Clustered | 10 | VC_1124_0 | 10 | 0.6268 | 1 | 0.6268 | 0 | 0 | 0 | 1 | Ref |
| 2867 | KT264802 | Unassigned | Unassigned | Unassigned | 1137_0 | Clustered | 3 | VC_1137_0 | 3 | 0.9176 | 0.93067968 | 0.854 | 0 | 0 | 0 | 1 | Ref |
| 2868 | KT264803 | Unassigned | Unassigned | Unassigned | 1136_0 | Clustered | 2 | VC_1136_0 | 2 | 0.58 | 0.88261817 | 0.5119 | 0 | 0 | 0 | 1 | Ref |
| 2869 | KT264805 | Unassigned | Unassigned | Unassigned | 1138_0 | Clustered | 4 | VC_1138_0 | 4 | 0.9437 | 0.98964986 | 0.934 | 0 | 0 | 0 | 1 | Ref |
| 2870 | KT264806 | Unassigned | Unassigned | Unassigned | 1139_0 | Clustered | 4 | VC_1139_0 | 4 | 1 | 1 | 1 | 0 | 0 | 0 | 1 | Ref |
| 2871 | KT264807 | Unassigned | Unassigned | Unassigned | 1056_0 | Clustered | 4 | VC_1056_0 | 4 | 0.169 | 0.84110627 | 0.1421 | 0 | 0 | 0 | 1 | Ref |
| 2872 | KT264808 | Unassigned | Unassigned | Unassigned | 1066_0 | Clustered | 52 | VC_1066_0 | 52 | 0.7682 | 1 | 0.7682 | 0 | 0 | 0 | 1 | Ref |
| 2873 | KT264809 | Unassigned | Unassigned | Unassigned | 1066_0 | Clustered | 52 | VC_1066_0 | 52 | 0.7682 | 1 | 0.7682 | 0 | 0 | 0 | 1 | Ref |
| 2874 | KT264810 | Unassigned | Unassigned | Unassigned | 1066_0 | Clustered | 52 | VC_1066_0 | 52 | 0.7682 | 1 | 0.7682 | 0 | 0 | 0 | 1 | Ref |
| 2875 | KT264811 | Unassigned | Unassigned | Unassigned | 398_0 | Clustered | 48 | VC_398_0 | 48 | 0.4813 | 1 | 0.4813 | 0 | 0 | 0 | 1 | Ref |
| 2876 | KT264812 | Unassigned | Unassigned | Unassigned | 1085_0 | Clustered | 2 | VC_1085_0 | 2 | 0.5907 | 0.9331928 | 0.5513 | 0 | 0 | 0 | 1 | Ref |
| 2877 | KT264813 | Unassigned | Unassigned | Unassigned | 995_0 | Clustered | 69 | VC_995_0 | 69 | 0.8547 | 1 | 0.8547 | 0 | 0 | 0 | 1 | Ref |
| 2878 | KT264814 | Unassigned | Unassigned | Unassigned | 995_0 | Clustered | 69 | VC_995_0 | 69 | 0.8547 | 1 | 0.8547 | 0 | 0 | 0 | 1 | Ref |
| 2879 | KT264815 | Unassigned | Unassigned | Unassigned | 995_0 | Clustered | 69 | VC_995_0 | 69 | 0.8547 | 1 | 0.8547 | 0 | 0 | 0 | 1 | Ref |
| 2880 | KT264818 | Unassigned | Unassigned | Unassigned | 1143_0 | Clustered | 9 | VC_1143_0 | 9 | 0.7268 | 1 | 0.7268 | 0 | 0 | 0 | 1 | Ref |
| 2881 | KT264819 | Unassigned | Unassigned | Unassigned | 1143_0 | Clustered | 9 | VC_1143_0 | 9 | 0.7268 | 1 | 0.7268 | 0 | 0 | 0 | 1 | Ref |
| 2882 | KT264820 | Unassigned | Unassigned | Unassigned | 1140_0 | Clustered | 4 | VC_1140_0 | 4 | 0.1325 | 0.66207199 | 0.0877 | 0 | 0 | 0 | 1 | Ref |
| 2883 | KT264821 | Unassigned | Unassigned | Unassigned | 1140_0 | Clustered | 4 | VC_1140_0 | 4 | 0.1325 | 0.66207199 | 0.0877 | 0 | 0 | 0 | 1 | Ref |
| 2884 | KT264822 | Unassigned | Unassigned | Unassigned | 1147_0 | Clustered | 3 | VC_1147_0 | 3 | 0.4334 | 0.65335909 | 0.2832 | 0 | 0 | 0 | 1 | Ref |
| 2885 | KT264823 | Unassigned | Unassigned | Unassigned | 1148_0 | Clustered | 16 | VC_1148_0 | 16 | 0.8821 | 1 | 0.8821 | 0 | 0 | 0 | 1 | Ref |
| 2886 | KT264824 | Unassigned | Unassigned | Unassigned | 1148_0 | Clustered | 16 | VC_1148_0 | 16 | 0.8821 | 1 | 0.8821 | 0 | 0 | 0 | 1 | Ref |
| 2887 | KT264825 | Unassigned | Unassigned | Unassigned | 1148_0 | Clustered | 16 | VC_1148_0 | 16 | 0.8821 | 1 | 0.8821 | 0 | 0 | 0 | 1 | Ref |
| 2888 | KT264826 | Unassigned | Unassigned | Unassigned | 1054_0 | Clustered | 12 | VC_1054_0 | 12 | 0.2863 | 0.99746644 | 0.2855 | 0 | 0 | 0 | 1 | Ref |
| 2889 | KT264827 | Unassigned | Unassigned | Unassigned | 1054_0 | Clustered | 12 | VC_1054_0 | 12 | 0.2863 | 0.99746644 | 0.2855 | 0 | 0 | 0 | 1 | Ref |
| 2890 | KT264828 | Unassigned | Unassigned | Unassigned | 1151_0 | Clustered | 9 | VC_1151_0 | 9 | 0.8548 | 1 | 0.8548 | 0 | 0 | 0 | 1 | Ref |
| 2891 | KT264829 | Unassigned | Unassigned | Unassigned | 1056_0 | Clustered | 4 | VC_1056_0 | 4 | 0.169 | 0.84110627 | 0.1421 | 0 | 0 | 0 | 1 | Ref |
| 2892 | KT264830 | Unassigned | Unassigned | Unassigned | 1147_0 | Clustered | 3 | VC_1147_0 | 3 | 0.4334 | 0.65335909 | 0.2832 | 0 | 0 | 0 | 1 | Ref |
| 2893 | KT264831 | Unassigned | Unassigned | Unassigned | 1153_0 | Clustered | 2 | VC_1153_0 | 2 | 0.073 | 0.80562642 | 0.0588 | 0 | 0 | 0 | 1 | Ref |
| 2894 | KT264833 | Unassigned | Unassigned | Unassigned | 1154_0 | Clustered | 5 | VC_1154_0 | 5 | 1 | 1 | 1 | 0 | 0 | 0 | 1 | Ref |
| 2895 | KT264834 | Unassigned | Unassigned | Unassigned | 1155_0 | Clustered | 2 | VC_1155_0 | 2 | 1 | 1 | 1 | 0 | 0 | 0 | 1 | Ref |
| 2896 | KT270441 | Unassigned | Unassigned | Unassigned | 372_0 | Clustered | 24 | VC_372_0 | 24 | 0.5539 | 1 | 0.5539 | 0 | 0 | 0 | 1 | Ref |
| 2897 | KT279576 | Unassigned | Unassigned | Unassigned | 520_60 | Clustered | 2 | VC_520_60 | 2 | 0.0091 | 0.95761233 | 0.0087 | 0 | 0 | 0 | 1 | Ref |
| 2898 | KT281790 | Unassigned | Unassigned | Unassigned | 558_11 | Clustered | 2 | VC_558_11 | 2 | 0.0032 | 0.78528587 | 0.0025 | 0 | 0 | 0 | 1 | Ref |
| 2899 | KT281791 | Unassigned | Unassigned | Unassigned | 889_0 | Clustered | 16 | VC_889_0 | 16 | 0.2969 | 1 | 0.2969 | 0 | 0 | 0 | 1 | Ref |
| 2900 | KT281792 | Unassigned | Unassigned | Unassigned | 520_27 | Clustered | 2 | VC_520_27 | 2 | 0.0143 | 0.95761635 | 0.0137 | 0 | 0 | 0 | 1 | Ref |
| 2901 | KT281793 | Unassigned | Unassigned | Unassigned | 520_13 | Clustered/Singleton | 1 | VC_520_13 | 1 | 0 | 1 | 0 | 0 | 0 | 0 | 1 | Ref |
| 2902 | KT281794 | Unassigned | Unassigned | Unassigned | 889_0 | Clustered | 16 | VC_889_0 | 16 | 0.2969 | 1 | 0.2969 | 0 | 0 | 0 | 1 | Ref |
| 2903 | KT281795 | Unassigned | Unassigned | Unassigned | 520_31 | Clustered | 11 | VC_520_31 | 11 | 0.0897 | 1 | 0.0897 | 0 | 0 | 0 | 0.6 | Ref |
| 2904 | KT281796 | Unassigned | Unassigned | Unassigned | 556_0 | Clustered | 21 | VC_556_0 | 21 | 0.9912 | 1 | 0.9912 | 0 | 0 | 0 | 1 | Ref |
| 2905 | KT285706 | Unassigned | Unassigned | Unassigned | 225_0 | Clustered | 32 | VC_225_0 | 32 | 0.848 | 1 | 0.848 | 0 | 0 | 0 | 0.996 | Ref |
| 2906 | KT287080 | Unassigned | Unassigned | Unassigned | 540_0 | Clustered | 15 | VC_540_0 | 15 | 0.2866 | 1 | 0.2866 | 0 | 0 | 0 | 1 | Ref |
| 2907 | KT307976 | Unassigned | Unassigned | Unassigned | 883_1 | Clustered | 12 | VC_883_1 | 12 | 0.2441 | 1 | 0.2441 | 0 | 0 | 0 | 1 | Ref |
| 2908 | KT309034 | Unassigned | Unassigned | Unassigned | 520_0 | Clustered/Singleton | 1 | VC_520_0 | 1 | 0 | 1 | 0 | 0 | 0 | 0 | 1 | Ref |
| 2909 | KT321314 | Unassigned | Unassigned | Unassigned | 388_0 | Clustered | 4 | VC_388_0 | 4 | 0.0345 | 0.99998812 | 0.0345 | 0 | 0 | 0 | 1 | Ref |
| 2910 | KT321315 | Unassigned | Unassigned | Unassigned | 668_0 | Clustered | 25 | VC_668_0 | 25 | 0.5539 | 1 | 0.5539 | 0 | 0 | 0 | 0.96 | Ref |
| 2911 | KT321316 | Unassigned | Unassigned | Unassigned | 594_0 | Clustered | 2 | VC_594_0 | 2 | 0.0737 | 0.95685859 | 0.0705 | 0 | 0 | 0 | 1 | Ref |
| 2912 | KT321317 | Unassigned | Unassigned | Unassigned | 779_1 | Clustered | 11 | VC_779_1 | 11 | 0.1587 | 1 | 0.1587 | 0 | 0 | 0 | 1 | Ref |
| 2913 | KT321476 | Unassigned | Unassigned | Unassigned | 558_50 | Clustered | 15 | VC_558_50 | 15 | 0.0468 | 1 | 0.0468 | 0 | 0 | 0 | 0.9238 | Ref |
| 2914 | KT326767 | Unassigned | Unassigned | Unassigned | 513_0 | Clustered | 32 | VC_513_0 | 32 | 0.6881 | 1 | 0.6881 | 0 | 0 | 0 | 1 | Ref |

| | | | | | | | | | | | | | | | | | |
|---|---|---|---|---|---|---|---|---|---|---|---|---|---|---|---|---|---|
| 2915 | KT326768 | Unassigned | Unassigned | Unassigned | 325_30 | Clustered | 4 | VC_325_30 | 4 | 0.0144 | 0.99998616 | 0.0144 | 0 | 0 | 0 | 0.8333 | Ref |
| 2916 | KT336320 | Unassigned | Unassigned | Unassigned | 632_0 | Clustered | 37 | VC_632_0 | 37 | 0.581 | 1 | 0.581 | 0 | 0 | 0 | 0.9234 | Ref |
| 2917 | KT336321 | Unassigned | Unassigned | Unassigned | 632_0 | Clustered | 37 | VC_632_0 | 37 | 0.581 | 1 | 0.581 | 0 | 0 | 0 | 0.9234 | Ref |
| 2918 | KT337339 | Unassigned | Unassigned | Unassigned | 414_0 | Clustered | 37 | VC_414_0 | 37 | 0.585 | 1 | 0.585 | 0 | 0 | 0 | 0.9474 | Ref |
| 2919 | KT337340 | Unassigned | Unassigned | Unassigned | 414_0 | Clustered | 37 | VC_414_0 | 37 | 0.585 | 1 | 0.585 | 0 | 0 | 0 | 0.9474 | Ref |
| 2920 | KT337341 | Unassigned | Unassigned | Unassigned | 414_0 | Clustered | 37 | VC_414_0 | 37 | 0.585 | 1 | 0.585 | 0 | 0 | 0 | 0.9474 | Ref |
| 2921 | KT337342 | Unassigned | Unassigned | Unassigned | 333_0 | Clustered | 25 | VC_333_0 | 25 | 0.4681 | 1 | 0.4681 | 0 | 0 | 0 | 1 | Ref |
| 2922 | KT337352 | Unassigned | Unassigned | Unassigned | 414_0 | Clustered | 37 | VC_414_0 | 37 | 0.585 | 1 | 0.585 | 0 | 0 | 0 | 0.9474 | Ref |
| 2923 | KT337354 | Unassigned | Unassigned | Unassigned | 414_0 | Clustered | 37 | VC_414_0 | 37 | 0.585 | 1 | 0.585 | 0 | 0 | 0 | 0.9474 | Ref |
| 2924 | KT337355 | Unassigned | Unassigned | Unassigned | 414_0 | Clustered | 37 | VC_414_0 | 37 | 0.585 | 1 | 0.585 | 0 | 0 | 0 | 0.9474 | Ref |
| 2925 | KT337356 | Unassigned | Unassigned | Unassigned | 414_0 | Clustered | 37 | VC_414_0 | 37 | 0.585 | 1 | 0.585 | 0 | 0 | 0 | 0.9474 | Ref |
| 2926 | KT337358 | Unassigned | Unassigned | Unassigned | 412_0 | Clustered | 39 | VC_412_0 | 39 | 0.845 | 1 | 0.845 | 0 | 0 | 0 | 1 | Ref |
| 2927 | KT337359 | Unassigned | Unassigned | Unassigned | 414_0 | Clustered | 37 | VC_414_0 | 37 | 0.585 | 1 | 0.585 | 0 | 0 | 0 | 0.9474 | Ref |
| 2928 | KT337360 | Unassigned | Unassigned | Unassigned | 317_2 | Clustered | 20 | VC_317_2 | 20 | 0.1976 | 1 | 0.1976 | 0 | 0 | 0 | 0.6842 | Ref |
| 2929 | KT337362 | Unassigned | Unassigned | Unassigned | 414_0 | Clustered | 37 | VC_414_0 | 37 | 0.585 | 1 | 0.585 | 0 | 0 | 0 | 0.9474 | Ref |
| 2930 | KT337364 | Unassigned | Unassigned | Unassigned | 412_0 | Clustered | 39 | VC_412_0 | 39 | 0.845 | 1 | 0.845 | 0 | 0 | 0 | 1 | Ref |
| 2931 | KT339321 | Unassigned | Unassigned | Unassigned | 699_0 | Clustered | 71 | VC_699_0 | 71 | 0.9352 | 1 | 0.9352 | 0 | 0 | 0 | 1 | Ref |
| 2932 | KT344895 | Unassigned | Unassigned | Unassigned | 12_0 | Clustered | 21 | VC_12_0 | 21 | 0.2274 | 1 | 0.2274 | 0 | 0 | 0 | 0.9571 | Ref |
| 2933 | KT345706 | Unassigned | Unassigned | Unassigned | 971_2 | Clustered/Singleton | 1 | VC_971_2 | 1 | 0 | 1 | 0 | 0 | 0 | 0 | 1 | Ref |
| 2934 | KT347313 | Unassigned | Unassigned | Unassigned | 519_0 | Clustered | 39 | VC_519_0 | 39 | 0.8156 | 1 | 0.8156 | 0 | 0 | 0 | 1 | Ref |
| 2935 | KT347314 | Unassigned | Unassigned | Unassigned | 761_0 | Clustered | 67 | VC_761_0 | 67 | 0.8947 | 1 | 0.8947 | 0 | 0 | 0 | 1 | Ref |
| 2936 | KT353109 | Unassigned | Unassigned | Unassigned | 849_5 | Clustered | 3 | VC_849_5 | 3 | 0.0492 | 0.97574727 | 0.048 | 0 | 0 | 0 | 1 | Ref |
| 2937 | KT355471 | Unassigned | Unassigned | Unassigned | 950_0 | Clustered | 38 | VC_950_0 | 38 | 0.9443 | 1 | 0.9443 | 0 | 0 | 0 | 1 | Ref |
| 2938 | KT355472 | Unassigned | Unassigned | Unassigned | 761_0 | Clustered | 67 | VC_761_0 | 67 | 0.8947 | 1 | 0.8947 | 0 | 0 | 0 | 1 | Ref |
| 2939 | KT355473 | Unassigned | Unassigned | Unassigned | 950_0 | Clustered | 38 | VC_950_0 | 38 | 0.9443 | 1 | 0.9443 | 0 | 0 | 0 | 1 | Ref |
| 2940 | KT355474 | Unassigned | Unassigned | Unassigned | 761_0 | Clustered | 67 | VC_761_0 | 67 | 0.8947 | 1 | 0.8947 | 0 | 0 | 0 | 1 | Ref |
| 2941 | KT355475 | Unassigned | Unassigned | Unassigned | 950_0 | Clustered | 38 | VC_950_0 | 38 | 0.9443 | 1 | 0.9443 | 0 | 0 | 0 | 1 | Ref |
| 2942 | KT359365 | Unassigned | Unassigned | Unassigned | 514_0 | Clustered | 75 | VC_514_0 | 75 | 0.9402 | 1 | 0.9402 | 0 | 0 | 0 | 1 | Ref |
| 2943 | KT361649 | Unassigned | Unassigned | Unassigned | 708_0 | Clustered | 47 | VC_708_0 | 47 | 0.8654 | 1 | 0.8654 | 0 | 0 | 0 | 1 | Ref |
| 2944 | KT361650 | Unassigned | Unassigned | Unassigned | 708_0 | Clustered | 47 | VC_708_0 | 47 | 0.8654 | 1 | 0.8654 | 0 | 0 | 0 | 1 | Ref |
| 2945 | KT361652 | Unassigned | Unassigned | Unassigned | 708_0 | Clustered | 47 | VC_708_0 | 47 | 0.8654 | 1 | 0.8654 | 0 | 0 | 0 | 1 | Ref |
| 2946 | KT361654 | Unassigned | Unassigned | Unassigned | 620_0 | Clustered | 27 | VC_620_0 | 27 | 0.879 | 1 | 0.879 | 0 | 0 | 0 | 1 | Ref |
| 2947 | KT361655 | Unassigned | Unassigned | Unassigned | 620_0 | Clustered | 27 | VC_620_0 | 27 | 0.879 | 1 | 0.879 | 0 | 0 | 0 | 1 | Ref |
| 2948 | KT361656 | Unassigned | Unassigned | Unassigned | 620_0 | Clustered | 27 | VC_620_0 | 27 | 0.879 | 1 | 0.879 | 0 | 0 | 0 | 1 | Ref |
| 2949 | KT361657 | Unassigned | Unassigned | Unassigned | 620_0 | Clustered | 27 | VC_620_0 | 27 | 0.879 | 1 | 0.879 | 0 | 0 | 0 | 1 | Ref |
| 2950 | KT361920 | Unassigned | Unassigned | Unassigned | 234_0 | Clustered | 21 | VC_234_0 | 21 | 0.8355 | 1 | 0.8355 | 0 | 0 | 0 | 1 | Ref |
| 2951 | KT363731 | Unassigned | Unassigned | Unassigned | 325_42 | Clustered | 4 | VC_325_42 | 4 | 0.0112 | 0.99980442 | 0.0112 | 0 | 0 | 0 | 0.6667 | Ref |
| 2952 | KT363872 | Unassigned | Unassigned | Unassigned | 330_0 | Clustered | 27 | VC_330_0 | 27 | 0.5088 | 1 | 0.5088 | 0 | 0 | 0 | 0.8661 | Ref |
| 2953 | KT364588 | Unassigned | Unassigned | Unassigned | 371_4 | Clustered | 5 | VC_371_4 | 5 | 0.0135 | 0.99999971 | 0.0135 | 0 | 0 | 0 | 1 | Ref |
| 2954 | KT365397 | Unassigned | Unassigned | Unassigned | 517_1 | Clustered | 108 | VC_517_1 | 108 | 0.8483 | 1 | 0.8483 | 0 | 0 | 0 | 0.9955 | Ref |
| 2955 | KT365398 | Unassigned | Unassigned | Unassigned | 558_6 | Clustered | 2 | VC_558_6 | 2 | 0.0032 | 0.73149279 | 0.0023 | 0 | 0 | 0 | 1 | Ref |
| 2956 | KT365399 | Unassigned | Unassigned | Unassigned | 761_0 | Clustered | 67 | VC_761_0 | 67 | 0.8947 | 1 | 0.8947 | 0 | 0 | 0 | 1 | Ref |
| 2957 | KT365400 | Unassigned | Unassigned | Unassigned | 558_37 | Clustered/Singleton | 1 | VC_558_37 | 1 | 0 | 1 | 0 | 0 | 0 | 0 | 1 | Ref |
| 2958 | KT365401 | Unassigned | Unassigned | Unassigned | 1073_0 | Clustered | 24 | VC_1073_0 | 24 | 0.8388 | 1 | 0.8388 | 0 | 0 | 0 | 1 | Ref |
| 2959 | KT365402 | Unassigned | Unassigned | Unassigned | 508_0 | Clustered | 32 | VC_508_0 | 32 | 0.906 | 1 | 0.906 | 0 | 0 | 0 | 1 | Ref |
| 2960 | KT367885 | Unassigned | Unassigned | Unassigned | 823_0 | Clustered | 124 | VC_823_0 | 124 | 0.9492 | 1 | 0.9492 | 0 | 0 | 0 | 1 | Ref |
| 2961 | KT367886 | Unassigned | Unassigned | Unassigned | 111_0 | Clustered | 91 | VC_111_0 | 91 | 0.9587 | 1 | 0.9587 | 0 | 0 | 0 | 1 | Ref |
| 2962 | KT367887 | Unassigned | Unassigned | Unassigned | 836_0 | Clustered | 9 | VC_836_0 | 9 | 0.7733 | 1 | 0.7733 | 0 | 0 | 0 | 1 | Ref |
| 2963 | KT372002 | Unassigned | Unassigned | Unassigned | 956_0 | Clustered | 39 | VC_956_0 | 39 | 1 | 1 | 1 | 0 | 0 | 0 | 1 | Ref |
| 2964 | KT372003 | Unassigned | Unassigned | Unassigned | 889_0 | Clustered | 16 | VC_889_0 | 16 | 0.2969 | 1 | 0.2969 | 0 | 0 | 0 | 1 | Ref |
| 2965 | KT372690 | Unassigned | Unassigned | Unassigned | 80_0 | Clustered | 41 | VC_80_0 | 41 | 0.5837 | 1 | 0.5837 | 0 | 0 | 0 | 0.9805 | Ref |
| 2966 | KT372691 | Unassigned | Unassigned | Unassigned | 80_0 | Clustered | 41 | VC_80_0 | 41 | 0.5837 | 1 | 0.5837 | 0 | 0 | 0 | 0.9805 | Ref |
| 2967 | KT372692 | Unassigned | Unassigned | Unassigned | 80_0 | Clustered | 41 | VC_80_0 | 41 | 0.5837 | 1 | 0.5837 | 0 | 0 | 0 | 0.9805 | Ref |
| 2968 | KT372693 | Unassigned | Unassigned | Unassigned | 80_0 | Clustered | 41 | VC_80_0 | 41 | 0.5837 | 1 | 0.5837 | 0 | 0 | 0 | 0.9805 | Ref |
| 2969 | KT372694 | Unassigned | Unassigned | Unassigned | 80_0 | Clustered | 41 | VC_80_0 | 41 | 0.5837 | 1 | 0.5837 | 0 | 0 | 0 | 0.9805 | Ref |
| 2970 | KT372695 | Unassigned | Unassigned | Unassigned | 80_0 | Clustered | 41 | VC_80_0 | 41 | 0.5837 | 1 | 0.5837 | 0 | 0 | 0 | 0.9805 | Ref |
| 2971 | KT372696 | Unassigned | Unassigned | Unassigned | 80_0 | Clustered | 41 | VC_80_0 | 41 | 0.5837 | 1 | 0.5837 | 0 | 0 | 0 | 0.9805 | Ref |
| 2972 | KT372697 | Unassigned | Unassigned | Unassigned | 80_0 | Clustered | 41 | VC_80_0 | 41 | 0.5837 | 1 | 0.5837 | 0 | 0 | 0 | 0.9805 | Ref |
| 2973 | KT372698 | Unassigned | Unassigned | Unassigned | 80_0 | Clustered | 41 | VC_80_0 | 41 | 0.5837 | 1 | 0.5837 | 0 | 0 | 0 | 0.9805 | Ref |
| 2974 | KT372714 | Unassigned | Unassigned | Unassigned | 20_0 | Clustered | 5 | VC_20_0 | 5 | 0.4387 | 0.99999934 | 0.4387 | 0 | 0 | 0 | 1 | Ref |
| 2975 | KT373978 | Unassigned | Unassigned | Unassigned | 551_7 | Clustered | 28 | VC_551_7 | 28 | 0.1661 | 1 | 0.1661 | 0 | 0 | 0 | 0.8519 | Ref |
| 2976 | KT375356 | Unassigned | Unassigned | Unassigned | 956_0 | Clustered | 39 | VC_956_0 | 39 | 1 | 1 | 1 | 0 | 0 | 0 | 1 | Ref |
| 2977 | KT381276 | Unassigned | Unassigned | Unassigned | 215_2 | Clustered | 5 | VC_215_2 | 5 | 0.0572 | 0.99999997 | 0.0572 | 0 | 0 | 0 | 1 | Ref |
| 2978 | KT381277 | Unassigned | Unassigned | Unassigned | 514_0 | Clustered | 75 | VC_514_0 | 75 | 0.9402 | 1 | 0.9402 | 0 | 0 | 0 | 1 | Ref |
| 2979 | KT381864 | Unassigned | Unassigned | Unassigned | 182_0 | Clustered | 5 | VC_182_0 | 5 | 0.6184 | 0.99999967 | 0.6184 | 0 | 0 | 0 | 1 | Ref |
| 2980 | KT381865 | Unassigned | Unassigned | Unassigned | 182_0 | Clustered | 5 | VC_182_0 | 5 | 0.6184 | 0.99999967 | 0.6184 | 0 | 0 | 0 | 1 | Ref |
| 2981 | KT381879 | Unassigned | Unassigned | Unassigned | 70_0 | Clustered | 27 | VC_70_0 | 27 | 0.5981 | 1 | 0.5981 | 0 | 0 | 0 | 0.9886 | Ref |
| 2982 | KT381880 | Unassigned | Unassigned | Unassigned | 669_0 | Clustered | 21 | VC_669_0 | 21 | 0.3623 | 1 | 0.3623 | 0 | 0 | 0 | 1 | Ref |
| 2983 | KT388014 | Unassigned | Unassigned | Unassigned | 325_53 | Clustered | 7 | VC_325_53 | 7 | 0.0242 | 1 | 0.0242 | 0 | 0 | 0 | 0.9048 | Ref |
| 2984 | KT388093 | Unassigned | Unassigned | Unassigned | 412_0 | Clustered | 39 | VC_412_0 | 39 | 0.845 | 1 | 0.845 | 0 | 0 | 0 | 1 | Ref |
| 2985 | KT388102 | Unassigned | Unassigned | Unassigned | 699_0 | Clustered | 71 | VC_699_0 | 71 | 0.9352 | 1 | 0.9352 | 0 | 0 | 0 | 1 | Ref |
| 2986 | KT388103 | Unassigned | Unassigned | Unassigned | 699_0 | Clustered | 71 | VC_699_0 | 71 | 0.9352 | 1 | 0.9352 | 0 | 0 | 0 | 1 | Ref |
| 2987 | KT429160 | Unassigned | Unassigned | Unassigned | 213_0 | Clustered | 2 | VC_213_0 | 2 | 0.4293 | 0.95300227 | 0.4091 | 0 | 0 | 0 | 1 | Ref |
| 2988 | KT438500 | Unassigned | Unassigned | Unassigned | 325_9 | Clustered | 2 | VC_325_9 | 2 | 0.0055 | 0.95750934 | 0.0053 | 0 | 0 | 0 | 1 | Ref |
| 2989 | KT438501 | Unassigned | Unassigned | Unassigned | 940_0 | Clustered | 34 | VC_940_0 | 34 | 0.7959 | 1 | 0.7959 | 0 | 0 | 0 | 1 | Ref |
| 2990 | KT454805 | Unassigned | Unassigned | Unassigned | 330_0 | Clustered | 27 | VC_330_0 | 27 | 0.5088 | 1 | 0.5088 | 0 | 0 | 0 | 0.8661 | Ref |
| 2991 | KT454806 | Unassigned | Unassigned | Unassigned | 330_0 | Clustered | 27 | VC_330_0 | 27 | 0.5088 | 1 | 0.5088 | 0 | 0 | 0 | 0.8661 | Ref |
| 2992 | KT454972 | Unassigned | Unassigned | Unassigned | 810_0 | Clustered | 18 | VC_810_0 | 18 | 0.9121 | 1 | 0.9121 | 0 | 0 | 0 | 1 | Ref |
| 2993 | KT588073 | Unassigned | Unassigned | Unassigned | 696_1 | Clustered | 8 | VC_696_1 | 8 | 0.5452 | 1 | 0.5452 | 0 | 0 | 0 | 0.9286 | Ref |
| 2994 | KT588074 | Unassigned | Unassigned | Unassigned | 242_0 | Clustered | 2 | VC_242_0 | 2 | 0.0867 | 0.95379254 | 0.0827 | 0 | 0 | 0 | 1 | Ref |
| 2995 | KT588075 | Unassigned | Unassigned | Unassigned | 696_0 | Clustered | 2 | VC_696_0 | 2 | 0.1624 | 0.94573679 | 0.1536 | 0 | 0 | 0 | 1 | Ref |
| 2996 | KT588442 | Unassigned | Unassigned | Unassigned | 216_6 | Clustered | 42 | VC_216_6 | 42 | 0.473 | 1 | 0.473 | 0 | 0 | 0 | 0.8351 | Ref |
| 2997 | KT591076 | Unassigned | Unassigned | Unassigned | 568_0 | Clustered | 8 | VC_568_0 | 8 | 0.5738 | 1 | 0.5738 | 0 | 0 | 0 | 1 | Ref |
| 2998 | KT591489 | Unassigned | Unassigned | Unassigned | 888_0 | Clustered | 28 | VC_888_0 | 28 | 0.5497 | 1 | 0.5497 | 0 | 0 | 0 | 1 | Ref |
| 2999 | KT591491 | Unassigned | Unassigned | Unassigned | 763_1 | Clustered | 9 | VC_763_1 | 9 | 0.3706 | 1 | 0.3706 | 0 | 0 | 0 | 1 | Ref |
| 3000 | KT599441 | Unassigned | Unassigned | Unassigned | 371_15 | Clustered | 44 | VC_371_15 | 44 | 0.1391 | 1 | 0.1391 | 0 | 0 | 0 | 0.7474 | Ref |
| 3001 | KT624200 | Unassigned | Unassigned | Unassigned | 88_0 | Clustered/Singleton | 1 | VC_88_0 | 1 | 0 | 1 | 0 | 0 | 0 | 0 | 1 | Ref |
| 3002 | KT626047 | Unassigned | Unassigned | Unassigned | 325_5 | Clustered | 11 | VC_325_5 | 11 | 0.047 | 1 | 0.047 | 0 | 0 | 0 | 0.7636 | Ref |
| 3003 | KT626446 | Unassigned | Unassigned | Unassigned | 143_0 | Clustered | 2 | VC_143_0 | 2 | 0.1189 | 0.94995671 | 0.1129 | 0 | 0 | 0 | 1 | Ref |
| 3004 | KT630644 | Unassigned | Unassigned | Unassigned | 228_0 | Clustered | 60 | VC_228_0 | 60 | 0.942 | 1 | 0.942 | 0 | 0 | 0 | 1 | Ref |
| 3005 | KT630647 | Unassigned | Unassigned | Unassigned | 6_0 | Clustered | 44 | VC_6_0 | 44 | 0.908 | 1 | 0.908 | 0 | 0 | 0 | 1 | Ref |
| 3006 | KT630649 | Unassigned | Unassigned | Unassigned | 242_1 | Clustered | 20 | VC_242_1 | 20 | 0.684 | 1 | 0.684 | 0 | 0 | 0 | 0.8368 | Ref |
| 3007 | KT716493 | Unassigned | Unassigned | Unassigned | 517_1 | Clustered | 108 | VC_517_1 | 108 | 0.8483 | 1 | 0.8483 | 0 | 0 | 0 | 0.9955 | Ref |
| 3008 | KT717083 | Unassigned | Unassigned | Unassigned | 252_0 | Clustered | 121 | VC_252_0 | 121 | 0.7376 | 1 | 0.7376 | 0 | 0 | 0 | 0.9999 | Ref |
| 3009 | KT717084 | Unassigned | Unassigned | Unassigned | 252_0 | Clustered | 121 | VC_252_0 | 121 | 0.7376 | 1 | 0.7376 | 0 | 0 | 0 | 0.9999 | Ref |
| 3010 | KT717085 | Unassigned | Unassigned | Unassigned | 252_0 | Clustered | 121 | VC_252_0 | 121 | 0.7376 | 1 | 0.7376 | 0 | 0 | 0 | 0.9999 | Ref |
| 3011 | KT725776 | Unassigned | Unassigned | Unassigned | 654_0 | Clustered | 4 | VC_654_0 | 4 | 0.2897 | 0.99998263 | 0.2897 | 0 | 0 | 0 | 1 | Ref |
| 3012 | KT728930 | Unassigned | Unassigned | Unassigned | 875_0 | Clustered | 10 | VC_875_0 | 10 | 0.988 | 0.99948433 | 0.9875 | 0 | 0 | 0 | 1 | Ref |
| 3013 | KT728931 | Unassigned | Unassigned | Unassigned | 875_0 | Clustered | 10 | VC_875_0 | 10 | 0.988 | 0.99948433 | 0.9875 | 0 | 0 | 0 | 1 | Ref |
| 3014 | KT734862 | Unassigned | Unassigned | Unassigned | 166_0 | Clustered | 26 | VC_166_0 | 26 | 0.7923 | 1 | 0.7923 | 0 | 0 | 0 | 1 | Ref |
| 3015 | KT736033 | Unassigned | Unassigned | Unassigned | 76_0 | Clustered | 37 | VC_76_0 | 37 | 0.9403 | 1 | 0.9403 | 0 | 0 | 0 | 1 | Ref |
| 3016 | KT755656 | Unassigned | Unassigned | Unassigned | 1156_0 | Clustered | 8 | VC_1156_0 | 8 | 0.8137 | 1 | 0.8137 | 0 | 0 | 0 | 1 | Ref |
| 3017 | KT783672 | Unassigned | Unassigned | Unassigned | 950_0 | Clustered | 38 | VC_950_0 | 38 | 0.9443 | 1 | 0.9443 | 0 | 0 | 0 | 1 | Ref |
| 3018 | KT804908 | Unassigned | Unassigned | Unassigned | 699_0 | Clustered | 71 | VC_699_0 | 71 | 0.9352 | 1 | 0.9352 | 0 | 0 | 0 | 1 | Ref |
| 3019 | KT804923 | Unassigned | Unassigned | Unassigned | 76_0 | Clustered | 37 | VC_76_0 | 37 | 0.9403 | 1 | 0.9403 | 0 | 0 | 0 | 1 | Ref |
| 3020 | KT809302 | Unassigned | Unassigned | Unassigned | 666_0 | Clustered | 4 | VC_666_0 | 4 | 1 | 1 | 1 | 0 | 0 | 0 | 1 | Ref |
| 3021 | KT809368 | Unassigned | Unassigned | Unassigned | 10_0 | Clustered | 25 | VC_10_0 | 25 | 0.3467 | 1 | 0.3467 | 0 | 0 | 0 | 0.9733 | Ref |
| 3022 | KT809369 | Unassigned | Unassigned | Unassigned | 10_0 | Clustered | 25 | VC_10_0 | 25 | 0.3467 | 1 | 0.3467 | 0 | 0 | 0 | 0.9733 | Ref |
| 3023 | KT818595 | Unassigned | Unassigned | Unassigned | 325_14 | Clustered | 3 | VC_325_14 | 3 | 0.0094 | 0.99859159 | 0.0094 | 0 | 0 | 0 | 1 | Ref |
| 3024 | KT825490 | Unassigned | Unassigned | Unassigned | 455_0 | Clustered | 18 | VC_455_0 | 18 | 0.4598 | 1 | 0.4598 | 0 | 0 | 0 | 1 | Ref |
| 3025 | KT852578 | Unassigned | Unassigned | Unassigned | 20_0 | Clustered | 5 | VC_20_0 | 5 | 0.4387 | 0.99999934 | 0.4387 | 0 | 0 | 0 | 1 | Ref |
| 3026 | KT870145 | Unassigned | Unassigned | Unassigned | 977_0 | Clustered | 2 | VC_977_0 | 2 | 0.0806 | 0.93920319 | 0.0757 | 0 | 0 | 0 | 1 | Ref |
| 3027 | KT876724 | Unassigned | Unassigned | Unassigned | 1157_0 | Clustered | 24 | VC_1157_0 | 24 | 1 | 1 | 1 | 0 | 0 | 0 | 1 | Ref |
| 3028 | KT876725 | Unassigned | Unassigned | Unassigned | 1040_0 | Clustered | 23 | VC_1040_0 | 23 | 0.9605 | 1 | 0.9605 | 0 | 0 | 0 | 1 | Ref |
| 3029 | KT876726 | Unassigned | Unassigned | Unassigned | 1157_0 | Clustered | 24 | VC_1157_0 | 24 | 1 | 1 | 1 | 0 | 0 | 0 | 1 | Ref |
| 3030 | KT878766 | Unassigned | Unassigned | Unassigned | 10_0 | Clustered | 25 | VC_10_0 | 25 | 0.3467 | 1 | 0.3467 | 0 | 0 | 0 | 0.9733 | Ref |
| 3031 | KT880194 | Unassigned | Unassigned | Unassigned | 508_0 | Clustered | 32 | VC_508_0 | 32 | 0.906 | 1 | 0.906 | 0 | 0 | 0 | 1 | Ref |
| 3032 | KT881477 | Unassigned | Unassigned | Unassigned | 637_0 | Clustered | 115 | VC_637_0 | 115 | 0.731 | 1 | 0.731 | 0 | 0 | 0 | 0.8764 | Ref |
| 3033 | KT887557 | Unassigned | Unassigned | Unassigned | 176_1 | Clustered | 2 | VC_176_1 | 2 | 0.0363 | 0.95657568 | 0.0347 | 0 | 0 | 0 | 1 | Ref |
| 3034 | KT887558 | Unassigned | Unassigned | Unassigned | 173_0 | Clustered | 2 | VC_173_0 | 2 | 0.0799 | 0.95489091 | 0.0763 | 0 | 0 | 0 | 1 | Ref |
| 3035 | KT887559 | Unassigned | Unassigned | Unassigned | 5_0 | Clustered | 30 | VC_5_0 | 30 | 0.9647 | 1 | 0.9647 | 0 | 0 | 0 | 1 | Ref |
| 3036 | KT895280 | Unassigned | Unassigned | Unassigned | 520_85 | Clustered | 2 | VC_520_85 | 2 | 0.0148 | 0.95762405 | 0.0142 | 0 | 0 | 0 | 1 | Ref |
| 3037 | KT895281 | Unassigned | Unassigned | Unassigned | 520_45 | Clustered | 7 | VC_520_45 | 7 | 0.0635 | 1 | 0.0635 | 0 | 0 | 0 | 0.8095 | Ref |
| 3038 | KT895374 | Unassigned | Unassigned | Unassigned | 448_1 | Clustered/Singleton | 1 | VC_448_1 | 1 | 0 | 1 | 0 | 0 | 0 | 0 | 1 | Ref |
| 3039 | KT919972 | Unassigned | Unassigned | Unassigned | 531_0 | Clustered | 9 | VC_531_0 | 9 | 0.26 | 1 | 0.26 | 0 | 0 | 0 | 1 | Ref |
| 3040 | KT919973 | Unassigned | Unassigned | Unassigned | 531_0 | Clustered | 9 | VC_531_0 | 9 | 0.26 | 1 | 0.26 | 0 | 0 | 0 | 1 | Ref |
| 3041 | KT932418 | Unassigned | Unassigned | Unassigned | 493_0 | Clustered/Singleton | 1 | VC_493_0 | 1 | 0 | 1 | 0 | 0 | 0 | 0 | 1 | Ref |
| 3042 | KT932699 | Unassigned | Unassigned | Unassigned | 108_0 | Clustered | 33 | VC_108_0 | 33 | 0.949 | 1 | 0.949 | 0 | 0 | 0 | 1 | Ref |
| 3043 | KT932700 | Unassigned | Unassigned | Unassigned | 967_0 | Clustered | 16 | VC_967_0 | 16 | 0.8971 | 1 | 0.8971 | 0 | 0 | 0 | 1 | Ref |
| 3044 | KT932701 | Unassigned | Unassigned | Unassigned | 695_0 | Clustered | 48 | VC_695_0 | 48 | 0.8162 | 1 | 0.8162 | 0 | 0 | 0 | 1 | Ref |
| 3045 | KT934381 | Unassigned | Unassigned | Unassigned | 765_0 | Clustered | 132 | VC_765_0 | 132 | 1 | 1 | 1 | 0 | 0 | 0 | 1 | Ref |
| 3046 | KT934943 | Unassigned | Unassigned | Unassigned | 582_3 | Clustered | 12 | VC_582_3 | 12 | 0.2357 | 1 | 0.2357 | 0 | 0 | 0 | 0.8636 | Ref |
| 3047 | KT945994 | Unassigned | Unassigned | Unassigned | 267_0 | Clustered | 2 | VC_267_0 | 2 | 0.0988 | 0.95557934 | 0.0945 | 0 | 0 | 0 | 1 | Ref |
| 3048 | KT945995 | Unassigned | Unassigned | Unassigned | 968_0 | Clustered | 4 | VC_968_0 | 4 | 0.2959 | 0.99997642 | 0.2959 | 0 | 0 | 0 | 1 | Ref |
| 3049 | KT949345 | Unassigned | Unassigned | Unassigned | 878_0 | Clustered | 32 | VC_878_0 | 32 | 0.9965 | 1 | 0.9965 | 0 | 0 | 0 | 1 | Ref |
| 3050 | KT959213 | Unassigned | Unassigned | Unassigned | 956_0 | Clustered | 39 | VC_956_0 | 39 | 1 | 1 | 1 | 0 | 0 | 0 | 1 | Ref |
| 3051 | KT962245 | Unassigned | Unassigned | Unassigned | 1017_0 | Clustered | 5 | VC_1017_0 | 5 | 0.9759 | 0.9999961 | 0.9759 | 0 | 0 | 0 | 1 | Ref |
| 3052 | KT962246 | Unassigned | Unassigned | Unassigned | 1017_0 | Clustered | 5 | VC_1017_0 | 5 | 0.9759 | 0.9999961 | 0.9759 | 0 | 0 | 0 | 1 | Ref |
| 3053 | KT962247 | Unassigned | Unassigned | Unassigned | 1017_0 | Clustered | 5 | VC_1017_0 | 5 | 0.9759 | 0.9999961 | 0.9759 | 0 | 0 | 0 | 1 | Ref |
| 3054 | KT962832 | Unassigned | Unassigned | Unassigned | 637_0 | Clustered | 115 | VC_637_0 | 115 | 0.731 | 1 | 0.731 | 0 | 0 | 0 | 0.8764 | Ref |
| 3055 | KT964103 | Unassigned | Unassigned | Unassigned | 111_0 | Clustered | 91 | VC_111_0 | 91 | 0.9587 | 1 | 0.9587 | 0 | 0 | 0 | 1 | Ref |
| 3056 | KT968831 | Unassigned | Unassigned | Unassigned | 176_3 | Clustered/Singleton | 1 | VC_176_3 | 1 | 0 | 1 | 0 | 0 | 0 | 0 | 1 | Ref |
| 3057 | KT968832 | Unassigned | Unassigned | Unassigned | 179_0 | Clustered | 53 | VC_179_0 | 53 | 0.9137 | 1 | 0.9137 | 0 | 0 | 0 | 1 | Ref |
| 3058 | KT989433 | Unassigned | Unassigned | Unassigned | 975_0 | Clustered | 11 | VC_975_0 | 11 | 0.9687 | 1 | 0.9687 | 0 | 0 | 0 | 1 | Ref |
| 3059 | KT990215 | Unassigned | Unassigned | Unassigned | 451_0 | Clustered | 80 | VC_451_0 | 80 | 0.9991 | 1 | 0.9991 | 0 | 0 | 0 | 1 | Ref |
| 3060 | KT990218 | Unassigned | Unassigned | Unassigned | 956_0 | Clustered | 39 | VC_956_0 | 39 | 1 | 1 | 1 | 0 | 0 | 0 | 1 | Ref |
| 3061 | KT995480 | Unassigned | Unassigned | Unassigned | 187_3 | Clustered | 9 | VC_187_3 | 9 | 0.1971 | 1 | 0.1971 | 0 | 0 | 0 | 0.8611 | Ref |
| 3062 | KT997847 | Unassigned | Unassigned | Unassigned | 481_0 | Clustered | 8 | VC_481_0 | 8 | 0.1348 | 1 | 0.1348 | 0 | 0 | 0 | 1 | Ref |
| 3063 | KU052037 | Unassigned | Unassigned | Unassigned | 37_1 | Clustered | 3 | VC_37_1 | 3 | 0.0833 | 0.9484635 | 0.079 | 0 | 0 | 0 | 1 | Ref |
| 3064 | KU052488 | Unassigned | Unassigned | Unassigned | 634_0 | Clustered | 3 | VC_634_0 | 3 | 0.2046 | 0.99837593 | 0.2043 | 0 | 0 | 0 | 1 | Ref |
| 3065 | KU055616 | Unassigned | Unassigned | Unassigned | 517_1 | Clustered | 108 | VC_517_1 | 108 | 0.8483 | 1 | 0.8483 | 0 | 0 | 0 | 0.9955 | Ref |
| 3066 | KU057941 | Unassigned | Unassigned | Unassigned | 733_0 | Clustered | 5 | VC_733_0 | 5 | 0.5111 | 0.99999994 | 0.5111 | 0 | 0 | 0 | 1 | Ref |
| 3067 | KU064779 | Unassigned | Unassigned | Unassigned | 129_0 | Clustered | 43 | VC_129_0 | 43 | 0.886 | 1 | 0.886 | 0 | 0 | 0 | 1 | Ref |
| 3068 | KU130126 | Unassigned | Unassigned | Unassigned | 78_1 | Clustered | 6 | VC_78_1 | 6 | 0.1362 | 1 | 0.1362 | 0 | 0 | 0 | 1 | Ref |
| 3069 | KU130127 | Unassigned | Unassigned | Unassigned | 78_1 | Clustered | 6 | VC_78_1 | 6 | 0.1362 | 1 | 0.1362 | 0 | 0 | 0 | 1 | Ref |
| 3070 | KU130128 | Unassigned | Unassigned | Unassigned | 78_1 | Clustered | 6 | VC_78_1 | 6 | 0.1362 | 1 | 0.1362 | 0 | 0 | 0 | 1 | Ref |
| 3071 | KU130129 | Unassigned | Unassigned | Unassigned | 78_1 | Clustered | 6 | VC_78_1 | 6 | 0.1362 | 1 | 0.1362 | 0 | 0 | 0 | 1 | Ref |
| 3072 | KU130130 | Unassigned | Unassigned | Unassigned | 78_1 | Clustered | 6 | VC_78_1 | 6 | 0.1362 | 1 | 0.1362 | 0 | 0 | 0 | 1 | Ref |
| 3073 | KU130131 | Unassigned | Unassigned | Unassigned | 78_1 | Clustered | 6 | VC_78_1 | 6 | 0.1362 | 1 | 0.1362 | 0 | 0 | 0 | 1 | Ref |
| 3074 | KU160638 | Unassigned | Unassigned | Unassigned | 857_0 | Clustered | 11 | VC_857_0 | 11 | 0.9712 | 1 | 0.9712 | 0 | 0 | 0 | 1 | Ref |
| 3075 | KU160639 | Unassigned | Unassigned | Unassigned | 857_0 | Clustered | 11 | VC_857_0 | 11 | 0.9712 | 1 | 0.9712 | 0 | 0 | 0 | 1 | Ref |
| 3076 | KU160640 | Unassigned | Unassigned | Unassigned | 1160_0 | Clustered | 73 | VC_1160_0 | 73 | 0.9835 | 1 | 0.9835 | 0 | 0 | 0 | 1 | Ref |

| | | | | | | | | | | | | | | | | | |
|---|---|---|---|---|---|---|---|---|---|---|---|---|---|---|---|---|---|
| 3077 | KU160641 | Unassigned | Unassigned | Unassigned | 1164_0 | Clustered | 25 | VC_1164_0 | 25 | 0.7312 | 1 | 0.7312 | 0 | 0 | 0 | 1 | Ref |
| 3078 | KU160642 | Unassigned | Unassigned | Unassigned | 802_0 | Clustered | 17 | VC_802_0 | 17 | 0.8025 | 1 | 0.8025 | 0 | 0 | 0 | 1 | Ref |
| 3079 | KU160643 | Unassigned | Unassigned | Unassigned | 1160_0 | Clustered | 73 | VC_1160_0 | 73 | 0.9835 | 1 | 0.9835 | 0 | 0 | 0 | 1 | Ref |
| 3080 | KU160644 | Unassigned | Unassigned | Unassigned | 1166_0 | Clustered | 20 | VC_1166_0 | 20 | 0.99 | 1 | 0.99 | 0 | 0 | 0 | 1 | Ref |
| 3081 | KU160645 | Unassigned | Unassigned | Unassigned | 1160_0 | Clustered | 73 | VC_1160_0 | 73 | 0.9835 | 1 | 0.9835 | 0 | 0 | 0 | 1 | Ref |
| 3082 | KU160646 | Unassigned | Unassigned | Unassigned | 1164_0 | Clustered | 25 | VC_1164_0 | 25 | 0.7312 | 1 | 0.7312 | 0 | 0 | 0 | 1 | Ref |
| 3083 | KU160647 | Unassigned | Unassigned | Unassigned | 857_0 | Clustered | 11 | VC_857_0 | 11 | 0.9712 | 1 | 0.9712 | 0 | 0 | 0 | 1 | Ref |
| 3084 | KU160648 | Unassigned | Unassigned | Unassigned | 1160_0 | Clustered | 73 | VC_1160_0 | 73 | 0.9835 | 1 | 0.9835 | 0 | 0 | 0 | 1 | Ref |
| 3085 | KU160649 | Unassigned | Unassigned | Unassigned | 1160_0 | Clustered | 73 | VC_1160_0 | 73 | 0.9835 | 1 | 0.9835 | 0 | 0 | 0 | 1 | Ref |
| 3086 | KU160650 | Unassigned | Unassigned | Unassigned | 1167_0 | Clustered | 6 | VC_1167_0 | 6 | 0.8642 | 0.99999946 | 0.8642 | 0 | 0 | 0 | 1 | Ref |
| 3087 | KU160651 | Unassigned | Unassigned | Unassigned | 1073_0 | Clustered | 24 | VC_1073_0 | 24 | 0.8388 | 1 | 0.8388 | 0 | 0 | 0 | 1 | Ref |
| 3088 | KU160652 | Unassigned | Unassigned | Unassigned | 1160_0 | Clustered | 73 | VC_1160_0 | 73 | 0.9835 | 1 | 0.9835 | 0 | 0 | 0 | 1 | Ref |
| 3089 | KU160653 | Unassigned | Unassigned | Unassigned | 1160_0 | Clustered | 73 | VC_1160_0 | 73 | 0.9835 | 1 | 0.9835 | 0 | 0 | 0 | 1 | Ref |
| 3090 | KU160654 | Unassigned | Unassigned | Unassigned | 1169_0 | Clustered | 7 | VC_1169_0 | 7 | 0.9811 | 0.99995613 | 0.981 | 0 | 0 | 0 | 1 | Ref |
| 3091 | KU160655 | Unassigned | Unassigned | Unassigned | 950_0 | Clustered | 38 | VC_950_0 | 38 | 0.9443 | 1 | 0.9443 | 0 | 0 | 0 | 1 | Ref |
| 3092 | KU160656 | Unassigned | Unassigned | Unassigned | 1073_0 | Clustered | 24 | VC_1073_0 | 24 | 0.8388 | 1 | 0.8388 | 0 | 0 | 0 | 1 | Ref |
| 3093 | KU160657 | Unassigned | Unassigned | Unassigned | 950_0 | Clustered | 38 | VC_950_0 | 38 | 0.9443 | 1 | 0.9443 | 0 | 0 | 0 | 1 | Ref |
| 3094 | KU160658 | Unassigned | Unassigned | Unassigned | 950_0 | Clustered | 38 | VC_950_0 | 38 | 0.9443 | 1 | 0.9443 | 0 | 0 | 0 | 1 | Ref |
| 3095 | KU160659 | Unassigned | Unassigned | Unassigned | 1160_0 | Clustered | 73 | VC_1160_0 | 73 | 0.9835 | 1 | 0.9835 | 0 | 0 | 0 | 1 | Ref |
| 3096 | KU160660 | Unassigned | Unassigned | Unassigned | 1074_0 | Clustered | 19 | VC_1074_0 | 19 | 0.9198 | 1 | 0.9198 | 0 | 0 | 0 | 1 | Ref |
| 3097 | KU160661 | Unassigned | Unassigned | Unassigned | 1160_0 | Clustered | 73 | VC_1160_0 | 73 | 0.9835 | 1 | 0.9835 | 0 | 0 | 0 | 1 | Ref |
| 3098 | KU160662 | Unassigned | Unassigned | Unassigned | 1160_0 | Clustered | 73 | VC_1160_0 | 73 | 0.9835 | 1 | 0.9835 | 0 | 0 | 0 | 1 | Ref |
| 3099 | KU160663 | Unassigned | Unassigned | Unassigned | 857_0 | Clustered | 11 | VC_857_0 | 11 | 0.9712 | 1 | 0.9712 | 0 | 0 | 0 | 1 | Ref |
| 3100 | KU160664 | Unassigned | Unassigned | Unassigned | 1169_0 | Clustered | 7 | VC_1169_0 | 7 | 0.9811 | 0.99995613 | 0.981 | 0 | 0 | 0 | 1 | Ref |
| 3101 | KU160665 | Unassigned | Unassigned | Unassigned | 1073_0 | Clustered | 24 | VC_1073_0 | 24 | 0.8388 | 1 | 0.8388 | 0 | 0 | 0 | 1 | Ref |
| 3102 | KU160666 | Unassigned | Unassigned | Unassigned | 857_0 | Clustered | 11 | VC_857_0 | 11 | 0.9712 | 1 | 0.9712 | 0 | 0 | 0 | 1 | Ref |
| 3103 | KU160667 | Unassigned | Unassigned | Unassigned | 950_0 | Clustered | 38 | VC_950_0 | 38 | 0.9443 | 1 | 0.9443 | 0 | 0 | 0 | 1 | Ref |
| 3104 | KU160668 | Unassigned | Unassigned | Unassigned | 1073_0 | Clustered | 24 | VC_1073_0 | 24 | 0.8388 | 1 | 0.8388 | 0 | 0 | 0 | 1 | Ref |
| 3105 | KU160669 | Unassigned | Unassigned | Unassigned | 1170_1 | Clustered | 3 | VC_1170_1 | 3 | 0.308 | 0.99836447 | 0.3075 | 0 | 0 | 0 | 1 | Ref |
| 3106 | KU160670 | Unassigned | Unassigned | Unassigned | 950_0 | Clustered | 38 | VC_950_0 | 38 | 0.9443 | 1 | 0.9443 | 0 | 0 | 0 | 1 | Ref |
| 3107 | KU160671 | Unassigned | Unassigned | Unassigned | 1160_0 | Clustered | 73 | VC_1160_0 | 73 | 0.9835 | 1 | 0.9835 | 0 | 0 | 0 | 1 | Ref |
| 3108 | KU160672 | Unassigned | Unassigned | Unassigned | 1160_0 | Clustered | 73 | VC_1160_0 | 73 | 0.9835 | 1 | 0.9835 | 0 | 0 | 0 | 1 | Ref |
| 3109 | KU160673 | Unassigned | Unassigned | Unassigned | 1170_1 | Clustered | 3 | VC_1170_1 | 3 | 0.308 | 0.99836447 | 0.3075 | 0 | 0 | 0 | 1 | Ref |
| 3110 | KU160674 | Unassigned | Unassigned | Unassigned | 950_0 | Clustered | 38 | VC_950_0 | 38 | 0.9443 | 1 | 0.9443 | 0 | 0 | 0 | 1 | Ref |
| 3111 | KU183006 | Unassigned | Unassigned | Unassigned | 823_0 | Clustered | 124 | VC_823_0 | 124 | 0.9492 | 1 | 0.9492 | 0 | 0 | 0 | 1 | Ref |
| 3112 | KU189325 | Unassigned | Unassigned | Unassigned | 975_0 | Clustered | 11 | VC_975_0 | 11 | 0.9687 | 1 | 0.9687 | 0 | 0 | 0 | 1 | Ref |
| 3113 | KU194205 | Unassigned | Unassigned | Unassigned | 575_0 | Clustered | 35 | VC_575_0 | 35 | 0.8815 | 1 | 0.8815 | 0 | 0 | 0 | 1 | Ref |
| 3114 | KU194206 | Unassigned | Unassigned | Unassigned | 575_0 | Clustered | 35 | VC_575_0 | 35 | 0.8815 | 1 | 0.8815 | 0 | 0 | 0 | 1 | Ref |
| 3115 | KU197013 | Unassigned | Unassigned | Unassigned | 74_0 | Clustered | 15 | VC_74_0 | 15 | 0.5204 | 1 | 0.5204 | 0 | 0 | 0 | 1 | Ref |
| 3116 | KU197014 | Unassigned | Unassigned | Unassigned | 749_0 | Clustered | 40 | VC_749_0 | 40 | 0.7517 | 1 | 0.7517 | 0 | 0 | 0 | 1 | Ref |
| 3117 | KU198331 | Unassigned | Unassigned | Unassigned | 79_7 | Clustered | 33 | VC_79_7 | 33 | 0.216 | 1 | 0.216 | 0 | 0 | 0 | 0.8409 | Ref |
| 3118 | KU199707 | Unassigned | Unassigned | Unassigned | 179_0 | Clustered | 53 | VC_179_0 | 53 | 0.9137 | 1 | 0.9137 | 0 | 0 | 0 | 1 | Ref |
| 3119 | KU199708 | Unassigned | Unassigned | Unassigned | 179_0 | Clustered | 53 | VC_179_0 | 53 | 0.9137 | 1 | 0.9137 | 0 | 0 | 0 | 1 | Ref |
| 3120 | KU199709 | Unassigned | Unassigned | Unassigned | 179_0 | Clustered | 53 | VC_179_0 | 53 | 0.9137 | 1 | 0.9137 | 0 | 0 | 0 | 1 | Ref |
| 3121 | KU199710 | Unassigned | Unassigned | Unassigned | 297_0 | Clustered | 5 | VC_297_0 | 5 | 0.1056 | 0.99999993 | 0.1056 | 0 | 0 | 0 | 0.9 | Ref |
| 3122 | KU204984 | Unassigned | Unassigned | Unassigned | 169_0 | Clustered | 20 | VC_169_0 | 20 | 0.7529 | 1 | 0.7529 | 0 | 0 | 0 | 1 | Ref |
| 3123 | KU230356 | Unassigned | Unassigned | Unassigned | 1173_0 | Clustered | 6 | VC_1173_0 | 6 | 0.9951 | 0.99999296 | 0.9951 | 0 | 0 | 0 | 1 | Ref |
| 3124 | KU234099 | Unassigned | Unassigned | Unassigned | 888_0 | Clustered | 28 | VC_888_0 | 28 | 0.5497 | 1 | 0.5497 | 0 | 0 | 0 | 1 | Ref |
| 3125 | KU234532 | Unassigned | Unassigned | Unassigned | 1174_0 | Clustered | 4 | VC_1174_0 | 4 | 1 | 1 | 1 | 0 | 0 | 0 | 1 | Ref |
| 3126 | KU234533 | Unassigned | Unassigned | Unassigned | 1174_0 | Clustered | 4 | VC_1174_0 | 4 | 1 | 1 | 1 | 0 | 0 | 0 | 1 | Ref |
| 3127 | KU238068 | Unassigned | Unassigned | Unassigned | 39_0 | Clustered | 35 | VC_39_0 | 35 | 0.4337 | 1 | 0.4337 | 0 | 0 | 0 | 1 | Ref |
| 3128 | KU238069 | Unassigned | Unassigned | Unassigned | 39_0 | Clustered | 35 | VC_39_0 | 35 | 0.4337 | 1 | 0.4337 | 0 | 0 | 0 | 1 | Ref |
| 3129 | KU238070 | Unassigned | Unassigned | Unassigned | 39_0 | Clustered | 35 | VC_39_0 | 35 | 0.4337 | 1 | 0.4337 | 0 | 0 | 0 | 1 | Ref |
| 3130 | KU245542 | Unassigned | Unassigned | Unassigned | 1028_0 | Clustered | 8 | VC_1028_0 | 8 | 0.9323 | 1 | 0.9323 | 0 | 0 | 0 | 1 | Ref |
| 3131 | KU253712 | Unassigned | Unassigned | Unassigned | 85_8 | Clustered/Singleton | 1 | VC_85_8 | 1 | 0 | 1 | 0 | 0 | 0 | 0 | 1 | Ref |
| 3132 | KU255188 | Unassigned | Unassigned | Unassigned | 514_0 | Clustered | 75 | VC_514_0 | 75 | 0.9402 | 1 | 0.9402 | 0 | 0 | 0 | 1 | Ref |
| 3133 | KU255730 | Unassigned | Unassigned | Unassigned | 286_8 | Clustered/Singleton | 1 | VC_286_8 | 1 | 0 | 1 | 0 | 0 | 0 | 0 | 1 | Ref |
| 3134 | KU297675 | Unassigned | Unassigned | Unassigned | 76_0 | Clustered | 37 | VC_76_0 | 37 | 0.9403 | 1 | 0.9403 | 0 | 0 | 0 | 1 | Ref |
| 3135 | KU297783 | Unassigned | Unassigned | Unassigned | 216_4 | Clustered | 13 | VC_216_4 | 13 | 0.1657 | 1 | 0.1657 | 0 | 0 | 0 | 1 | Ref |
| 3136 | KU298437 | Unassigned | Unassigned | Unassigned | 42_0 | Clustered | 49 | VC_42_0 | 49 | 0.4517 | 1 | 0.4517 | 0 | 0 | 0 | 0.8597 | Ref |
| 3137 | KU310943 | Unassigned | Unassigned | Unassigned | 297_1 | Clustered | 20 | VC_297_1 | 20 | 0.5653 | 1 | 0.5653 | 0 | 0 | 0 | 0.7684 | Ref |
| 3138 | KU356689 | Unassigned | Unassigned | Unassigned | 464_0 | Clustered | 8 | VC_464_0 | 8 | 0.2629 | 1 | 0.2629 | 0 | 0 | 0 | 1 | Ref |
| 3139 | KU356690 | Unassigned | Unassigned | Unassigned | 781_0 | Clustered | 2 | VC_781_0 | 2 | 0.0375 | 0.95691208 | 0.0359 | 0 | 0 | 0 | 1 | Ref |
| 3140 | KU356691 | Unassigned | Unassigned | Unassigned | 781_0 | Clustered | 2 | VC_781_0 | 2 | 0.0375 | 0.95691208 | 0.0359 | 0 | 0 | 0 | 1 | Ref |
| 3141 | KU497559 | Unassigned | Unassigned | Unassigned | 76_0 | Clustered | 37 | VC_76_0 | 37 | 0.9403 | 1 | 0.9403 | 0 | 0 | 0 | 1 | Ref |
| 3142 | KU504502 | Unassigned | Unassigned | Unassigned | 5_0 | Clustered | 30 | VC_5_0 | 30 | 0.9647 | 1 | 0.9647 | 0 | 0 | 0 | 1 | Ref |
| 3143 | KU510289 | Unassigned | Unassigned | Unassigned | 698_0 | Clustered | 26 | VC_698_0 | 26 | 0.9366 | 1 | 0.9366 | 0 | 0 | 0 | 1 | Ref |
| 3144 | KU517658 | Unassigned | Unassigned | Unassigned | 734_0 | Clustered | 6 | VC_734_0 | 6 | 0.1914 | 0.99996888 | 0.1914 | 0 | 0 | 0 | 1 | Ref |
| 3145 | KU521356 | Unassigned | Unassigned | Unassigned | 344_0 | Clustered | 21 | VC_344_0 | 21 | 0.5436 | 1 | 0.5436 | 0 | 0 | 0 | 1 | Ref |
| 3146 | KU522583 | Unassigned | Unassigned | Unassigned | 848_0 | Clustered | 28 | VC_848_0 | 28 | 0.7927 | 1 | 0.7927 | 0 | 0 | 0 | 1 | Ref |
| 3147 | KU530220 | Unassigned | Unassigned | Unassigned | 975_0 | Clustered | 11 | VC_975_0 | 11 | 0.9687 | 1 | 0.9687 | 0 | 0 | 0 | 1 | Ref |
| 3148 | KU535860 | Unassigned | Unassigned | Unassigned | 755_0 | Clustered | 9 | VC_755_0 | 9 | 0.4489 | 1 | 0.4489 | 0 | 0 | 0 | 1 | Ref |
| 3149 | KU568494 | Unassigned | Unassigned | Unassigned | 217_0 | Clustered | 5 | VC_217_0 | 5 | 0.069 | 0.99999998 | 0.069 | 0 | 0 | 0 | 1 | Ref |
| 3150 | KU574722 | Unassigned | Unassigned | Unassigned | 164_5 | Clustered/Singleton | 1 | VC_164_5 | 1 | 0 | 1 | 0 | 0 | 0 | 0 | 1 | Ref |
| 3151 | KU577463 | Unassigned | Unassigned | Unassigned | 187_3 | Clustered | 9 | VC_187_3 | 9 | 0.1971 | 1 | 0.1971 | 0 | 0 | 0 | 0.8611 | Ref |
| 3152 | KU578077 | Unassigned | Unassigned | Unassigned | 514_0 | Clustered | 75 | VC_514_0 | 75 | 0.9402 | 1 | 0.9402 | 0 | 0 | 0 | 1 | Ref |
| 3153 | KU594606 | Unassigned | Unassigned | Unassigned | 433_6 | Clustered/Singleton | 1 | VC_433_6 | 1 | 0 | 1 | 0 | 0 | 0 | 0 | 1 | Ref |
| 3154 | KU594607 | Unassigned | Unassigned | Unassigned | 433_4 | Clustered | 9 | VC_433_4 | 9 | 0.1285 | 1 | 0.1285 | 0 | 0 | 0 | 1 | Ref |
| 3155 | KU595432 | Unassigned | Unassigned | Unassigned | 74_0 | Clustered | 15 | VC_74_0 | 15 | 0.5204 | 1 | 0.5204 | 0 | 0 | 0 | 1 | Ref |
| 3156 | KU595433 | Unassigned | Unassigned | Unassigned | 74_0 | Clustered | 15 | VC_74_0 | 15 | 0.5204 | 1 | 0.5204 | 0 | 0 | 0 | 1 | Ref |
| 3157 | KU595434 | Unassigned | Unassigned | Unassigned | 637_0 | Clustered | 115 | VC_637_0 | 115 | 0.731 | 1 | 0.731 | 0 | 0 | 0 | 0.8764 | Ref |
| 3158 | KU599877 | Unassigned | Unassigned | Unassigned | 1157_0 | Clustered | 24 | VC_1157_0 | 24 | 1 | 1 | 1 | 0 | 0 | 0 | 1 | Ref |
| 3159 | KU599878 | Unassigned | Unassigned | Unassigned | 1157_0 | Clustered | 24 | VC_1157_0 | 24 | 1 | 1 | 1 | 0 | 0 | 0 | 1 | Ref |
| 3160 | KU599879 | Unassigned | Unassigned | Unassigned | 1157_0 | Clustered | 24 | VC_1157_0 | 24 | 1 | 1 | 1 | 0 | 0 | 0 | 1 | Ref |
| 3161 | KU599880 | Unassigned | Unassigned | Unassigned | 1040_0 | Clustered | 23 | VC_1040_0 | 23 | 0.9605 | 1 | 0.9605 | 0 | 0 | 0 | 1 | Ref |
| 3162 | KU599881 | Unassigned | Unassigned | Unassigned | 1040_0 | Clustered | 23 | VC_1040_0 | 23 | 0.9605 | 1 | 0.9605 | 0 | 0 | 0 | 1 | Ref |
| 3163 | KU599882 | Unassigned | Unassigned | Unassigned | 1040_0 | Clustered | 23 | VC_1040_0 | 23 | 0.9605 | 1 | 0.9605 | 0 | 0 | 0 | 1 | Ref |
| 3164 | KU599883 | Unassigned | Unassigned | Unassigned | 1040_0 | Clustered | 23 | VC_1040_0 | 23 | 0.9605 | 1 | 0.9605 | 0 | 0 | 0 | 1 | Ref |
| 3165 | KU599884 | Unassigned | Unassigned | Unassigned | 1040_0 | Clustered | 23 | VC_1040_0 | 23 | 0.9605 | 1 | 0.9605 | 0 | 0 | 0 | 1 | Ref |
| 3166 | KU599885 | Unassigned | Unassigned | Unassigned | 1040_0 | Clustered | 23 | VC_1040_0 | 23 | 0.9605 | 1 | 0.9605 | 0 | 0 | 0 | 1 | Ref |
| 3167 | KU599886 | Unassigned | Unassigned | Unassigned | 1157_0 | Clustered | 24 | VC_1157_0 | 24 | 1 | 1 | 1 | 0 | 0 | 0 | 1 | Ref |
| 3168 | KU599887 | Unassigned | Unassigned | Unassigned | 1023_0 | Clustered | 8 | VC_1023_0 | 8 | 0.8574 | 1 | 0.8574 | 0 | 0 | 0 | 1 | Ref |
| 3169 | KU599888 | Unassigned | Unassigned | Unassigned | 1023_0 | Clustered | 8 | VC_1023_0 | 8 | 0.8574 | 1 | 0.8574 | 0 | 0 | 0 | 1 | Ref |
| 3170 | KU599889 | Unassigned | Unassigned | Unassigned | 1023_0 | Clustered | 8 | VC_1023_0 | 8 | 0.8574 | 1 | 0.8574 | 0 | 0 | 0 | 1 | Ref |
| 3171 | KU613353 | Unassigned | Unassigned | Unassigned | 225_0 | Clustered | 32 | VC_225_0 | 32 | 0.848 | 1 | 0.848 | 0 | 0 | 0 | 0.996 | Ref |
| 3172 | KU640380 | Unassigned | Unassigned | Unassigned | 85_13 | Clustered/Singleton | 1 | VC_85_13 | 1 | 0 | 1 | 0 | 0 | 0 | 0 | 1 | Ref |
| 3173 | KU647626 | Unassigned | Unassigned | Unassigned | 1179_0 | Clustered | 3 | VC_1179_0 | 3 | 0.7588 | 0.96819872 | 0.7347 | 0 | 0 | 0 | 1 | Ref |
| 3174 | KU647627 | Unassigned | Unassigned | Unassigned | 1179_0 | Clustered | 3 | VC_1179_0 | 3 | 0.7588 | 0.96819872 | 0.7347 | 0 | 0 | 0 | 1 | Ref |
| 3175 | KU647628 | Unassigned | Unassigned | Unassigned | 802_0 | Clustered | 17 | VC_802_0 | 17 | 0.8025 | 1 | 0.8025 | 0 | 0 | 0 | 1 | Ref |
| 3176 | KU647629 | Unassigned | Unassigned | Unassigned | 1073_0 | Clustered | 24 | VC_1073_0 | 24 | 0.8388 | 1 | 0.8388 | 0 | 0 | 0 | 1 | Ref |
| 3177 | KU666550 | Unassigned | Unassigned | Unassigned | 111_0 | Clustered | 91 | VC_111_0 | 91 | 0.9587 | 1 | 0.9587 | 0 | 0 | 0 | 1 | Ref |
| 3178 | KU678389 | Unassigned | Unassigned | Unassigned | 256_0 | Clustered | 14 | VC_256_0 | 14 | 0.2368 | 1 | 0.2368 | 0 | 0 | 0 | 1 | Ref |
| 3179 | KU678390 | Unassigned | Unassigned | Unassigned | 256_0 | Clustered | 14 | VC_256_0 | 14 | 0.2368 | 1 | 0.2368 | 0 | 0 | 0 | 1 | Ref |
| 3180 | KU678391 | Unassigned | Unassigned | Unassigned | 256_0 | Clustered | 14 | VC_256_0 | 14 | 0.2368 | 1 | 0.2368 | 0 | 0 | 0 | 1 | Ref |
| 3181 | KU678392 | Unassigned | Unassigned | Unassigned | 256_0 | Clustered | 14 | VC_256_0 | 14 | 0.2368 | 1 | 0.2368 | 0 | 0 | 0 | 1 | Ref |
| 3182 | KU686192 | Unassigned | Unassigned | Unassigned | 433_0 | Clustered | 6 | VC_433_0 | 6 | 0.0787 | 1 | 0.0787 | 0 | 0 | 0 | 1 | Ref |
| 3183 | KU686193 | Unassigned | Unassigned | Unassigned | 433_0 | Clustered | 6 | VC_433_0 | 6 | 0.0787 | 1 | 0.0787 | 0 | 0 | 0 | 1 | Ref |
| 3184 | KU686194 | Unassigned | Unassigned | Unassigned | 433_0 | Clustered | 6 | VC_433_0 | 6 | 0.0787 | 1 | 0.0787 | 0 | 0 | 0 | 1 | Ref |
| 3185 | KU686195 | Unassigned | Unassigned | Unassigned | 433_0 | Clustered | 6 | VC_433_0 | 6 | 0.0787 | 1 | 0.0787 | 0 | 0 | 0 | 1 | Ref |
| 3186 | KU686196 | Unassigned | Unassigned | Unassigned | 433_0 | Clustered | 6 | VC_433_0 | 6 | 0.0787 | 1 | 0.0787 | 0 | 0 | 0 | 1 | Ref |
| 3187 | KU686197 | Unassigned | Unassigned | Unassigned | 433_3 | Clustered | 3 | VC_433_3 | 3 | 0.0292 | 0.9981281 | 0.0292 | 0 | 0 | 0 | 1 | Ref |
| 3188 | KU686198 | Unassigned | Unassigned | Unassigned | 433_3 | Clustered | 3 | VC_433_3 | 3 | 0.0292 | 0.9981281 | 0.0292 | 0 | 0 | 0 | 1 | Ref |
| 3189 | KU686199 | Unassigned | Unassigned | Unassigned | 433_3 | Clustered | 3 | VC_433_3 | 3 | 0.0292 | 0.9981281 | 0.0292 | 0 | 0 | 0 | 1 | Ref |
| 3190 | KU686200 | Unassigned | Unassigned | Unassigned | 433_1 | Clustered | 9 | VC_433_1 | 9 | 0.1288 | 1 | 0.1288 | 0 | 0 | 0 | 0.8611 | Ref |
| 3191 | KU686201 | Unassigned | Unassigned | Unassigned | 433_1 | Clustered | 9 | VC_433_1 | 9 | 0.1288 | 1 | 0.1288 | 0 | 0 | 0 | 0.8611 | Ref |
| 3192 | KU686202 | Unassigned | Unassigned | Unassigned | 433_1 | Clustered | 9 | VC_433_1 | 9 | 0.1288 | 1 | 0.1288 | 0 | 0 | 0 | 0.8611 | Ref |
| 3193 | KU686204 | Unassigned | Unassigned | Unassigned | 432_15 | Clustered | 3 | VC_432_15 | 3 | 0.0418 | 0.99855682 | 0.0417 | 0 | 0 | 0 | 1 | Ref |
| 3194 | KU686205 | Unassigned | Unassigned | Unassigned | 432_15 | Clustered | 3 | VC_432_15 | 3 | 0.0418 | 0.99855682 | 0.0417 | 0 | 0 | 0 | 1 | Ref |
| 3195 | KU686206 | Unassigned | Unassigned | Unassigned | 432_15 | Clustered | 3 | VC_432_15 | 3 | 0.0418 | 0.99855682 | 0.0417 | 0 | 0 | 0 | 1 | Ref |
| 3196 | KU686207 | Unassigned | Unassigned | Unassigned | 655_5 | Clustered | 3 | VC_655_5 | 3 | 0.0166 | 0.9986063 | 0.0165 | 0 | 0 | 0 | 1 | Ref |
| 3197 | KU686208 | Unassigned | Unassigned | Unassigned | 655_5 | Clustered | 3 | VC_655_5 | 3 | 0.0166 | 0.9986063 | 0.0165 | 0 | 0 | 0 | 1 | Ref |
| 3198 | KU686209 | Unassigned | Unassigned | Unassigned | 655_5 | Clustered | 3 | VC_655_5 | 3 | 0.0166 | 0.9986063 | 0.0165 | 0 | 0 | 0 | 1 | Ref |
| 3199 | KU686211 | Unassigned | Unassigned | Unassigned | 433_10 | Clustered/Singleton | 1 | VC_433_10 | 1 | 0 | 1 | 0 | 0 | 0 | 0 | 1 | Ref |
| 3200 | KU686212 | Unassigned | Unassigned | Unassigned | 657_0 | Clustered | 3 | VC_657_0 | 3 | 0.0217 | 0.92528126 | 0.0201 | 0 | 0 | 0 | 1 | Ref |
| 3201 | KU686213 | Unassigned | Unassigned | Unassigned | 657_0 | Clustered | 3 | VC_657_0 | 3 | 0.0217 | 0.92528126 | 0.0201 | 0 | 0 | 0 | 1 | Ref |
| 3202 | KU687347 | Unassigned | Unassigned | Unassigned | 385_0 | Clustered | 51 | VC_385_0 | 51 | 0.8754 | 1 | 0.8754 | 0 | 0 | 0 | 1 | Ref |
| 3203 | KU687348 | Unassigned | Unassigned | Unassigned | 385_0 | Clustered | 51 | VC_385_0 | 51 | 0.8754 | 1 | 0.8754 | 0 | 0 | 0 | 1 | Ref |
| 3204 | KU687349 | Unassigned | Unassigned | Unassigned | 451_0 | Clustered | 80 | VC_451_0 | 80 | 0.9991 | 1 | 0.9991 | 0 | 0 | 0 | 1 | Ref |
| 3205 | KU687350 | Unassigned | Unassigned | Unassigned | 451_0 | Clustered | 80 | VC_451_0 | 80 | 0.9991 | 1 | 0.9991 | 0 | 0 | 0 | 1 | Ref |
| 3206 | KU687351 | Unassigned | Unassigned | Unassigned | 451_0 | Clustered | 80 | VC_451_0 | 80 | 0.9991 | 1 | 0.9991 | 0 | 0 | 0 | 1 | Ref |
| 3207 | KU695581 | Unassigned | Unassigned | Unassigned | 221_0 | Clustered | 22 | VC_221_0 | 22 | 0.7447 | 1 | 0.7447 | 0 | 0 | 0 | 1 | Ref |
| 3208 | KU695582 | Unassigned | Unassigned | Unassigned | 223_0 | Clustered | 38 | VC_223_0 | 38 | 0.9095 | 1 | 0.9095 | 0 | 0 | 0 | 1 | Ref |
| 3209 | KU708004 | Unassigned | Unassigned | Unassigned | 298_0 | Clustered/Singleton | 1 | VC_298_0 | 1 | 0 | 1 | 0 | 0 | 0 | 0 | 1 | Ref |
| 3210 | KU716094 | Unassigned | Unassigned | Unassigned | 225_0 | Clustered | 32 | VC_225_0 | 32 | 0.848 | 1 | 0.848 | 0 | 0 | 0 | 0.996 | Ref |
| 3211 | KU716095 | Unassigned | Unassigned | Unassigned | 225_0 | Clustered | 32 | VC_225_0 | 32 | 0.848 | 1 | 0.848 | 0 | 0 | 0 | 0.996 | Ref |
| 3212 | KU726251 | Unassigned | Unassigned | Unassigned | 959_5 | Clustered | 19 | VC_959_5 | 19 | 0.2626 | 1 | 0.2626 | 0 | 0 | 0 | 1 | Ref |
| 3213 | KU728633 | Unassigned | Unassigned | Unassigned | 686_1 | Clustered | 2 | VC_686_1 | 2 | 0.2204 | 0.91327915 | 0.2013 | 0 | 0 | 0 | 1 | Ref |
| 3214 | KU737344 | Unassigned | Unassigned | Unassigned | 884_0 | Clustered | 28 | VC_884_0 | 28 | 0.6211 | 1 | 0.6211 | 0 | 0 | 0 | 0.9841 | Ref |
| 3215 | KU737345 | Unassigned | Unassigned | Unassigned | 883_1 | Clustered | 12 | VC_883_1 | 12 | 0.2441 | 1 | 0.2441 | 0 | 0 | 0 | 1 | Ref |
| 3216 | KU737346 | Unassigned | Unassigned | Unassigned | 883_0 | Clustered | 10 | VC_883_0 | 10 | 0.2015 | 1 | 0.2015 | 0 | 0 | 0 | 1 | Ref |
| 3217 | KU737347 | Unassigned | Unassigned | Unassigned | 883_1 | Clustered | 12 | VC_883_1 | 12 | 0.2441 | 1 | 0.2441 | 0 | 0 | 0 | 1 | Ref |
| 3218 | KU737348 | Unassigned | Unassigned | Unassigned | 884_0 | Clustered | 28 | VC_884_0 | 28 | 0.6211 | 1 | 0.6211 | 0 | 0 | 0 | 0.9841 | Ref |
| 3219 | KU737349 | Unassigned | Unassigned | Unassigned | 884_0 | Clustered | 28 | VC_884_0 | 28 | 0.6211 | 1 | 0.6211 | 0 | 0 | 0 | 0.9841 | Ref |
| 3220 | KU737350 | Unassigned | Unassigned | Unassigned | 884_0 | Clustered | 28 | VC_884_0 | 28 | 0.6211 | 1 | 0.6211 | 0 | 0 | 0 | 0.9841 | Ref |
| 3221 | KU737351 | Unassigned | Unassigned | Unassigned | 884_0 | Clustered | 28 | VC_884_0 | 28 | 0.6211 | 1 | 0.6211 | 0 | 0 | 0 | 0.9841 | Ref |
| 3222 | KU737352 | Unassigned | Unassigned | Unassigned | 884_0 | Clustered | 28 | VC_884_0 | 28 | 0.6211 | 1 | 0.6211 | 0 | 0 | 0 | 0.9841 | Ref |
| 3223 | KU743887 | Unassigned | Unassigned | Unassigned | 423_0 | Clustered | 45 | VC_423_0 | 45 | 0.9484 | 1 | 0.9484 | 0 | 0 | 0 | 1 | Ref |
| 3224 | KU747973 | Unassigned | Unassigned | Unassigned | 644_0 | Clustered | 5 | VC_644_0 | 5 | 0.6649 | 0.99999975 | 0.6649 | 0 | 0 | 0 | 1 | Ref |
| 3225 | KU760857 | Unassigned | Unassigned | Unassigned | 247_0 | Clustered | 14 | VC_247_0 | 14 | 0.9119 | 1 | 0.9119 | 0 | 0 | 0 | 0.967 | Ref |
| 3226 | KU761558 | Unassigned | Unassigned | Unassigned | 216_3 | Clustered | 8 | VC_216_3 | 8 | 0.0701 | 1 | 0.0701 | 0 | 0 | 0 | 0.8571 | Ref |
| 3227 | KU761559 | Unassigned | Unassigned | Unassigned | 216_3 | Clustered | 8 | VC_216_3 | 8 | 0.0701 | 1 | 0.0701 | 0 | 0 | 0 | 0.8571 | Ref |
| 3228 | KU761955 | Unassigned | Unassigned | Unassigned | 76_0 | Clustered | 37 | VC_76_0 | 37 | 0.9403 | 1 | 0.9403 | 0 | 0 | 0 | 1 | Ref |
| 3229 | KU831549 | Unassigned | Unassigned | Unassigned | 817_0 | Clustered | 14 | VC_817_0 | 14 | 0.6342 | 1 | 0.6342 | 0 | 0 | 0 | 1 | Ref |
| 3230 | KU836751 | Unassigned | Unassigned | Unassigned | 100_0 | Clustered | 15 | VC_100_0 | 15 | 0.6536 | 1 | 0.6536 | 0 | 0 | 0 | 1 | Ref |
| 3231 | KU847400 | Unassigned | Unassigned | Unassigned | 884_1 | Clustered | 8 | VC_884_1 | 8 | 0.1102 | 0.9989323 | 0.1101 | 0 | 0 | 0 | 0.75 | Ref |
| 3232 | KU848187 | Unassigned | Unassigned | Unassigned | 377_0 | Clustered | 3 | VC_377_0 | 3 | 0.3906 | 0.99785056 | 0.3898 | 0 | 0 | 0 | 1 | Ref |
| 3233 | KU862660 | Unassigned | Unassigned | Unassigned | 957_1 | Clustered/Singleton | 1 | VC_957_1 | 1 | 0 | 1 | 0 | 0 | 0 | 0 | 1 | Ref |
| 3234 | KU865302 | Unassigned | Unassigned | Unassigned | 520_40 | Clustered | 4 | VC_520_40 | 4 | 0.0396 | 0.99998776 | 0.0396 | 0 | 0 | 0 | 1 | Ref |
| 3235 | KU865303 | Unassigned | Unassigned | Unassigned | 551_2 | Clustered | 17 | VC_551_2 | 17 | 0.0973 | 1 | 0.0973 | 0 | 0 | 0 | 0.9779 | Ref |
| 3236 | KU867876 | Unassigned | Unassigned | Unassigned | 283_25 | Clustered/Singleton | 1 | VC_283_25 | 1 | 0 | 1 | 0 | 0 | 0 | 0 | 1 | Ref |
| 3237 | KU867906 | Unassigned | Unassigned | Unassigned | 517_1 | Clustered | 108 | VC_517_1 | 108 | 0.8483 | 1 | 0.8483 | 0 | 0 | 0 | 0.9955 | Ref |
| 3238 | KU867907 | Unassigned | Unassigned | Unassigned | 371_15 | Clustered | 44 | VC_371_15 | 44 | 0.1391 | 1 | 0.1391 | 0 | 0 | 0 | 0.7474 | Ref |

| | | | | | | | | | | | | | | | | | |
|---|---|---|---|---|---|---|---|---|---|---|---|---|---|---|---|---|---|
| 3239 | KU878088 | Unassigned | Unassigned | Unassigned | 448_0 | Clustered | 4 | VC_448_0 | 4 | 0.3059 | 0.99978853 | 0.3058 | 0 | 0 | 0 | 1 | Ref |
| 3240 | KU878967 | Unassigned | Unassigned | Unassigned | 28_1 | Clustered | 53 | VC_28_1 | 53 | 0.7164 | 1 | 0.7164 | 0 | 0 | 0 | 0.9325 | Ref |
| 3241 | KU878968 | Unassigned | Unassigned | Unassigned | 290_5 | Clustered | 14 | VC_290_5 | 14 | 0.1422 | 1 | 0.1422 | 0 | 0 | 0 | 0.9231 | Ref |
| 3242 | KU878969 | Unassigned | Unassigned | Unassigned | 290_5 | Clustered | 14 | VC_290_5 | 14 | 0.1422 | 1 | 0.1422 | 0 | 0 | 0 | 0.9231 | Ref |
| 3243 | KU884561 | Unassigned | Unassigned | Unassigned | 612_0 | Clustered | 9 | VC_612_0 | 9 | 0.9545 | 1 | 0.9545 | 0 | 0 | 0 | 1 | Ref |
| 3244 | KU884562 | Unassigned | Unassigned | Unassigned | 612_0 | Clustered | 9 | VC_612_0 | 9 | 0.9545 | 1 | 0.9545 | 0 | 0 | 0 | 1 | Ref |
| 3245 | KU884563 | Unassigned | Unassigned | Unassigned | 612_0 | Clustered | 9 | VC_612_0 | 9 | 0.9545 | 1 | 0.9545 | 0 | 0 | 0 | 1 | Ref |
| 3246 | KU884564 | Unassigned | Unassigned | Unassigned | 612_0 | Clustered | 9 | VC_612_0 | 9 | 0.9545 | 1 | 0.9545 | 0 | 0 | 0 | 1 | Ref |
| 3247 | KU884565 | Unassigned | Unassigned | Unassigned | 612_0 | Clustered | 9 | VC_612_0 | 9 | 0.9545 | 1 | 0.9545 | 0 | 0 | 0 | 1 | Ref |
| 3248 | KU885988 | Unassigned | Unassigned | Unassigned | 833_0 | Clustered | 16 | VC_833_0 | 16 | 0.9351 | 1 | 0.9351 | 0 | 0 | 0 | 1 | Ref |
| 3249 | KU885989 | Unassigned | Unassigned | Unassigned | 833_0 | Clustered | 16 | VC_833_0 | 16 | 0.9351 | 1 | 0.9351 | 0 | 0 | 0 | 1 | Ref |
| 3250 | KU885990 | Unassigned | Unassigned | Unassigned | 833_0 | Clustered | 16 | VC_833_0 | 16 | 0.9351 | 1 | 0.9351 | 0 | 0 | 0 | 1 | Ref |
| 3251 | KU886222 | Unassigned | Unassigned | Unassigned | 489_1 | Clustered | 11 | VC_489_1 | 11 | 0.4621 | 1 | 0.4621 | 0 | 0 | 0 | 1 | Ref |
| 3252 | KU886223 | Unassigned | Unassigned | Unassigned | 489_1 | Clustered | 11 | VC_489_1 | 11 | 0.4621 | 1 | 0.4621 | 0 | 0 | 0 | 1 | Ref |
| 3253 | KU886224 | Unassigned | Unassigned | Unassigned | 489_1 | Clustered | 11 | VC_489_1 | 11 | 0.4621 | 1 | 0.4621 | 0 | 0 | 0 | 1 | Ref |
| 3254 | KU886225 | Unassigned | Unassigned | Unassigned | 489_1 | Clustered | 11 | VC_489_1 | 11 | 0.4621 | 1 | 0.4621 | 0 | 0 | 0 | 1 | Ref |
| 3255 | KU925172 | Unassigned | Unassigned | Unassigned | 283_8 | Clustered/Singleton | 1 | VC_283_8 | 1 | 0 | 1 | 0 | 0 | 0 | 0 | 1 | Ref |
| 3256 | KU927491 | Unassigned | Unassigned | Unassigned | 28_1 | Clustered | 53 | VC_28_1 | 53 | 0.7164 | 1 | 0.7164 | 0 | 0 | 0 | 0.9325 | Ref |
| 3257 | KU927492 | Unassigned | Unassigned | Unassigned | 28_1 | Clustered | 53 | VC_28_1 | 53 | 0.7164 | 1 | 0.7164 | 0 | 0 | 0 | 0.9325 | Ref |
| 3258 | KU927493 | Unassigned | Unassigned | Unassigned | 45_0 | Clustered | 12 | VC_45_0 | 12 | 0.35 | 1 | 0.35 | 0 | 0 | 0 | 1 | Ref |
| 3259 | KU927494 | Unassigned | Unassigned | Unassigned | 28_1 | Clustered | 53 | VC_28_1 | 53 | 0.7164 | 1 | 0.7164 | 0 | 0 | 0 | 0.9325 | Ref |
| 3260 | KU927495 | Unassigned | Unassigned | Unassigned | 28_1 | Clustered | 53 | VC_28_1 | 53 | 0.7164 | 1 | 0.7164 | 0 | 0 | 0 | 0.9325 | Ref |
| 3261 | KU927496 | Unassigned | Unassigned | Unassigned | 28_1 | Clustered | 53 | VC_28_1 | 53 | 0.7164 | 1 | 0.7164 | 0 | 0 | 0 | 0.9325 | Ref |
| 3262 | KU927497 | Unassigned | Unassigned | Unassigned | 191_19 | Clustered | 12 | VC_191_19 | 12 | 0.0655 | 1 | 0.0655 | 0 | 0 | 0 | 0.8636 | Ref |
| 3263 | KU927498 | Unassigned | Unassigned | Unassigned | 28_1 | Clustered | 53 | VC_28_1 | 53 | 0.7164 | 1 | 0.7164 | 0 | 0 | 0 | 0.9325 | Ref |
| 3264 | KU927499 | Unassigned | Unassigned | Unassigned | 392_0 | Clustered | 126 | VC_392_0 | 126 | 0.9621 | 1 | 0.9621 | 0 | 0 | 0 | 1 | Ref |
| 3265 | KU927500 | Unassigned | Unassigned | Unassigned | 592_0 | Clustered | 50 | VC_592_0 | 50 | 0.7314 | 1 | 0.7314 | 0 | 0 | 0 | 1 | Ref |
| 3266 | KU935726 | Unassigned | Unassigned | Unassigned | 521_0 | Clustered | 37 | VC_521_0 | 37 | 0.912 | 1 | 0.912 | 0 | 0 | 0 | 1 | Ref |
| 3267 | KU935727 | Unassigned | Unassigned | Unassigned | 521_0 | Clustered | 37 | VC_521_0 | 37 | 0.912 | 1 | 0.912 | 0 | 0 | 0 | 1 | Ref |
| 3268 | KU935728 | Unassigned | Unassigned | Unassigned | 521_0 | Clustered | 37 | VC_521_0 | 37 | 0.912 | 1 | 0.912 | 0 | 0 | 0 | 1 | Ref |
| 3269 | KU935729 | Unassigned | Unassigned | Unassigned | 521_0 | Clustered | 37 | VC_521_0 | 37 | 0.912 | 1 | 0.912 | 0 | 0 | 0 | 1 | Ref |
| 3270 | KU935730 | Unassigned | Unassigned | Unassigned | 521_0 | Clustered | 37 | VC_521_0 | 37 | 0.912 | 1 | 0.912 | 0 | 0 | 0 | 1 | Ref |
| 3271 | KU935731 | Unassigned | Unassigned | Unassigned | 521_0 | Clustered | 37 | VC_521_0 | 37 | 0.912 | 1 | 0.912 | 0 | 0 | 0 | 1 | Ref |
| 3272 | KU946962 | Unassigned | Unassigned | Unassigned | 452_0 | Clustered | 66 | VC_452_0 | 66 | 0.9248 | 1 | 0.9248 | 0 | 0 | 0 | 1 | Ref |
| 3273 | KU948710 | Unassigned | Unassigned | Unassigned | 838_0 | Clustered | 29 | VC_838_0 | 29 | 0.8611 | 1 | 0.8611 | 0 | 0 | 0 | 1 | Ref |
| 3274 | KU951146 | Unassigned | Unassigned | Unassigned | 637_0 | Clustered | 115 | VC_637_0 | 115 | 0.731 | 1 | 0.731 | 0 | 0 | 0 | 0.8764 | Ref |
| 3275 | KU951147 | Unassigned | Unassigned | Unassigned | 637_0 | Clustered | 115 | VC_637_0 | 115 | 0.731 | 1 | 0.731 | 0 | 0 | 0 | 0.8764 | Ref |
| 3276 | KU958700 | Unassigned | Unassigned | Unassigned | 588_0 | Clustered | 2 | VC_588_0 | 2 | 0.0832 | 0.94395095 | 0.0785 | 0 | 0 | 0 | 1 | Ref |
| 3277 | KU963245 | Unassigned | Unassigned | Unassigned | 868_0 | Clustered | 25 | VC_868_0 | 25 | 0.5791 | 1 | 0.5791 | 0 | 0 | 0 | 1 | Ref |
| 3278 | KU963246 | Unassigned | Unassigned | Unassigned | 1175_0 | Clustered | 22 | VC_1175_0 | 22 | 0.4562 | 1 | 0.4562 | 0 | 0 | 0 | 1 | Ref |
| 3279 | KU963247 | Unassigned | Unassigned | Unassigned | 1175_0 | Clustered | 22 | VC_1175_0 | 22 | 0.4562 | 1 | 0.4562 | 0 | 0 | 0 | 1 | Ref |
| 3280 | KU963249 | Unassigned | Unassigned | Unassigned | 525_1 | Clustered | 5 | VC_525_1 | 5 | 0.0485 | 0.99999947 | 0.0485 | 0 | 0 | 0 | 0.9 | Ref |
| 3281 | KU963250 | Unassigned | Unassigned | Unassigned | 1093_0 | Clustered | 40 | VC_1093_0 | 40 | 0.8754 | 1 | 0.8754 | 0 | 0 | 0 | 1 | Ref |
| 3282 | KU963251 | Unassigned | Unassigned | Unassigned | 1176_0 | Clustered | 41 | VC_1176_0 | 41 | 0.9558 | 1 | 0.9558 | 0 | 0 | 0 | 0.9976 | Ref |
| 3283 | KU963252 | Unassigned | Unassigned | Unassigned | 1177_0 | Clustered | 14 | VC_1177_0 | 14 | 0.7847 | 1 | 0.7847 | 0 | 0 | 0 | 0.989 | Ref |
| 3284 | KU963253 | Unassigned | Unassigned | Unassigned | 1095_0 | Clustered | 16 | VC_1095_0 | 16 | 0.458 | 1 | 0.458 | 0 | 0 | 0 | 1 | Ref |
| 3285 | KU963254 | Unassigned | Unassigned | Unassigned | 1176_0 | Clustered | 41 | VC_1176_0 | 41 | 0.9558 | 1 | 0.9558 | 0 | 0 | 0 | 0.9976 | Ref |
| 3286 | KU963255 | Unassigned | Unassigned | Unassigned | 525_0 | Clustered | 25 | VC_525_0 | 25 | 0.4597 | 1 | 0.4597 | 0 | 0 | 0 | 1 | Ref |
| 3287 | KU963256 | Unassigned | Unassigned | Unassigned | 522_0 | Clustered | 3 | VC_522_0 | 3 | 0.0836 | 0.9985266 | 0.0835 | 0 | 0 | 0 | 1 | Ref |
| 3288 | KU963257 | Unassigned | Unassigned | Unassigned | 525_0 | Clustered | 25 | VC_525_0 | 25 | 0.4597 | 1 | 0.4597 | 0 | 0 | 0 | 1 | Ref |
| 3289 | KU963258 | Unassigned | Unassigned | Unassigned | 869_0 | Clustered | 9 | VC_869_0 | 9 | 0.2624 | 1 | 0.2624 | 0 | 0 | 0 | 1 | Ref |
| 3290 | KU963259 | Unassigned | Unassigned | Unassigned | 1176_0 | Clustered | 41 | VC_1176_0 | 41 | 0.9558 | 1 | 0.9558 | 0 | 0 | 0 | 0.9976 | Ref |
| 3291 | KU963260 | Unassigned | Unassigned | Unassigned | 919_0 | Clustered | 39 | VC_919_0 | 39 | 0.9069 | 1 | 0.9069 | 0 | 0 | 0 | 1 | Ref |
| 3292 | KU963261 | Unassigned | Unassigned | Unassigned | 954_0 | Clustered | 13 | VC_954_0 | 13 | 0.863 | 1 | 0.863 | 0 | 0 | 0 | 1 | Ref |
| 3293 | KU963262 | Unassigned | Unassigned | Unassigned | 869_0 | Clustered | 9 | VC_869_0 | 9 | 0.2624 | 1 | 0.2624 | 0 | 0 | 0 | 1 | Ref |
| 3294 | KU963263 | Unassigned | Unassigned | Unassigned | 525_1 | Clustered | 5 | VC_525_1 | 5 | 0.0485 | 0.99999947 | 0.0485 | 0 | 0 | 0 | 0.9 | Ref |
| 3295 | KU981050 | Unassigned | Unassigned | Unassigned | 269_0 | Clustered | 3 | VC_269_0 | 3 | 0.1948 | 0.99308177 | 0.1934 | 0 | 0 | 0 | 1 | Ref |
| 3296 | KU984914 | Unassigned | Unassigned | Unassigned | 514_0 | Clustered | 75 | VC_514_0 | 75 | 0.9402 | 1 | 0.9402 | 0 | 0 | 0 | 1 | Ref |
| 3297 | KU985090 | Unassigned | Unassigned | Unassigned | 519_0 | Clustered | 39 | VC_519_0 | 39 | 0.8156 | 1 | 0.8156 | 0 | 0 | 0 | 1 | Ref |
| 3298 | KU985091 | Unassigned | Unassigned | Unassigned | 514_0 | Clustered | 75 | VC_514_0 | 75 | 0.9402 | 1 | 0.9402 | 0 | 0 | 0 | 1 | Ref |
| 3299 | KU985092 | Unassigned | Unassigned | Unassigned | 517_1 | Clustered | 108 | VC_517_1 | 108 | 0.8483 | 1 | 0.8483 | 0 | 0 | 0 | 0.9955 | Ref |
| 3300 | KU985093 | Unassigned | Unassigned | Unassigned | 216_6 | Clustered | 42 | VC_216_6 | 42 | 0.473 | 1 | 0.473 | 0 | 0 | 0 | 0.8351 | Ref |
| 3301 | KU985094 | Unassigned | Unassigned | Unassigned | 517_1 | Clustered | 108 | VC_517_1 | 108 | 0.8483 | 1 | 0.8483 | 0 | 0 | 0 | 0.9955 | Ref |
| 3302 | KU985095 | Unassigned | Unassigned | Unassigned | 216_6 | Clustered | 42 | VC_216_6 | 42 | 0.473 | 1 | 0.473 | 0 | 0 | 0 | 0.8351 | Ref |
| 3303 | KU985096 | Unassigned | Unassigned | Unassigned | 223_0 | Clustered | 38 | VC_223_0 | 38 | 0.9095 | 1 | 0.9095 | 0 | 0 | 0 | 1 | Ref |
| 3304 | KU992911 | Unassigned | Unassigned | Unassigned | 96_0 | Clustered | 30 | VC_96_0 | 30 | 0.718 | 1 | 0.718 | 0 | 0 | 0 | 1 | Ref |
| 3305 | KU997639 | Unassigned | Unassigned | Unassigned | 888_0 | Clustered | 28 | VC_888_0 | 28 | 0.5497 | 1 | 0.5497 | 0 | 0 | 0 | 1 | Ref |
| 3306 | KU998234 | Unassigned | Unassigned | Unassigned | 1178_0 | Clustered | 24 | VC_1178_0 | 24 | 0.9524 | 1 | 0.9524 | 0 | 0 | 0 | 1 | Ref |
| 3307 | KU998235 | Unassigned | Unassigned | Unassigned | 1177_0 | Clustered | 14 | VC_1177_0 | 14 | 0.7847 | 1 | 0.7847 | 0 | 0 | 0 | 0.989 | Ref |
| 3308 | KU998236 | Unassigned | Unassigned | Unassigned | 1176_0 | Clustered | 41 | VC_1176_0 | 41 | 0.9558 | 1 | 0.9558 | 0 | 0 | 0 | 0.9976 | Ref |
| 3309 | KU998237 | Unassigned | Unassigned | Unassigned | 406_0 | Clustered | 11 | VC_406_0 | 11 | 0.9449 | 1 | 0.9449 | 0 | 0 | 0 | 1 | Ref |
| 3310 | KU998238 | Unassigned | Unassigned | Unassigned | 406_0 | Clustered | 11 | VC_406_0 | 11 | 0.9449 | 1 | 0.9449 | 0 | 0 | 0 | 1 | Ref |
| 3311 | KU998239 | Unassigned | Unassigned | Unassigned | 919_0 | Clustered | 39 | VC_919_0 | 39 | 0.9069 | 1 | 0.9069 | 0 | 0 | 0 | 1 | Ref |
| 3312 | KU998240 | Unassigned | Unassigned | Unassigned | 866_1 | Clustered | 20 | VC_866_1 | 20 | 0.4814 | 1 | 0.4814 | 0 | 0 | 0 | 1 | Ref |
| 3313 | KU998241 | Unassigned | Unassigned | Unassigned | 868_0 | Clustered | 25 | VC_868_0 | 25 | 0.5791 | 1 | 0.5791 | 0 | 0 | 0 | 1 | Ref |
| 3314 | KU998242 | Unassigned | Unassigned | Unassigned | 869_0 | Clustered | 9 | VC_869_0 | 9 | 0.2624 | 1 | 0.2624 | 0 | 0 | 0 | 1 | Ref |
| 3315 | KU998243 | Unassigned | Unassigned | Unassigned | 869_0 | Clustered | 9 | VC_869_0 | 9 | 0.2624 | 1 | 0.2624 | 0 | 0 | 0 | 1 | Ref |
| 3316 | KU998244 | Unassigned | Unassigned | Unassigned | 1181_1 | Clustered | 10 | VC_1181_1 | 10 | 0.5828 | 1 | 0.5828 | 0 | 0 | 0 | 1 | Ref |
| 3317 | KU998245 | Unassigned | Unassigned | Unassigned | 1181_0 | Clustered | 2 | VC_1181_0 | 2 | 0.0461 | 0.95126432 | 0.0439 | 0 | 0 | 0 | 1 | Ref |
| 3318 | KU998246 | Unassigned | Unassigned | Unassigned | 1181_1 | Clustered | 10 | VC_1181_1 | 10 | 0.5828 | 1 | 0.5828 | 0 | 0 | 0 | 1 | Ref |
| 3319 | KU998247 | Unassigned | Unassigned | Unassigned | 1181_1 | Clustered | 10 | VC_1181_1 | 10 | 0.5828 | 1 | 0.5828 | 0 | 0 | 0 | 1 | Ref |
| 3320 | KU998248 | Unassigned | Unassigned | Unassigned | 1176_0 | Clustered | 41 | VC_1176_0 | 41 | 0.9558 | 1 | 0.9558 | 0 | 0 | 0 | 0.9976 | Ref |
| 3321 | KU998249 | Unassigned | Unassigned | Unassigned | 224_0 | Clustered | 19 | VC_224_0 | 19 | 0.9416 | 1 | 0.9416 | 0 | 0 | 0 | 1 | Ref |
| 3322 | KU998250 | Unassigned | Unassigned | Unassigned | 224_0 | Clustered | 19 | VC_224_0 | 19 | 0.9416 | 1 | 0.9416 | 0 | 0 | 0 | 1 | Ref |
| 3323 | KU998251 | Unassigned | Unassigned | Unassigned | 224_0 | Clustered | 19 | VC_224_0 | 19 | 0.9416 | 1 | 0.9416 | 0 | 0 | 0 | 1 | Ref |
| 3324 | KU998252 | Unassigned | Unassigned | Unassigned | 870_0 | Clustered | 5 | VC_870_0 | 5 | 0.424 | 0.99999997 | 0.424 | 0 | 0 | 0 | 1 | Ref |
| 3325 | KU998253 | Unassigned | Unassigned | Unassigned | 870_0 | Clustered | 5 | VC_870_0 | 5 | 0.424 | 0.99999997 | 0.424 | 0 | 0 | 0 | 1 | Ref |
| 3326 | KU998254 | Unassigned | Unassigned | Unassigned | 870_0 | Clustered | 5 | VC_870_0 | 5 | 0.424 | 0.99999997 | 0.424 | 0 | 0 | 0 | 1 | Ref |
| 3327 | KU998255 | Unassigned | Unassigned | Unassigned | 949_0 | Clustered | 10 | VC_949_0 | 10 | 0.5977 | 1 | 0.5977 | 0 | 0 | 0 | 1 | Ref |
| 3328 | KU998256 | Unassigned | Unassigned | Unassigned | 949_0 | Clustered | 10 | VC_949_0 | 10 | 0.5977 | 1 | 0.5977 | 0 | 0 | 0 | 1 | Ref |
| 3329 | KX009778 | Unassigned | Unassigned | Unassigned | 283_139 | Clustered/Singleton | 1 | VC_283_139 | 1 | 0 | 1 | 0 | 0 | 0 | 0 | 1 | Ref |
| 3330 | KX011028 | Unassigned | Unassigned | Unassigned | 149_27 | Clustered/Singleton | 1 | VC_149_27 | 1 | 0 | 1 | 0 | 0 | 0 | 0 | 1 | Ref |
| 3331 | KX011169 | Unassigned | Unassigned | Unassigned | 884_1 | Clustered | 8 | VC_884_1 | 8 | 0.1102 | 0.9989323 | 0.1101 | 0 | 0 | 0 | 0.75 | Ref |
| 3332 | KX015770 | Unassigned | Unassigned | Unassigned | 584_0 | Clustered | 28 | VC_584_0 | 28 | 0.7704 | 1 | 0.7704 | 0 | 0 | 0 | 1 | Ref |
| 3333 | KX015771 | Unassigned | Unassigned | Unassigned | 584_0 | Clustered | 28 | VC_584_0 | 28 | 0.7704 | 1 | 0.7704 | 0 | 0 | 0 | 1 | Ref |
| 3334 | KX017520 | Unassigned | Unassigned | Unassigned | 461_0 | Clustered | 96 | VC_461_0 | 96 | 0.9307 | 1 | 0.9307 | 0 | 0 | 0 | 0.9575 | Ref |
| 3335 | KX017521 | Unassigned | Unassigned | Unassigned | 191_19 | Clustered | 12 | VC_191_19 | 12 | 0.0655 | 1 | 0.0655 | 0 | 0 | 0 | 0.8636 | Ref |
| 3336 | KX058879 | Unassigned | Unassigned | Unassigned | 5_0 | Clustered | 30 | VC_5_0 | 30 | 0.9647 | 1 | 0.9647 | 0 | 0 | 0 | 1 | Ref |
| 3337 | KX077179 | Unassigned | Unassigned | Unassigned | 182_0 | Clustered | 5 | VC_182_0 | 5 | 0.6184 | 0.99999967 | 0.6184 | 0 | 0 | 0 | 1 | Ref |
| 3338 | KX077892 | Unassigned | Unassigned | Unassigned | 1187_0 | Clustered | 3 | VC_1187_0 | 3 | 0.924 | 0.92554266 | 0.8552 | 0 | 0 | 0 | 1 | Ref |
| 3339 | KX077894 | Unassigned | Unassigned | Unassigned | 317_2 | Clustered | 20 | VC_317_2 | 20 | 0.1976 | 1 | 0.1976 | 0 | 0 | 0 | 0.6842 | Ref |
| 3340 | KX077896 | Unassigned | Unassigned | Unassigned | 632_0 | Clustered | 37 | VC_632_0 | 37 | 0.581 | 1 | 0.581 | 0 | 0 | 0 | 0.9234 | Ref |
| 3341 | KX078568 | Unassigned | Unassigned | Unassigned | 390_0 | Clustered | 4 | VC_390_0 | 4 | 0.0468 | 0.9999882 | 0.0468 | 0 | 0 | 0 | 1 | Ref |
| 3342 | KX078569 | Unassigned | Unassigned | Unassigned | 288_1 | Clustered/Singleton | 1 | VC_288_1 | 1 | 0 | 1 | 0 | 0 | 0 | 0 | 1 | Ref |
| 3343 | KX098389 | Unassigned | Unassigned | Unassigned | 779_2 | Clustered | 5 | VC_779_2 | 5 | 0.0876 | 0.99999997 | 0.0876 | 0 | 0 | 0 | 1 | Ref |
| 3344 | KX098390 | Unassigned | Unassigned | Unassigned | 779_2 | Clustered | 5 | VC_779_2 | 5 | 0.0876 | 0.99999997 | 0.0876 | 0 | 0 | 0 | 1 | Ref |
| 3345 | KX098391 | Unassigned | Unassigned | Unassigned | 779_2 | Clustered | 5 | VC_779_2 | 5 | 0.0876 | 0.99999997 | 0.0876 | 0 | 0 | 0 | 1 | Ref |
| 3346 | KX119174 | Unassigned | Unassigned | Unassigned | 99_0 | Clustered | 32 | VC_99_0 | 32 | 0.8828 | 1 | 0.8828 | 0 | 0 | 0 | 1 | Ref |
| 3347 | KX119175 | Unassigned | Unassigned | Unassigned | 99_0 | Clustered | 32 | VC_99_0 | 32 | 0.8828 | 1 | 0.8828 | 0 | 0 | 0 | 1 | Ref |
| 3348 | KX119176 | Unassigned | Unassigned | Unassigned | 99_0 | Clustered | 32 | VC_99_0 | 32 | 0.8828 | 1 | 0.8828 | 0 | 0 | 0 | 1 | Ref |
| 3349 | KX119177 | Unassigned | Unassigned | Unassigned | 99_0 | Clustered | 32 | VC_99_0 | 32 | 0.8828 | 1 | 0.8828 | 0 | 0 | 0 | 1 | Ref |
| 3350 | KX119182 | Unassigned | Unassigned | Unassigned | 99_0 | Clustered | 32 | VC_99_0 | 32 | 0.8828 | 1 | 0.8828 | 0 | 0 | 0 | 1 | Ref |
| 3351 | KX119188 | Unassigned | Unassigned | Unassigned | 99_0 | Clustered | 32 | VC_99_0 | 32 | 0.8828 | 1 | 0.8828 | 0 | 0 | 0 | 1 | Ref |
| 3352 | KX119189 | Unassigned | Unassigned | Unassigned | 99_0 | Clustered | 32 | VC_99_0 | 32 | 0.8828 | 1 | 0.8828 | 0 | 0 | 0 | 1 | Ref |
| 3353 | KX119190 | Unassigned | Unassigned | Unassigned | 99_0 | Clustered | 32 | VC_99_0 | 32 | 0.8828 | 1 | 0.8828 | 0 | 0 | 0 | 1 | Ref |
| 3354 | KX119191 | Unassigned | Unassigned | Unassigned | 99_0 | Clustered | 32 | VC_99_0 | 32 | 0.8828 | 1 | 0.8828 | 0 | 0 | 0 | 1 | Ref |
| 3355 | KX119192 | Unassigned | Unassigned | Unassigned | 99_0 | Clustered | 32 | VC_99_0 | 32 | 0.8828 | 1 | 0.8828 | 0 | 0 | 0 | 1 | Ref |
| 3356 | KX119193 | Unassigned | Unassigned | Unassigned | 99_0 | Clustered | 32 | VC_99_0 | 32 | 0.8828 | 1 | 0.8828 | 0 | 0 | 0 | 1 | Ref |
| 3357 | KX119194 | Unassigned | Unassigned | Unassigned | 99_0 | Clustered | 32 | VC_99_0 | 32 | 0.8828 | 1 | 0.8828 | 0 | 0 | 0 | 1 | Ref |
| 3358 | KX119195 | Unassigned | Unassigned | Unassigned | 99_0 | Clustered | 32 | VC_99_0 | 32 | 0.8828 | 1 | 0.8828 | 0 | 0 | 0 | 1 | Ref |
| 3359 | KX119196 | Unassigned | Unassigned | Unassigned | 99_0 | Clustered | 32 | VC_99_0 | 32 | 0.8828 | 1 | 0.8828 | 0 | 0 | 0 | 1 | Ref |
| 3360 | KX119197 | Unassigned | Unassigned | Unassigned | 99_0 | Clustered | 32 | VC_99_0 | 32 | 0.8828 | 1 | 0.8828 | 0 | 0 | 0 | 1 | Ref |
| 3361 | KX119198 | Unassigned | Unassigned | Unassigned | 99_0 | Clustered | 32 | VC_99_0 | 32 | 0.8828 | 1 | 0.8828 | 0 | 0 | 0 | 1 | Ref |
| 3362 | KX119199 | Unassigned | Unassigned | Unassigned | 99_0 | Clustered | 32 | VC_99_0 | 32 | 0.8828 | 1 | 0.8828 | 0 | 0 | 0 | 1 | Ref |
| 3363 | KX119200 | Unassigned | Unassigned | Unassigned | 99_0 | Clustered | 32 | VC_99_0 | 32 | 0.8828 | 1 | 0.8828 | 0 | 0 | 0 | 1 | Ref |
| 3364 | KX119201 | Unassigned | Unassigned | Unassigned | 99_0 | Clustered | 32 | VC_99_0 | 32 | 0.8828 | 1 | 0.8828 | 0 | 0 | 0 | 1 | Ref |
| 3365 | KX119202 | Unassigned | Unassigned | Unassigned | 99_0 | Clustered | 32 | VC_99_0 | 32 | 0.8828 | 1 | 0.8828 | 0 | 0 | 0 | 1 | Ref |
| 3366 | KX119203 | Unassigned | Unassigned | Unassigned | 99_0 | Clustered | 32 | VC_99_0 | 32 | 0.8828 | 1 | 0.8828 | 0 | 0 | 0 | 1 | Ref |
| 3367 | KX119204 | Unassigned | Unassigned | Unassigned | 99_0 | Clustered | 32 | VC_99_0 | 32 | 0.8828 | 1 | 0.8828 | 0 | 0 | 0 | 1 | Ref |
| 3368 | KX119205 | Unassigned | Unassigned | Unassigned | 99_0 | Clustered | 32 | VC_99_0 | 32 | 0.8828 | 1 | 0.8828 | 0 | 0 | 0 | 1 | Ref |
| 3369 | KX119206 | Unassigned | Unassigned | Unassigned | 99_0 | Clustered | 32 | VC_99_0 | 32 | 0.8828 | 1 | 0.8828 | 0 | 0 | 0 | 1 | Ref |
| 3370 | KX129925 | Unassigned | Unassigned | Unassigned | 465_0 | Clustered | 7 | VC_465_0 | 7 | 0.3322 | 1 | 0.3322 | 0 | 0 | 0 | 1 | Ref |
| 3371 | KX130726 | Unassigned | Unassigned | Unassigned | 385_0 | Clustered | 51 | VC_385_0 | 51 | 0.8754 | 1 | 0.8754 | 0 | 0 | 0 | 1 | Ref |
| 3372 | KX130727 | Unassigned | Unassigned | Unassigned | 668_0 | Clustered | 25 | VC_668_0 | 25 | 0.5539 | 1 | 0.5539 | 0 | 0 | 0 | 0.96 | Ref |
| 3373 | KX130861 | Unassigned | Unassigned | Unassigned | 283_114 | Clustered/Singleton | 1 | VC_283_114 | 1 | 0 | 1 | 0 | 0 | 0 | 0 | 1 | Ref |
| 3374 | KX130862 | Unassigned | Unassigned | Unassigned | 283_125 | Clustered/Singleton | 1 | VC_283_125 | 1 | 0 | 1 | 0 | 0 | 0 | 0 | 1 | Ref |
| 3375 | KX130863 | Unassigned | Unassigned | Unassigned | 190_0 | Clustered | 2 | VC_190_0 | 2 | 0.0087 | 0.95763473 | 0.0083 | 0 | 0 | 0 | 1 | Ref |
| 3376 | KX130864 | Unassigned | Unassigned | Unassigned | 283_58 | Clustered/Singleton | 1 | VC_283_58 | 1 | 0 | 1 | 0 | 0 | 0 | 0 | 1 | Ref |
| 3377 | KX130865 | Unassigned | Unassigned | Unassigned | 286_16 | Clustered/Singleton | 1 | VC_286_16 | 1 | 0 | 1 | 0 | 0 | 0 | 0 | 1 | Ref |
| 3378 | KX130960 | Unassigned | Unassigned | Unassigned | 456_0 | Clustered | 62 | VC_456_0 | 62 | 0.9026 | 1 | 0.9026 | 0 | 0 | 0 | 1 | Ref |
| 3379 | KX147096 | Unassigned | Unassigned | Unassigned | 132_1 | Clustered | 17 | VC_132_1 | 17 | 0.1738 | 1 | 0.1738 | 0 | 0 | 0 | 0.9853 | Ref |
| 3380 | KX147229 | Unassigned | Unassigned | Unassigned | 884_0 | Clustered | 28 | VC_884_0 | 28 | 0.6211 | 1 | 0.6211 | 0 | 0 | 0 | 0.9841 | Ref |
| 3381 | KX156152 | Unassigned | Unassigned | Unassigned | 884_0 | Clustered | 28 | VC_884_0 | 28 | 0.6211 | 1 | 0.6211 | 0 | 0 | 0 | 0.9841 | Ref |
| 3382 | KX159477 | Unassigned | Unassigned | Unassigned | 939_0 | Clustered | 15 | VC_939_0 | 15 | 1 | 0 | 0 | 0 | 0 | 0 | 1 | Ref |
| 3383 | KX160204 | Unassigned | Unassigned | Unassigned | 262_0 | Clustered | 5 | VC_262_0 | 5 | 0.0724 | 0.99999989 | 0.0724 | 0 | 0 | 0 | 1 | Ref |
| 3384 | KX160206 | Unassigned | Unassigned | Unassigned | 268_0 | Clustered | 7 | VC_268_0 | 7 | 0.3427 | 1 | 0.3427 | 0 | 0 | 0 | 1 | Ref |
| 3385 | KX160209 | Unassigned | Unassigned | Unassigned | 268_0 | Clustered | 7 | VC_268_0 | 7 | 0.3427 | 1 | 0.3427 | 0 | 0 | 0 | 1 | Ref |
| 3386 | KX160210 | Unassigned | Unassigned | Unassigned | 268_0 | Clustered | 7 | VC_268_0 | 7 | 0.3427 | 1 | 0.3427 | 0 | 0 | 0 | 1 | Ref |
| 3387 | KX160211 | Unassigned | Unassigned | Unassigned | 268_0 | Clustered | 7 | VC_268_0 | 7 | 0.3427 | 1 | 0.3427 | 0 | 0 | 0 | 1 | Ref |
| 3388 | KX160219 | Unassigned | Unassigned | Unassigned | 262_0 | Clustered | 5 | VC_262_0 | 5 | 0.0724 | 0.99999989 | 0.0724 | 0 | 0 | 0 | 1 | Ref |
| 3389 | KX160220 | Unassigned | Unassigned | Unassigned | 262_0 | Clustered | 5 | VC_262_0 | 5 | 0.0724 | 0.99999989 | 0.0724 | 0 | 0 | 0 | 1 | Ref |
| 3390 | KX171208 | Unassigned | Unassigned | Unassigned | 79_0 | Clustered | 64 | VC_79_0 | 64 | 0.4185 | 1 | 0.4185 | 0 | 0 | 0 | 0.7649 | Ref |
| 3391 | KX171209 | Unassigned | Unassigned | Unassigned | 838_0 | Clustered | 29 | VC_838_0 | 29 | 0.8611 | 1 | 0.8611 | 0 | 0 | 0 | 1 | Ref |
| 3392 | KX171210 | Unassigned | Unassigned | Unassigned | 838_0 | Clustered | 29 | VC_838_0 | 29 | 0.8611 | 1 | 0.8611 | 0 | 0 | 0 | 1 | Ref |
| 3393 | KX171211 | Unassigned | Unassigned | Unassigned | 133_9 | Clustered | 3 | VC_133_9 | 3 | 0.0099 | 0.98457109 | 0.0098 | 0 | 0 | 0 | 1 | Ref |
| 3394 | KX171212 | Unassigned | Unassigned | Unassigned | 151_0 | Clustered | 6 | VC_151_0 | 6 | 0.0735 | 1 | 0.0735 | 0 | 0 | 0 | 1 | Ref |
| 3395 | KX171213 | Unassigned | Unassigned | Unassigned | 151_0 | Clustered | 6 | VC_151_0 | 6 | 0.0735 | 1 | 0.0735 | 0 | 0 | 0 | 1 | Ref |
| 3396 | KX179905 | Unassigned | Unassigned | Unassigned | 52_0 | Clustered | 87 | VC_52_0 | 87 | 0.9858 | 1 | 0.9858 | 0 | 0 | 0 | 1 | Ref |
| 3397 | KX181651 | Unassigned | Unassigned | Unassigned | 177_0 | Clustered | 5 | VC_177_0 | 5 | 0.7468 | 0.99885646 | 0.746 | 0 | 0 | 0 | 1 | Ref |
| 3398 | KX190833 | Unassigned | Unassigned | Unassigned | 731_0 | Clustered | 2 | VC_731_0 | 2 | 0.3697 | 0.94979793 | 0.3512 | 0 | 0 | 0 | 1 | Ref |
| 3399 | KX190834 | Unassigned | Unassigned | Unassigned | 829_0 | Clustered | 5 | VC_829_0 | 5 | 0.8821 | 0.99999832 | 0.8821 | 0 | 0 | 0 | 1 | Ref |
| 3400 | KX190835 | Unassigned | Unassigned | Unassigned | 20_0 | Clustered | 5 | VC_20_0 | 5 | 0.4387 | 0.99999934 | 0.4387 | 0 | 0 | 0 | 1 | Ref |

| | | | | | | | | | | | | | | | | | |
|---|---|---|---|---|---|---|---|---|---|---|---|---|---|---|---|---|---|
| 3401 | KX196154 | Unassigned | Unassigned | Unassigned | 498_0 | Clustered | 8 | VC_498_0 | 8 | 0.3987 | 1 | 0.3987 | 0 | 0 | 0 | 1 | Ref |
| 3402 | KX198613 | Unassigned | Unassigned | Unassigned | 166_0 | Clustered | 26 | VC_166_0 | 26 | 0.7923 | 1 | 0.7923 | 0 | 0 | 0 | 1 | Ref |
| 3403 | KX198614 | Unassigned | Unassigned | Unassigned | 614_0 | Clustered | 5 | VC_614_0 | 5 | 0.2395 | 0.99999997 | 0.2395 | 0 | 0 | 0 | 1 | Ref |
| 3404 | KX198615 | Unassigned | Unassigned | Unassigned | 904_1 | Clustered | 4 | VC_904_1 | 4 | 0.1486 | 0.99997723 | 0.1486 | 0 | 0 | 0 | 1 | Ref |
| 3405 | KX211991 | Unassigned | Unassigned | Unassigned | 111_0 | Clustered | 91 | VC_111_0 | 91 | 0.9587 | 1 | 0.9587 | 0 | 0 | 0 | 1 | Ref |
| 3406 | KX223815 | Unassigned | Unassigned | Unassigned | 634_1 | Clustered | 6 | VC_634_1 | 6 | 0.4228 | 1 | 0.4228 | 0 | 0 | 0 | 1 | Ref |
| 3407 | KX227757 | Unassigned | Unassigned | Unassigned | 654_0 | Clustered | 4 | VC_654_0 | 4 | 0.2897 | 0.99998263 | 0.2897 | 0 | 0 | 0 | 1 | Ref |
| 3408 | KX227758 | Unassigned | Unassigned | Unassigned | 652_0 | Clustered | 4 | VC_652_0 | 4 | 0.4743 | 0.99998076 | 0.4743 | 0 | 0 | 0 | 1 | Ref |
| 3409 | KX227759 | Unassigned | Unassigned | Unassigned | 652_0 | Clustered | 4 | VC_652_0 | 4 | 0.4743 | 0.99998076 | 0.4743 | 0 | 0 | 0 | 1 | Ref |
| 3410 | KX227760 | Unassigned | Unassigned | Unassigned | 654_0 | Clustered | 4 | VC_654_0 | 4 | 0.2897 | 0.99998263 | 0.2897 | 0 | 0 | 0 | 1 | Ref |
| 3411 | KX229736 | Unassigned | Unassigned | Unassigned | 844_0 | Clustered | 27 | VC_844_0 | 27 | 0.9385 | 1 | 0.9385 | 0 | 0 | 0 | 1 | Ref |
| 3412 | KX231828 | Unassigned | Unassigned | Unassigned | 125_0 | Clustered | 17 | VC_125_0 | 17 | 0.9244 | 1 | 0.9244 | 0 | 0 | 0 | 1 | Ref |
| 3413 | KX231829 | Unassigned | Unassigned | Unassigned | 476_0 | Clustered | 19 | VC_476_0 | 19 | 0.7814 | 1 | 0.7814 | 0 | 0 | 0 | 1 | Ref |
| 3414 | KX232515 | Unassigned | Unassigned | Unassigned | 7_0 | Clustered | 28 | VC_7_0 | 28 | 0.3916 | 1 | 0.3916 | 0 | 0 | 0 | 0.9947 | Ref |
| 3415 | KX236333 | Unassigned | Unassigned | Unassigned | 844_0 | Clustered | 27 | VC_844_0 | 27 | 0.9385 | 1 | 0.9385 | 0 | 0 | 0 | 1 | Ref |
| 3416 | KX237514 | Unassigned | Unassigned | Unassigned | 111_0 | Clustered | 91 | VC_111_0 | 91 | 0.9587 | 1 | 0.9587 | 0 | 0 | 0 | 1 | Ref |
| 3417 | KX237515 | Unassigned | Unassigned | Unassigned | 937_0 | Clustered | 40 | VC_937_0 | 40 | 0.6809 | 1 | 0.6809 | 0 | 0 | 0 | 0.9359 | Ref |
| 3418 | KX237516 | Unassigned | Unassigned | Unassigned | 123_0 | Clustered | 23 | VC_123_0 | 23 | 0.817 | 1 | 0.817 | 0 | 0 | 0 | 1 | Ref |
| 3419 | KX241618 | Unassigned | Unassigned | Unassigned | 169_0 | Clustered | 20 | VC_169_0 | 20 | 0.7529 | 1 | 0.7529 | 0 | 0 | 0 | 1 | Ref |
| 3420 | KX245012 | Unassigned | Unassigned | Unassigned | 133_30 | Clustered | 2 | VC_133_30 | 2 | 0.0052 | 0.87738871 | 0.0046 | 0 | 0 | 0 | 1 | Ref |
| 3421 | KX245013 | Unassigned | Unassigned | Unassigned | 637_0 | Clustered | 115 | VC_637_0 | 115 | 0.731 | 1 | 0.731 | 0 | 0 | 0 | 0.8764 | Ref |
| 3422 | KX245890 | Unassigned | Unassigned | Unassigned | 669_1 | Clustered | 10 | VC_669_1 | 10 | 0.1268 | 1 | 0.1268 | 0 | 0 | 0 | 0.9778 | Ref |
| 3423 | KX257490 | Unassigned | Unassigned | Unassigned | 1026_0 | Clustered | 2 | VC_1026_0 | 2 | 0.2977 | 0.81445332 | 0.2425 | 0 | 0 | 0 | 1 | Ref |
| 3424 | KX258185 | Unassigned | Unassigned | Unassigned | 491_0 | Clustered | 36 | VC_491_0 | 36 | 0.5336 | 1 | 0.5336 | 0 | 0 | 0 | 0.9714 | Ref |
| 3425 | KX266300 | Unassigned | Unassigned | Unassigned | 360_0 | Clustered | 358 | VC_360_0 | 358 | 0.9615 | 1 | 0.9615 | 0 | 0 | 0 | 1 | Ref |
| 3426 | KX266301 | Unassigned | Unassigned | Unassigned | 360_0 | Clustered | 358 | VC_360_0 | 358 | 0.9615 | 1 | 0.9615 | 0 | 0 | 0 | 1 | Ref |
| 3427 | KX266302 | Unassigned | Unassigned | Unassigned | 360_0 | Clustered | 358 | VC_360_0 | 358 | 0.9615 | 1 | 0.9615 | 0 | 0 | 0 | 1 | Ref |
| 3428 | KX266303 | Unassigned | Unassigned | Unassigned | 360_0 | Clustered | 358 | VC_360_0 | 358 | 0.9615 | 1 | 0.9615 | 0 | 0 | 0 | 1 | Ref |
| 3429 | KX266304 | Unassigned | Unassigned | Unassigned | 360_0 | Clustered | 358 | VC_360_0 | 358 | 0.9615 | 1 | 0.9615 | 0 | 0 | 0 | 1 | Ref |
| 3430 | KX266305 | Unassigned | Unassigned | Unassigned | 360_0 | Clustered | 358 | VC_360_0 | 358 | 0.9615 | 1 | 0.9615 | 0 | 0 | 0 | 1 | Ref |
| 3431 | KX266306 | Unassigned | Unassigned | Unassigned | 360_0 | Clustered | 358 | VC_360_0 | 358 | 0.9615 | 1 | 0.9615 | 0 | 0 | 0 | 1 | Ref |
| 3432 | KX266307 | Unassigned | Unassigned | Unassigned | 360_0 | Clustered | 358 | VC_360_0 | 358 | 0.9615 | 1 | 0.9615 | 0 | 0 | 0 | 1 | Ref |
| 3433 | KX266308 | Unassigned | Unassigned | Unassigned | 360_0 | Clustered | 358 | VC_360_0 | 358 | 0.9615 | 1 | 0.9615 | 0 | 0 | 0 | 1 | Ref |
| 3434 | KX266309 | Unassigned | Unassigned | Unassigned | 360_0 | Clustered | 358 | VC_360_0 | 358 | 0.9615 | 1 | 0.9615 | 0 | 0 | 0 | 1 | Ref |
| 3435 | KX266310 | Unassigned | Unassigned | Unassigned | 360_0 | Clustered | 358 | VC_360_0 | 358 | 0.9615 | 1 | 0.9615 | 0 | 0 | 0 | 1 | Ref |
| 3436 | KX266311 | Unassigned | Unassigned | Unassigned | 360_0 | Clustered | 358 | VC_360_0 | 358 | 0.9615 | 1 | 0.9615 | 0 | 0 | 0 | 1 | Ref |
| 3437 | KX266312 | Unassigned | Unassigned | Unassigned | 360_0 | Clustered | 358 | VC_360_0 | 358 | 0.9615 | 1 | 0.9615 | 0 | 0 | 0 | 1 | Ref |
| 3438 | KX266313 | Unassigned | Unassigned | Unassigned | 360_0 | Clustered | 358 | VC_360_0 | 358 | 0.9615 | 1 | 0.9615 | 0 | 0 | 0 | 1 | Ref |
| 3439 | KX266314 | Unassigned | Unassigned | Unassigned | 360_0 | Clustered | 358 | VC_360_0 | 358 | 0.9615 | 1 | 0.9615 | 0 | 0 | 0 | 1 | Ref |
| 3440 | KX266315 | Unassigned | Unassigned | Unassigned | 360_0 | Clustered | 358 | VC_360_0 | 358 | 0.9615 | 1 | 0.9615 | 0 | 0 | 0 | 1 | Ref |
| 3441 | KX266316 | Unassigned | Unassigned | Unassigned | 360_0 | Clustered | 358 | VC_360_0 | 358 | 0.9615 | 1 | 0.9615 | 0 | 0 | 0 | 1 | Ref |
| 3442 | KX266317 | Unassigned | Unassigned | Unassigned | 360_0 | Clustered | 358 | VC_360_0 | 358 | 0.9615 | 1 | 0.9615 | 0 | 0 | 0 | 1 | Ref |
| 3443 | KX266318 | Unassigned | Unassigned | Unassigned | 360_0 | Clustered | 358 | VC_360_0 | 358 | 0.9615 | 1 | 0.9615 | 0 | 0 | 0 | 1 | Ref |
| 3444 | KX266319 | Unassigned | Unassigned | Unassigned | 360_0 | Clustered | 358 | VC_360_0 | 358 | 0.9615 | 1 | 0.9615 | 0 | 0 | 0 | 1 | Ref |
| 3445 | KX266320 | Unassigned | Unassigned | Unassigned | 360_0 | Clustered | 358 | VC_360_0 | 358 | 0.9615 | 1 | 0.9615 | 0 | 0 | 0 | 1 | Ref |
| 3446 | KX266321 | Unassigned | Unassigned | Unassigned | 360_0 | Clustered | 358 | VC_360_0 | 358 | 0.9615 | 1 | 0.9615 | 0 | 0 | 0 | 1 | Ref |
| 3447 | KX266322 | Unassigned | Unassigned | Unassigned | 360_0 | Clustered | 358 | VC_360_0 | 358 | 0.9615 | 1 | 0.9615 | 0 | 0 | 0 | 1 | Ref |
| 3448 | KX266323 | Unassigned | Unassigned | Unassigned | 360_0 | Clustered | 358 | VC_360_0 | 358 | 0.9615 | 1 | 0.9615 | 0 | 0 | 0 | 1 | Ref |
| 3449 | KX266324 | Unassigned | Unassigned | Unassigned | 360_0 | Clustered | 358 | VC_360_0 | 358 | 0.9615 | 1 | 0.9615 | 0 | 0 | 0 | 1 | Ref |
| 3450 | KX266325 | Unassigned | Unassigned | Unassigned | 360_0 | Clustered | 358 | VC_360_0 | 358 | 0.9615 | 1 | 0.9615 | 0 | 0 | 0 | 1 | Ref |
| 3451 | KX266326 | Unassigned | Unassigned | Unassigned | 360_0 | Clustered | 358 | VC_360_0 | 358 | 0.9615 | 1 | 0.9615 | 0 | 0 | 0 | 1 | Ref |
| 3452 | KX266327 | Unassigned | Unassigned | Unassigned | 360_0 | Clustered | 358 | VC_360_0 | 358 | 0.9615 | 1 | 0.9615 | 0 | 0 | 0 | 1 | Ref |
| 3453 | KX266328 | Unassigned | Unassigned | Unassigned | 360_0 | Clustered | 358 | VC_360_0 | 358 | 0.9615 | 1 | 0.9615 | 0 | 0 | 0 | 1 | Ref |
| 3454 | KX266329 | Unassigned | Unassigned | Unassigned | 360_0 | Clustered | 358 | VC_360_0 | 358 | 0.9615 | 1 | 0.9615 | 0 | 0 | 0 | 1 | Ref |
| 3455 | KX266330 | Unassigned | Unassigned | Unassigned | 360_0 | Clustered | 358 | VC_360_0 | 358 | 0.9615 | 1 | 0.9615 | 0 | 0 | 0 | 1 | Ref |
| 3456 | KX266331 | Unassigned | Unassigned | Unassigned | 360_0 | Clustered | 358 | VC_360_0 | 358 | 0.9615 | 1 | 0.9615 | 0 | 0 | 0 | 1 | Ref |
| 3457 | KX266332 | Unassigned | Unassigned | Unassigned | 360_0 | Clustered | 358 | VC_360_0 | 358 | 0.9615 | 1 | 0.9615 | 0 | 0 | 0 | 1 | Ref |
| 3458 | KX266333 | Unassigned | Unassigned | Unassigned | 360_0 | Clustered | 358 | VC_360_0 | 358 | 0.9615 | 1 | 0.9615 | 0 | 0 | 0 | 1 | Ref |
| 3459 | KX266334 | Unassigned | Unassigned | Unassigned | 360_0 | Clustered | 358 | VC_360_0 | 358 | 0.9615 | 1 | 0.9615 | 0 | 0 | 0 | 1 | Ref |
| 3460 | KX266335 | Unassigned | Unassigned | Unassigned | 360_0 | Clustered | 358 | VC_360_0 | 358 | 0.9615 | 1 | 0.9615 | 0 | 0 | 0 | 1 | Ref |
| 3461 | KX266336 | Unassigned | Unassigned | Unassigned | 360_0 | Clustered | 358 | VC_360_0 | 358 | 0.9615 | 1 | 0.9615 | 0 | 0 | 0 | 1 | Ref |
| 3462 | KX266337 | Unassigned | Unassigned | Unassigned | 360_0 | Clustered | 358 | VC_360_0 | 358 | 0.9615 | 1 | 0.9615 | 0 | 0 | 0 | 1 | Ref |
| 3463 | KX266338 | Unassigned | Unassigned | Unassigned | 360_0 | Clustered | 358 | VC_360_0 | 358 | 0.9615 | 1 | 0.9615 | 0 | 0 | 0 | 1 | Ref |
| 3464 | KX266339 | Unassigned | Unassigned | Unassigned | 360_0 | Clustered | 358 | VC_360_0 | 358 | 0.9615 | 1 | 0.9615 | 0 | 0 | 0 | 1 | Ref |
| 3465 | KX266340 | Unassigned | Unassigned | Unassigned | 360_0 | Clustered | 358 | VC_360_0 | 358 | 0.9615 | 1 | 0.9615 | 0 | 0 | 0 | 1 | Ref |
| 3466 | KX266341 | Unassigned | Unassigned | Unassigned | 360_0 | Clustered | 358 | VC_360_0 | 358 | 0.9615 | 1 | 0.9615 | 0 | 0 | 0 | 1 | Ref |
| 3467 | KX266342 | Unassigned | Unassigned | Unassigned | 360_0 | Clustered | 358 | VC_360_0 | 358 | 0.9615 | 1 | 0.9615 | 0 | 0 | 0 | 1 | Ref |
| 3468 | KX266343 | Unassigned | Unassigned | Unassigned | 360_0 | Clustered | 358 | VC_360_0 | 358 | 0.9615 | 1 | 0.9615 | 0 | 0 | 0 | 1 | Ref |
| 3469 | KX266344 | Unassigned | Unassigned | Unassigned | 360_0 | Clustered | 358 | VC_360_0 | 358 | 0.9615 | 1 | 0.9615 | 0 | 0 | 0 | 1 | Ref |
| 3470 | KX266345 | Unassigned | Unassigned | Unassigned | 360_0 | Clustered | 358 | VC_360_0 | 358 | 0.9615 | 1 | 0.9615 | 0 | 0 | 0 | 1 | Ref |
| 3471 | KX266346 | Unassigned | Unassigned | Unassigned | 360_0 | Clustered | 358 | VC_360_0 | 358 | 0.9615 | 1 | 0.9615 | 0 | 0 | 0 | 1 | Ref |
| 3472 | KX266347 | Unassigned | Unassigned | Unassigned | 360_0 | Clustered | 358 | VC_360_0 | 358 | 0.9615 | 1 | 0.9615 | 0 | 0 | 0 | 1 | Ref |
| 3473 | KX266348 | Unassigned | Unassigned | Unassigned | 360_0 | Clustered | 358 | VC_360_0 | 358 | 0.9615 | 1 | 0.9615 | 0 | 0 | 0 | 1 | Ref |
| 3474 | KX266349 | Unassigned | Unassigned | Unassigned | 360_0 | Clustered | 358 | VC_360_0 | 358 | 0.9615 | 1 | 0.9615 | 0 | 0 | 0 | 1 | Ref |
| 3475 | KX266350 | Unassigned | Unassigned | Unassigned | 360_0 | Clustered | 358 | VC_360_0 | 358 | 0.9615 | 1 | 0.9615 | 0 | 0 | 0 | 1 | Ref |
| 3476 | KX266351 | Unassigned | Unassigned | Unassigned | 360_0 | Clustered | 358 | VC_360_0 | 358 | 0.9615 | 1 | 0.9615 | 0 | 0 | 0 | 1 | Ref |
| 3477 | KX266352 | Unassigned | Unassigned | Unassigned | 360_0 | Clustered | 358 | VC_360_0 | 358 | 0.9615 | 1 | 0.9615 | 0 | 0 | 0 | 1 | Ref |
| 3478 | KX266353 | Unassigned | Unassigned | Unassigned | 360_0 | Clustered | 358 | VC_360_0 | 358 | 0.9615 | 1 | 0.9615 | 0 | 0 | 0 | 1 | Ref |
| 3479 | KX266354 | Unassigned | Unassigned | Unassigned | 360_0 | Clustered | 358 | VC_360_0 | 358 | 0.9615 | 1 | 0.9615 | 0 | 0 | 0 | 1 | Ref |
| 3480 | KX266355 | Unassigned | Unassigned | Unassigned | 360_0 | Clustered | 358 | VC_360_0 | 358 | 0.9615 | 1 | 0.9615 | 0 | 0 | 0 | 1 | Ref |
| 3481 | KX266356 | Unassigned | Unassigned | Unassigned | 360_0 | Clustered | 358 | VC_360_0 | 358 | 0.9615 | 1 | 0.9615 | 0 | 0 | 0 | 1 | Ref |
| 3482 | KX266357 | Unassigned | Unassigned | Unassigned | 360_0 | Clustered | 358 | VC_360_0 | 358 | 0.9615 | 1 | 0.9615 | 0 | 0 | 0 | 1 | Ref |
| 3483 | KX266358 | Unassigned | Unassigned | Unassigned | 360_0 | Clustered | 358 | VC_360_0 | 358 | 0.9615 | 1 | 0.9615 | 0 | 0 | 0 | 1 | Ref |
| 3484 | KX266359 | Unassigned | Unassigned | Unassigned | 360_0 | Clustered | 358 | VC_360_0 | 358 | 0.9615 | 1 | 0.9615 | 0 | 0 | 0 | 1 | Ref |
| 3485 | KX266360 | Unassigned | Unassigned | Unassigned | 360_0 | Clustered | 358 | VC_360_0 | 358 | 0.9615 | 1 | 0.9615 | 0 | 0 | 0 | 1 | Ref |
| 3486 | KX266361 | Unassigned | Unassigned | Unassigned | 360_0 | Clustered | 358 | VC_360_0 | 358 | 0.9615 | 1 | 0.9615 | 0 | 0 | 0 | 1 | Ref |
| 3487 | KX266362 | Unassigned | Unassigned | Unassigned | 360_0 | Clustered | 358 | VC_360_0 | 358 | 0.9615 | 1 | 0.9615 | 0 | 0 | 0 | 1 | Ref |
| 3488 | KX266363 | Unassigned | Unassigned | Unassigned | 360_0 | Clustered | 358 | VC_360_0 | 358 | 0.9615 | 1 | 0.9615 | 0 | 0 | 0 | 1 | Ref |
| 3489 | KX266364 | Unassigned | Unassigned | Unassigned | 360_0 | Clustered | 358 | VC_360_0 | 358 | 0.9615 | 1 | 0.9615 | 0 | 0 | 0 | 1 | Ref |
| 3490 | KX266365 | Unassigned | Unassigned | Unassigned | 360_0 | Clustered | 358 | VC_360_0 | 358 | 0.9615 | 1 | 0.9615 | 0 | 0 | 0 | 1 | Ref |
| 3491 | KX266366 | Unassigned | Unassigned | Unassigned | 360_0 | Clustered | 358 | VC_360_0 | 358 | 0.9615 | 1 | 0.9615 | 0 | 0 | 0 | 1 | Ref |
| 3492 | KX266367 | Unassigned | Unassigned | Unassigned | 360_0 | Clustered | 358 | VC_360_0 | 358 | 0.9615 | 1 | 0.9615 | 0 | 0 | 0 | 1 | Ref |
| 3493 | KX266368 | Unassigned | Unassigned | Unassigned | 360_0 | Clustered | 358 | VC_360_0 | 358 | 0.9615 | 1 | 0.9615 | 0 | 0 | 0 | 1 | Ref |
| 3494 | KX266369 | Unassigned | Unassigned | Unassigned | 360_0 | Clustered | 358 | VC_360_0 | 358 | 0.9615 | 1 | 0.9615 | 0 | 0 | 0 | 1 | Ref |
| 3495 | KX266370 | Unassigned | Unassigned | Unassigned | 360_0 | Clustered | 358 | VC_360_0 | 358 | 0.9615 | 1 | 0.9615 | 0 | 0 | 0 | 1 | Ref |
| 3496 | KX266371 | Unassigned | Unassigned | Unassigned | 360_0 | Clustered | 358 | VC_360_0 | 358 | 0.9615 | 1 | 0.9615 | 0 | 0 | 0 | 1 | Ref |
| 3497 | KX266372 | Unassigned | Unassigned | Unassigned | 360_0 | Clustered | 358 | VC_360_0 | 358 | 0.9615 | 1 | 0.9615 | 0 | 0 | 0 | 1 | Ref |
| 3498 | KX266373 | Unassigned | Unassigned | Unassigned | 360_0 | Clustered | 358 | VC_360_0 | 358 | 0.9615 | 1 | 0.9615 | 0 | 0 | 0 | 1 | Ref |
| 3499 | KX266374 | Unassigned | Unassigned | Unassigned | 360_0 | Clustered | 358 | VC_360_0 | 358 | 0.9615 | 1 | 0.9615 | 0 | 0 | 0 | 1 | Ref |
| 3500 | KX266375 | Unassigned | Unassigned | Unassigned | 360_0 | Clustered | 358 | VC_360_0 | 358 | 0.9615 | 1 | 0.9615 | 0 | 0 | 0 | 1 | Ref |
| 3501 | KX266376 | Unassigned | Unassigned | Unassigned | 360_0 | Clustered | 358 | VC_360_0 | 358 | 0.9615 | 1 | 0.9615 | 0 | 0 | 0 | 1 | Ref |
| 3502 | KX266377 | Unassigned | Unassigned | Unassigned | 360_0 | Clustered | 358 | VC_360_0 | 358 | 0.9615 | 1 | 0.9615 | 0 | 0 | 0 | 1 | Ref |
| 3503 | KX266378 | Unassigned | Unassigned | Unassigned | 360_0 | Clustered | 358 | VC_360_0 | 358 | 0.9615 | 1 | 0.9615 | 0 | 0 | 0 | 1 | Ref |
| 3504 | KX266379 | Unassigned | Unassigned | Unassigned | 360_0 | Clustered | 358 | VC_360_0 | 358 | 0.9615 | 1 | 0.9615 | 0 | 0 | 0 | 1 | Ref |
| 3505 | KX266380 | Unassigned | Unassigned | Unassigned | 360_0 | Clustered | 358 | VC_360_0 | 358 | 0.9615 | 1 | 0.9615 | 0 | 0 | 0 | 1 | Ref |
| 3506 | KX266381 | Unassigned | Unassigned | Unassigned | 360_0 | Clustered | 358 | VC_360_0 | 358 | 0.9615 | 1 | 0.9615 | 0 | 0 | 0 | 1 | Ref |
| 3507 | KX266382 | Unassigned | Unassigned | Unassigned | 360_0 | Clustered | 358 | VC_360_0 | 358 | 0.9615 | 1 | 0.9615 | 0 | 0 | 0 | 1 | Ref |
| 3508 | KX266383 | Unassigned | Unassigned | Unassigned | 360_0 | Clustered | 358 | VC_360_0 | 358 | 0.9615 | 1 | 0.9615 | 0 | 0 | 0 | 1 | Ref |
| 3509 | KX266384 | Unassigned | Unassigned | Unassigned | 360_0 | Clustered | 358 | VC_360_0 | 358 | 0.9615 | 1 | 0.9615 | 0 | 0 | 0 | 1 | Ref |
| 3510 | KX266385 | Unassigned | Unassigned | Unassigned | 360_0 | Clustered | 358 | VC_360_0 | 358 | 0.9615 | 1 | 0.9615 | 0 | 0 | 0 | 1 | Ref |
| 3511 | KX266386 | Unassigned | Unassigned | Unassigned | 360_0 | Clustered | 358 | VC_360_0 | 358 | 0.9615 | 1 | 0.9615 | 0 | 0 | 0 | 1 | Ref |
| 3512 | KX266387 | Unassigned | Unassigned | Unassigned | 360_0 | Clustered | 358 | VC_360_0 | 358 | 0.9615 | 1 | 0.9615 | 0 | 0 | 0 | 1 | Ref |
| 3513 | KX266388 | Unassigned | Unassigned | Unassigned | 360_0 | Clustered | 358 | VC_360_0 | 358 | 0.9615 | 1 | 0.9615 | 0 | 0 | 0 | 1 | Ref |
| 3514 | KX266389 | Unassigned | Unassigned | Unassigned | 360_0 | Clustered | 358 | VC_360_0 | 358 | 0.9615 | 1 | 0.9615 | 0 | 0 | 0 | 1 | Ref |
| 3515 | KX266390 | Unassigned | Unassigned | Unassigned | 360_0 | Clustered | 358 | VC_360_0 | 358 | 0.9615 | 1 | 0.9615 | 0 | 0 | 0 | 1 | Ref |
| 3516 | KX266391 | Unassigned | Unassigned | Unassigned | 360_0 | Clustered | 358 | VC_360_0 | 358 | 0.9615 | 1 | 0.9615 | 0 | 0 | 0 | 1 | Ref |
| 3517 | KX266392 | Unassigned | Unassigned | Unassigned | 360_0 | Clustered | 358 | VC_360_0 | 358 | 0.9615 | 1 | 0.9615 | 0 | 0 | 0 | 1 | Ref |
| 3518 | KX266393 | Unassigned | Unassigned | Unassigned | 360_0 | Clustered | 358 | VC_360_0 | 358 | 0.9615 | 1 | 0.9615 | 0 | 0 | 0 | 1 | Ref |
| 3519 | KX266394 | Unassigned | Unassigned | Unassigned | 360_0 | Clustered | 358 | VC_360_0 | 358 | 0.9615 | 1 | 0.9615 | 0 | 0 | 0 | 1 | Ref |
| 3520 | KX266395 | Unassigned | Unassigned | Unassigned | 360_0 | Clustered | 358 | VC_360_0 | 358 | 0.9615 | 1 | 0.9615 | 0 | 0 | 0 | 1 | Ref |
| 3521 | KX266396 | Unassigned | Unassigned | Unassigned | 360_0 | Clustered | 358 | VC_360_0 | 358 | 0.9615 | 1 | 0.9615 | 0 | 0 | 0 | 1 | Ref |
| 3522 | KX266397 | Unassigned | Unassigned | Unassigned | 360_0 | Clustered | 358 | VC_360_0 | 358 | 0.9615 | 1 | 0.9615 | 0 | 0 | 0 | 1 | Ref |
| 3523 | KX266398 | Unassigned | Unassigned | Unassigned | 360_0 | Clustered | 358 | VC_360_0 | 358 | 0.9615 | 1 | 0.9615 | 0 | 0 | 0 | 1 | Ref |
| 3524 | KX266399 | Unassigned | Unassigned | Unassigned | 360_0 | Clustered | 358 | VC_360_0 | 358 | 0.9615 | 1 | 0.9615 | 0 | 0 | 0 | 1 | Ref |
| 3525 | KX266400 | Unassigned | Unassigned | Unassigned | 360_0 | Clustered | 358 | VC_360_0 | 358 | 0.9615 | 1 | 0.9615 | 0 | 0 | 0 | 1 | Ref |
| 3526 | KX266401 | Unassigned | Unassigned | Unassigned | 360_0 | Clustered | 358 | VC_360_0 | 358 | 0.9615 | 1 | 0.9615 | 0 | 0 | 0 | 1 | Ref |
| 3527 | KX266402 | Unassigned | Unassigned | Unassigned | 360_0 | Clustered | 358 | VC_360_0 | 358 | 0.9615 | 1 | 0.9615 | 0 | 0 | 0 | 1 | Ref |
| 3528 | KX266403 | Unassigned | Unassigned | Unassigned | 360_0 | Clustered | 358 | VC_360_0 | 358 | 0.9615 | 1 | 0.9615 | 0 | 0 | 0 | 1 | Ref |
| 3529 | KX266404 | Unassigned | Unassigned | Unassigned | 360_0 | Clustered | 358 | VC_360_0 | 358 | 0.9615 | 1 | 0.9615 | 0 | 0 | 0 | 1 | Ref |
| 3530 | KX266405 | Unassigned | Unassigned | Unassigned | 360_0 | Clustered | 358 | VC_360_0 | 358 | 0.9615 | 1 | 0.9615 | 0 | 0 | 0 | 1 | Ref |
| 3531 | KX266406 | Unassigned | Unassigned | Unassigned | 360_0 | Clustered | 358 | VC_360_0 | 358 | 0.9615 | 1 | 0.9615 | 0 | 0 | 0 | 1 | Ref |
| 3532 | KX266407 | Unassigned | Unassigned | Unassigned | 360_0 | Clustered | 358 | VC_360_0 | 358 | 0.9615 | 1 | 0.9615 | 0 | 0 | 0 | 1 | Ref |
| 3533 | KX266408 | Unassigned | Unassigned | Unassigned | 360_0 | Clustered | 358 | VC_360_0 | 358 | 0.9615 | 1 | 0.9615 | 0 | 0 | 0 | 1 | Ref |
| 3534 | KX266409 | Unassigned | Unassigned | Unassigned | 360_0 | Clustered | 358 | VC_360_0 | 358 | 0.9615 | 1 | 0.9615 | 0 | 0 | 0 | 1 | Ref |
| 3535 | KX266410 | Unassigned | Unassigned | Unassigned | 360_0 | Clustered | 358 | VC_360_0 | 358 | 0.9615 | 1 | 0.9615 | 0 | 0 | 0 | 1 | Ref |
| 3536 | KX266411 | Unassigned | Unassigned | Unassigned | 360_0 | Clustered | 358 | VC_360_0 | 358 | 0.9615 | 1 | 0.9615 | 0 | 0 | 0 | 1 | Ref |
| 3537 | KX266412 | Unassigned | Unassigned | Unassigned | 360_0 | Clustered | 358 | VC_360_0 | 358 | 0.9615 | 1 | 0.9615 | 0 | 0 | 0 | 1 | Ref |
| 3538 | KX266413 | Unassigned | Unassigned | Unassigned | 360_0 | Clustered | 358 | VC_360_0 | 358 | 0.9615 | 1 | 0.9615 | 0 | 0 | 0 | 1 | Ref |
| 3539 | KX266414 | Unassigned | Unassigned | Unassigned | 360_0 | Clustered | 358 | VC_360_0 | 358 | 0.9615 | 1 | 0.9615 | 0 | 0 | 0 | 1 | Ref |
| 3540 | KX266415 | Unassigned | Unassigned | Unassigned | 360_0 | Clustered | 358 | VC_360_0 | 358 | 0.9615 | 1 | 0.9615 | 0 | 0 | 0 | 1 | Ref |
| 3541 | KX266416 | Unassigned | Unassigned | Unassigned | 360_0 | Clustered | 358 | VC_360_0 | 358 | 0.9615 | 1 | 0.9615 | 0 | 0 | 0 | 1 | Ref |
| 3542 | KX266417 | Unassigned | Unassigned | Unassigned | 360_0 | Clustered | 358 | VC_360_0 | 358 | 0.9615 | 1 | 0.9615 | 0 | 0 | 0 | 1 | Ref |
| 3543 | KX266418 | Unassigned | Unassigned | Unassigned | 360_0 | Clustered | 358 | VC_360_0 | 358 | 0.9615 | 1 | 0.9615 | 0 | 0 | 0 | 1 | Ref |
| 3544 | KX266419 | Unassigned | Unassigned | Unassigned | 360_0 | Clustered | 358 | VC_360_0 | 358 | 0.9615 | 1 | 0.9615 | 0 | 0 | 0 | 1 | Ref |
| 3545 | KX266420 | Unassigned | Unassigned | Unassigned | 360_0 | Clustered | 358 | VC_360_0 | 358 | 0.9615 | 1 | 0.9615 | 0 | 0 | 0 | 1 | Ref |
| 3546 | KX266421 | Unassigned | Unassigned | Unassigned | 360_0 | Clustered | 358 | VC_360_0 | 358 | 0.9615 | 1 | 0.9615 | 0 | 0 | 0 | 1 | Ref |
| 3547 | KX266422 | Unassigned | Unassigned | Unassigned | 360_0 | Clustered | 358 | VC_360_0 | 358 | 0.9615 | 1 | 0.9615 | 0 | 0 | 0 | 1 | Ref |
| 3548 | KX266423 | Unassigned | Unassigned | Unassigned | 360_0 | Clustered | 358 | VC_360_0 | 358 | 0.9615 | 1 | 0.9615 | 0 | 0 | 0 | 1 | Ref |
| 3549 | KX266424 | Unassigned | Unassigned | Unassigned | 360_0 | Clustered | 358 | VC_360_0 | 358 | 0.9615 | 1 | 0.9615 | 0 | 0 | 0 | 1 | Ref |
| 3550 | KX266425 | Unassigned | Unassigned | Unassigned | 360_0 | Clustered | 358 | VC_360_0 | 358 | 0.9615 | 1 | 0.9615 | 0 | 0 | 0 | 1 | Ref |
| 3551 | KX266426 | Unassigned | Unassigned | Unassigned | 360_0 | Clustered | 358 | VC_360_0 | 358 | 0.9615 | 1 | 0.9615 | 0 | 0 | 0 | 1 | Ref |
| 3552 | KX266427 | Unassigned | Unassigned | Unassigned | 360_0 | Clustered | 358 | VC_360_0 | 358 | 0.9615 | 1 | 0.9615 | 0 | 0 | 0 | 1 | Ref |
| 3553 | KX266428 | Unassigned | Unassigned | Unassigned | 360_0 | Clustered | 358 | VC_360_0 | 358 | 0.9615 | 1 | 0.9615 | 0 | 0 | 0 | 1 | Ref |
| 3554 | KX266429 | Unassigned | Unassigned | Unassigned | 360_0 | Clustered | 358 | VC_360_0 | 358 | 0.9615 | 1 | 0.9615 | 0 | 0 | 0 | 1 | Ref |
| 3555 | KX266430 | Unassigned | Unassigned | Unassigned | 360_0 | Clustered | 358 | VC_360_0 | 358 | 0.9615 | 1 | 0.9615 | 0 | 0 | 0 | 1 | Ref |
| 3556 | KX266431 | Unassigned | Unassigned | Unassigned | 360_0 | Clustered | 358 | VC_360_0 | 358 | 0.9615 | 1 | 0.9615 | 0 | 0 | 0 | 1 | Ref |
| 3557 | KX266432 | Unassigned | Unassigned | Unassigned | 360_0 | Clustered | 358 | VC_360_0 | 358 | 0.9615 | 1 | 0.9615 | 0 | 0 | 0 | 1 | Ref |
| 3558 | KX266433 | Unassigned | Unassigned | Unassigned | 360_0 | Clustered | 358 | VC_360_0 | 358 | 0.9615 | 1 | 0.9615 | 0 | 0 | 0 | 1 | Ref |
| 3559 | KX266434 | Unassigned | Unassigned | Unassigned | 360_0 | Clustered | 358 | VC_360_0 | 358 | 0.9615 | 1 | 0.9615 | 0 | 0 | 0 | 1 | Ref |
| 3560 | KX266435 | Unassigned | Unassigned | Unassigned | 360_0 | Clustered | 358 | VC_360_0 | 358 | 0.9615 | 1 | 0.9615 | 0 | 0 | 0 | 1 | Ref |
| 3561 | KX266436 | Unassigned | Unassigned | Unassigned | 360_0 | Clustered | 358 | VC_360_0 | 358 | 0.9615 | 1 | 0.9615 | 0 | 0 | 0 | 1 | Ref |
| 3562 | KX266437 | Unassigned | Unassigned | Unassigned | 360_0 | Clustered | 358 | VC_360_0 | 358 | 0.9615 | 1 | 0.9615 | 0 | 0 | 0 | 1 | Ref |

| | | | | | | | | | | | | | | | | | |
|---|---|---|---|---|---|---|---|---|---|---|---|---|---|---|---|---|---|
| 3563 | KX266438 | Unassigned | Unassigned | Unassigned | 360_0 | Clustered | 358 | VC_360_0 | 358 | 0.9615 | 1 | 0.9615 | 0 | 0 | 0 | 1 | Ref |
| 3564 | KX266439 | Unassigned | Unassigned | Unassigned | 360_0 | Clustered | 358 | VC_360_0 | 358 | 0.9615 | 1 | 0.9615 | 0 | 0 | 0 | 1 | Ref |
| 3565 | KX266440 | Unassigned | Unassigned | Unassigned | 360_0 | Clustered | 358 | VC_360_0 | 358 | 0.9615 | 1 | 0.9615 | 0 | 0 | 0 | 1 | Ref |
| 3566 | KX266441 | Unassigned | Unassigned | Unassigned | 360_0 | Clustered | 358 | VC_360_0 | 358 | 0.9615 | 1 | 0.9615 | 0 | 0 | 0 | 1 | Ref |
| 3567 | KX266442 | Unassigned | Unassigned | Unassigned | 360_0 | Clustered | 358 | VC_360_0 | 358 | 0.9615 | 1 | 0.9615 | 0 | 0 | 0 | 1 | Ref |
| 3568 | KX266443 | Unassigned | Unassigned | Unassigned | 360_0 | Clustered | 358 | VC_360_0 | 358 | 0.9615 | 1 | 0.9615 | 0 | 0 | 0 | 1 | Ref |
| 3569 | KX266444 | Unassigned | Unassigned | Unassigned | 360_0 | Clustered | 358 | VC_360_0 | 358 | 0.9615 | 1 | 0.9615 | 0 | 0 | 0 | 1 | Ref |
| 3570 | KX266445 | Unassigned | Unassigned | Unassigned | 360_0 | Clustered | 358 | VC_360_0 | 358 | 0.9615 | 1 | 0.9615 | 0 | 0 | 0 | 1 | Ref |
| 3571 | KX266446 | Unassigned | Unassigned | Unassigned | 360_0 | Clustered | 358 | VC_360_0 | 358 | 0.9615 | 1 | 0.9615 | 0 | 0 | 0 | 1 | Ref |
| 3572 | KX266447 | Unassigned | Unassigned | Unassigned | 360_0 | Clustered | 358 | VC_360_0 | 358 | 0.9615 | 1 | 0.9615 | 0 | 0 | 0 | 1 | Ref |
| 3573 | KX266448 | Unassigned | Unassigned | Unassigned | 360_0 | Clustered | 358 | VC_360_0 | 358 | 0.9615 | 1 | 0.9615 | 0 | 0 | 0 | 1 | Ref |
| 3574 | KX266449 | Unassigned | Unassigned | Unassigned | 360_0 | Clustered | 358 | VC_360_0 | 358 | 0.9615 | 1 | 0.9615 | 0 | 0 | 0 | 1 | Ref |
| 3575 | KX266450 | Unassigned | Unassigned | Unassigned | 360_0 | Clustered | 358 | VC_360_0 | 358 | 0.9615 | 1 | 0.9615 | 0 | 0 | 0 | 1 | Ref |
| 3576 | KX266451 | Unassigned | Unassigned | Unassigned | 360_0 | Clustered | 358 | VC_360_0 | 358 | 0.9615 | 1 | 0.9615 | 0 | 0 | 0 | 1 | Ref |
| 3577 | KX266452 | Unassigned | Unassigned | Unassigned | 360_0 | Clustered | 358 | VC_360_0 | 358 | 0.9615 | 1 | 0.9615 | 0 | 0 | 0 | 1 | Ref |
| 3578 | KX266453 | Unassigned | Unassigned | Unassigned | 360_0 | Clustered | 358 | VC_360_0 | 358 | 0.9615 | 1 | 0.9615 | 0 | 0 | 0 | 1 | Ref |
| 3579 | KX266454 | Unassigned | Unassigned | Unassigned | 360_0 | Clustered | 358 | VC_360_0 | 358 | 0.9615 | 1 | 0.9615 | 0 | 0 | 0 | 1 | Ref |
| 3580 | KX266455 | Unassigned | Unassigned | Unassigned | 360_0 | Clustered | 358 | VC_360_0 | 358 | 0.9615 | 1 | 0.9615 | 0 | 0 | 0 | 1 | Ref |
| 3581 | KX266456 | Unassigned | Unassigned | Unassigned | 360_0 | Clustered | 358 | VC_360_0 | 358 | 0.9615 | 1 | 0.9615 | 0 | 0 | 0 | 1 | Ref |
| 3582 | KX266457 | Unassigned | Unassigned | Unassigned | 360_0 | Clustered | 358 | VC_360_0 | 358 | 0.9615 | 1 | 0.9615 | 0 | 0 | 0 | 1 | Ref |
| 3583 | KX266458 | Unassigned | Unassigned | Unassigned | 360_0 | Clustered | 358 | VC_360_0 | 358 | 0.9615 | 1 | 0.9615 | 0 | 0 | 0 | 1 | Ref |
| 3584 | KX266459 | Unassigned | Unassigned | Unassigned | 360_0 | Clustered | 358 | VC_360_0 | 358 | 0.9615 | 1 | 0.9615 | 0 | 0 | 0 | 1 | Ref |
| 3585 | KX266460 | Unassigned | Unassigned | Unassigned | 360_0 | Clustered | 358 | VC_360_0 | 358 | 0.9615 | 1 | 0.9615 | 0 | 0 | 0 | 1 | Ref |
| 3586 | KX266461 | Unassigned | Unassigned | Unassigned | 360_0 | Clustered | 358 | VC_360_0 | 358 | 0.9615 | 1 | 0.9615 | 0 | 0 | 0 | 1 | Ref |
| 3587 | KX266462 | Unassigned | Unassigned | Unassigned | 360_0 | Clustered | 358 | VC_360_0 | 358 | 0.9615 | 1 | 0.9615 | 0 | 0 | 0 | 1 | Ref |
| 3588 | KX266463 | Unassigned | Unassigned | Unassigned | 360_0 | Clustered | 358 | VC_360_0 | 358 | 0.9615 | 1 | 0.9615 | 0 | 0 | 0 | 1 | Ref |
| 3589 | KX266464 | Unassigned | Unassigned | Unassigned | 360_0 | Clustered | 358 | VC_360_0 | 358 | 0.9615 | 1 | 0.9615 | 0 | 0 | 0 | 1 | Ref |
| 3590 | KX266465 | Unassigned | Unassigned | Unassigned | 360_0 | Clustered | 358 | VC_360_0 | 358 | 0.9615 | 1 | 0.9615 | 0 | 0 | 0 | 1 | Ref |
| 3591 | KX266466 | Unassigned | Unassigned | Unassigned | 360_0 | Clustered | 358 | VC_360_0 | 358 | 0.9615 | 1 | 0.9615 | 0 | 0 | 0 | 1 | Ref |
| 3592 | KX266467 | Unassigned | Unassigned | Unassigned | 360_0 | Clustered | 358 | VC_360_0 | 358 | 0.9615 | 1 | 0.9615 | 0 | 0 | 0 | 1 | Ref |
| 3593 | KX266468 | Unassigned | Unassigned | Unassigned | 360_0 | Clustered | 358 | VC_360_0 | 358 | 0.9615 | 1 | 0.9615 | 0 | 0 | 0 | 1 | Ref |
| 3594 | KX266469 | Unassigned | Unassigned | Unassigned | 360_0 | Clustered | 358 | VC_360_0 | 358 | 0.9615 | 1 | 0.9615 | 0 | 0 | 0 | 1 | Ref |
| 3595 | KX266470 | Unassigned | Unassigned | Unassigned | 360_0 | Clustered | 358 | VC_360_0 | 358 | 0.9615 | 1 | 0.9615 | 0 | 0 | 0 | 1 | Ref |
| 3596 | KX266471 | Unassigned | Unassigned | Unassigned | 360_0 | Clustered | 358 | VC_360_0 | 358 | 0.9615 | 1 | 0.9615 | 0 | 0 | 0 | 1 | Ref |
| 3597 | KX266472 | Unassigned | Unassigned | Unassigned | 360_0 | Clustered | 358 | VC_360_0 | 358 | 0.9615 | 1 | 0.9615 | 0 | 0 | 0 | 1 | Ref |
| 3598 | KX266473 | Unassigned | Unassigned | Unassigned | 360_0 | Clustered | 358 | VC_360_0 | 358 | 0.9615 | 1 | 0.9615 | 0 | 0 | 0 | 1 | Ref |
| 3599 | KX266474 | Unassigned | Unassigned | Unassigned | 360_0 | Clustered | 358 | VC_360_0 | 358 | 0.9615 | 1 | 0.9615 | 0 | 0 | 0 | 1 | Ref |
| 3600 | KX266475 | Unassigned | Unassigned | Unassigned | 360_0 | Clustered | 358 | VC_360_0 | 358 | 0.9615 | 1 | 0.9615 | 0 | 0 | 0 | 1 | Ref |
| 3601 | KX266476 | Unassigned | Unassigned | Unassigned | 360_0 | Clustered | 358 | VC_360_0 | 358 | 0.9615 | 1 | 0.9615 | 0 | 0 | 0 | 1 | Ref |
| 3602 | KX266477 | Unassigned | Unassigned | Unassigned | 360_0 | Clustered | 358 | VC_360_0 | 358 | 0.9615 | 1 | 0.9615 | 0 | 0 | 0 | 1 | Ref |
| 3603 | KX266478 | Unassigned | Unassigned | Unassigned | 360_0 | Clustered | 358 | VC_360_0 | 358 | 0.9615 | 1 | 0.9615 | 0 | 0 | 0 | 1 | Ref |
| 3604 | KX266479 | Unassigned | Unassigned | Unassigned | 360_0 | Clustered | 358 | VC_360_0 | 358 | 0.9615 | 1 | 0.9615 | 0 | 0 | 0 | 1 | Ref |
| 3605 | KX266480 | Unassigned | Unassigned | Unassigned | 360_0 | Clustered | 358 | VC_360_0 | 358 | 0.9615 | 1 | 0.9615 | 0 | 0 | 0 | 1 | Ref |
| 3606 | KX266481 | Unassigned | Unassigned | Unassigned | 360_0 | Clustered | 358 | VC_360_0 | 358 | 0.9615 | 1 | 0.9615 | 0 | 0 | 0 | 1 | Ref |
| 3607 | KX266482 | Unassigned | Unassigned | Unassigned | 360_0 | Clustered | 358 | VC_360_0 | 358 | 0.9615 | 1 | 0.9615 | 0 | 0 | 0 | 1 | Ref |
| 3608 | KX266483 | Unassigned | Unassigned | Unassigned | 360_0 | Clustered | 358 | VC_360_0 | 358 | 0.9615 | 1 | 0.9615 | 0 | 0 | 0 | 1 | Ref |
| 3609 | KX266484 | Unassigned | Unassigned | Unassigned | 360_0 | Clustered | 358 | VC_360_0 | 358 | 0.9615 | 1 | 0.9615 | 0 | 0 | 0 | 1 | Ref |
| 3610 | KX266485 | Unassigned | Unassigned | Unassigned | 360_0 | Clustered | 358 | VC_360_0 | 358 | 0.9615 | 1 | 0.9615 | 0 | 0 | 0 | 1 | Ref |
| 3611 | KX266486 | Unassigned | Unassigned | Unassigned | 360_0 | Clustered | 358 | VC_360_0 | 358 | 0.9615 | 1 | 0.9615 | 0 | 0 | 0 | 1 | Ref |
| 3612 | KX266487 | Unassigned | Unassigned | Unassigned | 360_0 | Clustered | 358 | VC_360_0 | 358 | 0.9615 | 1 | 0.9615 | 0 | 0 | 0 | 1 | Ref |
| 3613 | KX266488 | Unassigned | Unassigned | Unassigned | 360_0 | Clustered | 358 | VC_360_0 | 358 | 0.9615 | 1 | 0.9615 | 0 | 0 | 0 | 1 | Ref |
| 3614 | KX266489 | Unassigned | Unassigned | Unassigned | 360_0 | Clustered | 358 | VC_360_0 | 358 | 0.9615 | 1 | 0.9615 | 0 | 0 | 0 | 1 | Ref |
| 3615 | KX266490 | Unassigned | Unassigned | Unassigned | 360_0 | Clustered | 358 | VC_360_0 | 358 | 0.9615 | 1 | 0.9615 | 0 | 0 | 0 | 1 | Ref |
| 3616 | KX266491 | Unassigned | Unassigned | Unassigned | 360_0 | Clustered | 358 | VC_360_0 | 358 | 0.9615 | 1 | 0.9615 | 0 | 0 | 0 | 1 | Ref |
| 3617 | KX266492 | Unassigned | Unassigned | Unassigned | 360_0 | Clustered | 358 | VC_360_0 | 358 | 0.9615 | 1 | 0.9615 | 0 | 0 | 0 | 1 | Ref |
| 3618 | KX266493 | Unassigned | Unassigned | Unassigned | 360_0 | Clustered | 358 | VC_360_0 | 358 | 0.9615 | 1 | 0.9615 | 0 | 0 | 0 | 1 | Ref |
| 3619 | KX266494 | Unassigned | Unassigned | Unassigned | 360_0 | Clustered | 358 | VC_360_0 | 358 | 0.9615 | 1 | 0.9615 | 0 | 0 | 0 | 1 | Ref |
| 3620 | KX266495 | Unassigned | Unassigned | Unassigned | 360_0 | Clustered | 358 | VC_360_0 | 358 | 0.9615 | 1 | 0.9615 | 0 | 0 | 0 | 1 | Ref |
| 3621 | KX266496 | Unassigned | Unassigned | Unassigned | 360_0 | Clustered | 358 | VC_360_0 | 358 | 0.9615 | 1 | 0.9615 | 0 | 0 | 0 | 1 | Ref |
| 3622 | KX266497 | Unassigned | Unassigned | Unassigned | 360_0 | Clustered | 358 | VC_360_0 | 358 | 0.9615 | 1 | 0.9615 | 0 | 0 | 0 | 1 | Ref |
| 3623 | KX266498 | Unassigned | Unassigned | Unassigned | 360_0 | Clustered | 358 | VC_360_0 | 358 | 0.9615 | 1 | 0.9615 | 0 | 0 | 0 | 1 | Ref |
| 3624 | KX266499 | Unassigned | Unassigned | Unassigned | 360_0 | Clustered | 358 | VC_360_0 | 358 | 0.9615 | 1 | 0.9615 | 0 | 0 | 0 | 1 | Ref |
| 3625 | KX266500 | Unassigned | Unassigned | Unassigned | 360_0 | Clustered | 358 | VC_360_0 | 358 | 0.9615 | 1 | 0.9615 | 0 | 0 | 0 | 1 | Ref |
| 3626 | KX266501 | Unassigned | Unassigned | Unassigned | 360_0 | Clustered | 358 | VC_360_0 | 358 | 0.9615 | 1 | 0.9615 | 0 | 0 | 0 | 1 | Ref |
| 3627 | KX266502 | Unassigned | Unassigned | Unassigned | 360_0 | Clustered | 358 | VC_360_0 | 358 | 0.9615 | 1 | 0.9615 | 0 | 0 | 0 | 1 | Ref |
| 3628 | KX266503 | Unassigned | Unassigned | Unassigned | 360_0 | Clustered | 358 | VC_360_0 | 358 | 0.9615 | 1 | 0.9615 | 0 | 0 | 0 | 1 | Ref |
| 3629 | KX266504 | Unassigned | Unassigned | Unassigned | 360_0 | Clustered | 358 | VC_360_0 | 358 | 0.9615 | 1 | 0.9615 | 0 | 0 | 0 | 1 | Ref |
| 3630 | KX266505 | Unassigned | Unassigned | Unassigned | 360_0 | Clustered | 358 | VC_360_0 | 358 | 0.9615 | 1 | 0.9615 | 0 | 0 | 0 | 1 | Ref |
| 3631 | KX266506 | Unassigned | Unassigned | Unassigned | 360_0 | Clustered | 358 | VC_360_0 | 358 | 0.9615 | 1 | 0.9615 | 0 | 0 | 0 | 1 | Ref |
| 3632 | KX266507 | Unassigned | Unassigned | Unassigned | 360_0 | Clustered | 358 | VC_360_0 | 358 | 0.9615 | 1 | 0.9615 | 0 | 0 | 0 | 1 | Ref |
| 3633 | KX266508 | Unassigned | Unassigned | Unassigned | 360_0 | Clustered | 358 | VC_360_0 | 358 | 0.9615 | 1 | 0.9615 | 0 | 0 | 0 | 1 | Ref |
| 3634 | KX266509 | Unassigned | Unassigned | Unassigned | 360_0 | Clustered | 358 | VC_360_0 | 358 | 0.9615 | 1 | 0.9615 | 0 | 0 | 0 | 1 | Ref |
| 3635 | KX266510 | Unassigned | Unassigned | Unassigned | 360_0 | Clustered | 358 | VC_360_0 | 358 | 0.9615 | 1 | 0.9615 | 0 | 0 | 0 | 1 | Ref |
| 3636 | KX266511 | Unassigned | Unassigned | Unassigned | 360_0 | Clustered | 358 | VC_360_0 | 358 | 0.9615 | 1 | 0.9615 | 0 | 0 | 0 | 1 | Ref |
| 3637 | KX266512 | Unassigned | Unassigned | Unassigned | 360_0 | Clustered | 358 | VC_360_0 | 358 | 0.9615 | 1 | 0.9615 | 0 | 0 | 0 | 1 | Ref |
| 3638 | KX266513 | Unassigned | Unassigned | Unassigned | 360_0 | Clustered | 358 | VC_360_0 | 358 | 0.9615 | 1 | 0.9615 | 0 | 0 | 0 | 1 | Ref |
| 3639 | KX266514 | Unassigned | Unassigned | Unassigned | 360_0 | Clustered | 358 | VC_360_0 | 358 | 0.9615 | 1 | 0.9615 | 0 | 0 | 0 | 1 | Ref |
| 3640 | KX266515 | Unassigned | Unassigned | Unassigned | 360_0 | Clustered | 358 | VC_360_0 | 358 | 0.9615 | 1 | 0.9615 | 0 | 0 | 0 | 1 | Ref |
| 3641 | KX266516 | Unassigned | Unassigned | Unassigned | 360_0 | Clustered | 358 | VC_360_0 | 358 | 0.9615 | 1 | 0.9615 | 0 | 0 | 0 | 1 | Ref |
| 3642 | KX266517 | Unassigned | Unassigned | Unassigned | 360_0 | Clustered | 358 | VC_360_0 | 358 | 0.9615 | 1 | 0.9615 | 0 | 0 | 0 | 1 | Ref |
| 3643 | KX266518 | Unassigned | Unassigned | Unassigned | 360_0 | Clustered | 358 | VC_360_0 | 358 | 0.9615 | 1 | 0.9615 | 0 | 0 | 0 | 1 | Ref |
| 3644 | KX266519 | Unassigned | Unassigned | Unassigned | 360_0 | Clustered | 358 | VC_360_0 | 358 | 0.9615 | 1 | 0.9615 | 0 | 0 | 0 | 1 | Ref |
| 3645 | KX266520 | Unassigned | Unassigned | Unassigned | 360_0 | Clustered | 358 | VC_360_0 | 358 | 0.9615 | 1 | 0.9615 | 0 | 0 | 0 | 1 | Ref |
| 3646 | KX266521 | Unassigned | Unassigned | Unassigned | 360_0 | Clustered | 358 | VC_360_0 | 358 | 0.9615 | 1 | 0.9615 | 0 | 0 | 0 | 1 | Ref |
| 3647 | KX266522 | Unassigned | Unassigned | Unassigned | 360_0 | Clustered | 358 | VC_360_0 | 358 | 0.9615 | 1 | 0.9615 | 0 | 0 | 0 | 1 | Ref |
| 3648 | KX266523 | Unassigned | Unassigned | Unassigned | 360_0 | Clustered | 358 | VC_360_0 | 358 | 0.9615 | 1 | 0.9615 | 0 | 0 | 0 | 1 | Ref |
| 3649 | KX266524 | Unassigned | Unassigned | Unassigned | 360_0 | Clustered | 358 | VC_360_0 | 358 | 0.9615 | 1 | 0.9615 | 0 | 0 | 0 | 1 | Ref |
| 3650 | KX266525 | Unassigned | Unassigned | Unassigned | 360_0 | Clustered | 358 | VC_360_0 | 358 | 0.9615 | 1 | 0.9615 | 0 | 0 | 0 | 1 | Ref |
| 3651 | KX266526 | Unassigned | Unassigned | Unassigned | 360_0 | Clustered | 358 | VC_360_0 | 358 | 0.9615 | 1 | 0.9615 | 0 | 0 | 0 | 1 | Ref |
| 3652 | KX266527 | Unassigned | Unassigned | Unassigned | 360_0 | Clustered | 358 | VC_360_0 | 358 | 0.9615 | 1 | 0.9615 | 0 | 0 | 0 | 1 | Ref |
| 3653 | KX266528 | Unassigned | Unassigned | Unassigned | 360_0 | Clustered | 358 | VC_360_0 | 358 | 0.9615 | 1 | 0.9615 | 0 | 0 | 0 | 1 | Ref |
| 3654 | KX266529 | Unassigned | Unassigned | Unassigned | 360_0 | Clustered | 358 | VC_360_0 | 358 | 0.9615 | 1 | 0.9615 | 0 | 0 | 0 | 1 | Ref |
| 3655 | KX266530 | Unassigned | Unassigned | Unassigned | 360_0 | Clustered | 358 | VC_360_0 | 358 | 0.9615 | 1 | 0.9615 | 0 | 0 | 0 | 1 | Ref |
| 3656 | KX266531 | Unassigned | Unassigned | Unassigned | 360_0 | Clustered | 358 | VC_360_0 | 358 | 0.9615 | 1 | 0.9615 | 0 | 0 | 0 | 1 | Ref |
| 3657 | KX266532 | Unassigned | Unassigned | Unassigned | 360_0 | Clustered | 358 | VC_360_0 | 358 | 0.9615 | 1 | 0.9615 | 0 | 0 | 0 | 1 | Ref |
| 3658 | KX266533 | Unassigned | Unassigned | Unassigned | 360_0 | Clustered | 358 | VC_360_0 | 358 | 0.9615 | 1 | 0.9615 | 0 | 0 | 0 | 1 | Ref |
| 3659 | KX266534 | Unassigned | Unassigned | Unassigned | 360_0 | Clustered | 358 | VC_360_0 | 358 | 0.9615 | 1 | 0.9615 | 0 | 0 | 0 | 1 | Ref |
| 3660 | KX266535 | Unassigned | Unassigned | Unassigned | 360_0 | Clustered | 358 | VC_360_0 | 358 | 0.9615 | 1 | 0.9615 | 0 | 0 | 0 | 1 | Ref |
| 3661 | KX266536 | Unassigned | Unassigned | Unassigned | 360_0 | Clustered | 358 | VC_360_0 | 358 | 0.9615 | 1 | 0.9615 | 0 | 0 | 0 | 1 | Ref |
| 3662 | KX266537 | Unassigned | Unassigned | Unassigned | 360_0 | Clustered | 358 | VC_360_0 | 358 | 0.9615 | 1 | 0.9615 | 0 | 0 | 0 | 1 | Ref |
| 3663 | KX266538 | Unassigned | Unassigned | Unassigned | 360_0 | Clustered | 358 | VC_360_0 | 358 | 0.9615 | 1 | 0.9615 | 0 | 0 | 0 | 1 | Ref |
| 3664 | KX266539 | Unassigned | Unassigned | Unassigned | 360_0 | Clustered | 358 | VC_360_0 | 358 | 0.9615 | 1 | 0.9615 | 0 | 0 | 0 | 1 | Ref |
| 3665 | KX266540 | Unassigned | Unassigned | Unassigned | 360_0 | Clustered | 358 | VC_360_0 | 358 | 0.9615 | 1 | 0.9615 | 0 | 0 | 0 | 1 | Ref |
| 3666 | KX266541 | Unassigned | Unassigned | Unassigned | 360_0 | Clustered | 358 | VC_360_0 | 358 | 0.9615 | 1 | 0.9615 | 0 | 0 | 0 | 1 | Ref |
| 3667 | KX266542 | Unassigned | Unassigned | Unassigned | 360_0 | Clustered | 358 | VC_360_0 | 358 | 0.9615 | 1 | 0.9615 | 0 | 0 | 0 | 1 | Ref |
| 3668 | KX266543 | Unassigned | Unassigned | Unassigned | 360_0 | Clustered | 358 | VC_360_0 | 358 | 0.9615 | 1 | 0.9615 | 0 | 0 | 0 | 1 | Ref |
| 3669 | KX266544 | Unassigned | Unassigned | Unassigned | 360_0 | Clustered | 358 | VC_360_0 | 358 | 0.9615 | 1 | 0.9615 | 0 | 0 | 0 | 1 | Ref |
| 3670 | KX266545 | Unassigned | Unassigned | Unassigned | 360_0 | Clustered | 358 | VC_360_0 | 358 | 0.9615 | 1 | 0.9615 | 0 | 0 | 0 | 1 | Ref |
| 3671 | KX266546 | Unassigned | Unassigned | Unassigned | 360_0 | Clustered | 358 | VC_360_0 | 358 | 0.9615 | 1 | 0.9615 | 0 | 0 | 0 | 1 | Ref |
| 3672 | KX266547 | Unassigned | Unassigned | Unassigned | 360_0 | Clustered | 358 | VC_360_0 | 358 | 0.9615 | 1 | 0.9615 | 0 | 0 | 0 | 1 | Ref |
| 3673 | KX266548 | Unassigned | Unassigned | Unassigned | 360_0 | Clustered | 358 | VC_360_0 | 358 | 0.9615 | 1 | 0.9615 | 0 | 0 | 0 | 1 | Ref |
| 3674 | KX266549 | Unassigned | Unassigned | Unassigned | 360_0 | Clustered | 358 | VC_360_0 | 358 | 0.9615 | 1 | 0.9615 | 0 | 0 | 0 | 1 | Ref |
| 3675 | KX266550 | Unassigned | Unassigned | Unassigned | 360_0 | Clustered | 358 | VC_360_0 | 358 | 0.9615 | 1 | 0.9615 | 0 | 0 | 0 | 1 | Ref |
| 3676 | KX266551 | Unassigned | Unassigned | Unassigned | 360_0 | Clustered | 358 | VC_360_0 | 358 | 0.9615 | 1 | 0.9615 | 0 | 0 | 0 | 1 | Ref |
| 3677 | KX266552 | Unassigned | Unassigned | Unassigned | 360_0 | Clustered | 358 | VC_360_0 | 358 | 0.9615 | 1 | 0.9615 | 0 | 0 | 0 | 1 | Ref |
| 3678 | KX266553 | Unassigned | Unassigned | Unassigned | 360_0 | Clustered | 358 | VC_360_0 | 358 | 0.9615 | 1 | 0.9615 | 0 | 0 | 0 | 1 | Ref |
| 3679 | KX266554 | Unassigned | Unassigned | Unassigned | 360_0 | Clustered | 358 | VC_360_0 | 358 | 0.9615 | 1 | 0.9615 | 0 | 0 | 0 | 1 | Ref |
| 3680 | KX266555 | Unassigned | Unassigned | Unassigned | 360_0 | Clustered | 358 | VC_360_0 | 358 | 0.9615 | 1 | 0.9615 | 0 | 0 | 0 | 1 | Ref |
| 3681 | KX266556 | Unassigned | Unassigned | Unassigned | 360_0 | Clustered | 358 | VC_360_0 | 358 | 0.9615 | 1 | 0.9615 | 0 | 0 | 0 | 1 | Ref |
| 3682 | KX266557 | Unassigned | Unassigned | Unassigned | 360_0 | Clustered | 358 | VC_360_0 | 358 | 0.9615 | 1 | 0.9615 | 0 | 0 | 0 | 1 | Ref |
| 3683 | KX266558 | Unassigned | Unassigned | Unassigned | 360_0 | Clustered | 358 | VC_360_0 | 358 | 0.9615 | 1 | 0.9615 | 0 | 0 | 0 | 1 | Ref |
| 3684 | KX266559 | Unassigned | Unassigned | Unassigned | 360_0 | Clustered | 358 | VC_360_0 | 358 | 0.9615 | 1 | 0.9615 | 0 | 0 | 0 | 1 | Ref |
| 3685 | KX266560 | Unassigned | Unassigned | Unassigned | 360_0 | Clustered | 358 | VC_360_0 | 358 | 0.9615 | 1 | 0.9615 | 0 | 0 | 0 | 1 | Ref |
| 3686 | KX266561 | Unassigned | Unassigned | Unassigned | 360_0 | Clustered | 358 | VC_360_0 | 358 | 0.9615 | 1 | 0.9615 | 0 | 0 | 0 | 1 | Ref |
| 3687 | KX266562 | Unassigned | Unassigned | Unassigned | 360_0 | Clustered | 358 | VC_360_0 | 358 | 0.9615 | 1 | 0.9615 | 0 | 0 | 0 | 1 | Ref |
| 3688 | KX266563 | Unassigned | Unassigned | Unassigned | 360_0 | Clustered | 358 | VC_360_0 | 358 | 0.9615 | 1 | 0.9615 | 0 | 0 | 0 | 1 | Ref |
| 3689 | KX266564 | Unassigned | Unassigned | Unassigned | 360_0 | Clustered | 358 | VC_360_0 | 358 | 0.9615 | 1 | 0.9615 | 0 | 0 | 0 | 1 | Ref |
| 3690 | KX266565 | Unassigned | Unassigned | Unassigned | 360_0 | Clustered | 358 | VC_360_0 | 358 | 0.9615 | 1 | 0.9615 | 0 | 0 | 0 | 1 | Ref |
| 3691 | KX266566 | Unassigned | Unassigned | Unassigned | 360_0 | Clustered | 358 | VC_360_0 | 358 | 0.9615 | 1 | 0.9615 | 0 | 0 | 0 | 1 | Ref |
| 3692 | KX266567 | Unassigned | Unassigned | Unassigned | 360_0 | Clustered | 358 | VC_360_0 | 358 | 0.9615 | 1 | 0.9615 | 0 | 0 | 0 | 1 | Ref |
| 3693 | KX266568 | Unassigned | Unassigned | Unassigned | 360_0 | Clustered | 358 | VC_360_0 | 358 | 0.9615 | 1 | 0.9615 | 0 | 0 | 0 | 1 | Ref |
| 3694 | KX266569 | Unassigned | Unassigned | Unassigned | 360_0 | Clustered | 358 | VC_360_0 | 358 | 0.9615 | 1 | 0.9615 | 0 | 0 | 0 | 1 | Ref |
| 3695 | KX266570 | Unassigned | Unassigned | Unassigned | 360_0 | Clustered | 358 | VC_360_0 | 358 | 0.9615 | 1 | 0.9615 | 0 | 0 | 0 | 1 | Ref |
| 3696 | KX266571 | Unassigned | Unassigned | Unassigned | 360_0 | Clustered | 358 | VC_360_0 | 358 | 0.9615 | 1 | 0.9615 | 0 | 0 | 0 | 1 | Ref |
| 3697 | KX266572 | Unassigned | Unassigned | Unassigned | 360_0 | Clustered | 358 | VC_360_0 | 358 | 0.9615 | 1 | 0.9615 | 0 | 0 | 0 | 1 | Ref |
| 3698 | KX266573 | Unassigned | Unassigned | Unassigned | 360_0 | Clustered | 358 | VC_360_0 | 358 | 0.9615 | 1 | 0.9615 | 0 | 0 | 0 | 1 | Ref |
| 3699 | KX266574 | Unassigned | Unassigned | Unassigned | 360_0 | Clustered | 358 | VC_360_0 | 358 | 0.9615 | 1 | 0.9615 | 0 | 0 | 0 | 1 | Ref |
| 3700 | KX266575 | Unassigned | Unassigned | Unassigned | 360_0 | Clustered | 358 | VC_360_0 | 358 | 0.9615 | 1 | 0.9615 | 0 | 0 | 0 | 1 | Ref |
| 3701 | KX266576 | Unassigned | Unassigned | Unassigned | 360_0 | Clustered | 358 | VC_360_0 | 358 | 0.9615 | 1 | 0.9615 | 0 | 0 | 0 | 1 | Ref |
| 3702 | KX266577 | Unassigned | Unassigned | Unassigned | 360_0 | Clustered | 358 | VC_360_0 | 358 | 0.9615 | 1 | 0.9615 | 0 | 0 | 0 | 1 | Ref |
| 3703 | KX266578 | Unassigned | Unassigned | Unassigned | 360_0 | Clustered | 358 | VC_360_0 | 358 | 0.9615 | 1 | 0.9615 | 0 | 0 | 0 | 1 | Ref |
| 3704 | KX266579 | Unassigned | Unassigned | Unassigned | 360_0 | Clustered | 358 | VC_360_0 | 358 | 0.9615 | 1 | 0.9615 | 0 | 0 | 0 | 1 | Ref |
| 3705 | KX266580 | Unassigned | Unassigned | Unassigned | 360_0 | Clustered | 358 | VC_360_0 | 358 | 0.9615 | 1 | 0.9615 | 0 | 0 | 0 | 1 | Ref |
| 3706 | KX266581 | Unassigned | Unassigned | Unassigned | 360_0 | Clustered | 358 | VC_360_0 | 358 | 0.9615 | 1 | 0.9615 | 0 | 0 | 0 | 1 | Ref |
| 3707 | KX266582 | Unassigned | Unassigned | Unassigned | 360_0 | Clustered | 358 | VC_360_0 | 358 | 0.9615 | 1 | 0.9615 | 0 | 0 | 0 | 1 | Ref |
| 3708 | KX266583 | Unassigned | Unassigned | Unassigned | 360_0 | Clustered | 358 | VC_360_0 | 358 | 0.9615 | 1 | 0.9615 | 0 | 0 | 0 | 1 | Ref |
| 3709 | KX266584 | Unassigned | Unassigned | Unassigned | 360_0 | Clustered | 358 | VC_360_0 | 358 | 0.9615 | 1 | 0.9615 | 0 | 0 | 0 | 1 | Ref |
| 3710 | KX266585 | Unassigned | Unassigned | Unassigned | 360_0 | Clustered | 358 | VC_360_0 | 358 | 0.9615 | 1 | 0.9615 | 0 | 0 | 0 | 1 | Ref |
| 3711 | KX266586 | Unassigned | Unassigned | Unassigned | 360_0 | Clustered | 358 | VC_360_0 | 358 | 0.9615 | 1 | 0.9615 | 0 | 0 | 0 | 1 | Ref |
| 3712 | KX266587 | Unassigned | Unassigned | Unassigned | 360_0 | Clustered | 358 | VC_360_0 | 358 | 0.9615 | 1 | 0.9615 | 0 | 0 | 0 | 1 | Ref |
| 3713 | KX266588 | Unassigned | Unassigned | Unassigned | 360_0 | Clustered | 358 | VC_360_0 | 358 | 0.9615 | 1 | 0.9615 | 0 | 0 | 0 | 1 | Ref |
| 3714 | KX266589 | Unassigned | Unassigned | Unassigned | 360_0 | Clustered | 358 | VC_360_0 | 358 | 0.9615 | 1 | 0.9615 | 0 | 0 | 0 | 1 | Ref |
| 3715 | KX266590 | Unassigned | Unassigned | Unassigned | 360_0 | Clustered | 358 | VC_360_0 | 358 | 0.9615 | 1 | 0.9615 | 0 | 0 | 0 | 1 | Ref |
| 3716 | KX266591 | Unassigned | Unassigned | Unassigned | 360_0 | Clustered | 358 | VC_360_0 | 358 | 0.9615 | 1 | 0.9615 | 0 | 0 | 0 | 1 | Ref |
| 3717 | KX266592 | Unassigned | Unassigned | Unassigned | 360_0 | Clustered | 358 | VC_360_0 | 358 | 0.9615 | 1 | 0.9615 | 0 | 0 | 0 | 1 | Ref |
| 3718 | KX266593 | Unassigned | Unassigned | Unassigned | 360_0 | Clustered | 358 | VC_360_0 | 358 | 0.9615 | 1 | 0.9615 | 0 | 0 | 0 | 1 | Ref |
| 3719 | KX266594 | Unassigned | Unassigned | Unassigned | 360_0 | Clustered | 358 | VC_360_0 | 358 | 0.9615 | 1 | 0.9615 | 0 | 0 | 0 | 1 | Ref |
| 3720 | KX266595 | Unassigned | Unassigned | Unassigned | 360_0 | Clustered | 358 | VC_360_0 | 358 | 0.9615 | 1 | 0.9615 | 0 | 0 | 0 | 1 | Ref |
| 3721 | KX266596 | Unassigned | Unassigned | Unassigned | 360_0 | Clustered | 358 | VC_360_0 | 358 | 0.9615 | 1 | 0.9615 | 0 | 0 | 0 | 1 | Ref |
| 3722 | KX266597 | Unassigned | Unassigned | Unassigned | 360_0 | Clustered | 358 | VC_360_0 | 358 | 0.9615 | 1 | 0.9615 | 0 | 0 | 0 | 1 | Ref |
| 3723 | KX266598 | Unassigned | Unassigned | Unassigned | 360_0 | Clustered | 358 | VC_360_0 | 358 | 0.9615 | 1 | 0.9615 | 0 | 0 | 0 | 1 | Ref |
| 3724 | KX266599 | Unassigned | Unassigned | Unassigned | 360_0 | Clustered | 358 | VC_360_0 | 358 | 0.9615 | 1 | 0.9615 | 0 | 0 | 0 | 1 | Ref |

| | | | | | | | | | | | | | | | | | |
|---|---|---|---|---|---|---|---|---|---|---|---|---|---|---|---|---|---|
| 3725 | KX266600 | Unassigned | Unassigned | Unassigned | 360_0 | Clustered | 358 | VC_360_0 | 358 | 0.9615 | 1 | 0.9615 | 0 | 0 | 0 | 1 | Ref |
| 3726 | KX266601 | Unassigned | Unassigned | Unassigned | 360_0 | Clustered | 358 | VC_360_0 | 358 | 0.9615 | 1 | 0.9615 | 0 | 0 | 0 | 1 | Ref |
| 3727 | KX266602 | Unassigned | Unassigned | Unassigned | 360_0 | Clustered | 358 | VC_360_0 | 358 | 0.9615 | 1 | 0.9615 | 0 | 0 | 0 | 1 | Ref |
| 3728 | KX266603 | Unassigned | Unassigned | Unassigned | 360_0 | Clustered | 358 | VC_360_0 | 358 | 0.9615 | 1 | 0.9615 | 0 | 0 | 0 | 1 | Ref |
| 3729 | KX266604 | Unassigned | Unassigned | Unassigned | 360_0 | Clustered | 358 | VC_360_0 | 358 | 0.9615 | 1 | 0.9615 | 0 | 0 | 0 | 1 | Ref |
| 3730 | KX266605 | Unassigned | Unassigned | Unassigned | 360_0 | Clustered | 358 | VC_360_0 | 358 | 0.9615 | 1 | 0.9615 | 0 | 0 | 0 | 1 | Ref |
| 3731 | KX266606 | Unassigned | Unassigned | Unassigned | 360_0 | Clustered | 358 | VC_360_0 | 358 | 0.9615 | 1 | 0.9615 | 0 | 0 | 0 | 1 | Ref |
| 3732 | KX266607 | Unassigned | Unassigned | Unassigned | 360_0 | Clustered | 358 | VC_360_0 | 358 | 0.9615 | 1 | 0.9615 | 0 | 0 | 0 | 1 | Ref |
| 3733 | KX266608 | Unassigned | Unassigned | Unassigned | 360_0 | Clustered | 358 | VC_360_0 | 358 | 0.9615 | 1 | 0.9615 | 0 | 0 | 0 | 1 | Ref |
| 3734 | KX266609 | Unassigned | Unassigned | Unassigned | 360_0 | Clustered | 358 | VC_360_0 | 358 | 0.9615 | 1 | 0.9615 | 0 | 0 | 0 | 1 | Ref |
| 3735 | KX266610 | Unassigned | Unassigned | Unassigned | 360_0 | Clustered | 358 | VC_360_0 | 358 | 0.9615 | 1 | 0.9615 | 0 | 0 | 0 | 1 | Ref |
| 3736 | KX266611 | Unassigned | Unassigned | Unassigned | 360_0 | Clustered | 358 | VC_360_0 | 358 | 0.9615 | 1 | 0.9615 | 0 | 0 | 0 | 1 | Ref |
| 3737 | KX266612 | Unassigned | Unassigned | Unassigned | 360_0 | Clustered | 358 | VC_360_0 | 358 | 0.9615 | 1 | 0.9615 | 0 | 0 | 0 | 1 | Ref |
| 3738 | KX266613 | Unassigned | Unassigned | Unassigned | 360_0 | Clustered | 358 | VC_360_0 | 358 | 0.9615 | 1 | 0.9615 | 0 | 0 | 0 | 1 | Ref |
| 3739 | KX266614 | Unassigned | Unassigned | Unassigned | 360_0 | Clustered | 358 | VC_360_0 | 358 | 0.9615 | 1 | 0.9615 | 0 | 0 | 0 | 1 | Ref |
| 3740 | KX266615 | Unassigned | Unassigned | Unassigned | 360_0 | Clustered | 358 | VC_360_0 | 358 | 0.9615 | 1 | 0.9615 | 0 | 0 | 0 | 1 | Ref |
| 3741 | KX266616 | Unassigned | Unassigned | Unassigned | 360_0 | Clustered | 358 | VC_360_0 | 358 | 0.9615 | 1 | 0.9615 | 0 | 0 | 0 | 1 | Ref |
| 3742 | KX266617 | Unassigned | Unassigned | Unassigned | 360_0 | Clustered | 358 | VC_360_0 | 358 | 0.9615 | 1 | 0.9615 | 0 | 0 | 0 | 1 | Ref |
| 3743 | KX266618 | Unassigned | Unassigned | Unassigned | 360_0 | Clustered | 358 | VC_360_0 | 358 | 0.9615 | 1 | 0.9615 | 0 | 0 | 0 | 1 | Ref |
| 3744 | KX266619 | Unassigned | Unassigned | Unassigned | 360_0 | Clustered | 358 | VC_360_0 | 358 | 0.9615 | 1 | 0.9615 | 0 | 0 | 0 | 1 | Ref |
| 3745 | KX266620 | Unassigned | Unassigned | Unassigned | 360_0 | Clustered | 358 | VC_360_0 | 358 | 0.9615 | 1 | 0.9615 | 0 | 0 | 0 | 1 | Ref |
| 3746 | KX266621 | Unassigned | Unassigned | Unassigned | 360_0 | Clustered | 358 | VC_360_0 | 358 | 0.9615 | 1 | 0.9615 | 0 | 0 | 0 | 1 | Ref |
| 3747 | KX266622 | Unassigned | Unassigned | Unassigned | 360_0 | Clustered | 358 | VC_360_0 | 358 | 0.9615 | 1 | 0.9615 | 0 | 0 | 0 | 1 | Ref |
| 3748 | KX266623 | Unassigned | Unassigned | Unassigned | 360_0 | Clustered | 358 | VC_360_0 | 358 | 0.9615 | 1 | 0.9615 | 0 | 0 | 0 | 1 | Ref |
| 3749 | KX266624 | Unassigned | Unassigned | Unassigned | 360_0 | Clustered | 358 | VC_360_0 | 358 | 0.9615 | 1 | 0.9615 | 0 | 0 | 0 | 1 | Ref |
| 3750 | KX266625 | Unassigned | Unassigned | Unassigned | 360_0 | Clustered | 358 | VC_360_0 | 358 | 0.9615 | 1 | 0.9615 | 0 | 0 | 0 | 1 | Ref |
| 3751 | KX266626 | Unassigned | Unassigned | Unassigned | 360_0 | Clustered | 358 | VC_360_0 | 358 | 0.9615 | 1 | 0.9615 | 0 | 0 | 0 | 1 | Ref |
| 3752 | KX266627 | Unassigned | Unassigned | Unassigned | 360_0 | Clustered | 358 | VC_360_0 | 358 | 0.9615 | 1 | 0.9615 | 0 | 0 | 0 | 1 | Ref |
| 3753 | KX266628 | Unassigned | Unassigned | Unassigned | 360_0 | Clustered | 358 | VC_360_0 | 358 | 0.9615 | 1 | 0.9615 | 0 | 0 | 0 | 1 | Ref |
| 3754 | KX266629 | Unassigned | Unassigned | Unassigned | 360_0 | Clustered | 358 | VC_360_0 | 358 | 0.9615 | 1 | 0.9615 | 0 | 0 | 0 | 1 | Ref |
| 3755 | KX266630 | Unassigned | Unassigned | Unassigned | 360_0 | Clustered | 358 | VC_360_0 | 358 | 0.9615 | 1 | 0.9615 | 0 | 0 | 0 | 1 | Ref |
| 3756 | KX266631 | Unassigned | Unassigned | Unassigned | 360_0 | Clustered | 358 | VC_360_0 | 358 | 0.9615 | 1 | 0.9615 | 0 | 0 | 0 | 1 | Ref |
| 3757 | KX266632 | Unassigned | Unassigned | Unassigned | 360_0 | Clustered | 358 | VC_360_0 | 358 | 0.9615 | 1 | 0.9615 | 0 | 0 | 0 | 1 | Ref |
| 3758 | KX266633 | Unassigned | Unassigned | Unassigned | 360_0 | Clustered | 358 | VC_360_0 | 358 | 0.9615 | 1 | 0.9615 | 0 | 0 | 0 | 1 | Ref |
| 3759 | KX266634 | Unassigned | Unassigned | Unassigned | 360_0 | Clustered | 358 | VC_360_0 | 358 | 0.9615 | 1 | 0.9615 | 0 | 0 | 0 | 1 | Ref |
| 3760 | KX266635 | Unassigned | Unassigned | Unassigned | 360_0 | Clustered | 358 | VC_360_0 | 358 | 0.9615 | 1 | 0.9615 | 0 | 0 | 0 | 1 | Ref |
| 3761 | KX266636 | Unassigned | Unassigned | Unassigned | 360_0 | Clustered | 358 | VC_360_0 | 358 | 0.9615 | 1 | 0.9615 | 0 | 0 | 0 | 1 | Ref |
| 3762 | KX266637 | Unassigned | Unassigned | Unassigned | 360_0 | Clustered | 358 | VC_360_0 | 358 | 0.9615 | 1 | 0.9615 | 0 | 0 | 0 | 1 | Ref |
| 3763 | KX278418 | Unassigned | Unassigned | Unassigned | 984_0 | Clustered | 38 | VC_984_0 | 38 | 0.9758 | 1 | 0.9758 | 0 | 0 | 0 | 1 | Ref |
| 3764 | KX278419 | Unassigned | Unassigned | Unassigned | 984_0 | Clustered | 38 | VC_984_0 | 38 | 0.9758 | 1 | 0.9758 | 0 | 0 | 0 | 1 | Ref |
| 3765 | KX279892 | Unassigned | Unassigned | Unassigned | 452_0 | Clustered | 66 | VC_452_0 | 66 | 0.9248 | 1 | 0.9248 | 0 | 0 | 0 | 1 | Ref |
| 3766 | KX284704 | Unassigned | Unassigned | Unassigned | 695_0 | Clustered | 48 | VC_695_0 | 48 | 0.8162 | 1 | 0.8162 | 0 | 0 | 0 | 1 | Ref |
| 3767 | KX344445 | Unassigned | Unassigned | Unassigned | 982_0 | Clustered | 62 | VC_982_0 | 62 | 0.8646 | 1 | 0.8646 | 0 | 0 | 0 | 0.743 | Ref |
| 3768 | KX344510 | Unassigned | Unassigned | Unassigned | 961_0 | Clustered | 3 | VC_961_0 | 3 | 0.675 | 0.99054823 | 0.6686 | 0 | 0 | 0 | 1 | Ref |
| 3769 | KX346246 | Unassigned | Unassigned | Unassigned | 202_0 | Clustered | 102 | VC_202_0 | 102 | 0.4049 | 1 | 0.4049 | 0 | 0 | 0 | 0.7876 | Ref |
| 3770 | KX349226 | Unassigned | Unassigned | Unassigned | 662_0 | Clustered | 62 | VC_662_0 | 62 | 0.6976 | 1 | 0.6976 | 0 | 0 | 0 | 1 | Ref |
| 3771 | KX349227 | Unassigned | Unassigned | Unassigned | 662_0 | Clustered | 62 | VC_662_0 | 62 | 0.6976 | 1 | 0.6976 | 0 | 0 | 0 | 1 | Ref |
| 3772 | KX349228 | Unassigned | Unassigned | Unassigned | 662_0 | Clustered | 62 | VC_662_0 | 62 | 0.6976 | 1 | 0.6976 | 0 | 0 | 0 | 1 | Ref |
| 3773 | KX349229 | Unassigned | Unassigned | Unassigned | 662_0 | Clustered | 62 | VC_662_0 | 62 | 0.6976 | 1 | 0.6976 | 0 | 0 | 0 | 1 | Ref |
| 3774 | KX349230 | Unassigned | Unassigned | Unassigned | 662_0 | Clustered | 62 | VC_662_0 | 62 | 0.6976 | 1 | 0.6976 | 0 | 0 | 0 | 1 | Ref |
| 3775 | KX349231 | Unassigned | Unassigned | Unassigned | 662_0 | Clustered | 62 | VC_662_0 | 62 | 0.6976 | 1 | 0.6976 | 0 | 0 | 0 | 1 | Ref |
| 3776 | KX349232 | Unassigned | Unassigned | Unassigned | 662_0 | Clustered | 62 | VC_662_0 | 62 | 0.6976 | 1 | 0.6976 | 0 | 0 | 0 | 1 | Ref |
| 3777 | KX349233 | Unassigned | Unassigned | Unassigned | 662_0 | Clustered | 62 | VC_662_0 | 62 | 0.6976 | 1 | 0.6976 | 0 | 0 | 0 | 1 | Ref |
| 3778 | KX349234 | Unassigned | Unassigned | Unassigned | 662_0 | Clustered | 62 | VC_662_0 | 62 | 0.6976 | 1 | 0.6976 | 0 | 0 | 0 | 1 | Ref |
| 3779 | KX349235 | Unassigned | Unassigned | Unassigned | 662_0 | Clustered | 62 | VC_662_0 | 62 | 0.6976 | 1 | 0.6976 | 0 | 0 | 0 | 1 | Ref |
| 3780 | KX349236 | Unassigned | Unassigned | Unassigned | 662_0 | Clustered | 62 | VC_662_0 | 62 | 0.6976 | 1 | 0.6976 | 0 | 0 | 0 | 1 | Ref |
| 3781 | KX349237 | Unassigned | Unassigned | Unassigned | 662_0 | Clustered | 62 | VC_662_0 | 62 | 0.6976 | 1 | 0.6976 | 0 | 0 | 0 | 1 | Ref |
| 3782 | KX349238 | Unassigned | Unassigned | Unassigned | 662_0 | Clustered | 62 | VC_662_0 | 62 | 0.6976 | 1 | 0.6976 | 0 | 0 | 0 | 1 | Ref |
| 3783 | KX349239 | Unassigned | Unassigned | Unassigned | 662_0 | Clustered | 62 | VC_662_0 | 62 | 0.6976 | 1 | 0.6976 | 0 | 0 | 0 | 1 | Ref |
| 3784 | KX349240 | Unassigned | Unassigned | Unassigned | 662_0 | Clustered | 62 | VC_662_0 | 62 | 0.6976 | 1 | 0.6976 | 0 | 0 | 0 | 1 | Ref |
| 3785 | KX349241 | Unassigned | Unassigned | Unassigned | 662_0 | Clustered | 62 | VC_662_0 | 62 | 0.6976 | 1 | 0.6976 | 0 | 0 | 0 | 1 | Ref |
| 3786 | KX349242 | Unassigned | Unassigned | Unassigned | 662_0 | Clustered | 62 | VC_662_0 | 62 | 0.6976 | 1 | 0.6976 | 0 | 0 | 0 | 1 | Ref |
| 3787 | KX349243 | Unassigned | Unassigned | Unassigned | 662_0 | Clustered | 62 | VC_662_0 | 62 | 0.6976 | 1 | 0.6976 | 0 | 0 | 0 | 1 | Ref |
| 3788 | KX349244 | Unassigned | Unassigned | Unassigned | 662_0 | Clustered | 62 | VC_662_0 | 62 | 0.6976 | 1 | 0.6976 | 0 | 0 | 0 | 1 | Ref |
| 3789 | KX349245 | Unassigned | Unassigned | Unassigned | 662_0 | Clustered | 62 | VC_662_0 | 62 | 0.6976 | 1 | 0.6976 | 0 | 0 | 0 | 1 | Ref |
| 3790 | KX349246 | Unassigned | Unassigned | Unassigned | 662_0 | Clustered | 62 | VC_662_0 | 62 | 0.6976 | 1 | 0.6976 | 0 | 0 | 0 | 1 | Ref |
| 3791 | KX349247 | Unassigned | Unassigned | Unassigned | 662_0 | Clustered | 62 | VC_662_0 | 62 | 0.6976 | 1 | 0.6976 | 0 | 0 | 0 | 1 | Ref |
| 3792 | KX349248 | Unassigned | Unassigned | Unassigned | 662_0 | Clustered | 62 | VC_662_0 | 62 | 0.6976 | 1 | 0.6976 | 0 | 0 | 0 | 1 | Ref |
| 3793 | KX349249 | Unassigned | Unassigned | Unassigned | 662_0 | Clustered | 62 | VC_662_0 | 62 | 0.6976 | 1 | 0.6976 | 0 | 0 | 0 | 1 | Ref |
| 3794 | KX349250 | Unassigned | Unassigned | Unassigned | 662_0 | Clustered | 62 | VC_662_0 | 62 | 0.6976 | 1 | 0.6976 | 0 | 0 | 0 | 1 | Ref |
| 3795 | KX349251 | Unassigned | Unassigned | Unassigned | 662_0 | Clustered | 62 | VC_662_0 | 62 | 0.6976 | 1 | 0.6976 | 0 | 0 | 0 | 1 | Ref |
| 3796 | KX349252 | Unassigned | Unassigned | Unassigned | 662_0 | Clustered | 62 | VC_662_0 | 62 | 0.6976 | 1 | 0.6976 | 0 | 0 | 0 | 1 | Ref |
| 3797 | KX349253 | Unassigned | Unassigned | Unassigned | 662_0 | Clustered | 62 | VC_662_0 | 62 | 0.6976 | 1 | 0.6976 | 0 | 0 | 0 | 1 | Ref |
| 3798 | KX349254 | Unassigned | Unassigned | Unassigned | 662_0 | Clustered | 62 | VC_662_0 | 62 | 0.6976 | 1 | 0.6976 | 0 | 0 | 0 | 1 | Ref |
| 3799 | KX349255 | Unassigned | Unassigned | Unassigned | 662_0 | Clustered | 62 | VC_662_0 | 62 | 0.6976 | 1 | 0.6976 | 0 | 0 | 0 | 1 | Ref |
| 3800 | KX349256 | Unassigned | Unassigned | Unassigned | 662_0 | Clustered | 62 | VC_662_0 | 62 | 0.6976 | 1 | 0.6976 | 0 | 0 | 0 | 1 | Ref |
| 3801 | KX349257 | Unassigned | Unassigned | Unassigned | 662_0 | Clustered | 62 | VC_662_0 | 62 | 0.6976 | 1 | 0.6976 | 0 | 0 | 0 | 1 | Ref |
| 3802 | KX349258 | Unassigned | Unassigned | Unassigned | 662_0 | Clustered | 62 | VC_662_0 | 62 | 0.6976 | 1 | 0.6976 | 0 | 0 | 0 | 1 | Ref |
| 3803 | KX349259 | Unassigned | Unassigned | Unassigned | 662_0 | Clustered | 62 | VC_662_0 | 62 | 0.6976 | 1 | 0.6976 | 0 | 0 | 0 | 1 | Ref |
| 3804 | KX349260 | Unassigned | Unassigned | Unassigned | 662_0 | Clustered | 62 | VC_662_0 | 62 | 0.6976 | 1 | 0.6976 | 0 | 0 | 0 | 1 | Ref |
| 3805 | KX349261 | Unassigned | Unassigned | Unassigned | 662_0 | Clustered | 62 | VC_662_0 | 62 | 0.6976 | 1 | 0.6976 | 0 | 0 | 0 | 1 | Ref |
| 3806 | KX349262 | Unassigned | Unassigned | Unassigned | 662_0 | Clustered | 62 | VC_662_0 | 62 | 0.6976 | 1 | 0.6976 | 0 | 0 | 0 | 1 | Ref |
| 3807 | KX349263 | Unassigned | Unassigned | Unassigned | 662_0 | Clustered | 62 | VC_662_0 | 62 | 0.6976 | 1 | 0.6976 | 0 | 0 | 0 | 1 | Ref |
| 3808 | KX349264 | Unassigned | Unassigned | Unassigned | 662_0 | Clustered | 62 | VC_662_0 | 62 | 0.6976 | 1 | 0.6976 | 0 | 0 | 0 | 1 | Ref |
| 3809 | KX349265 | Unassigned | Unassigned | Unassigned | 662_0 | Clustered | 62 | VC_662_0 | 62 | 0.6976 | 1 | 0.6976 | 0 | 0 | 0 | 1 | Ref |
| 3810 | KX349266 | Unassigned | Unassigned | Unassigned | 662_0 | Clustered | 62 | VC_662_0 | 62 | 0.6976 | 1 | 0.6976 | 0 | 0 | 0 | 1 | Ref |
| 3811 | KX349267 | Unassigned | Unassigned | Unassigned | 662_0 | Clustered | 62 | VC_662_0 | 62 | 0.6976 | 1 | 0.6976 | 0 | 0 | 0 | 1 | Ref |
| 3812 | KX349268 | Unassigned | Unassigned | Unassigned | 662_0 | Clustered | 62 | VC_662_0 | 62 | 0.6976 | 1 | 0.6976 | 0 | 0 | 0 | 1 | Ref |
| 3813 | KX349269 | Unassigned | Unassigned | Unassigned | 662_0 | Clustered | 62 | VC_662_0 | 62 | 0.6976 | 1 | 0.6976 | 0 | 0 | 0 | 1 | Ref |
| 3814 | KX349270 | Unassigned | Unassigned | Unassigned | 662_0 | Clustered | 62 | VC_662_0 | 62 | 0.6976 | 1 | 0.6976 | 0 | 0 | 0 | 1 | Ref |
| 3815 | KX349271 | Unassigned | Unassigned | Unassigned | 662_0 | Clustered | 62 | VC_662_0 | 62 | 0.6976 | 1 | 0.6976 | 0 | 0 | 0 | 1 | Ref |
| 3816 | KX349272 | Unassigned | Unassigned | Unassigned | 662_0 | Clustered | 62 | VC_662_0 | 62 | 0.6976 | 1 | 0.6976 | 0 | 0 | 0 | 1 | Ref |
| 3817 | KX349273 | Unassigned | Unassigned | Unassigned | 662_0 | Clustered | 62 | VC_662_0 | 62 | 0.6976 | 1 | 0.6976 | 0 | 0 | 0 | 1 | Ref |
| 3818 | KX349274 | Unassigned | Unassigned | Unassigned | 662_0 | Clustered | 62 | VC_662_0 | 62 | 0.6976 | 1 | 0.6976 | 0 | 0 | 0 | 1 | Ref |
| 3819 | KX349275 | Unassigned | Unassigned | Unassigned | 662_0 | Clustered | 62 | VC_662_0 | 62 | 0.6976 | 1 | 0.6976 | 0 | 0 | 0 | 1 | Ref |
| 3820 | KX349276 | Unassigned | Unassigned | Unassigned | 662_0 | Clustered | 62 | VC_662_0 | 62 | 0.6976 | 1 | 0.6976 | 0 | 0 | 0 | 1 | Ref |
| 3821 | KX349277 | Unassigned | Unassigned | Unassigned | 662_0 | Clustered | 62 | VC_662_0 | 62 | 0.6976 | 1 | 0.6976 | 0 | 0 | 0 | 1 | Ref |
| 3822 | KX349278 | Unassigned | Unassigned | Unassigned | 662_0 | Clustered | 62 | VC_662_0 | 62 | 0.6976 | 1 | 0.6976 | 0 | 0 | 0 | 1 | Ref |
| 3823 | KX349279 | Unassigned | Unassigned | Unassigned | 662_0 | Clustered | 62 | VC_662_0 | 62 | 0.6976 | 1 | 0.6976 | 0 | 0 | 0 | 1 | Ref |
| 3824 | KX349280 | Unassigned | Unassigned | Unassigned | 662_0 | Clustered | 62 | VC_662_0 | 62 | 0.6976 | 1 | 0.6976 | 0 | 0 | 0 | 1 | Ref |
| 3825 | KX349281 | Unassigned | Unassigned | Unassigned | 662_0 | Clustered | 62 | VC_662_0 | 62 | 0.6976 | 1 | 0.6976 | 0 | 0 | 0 | 1 | Ref |
| 3826 | KX349282 | Unassigned | Unassigned | Unassigned | 662_0 | Clustered | 62 | VC_662_0 | 62 | 0.6976 | 1 | 0.6976 | 0 | 0 | 0 | 1 | Ref |
| 3827 | KX349283 | Unassigned | Unassigned | Unassigned | 662_0 | Clustered | 62 | VC_662_0 | 62 | 0.6976 | 1 | 0.6976 | 0 | 0 | 0 | 1 | Ref |
| 3828 | KX349284 | Unassigned | Unassigned | Unassigned | 662_0 | Clustered | 62 | VC_662_0 | 62 | 0.6976 | 1 | 0.6976 | 0 | 0 | 0 | 1 | Ref |
| 3829 | KX349291 | Unassigned | Unassigned | Unassigned | 433_4 | Clustered | 9 | VC_433_4 | 9 | 0.1285 | 1 | 0.1285 | 0 | 0 | 0 | 1 | Ref |
| 3830 | KX349292 | Unassigned | Unassigned | Unassigned | 433_4 | Clustered | 9 | VC_433_4 | 9 | 0.1285 | 1 | 0.1285 | 0 | 0 | 0 | 1 | Ref |
| 3831 | KX349293 | Unassigned | Unassigned | Unassigned | 433_4 | Clustered | 9 | VC_433_4 | 9 | 0.1285 | 1 | 0.1285 | 0 | 0 | 0 | 1 | Ref |
| 3832 | KX349294 | Unassigned | Unassigned | Unassigned | 433_4 | Clustered | 9 | VC_433_4 | 9 | 0.1285 | 1 | 0.1285 | 0 | 0 | 0 | 1 | Ref |
| 3833 | KX349295 | Unassigned | Unassigned | Unassigned | 433_4 | Clustered | 9 | VC_433_4 | 9 | 0.1285 | 1 | 0.1285 | 0 | 0 | 0 | 1 | Ref |
| 3834 | KX349296 | Unassigned | Unassigned | Unassigned | 433_4 | Clustered | 9 | VC_433_4 | 9 | 0.1285 | 1 | 0.1285 | 0 | 0 | 0 | 1 | Ref |
| 3835 | KX349297 | Unassigned | Unassigned | Unassigned | 433_4 | Clustered | 9 | VC_433_4 | 9 | 0.1285 | 1 | 0.1285 | 0 | 0 | 0 | 1 | Ref |
| 3836 | KX349899 | Unassigned | Unassigned | Unassigned | 818_0 | Clustered | 20 | VC_818_0 | 20 | 0.9601 | 1 | 0.9601 | 0 | 0 | 0 | 1 | Ref |
| 3837 | KX349900 | Unassigned | Unassigned | Unassigned | 818_0 | Clustered | 20 | VC_818_0 | 20 | 0.9601 | 1 | 0.9601 | 0 | 0 | 0 | 1 | Ref |
| 3838 | KX349901 | Unassigned | Unassigned | Unassigned | 818_0 | Clustered | 20 | VC_818_0 | 20 | 0.9601 | 1 | 0.9601 | 0 | 0 | 0 | 1 | Ref |
| 3839 | KX349902 | Unassigned | Unassigned | Unassigned | 884_0 | Clustered | 28 | VC_884_0 | 28 | 0.6211 | 1 | 0.6211 | 0 | 0 | 0 | 0.9841 | Ref |
| 3840 | KX349903 | Unassigned | Unassigned | Unassigned | 884_0 | Clustered | 28 | VC_884_0 | 28 | 0.6211 | 1 | 0.6211 | 0 | 0 | 0 | 0.9841 | Ref |
| 3841 | KX365747 | Unassigned | Unassigned | Unassigned | 117_0 | Clustered | 9 | VC_117_0 | 9 | 0.9977 | 0.98988637 | 0.9876 | 0 | 0 | 0 | 1 | Ref |
| 3842 | KX365748 | Unassigned | Unassigned | Unassigned | 895_0 | Clustered | 2 | VC_895_0 | 2 | 0.2788 | 0.94251254 | 0.2628 | 0 | 0 | 0 | 1 | Ref |
| 3843 | KX369583 | Unassigned | Unassigned | Unassigned | 558_23 | Clustered/Singleton | 1 | VC_558_23 | 1 | 0 | 1 | 0 | 0 | 0 | 0 | 1 | Ref |
| 3844 | KX369584 | Unassigned | Unassigned | Unassigned | 325_28 | Clustered/Singleton | 1 | VC_325_28 | 1 | 0 | 1 | 0 | 0 | 0 | 0 | 1 | Ref |
| 3845 | KX369585 | Unassigned | Unassigned | Unassigned | 371_16 | Clustered | 42 | VC_371_16 | 42 | 0.1341 | 1 | 0.1341 | 0 | 0 | 0 | 0.7282 | Ref |
| 3846 | KX369586 | Unassigned | Unassigned | Unassigned | 325_4 | Clustered/Singleton | 1 | VC_325_4 | 1 | 0 | 1 | 0 | 0 | 0 | 0 | 1 | Ref |
| 3847 | KX373685 | Unassigned | Unassigned | Unassigned | 690_0 | Clustered | 50 | VC_690_0 | 50 | 0.9805 | 1 | 0.9805 | 0 | 0 | 0 | 1 | Ref |
| 3848 | KX373686 | Unassigned | Unassigned | Unassigned | 690_0 | Clustered | 50 | VC_690_0 | 50 | 0.9805 | 1 | 0.9805 | 0 | 0 | 0 | 1 | Ref |
| 3849 | KX373687 | Unassigned | Unassigned | Unassigned | 690_0 | Clustered | 50 | VC_690_0 | 50 | 0.9805 | 1 | 0.9805 | 0 | 0 | 0 | 1 | Ref |
| 3850 | KX373688 | Unassigned | Unassigned | Unassigned | 690_0 | Clustered | 50 | VC_690_0 | 50 | 0.9805 | 1 | 0.9805 | 0 | 0 | 0 | 1 | Ref |
| 3851 | KX373689 | Unassigned | Unassigned | Unassigned | 690_0 | Clustered | 50 | VC_690_0 | 50 | 0.9805 | 1 | 0.9805 | 0 | 0 | 0 | 1 | Ref |
| 3852 | KX373690 | Unassigned | Unassigned | Unassigned | 690_0 | Clustered | 50 | VC_690_0 | 50 | 0.9805 | 1 | 0.9805 | 0 | 0 | 0 | 1 | Ref |
| 3853 | KX373691 | Unassigned | Unassigned | Unassigned | 690_0 | Clustered | 50 | VC_690_0 | 50 | 0.9805 | 1 | 0.9805 | 0 | 0 | 0 | 1 | Ref |
| 3854 | KX373692 | Unassigned | Unassigned | Unassigned | 690_0 | Clustered | 50 | VC_690_0 | 50 | 0.9805 | 1 | 0.9805 | 0 | 0 | 0 | 1 | Ref |
| 3855 | KX375815 | Unassigned | Unassigned | Unassigned | 223_0 | Clustered | 38 | VC_223_0 | 38 | 0.9095 | 1 | 0.9095 | 0 | 0 | 0 | 1 | Ref |
| 3856 | KX377933 | Unassigned | Unassigned | Unassigned | 329_3 | Clustered | 24 | VC_329_3 | 24 | 0.1206 | 1 | 0.1206 | 0 | 0 | 0 | 0.7283 | Ref |
| 3857 | KX379667 | Unassigned | Unassigned | Unassigned | 202_0 | Clustered | 102 | VC_202_0 | 102 | 0.4049 | 1 | 0.4049 | 0 | 0 | 0 | 0.7876 | Ref |
| 3858 | KX379668 | Unassigned | Unassigned | Unassigned | 202_0 | Clustered | 102 | VC_202_0 | 102 | 0.4049 | 1 | 0.4049 | 0 | 0 | 0 | 0.7876 | Ref |
| 3859 | KX379670 | Unassigned | Unassigned | Unassigned | 202_0 | Clustered | 102 | VC_202_0 | 102 | 0.4049 | 1 | 0.4049 | 0 | 0 | 0 | 0.7876 | Ref |
| 3860 | KX379671 | Unassigned | Unassigned | Unassigned | 202_0 | Clustered | 102 | VC_202_0 | 102 | 0.4049 | 1 | 0.4049 | 0 | 0 | 0 | 0.7876 | Ref |
| 3861 | KX397364 | Unassigned | Unassigned | Unassigned | 959_4 | Clustered | 3 | VC_959_4 | 3 | 0.0237 | 0.77138817 | 0.0183 | 0 | 0 | 0 | 1 | Ref |
| 3862 | KX397365 | Unassigned | Unassigned | Unassigned | 959_2 | Clustered | 5 | VC_959_2 | 5 | 0.0484 | 0.89346426 | 0.0432 | 0 | 0 | 0 | 1 | Ref |
| 3863 | KX397366 | Unassigned | Unassigned | Unassigned | 959_2 | Clustered | 5 | VC_959_2 | 5 | 0.0484 | 0.89346426 | 0.0432 | 0 | 0 | 0 | 1 | Ref |
| 3864 | KX397367 | Unassigned | Unassigned | Unassigned | 959_0 | Clustered | 5 | VC_959_0 | 5 | 0.0499 | 0.99999231 | 0.0499 | 0 | 0 | 0 | 1 | Ref |
| 3865 | KX397368 | Unassigned | Unassigned | Unassigned | 959_1 | Clustered | 8 | VC_959_1 | 8 | 0.0878 | 0.98435099 | 0.0864 | 0 | 0 | 0 | 1 | Ref |
| 3866 | KX397369 | Unassigned | Unassigned | Unassigned | 959_3 | Clustered | 3 | VC_959_3 | 3 | 0.0237 | 0.75911548 | 0.018 | 0 | 0 | 0 | 1 | Ref |
| 3867 | KX397370 | Unassigned | Unassigned | Unassigned | 959_1 | Clustered | 8 | VC_959_1 | 8 | 0.0878 | 0.98435099 | 0.0864 | 0 | 0 | 0 | 1 | Ref |
| 3868 | KX397371 | Unassigned | Unassigned | Unassigned | 959_1 | Clustered | 8 | VC_959_1 | 8 | 0.0878 | 0.98435099 | 0.0864 | 0 | 0 | 0 | 1 | Ref |
| 3869 | KX397372 | Unassigned | Unassigned | Unassigned | 959_0 | Clustered | 5 | VC_959_0 | 5 | 0.0499 | 0.99999231 | 0.0499 | 0 | 0 | 0 | 1 | Ref |
| 3870 | KX397373 | Unassigned | Unassigned | Unassigned | 959_4 | Clustered | 3 | VC_959_4 | 3 | 0.0237 | 0.77138817 | 0.0183 | 0 | 0 | 0 | 1 | Ref |
| 3871 | KX405002 | Unassigned | Unassigned | Unassigned | 637_0 | Clustered | 115 | VC_637_0 | 115 | 0.731 | 1 | 0.731 | 0 | 0 | 0 | 0.8764 | Ref |
| 3872 | KX405003 | Unassigned | Unassigned | Unassigned | 329_16 | Clustered | 4 | VC_329_16 | 4 | 0.0158 | 0.9999807 | 0.0158 | 0 | 0 | 0 | 1 | Ref |
| 3873 | KX431559 | Unassigned | Unassigned | Unassigned | 669_0 | Clustered | 21 | VC_669_0 | 21 | 0.3623 | 1 | 0.3623 | 0 | 0 | 0 | 1 | Ref |
| 3874 | KX431560 | Unassigned | Unassigned | Unassigned | 669_0 | Clustered | 21 | VC_669_0 | 21 | 0.3623 | 1 | 0.3623 | 0 | 0 | 0 | 1 | Ref |
| 3875 | KX431888 | Unassigned | Unassigned | Unassigned | 130_0 | Clustered | 19 | VC_130_0 | 19 | 0.6068 | 1 | 0.6068 | 0 | 0 | 0 | 1 | Ref |
| 3876 | KX438380 | Unassigned | Unassigned | Unassigned | 133_14 | Clustered | 3 | VC_133_14 | 3 | 0.0102 | 0.96327256 | 0.0098 | 0 | 0 | 0 | 1 | Ref |
| 3877 | KX443326 | Unassigned | Unassigned | Unassigned | 761_0 | Clustered | 67 | VC_761_0 | 67 | 0.8947 | 1 | 0.8947 | 0 | 0 | 0 | 1 | Ref |
| 3878 | KX443552 | Unassigned | Unassigned | Unassigned | 669_0 | Clustered | 21 | VC_669_0 | 21 | 0.3623 | 1 | 0.3623 | 0 | 0 | 0 | 1 | Ref |
| 3879 | KX443695 | Unassigned | Unassigned | Unassigned | 950_0 | Clustered | 38 | VC_950_0 | 38 | 0.9443 | 1 | 0.9443 | 0 | 0 | 0 | 1 | Ref |
| 3880 | KX443696 | Unassigned | Unassigned | Unassigned | 508_0 | Clustered | 32 | VC_508_0 | 32 | 0.906 | 1 | 0.906 | 0 | 0 | 0 | 1 | Ref |
| 3881 | KX449361 | Unassigned | Unassigned | Unassigned | 130_0 | Clustered | 19 | VC_130_0 | 19 | 0.6068 | 1 | 0.6068 | 0 | 0 | 0 | 1 | Ref |
| 3882 | KX449362 | Unassigned | Unassigned | Unassigned | 130_0 | Clustered | 19 | VC_130_0 | 19 | 0.6068 | 1 | 0.6068 | 0 | 0 | 0 | 1 | Ref |
| 3883 | KX449363 | Unassigned | Unassigned | Unassigned | 130_0 | Clustered | 19 | VC_130_0 | 19 | 0.6068 | 1 | 0.6068 | 0 | 0 | 0 | 1 | Ref |
| 3884 | KX452695 | Unassigned | Unassigned | Unassigned | 582_3 | Clustered | 12 | VC_582_3 | 12 | 0.2357 | 1 | 0.2357 | 0 | 0 | 0 | 0.8636 | Ref |
| 3885 | KX452696 | Unassigned | Unassigned | Unassigned | 385_0 | Clustered | 51 | VC_385_0 | 51 | 0.8754 | 1 | 0.8754 | 0 | 0 | 0 | 1 | Ref |
| 3886 | KX452697 | Unassigned | Unassigned | Unassigned | 132_0 | Clustered | 4 | VC_132_0 | 4 | 0.036 | 0.99998777 | 0.036 | 0 | 0 | 0 | 1 | Ref |

| | | | | | | | | | | | | | | | | | |
|---|---|---|---|---|---|---|---|---|---|---|---|---|---|---|---|---|---|
| 3887 | KX452698 | Unassigned | Unassigned | Unassigned | 283_143 | Clustered/Singleton | 1 | VC_283_143 | 1 | 0 | 1 | 0 | 0 | 0 | 0 | 1 | Ref |
| 3888 | KX452699 | Unassigned | Unassigned | Unassigned | 592_0 | Clustered | 50 | VC_592_0 | 50 | 0.7314 | 1 | 0.7314 | 0 | 0 | 0 | 1 | Ref |
| 3889 | KX452700 | Unassigned | Unassigned | Unassigned | 592_0 | Clustered | 50 | VC_592_0 | 50 | 0.7314 | 1 | 0.7314 | 0 | 0 | 0 | 1 | Ref |
| 3890 | KX455876 | Unassigned | Unassigned | Unassigned | 1189_0 | Clustered | 9 | VC_1189_0 | 9 | 0.968 | 1 | 0.968 | 0 | 0 | 0 | 1 | Ref |
| 3891 | KX456206 | Unassigned | Unassigned | Unassigned | 265_0 | Clustered | 12 | VC_265_0 | 12 | 0.4401 | 1 | 0.4401 | 0 | 0 | 0 | 1 | Ref |
| 3892 | KX456208 | Unassigned | Unassigned | Unassigned | 265_0 | Clustered | 12 | VC_265_0 | 12 | 0.4401 | 1 | 0.4401 | 0 | 0 | 0 | 1 | Ref |
| 3893 | KX456209 | Unassigned | Unassigned | Unassigned | 265_0 | Clustered | 12 | VC_265_0 | 12 | 0.4401 | 1 | 0.4401 | 0 | 0 | 0 | 1 | Ref |
| 3894 | KX456210 | Unassigned | Unassigned | Unassigned | 265_0 | Clustered | 12 | VC_265_0 | 12 | 0.4401 | 1 | 0.4401 | 0 | 0 | 0 | 1 | Ref |
| 3895 | KX456211 | Unassigned | Unassigned | Unassigned | 262_0 | Clustered | 5 | VC_262_0 | 5 | 0.0724 | 0.99999989 | 0.0724 | 0 | 0 | 0 | 1 | Ref |
| 3896 | KX456213 | Unassigned | Unassigned | Unassigned | 265_0 | Clustered | 12 | VC_265_0 | 12 | 0.4401 | 1 | 0.4401 | 0 | 0 | 0 | 1 | Ref |
| 3897 | KX458236 | Unassigned | Unassigned | Unassigned | 517_1 | Clustered | 108 | VC_517_1 | 108 | 0.8483 | 1 | 0.8483 | 0 | 0 | 0 | 0.9955 | Ref |
| 3898 | KX458237 | Unassigned | Unassigned | Unassigned | 514_0 | Clustered | 75 | VC_514_0 | 75 | 0.9402 | 1 | 0.9402 | 0 | 0 | 0 | 1 | Ref |
| 3899 | KX467643 | Unassigned | Unassigned | Unassigned | 349_0 | Clustered | 7 | VC_349_0 | 7 | 0.9363 | 0.99999996 | 0.9363 | 0 | 0 | 0 | 1 | Ref |
| 3900 | KX495186 | Unassigned | Unassigned | Unassigned | 187_3 | Clustered | 9 | VC_187_3 | 9 | 0.1971 | 1 | 0.1971 | 0 | 0 | 0 | 0.8611 | Ref |
| 3901 | KX507045 | Unassigned | Unassigned | Unassigned | 642_3 | Clustered | 9 | VC_642_3 | 9 | 0.2131 | 1 | 0.2131 | 0 | 0 | 0 | 0.6944 | Ref |
| 3902 | KX507046 | Unassigned | Unassigned | Unassigned | 493_11 | Clustered/Singleton | 1 | VC_493_11 | 1 | 0 | 1 | 0 | 0 | 0 | 0 | 1 | Ref |
| 3903 | KX507343 | Unassigned | Unassigned | Unassigned | 982_0 | Clustered | 62 | VC_982_0 | 62 | 0.8646 | 1 | 0.8646 | 0 | 0 | 0 | 0.743 | Ref |
| 3904 | KX507344 | Unassigned | Unassigned | Unassigned | 982_0 | Clustered | 62 | VC_982_0 | 62 | 0.8646 | 1 | 0.8646 | 0 | 0 | 0 | 0.743 | Ref |
| 3905 | KX507345 | Unassigned | Unassigned | Unassigned | 982_0 | Clustered | 62 | VC_982_0 | 62 | 0.8646 | 1 | 0.8646 | 0 | 0 | 0 | 0.743 | Ref |
| 3906 | KX507361 | Unassigned | Unassigned | Unassigned | 888_0 | Clustered | 28 | VC_888_0 | 28 | 0.5497 | 1 | 0.5497 | 0 | 0 | 0 | 1 | Ref |
| 3907 | KX507362 | Unassigned | Unassigned | Unassigned | 514_0 | Clustered | 75 | VC_514_0 | 75 | 0.9402 | 1 | 0.9402 | 0 | 0 | 0 | 1 | Ref |
| 3908 | KX513864 | Unassigned | Unassigned | Unassigned | 1123_0 | Clustered | 2 | VC_1123_0 | 2 | 0.3187 | 0.93270267 | 0.2972 | 0 | 0 | 0 | 1 | Ref |
| 3909 | KX513865 | Unassigned | Unassigned | Unassigned | 1104_0 | Clustered | 154 | VC_1104_0 | 154 | 0.8484 | 1 | 0.8484 | 0 | 0 | 0 | 1 | Ref |
| 3910 | KX513866 | Unassigned | Unassigned | Unassigned | 1119_0 | Clustered | 20 | VC_1119_0 | 20 | 0.6751 | 0.99997892 | 0.6751 | 0 | 0 | 0 | 1 | Ref |
| 3911 | KX513867 | Unassigned | Unassigned | Unassigned | 1143_0 | Clustered | 9 | VC_1143_0 | 9 | 0.7268 | 1 | 0.7268 | 0 | 0 | 0 | 1 | Ref |
| 3912 | KX513868 | Unassigned | Unassigned | Unassigned | 1127_0 | Clustered | 19 | VC_1127_0 | 19 | 0.7611 | 1 | 0.7611 | 0 | 0 | 0 | 1 | Ref |
| 3913 | KX513869 | Unassigned | Unassigned | Unassigned | 1056_0 | Clustered | 4 | VC_1056_0 | 4 | 0.169 | 0.84110627 | 0.1421 | 0 | 0 | 0 | 1 | Ref |
| 3914 | KX513870 | Unassigned | Unassigned | Unassigned | 1119_0 | Clustered | 20 | VC_1119_0 | 20 | 0.6751 | 0.99997892 | 0.6751 | 0 | 0 | 0 | 1 | Ref |
| 3915 | KX513871 | Unassigned | Unassigned | Unassigned | 1191_0 | Clustered | 3 | VC_1191_0 | 3 | 0.1408 | 0.98976974 | 0.1394 | 0 | 0 | 0 | 1 | Ref |
| 3916 | KX513872 | Unassigned | Unassigned | Unassigned | 1143_0 | Clustered | 9 | VC_1143_0 | 9 | 0.7268 | 1 | 0.7268 | 0 | 0 | 0 | 1 | Ref |
| 3917 | KX513873 | Unassigned | Unassigned | Unassigned | 1122_0 | Clustered | 10 | VC_1122_0 | 10 | 0.9643 | 0.99999697 | 0.9643 | 0 | 0 | 0 | 1 | Ref |
| 3918 | KX513874 | Unassigned | Unassigned | Unassigned | 1143_0 | Clustered | 9 | VC_1143_0 | 9 | 0.7268 | 1 | 0.7268 | 0 | 0 | 0 | 1 | Ref |
| 3919 | KX513875 | Unassigned | Unassigned | Unassigned | 1197_0 | Clustered | 15 | VC_1197_0 | 15 | 0.8991 | 0.76608394 | 0.6888 | 0 | 0 | 0 | 1 | Ref |
| 3920 | KX513877 | Unassigned | Unassigned | Unassigned | 1127_0 | Clustered | 19 | VC_1127_0 | 19 | 0.7611 | 1 | 0.7611 | 0 | 0 | 0 | 1 | Ref |
| 3921 | KX522565 | Unassigned | Unassigned | Unassigned | 22_0 | Clustered | 11 | VC_22_0 | 11 | 0.5163 | 1 | 0.5163 | 0 | 0 | 0 | 1 | Ref |
| 3922 | KX522649 | Unassigned | Unassigned | Unassigned | 325_21 | Clustered | 5 | VC_325_21 | 5 | 0.0209 | 0.99999976 | 0.0209 | 0 | 0 | 0 | 0.8 | Ref |
| 3923 | KX522650 | Unassigned | Unassigned | Unassigned | 1074_0 | Clustered | 19 | VC_1074_0 | 19 | 0.9198 | 1 | 0.9198 | 0 | 0 | 0 | 1 | Ref |
| 3924 | KX522943 | Unassigned | Unassigned | Unassigned | 325_21 | Clustered | 5 | VC_325_21 | 5 | 0.0209 | 0.99999976 | 0.0209 | 0 | 0 | 0 | 0.8 | Ref |
| 3925 | KX523125 | Unassigned | Unassigned | Unassigned | 514_0 | Clustered | 75 | VC_514_0 | 75 | 0.9402 | 1 | 0.9402 | 0 | 0 | 0 | 1 | Ref |
| 3926 | KX523699 | Unassigned | Unassigned | Unassigned | 461_0 | Clustered | 96 | VC_461_0 | 96 | 0.9307 | 1 | 0.9307 | 0 | 0 | 0 | 0.9575 | Ref |
| 3927 | KX532239 | Unassigned | Unassigned | Unassigned | 153_0 | Clustered | 25 | VC_153_0 | 25 | 0.7729 | 1 | 0.7729 | 0 | 0 | 0 | 1 | Ref |
| 3928 | KX534004 | Unassigned | Unassigned | Unassigned | 761_0 | Clustered | 67 | VC_761_0 | 67 | 0.8947 | 1 | 0.8947 | 0 | 0 | 0 | 1 | Ref |
| 3929 | KX534335 | Unassigned | Unassigned | Unassigned | 835_0 | Clustered | 67 | VC_835_0 | 67 | 0.9624 | 1 | 0.9624 | 0 | 0 | 0 | 1 | Ref |
| 3930 | KX534336 | Unassigned | Unassigned | Unassigned | 835_0 | Clustered | 67 | VC_835_0 | 67 | 0.9624 | 1 | 0.9624 | 0 | 0 | 0 | 1 | Ref |
| 3931 | KX534337 | Unassigned | Unassigned | Unassigned | 795_0 | Clustered | 19 | VC_795_0 | 19 | 0.6848 | 1 | 0.6848 | 0 | 0 | 0 | 1 | Ref |
| 3932 | KX534338 | Unassigned | Unassigned | Unassigned | 835_0 | Clustered | 67 | VC_835_0 | 67 | 0.9624 | 1 | 0.9624 | 0 | 0 | 0 | 1 | Ref |
| 3933 | KX534339 | Unassigned | Unassigned | Unassigned | 835_0 | Clustered | 67 | VC_835_0 | 67 | 0.9624 | 1 | 0.9624 | 0 | 0 | 0 | 1 | Ref |
| 3934 | KX534340 | Unassigned | Unassigned | Unassigned | 20_0 | Clustered | 5 | VC_20_0 | 5 | 0.4387 | 0.99999934 | 0.4387 | 0 | 0 | 0 | 1 | Ref |
| 3935 | KX534341 | Unassigned | Unassigned | Unassigned | 835_0 | Clustered | 67 | VC_835_0 | 67 | 0.9624 | 1 | 0.9624 | 0 | 0 | 0 | 1 | Ref |
| 3936 | KX550065 | Unassigned | Unassigned | Unassigned | 412_0 | Clustered | 39 | VC_412_0 | 39 | 0.845 | 1 | 0.845 | 0 | 0 | 0 | 1 | Ref |
| 3937 | KX550082 | Unassigned | Unassigned | Unassigned | 956_0 | Clustered | 39 | VC_956_0 | 39 | 1 | 1 | 1 | 0 | 0 | 0 | 1 | Ref |
| 3938 | KX550441 | Unassigned | Unassigned | Unassigned | 517_1 | Clustered | 108 | VC_517_1 | 108 | 0.8483 | 1 | 0.8483 | 0 | 0 | 0 | 0.9955 | Ref |
| 3939 | KX550442 | Unassigned | Unassigned | Unassigned | 517_0 | Clustered | 18 | VC_517_0 | 18 | 0.1599 | 1 | 0.1599 | 0 | 0 | 0 | 1 | Ref |
| 3940 | KX550443 | Unassigned | Unassigned | Unassigned | 325_53 | Clustered | 7 | VC_325_53 | 7 | 0.0242 | 1 | 0.0242 | 0 | 0 | 0 | 0.9048 | Ref |
| 3941 | KX552041 | Unassigned | Unassigned | Unassigned | 847_1 | Clustered | 26 | VC_847_1 | 26 | 0.6323 | 1 | 0.6323 | 0 | 0 | 0 | 0.9815 | Ref |
| 3942 | KX555527 | Unassigned | Unassigned | Unassigned | 861_0 | Clustered | 9 | VC_861_0 | 9 | 0.5869 | 1 | 0.5869 | 0 | 0 | 0 | 1 | Ref |
| 3943 | KX557234 | Unassigned | Unassigned | Unassigned | 517_1 | Clustered | 108 | VC_517_1 | 108 | 0.8483 | 1 | 0.8483 | 0 | 0 | 0 | 0.9955 | Ref |
| 3944 | KX557272 | Unassigned | Unassigned | Unassigned | 1182_0 | Clustered | 4 | VC_1182_0 | 4 | 0.2845 | 0.99997676 | 0.2845 | 0 | 0 | 0 | 1 | Ref |
| 3945 | KX557273 | Unassigned | Unassigned | Unassigned | 525_0 | Clustered | 25 | VC_525_0 | 25 | 0.4597 | 1 | 0.4597 | 0 | 0 | 0 | 1 | Ref |
| 3946 | KX557274 | Unassigned | Unassigned | Unassigned | 1176_0 | Clustered | 41 | VC_1176_0 | 41 | 0.9558 | 1 | 0.9558 | 0 | 0 | 0 | 0.9976 | Ref |
| 3947 | KX557275 | Unassigned | Unassigned | Unassigned | 1176_0 | Clustered | 41 | VC_1176_0 | 41 | 0.9558 | 1 | 0.9558 | 0 | 0 | 0 | 0.9976 | Ref |
| 3948 | KX557276 | Unassigned | Unassigned | Unassigned | 1181_1 | Clustered | 10 | VC_1181_1 | 10 | 0.5828 | 1 | 0.5828 | 0 | 0 | 0 | 1 | Ref |
| 3949 | KX557278 | Unassigned | Unassigned | Unassigned | 1161_0 | Clustered | 2 | VC_1161_0 | 2 | 0.0296 | 0.9573719 | 0.0283 | 0 | 0 | 0 | 1 | Ref |
| 3950 | KX557279 | Unassigned | Unassigned | Unassigned | 1177_0 | Clustered | 14 | VC_1177_0 | 14 | 0.7847 | 1 | 0.7847 | 0 | 0 | 0 | 0.989 | Ref |
| 3951 | KX557280 | Unassigned | Unassigned | Unassigned | 224_0 | Clustered | 19 | VC_224_0 | 19 | 0.9416 | 1 | 0.9416 | 0 | 0 | 0 | 1 | Ref |
| 3952 | KX557281 | Unassigned | Unassigned | Unassigned | 867_0 | Clustered | 6 | VC_867_0 | 6 | 0.3202 | 1 | 0.3202 | 0 | 0 | 0 | 1 | Ref |
| 3953 | KX557282 | Unassigned | Unassigned | Unassigned | 1184_0 | Clustered | 5 | VC_1184_0 | 5 | 0.6591 | 0.99999982 | 0.6591 | 0 | 0 | 0 | 1 | Ref |
| 3954 | KX557283 | Unassigned | Unassigned | Unassigned | 224_0 | Clustered | 19 | VC_224_0 | 19 | 0.9416 | 1 | 0.9416 | 0 | 0 | 0 | 1 | Ref |
| 3955 | KX557284 | Unassigned | Unassigned | Unassigned | 224_0 | Clustered | 19 | VC_224_0 | 19 | 0.9416 | 1 | 0.9416 | 0 | 0 | 0 | 1 | Ref |
| 3956 | KX557285 | Unassigned | Unassigned | Unassigned | 570_0 | Clustered | 9 | VC_570_0 | 9 | 0.499 | 1 | 0.499 | 0 | 0 | 0 | 1 | Ref |
| 3957 | KX557286 | Unassigned | Unassigned | Unassigned | 1178_0 | Clustered | 24 | VC_1178_0 | 24 | 0.9524 | 1 | 0.9524 | 0 | 0 | 0 | 1 | Ref |
| 3958 | KX557287 | Unassigned | Unassigned | Unassigned | 525_0 | Clustered | 25 | VC_525_0 | 25 | 0.4597 | 1 | 0.4597 | 0 | 0 | 0 | 1 | Ref |
| 3959 | KX574454 | Unassigned | Unassigned | Unassigned | 325_21 | Clustered | 5 | VC_325_21 | 5 | 0.0209 | 0.99999976 | 0.0209 | 0 | 0 | 0 | 0.8 | Ref |
| 3960 | KX574455 | Unassigned | Unassigned | Unassigned | 215_1 | Clustered | 19 | VC_215_1 | 19 | 0.4188 | 1 | 0.4188 | 0 | 0 | 0 | 1 | Ref |
| 3961 | KX576638 | Unassigned | Unassigned | Unassigned | 223_0 | Clustered | 38 | VC_223_0 | 38 | 0.9095 | 1 | 0.9095 | 0 | 0 | 0 | 1 | Ref |
| 3962 | KX576639 | Unassigned | Unassigned | Unassigned | 216_6 | Clustered | 42 | VC_216_6 | 42 | 0.473 | 1 | 0.473 | 0 | 0 | 0 | 0.8351 | Ref |
| 3963 | KX576640 | Unassigned | Unassigned | Unassigned | 1160_0 | Clustered | 73 | VC_1160_0 | 73 | 0.9835 | 1 | 0.9835 | 0 | 0 | 0 | 1 | Ref |
| 3964 | KX576641 | Unassigned | Unassigned | Unassigned | 1160_0 | Clustered | 73 | VC_1160_0 | 73 | 0.9835 | 1 | 0.9835 | 0 | 0 | 0 | 1 | Ref |
| 3965 | KX576642 | Unassigned | Unassigned | Unassigned | 950_0 | Clustered | 38 | VC_950_0 | 38 | 0.9443 | 1 | 0.9443 | 0 | 0 | 0 | 1 | Ref |
| 3966 | KX576643 | Unassigned | Unassigned | Unassigned | 371_18 | Clustered | 10 | VC_371_18 | 10 | 0.0302 | 1 | 0.0302 | 0 | 0 | 0 | 0.8 | Ref |
| 3967 | KX576644 | Unassigned | Unassigned | Unassigned | 520_77 | Clustered | 3 | VC_520_77 | 3 | 0.0218 | 0.99859911 | 0.0218 | 0 | 0 | 0 | 1 | Ref |
| 3968 | KX576645 | Unassigned | Unassigned | Unassigned | 371_13 | Clustered | 33 | VC_371_13 | 33 | 0.179 | 1 | 0.179 | 0 | 0 | 0 | 0.8769 | Ref |
| 3969 | KX576646 | Unassigned | Unassigned | Unassigned | 371_13 | Clustered | 33 | VC_371_13 | 33 | 0.179 | 1 | 0.179 | 0 | 0 | 0 | 0.8769 | Ref |
| 3970 | KX576647 | Unassigned | Unassigned | Unassigned | 371_13 | Clustered | 33 | VC_371_13 | 33 | 0.179 | 1 | 0.179 | 0 | 0 | 0 | 0.8769 | Ref |
| 3971 | KX578042 | Unassigned | Unassigned | Unassigned | 862_0 | Clustered | 11 | VC_862_0 | 11 | 0.6745 | 1 | 0.6745 | 0 | 0 | 0 | 1 | Ref |
| 3972 | KX578043 | Unassigned | Unassigned | Unassigned | 862_0 | Clustered | 11 | VC_862_0 | 11 | 0.6745 | 1 | 0.6745 | 0 | 0 | 0 | 1 | Ref |
| 3973 | KX578044 | Unassigned | Unassigned | Unassigned | 862_0 | Clustered | 11 | VC_862_0 | 11 | 0.6745 | 1 | 0.6745 | 0 | 0 | 0 | 1 | Ref |
| 3974 | KX578071 | Unassigned | Unassigned | Unassigned | 371_15 | Clustered | 44 | VC_371_15 | 44 | 0.1391 | 1 | 0.1391 | 0 | 0 | 0 | 0.7474 | Ref |
| 3975 | KX579975 | Unassigned | Unassigned | Unassigned | 517_0 | Clustered | 18 | VC_517_0 | 18 | 0.1599 | 1 | 0.1599 | 0 | 0 | 0 | 1 | Ref |
| 3976 | KX580961 | Unassigned | Unassigned | Unassigned | 888_0 | Clustered | 28 | VC_888_0 | 28 | 0.5497 | 1 | 0.5497 | 0 | 0 | 0 | 1 | Ref |
| 3977 | KX580962 | Unassigned | Unassigned | Unassigned | 888_0 | Clustered | 28 | VC_888_0 | 28 | 0.5497 | 1 | 0.5497 | 0 | 0 | 0 | 1 | Ref |
| 3978 | KX581090 | Unassigned | Unassigned | Unassigned | 299_0 | Clustered | 24 | VC_299_0 | 24 | 0.9905 | 1 | 0.9905 | 0 | 0 | 0 | 1 | Ref |
| 3979 | KX581091 | Unassigned | Unassigned | Unassigned | 299_0 | Clustered | 24 | VC_299_0 | 24 | 0.9905 | 1 | 0.9905 | 0 | 0 | 0 | 1 | Ref |
| 3980 | KX581092 | Unassigned | Unassigned | Unassigned | 299_0 | Clustered | 24 | VC_299_0 | 24 | 0.9905 | 1 | 0.9905 | 0 | 0 | 0 | 1 | Ref |
| 3981 | KX581093 | Unassigned | Unassigned | Unassigned | 299_0 | Clustered | 24 | VC_299_0 | 24 | 0.9905 | 1 | 0.9905 | 0 | 0 | 0 | 1 | Ref |
| 3982 | KX581094 | Unassigned | Unassigned | Unassigned | 299_0 | Clustered | 24 | VC_299_0 | 24 | 0.9905 | 1 | 0.9905 | 0 | 0 | 0 | 1 | Ref |
| 3983 | KX581095 | Unassigned | Unassigned | Unassigned | 299_0 | Clustered | 24 | VC_299_0 | 24 | 0.9905 | 1 | 0.9905 | 0 | 0 | 0 | 1 | Ref |
| 3984 | KX581096 | Unassigned | Unassigned | Unassigned | 299_0 | Clustered | 24 | VC_299_0 | 24 | 0.9905 | 1 | 0.9905 | 0 | 0 | 0 | 1 | Ref |
| 3985 | KX581097 | Unassigned | Unassigned | Unassigned | 299_0 | Clustered | 24 | VC_299_0 | 24 | 0.9905 | 1 | 0.9905 | 0 | 0 | 0 | 1 | Ref |
| 3986 | KX581098 | Unassigned | Unassigned | Unassigned | 299_0 | Clustered | 24 | VC_299_0 | 24 | 0.9905 | 1 | 0.9905 | 0 | 0 | 0 | 1 | Ref |
| 3987 | KX581099 | Unassigned | Unassigned | Unassigned | 299_0 | Clustered | 24 | VC_299_0 | 24 | 0.9905 | 1 | 0.9905 | 0 | 0 | 0 | 1 | Ref |
| 3988 | KX581100 | Unassigned | Unassigned | Unassigned | 299_0 | Clustered | 24 | VC_299_0 | 24 | 0.9905 | 1 | 0.9905 | 0 | 0 | 0 | 1 | Ref |
| 3989 | KX585251 | Unassigned | Unassigned | Unassigned | 238_0 | Clustered | 17 | VC_238_0 | 17 | 0.6109 | 1 | 0.6109 | 0 | 0 | 0 | 1 | Ref |
| 3990 | KX585252 | Unassigned | Unassigned | Unassigned | 225_0 | Clustered | 32 | VC_225_0 | 32 | 0.848 | 1 | 0.848 | 0 | 0 | 0 | 0.996 | Ref |
| 3991 | KX585253 | Unassigned | Unassigned | Unassigned | 236_0 | Clustered | 72 | VC_236_0 | 72 | 0.8104 | 1 | 0.8104 | 0 | 0 | 0 | 1 | Ref |
| 3992 | KX587949 | Unassigned | Unassigned | Unassigned | 111_0 | Clustered | 91 | VC_111_0 | 91 | 0.9587 | 1 | 0.9587 | 0 | 0 | 0 | 1 | Ref |
| 3993 | KX588251 | Unassigned | Unassigned | Unassigned | 761_0 | Clustered | 67 | VC_761_0 | 67 | 0.8947 | 1 | 0.8947 | 0 | 0 | 0 | 1 | Ref |
| 3994 | KX589269 | Unassigned | Unassigned | Unassigned | 372_0 | Clustered | 24 | VC_372_0 | 24 | 0.5539 | 1 | 0.5539 | 0 | 0 | 0 | 1 | Ref |
| 3995 | KX591654 | Unassigned | Unassigned | Unassigned | 823_0 | Clustered | 124 | VC_823_0 | 124 | 0.9492 | 1 | 0.9492 | 0 | 0 | 0 | 1 | Ref |
| 3996 | KX592589 | Unassigned | Unassigned | Unassigned | 371_11 | Clustered | 31 | VC_371_11 | 31 | 0.0927 | 1 | 0.0927 | 0 | 0 | 0 | 0.7011 | Ref |
| 3997 | KX607101 | Unassigned | Unassigned | Unassigned | 447_0 | Clustered | 3 | VC_447_0 | 3 | 0.8268 | 0.99509929 | 0.8227 | 0 | 0 | 0 | 1 | Ref |
| 3998 | KX607102 | Unassigned | Unassigned | Unassigned | 447_0 | Clustered | 3 | VC_447_0 | 3 | 0.8268 | 0.99509929 | 0.8227 | 0 | 0 | 0 | 1 | Ref |
| 3999 | KX610764 | Unassigned | Unassigned | Unassigned | 520_83 | Clustered/Singleton | 1 | VC_520_83 | 1 | 0 | 1 | 0 | 0 | 0 | 0 | 1 | Ref |
| 4000 | KX610765 | Unassigned | Unassigned | Unassigned | 950_0 | Clustered | 38 | VC_950_0 | 38 | 0.9443 | 1 | 0.9443 | 0 | 0 | 0 | 1 | Ref |
| 4001 | KX611788 | Unassigned | Unassigned | Unassigned | 956_0 | Clustered | 39 | VC_956_0 | 39 | 1 | 1 | 1 | 0 | 0 | 0 | 1 | Ref |
| 4002 | KX611831 | Unassigned | Unassigned | Unassigned | 551_6 | Clustered | 22 | VC_551_6 | 22 | 0.1214 | 1 | 0.1214 | 0 | 0 | 0 | 0.8095 | Ref |
| 4003 | KX619650 | Unassigned | Unassigned | Unassigned | 216_6 | Clustered | 42 | VC_216_6 | 42 | 0.473 | 1 | 0.473 | 0 | 0 | 0 | 0.8351 | Ref |
| 4004 | KX619651 | Unassigned | Unassigned | Unassigned | 517_1 | Clustered | 108 | VC_517_1 | 108 | 0.8483 | 1 | 0.8483 | 0 | 0 | 0 | 0.9955 | Ref |
| 4005 | KX620750 | Unassigned | Unassigned | Unassigned | 1186_0 | Clustered | 4 | VC_1186_0 | 4 | 0.4582 | 0.99991945 | 0.4582 | 0 | 0 | 0 | 1 | Ref |
| 4006 | KX620751 | Unassigned | Unassigned | Unassigned | 1186_0 | Clustered | 4 | VC_1186_0 | 4 | 0.4582 | 0.99991945 | 0.4582 | 0 | 0 | 0 | 1 | Ref |
| 4007 | KX620753 | Unassigned | Unassigned | Unassigned | 1186_0 | Clustered | 4 | VC_1186_0 | 4 | 0.4582 | 0.99991945 | 0.4582 | 0 | 0 | 0 | 1 | Ref |
| 4008 | KX620754 | Unassigned | Unassigned | Unassigned | 1186_0 | Clustered | 4 | VC_1186_0 | 4 | 0.4582 | 0.99991945 | 0.4582 | 0 | 0 | 0 | 1 | Ref |
| 4009 | KX620786 | Unassigned | Unassigned | Unassigned | 371_16 | Clustered | 42 | VC_371_16 | 42 | 0.1341 | 1 | 0.1341 | 0 | 0 | 0 | 0.7282 | Ref |
| 4010 | KX621004 | Unassigned | Unassigned | Unassigned | 1160_0 | Clustered | 73 | VC_1160_0 | 73 | 0.9835 | 1 | 0.9835 | 0 | 0 | 0 | 1 | Ref |
| 4011 | KX621005 | Unassigned | Unassigned | Unassigned | 1160_0 | Clustered | 73 | VC_1160_0 | 73 | 0.9835 | 1 | 0.9835 | 0 | 0 | 0 | 1 | Ref |
| 4012 | KX621007 | Unassigned | Unassigned | Unassigned | 234_0 | Clustered | 21 | VC_234_0 | 21 | 0.8355 | 1 | 0.8355 | 0 | 0 | 0 | 1 | Ref |
| 4013 | KX636165 | Unassigned | Unassigned | Unassigned | 238_0 | Clustered | 17 | VC_238_0 | 17 | 0.6109 | 1 | 0.6109 | 0 | 0 | 0 | 1 | Ref |
| 4014 | KX640831 | Unassigned | Unassigned | Unassigned | 513_0 | Clustered | 32 | VC_513_0 | 32 | 0.6881 | 1 | 0.6881 | 0 | 0 | 0 | 1 | Ref |
| 4015 | KX641260 | Unassigned | Unassigned | Unassigned | 517_0 | Clustered | 18 | VC_517_0 | 18 | 0.1599 | 1 | 0.1599 | 0 | 0 | 0 | 1 | Ref |
| 4016 | KX641261 | Unassigned | Unassigned | Unassigned | 223_0 | Clustered | 38 | VC_223_0 | 38 | 0.9095 | 1 | 0.9095 | 0 | 0 | 0 | 1 | Ref |
| 4017 | KX641262 | Unassigned | Unassigned | Unassigned | 519_0 | Clustered | 39 | VC_519_0 | 39 | 0.8156 | 1 | 0.8156 | 0 | 0 | 0 | 1 | Ref |
| 4018 | KX641263 | Unassigned | Unassigned | Unassigned | 216_3 | Clustered | 8 | VC_216_3 | 8 | 0.0701 | 1 | 0.0701 | 0 | 0 | 0 | 0.8571 | Ref |
| 4019 | KX641264 | Unassigned | Unassigned | Unassigned | 236_0 | Clustered | 72 | VC_236_0 | 72 | 0.8104 | 1 | 0.8104 | 0 | 0 | 0 | 1 | Ref |
| 4020 | KX641265 | Unassigned | Unassigned | Unassigned | 761_0 | Clustered | 67 | VC_761_0 | 67 | 0.8947 | 1 | 0.8947 | 0 | 0 | 0 | 1 | Ref |
| 4021 | KX641266 | Unassigned | Unassigned | Unassigned | 761_0 | Clustered | 67 | VC_761_0 | 67 | 0.8947 | 1 | 0.8947 | 0 | 0 | 0 | 1 | Ref |
| 4022 | KX648373 | Unassigned | Unassigned | Unassigned | 517_1 | Clustered | 108 | VC_517_1 | 108 | 0.8483 | 1 | 0.8483 | 0 | 0 | 0 | 0.9955 | Ref |
| 4023 | KX649889 | Unassigned | Unassigned | Unassigned | 637_0 | Clustered | 115 | VC_637_0 | 115 | 0.731 | 1 | 0.731 | 0 | 0 | 0 | 0.8764 | Ref |
| 4024 | KX656785 | Unassigned | Unassigned | Unassigned | 687_0 | Clustered | 3 | VC_687_0 | 3 | 0.5872 | 0.99252665 | 0.5828 | 0 | 0 | 0 | 1 | Ref |
| 4025 | KX657793 | Unassigned | Unassigned | Unassigned | 222_0 | Clustered | 5 | VC_222_0 | 5 | 0.0565 | 0.99999997 | 0.0565 | 0 | 0 | 0 | 1 | Ref |
| 4026 | KX657794 | Unassigned | Unassigned | Unassigned | 236_0 | Clustered | 72 | VC_236_0 | 72 | 0.8104 | 1 | 0.8104 | 0 | 0 | 0 | 1 | Ref |
| 4027 | KX657795 | Unassigned | Unassigned | Unassigned | 236_0 | Clustered | 72 | VC_236_0 | 72 | 0.8104 | 1 | 0.8104 | 0 | 0 | 0 | 1 | Ref |
| 4028 | KX657796 | Unassigned | Unassigned | Unassigned | 236_0 | Clustered | 72 | VC_236_0 | 72 | 0.8104 | 1 | 0.8104 | 0 | 0 | 0 | 1 | Ref |
| 4029 | KX660669 | Unassigned | Unassigned | Unassigned | 70_0 | Clustered | 27 | VC_70_0 | 27 | 0.5981 | 1 | 0.5981 | 0 | 0 | 0 | 0.9886 | Ref |
| 4030 | KX664447 | Unassigned | Unassigned | Unassigned | 225_0 | Clustered | 32 | VC_225_0 | 32 | 0.848 | 1 | 0.848 | 0 | 0 | 0 | 0.996 | Ref |
| 4031 | KX664448 | Unassigned | Unassigned | Unassigned | 514_0 | Clustered | 75 | VC_514_0 | 75 | 0.9402 | 1 | 0.9402 | 0 | 0 | 0 | 1 | Ref |
| 4032 | KX664455 | Unassigned | Unassigned | Unassigned | 761_0 | Clustered | 67 | VC_761_0 | 67 | 0.8947 | 1 | 0.8947 | 0 | 0 | 0 | 1 | Ref |
| 4033 | KX664695 | Unassigned | Unassigned | Unassigned | 847_1 | Clustered | 26 | VC_847_1 | 26 | 0.6323 | 1 | 0.6323 | 0 | 0 | 0 | 0.9815 | Ref |
| 4034 | KX669227 | Unassigned | Unassigned | Unassigned | 451_0 | Clustered | 80 | VC_451_0 | 80 | 0.9991 | 1 | 0.9991 | 0 | 0 | 0 | 1 | Ref |
| 4035 | KX670786 | Unassigned | Unassigned | Unassigned | 1074_0 | Clustered | 19 | VC_1074_0 | 19 | 0.9198 | 1 | 0.9198 | 0 | 0 | 0 | 1 | Ref |
| 4036 | KX670787 | Unassigned | Unassigned | Unassigned | 1074_0 | Clustered | 19 | VC_1074_0 | 19 | 0.9198 | 1 | 0.9198 | 0 | 0 | 0 | 1 | Ref |
| 4037 | KX670788 | Unassigned | Unassigned | Unassigned | 514_0 | Clustered | 75 | VC_514_0 | 75 | 0.9402 | 1 | 0.9402 | 0 | 0 | 0 | 1 | Ref |
| 4038 | KX670789 | Unassigned | Unassigned | Unassigned | 872_0 | Clustered | 31 | VC_872_0 | 31 | 0.8151 | 1 | 0.8151 | 0 | 0 | 0 | 0.8194 | Ref |
| 4039 | KX670790 | Unassigned | Unassigned | Unassigned | 872_0 | Clustered | 31 | VC_872_0 | 31 | 0.8151 | 1 | 0.8151 | 0 | 0 | 0 | 0.8194 | Ref |
| 4040 | KX670813 | Unassigned | Unassigned | Unassigned | 371_15 | Clustered | 44 | VC_371_15 | 44 | 0.1391 | 1 | 0.1391 | 0 | 0 | 0 | 0.7474 | Ref |
| 4041 | KX670828 | Unassigned | Unassigned | Unassigned | 513_0 | Clustered | 32 | VC_513_0 | 32 | 0.6881 | 1 | 0.6881 | 0 | 0 | 0 | 1 | Ref |
| 4042 | KX683292 | Unassigned | Unassigned | Unassigned | 371_15 | Clustered | 44 | VC_371_15 | 44 | 0.1391 | 1 | 0.1391 | 0 | 0 | 0 | 0.7474 | Ref |
| 4043 | KX683293 | Unassigned | Unassigned | Unassigned | 371_15 | Clustered | 44 | VC_371_15 | 44 | 0.1391 | 1 | 0.1391 | 0 | 0 | 0 | 0.7474 | Ref |
| 4044 | KX683423 | Unassigned | Unassigned | Unassigned | 513_0 | Clustered | 32 | VC_513_0 | 32 | 0.6881 | 1 | 0.6881 | 0 | 0 | 0 | 1 | Ref |
| 4045 | KX683428 | Unassigned | Unassigned | Unassigned | 237_1 | Clustered | 13 | VC_237_1 | 13 | 0.1773 | 1 | 0.1773 | 0 | 0 | 0 | 0.7308 | Ref |
| 4046 | KX683875 | Unassigned | Unassigned | Unassigned | 216_4 | Clustered | 13 | VC_216_4 | 13 | 0.1657 | 1 | 0.1657 | 0 | 0 | 0 | 1 | Ref |
| 4047 | KX683876 | Unassigned | Unassigned | Unassigned | 325_38 | Clustered | 6 | VC_325_38 | 6 | 0.0267 | 1 | 0.0267 | 0 | 0 | 0 | 0.8 | Ref |
| 4048 | KX685355 | Unassigned | Unassigned | Unassigned | 225_0 | Clustered | 32 | VC_225_0 | 32 | 0.848 | 1 | 0.848 | 0 | 0 | 0 | 0.996 | Ref |

| | | | | | | | | | | | | | | | | | |
|---|---|---|---|---|---|---|---|---|---|---|---|---|---|---|---|---|---|
| 4049 | KX688102 | Unassigned | Unassigned | Unassigned | 1160_0 | Clustered | 73 | VC_1160_0 | 73 | 0.9835 | 1 | 0.9835 | 0 | 0 | 0 | 1 | Ref |
| 4050 | KX688103 | Unassigned | Unassigned | Unassigned | 1160_0 | Clustered | 73 | VC_1160_0 | 73 | 0.9835 | 1 | 0.9835 | 0 | 0 | 0 | 1 | Ref |
| 4051 | KX689784 | Unassigned | Unassigned | Unassigned | 451_0 | Clustered | 80 | VC_451_0 | 80 | 0.9991 | 1 | 0.9991 | 0 | 0 | 0 | 1 | Ref |
| 4052 | KX702319 | Unassigned | Unassigned | Unassigned | 371_13 | Clustered | 33 | VC_371_13 | 33 | 0.179 | 1 | 0.179 | 0 | 0 | 0 | 0.8769 | Ref |
| 4053 | KX702320 | Unassigned | Unassigned | Unassigned | 325_38 | Clustered | 6 | VC_325_38 | 6 | 0.0267 | 1 | 0.0267 | 0 | 0 | 0 | 0.8 | Ref |
| 4054 | KX711710 | Unassigned | Unassigned | Unassigned | 423_0 | Clustered | 45 | VC_423_0 | 45 | 0.9484 | 1 | 0.9484 | 0 | 0 | 0 | 1 | Ref |
| 4055 | KX712070 | Unassigned | Unassigned | Unassigned | 823_0 | Clustered | 124 | VC_823_0 | 124 | 0.9492 | 1 | 0.9492 | 0 | 0 | 0 | 1 | Ref |
| 4056 | KX712071 | Unassigned | Unassigned | Unassigned | 823_0 | Clustered | 124 | VC_823_0 | 124 | 0.9492 | 1 | 0.9492 | 0 | 0 | 0 | 1 | Ref |
| 4057 | KX712143 | Unassigned | Unassigned | Unassigned | 348_0 | Clustered | 22 | VC_348_0 | 22 | 0.9602 | 1 | 0.9602 | 0 | 0 | 0 | 1 | Ref |
| 4058 | KX712236 | Unassigned | Unassigned | Unassigned | 956_0 | Clustered | 39 | VC_956_0 | 39 | 1 | 1 | 1 | 0 | 0 | 0 | 1 | Ref |
| 4059 | KX712237 | Unassigned | Unassigned | Unassigned | 956_0 | Clustered | 39 | VC_956_0 | 39 | 1 | 1 | 1 | 0 | 0 | 0 | 1 | Ref |
| 4060 | KX712238 | Unassigned | Unassigned | Unassigned | 325_43 | Clustered/Singleton | 1 | VC_325_43 | 1 | 0 | 1 | 0 | 0 | 0 | 0 | 1 | Ref |
| 4061 | KX721255 | Unassigned | Unassigned | Unassigned | 558_49 | Clustered | 7 | VC_558_49 | 7 | 0.0194 | 0.99510369 | 0.0193 | 0 | 0 | 0 | 0.8571 | Ref |
| 4062 | KX721256 | Unassigned | Unassigned | Unassigned | 558_48 | Clustered | 2 | VC_558_48 | 2 | 0.0032 | 0.69466126 | 0.0022 | 0 | 0 | 0 | 1 | Ref |
| 4063 | KX752698 | Unassigned | Unassigned | Unassigned | 558_81 | Clustered/Singleton | 1 | VC_558_81 | 1 | 0 | 1 | 0 | 0 | 0 | 0 | 1 | Ref |
| 4064 | KX756439 | Unassigned | Unassigned | Unassigned | 521_0 | Clustered | 37 | VC_521_0 | 37 | 0.912 | 1 | 0.912 | 0 | 0 | 0 | 1 | Ref |
| 4065 | KX756572 | Unassigned | Unassigned | Unassigned | 878_0 | Clustered | 32 | VC_878_0 | 32 | 0.9965 | 1 | 0.9965 | 0 | 0 | 0 | 1 | Ref |
| 4066 | KX758538 | Unassigned | Unassigned | Unassigned | 215_1 | Clustered | 19 | VC_215_1 | 19 | 0.4188 | 1 | 0.4188 | 0 | 0 | 0 | 1 | Ref |
| 4067 | KX758539 | Unassigned | Unassigned | Unassigned | 217_0 | Clustered | 5 | VC_217_0 | 5 | 0.069 | 0.99999998 | 0.069 | 0 | 0 | 0 | 1 | Ref |
| 4068 | KX765862 | Unassigned | Unassigned | Unassigned | 592_0 | Clustered | 50 | VC_592_0 | 50 | 0.7314 | 1 | 0.7314 | 0 | 0 | 0 | 1 | Ref |
| 4069 | KX765863 | Unassigned | Unassigned | Unassigned | 592_0 | Clustered | 50 | VC_592_0 | 50 | 0.7314 | 1 | 0.7314 | 0 | 0 | 0 | 1 | Ref |
| 4070 | KX765864 | Unassigned | Unassigned | Unassigned | 592_0 | Clustered | 50 | VC_592_0 | 50 | 0.7314 | 1 | 0.7314 | 0 | 0 | 0 | 1 | Ref |
| 4071 | KX765865 | Unassigned | Unassigned | Unassigned | 133_15 | Clustered | 2 | VC_133_15 | 2 | 0.0051 | 0.84786437 | 0.0044 | 0 | 0 | 0 | 1 | Ref |
| 4072 | KX774321 | Unassigned | Unassigned | Unassigned | 1070_1 | Clustered/Singleton | 1 | VC_1070_1 | 1 | 0 | 1 | 0 | 0 | 0 | 0 | 1 | Ref |
| 4073 | KX774374 | Unassigned | Unassigned | Unassigned | 5_0 | Clustered | 30 | VC_5_0 | 30 | 0.9647 | 1 | 0.9647 | 0 | 0 | 0 | 1 | Ref |
| 4074 | KX776161 | Unassigned | Unassigned | Unassigned | 28_1 | Clustered | 53 | VC_28_1 | 53 | 0.7164 | 1 | 0.7164 | 0 | 0 | 0 | 0.9325 | Ref |
| 4075 | KX778611 | Unassigned | Unassigned | Unassigned | 385_0 | Clustered | 51 | VC_385_0 | 51 | 0.8754 | 1 | 0.8754 | 0 | 0 | 0 | 1 | Ref |
| 4076 | KX781992 | Unassigned | Unassigned | Unassigned | 558_9 | Clustered | 3 | VC_558_9 | 3 | 0.0064 | 0.82911529 | 0.0053 | 0 | 0 | 0 | 0.6667 | Ref |
| 4077 | KX808129 | Unassigned | Unassigned | Unassigned | 514_0 | Clustered | 75 | VC_514_0 | 75 | 0.9402 | 1 | 0.9402 | 0 | 0 | 0 | 1 | Ref |
| 4078 | KX808130 | Unassigned | Unassigned | Unassigned | 517_1 | Clustered | 108 | VC_517_1 | 108 | 0.8483 | 1 | 0.8483 | 0 | 0 | 0 | 0.9955 | Ref |
| 4079 | KX808131 | Unassigned | Unassigned | Unassigned | 520_16 | Clustered/Singleton | 1 | VC_520_16 | 1 | 0 | 1 | 0 | 0 | 0 | 0 | 1 | Ref |
| 4080 | KX815338 | Unassigned | Unassigned | Unassigned | 982_0 | Clustered | 62 | VC_982_0 | 62 | 0.8646 | 1 | 0.8646 | 0 | 0 | 0 | 0.743 | Ref |
| 4081 | KX817173 | Unassigned | Unassigned | Unassigned | 551_2 | Clustered | 17 | VC_551_2 | 17 | 0.0973 | 1 | 0.0973 | 0 | 0 | 0 | 0.9779 | Ref |
| 4082 | KX817174 | Unassigned | Unassigned | Unassigned | 517_1 | Clustered | 108 | VC_517_1 | 108 | 0.8483 | 1 | 0.8483 | 0 | 0 | 0 | 0.9955 | Ref |
| 4083 | KX817175 | Unassigned | Unassigned | Unassigned | 517_1 | Clustered | 108 | VC_517_1 | 108 | 0.8483 | 1 | 0.8483 | 0 | 0 | 0 | 0.9955 | Ref |
| 4084 | KX822733 | Unassigned | Unassigned | Unassigned | 496_3 | Clustered/Singleton | 1 | VC_496_3 | 1 | 0 | 1 | 0 | 0 | 0 | 0 | 1 | Ref |
| 4085 | KX826077 | Unassigned | Unassigned | Unassigned | 428_0 | Clustered | 33 | VC_428_0 | 33 | 0.9915 | 1 | 0.9915 | 0 | 0 | 0 | 1 | Ref |
| 4086 | KX827368 | Unassigned | Unassigned | Unassigned | 11_0 | Clustered | 19 | VC_11_0 | 19 | 0.7562 | 1 | 0.7562 | 0 | 0 | 0 | 1 | Ref |
| 4087 | KX827369 | Unassigned | Unassigned | Unassigned | 11_0 | Clustered | 19 | VC_11_0 | 19 | 0.7562 | 1 | 0.7562 | 0 | 0 | 0 | 1 | Ref |
| 4088 | KX827370 | Unassigned | Unassigned | Unassigned | 11_0 | Clustered | 19 | VC_11_0 | 19 | 0.7562 | 1 | 0.7562 | 0 | 0 | 0 | 1 | Ref |
| 4089 | KX827371 | Unassigned | Unassigned | Unassigned | 15_0 | Clustered | 3 | VC_15_0 | 3 | 0.0481 | 0.9985346 | 0.048 | 0 | 0 | 0 | 1 | Ref |
| 4090 | KX828710 | Unassigned | Unassigned | Unassigned | 575_0 | Clustered | 35 | VC_575_0 | 35 | 0.8815 | 1 | 0.8815 | 0 | 0 | 0 | 1 | Ref |
| 4091 | KX828711 | Unassigned | Unassigned | Unassigned | 283_85 | Clustered/Singleton | 1 | VC_283_85 | 1 | 0 | 1 | 0 | 0 | 0 | 0 | 1 | Ref |
| 4092 | KX831080 | Unassigned | Unassigned | Unassigned | 558_82 | Clustered | 3 | VC_558_82 | 3 | 0.0064 | 0.88433874 | 0.0057 | 0 | 0 | 0 | 0.6667 | Ref |
| 4093 | KX833211 | Unassigned | Unassigned | Unassigned | 228_0 | Clustered | 60 | VC_228_0 | 60 | 0.942 | 1 | 0.942 | 0 | 0 | 0 | 1 | Ref |
| 4094 | KX834009 | Unassigned | Unassigned | Unassigned | 551_6 | Clustered | 22 | VC_551_6 | 22 | 0.1214 | 1 | 0.1214 | 0 | 0 | 0 | 0.8095 | Ref |
| 4095 | KX845404 | Unassigned | Unassigned | Unassigned | 192_0 | Clustered | 15 | VC_192_0 | 15 | 0.5071 | 1 | 0.5071 | 0 | 0 | 0 | 1 | Ref |
| 4096 | KX855961 | Unassigned | Unassigned | Unassigned | 1074_0 | Clustered | 19 | VC_1074_0 | 19 | 0.9198 | 1 | 0.9198 | 0 | 0 | 0 | 1 | Ref |
| 4097 | KX855962 | Unassigned | Unassigned | Unassigned | 1074_0 | Clustered | 19 | VC_1074_0 | 19 | 0.9198 | 1 | 0.9198 | 0 | 0 | 0 | 1 | Ref |
| 4098 | KX856662 | Unassigned | Unassigned | Unassigned | 111_0 | Clustered | 91 | VC_111_0 | 91 | 0.9587 | 1 | 0.9587 | 0 | 0 | 0 | 1 | Ref |
| 4099 | KX879601 | Unassigned | Unassigned | Unassigned | 730_0 | Clustered | 9 | VC_730_0 | 9 | 1 | 1 | 1 | 0 | 0 | 0 | 1 | Ref |
| 4100 | KX879602 | Unassigned | Unassigned | Unassigned | 730_0 | Clustered | 9 | VC_730_0 | 9 | 1 | 1 | 1 | 0 | 0 | 0 | 1 | Ref |
| 4101 | KX879627 | Unassigned | Unassigned | Unassigned | 844_0 | Clustered | 27 | VC_844_0 | 27 | 0.9385 | 1 | 0.9385 | 0 | 0 | 0 | 1 | Ref |
| 4102 | KX879641 | Unassigned | Unassigned | Unassigned | 256_0 | Clustered | 14 | VC_256_0 | 14 | 0.2368 | 1 | 0.2368 | 0 | 0 | 0 | 1 | Ref |
| 4103 | KX879642 | Unassigned | Unassigned | Unassigned | 256_0 | Clustered | 14 | VC_256_0 | 14 | 0.2368 | 1 | 0.2368 | 0 | 0 | 0 | 1 | Ref |
| 4104 | KX879643 | Unassigned | Unassigned | Unassigned | 255_0 | Clustered | 13 | VC_255_0 | 13 | 0.2048 | 1 | 0.2048 | 0 | 0 | 0 | 1 | Ref |
| 4105 | KX889068 | Unassigned | Unassigned | Unassigned | 930_0 | Clustered | 7 | VC_930_0 | 7 | 0.519 | 1 | 0.519 | 0 | 0 | 0 | 1 | Ref |
| 4106 | KX897981 | Unassigned | Unassigned | Unassigned | 215_1 | Clustered | 19 | VC_215_1 | 19 | 0.4188 | 1 | 0.4188 | 0 | 0 | 0 | 1 | Ref |
| 4107 | KX898399 | Unassigned | Unassigned | Unassigned | 465_0 | Clustered | 7 | VC_465_0 | 7 | 0.3322 | 1 | 0.3322 | 0 | 0 | 0 | 1 | Ref |
| 4108 | KX898400 | Unassigned | Unassigned | Unassigned | 465_0 | Clustered | 7 | VC_465_0 | 7 | 0.3322 | 1 | 0.3322 | 0 | 0 | 0 | 1 | Ref |
| 4109 | KX912252 | Unassigned | Unassigned | Unassigned | 896_0 | Clustered | 5 | VC_896_0 | 5 | 0.4981 | 0.99999977 | 0.4981 | 0 | 0 | 0 | 1 | Ref |
| 4110 | KX925554 | Unassigned | Unassigned | Unassigned | 960_1 | Clustered/Singleton | 1 | VC_960_1 | 1 | 0 | 1 | 0 | 0 | 0 | 0 | 1 | Ref |
| 4111 | KX961385 | Unassigned | Unassigned | Unassigned | 166_0 | Clustered | 26 | VC_166_0 | 26 | 0.7923 | 1 | 0.7923 | 0 | 0 | 0 | 1 | Ref |
| 4112 | KX961629 | Unassigned | Unassigned | Unassigned | 884_0 | Clustered | 28 | VC_884_0 | 28 | 0.6211 | 1 | 0.6211 | 0 | 0 | 0 | 0.9841 | Ref |
| 4113 | KX961630 | Unassigned | Unassigned | Unassigned | 187_0 | Clustered | 4 | VC_187_0 | 4 | 0.0661 | 0.99994098 | 0.0661 | 0 | 0 | 0 | 1 | Ref |
| 4114 | KX961631 | Unassigned | Unassigned | Unassigned | 818_0 | Clustered | 20 | VC_818_0 | 20 | 0.9601 | 1 | 0.9601 | 0 | 0 | 0 | 1 | Ref |
| 4115 | KX961632 | Unassigned | Unassigned | Unassigned | 884_0 | Clustered | 28 | VC_884_0 | 28 | 0.6211 | 1 | 0.6211 | 0 | 0 | 0 | 0.9841 | Ref |
| 4116 | KX987127 | Unassigned | Unassigned | Unassigned | 805_0 | Clustered | 4 | VC_805_0 | 4 | 0.9081 | 0.99622688 | 0.9046 | 0 | 0 | 0 | 1 | Ref |
| 4117 | KX987158 | Unassigned | Unassigned | Unassigned | 1049_0 | Clustered | 8 | VC_1049_0 | 8 | 0.6413 | 1 | 0.6413 | 0 | 0 | 0 | 1 | Ref |
| 4118 | KX987999 | Unassigned | Unassigned | Unassigned | 187_1 | Clustered | 7 | VC_187_1 | 7 | 0.1423 | 1 | 0.1423 | 0 | 0 | 0 | 1 | Ref |
| 4119 | KY000003 | Unassigned | Unassigned | Unassigned | 133_30 | Clustered | 2 | VC_133_30 | 2 | 0.0052 | 0.87738871 | 0.0046 | 0 | 0 | 0 | 1 | Ref |
| 4120 | KY000079 | Unassigned | Unassigned | Unassigned | 702_0 | Clustered | 5 | VC_702_0 | 5 | 0.7794 | 0.99999988 | 0.7794 | 0 | 0 | 0 | 1 | Ref |
| 4121 | KY000080 | Unassigned | Unassigned | Unassigned | 491_0 | Clustered | 36 | VC_491_0 | 36 | 0.5336 | 1 | 0.5336 | 0 | 0 | 0 | 0.9714 | Ref |
| 4122 | KY000081 | Unassigned | Unassigned | Unassigned | 111_0 | Clustered | 91 | VC_111_0 | 91 | 0.9587 | 1 | 0.9587 | 0 | 0 | 0 | 1 | Ref |
| 4123 | KY000082 | Unassigned | Unassigned | Unassigned | 79_11 | Clustered/Singleton | 1 | VC_79_11 | 1 | 0 | 1 | 0 | 0 | 0 | 0 | 1 | Ref |
| 4124 | KY000083 | Unassigned | Unassigned | Unassigned | 76_0 | Clustered | 37 | VC_76_0 | 37 | 0.9403 | 1 | 0.9403 | 0 | 0 | 0 | 1 | Ref |
| 4125 | KY000084 | Unassigned | Unassigned | Unassigned | 96_0 | Clustered | 30 | VC_96_0 | 30 | 0.718 | 1 | 0.718 | 0 | 0 | 0 | 1 | Ref |
| 4126 | KY000085 | Unassigned | Unassigned | Unassigned | 96_0 | Clustered | 30 | VC_96_0 | 30 | 0.718 | 1 | 0.718 | 0 | 0 | 0 | 1 | Ref |
| 4127 | KY000218 | Unassigned | Unassigned | Unassigned | 166_0 | Clustered | 26 | VC_166_0 | 26 | 0.7923 | 1 | 0.7923 | 0 | 0 | 0 | 1 | Ref |
| 4128 | KY000219 | Unassigned | Unassigned | Unassigned | 166_0 | Clustered | 26 | VC_166_0 | 26 | 0.7923 | 1 | 0.7923 | 0 | 0 | 0 | 1 | Ref |
| 4129 | KY000220 | Unassigned | Unassigned | Unassigned | 166_0 | Clustered | 26 | VC_166_0 | 26 | 0.7923 | 1 | 0.7923 | 0 | 0 | 0 | 1 | Ref |
| 4130 | KY000221 | Unassigned | Unassigned | Unassigned | 166_0 | Clustered | 26 | VC_166_0 | 26 | 0.7923 | 1 | 0.7923 | 0 | 0 | 0 | 1 | Ref |
| 4131 | KY002061 | Unassigned | Unassigned | Unassigned | 1049_0 | Clustered | 8 | VC_1049_0 | 8 | 0.6413 | 1 | 0.6413 | 0 | 0 | 0 | 1 | Ref |
| 4132 | KY006473 | Unassigned | Unassigned | Unassigned | 517_1 | Clustered | 108 | VC_517_1 | 108 | 0.8483 | 1 | 0.8483 | 0 | 0 | 0 | 0.9955 | Ref |
| 4133 | KY006474 | Unassigned | Unassigned | Unassigned | 371_1 | Clustered | 11 | VC_371_1 | 11 | 0.0379 | 1 | 0.0379 | 0 | 0 | 0 | 0.8545 | Ref |
| 4134 | KY006853 | Unassigned | Unassigned | Unassigned | 170_1 | Clustered/Singleton | 1 | VC_170_1 | 1 | 0 | 1 | 0 | 0 | 0 | 0 | 1 | Ref |
| 4135 | KY014601 | Unassigned | Unassigned | Unassigned | 592_0 | Clustered | 50 | VC_592_0 | 50 | 0.7314 | 1 | 0.7314 | 0 | 0 | 0 | 1 | Ref |
| 4136 | KY022433 | Unassigned | Unassigned | Unassigned | 904_2 | Clustered | 7 | VC_904_2 | 7 | 0.2857 | 1 | 0.2857 | 0 | 0 | 0 | 1 | Ref |
| 4137 | KY030782 | Unassigned | Unassigned | Unassigned | 89_2 | Clustered | 5 | VC_89_2 | 5 | 0.2033 | 0.99999643 | 0.2033 | 0 | 0 | 0 | 1 | Ref |
| 4138 | KY045851 | Unassigned | Unassigned | Unassigned | 5_0 | Clustered | 30 | VC_5_0 | 30 | 0.9647 | 1 | 0.9647 | 0 | 0 | 0 | 1 | Ref |
| 4139 | KY056619 | Unassigned | Unassigned | Unassigned | 879_0 | Clustered | 36 | VC_879_0 | 36 | 0.994 | 1 | 0.994 | 0 | 0 | 0 | 1 | Ref |
| 4140 | KY056620 | Unassigned | Unassigned | Unassigned | 8_0 | Clustered | 58 | VC_8_0 | 58 | 0.8306 | 1 | 0.8306 | 0 | 0 | 0 | 0.9946 | Ref |
| 4141 | KY065147 | Unassigned | Unassigned | Unassigned | 495_1 | Clustered | 41 | VC_495_1 | 41 | 0.328 | 1 | 0.328 | 0 | 0 | 0 | 0.911 | Ref |
| 4142 | KY065148 | Unassigned | Unassigned | Unassigned | 784_0 | Clustered | 6 | VC_784_0 | 6 | 0.5517 | 1 | 0.5517 | 0 | 0 | 0 | 1 | Ref |
| 4143 | KY065149 | Unassigned | Unassigned | Unassigned | 989_0 | Clustered | 2 | VC_989_0 | 2 | 0.1205 | 0.95251219 | 0.1148 | 0 | 0 | 0 | 1 | Ref |
| 4144 | KY065449 | Unassigned | Unassigned | Unassigned | 415_0 | Clustered/Singleton | 1 | VC_415_0 | 1 | 0 | 1 | 0 | 0 | 0 | 0 | 1 | Ref |
| 4145 | KY065450 | Unassigned | Unassigned | Unassigned | 414_0 | Clustered | 37 | VC_414_0 | 37 | 0.585 | 1 | 0.585 | 0 | 0 | 0 | 0.9474 | Ref |
| 4146 | KY065451 | Unassigned | Unassigned | Unassigned | 414_0 | Clustered | 37 | VC_414_0 | 37 | 0.585 | 1 | 0.585 | 0 | 0 | 0 | 0.9474 | Ref |
| 4147 | KY065452 | Unassigned | Unassigned | Unassigned | 414_0 | Clustered | 37 | VC_414_0 | 37 | 0.585 | 1 | 0.585 | 0 | 0 | 0 | 0.9474 | Ref |
| 4148 | KY065453 | Unassigned | Unassigned | Unassigned | 333_0 | Clustered | 25 | VC_333_0 | 25 | 0.4681 | 1 | 0.4681 | 0 | 0 | 0 | 1 | Ref |
| 4149 | KY065454 | Unassigned | Unassigned | Unassigned | 333_0 | Clustered | 25 | VC_333_0 | 25 | 0.4681 | 1 | 0.4681 | 0 | 0 | 0 | 1 | Ref |
| 4150 | KY065455 | Unassigned | Unassigned | Unassigned | 412_0 | Clustered | 39 | VC_412_0 | 39 | 0.845 | 1 | 0.845 | 0 | 0 | 0 | 1 | Ref |
| 4151 | KY065456 | Unassigned | Unassigned | Unassigned | 414_0 | Clustered | 37 | VC_414_0 | 37 | 0.585 | 1 | 0.585 | 0 | 0 | 0 | 0.9474 | Ref |
| 4152 | KY065458 | Unassigned | Unassigned | Unassigned | 333_0 | Clustered | 25 | VC_333_0 | 25 | 0.4681 | 1 | 0.4681 | 0 | 0 | 0 | 1 | Ref |
| 4153 | KY065459 | Unassigned | Unassigned | Unassigned | 333_0 | Clustered | 25 | VC_333_0 | 25 | 0.4681 | 1 | 0.4681 | 0 | 0 | 0 | 1 | Ref |
| 4154 | KY065460 | Unassigned | Unassigned | Unassigned | 333_0 | Clustered | 25 | VC_333_0 | 25 | 0.4681 | 1 | 0.4681 | 0 | 0 | 0 | 1 | Ref |
| 4155 | KY065461 | Unassigned | Unassigned | Unassigned | 333_0 | Clustered | 25 | VC_333_0 | 25 | 0.4681 | 1 | 0.4681 | 0 | 0 | 0 | 1 | Ref |
| 4156 | KY065462 | Unassigned | Unassigned | Unassigned | 333_0 | Clustered | 25 | VC_333_0 | 25 | 0.4681 | 1 | 0.4681 | 0 | 0 | 0 | 1 | Ref |
| 4157 | KY065463 | Unassigned | Unassigned | Unassigned | 333_0 | Clustered | 25 | VC_333_0 | 25 | 0.4681 | 1 | 0.4681 | 0 | 0 | 0 | 1 | Ref |
| 4158 | KY065465 | Unassigned | Unassigned | Unassigned | 413_0 | Clustered | 3 | VC_413_0 | 3 | 0.0415 | 0.99841754 | 0.0415 | 0 | 0 | 0 | 1 | Ref |
| 4159 | KY065466 | Unassigned | Unassigned | Unassigned | 318_0 | Clustered | 3 | VC_318_0 | 3 | 0.0414 | 0.98579852 | 0.0408 | 0 | 0 | 0 | 1 | Ref |
| 4160 | KY065468 | Unassigned | Unassigned | Unassigned | 318_0 | Clustered | 3 | VC_318_0 | 3 | 0.0414 | 0.98579852 | 0.0408 | 0 | 0 | 0 | 1 | Ref |
| 4161 | KY065469 | Unassigned | Unassigned | Unassigned | 333_0 | Clustered | 25 | VC_333_0 | 25 | 0.4681 | 1 | 0.4681 | 0 | 0 | 0 | 1 | Ref |
| 4162 | KY065470 | Unassigned | Unassigned | Unassigned | 333_0 | Clustered | 25 | VC_333_0 | 25 | 0.4681 | 1 | 0.4681 | 0 | 0 | 0 | 1 | Ref |
| 4163 | KY065471 | Unassigned | Unassigned | Unassigned | 333_0 | Clustered | 25 | VC_333_0 | 25 | 0.4681 | 1 | 0.4681 | 0 | 0 | 0 | 1 | Ref |
| 4164 | KY065472 | Unassigned | Unassigned | Unassigned | 414_0 | Clustered | 37 | VC_414_0 | 37 | 0.585 | 1 | 0.585 | 0 | 0 | 0 | 0.9474 | Ref |
| 4165 | KY065473 | Unassigned | Unassigned | Unassigned | 414_0 | Clustered | 37 | VC_414_0 | 37 | 0.585 | 1 | 0.585 | 0 | 0 | 0 | 0.9474 | Ref |
| 4166 | KY065474 | Unassigned | Unassigned | Unassigned | 414_0 | Clustered | 37 | VC_414_0 | 37 | 0.585 | 1 | 0.585 | 0 | 0 | 0 | 0.9474 | Ref |
| 4167 | KY065475 | Unassigned | Unassigned | Unassigned | 414_0 | Clustered | 37 | VC_414_0 | 37 | 0.585 | 1 | 0.585 | 0 | 0 | 0 | 0.9474 | Ref |
| 4168 | KY065476 | Unassigned | Unassigned | Unassigned | 414_0 | Clustered | 37 | VC_414_0 | 37 | 0.585 | 1 | 0.585 | 0 | 0 | 0 | 0.9474 | Ref |
| 4169 | KY065477 | Unassigned | Unassigned | Unassigned | 414_0 | Clustered | 37 | VC_414_0 | 37 | 0.585 | 1 | 0.585 | 0 | 0 | 0 | 0.9474 | Ref |
| 4170 | KY065478 | Unassigned | Unassigned | Unassigned | 414_0 | Clustered | 37 | VC_414_0 | 37 | 0.585 | 1 | 0.585 | 0 | 0 | 0 | 0.9474 | Ref |
| 4171 | KY065479 | Unassigned | Unassigned | Unassigned | 412_0 | Clustered | 39 | VC_412_0 | 39 | 0.845 | 1 | 0.845 | 0 | 0 | 0 | 1 | Ref |
| 4172 | KY065480 | Unassigned | Unassigned | Unassigned | 414_0 | Clustered | 37 | VC_414_0 | 37 | 0.585 | 1 | 0.585 | 0 | 0 | 0 | 0.9474 | Ref |
| 4173 | KY065485 | Unassigned | Unassigned | Unassigned | 317_2 | Clustered | 20 | VC_317_2 | 20 | 0.1976 | 1 | 0.1976 | 0 | 0 | 0 | 0.6842 | Ref |
| 4174 | KY065486 | Unassigned | Unassigned | Unassigned | 414_0 | Clustered | 37 | VC_414_0 | 37 | 0.585 | 1 | 0.585 | 0 | 0 | 0 | 0.9474 | Ref |
| 4175 | KY065487 | Unassigned | Unassigned | Unassigned | 412_0 | Clustered | 39 | VC_412_0 | 39 | 0.845 | 1 | 0.845 | 0 | 0 | 0 | 1 | Ref |
| 4176 | KY065488 | Unassigned | Unassigned | Unassigned | 414_0 | Clustered | 37 | VC_414_0 | 37 | 0.585 | 1 | 0.585 | 0 | 0 | 0 | 0.9474 | Ref |
| 4177 | KY065490 | Unassigned | Unassigned | Unassigned | 412_0 | Clustered | 39 | VC_412_0 | 39 | 0.845 | 1 | 0.845 | 0 | 0 | 0 | 1 | Ref |
| 4178 | KY065491 | Unassigned | Unassigned | Unassigned | 412_0 | Clustered | 39 | VC_412_0 | 39 | 0.845 | 1 | 0.845 | 0 | 0 | 0 | 1 | Ref |
| 4179 | KY065492 | Unassigned | Unassigned | Unassigned | 412_0 | Clustered | 39 | VC_412_0 | 39 | 0.845 | 1 | 0.845 | 0 | 0 | 0 | 1 | Ref |
| 4180 | KY065493 | Unassigned | Unassigned | Unassigned | 412_0 | Clustered | 39 | VC_412_0 | 39 | 0.845 | 1 | 0.845 | 0 | 0 | 0 | 1 | Ref |
| 4181 | KY065494 | Unassigned | Unassigned | Unassigned | 333_0 | Clustered | 25 | VC_333_0 | 25 | 0.4681 | 1 | 0.4681 | 0 | 0 | 0 | 1 | Ref |
| 4182 | KY065495 | Unassigned | Unassigned | Unassigned | 333_0 | Clustered | 25 | VC_333_0 | 25 | 0.4681 | 1 | 0.4681 | 0 | 0 | 0 | 1 | Ref |
| 4183 | KY065496 | Unassigned | Unassigned | Unassigned | 414_0 | Clustered | 37 | VC_414_0 | 37 | 0.585 | 1 | 0.585 | 0 | 0 | 0 | 0.9474 | Ref |
| 4184 | KY065497 | Unassigned | Unassigned | Unassigned | 414_0 | Clustered | 37 | VC_414_0 | 37 | 0.585 | 1 | 0.585 | 0 | 0 | 0 | 0.9474 | Ref |
| 4185 | KY065498 | Unassigned | Unassigned | Unassigned | 323_1 | Clustered | 9 | VC_323_1 | 9 | 0.1798 | 1 | 0.1798 | 0 | 0 | 0 | 0.75 | Ref |
| 4186 | KY065499 | Unassigned | Unassigned | Unassigned | 333_0 | Clustered | 25 | VC_333_0 | 25 | 0.4681 | 1 | 0.4681 | 0 | 0 | 0 | 1 | Ref |
| 4187 | KY065500 | Unassigned | Unassigned | Unassigned | 414_0 | Clustered | 37 | VC_414_0 | 37 | 0.585 | 1 | 0.585 | 0 | 0 | 0 | 0.9474 | Ref |
| 4188 | KY065501 | Unassigned | Unassigned | Unassigned | 412_0 | Clustered | 39 | VC_412_0 | 39 | 0.845 | 1 | 0.845 | 0 | 0 | 0 | 1 | Ref |
| 4189 | KY065502 | Unassigned | Unassigned | Unassigned | 412_0 | Clustered | 39 | VC_412_0 | 39 | 0.845 | 1 | 0.845 | 0 | 0 | 0 | 1 | Ref |
| 4190 | KY065503 | Unassigned | Unassigned | Unassigned | 317_2 | Clustered | 20 | VC_317_2 | 20 | 0.1976 | 1 | 0.1976 | 0 | 0 | 0 | 0.6842 | Ref |
| 4191 | KY065504 | Unassigned | Unassigned | Unassigned | 317_2 | Clustered | 20 | VC_317_2 | 20 | 0.1976 | 1 | 0.1976 | 0 | 0 | 0 | 0.6842 | Ref |
| 4192 | KY065505 | Unassigned | Unassigned | Unassigned | 414_0 | Clustered | 37 | VC_414_0 | 37 | 0.585 | 1 | 0.585 | 0 | 0 | 0 | 0.9474 | Ref |
| 4193 | KY073123 | Unassigned | Unassigned | Unassigned | 132_1 | Clustered | 17 | VC_132_1 | 17 | 0.1738 | 1 | 0.1738 | 0 | 0 | 0 | 0.9853 | Ref |
| 4194 | KY073228 | Unassigned | Unassigned | Unassigned | 76_0 | Clustered | 37 | VC_76_0 | 37 | 0.9403 | 1 | 0.9403 | 0 | 0 | 0 | 1 | Ref |
| 4195 | KY082667 | Unassigned | Unassigned | Unassigned | 699_0 | Clustered | 71 | VC_699_0 | 71 | 0.9352 | 1 | 0.9352 | 0 | 0 | 0 | 1 | Ref |
| 4196 | KY083058 | Unassigned | Unassigned | Unassigned | 517_1 | Clustered | 108 | VC_517_1 | 108 | 0.8483 | 1 | 0.8483 | 0 | 0 | 0 | 0.9955 | Ref |
| 4197 | KY083059 | Unassigned | Unassigned | Unassigned | 517_0 | Clustered | 18 | VC_517_0 | 18 | 0.1599 | 1 | 0.1599 | 0 | 0 | 0 | 1 | Ref |
| 4198 | KY083060 | Unassigned | Unassigned | Unassigned | 517_1 | Clustered | 108 | VC_517_1 | 108 | 0.8483 | 1 | 0.8483 | 0 | 0 | 0 | 0.9955 | Ref |
| 4199 | KY083061 | Unassigned | Unassigned | Unassigned | 517_1 | Clustered | 108 | VC_517_1 | 108 | 0.8483 | 1 | 0.8483 | 0 | 0 | 0 | 0.9955 | Ref |
| 4200 | KY083062 | Unassigned | Unassigned | Unassigned | 517_1 | Clustered | 108 | VC_517_1 | 108 | 0.8483 | 1 | 0.8483 | 0 | 0 | 0 | 0.9955 | Ref |
| 4201 | KY083063 | Unassigned | Unassigned | Unassigned | 517_1 | Clustered | 108 | VC_517_1 | 108 | 0.8483 | 1 | 0.8483 | 0 | 0 | 0 | 0.9955 | Ref |
| 4202 | KY083726 | Unassigned | Unassigned | Unassigned | 841_0 | Clustered | 10 | VC_841_0 | 10 | 0.5997 | 1 | 0.5997 | 0 | 0 | 0 | 1 | Ref |
| 4203 | KY084243 | Unassigned | Unassigned | Unassigned | 386_0 | Clustered | 49 | VC_386_0 | 49 | 0.7941 | 1 | 0.7941 | 0 | 0 | 0 | 1 | Ref |
| 4204 | KY087898 | Unassigned | Unassigned | Unassigned | 792_0 | Clustered | 35 | VC_792_0 | 35 | 0.9828 | 1 | 0.9828 | 0 | 0 | 0 | 1 | Ref |
| 4205 | KY087992 | Unassigned | Unassigned | Unassigned | 234_0 | Clustered | 21 | VC_234_0 | 21 | 0.8355 | 1 | 0.8355 | 0 | 0 | 0 | 1 | Ref |
| 4206 | KY087993 | Unassigned | Unassigned | Unassigned | 237_1 | Clustered | 13 | VC_237_1 | 13 | 0.1773 | 1 | 0.1773 | 0 | 0 | 0 | 0.7308 | Ref |
| 4207 | KY092479 | Unassigned | Unassigned | Unassigned | 727_0 | Clustered | 11 | VC_727_0 | 11 | 0.5054 | 1 | 0.5054 | 0 | 0 | 0 | 1 | Ref |
| 4208 | KY092480 | Unassigned | Unassigned | Unassigned | 727_0 | Clustered | 11 | VC_727_0 | 11 | 0.5054 | 1 | 0.5054 | 0 | 0 | 0 | 1 | Ref |
| 4209 | KY092481 | Unassigned | Unassigned | Unassigned | 727_0 | Clustered | 11 | VC_727_0 | 11 | 0.5054 | 1 | 0.5054 | 0 | 0 | 0 | 1 | Ref |
| 4210 | KY092482 | Unassigned | Unassigned | Unassigned | 727_0 | Clustered | 11 | VC_727_0 | 11 | 0.5054 | 1 | 0.5054 | 0 | 0 | 0 | 1 | Ref |

| | | | | | | | | | | | | | | | | | |
|---|---|---|---|---|---|---|---|---|---|---|---|---|---|---|---|---|---|
| 4211 | KY092483 | Unassigned | Unassigned | Unassigned | 727_0 | Clustered | 11 | VC_727_0 | 11 | 0.5054 | 1 | 0.5054 | 0 | 0 | 0 | 1 | Ref |
| 4212 | KY092484 | Unassigned | Unassigned | Unassigned | 727_0 | Clustered | 11 | VC_727_0 | 11 | 0.5054 | 1 | 0.5054 | 0 | 0 | 0 | 1 | Ref |
| 4213 | KY114934 | Unassigned | Unassigned | Unassigned | 190_21 | Clustered/Singleton | 1 | VC_190_21 | 1 | 0 | 1 | 0 | 0 | 0 | 0 | 1 | Ref |
| 4214 | KY117485 | Unassigned | Unassigned | Unassigned | 154_0 | Clustered | 4 | VC_154_0 | 4 | 0.1766 | 0.99995106 | 0.1766 | 0 | 0 | 0 | 1 | Ref |
| 4215 | KY130461 | Unassigned | Unassigned | Unassigned | 507_0 | Clustered | 8 | VC_507_0 | 8 | 0.4466 | 1 | 0.4466 | 0 | 0 | 0 | 1 | Ref |
| 4216 | KY176369 | Unassigned | Unassigned | Unassigned | 637_1 | Clustered | 27 | VC_637_1 | 27 | 0.2578 | 1 | 0.2578 | 0 | 0 | 0 | 0.9858 | Ref |
| 4217 | KY203335 | Unassigned | Unassigned | Unassigned | 409_0 | Clustered | 3 | VC_409_0 | 3 | 0.1496 | 0.98294101 | 0.147 | 0 | 0 | 0 | 1 | Ref |
| 4218 | KY204244 | Unassigned | Unassigned | Unassigned | 517_1 | Clustered | 108 | VC_517_1 | 108 | 0.8483 | 1 | 0.8483 | 0 | 0 | 0 | 0.9955 | Ref |
| 4219 | KY204245 | Unassigned | Unassigned | Unassigned | 517_1 | Clustered | 108 | VC_517_1 | 108 | 0.8483 | 1 | 0.8483 | 0 | 0 | 0 | 0.9955 | Ref |
| 4220 | KY204246 | Unassigned | Unassigned | Unassigned | 517_1 | Clustered | 108 | VC_517_1 | 108 | 0.8483 | 1 | 0.8483 | 0 | 0 | 0 | 0.9955 | Ref |
| 4221 | KY204247 | Unassigned | Unassigned | Unassigned | 517_1 | Clustered | 108 | VC_517_1 | 108 | 0.8483 | 1 | 0.8483 | 0 | 0 | 0 | 0.9955 | Ref |
| 4222 | KY204248 | Unassigned | Unassigned | Unassigned | 517_1 | Clustered | 108 | VC_517_1 | 108 | 0.8483 | 1 | 0.8483 | 0 | 0 | 0 | 0.9955 | Ref |
| 4223 | KY204249 | Unassigned | Unassigned | Unassigned | 517_1 | Clustered | 108 | VC_517_1 | 108 | 0.8483 | 1 | 0.8483 | 0 | 0 | 0 | 0.9955 | Ref |
| 4224 | KY204250 | Unassigned | Unassigned | Unassigned | 517_1 | Clustered | 108 | VC_517_1 | 108 | 0.8483 | 1 | 0.8483 | 0 | 0 | 0 | 0.9955 | Ref |
| 4225 | KY204251 | Unassigned | Unassigned | Unassigned | 517_1 | Clustered | 108 | VC_517_1 | 108 | 0.8483 | 1 | 0.8483 | 0 | 0 | 0 | 0.9955 | Ref |
| 4226 | KY206887 | Unassigned | Unassigned | Unassigned | 1022_0 | Clustered | 4 | VC_1022_0 | 4 | 0.8047 | 0.99996089 | 0.8046 | 0 | 0 | 0 | 1 | Ref |
| 4227 | KY210139 | Unassigned | Unassigned | Unassigned | 364_0 | Clustered | 16 | VC_364_0 | 16 | 0.6491 | 1 | 0.6491 | 0 | 0 | 0 | 1 | Ref |
| 4228 | KY213952 | Unassigned | Unassigned | Unassigned | 325_48 | Clustered | 2 | VC_325_48 | 2 | 0.0039 | 0.95008941 | 0.0037 | 0 | 0 | 0 | 1 | Ref |
| 4229 | KY223999 | Unassigned | Unassigned | Unassigned | 763_2 | Clustered | 7 | VC_763_2 | 7 | 0.2612 | 1 | 0.2612 | 0 | 0 | 0 | 1 | Ref |
| 4230 | KY224000 | Unassigned | Unassigned | Unassigned | 372_1 | Clustered | 9 | VC_372_1 | 9 | 0.194 | 1 | 0.194 | 0 | 0 | 0 | 1 | Ref |
| 4231 | KY224001 | Unassigned | Unassigned | Unassigned | 325_54 | Clustered/Singleton | 1 | VC_325_54 | 1 | 0 | 1 | 0 | 0 | 0 | 0 | 1 | Ref |
| 4232 | KY249644 | Unassigned | Unassigned | Unassigned | 598_0 | Clustered | 4 | VC_598_0 | 4 | 0.0597 | 0.99355358 | 0.0593 | 0 | 0 | 0 | 1 | Ref |
| 4233 | KY250482 | Unassigned | Unassigned | Unassigned | 409_0 | Clustered | 3 | VC_409_0 | 3 | 0.1496 | 0.98294101 | 0.147 | 0 | 0 | 0 | 1 | Ref |
| 4234 | KY264020 | Unassigned | Unassigned | Unassigned | 961_0 | Clustered | 3 | VC_961_0 | 3 | 0.675 | 0.99054823 | 0.6686 | 0 | 0 | 0 | 1 | Ref |
| 4235 | KY268295 | Unassigned | Unassigned | Unassigned | 699_0 | Clustered | 71 | VC_699_0 | 71 | 0.9352 | 1 | 0.9352 | 0 | 0 | 0 | 1 | Ref |
| 4236 | KY268296 | Unassigned | Unassigned | Unassigned | 699_0 | Clustered | 71 | VC_699_0 | 71 | 0.9352 | 1 | 0.9352 | 0 | 0 | 0 | 1 | Ref |
| 4237 | KY271395 | Unassigned | Unassigned | Unassigned | 31_0 | Clustered | 9 | VC_31_0 | 9 | 0.3435 | 1 | 0.3435 | 0 | 0 | 0 | 0.6944 | Ref |
| 4238 | KY271396 | Unassigned | Unassigned | Unassigned | 31_0 | Clustered | 9 | VC_31_0 | 9 | 0.3435 | 1 | 0.3435 | 0 | 0 | 0 | 0.6944 | Ref |
| 4239 | KY271397 | Unassigned | Unassigned | Unassigned | 229_0 | Clustered | 32 | VC_229_0 | 32 | 0.9119 | 1 | 0.9119 | 0 | 0 | 0 | 1 | Ref |
| 4240 | KY271398 | Unassigned | Unassigned | Unassigned | 6_0 | Clustered | 44 | VC_6_0 | 44 | 0.908 | 1 | 0.908 | 0 | 0 | 0 | 1 | Ref |
| 4241 | KY271399 | Unassigned | Unassigned | Unassigned | 35_0 | Clustered | 6 | VC_35_0 | 6 | 0.2254 | 1 | 0.2254 | 0 | 0 | 0 | 0.9333 | Ref |
| 4242 | KY271401 | Unassigned | Unassigned | Unassigned | 244_0 | Clustered | 12 | VC_244_0 | 12 | 0.4039 | 1 | 0.4039 | 0 | 0 | 0 | 1 | Ref |
| 4243 | KY271402 | Unassigned | Unassigned | Unassigned | 34_0 | Clustered | 4 | VC_34_0 | 4 | 0.4349 | 0.99998062 | 0.4349 | 0 | 0 | 0 | 1 | Ref |
| 4244 | KY290756 | Unassigned | Unassigned | Unassigned | 1027_0 | Clustered | 20 | VC_1027_0 | 20 | 0.7114 | 1 | 0.7114 | 0 | 0 | 0 | 1 | Ref |
| 4245 | KY290947 | Unassigned | Unassigned | Unassigned | 1189_0 | Clustered | 9 | VC_1189_0 | 9 | 0.968 | 1 | 0.968 | 0 | 0 | 0 | 1 | Ref |
| 4246 | KY290948 | Unassigned | Unassigned | Unassigned | 600_1 | Clustered | 7 | VC_600_1 | 7 | 0.1837 | 0.99999997 | 0.1837 | 0 | 0 | 0 | 1 | Ref |
| 4247 | KY290949 | Unassigned | Unassigned | Unassigned | 1189_0 | Clustered | 9 | VC_1189_0 | 9 | 0.968 | 1 | 0.968 | 0 | 0 | 0 | 1 | Ref |
| 4248 | KY290950 | Unassigned | Unassigned | Unassigned | 1189_0 | Clustered | 9 | VC_1189_0 | 9 | 0.968 | 1 | 0.968 | 0 | 0 | 0 | 1 | Ref |
| 4249 | KY290951 | Unassigned | Unassigned | Unassigned | 600_1 | Clustered | 7 | VC_600_1 | 7 | 0.1837 | 0.99999997 | 0.1837 | 0 | 0 | 0 | 1 | Ref |
| 4250 | KY290952 | Unassigned | Unassigned | Unassigned | 1189_0 | Clustered | 9 | VC_1189_0 | 9 | 0.968 | 1 | 0.968 | 0 | 0 | 0 | 1 | Ref |
| 4251 | KY290953 | Unassigned | Unassigned | Unassigned | 369_0 | Clustered | 6 | VC_369_0 | 6 | 0.5924 | 1 | 0.5924 | 0 | 0 | 0 | 1 | Ref |
| 4252 | KY290954 | Unassigned | Unassigned | Unassigned | 369_0 | Clustered | 6 | VC_369_0 | 6 | 0.5924 | 1 | 0.5924 | 0 | 0 | 0 | 1 | Ref |
| 4253 | KY290955 | Unassigned | Unassigned | Unassigned | 443_1 | Clustered | 2 | VC_443_1 | 2 | 0.0287 | 0.89347378 | 0.0257 | 0 | 0 | 0 | 1 | Ref |
| 4254 | KY290956 | Unassigned | Unassigned | Unassigned | 600_1 | Clustered | 7 | VC_600_1 | 7 | 0.1837 | 0.99999997 | 0.1837 | 0 | 0 | 0 | 1 | Ref |
| 4255 | KY290957 | Unassigned | Unassigned | Unassigned | 600_1 | Clustered | 7 | VC_600_1 | 7 | 0.1837 | 0.99999997 | 0.1837 | 0 | 0 | 0 | 1 | Ref |
| 4256 | KY290958 | Unassigned | Unassigned | Unassigned | 600_1 | Clustered | 7 | VC_600_1 | 7 | 0.1837 | 0.99999997 | 0.1837 | 0 | 0 | 0 | 1 | Ref |
| 4257 | KY290975 | Unassigned | Unassigned | Unassigned | 283_111 | Clustered/Singleton | 1 | VC_283_111 | 1 | 0 | 1 | 0 | 0 | 0 | 0 | 1 | Ref |
| 4258 | KY295891 | Unassigned | Unassigned | Unassigned | 452_0 | Clustered | 66 | VC_452_0 | 66 | 0.9248 | 1 | 0.9248 | 0 | 0 | 0 | 1 | Ref |
| 4259 | KY295892 | Unassigned | Unassigned | Unassigned | 452_0 | Clustered | 66 | VC_452_0 | 66 | 0.9248 | 1 | 0.9248 | 0 | 0 | 0 | 1 | Ref |
| 4260 | KY295893 | Unassigned | Unassigned | Unassigned | 452_0 | Clustered | 66 | VC_452_0 | 66 | 0.9248 | 1 | 0.9248 | 0 | 0 | 0 | 1 | Ref |
| 4261 | KY295894 | Unassigned | Unassigned | Unassigned | 451_0 | Clustered | 80 | VC_451_0 | 80 | 0.9991 | 1 | 0.9991 | 0 | 0 | 0 | 1 | Ref |
| 4262 | KY295895 | Unassigned | Unassigned | Unassigned | 638_0 | Clustered | 45 | VC_638_0 | 45 | 0.7741 | 1 | 0.7741 | 0 | 0 | 0 | 1 | Ref |
| 4263 | KY295896 | Unassigned | Unassigned | Unassigned | 638_0 | Clustered | 45 | VC_638_0 | 45 | 0.7741 | 1 | 0.7741 | 0 | 0 | 0 | 1 | Ref |
| 4264 | KY295897 | Unassigned | Unassigned | Unassigned | 452_0 | Clustered | 66 | VC_452_0 | 66 | 0.9248 | 1 | 0.9248 | 0 | 0 | 0 | 1 | Ref |
| 4265 | KY295898 | Unassigned | Unassigned | Unassigned | 638_0 | Clustered | 45 | VC_638_0 | 45 | 0.7741 | 1 | 0.7741 | 0 | 0 | 0 | 1 | Ref |
| 4266 | KY295899 | Unassigned | Unassigned | Unassigned | 452_0 | Clustered | 66 | VC_452_0 | 66 | 0.9248 | 1 | 0.9248 | 0 | 0 | 0 | 1 | Ref |
| 4267 | KY296500 | Unassigned | Unassigned | Unassigned | 814_0 | Clustered | 3 | VC_814_0 | 3 | 1 | 1 | 1 | 0 | 0 | 0 | 1 | Ref |
| 4268 | KY296501 | Unassigned | Unassigned | Unassigned | 814_0 | Clustered | 3 | VC_814_0 | 3 | 1 | 1 | 1 | 0 | 0 | 0 | 1 | Ref |
| 4269 | KY303907 | Unassigned | Unassigned | Unassigned | 108_0 | Clustered | 33 | VC_108_0 | 33 | 0.949 | 1 | 0.949 | 0 | 0 | 0 | 1 | Ref |
| 4270 | KY316062 | Unassigned | Unassigned | Unassigned | 70_0 | Clustered | 27 | VC_70_0 | 27 | 0.5981 | 1 | 0.5981 | 0 | 0 | 0 | 0.9886 | Ref |
| 4271 | KY318515 | Unassigned | Unassigned | Unassigned | 878_0 | Clustered | 32 | VC_878_0 | 32 | 0.9965 | 1 | 0.9965 | 0 | 0 | 0 | 1 | Ref |
| 4272 | KY319168 | Unassigned | Unassigned | Unassigned | 551_2 | Clustered | 17 | VC_551_2 | 17 | 0.0973 | 1 | 0.0973 | 0 | 0 | 0 | 0.9779 | Ref |
| 4273 | KY348865 | Unassigned | Unassigned | Unassigned | 520_44 | Clustered/Singleton | 1 | VC_520_44 | 1 | 0 | 1 | 0 | 0 | 0 | 0 | 1 | Ref |
| 4274 | KY349816 | Unassigned | Unassigned | Unassigned | 412_0 | Clustered | 39 | VC_412_0 | 39 | 0.845 | 1 | 0.845 | 0 | 0 | 0 | 1 | Ref |
| 4275 | KY363465 | Unassigned | Unassigned | Unassigned | 193_0 | Clustered | 2 | VC_193_0 | 2 | 0.0321 | 0.95762656 | 0.0308 | 0 | 0 | 0 | 1 | Ref |
| 4276 | KY365739 | Unassigned | Unassigned | Unassigned | 975_0 | Clustered | 11 | VC_975_0 | 11 | 0.9687 | 1 | 0.9687 | 0 | 0 | 0 | 1 | Ref |
| 4277 | KY368639 | Unassigned | Unassigned | Unassigned | 87_0 | Clustered | 8 | VC_87_0 | 8 | 0.5545 | 1 | 0.5545 | 0 | 0 | 0 | 1 | Ref |
| 4278 | KY368640 | Unassigned | Unassigned | Unassigned | 85_4 | Clustered | 3 | VC_85_4 | 3 | 0.0294 | 0.98125255 | 0.0288 | 0 | 0 | 0 | 1 | Ref |
| 4279 | KY379511 | Unassigned | Unassigned | Unassigned | 896_0 | Clustered | 5 | VC_896_0 | 5 | 0.4981 | 0.99999977 | 0.4981 | 0 | 0 | 0 | 1 | Ref |
| 4280 | KY379853 | Unassigned | Unassigned | Unassigned | 637_0 | Clustered | 115 | VC_637_0 | 115 | 0.731 | 1 | 0.731 | 0 | 0 | 0 | 0.8764 | Ref |
| 4281 | KY380102 | Unassigned | Unassigned | Unassigned | 236_0 | Clustered | 72 | VC_236_0 | 72 | 0.8104 | 1 | 0.8104 | 0 | 0 | 0 | 1 | Ref |
| 4282 | KY381600 | Unassigned | Unassigned | Unassigned | 634_1 | Clustered | 6 | VC_634_1 | 6 | 0.4228 | 1 | 0.4228 | 0 | 0 | 0 | 1 | Ref |
| 4283 | KY385380 | Unassigned | Unassigned | Unassigned | 371_13 | Clustered | 33 | VC_371_13 | 33 | 0.179 | 1 | 0.179 | 0 | 0 | 0 | 0.8769 | Ref |
| 4284 | KY385381 | Unassigned | Unassigned | Unassigned | 764_0 | Clustered | 4 | VC_764_0 | 4 | 0.3556 | 0.99997469 | 0.3556 | 0 | 0 | 0 | 1 | Ref |
| 4285 | KY385382 | Unassigned | Unassigned | Unassigned | 371_13 | Clustered | 33 | VC_371_13 | 33 | 0.179 | 1 | 0.179 | 0 | 0 | 0 | 0.8769 | Ref |
| 4286 | KY385383 | Unassigned | Unassigned | Unassigned | 371_13 | Clustered | 33 | VC_371_13 | 33 | 0.179 | 1 | 0.179 | 0 | 0 | 0 | 0.8769 | Ref |
| 4287 | KY385384 | Unassigned | Unassigned | Unassigned | 520_53 | Clustered/Singleton | 1 | VC_520_53 | 1 | 0 | 1 | 0 | 0 | 0 | 0 | 1 | Ref |
| 4288 | KY385423 | Unassigned | Unassigned | Unassigned | 111_0 | Clustered | 91 | VC_111_0 | 91 | 0.9587 | 1 | 0.9587 | 0 | 0 | 0 | 1 | Ref |
| 4289 | KY389063 | Unassigned | Unassigned | Unassigned | 7_0 | Clustered | 28 | VC_7_0 | 28 | 0.3916 | 1 | 0.3916 | 0 | 0 | 0 | 0.9947 | Ref |
| 4290 | KY389064 | Unassigned | Unassigned | Unassigned | 7_0 | Clustered | 28 | VC_7_0 | 28 | 0.3916 | 1 | 0.3916 | 0 | 0 | 0 | 0.9947 | Ref |
| 4291 | KY389315 | Unassigned | Unassigned | Unassigned | 823_0 | Clustered | 124 | VC_823_0 | 124 | 0.9492 | 1 | 0.9492 | 0 | 0 | 0 | 1 | Ref |
| 4292 | KY389316 | Unassigned | Unassigned | Unassigned | 823_0 | Clustered | 124 | VC_823_0 | 124 | 0.9492 | 1 | 0.9492 | 0 | 0 | 0 | 1 | Ref |
| 4293 | KY398841 | Unassigned | Unassigned | Unassigned | 456_0 | Clustered | 62 | VC_456_0 | 62 | 0.9026 | 1 | 0.9026 | 0 | 0 | 0 | 1 | Ref |
| 4294 | KY434670 | Unassigned | Unassigned | Unassigned | 950_0 | Clustered | 38 | VC_950_0 | 38 | 0.9443 | 1 | 0.9443 | 0 | 0 | 0 | 1 | Ref |
| 4295 | KY435490 | Unassigned | Unassigned | Unassigned | 452_0 | Clustered | 66 | VC_452_0 | 66 | 0.9248 | 1 | 0.9248 | 0 | 0 | 0 | 1 | Ref |
| 4296 | KY448244 | Unassigned | Unassigned | Unassigned | 67_0 | Clustered | 2 | VC_67_0 | 2 | 0.014 | 0.69357611 | 0.0097 | 0 | 0 | 0 | 1 | Ref |
| 4297 | KY450753 | Unassigned | Unassigned | Unassigned | 823_0 | Clustered | 124 | VC_823_0 | 124 | 0.9492 | 1 | 0.9492 | 0 | 0 | 0 | 1 | Ref |
| 4298 | KY464836 | Unassigned | Unassigned | Unassigned | 70_0 | Clustered | 27 | VC_70_0 | 27 | 0.5981 | 1 | 0.5981 | 0 | 0 | 0 | 0.9886 | Ref |
| 4299 | KY464936 | Unassigned | Unassigned | Unassigned | 513_0 | Clustered | 32 | VC_513_0 | 32 | 0.6881 | 1 | 0.6881 | 0 | 0 | 0 | 1 | Ref |
| 4300 | KY471266 | Unassigned | Unassigned | Unassigned | 517_1 | Clustered | 108 | VC_517_1 | 108 | 0.8483 | 1 | 0.8483 | 0 | 0 | 0 | 0.9955 | Ref |
| 4301 | KY471267 | Unassigned | Unassigned | Unassigned | 520_27 | Clustered | 2 | VC_520_27 | 2 | 0.0143 | 0.95761635 | 0.0137 | 0 | 0 | 0 | 1 | Ref |
| 4302 | KY471268 | Unassigned | Unassigned | Unassigned | 406_0 | Clustered | 11 | VC_406_0 | 11 | 0.9449 | 1 | 0.9449 | 0 | 0 | 0 | 1 | Ref |
| 4303 | KY471269 | Unassigned | Unassigned | Unassigned | 406_0 | Clustered | 11 | VC_406_0 | 11 | 0.9449 | 1 | 0.9449 | 0 | 0 | 0 | 1 | Ref |
| 4304 | KY471460 | Unassigned | Unassigned | Unassigned | 221_0 | Clustered | 22 | VC_221_0 | 22 | 0.7447 | 1 | 0.7447 | 0 | 0 | 0 | 1 | Ref |
| 4305 | KY472224 | Unassigned | Unassigned | Unassigned | 108_0 | Clustered | 33 | VC_108_0 | 33 | 0.949 | 1 | 0.949 | 0 | 0 | 0 | 1 | Ref |
| 4306 | KY514263 | Unassigned | Unassigned | Unassigned | 849_2 | Clustered | 2 | VC_849_2 | 2 | 0.0246 | 0.86513761 | 0.0213 | 0 | 0 | 0 | 1 | Ref |
| 4307 | KY514264 | Unassigned | Unassigned | Unassigned | 841_0 | Clustered | 10 | VC_841_0 | 10 | 0.5997 | 1 | 0.5997 | 0 | 0 | 0 | 1 | Ref |
| 4308 | KY514265 | Unassigned | Unassigned | Unassigned | 841_0 | Clustered | 10 | VC_841_0 | 10 | 0.5997 | 1 | 0.5997 | 0 | 0 | 0 | 1 | Ref |
| 4309 | KY549152 | Unassigned | Unassigned | Unassigned | 551_7 | Clustered | 28 | VC_551_7 | 28 | 0.1661 | 1 | 0.1661 | 0 | 0 | 0 | 0.8519 | Ref |
| 4310 | KY549153 | Unassigned | Unassigned | Unassigned | 956_0 | Clustered | 39 | VC_956_0 | 39 | 1 | 1 | 1 | 0 | 0 | 0 | 1 | Ref |
| 4311 | KY549154 | Unassigned | Unassigned | Unassigned | 956_0 | Clustered | 39 | VC_956_0 | 39 | 1 | 1 | 1 | 0 | 0 | 0 | 1 | Ref |
| 4312 | KY549155 | Unassigned | Unassigned | Unassigned | 517_1 | Clustered | 108 | VC_517_1 | 108 | 0.8483 | 1 | 0.8483 | 0 | 0 | 0 | 0.9955 | Ref |
| 4313 | KY549443 | Unassigned | Unassigned | Unassigned | 92_0 | Clustered | 22 | VC_92_0 | 22 | 0.5639 | 1 | 0.5639 | 0 | 0 | 0 | 1 | Ref |
| 4314 | KY549659 | Unassigned | Unassigned | Unassigned | 841_0 | Clustered | 10 | VC_841_0 | 10 | 0.5997 | 1 | 0.5997 | 0 | 0 | 0 | 1 | Ref |
| 4315 | KY554760 | Unassigned | Unassigned | Unassigned | 202_0 | Clustered | 102 | VC_202_0 | 102 | 0.4049 | 1 | 0.4049 | 0 | 0 | 0 | 0.7876 | Ref |
| 4316 | KY554761 | Unassigned | Unassigned | Unassigned | 202_0 | Clustered | 102 | VC_202_0 | 102 | 0.4049 | 1 | 0.4049 | 0 | 0 | 0 | 0.7876 | Ref |
| 4317 | KY554762 | Unassigned | Unassigned | Unassigned | 534_0 | Clustered | 9 | VC_534_0 | 9 | 0.99 | 1 | 0.99 | 0 | 0 | 0 | 1 | Ref |
| 4318 | KY554763 | Unassigned | Unassigned | Unassigned | 534_0 | Clustered | 9 | VC_534_0 | 9 | 0.99 | 1 | 0.99 | 0 | 0 | 0 | 1 | Ref |
| 4319 | KY554764 | Unassigned | Unassigned | Unassigned | 534_0 | Clustered | 9 | VC_534_0 | 9 | 0.99 | 1 | 0.99 | 0 | 0 | 0 | 1 | Ref |
| 4320 | KY554765 | Unassigned | Unassigned | Unassigned | 534_0 | Clustered | 9 | VC_534_0 | 9 | 0.99 | 1 | 0.99 | 0 | 0 | 0 | 1 | Ref |
| 4321 | KY554766 | Unassigned | Unassigned | Unassigned | 534_0 | Clustered | 9 | VC_534_0 | 9 | 0.99 | 1 | 0.99 | 0 | 0 | 0 | 1 | Ref |
| 4322 | KY554767 | Unassigned | Unassigned | Unassigned | 534_0 | Clustered | 9 | VC_534_0 | 9 | 0.99 | 1 | 0.99 | 0 | 0 | 0 | 1 | Ref |
| 4323 | KY554768 | Unassigned | Unassigned | Unassigned | 474_0 | Clustered | 14 | VC_474_0 | 14 | 0.8466 | 1 | 0.8466 | 0 | 0 | 0 | 1 | Ref |
| 4324 | KY554769 | Unassigned | Unassigned | Unassigned | 474_0 | Clustered | 14 | VC_474_0 | 14 | 0.8466 | 1 | 0.8466 | 0 | 0 | 0 | 1 | Ref |
| 4325 | KY554770 | Unassigned | Unassigned | Unassigned | 474_0 | Clustered | 14 | VC_474_0 | 14 | 0.8466 | 1 | 0.8466 | 0 | 0 | 0 | 1 | Ref |
| 4326 | KY554771 | Unassigned | Unassigned | Unassigned | 474_0 | Clustered | 14 | VC_474_0 | 14 | 0.8466 | 1 | 0.8466 | 0 | 0 | 0 | 1 | Ref |
| 4327 | KY554772 | Unassigned | Unassigned | Unassigned | 474_0 | Clustered | 14 | VC_474_0 | 14 | 0.8466 | 1 | 0.8466 | 0 | 0 | 0 | 1 | Ref |
| 4328 | KY554773 | Unassigned | Unassigned | Unassigned | 474_0 | Clustered | 14 | VC_474_0 | 14 | 0.8466 | 1 | 0.8466 | 0 | 0 | 0 | 1 | Ref |
| 4329 | KY554774 | Unassigned | Unassigned | Unassigned | 474_0 | Clustered | 14 | VC_474_0 | 14 | 0.8466 | 1 | 0.8466 | 0 | 0 | 0 | 1 | Ref |
| 4330 | KY554775 | Unassigned | Unassigned | Unassigned | 474_0 | Clustered | 14 | VC_474_0 | 14 | 0.8466 | 1 | 0.8466 | 0 | 0 | 0 | 1 | Ref |
| 4331 | KY554776 | Unassigned | Unassigned | Unassigned | 474_0 | Clustered | 14 | VC_474_0 | 14 | 0.8466 | 1 | 0.8466 | 0 | 0 | 0 | 1 | Ref |
| 4332 | KY554777 | Unassigned | Unassigned | Unassigned | 474_0 | Clustered | 14 | VC_474_0 | 14 | 0.8466 | 1 | 0.8466 | 0 | 0 | 0 | 1 | Ref |
| 4333 | KY555142 | Unassigned | Unassigned | Unassigned | 976_4 | Clustered | 11 | VC_976_4 | 11 | 0.3364 | 1 | 0.3364 | 0 | 0 | 0 | 1 | Ref |
| 4334 | KY555143 | Unassigned | Unassigned | Unassigned | 976_4 | Clustered | 11 | VC_976_4 | 11 | 0.3364 | 1 | 0.3364 | 0 | 0 | 0 | 1 | Ref |
| 4335 | KY555144 | Unassigned | Unassigned | Unassigned | 976_4 | Clustered | 11 | VC_976_4 | 11 | 0.3364 | 1 | 0.3364 | 0 | 0 | 0 | 1 | Ref |
| 4336 | KY555145 | Unassigned | Unassigned | Unassigned | 976_4 | Clustered | 11 | VC_976_4 | 11 | 0.3364 | 1 | 0.3364 | 0 | 0 | 0 | 1 | Ref |
| 4337 | KY555146 | Unassigned | Unassigned | Unassigned | 976_4 | Clustered | 11 | VC_976_4 | 11 | 0.3364 | 1 | 0.3364 | 0 | 0 | 0 | 1 | Ref |
| 4338 | KY555147 | Unassigned | Unassigned | Unassigned | 976_4 | Clustered | 11 | VC_976_4 | 11 | 0.3364 | 1 | 0.3364 | 0 | 0 | 0 | 1 | Ref |
| 4339 | KY563228 | Unassigned | Unassigned | Unassigned | 599_0 | Clustered | 50 | VC_599_0 | 50 | 0.9989 | 1 | 0.9989 | 0 | 0 | 0 | 1 | Ref |
| 4340 | KY565347 | Unassigned | Unassigned | Unassigned | 101_0 | Clustered | 3 | VC_101_0 | 3 | 0.3621 | 0.99764884 | 0.3612 | 0 | 0 | 0 | 1 | Ref |
| 4341 | KY566218 | Unassigned | Unassigned | Unassigned | 408_0 | Clustered | 9 | VC_408_0 | 9 | 0.86 | 1 | 0.86 | 0 | 0 | 0 | 1 | Ref |
| 4342 | KY575286 | Unassigned | Unassigned | Unassigned | 938_0 | Clustered | 99 | VC_938_0 | 99 | 0.9119 | 1 | 0.9119 | 0 | 0 | 0 | 0.9928 | Ref |
| 4343 | KY579375 | Unassigned | Unassigned | Unassigned | 599_0 | Clustered | 50 | VC_599_0 | 50 | 0.9989 | 1 | 0.9989 | 0 | 0 | 0 | 1 | Ref |
| 4344 | KY581279 | Unassigned | Unassigned | Unassigned | 149_8 | Clustered | 66 | VC_149_8 | 66 | 0.4565 | 1 | 0.4565 | 0 | 0 | 0 | 0.8182 | Ref |
| 4345 | KY593455 | Unassigned | Unassigned | Unassigned | 490_3 | Clustered | 2 | VC_490_3 | 2 | 0.0135 | 0.95703149 | 0.0129 | 0 | 0 | 0 | 1 | Ref |
| 4346 | KY608966 | Unassigned | Unassigned | Unassigned | 428_0 | Clustered | 33 | VC_428_0 | 33 | 0.9915 | 1 | 0.9915 | 0 | 0 | 0 | 1 | Ref |
| 4347 | KY608967 | Unassigned | Unassigned | Unassigned | 283_109 | Clustered | 2 | VC_283_109 | 2 | 0.0027 | 0.91535723 | 0.0024 | 0 | 0 | 0 | 1 | Ref |
| 4348 | KY612839 | Unassigned | Unassigned | Unassigned | 784_0 | Clustered | 6 | VC_784_0 | 6 | 0.5517 | 1 | 0.5517 | 0 | 0 | 0 | 1 | Ref |
| 4349 | KY613597 | Unassigned | Unassigned | Unassigned | 408_0 | Clustered | 9 | VC_408_0 | 9 | 0.86 | 1 | 0.86 | 0 | 0 | 0 | 1 | Ref |
| 4350 | KY618819 | Unassigned | Unassigned | Unassigned | 423_0 | Clustered | 45 | VC_423_0 | 45 | 0.9484 | 1 | 0.9484 | 0 | 0 | 0 | 1 | Ref |
| 4351 | KY620117 | Unassigned | Unassigned | Unassigned | 592_0 | Clustered | 50 | VC_592_0 | 50 | 0.7314 | 1 | 0.7314 | 0 | 0 | 0 | 1 | Ref |
| 4352 | KY622015 | Unassigned | Unassigned | Unassigned | 498_0 | Clustered | 8 | VC_498_0 | 8 | 0.3987 | 1 | 0.3987 | 0 | 0 | 0 | 1 | Ref |
| 4353 | KY624610 | Unassigned | Unassigned | Unassigned | 408_0 | Clustered | 9 | VC_408_0 | 9 | 0.86 | 1 | 0.86 | 0 | 0 | 0 | 1 | Ref |
| 4354 | KY624611 | Unassigned | Unassigned | Unassigned | 408_0 | Clustered | 9 | VC_408_0 | 9 | 0.86 | 1 | 0.86 | 0 | 0 | 0 | 1 | Ref |
| 4355 | KY624612 | Unassigned | Unassigned | Unassigned | 408_0 | Clustered | 9 | VC_408_0 | 9 | 0.86 | 1 | 0.86 | 0 | 0 | 0 | 1 | Ref |
| 4356 | KY624613 | Unassigned | Unassigned | Unassigned | 408_0 | Clustered | 9 | VC_408_0 | 9 | 0.86 | 1 | 0.86 | 0 | 0 | 0 | 1 | Ref |
| 4357 | KY624614 | Unassigned | Unassigned | Unassigned | 408_0 | Clustered | 9 | VC_408_0 | 9 | 0.86 | 1 | 0.86 | 0 | 0 | 0 | 1 | Ref |
| 4358 | KY624615 | Unassigned | Unassigned | Unassigned | 408_0 | Clustered | 9 | VC_408_0 | 9 | 0.86 | 1 | 0.86 | 0 | 0 | 0 | 1 | Ref |
| 4359 | KY624616 | Unassigned | Unassigned | Unassigned | 408_0 | Clustered | 9 | VC_408_0 | 9 | 0.86 | 1 | 0.86 | 0 | 0 | 0 | 1 | Ref |
| 4360 | KY626162 | Unassigned | Unassigned | Unassigned | 329_1 | Clustered | 17 | VC_329_1 | 17 | 0.0867 | 1 | 0.0867 | 0 | 0 | 0 | 0.8235 | Ref |
| 4361 | KY626163 | Unassigned | Unassigned | Unassigned | 637_0 | Clustered | 115 | VC_637_0 | 115 | 0.731 | 1 | 0.731 | 0 | 0 | 0 | 0.8764 | Ref |
| 4362 | KY626176 | Unassigned | Unassigned | Unassigned | 1027_0 | Clustered | 20 | VC_1027_0 | 20 | 0.7114 | 1 | 0.7114 | 0 | 0 | 0 | 1 | Ref |
| 4363 | KY626177 | Unassigned | Unassigned | Unassigned | 1027_0 | Clustered | 20 | VC_1027_0 | 20 | 0.7114 | 1 | 0.7114 | 0 | 0 | 0 | 1 | Ref |
| 4364 | KY629563 | Unassigned | Unassigned | Unassigned | 978_0 | Clustered | 2 | VC_978_0 | 2 | 0.0595 | 0.95411371 | 0.0568 | 0 | 0 | 0 | 1 | Ref |
| 4365 | KY629621 | Unassigned | Unassigned | Unassigned | 891_0 | Clustered | 2 | VC_891_0 | 2 | 0.5746 | 0.92263554 | 0.5301 | 0 | 0 | 0 | 1 | Ref |
| 4366 | KY630163 | Unassigned | Unassigned | Unassigned | 133_34 | Clustered | 3 | VC_133_34 | 3 | 0.0102 | 0.97369044 | 0.01 | 0 | 0 | 0 | 0.6667 | Ref |
| 4367 | KY630187 | Unassigned | Unassigned | Unassigned | 164_7 | Clustered | 7 | VC_164_7 | 7 | 0.0956 | 0.99998 | 0.0956 | 0 | 0 | 0 | 1 | Ref |
| 4368 | KY652723 | Unassigned | Unassigned | Unassigned | 823_0 | Clustered | 124 | VC_823_0 | 124 | 0.9492 | 1 | 0.9492 | 0 | 0 | 0 | 1 | Ref |
| 4369 | KY652724 | Unassigned | Unassigned | Unassigned | 823_0 | Clustered | 124 | VC_823_0 | 124 | 0.9492 | 1 | 0.9492 | 0 | 0 | 0 | 1 | Ref |
| 4370 | KY652725 | Unassigned | Unassigned | Unassigned | 823_0 | Clustered | 124 | VC_823_0 | 124 | 0.9492 | 1 | 0.9492 | 0 | 0 | 0 | 1 | Ref |
| 4371 | KY652726 | Unassigned | Unassigned | Unassigned | 582_3 | Clustered | 12 | VC_582_3 | 12 | 0.2357 | 1 | 0.2357 | 0 | 0 | 0 | 0.8636 | Ref |
| 4372 | KY653119 | Unassigned | Unassigned | Unassigned | 245_0 | Clustered | 10 | VC_245_0 | 10 | 0.1805 | 1 | 0.1805 | 0 | 0 | 0 | 0.9778 | Ref |

| | | | | | | | | | | | | | | | | | |
|---|---|---|---|---|---|---|---|---|---|---|---|---|---|---|---|---|---|
| 4373 | KY653123 | Unassigned | Unassigned | Unassigned | 10_0 | Clustered | 25 | VC_10_0 | 25 | 0.3467 | 1 | 0.3467 | 0 | 0 | 0 | 0.9733 | Ref |
| 4374 | KY653125 | Unassigned | Unassigned | Unassigned | 7_0 | Clustered | 28 | VC_7_0 | 28 | 0.3916 | 1 | 0.3916 | 0 | 0 | 0 | 0.9947 | Ref |
| 4375 | KY653237 | Unassigned | Unassigned | Unassigned | 303_0 | Clustered | 155 | VC_303_0 | 155 | 0.9979 | 1 | 0.9979 | 0 | 0 | 0 | 1 | Ref |
| 4376 | KY654690 | Unassigned | Unassigned | Unassigned | 330_0 | Clustered | 27 | VC_330_0 | 27 | 0.5088 | 1 | 0.5088 | 0 | 0 | 0 | 0.8661 | Ref |
| 4377 | KY657202 | Unassigned | Unassigned | Unassigned | 584_0 | Clustered | 28 | VC_584_0 | 28 | 0.7704 | 1 | 0.7704 | 0 | 0 | 0 | 1 | Ref |
| 4378 | KY658673 | Unassigned | Unassigned | Unassigned | 299_0 | Clustered | 24 | VC_299_0 | 24 | 0.9905 | 1 | 0.9905 | 0 | 0 | 0 | 1 | Ref |
| 4379 | KY658674 | Unassigned | Unassigned | Unassigned | 299_0 | Clustered | 24 | VC_299_0 | 24 | 0.9905 | 1 | 0.9905 | 0 | 0 | 0 | 1 | Ref |
| 4380 | KY658675 | Unassigned | Unassigned | Unassigned | 299_0 | Clustered | 24 | VC_299_0 | 24 | 0.9905 | 1 | 0.9905 | 0 | 0 | 0 | 1 | Ref |
| 4381 | KY658676 | Unassigned | Unassigned | Unassigned | 299_0 | Clustered | 24 | VC_299_0 | 24 | 0.9905 | 1 | 0.9905 | 0 | 0 | 0 | 1 | Ref |
| 4382 | KY658677 | Unassigned | Unassigned | Unassigned | 299_0 | Clustered | 24 | VC_299_0 | 24 | 0.9905 | 1 | 0.9905 | 0 | 0 | 0 | 1 | Ref |
| 4383 | KY658678 | Unassigned | Unassigned | Unassigned | 299_0 | Clustered | 24 | VC_299_0 | 24 | 0.9905 | 1 | 0.9905 | 0 | 0 | 0 | 1 | Ref |
| 4384 | KY658679 | Unassigned | Unassigned | Unassigned | 299_0 | Clustered | 24 | VC_299_0 | 24 | 0.9905 | 1 | 0.9905 | 0 | 0 | 0 | 1 | Ref |
| 4385 | KY658680 | Unassigned | Unassigned | Unassigned | 299_0 | Clustered | 24 | VC_299_0 | 24 | 0.9905 | 1 | 0.9905 | 0 | 0 | 0 | 1 | Ref |
| 4386 | KY661963 | Unassigned | Unassigned | Unassigned | 730_0 | Clustered | 9 | VC_730_0 | 9 | 1 | 1 | 1 | 0 | 0 | 0 | 1 | Ref |
| 4387 | KY670595 | Unassigned | Unassigned | Unassigned | 698_0 | Clustered | 26 | VC_698_0 | 26 | 0.9366 | 1 | 0.9366 | 0 | 0 | 0 | 1 | Ref |
| 4388 | KY676783 | Unassigned | Unassigned | Unassigned | 371_15 | Clustered | 44 | VC_371_15 | 44 | 0.1391 | 1 | 0.1391 | 0 | 0 | 0 | 0.7474 | Ref |
| 4389 | KY676784 | Unassigned | Unassigned | Unassigned | 727_0 | Clustered | 11 | VC_727_0 | 11 | 0.5054 | 1 | 0.5054 | 0 | 0 | 0 | 1 | Ref |
| 4390 | KY677846 | Unassigned | Unassigned | Unassigned | 190_15 | Clustered | 3 | VC_190_15 | 3 | 0.0146 | 0.99759465 | 0.0145 | 0 | 0 | 0 | 0.6667 | Ref |
| 4391 | KY683735 | Unassigned | Unassigned | Unassigned | 590_0 | Clustered | 27 | VC_590_0 | 27 | 0.9947 | 1 | 0.9947 | 0 | 0 | 0 | 1 | Ref |
| 4392 | KY683736 | Unassigned | Unassigned | Unassigned | 772_9 | Clustered | 7 | VC_772_9 | 7 | 0.0369 | 0.87705503 | 0.0324 | 0 | 0 | 0 | 1 | Ref |
| 4393 | KY684082 | Unassigned | Unassigned | Unassigned | 111_0 | Clustered | 91 | VC_111_0 | 91 | 0.9587 | 1 | 0.9587 | 0 | 0 | 0 | 1 | Ref |
| 4394 | KY694971 | Unassigned | Unassigned | Unassigned | 584_0 | Clustered | 28 | VC_584_0 | 28 | 0.7704 | 1 | 0.7704 | 0 | 0 | 0 | 1 | Ref |
| 4395 | KY695153 | Unassigned | Unassigned | Unassigned | 7_0 | Clustered | 28 | VC_7_0 | 28 | 0.3916 | 1 | 0.3916 | 0 | 0 | 0 | 0.9947 | Ref |
| 4396 | KY703222 | Unassigned | Unassigned | Unassigned | 286_10 | Clustered | 9 | VC_286_10 | 9 | 0.0615 | 0.99999878 | 0.0615 | 0 | 0 | 0 | 0.6667 | Ref |
| 4397 | KY705251 | Unassigned | Unassigned | Unassigned | 252_0 | Clustered | 121 | VC_252_0 | 121 | 0.7376 | 1 | 0.7376 | 0 | 0 | 0 | 0.9999 | Ref |
| 4398 | KY705252 | Unassigned | Unassigned | Unassigned | 255_0 | Clustered | 13 | VC_255_0 | 13 | 0.2048 | 1 | 0.2048 | 0 | 0 | 0 | 1 | Ref |
| 4399 | KY705253 | Unassigned | Unassigned | Unassigned | 255_0 | Clustered | 13 | VC_255_0 | 13 | 0.2048 | 1 | 0.2048 | 0 | 0 | 0 | 1 | Ref |
| 4400 | KY705254 | Unassigned | Unassigned | Unassigned | 255_0 | Clustered | 13 | VC_255_0 | 13 | 0.2048 | 1 | 0.2048 | 0 | 0 | 0 | 1 | Ref |
| 4401 | KY705255 | Unassigned | Unassigned | Unassigned | 255_0 | Clustered | 13 | VC_255_0 | 13 | 0.2048 | 1 | 0.2048 | 0 | 0 | 0 | 1 | Ref |
| 4402 | KY705256 | Unassigned | Unassigned | Unassigned | 252_0 | Clustered | 121 | VC_252_0 | 121 | 0.7376 | 1 | 0.7376 | 0 | 0 | 0 | 0.9999 | Ref |
| 4403 | KY705257 | Unassigned | Unassigned | Unassigned | 252_0 | Clustered | 121 | VC_252_0 | 121 | 0.7376 | 1 | 0.7376 | 0 | 0 | 0 | 0.9999 | Ref |
| 4404 | KY705258 | Unassigned | Unassigned | Unassigned | 254_0 | Clustered | 46 | VC_254_0 | 46 | 0.5159 | 1 | 0.5159 | 0 | 0 | 0 | 1 | Ref |
| 4405 | KY705259 | Unassigned | Unassigned | Unassigned | 252_0 | Clustered | 121 | VC_252_0 | 121 | 0.7376 | 1 | 0.7376 | 0 | 0 | 0 | 0.9999 | Ref |
| 4406 | KY705260 | Unassigned | Unassigned | Unassigned | 252_0 | Clustered | 121 | VC_252_0 | 121 | 0.7376 | 1 | 0.7376 | 0 | 0 | 0 | 0.9999 | Ref |
| 4407 | KY705261 | Unassigned | Unassigned | Unassigned | 252_0 | Clustered | 121 | VC_252_0 | 121 | 0.7376 | 1 | 0.7376 | 0 | 0 | 0 | 0.9999 | Ref |
| 4408 | KY705262 | Unassigned | Unassigned | Unassigned | 252_0 | Clustered | 121 | VC_252_0 | 121 | 0.7376 | 1 | 0.7376 | 0 | 0 | 0 | 0.9999 | Ref |
| 4409 | KY705263 | Unassigned | Unassigned | Unassigned | 252_0 | Clustered | 121 | VC_252_0 | 121 | 0.7376 | 1 | 0.7376 | 0 | 0 | 0 | 0.9999 | Ref |
| 4410 | KY705264 | Unassigned | Unassigned | Unassigned | 252_0 | Clustered | 121 | VC_252_0 | 121 | 0.7376 | 1 | 0.7376 | 0 | 0 | 0 | 0.9999 | Ref |
| 4411 | KY705265 | Unassigned | Unassigned | Unassigned | 252_0 | Clustered | 121 | VC_252_0 | 121 | 0.7376 | 1 | 0.7376 | 0 | 0 | 0 | 0.9999 | Ref |
| 4412 | KY705266 | Unassigned | Unassigned | Unassigned | 252_0 | Clustered | 121 | VC_252_0 | 121 | 0.7376 | 1 | 0.7376 | 0 | 0 | 0 | 0.9999 | Ref |
| 4413 | KY705267 | Unassigned | Unassigned | Unassigned | 252_0 | Clustered | 121 | VC_252_0 | 121 | 0.7376 | 1 | 0.7376 | 0 | 0 | 0 | 0.9999 | Ref |
| 4414 | KY705268 | Unassigned | Unassigned | Unassigned | 254_0 | Clustered | 46 | VC_254_0 | 46 | 0.5159 | 1 | 0.5159 | 0 | 0 | 0 | 1 | Ref |
| 4415 | KY705269 | Unassigned | Unassigned | Unassigned | 254_0 | Clustered | 46 | VC_254_0 | 46 | 0.5159 | 1 | 0.5159 | 0 | 0 | 0 | 1 | Ref |
| 4416 | KY705270 | Unassigned | Unassigned | Unassigned | 252_0 | Clustered | 121 | VC_252_0 | 121 | 0.7376 | 1 | 0.7376 | 0 | 0 | 0 | 0.9999 | Ref |
| 4417 | KY705271 | Unassigned | Unassigned | Unassigned | 252_0 | Clustered | 121 | VC_252_0 | 121 | 0.7376 | 1 | 0.7376 | 0 | 0 | 0 | 0.9999 | Ref |
| 4418 | KY705272 | Unassigned | Unassigned | Unassigned | 252_0 | Clustered | 121 | VC_252_0 | 121 | 0.7376 | 1 | 0.7376 | 0 | 0 | 0 | 0.9999 | Ref |
| 4419 | KY705273 | Unassigned | Unassigned | Unassigned | 252_0 | Clustered | 121 | VC_252_0 | 121 | 0.7376 | 1 | 0.7376 | 0 | 0 | 0 | 0.9999 | Ref |
| 4420 | KY705274 | Unassigned | Unassigned | Unassigned | 252_0 | Clustered | 121 | VC_252_0 | 121 | 0.7376 | 1 | 0.7376 | 0 | 0 | 0 | 0.9999 | Ref |
| 4421 | KY705275 | Unassigned | Unassigned | Unassigned | 252_0 | Clustered | 121 | VC_252_0 | 121 | 0.7376 | 1 | 0.7376 | 0 | 0 | 0 | 0.9999 | Ref |
| 4422 | KY705276 | Unassigned | Unassigned | Unassigned | 252_0 | Clustered | 121 | VC_252_0 | 121 | 0.7376 | 1 | 0.7376 | 0 | 0 | 0 | 0.9999 | Ref |
| 4423 | KY705277 | Unassigned | Unassigned | Unassigned | 254_0 | Clustered | 46 | VC_254_0 | 46 | 0.5159 | 1 | 0.5159 | 0 | 0 | 0 | 1 | Ref |
| 4424 | KY705278 | Unassigned | Unassigned | Unassigned | 254_0 | Clustered | 46 | VC_254_0 | 46 | 0.5159 | 1 | 0.5159 | 0 | 0 | 0 | 1 | Ref |
| 4425 | KY705279 | Unassigned | Unassigned | Unassigned | 254_0 | Clustered | 46 | VC_254_0 | 46 | 0.5159 | 1 | 0.5159 | 0 | 0 | 0 | 1 | Ref |
| 4426 | KY705280 | Unassigned | Unassigned | Unassigned | 254_0 | Clustered | 46 | VC_254_0 | 46 | 0.5159 | 1 | 0.5159 | 0 | 0 | 0 | 1 | Ref |
| 4427 | KY705281 | Unassigned | Unassigned | Unassigned | 254_0 | Clustered | 46 | VC_254_0 | 46 | 0.5159 | 1 | 0.5159 | 0 | 0 | 0 | 1 | Ref |
| 4428 | KY705282 | Unassigned | Unassigned | Unassigned | 252_0 | Clustered | 121 | VC_252_0 | 121 | 0.7376 | 1 | 0.7376 | 0 | 0 | 0 | 0.9999 | Ref |
| 4429 | KY705283 | Unassigned | Unassigned | Unassigned | 252_0 | Clustered | 121 | VC_252_0 | 121 | 0.7376 | 1 | 0.7376 | 0 | 0 | 0 | 0.9999 | Ref |
| 4430 | KY705284 | Unassigned | Unassigned | Unassigned | 252_0 | Clustered | 121 | VC_252_0 | 121 | 0.7376 | 1 | 0.7376 | 0 | 0 | 0 | 0.9999 | Ref |
| 4431 | KY705285 | Unassigned | Unassigned | Unassigned | 254_0 | Clustered | 46 | VC_254_0 | 46 | 0.5159 | 1 | 0.5159 | 0 | 0 | 0 | 1 | Ref |
| 4432 | KY705286 | Unassigned | Unassigned | Unassigned | 254_0 | Clustered | 46 | VC_254_0 | 46 | 0.5159 | 1 | 0.5159 | 0 | 0 | 0 | 1 | Ref |
| 4433 | KY705287 | Unassigned | Unassigned | Unassigned | 252_0 | Clustered | 121 | VC_252_0 | 121 | 0.7376 | 1 | 0.7376 | 0 | 0 | 0 | 0.9999 | Ref |
| 4434 | KY705288 | Unassigned | Unassigned | Unassigned | 252_0 | Clustered | 121 | VC_252_0 | 121 | 0.7376 | 1 | 0.7376 | 0 | 0 | 0 | 0.9999 | Ref |
| 4435 | KY705289 | Unassigned | Unassigned | Unassigned | 252_0 | Clustered | 121 | VC_252_0 | 121 | 0.7376 | 1 | 0.7376 | 0 | 0 | 0 | 0.9999 | Ref |
| 4436 | KY705290 | Unassigned | Unassigned | Unassigned | 252_0 | Clustered | 121 | VC_252_0 | 121 | 0.7376 | 1 | 0.7376 | 0 | 0 | 0 | 0.9999 | Ref |
| 4437 | KY705409 | Unassigned | Unassigned | Unassigned | 125_0 | Clustered | 17 | VC_125_0 | 17 | 0.9244 | 1 | 0.9244 | 0 | 0 | 0 | 1 | Ref |
| 4438 | KY707339 | Unassigned | Unassigned | Unassigned | 747_0 | Clustered | 23 | VC_747_0 | 23 | 0.8202 | 1 | 0.8202 | 0 | 0 | 0 | 1 | Ref |
| 4439 | KY709296 | Unassigned | Unassigned | Unassigned | 793_0 | Clustered | 12 | VC_793_0 | 12 | 0.8531 | 1 | 0.8531 | 0 | 0 | 0 | 1 | Ref |
| 4440 | KY744228 | Unassigned | Unassigned | Unassigned | 348_0 | Clustered | 22 | VC_348_0 | 22 | 0.9602 | 1 | 0.9602 | 0 | 0 | 0 | 1 | Ref |
| 4441 | KY744229 | Unassigned | Unassigned | Unassigned | 348_0 | Clustered | 22 | VC_348_0 | 22 | 0.9602 | 1 | 0.9602 | 0 | 0 | 0 | 1 | Ref |
| 4442 | KY744230 | Unassigned | Unassigned | Unassigned | 348_0 | Clustered | 22 | VC_348_0 | 22 | 0.9602 | 1 | 0.9602 | 0 | 0 | 0 | 1 | Ref |
| 4443 | KY744231 | Unassigned | Unassigned | Unassigned | 348_0 | Clustered | 22 | VC_348_0 | 22 | 0.9602 | 1 | 0.9602 | 0 | 0 | 0 | 1 | Ref |
| 4444 | KY744232 | Unassigned | Unassigned | Unassigned | 348_0 | Clustered | 22 | VC_348_0 | 22 | 0.9602 | 1 | 0.9602 | 0 | 0 | 0 | 1 | Ref |
| 4445 | KY744233 | Unassigned | Unassigned | Unassigned | 348_0 | Clustered | 22 | VC_348_0 | 22 | 0.9602 | 1 | 0.9602 | 0 | 0 | 0 | 1 | Ref |
| 4446 | KY744234 | Unassigned | Unassigned | Unassigned | 348_0 | Clustered | 22 | VC_348_0 | 22 | 0.9602 | 1 | 0.9602 | 0 | 0 | 0 | 1 | Ref |
| 4447 | KY744235 | Unassigned | Unassigned | Unassigned | 348_0 | Clustered | 22 | VC_348_0 | 22 | 0.9602 | 1 | 0.9602 | 0 | 0 | 0 | 1 | Ref |
| 4448 | KY744240 | Unassigned | Unassigned | Unassigned | 269_0 | Clustered | 3 | VC_269_0 | 3 | 0.1948 | 0.99308177 | 0.1934 | 0 | 0 | 0 | 1 | Ref |
| 4449 | KY744566 | Unassigned | Unassigned | Unassigned | 452_0 | Clustered | 66 | VC_452_0 | 66 | 0.9248 | 1 | 0.9248 | 0 | 0 | 0 | 1 | Ref |
| 4450 | KY769270 | Unassigned | Unassigned | Unassigned | 841_0 | Clustered | 10 | VC_841_0 | 10 | 0.5997 | 1 | 0.5997 | 0 | 0 | 0 | 1 | Ref |
| 4451 | KY779848 | Unassigned | Unassigned | Unassigned | 153_0 | Clustered | 25 | VC_153_0 | 25 | 0.7729 | 1 | 0.7729 | 0 | 0 | 0 | 1 | Ref |
| 4452 | KY779849 | Unassigned | Unassigned | Unassigned | 149_15 | Clustered | 16 | VC_149_15 | 16 | 0.0788 | 1 | 0.0788 | 0 | 0 | 0 | 0.8083 | Ref |
| 4453 | KY780482 | Unassigned | Unassigned | Unassigned | 937_0 | Clustered | 40 | VC_937_0 | 40 | 0.6809 | 1 | 0.6809 | 0 | 0 | 0 | 0.9359 | Ref |
| 4454 | KY783914 | Unassigned | Unassigned | Unassigned | 763_2 | Clustered | 7 | VC_763_2 | 7 | 0.2612 | 1 | 0.2612 | 0 | 0 | 0 | 1 | Ref |
| 4455 | KY787212 | Unassigned | Unassigned | Unassigned | 191_16 | Clustered | 12 | VC_191_16 | 12 | 0.0643 | 1 | 0.0643 | 0 | 0 | 0 | 0.9394 | Ref |
| 4456 | KY787213 | Unassigned | Unassigned | Unassigned | 133_34 | Clustered | 3 | VC_133_34 | 3 | 0.0102 | 0.97369044 | 0.01 | 0 | 0 | 0 | 0.6667 | Ref |
| 4457 | KY794641 | Unassigned | Unassigned | Unassigned | 149_10 | Clustered/Singleton | 1 | VC_149_10 | 1 | 0 | 1 | 0 | 0 | 0 | 0 | 1 | Ref |
| 4458 | KY794642 | Unassigned | Unassigned | Unassigned | 149_29 | Clustered | 3 | VC_149_29 | 3 | 0.012 | 0.99448577 | 0.012 | 0 | 0 | 0 | 1 | Ref |
| 4459 | KY794643 | Unassigned | Unassigned | Unassigned | 149_29 | Clustered | 3 | VC_149_29 | 3 | 0.012 | 0.99448577 | 0.012 | 0 | 0 | 0 | 1 | Ref |
| 4460 | KY798120 | Unassigned | Unassigned | Unassigned | 129_0 | Clustered | 43 | VC_129_0 | 43 | 0.886 | 1 | 0.886 | 0 | 0 | 0 | 1 | Ref |
| 4461 | KY798121 | Unassigned | Unassigned | Unassigned | 129_0 | Clustered | 43 | VC_129_0 | 43 | 0.886 | 1 | 0.886 | 0 | 0 | 0 | 1 | Ref |
| 4462 | KY817360 | Unassigned | Unassigned | Unassigned | 1201_0 | Clustered | 2 | VC_1201_0 | 2 | 0.8919 | 0.81445332 | 0.7264 | 0 | 0 | 0 | 1 | Ref |
| 4463 | KY829116 | Unassigned | Unassigned | Unassigned | 698_0 | Clustered | 26 | VC_698_0 | 26 | 0.9366 | 1 | 0.9366 | 0 | 0 | 0 | 1 | Ref |
| 4464 | KY853667 | Unassigned | Unassigned | Unassigned | 177_0 | Clustered | 5 | VC_177_0 | 5 | 0.7468 | 0.99885646 | 0.746 | 0 | 0 | 0 | 1 | Ref |
| 4465 | KY860935 | Unassigned | Unassigned | Unassigned | 461_0 | Clustered | 96 | VC_461_0 | 96 | 0.9307 | 1 | 0.9307 | 0 | 0 | 0 | 0.9575 | Ref |
| 4466 | KY883634 | Unassigned | Unassigned | Unassigned | 495_1 | Clustered | 41 | VC_495_1 | 41 | 0.328 | 1 | 0.328 | 0 | 0 | 0 | 0.911 | Ref |
| 4467 | KY883635 | Unassigned | Unassigned | Unassigned | 495_1 | Clustered | 41 | VC_495_1 | 41 | 0.328 | 1 | 0.328 | 0 | 0 | 0 | 0.911 | Ref |
| 4468 | KY883636 | Unassigned | Unassigned | Unassigned | 495_1 | Clustered | 41 | VC_495_1 | 41 | 0.328 | 1 | 0.328 | 0 | 0 | 0 | 0.911 | Ref |
| 4469 | KY883637 | Unassigned | Unassigned | Unassigned | 495_1 | Clustered | 41 | VC_495_1 | 41 | 0.328 | 1 | 0.328 | 0 | 0 | 0 | 0.911 | Ref |
| 4470 | KY883638 | Unassigned | Unassigned | Unassigned | 495_0 | Clustered | 39 | VC_495_0 | 39 | 0.308 | 1 | 0.308 | 0 | 0 | 0 | 1 | Ref |
| 4471 | KY883639 | Unassigned | Unassigned | Unassigned | 495_0 | Clustered | 39 | VC_495_0 | 39 | 0.308 | 1 | 0.308 | 0 | 0 | 0 | 1 | Ref |
| 4472 | KY883640 | Unassigned | Unassigned | Unassigned | 495_0 | Clustered | 39 | VC_495_0 | 39 | 0.308 | 1 | 0.308 | 0 | 0 | 0 | 1 | Ref |
| 4473 | KY883641 | Unassigned | Unassigned | Unassigned | 82_0 | Clustered | 24 | VC_82_0 | 24 | 0.7351 | 1 | 0.7351 | 0 | 0 | 0 | 1 | Ref |
| 4474 | KY883642 | Unassigned | Unassigned | Unassigned | 607_0 | Clustered | 16 | VC_607_0 | 16 | 0.9179 | 1 | 0.9179 | 0 | 0 | 0 | 1 | Ref |
| 4475 | KY883643 | Unassigned | Unassigned | Unassigned | 82_0 | Clustered | 24 | VC_82_0 | 24 | 0.7351 | 1 | 0.7351 | 0 | 0 | 0 | 1 | Ref |
| 4476 | KY883644 | Unassigned | Unassigned | Unassigned | 82_0 | Clustered | 24 | VC_82_0 | 24 | 0.7351 | 1 | 0.7351 | 0 | 0 | 0 | 1 | Ref |
| 4477 | KY883645 | Unassigned | Unassigned | Unassigned | 82_0 | Clustered | 24 | VC_82_0 | 24 | 0.7351 | 1 | 0.7351 | 0 | 0 | 0 | 1 | Ref |
| 4478 | KY883646 | Unassigned | Unassigned | Unassigned | 82_0 | Clustered | 24 | VC_82_0 | 24 | 0.7351 | 1 | 0.7351 | 0 | 0 | 0 | 1 | Ref |
| 4479 | KY883647 | Unassigned | Unassigned | Unassigned | 607_0 | Clustered | 16 | VC_607_0 | 16 | 0.9179 | 1 | 0.9179 | 0 | 0 | 0 | 1 | Ref |
| 4480 | KY883648 | Unassigned | Unassigned | Unassigned | 82_0 | Clustered | 24 | VC_82_0 | 24 | 0.7351 | 1 | 0.7351 | 0 | 0 | 0 | 1 | Ref |
| 4481 | KY883649 | Unassigned | Unassigned | Unassigned | 82_0 | Clustered | 24 | VC_82_0 | 24 | 0.7351 | 1 | 0.7351 | 0 | 0 | 0 | 1 | Ref |
| 4482 | KY883650 | Unassigned | Unassigned | Unassigned | 82_0 | Clustered | 24 | VC_82_0 | 24 | 0.7351 | 1 | 0.7351 | 0 | 0 | 0 | 1 | Ref |
| 4483 | KY883651 | Unassigned | Unassigned | Unassigned | 82_0 | Clustered | 24 | VC_82_0 | 24 | 0.7351 | 1 | 0.7351 | 0 | 0 | 0 | 1 | Ref |
| 4484 | KY883652 | Unassigned | Unassigned | Unassigned | 82_0 | Clustered | 24 | VC_82_0 | 24 | 0.7351 | 1 | 0.7351 | 0 | 0 | 0 | 1 | Ref |
| 4485 | KY883653 | Unassigned | Unassigned | Unassigned | 82_0 | Clustered | 24 | VC_82_0 | 24 | 0.7351 | 1 | 0.7351 | 0 | 0 | 0 | 1 | Ref |
| 4486 | KY883654 | Unassigned | Unassigned | Unassigned | 971_0 | Clustered | 3 | VC_971_0 | 3 | 0.0548 | 0.99838914 | 0.0547 | 0 | 0 | 0 | 1 | Ref |
| 4487 | KY883655 | Unassigned | Unassigned | Unassigned | 971_0 | Clustered | 3 | VC_971_0 | 3 | 0.0548 | 0.99838914 | 0.0547 | 0 | 0 | 0 | 1 | Ref |
| 4488 | KY883656 | Unassigned | Unassigned | Unassigned | 607_0 | Clustered | 16 | VC_607_0 | 16 | 0.9179 | 1 | 0.9179 | 0 | 0 | 0 | 1 | Ref |
| 4489 | KY883657 | Unassigned | Unassigned | Unassigned | 754_0 | Clustered | 8 | VC_754_0 | 8 | 0.752 | 1 | 0.752 | 0 | 0 | 0 | 1 | Ref |
| 4490 | KY883658 | Unassigned | Unassigned | Unassigned | 754_0 | Clustered | 8 | VC_754_0 | 8 | 0.752 | 1 | 0.752 | 0 | 0 | 0 | 1 | Ref |
| 4491 | KY888143 | Unassigned | Unassigned | Unassigned | 209_0 | Clustered | 4 | VC_209_0 | 4 | 0.1136 | 0.99998779 | 0.1136 | 0 | 0 | 0 | 1 | Ref |
| 4492 | KY888680 | Unassigned | Unassigned | Unassigned | 699_0 | Clustered | 71 | VC_699_0 | 71 | 0.9352 | 1 | 0.9352 | 0 | 0 | 0 | 1 | Ref |
| 4493 | KY888882 | Unassigned | Unassigned | Unassigned | 883_3 | Clustered | 2 | VC_883_3 | 2 | 0.0182 | 0.87900236 | 0.016 | 0 | 0 | 0 | 1 | Ref |
| 4494 | KY914478 | Unassigned | Unassigned | Unassigned | 42_0 | Clustered | 49 | VC_42_0 | 49 | 0.4517 | 1 | 0.4517 | 0 | 0 | 0 | 0.8597 | Ref |
| 4495 | KY926698 | Unassigned | Unassigned | Unassigned | 209_0 | Clustered | 4 | VC_209_0 | 4 | 0.1136 | 0.99998779 | 0.1136 | 0 | 0 | 0 | 1 | Ref |
| 4496 | KY926699 | Unassigned | Unassigned | Unassigned | 209_0 | Clustered | 4 | VC_209_0 | 4 | 0.1136 | 0.99998779 | 0.1136 | 0 | 0 | 0 | 1 | Ref |
| 4497 | KY926791 | Unassigned | Unassigned | Unassigned | 722_0 | Clustered | 13 | VC_722_0 | 13 | 0.5104 | 1 | 0.5104 | 0 | 0 | 0 | 1 | Ref |
| 4498 | KY926792 | Unassigned | Unassigned | Unassigned | 765_0 | Clustered | 132 | VC_765_0 | 132 | 1 | 1 | 1 | 0 | 0 | 0 | 1 | Ref |
| 4499 | KY926793 | Unassigned | Unassigned | Unassigned | 765_0 | Clustered | 132 | VC_765_0 | 132 | 1 | 1 | 1 | 0 | 0 | 0 | 1 | Ref |
| 4500 | KY942056 | Unassigned | Unassigned | Unassigned | 131_0 | Clustered | 17 | VC_131_0 | 17 | 0.2671 | 1 | 0.2671 | 0 | 0 | 0 | 1 | Ref |
| 4501 | KY942057 | Unassigned | Unassigned | Unassigned | 131_0 | Clustered | 17 | VC_131_0 | 17 | 0.2671 | 1 | 0.2671 | 0 | 0 | 0 | 1 | Ref |
| 4502 | KY945355 | Unassigned | Unassigned | Unassigned | 237_2 | Clustered | 11 | VC_237_2 | 11 | 0.1409 | 1 | 0.1409 | 0 | 0 | 0 | 0.9818 | Ref |
| 4503 | KY947509 | Unassigned | Unassigned | Unassigned | 818_0 | Clustered | 20 | VC_818_0 | 20 | 0.9601 | 1 | 0.9601 | 0 | 0 | 0 | 1 | Ref |
| 4504 | KY951963 | Unassigned | Unassigned | Unassigned | 912_0 | Clustered | 76 | VC_912_0 | 76 | 0.979 | 1 | 0.979 | 0 | 0 | 0 | 1 | Ref |
| 4505 | KY951964 | Unassigned | Unassigned | Unassigned | 912_0 | Clustered | 76 | VC_912_0 | 76 | 0.979 | 1 | 0.979 | 0 | 0 | 0 | 1 | Ref |
| 4506 | KY953156 | Unassigned | Unassigned | Unassigned | 984_0 | Clustered | 38 | VC_984_0 | 38 | 0.9758 | 1 | 0.9758 | 0 | 0 | 0 | 1 | Ref |
| 4507 | KY962008 | Unassigned | Unassigned | Unassigned | 451_0 | Clustered | 80 | VC_451_0 | 80 | 0.9991 | 1 | 0.9991 | 0 | 0 | 0 | 1 | Ref |
| 4508 | KY963369 | Unassigned | Unassigned | Unassigned | 704_0 | Clustered | 16 | VC_704_0 | 16 | 0.7274 | 1 | 0.7274 | 0 | 0 | 0 | 1 | Ref |
| 4509 | KY963370 | Unassigned | Unassigned | Unassigned | 704_0 | Clustered | 16 | VC_704_0 | 16 | 0.7274 | 1 | 0.7274 | 0 | 0 | 0 | 1 | Ref |
| 4510 | KY963371 | Unassigned | Unassigned | Unassigned | 704_0 | Clustered | 16 | VC_704_0 | 16 | 0.7274 | 1 | 0.7274 | 0 | 0 | 0 | 1 | Ref |
| 4511 | KY963424 | Unassigned | Unassigned | Unassigned | 190_23 | Clustered | 10 | VC_190_23 | 10 | 0.0704 | 1 | 0.0704 | 0 | 0 | 0 | 0.8444 | Ref |
| 4512 | KY965063 | Unassigned | Unassigned | Unassigned | 514_0 | Clustered | 75 | VC_514_0 | 75 | 0.9402 | 1 | 0.9402 | 0 | 0 | 0 | 1 | Ref |
| 4513 | KY965064 | Unassigned | Unassigned | Unassigned | 520_40 | Clustered | 4 | VC_520_40 | 4 | 0.0396 | 0.99998776 | 0.0396 | 0 | 0 | 0 | 1 | Ref |
| 4514 | KY965065 | Unassigned | Unassigned | Unassigned | 216_6 | Clustered | 42 | VC_216_6 | 42 | 0.473 | 1 | 0.473 | 0 | 0 | 0 | 0.8351 | Ref |
| 4515 | KY965066 | Unassigned | Unassigned | Unassigned | 371_18 | Clustered | 10 | VC_371_18 | 10 | 0.0302 | 1 | 0.0302 | 0 | 0 | 0 | 0.8 | Ref |
| 4516 | KY965934 | Unassigned | Unassigned | Unassigned | 963_0 | Clustered | 4 | VC_963_0 | 4 | 0.8505 | 0.99879268 | 0.8495 | 0 | 0 | 0 | 1 | Ref |
| 4517 | KY969628 | Unassigned | Unassigned | Unassigned | 568_0 | Clustered | 8 | VC_568_0 | 8 | 0.5738 | 1 | 0.5738 | 0 | 0 | 0 | 1 | Ref |
| 4518 | KY971610 | Unassigned | Unassigned | Unassigned | 287_1 | Clustered/Singleton | 1 | VC_287_1 | 1 | 0 | 1 | 0 | 0 | 0 | 0 | 1 | Ref |
| 4519 | KY971611 | Unassigned | Unassigned | Unassigned | 129_0 | Clustered | 43 | VC_129_0 | 43 | 0.886 | 1 | 0.886 | 0 | 0 | 0 | 1 | Ref |
| 4520 | KY979235 | Unassigned | Unassigned | Unassigned | 912_0 | Clustered | 76 | VC_912_0 | 76 | 0.979 | 1 | 0.979 | 0 | 0 | 0 | 1 | Ref |
| 4521 | KY979236 | Unassigned | Unassigned | Unassigned | 912_0 | Clustered | 76 | VC_912_0 | 76 | 0.979 | 1 | 0.979 | 0 | 0 | 0 | 1 | Ref |
| 4522 | KY979237 | Unassigned | Unassigned | Unassigned | 912_0 | Clustered | 76 | VC_912_0 | 76 | 0.979 | 1 | 0.979 | 0 | 0 | 0 | 1 | Ref |
| 4523 | KY979238 | Unassigned | Unassigned | Unassigned | 912_0 | Clustered | 76 | VC_912_0 | 76 | 0.979 | 1 | 0.979 | 0 | 0 | 0 | 1 | Ref |
| 4524 | KY979239 | Unassigned | Unassigned | Unassigned | 912_0 | Clustered | 76 | VC_912_0 | 76 | 0.979 | 1 | 0.979 | 0 | 0 | 0 | 1 | Ref |
| 4525 | KY979240 | Unassigned | Unassigned | Unassigned | 912_0 | Clustered | 76 | VC_912_0 | 76 | 0.979 | 1 | 0.979 | 0 | 0 | 0 | 1 | Ref |
| 4526 | KY979241 | Unassigned | Unassigned | Unassigned | 912_0 | Clustered | 76 | VC_912_0 | 76 | 0.979 | 1 | 0.979 | 0 | 0 | 0 | 1 | Ref |
| 4527 | KY979242 | Unassigned | Unassigned | Unassigned | 912_0 | Clustered | 76 | VC_912_0 | 76 | 0.979 | 1 | 0.979 | 0 | 0 | 0 | 1 | Ref |
| 4528 | KY979243 | Unassigned | Unassigned | Unassigned | 912_0 | Clustered | 76 | VC_912_0 | 76 | 0.979 | 1 | 0.979 | 0 | 0 | 0 | 1 | Ref |
| 4529 | KY979244 | Unassigned | Unassigned | Unassigned | 912_0 | Clustered | 76 | VC_912_0 | 76 | 0.979 | 1 | 0.979 | 0 | 0 | 0 | 1 | Ref |
| 4530 | KY979245 | Unassigned | Unassigned | Unassigned | 912_0 | Clustered | 76 | VC_912_0 | 76 | 0.979 | 1 | 0.979 | 0 | 0 | 0 | 1 | Ref |
| 4531 | KY979246 | Unassigned | Unassigned | Unassigned | 912_0 | Clustered | 76 | VC_912_0 | 76 | 0.979 | 1 | 0.979 | 0 | 0 | 0 | 1 | Ref |
| 4532 | KY979247 | Unassigned | Unassigned | Unassigned | 912_0 | Clustered | 76 | VC_912_0 | 76 | 0.979 | 1 | 0.979 | 0 | 0 | 0 | 1 | Ref |
| 4533 | KY981272 | Unassigned | Unassigned | Unassigned | 70_0 | Clustered | 27 | VC_70_0 | 27 | 0.5981 | 1 | 0.5981 | 0 | 0 | 0 | 0.9886 | Ref |
| 4534 | KY984068 | Unassigned | Unassigned | Unassigned | 67_2 | Clustered | 2 | VC_67_2 | 2 | 0.014 | 0.69357611 | 0.0097 | 0 | 0 | 0 | 1 | Ref |

| | | | | | | | | | | | | | | | | | |
|---|---|---|---|---|---|---|---|---|---|---|---|---|---|---|---|---|---|
| 4535 | KY985004 | Unassigned | Unassigned | Unassigned | 575_0 | Clustered | 35 | VC_575_0 | 35 | 0.8815 | 1 | 0.8815 | 0 | 0 | 0 | 1 | Ref |
| 4536 | KY992519 | Unassigned | Unassigned | Unassigned | 912_0 | Clustered | 76 | VC_912_0 | 76 | 0.979 | 1 | 0.979 | 0 | 0 | 0 | 1 | Ref |
| 4537 | KY992520 | Unassigned | Unassigned | Unassigned | 912_0 | Clustered | 76 | VC_912_0 | 76 | 0.979 | 1 | 0.979 | 0 | 0 | 0 | 1 | Ref |
| 4538 | KY994101 | Unassigned | Unassigned | Unassigned | 77_0 | Clustered | 20 | VC_77_0 | 20 | 0.9657 | 1 | 0.9657 | 0 | 0 | 0 | 1 | Ref |
| 4539 | KY996523 | Unassigned | Unassigned | Unassigned | 897_0 | Clustered | 6 | VC_897_0 | 6 | 0.4156 | 1 | 0.4156 | 0 | 0 | 0 | 1 | Ref |
| 4540 | L67CG | Unassigned | Unassigned | Unassigned | 690_0 | Clustered | 50 | VC_690_0 | 50 | 0.9805 | 1 | 0.9805 | 0 | 0 | 0 | 1 | Ref |
| 4541 | LAMCG | Unassigned | Unassigned | Unassigned | 43_0 | Clustered | 38 | VC_43_0 | 38 | 0.5885 | 1 | 0.5885 | 0 | 0 | 0 | 0.9659 | Ref |
| 4542 | LC035386 | Unassigned | Unassigned | Unassigned | 1202_0 | Clustered | 2 | VC_1202_0 | 2 | 1 | 1 | 1 | 0 | 0 | 0 | 1 | Ref |
| 4543 | LC063634 | Unassigned | Unassigned | Unassigned | 984_0 | Clustered | 38 | VC_984_0 | 38 | 0.9758 | 1 | 0.9758 | 0 | 0 | 0 | 1 | Ref |
| 4544 | LC064302 | Unassigned | Unassigned | Unassigned | 839_0 | Clustered | 5 | VC_839_0 | 5 | 0.165 | 0.99999995 | 0.165 | 0 | 0 | 0 | 1 | Ref |
| 4545 | LC066596 | Unassigned | Unassigned | Unassigned | 54_0 | Clustered | 6 | VC_54_0 | 6 | 0.6725 | 0.9999999 | 0.6725 | 0 | 0 | 0 | 1 | Ref |
| 4546 | LC102729 | Unassigned | Unassigned | Unassigned | 174_0 | Clustered | 3 | VC_174_0 | 3 | 0.9624 | 0.99333557 | 0.9559 | 0 | 0 | 0 | 1 | Ref |
| 4547 | LC102730 | Unassigned | Unassigned | Unassigned | 79_0 | Clustered | 64 | VC_79_0 | 64 | 0.4185 | 1 | 0.4185 | 0 | 0 | 0 | 0.7649 | Ref |
| 4548 | LC105987 | Unassigned | Unassigned | Unassigned | 79_0 | Clustered | 64 | VC_79_0 | 64 | 0.4185 | 1 | 0.4185 | 0 | 0 | 0 | 0.7649 | Ref |
| 4549 | LC105988 | Unassigned | Unassigned | Unassigned | 79_0 | Clustered | 64 | VC_79_0 | 64 | 0.4185 | 1 | 0.4185 | 0 | 0 | 0 | 0.7649 | Ref |
| 4550 | LC105989 | Unassigned | Unassigned | Unassigned | 79_0 | Clustered | 64 | VC_79_0 | 64 | 0.4185 | 1 | 0.4185 | 0 | 0 | 0 | 0.7649 | Ref |
| 4551 | LC105990 | Unassigned | Unassigned | Unassigned | 79_0 | Clustered | 64 | VC_79_0 | 64 | 0.4185 | 1 | 0.4185 | 0 | 0 | 0 | 0.7649 | Ref |
| 4552 | LC168164 | Unassigned | Unassigned | Unassigned | 532_1 | Clustered/Singleton | 1 | VC_532_1 | 1 | 0 | 1 | 0 | 0 | 0 | 0 | 1 | Ref |
| 4553 | LC210520 | Unassigned | Unassigned | Unassigned | 1202_0 | Clustered | 2 | VC_1202_0 | 2 | 1 | 1 | 1 | 0 | 0 | 0 | 1 | Ref |
| 4554 | LC348379 | Unassigned | Unassigned | Unassigned | 283_182 | Clustered/Singleton | 1 | VC_283_182 | 1 | 0 | 1 | 0 | 0 | 0 | 0 | 1 | Ref |
| 4555 | LC348380 | Unassigned | Unassigned | Unassigned | 283_92 | Clustered | 4 | VC_283_92 | 4 | 0.0079 | 0.9999582 | 0.0079 | 0 | 0 | 0 | 1 | Ref |
| 4556 | LC373201 | Unassigned | Unassigned | Unassigned | 131_2 | Clustered | 7 | VC_131_2 | 7 | 0.0685 | 1 | 0.0685 | 0 | 0 | 0 | 0.9048 | Ref |
| 4557 | LC375533 | Unassigned | Unassigned | Unassigned | 984_0 | Clustered | 38 | VC_984_0 | 38 | 0.9758 | 1 | 0.9758 | 0 | 0 | 0 | 1 | Ref |
| 4558 | LC413193 | Unassigned | Unassigned | Unassigned | 823_0 | Clustered | 124 | VC_823_0 | 124 | 0.9492 | 1 | 0.9492 | 0 | 0 | 0 | 1 | Ref |
| 4559 | LC413194 | Unassigned | Unassigned | Unassigned | 823_0 | Clustered | 124 | VC_823_0 | 124 | 0.9492 | 1 | 0.9492 | 0 | 0 | 0 | 1 | Ref |
| 4560 | LC413195 | Unassigned | Unassigned | Unassigned | 823_0 | Clustered | 124 | VC_823_0 | 124 | 0.9492 | 1 | 0.9492 | 0 | 0 | 0 | 1 | Ref |
| 4561 | LC465543 | Unassigned | Unassigned | Unassigned | 283_15 | Clustered/Singleton | 1 | VC_283_15 | 1 | 0 | 1 | 0 | 0 | 0 | 0 | 1 | Ref |
| 4562 | LC472881 | Unassigned | Unassigned | Unassigned | 79_1 | Clustered | 2 | VC_79_1 | 2 | 0.006 | 0.95748743 | 0.0057 | 0 | 0 | 0 | 1 | Ref |
| 4563 | LC472882 | Unassigned | Unassigned | Unassigned | 80_0 | Clustered | 41 | VC_80_0 | 41 | 0.5837 | 1 | 0.5837 | 0 | 0 | 0 | 0.9805 | Ref |
| 4564 | LC472883 | Unassigned | Unassigned | Unassigned | 79_0 | Clustered | 64 | VC_79_0 | 64 | 0.4185 | 1 | 0.4185 | 0 | 0 | 0 | 0.7649 | Ref |
| 4565 | LC472884 | Unassigned | Unassigned | Unassigned | 79_0 | Clustered | 64 | VC_79_0 | 64 | 0.4185 | 1 | 0.4185 | 0 | 0 | 0 | 0.7649 | Ref |
| 4566 | LC473039 | Unassigned | Unassigned | Unassigned | 329_8 | Clustered | 2 | VC_329_8 | 2 | 0.0048 | 0.95654014 | 0.0046 | 0 | 0 | 0 | 1 | Ref |
| 4567 | LC487410 | Unassigned | Unassigned | Unassigned | 971_1 | Clustered | 24 | VC_971_1 | 24 | 0.7393 | 1 | 0.7393 | 0 | 0 | 0 | 0.8913 | Ref |
| 4568 | LC492751 | Unassigned | Unassigned | Unassigned | 153_0 | Clustered | 25 | VC_153_0 | 25 | 0.7729 | 1 | 0.7729 | 0 | 0 | 0 | 1 | Ref |
| 4569 | LC492752 | Unassigned | Unassigned | Unassigned | 153_0 | Clustered | 25 | VC_153_0 | 25 | 0.7729 | 1 | 0.7729 | 0 | 0 | 0 | 1 | Ref |
| 4570 | LC494302 | Unassigned | Unassigned | Unassigned | 164_11 | Clustered | 10 | VC_164_11 | 10 | 0.1507 | 1 | 0.1507 | 0 | 0 | 0 | 0.9111 | Ref |
| 4571 | LC507823 | Unassigned | Unassigned | Unassigned | 637_0 | Clustered | 115 | VC_637_0 | 115 | 0.731 | 1 | 0.731 | 0 | 0 | 0 | 0.8764 | Ref |
| 4572 | LC516895 | Unassigned | Unassigned | Unassigned | 575_0 | Clustered | 35 | VC_575_0 | 35 | 0.8815 | 1 | 0.8815 | 0 | 0 | 0 | 1 | Ref |
| 4573 | LC519489 | Unassigned | Unassigned | Unassigned | 329_7 | Clustered | 18 | VC_329_7 | 18 | 0.0938 | 1 | 0.0938 | 0 | 0 | 0 | 0.7778 | Ref |
| 4574 | LC519601 | Unassigned | Unassigned | Unassigned | 812_1 | Clustered | 4 | VC_812_1 | 4 | 0.2266 | 0.99998426 | 0.2266 | 0 | 0 | 0 | 1 | Ref |
| 4575 | LC528882 | Unassigned | Unassigned | Unassigned | 3_0 | Clustered | 17 | VC_3_0 | 17 | 0.5639 | 1 | 0.5639 | 0 | 0 | 0 | 1 | Ref |
| 4576 | LC541428 | Unassigned | Unassigned | Unassigned | 8_0 | Clustered | 58 | VC_8_0 | 58 | 0.8306 | 1 | 0.8306 | 0 | 0 | 0 | 0.9946 | Ref |
| 4577 | LC545443 | Unassigned | Unassigned | Unassigned | 118_0 | Clustered | 3 | VC_118_0 | 3 | 0.7119 | 0.99211494 | 0.7063 | 0 | 0 | 0 | 1 | Ref |
| 4578 | LC545444 | Unassigned | Unassigned | Unassigned | 118_0 | Clustered | 3 | VC_118_0 | 3 | 0.7119 | 0.99211494 | 0.7063 | 0 | 0 | 0 | 1 | Ref |
| 4579 | LC545445 | Unassigned | Unassigned | Unassigned | 119_0 | Clustered | 2 | VC_119_0 | 2 | 0.5516 | 0.91840661 | 0.5066 | 0 | 0 | 0 | 1 | Ref |
| 4580 | LC545446 | Unassigned | Unassigned | Unassigned | 119_0 | Clustered | 2 | VC_119_0 | 2 | 0.5516 | 0.91840661 | 0.5066 | 0 | 0 | 0 | 1 | Ref |
| 4581 | LC547238 | Unassigned | Unassigned | Unassigned | 108_0 | Clustered | 33 | VC_108_0 | 33 | 0.949 | 1 | 0.949 | 0 | 0 | 0 | 1 | Ref |
| 4582 | LC552830 | Unassigned | Unassigned | Unassigned | 749_0 | Clustered | 40 | VC_749_0 | 40 | 0.7517 | 1 | 0.7517 | 0 | 0 | 0 | 1 | Ref |
| 4583 | LC553734 | Unassigned | Unassigned | Unassigned | 813_0 | Clustered | 12 | VC_813_0 | 12 | 0.2347 | 1 | 0.2347 | 0 | 0 | 0 | 1 | Ref |
| 4584 | LC553735 | Unassigned | Unassigned | Unassigned | 813_1 | Clustered | 24 | VC_813_1 | 24 | 0.535 | 1 | 0.535 | 0 | 0 | 0 | 1 | Ref |
| 4585 | LC553736 | Unassigned | Unassigned | Unassigned | 592_0 | Clustered | 50 | VC_592_0 | 50 | 0.7314 | 1 | 0.7314 | 0 | 0 | 0 | 1 | Ref |
| 4586 | LC567830 | Unassigned | Unassigned | Unassigned | 40_0 | Clustered | 5 | VC_40_0 | 5 | 0.0844 | 0.99999995 | 0.0844 | 0 | 0 | 0 | 1 | Ref |
| 4587 | LC567838 | Unassigned | Unassigned | Unassigned | 40_0 | Clustered | 5 | VC_40_0 | 5 | 0.0844 | 0.99999995 | 0.0844 | 0 | 0 | 0 | 1 | Ref |
| 4588 | LC567842 | Unassigned | Unassigned | Unassigned | 40_0 | Clustered | 5 | VC_40_0 | 5 | 0.0844 | 0.99999995 | 0.0844 | 0 | 0 | 0 | 1 | Ref |
| 4589 | LC574321 | Unassigned | Unassigned | Unassigned | 149_15 | Clustered | 16 | VC_149_15 | 16 | 0.0788 | 1 | 0.0788 | 0 | 0 | 0 | 0.8083 | Ref |
| 4590 | LC576631 | Unassigned | Unassigned | Unassigned | 153_1 | Clustered/Singleton | 1 | VC_153_1 | 1 | 0 | 1 | 0 | 0 | 0 | 0 | 1 | Ref |
| 4591 | LC589952 | Unassigned | Unassigned | Unassigned | 669_0 | Clustered | 21 | VC_669_0 | 21 | 0.3623 | 1 | 0.3623 | 0 | 0 | 0 | 1 | Ref |
| 4592 | LC596377 | Unassigned | Unassigned | Unassigned | 91_0 | Clustered | 22 | VC_91_0 | 22 | 0.5318 | 1 | 0.5318 | 0 | 0 | 0 | 1 | Ref |
| 4593 | LC596378 | Unassigned | Unassigned | Unassigned | 91_0 | Clustered | 22 | VC_91_0 | 22 | 0.5318 | 1 | 0.5318 | 0 | 0 | 0 | 1 | Ref |
| 4594 | LC596379 | Unassigned | Unassigned | Unassigned | 91_0 | Clustered | 22 | VC_91_0 | 22 | 0.5318 | 1 | 0.5318 | 0 | 0 | 0 | 1 | Ref |
| 4595 | LC597490 | Unassigned | Unassigned | Unassigned | 90_0 | Clustered/Singleton | 1 | VC_90_0 | 1 | 0 | 1 | 0 | 0 | 0 | 0 | 1 | Ref |
| 4596 | LC616031 | Unassigned | Unassigned | Unassigned | 47_0 | Clustered | 48 | VC_47_0 | 48 | 0.7783 | 1 | 0.7783 | 0 | 0 | 0 | 1 | Ref |
| 4597 | LC616032 | Unassigned | Unassigned | Unassigned | 47_0 | Clustered | 48 | VC_47_0 | 48 | 0.7783 | 1 | 0.7783 | 0 | 0 | 0 | 1 | Ref |
| 4598 | LC616033 | Unassigned | Unassigned | Unassigned | 47_0 | Clustered | 48 | VC_47_0 | 48 | 0.7783 | 1 | 0.7783 | 0 | 0 | 0 | 1 | Ref |
| 4599 | LC616034 | Unassigned | Unassigned | Unassigned | 47_0 | Clustered | 48 | VC_47_0 | 48 | 0.7783 | 1 | 0.7783 | 0 | 0 | 0 | 1 | Ref |
| 4600 | LC616035 | Unassigned | Unassigned | Unassigned | 47_0 | Clustered | 48 | VC_47_0 | 48 | 0.7783 | 1 | 0.7783 | 0 | 0 | 0 | 1 | Ref |
| 4601 | LC616036 | Unassigned | Unassigned | Unassigned | 47_0 | Clustered | 48 | VC_47_0 | 48 | 0.7783 | 1 | 0.7783 | 0 | 0 | 0 | 1 | Ref |
| 4602 | LC616037 | Unassigned | Unassigned | Unassigned | 47_0 | Clustered | 48 | VC_47_0 | 48 | 0.7783 | 1 | 0.7783 | 0 | 0 | 0 | 1 | Ref |
| 4603 | LC616038 | Unassigned | Unassigned | Unassigned | 47_0 | Clustered | 48 | VC_47_0 | 48 | 0.7783 | 1 | 0.7783 | 0 | 0 | 0 | 1 | Ref |
| 4604 | LC616039 | Unassigned | Unassigned | Unassigned | 47_0 | Clustered | 48 | VC_47_0 | 48 | 0.7783 | 1 | 0.7783 | 0 | 0 | 0 | 1 | Ref |
| 4605 | LC616040 | Unassigned | Unassigned | Unassigned | 47_0 | Clustered | 48 | VC_47_0 | 48 | 0.7783 | 1 | 0.7783 | 0 | 0 | 0 | 1 | Ref |
| 4606 | LC616041 | Unassigned | Unassigned | Unassigned | 47_0 | Clustered | 48 | VC_47_0 | 48 | 0.7783 | 1 | 0.7783 | 0 | 0 | 0 | 1 | Ref |
| 4607 | LC616042 | Unassigned | Unassigned | Unassigned | 47_0 | Clustered | 48 | VC_47_0 | 48 | 0.7783 | 1 | 0.7783 | 0 | 0 | 0 | 1 | Ref |
| 4608 | LC616043 | Unassigned | Unassigned | Unassigned | 47_0 | Clustered | 48 | VC_47_0 | 48 | 0.7783 | 1 | 0.7783 | 0 | 0 | 0 | 1 | Ref |
| 4609 | LC616044 | Unassigned | Unassigned | Unassigned | 47_0 | Clustered | 48 | VC_47_0 | 48 | 0.7783 | 1 | 0.7783 | 0 | 0 | 0 | 1 | Ref |
| 4610 | LC616045 | Unassigned | Unassigned | Unassigned | 47_0 | Clustered | 48 | VC_47_0 | 48 | 0.7783 | 1 | 0.7783 | 0 | 0 | 0 | 1 | Ref |
| 4611 | LC616046 | Unassigned | Unassigned | Unassigned | 47_0 | Clustered | 48 | VC_47_0 | 48 | 0.7783 | 1 | 0.7783 | 0 | 0 | 0 | 1 | Ref |
| 4612 | LC616047 | Unassigned | Unassigned | Unassigned | 47_0 | Clustered | 48 | VC_47_0 | 48 | 0.7783 | 1 | 0.7783 | 0 | 0 | 0 | 1 | Ref |
| 4613 | LC616048 | Unassigned | Unassigned | Unassigned | 47_0 | Clustered | 48 | VC_47_0 | 48 | 0.7783 | 1 | 0.7783 | 0 | 0 | 0 | 1 | Ref |
| 4614 | LC616049 | Unassigned | Unassigned | Unassigned | 47_0 | Clustered | 48 | VC_47_0 | 48 | 0.7783 | 1 | 0.7783 | 0 | 0 | 0 | 1 | Ref |
| 4615 | LC616050 | Unassigned | Unassigned | Unassigned | 47_0 | Clustered | 48 | VC_47_0 | 48 | 0.7783 | 1 | 0.7783 | 0 | 0 | 0 | 1 | Ref |
| 4616 | LC616051 | Unassigned | Unassigned | Unassigned | 47_0 | Clustered | 48 | VC_47_0 | 48 | 0.7783 | 1 | 0.7783 | 0 | 0 | 0 | 1 | Ref |
| 4617 | LC616052 | Unassigned | Unassigned | Unassigned | 47_0 | Clustered | 48 | VC_47_0 | 48 | 0.7783 | 1 | 0.7783 | 0 | 0 | 0 | 1 | Ref |
| 4618 | LC616053 | Unassigned | Unassigned | Unassigned | 47_0 | Clustered | 48 | VC_47_0 | 48 | 0.7783 | 1 | 0.7783 | 0 | 0 | 0 | 1 | Ref |
| 4619 | LC616054 | Unassigned | Unassigned | Unassigned | 47_0 | Clustered | 48 | VC_47_0 | 48 | 0.7783 | 1 | 0.7783 | 0 | 0 | 0 | 1 | Ref |
| 4620 | LC616055 | Unassigned | Unassigned | Unassigned | 47_0 | Clustered | 48 | VC_47_0 | 48 | 0.7783 | 1 | 0.7783 | 0 | 0 | 0 | 1 | Ref |
| 4621 | LC616056 | Unassigned | Unassigned | Unassigned | 47_0 | Clustered | 48 | VC_47_0 | 48 | 0.7783 | 1 | 0.7783 | 0 | 0 | 0 | 1 | Ref |
| 4622 | LC616057 | Unassigned | Unassigned | Unassigned | 47_0 | Clustered | 48 | VC_47_0 | 48 | 0.7783 | 1 | 0.7783 | 0 | 0 | 0 | 1 | Ref |
| 4623 | LC616058 | Unassigned | Unassigned | Unassigned | 47_0 | Clustered | 48 | VC_47_0 | 48 | 0.7783 | 1 | 0.7783 | 0 | 0 | 0 | 1 | Ref |
| 4624 | LC616059 | Unassigned | Unassigned | Unassigned | 47_0 | Clustered | 48 | VC_47_0 | 48 | 0.7783 | 1 | 0.7783 | 0 | 0 | 0 | 1 | Ref |
| 4625 | LC616060 | Unassigned | Unassigned | Unassigned | 47_0 | Clustered | 48 | VC_47_0 | 48 | 0.7783 | 1 | 0.7783 | 0 | 0 | 0 | 1 | Ref |
| 4626 | LC616061 | Unassigned | Unassigned | Unassigned | 47_0 | Clustered | 48 | VC_47_0 | 48 | 0.7783 | 1 | 0.7783 | 0 | 0 | 0 | 1 | Ref |
| 4627 | LC616062 | Unassigned | Unassigned | Unassigned | 47_0 | Clustered | 48 | VC_47_0 | 48 | 0.7783 | 1 | 0.7783 | 0 | 0 | 0 | 1 | Ref |
| 4628 | LC616063 | Unassigned | Unassigned | Unassigned | 47_0 | Clustered | 48 | VC_47_0 | 48 | 0.7783 | 1 | 0.7783 | 0 | 0 | 0 | 1 | Ref |
| 4629 | LC616064 | Unassigned | Unassigned | Unassigned | 47_0 | Clustered | 48 | VC_47_0 | 48 | 0.7783 | 1 | 0.7783 | 0 | 0 | 0 | 1 | Ref |
| 4630 | LC616065 | Unassigned | Unassigned | Unassigned | 47_0 | Clustered | 48 | VC_47_0 | 48 | 0.7783 | 1 | 0.7783 | 0 | 0 | 0 | 1 | Ref |
| 4631 | LC616066 | Unassigned | Unassigned | Unassigned | 47_0 | Clustered | 48 | VC_47_0 | 48 | 0.7783 | 1 | 0.7783 | 0 | 0 | 0 | 1 | Ref |
| 4632 | LC616067 | Unassigned | Unassigned | Unassigned | 47_0 | Clustered | 48 | VC_47_0 | 48 | 0.7783 | 1 | 0.7783 | 0 | 0 | 0 | 1 | Ref |
| 4633 | LC616068 | Unassigned | Unassigned | Unassigned | 47_0 | Clustered | 48 | VC_47_0 | 48 | 0.7783 | 1 | 0.7783 | 0 | 0 | 0 | 1 | Ref |
| 4634 | LC616069 | Unassigned | Unassigned | Unassigned | 47_0 | Clustered | 48 | VC_47_0 | 48 | 0.7783 | 1 | 0.7783 | 0 | 0 | 0 | 1 | Ref |
| 4635 | LC616070 | Unassigned | Unassigned | Unassigned | 47_0 | Clustered | 48 | VC_47_0 | 48 | 0.7783 | 1 | 0.7783 | 0 | 0 | 0 | 1 | Ref |
| 4636 | LC616071 | Unassigned | Unassigned | Unassigned | 47_0 | Clustered | 48 | VC_47_0 | 48 | 0.7783 | 1 | 0.7783 | 0 | 0 | 0 | 1 | Ref |
| 4637 | LC616072 | Unassigned | Unassigned | Unassigned | 47_0 | Clustered | 48 | VC_47_0 | 48 | 0.7783 | 1 | 0.7783 | 0 | 0 | 0 | 1 | Ref |
| 4638 | LC616073 | Unassigned | Unassigned | Unassigned | 47_0 | Clustered | 48 | VC_47_0 | 48 | 0.7783 | 1 | 0.7783 | 0 | 0 | 0 | 1 | Ref |
| 4639 | LC616074 | Unassigned | Unassigned | Unassigned | 47_0 | Clustered | 48 | VC_47_0 | 48 | 0.7783 | 1 | 0.7783 | 0 | 0 | 0 | 1 | Ref |
| 4640 | LC616075 | Unassigned | Unassigned | Unassigned | 47_0 | Clustered | 48 | VC_47_0 | 48 | 0.7783 | 1 | 0.7783 | 0 | 0 | 0 | 1 | Ref |
| 4641 | LC616076 | Unassigned | Unassigned | Unassigned | 47_0 | Clustered | 48 | VC_47_0 | 48 | 0.7783 | 1 | 0.7783 | 0 | 0 | 0 | 1 | Ref |
| 4642 | LC616077 | Unassigned | Unassigned | Unassigned | 47_0 | Clustered | 48 | VC_47_0 | 48 | 0.7783 | 1 | 0.7783 | 0 | 0 | 0 | 1 | Ref |
| 4643 | LC616078 | Unassigned | Unassigned | Unassigned | 47_0 | Clustered | 48 | VC_47_0 | 48 | 0.7783 | 1 | 0.7783 | 0 | 0 | 0 | 1 | Ref |
| 4644 | LC616079 | Unassigned | Unassigned | Unassigned | 40_0 | Clustered | 5 | VC_40_0 | 5 | 0.0844 | 0.99999995 | 0.0844 | 0 | 0 | 0 | 1 | Ref |
| 4645 | LC620505 | Unassigned | Unassigned | Unassigned | 283_5 | Clustered/Singleton | 1 | VC_283_5 | 1 | 0 | 1 | 0 | 0 | 0 | 0 | 1 | Ref |
| 4646 | LC623721 | Unassigned | Unassigned | Unassigned | 695_0 | Clustered | 48 | VC_695_0 | 48 | 0.8162 | 1 | 0.8162 | 0 | 0 | 0 | 1 | Ref |
| 4647 | LC625742 | Unassigned | Unassigned | Unassigned | 377_0 | Clustered | 3 | VC_377_0 | 3 | 0.3906 | 0.99785056 | 0.3898 | 0 | 0 | 0 | 1 | Ref |
| 4648 | LC628082 | Unassigned | Unassigned | Unassigned | 97_0 | Clustered | 5 | VC_97_0 | 5 | 0.0696 | 0.98113835 | 0.0683 | 0 | 0 | 0 | 1 | Ref |
| 4649 | LC644557 | Unassigned | Unassigned | Unassigned | 97_0 | Clustered | 5 | VC_97_0 | 5 | 0.0696 | 0.98113835 | 0.0683 | 0 | 0 | 0 | 1 | Ref |
| 4650 | LC644972 | Unassigned | Unassigned | Unassigned | 411_0 | Clustered | 3 | VC_411_0 | 3 | 0.1239 | 0.98986341 | 0.1227 | 0 | 0 | 0 | 1 | Ref |
| 4651 | LC644974 | Unassigned | Unassigned | Unassigned | 411_0 | Clustered | 3 | VC_411_0 | 3 | 0.1239 | 0.98986341 | 0.1227 | 0 | 0 | 0 | 1 | Ref |
| 4652 | LC644975 | Unassigned | Unassigned | Unassigned | 1204_0 | Clustered | 2 | VC_1204_0 | 2 | 0.883 | 0.9197097 | 0.8121 | 0 | 0 | 0 | 1 | Ref |
| 4653 | LC644976 | Unassigned | Unassigned | Unassigned | 1204_0 | Clustered | 2 | VC_1204_0 | 2 | 0.883 | 0.9197097 | 0.8121 | 0 | 0 | 0 | 1 | Ref |
| 4654 | LC645408 | Unassigned | Unassigned | Unassigned | 41_1 | Clustered | 10 | VC_41_1 | 10 | 0.1178 | 1 | 0.1178 | 0 | 0 | 0 | 0.8222 | Ref |
| 4655 | LC645409 | Unassigned | Unassigned | Unassigned | 42_0 | Clustered | 49 | VC_42_0 | 49 | 0.4517 | 1 | 0.4517 | 0 | 0 | 0 | 0.8597 | Ref |
| 4656 | LC645410 | Unassigned | Unassigned | Unassigned | 42_0 | Clustered | 49 | VC_42_0 | 49 | 0.4517 | 1 | 0.4517 | 0 | 0 | 0 | 0.8597 | Ref |
| 4657 | LC645411 | Unassigned | Unassigned | Unassigned | 42_0 | Clustered | 49 | VC_42_0 | 49 | 0.4517 | 1 | 0.4517 | 0 | 0 | 0 | 0.8597 | Ref |
| 4658 | LC645412 | Unassigned | Unassigned | Unassigned | 42_0 | Clustered | 49 | VC_42_0 | 49 | 0.4517 | 1 | 0.4517 | 0 | 0 | 0 | 0.8597 | Ref |
| 4659 | LC645413 | Unassigned | Unassigned | Unassigned | 42_0 | Clustered | 49 | VC_42_0 | 49 | 0.4517 | 1 | 0.4517 | 0 | 0 | 0 | 0.8597 | Ref |
| 4660 | LC645414 | Unassigned | Unassigned | Unassigned | 42_0 | Clustered | 49 | VC_42_0 | 49 | 0.4517 | 1 | 0.4517 | 0 | 0 | 0 | 0.8597 | Ref |
| 4661 | LC645415 | Unassigned | Unassigned | Unassigned | 42_0 | Clustered | 49 | VC_42_0 | 49 | 0.4517 | 1 | 0.4517 | 0 | 0 | 0 | 0.8597 | Ref |
| 4662 | LC645416 | Unassigned | Unassigned | Unassigned | 42_0 | Clustered | 49 | VC_42_0 | 49 | 0.4517 | 1 | 0.4517 | 0 | 0 | 0 | 0.8597 | Ref |
| 4663 | LC645417 | Unassigned | Unassigned | Unassigned | 42_0 | Clustered | 49 | VC_42_0 | 49 | 0.4517 | 1 | 0.4517 | 0 | 0 | 0 | 0.8597 | Ref |
| 4664 | LC645418 | Unassigned | Unassigned | Unassigned | 42_0 | Clustered | 49 | VC_42_0 | 49 | 0.4517 | 1 | 0.4517 | 0 | 0 | 0 | 0.8597 | Ref |
| 4665 | LC645419 | Unassigned | Unassigned | Unassigned | 42_0 | Clustered | 49 | VC_42_0 | 49 | 0.4517 | 1 | 0.4517 | 0 | 0 | 0 | 0.8597 | Ref |
| 4666 | LC645420 | Unassigned | Unassigned | Unassigned | 42_0 | Clustered | 49 | VC_42_0 | 49 | 0.4517 | 1 | 0.4517 | 0 | 0 | 0 | 0.8597 | Ref |
| 4667 | LC645421 | Unassigned | Unassigned | Unassigned | 42_0 | Clustered | 49 | VC_42_0 | 49 | 0.4517 | 1 | 0.4517 | 0 | 0 | 0 | 0.8597 | Ref |
| 4668 | LC645422 | Unassigned | Unassigned | Unassigned | 42_0 | Clustered | 49 | VC_42_0 | 49 | 0.4517 | 1 | 0.4517 | 0 | 0 | 0 | 0.8597 | Ref |
| 4669 | LC645423 | Unassigned | Unassigned | Unassigned | 42_0 | Clustered | 49 | VC_42_0 | 49 | 0.4517 | 1 | 0.4517 | 0 | 0 | 0 | 0.8597 | Ref |
| 4670 | LC645424 | Unassigned | Unassigned | Unassigned | 42_0 | Clustered | 49 | VC_42_0 | 49 | 0.4517 | 1 | 0.4517 | 0 | 0 | 0 | 0.8597 | Ref |
| 4671 | LC645425 | Unassigned | Unassigned | Unassigned | 41_1 | Clustered | 10 | VC_41_1 | 10 | 0.1178 | 1 | 0.1178 | 0 | 0 | 0 | 0.8222 | Ref |
| 4672 | LC645426 | Unassigned | Unassigned | Unassigned | 41_1 | Clustered | 10 | VC_41_1 | 10 | 0.1178 | 1 | 0.1178 | 0 | 0 | 0 | 0.8222 | Ref |
| 4673 | LC645427 | Unassigned | Unassigned | Unassigned | 42_0 | Clustered | 49 | VC_42_0 | 49 | 0.4517 | 1 | 0.4517 | 0 | 0 | 0 | 0.8597 | Ref |
| 4674 | LC645428 | Unassigned | Unassigned | Unassigned | 48_0 | Clustered | 10 | VC_48_0 | 10 | 0.174 | 1 | 0.174 | 0 | 0 | 0 | 1 | Ref |
| 4675 | LC645429 | Unassigned | Unassigned | Unassigned | 48_0 | Clustered | 10 | VC_48_0 | 10 | 0.174 | 1 | 0.174 | 0 | 0 | 0 | 1 | Ref |
| 4676 | LC645430 | Unassigned | Unassigned | Unassigned | 48_0 | Clustered | 10 | VC_48_0 | 10 | 0.174 | 1 | 0.174 | 0 | 0 | 0 | 1 | Ref |
| 4677 | LC645431 | Unassigned | Unassigned | Unassigned | 48_0 | Clustered | 10 | VC_48_0 | 10 | 0.174 | 1 | 0.174 | 0 | 0 | 0 | 1 | Ref |
| 4678 | LC645432 | Unassigned | Unassigned | Unassigned | 48_0 | Clustered | 10 | VC_48_0 | 10 | 0.174 | 1 | 0.174 | 0 | 0 | 0 | 1 | Ref |
| 4679 | LC645433 | Unassigned | Unassigned | Unassigned | 48_0 | Clustered | 10 | VC_48_0 | 10 | 0.174 | 1 | 0.174 | 0 | 0 | 0 | 1 | Ref |
| 4680 | LC645434 | Unassigned | Unassigned | Unassigned | 48_0 | Clustered | 10 | VC_48_0 | 10 | 0.174 | 1 | 0.174 | 0 | 0 | 0 | 1 | Ref |
| 4681 | LC645435 | Unassigned | Unassigned | Unassigned | 48_0 | Clustered | 10 | VC_48_0 | 10 | 0.174 | 1 | 0.174 | 0 | 0 | 0 | 1 | Ref |
| 4682 | LC645436 | Unassigned | Unassigned | Unassigned | 48_0 | Clustered | 10 | VC_48_0 | 10 | 0.174 | 1 | 0.174 | 0 | 0 | 0 | 1 | Ref |
| 4683 | LC645437 | Unassigned | Unassigned | Unassigned | 48_0 | Clustered | 10 | VC_48_0 | 10 | 0.174 | 1 | 0.174 | 0 | 0 | 0 | 1 | Ref |
| 4684 | LC645438 | Unassigned | Unassigned | Unassigned | 42_0 | Clustered | 49 | VC_42_0 | 49 | 0.4517 | 1 | 0.4517 | 0 | 0 | 0 | 0.8597 | Ref |
| 4685 | LC645442 | Unassigned | Unassigned | Unassigned | 42_0 | Clustered | 49 | VC_42_0 | 49 | 0.4517 | 1 | 0.4517 | 0 | 0 | 0 | 0.8597 | Ref |
| 4686 | LC645443 | Unassigned | Unassigned | Unassigned | 42_0 | Clustered | 49 | VC_42_0 | 49 | 0.4517 | 1 | 0.4517 | 0 | 0 | 0 | 0.8597 | Ref |
| 4687 | LC645444 | Unassigned | Unassigned | Unassigned | 42_0 | Clustered | 49 | VC_42_0 | 49 | 0.4517 | 1 | 0.4517 | 0 | 0 | 0 | 0.8597 | Ref |
| 4688 | LC645445 | Unassigned | Unassigned | Unassigned | 42_0 | Clustered | 49 | VC_42_0 | 49 | 0.4517 | 1 | 0.4517 | 0 | 0 | 0 | 0.8597 | Ref |
| 4689 | LC645446 | Unassigned | Unassigned | Unassigned | 42_0 | Clustered | 49 | VC_42_0 | 49 | 0.4517 | 1 | 0.4517 | 0 | 0 | 0 | 0.8597 | Ref |
| 4690 | LC645447 | Unassigned | Unassigned | Unassigned | 42_0 | Clustered | 49 | VC_42_0 | 49 | 0.4517 | 1 | 0.4517 | 0 | 0 | 0 | 0.8597 | Ref |
| 4691 | LC645448 | Unassigned | Unassigned | Unassigned | 42_0 | Clustered | 49 | VC_42_0 | 49 | 0.4517 | 1 | 0.4517 | 0 | 0 | 0 | 0.8597 | Ref |
| 4692 | LC645449 | Unassigned | Unassigned | Unassigned | 42_0 | Clustered | 49 | VC_42_0 | 49 | 0.4517 | 1 | 0.4517 | 0 | 0 | 0 | 0.8597 | Ref |
| 4693 | LC645450 | Unassigned | Unassigned | Unassigned | 42_0 | Clustered | 49 | VC_42_0 | 49 | 0.4517 | 1 | 0.4517 | 0 | 0 | 0 | 0.8597 | Ref |
| 4694 | LC645451 | Unassigned | Unassigned | Unassigned | 42_0 | Clustered | 49 | VC_42_0 | 49 | 0.4517 | 1 | 0.4517 | 0 | 0 | 0 | 0.8597 | Ref |
| 4695 | LC647030 | Unassigned | Unassigned | Unassigned | 152_0 | Clustered/Singleton | 1 | VC_152_0 | 1 | 0 | 1 | 0 | 0 | 0 | 0 | 1 | Ref |
| 4696 | LC648442 | Unassigned | Unassigned | Unassigned | 149_7 | Clustered/Singleton | 1 | VC_149_7 | 1 | 0 | 1 | 0 | 0 | 0 | 0 | 1 | Ref |

| | | | | | | | | | | | | | | | | | |
|---|---|---|---|---|---|---|---|---|---|---|---|---|---|---|---|---|---|
| 4697 | LC648443 | Unassigned | Unassigned | Unassigned | 472_0 | Clustered | 47 | VC_472_0 | 47 | 0.9025 | 1 | 0.9025 | 0 | 0 | 0 | 1 | Ref |
| 4698 | LC659915 | Unassigned | Unassigned | Unassigned | 191_19 | Clustered | 12 | VC_191_19 | 12 | 0.0655 | 1 | 0.0655 | 0 | 0 | 0 | 0.8636 | Ref |
| 4699 | LC667452 | Unassigned | Unassigned | Unassigned | 3_0 | Clustered | 17 | VC_3_0 | 17 | 0.5639 | 1 | 0.5639 | 0 | 0 | 0 | 1 | Ref |
| 4700 | LC680884 | Unassigned | Unassigned | Unassigned | 448_0 | Clustered | 4 | VC_448_0 | 4 | 0.3059 | 0.99978853 | 0.3058 | 0 | 0 | 0 | 1 | Ref |
| 4701 | LC680885 | Unassigned | Unassigned | Unassigned | 449_0 | Clustered | 7 | VC_449_0 | 7 | 0.6015 | 1 | 0.6015 | 0 | 0 | 0 | 1 | Ref |
| 4702 | LC709172 | Unassigned | Unassigned | Unassigned | 96_0 | Clustered | 30 | VC_96_0 | 30 | 0.718 | 1 | 0.718 | 0 | 0 | 0 | 1 | Ref |
| 4703 | LC727695 | Unassigned | Unassigned | Unassigned | 344_0 | Clustered | 21 | VC_344_0 | 21 | 0.5436 | 1 | 0.5436 | 0 | 0 | 0 | 1 | Ref |
| 4704 | LC727696 | Unassigned | Unassigned | Unassigned | 344_0 | Clustered | 21 | VC_344_0 | 21 | 0.5436 | 1 | 0.5436 | 0 | 0 | 0 | 1 | Ref |
| 4705 | LC727697 | Unassigned | Unassigned | Unassigned | 165_0 | Clustered | 4 | VC_165_0 | 4 | 0.2247 | 0.99998328 | 0.2247 | 0 | 0 | 0 | 1 | Ref |
| 4706 | LC727698 | Unassigned | Unassigned | Unassigned | 165_0 | Clustered | 4 | VC_165_0 | 4 | 0.2247 | 0.99998328 | 0.2247 | 0 | 0 | 0 | 1 | Ref |
| 4707 | LC727699 | Unassigned | Unassigned | Unassigned | 344_0 | Clustered | 21 | VC_344_0 | 21 | 0.5436 | 1 | 0.5436 | 0 | 0 | 0 | 1 | Ref |
| 4708 | LC727700 | Unassigned | Unassigned | Unassigned | 423_0 | Clustered | 45 | VC_423_0 | 45 | 0.9484 | 1 | 0.9484 | 0 | 0 | 0 | 1 | Ref |
| 4709 | LC727701 | Unassigned | Unassigned | Unassigned | 344_0 | Clustered | 21 | VC_344_0 | 21 | 0.5436 | 1 | 0.5436 | 0 | 0 | 0 | 1 | Ref |
| 4710 | LC727840 | Unassigned | Unassigned | Unassigned | 49_0 | Clustered | 36 | VC_49_0 | 36 | 0.611 | 1 | 0.611 | 0 | 0 | 0 | 1 | Ref |
| 4711 | LC727841 | Unassigned | Unassigned | Unassigned | 49_0 | Clustered | 36 | VC_49_0 | 36 | 0.611 | 1 | 0.611 | 0 | 0 | 0 | 1 | Ref |
| 4712 | LC727842 | Unassigned | Unassigned | Unassigned | 49_0 | Clustered | 36 | VC_49_0 | 36 | 0.611 | 1 | 0.611 | 0 | 0 | 0 | 1 | Ref |
| 4713 | LC727843 | Unassigned | Unassigned | Unassigned | 49_0 | Clustered | 36 | VC_49_0 | 36 | 0.611 | 1 | 0.611 | 0 | 0 | 0 | 1 | Ref |
| 4714 | LC727844 | Unassigned | Unassigned | Unassigned | 49_0 | Clustered | 36 | VC_49_0 | 36 | 0.611 | 1 | 0.611 | 0 | 0 | 0 | 1 | Ref |
| 4715 | LC727845 | Unassigned | Unassigned | Unassigned | 49_0 | Clustered | 36 | VC_49_0 | 36 | 0.611 | 1 | 0.611 | 0 | 0 | 0 | 1 | Ref |
| 4716 | LC727846 | Unassigned | Unassigned | Unassigned | 49_0 | Clustered | 36 | VC_49_0 | 36 | 0.611 | 1 | 0.611 | 0 | 0 | 0 | 1 | Ref |
| 4717 | LC727847 | Unassigned | Unassigned | Unassigned | 49_0 | Clustered | 36 | VC_49_0 | 36 | 0.611 | 1 | 0.611 | 0 | 0 | 0 | 1 | Ref |
| 4718 | LC727848 | Unassigned | Unassigned | Unassigned | 49_0 | Clustered | 36 | VC_49_0 | 36 | 0.611 | 1 | 0.611 | 0 | 0 | 0 | 1 | Ref |
| 4719 | LC727849 | Unassigned | Unassigned | Unassigned | 49_0 | Clustered | 36 | VC_49_0 | 36 | 0.611 | 1 | 0.611 | 0 | 0 | 0 | 1 | Ref |
| 4720 | LC727850 | Unassigned | Unassigned | Unassigned | 49_0 | Clustered | 36 | VC_49_0 | 36 | 0.611 | 1 | 0.611 | 0 | 0 | 0 | 1 | Ref |
| 4721 | LC727851 | Unassigned | Unassigned | Unassigned | 49_0 | Clustered | 36 | VC_49_0 | 36 | 0.611 | 1 | 0.611 | 0 | 0 | 0 | 1 | Ref |
| 4722 | LC727852 | Unassigned | Unassigned | Unassigned | 49_0 | Clustered | 36 | VC_49_0 | 36 | 0.611 | 1 | 0.611 | 0 | 0 | 0 | 1 | Ref |
| 4723 | LC727853 | Unassigned | Unassigned | Unassigned | 49_0 | Clustered | 36 | VC_49_0 | 36 | 0.611 | 1 | 0.611 | 0 | 0 | 0 | 1 | Ref |
| 4724 | LC727854 | Unassigned | Unassigned | Unassigned | 49_0 | Clustered | 36 | VC_49_0 | 36 | 0.611 | 1 | 0.611 | 0 | 0 | 0 | 1 | Ref |
| 4725 | LC727855 | Unassigned | Unassigned | Unassigned | 49_0 | Clustered | 36 | VC_49_0 | 36 | 0.611 | 1 | 0.611 | 0 | 0 | 0 | 1 | Ref |
| 4726 | LC727856 | Unassigned | Unassigned | Unassigned | 49_0 | Clustered | 36 | VC_49_0 | 36 | 0.611 | 1 | 0.611 | 0 | 0 | 0 | 1 | Ref |
| 4727 | LC727857 | Unassigned | Unassigned | Unassigned | 49_0 | Clustered | 36 | VC_49_0 | 36 | 0.611 | 1 | 0.611 | 0 | 0 | 0 | 1 | Ref |
| 4728 | LC727858 | Unassigned | Unassigned | Unassigned | 49_0 | Clustered | 36 | VC_49_0 | 36 | 0.611 | 1 | 0.611 | 0 | 0 | 0 | 1 | Ref |
| 4729 | LC727859 | Unassigned | Unassigned | Unassigned | 49_0 | Clustered | 36 | VC_49_0 | 36 | 0.611 | 1 | 0.611 | 0 | 0 | 0 | 1 | Ref |
| 4730 | LC727860 | Unassigned | Unassigned | Unassigned | 49_0 | Clustered | 36 | VC_49_0 | 36 | 0.611 | 1 | 0.611 | 0 | 0 | 0 | 1 | Ref |
| 4731 | LC727861 | Unassigned | Unassigned | Unassigned | 49_0 | Clustered | 36 | VC_49_0 | 36 | 0.611 | 1 | 0.611 | 0 | 0 | 0 | 1 | Ref |
| 4732 | LC727862 | Unassigned | Unassigned | Unassigned | 49_0 | Clustered | 36 | VC_49_0 | 36 | 0.611 | 1 | 0.611 | 0 | 0 | 0 | 1 | Ref |
| 4733 | LC727863 | Unassigned | Unassigned | Unassigned | 49_0 | Clustered | 36 | VC_49_0 | 36 | 0.611 | 1 | 0.611 | 0 | 0 | 0 | 1 | Ref |
| 4734 | LC727864 | Unassigned | Unassigned | Unassigned | 49_0 | Clustered | 36 | VC_49_0 | 36 | 0.611 | 1 | 0.611 | 0 | 0 | 0 | 1 | Ref |
| 4735 | LC727865 | Unassigned | Unassigned | Unassigned | 49_0 | Clustered | 36 | VC_49_0 | 36 | 0.611 | 1 | 0.611 | 0 | 0 | 0 | 1 | Ref |
| 4736 | LC727866 | Unassigned | Unassigned | Unassigned | 49_0 | Clustered | 36 | VC_49_0 | 36 | 0.611 | 1 | 0.611 | 0 | 0 | 0 | 1 | Ref |
| 4737 | LC727867 | Unassigned | Unassigned | Unassigned | 49_0 | Clustered | 36 | VC_49_0 | 36 | 0.611 | 1 | 0.611 | 0 | 0 | 0 | 1 | Ref |
| 4738 | LC727868 | Unassigned | Unassigned | Unassigned | 49_0 | Clustered | 36 | VC_49_0 | 36 | 0.611 | 1 | 0.611 | 0 | 0 | 0 | 1 | Ref |
| 4739 | LC727869 | Unassigned | Unassigned | Unassigned | 49_0 | Clustered | 36 | VC_49_0 | 36 | 0.611 | 1 | 0.611 | 0 | 0 | 0 | 1 | Ref |
| 4740 | LC727870 | Unassigned | Unassigned | Unassigned | 49_0 | Clustered | 36 | VC_49_0 | 36 | 0.611 | 1 | 0.611 | 0 | 0 | 0 | 1 | Ref |
| 4741 | LC727871 | Unassigned | Unassigned | Unassigned | 49_0 | Clustered | 36 | VC_49_0 | 36 | 0.611 | 1 | 0.611 | 0 | 0 | 0 | 1 | Ref |
| 4742 | LC727872 | Unassigned | Unassigned | Unassigned | 49_0 | Clustered | 36 | VC_49_0 | 36 | 0.611 | 1 | 0.611 | 0 | 0 | 0 | 1 | Ref |
| 4743 | LC727873 | Unassigned | Unassigned | Unassigned | 49_0 | Clustered | 36 | VC_49_0 | 36 | 0.611 | 1 | 0.611 | 0 | 0 | 0 | 1 | Ref |
| 4744 | LC727874 | Unassigned | Unassigned | Unassigned | 49_0 | Clustered | 36 | VC_49_0 | 36 | 0.611 | 1 | 0.611 | 0 | 0 | 0 | 1 | Ref |
| 4745 | LC727875 | Unassigned | Unassigned | Unassigned | 49_0 | Clustered | 36 | VC_49_0 | 36 | 0.611 | 1 | 0.611 | 0 | 0 | 0 | 1 | Ref |
| 4746 | LC727891 | Unassigned | Unassigned | Unassigned | 41_0 | Clustered | 16 | VC_41_0 | 16 | 0.2025 | 1 | 0.2025 | 0 | 0 | 0 | 0.9667 | Ref |
| 4747 | LC727892 | Unassigned | Unassigned | Unassigned | 41_0 | Clustered | 16 | VC_41_0 | 16 | 0.2025 | 1 | 0.2025 | 0 | 0 | 0 | 0.9667 | Ref |
| 4748 | LC727893 | Unassigned | Unassigned | Unassigned | 41_0 | Clustered | 16 | VC_41_0 | 16 | 0.2025 | 1 | 0.2025 | 0 | 0 | 0 | 0.9667 | Ref |
| 4749 | LC727894 | Unassigned | Unassigned | Unassigned | 41_0 | Clustered | 16 | VC_41_0 | 16 | 0.2025 | 1 | 0.2025 | 0 | 0 | 0 | 0.9667 | Ref |
| 4750 | LC727895 | Unassigned | Unassigned | Unassigned | 41_0 | Clustered | 16 | VC_41_0 | 16 | 0.2025 | 1 | 0.2025 | 0 | 0 | 0 | 0.9667 | Ref |
| 4751 | LC727896 | Unassigned | Unassigned | Unassigned | 41_0 | Clustered | 16 | VC_41_0 | 16 | 0.2025 | 1 | 0.2025 | 0 | 0 | 0 | 0.9667 | Ref |
| 4752 | LC727897 | Unassigned | Unassigned | Unassigned | 41_0 | Clustered | 16 | VC_41_0 | 16 | 0.2025 | 1 | 0.2025 | 0 | 0 | 0 | 0.9667 | Ref |
| 4753 | LC727898 | Unassigned | Unassigned | Unassigned | 41_0 | Clustered | 16 | VC_41_0 | 16 | 0.2025 | 1 | 0.2025 | 0 | 0 | 0 | 0.9667 | Ref |
| 4754 | LC727899 | Unassigned | Unassigned | Unassigned | 41_0 | Clustered | 16 | VC_41_0 | 16 | 0.2025 | 1 | 0.2025 | 0 | 0 | 0 | 0.9667 | Ref |
| 4755 | LC727900 | Unassigned | Unassigned | Unassigned | 41_0 | Clustered | 16 | VC_41_0 | 16 | 0.2025 | 1 | 0.2025 | 0 | 0 | 0 | 0.9667 | Ref |
| 4756 | LC727901 | Unassigned | Unassigned | Unassigned | 41_0 | Clustered | 16 | VC_41_0 | 16 | 0.2025 | 1 | 0.2025 | 0 | 0 | 0 | 0.9667 | Ref |
| 4757 | LC727902 | Unassigned | Unassigned | Unassigned | 41_0 | Clustered | 16 | VC_41_0 | 16 | 0.2025 | 1 | 0.2025 | 0 | 0 | 0 | 0.9667 | Ref |
| 4758 | LC727903 | Unassigned | Unassigned | Unassigned | 42_0 | Clustered | 49 | VC_42_0 | 49 | 0.4517 | 1 | 0.4517 | 0 | 0 | 0 | 0.8597 | Ref |
| 4759 | LC730317 | Unassigned | Unassigned | Unassigned | 513_0 | Clustered | 32 | VC_513_0 | 32 | 0.6881 | 1 | 0.6881 | 0 | 0 | 0 | 1 | Ref |
| 4760 | LC730318 | Unassigned | Unassigned | Unassigned | 215_1 | Clustered | 19 | VC_215_1 | 19 | 0.4188 | 1 | 0.4188 | 0 | 0 | 0 | 1 | Ref |
| 4761 | LC730319 | Unassigned | Unassigned | Unassigned | 129_0 | Clustered | 43 | VC_129_0 | 43 | 0.886 | 1 | 0.886 | 0 | 0 | 0 | 1 | Ref |
| 4762 | LC730320 | Unassigned | Unassigned | Unassigned | 216_6 | Clustered | 42 | VC_216_6 | 42 | 0.473 | 1 | 0.473 | 0 | 0 | 0 | 0.8351 | Ref |
| 4763 | LC730321 | Unassigned | Unassigned | Unassigned | 43_0 | Clustered | 38 | VC_43_0 | 38 | 0.5885 | 1 | 0.5885 | 0 | 0 | 0 | 0.9659 | Ref |
| 4764 | LC730322 | Unassigned | Unassigned | Unassigned | 28_1 | Clustered | 53 | VC_28_1 | 53 | 0.7164 | 1 | 0.7164 | 0 | 0 | 0 | 0.9325 | Ref |
| 4765 | LC730323 | Unassigned | Unassigned | Unassigned | 452_0 | Clustered | 66 | VC_452_0 | 66 | 0.9248 | 1 | 0.9248 | 0 | 0 | 0 | 1 | Ref |
| 4766 | LC730324 | Unassigned | Unassigned | Unassigned | 385_0 | Clustered | 51 | VC_385_0 | 51 | 0.8754 | 1 | 0.8754 | 0 | 0 | 0 | 1 | Ref |
| 4767 | LC730325 | Unassigned | Unassigned | Unassigned | 392_0 | Clustered | 126 | VC_392_0 | 126 | 0.9621 | 1 | 0.9621 | 0 | 0 | 0 | 1 | Ref |
| 4768 | LC739530 | Unassigned | Unassigned | Unassigned | 329_20 | Clustered | 4 | VC_329_20 | 4 | 0.0174 | 0.99998777 | 0.0174 | 0 | 0 | 0 | 1 | Ref |
| 4769 | LC739531 | Unassigned | Unassigned | Unassigned | 329_20 | Clustered | 4 | VC_329_20 | 4 | 0.0174 | 0.99998777 | 0.0174 | 0 | 0 | 0 | 1 | Ref |
| 4770 | LC739532 | Unassigned | Unassigned | Unassigned | 329_20 | Clustered | 4 | VC_329_20 | 4 | 0.0174 | 0.99998777 | 0.0174 | 0 | 0 | 0 | 1 | Ref |
| 4771 | LC739533 | Unassigned | Unassigned | Unassigned | 847_0 | Clustered | 5 | VC_847_0 | 5 | 0.0621 | 0.99921937 | 0.0621 | 0 | 0 | 0 | 1 | Ref |
| 4772 | LC739534 | Unassigned | Unassigned | Unassigned | 847_0 | Clustered | 5 | VC_847_0 | 5 | 0.0621 | 0.99921937 | 0.0621 | 0 | 0 | 0 | 1 | Ref |
| 4773 | LC739535 | Unassigned | Unassigned | Unassigned | 847_0 | Clustered | 5 | VC_847_0 | 5 | 0.0621 | 0.99921937 | 0.0621 | 0 | 0 | 0 | 1 | Ref |
| 4774 | LC739536 | Unassigned | Unassigned | Unassigned | 848_0 | Clustered | 28 | VC_848_0 | 28 | 0.7927 | 1 | 0.7927 | 0 | 0 | 0 | 1 | Ref |
| 4775 | LC739537 | Unassigned | Unassigned | Unassigned | 847_0 | Clustered | 5 | VC_847_0 | 5 | 0.0621 | 0.99921937 | 0.0621 | 0 | 0 | 0 | 1 | Ref |
| 4776 | LC739538 | Unassigned | Unassigned | Unassigned | 772_16 | Clustered | 10 | VC_772_16 | 10 | 0.0563 | 0.9434928 | 0.0531 | 0 | 0 | 0 | 0.8667 | Ref |
| 4777 | LC739539 | Unassigned | Unassigned | Unassigned | 847_0 | Clustered | 5 | VC_847_0 | 5 | 0.0621 | 0.99921937 | 0.0621 | 0 | 0 | 0 | 1 | Ref |
| 4778 | LC739540 | Unassigned | Unassigned | Unassigned | 772_16 | Clustered | 10 | VC_772_16 | 10 | 0.0563 | 0.9434928 | 0.0531 | 0 | 0 | 0 | 0.8667 | Ref |
| 4779 | LC739541 | Unassigned | Unassigned | Unassigned | 772_11 | Clustered | 6 | VC_772_11 | 6 | 0.0305 | 0.84253803 | 0.0257 | 0 | 0 | 0 | 0.8 | Ref |
| 4780 | LC739542 | Unassigned | Unassigned | Unassigned | 772_11 | Clustered | 6 | VC_772_11 | 6 | 0.0305 | 0.84253803 | 0.0257 | 0 | 0 | 0 | 0.8 | Ref |
| 4781 | LK392619 | Unassigned | Unassigned | Unassigned | 1048_0 | Clustered | 3 | VC_1048_0 | 3 | 1 | 1 | 1 | 0 | 0 | 0 | 1 | Ref |
| 4782 | LK985321 | Unassigned | Unassigned | Unassigned | 647_0 | Clustered | 11 | VC_647_0 | 11 | 0.598 | 1 | 0.598 | 0 | 0 | 0 | 1 | Ref |
| 4783 | LK985322 | Unassigned | Unassigned | Unassigned | 139_0 | Clustered | 2 | VC_139_0 | 2 | 0.0701 | 0.95510185 | 0.067 | 0 | 0 | 0 | 1 | Ref |
| 4784 | LN610572 | Unassigned | Unassigned | Unassigned | 76_0 | Clustered | 37 | VC_76_0 | 37 | 0.9403 | 1 | 0.9403 | 0 | 0 | 0 | 1 | Ref |
| 4785 | LN610573 | Unassigned | Unassigned | Unassigned | 77_0 | Clustered | 20 | VC_77_0 | 20 | 0.9657 | 1 | 0.9657 | 0 | 0 | 0 | 1 | Ref |
| 4786 | LN610574 | Unassigned | Unassigned | Unassigned | 423_0 | Clustered | 45 | VC_423_0 | 45 | 0.9484 | 1 | 0.9484 | 0 | 0 | 0 | 1 | Ref |
| 4787 | LN610575 | Unassigned | Unassigned | Unassigned | 76_0 | Clustered | 37 | VC_76_0 | 37 | 0.9403 | 1 | 0.9403 | 0 | 0 | 0 | 1 | Ref |
| 4788 | LN610576 | Unassigned | Unassigned | Unassigned | 77_0 | Clustered | 20 | VC_77_0 | 20 | 0.9657 | 1 | 0.9657 | 0 | 0 | 0 | 1 | Ref |
| 4789 | LN610577 | Unassigned | Unassigned | Unassigned | 464_0 | Clustered | 8 | VC_464_0 | 8 | 0.2629 | 1 | 0.2629 | 0 | 0 | 0 | 1 | Ref |
| 4790 | LN610578 | Unassigned | Unassigned | Unassigned | 472_0 | Clustered | 47 | VC_472_0 | 47 | 0.9025 | 1 | 0.9025 | 0 | 0 | 0 | 1 | Ref |
| 4791 | LN610579 | Unassigned | Unassigned | Unassigned | 79_0 | Clustered | 64 | VC_79_0 | 64 | 0.4185 | 1 | 0.4185 | 0 | 0 | 0 | 0.7649 | Ref |
| 4792 | LN610580 | Unassigned | Unassigned | Unassigned | 423_0 | Clustered | 45 | VC_423_0 | 45 | 0.9484 | 1 | 0.9484 | 0 | 0 | 0 | 1 | Ref |
| 4793 | LN610581 | Unassigned | Unassigned | Unassigned | 77_0 | Clustered | 20 | VC_77_0 | 20 | 0.9657 | 1 | 0.9657 | 0 | 0 | 0 | 1 | Ref |
| 4794 | LN610582 | Unassigned | Unassigned | Unassigned | 77_0 | Clustered | 20 | VC_77_0 | 20 | 0.9657 | 1 | 0.9657 | 0 | 0 | 0 | 1 | Ref |
| 4795 | LN610583 | Unassigned | Unassigned | Unassigned | 77_0 | Clustered | 20 | VC_77_0 | 20 | 0.9657 | 1 | 0.9657 | 0 | 0 | 0 | 1 | Ref |
| 4796 | LN610584 | Unassigned | Unassigned | Unassigned | 464_0 | Clustered | 8 | VC_464_0 | 8 | 0.2629 | 1 | 0.2629 | 0 | 0 | 0 | 1 | Ref |
| 4797 | LN610585 | Unassigned | Unassigned | Unassigned | 464_0 | Clustered | 8 | VC_464_0 | 8 | 0.2629 | 1 | 0.2629 | 0 | 0 | 0 | 1 | Ref |
| 4798 | LN610586 | Unassigned | Unassigned | Unassigned | 76_0 | Clustered | 37 | VC_76_0 | 37 | 0.9403 | 1 | 0.9403 | 0 | 0 | 0 | 1 | Ref |
| 4799 | LN610587 | Unassigned | Unassigned | Unassigned | 76_0 | Clustered | 37 | VC_76_0 | 37 | 0.9403 | 1 | 0.9403 | 0 | 0 | 0 | 1 | Ref |
| 4800 | LN610588 | Unassigned | Unassigned | Unassigned | 79_0 | Clustered | 64 | VC_79_0 | 64 | 0.4185 | 1 | 0.4185 | 0 | 0 | 0 | 0.7649 | Ref |
| 4801 | LN610589 | Unassigned | Unassigned | Unassigned | 80_0 | Clustered | 41 | VC_80_0 | 41 | 0.5837 | 1 | 0.5837 | 0 | 0 | 0 | 0.9805 | Ref |
| 4802 | LN610590 | Unassigned | Unassigned | Unassigned | 179_0 | Clustered | 53 | VC_179_0 | 53 | 0.9137 | 1 | 0.9137 | 0 | 0 | 0 | 1 | Ref |
| 4803 | LN681534 | Unassigned | Unassigned | Unassigned | 766_0 | Clustered | 2 | VC_766_0 | 2 | 0.2674 | 0.94916107 | 0.2538 | 0 | 0 | 0 | 1 | Ref |
| 4804 | LN681535 | Unassigned | Unassigned | Unassigned | 733_0 | Clustered | 5 | VC_733_0 | 5 | 0.5111 | 0.99999994 | 0.5111 | 0 | 0 | 0 | 1 | Ref |
| 4805 | LN681536 | Unassigned | Unassigned | Unassigned | 733_0 | Clustered | 5 | VC_733_0 | 5 | 0.5111 | 0.99999994 | 0.5111 | 0 | 0 | 0 | 1 | Ref |
| 4806 | LN681537 | Unassigned | Unassigned | Unassigned | 473_2 | Clustered | 2 | VC_473_2 | 2 | 0.1022 | 0.94602228 | 0.0966 | 0 | 0 | 0 | 1 | Ref |
| 4807 | LN681538 | Unassigned | Unassigned | Unassigned | 647_0 | Clustered | 11 | VC_647_0 | 11 | 0.598 | 1 | 0.598 | 0 | 0 | 0 | 1 | Ref |
| 4808 | LN681540 | Unassigned | Unassigned | Unassigned | 647_0 | Clustered | 11 | VC_647_0 | 11 | 0.598 | 1 | 0.598 | 0 | 0 | 0 | 1 | Ref |
| 4809 | LN681541 | Unassigned | Unassigned | Unassigned | 139_1 | Clustered | 4 | VC_139_1 | 4 | 0.1775 | 0.99998609 | 0.1775 | 0 | 0 | 0 | 1 | Ref |
| 4810 | LN681542 | Unassigned | Unassigned | Unassigned | 139_1 | Clustered | 4 | VC_139_1 | 4 | 0.1775 | 0.99998609 | 0.1775 | 0 | 0 | 0 | 1 | Ref |
| 4811 | LN828717 | Unassigned | Unassigned | Unassigned | 433_5 | Clustered | 2 | VC_433_5 | 2 | 0.0173 | 0.93642158 | 0.0162 | 0 | 0 | 0 | 1 | Ref |
| 4812 | LN866626 | Unassigned | Unassigned | Unassigned | 823_0 | Clustered | 124 | VC_823_0 | 124 | 0.9492 | 1 | 0.9492 | 0 | 0 | 0 | 1 | Ref |
| 4813 | LN878149 | Unassigned | Unassigned | Unassigned | 878_0 | Clustered | 32 | VC_878_0 | 32 | 0.9965 | 1 | 0.9965 | 0 | 0 | 0 | 1 | Ref |
| 4814 | LN881725 | Unassigned | Unassigned | Unassigned | 795_0 | Clustered | 19 | VC_795_0 | 19 | 0.6848 | 1 | 0.6848 | 0 | 0 | 0 | 1 | Ref |
| 4815 | LN881726 | Unassigned | Unassigned | Unassigned | 283_33 | Clustered | 4 | VC_283_33 | 4 | 0.0075 | 0.99979471 | 0.0075 | 0 | 0 | 0 | 1 | Ref |
| 4816 | LN881727 | Unassigned | Unassigned | Unassigned | 772_9 | Clustered | 7 | VC_772_9 | 7 | 0.0369 | 0.87705503 | 0.0324 | 0 | 0 | 0 | 1 | Ref |
| 4817 | LN881728 | Unassigned | Unassigned | Unassigned | 283_73 | Clustered | 3 | VC_283_73 | 3 | 0.0052 | 0.9974535 | 0.0051 | 0 | 0 | 0 | 1 | Ref |
| 4818 | LN881729 | Unassigned | Unassigned | Unassigned | 283_33 | Clustered | 4 | VC_283_33 | 4 | 0.0075 | 0.99979471 | 0.0075 | 0 | 0 | 0 | 1 | Ref |
| 4819 | LN881730 | Unassigned | Unassigned | Unassigned | 835_0 | Clustered | 67 | VC_835_0 | 67 | 0.9624 | 1 | 0.9624 | 0 | 0 | 0 | 1 | Ref |
| 4820 | LN881731 | Unassigned | Unassigned | Unassigned | 835_0 | Clustered | 67 | VC_835_0 | 67 | 0.9624 | 1 | 0.9624 | 0 | 0 | 0 | 1 | Ref |
| 4821 | LN881732 | Unassigned | Unassigned | Unassigned | 283_37 | Clustered/Singleton | 1 | VC_283_37 | 1 | 0 | 1 | 0 | 0 | 0 | 0 | 1 | Ref |
| 4822 | LN881733 | Unassigned | Unassigned | Unassigned | 283_73 | Clustered | 3 | VC_283_73 | 3 | 0.0052 | 0.9974535 | 0.0051 | 0 | 0 | 0 | 1 | Ref |
| 4823 | LN881734 | Unassigned | Unassigned | Unassigned | 283_33 | Clustered | 4 | VC_283_33 | 4 | 0.0075 | 0.99979471 | 0.0075 | 0 | 0 | 0 | 1 | Ref |
| 4824 | LN881735 | Unassigned | Unassigned | Unassigned | 772_9 | Clustered | 7 | VC_772_9 | 7 | 0.0369 | 0.87705503 | 0.0324 | 0 | 0 | 0 | 1 | Ref |
| 4825 | LN881736 | Unassigned | Unassigned | Unassigned | 283_73 | Clustered | 3 | VC_283_73 | 3 | 0.0052 | 0.9974535 | 0.0051 | 0 | 0 | 0 | 1 | Ref |
| 4826 | LN881737 | Unassigned | Unassigned | Unassigned | 283_33 | Clustered | 4 | VC_283_33 | 4 | 0.0075 | 0.99979471 | 0.0075 | 0 | 0 | 0 | 1 | Ref |
| 4827 | LN881738 | Unassigned | Unassigned | Unassigned | 733_0 | Clustered | 5 | VC_733_0 | 5 | 0.5111 | 0.99999994 | 0.5111 | 0 | 0 | 0 | 1 | Ref |
| 4828 | LN887844 | Unassigned | Unassigned | Unassigned | 78_5 | Clustered | 6 | VC_78_5 | 6 | 0.1087 | 0.99978082 | 0.1087 | 0 | 0 | 0 | 1 | Ref |
| 4829 | LN887948 | Unassigned | Unassigned | Unassigned | 190_20 | Clustered | 4 | VC_190_20 | 4 | 0.0229 | 0.99997885 | 0.0229 | 0 | 0 | 0 | 1 | Ref |
| 4830 | LN889756 | Unassigned | Unassigned | Unassigned | 129_0 | Clustered | 43 | VC_129_0 | 43 | 0.886 | 1 | 0.886 | 0 | 0 | 0 | 1 | Ref |
| 4831 | LN889995 | Unassigned | Unassigned | Unassigned | 129_0 | Clustered | 43 | VC_129_0 | 43 | 0.886 | 1 | 0.886 | 0 | 0 | 0 | 1 | Ref |
| 4832 | LN898172 | Unassigned | Unassigned | Unassigned | 251_0 | Clustered | 22 | VC_251_0 | 22 | 0.6508 | 1 | 0.6508 | 0 | 0 | 0 | 1 | Ref |
| 4833 | LN907801 | Unassigned | Unassigned | Unassigned | 179_0 | Clustered | 53 | VC_179_0 | 53 | 0.9137 | 1 | 0.9137 | 0 | 0 | 0 | 1 | Ref |
| 4834 | LN913017 | Unassigned | Unassigned | Unassigned | 129_0 | Clustered | 43 | VC_129_0 | 43 | 0.886 | 1 | 0.886 | 0 | 0 | 0 | 1 | Ref |
| 4835 | LN997803 | Unassigned | Unassigned | Unassigned | 245_0 | Clustered | 10 | VC_245_0 | 10 | 0.1805 | 1 | 0.1805 | 0 | 0 | 0 | 0.9778 | Ref |
| 4836 | LR025195 | Unassigned | Unassigned | Unassigned | 795_0 | Clustered | 19 | VC_795_0 | 19 | 0.6848 | 1 | 0.6848 | 0 | 0 | 0 | 1 | Ref |
| 4837 | LR025196 | Unassigned | Unassigned | Unassigned | 795_0 | Clustered | 19 | VC_795_0 | 19 | 0.6848 | 1 | 0.6848 | 0 | 0 | 0 | 1 | Ref |
| 4838 | LR025197 | Unassigned | Unassigned | Unassigned | 795_0 | Clustered | 19 | VC_795_0 | 19 | 0.6848 | 1 | 0.6848 | 0 | 0 | 0 | 1 | Ref |
| 4839 | LR025198 | Unassigned | Unassigned | Unassigned | 795_0 | Clustered | 19 | VC_795_0 | 19 | 0.6848 | 1 | 0.6848 | 0 | 0 | 0 | 1 | Ref |
| 4840 | LR026998 | Unassigned | Unassigned | Unassigned | 592_0 | Clustered | 50 | VC_592_0 | 50 | 0.7314 | 1 | 0.7314 | 0 | 0 | 0 | 1 | Ref |
| 4841 | LR027383 | Unassigned | Unassigned | Unassigned | 286_1 | Clustered | 21 | VC_286_1 | 21 | 0.1696 | 1 | 0.1696 | 0 | 0 | 0 | 0.9048 | Ref |
| 4842 | LR027384 | Unassigned | Unassigned | Unassigned | 190_12 | Clustered/Singleton | 1 | VC_190_12 | 1 | 0 | 1 | 0 | 0 | 0 | 0 | 1 | Ref |
| 4843 | LR027385 | Unassigned | Unassigned | Unassigned | 942_0 | Clustered | 36 | VC_942_0 | 36 | 0.7848 | 1 | 0.7848 | 0 | 0 | 0 | 1 | Ref |
| 4844 | LR027386 | Unassigned | Unassigned | Unassigned | 286_1 | Clustered | 21 | VC_286_1 | 21 | 0.1696 | 1 | 0.1696 | 0 | 0 | 0 | 0.9048 | Ref |
| 4845 | LR027387 | Unassigned | Unassigned | Unassigned | 286_1 | Clustered | 21 | VC_286_1 | 21 | 0.1696 | 1 | 0.1696 | 0 | 0 | 0 | 0.9048 | Ref |
| 4846 | LR027388 | Unassigned | Unassigned | Unassigned | 942_0 | Clustered | 36 | VC_942_0 | 36 | 0.7848 | 1 | 0.7848 | 0 | 0 | 0 | 1 | Ref |
| 4847 | LR027389 | Unassigned | Unassigned | Unassigned | 191_14 | Clustered/Singleton | 1 | VC_191_14 | 1 | 0 | 1 | 0 | 0 | 0 | 0 | 1 | Ref |
| 4848 | LR027390 | Unassigned | Unassigned | Unassigned | 286_1 | Clustered | 21 | VC_286_1 | 21 | 0.1696 | 1 | 0.1696 | 0 | 0 | 0 | 0.9048 | Ref |
| 4849 | LR027391 | Unassigned | Unassigned | Unassigned | 286_1 | Clustered | 21 | VC_286_1 | 21 | 0.1696 | 1 | 0.1696 | 0 | 0 | 0 | 0.9048 | Ref |
| 4850 | LR031359 | Unassigned | Unassigned | Unassigned | 91_0 | Clustered | 22 | VC_91_0 | 22 | 0.5318 | 1 | 0.5318 | 0 | 0 | 0 | 1 | Ref |
| 4851 | LR215718 | Unassigned | Unassigned | Unassigned | 149_15 | Clustered | 16 | VC_149_15 | 16 | 0.0788 | 1 | 0.0788 | 0 | 0 | 0 | 0.8083 | Ref |
| 4852 | LR215719 | Unassigned | Unassigned | Unassigned | 149_13 | Clustered | 3 | VC_149_13 | 3 | 0.0087 | 0.95178903 | 0.0083 | 0 | 0 | 0 | 1 | Ref |
| 4853 | LR215720 | Unassigned | Unassigned | Unassigned | 149_2 | Clustered/Singleton | 1 | VC_149_2 | 1 | 0 | 1 | 0 | 0 | 0 | 0 | 1 | Ref |
| 4854 | LR215721 | Unassigned | Unassigned | Unassigned | 149_29 | Clustered | 3 | VC_149_29 | 3 | 0.012 | 0.99448577 | 0.012 | 0 | 0 | 0 | 1 | Ref |
| 4855 | LR215722 | Unassigned | Unassigned | Unassigned | 283_97 | Clustered/Singleton | 1 | VC_283_97 | 1 | 0 | 1 | 0 | 0 | 0 | 0 | 1 | Ref |
| 4856 | LR215723 | Unassigned | Unassigned | Unassigned | 283_71 | Clustered/Singleton | 1 | VC_283_71 | 1 | 0 | 1 | 0 | 0 | 0 | 0 | 1 | Ref |
| 4857 | LR215724 | Unassigned | Unassigned | Unassigned | 283_113 | Clustered/Singleton | 1 | VC_283_113 | 1 | 0 | 1 | 0 | 0 | 0 | 0 | 1 | Ref |
| 4858 | LR535901 | Unassigned | Unassigned | Unassigned | 1208_0 | Clustered | 11 | VC_1208_0 | 11 | 1 | 0 | 0 | 0 | 0 | 0 | 1 | Ref |

| | | | | | | | | | | | | | | | | | |
|---|---|---|---|---|---|---|---|---|---|---|---|---|---|---|---|---|---|
| 4859 | LR535902 | Unassigned | Unassigned | Unassigned | 959_5 | Clustered | 19 | VC_959_5 | 19 | 0.2626 | 1 | 0.2626 | 0 | 0 | 0 | 1 | Ref |
| 4860 | LR535903 | Unassigned | Unassigned | Unassigned | 1208_0 | Clustered | 11 | VC_1208_0 | 11 | 1 | 0 | 0 | 0 | 0 | 0 | 1 | Ref |
| 4861 | LR535904 | Unassigned | Unassigned | Unassigned | 1208_0 | Clustered | 11 | VC_1208_0 | 11 | 1 | 0 | 0 | 0 | 0 | 0 | 1 | Ref |
| 4862 | LR535905 | Unassigned | Unassigned | Unassigned | 1208_0 | Clustered | 11 | VC_1208_0 | 11 | 1 | 0 | 0 | 0 | 0 | 0 | 1 | Ref |
| 4863 | LR535906 | Unassigned | Unassigned | Unassigned | 1208_0 | Clustered | 11 | VC_1208_0 | 11 | 1 | 0 | 0 | 0 | 0 | 0 | 1 | Ref |
| 4864 | LR535907 | Unassigned | Unassigned | Unassigned | 1208_0 | Clustered | 11 | VC_1208_0 | 11 | 1 | 0 | 0 | 0 | 0 | 0 | 1 | Ref |
| 4865 | LR535908 | Unassigned | Unassigned | Unassigned | 1208_0 | Clustered | 11 | VC_1208_0 | 11 | 1 | 0 | 0 | 0 | 0 | 0 | 1 | Ref |
| 4866 | LR535909 | Unassigned | Unassigned | Unassigned | 959_5 | Clustered | 19 | VC_959_5 | 19 | 0.2626 | 1 | 0.2626 | 0 | 0 | 0 | 1 | Ref |
| 4867 | LR535910 | Unassigned | Unassigned | Unassigned | 959_5 | Clustered | 19 | VC_959_5 | 19 | 0.2626 | 1 | 0.2626 | 0 | 0 | 0 | 1 | Ref |
| 4868 | LR535911 | Unassigned | Unassigned | Unassigned | 1208_0 | Clustered | 11 | VC_1208_0 | 11 | 1 | 0 | 0 | 0 | 0 | 0 | 1 | Ref |
| 4869 | LR535912 | Unassigned | Unassigned | Unassigned | 1208_0 | Clustered | 11 | VC_1208_0 | 11 | 1 | 0 | 0 | 0 | 0 | 0 | 1 | Ref |
| 4870 | LR535913 | Unassigned | Unassigned | Unassigned | 1208_0 | Clustered | 11 | VC_1208_0 | 11 | 1 | 0 | 0 | 0 | 0 | 0 | 1 | Ref |
| 4871 | LR535914 | Unassigned | Unassigned | Unassigned | 959_5 | Clustered | 19 | VC_959_5 | 19 | 0.2626 | 1 | 0.2626 | 0 | 0 | 0 | 1 | Ref |
| 4872 | LR535915 | Unassigned | Unassigned | Unassigned | 959_5 | Clustered | 19 | VC_959_5 | 19 | 0.2626 | 1 | 0.2626 | 0 | 0 | 0 | 1 | Ref |
| 4873 | LR535916 | Unassigned | Unassigned | Unassigned | 959_5 | Clustered | 19 | VC_959_5 | 19 | 0.2626 | 1 | 0.2626 | 0 | 0 | 0 | 1 | Ref |
| 4874 | LR535917 | Unassigned | Unassigned | Unassigned | 1208_0 | Clustered | 11 | VC_1208_0 | 11 | 1 | 0 | 0 | 0 | 0 | 0 | 1 | Ref |
| 4875 | LR535918 | Unassigned | Unassigned | Unassigned | 959_5 | Clustered | 19 | VC_959_5 | 19 | 0.2626 | 1 | 0.2626 | 0 | 0 | 0 | 1 | Ref |
| 4876 | LR535919 | Unassigned | Unassigned | Unassigned | 959_5 | Clustered | 19 | VC_959_5 | 19 | 0.2626 | 1 | 0.2626 | 0 | 0 | 0 | 1 | Ref |
| 4877 | LR535920 | Unassigned | Unassigned | Unassigned | 959_5 | Clustered | 19 | VC_959_5 | 19 | 0.2626 | 1 | 0.2626 | 0 | 0 | 0 | 1 | Ref |
| 4878 | LR535921 | Unassigned | Unassigned | Unassigned | 959_5 | Clustered | 19 | VC_959_5 | 19 | 0.2626 | 1 | 0.2626 | 0 | 0 | 0 | 1 | Ref |
| 4879 | LR588166 | Unassigned | Unassigned | Unassigned | 165_0 | Clustered | 4 | VC_165_0 | 4 | 0.2247 | 0.99998328 | 0.2247 | 0 | 0 | 0 | 1 | Ref |
| 4880 | LR595850 | Unassigned | Unassigned | Unassigned | 43_0 | Clustered | 38 | VC_43_0 | 38 | 0.5885 | 1 | 0.5885 | 0 | 0 | 0 | 0.9659 | Ref |
| 4881 | LR595859 | Unassigned | Unassigned | Unassigned | 43_0 | Clustered | 38 | VC_43_0 | 38 | 0.5885 | 1 | 0.5885 | 0 | 0 | 0 | 0.9659 | Ref |
| 4882 | LR595860 | Unassigned | Unassigned | Unassigned | 43_0 | Clustered | 38 | VC_43_0 | 38 | 0.5885 | 1 | 0.5885 | 0 | 0 | 0 | 0.9659 | Ref |
| 4883 | LR595861 | Unassigned | Unassigned | Unassigned | 43_0 | Clustered | 38 | VC_43_0 | 38 | 0.5885 | 1 | 0.5885 | 0 | 0 | 0 | 0.9659 | Ref |
| 4884 | LR595863 | Unassigned | Unassigned | Unassigned | 43_0 | Clustered | 38 | VC_43_0 | 38 | 0.5885 | 1 | 0.5885 | 0 | 0 | 0 | 0.9659 | Ref |
| 4885 | LR595864 | Unassigned | Unassigned | Unassigned | 43_0 | Clustered | 38 | VC_43_0 | 38 | 0.5885 | 1 | 0.5885 | 0 | 0 | 0 | 0.9659 | Ref |
| 4886 | LR595865 | Unassigned | Unassigned | Unassigned | 43_0 | Clustered | 38 | VC_43_0 | 38 | 0.5885 | 1 | 0.5885 | 0 | 0 | 0 | 0.9659 | Ref |
| 4887 | LR595866 | Unassigned | Unassigned | Unassigned | 43_0 | Clustered | 38 | VC_43_0 | 38 | 0.5885 | 1 | 0.5885 | 0 | 0 | 0 | 0.9659 | Ref |
| 4888 | LR595867 | Unassigned | Unassigned | Unassigned | 50_0 | Clustered | 9 | VC_50_0 | 9 | 0.5531 | 1 | 0.5531 | 0 | 0 | 0 | 1 | Ref |
| 4889 | LR595868 | Unassigned | Unassigned | Unassigned | 51_0 | Clustered | 10 | VC_51_0 | 10 | 0.1741 | 1 | 0.1741 | 0 | 0 | 0 | 1 | Ref |
| 4890 | LR595869 | Unassigned | Unassigned | Unassigned | 228_0 | Clustered | 60 | VC_228_0 | 60 | 0.942 | 1 | 0.942 | 0 | 0 | 0 | 1 | Ref |
| 4891 | LR595870 | Unassigned | Unassigned | Unassigned | 228_0 | Clustered | 60 | VC_228_0 | 60 | 0.942 | 1 | 0.942 | 0 | 0 | 0 | 1 | Ref |
| 4892 | LR595873 | Unassigned | Unassigned | Unassigned | 228_0 | Clustered | 60 | VC_228_0 | 60 | 0.942 | 1 | 0.942 | 0 | 0 | 0 | 1 | Ref |
| 4893 | LR595883 | Unassigned | Unassigned | Unassigned | 228_0 | Clustered | 60 | VC_228_0 | 60 | 0.942 | 1 | 0.942 | 0 | 0 | 0 | 1 | Ref |
| 4894 | LR595885 | Unassigned | Unassigned | Unassigned | 228_0 | Clustered | 60 | VC_228_0 | 60 | 0.942 | 1 | 0.942 | 0 | 0 | 0 | 1 | Ref |
| 4895 | LR595887 | Unassigned | Unassigned | Unassigned | 228_0 | Clustered | 60 | VC_228_0 | 60 | 0.942 | 1 | 0.942 | 0 | 0 | 0 | 1 | Ref |
| 4896 | LR595891 | Unassigned | Unassigned | Unassigned | 229_0 | Clustered | 32 | VC_229_0 | 32 | 0.9119 | 1 | 0.9119 | 0 | 0 | 0 | 1 | Ref |
| 4897 | LR596614 | Unassigned | Unassigned | Unassigned | 942_0 | Clustered | 36 | VC_942_0 | 36 | 0.7848 | 1 | 0.7848 | 0 | 0 | 0 | 1 | Ref |
| 4898 | LR596615 | Unassigned | Unassigned | Unassigned | 164_8 | Clustered | 3 | VC_164_8 | 3 | 0.0299 | 0.84944649 | 0.0254 | 0 | 0 | 0 | 1 | Ref |
| 4899 | LR596902 | Unassigned | Unassigned | Unassigned | 418_0 | Clustered | 6 | VC_418_0 | 6 | 0.4575 | 0.9999995 | 0.4575 | 0 | 0 | 0 | 0.8 | Ref |
| 4900 | LR596903 | Unassigned | Unassigned | Unassigned | 717_0 | Clustered | 7 | VC_717_0 | 7 | 0.5813 | 1 | 0.5813 | 0 | 0 | 0 | 0.9048 | Ref |
| 4901 | LR597635 | Unassigned | Unassigned | Unassigned | 43_0 | Clustered | 38 | VC_43_0 | 38 | 0.5885 | 1 | 0.5885 | 0 | 0 | 0 | 0.9659 | Ref |
| 4902 | LR597636 | Unassigned | Unassigned | Unassigned | 43_0 | Clustered | 38 | VC_43_0 | 38 | 0.5885 | 1 | 0.5885 | 0 | 0 | 0 | 0.9659 | Ref |
| 4903 | LR597637 | Unassigned | Unassigned | Unassigned | 5_0 | Clustered | 30 | VC_5_0 | 30 | 0.9647 | 1 | 0.9647 | 0 | 0 | 0 | 1 | Ref |
| 4904 | LR597638 | Unassigned | Unassigned | Unassigned | 5_0 | Clustered | 30 | VC_5_0 | 30 | 0.9647 | 1 | 0.9647 | 0 | 0 | 0 | 1 | Ref |
| 4905 | LR597639 | Unassigned | Unassigned | Unassigned | 43_0 | Clustered | 38 | VC_43_0 | 38 | 0.5885 | 1 | 0.5885 | 0 | 0 | 0 | 0.9659 | Ref |
| 4906 | LR597640 | Unassigned | Unassigned | Unassigned | 5_0 | Clustered | 30 | VC_5_0 | 30 | 0.9647 | 1 | 0.9647 | 0 | 0 | 0 | 1 | Ref |
| 4907 | LR597641 | Unassigned | Unassigned | Unassigned | 51_0 | Clustered | 10 | VC_51_0 | 10 | 0.1741 | 1 | 0.1741 | 0 | 0 | 0 | 1 | Ref |
| 4908 | LR597642 | Unassigned | Unassigned | Unassigned | 245_0 | Clustered | 10 | VC_245_0 | 10 | 0.1805 | 1 | 0.1805 | 0 | 0 | 0 | 0.9778 | Ref |
| 4909 | LR597643 | Unassigned | Unassigned | Unassigned | 43_0 | Clustered | 38 | VC_43_0 | 38 | 0.5885 | 1 | 0.5885 | 0 | 0 | 0 | 0.9659 | Ref |
| 4910 | LR597644 | Unassigned | Unassigned | Unassigned | 43_0 | Clustered | 38 | VC_43_0 | 38 | 0.5885 | 1 | 0.5885 | 0 | 0 | 0 | 0.9659 | Ref |
| 4911 | LR597646 | Unassigned | Unassigned | Unassigned | 835_0 | Clustered | 67 | VC_835_0 | 67 | 0.9624 | 1 | 0.9624 | 0 | 0 | 0 | 1 | Ref |
| 4912 | LR597647 | Unassigned | Unassigned | Unassigned | 772_9 | Clustered | 7 | VC_772_9 | 7 | 0.0369 | 0.87705503 | 0.0324 | 0 | 0 | 0 | 1 | Ref |
| 4913 | LR597648 | Unassigned | Unassigned | Unassigned | 43_0 | Clustered | 38 | VC_43_0 | 38 | 0.5885 | 1 | 0.5885 | 0 | 0 | 0 | 0.9659 | Ref |
| 4914 | LR597649 | Unassigned | Unassigned | Unassigned | 5_0 | Clustered | 30 | VC_5_0 | 30 | 0.9647 | 1 | 0.9647 | 0 | 0 | 0 | 1 | Ref |
| 4915 | LR597650 | Unassigned | Unassigned | Unassigned | 43_0 | Clustered | 38 | VC_43_0 | 38 | 0.5885 | 1 | 0.5885 | 0 | 0 | 0 | 0.9659 | Ref |
| 4916 | LR597651 | Unassigned | Unassigned | Unassigned | 43_0 | Clustered | 38 | VC_43_0 | 38 | 0.5885 | 1 | 0.5885 | 0 | 0 | 0 | 0.9659 | Ref |
| 4917 | LR597652 | Unassigned | Unassigned | Unassigned | 43_0 | Clustered | 38 | VC_43_0 | 38 | 0.5885 | 1 | 0.5885 | 0 | 0 | 0 | 0.9659 | Ref |
| 4918 | LR597653 | Unassigned | Unassigned | Unassigned | 43_0 | Clustered | 38 | VC_43_0 | 38 | 0.5885 | 1 | 0.5885 | 0 | 0 | 0 | 0.9659 | Ref |
| 4919 | LR597654 | Unassigned | Unassigned | Unassigned | 847_1 | Clustered | 26 | VC_847_1 | 26 | 0.6323 | 1 | 0.6323 | 0 | 0 | 0 | 0.9815 | Ref |
| 4920 | LR597655 | Unassigned | Unassigned | Unassigned | 190_14 | Clustered | 3 | VC_190_14 | 3 | 0.0144 | 0.99696852 | 0.0143 | 0 | 0 | 0 | 0.6667 | Ref |
| 4921 | LR597656 | Unassigned | Unassigned | Unassigned | 584_0 | Clustered | 28 | VC_584_0 | 28 | 0.7704 | 1 | 0.7704 | 0 | 0 | 0 | 1 | Ref |
| 4922 | LR597657 | Unassigned | Unassigned | Unassigned | 283_137 | Clustered/Singleton | 1 | VC_283_137 | 1 | 0 | 1 | 0 | 0 | 0 | 0 | 1 | Ref |
| 4923 | LR597658 | Unassigned | Unassigned | Unassigned | 329_6 | Clustered | 3 | VC_329_6 | 3 | 0.0107 | 0.99833469 | 0.0107 | 0 | 0 | 0 | 1 | Ref |
| 4924 | LR597659 | Unassigned | Unassigned | Unassigned | 190_24 | Clustered | 5 | VC_190_24 | 5 | 0.0293 | 0.9999996 | 0.0293 | 0 | 0 | 0 | 0.7 | Ref |
| 4925 | LR597660 | Unassigned | Unassigned | Unassigned | 283_101 | Clustered/Singleton | 1 | VC_283_101 | 1 | 0 | 1 | 0 | 0 | 0 | 0 | 1 | Ref |
| 4926 | LR597663 | Unassigned | Unassigned | Unassigned | 329_9 | Clustered/Singleton | 1 | VC_329_9 | 1 | 0 | 1 | 0 | 0 | 0 | 0 | 1 | Ref |
| 4927 | LR694165 | Unassigned | Unassigned | Unassigned | 772_9 | Clustered | 7 | VC_772_9 | 7 | 0.0369 | 0.87705503 | 0.0324 | 0 | 0 | 0 | 1 | Ref |
| 4928 | LR694610 | Unassigned | Unassigned | Unassigned | 772_7 | Clustered | 2 | VC_772_7 | 2 | 0.006 | 0.58868714 | 0.0035 | 0 | 0 | 0 | 1 | Ref |
| 4929 | LR694611 | Unassigned | Unassigned | Unassigned | 772_7 | Clustered | 2 | VC_772_7 | 2 | 0.006 | 0.58868714 | 0.0035 | 0 | 0 | 0 | 1 | Ref |
| 4930 | LR699048 | Unassigned | Unassigned | Unassigned | 329_7 | Clustered | 18 | VC_329_7 | 18 | 0.0938 | 1 | 0.0938 | 0 | 0 | 0 | 0.7778 | Ref |
| 4931 | LR699804 | Unassigned | Unassigned | Unassigned | 772_19 | Clustered | 16 | VC_772_19 | 16 | 0.0975 | 0.9970466 | 0.0972 | 0 | 0 | 0 | 0.8083 | Ref |
| 4932 | LR743523 | Unassigned | Unassigned | Unassigned | 116_0 | Clustered | 13 | VC_116_0 | 13 | 1 | 1 | 1 | 0 | 0 | 0 | 1 | Ref |
| 4933 | LR743524 | Unassigned | Unassigned | Unassigned | 172_0 | Clustered | 5 | VC_172_0 | 5 | 0.3579 | 0.99999997 | 0.3579 | 0 | 0 | 0 | 1 | Ref |
| 4934 | LR743528 | Unassigned | Unassigned | Unassigned | 116_0 | Clustered | 13 | VC_116_0 | 13 | 1 | 1 | 1 | 0 | 0 | 0 | 1 | Ref |
| 4935 | LR743529 | Unassigned | Unassigned | Unassigned | 116_0 | Clustered | 13 | VC_116_0 | 13 | 1 | 1 | 1 | 0 | 0 | 0 | 1 | Ref |
| 4936 | LR743530 | Unassigned | Unassigned | Unassigned | 749_0 | Clustered | 40 | VC_749_0 | 40 | 0.7517 | 1 | 0.7517 | 0 | 0 | 0 | 1 | Ref |
| 4937 | LR743531 | Unassigned | Unassigned | Unassigned | 116_0 | Clustered | 13 | VC_116_0 | 13 | 1 | 1 | 1 | 0 | 0 | 0 | 1 | Ref |
| 4938 | LR743532 | Unassigned | Unassigned | Unassigned | 169_0 | Clustered | 20 | VC_169_0 | 20 | 0.7529 | 1 | 0.7529 | 0 | 0 | 0 | 1 | Ref |
| 4939 | LR745206 | Unassigned | Unassigned | Unassigned | 533_1 | Clustered/Singleton | 1 | VC_533_1 | 1 | 0 | 1 | 0 | 0 | 0 | 0 | 1 | Ref |
| 4940 | LR745706 | Unassigned | Unassigned | Unassigned | 392_0 | Clustered | 126 | VC_392_0 | 126 | 0.9621 | 1 | 0.9621 | 0 | 0 | 0 | 1 | Ref |
| 4941 | LR745707 | Unassigned | Unassigned | Unassigned | 392_0 | Clustered | 126 | VC_392_0 | 126 | 0.9621 | 1 | 0.9621 | 0 | 0 | 0 | 1 | Ref |
| 4942 | LR745708 | Unassigned | Unassigned | Unassigned | 392_0 | Clustered | 126 | VC_392_0 | 126 | 0.9621 | 1 | 0.9621 | 0 | 0 | 0 | 1 | Ref |
| 4943 | LR745709 | Unassigned | Unassigned | Unassigned | 392_0 | Clustered | 126 | VC_392_0 | 126 | 0.9621 | 1 | 0.9621 | 0 | 0 | 0 | 1 | Ref |
| 4944 | LR745710 | Unassigned | Unassigned | Unassigned | 392_0 | Clustered | 126 | VC_392_0 | 126 | 0.9621 | 1 | 0.9621 | 0 | 0 | 0 | 1 | Ref |
| 4945 | LR746137 | Unassigned | Unassigned | Unassigned | 847_1 | Clustered | 26 | VC_847_1 | 26 | 0.6323 | 1 | 0.6323 | 0 | 0 | 0 | 0.9815 | Ref |
| 4946 | LR746310 | Unassigned | Unassigned | Unassigned | 668_0 | Clustered | 25 | VC_668_0 | 25 | 0.5539 | 1 | 0.5539 | 0 | 0 | 0 | 0.96 | Ref |
| 4947 | LR746311 | Unassigned | Unassigned | Unassigned | 283_141 | Clustered/Singleton | 1 | VC_283_141 | 1 | 0 | 1 | 0 | 0 | 0 | 0 | 1 | Ref |
| 4948 | LR756501 | Unassigned | Unassigned | Unassigned | 533_0 | Clustered | 2 | VC_533_0 | 2 | 0.2712 | 0.93085693 | 0.2525 | 0 | 0 | 0 | 1 | Ref |
| 4949 | LR756502 | Unassigned | Unassigned | Unassigned | 533_2 | Clustered/Singleton | 1 | VC_533_2 | 1 | 0 | 1 | 0 | 0 | 0 | 0 | 1 | Ref |
| 4950 | LR756503 | Unassigned | Unassigned | Unassigned | 533_0 | Clustered | 2 | VC_533_0 | 2 | 0.2712 | 0.93085693 | 0.2525 | 0 | 0 | 0 | 1 | Ref |
| 4951 | LR756504 | Unassigned | Unassigned | Unassigned | 533_3 | Clustered/Singleton | 1 | VC_533_3 | 1 | 0 | 1 | 0 | 0 | 0 | 0 | 1 | Ref |
| 4952 | LR757892 | Unassigned | Unassigned | Unassigned | 938_0 | Clustered | 99 | VC_938_0 | 99 | 0.9119 | 1 | 0.9119 | 0 | 0 | 0 | 0.9928 | Ref |
| 4953 | LR778216 | Unassigned | Unassigned | Unassigned | 74_0 | Clustered | 15 | VC_74_0 | 15 | 0.5204 | 1 | 0.5204 | 0 | 0 | 0 | 1 | Ref |
| 4954 | LR794124 | Unassigned | Unassigned | Unassigned | 837_0 | Clustered | 29 | VC_837_0 | 29 | 0.9955 | 1 | 0.9955 | 0 | 0 | 0 | 1 | Ref |
| 4955 | LR794146 | Unassigned | Unassigned | Unassigned | 29_0 | Clustered | 10 | VC_29_0 | 10 | 0.5208 | 1 | 0.5208 | 0 | 0 | 0 | 1 | Ref |
| 4956 | LR794147 | Unassigned | Unassigned | Unassigned | 29_0 | Clustered | 10 | VC_29_0 | 10 | 0.5208 | 1 | 0.5208 | 0 | 0 | 0 | 1 | Ref |
| 4957 | LR794148 | Unassigned | Unassigned | Unassigned | 29_0 | Clustered | 10 | VC_29_0 | 10 | 0.5208 | 1 | 0.5208 | 0 | 0 | 0 | 1 | Ref |
| 4958 | LR794149 | Unassigned | Unassigned | Unassigned | 29_0 | Clustered | 10 | VC_29_0 | 10 | 0.5208 | 1 | 0.5208 | 0 | 0 | 0 | 1 | Ref |
| 4959 | LR794150 | Unassigned | Unassigned | Unassigned | 29_0 | Clustered | 10 | VC_29_0 | 10 | 0.5208 | 1 | 0.5208 | 0 | 0 | 0 | 1 | Ref |
| 4960 | LR794151 | Unassigned | Unassigned | Unassigned | 29_0 | Clustered | 10 | VC_29_0 | 10 | 0.5208 | 1 | 0.5208 | 0 | 0 | 0 | 1 | Ref |
| 4961 | LR794152 | Unassigned | Unassigned | Unassigned | 29_0 | Clustered | 10 | VC_29_0 | 10 | 0.5208 | 1 | 0.5208 | 0 | 0 | 0 | 1 | Ref |
| 4962 | LR797314 | Unassigned | Unassigned | Unassigned | 185_0 | Clustered | 3 | VC_185_0 | 3 | 0.4487 | 0.97148006 | 0.4359 | 0 | 0 | 0 | 1 | Ref |
| 4963 | LR797923 | Unassigned | Unassigned | Unassigned | 29_0 | Clustered | 10 | VC_29_0 | 10 | 0.5208 | 1 | 0.5208 | 0 | 0 | 0 | 1 | Ref |
| 4964 | LR800396 | Unassigned | Unassigned | Unassigned | 133_13 | Clustered | 2 | VC_133_13 | 2 | 0.0051 | 0.86794768 | 0.0044 | 0 | 0 | 0 | 1 | Ref |
| 4965 | LR811967 | Unassigned | Unassigned | Unassigned | 133_28 | Clustered | 3 | VC_133_28 | 3 | 0.0105 | 0.97661637 | 0.0103 | 0 | 0 | 0 | 0.6667 | Ref |
| 4966 | LR811968 | Unassigned | Unassigned | Unassigned | 133_4 | Clustered/Singleton | 1 | VC_133_4 | 1 | 0 | 1 | 0 | 0 | 0 | 0 | 1 | Ref |
| 4967 | LR811969 | Unassigned | Unassigned | Unassigned | 133_28 | Clustered | 3 | VC_133_28 | 3 | 0.0105 | 0.97661637 | 0.0103 | 0 | 0 | 0 | 0.6667 | Ref |
| 4968 | LR813619 | Unassigned | Unassigned | Unassigned | 793_0 | Clustered | 12 | VC_793_0 | 12 | 0.8531 | 1 | 0.8531 | 0 | 0 | 0 | 1 | Ref |
| 4969 | LR865361 | Unassigned | Unassigned | Unassigned | 848_0 | Clustered | 28 | VC_848_0 | 28 | 0.7927 | 1 | 0.7927 | 0 | 0 | 0 | 1 | Ref |
| 4970 | LR877331 | Unassigned | Unassigned | Unassigned | 491_0 | Clustered | 36 | VC_491_0 | 36 | 0.5336 | 1 | 0.5336 | 0 | 0 | 0 | 0.9714 | Ref |
| 4971 | LR877332 | Unassigned | Unassigned | Unassigned | 283_67 | Clustered | 2 | VC_283_67 | 2 | 0.0027 | 0.90726003 | 0.0025 | 0 | 0 | 0 | 1 | Ref |
| 4972 | LR880803 | Unassigned | Unassigned | Unassigned | 329_3 | Clustered | 24 | VC_329_3 | 24 | 0.1206 | 1 | 0.1206 | 0 | 0 | 0 | 0.7283 | Ref |
| 4973 | LR881104 | Unassigned | Unassigned | Unassigned | 347_0 | Clustered | 3 | VC_347_0 | 3 | 0.1545 | 0.9984446 | 0.1543 | 0 | 0 | 0 | 1 | Ref |
| 4974 | LR990702 | Unassigned | Unassigned | Unassigned | 835_0 | Clustered | 67 | VC_835_0 | 67 | 0.9624 | 1 | 0.9624 | 0 | 0 | 0 | 1 | Ref |
| 4975 | LR990704 | Unassigned | Unassigned | Unassigned | 283_38 | Clustered/Singleton | 1 | VC_283_38 | 1 | 0 | 1 | 0 | 0 | 0 | 0 | 1 | Ref |
| 4976 | LR990835 | Unassigned | Unassigned | Unassigned | 92_0 | Clustered | 22 | VC_92_0 | 22 | 0.5639 | 1 | 0.5639 | 0 | 0 | 0 | 1 | Ref |
| 4977 | LR999870 | Unassigned | Unassigned | Unassigned | 835_0 | Clustered | 67 | VC_835_0 | 67 | 0.9624 | 1 | 0.9624 | 0 | 0 | 0 | 1 | Ref |
| 4978 | LR999871 | Unassigned | Unassigned | Unassigned | 779_0 | Clustered | 37 | VC_779_0 | 37 | 0.5682 | 1 | 0.5682 | 0 | 0 | 0 | 1 | Ref |
| 4979 | LT546029 | Unassigned | Unassigned | Unassigned | 691_1 | Clustered | 5 | VC_691_1 | 5 | 0.4358 | 0.99999993 | 0.4358 | 0 | 0 | 0 | 1 | Ref |
| 4980 | LT594300 | Unassigned | Unassigned | Unassigned | 451_0 | Clustered | 80 | VC_451_0 | 80 | 0.9991 | 1 | 0.9991 | 0 | 0 | 0 | 1 | Ref |
| 4981 | LT598654 | Unassigned | Unassigned | Unassigned | 67_5 | Clustered/Singleton | 1 | VC_67_5 | 1 | 0 | 1 | 0 | 0 | 0 | 0 | 1 | Ref |
| 4982 | LT600745 | Unassigned | Unassigned | Unassigned | 822_0 | Clustered | 2 | VC_822_0 | 2 | 0.038 | 0.89489263 | 0.034 | 0 | 0 | 0 | 1 | Ref |
| 4983 | LT603033 | Unassigned | Unassigned | Unassigned | 164_11 | Clustered | 10 | VC_164_11 | 10 | 0.1507 | 1 | 0.1507 | 0 | 0 | 0 | 0.9111 | Ref |
| 4984 | LT603684 | Unassigned | Unassigned | Unassigned | 630_0 | Clustered | 20 | VC_630_0 | 20 | 0.8513 | 1 | 0.8513 | 0 | 0 | 0 | 1 | Ref |
| 4985 | LT607758 | Unassigned | Unassigned | Unassigned | 668_0 | Clustered | 25 | VC_668_0 | 25 | 0.5539 | 1 | 0.5539 | 0 | 0 | 0 | 0.96 | Ref |
| 4986 | LT608331 | Unassigned | Unassigned | Unassigned | 179_0 | Clustered | 53 | VC_179_0 | 53 | 0.9137 | 1 | 0.9137 | 0 | 0 | 0 | 1 | Ref |
| 4987 | LT615366 | Unassigned | Unassigned | Unassigned | 691_1 | Clustered | 5 | VC_691_1 | 5 | 0.4358 | 0.99999993 | 0.4358 | 0 | 0 | 0 | 1 | Ref |
| 4988 | LT630001 | Unassigned | Unassigned | Unassigned | 967_0 | Clustered | 16 | VC_967_0 | 16 | 0.8971 | 1 | 0.8971 | 0 | 0 | 0 | 1 | Ref |
| 4989 | LT714109 | Unassigned | Unassigned | Unassigned | 28_1 | Clustered | 53 | VC_28_1 | 53 | 0.7164 | 1 | 0.7164 | 0 | 0 | 0 | 0.9325 | Ref |
| 4990 | LT841304 | Unassigned | Unassigned | Unassigned | 942_0 | Clustered | 36 | VC_942_0 | 36 | 0.7848 | 1 | 0.7848 | 0 | 0 | 0 | 1 | Ref |
| 4991 | LT898437 | Unassigned | Unassigned | Unassigned | 315_0 | Clustered | 12 | VC_315_0 | 12 | 0.8947 | 1 | 0.8947 | 0 | 0 | 0 | 1 | Ref |
| 4992 | LT907986 | Unassigned | Unassigned | Unassigned | 795_0 | Clustered | 19 | VC_795_0 | 19 | 0.6848 | 1 | 0.6848 | 0 | 0 | 0 | 1 | Ref |
| 4993 | LT960551 | Unassigned | Unassigned | Unassigned | 164_8 | Clustered | 3 | VC_164_8 | 3 | 0.0299 | 0.84944649 | 0.0254 | 0 | 0 | 0 | 1 | Ref |
| 4994 | LT960552 | Unassigned | Unassigned | Unassigned | 164_8 | Clustered | 3 | VC_164_8 | 3 | 0.0299 | 0.84944649 | 0.0254 | 0 | 0 | 0 | 1 | Ref |
| 4995 | LT960606 | Unassigned | Unassigned | Unassigned | 386_0 | Clustered | 49 | VC_386_0 | 49 | 0.7941 | 1 | 0.7941 | 0 | 0 | 0 | 1 | Ref |
| 4996 | LT960608 | Unassigned | Unassigned | Unassigned | 85_3 | Clustered | 19 | VC_85_3 | 19 | 0.3671 | 1 | 0.3671 | 0 | 0 | 0 | 0.807 | Ref |
| 4997 | LT960609 | Unassigned | Unassigned | Unassigned | 835_0 | Clustered | 67 | VC_835_0 | 67 | 0.9624 | 1 | 0.9624 | 0 | 0 | 0 | 1 | Ref |
| 4998 | LT961732 | Unassigned | Unassigned | Unassigned | 942_0 | Clustered | 36 | VC_942_0 | 36 | 0.7848 | 1 | 0.7848 | 0 | 0 | 0 | 1 | Ref |
| 4999 | LT961836 | Unassigned | Unassigned | Unassigned | 386_0 | Clustered | 49 | VC_386_0 | 49 | 0.7941 | 1 | 0.7941 | 0 | 0 | 0 | 1 | Ref |
| 5000 | LT961837 | Unassigned | Unassigned | Unassigned | 386_0 | Clustered | 49 | VC_386_0 | 49 | 0.7941 | 1 | 0.7941 | 0 | 0 | 0 | 1 | Ref |
| 5001 | LT961838 | Unassigned | Unassigned | Unassigned | 386_0 | Clustered | 49 | VC_386_0 | 49 | 0.7941 | 1 | 0.7941 | 0 | 0 | 0 | 1 | Ref |
| 5002 | LT961840 | Unassigned | Unassigned | Unassigned | 386_0 | Clustered | 49 | VC_386_0 | 49 | 0.7941 | 1 | 0.7941 | 0 | 0 | 0 | 1 | Ref |
| 5003 | LT961841 | Unassigned | Unassigned | Unassigned | 386_0 | Clustered | 49 | VC_386_0 | 49 | 0.7941 | 1 | 0.7941 | 0 | 0 | 0 | 1 | Ref |
| 5004 | LT961842 | Unassigned | Unassigned | Unassigned | 386_0 | Clustered | 49 | VC_386_0 | 49 | 0.7941 | 1 | 0.7941 | 0 | 0 | 0 | 1 | Ref |
| 5005 | LT961843 | Unassigned | Unassigned | Unassigned | 386_0 | Clustered | 49 | VC_386_0 | 49 | 0.7941 | 1 | 0.7941 | 0 | 0 | 0 | 1 | Ref |
| 5006 | LT961844 | Unassigned | Unassigned | Unassigned | 386_0 | Clustered | 49 | VC_386_0 | 49 | 0.7941 | 1 | 0.7941 | 0 | 0 | 0 | 1 | Ref |
| 5007 | LT961845 | Unassigned | Unassigned | Unassigned | 386_0 | Clustered | 49 | VC_386_0 | 49 | 0.7941 | 1 | 0.7941 | 0 | 0 | 0 | 1 | Ref |
| 5008 | LT961846 | Unassigned | Unassigned | Unassigned | 386_0 | Clustered | 49 | VC_386_0 | 49 | 0.7941 | 1 | 0.7941 | 0 | 0 | 0 | 1 | Ref |
| 5009 | LT962379 | Unassigned | Unassigned | Unassigned | 386_0 | Clustered | 49 | VC_386_0 | 49 | 0.7941 | 1 | 0.7941 | 0 | 0 | 0 | 1 | Ref |
| 5010 | LT962380 | Unassigned | Unassigned | Unassigned | 386_0 | Clustered | 49 | VC_386_0 | 49 | 0.7941 | 1 | 0.7941 | 0 | 0 | 0 | 1 | Ref |
| 5011 | LT962475 | Unassigned | Unassigned | Unassigned | 386_0 | Clustered | 49 | VC_386_0 | 49 | 0.7941 | 1 | 0.7941 | 0 | 0 | 0 | 1 | Ref |
| 5012 | LT962906 | Unassigned | Unassigned | Unassigned | 386_0 | Clustered | 49 | VC_386_0 | 49 | 0.7941 | 1 | 0.7941 | 0 | 0 | 0 | 1 | Ref |
| 5013 | LT962907 | Unassigned | Unassigned | Unassigned | 386_0 | Clustered | 49 | VC_386_0 | 49 | 0.7941 | 1 | 0.7941 | 0 | 0 | 0 | 1 | Ref |
| 5014 | LT985568 | Unassigned | Unassigned | Unassigned | 260_0 | Clustered | 13 | VC_260_0 | 13 | 0.3705 | 1 | 0.3705 | 0 | 0 | 0 | 1 | Ref |
| 5015 | LT985569 | Unassigned | Unassigned | Unassigned | 263_0 | Clustered | 7 | VC_263_0 | 7 | 0.2364 | 1 | 0.2364 | 0 | 0 | 0 | 1 | Ref |
| 5016 | LT985571 | Unassigned | Unassigned | Unassigned | 260_0 | Clustered | 13 | VC_260_0 | 13 | 0.3705 | 1 | 0.3705 | 0 | 0 | 0 | 1 | Ref |
| 5017 | LT985572 | Unassigned | Unassigned | Unassigned | 339_0 | Clustered | 9 | VC_339_0 | 9 | 0.9405 | 0.99999771 | 0.9405 | 0 | 0 | 0 | 1 | Ref |
| 5018 | LT985574 | Unassigned | Unassigned | Unassigned | 263_0 | Clustered | 7 | VC_263_0 | 7 | 0.2364 | 1 | 0.2364 | 0 | 0 | 0 | 1 | Ref |
| 5019 | LT985575 | Unassigned | Unassigned | Unassigned | 265_0 | Clustered | 12 | VC_265_0 | 12 | 0.4401 | 1 | 0.4401 | 0 | 0 | 0 | 1 | Ref |
| 5020 | LT985576 | Unassigned | Unassigned | Unassigned | 265_0 | Clustered | 12 | VC_265_0 | 12 | 0.4401 | 1 | 0.4401 | 0 | 0 | 0 | 1 | Ref |

| | | | | | | | | | | | | | | | | | |
|---|---|---|---|---|---|---|---|---|---|---|---|---|---|---|---|---|---|
| 5021 | LT985578 | Unassigned | Unassigned | Unassigned | 263_0 | Clustered | 7 | VC_263_0 | 7 | 0.2364 | 1 | 0.2364 | 0 | 0 | 0 | 1 | Ref |
| 5022 | LT985581 | Unassigned | Unassigned | Unassigned | 339_0 | Clustered | 9 | VC_339_0 | 9 | 0.9405 | 0.99999771 | 0.9405 | 0 | 0 | 0 | 1 | Ref |
| 5023 | LT985583 | Unassigned | Unassigned | Unassigned | 263_0 | Clustered | 7 | VC_263_0 | 7 | 0.2364 | 1 | 0.2364 | 0 | 0 | 0 | 1 | Ref |
| 5024 | LT985584 | Unassigned | Unassigned | Unassigned | 339_0 | Clustered | 9 | VC_339_0 | 9 | 0.9405 | 0.99999771 | 0.9405 | 0 | 0 | 0 | 1 | Ref |
| 5025 | LT985586 | Unassigned | Unassigned | Unassigned | 339_0 | Clustered | 9 | VC_339_0 | 9 | 0.9405 | 0.99999771 | 0.9405 | 0 | 0 | 0 | 1 | Ref |
| 5026 | LT985587 | Unassigned | Unassigned | Unassigned | 339_0 | Clustered | 9 | VC_339_0 | 9 | 0.9405 | 0.99999771 | 0.9405 | 0 | 0 | 0 | 1 | Ref |
| 5027 | LT985589 | Unassigned | Unassigned | Unassigned | 301_0 | Clustered | 2 | VC_301_0 | 2 | 0.1292 | 0.94930205 | 0.1226 | 0 | 0 | 0 | 1 | Ref |
| 5028 | LT985593 | Unassigned | Unassigned | Unassigned | 339_0 | Clustered | 9 | VC_339_0 | 9 | 0.9405 | 0.99999771 | 0.9405 | 0 | 0 | 0 | 1 | Ref |
| 5029 | LT986460 | Unassigned | Unassigned | Unassigned | 755_0 | Clustered | 9 | VC_755_0 | 9 | 0.4489 | 1 | 0.4489 | 0 | 0 | 0 | 1 | Ref |
| 5030 | LT986678 | Unassigned | Unassigned | Unassigned | 1211_0 | Clustered | 25 | VC_1211_0 | 25 | 1 | 1 | 1 | 0 | 0 | 0 | 1 | Ref |
| 5031 | LT993224 | Unassigned | Unassigned | Unassigned | 260_0 | Clustered | 13 | VC_260_0 | 13 | 0.3705 | 1 | 0.3705 | 0 | 0 | 0 | 1 | Ref |
| 5032 | LT993227 | Unassigned | Unassigned | Unassigned | 339_0 | Clustered | 9 | VC_339_0 | 9 | 0.9405 | 0.99999771 | 0.9405 | 0 | 0 | 0 | 1 | Ref |
| 5033 | LT993229 | Unassigned | Unassigned | Unassigned | 263_0 | Clustered | 7 | VC_263_0 | 7 | 0.2364 | 1 | 0.2364 | 0 | 0 | 0 | 1 | Ref |
| 5034 | LT993247 | Unassigned | Unassigned | Unassigned | 301_0 | Clustered | 2 | VC_301_0 | 2 | 0.1292 | 0.94930205 | 0.1226 | 0 | 0 | 0 | 1 | Ref |
| 5035 | LT996080 | Unassigned | Unassigned | Unassigned | 339_0 | Clustered | 9 | VC_339_0 | 9 | 0.9405 | 0.99999771 | 0.9405 | 0 | 0 | 0 | 1 | Ref |
| 5036 | LT996081 | Unassigned | Unassigned | Unassigned | 265_0 | Clustered | 12 | VC_265_0 | 12 | 0.4401 | 1 | 0.4401 | 0 | 0 | 0 | 1 | Ref |
| 5037 | LT996082 | Unassigned | Unassigned | Unassigned | 263_0 | Clustered | 7 | VC_263_0 | 7 | 0.2364 | 1 | 0.2364 | 0 | 0 | 0 | 1 | Ref |
| 5038 | LT999987 | Unassigned | Unassigned | Unassigned | 179_0 | Clustered | 53 | VC_179_0 | 53 | 0.9137 | 1 | 0.9137 | 0 | 0 | 0 | 1 | Ref |
| 5039 | MF001354 | Unassigned | Unassigned | Unassigned | 283_79 | Clustered/Singleton | 1 | VC_283_79 | 1 | 0 | 1 | 0 | 0 | 0 | 0 | 1 | Ref |
| 5040 | MF001355 | Unassigned | Unassigned | Unassigned | 288_3 | Clustered | 3 | VC_288_3 | 3 | 0.0452 | 0.99844025 | 0.0452 | 0 | 0 | 0 | 1 | Ref |
| 5041 | MF001356 | Unassigned | Unassigned | Unassigned | 637_0 | Clustered | 115 | VC_637_0 | 115 | 0.731 | 1 | 0.731 | 0 | 0 | 0 | 0.8764 | Ref |
| 5042 | MF001358 | Unassigned | Unassigned | Unassigned | 692_0 | Clustered | 2 | VC_692_0 | 2 | 0.4503 | 0.95340726 | 0.4294 | 0 | 0 | 0 | 1 | Ref |
| 5043 | MF001359 | Unassigned | Unassigned | Unassigned | 283_183 | Clustered/Singleton | 1 | VC_283_183 | 1 | 0 | 1 | 0 | 0 | 0 | 0 | 1 | Ref |
| 5044 | MF001360 | Unassigned | Unassigned | Unassigned | 283_14 | Clustered/Singleton | 1 | VC_283_14 | 1 | 0 | 1 | 0 | 0 | 0 | 0 | 1 | Ref |
| 5045 | MF001361 | Unassigned | Unassigned | Unassigned | 692_0 | Clustered | 2 | VC_692_0 | 2 | 0.4503 | 0.95340726 | 0.4294 | 0 | 0 | 0 | 1 | Ref |
| 5046 | MF001362 | Unassigned | Unassigned | Unassigned | 133_24 | Clustered/Singleton | 1 | VC_133_24 | 1 | 0 | 1 | 0 | 0 | 0 | 0 | 1 | Ref |
| 5047 | MF001363 | Unassigned | Unassigned | Unassigned | 191_16 | Clustered | 12 | VC_191_16 | 12 | 0.0643 | 1 | 0.0643 | 0 | 0 | 0 | 0.9394 | Ref |
| 5048 | MF001365 | Unassigned | Unassigned | Unassigned | 149_15 | Clustered | 16 | VC_149_15 | 16 | 0.0788 | 1 | 0.0788 | 0 | 0 | 0 | 0.8083 | Ref |
| 5049 | MF001366 | Unassigned | Unassigned | Unassigned | 637_0 | Clustered | 115 | VC_637_0 | 115 | 0.731 | 1 | 0.731 | 0 | 0 | 0 | 0.8764 | Ref |
| 5050 | MF033347 | Unassigned | Unassigned | Unassigned | 699_0 | Clustered | 71 | VC_699_0 | 71 | 0.9352 | 1 | 0.9352 | 0 | 0 | 0 | 1 | Ref |
| 5051 | MF033348 | Unassigned | Unassigned | Unassigned | 699_0 | Clustered | 71 | VC_699_0 | 71 | 0.9352 | 1 | 0.9352 | 0 | 0 | 0 | 1 | Ref |
| 5052 | MF033349 | Unassigned | Unassigned | Unassigned | 699_0 | Clustered | 71 | VC_699_0 | 71 | 0.9352 | 1 | 0.9352 | 0 | 0 | 0 | 1 | Ref |
| 5053 | MF033350 | Unassigned | Unassigned | Unassigned | 699_0 | Clustered | 71 | VC_699_0 | 71 | 0.9352 | 1 | 0.9352 | 0 | 0 | 0 | 1 | Ref |
| 5054 | MF033351 | Unassigned | Unassigned | Unassigned | 699_0 | Clustered | 71 | VC_699_0 | 71 | 0.9352 | 1 | 0.9352 | 0 | 0 | 0 | 1 | Ref |
| 5055 | MF034659 | Unassigned | Unassigned | Unassigned | 422_0 | Clustered | 5 | VC_422_0 | 5 | 0.2592 | 0.99999997 | 0.2592 | 0 | 0 | 0 | 1 | Ref |
| 5056 | MF038790 | Unassigned | Unassigned | Unassigned | 1164_0 | Clustered | 25 | VC_1164_0 | 25 | 0.7312 | 1 | 0.7312 | 0 | 0 | 0 | 1 | Ref |
| 5057 | MF038791 | Unassigned | Unassigned | Unassigned | 1164_0 | Clustered | 25 | VC_1164_0 | 25 | 0.7312 | 1 | 0.7312 | 0 | 0 | 0 | 1 | Ref |
| 5058 | MF041990 | Unassigned | Unassigned | Unassigned | 309_0 | Clustered | 3 | VC_309_0 | 3 | 0.1265 | 0.99498896 | 0.1258 | 0 | 0 | 0 | 1 | Ref |
| 5059 | MF042360 | Unassigned | Unassigned | Unassigned | 344_5 | Clustered/Singleton | 1 | VC_344_5 | 1 | 0 | 1 | 0 | 0 | 0 | 0 | 1 | Ref |
| 5060 | MF044457 | Unassigned | Unassigned | Unassigned | 286_13 | Clustered | 10 | VC_286_13 | 10 | 0.0698 | 0.99999929 | 0.0698 | 0 | 0 | 0 | 0.9556 | Ref |
| 5061 | MF044458 | Unassigned | Unassigned | Unassigned | 792_0 | Clustered | 35 | VC_792_0 | 35 | 0.9828 | 1 | 0.9828 | 0 | 0 | 0 | 1 | Ref |
| 5062 | MF063068 | Unassigned | Unassigned | Unassigned | 344_2 | Clustered/Singleton | 1 | VC_344_2 | 1 | 0 | 1 | 0 | 0 | 0 | 0 | 1 | Ref |
| 5063 | MF072690 | Unassigned | Unassigned | Unassigned | 546_3 | Clustered | 4 | VC_546_3 | 4 | 0.0418 | 0.92194988 | 0.0385 | 0 | 0 | 0 | 1 | Ref |
| 5064 | MF074189 | Unassigned | Unassigned | Unassigned | 609_0 | Clustered | 4 | VC_609_0 | 4 | 0.489 | 0.99992419 | 0.489 | 0 | 0 | 0 | 1 | Ref |
| 5065 | MF093736 | Unassigned | Unassigned | Unassigned | 575_0 | Clustered | 35 | VC_575_0 | 35 | 0.8815 | 1 | 0.8815 | 0 | 0 | 0 | 1 | Ref |
| 5066 | MF098558 | Unassigned | Unassigned | Unassigned | 1090_0 | Clustered | 4 | VC_1090_0 | 4 | 1 | 1 | 1 | 0 | 0 | 0 | 1 | Ref |
| 5067 | MF101922 | Unassigned | Unassigned | Unassigned | 1212_0 | Clustered | 2 | VC_1212_0 | 2 | 1 | 1 | 1 | 0 | 0 | 0 | 1 | Ref |
| 5068 | MF133445 | Unassigned | Unassigned | Unassigned | 546_3 | Clustered | 4 | VC_546_3 | 4 | 0.0418 | 0.92194988 | 0.0385 | 0 | 0 | 0 | 1 | Ref |
| 5069 | MF133446 | Unassigned | Unassigned | Unassigned | 519_0 | Clustered | 39 | VC_519_0 | 39 | 0.8156 | 1 | 0.8156 | 0 | 0 | 0 | 1 | Ref |
| 5070 | MF140397 | Unassigned | Unassigned | Unassigned | 1166_0 | Clustered | 20 | VC_1166_0 | 20 | 0.99 | 1 | 0.99 | 0 | 0 | 0 | 1 | Ref |
| 5071 | MF140398 | Unassigned | Unassigned | Unassigned | 237_1 | Clustered | 13 | VC_237_1 | 13 | 0.1773 | 1 | 0.1773 | 0 | 0 | 0 | 0.7308 | Ref |
| 5072 | MF140399 | Unassigned | Unassigned | Unassigned | 519_0 | Clustered | 39 | VC_519_0 | 39 | 0.8156 | 1 | 0.8156 | 0 | 0 | 0 | 1 | Ref |
| 5073 | MF140400 | Unassigned | Unassigned | Unassigned | 1160_0 | Clustered | 73 | VC_1160_0 | 73 | 0.9835 | 1 | 0.9835 | 0 | 0 | 0 | 1 | Ref |
| 5074 | MF140401 | Unassigned | Unassigned | Unassigned | 1164_0 | Clustered | 25 | VC_1164_0 | 25 | 0.7312 | 1 | 0.7312 | 0 | 0 | 0 | 1 | Ref |
| 5075 | MF140402 | Unassigned | Unassigned | Unassigned | 234_0 | Clustered | 21 | VC_234_0 | 21 | 0.8355 | 1 | 0.8355 | 0 | 0 | 0 | 1 | Ref |
| 5076 | MF140403 | Unassigned | Unassigned | Unassigned | 216_3 | Clustered | 8 | VC_216_3 | 8 | 0.0701 | 1 | 0.0701 | 0 | 0 | 0 | 0.8571 | Ref |
| 5077 | MF140404 | Unassigned | Unassigned | Unassigned | 1160_0 | Clustered | 73 | VC_1160_0 | 73 | 0.9835 | 1 | 0.9835 | 0 | 0 | 0 | 1 | Ref |
| 5078 | MF140405 | Unassigned | Unassigned | Unassigned | 889_0 | Clustered | 16 | VC_889_0 | 16 | 0.2969 | 1 | 0.2969 | 0 | 0 | 0 | 1 | Ref |
| 5079 | MF140406 | Unassigned | Unassigned | Unassigned | 237_1 | Clustered | 13 | VC_237_1 | 13 | 0.1773 | 1 | 0.1773 | 0 | 0 | 0 | 0.7308 | Ref |
| 5080 | MF140407 | Unassigned | Unassigned | Unassigned | 1160_0 | Clustered | 73 | VC_1160_0 | 73 | 0.9835 | 1 | 0.9835 | 0 | 0 | 0 | 1 | Ref |
| 5081 | MF140409 | Unassigned | Unassigned | Unassigned | 950_0 | Clustered | 38 | VC_950_0 | 38 | 0.9443 | 1 | 0.9443 | 0 | 0 | 0 | 1 | Ref |
| 5082 | MF140410 | Unassigned | Unassigned | Unassigned | 221_0 | Clustered | 22 | VC_221_0 | 22 | 0.7447 | 1 | 0.7447 | 0 | 0 | 0 | 1 | Ref |
| 5083 | MF140412 | Unassigned | Unassigned | Unassigned | 236_0 | Clustered | 72 | VC_236_0 | 72 | 0.8104 | 1 | 0.8104 | 0 | 0 | 0 | 1 | Ref |
| 5084 | MF140413 | Unassigned | Unassigned | Unassigned | 889_0 | Clustered | 16 | VC_889_0 | 16 | 0.2969 | 1 | 0.2969 | 0 | 0 | 0 | 1 | Ref |
| 5085 | MF140414 | Unassigned | Unassigned | Unassigned | 889_0 | Clustered | 16 | VC_889_0 | 16 | 0.2969 | 1 | 0.2969 | 0 | 0 | 0 | 1 | Ref |
| 5086 | MF140415 | Unassigned | Unassigned | Unassigned | 950_0 | Clustered | 38 | VC_950_0 | 38 | 0.9443 | 1 | 0.9443 | 0 | 0 | 0 | 1 | Ref |
| 5087 | MF140416 | Unassigned | Unassigned | Unassigned | 236_1 | Clustered | 5 | VC_236_1 | 5 | 0.0358 | 0.99999997 | 0.0358 | 0 | 0 | 0 | 1 | Ref |
| 5088 | MF140417 | Unassigned | Unassigned | Unassigned | 950_0 | Clustered | 38 | VC_950_0 | 38 | 0.9443 | 1 | 0.9443 | 0 | 0 | 0 | 1 | Ref |
| 5089 | MF140418 | Unassigned | Unassigned | Unassigned | 1169_0 | Clustered | 7 | VC_1169_0 | 7 | 0.9811 | 0.99995613 | 0.981 | 0 | 0 | 0 | 1 | Ref |
| 5090 | MF140419 | Unassigned | Unassigned | Unassigned | 950_0 | Clustered | 38 | VC_950_0 | 38 | 0.9443 | 1 | 0.9443 | 0 | 0 | 0 | 1 | Ref |
| 5091 | MF140420 | Unassigned | Unassigned | Unassigned | 950_0 | Clustered | 38 | VC_950_0 | 38 | 0.9443 | 1 | 0.9443 | 0 | 0 | 0 | 1 | Ref |
| 5092 | MF140421 | Unassigned | Unassigned | Unassigned | 889_0 | Clustered | 16 | VC_889_0 | 16 | 0.2969 | 1 | 0.2969 | 0 | 0 | 0 | 1 | Ref |
| 5093 | MF140422 | Unassigned | Unassigned | Unassigned | 888_0 | Clustered | 28 | VC_888_0 | 28 | 0.5497 | 1 | 0.5497 | 0 | 0 | 0 | 1 | Ref |
| 5094 | MF140423 | Unassigned | Unassigned | Unassigned | 1164_0 | Clustered | 25 | VC_1164_0 | 25 | 0.7312 | 1 | 0.7312 | 0 | 0 | 0 | 1 | Ref |
| 5095 | MF140424 | Unassigned | Unassigned | Unassigned | 1160_0 | Clustered | 73 | VC_1160_0 | 73 | 0.9835 | 1 | 0.9835 | 0 | 0 | 0 | 1 | Ref |
| 5096 | MF140425 | Unassigned | Unassigned | Unassigned | 1160_0 | Clustered | 73 | VC_1160_0 | 73 | 0.9835 | 1 | 0.9835 | 0 | 0 | 0 | 1 | Ref |
| 5097 | MF140426 | Unassigned | Unassigned | Unassigned | 950_0 | Clustered | 38 | VC_950_0 | 38 | 0.9443 | 1 | 0.9443 | 0 | 0 | 0 | 1 | Ref |
| 5098 | MF140427 | Unassigned | Unassigned | Unassigned | 1169_0 | Clustered | 7 | VC_1169_0 | 7 | 0.9811 | 0.99995613 | 0.981 | 0 | 0 | 0 | 1 | Ref |
| 5099 | MF140428 | Unassigned | Unassigned | Unassigned | 236_0 | Clustered | 72 | VC_236_0 | 72 | 0.8104 | 1 | 0.8104 | 0 | 0 | 0 | 1 | Ref |
| 5100 | MF140429 | Unassigned | Unassigned | Unassigned | 950_0 | Clustered | 38 | VC_950_0 | 38 | 0.9443 | 1 | 0.9443 | 0 | 0 | 0 | 1 | Ref |
| 5101 | MF140430 | Unassigned | Unassigned | Unassigned | 236_0 | Clustered | 72 | VC_236_0 | 72 | 0.8104 | 1 | 0.8104 | 0 | 0 | 0 | 1 | Ref |
| 5102 | MF140431 | Unassigned | Unassigned | Unassigned | 950_0 | Clustered | 38 | VC_950_0 | 38 | 0.9443 | 1 | 0.9443 | 0 | 0 | 0 | 1 | Ref |
| 5103 | MF140432 | Unassigned | Unassigned | Unassigned | 1164_0 | Clustered | 25 | VC_1164_0 | 25 | 0.7312 | 1 | 0.7312 | 0 | 0 | 0 | 1 | Ref |
| 5104 | MF140433 | Unassigned | Unassigned | Unassigned | 950_0 | Clustered | 38 | VC_950_0 | 38 | 0.9443 | 1 | 0.9443 | 0 | 0 | 0 | 1 | Ref |
| 5105 | MF140434 | Unassigned | Unassigned | Unassigned | 1169_0 | Clustered | 7 | VC_1169_0 | 7 | 0.9811 | 0.99995613 | 0.981 | 0 | 0 | 0 | 1 | Ref |
| 5106 | MF140435 | Unassigned | Unassigned | Unassigned | 234_0 | Clustered | 21 | VC_234_0 | 21 | 0.8355 | 1 | 0.8355 | 0 | 0 | 0 | 1 | Ref |
| 5107 | MF141540 | Unassigned | Unassigned | Unassigned | 762_0 | Clustered | 9 | VC_762_0 | 9 | 0.0885 | 1 | 0.0885 | 0 | 0 | 0 | 1 | Ref |
| 5108 | MF150911 | Unassigned | Unassigned | Unassigned | 160_0 | Clustered | 2 | VC_160_0 | 2 | 0.4956 | 0.94363853 | 0.4677 | 0 | 0 | 0 | 1 | Ref |
| 5109 | MF153391 | Unassigned | Unassigned | Unassigned | 638_0 | Clustered | 45 | VC_638_0 | 45 | 0.7741 | 1 | 0.7741 | 0 | 0 | 0 | 1 | Ref |
| 5110 | MF155889 | Unassigned | Unassigned | Unassigned | 875_0 | Clustered | 10 | VC_875_0 | 10 | 0.988 | 0.99948433 | 0.9875 | 0 | 0 | 0 | 1 | Ref |
| 5111 | MF155936 | Unassigned | Unassigned | Unassigned | 1046_0 | Clustered | 7 | VC_1046_0 | 7 | 0.9413 | 1 | 0.9413 | 0 | 0 | 0 | 1 | Ref |
| 5112 | MF155946 | Unassigned | Unassigned | Unassigned | 1071_2 | Clustered | 5 | VC_1071_2 | 5 | 0.0534 | 0.99997362 | 0.0534 | 0 | 0 | 0 | 0.9 | Ref |
| 5113 | MF155947 | Unassigned | Unassigned | Unassigned | 371_11 | Clustered | 31 | VC_371_11 | 31 | 0.0927 | 1 | 0.0927 | 0 | 0 | 0 | 0.7011 | Ref |
| 5114 | MF156577 | Unassigned | Unassigned | Unassigned | 818_0 | Clustered | 20 | VC_818_0 | 20 | 0.9601 | 1 | 0.9601 | 0 | 0 | 0 | 1 | Ref |
| 5115 | MF156578 | Unassigned | Unassigned | Unassigned | 818_0 | Clustered | 20 | VC_818_0 | 20 | 0.9601 | 1 | 0.9601 | 0 | 0 | 0 | 1 | Ref |
| 5116 | MF156580 | Unassigned | Unassigned | Unassigned | 818_0 | Clustered | 20 | VC_818_0 | 20 | 0.9601 | 1 | 0.9601 | 0 | 0 | 0 | 1 | Ref |
| 5117 | MF158036 | Unassigned | Unassigned | Unassigned | 540_0 | Clustered | 15 | VC_540_0 | 15 | 0.2866 | 1 | 0.2866 | 0 | 0 | 0 | 1 | Ref |
| 5118 | MF158037 | Unassigned | Unassigned | Unassigned | 540_0 | Clustered | 15 | VC_540_0 | 15 | 0.2866 | 1 | 0.2866 | 0 | 0 | 0 | 1 | Ref |
| 5119 | MF158038 | Unassigned | Unassigned | Unassigned | 461_0 | Clustered | 96 | VC_461_0 | 96 | 0.9307 | 1 | 0.9307 | 0 | 0 | 0 | 0.9575 | Ref |
| 5120 | MF158039 | Unassigned | Unassigned | Unassigned | 455_0 | Clustered | 18 | VC_455_0 | 18 | 0.4598 | 1 | 0.4598 | 0 | 0 | 0 | 1 | Ref |
| 5121 | MF158040 | Unassigned | Unassigned | Unassigned | 330_0 | Clustered | 27 | VC_330_0 | 27 | 0.5088 | 1 | 0.5088 | 0 | 0 | 0 | 0.8661 | Ref |
| 5122 | MF158041 | Unassigned | Unassigned | Unassigned | 330_0 | Clustered | 27 | VC_330_0 | 27 | 0.5088 | 1 | 0.5088 | 0 | 0 | 0 | 0.8661 | Ref |
| 5123 | MF158042 | Unassigned | Unassigned | Unassigned | 455_0 | Clustered | 18 | VC_455_0 | 18 | 0.4598 | 1 | 0.4598 | 0 | 0 | 0 | 1 | Ref |
| 5124 | MF158043 | Unassigned | Unassigned | Unassigned | 330_0 | Clustered | 27 | VC_330_0 | 27 | 0.5088 | 1 | 0.5088 | 0 | 0 | 0 | 0.8661 | Ref |
| 5125 | MF158044 | Unassigned | Unassigned | Unassigned | 330_0 | Clustered | 27 | VC_330_0 | 27 | 0.5088 | 1 | 0.5088 | 0 | 0 | 0 | 0.8661 | Ref |
| 5126 | MF158045 | Unassigned | Unassigned | Unassigned | 283_64 | Clustered/Singleton | 1 | VC_283_64 | 1 | 0 | 1 | 0 | 0 | 0 | 0 | 1 | Ref |
| 5127 | MF158046 | Unassigned | Unassigned | Unassigned | 283_189 | Clustered/Singleton | 1 | VC_283_189 | 1 | 0 | 1 | 0 | 0 | 0 | 0 | 1 | Ref |
| 5128 | MF161328 | Unassigned | Unassigned | Unassigned | 252_0 | Clustered | 121 | VC_252_0 | 121 | 0.7376 | 1 | 0.7376 | 0 | 0 | 0 | 0.9999 | Ref |
| 5129 | MF166859 | Unassigned | Unassigned | Unassigned | 422_0 | Clustered | 5 | VC_422_0 | 5 | 0.2592 | 0.99999997 | 0.2592 | 0 | 0 | 0 | 1 | Ref |
| 5130 | MF172979 | Unassigned | Unassigned | Unassigned | 626_0 | Clustered | 23 | VC_626_0 | 23 | 0.66 | 1 | 0.66 | 0 | 0 | 0 | 0.8379 | Ref |
| 5131 | MF176161 | Unassigned | Unassigned | Unassigned | 622_0 | Clustered | 2 | VC_622_0 | 2 | 0.0507 | 0.6259393 | 0.0317 | 0 | 0 | 0 | 1 | Ref |
| 5132 | MF185717 | Unassigned | Unassigned | Unassigned | 238_0 | Clustered | 17 | VC_238_0 | 17 | 0.6109 | 1 | 0.6109 | 0 | 0 | 0 | 1 | Ref |
| 5133 | MF185718 | Unassigned | Unassigned | Unassigned | 1074_0 | Clustered | 19 | VC_1074_0 | 19 | 0.9198 | 1 | 0.9198 | 0 | 0 | 0 | 1 | Ref |
| 5134 | MF185719 | Unassigned | Unassigned | Unassigned | 238_0 | Clustered | 17 | VC_238_0 | 17 | 0.6109 | 1 | 0.6109 | 0 | 0 | 0 | 1 | Ref |
| 5135 | MF185720 | Unassigned | Unassigned | Unassigned | 238_0 | Clustered | 17 | VC_238_0 | 17 | 0.6109 | 1 | 0.6109 | 0 | 0 | 0 | 1 | Ref |
| 5136 | MF185721 | Unassigned | Unassigned | Unassigned | 1074_0 | Clustered | 19 | VC_1074_0 | 19 | 0.9198 | 1 | 0.9198 | 0 | 0 | 0 | 1 | Ref |
| 5137 | MF185722 | Unassigned | Unassigned | Unassigned | 237_0 | Clustered | 4 | VC_237_0 | 4 | 0.0429 | 0.99998759 | 0.0429 | 0 | 0 | 0 | 1 | Ref |
| 5138 | MF185723 | Unassigned | Unassigned | Unassigned | 1074_0 | Clustered | 19 | VC_1074_0 | 19 | 0.9198 | 1 | 0.9198 | 0 | 0 | 0 | 1 | Ref |
| 5139 | MF185724 | Unassigned | Unassigned | Unassigned | 1074_0 | Clustered | 19 | VC_1074_0 | 19 | 0.9198 | 1 | 0.9198 | 0 | 0 | 0 | 1 | Ref |
| 5140 | MF185725 | Unassigned | Unassigned | Unassigned | 1074_0 | Clustered | 19 | VC_1074_0 | 19 | 0.9198 | 1 | 0.9198 | 0 | 0 | 0 | 1 | Ref |
| 5141 | MF185726 | Unassigned | Unassigned | Unassigned | 887_0 | Clustered | 18 | VC_887_0 | 18 | 0.3663 | 1 | 0.3663 | 0 | 0 | 0 | 1 | Ref |
| 5142 | MF185727 | Unassigned | Unassigned | Unassigned | 216_0 | Clustered | 4 | VC_216_0 | 4 | 0.048 | 0.9999878 | 0.048 | 0 | 0 | 0 | 1 | Ref |
| 5143 | MF185728 | Unassigned | Unassigned | Unassigned | 888_0 | Clustered | 28 | VC_888_0 | 28 | 0.5497 | 1 | 0.5497 | 0 | 0 | 0 | 1 | Ref |
| 5144 | MF185729 | Unassigned | Unassigned | Unassigned | 514_0 | Clustered | 75 | VC_514_0 | 75 | 0.9402 | 1 | 0.9402 | 0 | 0 | 0 | 1 | Ref |
| 5145 | MF185730 | Unassigned | Unassigned | Unassigned | 888_0 | Clustered | 28 | VC_888_0 | 28 | 0.5497 | 1 | 0.5497 | 0 | 0 | 0 | 1 | Ref |
| 5146 | MF185731 | Unassigned | Unassigned | Unassigned | 857_0 | Clustered | 11 | VC_857_0 | 11 | 0.9712 | 1 | 0.9712 | 0 | 0 | 0 | 1 | Ref |
| 5147 | MF185732 | Unassigned | Unassigned | Unassigned | 514_0 | Clustered | 75 | VC_514_0 | 75 | 0.9402 | 1 | 0.9402 | 0 | 0 | 0 | 1 | Ref |
| 5148 | MF185733 | Unassigned | Unassigned | Unassigned | 514_0 | Clustered | 75 | VC_514_0 | 75 | 0.9402 | 1 | 0.9402 | 0 | 0 | 0 | 1 | Ref |
| 5149 | MF188997 | Unassigned | Unassigned | Unassigned | 28_1 | Clustered | 53 | VC_28_1 | 53 | 0.7164 | 1 | 0.7164 | 0 | 0 | 0 | 0.9325 | Ref |
| 5150 | MF189170 | Unassigned | Unassigned | Unassigned | 802_0 | Clustered | 17 | VC_802_0 | 17 | 0.8025 | 1 | 0.8025 | 0 | 0 | 0 | 1 | Ref |
| 5151 | MF189171 | Unassigned | Unassigned | Unassigned | 802_0 | Clustered | 17 | VC_802_0 | 17 | 0.8025 | 1 | 0.8025 | 0 | 0 | 0 | 1 | Ref |
| 5152 | MF189172 | Unassigned | Unassigned | Unassigned | 802_0 | Clustered | 17 | VC_802_0 | 17 | 0.8025 | 1 | 0.8025 | 0 | 0 | 0 | 1 | Ref |
| 5153 | MF189173 | Unassigned | Unassigned | Unassigned | 802_0 | Clustered | 17 | VC_802_0 | 17 | 0.8025 | 1 | 0.8025 | 0 | 0 | 0 | 1 | Ref |
| 5154 | MF189174 | Unassigned | Unassigned | Unassigned | 802_0 | Clustered | 17 | VC_802_0 | 17 | 0.8025 | 1 | 0.8025 | 0 | 0 | 0 | 1 | Ref |
| 5155 | MF189175 | Unassigned | Unassigned | Unassigned | 802_0 | Clustered | 17 | VC_802_0 | 17 | 0.8025 | 1 | 0.8025 | 0 | 0 | 0 | 1 | Ref |
| 5156 | MF189176 | Unassigned | Unassigned | Unassigned | 1073_0 | Clustered | 24 | VC_1073_0 | 24 | 0.8388 | 1 | 0.8388 | 0 | 0 | 0 | 1 | Ref |
| 5157 | MF189177 | Unassigned | Unassigned | Unassigned | 1073_0 | Clustered | 24 | VC_1073_0 | 24 | 0.8388 | 1 | 0.8388 | 0 | 0 | 0 | 1 | Ref |
| 5158 | MF189178 | Unassigned | Unassigned | Unassigned | 1073_0 | Clustered | 24 | VC_1073_0 | 24 | 0.8388 | 1 | 0.8388 | 0 | 0 | 0 | 1 | Ref |
| 5159 | MF189179 | Unassigned | Unassigned | Unassigned | 956_0 | Clustered | 39 | VC_956_0 | 39 | 1 | 1 | 1 | 0 | 0 | 0 | 1 | Ref |
| 5160 | MF197383 | Unassigned | Unassigned | Unassigned | 1200_0 | Clustered | 4 | VC_1200_0 | 4 | 0.8691 | 0.9999992 | 0.8691 | 0 | 0 | 0 | 1 | Ref |
| 5161 | MF276773 | Unassigned | Unassigned | Unassigned | 228_0 | Clustered | 60 | VC_228_0 | 60 | 0.942 | 1 | 0.942 | 0 | 0 | 0 | 1 | Ref |
| 5162 | MF278336 | Unassigned | Unassigned | Unassigned | 988_0 | Clustered | 2 | VC_988_0 | 2 | 0.1586 | 0.95268934 | 0.1511 | 0 | 0 | 0 | 1 | Ref |
| 5163 | MF281061 | Unassigned | Unassigned | Unassigned | 519_0 | Clustered | 39 | VC_519_0 | 39 | 0.8156 | 1 | 0.8156 | 0 | 0 | 0 | 1 | Ref |
| 5164 | MF285615 | Unassigned | Unassigned | Unassigned | 823_0 | Clustered | 124 | VC_823_0 | 124 | 0.9492 | 1 | 0.9492 | 0 | 0 | 0 | 1 | Ref |
| 5165 | MF285616 | Unassigned | Unassigned | Unassigned | 385_0 | Clustered | 51 | VC_385_0 | 51 | 0.8754 | 1 | 0.8754 | 0 | 0 | 0 | 1 | Ref |
| 5166 | MF285617 | Unassigned | Unassigned | Unassigned | 385_0 | Clustered | 51 | VC_385_0 | 51 | 0.8754 | 1 | 0.8754 | 0 | 0 | 0 | 1 | Ref |
| 5167 | MF285618 | Unassigned | Unassigned | Unassigned | 488_1 | Clustered | 8 | VC_488_1 | 8 | 0.2927 | 1 | 0.2927 | 0 | 0 | 0 | 1 | Ref |
| 5168 | MF285619 | Unassigned | Unassigned | Unassigned | 132_0 | Clustered | 4 | VC_132_0 | 4 | 0.036 | 0.99998777 | 0.036 | 0 | 0 | 0 | 1 | Ref |
| 5169 | MF285620 | Unassigned | Unassigned | Unassigned | 385_0 | Clustered | 51 | VC_385_0 | 51 | 0.8754 | 1 | 0.8754 | 0 | 0 | 0 | 1 | Ref |
| 5170 | MF288917 | Unassigned | Unassigned | Unassigned | 884_0 | Clustered | 28 | VC_884_0 | 28 | 0.6211 | 1 | 0.6211 | 0 | 0 | 0 | 0.9841 | Ref |
| 5171 | MF288918 | Unassigned | Unassigned | Unassigned | 884_0 | Clustered | 28 | VC_884_0 | 28 | 0.6211 | 1 | 0.6211 | 0 | 0 | 0 | 0.9841 | Ref |
| 5172 | MF288919 | Unassigned | Unassigned | Unassigned | 884_0 | Clustered | 28 | VC_884_0 | 28 | 0.6211 | 1 | 0.6211 | 0 | 0 | 0 | 0.9841 | Ref |
| 5173 | MF288920 | Unassigned | Unassigned | Unassigned | 884_0 | Clustered | 28 | VC_884_0 | 28 | 0.6211 | 1 | 0.6211 | 0 | 0 | 0 | 0.9841 | Ref |
| 5174 | MF288921 | Unassigned | Unassigned | Unassigned | 883_0 | Clustered | 10 | VC_883_0 | 10 | 0.2015 | 1 | 0.2015 | 0 | 0 | 0 | 1 | Ref |
| 5175 | MF288922 | Unassigned | Unassigned | Unassigned | 883_0 | Clustered | 10 | VC_883_0 | 10 | 0.2015 | 1 | 0.2015 | 0 | 0 | 0 | 1 | Ref |
| 5176 | MF319184 | Unassigned | Unassigned | Unassigned | 763_2 | Clustered | 7 | VC_763_2 | 7 | 0.2612 | 1 | 0.2612 | 0 | 0 | 0 | 1 | Ref |
| 5177 | MF324898 | Unassigned | Unassigned | Unassigned | 956_0 | Clustered | 39 | VC_956_0 | 39 | 1 | 1 | 1 | 0 | 0 | 0 | 1 | Ref |
| 5178 | MF324899 | Unassigned | Unassigned | Unassigned | 216_0 | Clustered | 4 | VC_216_0 | 4 | 0.048 | 0.9999878 | 0.048 | 0 | 0 | 0 | 1 | Ref |
| 5179 | MF324900 | Unassigned | Unassigned | Unassigned | 956_0 | Clustered | 39 | VC_956_0 | 39 | 1 | 1 | 1 | 0 | 0 | 0 | 1 | Ref |
| 5180 | MF324901 | Unassigned | Unassigned | Unassigned | 956_0 | Clustered | 39 | VC_956_0 | 39 | 1 | 1 | 1 | 0 | 0 | 0 | 1 | Ref |
| 5181 | MF324902 | Unassigned | Unassigned | Unassigned | 956_0 | Clustered | 39 | VC_956_0 | 39 | 1 | 1 | 1 | 0 | 0 | 0 | 1 | Ref |
| 5182 | MF324903 | Unassigned | Unassigned | Unassigned | 956_0 | Clustered | 39 | VC_956_0 | 39 | 1 | 1 | 1 | 0 | 0 | 0 | 1 | Ref |

| | | | | | | | | | | | | | | | | | |
|---|---|---|---|---|---|---|---|---|---|---|---|---|---|---|---|---|---|
| 5183 | MF324904 | Unassigned | Unassigned | Unassigned | 956_0 | Clustered | 39 | VC_956_0 | 39 | 1 | 1 | 1 | 0 | 0 | 0 | 1 | Ref |
| 5184 | MF324905 | Unassigned | Unassigned | Unassigned | 956_0 | Clustered | 39 | VC_956_0 | 39 | 1 | 1 | 1 | 0 | 0 | 0 | 1 | Ref |
| 5185 | MF324906 | Unassigned | Unassigned | Unassigned | 802_0 | Clustered | 17 | VC_802_0 | 17 | 0.8025 | 1 | 0.8025 | 0 | 0 | 0 | 1 | Ref |
| 5186 | MF324907 | Unassigned | Unassigned | Unassigned | 1073_0 | Clustered | 24 | VC_1073_0 | 24 | 0.8388 | 1 | 0.8388 | 0 | 0 | 0 | 1 | Ref |
| 5187 | MF324908 | Unassigned | Unassigned | Unassigned | 237_2 | Clustered | 11 | VC_237_2 | 11 | 0.1409 | 1 | 0.1409 | 0 | 0 | 0 | 0.9818 | Ref |
| 5188 | MF324909 | Unassigned | Unassigned | Unassigned | 237_2 | Clustered | 11 | VC_237_2 | 11 | 0.1409 | 1 | 0.1409 | 0 | 0 | 0 | 0.9818 | Ref |
| 5189 | MF324910 | Unassigned | Unassigned | Unassigned | 888_0 | Clustered | 28 | VC_888_0 | 28 | 0.5497 | 1 | 0.5497 | 0 | 0 | 0 | 1 | Ref |
| 5190 | MF324911 | Unassigned | Unassigned | Unassigned | 237_2 | Clustered | 11 | VC_237_2 | 11 | 0.1409 | 1 | 0.1409 | 0 | 0 | 0 | 0.9818 | Ref |
| 5191 | MF324912 | Unassigned | Unassigned | Unassigned | 237_2 | Clustered | 11 | VC_237_2 | 11 | 0.1409 | 1 | 0.1409 | 0 | 0 | 0 | 0.9818 | Ref |
| 5192 | MF324913 | Unassigned | Unassigned | Unassigned | 237_2 | Clustered | 11 | VC_237_2 | 11 | 0.1409 | 1 | 0.1409 | 0 | 0 | 0 | 0.9818 | Ref |
| 5193 | MF324914 | Unassigned | Unassigned | Unassigned | 237_2 | Clustered | 11 | VC_237_2 | 11 | 0.1409 | 1 | 0.1409 | 0 | 0 | 0 | 0.9818 | Ref |
| 5194 | MF324915 | Unassigned | Unassigned | Unassigned | 237_1 | Clustered | 13 | VC_237_1 | 13 | 0.1773 | 1 | 0.1773 | 0 | 0 | 0 | 0.7308 | Ref |
| 5195 | MF327003 | Unassigned | Unassigned | Unassigned | 330_0 | Clustered | 27 | VC_330_0 | 27 | 0.5088 | 1 | 0.5088 | 0 | 0 | 0 | 0.8661 | Ref |
| 5196 | MF327004 | Unassigned | Unassigned | Unassigned | 330_0 | Clustered | 27 | VC_330_0 | 27 | 0.5088 | 1 | 0.5088 | 0 | 0 | 0 | 0.8661 | Ref |
| 5197 | MF327005 | Unassigned | Unassigned | Unassigned | 330_0 | Clustered | 27 | VC_330_0 | 27 | 0.5088 | 1 | 0.5088 | 0 | 0 | 0 | 0.8661 | Ref |
| 5198 | MF327006 | Unassigned | Unassigned | Unassigned | 590_0 | Clustered | 27 | VC_590_0 | 27 | 0.9947 | 1 | 0.9947 | 0 | 0 | 0 | 1 | Ref |
| 5199 | MF327007 | Unassigned | Unassigned | Unassigned | 283_158 | Clustered/Singleton | 1 | VC_283_158 | 1 | 0 | 1 | 0 | 0 | 0 | 0 | 1 | Ref |
| 5200 | MF327008 | Unassigned | Unassigned | Unassigned | 283_19 | Clustered/Singleton | 1 | VC_283_19 | 1 | 0 | 1 | 0 | 0 | 0 | 0 | 1 | Ref |
| 5201 | MF327009 | Unassigned | Unassigned | Unassigned | 283_20 | Clustered/Singleton | 1 | VC_283_20 | 1 | 0 | 1 | 0 | 0 | 0 | 0 | 1 | Ref |
| 5202 | MF346584 | Unassigned | Unassigned | Unassigned | 698_0 | Clustered | 26 | VC_698_0 | 26 | 0.9366 | 1 | 0.9366 | 0 | 0 | 0 | 1 | Ref |
| 5203 | MF346933 | Unassigned | Unassigned | Unassigned | 202_0 | Clustered | 102 | VC_202_0 | 102 | 0.4049 | 1 | 0.4049 | 0 | 0 | 0 | 0.7876 | Ref |
| 5204 | MF347636 | Unassigned | Unassigned | Unassigned | 1071_3 | Clustered | 16 | VC_1071_3 | 16 | 0.2354 | 1 | 0.2354 | 0 | 0 | 0 | 0.9667 | Ref |
| 5205 | MF347637 | Unassigned | Unassigned | Unassigned | 1071_3 | Clustered | 16 | VC_1071_3 | 16 | 0.2354 | 1 | 0.2354 | 0 | 0 | 0 | 0.9667 | Ref |
| 5206 | MF347638 | Unassigned | Unassigned | Unassigned | 1071_3 | Clustered | 16 | VC_1071_3 | 16 | 0.2354 | 1 | 0.2354 | 0 | 0 | 0 | 0.9667 | Ref |
| 5207 | MF347639 | Unassigned | Unassigned | Unassigned | 1071_3 | Clustered | 16 | VC_1071_3 | 16 | 0.2354 | 1 | 0.2354 | 0 | 0 | 0 | 0.9667 | Ref |
| 5208 | MF351863 | Unassigned | Unassigned | Unassigned | 432_3 | Clustered/Singleton | 1 | VC_432_3 | 1 | 0 | 1 | 0 | 0 | 0 | 0 | 1 | Ref |
| 5209 | MF356679 | Unassigned | Unassigned | Unassigned | 247_1 | Clustered/Singleton | 1 | VC_247_1 | 1 | 0 | 1 | 0 | 0 | 0 | 0 | 1 | Ref |
| 5210 | MF358541 | Unassigned | Unassigned | Unassigned | 1071_3 | Clustered | 16 | VC_1071_3 | 16 | 0.2354 | 1 | 0.2354 | 0 | 0 | 0 | 0.9667 | Ref |
| 5211 | MF358542 | Unassigned | Unassigned | Unassigned | 1071_3 | Clustered | 16 | VC_1071_3 | 16 | 0.2354 | 1 | 0.2354 | 0 | 0 | 0 | 0.9667 | Ref |
| 5212 | MF360957 | Unassigned | Unassigned | Unassigned | 448_0 | Clustered | 4 | VC_448_0 | 4 | 0.3059 | 0.99978853 | 0.3058 | 0 | 0 | 0 | 1 | Ref |
| 5213 | MF370225 | Unassigned | Unassigned | Unassigned | 329_1 | Clustered | 17 | VC_329_1 | 17 | 0.0867 | 1 | 0.0867 | 0 | 0 | 0 | 0.8235 | Ref |
| 5214 | MF370964 | Unassigned | Unassigned | Unassigned | 496_1 | Clustered/Singleton | 1 | VC_496_1 | 1 | 0 | 1 | 0 | 0 | 0 | 0 | 1 | Ref |
| 5215 | MF370965 | Unassigned | Unassigned | Unassigned | 896_0 | Clustered | 5 | VC_896_0 | 5 | 0.4981 | 0.99999977 | 0.4981 | 0 | 0 | 0 | 1 | Ref |
| 5216 | MF373840 | Unassigned | Unassigned | Unassigned | 513_0 | Clustered | 32 | VC_513_0 | 32 | 0.6881 | 1 | 0.6881 | 0 | 0 | 0 | 1 | Ref |
| 5217 | MF373841 | Unassigned | Unassigned | Unassigned | 237_1 | Clustered | 13 | VC_237_1 | 13 | 0.1773 | 1 | 0.1773 | 0 | 0 | 0 | 0.7308 | Ref |
| 5218 | MF373842 | Unassigned | Unassigned | Unassigned | 956_0 | Clustered | 39 | VC_956_0 | 39 | 1 | 1 | 1 | 0 | 0 | 0 | 1 | Ref |
| 5219 | MF373843 | Unassigned | Unassigned | Unassigned | 519_0 | Clustered | 39 | VC_519_0 | 39 | 0.8156 | 1 | 0.8156 | 0 | 0 | 0 | 1 | Ref |
| 5220 | MF374379 | Unassigned | Unassigned | Unassigned | 43_0 | Clustered | 38 | VC_43_0 | 38 | 0.5885 | 1 | 0.5885 | 0 | 0 | 0 | 0.9659 | Ref |
| 5221 | MF375456 | Unassigned | Unassigned | Unassigned | 74_0 | Clustered | 15 | VC_74_0 | 15 | 0.5204 | 1 | 0.5204 | 0 | 0 | 0 | 1 | Ref |
| 5222 | MF377440 | Unassigned | Unassigned | Unassigned | 236_0 | Clustered | 72 | VC_236_0 | 72 | 0.8104 | 1 | 0.8104 | 0 | 0 | 0 | 1 | Ref |
| 5223 | MF377441 | Unassigned | Unassigned | Unassigned | 1073_0 | Clustered | 24 | VC_1073_0 | 24 | 0.8388 | 1 | 0.8388 | 0 | 0 | 0 | 1 | Ref |
| 5224 | MF377442 | Unassigned | Unassigned | Unassigned | 1073_0 | Clustered | 24 | VC_1073_0 | 24 | 0.8388 | 1 | 0.8388 | 0 | 0 | 0 | 1 | Ref |
| 5225 | MF398190 | Unassigned | Unassigned | Unassigned | 149_8 | Clustered | 66 | VC_149_8 | 66 | 0.4565 | 1 | 0.4565 | 0 | 0 | 0 | 0.8182 | Ref |
| 5226 | MF402939 | Unassigned | Unassigned | Unassigned | 190_18 | Clustered/Singleton | 1 | VC_190_18 | 1 | 0 | 1 | 0 | 0 | 0 | 0 | 1 | Ref |
| 5227 | MF403005 | Unassigned | Unassigned | Unassigned | 987_0 | Clustered | 22 | VC_987_0 | 22 | 0.8955 | 1 | 0.8955 | 0 | 0 | 0 | 1 | Ref |
| 5228 | MF403006 | Unassigned | Unassigned | Unassigned | 987_0 | Clustered | 22 | VC_987_0 | 22 | 0.8955 | 1 | 0.8955 | 0 | 0 | 0 | 1 | Ref |
| 5229 | MF403007 | Unassigned | Unassigned | Unassigned | 1052_2 | Clustered/Singleton | 1 | VC_1052_2 | 1 | 0 | 1 | 0 | 0 | 0 | 0 | 1 | Ref |
| 5230 | MF405094 | Unassigned | Unassigned | Unassigned | 149_8 | Clustered | 66 | VC_149_8 | 66 | 0.4565 | 1 | 0.4565 | 0 | 0 | 0 | 0.8182 | Ref |
| 5231 | MF407276 | Unassigned | Unassigned | Unassigned | 817_0 | Clustered | 14 | VC_817_0 | 14 | 0.6342 | 1 | 0.6342 | 0 | 0 | 0 | 1 | Ref |
| 5232 | MF415412 | Unassigned | Unassigned | Unassigned | 938_0 | Clustered | 99 | VC_938_0 | 99 | 0.9119 | 1 | 0.9119 | 0 | 0 | 0 | 0.9928 | Ref |
| 5233 | MF418016 | Unassigned | Unassigned | Unassigned | 88_1 | Clustered/Singleton | 1 | VC_88_1 | 1 | 0 | 1 | 0 | 0 | 0 | 0 | 1 | Ref |
| 5234 | MF422185 | Unassigned | Unassigned | Unassigned | 85_3 | Clustered | 19 | VC_85_3 | 19 | 0.3671 | 1 | 0.3671 | 0 | 0 | 0 | 0.807 | Ref |
| 5235 | MF428478 | Unassigned | Unassigned | Unassigned | 11_0 | Clustered | 19 | VC_11_0 | 19 | 0.7562 | 1 | 0.7562 | 0 | 0 | 0 | 1 | Ref |
| 5236 | MF428479 | Unassigned | Unassigned | Unassigned | 11_0 | Clustered | 19 | VC_11_0 | 19 | 0.7562 | 1 | 0.7562 | 0 | 0 | 0 | 1 | Ref |
| 5237 | MF428480 | Unassigned | Unassigned | Unassigned | 11_0 | Clustered | 19 | VC_11_0 | 19 | 0.7562 | 1 | 0.7562 | 0 | 0 | 0 | 1 | Ref |
| 5238 | MF428481 | Unassigned | Unassigned | Unassigned | 11_0 | Clustered | 19 | VC_11_0 | 19 | 0.7562 | 1 | 0.7562 | 0 | 0 | 0 | 1 | Ref |
| 5239 | MF431252 | Unassigned | Unassigned | Unassigned | 637_0 | Clustered | 115 | VC_637_0 | 115 | 0.731 | 1 | 0.731 | 0 | 0 | 0 | 0.8764 | Ref |
| 5240 | MF431615 | Unassigned | Unassigned | Unassigned | 304_0 | Clustered | 11 | VC_304_0 | 11 | 0.7869 | 1 | 0.7869 | 0 | 0 | 0 | 1 | Ref |
| 5241 | MF431616 | Unassigned | Unassigned | Unassigned | 304_0 | Clustered | 11 | VC_304_0 | 11 | 0.7869 | 1 | 0.7869 | 0 | 0 | 0 | 1 | Ref |
| 5242 | MF431617 | Unassigned | Unassigned | Unassigned | 304_0 | Clustered | 11 | VC_304_0 | 11 | 0.7869 | 1 | 0.7869 | 0 | 0 | 0 | 1 | Ref |
| 5243 | MF431726 | Unassigned | Unassigned | Unassigned | 530_0 | Clustered | 9 | VC_530_0 | 9 | 0.8027 | 1 | 0.8027 | 0 | 0 | 0 | 1 | Ref |
| 5244 | MF431730 | Unassigned | Unassigned | Unassigned | 190_7 | Clustered | 7 | VC_190_7 | 7 | 0.0476 | 1 | 0.0476 | 0 | 0 | 0 | 1 | Ref |
| 5245 | MF431731 | Unassigned | Unassigned | Unassigned | 190_7 | Clustered | 7 | VC_190_7 | 7 | 0.0476 | 1 | 0.0476 | 0 | 0 | 0 | 1 | Ref |
| 5246 | MF431732 | Unassigned | Unassigned | Unassigned | 190_7 | Clustered | 7 | VC_190_7 | 7 | 0.0476 | 1 | 0.0476 | 0 | 0 | 0 | 1 | Ref |
| 5247 | MF431733 | Unassigned | Unassigned | Unassigned | 190_7 | Clustered | 7 | VC_190_7 | 7 | 0.0476 | 1 | 0.0476 | 0 | 0 | 0 | 1 | Ref |
| 5248 | MF431734 | Unassigned | Unassigned | Unassigned | 190_7 | Clustered | 7 | VC_190_7 | 7 | 0.0476 | 1 | 0.0476 | 0 | 0 | 0 | 1 | Ref |
| 5249 | MF431735 | Unassigned | Unassigned | Unassigned | 190_7 | Clustered | 7 | VC_190_7 | 7 | 0.0476 | 1 | 0.0476 | 0 | 0 | 0 | 1 | Ref |
| 5250 | MF431736 | Unassigned | Unassigned | Unassigned | 191_32 | Clustered | 12 | VC_191_32 | 12 | 0.0683 | 1 | 0.0683 | 0 | 0 | 0 | 0.7576 | Ref |
| 5251 | MF431737 | Unassigned | Unassigned | Unassigned | 191_32 | Clustered | 12 | VC_191_32 | 12 | 0.0683 | 1 | 0.0683 | 0 | 0 | 0 | 0.7576 | Ref |
| 5252 | MF431738 | Unassigned | Unassigned | Unassigned | 191_32 | Clustered | 12 | VC_191_32 | 12 | 0.0683 | 1 | 0.0683 | 0 | 0 | 0 | 0.7576 | Ref |
| 5253 | MF431739 | Unassigned | Unassigned | Unassigned | 191_32 | Clustered | 12 | VC_191_32 | 12 | 0.0683 | 1 | 0.0683 | 0 | 0 | 0 | 0.7576 | Ref |
| 5254 | MF431740 | Unassigned | Unassigned | Unassigned | 191_32 | Clustered | 12 | VC_191_32 | 12 | 0.0683 | 1 | 0.0683 | 0 | 0 | 0 | 0.7576 | Ref |
| 5255 | MF431741 | Unassigned | Unassigned | Unassigned | 191_32 | Clustered | 12 | VC_191_32 | 12 | 0.0683 | 1 | 0.0683 | 0 | 0 | 0 | 0.7576 | Ref |
| 5256 | MF443119 | Unassigned | Unassigned | Unassigned | 690_0 | Clustered | 50 | VC_690_0 | 50 | 0.9805 | 1 | 0.9805 | 0 | 0 | 0 | 1 | Ref |
| 5257 | MF443120 | Unassigned | Unassigned | Unassigned | 690_0 | Clustered | 50 | VC_690_0 | 50 | 0.9805 | 1 | 0.9805 | 0 | 0 | 0 | 1 | Ref |
| 5258 | MF443121 | Unassigned | Unassigned | Unassigned | 690_0 | Clustered | 50 | VC_690_0 | 50 | 0.9805 | 1 | 0.9805 | 0 | 0 | 0 | 1 | Ref |
| 5259 | MF443122 | Unassigned | Unassigned | Unassigned | 690_0 | Clustered | 50 | VC_690_0 | 50 | 0.9805 | 1 | 0.9805 | 0 | 0 | 0 | 1 | Ref |
| 5260 | MF443123 | Unassigned | Unassigned | Unassigned | 690_0 | Clustered | 50 | VC_690_0 | 50 | 0.9805 | 1 | 0.9805 | 0 | 0 | 0 | 1 | Ref |
| 5261 | MF443124 | Unassigned | Unassigned | Unassigned | 690_0 | Clustered | 50 | VC_690_0 | 50 | 0.9805 | 1 | 0.9805 | 0 | 0 | 0 | 1 | Ref |
| 5262 | MF443125 | Unassigned | Unassigned | Unassigned | 690_0 | Clustered | 50 | VC_690_0 | 50 | 0.9805 | 1 | 0.9805 | 0 | 0 | 0 | 1 | Ref |
| 5263 | MF443126 | Unassigned | Unassigned | Unassigned | 690_0 | Clustered | 50 | VC_690_0 | 50 | 0.9805 | 1 | 0.9805 | 0 | 0 | 0 | 1 | Ref |
| 5264 | MF443783 | Unassigned | Unassigned | Unassigned | 1213_0 | Clustered | 2 | VC_1213_0 | 2 | 1 | 1 | 1 | 0 | 0 | 0 | 1 | Ref |
| 5265 | MF443784 | Unassigned | Unassigned | Unassigned | 1213_0 | Clustered | 2 | VC_1213_0 | 2 | 1 | 1 | 1 | 0 | 0 | 0 | 1 | Ref |
| 5266 | MF448340 | Unassigned | Unassigned | Unassigned | 443_2 | Clustered | 10 | VC_443_2 | 10 | 0.3344 | 1 | 0.3344 | 0 | 0 | 0 | 0.9333 | Ref |
| 5267 | MF448554 | Unassigned | Unassigned | Unassigned | 202_0 | Clustered | 102 | VC_202_0 | 102 | 0.4049 | 1 | 0.4049 | 0 | 0 | 0 | 0.7876 | Ref |
| 5268 | MF448555 | Unassigned | Unassigned | Unassigned | 202_0 | Clustered | 102 | VC_202_0 | 102 | 0.4049 | 1 | 0.4049 | 0 | 0 | 0 | 0.7876 | Ref |
| 5269 | MF448556 | Unassigned | Unassigned | Unassigned | 202_0 | Clustered | 102 | VC_202_0 | 102 | 0.4049 | 1 | 0.4049 | 0 | 0 | 0 | 0.7876 | Ref |
| 5270 | MF448557 | Unassigned | Unassigned | Unassigned | 202_0 | Clustered | 102 | VC_202_0 | 102 | 0.4049 | 1 | 0.4049 | 0 | 0 | 0 | 0.7876 | Ref |
| 5271 | MF448558 | Unassigned | Unassigned | Unassigned | 202_0 | Clustered | 102 | VC_202_0 | 102 | 0.4049 | 1 | 0.4049 | 0 | 0 | 0 | 0.7876 | Ref |
| 5272 | MF448559 | Unassigned | Unassigned | Unassigned | 202_0 | Clustered | 102 | VC_202_0 | 102 | 0.4049 | 1 | 0.4049 | 0 | 0 | 0 | 0.7876 | Ref |
| 5273 | MF448560 | Unassigned | Unassigned | Unassigned | 202_0 | Clustered | 102 | VC_202_0 | 102 | 0.4049 | 1 | 0.4049 | 0 | 0 | 0 | 0.7876 | Ref |
| 5274 | MF448561 | Unassigned | Unassigned | Unassigned | 202_0 | Clustered | 102 | VC_202_0 | 102 | 0.4049 | 1 | 0.4049 | 0 | 0 | 0 | 0.7876 | Ref |
| 5275 | MF448562 | Unassigned | Unassigned | Unassigned | 202_0 | Clustered | 102 | VC_202_0 | 102 | 0.4049 | 1 | 0.4049 | 0 | 0 | 0 | 0.7876 | Ref |
| 5276 | MF448563 | Unassigned | Unassigned | Unassigned | 202_0 | Clustered | 102 | VC_202_0 | 102 | 0.4049 | 1 | 0.4049 | 0 | 0 | 0 | 0.7876 | Ref |
| 5277 | MF448564 | Unassigned | Unassigned | Unassigned | 202_0 | Clustered | 102 | VC_202_0 | 102 | 0.4049 | 1 | 0.4049 | 0 | 0 | 0 | 0.7876 | Ref |
| 5278 | MF448565 | Unassigned | Unassigned | Unassigned | 202_0 | Clustered | 102 | VC_202_0 | 102 | 0.4049 | 1 | 0.4049 | 0 | 0 | 0 | 0.7876 | Ref |
| 5279 | MF448566 | Unassigned | Unassigned | Unassigned | 202_0 | Clustered | 102 | VC_202_0 | 102 | 0.4049 | 1 | 0.4049 | 0 | 0 | 0 | 0.7876 | Ref |
| 5280 | MF448567 | Unassigned | Unassigned | Unassigned | 202_0 | Clustered | 102 | VC_202_0 | 102 | 0.4049 | 1 | 0.4049 | 0 | 0 | 0 | 0.7876 | Ref |
| 5281 | MF448568 | Unassigned | Unassigned | Unassigned | 202_0 | Clustered | 102 | VC_202_0 | 102 | 0.4049 | 1 | 0.4049 | 0 | 0 | 0 | 0.7876 | Ref |
| 5282 | MF448569 | Unassigned | Unassigned | Unassigned | 202_0 | Clustered | 102 | VC_202_0 | 102 | 0.4049 | 1 | 0.4049 | 0 | 0 | 0 | 0.7876 | Ref |
| 5283 | MF448570 | Unassigned | Unassigned | Unassigned | 202_0 | Clustered | 102 | VC_202_0 | 102 | 0.4049 | 1 | 0.4049 | 0 | 0 | 0 | 0.7876 | Ref |
| 5284 | MF448571 | Unassigned | Unassigned | Unassigned | 202_0 | Clustered | 102 | VC_202_0 | 102 | 0.4049 | 1 | 0.4049 | 0 | 0 | 0 | 0.7876 | Ref |
| 5285 | MF448572 | Unassigned | Unassigned | Unassigned | 202_0 | Clustered | 102 | VC_202_0 | 102 | 0.4049 | 1 | 0.4049 | 0 | 0 | 0 | 0.7876 | Ref |
| 5286 | MF467948 | Unassigned | Unassigned | Unassigned | 872_0 | Clustered | 31 | VC_872_0 | 31 | 0.8151 | 1 | 0.8151 | 0 | 0 | 0 | 0.8194 | Ref |
| 5287 | MF467949 | Unassigned | Unassigned | Unassigned | 872_0 | Clustered | 31 | VC_872_0 | 31 | 0.8151 | 1 | 0.8151 | 0 | 0 | 0 | 0.8194 | Ref |
| 5288 | MF468274 | Unassigned | Unassigned | Unassigned | 575_0 | Clustered | 35 | VC_575_0 | 35 | 0.8815 | 1 | 0.8815 | 0 | 0 | 0 | 1 | Ref |
| 5289 | MF472892 | Unassigned | Unassigned | Unassigned | 236_0 | Clustered | 72 | VC_236_0 | 72 | 0.8104 | 1 | 0.8104 | 0 | 0 | 0 | 1 | Ref |
| 5290 | MF472894 | Unassigned | Unassigned | Unassigned | 519_0 | Clustered | 39 | VC_519_0 | 39 | 0.8156 | 1 | 0.8156 | 0 | 0 | 0 | 1 | Ref |
| 5291 | MF472896 | Unassigned | Unassigned | Unassigned | 216_6 | Clustered | 42 | VC_216_6 | 42 | 0.473 | 1 | 0.473 | 0 | 0 | 0 | 0.8351 | Ref |
| 5292 | MF476925 | Unassigned | Unassigned | Unassigned | 598_0 | Clustered | 4 | VC_598_0 | 4 | 0.0597 | 0.99355358 | 0.0593 | 0 | 0 | 0 | 1 | Ref |
| 5293 | MF479730 | Unassigned | Unassigned | Unassigned | 600_2 | Clustered | 10 | VC_600_2 | 10 | 0.3041 | 1 | 0.3041 | 0 | 0 | 0 | 1 | Ref |
| 5294 | MF481197 | Unassigned | Unassigned | Unassigned | 1027_0 | Clustered | 20 | VC_1027_0 | 20 | 0.7114 | 1 | 0.7114 | 0 | 0 | 0 | 1 | Ref |
| 5295 | MF490236 | Unassigned | Unassigned | Unassigned | 838_0 | Clustered | 29 | VC_838_0 | 29 | 0.8611 | 1 | 0.8611 | 0 | 0 | 0 | 1 | Ref |
| 5296 | MF490237 | Unassigned | Unassigned | Unassigned | 630_0 | Clustered | 20 | VC_630_0 | 20 | 0.8513 | 1 | 0.8513 | 0 | 0 | 0 | 1 | Ref |
| 5297 | MF490238 | Unassigned | Unassigned | Unassigned | 838_0 | Clustered | 29 | VC_838_0 | 29 | 0.8611 | 1 | 0.8611 | 0 | 0 | 0 | 1 | Ref |
| 5298 | MF490239 | Unassigned | Unassigned | Unassigned | 166_0 | Clustered | 26 | VC_166_0 | 26 | 0.7923 | 1 | 0.7923 | 0 | 0 | 0 | 1 | Ref |
| 5299 | MF490240 | Unassigned | Unassigned | Unassigned | 79_0 | Clustered | 64 | VC_79_0 | 64 | 0.4185 | 1 | 0.4185 | 0 | 0 | 0 | 0.7649 | Ref |
| 5300 | MF490241 | Unassigned | Unassigned | Unassigned | 79_7 | Clustered | 33 | VC_79_7 | 33 | 0.216 | 1 | 0.216 | 0 | 0 | 0 | 0.8409 | Ref |
| 5301 | MF498773 | Unassigned | Unassigned | Unassigned | 443_2 | Clustered | 10 | VC_443_2 | 10 | 0.3344 | 1 | 0.3344 | 0 | 0 | 0 | 0.9333 | Ref |
| 5302 | MF498775 | Unassigned | Unassigned | Unassigned | 443_2 | Clustered | 10 | VC_443_2 | 10 | 0.3344 | 1 | 0.3344 | 0 | 0 | 0 | 0.9333 | Ref |
| 5303 | MF498901 | Unassigned | Unassigned | Unassigned | 883_0 | Clustered | 10 | VC_883_0 | 10 | 0.2015 | 1 | 0.2015 | 0 | 0 | 0 | 1 | Ref |
| 5304 | MF537628 | Unassigned | Unassigned | Unassigned | 956_0 | Clustered | 39 | VC_956_0 | 39 | 1 | 1 | 1 | 0 | 0 | 0 | 1 | Ref |
| 5305 | MF541403 | Unassigned | Unassigned | Unassigned | 982_0 | Clustered | 62 | VC_982_0 | 62 | 0.8646 | 1 | 0.8646 | 0 | 0 | 0 | 0.743 | Ref |
| 5306 | MF541404 | Unassigned | Unassigned | Unassigned | 982_0 | Clustered | 62 | VC_982_0 | 62 | 0.8646 | 1 | 0.8646 | 0 | 0 | 0 | 0.743 | Ref |
| 5307 | MF541405 | Unassigned | Unassigned | Unassigned | 982_0 | Clustered | 62 | VC_982_0 | 62 | 0.8646 | 1 | 0.8646 | 0 | 0 | 0 | 0.743 | Ref |
| 5308 | MF541406 | Unassigned | Unassigned | Unassigned | 982_0 | Clustered | 62 | VC_982_0 | 62 | 0.8646 | 1 | 0.8646 | 0 | 0 | 0 | 0.743 | Ref |
| 5309 | MF541407 | Unassigned | Unassigned | Unassigned | 982_0 | Clustered | 62 | VC_982_0 | 62 | 0.8646 | 1 | 0.8646 | 0 | 0 | 0 | 0.743 | Ref |
| 5310 | MF541408 | Unassigned | Unassigned | Unassigned | 982_0 | Clustered | 62 | VC_982_0 | 62 | 0.8646 | 1 | 0.8646 | 0 | 0 | 0 | 0.743 | Ref |
| 5311 | MF541409 | Unassigned | Unassigned | Unassigned | 982_0 | Clustered | 62 | VC_982_0 | 62 | 0.8646 | 1 | 0.8646 | 0 | 0 | 0 | 0.743 | Ref |
| 5312 | MF541410 | Unassigned | Unassigned | Unassigned | 982_0 | Clustered | 62 | VC_982_0 | 62 | 0.8646 | 1 | 0.8646 | 0 | 0 | 0 | 0.743 | Ref |
| 5313 | MF547662 | Unassigned | Unassigned | Unassigned | 473_3 | Clustered | 2 | VC_473_3 | 2 | 0.1063 | 0.94203251 | 0.1001 | 0 | 0 | 0 | 1 | Ref |
| 5314 | MF547663 | Unassigned | Unassigned | Unassigned | 473_3 | Clustered | 2 | VC_473_3 | 2 | 0.1063 | 0.94203251 | 0.1001 | 0 | 0 | 0 | 1 | Ref |
| 5315 | MF564201 | Unassigned | Unassigned | Unassigned | 942_0 | Clustered | 36 | VC_942_0 | 36 | 0.7848 | 1 | 0.7848 | 0 | 0 | 0 | 1 | Ref |
| 5316 | MF574151 | Unassigned | Unassigned | Unassigned | 82_0 | Clustered | 24 | VC_82_0 | 24 | 0.7351 | 1 | 0.7351 | 0 | 0 | 0 | 1 | Ref |
| 5317 | MF580410 | Unassigned | Unassigned | Unassigned | 8_0 | Clustered | 58 | VC_8_0 | 58 | 0.8306 | 1 | 0.8306 | 0 | 0 | 0 | 0.9946 | Ref |
| 5318 | MF580759 | Unassigned | Unassigned | Unassigned | 252_0 | Clustered | 121 | VC_252_0 | 121 | 0.7376 | 1 | 0.7376 | 0 | 0 | 0 | 0.9999 | Ref |
| 5319 | MF580760 | Unassigned | Unassigned | Unassigned | 252_0 | Clustered | 121 | VC_252_0 | 121 | 0.7376 | 1 | 0.7376 | 0 | 0 | 0 | 0.9999 | Ref |
| 5320 | MF580761 | Unassigned | Unassigned | Unassigned | 252_0 | Clustered | 121 | VC_252_0 | 121 | 0.7376 | 1 | 0.7376 | 0 | 0 | 0 | 0.9999 | Ref |
| 5321 | MF580762 | Unassigned | Unassigned | Unassigned | 252_0 | Clustered | 121 | VC_252_0 | 121 | 0.7376 | 1 | 0.7376 | 0 | 0 | 0 | 0.9999 | Ref |
| 5322 | MF580763 | Unassigned | Unassigned | Unassigned | 252_0 | Clustered | 121 | VC_252_0 | 121 | 0.7376 | 1 | 0.7376 | 0 | 0 | 0 | 0.9999 | Ref |
| 5323 | MF580764 | Unassigned | Unassigned | Unassigned | 252_0 | Clustered | 121 | VC_252_0 | 121 | 0.7376 | 1 | 0.7376 | 0 | 0 | 0 | 0.9999 | Ref |
| 5324 | MF580765 | Unassigned | Unassigned | Unassigned | 252_0 | Clustered | 121 | VC_252_0 | 121 | 0.7376 | 1 | 0.7376 | 0 | 0 | 0 | 0.9999 | Ref |
| 5325 | MF580766 | Unassigned | Unassigned | Unassigned | 252_0 | Clustered | 121 | VC_252_0 | 121 | 0.7376 | 1 | 0.7376 | 0 | 0 | 0 | 0.9999 | Ref |
| 5326 | MF580767 | Unassigned | Unassigned | Unassigned | 252_0 | Clustered | 121 | VC_252_0 | 121 | 0.7376 | 1 | 0.7376 | 0 | 0 | 0 | 0.9999 | Ref |
| 5327 | MF580768 | Unassigned | Unassigned | Unassigned | 252_0 | Clustered | 121 | VC_252_0 | 121 | 0.7376 | 1 | 0.7376 | 0 | 0 | 0 | 0.9999 | Ref |
| 5328 | MF580769 | Unassigned | Unassigned | Unassigned | 252_0 | Clustered | 121 | VC_252_0 | 121 | 0.7376 | 1 | 0.7376 | 0 | 0 | 0 | 0.9999 | Ref |
| 5329 | MF580770 | Unassigned | Unassigned | Unassigned | 252_0 | Clustered | 121 | VC_252_0 | 121 | 0.7376 | 1 | 0.7376 | 0 | 0 | 0 | 0.9999 | Ref |
| 5330 | MF580771 | Unassigned | Unassigned | Unassigned | 252_0 | Clustered | 121 | VC_252_0 | 121 | 0.7376 | 1 | 0.7376 | 0 | 0 | 0 | 0.9999 | Ref |
| 5331 | MF580772 | Unassigned | Unassigned | Unassigned | 252_0 | Clustered | 121 | VC_252_0 | 121 | 0.7376 | 1 | 0.7376 | 0 | 0 | 0 | 0.9999 | Ref |
| 5332 | MF580773 | Unassigned | Unassigned | Unassigned | 252_0 | Clustered | 121 | VC_252_0 | 121 | 0.7376 | 1 | 0.7376 | 0 | 0 | 0 | 0.9999 | Ref |
| 5333 | MF580774 | Unassigned | Unassigned | Unassigned | 252_0 | Clustered | 121 | VC_252_0 | 121 | 0.7376 | 1 | 0.7376 | 0 | 0 | 0 | 0.9999 | Ref |
| 5334 | MF580775 | Unassigned | Unassigned | Unassigned | 252_0 | Clustered | 121 | VC_252_0 | 121 | 0.7376 | 1 | 0.7376 | 0 | 0 | 0 | 0.9999 | Ref |
| 5335 | MF580955 | Unassigned | Unassigned | Unassigned | 1215_0 | Clustered | 3 | VC_1215_0 | 3 | 1 | 1 | 1 | 0 | 0 | 0 | 1 | Ref |
| 5336 | MF580956 | Unassigned | Unassigned | Unassigned | 1216_0 | Clustered | 3 | VC_1216_0 | 3 | 0.8134 | 0.99225442 | 0.8071 | 0 | 0 | 0 | 1 | Ref |
| 5337 | MF580957 | Unassigned | Unassigned | Unassigned | 1215_0 | Clustered | 3 | VC_1215_0 | 3 | 1 | 1 | 1 | 0 | 0 | 0 | 1 | Ref |
| 5338 | MF580958 | Unassigned | Unassigned | Unassigned | 1215_0 | Clustered | 3 | VC_1215_0 | 3 | 1 | 1 | 1 | 0 | 0 | 0 | 1 | Ref |
| 5339 | MF580959 | Unassigned | Unassigned | Unassigned | 1216_0 | Clustered | 3 | VC_1216_0 | 3 | 0.8134 | 0.99225442 | 0.8071 | 0 | 0 | 0 | 1 | Ref |
| 5340 | MF580960 | Unassigned | Unassigned | Unassigned | 1216_0 | Clustered | 3 | VC_1216_0 | 3 | 0.8134 | 0.99225442 | 0.8071 | 0 | 0 | 0 | 1 | Ref |
| 5341 | MF580961 | Unassigned | Unassigned | Unassigned | 1217_0 | Clustered | 3 | VC_1217_0 | 3 | 0.7992 | 0.99216275 | 0.793 | 0 | 0 | 0 | 1 | Ref |
| 5342 | MF580962 | Unassigned | Unassigned | Unassigned | 1217_0 | Clustered | 3 | VC_1217_0 | 3 | 0.7992 | 0.99216275 | 0.793 | 0 | 0 | 0 | 1 | Ref |
| 5343 | MF595878 | Unassigned | Unassigned | Unassigned | 734_0 | Clustered | 6 | VC_734_0 | 6 | 0.1914 | 0.99996888 | 0.1914 | 0 | 0 | 0 | 1 | Ref |
| 5344 | MF612071 | Unassigned | Unassigned | Unassigned | 111_0 | Clustered | 91 | VC_111_0 | 91 | 0.9587 | 1 | 0.9587 | 0 | 0 | 0 | 1 | Ref |

| | | | | | | | | | | | | | | | | | |
|---|---|---|---|---|---|---|---|---|---|---|---|---|---|---|---|---|---|
| 5345 | MF612072 | Unassigned | Unassigned | Unassigned | 938_0 | Clustered | 99 | VC_938_0 | 99 | 0.9119 | 1 | 0.9119 | 0 | 0 | 0 | 0.9928 | Ref |
| 5346 | MF612073 | Unassigned | Unassigned | Unassigned | 359_0 | Clustered | 7 | VC_359_0 | 7 | 0.3703 | 1 | 0.3703 | 0 | 0 | 0 | 1 | Ref |
| 5347 | MF614100 | Unassigned | Unassigned | Unassigned | 705_0 | Clustered | 11 | VC_705_0 | 11 | 0.8861 | 1 | 0.8861 | 0 | 0 | 0 | 1 | Ref |
| 5348 | MF614627 | Unassigned | Unassigned | Unassigned | 21_0 | Clustered | 4 | VC_21_0 | 4 | 0.5696 | 0.87262442 | 0.4971 | 0 | 0 | 0 | 0.6667 | Ref |
| 5349 | MF614628 | Unassigned | Unassigned | Unassigned | 21_0 | Clustered | 4 | VC_21_0 | 4 | 0.5696 | 0.87262442 | 0.4971 | 0 | 0 | 0 | 0.6667 | Ref |
| 5350 | MF621978 | Unassigned | Unassigned | Unassigned | 70_0 | Clustered | 27 | VC_70_0 | 27 | 0.5981 | 1 | 0.5981 | 0 | 0 | 0 | 0.9886 | Ref |
| 5351 | MF623055 | Unassigned | Unassigned | Unassigned | 79_4 | Clustered | 3 | VC_79_4 | 3 | 0.0106 | 0.99632092 | 0.0105 | 0 | 0 | 0 | 0.6667 | Ref |
| 5352 | MF629150 | Unassigned | Unassigned | Unassigned | 1217_0 | Clustered | 3 | VC_1217_0 | 3 | 0.7992 | 0.99216275 | 0.793 | 0 | 0 | 0 | 1 | Ref |
| 5353 | MF630921 | Unassigned | Unassigned | Unassigned | 451_0 | Clustered | 80 | VC_451_0 | 80 | 0.9991 | 1 | 0.9991 | 0 | 0 | 0 | 1 | Ref |
| 5354 | MF663761 | Unassigned | Unassigned | Unassigned | 123_0 | Clustered | 23 | VC_123_0 | 23 | 0.817 | 1 | 0.817 | 0 | 0 | 0 | 1 | Ref |
| 5355 | MF663786 | Unassigned | Unassigned | Unassigned | 127_0 | Clustered/Singleton | 1 | VC_127_0 | 1 | 0 | 1 | 0 | 0 | 0 | 0 | 1 | Ref |
| 5356 | MF668266 | Unassigned | Unassigned | Unassigned | 1167_0 | Clustered | 6 | VC_1167_0 | 6 | 0.8642 | 0.99999946 | 0.8642 | 0 | 0 | 0 | 1 | Ref |
| 5357 | MF668267 | Unassigned | Unassigned | Unassigned | 236_0 | Clustered | 72 | VC_236_0 | 72 | 0.8104 | 1 | 0.8104 | 0 | 0 | 0 | 1 | Ref |
| 5358 | MF668268 | Unassigned | Unassigned | Unassigned | 761_0 | Clustered | 67 | VC_761_0 | 67 | 0.8947 | 1 | 0.8947 | 0 | 0 | 0 | 1 | Ref |
| 5359 | MF668269 | Unassigned | Unassigned | Unassigned | 216_4 | Clustered | 13 | VC_216_4 | 13 | 0.1657 | 1 | 0.1657 | 0 | 0 | 0 | 1 | Ref |
| 5360 | MF668270 | Unassigned | Unassigned | Unassigned | 520_15 | Clustered | 2 | VC_520_15 | 2 | 0.0078 | 0.957604 | 0.0075 | 0 | 0 | 0 | 0 | Ref |
| 5361 | MF668272 | Unassigned | Unassigned | Unassigned | 761_0 | Clustered | 67 | VC_761_0 | 67 | 0.8947 | 1 | 0.8947 | 0 | 0 | 0 | 1 | Ref |
| 5362 | MF668273 | Unassigned | Unassigned | Unassigned | 1167_0 | Clustered | 6 | VC_1167_0 | 6 | 0.8642 | 0.99999946 | 0.8642 | 0 | 0 | 0 | 1 | Ref |
| 5363 | MF668274 | Unassigned | Unassigned | Unassigned | 1074_0 | Clustered | 19 | VC_1074_0 | 19 | 0.9198 | 1 | 0.9198 | 0 | 0 | 0 | 1 | Ref |
| 5364 | MF668275 | Unassigned | Unassigned | Unassigned | 1159_0 | Clustered | 3 | VC_1159_0 | 3 | 0.3304 | 0.98548359 | 0.3256 | 0 | 0 | 0 | 1 | Ref |
| 5365 | MF668276 | Unassigned | Unassigned | Unassigned | 371_11 | Clustered | 31 | VC_371_11 | 31 | 0.0927 | 1 | 0.0927 | 0 | 0 | 0 | 0.7011 | Ref |
| 5366 | MF668277 | Unassigned | Unassigned | Unassigned | 551_7 | Clustered | 28 | VC_551_7 | 28 | 0.1661 | 1 | 0.1661 | 0 | 0 | 0 | 0.8519 | Ref |
| 5367 | MF668278 | Unassigned | Unassigned | Unassigned | 517_0 | Clustered | 18 | VC_517_0 | 18 | 0.1599 | 1 | 0.1599 | 0 | 0 | 0 | 1 | Ref |
| 5368 | MF668279 | Unassigned | Unassigned | Unassigned | 1167_0 | Clustered | 6 | VC_1167_0 | 6 | 0.8642 | 0.99999946 | 0.8642 | 0 | 0 | 0 | 1 | Ref |
| 5369 | MF668280 | Unassigned | Unassigned | Unassigned | 559_0 | Clustered | 2 | VC_559_0 | 2 | 0.0112 | 0.95651281 | 0.0107 | 0 | 0 | 0 | 1 | Ref |
| 5370 | MF668282 | Unassigned | Unassigned | Unassigned | 224_0 | Clustered | 19 | VC_224_0 | 19 | 0.9416 | 1 | 0.9416 | 0 | 0 | 0 | 1 | Ref |
| 5371 | MF668283 | Unassigned | Unassigned | Unassigned | 325_35 | Clustered | 17 | VC_325_35 | 17 | 0.0732 | 1 | 0.0732 | 0 | 0 | 0 | 0.7721 | Ref |
| 5372 | MF668284 | Unassigned | Unassigned | Unassigned | 546_3 | Clustered | 4 | VC_546_3 | 4 | 0.0418 | 0.92194988 | 0.0385 | 0 | 0 | 0 | 1 | Ref |
| 5373 | MF668285 | Unassigned | Unassigned | Unassigned | 1160_0 | Clustered | 73 | VC_1160_0 | 73 | 0.9835 | 1 | 0.9835 | 0 | 0 | 0 | 1 | Ref |
| 5374 | MF668287 | Unassigned | Unassigned | Unassigned | 520_3 | Clustered/Singleton | 1 | VC_520_3 | 1 | 0 | 1 | 0 | 0 | 0 | 0 | 1 | Ref |
| 5375 | MF668288 | Unassigned | Unassigned | Unassigned | 223_0 | Clustered | 38 | VC_223_0 | 38 | 0.9095 | 1 | 0.9095 | 0 | 0 | 0 | 1 | Ref |
| 5376 | MF678788 | Unassigned | Unassigned | Unassigned | 695_0 | Clustered | 48 | VC_695_0 | 48 | 0.8162 | 1 | 0.8162 | 0 | 0 | 0 | 1 | Ref |
| 5377 | MF678789 | Unassigned | Unassigned | Unassigned | 695_0 | Clustered | 48 | VC_695_0 | 48 | 0.8162 | 1 | 0.8162 | 0 | 0 | 0 | 1 | Ref |
| 5378 | MF678790 | Unassigned | Unassigned | Unassigned | 695_0 | Clustered | 48 | VC_695_0 | 48 | 0.8162 | 1 | 0.8162 | 0 | 0 | 0 | 1 | Ref |
| 5379 | MF681663 | Unassigned | Unassigned | Unassigned | 191_11 | Clustered/Singleton | 1 | VC_191_11 | 1 | 0 | 1 | 0 | 0 | 0 | 0 | 1 | Ref |
| 5380 | MF683623 | Unassigned | Unassigned | Unassigned | 878_0 | Clustered | 32 | VC_878_0 | 32 | 0.9965 | 1 | 0.9965 | 0 | 0 | 0 | 1 | Ref |
| 5381 | MF695815 | Unassigned | Unassigned | Unassigned | 243_0 | Clustered | 2 | VC_243_0 | 2 | 0.0637 | 0.95609892 | 0.0609 | 0 | 0 | 0 | 1 | Ref |
| 5382 | MF716957 | Unassigned | Unassigned | Unassigned | 52_0 | Clustered | 87 | VC_52_0 | 87 | 0.9858 | 1 | 0.9858 | 0 | 0 | 0 | 1 | Ref |
| 5383 | MF740800 | Unassigned | Unassigned | Unassigned | 428_0 | Clustered | 33 | VC_428_0 | 33 | 0.9915 | 1 | 0.9915 | 0 | 0 | 0 | 1 | Ref |
| 5384 | MF754111 | Unassigned | Unassigned | Unassigned | 837_0 | Clustered | 29 | VC_837_0 | 29 | 0.9955 | 1 | 0.9955 | 0 | 0 | 0 | 1 | Ref |
| 5385 | MF754112 | Unassigned | Unassigned | Unassigned | 837_0 | Clustered | 29 | VC_837_0 | 29 | 0.9955 | 1 | 0.9955 | 0 | 0 | 0 | 1 | Ref |
| 5386 | MF754113 | Unassigned | Unassigned | Unassigned | 882_0 | Clustered | 20 | VC_882_0 | 20 | 0.9917 | 1 | 0.9917 | 0 | 0 | 0 | 1 | Ref |
| 5387 | MF754114 | Unassigned | Unassigned | Unassigned | 882_0 | Clustered | 20 | VC_882_0 | 20 | 0.9917 | 1 | 0.9917 | 0 | 0 | 0 | 1 | Ref |
| 5388 | MF754115 | Unassigned | Unassigned | Unassigned | 882_0 | Clustered | 20 | VC_882_0 | 20 | 0.9917 | 1 | 0.9917 | 0 | 0 | 0 | 1 | Ref |
| 5389 | MF754116 | Unassigned | Unassigned | Unassigned | 882_0 | Clustered | 20 | VC_882_0 | 20 | 0.9917 | 1 | 0.9917 | 0 | 0 | 0 | 1 | Ref |
| 5390 | MF765814 | Unassigned | Unassigned | Unassigned | 883_1 | Clustered | 12 | VC_883_1 | 12 | 0.2441 | 1 | 0.2441 | 0 | 0 | 0 | 1 | Ref |
| 5391 | MF766044 | Unassigned | Unassigned | Unassigned | 981_1 | Clustered | 26 | VC_981_1 | 26 | 0.7009 | 1 | 0.7009 | 0 | 0 | 0 | 1 | Ref |
| 5392 | MF766045 | Unassigned | Unassigned | Unassigned | 981_1 | Clustered | 26 | VC_981_1 | 26 | 0.7009 | 1 | 0.7009 | 0 | 0 | 0 | 1 | Ref |
| 5393 | MF766046 | Unassigned | Unassigned | Unassigned | 981_1 | Clustered | 26 | VC_981_1 | 26 | 0.7009 | 1 | 0.7009 | 0 | 0 | 0 | 1 | Ref |
| 5394 | MF766047 | Unassigned | Unassigned | Unassigned | 981_1 | Clustered | 26 | VC_981_1 | 26 | 0.7009 | 1 | 0.7009 | 0 | 0 | 0 | 1 | Ref |
| 5395 | MF766048 | Unassigned | Unassigned | Unassigned | 981_1 | Clustered | 26 | VC_981_1 | 26 | 0.7009 | 1 | 0.7009 | 0 | 0 | 0 | 1 | Ref |
| 5396 | MF768469 | Unassigned | Unassigned | Unassigned | 472_0 | Clustered | 47 | VC_472_0 | 47 | 0.9025 | 1 | 0.9025 | 0 | 0 | 0 | 1 | Ref |
| 5397 | MF768470 | Unassigned | Unassigned | Unassigned | 79_7 | Clustered | 33 | VC_79_7 | 33 | 0.216 | 1 | 0.216 | 0 | 0 | 0 | 0.8409 | Ref |
| 5398 | MF773750 | Unassigned | Unassigned | Unassigned | 515_0 | Clustered | 16 | VC_515_0 | 16 | 0.3793 | 1 | 0.3793 | 0 | 0 | 0 | 1 | Ref |
| 5399 | MF774688 | Unassigned | Unassigned | Unassigned | 190_30 | Clustered/Singleton | 1 | VC_190_30 | 1 | 0 | 1 | 0 | 0 | 0 | 0 | 1 | Ref |
| 5400 | MF775703 | Unassigned | Unassigned | Unassigned | 202_0 | Clustered | 102 | VC_202_0 | 102 | 0.4049 | 1 | 0.4049 | 0 | 0 | 0 | 0.7876 | Ref |
| 5401 | MF775704 | Unassigned | Unassigned | Unassigned | 202_0 | Clustered | 102 | VC_202_0 | 102 | 0.4049 | 1 | 0.4049 | 0 | 0 | 0 | 0.7876 | Ref |
| 5402 | MF775705 | Unassigned | Unassigned | Unassigned | 202_0 | Clustered | 102 | VC_202_0 | 102 | 0.4049 | 1 | 0.4049 | 0 | 0 | 0 | 0.7876 | Ref |
| 5403 | MF787246 | Unassigned | Unassigned | Unassigned | 526_1 | Clustered/Singleton | 1 | VC_526_1 | 1 | 0 | 1 | 0 | 0 | 0 | 0 | 1 | Ref |
| 5404 | MF788075 | Unassigned | Unassigned | Unassigned | 755_0 | Clustered | 9 | VC_755_0 | 9 | 0.4489 | 1 | 0.4489 | 0 | 0 | 0 | 1 | Ref |
| 5405 | MF805716 | Unassigned | Unassigned | Unassigned | 344_0 | Clustered | 21 | VC_344_0 | 21 | 0.5436 | 1 | 0.5436 | 0 | 0 | 0 | 1 | Ref |
| 5406 | MF805809 | Unassigned | Unassigned | Unassigned | 848_0 | Clustered | 28 | VC_848_0 | 28 | 0.7927 | 1 | 0.7927 | 0 | 0 | 0 | 1 | Ref |
| 5407 | MF807953 | Unassigned | Unassigned | Unassigned | 51_0 | Clustered | 10 | VC_51_0 | 10 | 0.1741 | 1 | 0.1741 | 0 | 0 | 0 | 1 | Ref |
| 5408 | MF893340 | Unassigned | Unassigned | Unassigned | 78_6 | Clustered/Singleton | 1 | VC_78_6 | 1 | 0 | 1 | 0 | 0 | 0 | 0 | 1 | Ref |
| 5409 | MF919490 | Unassigned | Unassigned | Unassigned | 216_6 | Clustered | 42 | VC_216_6 | 42 | 0.473 | 1 | 0.473 | 0 | 0 | 0 | 0.8351 | Ref |
| 5410 | MF919491 | Unassigned | Unassigned | Unassigned | 765_0 | Clustered | 132 | VC_765_0 | 132 | 1 | 1 | 1 | 0 | 0 | 0 | 1 | Ref |
| 5411 | MF919492 | Unassigned | Unassigned | Unassigned | 519_0 | Clustered | 39 | VC_519_0 | 39 | 0.8156 | 1 | 0.8156 | 0 | 0 | 0 | 1 | Ref |
| 5412 | MF919493 | Unassigned | Unassigned | Unassigned | 558_20 | Clustered | 17 | VC_558_20 | 17 | 0.054 | 1 | 0.054 | 0 | 0 | 0 | 0.9926 | Ref |
| 5413 | MF919494 | Unassigned | Unassigned | Unassigned | 558_55 | Clustered/Singleton | 1 | VC_558_55 | 1 | 0 | 1 | 0 | 0 | 0 | 0 | 1 | Ref |
| 5414 | MF919495 | Unassigned | Unassigned | Unassigned | 558_74 | Clustered/Singleton | 1 | VC_558_74 | 1 | 0 | 1 | 0 | 0 | 0 | 0 | 1 | Ref |
| 5415 | MF919496 | Unassigned | Unassigned | Unassigned | 517_1 | Clustered | 108 | VC_517_1 | 108 | 0.8483 | 1 | 0.8483 | 0 | 0 | 0 | 0.9955 | Ref |
| 5416 | MF919497 | Unassigned | Unassigned | Unassigned | 761_0 | Clustered | 67 | VC_761_0 | 67 | 0.8947 | 1 | 0.8947 | 0 | 0 | 0 | 1 | Ref |
| 5417 | MF919499 | Unassigned | Unassigned | Unassigned | 558_50 | Clustered | 15 | VC_558_50 | 15 | 0.0468 | 1 | 0.0468 | 0 | 0 | 0 | 0.9238 | Ref |
| 5418 | MF919500 | Unassigned | Unassigned | Unassigned | 216_6 | Clustered | 42 | VC_216_6 | 42 | 0.473 | 1 | 0.473 | 0 | 0 | 0 | 0.8351 | Ref |
| 5419 | MF919501 | Unassigned | Unassigned | Unassigned | 520_63 | Clustered/Singleton | 1 | VC_520_63 | 1 | 0 | 1 | 0 | 0 | 0 | 0 | 1 | Ref |
| 5420 | MF919503 | Unassigned | Unassigned | Unassigned | 371_3 | Clustered | 22 | VC_371_3 | 22 | 0.0787 | 1 | 0.0787 | 0 | 0 | 0 | 0.9048 | Ref |
| 5421 | MF919504 | Unassigned | Unassigned | Unassigned | 546_5 | Clustered/Singleton | 1 | VC_546_5 | 1 | 0 | 1 | 0 | 0 | 0 | 0 | 1 | Ref |
| 5422 | MF919505 | Unassigned | Unassigned | Unassigned | 765_0 | Clustered | 132 | VC_765_0 | 132 | 1 | 1 | 1 | 0 | 0 | 0 | 1 | Ref |
| 5423 | MF919506 | Unassigned | Unassigned | Unassigned | 551_8 | Clustered | 19 | VC_551_8 | 19 | 0.1039 | 1 | 0.1039 | 0 | 0 | 0 | 0.8713 | Ref |
| 5424 | MF919507 | Unassigned | Unassigned | Unassigned | 371_3 | Clustered | 22 | VC_371_3 | 22 | 0.0787 | 1 | 0.0787 | 0 | 0 | 0 | 0.9048 | Ref |
| 5425 | MF919508 | Unassigned | Unassigned | Unassigned | 325_56 | Clustered | 2 | VC_325_56 | 2 | 0.0036 | 0.94604798 | 0.0034 | 0 | 0 | 0 | 0 | Ref |
| 5426 | MF919509 | Unassigned | Unassigned | Unassigned | 517_1 | Clustered | 108 | VC_517_1 | 108 | 0.8483 | 1 | 0.8483 | 0 | 0 | 0 | 0.9955 | Ref |
| 5427 | MF919510 | Unassigned | Unassigned | Unassigned | 767_0 | Clustered | 34 | VC_767_0 | 34 | 0.9173 | 1 | 0.9173 | 0 | 0 | 0 | 1 | Ref |
| 5428 | MF919511 | Unassigned | Unassigned | Unassigned | 371_13 | Clustered | 33 | VC_371_13 | 33 | 0.179 | 1 | 0.179 | 0 | 0 | 0 | 0.8769 | Ref |
| 5429 | MF919512 | Unassigned | Unassigned | Unassigned | 546_6 | Clustered | 2 | VC_546_6 | 2 | 0.0135 | 0.7810929 | 0.0105 | 0 | 0 | 0 | 1 | Ref |
| 5430 | MF919513 | Unassigned | Unassigned | Unassigned | 558_77 | Clustered | 3 | VC_558_77 | 3 | 0.0065 | 0.90877939 | 0.0059 | 0 | 0 | 0 | 0.6667 | Ref |
| 5431 | MF919514 | Unassigned | Unassigned | Unassigned | 1093_0 | Clustered | 40 | VC_1093_0 | 40 | 0.8754 | 1 | 0.8754 | 0 | 0 | 0 | 1 | Ref |
| 5432 | MF919515 | Unassigned | Unassigned | Unassigned | 765_0 | Clustered | 132 | VC_765_0 | 132 | 1 | 1 | 1 | 0 | 0 | 0 | 1 | Ref |
| 5433 | MF919516 | Unassigned | Unassigned | Unassigned | 765_0 | Clustered | 132 | VC_765_0 | 132 | 1 | 1 | 1 | 0 | 0 | 0 | 1 | Ref |
| 5434 | MF919517 | Unassigned | Unassigned | Unassigned | 810_0 | Clustered | 18 | VC_810_0 | 18 | 0.9121 | 1 | 0.9121 | 0 | 0 | 0 | 1 | Ref |
| 5435 | MF919519 | Unassigned | Unassigned | Unassigned | 371_6 | Clustered/Singleton | 1 | VC_371_6 | 1 | 0 | 1 | 0 | 0 | 0 | 0 | 1 | Ref |
| 5436 | MF919520 | Unassigned | Unassigned | Unassigned | 1181_1 | Clustered | 10 | VC_1181_1 | 10 | 0.5828 | 1 | 0.5828 | 0 | 0 | 0 | 1 | Ref |
| 5437 | MF919521 | Unassigned | Unassigned | Unassigned | 1176_0 | Clustered | 41 | VC_1176_0 | 41 | 0.9558 | 1 | 0.9558 | 0 | 0 | 0 | 0.9976 | Ref |
| 5438 | MF919522 | Unassigned | Unassigned | Unassigned | 765_0 | Clustered | 132 | VC_765_0 | 132 | 1 | 1 | 1 | 0 | 0 | 0 | 1 | Ref |
| 5439 | MF919523 | Unassigned | Unassigned | Unassigned | 371_3 | Clustered | 22 | VC_371_3 | 22 | 0.0787 | 1 | 0.0787 | 0 | 0 | 0 | 0.9048 | Ref |
| 5440 | MF919524 | Unassigned | Unassigned | Unassigned | 551_8 | Clustered | 19 | VC_551_8 | 19 | 0.1039 | 1 | 0.1039 | 0 | 0 | 0 | 0.8713 | Ref |
| 5441 | MF919525 | Unassigned | Unassigned | Unassigned | 551_8 | Clustered | 19 | VC_551_8 | 19 | 0.1039 | 1 | 0.1039 | 0 | 0 | 0 | 0.8713 | Ref |
| 5442 | MF919526 | Unassigned | Unassigned | Unassigned | 520_81 | Clustered/Singleton | 1 | VC_520_81 | 1 | 0 | 1 | 0 | 0 | 0 | 0 | 1 | Ref |
| 5443 | MF919527 | Unassigned | Unassigned | Unassigned | 558_82 | Clustered | 3 | VC_558_82 | 3 | 0.0064 | 0.88433874 | 0.0057 | 0 | 0 | 0 | 0.6667 | Ref |
| 5444 | MF919528 | Unassigned | Unassigned | Unassigned | 371_11 | Clustered | 31 | VC_371_11 | 31 | 0.0927 | 1 | 0.0927 | 0 | 0 | 0 | 0.7011 | Ref |
| 5445 | MF919529 | Unassigned | Unassigned | Unassigned | 551_8 | Clustered | 19 | VC_551_8 | 19 | 0.1039 | 1 | 0.1039 | 0 | 0 | 0 | 0.8713 | Ref |
| 5446 | MF919530 | Unassigned | Unassigned | Unassigned | 371_3 | Clustered | 22 | VC_371_3 | 22 | 0.0787 | 1 | 0.0787 | 0 | 0 | 0 | 0.9048 | Ref |
| 5447 | MF919531 | Unassigned | Unassigned | Unassigned | 216_6 | Clustered | 42 | VC_216_6 | 42 | 0.473 | 1 | 0.473 | 0 | 0 | 0 | 0.8351 | Ref |
| 5448 | MF919532 | Unassigned | Unassigned | Unassigned | 236_0 | Clustered | 72 | VC_236_0 | 72 | 0.8104 | 1 | 0.8104 | 0 | 0 | 0 | 1 | Ref |
| 5449 | MF919533 | Unassigned | Unassigned | Unassigned | 765_0 | Clustered | 132 | VC_765_0 | 132 | 1 | 1 | 1 | 0 | 0 | 0 | 1 | Ref |
| 5450 | MF919534 | Unassigned | Unassigned | Unassigned | 546_2 | Clustered | 3 | VC_546_2 | 3 | 0.0275 | 0.85260342 | 0.0234 | 0 | 0 | 0 | 1 | Ref |
| 5451 | MF919535 | Unassigned | Unassigned | Unassigned | 551_8 | Clustered | 19 | VC_551_8 | 19 | 0.1039 | 1 | 0.1039 | 0 | 0 | 0 | 0.8713 | Ref |
| 5452 | MF919536 | Unassigned | Unassigned | Unassigned | 215_2 | Clustered | 5 | VC_215_2 | 5 | 0.0572 | 0.99999997 | 0.0572 | 0 | 0 | 0 | 1 | Ref |
| 5453 | MF919537 | Unassigned | Unassigned | Unassigned | 1181_1 | Clustered | 10 | VC_1181_1 | 10 | 0.5828 | 1 | 0.5828 | 0 | 0 | 0 | 1 | Ref |
| 5454 | MF919538 | Unassigned | Unassigned | Unassigned | 216_4 | Clustered | 13 | VC_216_4 | 13 | 0.1657 | 1 | 0.1657 | 0 | 0 | 0 | 1 | Ref |
| 5455 | MF919539 | Unassigned | Unassigned | Unassigned | 371_11 | Clustered | 31 | VC_371_11 | 31 | 0.0927 | 1 | 0.0927 | 0 | 0 | 0 | 0.7011 | Ref |
| 5456 | MF919540 | Unassigned | Unassigned | Unassigned | 551_8 | Clustered | 19 | VC_551_8 | 19 | 0.1039 | 1 | 0.1039 | 0 | 0 | 0 | 0.8713 | Ref |
| 5457 | MF919541 | Unassigned | Unassigned | Unassigned | 551_1 | Clustered | 4 | VC_551_1 | 4 | 0.0159 | 0.99968337 | 0.0159 | 0 | 0 | 0 | 0.6667 | Ref |
| 5458 | MF919542 | Unassigned | Unassigned | Unassigned | 767_0 | Clustered | 34 | VC_767_0 | 34 | 0.9173 | 1 | 0.9173 | 0 | 0 | 0 | 1 | Ref |
| 5459 | MF959999 | Unassigned | Unassigned | Unassigned | 185_0 | Clustered | 3 | VC_185_0 | 3 | 0.4487 | 0.97148006 | 0.4359 | 0 | 0 | 0 | 1 | Ref |
| 5460 | MF974178 | Unassigned | Unassigned | Unassigned | 76_0 | Clustered | 37 | VC_76_0 | 37 | 0.9403 | 1 | 0.9403 | 0 | 0 | 0 | 1 | Ref |
| 5461 | MF974396 | Unassigned | Unassigned | Unassigned | 688_0 | Clustered | 2 | VC_688_0 | 2 | 0.9946 | 0.5 | 0.4973 | 0 | 0 | 0 | 1 | Ref |
| 5462 | MF974397 | Unassigned | Unassigned | Unassigned | 688_0 | Clustered | 2 | VC_688_0 | 2 | 0.9946 | 0.5 | 0.4973 | 0 | 0 | 0 | 1 | Ref |
| 5463 | MF975638 | Unassigned | Unassigned | Unassigned | 975_0 | Clustered | 11 | VC_975_0 | 11 | 0.9687 | 1 | 0.9687 | 0 | 0 | 0 | 1 | Ref |
| 5464 | MF975720 | Unassigned | Unassigned | Unassigned | 241_0 | Clustered | 2 | VC_241_0 | 2 | 0.072 | 0.95433582 | 0.0687 | 0 | 0 | 0 | 1 | Ref |
| 5465 | MF979560 | Unassigned | Unassigned | Unassigned | 849_3 | Clustered | 2 | VC_849_3 | 2 | 0.0243 | 0.85188229 | 0.0207 | 0 | 0 | 0 | 1 | Ref |
| 5466 | MF979561 | Unassigned | Unassigned | Unassigned | 389_0 | Clustered | 3 | VC_389_0 | 3 | 0.0515 | 0.99861557 | 0.0514 | 0 | 0 | 0 | 1 | Ref |
| 5467 | MF979562 | Unassigned | Unassigned | Unassigned | 1218_0 | Clustered | 4 | VC_1218_0 | 4 | 0.8905 | 0.99530591 | 0.8863 | 0 | 0 | 0 | 1 | Ref |
| 5468 | MF979563 | Unassigned | Unassigned | Unassigned | 849_3 | Clustered | 2 | VC_849_3 | 2 | 0.0243 | 0.85188229 | 0.0207 | 0 | 0 | 0 | 1 | Ref |
| 5469 | MF996376 | Unassigned | Unassigned | Unassigned | 576_0 | Clustered | 4 | VC_576_0 | 4 | 0.0606 | 0.99998766 | 0.0606 | 0 | 0 | 0 | 1 | Ref |
| 5470 | MF999224 | Unassigned | Unassigned | Unassigned | 272_0 | Clustered | 11 | VC_272_0 | 11 | 0.4915 | 1 | 0.4915 | 0 | 0 | 0 | 0.8727 | Ref |
| 5471 | MG000860 | Unassigned | Unassigned | Unassigned | 85_3 | Clustered | 19 | VC_85_3 | 19 | 0.3671 | 1 | 0.3671 | 0 | 0 | 0 | 0.807 | Ref |
| 5472 | MG004687 | Unassigned | Unassigned | Unassigned | 452_0 | Clustered | 66 | VC_452_0 | 66 | 0.9248 | 1 | 0.9248 | 0 | 0 | 0 | 1 | Ref |
| 5473 | MG018224 | Unassigned | Unassigned | Unassigned | 169_0 | Clustered | 20 | VC_169_0 | 20 | 0.7529 | 1 | 0.7529 | 0 | 0 | 0 | 1 | Ref |
| 5474 | MG018927 | Unassigned | Unassigned | Unassigned | 725_0 | Clustered | 4 | VC_725_0 | 4 | 0.4268 | 0.99997355 | 0.4268 | 0 | 0 | 0 | 1 | Ref |
| 5475 | MG018928 | Unassigned | Unassigned | Unassigned | 779_1 | Clustered | 11 | VC_779_1 | 11 | 0.1587 | 1 | 0.1587 | 0 | 0 | 0 | 1 | Ref |
| 5476 | MG018929 | Unassigned | Unassigned | Unassigned | 1027_0 | Clustered | 20 | VC_1027_0 | 20 | 0.7114 | 1 | 0.7114 | 0 | 0 | 0 | 1 | Ref |
| 5477 | MG018930 | Unassigned | Unassigned | Unassigned | 78_0 | Clustered | 2 | VC_78_0 | 2 | 0.0306 | 0.95617387 | 0.0293 | 0 | 0 | 0 | 1 | Ref |
| 5478 | MG020111 | Unassigned | Unassigned | Unassigned | 578_0 | Clustered | 2 | VC_578_0 | 2 | 0.1712 | 0.93973606 | 0.1608 | 0 | 0 | 0 | 1 | Ref |
| 5479 | MG022439 | Unassigned | Unassigned | Unassigned | 772_9 | Clustered | 7 | VC_772_9 | 7 | 0.0369 | 0.87705503 | 0.0324 | 0 | 0 | 0 | 1 | Ref |
| 5480 | MG022440 | Unassigned | Unassigned | Unassigned | 772_9 | Clustered | 7 | VC_772_9 | 7 | 0.0369 | 0.87705503 | 0.0324 | 0 | 0 | 0 | 1 | Ref |
| 5481 | MG025518 | Unassigned | Unassigned | Unassigned | 1104_0 | Clustered | 154 | VC_1104_0 | 154 | 0.8484 | 1 | 0.8484 | 0 | 0 | 0 | 1 | Ref |
| 5482 | MG029509 | Unassigned | Unassigned | Unassigned | 8_0 | Clustered | 58 | VC_8_0 | 58 | 0.8306 | 1 | 0.8306 | 0 | 0 | 0 | 0.9946 | Ref |
| 5483 | MG029510 | Unassigned | Unassigned | Unassigned | 10_0 | Clustered | 25 | VC_10_0 | 25 | 0.3467 | 1 | 0.3467 | 0 | 0 | 0 | 0.9733 | Ref |
| 5484 | MG029511 | Unassigned | Unassigned | Unassigned | 8_0 | Clustered | 58 | VC_8_0 | 58 | 0.8306 | 1 | 0.8306 | 0 | 0 | 0 | 0.9946 | Ref |
| 5485 | MG029512 | Unassigned | Unassigned | Unassigned | 10_0 | Clustered | 25 | VC_10_0 | 25 | 0.3467 | 1 | 0.3467 | 0 | 0 | 0 | 0.9733 | Ref |
| 5486 | MG029513 | Unassigned | Unassigned | Unassigned | 8_0 | Clustered | 58 | VC_8_0 | 58 | 0.8306 | 1 | 0.8306 | 0 | 0 | 0 | 0.9946 | Ref |
| 5487 | MG029514 | Unassigned | Unassigned | Unassigned | 8_0 | Clustered | 58 | VC_8_0 | 58 | 0.8306 | 1 | 0.8306 | 0 | 0 | 0 | 0.9946 | Ref |
| 5488 | MG029515 | Unassigned | Unassigned | Unassigned | 8_0 | Clustered | 58 | VC_8_0 | 58 | 0.8306 | 1 | 0.8306 | 0 | 0 | 0 | 0.9946 | Ref |
| 5489 | MG029516 | Unassigned | Unassigned | Unassigned | 8_0 | Clustered | 58 | VC_8_0 | 58 | 0.8306 | 1 | 0.8306 | 0 | 0 | 0 | 0.9946 | Ref |
| 5490 | MG029517 | Unassigned | Unassigned | Unassigned | 8_0 | Clustered | 58 | VC_8_0 | 58 | 0.8306 | 1 | 0.8306 | 0 | 0 | 0 | 0.9946 | Ref |
| 5491 | MG029518 | Unassigned | Unassigned | Unassigned | 8_0 | Clustered | 58 | VC_8_0 | 58 | 0.8306 | 1 | 0.8306 | 0 | 0 | 0 | 0.9946 | Ref |
| 5492 | MG030346 | Unassigned | Unassigned | Unassigned | 591_0 | Clustered | 14 | VC_591_0 | 14 | 0.5184 | 1 | 0.5184 | 0 | 0 | 0 | 1 | Ref |
| 5493 | MG049919 | Unassigned | Unassigned | Unassigned | 575_0 | Clustered | 35 | VC_575_0 | 35 | 0.8815 | 1 | 0.8815 | 0 | 0 | 0 | 1 | Ref |
| 5494 | MG050172 | Unassigned | Unassigned | Unassigned | 456_0 | Clustered | 62 | VC_456_0 | 62 | 0.9026 | 1 | 0.9026 | 0 | 0 | 0 | 1 | Ref |
| 5495 | MG065635 | Unassigned | Unassigned | Unassigned | 772_19 | Clustered | 16 | VC_772_19 | 16 | 0.0975 | 0.9970466 | 0.0972 | 0 | 0 | 0 | 0.8083 | Ref |
| 5496 | MG065636 | Unassigned | Unassigned | Unassigned | 941_0 | Clustered | 40 | VC_941_0 | 40 | 0.6903 | 1 | 0.6903 | 0 | 0 | 0 | 1 | Ref |
| 5497 | MG065639 | Unassigned | Unassigned | Unassigned | 848_0 | Clustered | 28 | VC_848_0 | 28 | 0.7927 | 1 | 0.7927 | 0 | 0 | 0 | 1 | Ref |
| 5498 | MG065640 | Unassigned | Unassigned | Unassigned | 941_0 | Clustered | 40 | VC_941_0 | 40 | 0.6903 | 1 | 0.6903 | 0 | 0 | 0 | 1 | Ref |
| 5499 | MG065641 | Unassigned | Unassigned | Unassigned | 941_0 | Clustered | 40 | VC_941_0 | 40 | 0.6903 | 1 | 0.6903 | 0 | 0 | 0 | 1 | Ref |
| 5500 | MG065642 | Unassigned | Unassigned | Unassigned | 191_19 | Clustered | 12 | VC_191_19 | 12 | 0.0655 | 1 | 0.0655 | 0 | 0 | 0 | 0.8636 | Ref |
| 5501 | MG065643 | Unassigned | Unassigned | Unassigned | 941_0 | Clustered | 40 | VC_941_0 | 40 | 0.6903 | 1 | 0.6903 | 0 | 0 | 0 | 1 | Ref |
| 5502 | MG065644 | Unassigned | Unassigned | Unassigned | 772_0 | Clustered | 6 | VC_772_0 | 6 | 0.0305 | 0.8101283 | 0.0247 | 0 | 0 | 0 | 1 | Ref |
| 5503 | MG065645 | Unassigned | Unassigned | Unassigned | 848_0 | Clustered | 28 | VC_848_0 | 28 | 0.7927 | 1 | 0.7927 | 0 | 0 | 0 | 1 | Ref |
| 5504 | MG065646 | Unassigned | Unassigned | Unassigned | 772_16 | Clustered | 10 | VC_772_16 | 10 | 0.0563 | 0.9434928 | 0.0531 | 0 | 0 | 0 | 0.8667 | Ref |
| 5505 | MG065647 | Unassigned | Unassigned | Unassigned | 772_16 | Clustered | 10 | VC_772_16 | 10 | 0.0563 | 0.9434928 | 0.0531 | 0 | 0 | 0 | 0.8667 | Ref |
| 5506 | MG065648 | Unassigned | Unassigned | Unassigned | 835_0 | Clustered | 67 | VC_835_0 | 67 | 0.9624 | 1 | 0.9624 | 0 | 0 | 0 | 1 | Ref |

| | | | | | | | | | | | | | | | | | |
|---|---|---|---|---|---|---|---|---|---|---|---|---|---|---|---|---|---|
| 5507 | MG065649 | Unassigned | Unassigned | Unassigned | 941_0 | Clustered | 40 | VC_941_0 | 40 | 0.6903 | 1 | 0.6903 | 0 | 0 | 0 | 1 | Ref |
| 5508 | MG065650 | Unassigned | Unassigned | Unassigned | 847_1 | Clustered | 26 | VC_847_1 | 26 | 0.6323 | 1 | 0.6323 | 0 | 0 | 0 | 0.9815 | Ref |
| 5509 | MG065653 | Unassigned | Unassigned | Unassigned | 772_13 | Clustered/Singleton | 1 | VC_772_13 | 1 | 0 | 1 | 0 | 0 | 0 | 0 | 1 | Ref |
| 5510 | MG065654 | Unassigned | Unassigned | Unassigned | 772_14 | Clustered | 3 | VC_772_14 | 3 | 0.012 | 0.63979507 | 0.0077 | 0 | 0 | 0 | 1 | Ref |
| 5511 | MG065655 | Unassigned | Unassigned | Unassigned | 772_14 | Clustered | 3 | VC_772_14 | 3 | 0.012 | 0.63979507 | 0.0077 | 0 | 0 | 0 | 1 | Ref |
| 5512 | MG065656 | Unassigned | Unassigned | Unassigned | 848_0 | Clustered | 28 | VC_848_0 | 28 | 0.7927 | 1 | 0.7927 | 0 | 0 | 0 | 1 | Ref |
| 5513 | MG065657 | Unassigned | Unassigned | Unassigned | 941_0 | Clustered | 40 | VC_941_0 | 40 | 0.6903 | 1 | 0.6903 | 0 | 0 | 0 | 1 | Ref |
| 5514 | MG065658 | Unassigned | Unassigned | Unassigned | 772_16 | Clustered | 10 | VC_772_16 | 10 | 0.0563 | 0.9434928 | 0.0531 | 0 | 0 | 0 | 0.8667 | Ref |
| 5515 | MG065659 | Unassigned | Unassigned | Unassigned | 772_12 | Clustered/Singleton | 1 | VC_772_12 | 1 | 0 | 1 | 0 | 0 | 0 | 0 | 1 | Ref |
| 5516 | MG065661 | Unassigned | Unassigned | Unassigned | 772_16 | Clustered | 10 | VC_772_16 | 10 | 0.0563 | 0.9434928 | 0.0531 | 0 | 0 | 0 | 0.8667 | Ref |
| 5517 | MG065662 | Unassigned | Unassigned | Unassigned | 772_0 | Clustered | 6 | VC_772_0 | 6 | 0.0305 | 0.8101283 | 0.0247 | 0 | 0 | 0 | 1 | Ref |
| 5518 | MG065663 | Unassigned | Unassigned | Unassigned | 941_0 | Clustered | 40 | VC_941_0 | 40 | 0.6903 | 1 | 0.6903 | 0 | 0 | 0 | 1 | Ref |
| 5519 | MG065664 | Unassigned | Unassigned | Unassigned | 329_12 | Clustered | 2 | VC_329_12 | 2 | 0.0041 | 0.72605466 | 0.003 | 0 | 0 | 0 | 1 | Ref |
| 5520 | MG065665 | Unassigned | Unassigned | Unassigned | 941_0 | Clustered | 40 | VC_941_0 | 40 | 0.6903 | 1 | 0.6903 | 0 | 0 | 0 | 1 | Ref |
| 5521 | MG065666 | Unassigned | Unassigned | Unassigned | 772_14 | Clustered | 3 | VC_772_14 | 3 | 0.012 | 0.63979507 | 0.0077 | 0 | 0 | 0 | 1 | Ref |
| 5522 | MG065667 | Unassigned | Unassigned | Unassigned | 941_0 | Clustered | 40 | VC_941_0 | 40 | 0.6903 | 1 | 0.6903 | 0 | 0 | 0 | 1 | Ref |
| 5523 | MG065669 | Unassigned | Unassigned | Unassigned | 941_0 | Clustered | 40 | VC_941_0 | 40 | 0.6903 | 1 | 0.6903 | 0 | 0 | 0 | 1 | Ref |
| 5524 | MG065670 | Unassigned | Unassigned | Unassigned | 941_0 | Clustered | 40 | VC_941_0 | 40 | 0.6903 | 1 | 0.6903 | 0 | 0 | 0 | 1 | Ref |
| 5525 | MG065671 | Unassigned | Unassigned | Unassigned | 941_0 | Clustered | 40 | VC_941_0 | 40 | 0.6903 | 1 | 0.6903 | 0 | 0 | 0 | 1 | Ref |
| 5526 | MG065672 | Unassigned | Unassigned | Unassigned | 772_0 | Clustered | 6 | VC_772_0 | 6 | 0.0305 | 0.8101283 | 0.0247 | 0 | 0 | 0 | 1 | Ref |
| 5527 | MG065673 | Unassigned | Unassigned | Unassigned | 941_0 | Clustered | 40 | VC_941_0 | 40 | 0.6903 | 1 | 0.6903 | 0 | 0 | 0 | 1 | Ref |
| 5528 | MG065674 | Unassigned | Unassigned | Unassigned | 772_0 | Clustered | 6 | VC_772_0 | 6 | 0.0305 | 0.8101283 | 0.0247 | 0 | 0 | 0 | 1 | Ref |
| 5529 | MG065675 | Unassigned | Unassigned | Unassigned | 941_0 | Clustered | 40 | VC_941_0 | 40 | 0.6903 | 1 | 0.6903 | 0 | 0 | 0 | 1 | Ref |
| 5530 | MG065677 | Unassigned | Unassigned | Unassigned | 772_0 | Clustered | 6 | VC_772_0 | 6 | 0.0305 | 0.8101283 | 0.0247 | 0 | 0 | 0 | 1 | Ref |
| 5531 | MG065679 | Unassigned | Unassigned | Unassigned | 941_0 | Clustered | 40 | VC_941_0 | 40 | 0.6903 | 1 | 0.6903 | 0 | 0 | 0 | 1 | Ref |
| 5532 | MG065680 | Unassigned | Unassigned | Unassigned | 941_0 | Clustered | 40 | VC_941_0 | 40 | 0.6903 | 1 | 0.6903 | 0 | 0 | 0 | 1 | Ref |
| 5533 | MG065681 | Unassigned | Unassigned | Unassigned | 941_0 | Clustered | 40 | VC_941_0 | 40 | 0.6903 | 1 | 0.6903 | 0 | 0 | 0 | 1 | Ref |
| 5534 | MG065682 | Unassigned | Unassigned | Unassigned | 772_16 | Clustered | 10 | VC_772_16 | 10 | 0.0563 | 0.9434928 | 0.0531 | 0 | 0 | 0 | 0.8667 | Ref |
| 5535 | MG065687 | Unassigned | Unassigned | Unassigned | 941_0 | Clustered | 40 | VC_941_0 | 40 | 0.6903 | 1 | 0.6903 | 0 | 0 | 0 | 1 | Ref |
| 5536 | MG065688 | Unassigned | Unassigned | Unassigned | 456_0 | Clustered | 62 | VC_456_0 | 62 | 0.9026 | 1 | 0.9026 | 0 | 0 | 0 | 1 | Ref |
| 5537 | MG065689 | Unassigned | Unassigned | Unassigned | 848_0 | Clustered | 28 | VC_848_0 | 28 | 0.7927 | 1 | 0.7927 | 0 | 0 | 0 | 1 | Ref |
| 5538 | MG065690 | Unassigned | Unassigned | Unassigned | 772_0 | Clustered | 6 | VC_772_0 | 6 | 0.0305 | 0.8101283 | 0.0247 | 0 | 0 | 0 | 1 | Ref |
| 5539 | MG065691 | Unassigned | Unassigned | Unassigned | 1219_0 | Clustered/Singleton | 1 | VC_1219_0 | 1 | 0 | 1 | 0 | 0 | 0 | 0 | 1 | Ref |
| 5540 | MG065692 | Unassigned | Unassigned | Unassigned | 190_31 | Clustered/Singleton | 1 | VC_190_31 | 1 | 0 | 1 | 0 | 0 | 0 | 0 | 1 | Ref |
| 5541 | MG099933 | Unassigned | Unassigned | Unassigned | 638_0 | Clustered | 45 | VC_638_0 | 45 | 0.7741 | 1 | 0.7741 | 0 | 0 | 0 | 1 | Ref |
| 5542 | MG099934 | Unassigned | Unassigned | Unassigned | 940_0 | Clustered | 34 | VC_940_0 | 34 | 0.7959 | 1 | 0.7959 | 0 | 0 | 0 | 1 | Ref |
| 5543 | MG099935 | Unassigned | Unassigned | Unassigned | 866_1 | Clustered | 20 | VC_866_1 | 20 | 0.4814 | 1 | 0.4814 | 0 | 0 | 0 | 1 | Ref |
| 5544 | MG099936 | Unassigned | Unassigned | Unassigned | 521_0 | Clustered | 37 | VC_521_0 | 37 | 0.912 | 1 | 0.912 | 0 | 0 | 0 | 1 | Ref |
| 5545 | MG099937 | Unassigned | Unassigned | Unassigned | 940_0 | Clustered | 34 | VC_940_0 | 34 | 0.7959 | 1 | 0.7959 | 0 | 0 | 0 | 1 | Ref |
| 5546 | MG099938 | Unassigned | Unassigned | Unassigned | 940_0 | Clustered | 34 | VC_940_0 | 34 | 0.7959 | 1 | 0.7959 | 0 | 0 | 0 | 1 | Ref |
| 5547 | MG099939 | Unassigned | Unassigned | Unassigned | 557_1 | Clustered | 11 | VC_557_1 | 11 | 0.4945 | 1 | 0.4945 | 0 | 0 | 0 | 0.7818 | Ref |
| 5548 | MG099940 | Unassigned | Unassigned | Unassigned | 868_0 | Clustered | 25 | VC_868_0 | 25 | 0.5791 | 1 | 0.5791 | 0 | 0 | 0 | 1 | Ref |
| 5549 | MG099941 | Unassigned | Unassigned | Unassigned | 1175_0 | Clustered | 22 | VC_1175_0 | 22 | 0.4562 | 1 | 0.4562 | 0 | 0 | 0 | 1 | Ref |
| 5550 | MG099942 | Unassigned | Unassigned | Unassigned | 517_1 | Clustered | 108 | VC_517_1 | 108 | 0.8483 | 1 | 0.8483 | 0 | 0 | 0 | 0.9955 | Ref |
| 5551 | MG099943 | Unassigned | Unassigned | Unassigned | 556_0 | Clustered | 21 | VC_556_0 | 21 | 0.9912 | 1 | 0.9912 | 0 | 0 | 0 | 1 | Ref |
| 5552 | MG099944 | Unassigned | Unassigned | Unassigned | 940_0 | Clustered | 34 | VC_940_0 | 34 | 0.7959 | 1 | 0.7959 | 0 | 0 | 0 | 1 | Ref |
| 5553 | MG099945 | Unassigned | Unassigned | Unassigned | 223_0 | Clustered | 38 | VC_223_0 | 38 | 0.9095 | 1 | 0.9095 | 0 | 0 | 0 | 1 | Ref |
| 5554 | MG099946 | Unassigned | Unassigned | Unassigned | 761_0 | Clustered | 67 | VC_761_0 | 67 | 0.8947 | 1 | 0.8947 | 0 | 0 | 0 | 1 | Ref |
| 5555 | MG099947 | Unassigned | Unassigned | Unassigned | 215_0 | Clustered | 5 | VC_215_0 | 5 | 0.0724 | 0.99999997 | 0.0724 | 0 | 0 | 0 | 1 | Ref |
| 5556 | MG099948 | Unassigned | Unassigned | Unassigned | 521_0 | Clustered | 37 | VC_521_0 | 37 | 0.912 | 1 | 0.912 | 0 | 0 | 0 | 1 | Ref |
| 5557 | MG099949 | Unassigned | Unassigned | Unassigned | 940_0 | Clustered | 34 | VC_940_0 | 34 | 0.7959 | 1 | 0.7959 | 0 | 0 | 0 | 1 | Ref |
| 5558 | MG099950 | Unassigned | Unassigned | Unassigned | 1169_0 | Clustered | 7 | VC_1169_0 | 7 | 0.9811 | 0.99995613 | 0.981 | 0 | 0 | 0 | 1 | Ref |
| 5559 | MG099951 | Unassigned | Unassigned | Unassigned | 325_38 | Clustered | 6 | VC_325_38 | 6 | 0.0267 | 1 | 0.0267 | 0 | 0 | 0 | 0.8 | Ref |
| 5560 | MG099952 | Unassigned | Unassigned | Unassigned | 223_0 | Clustered | 38 | VC_223_0 | 38 | 0.9095 | 1 | 0.9095 | 0 | 0 | 0 | 1 | Ref |
| 5561 | MG099953 | Unassigned | Unassigned | Unassigned | 551_8 | Clustered | 19 | VC_551_8 | 19 | 0.1039 | 1 | 0.1039 | 0 | 0 | 0 | 0.8713 | Ref |
| 5562 | MG189906 | Unassigned | Unassigned | Unassigned | 905_0 | Clustered | 3 | VC_905_0 | 3 | 0.3424 | 0.99823775 | 0.3417 | 0 | 0 | 0 | 1 | Ref |
| 5563 | MG197810 | Unassigned | Unassigned | Unassigned | 123_0 | Clustered | 23 | VC_123_0 | 23 | 0.817 | 1 | 0.817 | 0 | 0 | 0 | 1 | Ref |
| 5564 | MG198776 | Unassigned | Unassigned | Unassigned | 768_0 | Clustered | 8 | VC_768_0 | 8 | 0.7012 | 1 | 0.7012 | 0 | 0 | 0 | 1 | Ref |
| 5565 | MG198777 | Unassigned | Unassigned | Unassigned | 768_0 | Clustered | 8 | VC_768_0 | 8 | 0.7012 | 1 | 0.7012 | 0 | 0 | 0 | 1 | Ref |
| 5566 | MG198778 | Unassigned | Unassigned | Unassigned | 768_0 | Clustered | 8 | VC_768_0 | 8 | 0.7012 | 1 | 0.7012 | 0 | 0 | 0 | 1 | Ref |
| 5567 | MG198779 | Unassigned | Unassigned | Unassigned | 1160_0 | Clustered | 73 | VC_1160_0 | 73 | 0.9835 | 1 | 0.9835 | 0 | 0 | 0 | 1 | Ref |
| 5568 | MG198780 | Unassigned | Unassigned | Unassigned | 768_0 | Clustered | 8 | VC_768_0 | 8 | 0.7012 | 1 | 0.7012 | 0 | 0 | 0 | 1 | Ref |
| 5569 | MG198781 | Unassigned | Unassigned | Unassigned | 1160_0 | Clustered | 73 | VC_1160_0 | 73 | 0.9835 | 1 | 0.9835 | 0 | 0 | 0 | 1 | Ref |
| 5570 | MG198782 | Unassigned | Unassigned | Unassigned | 1160_0 | Clustered | 73 | VC_1160_0 | 73 | 0.9835 | 1 | 0.9835 | 0 | 0 | 0 | 1 | Ref |
| 5571 | MG198783 | Unassigned | Unassigned | Unassigned | 410_0 | Clustered | 7 | VC_410_0 | 7 | 0.7905 | 1 | 0.7905 | 0 | 0 | 0 | 1 | Ref |
| 5572 | MG198784 | Unassigned | Unassigned | Unassigned | 410_0 | Clustered | 7 | VC_410_0 | 7 | 0.7905 | 1 | 0.7905 | 0 | 0 | 0 | 1 | Ref |
| 5573 | MG198785 | Unassigned | Unassigned | Unassigned | 519_0 | Clustered | 39 | VC_519_0 | 39 | 0.8156 | 1 | 0.8156 | 0 | 0 | 0 | 1 | Ref |
| 5574 | MG201401 | Unassigned | Unassigned | Unassigned | 72_0 | Clustered | 2 | VC_72_0 | 2 | 0.0777 | 0.95616461 | 0.0743 | 0 | 0 | 0 | 1 | Ref |
| 5575 | MG208881 | Unassigned | Unassigned | Unassigned | 779_0 | Clustered | 37 | VC_779_0 | 37 | 0.5682 | 1 | 0.5682 | 0 | 0 | 0 | 1 | Ref |
| 5576 | MG209611 | Unassigned | Unassigned | Unassigned | 932_0 | Clustered | 2 | VC_932_0 | 2 | 0.4438 | 0.89435023 | 0.3969 | 0 | 0 | 0 | 1 | Ref |
| 5577 | MG210949 | Unassigned | Unassigned | Unassigned | 1160_0 | Clustered | 73 | VC_1160_0 | 73 | 0.9835 | 1 | 0.9835 | 0 | 0 | 0 | 1 | Ref |
| 5578 | MG214764 | Unassigned | Unassigned | Unassigned | 1222_0 | Clustered | 3 | VC_1222_0 | 3 | 1 | 0 | 0 | 0 | 0 | 0 | 1 | Ref |
| 5579 | MG241338 | Unassigned | Unassigned | Unassigned | 584_0 | Clustered | 28 | VC_584_0 | 28 | 0.7704 | 1 | 0.7704 | 0 | 0 | 0 | 1 | Ref |
| 5580 | MG250485 | Unassigned | Unassigned | Unassigned | 129_0 | Clustered | 43 | VC_129_0 | 43 | 0.886 | 1 | 0.886 | 0 | 0 | 0 | 1 | Ref |
| 5581 | MG250486 | Unassigned | Unassigned | Unassigned | 582_3 | Clustered | 12 | VC_582_3 | 12 | 0.2357 | 1 | 0.2357 | 0 | 0 | 0 | 0.8636 | Ref |
| 5582 | MG251390 | Unassigned | Unassigned | Unassigned | 452_0 | Clustered | 66 | VC_452_0 | 66 | 0.9248 | 1 | 0.9248 | 0 | 0 | 0 | 1 | Ref |
| 5583 | MG251391 | Unassigned | Unassigned | Unassigned | 329_1 | Clustered | 17 | VC_329_1 | 17 | 0.0867 | 1 | 0.0867 | 0 | 0 | 0 | 0.8235 | Ref |
| 5584 | MG251392 | Unassigned | Unassigned | Unassigned | 329_1 | Clustered | 17 | VC_329_1 | 17 | 0.0867 | 1 | 0.0867 | 0 | 0 | 0 | 0.8235 | Ref |
| 5585 | MG252615 | Unassigned | Unassigned | Unassigned | 638_0 | Clustered | 45 | VC_638_0 | 45 | 0.7741 | 1 | 0.7741 | 0 | 0 | 0 | 1 | Ref |
| 5586 | MG252693 | Unassigned | Unassigned | Unassigned | 634_0 | Clustered | 3 | VC_634_0 | 3 | 0.2046 | 0.99837593 | 0.2043 | 0 | 0 | 0 | 1 | Ref |
| 5587 | MG253647 | Unassigned | Unassigned | Unassigned | 202_0 | Clustered | 102 | VC_202_0 | 102 | 0.4049 | 1 | 0.4049 | 0 | 0 | 0 | 0.7876 | Ref |
| 5588 | MG253648 | Unassigned | Unassigned | Unassigned | 202_0 | Clustered | 102 | VC_202_0 | 102 | 0.4049 | 1 | 0.4049 | 0 | 0 | 0 | 0.7876 | Ref |
| 5589 | MG253649 | Unassigned | Unassigned | Unassigned | 202_0 | Clustered | 102 | VC_202_0 | 102 | 0.4049 | 1 | 0.4049 | 0 | 0 | 0 | 0.7876 | Ref |
| 5590 | MG253650 | Unassigned | Unassigned | Unassigned | 202_0 | Clustered | 102 | VC_202_0 | 102 | 0.4049 | 1 | 0.4049 | 0 | 0 | 0 | 0.7876 | Ref |
| 5591 | MG253651 | Unassigned | Unassigned | Unassigned | 202_0 | Clustered | 102 | VC_202_0 | 102 | 0.4049 | 1 | 0.4049 | 0 | 0 | 0 | 0.7876 | Ref |
| 5592 | MG253652 | Unassigned | Unassigned | Unassigned | 202_0 | Clustered | 102 | VC_202_0 | 102 | 0.4049 | 1 | 0.4049 | 0 | 0 | 0 | 0.7876 | Ref |
| 5593 | MG253653 | Unassigned | Unassigned | Unassigned | 202_0 | Clustered | 102 | VC_202_0 | 102 | 0.4049 | 1 | 0.4049 | 0 | 0 | 0 | 0.7876 | Ref |
| 5594 | MG264739 | Unassigned | Unassigned | Unassigned | 695_0 | Clustered | 48 | VC_695_0 | 48 | 0.8162 | 1 | 0.8162 | 0 | 0 | 0 | 1 | Ref |
| 5595 | MG266157 | Unassigned | Unassigned | Unassigned | 131_0 | Clustered | 17 | VC_131_0 | 17 | 0.2671 | 1 | 0.2671 | 0 | 0 | 0 | 1 | Ref |
| 5596 | MG269973 | Unassigned | Unassigned | Unassigned | 1223_0 | Clustered | 2 | VC_1223_0 | 2 | 1 | 1 | 1 | 0 | 0 | 0 | 1 | Ref |
| 5597 | MG271909 | Unassigned | Unassigned | Unassigned | 873_0 | Clustered | 3 | VC_873_0 | 3 | 0.9107 | 0.98376334 | 0.8959 | 0 | 0 | 0 | 1 | Ref |
| 5598 | MG280946 | Unassigned | Unassigned | Unassigned | 592_0 | Clustered | 50 | VC_592_0 | 50 | 0.7314 | 1 | 0.7314 | 0 | 0 | 0 | 1 | Ref |
| 5599 | MG298964 | Unassigned | Unassigned | Unassigned | 982_0 | Clustered | 62 | VC_982_0 | 62 | 0.8646 | 1 | 0.8646 | 0 | 0 | 0 | 0.743 | Ref |
| 5600 | MG309717 | Unassigned | Unassigned | Unassigned | 202_0 | Clustered | 102 | VC_202_0 | 102 | 0.4049 | 1 | 0.4049 | 0 | 0 | 0 | 0.7876 | Ref |
| 5601 | MG309718 | Unassigned | Unassigned | Unassigned | 202_0 | Clustered | 102 | VC_202_0 | 102 | 0.4049 | 1 | 0.4049 | 0 | 0 | 0 | 0.7876 | Ref |
| 5602 | MG324353 | Unassigned | Unassigned | Unassigned | 407_0 | Clustered | 3 | VC_407_0 | 3 | 0.3352 | 0.99824791 | 0.3346 | 0 | 0 | 0 | 1 | Ref |
| 5603 | MG324354 | Unassigned | Unassigned | Unassigned | 407_0 | Clustered | 3 | VC_407_0 | 3 | 0.3352 | 0.99824791 | 0.3346 | 0 | 0 | 0 | 1 | Ref |
| 5604 | MG366115 | Unassigned | Unassigned | Unassigned | 750_0 | Clustered/Singleton | 1 | VC_750_0 | 1 | 0 | 1 | 0 | 0 | 0 | 0 | 1 | Ref |
| 5605 | MG383452 | Unassigned | Unassigned | Unassigned | 133_27 | Clustered/Singleton | 1 | VC_133_27 | 1 | 0 | 1 | 0 | 0 | 0 | 0 | 1 | Ref |
| 5606 | MG387042 | Unassigned | Unassigned | Unassigned | 190_26 | Clustered/Singleton | 1 | VC_190_26 | 1 | 0 | 1 | 0 | 0 | 0 | 0 | 1 | Ref |
| 5607 | MG407615 | Unassigned | Unassigned | Unassigned | 28_1 | Clustered | 53 | VC_28_1 | 53 | 0.7164 | 1 | 0.7164 | 0 | 0 | 0 | 0.9325 | Ref |
| 5608 | MG428990 | Unassigned | Unassigned | Unassigned | 132_1 | Clustered | 17 | VC_132_1 | 17 | 0.1738 | 1 | 0.1738 | 0 | 0 | 0 | 0.9853 | Ref |
| 5609 | MG428991 | Unassigned | Unassigned | Unassigned | 132_1 | Clustered | 17 | VC_132_1 | 17 | 0.1738 | 1 | 0.1738 | 0 | 0 | 0 | 0.9853 | Ref |
| 5610 | MG428992 | Unassigned | Unassigned | Unassigned | 133_1 | Clustered/Singleton | 1 | VC_133_1 | 1 | 0 | 1 | 0 | 0 | 0 | 0 | 1 | Ref |
| 5611 | MG432137 | Unassigned | Unassigned | Unassigned | 123_0 | Clustered | 23 | VC_123_0 | 23 | 0.817 | 1 | 0.817 | 0 | 0 | 0 | 1 | Ref |
| 5612 | MG432151 | Unassigned | Unassigned | Unassigned | 472_0 | Clustered | 47 | VC_472_0 | 47 | 0.9025 | 1 | 0.9025 | 0 | 0 | 0 | 1 | Ref |
| 5613 | MG459218 | Unassigned | Unassigned | Unassigned | 699_0 | Clustered | 71 | VC_699_0 | 71 | 0.9352 | 1 | 0.9352 | 0 | 0 | 0 | 1 | Ref |
| 5614 | MG459987 | Unassigned | Unassigned | Unassigned | 192_0 | Clustered | 15 | VC_192_0 | 15 | 0.5071 | 1 | 0.5071 | 0 | 0 | 0 | 1 | Ref |
| 5615 | MG471392 | Unassigned | Unassigned | Unassigned | 386_0 | Clustered | 49 | VC_386_0 | 49 | 0.7941 | 1 | 0.7941 | 0 | 0 | 0 | 1 | Ref |
| 5616 | MG488277 | Unassigned | Unassigned | Unassigned | 392_0 | Clustered | 126 | VC_392_0 | 126 | 0.9621 | 1 | 0.9621 | 0 | 0 | 0 | 1 | Ref |
| 5617 | MG515223 | Unassigned | Unassigned | Unassigned | 1224_0 | Clustered | 19 | VC_1224_0 | 19 | 1 | 1 | 1 | 0 | 0 | 0 | 1 | Ref |
| 5618 | MG518519 | Unassigned | Unassigned | Unassigned | 1224_0 | Clustered | 19 | VC_1224_0 | 19 | 1 | 1 | 1 | 0 | 0 | 0 | 1 | Ref |
| 5619 | MG518520 | Unassigned | Unassigned | Unassigned | 1224_0 | Clustered | 19 | VC_1224_0 | 19 | 1 | 1 | 1 | 0 | 0 | 0 | 1 | Ref |
| 5620 | MG543995 | Unassigned | Unassigned | Unassigned | 12_1 | Clustered | 2 | VC_12_1 | 2 | 0.0192 | 0.95739495 | 0.0184 | 0 | 0 | 0 | 1 | Ref |
| 5621 | MG545917 | Unassigned | Unassigned | Unassigned | 1027_0 | Clustered | 20 | VC_1027_0 | 20 | 0.7114 | 1 | 0.7114 | 0 | 0 | 0 | 1 | Ref |
| 5622 | MG550110 | Unassigned | Unassigned | Unassigned | 963_0 | Clustered | 4 | VC_963_0 | 4 | 0.8505 | 0.99879268 | 0.8495 | 0 | 0 | 0 | 1 | Ref |
| 5623 | MG550111 | Unassigned | Unassigned | Unassigned | 963_0 | Clustered | 4 | VC_963_0 | 4 | 0.8505 | 0.99879268 | 0.8495 | 0 | 0 | 0 | 1 | Ref |
| 5624 | MG550112 | Unassigned | Unassigned | Unassigned | 459_0 | Clustered | 3 | VC_459_0 | 3 | 0.8542 | 0.99054823 | 0.8461 | 0 | 0 | 0 | 1 | Ref |
| 5625 | MG550113 | Unassigned | Unassigned | Unassigned | 963_0 | Clustered | 4 | VC_963_0 | 4 | 0.8505 | 0.99879268 | 0.8495 | 0 | 0 | 0 | 1 | Ref |
| 5626 | MG552615 | Unassigned | Unassigned | Unassigned | 938_0 | Clustered | 99 | VC_938_0 | 99 | 0.9119 | 1 | 0.9119 | 0 | 0 | 0 | 0.9928 | Ref |
| 5627 | MG557618 | Unassigned | Unassigned | Unassigned | 149_15 | Clustered | 16 | VC_149_15 | 16 | 0.0788 | 1 | 0.0788 | 0 | 0 | 0 | 0.8083 | Ref |
| 5628 | MG557619 | Unassigned | Unassigned | Unassigned | 9_0 | Clustered | 38 | VC_9_0 | 38 | 0.4895 | 1 | 0.4895 | 0 | 0 | 0 | 1 | Ref |
| 5629 | MG557979 | Unassigned | Unassigned | Unassigned | 526_0 | Clustered | 6 | VC_526_0 | 6 | 0.4386 | 1 | 0.4386 | 0 | 0 | 0 | 1 | Ref |
| 5630 | MG563770 | Unassigned | Unassigned | Unassigned | 360_0 | Clustered | 358 | VC_360_0 | 358 | 0.9615 | 1 | 0.9615 | 0 | 0 | 0 | 1 | Ref |
| 5631 | MG564297 | Unassigned | Unassigned | Unassigned | 12_1 | Clustered | 2 | VC_12_1 | 2 | 0.0192 | 0.95739495 | 0.0184 | 0 | 0 | 0 | 1 | Ref |
| 5632 | MG575418 | Unassigned | Unassigned | Unassigned | 112_0 | Clustered | 8 | VC_112_0 | 8 | 0.2537 | 1 | 0.2537 | 0 | 0 | 0 | 1 | Ref |
| 5633 | MG575419 | Unassigned | Unassigned | Unassigned | 112_0 | Clustered | 8 | VC_112_0 | 8 | 0.2537 | 1 | 0.2537 | 0 | 0 | 0 | 1 | Ref |
| 5634 | MG575421 | Unassigned | Unassigned | Unassigned | 426_1 | Clustered | 3 | VC_426_1 | 3 | 0.1152 | 0.99804615 | 0.1149 | 0 | 0 | 0 | 1 | Ref |
| 5635 | MG581355 | Unassigned | Unassigned | Unassigned | 835_0 | Clustered | 67 | VC_835_0 | 67 | 0.9624 | 1 | 0.9624 | 0 | 0 | 0 | 1 | Ref |
| 5636 | MG589383 | Unassigned | Unassigned | Unassigned | 286_12 | Clustered | 3 | VC_286_12 | 3 | 0.0146 | 0.92328265 | 0.0135 | 0 | 0 | 0 | 1 | Ref |
| 5637 | MG589384 | Unassigned | Unassigned | Unassigned | 328_0 | Clustered | 11 | VC_328_0 | 11 | 0.4707 | 1 | 0.4707 | 0 | 0 | 0 | 1 | Ref |
| 5638 | MG589385 | Unassigned | Unassigned | Unassigned | 472_0 | Clustered | 47 | VC_472_0 | 47 | 0.9025 | 1 | 0.9025 | 0 | 0 | 0 | 1 | Ref |
| 5639 | MG589386 | Unassigned | Unassigned | Unassigned | 472_0 | Clustered | 47 | VC_472_0 | 47 | 0.9025 | 1 | 0.9025 | 0 | 0 | 0 | 1 | Ref |
| 5640 | MG589387 | Unassigned | Unassigned | Unassigned | 229_0 | Clustered | 32 | VC_229_0 | 32 | 0.9119 | 1 | 0.9119 | 0 | 0 | 0 | 1 | Ref |
| 5641 | MG592390 | Unassigned | Unassigned | Unassigned | 645_0 | Clustered | 8 | VC_645_0 | 8 | 0.3138 | 1 | 0.3138 | 0 | 0 | 0 | 1 | Ref |
| 5642 | MG592391 | Unassigned | Unassigned | Unassigned | 1041_0 | Clustered | 9 | VC_1041_0 | 9 | 0.6541 | 1 | 0.6541 | 0 | 0 | 0 | 1 | Ref |
| 5643 | MG592392 | Unassigned | Unassigned | Unassigned | 499_0 | Clustered | 24 | VC_499_0 | 24 | 0.8569 | 1 | 0.8569 | 0 | 0 | 0 | 0.9529 | Ref |
| 5644 | MG592393 | Unassigned | Unassigned | Unassigned | 646_0 | Clustered | 29 | VC_646_0 | 29 | 0.9131 | 1 | 0.9131 | 0 | 0 | 0 | 1 | Ref |
| 5645 | MG592394 | Unassigned | Unassigned | Unassigned | 1225_0 | Clustered | 4 | VC_1225_0 | 4 | 0.9619 | 0.99374885 | 0.9559 | 0 | 0 | 0 | 1 | Ref |
| 5646 | MG592395 | Unassigned | Unassigned | Unassigned | 498_0 | Clustered | 8 | VC_498_0 | 8 | 0.3987 | 1 | 0.3987 | 0 | 0 | 0 | 1 | Ref |
| 5647 | MG592397 | Unassigned | Unassigned | Unassigned | 1050_1 | Clustered | 29 | VC_1050_1 | 29 | 0.4861 | 1 | 0.4861 | 0 | 0 | 0 | 0.9581 | Ref |
| 5648 | MG592398 | Unassigned | Unassigned | Unassigned | 944_0 | Clustered | 6 | VC_944_0 | 6 | 0.8196 | 1 | 0.8196 | 0 | 0 | 0 | 1 | Ref |
| 5649 | MG592399 | Unassigned | Unassigned | Unassigned | 543_0 | Clustered | 6 | VC_543_0 | 6 | 0.864 | 0.99999948 | 0.864 | 0 | 0 | 0 | 1 | Ref |
| 5650 | MG592400 | Unassigned | Unassigned | Unassigned | 122_0 | Clustered | 5 | VC_122_0 | 5 | 0.2073 | 0.99999996 | 0.2073 | 0 | 0 | 0 | 1 | Ref |
| 5651 | MG592401 | Unassigned | Unassigned | Unassigned | 499_0 | Clustered | 24 | VC_499_0 | 24 | 0.8569 | 1 | 0.8569 | 0 | 0 | 0 | 0.9529 | Ref |
| 5652 | MG592402 | Unassigned | Unassigned | Unassigned | 1225_0 | Clustered | 4 | VC_1225_0 | 4 | 0.9619 | 0.99374885 | 0.9559 | 0 | 0 | 0 | 1 | Ref |
| 5653 | MG592403 | Unassigned | Unassigned | Unassigned | 296_0 | Clustered | 6 | VC_296_0 | 6 | 0.5315 | 1 | 0.5315 | 0 | 0 | 0 | 1 | Ref |
| 5654 | MG592404 | Unassigned | Unassigned | Unassigned | 296_0 | Clustered | 6 | VC_296_0 | 6 | 0.5315 | 1 | 0.5315 | 0 | 0 | 0 | 1 | Ref |
| 5655 | MG592405 | Unassigned | Unassigned | Unassigned | 296_0 | Clustered | 6 | VC_296_0 | 6 | 0.5315 | 1 | 0.5315 | 0 | 0 | 0 | 1 | Ref |
| 5656 | MG592406 | Unassigned | Unassigned | Unassigned | 1050_1 | Clustered | 29 | VC_1050_1 | 29 | 0.4861 | 1 | 0.4861 | 0 | 0 | 0 | 0.9581 | Ref |
| 5657 | MG592407 | Unassigned | Unassigned | Unassigned | 1050_1 | Clustered | 29 | VC_1050_1 | 29 | 0.4861 | 1 | 0.4861 | 0 | 0 | 0 | 0.9581 | Ref |
| 5658 | MG592408 | Unassigned | Unassigned | Unassigned | 1050_1 | Clustered | 29 | VC_1050_1 | 29 | 0.4861 | 1 | 0.4861 | 0 | 0 | 0 | 0.9581 | Ref |
| 5659 | MG592409 | Unassigned | Unassigned | Unassigned | 930_0 | Clustered | 7 | VC_930_0 | 7 | 0.519 | 1 | 0.519 | 0 | 0 | 0 | 1 | Ref |
| 5660 | MG592410 | Unassigned | Unassigned | Unassigned | 930_0 | Clustered | 7 | VC_930_0 | 7 | 0.519 | 1 | 0.519 | 0 | 0 | 0 | 1 | Ref |
| 5661 | MG592411 | Unassigned | Unassigned | Unassigned | 1050_0 | Clustered | 24 | VC_1050_0 | 24 | 0.5274 | 1 | 0.5274 | 0 | 0 | 0 | 1 | Ref |
| 5662 | MG592412 | Unassigned | Unassigned | Unassigned | 248_0 | Clustered | 11 | VC_248_0 | 11 | 0.7031 | 1 | 0.7031 | 0 | 0 | 0 | 1 | Ref |
| 5663 | MG592414 | Unassigned | Unassigned | Unassigned | 499_0 | Clustered | 24 | VC_499_0 | 24 | 0.8569 | 1 | 0.8569 | 0 | 0 | 0 | 0.9529 | Ref |
| 5664 | MG592415 | Unassigned | Unassigned | Unassigned | 478_0 | Clustered | 2 | VC_478_0 | 2 | 0.3505 | 0.94766489 | 0.3321 | 0 | 0 | 0 | 1 | Ref |
| 5665 | MG592416 | Unassigned | Unassigned | Unassigned | 1050_1 | Clustered | 29 | VC_1050_1 | 29 | 0.4861 | 1 | 0.4861 | 0 | 0 | 0 | 0.9581 | Ref |
| 5666 | MG592417 | Unassigned | Unassigned | Unassigned | 1050_1 | Clustered | 29 | VC_1050_1 | 29 | 0.4861 | 1 | 0.4861 | 0 | 0 | 0 | 0.9581 | Ref |
| 5667 | MG592418 | Unassigned | Unassigned | Unassigned | 543_0 | Clustered | 6 | VC_543_0 | 6 | 0.864 | 0.99999948 | 0.864 | 0 | 0 | 0 | 1 | Ref |
| 5668 | MG592419 | Unassigned | Unassigned | Unassigned | 543_0 | Clustered | 6 | VC_543_0 | 6 | 0.864 | 0.99999948 | 0.864 | 0 | 0 | 0 | 1 | Ref |

| | | | | | | | | | | | | | | | | | |
|---|---|---|---|---|---|---|---|---|---|---|---|---|---|---|---|---|---|
| 5669 | MG592420 | Unassigned | Unassigned | Unassigned | 645_0 | Clustered | 8 | VC_645_0 | 8 | 0.3138 | 1 | 0.3138 | 0 | 0 | 0 | 1 | Ref |
| 5670 | MG592421 | Unassigned | Unassigned | Unassigned | 1041_0 | Clustered | 9 | VC_1041_0 | 9 | 0.6541 | 1 | 0.6541 | 0 | 0 | 0 | 1 | Ref |
| 5671 | MG592422 | Unassigned | Unassigned | Unassigned | 122_0 | Clustered | 5 | VC_122_0 | 5 | 0.2073 | 0.99999996 | 0.2073 | 0 | 0 | 0 | 1 | Ref |
| 5672 | MG592423 | Unassigned | Unassigned | Unassigned | 645_0 | Clustered | 8 | VC_645_0 | 8 | 0.3138 | 1 | 0.3138 | 0 | 0 | 0 | 1 | Ref |
| 5673 | MG592425 | Unassigned | Unassigned | Unassigned | 122_0 | Clustered | 5 | VC_122_0 | 5 | 0.2073 | 0.99999996 | 0.2073 | 0 | 0 | 0 | 1 | Ref |
| 5674 | MG592427 | Unassigned | Unassigned | Unassigned | 1225_0 | Clustered | 4 | VC_1225_0 | 4 | 0.9619 | 0.99374885 | 0.9559 | 0 | 0 | 0 | 1 | Ref |
| 5675 | MG592428 | Unassigned | Unassigned | Unassigned | 1050_1 | Clustered | 29 | VC_1050_1 | 29 | 0.4861 | 1 | 0.4861 | 0 | 0 | 0 | 0.9581 | Ref |
| 5676 | MG592429 | Unassigned | Unassigned | Unassigned | 948_0 | Clustered | 9 | VC_948_0 | 9 | 0.5443 | 1 | 0.5443 | 0 | 0 | 0 | 1 | Ref |
| 5677 | MG592431 | Unassigned | Unassigned | Unassigned | 827_0 | Clustered | 6 | VC_827_0 | 6 | 0.7493 | 1 | 0.7493 | 0 | 0 | 0 | 1 | Ref |
| 5678 | MG592432 | Unassigned | Unassigned | Unassigned | 827_0 | Clustered | 6 | VC_827_0 | 6 | 0.7493 | 1 | 0.7493 | 0 | 0 | 0 | 1 | Ref |
| 5679 | MG592434 | Unassigned | Unassigned | Unassigned | 122_1 | Clustered/Singleton | 1 | VC_122_1 | 1 | 0 | 1 | 0 | 0 | 0 | 0 | 1 | Ref |
| 5680 | MG592435 | Unassigned | Unassigned | Unassigned | 827_0 | Clustered | 6 | VC_827_0 | 6 | 0.7493 | 1 | 0.7493 | 0 | 0 | 0 | 1 | Ref |
| 5681 | MG592436 | Unassigned | Unassigned | Unassigned | 1041_0 | Clustered | 9 | VC_1041_0 | 9 | 0.6541 | 1 | 0.6541 | 0 | 0 | 0 | 1 | Ref |
| 5682 | MG592438 | Unassigned | Unassigned | Unassigned | 948_0 | Clustered | 9 | VC_948_0 | 9 | 0.5443 | 1 | 0.5443 | 0 | 0 | 0 | 1 | Ref |
| 5683 | MG592439 | Unassigned | Unassigned | Unassigned | 645_0 | Clustered | 8 | VC_645_0 | 8 | 0.3138 | 1 | 0.3138 | 0 | 0 | 0 | 1 | Ref |
| 5684 | MG592441 | Unassigned | Unassigned | Unassigned | 493_8 | Clustered/Singleton | 1 | VC_493_8 | 1 | 0 | 1 | 0 | 0 | 0 | 0 | 1 | Ref |
| 5685 | MG592442 | Unassigned | Unassigned | Unassigned | 924_0 | Clustered | 34 | VC_924_0 | 34 | 0.8942 | 1 | 0.8942 | 0 | 0 | 0 | 0.9768 | Ref |
| 5686 | MG592443 | Unassigned | Unassigned | Unassigned | 1041_0 | Clustered | 9 | VC_1041_0 | 9 | 0.6541 | 1 | 0.6541 | 0 | 0 | 0 | 1 | Ref |
| 5687 | MG592444 | Unassigned | Unassigned | Unassigned | 645_0 | Clustered | 8 | VC_645_0 | 8 | 0.3138 | 1 | 0.3138 | 0 | 0 | 0 | 1 | Ref |
| 5688 | MG592445 | Unassigned | Unassigned | Unassigned | 1041_0 | Clustered | 9 | VC_1041_0 | 9 | 0.6541 | 1 | 0.6541 | 0 | 0 | 0 | 1 | Ref |
| 5689 | MG592447 | Unassigned | Unassigned | Unassigned | 544_0 | Clustered | 2 | VC_544_0 | 2 | 0.3517 | 0.94059057 | 0.3308 | 0 | 0 | 0 | 1 | Ref |
| 5690 | MG592448 | Unassigned | Unassigned | Unassigned | 122_0 | Clustered | 5 | VC_122_0 | 5 | 0.2073 | 0.99999996 | 0.2073 | 0 | 0 | 0 | 1 | Ref |
| 5691 | MG592449 | Unassigned | Unassigned | Unassigned | 1041_0 | Clustered | 9 | VC_1041_0 | 9 | 0.6541 | 1 | 0.6541 | 0 | 0 | 0 | 1 | Ref |
| 5692 | MG592450 | Unassigned | Unassigned | Unassigned | 924_0 | Clustered | 34 | VC_924_0 | 34 | 0.8942 | 1 | 0.8942 | 0 | 0 | 0 | 0.9768 | Ref |
| 5693 | MG592451 | Unassigned | Unassigned | Unassigned | 499_0 | Clustered | 24 | VC_499_0 | 24 | 0.8569 | 1 | 0.8569 | 0 | 0 | 0 | 0.9529 | Ref |
| 5694 | MG592453 | Unassigned | Unassigned | Unassigned | 544_0 | Clustered | 2 | VC_544_0 | 2 | 0.3517 | 0.94059057 | 0.3308 | 0 | 0 | 0 | 1 | Ref |
| 5695 | MG592454 | Unassigned | Unassigned | Unassigned | 645_0 | Clustered | 8 | VC_645_0 | 8 | 0.3138 | 1 | 0.3138 | 0 | 0 | 0 | 1 | Ref |
| 5696 | MG592455 | Unassigned | Unassigned | Unassigned | 1225_0 | Clustered | 4 | VC_1225_0 | 4 | 0.9619 | 0.99374885 | 0.9559 | 0 | 0 | 0 | 1 | Ref |
| 5697 | MG592456 | Unassigned | Unassigned | Unassigned | 531_4 | Clustered/Singleton | 1 | VC_531_4 | 1 | 0 | 1 | 0 | 0 | 0 | 0 | 1 | Ref |
| 5698 | MG592457 | Unassigned | Unassigned | Unassigned | 924_0 | Clustered | 34 | VC_924_0 | 34 | 0.8942 | 1 | 0.8942 | 0 | 0 | 0 | 0.9768 | Ref |
| 5699 | MG592458 | Unassigned | Unassigned | Unassigned | 827_0 | Clustered | 6 | VC_827_0 | 6 | 0.7493 | 1 | 0.7493 | 0 | 0 | 0 | 1 | Ref |
| 5700 | MG592459 | Unassigned | Unassigned | Unassigned | 497_0 | Clustered/Singleton | 1 | VC_497_0 | 1 | 0 | 1 | 0 | 0 | 0 | 0 | 1 | Ref |
| 5701 | MG592460 | Unassigned | Unassigned | Unassigned | 924_0 | Clustered | 34 | VC_924_0 | 34 | 0.8942 | 1 | 0.8942 | 0 | 0 | 0 | 0.9768 | Ref |
| 5702 | MG592461 | Unassigned | Unassigned | Unassigned | 499_0 | Clustered | 24 | VC_499_0 | 24 | 0.8569 | 1 | 0.8569 | 0 | 0 | 0 | 0.9529 | Ref |
| 5703 | MG592462 | Unassigned | Unassigned | Unassigned | 499_0 | Clustered | 24 | VC_499_0 | 24 | 0.8569 | 1 | 0.8569 | 0 | 0 | 0 | 0.9529 | Ref |
| 5704 | MG592463 | Unassigned | Unassigned | Unassigned | 1050_1 | Clustered | 29 | VC_1050_1 | 29 | 0.4861 | 1 | 0.4861 | 0 | 0 | 0 | 0.9581 | Ref |
| 5705 | MG592464 | Unassigned | Unassigned | Unassigned | 1050_1 | Clustered | 29 | VC_1050_1 | 29 | 0.4861 | 1 | 0.4861 | 0 | 0 | 0 | 0.9581 | Ref |
| 5706 | MG592465 | Unassigned | Unassigned | Unassigned | 645_0 | Clustered | 8 | VC_645_0 | 8 | 0.3138 | 1 | 0.3138 | 0 | 0 | 0 | 1 | Ref |
| 5707 | MG592466 | Unassigned | Unassigned | Unassigned | 645_0 | Clustered | 8 | VC_645_0 | 8 | 0.3138 | 1 | 0.3138 | 0 | 0 | 0 | 1 | Ref |
| 5708 | MG592467 | Unassigned | Unassigned | Unassigned | 499_0 | Clustered | 24 | VC_499_0 | 24 | 0.8569 | 1 | 0.8569 | 0 | 0 | 0 | 0.9529 | Ref |
| 5709 | MG592468 | Unassigned | Unassigned | Unassigned | 499_0 | Clustered | 24 | VC_499_0 | 24 | 0.8569 | 1 | 0.8569 | 0 | 0 | 0 | 0.9529 | Ref |
| 5710 | MG592470 | Unassigned | Unassigned | Unassigned | 930_0 | Clustered | 7 | VC_930_0 | 7 | 0.519 | 1 | 0.519 | 0 | 0 | 0 | 1 | Ref |
| 5711 | MG592471 | Unassigned | Unassigned | Unassigned | 1050_1 | Clustered | 29 | VC_1050_1 | 29 | 0.4861 | 1 | 0.4861 | 0 | 0 | 0 | 0.9581 | Ref |
| 5712 | MG592472 | Unassigned | Unassigned | Unassigned | 499_0 | Clustered | 24 | VC_499_0 | 24 | 0.8569 | 1 | 0.8569 | 0 | 0 | 0 | 0.9529 | Ref |
| 5713 | MG592473 | Unassigned | Unassigned | Unassigned | 497_1 | Clustered/Singleton | 1 | VC_497_1 | 1 | 0 | 1 | 0 | 0 | 0 | 0 | 1 | Ref |
| 5714 | MG592475 | Unassigned | Unassigned | Unassigned | 1050_1 | Clustered | 29 | VC_1050_1 | 29 | 0.4861 | 1 | 0.4861 | 0 | 0 | 0 | 0.9581 | Ref |
| 5715 | MG592476 | Unassigned | Unassigned | Unassigned | 1050_1 | Clustered | 29 | VC_1050_1 | 29 | 0.4861 | 1 | 0.4861 | 0 | 0 | 0 | 0.9581 | Ref |
| 5716 | MG592477 | Unassigned | Unassigned | Unassigned | 646_0 | Clustered | 29 | VC_646_0 | 29 | 0.9131 | 1 | 0.9131 | 0 | 0 | 0 | 1 | Ref |
| 5717 | MG592478 | Unassigned | Unassigned | Unassigned | 646_0 | Clustered | 29 | VC_646_0 | 29 | 0.9131 | 1 | 0.9131 | 0 | 0 | 0 | 1 | Ref |
| 5718 | MG592482 | Unassigned | Unassigned | Unassigned | 924_0 | Clustered | 34 | VC_924_0 | 34 | 0.8942 | 1 | 0.8942 | 0 | 0 | 0 | 0.9768 | Ref |
| 5719 | MG592483 | Unassigned | Unassigned | Unassigned | 924_0 | Clustered | 34 | VC_924_0 | 34 | 0.8942 | 1 | 0.8942 | 0 | 0 | 0 | 0.9768 | Ref |
| 5720 | MG592484 | Unassigned | Unassigned | Unassigned | 1190_0 | Clustered | 2 | VC_1190_0 | 2 | 0.5104 | 0.94764538 | 0.4837 | 0 | 0 | 0 | 1 | Ref |
| 5721 | MG592485 | Unassigned | Unassigned | Unassigned | 1190_0 | Clustered | 2 | VC_1190_0 | 2 | 0.5104 | 0.94764538 | 0.4837 | 0 | 0 | 0 | 1 | Ref |
| 5722 | MG592486 | Unassigned | Unassigned | Unassigned | 122_0 | Clustered | 5 | VC_122_0 | 5 | 0.2073 | 0.99999996 | 0.2073 | 0 | 0 | 0 | 1 | Ref |
| 5723 | MG592487 | Unassigned | Unassigned | Unassigned | 948_0 | Clustered | 9 | VC_948_0 | 9 | 0.5443 | 1 | 0.5443 | 0 | 0 | 0 | 1 | Ref |
| 5724 | MG592488 | Unassigned | Unassigned | Unassigned | 924_0 | Clustered | 34 | VC_924_0 | 34 | 0.8942 | 1 | 0.8942 | 0 | 0 | 0 | 0.9768 | Ref |
| 5725 | MG592489 | Unassigned | Unassigned | Unassigned | 924_0 | Clustered | 34 | VC_924_0 | 34 | 0.8942 | 1 | 0.8942 | 0 | 0 | 0 | 0.9768 | Ref |
| 5726 | MG592490 | Unassigned | Unassigned | Unassigned | 924_0 | Clustered | 34 | VC_924_0 | 34 | 0.8942 | 1 | 0.8942 | 0 | 0 | 0 | 0.9768 | Ref |
| 5727 | MG592492 | Unassigned | Unassigned | Unassigned | 1050_1 | Clustered | 29 | VC_1050_1 | 29 | 0.4861 | 1 | 0.4861 | 0 | 0 | 0 | 0.9581 | Ref |
| 5728 | MG592493 | Unassigned | Unassigned | Unassigned | 1050_1 | Clustered | 29 | VC_1050_1 | 29 | 0.4861 | 1 | 0.4861 | 0 | 0 | 0 | 0.9581 | Ref |
| 5729 | MG592494 | Unassigned | Unassigned | Unassigned | 642_3 | Clustered | 9 | VC_642_3 | 9 | 0.2131 | 1 | 0.2131 | 0 | 0 | 0 | 0.6944 | Ref |
| 5730 | MG592495 | Unassigned | Unassigned | Unassigned | 493_6 | Clustered/Singleton | 1 | VC_493_6 | 1 | 0 | 1 | 0 | 0 | 0 | 0 | 1 | Ref |
| 5731 | MG592496 | Unassigned | Unassigned | Unassigned | 944_0 | Clustered | 6 | VC_944_0 | 6 | 0.8196 | 1 | 0.8196 | 0 | 0 | 0 | 1 | Ref |
| 5732 | MG592497 | Unassigned | Unassigned | Unassigned | 944_0 | Clustered | 6 | VC_944_0 | 6 | 0.8196 | 1 | 0.8196 | 0 | 0 | 0 | 1 | Ref |
| 5733 | MG592499 | Unassigned | Unassigned | Unassigned | 499_0 | Clustered | 24 | VC_499_0 | 24 | 0.8569 | 1 | 0.8569 | 0 | 0 | 0 | 0.9529 | Ref |
| 5734 | MG592501 | Unassigned | Unassigned | Unassigned | 642_3 | Clustered | 9 | VC_642_3 | 9 | 0.2131 | 1 | 0.2131 | 0 | 0 | 0 | 0.6944 | Ref |
| 5735 | MG592502 | Unassigned | Unassigned | Unassigned | 924_0 | Clustered | 34 | VC_924_0 | 34 | 0.8942 | 1 | 0.8942 | 0 | 0 | 0 | 0.9768 | Ref |
| 5736 | MG592503 | Unassigned | Unassigned | Unassigned | 924_0 | Clustered | 34 | VC_924_0 | 34 | 0.8942 | 1 | 0.8942 | 0 | 0 | 0 | 0.9768 | Ref |
| 5737 | MG592504 | Unassigned | Unassigned | Unassigned | 924_0 | Clustered | 34 | VC_924_0 | 34 | 0.8942 | 1 | 0.8942 | 0 | 0 | 0 | 0.9768 | Ref |
| 5738 | MG592505 | Unassigned | Unassigned | Unassigned | 924_0 | Clustered | 34 | VC_924_0 | 34 | 0.8942 | 1 | 0.8942 | 0 | 0 | 0 | 0.9768 | Ref |
| 5739 | MG592506 | Unassigned | Unassigned | Unassigned | 642_0 | Clustered | 7 | VC_642_0 | 7 | 0.1508 | 1 | 0.1508 | 0 | 0 | 0 | 0.7619 | Ref |
| 5740 | MG592507 | Unassigned | Unassigned | Unassigned | 248_0 | Clustered | 11 | VC_248_0 | 11 | 0.7031 | 1 | 0.7031 | 0 | 0 | 0 | 1 | Ref |
| 5741 | MG592508 | Unassigned | Unassigned | Unassigned | 642_3 | Clustered | 9 | VC_642_3 | 9 | 0.2131 | 1 | 0.2131 | 0 | 0 | 0 | 0.6944 | Ref |
| 5742 | MG592509 | Unassigned | Unassigned | Unassigned | 501_0 | Clustered | 16 | VC_501_0 | 16 | 0.9985 | 1 | 0.9985 | 0 | 0 | 0 | 1 | Ref |
| 5743 | MG592510 | Unassigned | Unassigned | Unassigned | 543_0 | Clustered | 6 | VC_543_0 | 6 | 0.864 | 0.99999948 | 0.864 | 0 | 0 | 0 | 1 | Ref |
| 5744 | MG592511 | Unassigned | Unassigned | Unassigned | 543_0 | Clustered | 6 | VC_543_0 | 6 | 0.864 | 0.99999948 | 0.864 | 0 | 0 | 0 | 1 | Ref |
| 5745 | MG592513 | Unassigned | Unassigned | Unassigned | 248_0 | Clustered | 11 | VC_248_0 | 11 | 0.7031 | 1 | 0.7031 | 0 | 0 | 0 | 1 | Ref |
| 5746 | MG592514 | Unassigned | Unassigned | Unassigned | 642_3 | Clustered | 9 | VC_642_3 | 9 | 0.2131 | 1 | 0.2131 | 0 | 0 | 0 | 0.6944 | Ref |
| 5747 | MG592516 | Unassigned | Unassigned | Unassigned | 924_0 | Clustered | 34 | VC_924_0 | 34 | 0.8942 | 1 | 0.8942 | 0 | 0 | 0 | 0.9768 | Ref |
| 5748 | MG592517 | Unassigned | Unassigned | Unassigned | 924_0 | Clustered | 34 | VC_924_0 | 34 | 0.8942 | 1 | 0.8942 | 0 | 0 | 0 | 0.9768 | Ref |
| 5749 | MG592518 | Unassigned | Unassigned | Unassigned | 646_0 | Clustered | 29 | VC_646_0 | 29 | 0.9131 | 1 | 0.9131 | 0 | 0 | 0 | 1 | Ref |
| 5750 | MG592519 | Unassigned | Unassigned | Unassigned | 930_0 | Clustered | 7 | VC_930_0 | 7 | 0.519 | 1 | 0.519 | 0 | 0 | 0 | 1 | Ref |
| 5751 | MG592520 | Unassigned | Unassigned | Unassigned | 926_0 | Clustered | 3 | VC_926_0 | 3 | 0.9445 | 0.99430386 | 0.9391 | 0 | 0 | 0 | 1 | Ref |
| 5752 | MG592521 | Unassigned | Unassigned | Unassigned | 930_0 | Clustered | 7 | VC_930_0 | 7 | 0.519 | 1 | 0.519 | 0 | 0 | 0 | 1 | Ref |
| 5753 | MG592522 | Unassigned | Unassigned | Unassigned | 909_0 | Clustered | 5 | VC_909_0 | 5 | 0.7406 | 0.9999861 | 0.7406 | 0 | 0 | 0 | 1 | Ref |
| 5754 | MG592523 | Unassigned | Unassigned | Unassigned | 115_0 | Clustered | 2 | VC_115_0 | 2 | 0.4117 | 0.90744399 | 0.3736 | 0 | 0 | 0 | 1 | Ref |
| 5755 | MG592524 | Unassigned | Unassigned | Unassigned | 248_0 | Clustered | 11 | VC_248_0 | 11 | 0.7031 | 1 | 0.7031 | 0 | 0 | 0 | 1 | Ref |
| 5756 | MG592525 | Unassigned | Unassigned | Unassigned | 924_0 | Clustered | 34 | VC_924_0 | 34 | 0.8942 | 1 | 0.8942 | 0 | 0 | 0 | 0.9768 | Ref |
| 5757 | MG592526 | Unassigned | Unassigned | Unassigned | 499_0 | Clustered | 24 | VC_499_0 | 24 | 0.8569 | 1 | 0.8569 | 0 | 0 | 0 | 0.9529 | Ref |
| 5758 | MG592527 | Unassigned | Unassigned | Unassigned | 248_0 | Clustered | 11 | VC_248_0 | 11 | 0.7031 | 1 | 0.7031 | 0 | 0 | 0 | 1 | Ref |
| 5759 | MG592528 | Unassigned | Unassigned | Unassigned | 1050_1 | Clustered | 29 | VC_1050_1 | 29 | 0.4861 | 1 | 0.4861 | 0 | 0 | 0 | 0.9581 | Ref |
| 5760 | MG592529 | Unassigned | Unassigned | Unassigned | 493_2 | Clustered/Singleton | 1 | VC_493_2 | 1 | 0 | 1 | 0 | 0 | 0 | 0 | 1 | Ref |
| 5761 | MG592530 | Unassigned | Unassigned | Unassigned | 924_0 | Clustered | 34 | VC_924_0 | 34 | 0.8942 | 1 | 0.8942 | 0 | 0 | 0 | 0.9768 | Ref |
| 5762 | MG592531 | Unassigned | Unassigned | Unassigned | 499_0 | Clustered | 24 | VC_499_0 | 24 | 0.8569 | 1 | 0.8569 | 0 | 0 | 0 | 0.9529 | Ref |
| 5763 | MG592532 | Unassigned | Unassigned | Unassigned | 924_0 | Clustered | 34 | VC_924_0 | 34 | 0.8942 | 1 | 0.8942 | 0 | 0 | 0 | 0.9768 | Ref |
| 5764 | MG592533 | Unassigned | Unassigned | Unassigned | 1226_0 | Clustered | 2 | VC_1226_0 | 2 | 0.6497 | 0.86822376 | 0.564 | 0 | 0 | 0 | 1 | Ref |
| 5765 | MG592534 | Unassigned | Unassigned | Unassigned | 924_0 | Clustered | 34 | VC_924_0 | 34 | 0.8942 | 1 | 0.8942 | 0 | 0 | 0 | 0.9768 | Ref |
| 5766 | MG592535 | Unassigned | Unassigned | Unassigned | 924_0 | Clustered | 34 | VC_924_0 | 34 | 0.8942 | 1 | 0.8942 | 0 | 0 | 0 | 0.9768 | Ref |
| 5767 | MG592536 | Unassigned | Unassigned | Unassigned | 1227_0 | Clustered | 6 | VC_1227_0 | 6 | 0.9303 | 0.99999969 | 0.9303 | 0 | 0 | 0 | 1 | Ref |
| 5768 | MG592538 | Unassigned | Unassigned | Unassigned | 1050_0 | Clustered | 24 | VC_1050_0 | 24 | 0.5274 | 1 | 0.5274 | 0 | 0 | 0 | 1 | Ref |
| 5769 | MG592539 | Unassigned | Unassigned | Unassigned | 924_0 | Clustered | 34 | VC_924_0 | 34 | 0.8942 | 1 | 0.8942 | 0 | 0 | 0 | 0.9768 | Ref |
| 5770 | MG592540 | Unassigned | Unassigned | Unassigned | 646_0 | Clustered | 29 | VC_646_0 | 29 | 0.9131 | 1 | 0.9131 | 0 | 0 | 0 | 1 | Ref |
| 5771 | MG592541 | Unassigned | Unassigned | Unassigned | 499_0 | Clustered | 24 | VC_499_0 | 24 | 0.8569 | 1 | 0.8569 | 0 | 0 | 0 | 0.9529 | Ref |
| 5772 | MG592542 | Unassigned | Unassigned | Unassigned | 499_0 | Clustered | 24 | VC_499_0 | 24 | 0.8569 | 1 | 0.8569 | 0 | 0 | 0 | 0.9529 | Ref |
| 5773 | MG592543 | Unassigned | Unassigned | Unassigned | 924_0 | Clustered | 34 | VC_924_0 | 34 | 0.8942 | 1 | 0.8942 | 0 | 0 | 0 | 0.9768 | Ref |
| 5774 | MG592544 | Unassigned | Unassigned | Unassigned | 642_5 | Clustered | 4 | VC_642_5 | 4 | 0.0902 | 0.99996664 | 0.0902 | 0 | 0 | 0 | 1 | Ref |
| 5775 | MG592545 | Unassigned | Unassigned | Unassigned | 642_0 | Clustered | 7 | VC_642_0 | 7 | 0.1508 | 1 | 0.1508 | 0 | 0 | 0 | 0.7619 | Ref |
| 5776 | MG592546 | Unassigned | Unassigned | Unassigned | 501_0 | Clustered | 16 | VC_501_0 | 16 | 0.9985 | 1 | 0.9985 | 0 | 0 | 0 | 1 | Ref |
| 5777 | MG592547 | Unassigned | Unassigned | Unassigned | 499_0 | Clustered | 24 | VC_499_0 | 24 | 0.8569 | 1 | 0.8569 | 0 | 0 | 0 | 0.9529 | Ref |
| 5778 | MG592549 | Unassigned | Unassigned | Unassigned | 807_0 | Clustered | 17 | VC_807_0 | 17 | 0.907 | 1 | 0.907 | 0 | 0 | 0 | 1 | Ref |
| 5779 | MG592550 | Unassigned | Unassigned | Unassigned | 807_0 | Clustered | 17 | VC_807_0 | 17 | 0.907 | 1 | 0.907 | 0 | 0 | 0 | 1 | Ref |
| 5780 | MG592551 | Unassigned | Unassigned | Unassigned | 296_0 | Clustered | 6 | VC_296_0 | 6 | 0.5315 | 1 | 0.5315 | 0 | 0 | 0 | 1 | Ref |
| 5781 | MG592552 | Unassigned | Unassigned | Unassigned | 646_0 | Clustered | 29 | VC_646_0 | 29 | 0.9131 | 1 | 0.9131 | 0 | 0 | 0 | 1 | Ref |
| 5782 | MG592553 | Unassigned | Unassigned | Unassigned | 493_16 | Clustered/Singleton | 1 | VC_493_16 | 1 | 0 | 1 | 0 | 0 | 0 | 0 | 1 | Ref |
| 5783 | MG592554 | Unassigned | Unassigned | Unassigned | 1227_0 | Clustered | 6 | VC_1227_0 | 6 | 0.9303 | 0.99999969 | 0.9303 | 0 | 0 | 0 | 1 | Ref |
| 5784 | MG592555 | Unassigned | Unassigned | Unassigned | 1227_0 | Clustered | 6 | VC_1227_0 | 6 | 0.9303 | 0.99999969 | 0.9303 | 0 | 0 | 0 | 1 | Ref |
| 5785 | MG592556 | Unassigned | Unassigned | Unassigned | 1227_0 | Clustered | 6 | VC_1227_0 | 6 | 0.9303 | 0.99999969 | 0.9303 | 0 | 0 | 0 | 1 | Ref |
| 5786 | MG592557 | Unassigned | Unassigned | Unassigned | 924_0 | Clustered | 34 | VC_924_0 | 34 | 0.8942 | 1 | 0.8942 | 0 | 0 | 0 | 0.9768 | Ref |
| 5787 | MG592558 | Unassigned | Unassigned | Unassigned | 924_0 | Clustered | 34 | VC_924_0 | 34 | 0.8942 | 1 | 0.8942 | 0 | 0 | 0 | 0.9768 | Ref |
| 5788 | MG592559 | Unassigned | Unassigned | Unassigned | 924_0 | Clustered | 34 | VC_924_0 | 34 | 0.8942 | 1 | 0.8942 | 0 | 0 | 0 | 0.9768 | Ref |
| 5789 | MG592560 | Unassigned | Unassigned | Unassigned | 1226_0 | Clustered | 2 | VC_1226_0 | 2 | 0.6497 | 0.86822376 | 0.564 | 0 | 0 | 0 | 1 | Ref |
| 5790 | MG592561 | Unassigned | Unassigned | Unassigned | 499_0 | Clustered | 24 | VC_499_0 | 24 | 0.8569 | 1 | 0.8569 | 0 | 0 | 0 | 0.9529 | Ref |
| 5791 | MG592562 | Unassigned | Unassigned | Unassigned | 493_1 | Clustered/Singleton | 1 | VC_493_1 | 1 | 0 | 1 | 0 | 0 | 0 | 0 | 1 | Ref |
| 5792 | MG592563 | Unassigned | Unassigned | Unassigned | 1050_0 | Clustered | 24 | VC_1050_0 | 24 | 0.5274 | 1 | 0.5274 | 0 | 0 | 0 | 1 | Ref |
| 5793 | MG592564 | Unassigned | Unassigned | Unassigned | 1050_0 | Clustered | 24 | VC_1050_0 | 24 | 0.5274 | 1 | 0.5274 | 0 | 0 | 0 | 1 | Ref |
| 5794 | MG592565 | Unassigned | Unassigned | Unassigned | 1050_0 | Clustered | 24 | VC_1050_0 | 24 | 0.5274 | 1 | 0.5274 | 0 | 0 | 0 | 1 | Ref |
| 5795 | MG592566 | Unassigned | Unassigned | Unassigned | 948_0 | Clustered | 9 | VC_948_0 | 9 | 0.5443 | 1 | 0.5443 | 0 | 0 | 0 | 1 | Ref |
| 5796 | MG592567 | Unassigned | Unassigned | Unassigned | 642_0 | Clustered | 7 | VC_642_0 | 7 | 0.1508 | 1 | 0.1508 | 0 | 0 | 0 | 0.7619 | Ref |
| 5797 | MG592568 | Unassigned | Unassigned | Unassigned | 642_0 | Clustered | 7 | VC_642_0 | 7 | 0.1508 | 1 | 0.1508 | 0 | 0 | 0 | 0.7619 | Ref |
| 5798 | MG592569 | Unassigned | Unassigned | Unassigned | 646_0 | Clustered | 29 | VC_646_0 | 29 | 0.9131 | 1 | 0.9131 | 0 | 0 | 0 | 1 | Ref |
| 5799 | MG592570 | Unassigned | Unassigned | Unassigned | 646_0 | Clustered | 29 | VC_646_0 | 29 | 0.9131 | 1 | 0.9131 | 0 | 0 | 0 | 1 | Ref |
| 5800 | MG592571 | Unassigned | Unassigned | Unassigned | 1050_0 | Clustered | 24 | VC_1050_0 | 24 | 0.5274 | 1 | 0.5274 | 0 | 0 | 0 | 1 | Ref |
| 5801 | MG592572 | Unassigned | Unassigned | Unassigned | 499_0 | Clustered | 24 | VC_499_0 | 24 | 0.8569 | 1 | 0.8569 | 0 | 0 | 0 | 0.9529 | Ref |
| 5802 | MG592573 | Unassigned | Unassigned | Unassigned | 5_0 | Clustered | 30 | VC_5_0 | 30 | 0.9647 | 1 | 0.9647 | 0 | 0 | 0 | 1 | Ref |
| 5803 | MG592576 | Unassigned | Unassigned | Unassigned | 924_0 | Clustered | 34 | VC_924_0 | 34 | 0.8942 | 1 | 0.8942 | 0 | 0 | 0 | 0.9768 | Ref |
| 5804 | MG592578 | Unassigned | Unassigned | Unassigned | 175_0 | Clustered | 4 | VC_175_0 | 4 | 0.3847 | 0.99998259 | 0.3846 | 0 | 0 | 0 | 1 | Ref |
| 5805 | MG592579 | Unassigned | Unassigned | Unassigned | 543_0 | Clustered | 6 | VC_543_0 | 6 | 0.864 | 0.99999948 | 0.864 | 0 | 0 | 0 | 1 | Ref |
| 5806 | MG592581 | Unassigned | Unassigned | Unassigned | 807_0 | Clustered | 17 | VC_807_0 | 17 | 0.907 | 1 | 0.907 | 0 | 0 | 0 | 1 | Ref |
| 5807 | MG592582 | Unassigned | Unassigned | Unassigned | 807_0 | Clustered | 17 | VC_807_0 | 17 | 0.907 | 1 | 0.907 | 0 | 0 | 0 | 1 | Ref |
| 5808 | MG592583 | Unassigned | Unassigned | Unassigned | 642_3 | Clustered | 9 | VC_642_3 | 9 | 0.2131 | 1 | 0.2131 | 0 | 0 | 0 | 0.6944 | Ref |
| 5809 | MG592584 | Unassigned | Unassigned | Unassigned | 944_0 | Clustered | 6 | VC_944_0 | 6 | 0.8196 | 1 | 0.8196 | 0 | 0 | 0 | 1 | Ref |
| 5810 | MG592585 | Unassigned | Unassigned | Unassigned | 904_0 | Clustered | 6 | VC_904_0 | 6 | 0.233 | 1 | 0.233 | 0 | 0 | 0 | 1 | Ref |
| 5811 | MG592586 | Unassigned | Unassigned | Unassigned | 904_0 | Clustered | 6 | VC_904_0 | 6 | 0.233 | 1 | 0.233 | 0 | 0 | 0 | 1 | Ref |
| 5812 | MG592587 | Unassigned | Unassigned | Unassigned | 642_3 | Clustered | 9 | VC_642_3 | 9 | 0.2131 | 1 | 0.2131 | 0 | 0 | 0 | 0.6944 | Ref |
| 5813 | MG592588 | Unassigned | Unassigned | Unassigned | 248_0 | Clustered | 11 | VC_248_0 | 11 | 0.7031 | 1 | 0.7031 | 0 | 0 | 0 | 1 | Ref |
| 5814 | MG592589 | Unassigned | Unassigned | Unassigned | 248_0 | Clustered | 11 | VC_248_0 | 11 | 0.7031 | 1 | 0.7031 | 0 | 0 | 0 | 1 | Ref |
| 5815 | MG592590 | Unassigned | Unassigned | Unassigned | 499_0 | Clustered | 24 | VC_499_0 | 24 | 0.8569 | 1 | 0.8569 | 0 | 0 | 0 | 0.9529 | Ref |
| 5816 | MG592591 | Unassigned | Unassigned | Unassigned | 1227_0 | Clustered | 6 | VC_1227_0 | 6 | 0.9303 | 0.99999969 | 0.9303 | 0 | 0 | 0 | 1 | Ref |
| 5817 | MG592592 | Unassigned | Unassigned | Unassigned | 499_0 | Clustered | 24 | VC_499_0 | 24 | 0.8569 | 1 | 0.8569 | 0 | 0 | 0 | 0.9529 | Ref |
| 5818 | MG592593 | Unassigned | Unassigned | Unassigned | 924_0 | Clustered | 34 | VC_924_0 | 34 | 0.8942 | 1 | 0.8942 | 0 | 0 | 0 | 0.9768 | Ref |
| 5819 | MG592594 | Unassigned | Unassigned | Unassigned | 924_0 | Clustered | 34 | VC_924_0 | 34 | 0.8942 | 1 | 0.8942 | 0 | 0 | 0 | 0.9768 | Ref |
| 5820 | MG592595 | Unassigned | Unassigned | Unassigned | 642_3 | Clustered | 9 | VC_642_3 | 9 | 0.2131 | 1 | 0.2131 | 0 | 0 | 0 | 0.6944 | Ref |
| 5821 | MG592596 | Unassigned | Unassigned | Unassigned | 1229_0 | Clustered | 7 | VC_1229_0 | 7 | 0.9924 | 0.99999987 | 0.9924 | 0 | 0 | 0 | 1 | Ref |
| 5822 | MG592597 | Unassigned | Unassigned | Unassigned | 924_0 | Clustered | 34 | VC_924_0 | 34 | 0.8942 | 1 | 0.8942 | 0 | 0 | 0 | 0.9768 | Ref |
| 5823 | MG592598 | Unassigned | Unassigned | Unassigned | 924_0 | Clustered | 34 | VC_924_0 | 34 | 0.8942 | 1 | 0.8942 | 0 | 0 | 0 | 0.9768 | Ref |
| 5824 | MG592599 | Unassigned | Unassigned | Unassigned | 757_0 | Clustered | 2 | VC_757_0 | 2 | 0.0727 | 0.95527362 | 0.0695 | 0 | 0 | 0 | 1 | Ref |
| 5825 | MG592600 | Unassigned | Unassigned | Unassigned | 642_0 | Clustered | 7 | VC_642_0 | 7 | 0.1508 | 1 | 0.1508 | 0 | 0 | 0 | 0.7619 | Ref |
| 5826 | MG592601 | Unassigned | Unassigned | Unassigned | 1050_1 | Clustered | 29 | VC_1050_1 | 29 | 0.4861 | 1 | 0.4861 | 0 | 0 | 0 | 0.9581 | Ref |
| 5827 | MG592602 | Unassigned | Unassigned | Unassigned | 1050_1 | Clustered | 29 | VC_1050_1 | 29 | 0.4861 | 1 | 0.4861 | 0 | 0 | 0 | 0.9581 | Ref |
| 5828 | MG592603 | Unassigned | Unassigned | Unassigned | 782_0 | Clustered | 3 | VC_782_0 | 3 | 0.296 | 0.99856285 | 0.2956 | 0 | 0 | 0 | 1 | Ref |
| 5829 | MG592604 | Unassigned | Unassigned | Unassigned | 782_0 | Clustered | 3 | VC_782_0 | 3 | 0.296 | 0.99856285 | 0.2956 | 0 | 0 | 0 | 1 | Ref |
| 5830 | MG592605 | Unassigned | Unassigned | Unassigned | 924_0 | Clustered | 34 | VC_924_0 | 34 | 0.8942 | 1 | 0.8942 | 0 | 0 | 0 | 0.9768 | Ref |

| | | | | | | | | | | | | | | | | | |
|---|---|---|---|---|---|---|---|---|---|---|---|---|---|---|---|---|---|
| 5831 | MG592606 | Unassigned | Unassigned | Unassigned | 924_0 | Clustered | 34 | VC_924_0 | 34 | 0.8942 | 1 | 0.8942 | 0 | 0 | 0 | 0.9768 | Ref |
| 5832 | MG592607 | Unassigned | Unassigned | Unassigned | 924_0 | Clustered | 34 | VC_924_0 | 34 | 0.8942 | 1 | 0.8942 | 0 | 0 | 0 | 0.9768 | Ref |
| 5833 | MG592608 | Unassigned | Unassigned | Unassigned | 646_0 | Clustered | 29 | VC_646_0 | 29 | 0.9131 | 1 | 0.9131 | 0 | 0 | 0 | 1 | Ref |
| 5834 | MG592609 | Unassigned | Unassigned | Unassigned | 135_0 | Clustered | 2 | VC_135_0 | 2 | 0.0302 | 0.95557467 | 0.0288 | 0 | 0 | 0 | 1 | Ref |
| 5835 | MG592610 | Unassigned | Unassigned | Unassigned | 782_0 | Clustered | 3 | VC_782_0 | 3 | 0.296 | 0.99856285 | 0.2956 | 0 | 0 | 0 | 1 | Ref |
| 5836 | MG592611 | Unassigned | Unassigned | Unassigned | 499_0 | Clustered | 24 | VC_499_0 | 24 | 0.8569 | 1 | 0.8569 | 0 | 0 | 0 | 0.9529 | Ref |
| 5837 | MG592612 | Unassigned | Unassigned | Unassigned | 944_0 | Clustered | 6 | VC_944_0 | 6 | 0.8196 | 1 | 0.8196 | 0 | 0 | 0 | 1 | Ref |
| 5838 | MG592613 | Unassigned | Unassigned | Unassigned | 944_0 | Clustered | 6 | VC_944_0 | 6 | 0.8196 | 1 | 0.8196 | 0 | 0 | 0 | 1 | Ref |
| 5839 | MG592614 | Unassigned | Unassigned | Unassigned | 646_0 | Clustered | 29 | VC_646_0 | 29 | 0.9131 | 1 | 0.9131 | 0 | 0 | 0 | 1 | Ref |
| 5840 | MG592617 | Unassigned | Unassigned | Unassigned | 1050_1 | Clustered | 29 | VC_1050_1 | 29 | 0.4861 | 1 | 0.4861 | 0 | 0 | 0 | 0.9581 | Ref |
| 5841 | MG592618 | Unassigned | Unassigned | Unassigned | 1050_1 | Clustered | 29 | VC_1050_1 | 29 | 0.4861 | 1 | 0.4861 | 0 | 0 | 0 | 0.9581 | Ref |
| 5842 | MG592619 | Unassigned | Unassigned | Unassigned | 757_0 | Clustered | 2 | VC_757_0 | 2 | 0.0727 | 0.95527362 | 0.0695 | 0 | 0 | 0 | 1 | Ref |
| 5843 | MG592620 | Unassigned | Unassigned | Unassigned | 501_0 | Clustered | 16 | VC_501_0 | 16 | 0.9985 | 1 | 0.9985 | 0 | 0 | 0 | 1 | Ref |
| 5844 | MG592621 | Unassigned | Unassigned | Unassigned | 135_0 | Clustered | 2 | VC_135_0 | 2 | 0.0302 | 0.95557467 | 0.0288 | 0 | 0 | 0 | 1 | Ref |
| 5845 | MG592622 | Unassigned | Unassigned | Unassigned | 646_0 | Clustered | 29 | VC_646_0 | 29 | 0.9131 | 1 | 0.9131 | 0 | 0 | 0 | 1 | Ref |
| 5846 | MG592623 | Unassigned | Unassigned | Unassigned | 501_0 | Clustered | 16 | VC_501_0 | 16 | 0.9985 | 1 | 0.9985 | 0 | 0 | 0 | 1 | Ref |
| 5847 | MG592625 | Unassigned | Unassigned | Unassigned | 1227_0 | Clustered | 6 | VC_1227_0 | 6 | 0.9303 | 0.99999969 | 0.9303 | 0 | 0 | 0 | 1 | Ref |
| 5848 | MG592626 | Unassigned | Unassigned | Unassigned | 530_0 | Clustered | 9 | VC_530_0 | 9 | 0.8027 | 1 | 0.8027 | 0 | 0 | 0 | 1 | Ref |
| 5849 | MG592627 | Unassigned | Unassigned | Unassigned | 646_0 | Clustered | 29 | VC_646_0 | 29 | 0.9131 | 1 | 0.9131 | 0 | 0 | 0 | 1 | Ref |
| 5850 | MG592628 | Unassigned | Unassigned | Unassigned | 646_0 | Clustered | 29 | VC_646_0 | 29 | 0.9131 | 1 | 0.9131 | 0 | 0 | 0 | 1 | Ref |
| 5851 | MG592629 | Unassigned | Unassigned | Unassigned | 758_0 | Clustered | 18 | VC_758_0 | 18 | 0.738 | 1 | 0.738 | 0 | 0 | 0 | 1 | Ref |
| 5852 | MG592630 | Unassigned | Unassigned | Unassigned | 642_0 | Clustered | 7 | VC_642_0 | 7 | 0.1508 | 1 | 0.1508 | 0 | 0 | 0 | 0.7619 | Ref |
| 5853 | MG592631 | Unassigned | Unassigned | Unassigned | 924_0 | Clustered | 34 | VC_924_0 | 34 | 0.8942 | 1 | 0.8942 | 0 | 0 | 0 | 0.9768 | Ref |
| 5854 | MG592632 | Unassigned | Unassigned | Unassigned | 1050_0 | Clustered | 24 | VC_1050_0 | 24 | 0.5274 | 1 | 0.5274 | 0 | 0 | 0 | 1 | Ref |
| 5855 | MG592633 | Unassigned | Unassigned | Unassigned | 1050_0 | Clustered | 24 | VC_1050_0 | 24 | 0.5274 | 1 | 0.5274 | 0 | 0 | 0 | 1 | Ref |
| 5856 | MG592634 | Unassigned | Unassigned | Unassigned | 1050_0 | Clustered | 24 | VC_1050_0 | 24 | 0.5274 | 1 | 0.5274 | 0 | 0 | 0 | 1 | Ref |
| 5857 | MG592635 | Unassigned | Unassigned | Unassigned | 1050_0 | Clustered | 24 | VC_1050_0 | 24 | 0.5274 | 1 | 0.5274 | 0 | 0 | 0 | 1 | Ref |
| 5858 | MG592636 | Unassigned | Unassigned | Unassigned | 1050_0 | Clustered | 24 | VC_1050_0 | 24 | 0.5274 | 1 | 0.5274 | 0 | 0 | 0 | 1 | Ref |
| 5859 | MG592637 | Unassigned | Unassigned | Unassigned | 1050_0 | Clustered | 24 | VC_1050_0 | 24 | 0.5274 | 1 | 0.5274 | 0 | 0 | 0 | 1 | Ref |
| 5860 | MG592638 | Unassigned | Unassigned | Unassigned | 1050_0 | Clustered | 24 | VC_1050_0 | 24 | 0.5274 | 1 | 0.5274 | 0 | 0 | 0 | 1 | Ref |
| 5861 | MG592639 | Unassigned | Unassigned | Unassigned | 1050_0 | Clustered | 24 | VC_1050_0 | 24 | 0.5274 | 1 | 0.5274 | 0 | 0 | 0 | 1 | Ref |
| 5862 | MG592640 | Unassigned | Unassigned | Unassigned | 1050_0 | Clustered | 24 | VC_1050_0 | 24 | 0.5274 | 1 | 0.5274 | 0 | 0 | 0 | 1 | Ref |
| 5863 | MG592641 | Unassigned | Unassigned | Unassigned | 646_0 | Clustered | 29 | VC_646_0 | 29 | 0.9131 | 1 | 0.9131 | 0 | 0 | 0 | 1 | Ref |
| 5864 | MG592642 | Unassigned | Unassigned | Unassigned | 1050_0 | Clustered | 24 | VC_1050_0 | 24 | 0.5274 | 1 | 0.5274 | 0 | 0 | 0 | 1 | Ref |
| 5865 | MG592644 | Unassigned | Unassigned | Unassigned | 642_3 | Clustered | 9 | VC_642_3 | 9 | 0.2131 | 1 | 0.2131 | 0 | 0 | 0 | 0.6944 | Ref |
| 5866 | MG592645 | Unassigned | Unassigned | Unassigned | 1050_0 | Clustered | 24 | VC_1050_0 | 24 | 0.5274 | 1 | 0.5274 | 0 | 0 | 0 | 1 | Ref |
| 5867 | MG592646 | Unassigned | Unassigned | Unassigned | 1050_0 | Clustered | 24 | VC_1050_0 | 24 | 0.5274 | 1 | 0.5274 | 0 | 0 | 0 | 1 | Ref |
| 5868 | MG592647 | Unassigned | Unassigned | Unassigned | 646_0 | Clustered | 29 | VC_646_0 | 29 | 0.9131 | 1 | 0.9131 | 0 | 0 | 0 | 1 | Ref |
| 5869 | MG592648 | Unassigned | Unassigned | Unassigned | 1050_0 | Clustered | 24 | VC_1050_0 | 24 | 0.5274 | 1 | 0.5274 | 0 | 0 | 0 | 1 | Ref |
| 5870 | MG592649 | Unassigned | Unassigned | Unassigned | 1050_0 | Clustered | 24 | VC_1050_0 | 24 | 0.5274 | 1 | 0.5274 | 0 | 0 | 0 | 1 | Ref |
| 5871 | MG592650 | Unassigned | Unassigned | Unassigned | 1050_0 | Clustered | 24 | VC_1050_0 | 24 | 0.5274 | 1 | 0.5274 | 0 | 0 | 0 | 1 | Ref |
| 5872 | MG592651 | Unassigned | Unassigned | Unassigned | 1050_0 | Clustered | 24 | VC_1050_0 | 24 | 0.5274 | 1 | 0.5274 | 0 | 0 | 0 | 1 | Ref |
| 5873 | MG592652 | Unassigned | Unassigned | Unassigned | 1050_0 | Clustered | 24 | VC_1050_0 | 24 | 0.5274 | 1 | 0.5274 | 0 | 0 | 0 | 1 | Ref |
| 5874 | MG592653 | Unassigned | Unassigned | Unassigned | 1050_0 | Clustered | 24 | VC_1050_0 | 24 | 0.5274 | 1 | 0.5274 | 0 | 0 | 0 | 1 | Ref |
| 5875 | MG592654 | Unassigned | Unassigned | Unassigned | 948_0 | Clustered | 9 | VC_948_0 | 9 | 0.5443 | 1 | 0.5443 | 0 | 0 | 0 | 1 | Ref |
| 5876 | MG592655 | Unassigned | Unassigned | Unassigned | 1050_1 | Clustered | 29 | VC_1050_1 | 29 | 0.4861 | 1 | 0.4861 | 0 | 0 | 0 | 0.9581 | Ref |
| 5877 | MG592656 | Unassigned | Unassigned | Unassigned | 646_0 | Clustered | 29 | VC_646_0 | 29 | 0.9131 | 1 | 0.9131 | 0 | 0 | 0 | 1 | Ref |
| 5878 | MG592657 | Unassigned | Unassigned | Unassigned | 948_0 | Clustered | 9 | VC_948_0 | 9 | 0.5443 | 1 | 0.5443 | 0 | 0 | 0 | 1 | Ref |
| 5879 | MG592658 | Unassigned | Unassigned | Unassigned | 948_0 | Clustered | 9 | VC_948_0 | 9 | 0.5443 | 1 | 0.5443 | 0 | 0 | 0 | 1 | Ref |
| 5880 | MG592659 | Unassigned | Unassigned | Unassigned | 948_0 | Clustered | 9 | VC_948_0 | 9 | 0.5443 | 1 | 0.5443 | 0 | 0 | 0 | 1 | Ref |
| 5881 | MG592660 | Unassigned | Unassigned | Unassigned | 175_0 | Clustered | 4 | VC_175_0 | 4 | 0.3847 | 0.99998259 | 0.3846 | 0 | 0 | 0 | 1 | Ref |
| 5882 | MG592661 | Unassigned | Unassigned | Unassigned | 1041_0 | Clustered | 9 | VC_1041_0 | 9 | 0.6541 | 1 | 0.6541 | 0 | 0 | 0 | 1 | Ref |
| 5883 | MG592662 | Unassigned | Unassigned | Unassigned | 902_0 | Clustered | 3 | VC_902_0 | 3 | 0.9978 | 0.87589346 | 0.8739 | 0 | 0 | 0 | 1 | Ref |
| 5884 | MG592664 | Unassigned | Unassigned | Unassigned | 642_5 | Clustered | 4 | VC_642_5 | 4 | 0.0902 | 0.99996664 | 0.0902 | 0 | 0 | 0 | 1 | Ref |
| 5885 | MG592665 | Unassigned | Unassigned | Unassigned | 642_5 | Clustered | 4 | VC_642_5 | 4 | 0.0902 | 0.99996664 | 0.0902 | 0 | 0 | 0 | 1 | Ref |
| 5886 | MG592666 | Unassigned | Unassigned | Unassigned | 827_0 | Clustered | 6 | VC_827_0 | 6 | 0.7493 | 1 | 0.7493 | 0 | 0 | 0 | 1 | Ref |
| 5887 | MG592668 | Unassigned | Unassigned | Unassigned | 930_0 | Clustered | 7 | VC_930_0 | 7 | 0.519 | 1 | 0.519 | 0 | 0 | 0 | 1 | Ref |
| 5888 | MG592669 | Unassigned | Unassigned | Unassigned | 248_0 | Clustered | 11 | VC_248_0 | 11 | 0.7031 | 1 | 0.7031 | 0 | 0 | 0 | 1 | Ref |
| 5889 | MG592670 | Unassigned | Unassigned | Unassigned | 248_0 | Clustered | 11 | VC_248_0 | 11 | 0.7031 | 1 | 0.7031 | 0 | 0 | 0 | 1 | Ref |
| 5890 | MG592671 | Unassigned | Unassigned | Unassigned | 164_9 | Clustered/Singleton | 1 | VC_164_9 | 1 | 0 | 1 | 0 | 0 | 0 | 0 | 1 | Ref |
| 5891 | MG592672 | Unassigned | Unassigned | Unassigned | 902_0 | Clustered | 3 | VC_902_0 | 3 | 0.9978 | 0.87589346 | 0.8739 | 0 | 0 | 0 | 1 | Ref |
| 5892 | MG593800 | Unassigned | Unassigned | Unassigned | 375_0 | Clustered | 3 | VC_375_0 | 3 | 0.2479 | 0.99807286 | 0.2474 | 0 | 0 | 0 | 1 | Ref |
| 5893 | MG593802 | Unassigned | Unassigned | Unassigned | 981_1 | Clustered | 26 | VC_981_1 | 26 | 0.7009 | 1 | 0.7009 | 0 | 0 | 0 | 1 | Ref |
| 5894 | MG593803 | Unassigned | Unassigned | Unassigned | 375_0 | Clustered | 3 | VC_375_0 | 3 | 0.2479 | 0.99807286 | 0.2474 | 0 | 0 | 0 | 1 | Ref |
| 5895 | MG596799 | Unassigned | Unassigned | Unassigned | 1030_0 | Clustered | 3 | VC_1030_0 | 3 | 0.7895 | 0.99542416 | 0.7859 | 0 | 0 | 0 | 1 | Ref |
| 5896 | MG596800 | Unassigned | Unassigned | Unassigned | 965_0 | Clustered | 2 | VC_965_0 | 2 | 0.2408 | 0.95689931 | 0.2304 | 0 | 0 | 0 | 1 | Ref |
| 5897 | MG599035 | Unassigned | Unassigned | Unassigned | 699_0 | Clustered | 71 | VC_699_0 | 71 | 0.9352 | 1 | 0.9352 | 0 | 0 | 0 | 1 | Ref |
| 5898 | MG602476 | Unassigned | Unassigned | Unassigned | 882_0 | Clustered | 20 | VC_882_0 | 20 | 0.9917 | 1 | 0.9917 | 0 | 0 | 0 | 1 | Ref |
| 5899 | MG602477 | Unassigned | Unassigned | Unassigned | 187_3 | Clustered | 9 | VC_187_3 | 9 | 0.1971 | 1 | 0.1971 | 0 | 0 | 0 | 0.8611 | Ref |
| 5900 | MG640035 | Unassigned | Unassigned | Unassigned | 909_0 | Clustered | 5 | VC_909_0 | 5 | 0.7406 | 0.9999861 | 0.7406 | 0 | 0 | 0 | 1 | Ref |
| 5901 | MG641179 | Unassigned | Unassigned | Unassigned | 1104_0 | Clustered | 154 | VC_1104_0 | 154 | 0.8484 | 1 | 0.8484 | 0 | 0 | 0 | 1 | Ref |
| 5902 | MG641180 | Unassigned | Unassigned | Unassigned | 1104_0 | Clustered | 154 | VC_1104_0 | 154 | 0.8484 | 1 | 0.8484 | 0 | 0 | 0 | 1 | Ref |
| 5903 | MG641183 | Unassigned | Unassigned | Unassigned | 197_0 | Clustered | 5 | VC_197_0 | 5 | 0.1324 | 0.84698401 | 0.1122 | 0 | 0 | 0 | 1 | Ref |
| 5904 | MG641184 | Unassigned | Unassigned | Unassigned | 1133_0 | Clustered | 4 | VC_1133_0 | 4 | 0.1253 | 0.975067 | 0.1222 | 0 | 0 | 0 | 1 | Ref |
| 5905 | MG641185 | Unassigned | Unassigned | Unassigned | 1232_0 | Clustered | 2 | VC_1232_0 | 2 | 0.4709 | 0.85557782 | 0.4029 | 0 | 0 | 0 | 1 | Ref |
| 5906 | MG641186 | Unassigned | Unassigned | Unassigned | 1104_0 | Clustered | 154 | VC_1104_0 | 154 | 0.8484 | 1 | 0.8484 | 0 | 0 | 0 | 1 | Ref |
| 5907 | MG641188 | Unassigned | Unassigned | Unassigned | 1233_0 | Clustered | 16 | VC_1233_0 | 16 | 0.8971 | 1 | 0.8971 | 0 | 0 | 0 | 1 | Ref |
| 5908 | MG646668 | Unassigned | Unassigned | Unassigned | 637_0 | Clustered | 115 | VC_637_0 | 115 | 0.731 | 1 | 0.731 | 0 | 0 | 0 | 0.8764 | Ref |
| 5909 | MG646669 | Unassigned | Unassigned | Unassigned | 329_11 | Clustered | 15 | VC_329_11 | 15 | 0.0791 | 1 | 0.0791 | 0 | 0 | 0 | 0.9333 | Ref |
| 5910 | MG646670 | Unassigned | Unassigned | Unassigned | 329_11 | Clustered | 15 | VC_329_11 | 15 | 0.0791 | 1 | 0.0791 | 0 | 0 | 0 | 0.9333 | Ref |
| 5911 | MG646671 | Unassigned | Unassigned | Unassigned | 329_11 | Clustered | 15 | VC_329_11 | 15 | 0.0791 | 1 | 0.0791 | 0 | 0 | 0 | 0.9333 | Ref |
| 5912 | MG646672 | Unassigned | Unassigned | Unassigned | 329_0 | Clustered | 2 | VC_329_0 | 2 | 0.0051 | 0.95464582 | 0.0048 | 0 | 0 | 0 | 1 | Ref |
| 5913 | MG649966 | Unassigned | Unassigned | Unassigned | 971_1 | Clustered | 24 | VC_971_1 | 24 | 0.7393 | 1 | 0.7393 | 0 | 0 | 0 | 0.8913 | Ref |
| 5914 | MG649967 | Unassigned | Unassigned | Unassigned | 971_1 | Clustered | 24 | VC_971_1 | 24 | 0.7393 | 1 | 0.7393 | 0 | 0 | 0 | 0.8913 | Ref |
| 5915 | MG655267 | Unassigned | Unassigned | Unassigned | 489_1 | Clustered | 11 | VC_489_1 | 11 | 0.4621 | 1 | 0.4621 | 0 | 0 | 0 | 1 | Ref |
| 5916 | MG655268 | Unassigned | Unassigned | Unassigned | 489_1 | Clustered | 11 | VC_489_1 | 11 | 0.4621 | 1 | 0.4621 | 0 | 0 | 0 | 1 | Ref |
| 5917 | MG655269 | Unassigned | Unassigned | Unassigned | 489_1 | Clustered | 11 | VC_489_1 | 11 | 0.4621 | 1 | 0.4621 | 0 | 0 | 0 | 1 | Ref |
| 5918 | MG655270 | Unassigned | Unassigned | Unassigned | 489_1 | Clustered | 11 | VC_489_1 | 11 | 0.4621 | 1 | 0.4621 | 0 | 0 | 0 | 1 | Ref |
| 5919 | MG656408 | Unassigned | Unassigned | Unassigned | 149_8 | Clustered | 66 | VC_149_8 | 66 | 0.4565 | 1 | 0.4565 | 0 | 0 | 0 | 0.8182 | Ref |
| 5920 | MG663582 | Unassigned | Unassigned | Unassigned | 1224_0 | Clustered | 19 | VC_1224_0 | 19 | 1 | 1 | 1 | 0 | 0 | 0 | 1 | Ref |
| 5921 | MG663583 | Unassigned | Unassigned | Unassigned | 1224_0 | Clustered | 19 | VC_1224_0 | 19 | 1 | 1 | 1 | 0 | 0 | 0 | 1 | Ref |
| 5922 | MG663584 | Unassigned | Unassigned | Unassigned | 1224_0 | Clustered | 19 | VC_1224_0 | 19 | 1 | 1 | 1 | 0 | 0 | 0 | 1 | Ref |
| 5923 | MG663585 | Unassigned | Unassigned | Unassigned | 1224_0 | Clustered | 19 | VC_1224_0 | 19 | 1 | 1 | 1 | 0 | 0 | 0 | 1 | Ref |
| 5924 | MG663586 | Unassigned | Unassigned | Unassigned | 1224_0 | Clustered | 19 | VC_1224_0 | 19 | 1 | 1 | 1 | 0 | 0 | 0 | 1 | Ref |
| 5925 | MG670586 | Unassigned | Unassigned | Unassigned | 1234_0 | Clustered | 44 | VC_1234_0 | 44 | 1 | 1 | 1 | 0 | 0 | 0 | 1 | Ref |
| 5926 | MG673519 | Unassigned | Unassigned | Unassigned | 813_1 | Clustered | 24 | VC_813_1 | 24 | 0.535 | 1 | 0.535 | 0 | 0 | 0 | 1 | Ref |
| 5927 | MG674163 | Unassigned | Unassigned | Unassigned | 608_0 | Clustered | 5 | VC_608_0 | 5 | 0.4331 | 0.99999996 | 0.4331 | 0 | 0 | 0 | 1 | Ref |
| 5928 | MG676223 | Unassigned | Unassigned | Unassigned | 642_2 | Clustered | 6 | VC_642_2 | 6 | 0.1679 | 1 | 0.1679 | 0 | 0 | 0 | 1 | Ref |
| 5929 | MG676224 | Unassigned | Unassigned | Unassigned | 195_0 | Clustered | 2 | VC_195_0 | 2 | 0.1201 | 0.95680007 | 0.1149 | 0 | 0 | 0 | 1 | Ref |
| 5930 | MG676225 | Unassigned | Unassigned | Unassigned | 195_0 | Clustered | 2 | VC_195_0 | 2 | 0.1201 | 0.95680007 | 0.1149 | 0 | 0 | 0 | 1 | Ref |
| 5931 | MG676466 | Unassigned | Unassigned | Unassigned | 755_0 | Clustered | 9 | VC_755_0 | 9 | 0.4489 | 1 | 0.4489 | 0 | 0 | 0 | 1 | Ref |
| 5932 | MG696114 | Unassigned | Unassigned | Unassigned | 288_2 | Clustered | 2 | VC_288_2 | 2 | 0.0228 | 0.95538559 | 0.0218 | 0 | 0 | 0 | 1 | Ref |
| 5933 | MG707188 | Unassigned | Unassigned | Unassigned | 173_0 | Clustered | 2 | VC_173_0 | 2 | 0.0799 | 0.95489091 | 0.0763 | 0 | 0 | 0 | 1 | Ref |
| 5934 | MG708273 | Unassigned | Unassigned | Unassigned | 252_0 | Clustered | 121 | VC_252_0 | 121 | 0.7376 | 1 | 0.7376 | 0 | 0 | 0 | 0.9999 | Ref |
| 5935 | MG708274 | Unassigned | Unassigned | Unassigned | 252_0 | Clustered | 121 | VC_252_0 | 121 | 0.7376 | 1 | 0.7376 | 0 | 0 | 0 | 0.9999 | Ref |
| 5936 | MG708275 | Unassigned | Unassigned | Unassigned | 252_0 | Clustered | 121 | VC_252_0 | 121 | 0.7376 | 1 | 0.7376 | 0 | 0 | 0 | 0.9999 | Ref |
| 5937 | MG708276 | Unassigned | Unassigned | Unassigned | 695_0 | Clustered | 48 | VC_695_0 | 48 | 0.8162 | 1 | 0.8162 | 0 | 0 | 0 | 1 | Ref |
| 5938 | MG710528 | Unassigned | Unassigned | Unassigned | 42_0 | Clustered | 49 | VC_42_0 | 49 | 0.4517 | 1 | 0.4517 | 0 | 0 | 0 | 0.8597 | Ref |
| 5939 | MG711460 | Unassigned | Unassigned | Unassigned | 1235_0 | Clustered | 12 | VC_1235_0 | 12 | 0.7997 | 1 | 0.7997 | 0 | 0 | 0 | 1 | Ref |
| 5940 | MG711461 | Unassigned | Unassigned | Unassigned | 717_0 | Clustered | 7 | VC_717_0 | 7 | 0.5813 | 1 | 0.5813 | 0 | 0 | 0 | 0.9048 | Ref |
| 5941 | MG711462 | Unassigned | Unassigned | Unassigned | 707_1 | Clustered | 9 | VC_707_1 | 9 | 0.2702 | 1 | 0.2702 | 0 | 0 | 0 | 0.8889 | Ref |
| 5942 | MG711463 | Unassigned | Unassigned | Unassigned | 804_0 | Clustered | 2 | VC_804_0 | 2 | 0.0559 | 0.95690755 | 0.0535 | 0 | 0 | 0 | 1 | Ref |
| 5943 | MG711464 | Unassigned | Unassigned | Unassigned | 621_0 | Clustered | 7 | VC_621_0 | 7 | 0.5783 | 1 | 0.5783 | 0 | 0 | 0 | 1 | Ref |
| 5944 | MG711465 | Unassigned | Unassigned | Unassigned | 606_0 | Clustered | 4 | VC_606_0 | 4 | 0.4507 | 0.99995146 | 0.4507 | 0 | 0 | 0 | 1 | Ref |
| 5945 | MG711466 | Unassigned | Unassigned | Unassigned | 710_0 | Clustered | 10 | VC_710_0 | 10 | 0.2446 | 1 | 0.2446 | 0 | 0 | 0 | 0.8222 | Ref |
| 5946 | MG711467 | Unassigned | Unassigned | Unassigned | 711_0 | Clustered | 3 | VC_711_0 | 3 | 0.2312 | 0.99842495 | 0.2309 | 0 | 0 | 0 | 1 | Ref |
| 5947 | MG720309 | Unassigned | Unassigned | Unassigned | 904_0 | Clustered | 6 | VC_904_0 | 6 | 0.233 | 1 | 0.233 | 0 | 0 | 0 | 1 | Ref |
| 5948 | MG721208 | Unassigned | Unassigned | Unassigned | 149_21 | Clustered/Singleton | 1 | VC_149_21 | 1 | 0 | 1 | 0 | 0 | 0 | 0 | 1 | Ref |
| 5949 | MG727695 | Unassigned | Unassigned | Unassigned | 708_0 | Clustered | 47 | VC_708_0 | 47 | 0.8654 | 1 | 0.8654 | 0 | 0 | 0 | 1 | Ref |
| 5950 | MG727696 | Unassigned | Unassigned | Unassigned | 708_0 | Clustered | 47 | VC_708_0 | 47 | 0.8654 | 1 | 0.8654 | 0 | 0 | 0 | 1 | Ref |
| 5951 | MG727698 | Unassigned | Unassigned | Unassigned | 708_0 | Clustered | 47 | VC_708_0 | 47 | 0.8654 | 1 | 0.8654 | 0 | 0 | 0 | 1 | Ref |
| 5952 | MG727699 | Unassigned | Unassigned | Unassigned | 708_0 | Clustered | 47 | VC_708_0 | 47 | 0.8654 | 1 | 0.8654 | 0 | 0 | 0 | 1 | Ref |
| 5953 | MG727700 | Unassigned | Unassigned | Unassigned | 708_0 | Clustered | 47 | VC_708_0 | 47 | 0.8654 | 1 | 0.8654 | 0 | 0 | 0 | 1 | Ref |
| 5954 | MG727701 | Unassigned | Unassigned | Unassigned | 708_0 | Clustered | 47 | VC_708_0 | 47 | 0.8654 | 1 | 0.8654 | 0 | 0 | 0 | 1 | Ref |
| 5955 | MG727702 | Unassigned | Unassigned | Unassigned | 708_0 | Clustered | 47 | VC_708_0 | 47 | 0.8654 | 1 | 0.8654 | 0 | 0 | 0 | 1 | Ref |
| 5956 | MG736918 | Unassigned | Unassigned | Unassigned | 453_0 | Clustered | 6 | VC_453_0 | 6 | 0.114 | 1 | 0.114 | 0 | 0 | 0 | 1 | Ref |
| 5957 | MG744354 | Unassigned | Unassigned | Unassigned | 634_1 | Clustered | 6 | VC_634_1 | 6 | 0.4228 | 1 | 0.4228 | 0 | 0 | 0 | 1 | Ref |
| 5958 | MG747435 | Unassigned | Unassigned | Unassigned | 1_0 | Clustered | 54 | VC_1_0 | 54 | 0.8527 | 1 | 0.8527 | 0 | 0 | 0 | 1 | Ref |
| 5959 | MG748547 | Unassigned | Unassigned | Unassigned | 133_5 | Clustered/Singleton | 1 | VC_133_5 | 1 | 0 | 1 | 0 | 0 | 0 | 0 | 1 | Ref |
| 5960 | MG748548 | Unassigned | Unassigned | Unassigned | 813_1 | Clustered | 24 | VC_813_1 | 24 | 0.535 | 1 | 0.535 | 0 | 0 | 0 | 1 | Ref |
| 5961 | MG751100 | Unassigned | Unassigned | Unassigned | 491_0 | Clustered | 36 | VC_491_0 | 36 | 0.5336 | 1 | 0.5336 | 0 | 0 | 0 | 0.9714 | Ref |
| 5962 | MG757152 | Unassigned | Unassigned | Unassigned | 868_0 | Clustered | 25 | VC_868_0 | 25 | 0.5791 | 1 | 0.5791 | 0 | 0 | 0 | 1 | Ref |
| 5963 | MG757153 | Unassigned | Unassigned | Unassigned | 1072_0 | Clustered | 2 | VC_1072_0 | 2 | 0.0264 | 0.95117401 | 0.0251 | 0 | 0 | 0 | 1 | Ref |
| 5964 | MG757154 | Unassigned | Unassigned | Unassigned | 872_0 | Clustered | 31 | VC_872_0 | 31 | 0.8151 | 1 | 0.8151 | 0 | 0 | 0 | 0.8194 | Ref |
| 5965 | MG757155 | Unassigned | Unassigned | Unassigned | 868_0 | Clustered | 25 | VC_868_0 | 25 | 0.5791 | 1 | 0.5791 | 0 | 0 | 0 | 1 | Ref |
| 5966 | MG757156 | Unassigned | Unassigned | Unassigned | 571_0 | Clustered | 4 | VC_571_0 | 4 | 0.1921 | 0.99998591 | 0.1921 | 0 | 0 | 0 | 1 | Ref |
| 5967 | MG757157 | Unassigned | Unassigned | Unassigned | 767_0 | Clustered | 34 | VC_767_0 | 34 | 0.9173 | 1 | 0.9173 | 0 | 0 | 0 | 1 | Ref |
| 5968 | MG757158 | Unassigned | Unassigned | Unassigned | 371_16 | Clustered | 42 | VC_371_16 | 42 | 0.1341 | 1 | 0.1341 | 0 | 0 | 0 | 0.7282 | Ref |
| 5969 | MG757159 | Unassigned | Unassigned | Unassigned | 371_11 | Clustered | 31 | VC_371_11 | 31 | 0.0927 | 1 | 0.0927 | 0 | 0 | 0 | 0.7011 | Ref |
| 5970 | MG757160 | Unassigned | Unassigned | Unassigned | 551_6 | Clustered | 22 | VC_551_6 | 22 | 0.1214 | 1 | 0.1214 | 0 | 0 | 0 | 0.8095 | Ref |
| 5971 | MG757161 | Unassigned | Unassigned | Unassigned | 514_0 | Clustered | 75 | VC_514_0 | 75 | 0.9402 | 1 | 0.9402 | 0 | 0 | 0 | 1 | Ref |
| 5972 | MG757162 | Unassigned | Unassigned | Unassigned | 508_0 | Clustered | 32 | VC_508_0 | 32 | 0.906 | 1 | 0.906 | 0 | 0 | 0 | 1 | Ref |
| 5973 | MG757163 | Unassigned | Unassigned | Unassigned | 982_0 | Clustered | 62 | VC_982_0 | 62 | 0.8646 | 1 | 0.8646 | 0 | 0 | 0 | 0.743 | Ref |
| 5974 | MG757164 | Unassigned | Unassigned | Unassigned | 371_13 | Clustered | 33 | VC_371_13 | 33 | 0.179 | 1 | 0.179 | 0 | 0 | 0 | 0.8769 | Ref |
| 5975 | MG757165 | Unassigned | Unassigned | Unassigned | 371_13 | Clustered | 33 | VC_371_13 | 33 | 0.179 | 1 | 0.179 | 0 | 0 | 0 | 0.8769 | Ref |
| 5976 | MG757166 | Unassigned | Unassigned | Unassigned | 767_0 | Clustered | 34 | VC_767_0 | 34 | 0.9173 | 1 | 0.9173 | 0 | 0 | 0 | 1 | Ref |
| 5977 | MG757167 | Unassigned | Unassigned | Unassigned | 939_0 | Clustered | 15 | VC_939_0 | 15 | 1 | 0 | 0 | 0 | 0 | 0 | 1 | Ref |
| 5978 | MG763894 | Unassigned | Unassigned | Unassigned | 884_0 | Clustered | 28 | VC_884_0 | 28 | 0.6211 | 1 | 0.6211 | 0 | 0 | 0 | 0.9841 | Ref |
| 5979 | MG765274 | Unassigned | Unassigned | Unassigned | 634_1 | Clustered | 6 | VC_634_1 | 6 | 0.4228 | 1 | 0.4228 | 0 | 0 | 0 | 1 | Ref |
| 5980 | MG765275 | Unassigned | Unassigned | Unassigned | 577_0 | Clustered | 6 | VC_577_0 | 6 | 0.9267 | 0.99999993 | 0.9267 | 0 | 0 | 0 | 1 | Ref |
| 5981 | MG765276 | Unassigned | Unassigned | Unassigned | 634_0 | Clustered | 3 | VC_634_0 | 3 | 0.2046 | 0.99837593 | 0.2043 | 0 | 0 | 0 | 1 | Ref |
| 5982 | MG765277 | Unassigned | Unassigned | Unassigned | 526_0 | Clustered | 6 | VC_526_0 | 6 | 0.4386 | 1 | 0.4386 | 0 | 0 | 0 | 1 | Ref |
| 5983 | MG765278 | Unassigned | Unassigned | Unassigned | 577_0 | Clustered | 6 | VC_577_0 | 6 | 0.9267 | 0.99999993 | 0.9267 | 0 | 0 | 0 | 1 | Ref |
| 5984 | MG765279 | Unassigned | Unassigned | Unassigned | 526_0 | Clustered | 6 | VC_526_0 | 6 | 0.4386 | 1 | 0.4386 | 0 | 0 | 0 | 1 | Ref |
| 5985 | MG766218 | Unassigned | Unassigned | Unassigned | 96_0 | Clustered | 30 | VC_96_0 | 30 | 0.718 | 1 | 0.718 | 0 | 0 | 0 | 1 | Ref |
| 5986 | MG766219 | Unassigned | Unassigned | Unassigned | 96_0 | Clustered | 30 | VC_96_0 | 30 | 0.718 | 1 | 0.718 | 0 | 0 | 0 | 1 | Ref |
| 5987 | MG770211 | Unassigned | Unassigned | Unassigned | 868_0 | Clustered | 25 | VC_868_0 | 25 | 0.5791 | 1 | 0.5791 | 0 | 0 | 0 | 1 | Ref |
| 5988 | MG770212 | Unassigned | Unassigned | Unassigned | 371_15 | Clustered | 44 | VC_371_15 | 44 | 0.1391 | 1 | 0.1391 | 0 | 0 | 0 | 0.7474 | Ref |
| 5989 | MG770214 | Unassigned | Unassigned | Unassigned | 868_0 | Clustered | 25 | VC_868_0 | 25 | 0.5791 | 1 | 0.5791 | 0 | 0 | 0 | 1 | Ref |
| 5990 | MG770215 | Unassigned | Unassigned | Unassigned | 919_0 | Clustered | 39 | VC_919_0 | 39 | 0.9069 | 1 | 0.9069 | 0 | 0 | 0 | 1 | Ref |
| 5991 | MG770216 | Unassigned | Unassigned | Unassigned | 809_0 | Clustered | 2 | VC_809_0 | 2 | 0.1799 | 0.96299906 | 0.1733 | 0 | 0 | 0 | 1 | Ref |
| 5992 | MG770228 | Unassigned | Unassigned | Unassigned | 779_0 | Clustered | 37 | VC_779_0 | 37 | 0.5682 | 1 | 0.5682 | 0 | 0 | 0 | 1 | Ref |

| | | | | | | | | | | | | | | | | | |
|---|---|---|---|---|---|---|---|---|---|---|---|---|---|---|---|---|---|
| 5993 | MG770379 | Unassigned | Unassigned | Unassigned | 132_1 | Clustered | 17 | VC_132_1 | 17 | 0.1738 | 1 | 0.1738 | 0 | 0 | 0 | 0.9853 | Ref |
| 5994 | MG770897 | Unassigned | Unassigned | Unassigned | 8_0 | Clustered | 58 | VC_8_0 | 58 | 0.8306 | 1 | 0.8306 | 0 | 0 | 0 | 0.9946 | Ref |
| 5995 | MG775042 | Unassigned | Unassigned | Unassigned | 452_0 | Clustered | 66 | VC_452_0 | 66 | 0.9248 | 1 | 0.9248 | 0 | 0 | 0 | 1 | Ref |
| 5996 | MG775043 | Unassigned | Unassigned | Unassigned | 283_24 | Clustered/Singleton | 1 | VC_283_24 | 1 | 0 | 1 | 0 | 0 | 0 | 0 | 1 | Ref |
| 5997 | MG775258 | Unassigned | Unassigned | Unassigned | 129_0 | Clustered | 43 | VC_129_0 | 43 | 0.886 | 1 | 0.886 | 0 | 0 | 0 | 1 | Ref |
| 5998 | MG775260 | Unassigned | Unassigned | Unassigned | 842_0 | Clustered | 2 | VC_842_0 | 2 | 0.0559 | 0.95564851 | 0.0534 | 0 | 0 | 0 | 1 | Ref |
| 5999 | MG781190 | Unassigned | Unassigned | Unassigned | 283_89 | Clustered | 2 | VC_283_89 | 2 | 0.0027 | 0.91264761 | 0.0025 | 0 | 0 | 0 | 1 | Ref |
| 6000 | MG781191 | Unassigned | Unassigned | Unassigned | 283_89 | Clustered | 2 | VC_283_89 | 2 | 0.0027 | 0.91264761 | 0.0025 | 0 | 0 | 0 | 1 | Ref |
| 6001 | MG784342 | Unassigned | Unassigned | Unassigned | 828_0 | Clustered | 3 | VC_828_0 | 3 | 0.4748 | 0.99843555 | 0.4741 | 0 | 0 | 0 | 1 | Ref |
| 6002 | MG788324 | Unassigned | Unassigned | Unassigned | 300_0 | Clustered | 3 | VC_300_0 | 3 | 0.1218 | 0.99841701 | 0.1216 | 0 | 0 | 0 | 0.6667 | Ref |
| 6003 | MG793454 | Unassigned | Unassigned | Unassigned | 219_0 | Clustered | 5 | VC_219_0 | 5 | 0.08 | 0.99999998 | 0.08 | 0 | 0 | 0 | 1 | Ref |
| 6004 | MG812486 | Unassigned | Unassigned | Unassigned | 325_50 | Clustered | 4 | VC_325_50 | 4 | 0.0182 | 0.99998785 | 0.0182 | 0 | 0 | 0 | 1 | Ref |
| 6005 | MG812487 | Unassigned | Unassigned | Unassigned | 1093_0 | Clustered | 40 | VC_1093_0 | 40 | 0.8754 | 1 | 0.8754 | 0 | 0 | 0 | 1 | Ref |
| 6006 | MG812488 | Unassigned | Unassigned | Unassigned | 520_98 | Clustered/Singleton | 1 | VC_520_98 | 1 | 0 | 1 | 0 | 0 | 0 | 0 | 1 | Ref |
| 6007 | MG812489 | Unassigned | Unassigned | Unassigned | 325_50 | Clustered | 4 | VC_325_50 | 4 | 0.0182 | 0.99998785 | 0.0182 | 0 | 0 | 0 | 1 | Ref |
| 6008 | MG812490 | Unassigned | Unassigned | Unassigned | 508_0 | Clustered | 32 | VC_508_0 | 32 | 0.906 | 1 | 0.906 | 0 | 0 | 0 | 1 | Ref |
| 6009 | MG812491 | Unassigned | Unassigned | Unassigned | 508_0 | Clustered | 32 | VC_508_0 | 32 | 0.906 | 1 | 0.906 | 0 | 0 | 0 | 1 | Ref |
| 6010 | MG812492 | Unassigned | Unassigned | Unassigned | 513_0 | Clustered | 32 | VC_513_0 | 32 | 0.6881 | 1 | 0.6881 | 0 | 0 | 0 | 1 | Ref |
| 6011 | MG812493 | Unassigned | Unassigned | Unassigned | 558_72 | Clustered/Singleton | 1 | VC_558_72 | 1 | 0 | 1 | 0 | 0 | 0 | 0 | 1 | Ref |
| 6012 | MG812494 | Unassigned | Unassigned | Unassigned | 558_68 | Clustered/Singleton | 1 | VC_558_68 | 1 | 0 | 1 | 0 | 0 | 0 | 0 | 1 | Ref |
| 6013 | MG812495 | Unassigned | Unassigned | Unassigned | 325_50 | Clustered | 4 | VC_325_50 | 4 | 0.0182 | 0.99998785 | 0.0182 | 0 | 0 | 0 | 1 | Ref |
| 6014 | MG812496 | Unassigned | Unassigned | Unassigned | 949_0 | Clustered | 10 | VC_949_0 | 10 | 0.5977 | 1 | 0.5977 | 0 | 0 | 0 | 1 | Ref |
| 6015 | MG820632 | Unassigned | Unassigned | Unassigned | 765_0 | Clustered | 132 | VC_765_0 | 132 | 1 | 1 | 1 | 0 | 0 | 0 | 1 | Ref |
| 6016 | MG820633 | Unassigned | Unassigned | Unassigned | 765_0 | Clustered | 132 | VC_765_0 | 132 | 1 | 1 | 1 | 0 | 0 | 0 | 1 | Ref |
| 6017 | MG820634 | Unassigned | Unassigned | Unassigned | 765_0 | Clustered | 132 | VC_765_0 | 132 | 1 | 1 | 1 | 0 | 0 | 0 | 1 | Ref |
| 6018 | MG820635 | Unassigned | Unassigned | Unassigned | 765_0 | Clustered | 132 | VC_765_0 | 132 | 1 | 1 | 1 | 0 | 0 | 0 | 1 | Ref |
| 6019 | MG820636 | Unassigned | Unassigned | Unassigned | 765_0 | Clustered | 132 | VC_765_0 | 132 | 1 | 1 | 1 | 0 | 0 | 0 | 1 | Ref |
| 6020 | MG820637 | Unassigned | Unassigned | Unassigned | 765_0 | Clustered | 132 | VC_765_0 | 132 | 1 | 1 | 1 | 0 | 0 | 0 | 1 | Ref |
| 6021 | MG820638 | Unassigned | Unassigned | Unassigned | 765_0 | Clustered | 132 | VC_765_0 | 132 | 1 | 1 | 1 | 0 | 0 | 0 | 1 | Ref |
| 6022 | MG820639 | Unassigned | Unassigned | Unassigned | 765_0 | Clustered | 132 | VC_765_0 | 132 | 1 | 1 | 1 | 0 | 0 | 0 | 1 | Ref |
| 6023 | MG820640 | Unassigned | Unassigned | Unassigned | 765_0 | Clustered | 132 | VC_765_0 | 132 | 1 | 1 | 1 | 0 | 0 | 0 | 1 | Ref |
| 6024 | MG820641 | Unassigned | Unassigned | Unassigned | 765_0 | Clustered | 132 | VC_765_0 | 132 | 1 | 1 | 1 | 0 | 0 | 0 | 1 | Ref |
| 6025 | MG820642 | Unassigned | Unassigned | Unassigned | 765_0 | Clustered | 132 | VC_765_0 | 132 | 1 | 1 | 1 | 0 | 0 | 0 | 1 | Ref |
| 6026 | MG820643 | Unassigned | Unassigned | Unassigned | 765_0 | Clustered | 132 | VC_765_0 | 132 | 1 | 1 | 1 | 0 | 0 | 0 | 1 | Ref |
| 6027 | MG820644 | Unassigned | Unassigned | Unassigned | 765_0 | Clustered | 132 | VC_765_0 | 132 | 1 | 1 | 1 | 0 | 0 | 0 | 1 | Ref |
| 6028 | MG820645 | Unassigned | Unassigned | Unassigned | 765_0 | Clustered | 132 | VC_765_0 | 132 | 1 | 1 | 1 | 0 | 0 | 0 | 1 | Ref |
| 6029 | MG832642 | Unassigned | Unassigned | Unassigned | 392_0 | Clustered | 126 | VC_392_0 | 126 | 0.9621 | 1 | 0.9621 | 0 | 0 | 0 | 1 | Ref |
| 6030 | MG833025 | Unassigned | Unassigned | Unassigned | 392_0 | Clustered | 126 | VC_392_0 | 126 | 0.9621 | 1 | 0.9621 | 0 | 0 | 0 | 1 | Ref |
| 6031 | MG835568 | Unassigned | Unassigned | Unassigned | 823_0 | Clustered | 124 | VC_823_0 | 124 | 0.9492 | 1 | 0.9492 | 0 | 0 | 0 | 1 | Ref |
| 6032 | MG839014 | Unassigned | Unassigned | Unassigned | 509_0 | Clustered | 37 | VC_509_0 | 37 | 0.9689 | 1 | 0.9689 | 0 | 0 | 0 | 1 | Ref |
| 6033 | MG839015 | Unassigned | Unassigned | Unassigned | 1240_0 | Clustered | 82 | VC_1240_0 | 82 | 0.7945 | 1 | 0.7945 | 0 | 0 | 0 | 1 | Ref |
| 6034 | MG839016 | Unassigned | Unassigned | Unassigned | 1240_0 | Clustered | 82 | VC_1240_0 | 82 | 0.7945 | 1 | 0.7945 | 0 | 0 | 0 | 1 | Ref |
| 6035 | MG839017 | Unassigned | Unassigned | Unassigned | 1240_0 | Clustered | 82 | VC_1240_0 | 82 | 0.7945 | 1 | 0.7945 | 0 | 0 | 0 | 1 | Ref |
| 6036 | MG839018 | Unassigned | Unassigned | Unassigned | 1240_0 | Clustered | 82 | VC_1240_0 | 82 | 0.7945 | 1 | 0.7945 | 0 | 0 | 0 | 1 | Ref |
| 6037 | MG839020 | Unassigned | Unassigned | Unassigned | 1240_0 | Clustered | 82 | VC_1240_0 | 82 | 0.7945 | 1 | 0.7945 | 0 | 0 | 0 | 1 | Ref |
| 6038 | MG839021 | Unassigned | Unassigned | Unassigned | 1240_0 | Clustered | 82 | VC_1240_0 | 82 | 0.7945 | 1 | 0.7945 | 0 | 0 | 0 | 1 | Ref |
| 6039 | MG839022 | Unassigned | Unassigned | Unassigned | 1240_0 | Clustered | 82 | VC_1240_0 | 82 | 0.7945 | 1 | 0.7945 | 0 | 0 | 0 | 1 | Ref |
| 6040 | MG839023 | Unassigned | Unassigned | Unassigned | 1240_0 | Clustered | 82 | VC_1240_0 | 82 | 0.7945 | 1 | 0.7945 | 0 | 0 | 0 | 1 | Ref |
| 6041 | MG839026 | Unassigned | Unassigned | Unassigned | 1240_0 | Clustered | 82 | VC_1240_0 | 82 | 0.7945 | 1 | 0.7945 | 0 | 0 | 0 | 1 | Ref |
| 6042 | MG839027 | Unassigned | Unassigned | Unassigned | 1163_0 | Clustered | 28 | VC_1163_0 | 28 | 0.4923 | 1 | 0.4923 | 0 | 0 | 0 | 1 | Ref |
| 6043 | MG839028 | Unassigned | Unassigned | Unassigned | 1240_0 | Clustered | 82 | VC_1240_0 | 82 | 0.7945 | 1 | 0.7945 | 0 | 0 | 0 | 1 | Ref |
| 6044 | MG839029 | Unassigned | Unassigned | Unassigned | 1240_0 | Clustered | 82 | VC_1240_0 | 82 | 0.7945 | 1 | 0.7945 | 0 | 0 | 0 | 1 | Ref |
| 6045 | MG845684 | Unassigned | Unassigned | Unassigned | 472_0 | Clustered | 47 | VC_472_0 | 47 | 0.9025 | 1 | 0.9025 | 0 | 0 | 0 | 1 | Ref |
| 6046 | MG845865 | Unassigned | Unassigned | Unassigned | 451_0 | Clustered | 80 | VC_451_0 | 80 | 0.9991 | 1 | 0.9991 | 0 | 0 | 0 | 1 | Ref |
| 6047 | MG852086 | Unassigned | Unassigned | Unassigned | 638_0 | Clustered | 45 | VC_638_0 | 45 | 0.7741 | 1 | 0.7741 | 0 | 0 | 0 | 1 | Ref |
| 6048 | MG867727 | Unassigned | Unassigned | Unassigned | 283_138 | Clustered/Singleton | 1 | VC_283_138 | 1 | 0 | 1 | 0 | 0 | 0 | 0 | 1 | Ref |
| 6049 | MG872831 | Unassigned | Unassigned | Unassigned | 551_0 | Clustered | 16 | VC_551_0 | 16 | 0.0915 | 1 | 0.0915 | 0 | 0 | 0 | 0.6667 | Ref |
| 6050 | MG872832 | Unassigned | Unassigned | Unassigned | 551_0 | Clustered | 16 | VC_551_0 | 16 | 0.0915 | 1 | 0.0915 | 0 | 0 | 0 | 0.6667 | Ref |
| 6051 | MG872833 | Unassigned | Unassigned | Unassigned | 325_5 | Clustered | 11 | VC_325_5 | 11 | 0.047 | 1 | 0.047 | 0 | 0 | 0 | 0.7636 | Ref |
| 6052 | MG872834 | Unassigned | Unassigned | Unassigned | 685_0 | Clustered | 16 | VC_685_0 | 16 | 0.2972 | 1 | 0.2972 | 0 | 0 | 0 | 0.9167 | Ref |
| 6053 | MG872835 | Unassigned | Unassigned | Unassigned | 225_0 | Clustered | 32 | VC_225_0 | 32 | 0.848 | 1 | 0.848 | 0 | 0 | 0 | 0.996 | Ref |
| 6054 | MG872836 | Unassigned | Unassigned | Unassigned | 325_50 | Clustered | 4 | VC_325_50 | 4 | 0.0182 | 0.99998785 | 0.0182 | 0 | 0 | 0 | 1 | Ref |
| 6055 | MG872837 | Unassigned | Unassigned | Unassigned | 551_6 | Clustered | 22 | VC_551_6 | 22 | 0.1214 | 1 | 0.1214 | 0 | 0 | 0 | 0.8095 | Ref |
| 6056 | MG872838 | Unassigned | Unassigned | Unassigned | 556_0 | Clustered | 21 | VC_556_0 | 21 | 0.9912 | 1 | 0.9912 | 0 | 0 | 0 | 1 | Ref |
| 6057 | MG872839 | Unassigned | Unassigned | Unassigned | 558_14 | Clustered | 8 | VC_558_14 | 8 | 0.0228 | 0.99945246 | 0.0228 | 0 | 0 | 0 | 0.8214 | Ref |
| 6058 | MG872840 | Unassigned | Unassigned | Unassigned | 225_0 | Clustered | 32 | VC_225_0 | 32 | 0.848 | 1 | 0.848 | 0 | 0 | 0 | 0.996 | Ref |
| 6059 | MG872841 | Unassigned | Unassigned | Unassigned | 520_45 | Clustered | 7 | VC_520_45 | 7 | 0.0635 | 1 | 0.0635 | 0 | 0 | 0 | 0.8095 | Ref |
| 6060 | MG872842 | Unassigned | Unassigned | Unassigned | 225_0 | Clustered | 32 | VC_225_0 | 32 | 0.848 | 1 | 0.848 | 0 | 0 | 0 | 0.996 | Ref |
| 6061 | MG872843 | Unassigned | Unassigned | Unassigned | 551_0 | Clustered | 16 | VC_551_0 | 16 | 0.0915 | 1 | 0.0915 | 0 | 0 | 0 | 0.6667 | Ref |
| 6062 | MG873442 | Unassigned | Unassigned | Unassigned | 812_0 | Clustered | 3 | VC_812_0 | 3 | 0.1174 | 0.99842379 | 0.1172 | 0 | 0 | 0 | 1 | Ref |
| 6063 | MG878892 | Unassigned | Unassigned | Unassigned | 878_0 | Clustered | 32 | VC_878_0 | 32 | 0.9965 | 1 | 0.9965 | 0 | 0 | 0 | 1 | Ref |
| 6064 | MG883709 | Unassigned | Unassigned | Unassigned | 1243_0 | Clustered | 52 | VC_1243_0 | 52 | 0.9126 | 0.92708311 | 0.8461 | 0 | 0 | 0 | 1 | Ref |
| 6065 | MG883710 | Unassigned | Unassigned | Unassigned | 1245_0 | Clustered | 2 | VC_1245_0 | 2 | 1 | 1 | 1 | 0 | 0 | 0 | 1 | Ref |
| 6066 | MG883712 | Unassigned | Unassigned | Unassigned | 1197_0 | Clustered | 15 | VC_1197_0 | 15 | 0.8991 | 0.76608394 | 0.6888 | 0 | 0 | 0 | 1 | Ref |
| 6067 | MG883713 | Unassigned | Unassigned | Unassigned | 1246_0 | Clustered | 2 | VC_1246_0 | 2 | 1 | 1 | 1 | 0 | 0 | 0 | 1 | Ref |
| 6068 | MG883714 | Unassigned | Unassigned | Unassigned | 1144_0 | Clustered | 7 | VC_1144_0 | 7 | 0.7076 | 1 | 0.7076 | 0 | 0 | 0 | 1 | Ref |
| 6069 | MG883715 | Unassigned | Unassigned | Unassigned | 672_0 | Clustered | 12 | VC_672_0 | 12 | 1 | 1 | 1 | 0 | 0 | 0 | 1 | Ref |
| 6070 | MG883717 | Unassigned | Unassigned | Unassigned | 1104_0 | Clustered | 154 | VC_1104_0 | 154 | 0.8484 | 1 | 0.8484 | 0 | 0 | 0 | 1 | Ref |
| 6071 | MG883718 | Unassigned | Unassigned | Unassigned | 1108_0 | Clustered | 4 | VC_1108_0 | 4 | 0.0485 | 0.99999021 | 0.0485 | 0 | 0 | 0 | 1 | Ref |
| 6072 | MG883719 | Unassigned | Unassigned | Unassigned | 1116_0 | Clustered | 14 | VC_1116_0 | 14 | 0.7927 | 1 | 0.7927 | 0 | 0 | 0 | 1 | Ref |
| 6073 | MG883720 | Unassigned | Unassigned | Unassigned | 1247_0 | Clustered | 7 | VC_1247_0 | 7 | 0.9086 | 0.99998959 | 0.9086 | 0 | 0 | 0 | 1 | Ref |
| 6074 | MG883722 | Unassigned | Unassigned | Unassigned | 1125_0 | Clustered | 20 | VC_1125_0 | 20 | 0.5599 | 1 | 0.5599 | 0 | 0 | 0 | 1 | Ref |
| 6075 | MG883723 | Unassigned | Unassigned | Unassigned | 198_0 | Clustered | 28 | VC_198_0 | 28 | 0.2429 | 0.99971175 | 0.2429 | 0 | 0 | 0 | 1 | Ref |
| 6076 | MG883724 | Unassigned | Unassigned | Unassigned | 1250_0 | Clustered | 5 | VC_1250_0 | 5 | 0.6384 | 0.99918945 | 0.6379 | 0 | 0 | 0 | 1 | Ref |
| 6077 | MG883725 | Unassigned | Unassigned | Unassigned | 672_0 | Clustered | 12 | VC_672_0 | 12 | 1 | 1 | 1 | 0 | 0 | 0 | 1 | Ref |
| 6078 | MG883726 | Unassigned | Unassigned | Unassigned | 1077_0 | Clustered | 48 | VC_1077_0 | 48 | 0.8525 | 1 | 0.8525 | 0 | 0 | 0 | 1 | Ref |
| 6079 | MG893203 | Unassigned | Unassigned | Unassigned | 882_0 | Clustered | 20 | VC_882_0 | 20 | 0.9917 | 1 | 0.9917 | 0 | 0 | 0 | 1 | Ref |
| 6080 | MG894052 | Unassigned | Unassigned | Unassigned | 938_0 | Clustered | 99 | VC_938_0 | 99 | 0.9119 | 1 | 0.9119 | 0 | 0 | 0 | 0.9928 | Ref |
| 6081 | MG897799 | Unassigned | Unassigned | Unassigned | 79_0 | Clustered | 64 | VC_79_0 | 64 | 0.4185 | 1 | 0.4185 | 0 | 0 | 0 | 0.7649 | Ref |
| 6082 | MG897800 | Unassigned | Unassigned | Unassigned | 749_0 | Clustered | 40 | VC_749_0 | 40 | 0.7517 | 1 | 0.7517 | 0 | 0 | 0 | 1 | Ref |
| 6083 | MG913376 | Unassigned | Unassigned | Unassigned | 640_0 | Clustered | 7 | VC_640_0 | 7 | 0.9025 | 1 | 0.9025 | 0 | 0 | 0 | 1 | Ref |
| 6084 | MG920059 | Unassigned | Unassigned | Unassigned | 509_0 | Clustered | 37 | VC_509_0 | 37 | 0.9689 | 1 | 0.9689 | 0 | 0 | 0 | 1 | Ref |
| 6085 | MG920060 | Unassigned | Unassigned | Unassigned | 325_23 | Clustered | 17 | VC_325_23 | 17 | 0.0716 | 1 | 0.0716 | 0 | 0 | 0 | 0.6397 | Ref |
| 6086 | MG920061 | Unassigned | Unassigned | Unassigned | 1240_0 | Clustered | 82 | VC_1240_0 | 82 | 0.7945 | 1 | 0.7945 | 0 | 0 | 0 | 1 | Ref |
| 6087 | MG922974 | Unassigned | Unassigned | Unassigned | 779_0 | Clustered | 37 | VC_779_0 | 37 | 0.5682 | 1 | 0.5682 | 0 | 0 | 0 | 1 | Ref |
| 6088 | MG925338 | Unassigned | Unassigned | Unassigned | 509_0 | Clustered | 37 | VC_509_0 | 37 | 0.9689 | 1 | 0.9689 | 0 | 0 | 0 | 1 | Ref |
| 6089 | MG925339 | Unassigned | Unassigned | Unassigned | 371_3 | Clustered | 22 | VC_371_3 | 22 | 0.0787 | 1 | 0.0787 | 0 | 0 | 0 | 0.9048 | Ref |
| 6090 | MG925340 | Unassigned | Unassigned | Unassigned | 325_61 | Clustered | 8 | VC_325_61 | 8 | 0.0395 | 1 | 0.0395 | 0 | 0 | 0 | 0.7857 | Ref |
| 6091 | MG925341 | Unassigned | Unassigned | Unassigned | 514_0 | Clustered | 75 | VC_514_0 | 75 | 0.9402 | 1 | 0.9402 | 0 | 0 | 0 | 1 | Ref |
| 6092 | MG925342 | Unassigned | Unassigned | Unassigned | 325_61 | Clustered | 8 | VC_325_61 | 8 | 0.0395 | 1 | 0.0395 | 0 | 0 | 0 | 0.7857 | Ref |
| 6093 | MG925343 | Unassigned | Unassigned | Unassigned | 1163_0 | Clustered | 28 | VC_1163_0 | 28 | 0.4923 | 1 | 0.4923 | 0 | 0 | 0 | 1 | Ref |
| 6094 | MG925344 | Unassigned | Unassigned | Unassigned | 325_5 | Clustered | 11 | VC_325_5 | 11 | 0.047 | 1 | 0.047 | 0 | 0 | 0 | 0.7636 | Ref |
| 6095 | MG925345 | Unassigned | Unassigned | Unassigned | 1163_0 | Clustered | 28 | VC_1163_0 | 28 | 0.4923 | 1 | 0.4923 | 0 | 0 | 0 | 1 | Ref |
| 6096 | MG925346 | Unassigned | Unassigned | Unassigned | 371_15 | Clustered | 44 | VC_371_15 | 44 | 0.1391 | 1 | 0.1391 | 0 | 0 | 0 | 0.7474 | Ref |
| 6097 | MG925347 | Unassigned | Unassigned | Unassigned | 1163_0 | Clustered | 28 | VC_1163_0 | 28 | 0.4923 | 1 | 0.4923 | 0 | 0 | 0 | 1 | Ref |
| 6098 | MG925348 | Unassigned | Unassigned | Unassigned | 371_16 | Clustered | 42 | VC_371_16 | 42 | 0.1341 | 1 | 0.1341 | 0 | 0 | 0 | 0.7282 | Ref |
| 6099 | MG925349 | Unassigned | Unassigned | Unassigned | 1046_0 | Clustered | 7 | VC_1046_0 | 7 | 0.9413 | 1 | 0.9413 | 0 | 0 | 0 | 1 | Ref |
| 6100 | MG925350 | Unassigned | Unassigned | Unassigned | 371_15 | Clustered | 44 | VC_371_15 | 44 | 0.1391 | 1 | 0.1391 | 0 | 0 | 0 | 0.7474 | Ref |
| 6101 | MG925351 | Unassigned | Unassigned | Unassigned | 325_23 | Clustered | 17 | VC_325_23 | 17 | 0.0716 | 1 | 0.0716 | 0 | 0 | 0 | 0.6397 | Ref |
| 6102 | MG925352 | Unassigned | Unassigned | Unassigned | 1240_0 | Clustered | 82 | VC_1240_0 | 82 | 0.7945 | 1 | 0.7945 | 0 | 0 | 0 | 1 | Ref |
| 6103 | MG925353 | Unassigned | Unassigned | Unassigned | 509_0 | Clustered | 37 | VC_509_0 | 37 | 0.9689 | 1 | 0.9689 | 0 | 0 | 0 | 1 | Ref |
| 6104 | MG925354 | Unassigned | Unassigned | Unassigned | 520_97 | Clustered/Singleton | 1 | VC_520_97 | 1 | 0 | 1 | 0 | 0 | 0 | 0 | 1 | Ref |
| 6105 | MG925355 | Unassigned | Unassigned | Unassigned | 514_0 | Clustered | 75 | VC_514_0 | 75 | 0.9402 | 1 | 0.9402 | 0 | 0 | 0 | 1 | Ref |
| 6106 | MG925356 | Unassigned | Unassigned | Unassigned | 371_2 | Clustered | 8 | VC_371_2 | 8 | 0.0232 | 1 | 0.0232 | 0 | 0 | 0 | 0.8571 | Ref |
| 6107 | MG941013 | Unassigned | Unassigned | Unassigned | 853_0 | Clustered | 2 | VC_853_0 | 2 | 0.27 | 0.72985431 | 0.1971 | 0 | 0 | 0 | 1 | Ref |
| 6108 | MG944216 | Unassigned | Unassigned | Unassigned | 1163_0 | Clustered | 28 | VC_1163_0 | 28 | 0.4923 | 1 | 0.4923 | 0 | 0 | 0 | 1 | Ref |
| 6109 | MG944217 | Unassigned | Unassigned | Unassigned | 517_1 | Clustered | 108 | VC_517_1 | 108 | 0.8483 | 1 | 0.8483 | 0 | 0 | 0 | 0.9955 | Ref |
| 6110 | MG944218 | Unassigned | Unassigned | Unassigned | 1163_0 | Clustered | 28 | VC_1163_0 | 28 | 0.4923 | 1 | 0.4923 | 0 | 0 | 0 | 1 | Ref |
| 6111 | MG944219 | Unassigned | Unassigned | Unassigned | 1240_0 | Clustered | 82 | VC_1240_0 | 82 | 0.7945 | 1 | 0.7945 | 0 | 0 | 0 | 1 | Ref |
| 6112 | MG944220 | Unassigned | Unassigned | Unassigned | 325_26 | Clustered/Singleton | 1 | VC_325_26 | 1 | 0 | 1 | 0 | 0 | 0 | 0 | 1 | Ref |
| 6113 | MG944221 | Unassigned | Unassigned | Unassigned | 325_7 | Clustered | 3 | VC_325_7 | 3 | 0.0085 | 0.99739782 | 0.0085 | 0 | 0 | 0 | 0.6667 | Ref |
| 6114 | MG944222 | Unassigned | Unassigned | Unassigned | 1240_0 | Clustered | 82 | VC_1240_0 | 82 | 0.7945 | 1 | 0.7945 | 0 | 0 | 0 | 1 | Ref |
| 6115 | MG944223 | Unassigned | Unassigned | Unassigned | 371_13 | Clustered | 33 | VC_371_13 | 33 | 0.179 | 1 | 0.179 | 0 | 0 | 0 | 0.8769 | Ref |
| 6116 | MG944224 | Unassigned | Unassigned | Unassigned | 517_1 | Clustered | 108 | VC_517_1 | 108 | 0.8483 | 1 | 0.8483 | 0 | 0 | 0 | 0.9955 | Ref |
| 6117 | MG944225 | Unassigned | Unassigned | Unassigned | 371_15 | Clustered | 44 | VC_371_15 | 44 | 0.1391 | 1 | 0.1391 | 0 | 0 | 0 | 0.7474 | Ref |
| 6118 | MG944226 | Unassigned | Unassigned | Unassigned | 1163_0 | Clustered | 28 | VC_1163_0 | 28 | 0.4923 | 1 | 0.4923 | 0 | 0 | 0 | 1 | Ref |
| 6119 | MG944227 | Unassigned | Unassigned | Unassigned | 364_0 | Clustered | 16 | VC_364_0 | 16 | 0.6491 | 1 | 0.6491 | 0 | 0 | 0 | 1 | Ref |
| 6120 | MG944228 | Unassigned | Unassigned | Unassigned | 364_0 | Clustered | 16 | VC_364_0 | 16 | 0.6491 | 1 | 0.6491 | 0 | 0 | 0 | 1 | Ref |
| 6121 | MG944229 | Unassigned | Unassigned | Unassigned | 364_0 | Clustered | 16 | VC_364_0 | 16 | 0.6491 | 1 | 0.6491 | 0 | 0 | 0 | 1 | Ref |
| 6122 | MG944230 | Unassigned | Unassigned | Unassigned | 364_0 | Clustered | 16 | VC_364_0 | 16 | 0.6491 | 1 | 0.6491 | 0 | 0 | 0 | 1 | Ref |
| 6123 | MG944231 | Unassigned | Unassigned | Unassigned | 364_0 | Clustered | 16 | VC_364_0 | 16 | 0.6491 | 1 | 0.6491 | 0 | 0 | 0 | 1 | Ref |
| 6124 | MG944232 | Unassigned | Unassigned | Unassigned | 364_0 | Clustered | 16 | VC_364_0 | 16 | 0.6491 | 1 | 0.6491 | 0 | 0 | 0 | 1 | Ref |
| 6125 | MG944233 | Unassigned | Unassigned | Unassigned | 364_0 | Clustered | 16 | VC_364_0 | 16 | 0.6491 | 1 | 0.6491 | 0 | 0 | 0 | 1 | Ref |
| 6126 | MG944234 | Unassigned | Unassigned | Unassigned | 364_0 | Clustered | 16 | VC_364_0 | 16 | 0.6491 | 1 | 0.6491 | 0 | 0 | 0 | 1 | Ref |
| 6127 | MG944235 | Unassigned | Unassigned | Unassigned | 364_0 | Clustered | 16 | VC_364_0 | 16 | 0.6491 | 1 | 0.6491 | 0 | 0 | 0 | 1 | Ref |
| 6128 | MG944236 | Unassigned | Unassigned | Unassigned | 364_0 | Clustered | 16 | VC_364_0 | 16 | 0.6491 | 1 | 0.6491 | 0 | 0 | 0 | 1 | Ref |
| 6129 | MG945128 | Unassigned | Unassigned | Unassigned | 1251_0 | Clustered | 15 | VC_1251_0 | 15 | 0.9082 | 1 | 0.9082 | 0 | 0 | 0 | 1 | Ref |
| 6130 | MG945129 | Unassigned | Unassigned | Unassigned | 1077_0 | Clustered | 48 | VC_1077_0 | 48 | 0.8525 | 1 | 0.8525 | 0 | 0 | 0 | 1 | Ref |
| 6131 | MG945130 | Unassigned | Unassigned | Unassigned | 1233_0 | Clustered | 16 | VC_1233_0 | 16 | 0.8971 | 1 | 0.8971 | 0 | 0 | 0 | 1 | Ref |
| 6132 | MG945131 | Unassigned | Unassigned | Unassigned | 1114_0 | Clustered | 2 | VC_1114_0 | 2 | 0.0522 | 0.94760452 | 0.0495 | 0 | 0 | 0 | 1 | Ref |
| 6133 | MG945135 | Unassigned | Unassigned | Unassigned | 1124_0 | Clustered | 10 | VC_1124_0 | 10 | 0.6268 | 1 | 0.6268 | 0 | 0 | 0 | 1 | Ref |
| 6134 | MG945136 | Unassigned | Unassigned | Unassigned | 1124_0 | Clustered | 10 | VC_1124_0 | 10 | 0.6268 | 1 | 0.6268 | 0 | 0 | 0 | 1 | Ref |
| 6135 | MG945138 | Unassigned | Unassigned | Unassigned | 1126_0 | Clustered | 8 | VC_1126_0 | 8 | 0.2864 | 1 | 0.2864 | 0 | 0 | 0 | 1 | Ref |
| 6136 | MG945139 | Unassigned | Unassigned | Unassigned | 1116_0 | Clustered | 14 | VC_1116_0 | 14 | 0.7927 | 1 | 0.7927 | 0 | 0 | 0 | 1 | Ref |
| 6137 | MG945140 | Unassigned | Unassigned | Unassigned | 1257_0 | Clustered | 6 | VC_1257_0 | 6 | 0.6517 | 0.99999969 | 0.6517 | 0 | 0 | 0 | 1 | Ref |
| 6138 | MG945141 | Unassigned | Unassigned | Unassigned | 1211_0 | Clustered | 25 | VC_1211_0 | 25 | 1 | 1 | 1 | 0 | 0 | 0 | 1 | Ref |
| 6139 | MG945142 | Unassigned | Unassigned | Unassigned | 1136_0 | Clustered | 2 | VC_1136_0 | 2 | 0.58 | 0.88261817 | 0.5119 | 0 | 0 | 0 | 1 | Ref |
| 6140 | MG945153 | Unassigned | Unassigned | Unassigned | 1104_0 | Clustered | 154 | VC_1104_0 | 154 | 0.8484 | 1 | 0.8484 | 0 | 0 | 0 | 1 | Ref |
| 6141 | MG945154 | Unassigned | Unassigned | Unassigned | 1119_0 | Clustered | 20 | VC_1119_0 | 20 | 0.6751 | 0.99997892 | 0.6751 | 0 | 0 | 0 | 1 | Ref |
| 6142 | MG945157 | Unassigned | Unassigned | Unassigned | 1124_0 | Clustered | 10 | VC_1124_0 | 10 | 0.6268 | 1 | 0.6268 | 0 | 0 | 0 | 1 | Ref |
| 6143 | MG945158 | Unassigned | Unassigned | Unassigned | 1104_0 | Clustered | 154 | VC_1104_0 | 154 | 0.8484 | 1 | 0.8484 | 0 | 0 | 0 | 1 | Ref |
| 6144 | MG945159 | Unassigned | Unassigned | Unassigned | 1251_0 | Clustered | 15 | VC_1251_0 | 15 | 0.9082 | 1 | 0.9082 | 0 | 0 | 0 | 1 | Ref |
| 6145 | MG945160 | Unassigned | Unassigned | Unassigned | 1191_0 | Clustered | 3 | VC_1191_0 | 3 | 0.1408 | 0.98976974 | 0.1394 | 0 | 0 | 0 | 1 | Ref |
| 6146 | MG945161 | Unassigned | Unassigned | Unassigned | 1000_0 | Clustered | 21 | VC_1000_0 | 21 | 0.8437 | 0.99999998 | 0.8437 | 0 | 0 | 0 | 1 | Ref |
| 6147 | MG945162 | Unassigned | Unassigned | Unassigned | 1127_0 | Clustered | 19 | VC_1127_0 | 19 | 0.7611 | 1 | 0.7611 | 0 | 0 | 0 | 1 | Ref |
| 6148 | MG945163 | Unassigned | Unassigned | Unassigned | 1104_0 | Clustered | 154 | VC_1104_0 | 154 | 0.8484 | 1 | 0.8484 | 0 | 0 | 0 | 1 | Ref |
| 6149 | MG945164 | Unassigned | Unassigned | Unassigned | 398_0 | Clustered | 48 | VC_398_0 | 48 | 0.4813 | 1 | 0.4813 | 0 | 0 | 0 | 1 | Ref |
| 6150 | MG945165 | Unassigned | Unassigned | Unassigned | 1260_0 | Clustered | 2 | VC_1260_0 | 2 | 1 | 1 | 1 | 0 | 0 | 0 | 1 | Ref |
| 6151 | MG945166 | Unassigned | Unassigned | Unassigned | 1259_0 | Clustered | 8 | VC_1259_0 | 8 | 0.8225 | 1 | 0.8225 | 0 | 0 | 0 | 1 | Ref |
| 6152 | MG945177 | Unassigned | Unassigned | Unassigned | 1261_0 | Clustered | 5 | VC_1261_0 | 5 | 0.765 | 0.99993003 | 0.7649 | 0 | 0 | 0 | 1 | Ref |
| 6153 | MG945178 | Unassigned | Unassigned | Unassigned | 1062_0 | Clustered | 45 | VC_1062_0 | 45 | 0.9734 | 0.50949847 | 0.4959 | 0 | 0 | 0 | 1 | Ref |
| 6154 | MG945179 | Unassigned | Unassigned | Unassigned | 1148_0 | Clustered | 16 | VC_1148_0 | 16 | 0.8821 | 1 | 0.8821 | 0 | 0 | 0 | 1 | Ref |

| | | | | | | | | | | | | | | | | | |
|---|---|---|---|---|---|---|---|---|---|---|---|---|---|---|---|---|---|
| 6155 | MG945181 | Unassigned | Unassigned | Unassigned | 1110_0 | Clustered | 8 | VC_1110_0 | 8 | 0.6915 | 1 | 0.6915 | 0 | 0 | 0 | 1 | Ref |
| 6156 | MG945183 | Unassigned | Unassigned | Unassigned | 1263_0 | Clustered | 2 | VC_1263_0 | 2 | 1 | 1 | 1 | 0 | 0 | 0 | 1 | Ref |
| 6157 | MG945185 | Unassigned | Unassigned | Unassigned | 1191_0 | Clustered | 3 | VC_1191_0 | 3 | 0.1408 | 0.98976974 | 0.1394 | 0 | 0 | 0 | 1 | Ref |
| 6158 | MG945186 | Unassigned | Unassigned | Unassigned | 1122_0 | Clustered | 10 | VC_1122_0 | 10 | 0.9643 | 0.99999697 | 0.9643 | 0 | 0 | 0 | 1 | Ref |
| 6159 | MG945188 | Unassigned | Unassigned | Unassigned | 1251_0 | Clustered | 15 | VC_1251_0 | 15 | 0.9082 | 1 | 0.9082 | 0 | 0 | 0 | 1 | Ref |
| 6160 | MG945189 | Unassigned | Unassigned | Unassigned | 1266_0 | Clustered | 5 | VC_1266_0 | 5 | 1 | 1 | 1 | 0 | 0 | 0 | 1 | Ref |
| 6161 | MG945190 | Unassigned | Unassigned | Unassigned | 1104_0 | Clustered | 154 | VC_1104_0 | 154 | 0.8484 | 1 | 0.8484 | 0 | 0 | 0 | 1 | Ref |
| 6162 | MG945202 | Unassigned | Unassigned | Unassigned | 995_0 | Clustered | 69 | VC_995_0 | 69 | 0.8547 | 1 | 0.8547 | 0 | 0 | 0 | 1 | Ref |
| 6163 | MG945203 | Unassigned | Unassigned | Unassigned | 1104_0 | Clustered | 154 | VC_1104_0 | 154 | 0.8484 | 1 | 0.8484 | 0 | 0 | 0 | 1 | Ref |
| 6164 | MG945204 | Unassigned | Unassigned | Unassigned | 1152_0 | Clustered | 4 | VC_1152_0 | 4 | 0.7206 | 0.99971165 | 0.7204 | 0 | 0 | 0 | 1 | Ref |
| 6165 | MG945205 | Unassigned | Unassigned | Unassigned | 1104_0 | Clustered | 154 | VC_1104_0 | 154 | 0.8484 | 1 | 0.8484 | 0 | 0 | 0 | 1 | Ref |
| 6166 | MG945206 | Unassigned | Unassigned | Unassigned | 1077_0 | Clustered | 48 | VC_1077_0 | 48 | 0.8525 | 1 | 0.8525 | 0 | 0 | 0 | 1 | Ref |
| 6167 | MG945207 | Unassigned | Unassigned | Unassigned | 1104_0 | Clustered | 154 | VC_1104_0 | 154 | 0.8484 | 1 | 0.8484 | 0 | 0 | 0 | 1 | Ref |
| 6168 | MG945208 | Unassigned | Unassigned | Unassigned | 1259_0 | Clustered | 8 | VC_1259_0 | 8 | 0.8225 | 1 | 0.8225 | 0 | 0 | 0 | 1 | Ref |
| 6169 | MG945209 | Unassigned | Unassigned | Unassigned | 1267_0 | Clustered | 7 | VC_1267_0 | 7 | 1 | 1 | 1 | 0 | 0 | 0 | 1 | Ref |
| 6170 | MG945210 | Unassigned | Unassigned | Unassigned | 1194_0 | Clustered | 5 | VC_1194_0 | 5 | 0.5236 | 0.99999899 | 0.5236 | 0 | 0 | 0 | 1 | Ref |
| 6171 | MG945211 | Unassigned | Unassigned | Unassigned | 1110_0 | Clustered | 8 | VC_1110_0 | 8 | 0.6915 | 1 | 0.6915 | 0 | 0 | 0 | 1 | Ref |
| 6172 | MG945212 | Unassigned | Unassigned | Unassigned | 1268_0 | Clustered | 3 | VC_1268_0 | 3 | 0.0807 | 0.6862532 | 0.0554 | 0 | 0 | 0 | 1 | Ref |
| 6173 | MG945213 | Unassigned | Unassigned | Unassigned | 1104_0 | Clustered | 154 | VC_1104_0 | 154 | 0.8484 | 1 | 0.8484 | 0 | 0 | 0 | 1 | Ref |
| 6174 | MG945214 | Unassigned | Unassigned | Unassigned | 1270_0 | Clustered | 3 | VC_1270_0 | 3 | 0.6808 | 0.98688028 | 0.6719 | 0 | 0 | 0 | 1 | Ref |
| 6175 | MG945215 | Unassigned | Unassigned | Unassigned | 672_0 | Clustered | 12 | VC_672_0 | 12 | 1 | 1 | 1 | 0 | 0 | 0 | 1 | Ref |
| 6176 | MG945227 | Unassigned | Unassigned | Unassigned | 1104_0 | Clustered | 154 | VC_1104_0 | 154 | 0.8484 | 1 | 0.8484 | 0 | 0 | 0 | 1 | Ref |
| 6177 | MG945228 | Unassigned | Unassigned | Unassigned | 1151_0 | Clustered | 9 | VC_1151_0 | 9 | 0.8548 | 1 | 0.8548 | 0 | 0 | 0 | 1 | Ref |
| 6178 | MG945229 | Unassigned | Unassigned | Unassigned | 1251_0 | Clustered | 15 | VC_1251_0 | 15 | 0.9082 | 1 | 0.9082 | 0 | 0 | 0 | 1 | Ref |
| 6179 | MG945230 | Unassigned | Unassigned | Unassigned | 1271_0 | Clustered | 2 | VC_1271_0 | 2 | 1 | 1 | 1 | 0 | 0 | 0 | 1 | Ref |
| 6180 | MG945231 | Unassigned | Unassigned | Unassigned | 1128_0 | Clustered | 2 | VC_1128_0 | 2 | 0.0904 | 0.95143313 | 0.086 | 0 | 0 | 0 | 1 | Ref |
| 6181 | MG945232 | Unassigned | Unassigned | Unassigned | 1110_0 | Clustered | 8 | VC_1110_0 | 8 | 0.6915 | 1 | 0.6915 | 0 | 0 | 0 | 1 | Ref |
| 6182 | MG945233 | Unassigned | Unassigned | Unassigned | 1127_0 | Clustered | 19 | VC_1127_0 | 19 | 0.7611 | 1 | 0.7611 | 0 | 0 | 0 | 1 | Ref |
| 6183 | MG945234 | Unassigned | Unassigned | Unassigned | 1146_0 | Clustered | 3 | VC_1146_0 | 3 | 0.3641 | 0.99867077 | 0.3636 | 0 | 0 | 0 | 1 | Ref |
| 6184 | MG945235 | Unassigned | Unassigned | Unassigned | 1272_0 | Clustered | 5 | VC_1272_0 | 5 | 1 | 1 | 1 | 0 | 0 | 0 | 1 | Ref |
| 6185 | MG945236 | Unassigned | Unassigned | Unassigned | 1273_0 | Clustered | 6 | VC_1273_0 | 6 | 0.7385 | 0.9999804 | 0.7385 | 0 | 0 | 0 | 1 | Ref |
| 6186 | MG945237 | Unassigned | Unassigned | Unassigned | 1104_0 | Clustered | 154 | VC_1104_0 | 154 | 0.8484 | 1 | 0.8484 | 0 | 0 | 0 | 1 | Ref |
| 6187 | MG945238 | Unassigned | Unassigned | Unassigned | 1104_0 | Clustered | 154 | VC_1104_0 | 154 | 0.8484 | 1 | 0.8484 | 0 | 0 | 0 | 1 | Ref |
| 6188 | MG945241 | Unassigned | Unassigned | Unassigned | 1256_0 | Clustered | 4 | VC_1256_0 | 4 | 0.581 | 0.94751743 | 0.5505 | 0 | 0 | 0 | 1 | Ref |
| 6189 | MG945252 | Unassigned | Unassigned | Unassigned | 1275_0 | Clustered | 4 | VC_1275_0 | 4 | 1 | 1 | 1 | 0 | 0 | 0 | 1 | Ref |
| 6190 | MG945254 | Unassigned | Unassigned | Unassigned | 743_0 | Clustered | 15 | VC_743_0 | 15 | 1 | 1 | 1 | 0 | 0 | 0 | 1 | Ref |
| 6191 | MG945255 | Unassigned | Unassigned | Unassigned | 1251_0 | Clustered | 15 | VC_1251_0 | 15 | 0.9082 | 1 | 0.9082 | 0 | 0 | 0 | 1 | Ref |
| 6192 | MG945256 | Unassigned | Unassigned | Unassigned | 1233_0 | Clustered | 16 | VC_1233_0 | 16 | 0.8971 | 1 | 0.8971 | 0 | 0 | 0 | 1 | Ref |
| 6193 | MG945257 | Unassigned | Unassigned | Unassigned | 1257_0 | Clustered | 6 | VC_1257_0 | 6 | 0.6517 | 0.99999969 | 0.6517 | 0 | 0 | 0 | 1 | Ref |
| 6194 | MG945258 | Unassigned | Unassigned | Unassigned | 1279_0 | Clustered | 4 | VC_1279_0 | 4 | 0.6058 | 0.76972092 | 0.4663 | 0 | 0 | 0 | 1 | Ref |
| 6195 | MG945260 | Unassigned | Unassigned | Unassigned | 1243_0 | Clustered | 52 | VC_1243_0 | 52 | 0.9126 | 0.92708311 | 0.8461 | 0 | 0 | 0 | 1 | Ref |
| 6196 | MG945261 | Unassigned | Unassigned | Unassigned | 1264_0 | Clustered | 6 | VC_1264_0 | 6 | 0.7062 | 0.92055725 | 0.6501 | 0 | 0 | 0 | 1 | Ref |
| 6197 | MG945263 | Unassigned | Unassigned | Unassigned | 1267_0 | Clustered | 7 | VC_1267_0 | 7 | 1 | 1 | 1 | 0 | 0 | 0 | 1 | Ref |
| 6198 | MG945275 | Unassigned | Unassigned | Unassigned | 1266_0 | Clustered | 5 | VC_1266_0 | 5 | 1 | 1 | 1 | 0 | 0 | 0 | 1 | Ref |
| 6199 | MG945276 | Unassigned | Unassigned | Unassigned | 1256_0 | Clustered | 4 | VC_1256_0 | 4 | 0.581 | 0.94751743 | 0.5505 | 0 | 0 | 0 | 1 | Ref |
| 6200 | MG945278 | Unassigned | Unassigned | Unassigned | 1281_0 | Clustered | 7 | VC_1281_0 | 7 | 0.9212 | 0.99995294 | 0.9212 | 0 | 0 | 0 | 1 | Ref |
| 6201 | MG945279 | Unassigned | Unassigned | Unassigned | 1267_0 | Clustered | 7 | VC_1267_0 | 7 | 1 | 1 | 1 | 0 | 0 | 0 | 1 | Ref |
| 6202 | MG945280 | Unassigned | Unassigned | Unassigned | 1282_0 | Clustered | 3 | VC_1282_0 | 3 | 1 | 1 | 1 | 0 | 0 | 0 | 1 | Ref |
| 6203 | MG945282 | Unassigned | Unassigned | Unassigned | 1243_0 | Clustered | 52 | VC_1243_0 | 52 | 0.9126 | 0.92708311 | 0.8461 | 0 | 0 | 0 | 1 | Ref |
| 6204 | MG945284 | Unassigned | Unassigned | Unassigned | 1197_0 | Clustered | 15 | VC_1197_0 | 15 | 0.8991 | 0.76608394 | 0.6888 | 0 | 0 | 0 | 1 | Ref |
| 6205 | MG945285 | Unassigned | Unassigned | Unassigned | 1281_0 | Clustered | 7 | VC_1281_0 | 7 | 0.9212 | 0.99995294 | 0.9212 | 0 | 0 | 0 | 1 | Ref |
| 6206 | MG945286 | Unassigned | Unassigned | Unassigned | 1125_0 | Clustered | 20 | VC_1125_0 | 20 | 0.5599 | 1 | 0.5599 | 0 | 0 | 0 | 1 | Ref |
| 6207 | MG945309 | Unassigned | Unassigned | Unassigned | 1197_0 | Clustered | 15 | VC_1197_0 | 15 | 0.8991 | 0.76608394 | 0.6888 | 0 | 0 | 0 | 1 | Ref |
| 6208 | MG945314 | Unassigned | Unassigned | Unassigned | 1263_0 | Clustered | 2 | VC_1263_0 | 2 | 1 | 1 | 1 | 0 | 0 | 0 | 1 | Ref |
| 6209 | MG945316 | Unassigned | Unassigned | Unassigned | 1285_0 | Clustered | 25 | VC_1285_0 | 25 | 1 | 1 | 1 | 0 | 0 | 0 | 1 | Ref |
| 6210 | MG945318 | Unassigned | Unassigned | Unassigned | 1104_0 | Clustered | 154 | VC_1104_0 | 154 | 0.8484 | 1 | 0.8484 | 0 | 0 | 0 | 1 | Ref |
| 6211 | MG945319 | Unassigned | Unassigned | Unassigned | 1125_0 | Clustered | 20 | VC_1125_0 | 20 | 0.5599 | 1 | 0.5599 | 0 | 0 | 0 | 1 | Ref |
| 6212 | MG945320 | Unassigned | Unassigned | Unassigned | 1139_0 | Clustered | 4 | VC_1139_0 | 4 | 1 | 1 | 1 | 0 | 0 | 0 | 1 | Ref |
| 6213 | MG945323 | Unassigned | Unassigned | Unassigned | 1251_0 | Clustered | 15 | VC_1251_0 | 15 | 0.9082 | 1 | 0.9082 | 0 | 0 | 0 | 1 | Ref |
| 6214 | MG945324 | Unassigned | Unassigned | Unassigned | 198_0 | Clustered | 28 | VC_198_0 | 28 | 0.2429 | 0.99971175 | 0.2429 | 0 | 0 | 0 | 1 | Ref |
| 6215 | MG945325 | Unassigned | Unassigned | Unassigned | 1104_0 | Clustered | 154 | VC_1104_0 | 154 | 0.8484 | 1 | 0.8484 | 0 | 0 | 0 | 1 | Ref |
| 6216 | MG945327 | Unassigned | Unassigned | Unassigned | 1194_0 | Clustered | 5 | VC_1194_0 | 5 | 0.5236 | 0.99999899 | 0.5236 | 0 | 0 | 0 | 1 | Ref |
| 6217 | MG945328 | Unassigned | Unassigned | Unassigned | 1286_0 | Clustered/Singleton | 1 | VC_1286_0 | 1 | 0 | 1 | 0 | 0 | 0 | 0 | 1 | Ref |
| 6218 | MG945329 | Unassigned | Unassigned | Unassigned | 1259_0 | Clustered | 8 | VC_1259_0 | 8 | 0.8225 | 1 | 0.8225 | 0 | 0 | 0 | 1 | Ref |
| 6219 | MG945330 | Unassigned | Unassigned | Unassigned | 1104_0 | Clustered | 154 | VC_1104_0 | 154 | 0.8484 | 1 | 0.8484 | 0 | 0 | 0 | 1 | Ref |
| 6220 | MG945331 | Unassigned | Unassigned | Unassigned | 1287_0 | Clustered | 7 | VC_1287_0 | 7 | 1 | 1 | 1 | 0 | 0 | 0 | 1 | Ref |
| 6221 | MG945332 | Unassigned | Unassigned | Unassigned | 1251_0 | Clustered | 15 | VC_1251_0 | 15 | 0.9082 | 1 | 0.9082 | 0 | 0 | 0 | 1 | Ref |
| 6222 | MG945333 | Unassigned | Unassigned | Unassigned | 1104_0 | Clustered | 154 | VC_1104_0 | 154 | 0.8484 | 1 | 0.8484 | 0 | 0 | 0 | 1 | Ref |
| 6223 | MG945334 | Unassigned | Unassigned | Unassigned | 1127_0 | Clustered | 19 | VC_1127_0 | 19 | 0.7611 | 1 | 0.7611 | 0 | 0 | 0 | 1 | Ref |
| 6224 | MG945335 | Unassigned | Unassigned | Unassigned | 1288_0 | Clustered | 3 | VC_1288_0 | 3 | 1 | 1 | 1 | 0 | 0 | 0 | 1 | Ref |
| 6225 | MG945336 | Unassigned | Unassigned | Unassigned | 1289_0 | Clustered | 2 | VC_1289_0 | 2 | 0.4661 | 0.85557782 | 0.3987 | 0 | 0 | 0 | 1 | Ref |
| 6226 | MG945337 | Unassigned | Unassigned | Unassigned | 1290_0 | Clustered | 2 | VC_1290_0 | 2 | 1 | 1 | 1 | 0 | 0 | 0 | 1 | Ref |
| 6227 | MG945338 | Unassigned | Unassigned | Unassigned | 1125_0 | Clustered | 20 | VC_1125_0 | 20 | 0.5599 | 1 | 0.5599 | 0 | 0 | 0 | 1 | Ref |
| 6228 | MG945339 | Unassigned | Unassigned | Unassigned | 1116_0 | Clustered | 14 | VC_1116_0 | 14 | 0.7927 | 1 | 0.7927 | 0 | 0 | 0 | 1 | Ref |
| 6229 | MG945341 | Unassigned | Unassigned | Unassigned | 672_0 | Clustered | 12 | VC_672_0 | 12 | 1 | 1 | 1 | 0 | 0 | 0 | 1 | Ref |
| 6230 | MG945343 | Unassigned | Unassigned | Unassigned | 1287_0 | Clustered | 7 | VC_1287_0 | 7 | 1 | 1 | 1 | 0 | 0 | 0 | 1 | Ref |
| 6231 | MG945344 | Unassigned | Unassigned | Unassigned | 1285_0 | Clustered | 25 | VC_1285_0 | 25 | 1 | 1 | 1 | 0 | 0 | 0 | 1 | Ref |
| 6232 | MG945345 | Unassigned | Unassigned | Unassigned | 1291_0 | Clustered | 2 | VC_1291_0 | 2 | 1 | 1 | 1 | 0 | 0 | 0 | 1 | Ref |
| 6233 | MG945346 | Unassigned | Unassigned | Unassigned | 1104_0 | Clustered | 154 | VC_1104_0 | 154 | 0.8484 | 1 | 0.8484 | 0 | 0 | 0 | 1 | Ref |
| 6234 | MG945347 | Unassigned | Unassigned | Unassigned | 1125_0 | Clustered | 20 | VC_1125_0 | 20 | 0.5599 | 1 | 0.5599 | 0 | 0 | 0 | 1 | Ref |
| 6235 | MG945348 | Unassigned | Unassigned | Unassigned | 1292_0 | Clustered | 3 | VC_1292_0 | 3 | 1 | 1 | 1 | 0 | 0 | 0 | 1 | Ref |
| 6236 | MG945349 | Unassigned | Unassigned | Unassigned | 1261_0 | Clustered | 5 | VC_1261_0 | 5 | 0.765 | 0.99993003 | 0.7649 | 0 | 0 | 0 | 1 | Ref |
| 6237 | MG945350 | Unassigned | Unassigned | Unassigned | 1293_0 | Clustered | 4 | VC_1293_0 | 4 | 0.7847 | 0.99289298 | 0.7791 | 0 | 0 | 0 | 1 | Ref |
| 6238 | MG945351 | Unassigned | Unassigned | Unassigned | 1104_0 | Clustered | 154 | VC_1104_0 | 154 | 0.8484 | 1 | 0.8484 | 0 | 0 | 0 | 1 | Ref |
| 6239 | MG945352 | Unassigned | Unassigned | Unassigned | 1193_0 | Clustered | 3 | VC_1193_0 | 3 | 0.5473 | 0.99523779 | 0.5447 | 0 | 0 | 0 | 1 | Ref |
| 6240 | MG945354 | Unassigned | Unassigned | Unassigned | 1294_0 | Clustered | 6 | VC_1294_0 | 6 | 0.6626 | 0.99999272 | 0.6626 | 0 | 0 | 0 | 1 | Ref |
| 6241 | MG945355 | Unassigned | Unassigned | Unassigned | 1104_0 | Clustered | 154 | VC_1104_0 | 154 | 0.8484 | 1 | 0.8484 | 0 | 0 | 0 | 1 | Ref |
| 6242 | MG945357 | Unassigned | Unassigned | Unassigned | 1148_0 | Clustered | 16 | VC_1148_0 | 16 | 0.8821 | 1 | 0.8821 | 0 | 0 | 0 | 1 | Ref |
| 6243 | MG945360 | Unassigned | Unassigned | Unassigned | 1295_0 | Clustered | 3 | VC_1295_0 | 3 | 1 | 0 | 0 | 0 | 0 | 0 | 1 | Ref |
| 6244 | MG945361 | Unassigned | Unassigned | Unassigned | 1104_0 | Clustered | 154 | VC_1104_0 | 154 | 0.8484 | 1 | 0.8484 | 0 | 0 | 0 | 1 | Ref |
| 6245 | MG945362 | Unassigned | Unassigned | Unassigned | 1296_0 | Clustered | 4 | VC_1296_0 | 4 | 0.8684 | 0.91151676 | 0.7915 | 0 | 0 | 0 | 1 | Ref |
| 6246 | MG945363 | Unassigned | Unassigned | Unassigned | 1104_0 | Clustered | 154 | VC_1104_0 | 154 | 0.8484 | 1 | 0.8484 | 0 | 0 | 0 | 1 | Ref |
| 6247 | MG945364 | Unassigned | Unassigned | Unassigned | 1144_0 | Clustered | 7 | VC_1144_0 | 7 | 0.7076 | 1 | 0.7076 | 0 | 0 | 0 | 1 | Ref |
| 6248 | MG945365 | Unassigned | Unassigned | Unassigned | 1259_0 | Clustered | 8 | VC_1259_0 | 8 | 0.8225 | 1 | 0.8225 | 0 | 0 | 0 | 1 | Ref |
| 6249 | MG945381 | Unassigned | Unassigned | Unassigned | 1126_0 | Clustered | 8 | VC_1126_0 | 8 | 0.2864 | 1 | 0.2864 | 0 | 0 | 0 | 1 | Ref |
| 6250 | MG945382 | Unassigned | Unassigned | Unassigned | 1126_0 | Clustered | 8 | VC_1126_0 | 8 | 0.2864 | 1 | 0.2864 | 0 | 0 | 0 | 1 | Ref |
| 6251 | MG945383 | Unassigned | Unassigned | Unassigned | 1127_0 | Clustered | 19 | VC_1127_0 | 19 | 0.7611 | 1 | 0.7611 | 0 | 0 | 0 | 1 | Ref |
| 6252 | MG945384 | Unassigned | Unassigned | Unassigned | 1247_0 | Clustered | 7 | VC_1247_0 | 7 | 0.9086 | 0.99998959 | 0.9086 | 0 | 0 | 0 | 1 | Ref |
| 6253 | MG945385 | Unassigned | Unassigned | Unassigned | 1125_0 | Clustered | 20 | VC_1125_0 | 20 | 0.5599 | 1 | 0.5599 | 0 | 0 | 0 | 1 | Ref |
| 6254 | MG945386 | Unassigned | Unassigned | Unassigned | 1108_0 | Clustered | 4 | VC_1108_0 | 4 | 0.0485 | 0.99999021 | 0.0485 | 0 | 0 | 0 | 1 | Ref |
| 6255 | MG945387 | Unassigned | Unassigned | Unassigned | 1104_0 | Clustered | 154 | VC_1104_0 | 154 | 0.8484 | 1 | 0.8484 | 0 | 0 | 0 | 1 | Ref |
| 6256 | MG945388 | Unassigned | Unassigned | Unassigned | 1127_0 | Clustered | 19 | VC_1127_0 | 19 | 0.7611 | 1 | 0.7611 | 0 | 0 | 0 | 1 | Ref |
| 6257 | MG945389 | Unassigned | Unassigned | Unassigned | 1125_0 | Clustered | 20 | VC_1125_0 | 20 | 0.5599 | 1 | 0.5599 | 0 | 0 | 0 | 1 | Ref |
| 6258 | MG945390 | Unassigned | Unassigned | Unassigned | 1243_0 | Clustered | 52 | VC_1243_0 | 52 | 0.9126 | 0.92708311 | 0.8461 | 0 | 0 | 0 | 1 | Ref |
| 6259 | MG945391 | Unassigned | Unassigned | Unassigned | 1124_0 | Clustered | 10 | VC_1124_0 | 10 | 0.6268 | 1 | 0.6268 | 0 | 0 | 0 | 1 | Ref |
| 6260 | MG945392 | Unassigned | Unassigned | Unassigned | 1272_0 | Clustered | 5 | VC_1272_0 | 5 | 1 | 1 | 1 | 0 | 0 | 0 | 1 | Ref |
| 6261 | MG945393 | Unassigned | Unassigned | Unassigned | 1104_0 | Clustered | 154 | VC_1104_0 | 154 | 0.8484 | 1 | 0.8484 | 0 | 0 | 0 | 1 | Ref |
| 6262 | MG945394 | Unassigned | Unassigned | Unassigned | 1116_0 | Clustered | 14 | VC_1116_0 | 14 | 0.7927 | 1 | 0.7927 | 0 | 0 | 0 | 1 | Ref |
| 6263 | MG945395 | Unassigned | Unassigned | Unassigned | 1297_0 | Clustered | 7 | VC_1297_0 | 7 | 1 | 1 | 1 | 0 | 0 | 0 | 1 | Ref |
| 6264 | MG945396 | Unassigned | Unassigned | Unassigned | 1243_0 | Clustered | 52 | VC_1243_0 | 52 | 0.9126 | 0.92708311 | 0.8461 | 0 | 0 | 0 | 1 | Ref |
| 6265 | MG945397 | Unassigned | Unassigned | Unassigned | 1298_0 | Clustered | 3 | VC_1298_0 | 3 | 0.4889 | 0.98167473 | 0.4799 | 0 | 0 | 0 | 1 | Ref |
| 6266 | MG945398 | Unassigned | Unassigned | Unassigned | 1104_0 | Clustered | 154 | VC_1104_0 | 154 | 0.8484 | 1 | 0.8484 | 0 | 0 | 0 | 1 | Ref |
| 6267 | MG945401 | Unassigned | Unassigned | Unassigned | 1251_0 | Clustered | 15 | VC_1251_0 | 15 | 0.9082 | 1 | 0.9082 | 0 | 0 | 0 | 1 | Ref |
| 6268 | MG945402 | Unassigned | Unassigned | Unassigned | 1104_0 | Clustered | 154 | VC_1104_0 | 154 | 0.8484 | 1 | 0.8484 | 0 | 0 | 0 | 1 | Ref |
| 6269 | MG945403 | Unassigned | Unassigned | Unassigned | 1116_0 | Clustered | 14 | VC_1116_0 | 14 | 0.7927 | 1 | 0.7927 | 0 | 0 | 0 | 1 | Ref |
| 6270 | MG945404 | Unassigned | Unassigned | Unassigned | 1267_0 | Clustered | 7 | VC_1267_0 | 7 | 1 | 1 | 1 | 0 | 0 | 0 | 1 | Ref |
| 6271 | MG945405 | Unassigned | Unassigned | Unassigned | 1146_0 | Clustered | 3 | VC_1146_0 | 3 | 0.3641 | 0.99867077 | 0.3636 | 0 | 0 | 0 | 1 | Ref |
| 6272 | MG945407 | Unassigned | Unassigned | Unassigned | 1104_0 | Clustered | 154 | VC_1104_0 | 154 | 0.8484 | 1 | 0.8484 | 0 | 0 | 0 | 1 | Ref |
| 6273 | MG945408 | Unassigned | Unassigned | Unassigned | 1266_0 | Clustered | 5 | VC_1266_0 | 5 | 1 | 1 | 1 | 0 | 0 | 0 | 1 | Ref |
| 6274 | MG945409 | Unassigned | Unassigned | Unassigned | 1299_0 | Clustered | 2 | VC_1299_0 | 2 | 1 | 1 | 1 | 0 | 0 | 0 | 1 | Ref |
| 6275 | MG945410 | Unassigned | Unassigned | Unassigned | 1125_0 | Clustered | 20 | VC_1125_0 | 20 | 0.5599 | 1 | 0.5599 | 0 | 0 | 0 | 1 | Ref |
| 6276 | MG945411 | Unassigned | Unassigned | Unassigned | 1261_0 | Clustered | 5 | VC_1261_0 | 5 | 0.765 | 0.99993003 | 0.7649 | 0 | 0 | 0 | 1 | Ref |
| 6277 | MG945412 | Unassigned | Unassigned | Unassigned | 1300_0 | Clustered | 2 | VC_1300_0 | 2 | 0.664 | 0.72985431 | 0.4846 | 0 | 0 | 0 | 1 | Ref |
| 6278 | MG945413 | Unassigned | Unassigned | Unassigned | 1148_0 | Clustered | 16 | VC_1148_0 | 16 | 0.8821 | 1 | 0.8821 | 0 | 0 | 0 | 1 | Ref |
| 6279 | MG945414 | Unassigned | Unassigned | Unassigned | 1253_0 | Clustered | 6 | VC_1253_0 | 6 | 0.6097 | 1 | 0.6097 | 0 | 0 | 0 | 1 | Ref |
| 6280 | MG945415 | Unassigned | Unassigned | Unassigned | 1104_0 | Clustered | 154 | VC_1104_0 | 154 | 0.8484 | 1 | 0.8484 | 0 | 0 | 0 | 1 | Ref |
| 6281 | MG945416 | Unassigned | Unassigned | Unassigned | 1125_0 | Clustered | 20 | VC_1125_0 | 20 | 0.5599 | 1 | 0.5599 | 0 | 0 | 0 | 1 | Ref |
| 6282 | MG945417 | Unassigned | Unassigned | Unassigned | 1301_0 | Clustered | 2 | VC_1301_0 | 2 | 1 | 1 | 1 | 0 | 0 | 0 | 1 | Ref |
| 6283 | MG945418 | Unassigned | Unassigned | Unassigned | 1154_0 | Clustered | 5 | VC_1154_0 | 5 | 1 | 1 | 1 | 0 | 0 | 0 | 1 | Ref |
| 6284 | MG945419 | Unassigned | Unassigned | Unassigned | 1301_0 | Clustered | 2 | VC_1301_0 | 2 | 1 | 1 | 1 | 0 | 0 | 0 | 1 | Ref |
| 6285 | MG945420 | Unassigned | Unassigned | Unassigned | 1264_0 | Clustered | 6 | VC_1264_0 | 6 | 0.7062 | 0.92055725 | 0.6501 | 0 | 0 | 0 | 1 | Ref |
| 6286 | MG945421 | Unassigned | Unassigned | Unassigned | 1297_0 | Clustered | 7 | VC_1297_0 | 7 | 1 | 1 | 1 | 0 | 0 | 0 | 1 | Ref |
| 6287 | MG945424 | Unassigned | Unassigned | Unassigned | 1104_0 | Clustered | 154 | VC_1104_0 | 154 | 0.8484 | 1 | 0.8484 | 0 | 0 | 0 | 1 | Ref |
| 6288 | MG945425 | Unassigned | Unassigned | Unassigned | 1278_0 | Clustered | 3 | VC_1278_0 | 3 | 0.3961 | 0.99705286 | 0.395 | 0 | 0 | 0 | 1 | Ref |
| 6289 | MG945426 | Unassigned | Unassigned | Unassigned | 1151_0 | Clustered | 9 | VC_1151_0 | 9 | 0.8548 | 1 | 0.8548 | 0 | 0 | 0 | 1 | Ref |
| 6290 | MG945427 | Unassigned | Unassigned | Unassigned | 1302_0 | Clustered | 3 | VC_1302_0 | 3 | 1 | 1 | 1 | 0 | 0 | 0 | 1 | Ref |
| 6291 | MG945429 | Unassigned | Unassigned | Unassigned | 1104_0 | Clustered | 154 | VC_1104_0 | 154 | 0.8484 | 1 | 0.8484 | 0 | 0 | 0 | 1 | Ref |
| 6292 | MG945430 | Unassigned | Unassigned | Unassigned | 1127_0 | Clustered | 19 | VC_1127_0 | 19 | 0.7611 | 1 | 0.7611 | 0 | 0 | 0 | 1 | Ref |
| 6293 | MG945432 | Unassigned | Unassigned | Unassigned | 1293_0 | Clustered | 4 | VC_1293_0 | 4 | 0.7847 | 0.99289298 | 0.7791 | 0 | 0 | 0 | 1 | Ref |
| 6294 | MG945433 | Unassigned | Unassigned | Unassigned | 1197_0 | Clustered | 15 | VC_1197_0 | 15 | 0.8991 | 0.76608394 | 0.6888 | 0 | 0 | 0 | 1 | Ref |
| 6295 | MG945434 | Unassigned | Unassigned | Unassigned | 1279_0 | Clustered | 4 | VC_1279_0 | 4 | 0.6058 | 0.76972092 | 0.4663 | 0 | 0 | 0 | 1 | Ref |
| 6296 | MG945435 | Unassigned | Unassigned | Unassigned | 1104_0 | Clustered | 154 | VC_1104_0 | 154 | 0.8484 | 1 | 0.8484 | 0 | 0 | 0 | 1 | Ref |
| 6297 | MG945437 | Unassigned | Unassigned | Unassigned | 1125_0 | Clustered | 20 | VC_1125_0 | 20 | 0.5599 | 1 | 0.5599 | 0 | 0 | 0 | 1 | Ref |
| 6298 | MG945438 | Unassigned | Unassigned | Unassigned | 1104_0 | Clustered | 154 | VC_1104_0 | 154 | 0.8484 | 1 | 0.8484 | 0 | 0 | 0 | 1 | Ref |
| 6299 | MG945439 | Unassigned | Unassigned | Unassigned | 1303_0 | Clustered | 6 | VC_1303_0 | 6 | 1 | 1 | 1 | 0 | 0 | 0 | 1 | Ref |
| 6300 | MG945441 | Unassigned | Unassigned | Unassigned | 1149_0 | Clustered | 5 | VC_1149_0 | 5 | 0.4768 | 0.9999998 | 0.4768 | 0 | 0 | 0 | 1 | Ref |
| 6301 | MG945444 | Unassigned | Unassigned | Unassigned | 1104_0 | Clustered | 154 | VC_1104_0 | 154 | 0.8484 | 1 | 0.8484 | 0 | 0 | 0 | 1 | Ref |
| 6302 | MG945445 | Unassigned | Unassigned | Unassigned | 1253_0 | Clustered | 6 | VC_1253_0 | 6 | 0.6097 | 1 | 0.6097 | 0 | 0 | 0 | 1 | Ref |
| 6303 | MG945447 | Unassigned | Unassigned | Unassigned | 1127_0 | Clustered | 19 | VC_1127_0 | 19 | 0.7611 | 1 | 0.7611 | 0 | 0 | 0 | 1 | Ref |
| 6304 | MG945448 | Unassigned | Unassigned | Unassigned | 1256_0 | Clustered | 4 | VC_1256_0 | 4 | 0.581 | 0.94751743 | 0.5505 | 0 | 0 | 0 | 1 | Ref |
| 6305 | MG945449 | Unassigned | Unassigned | Unassigned | 1257_0 | Clustered | 6 | VC_1257_0 | 6 | 0.6517 | 0.99999969 | 0.6517 | 0 | 0 | 0 | 1 | Ref |
| 6306 | MG945452 | Unassigned | Unassigned | Unassigned | 1243_0 | Clustered | 52 | VC_1243_0 | 52 | 0.9126 | 0.92708311 | 0.8461 | 0 | 0 | 0 | 1 | Ref |
| 6307 | MG945453 | Unassigned | Unassigned | Unassigned | 1211_0 | Clustered | 25 | VC_1211_0 | 25 | 1 | 1 | 1 | 0 | 0 | 0 | 1 | Ref |
| 6308 | MG945455 | Unassigned | Unassigned | Unassigned | 1211_0 | Clustered | 25 | VC_1211_0 | 25 | 1 | 1 | 1 | 0 | 0 | 0 | 1 | Ref |
| 6309 | MG945456 | Unassigned | Unassigned | Unassigned | 1211_0 | Clustered | 25 | VC_1211_0 | 25 | 1 | 1 | 1 | 0 | 0 | 0 | 1 | Ref |
| 6310 | MG945457 | Unassigned | Unassigned | Unassigned | 1233_0 | Clustered | 16 | VC_1233_0 | 16 | 0.8971 | 1 | 0.8971 | 0 | 0 | 0 | 1 | Ref |
| 6311 | MG945458 | Unassigned | Unassigned | Unassigned | 1257_0 | Clustered | 6 | VC_1257_0 | 6 | 0.6517 | 0.99999969 | 0.6517 | 0 | 0 | 0 | 1 | Ref |
| 6312 | MG945459 | Unassigned | Unassigned | Unassigned | 1104_0 | Clustered | 154 | VC_1104_0 | 154 | 0.8484 | 1 | 0.8484 | 0 | 0 | 0 | 1 | Ref |
| 6313 | MG945460 | Unassigned | Unassigned | Unassigned | 1255_0 | Clustered | 4 | VC_1255_0 | 4 | 0.248 | 0.99828153 | 0.2476 | 0 | 0 | 0 | 1 | Ref |
| 6314 | MG945461 | Unassigned | Unassigned | Unassigned | 1104_0 | Clustered | 154 | VC_1104_0 | 154 | 0.8484 | 1 | 0.8484 | 0 | 0 | 0 | 1 | Ref |
| 6315 | MG945462 | Unassigned | Unassigned | Unassigned | 1261_0 | Clustered | 5 | VC_1261_0 | 5 | 0.765 | 0.99993003 | 0.7649 | 0 | 0 | 0 | 1 | Ref |
| 6316 | MG945463 | Unassigned | Unassigned | Unassigned | 1062_0 | Clustered | 45 | VC_1062_0 | 45 | 0.9734 | 0.50949847 | 0.4959 | 0 | 0 | 0 | 1 | Ref |

| | | | | | | | | | | | | | | | | | |
|---|---|---|---|---|---|---|---|---|---|---|---|---|---|---|---|---|---|
| 6317 | MG945464 | Unassigned | Unassigned | Unassigned | 1245_0 | Clustered | 2 | VC_1245_0 | 2 | 1 | 1 | 1 | 0 | 0 | 0 | 1 | Ref |
| 6318 | MG945465 | Unassigned | Unassigned | Unassigned | 995_0 | Clustered | 69 | VC_995_0 | 69 | 0.8547 | 1 | 0.8547 | 0 | 0 | 0 | 1 | Ref |
| 6319 | MG945467 | Unassigned | Unassigned | Unassigned | 1104_0 | Clustered | 154 | VC_1104_0 | 154 | 0.8484 | 1 | 0.8484 | 0 | 0 | 0 | 1 | Ref |
| 6320 | MG945468 | Unassigned | Unassigned | Unassigned | 1285_0 | Clustered | 25 | VC_1285_0 | 25 | 1 | 1 | 1 | 0 | 0 | 0 | 1 | Ref |
| 6321 | MG945469 | Unassigned | Unassigned | Unassigned | 1305_0 | Clustered | 5 | VC_1305_0 | 5 | 0.7701 | 0.66985041 | 0.5159 | 0 | 0 | 0 | 1 | Ref |
| 6322 | MG945470 | Unassigned | Unassigned | Unassigned | 1264_0 | Clustered | 6 | VC_1264_0 | 6 | 0.7062 | 0.92055725 | 0.6501 | 0 | 0 | 0 | 1 | Ref |
| 6323 | MG945472 | Unassigned | Unassigned | Unassigned | 1119_0 | Clustered | 20 | VC_1119_0 | 20 | 0.6751 | 0.99997892 | 0.6751 | 0 | 0 | 0 | 1 | Ref |
| 6324 | MG945473 | Unassigned | Unassigned | Unassigned | 1054_0 | Clustered | 12 | VC_1054_0 | 12 | 0.2863 | 0.99746644 | 0.2855 | 0 | 0 | 0 | 1 | Ref |
| 6325 | MG945475 | Unassigned | Unassigned | Unassigned | 1085_0 | Clustered | 2 | VC_1085_0 | 2 | 0.5907 | 0.9331928 | 0.5513 | 0 | 0 | 0 | 1 | Ref |
| 6326 | MG945476 | Unassigned | Unassigned | Unassigned | 1104_0 | Clustered | 154 | VC_1104_0 | 154 | 0.8484 | 1 | 0.8484 | 0 | 0 | 0 | 1 | Ref |
| 6327 | MG945477 | Unassigned | Unassigned | Unassigned | 995_0 | Clustered | 69 | VC_995_0 | 69 | 0.8547 | 1 | 0.8547 | 0 | 0 | 0 | 1 | Ref |
| 6328 | MG945478 | Unassigned | Unassigned | Unassigned | 1295_0 | Clustered | 3 | VC_1295_0 | 3 | 1 | 0 | 0 | 0 | 0 | 0 | 1 | Ref |
| 6329 | MG945480 | Unassigned | Unassigned | Unassigned | 1297_0 | Clustered | 7 | VC_1297_0 | 7 | 1 | 1 | 1 | 0 | 0 | 0 | 1 | Ref |
| 6330 | MG945481 | Unassigned | Unassigned | Unassigned | 1117_0 | Clustered | 5 | VC_1117_0 | 5 | 0.4688 | 0.99999974 | 0.4688 | 0 | 0 | 0 | 1 | Ref |
| 6331 | MG945482 | Unassigned | Unassigned | Unassigned | 1247_0 | Clustered | 7 | VC_1247_0 | 7 | 0.9086 | 0.99998959 | 0.9086 | 0 | 0 | 0 | 1 | Ref |
| 6332 | MG945483 | Unassigned | Unassigned | Unassigned | 1306_0 | Clustered | 2 | VC_1306_0 | 2 | 1 | 1 | 1 | 0 | 0 | 0 | 1 | Ref |
| 6333 | MG945485 | Unassigned | Unassigned | Unassigned | 1243_0 | Clustered | 52 | VC_1243_0 | 52 | 0.9126 | 0.92708311 | 0.8461 | 0 | 0 | 0 | 1 | Ref |
| 6334 | MG945486 | Unassigned | Unassigned | Unassigned | 1104_0 | Clustered | 154 | VC_1104_0 | 154 | 0.8484 | 1 | 0.8484 | 0 | 0 | 0 | 1 | Ref |
| 6335 | MG945487 | Unassigned | Unassigned | Unassigned | 1149_0 | Clustered | 5 | VC_1149_0 | 5 | 0.4768 | 0.9999998 | 0.4768 | 0 | 0 | 0 | 1 | Ref |
| 6336 | MG945488 | Unassigned | Unassigned | Unassigned | 1276_0 | Clustered | 7 | VC_1276_0 | 7 | 0.8695 | 0.99982476 | 0.8693 | 0 | 0 | 0 | 1 | Ref |
| 6337 | MG945489 | Unassigned | Unassigned | Unassigned | 1111_0 | Clustered | 2 | VC_1111_0 | 2 | 0.2635 | 0.89847992 | 0.2367 | 0 | 0 | 0 | 1 | Ref |
| 6338 | MG945490 | Unassigned | Unassigned | Unassigned | 1287_0 | Clustered | 7 | VC_1287_0 | 7 | 1 | 1 | 1 | 0 | 0 | 0 | 1 | Ref |
| 6339 | MG945492 | Unassigned | Unassigned | Unassigned | 1281_0 | Clustered | 7 | VC_1281_0 | 7 | 0.9212 | 0.99995294 | 0.9212 | 0 | 0 | 0 | 1 | Ref |
| 6340 | MG945494 | Unassigned | Unassigned | Unassigned | 1277_0 | Clustered | 14 | VC_1277_0 | 14 | 0.8218 | 1 | 0.8218 | 0 | 0 | 0 | 1 | Ref |
| 6341 | MG945495 | Unassigned | Unassigned | Unassigned | 1261_0 | Clustered | 5 | VC_1261_0 | 5 | 0.765 | 0.99993003 | 0.7649 | 0 | 0 | 0 | 1 | Ref |
| 6342 | MG945496 | Unassigned | Unassigned | Unassigned | 1255_0 | Clustered | 4 | VC_1255_0 | 4 | 0.248 | 0.99828153 | 0.2476 | 0 | 0 | 0 | 1 | Ref |
| 6343 | MG945498 | Unassigned | Unassigned | Unassigned | 1211_0 | Clustered | 25 | VC_1211_0 | 25 | 1 | 1 | 1 | 0 | 0 | 0 | 1 | Ref |
| 6344 | MG945499 | Unassigned | Unassigned | Unassigned | 1104_0 | Clustered | 154 | VC_1104_0 | 154 | 0.8484 | 1 | 0.8484 | 0 | 0 | 0 | 1 | Ref |
| 6345 | MG945500 | Unassigned | Unassigned | Unassigned | 1308_0 | Clustered | 12 | VC_1308_0 | 12 | 0.8184 | 0.9810731 | 0.8029 | 0 | 0 | 0 | 1 | Ref |
| 6346 | MG945501 | Unassigned | Unassigned | Unassigned | 1287_0 | Clustered | 7 | VC_1287_0 | 7 | 1 | 1 | 1 | 0 | 0 | 0 | 1 | Ref |
| 6347 | MG945502 | Unassigned | Unassigned | Unassigned | 1197_0 | Clustered | 15 | VC_1197_0 | 15 | 0.8991 | 0.76608394 | 0.6888 | 0 | 0 | 0 | 1 | Ref |
| 6348 | MG945504 | Unassigned | Unassigned | Unassigned | 1310_0 | Clustered | 2 | VC_1310_0 | 2 | 1 | 1 | 1 | 0 | 0 | 0 | 1 | Ref |
| 6349 | MG945505 | Unassigned | Unassigned | Unassigned | 1149_0 | Clustered | 5 | VC_1149_0 | 5 | 0.4768 | 0.9999998 | 0.4768 | 0 | 0 | 0 | 1 | Ref |
| 6350 | MG945506 | Unassigned | Unassigned | Unassigned | 1148_0 | Clustered | 16 | VC_1148_0 | 16 | 0.8821 | 1 | 0.8821 | 0 | 0 | 0 | 1 | Ref |
| 6351 | MG945507 | Unassigned | Unassigned | Unassigned | 1148_0 | Clustered | 16 | VC_1148_0 | 16 | 0.8821 | 1 | 0.8821 | 0 | 0 | 0 | 1 | Ref |
| 6352 | MG945508 | Unassigned | Unassigned | Unassigned | 1104_0 | Clustered | 154 | VC_1104_0 | 154 | 0.8484 | 1 | 0.8484 | 0 | 0 | 0 | 1 | Ref |
| 6353 | MG945509 | Unassigned | Unassigned | Unassigned | 1211_0 | Clustered | 25 | VC_1211_0 | 25 | 1 | 1 | 1 | 0 | 0 | 0 | 1 | Ref |
| 6354 | MG945511 | Unassigned | Unassigned | Unassigned | 1243_0 | Clustered | 52 | VC_1243_0 | 52 | 0.9126 | 0.92708311 | 0.8461 | 0 | 0 | 0 | 1 | Ref |
| 6355 | MG945512 | Unassigned | Unassigned | Unassigned | 1259_0 | Clustered | 8 | VC_1259_0 | 8 | 0.8225 | 1 | 0.8225 | 0 | 0 | 0 | 1 | Ref |
| 6356 | MG945514 | Unassigned | Unassigned | Unassigned | 1127_0 | Clustered | 19 | VC_1127_0 | 19 | 0.7611 | 1 | 0.7611 | 0 | 0 | 0 | 1 | Ref |
| 6357 | MG945515 | Unassigned | Unassigned | Unassigned | 1243_0 | Clustered | 52 | VC_1243_0 | 52 | 0.9126 | 0.92708311 | 0.8461 | 0 | 0 | 0 | 1 | Ref |
| 6358 | MG945517 | Unassigned | Unassigned | Unassigned | 1104_0 | Clustered | 154 | VC_1104_0 | 154 | 0.8484 | 1 | 0.8484 | 0 | 0 | 0 | 1 | Ref |
| 6359 | MG945518 | Unassigned | Unassigned | Unassigned | 1264_0 | Clustered | 6 | VC_1264_0 | 6 | 0.7062 | 0.92055725 | 0.6501 | 0 | 0 | 0 | 1 | Ref |
| 6360 | MG945519 | Unassigned | Unassigned | Unassigned | 1138_0 | Clustered | 4 | VC_1138_0 | 4 | 0.9437 | 0.98964986 | 0.934 | 0 | 0 | 0 | 1 | Ref |
| 6361 | MG945520 | Unassigned | Unassigned | Unassigned | 672_0 | Clustered | 12 | VC_672_0 | 12 | 1 | 1 | 1 | 0 | 0 | 0 | 1 | Ref |
| 6362 | MG945523 | Unassigned | Unassigned | Unassigned | 1230_0 | Clustered | 3 | VC_1230_0 | 3 | 0.4802 | 0.82711071 | 0.3971 | 0 | 0 | 0 | 1 | Ref |
| 6363 | MG945524 | Unassigned | Unassigned | Unassigned | 1285_0 | Clustered | 25 | VC_1285_0 | 25 | 1 | 1 | 1 | 0 | 0 | 0 | 1 | Ref |
| 6364 | MG945525 | Unassigned | Unassigned | Unassigned | 1144_0 | Clustered | 7 | VC_1144_0 | 7 | 0.7076 | 1 | 0.7076 | 0 | 0 | 0 | 1 | Ref |
| 6365 | MG945526 | Unassigned | Unassigned | Unassigned | 1104_0 | Clustered | 154 | VC_1104_0 | 154 | 0.8484 | 1 | 0.8484 | 0 | 0 | 0 | 1 | Ref |
| 6366 | MG945529 | Unassigned | Unassigned | Unassigned | 1311_0 | Clustered | 5 | VC_1311_0 | 5 | 0.8316 | 0.99879053 | 0.8306 | 0 | 0 | 0 | 1 | Ref |
| 6367 | MG945533 | Unassigned | Unassigned | Unassigned | 1312_0 | Clustered | 3 | VC_1312_0 | 3 | 1 | 1 | 1 | 0 | 0 | 0 | 1 | Ref |
| 6368 | MG945534 | Unassigned | Unassigned | Unassigned | 1119_0 | Clustered | 20 | VC_1119_0 | 20 | 0.6751 | 0.99997892 | 0.6751 | 0 | 0 | 0 | 1 | Ref |
| 6369 | MG945538 | Unassigned | Unassigned | Unassigned | 1125_0 | Clustered | 20 | VC_1125_0 | 20 | 0.5599 | 1 | 0.5599 | 0 | 0 | 0 | 1 | Ref |
| 6370 | MG945539 | Unassigned | Unassigned | Unassigned | 1297_0 | Clustered | 7 | VC_1297_0 | 7 | 1 | 1 | 1 | 0 | 0 | 0 | 1 | Ref |
| 6371 | MG945540 | Unassigned | Unassigned | Unassigned | 1259_0 | Clustered | 8 | VC_1259_0 | 8 | 0.8225 | 1 | 0.8225 | 0 | 0 | 0 | 1 | Ref |
| 6372 | MG945541 | Unassigned | Unassigned | Unassigned | 1104_0 | Clustered | 154 | VC_1104_0 | 154 | 0.8484 | 1 | 0.8484 | 0 | 0 | 0 | 1 | Ref |
| 6373 | MG945542 | Unassigned | Unassigned | Unassigned | 1116_0 | Clustered | 14 | VC_1116_0 | 14 | 0.7927 | 1 | 0.7927 | 0 | 0 | 0 | 1 | Ref |
| 6374 | MG945544 | Unassigned | Unassigned | Unassigned | 1252_0 | Clustered | 3 | VC_1252_0 | 3 | 0.3237 | 0.99790393 | 0.323 | 0 | 0 | 0 | 1 | Ref |
| 6375 | MG945546 | Unassigned | Unassigned | Unassigned | 198_0 | Clustered | 28 | VC_198_0 | 28 | 0.2429 | 0.99971175 | 0.2429 | 0 | 0 | 0 | 1 | Ref |
| 6376 | MG945548 | Unassigned | Unassigned | Unassigned | 1104_0 | Clustered | 154 | VC_1104_0 | 154 | 0.8484 | 1 | 0.8484 | 0 | 0 | 0 | 1 | Ref |
| 6377 | MG945549 | Unassigned | Unassigned | Unassigned | 1154_0 | Clustered | 5 | VC_1154_0 | 5 | 1 | 1 | 1 | 0 | 0 | 0 | 1 | Ref |
| 6378 | MG945550 | Unassigned | Unassigned | Unassigned | 1116_0 | Clustered | 14 | VC_1116_0 | 14 | 0.7927 | 1 | 0.7927 | 0 | 0 | 0 | 1 | Ref |
| 6379 | MG945551 | Unassigned | Unassigned | Unassigned | 672_0 | Clustered | 12 | VC_672_0 | 12 | 1 | 1 | 1 | 0 | 0 | 0 | 1 | Ref |
| 6380 | MG945554 | Unassigned | Unassigned | Unassigned | 1319_0 | Clustered | 2 | VC_1319_0 | 2 | 1 | 1 | 1 | 0 | 0 | 0 | 1 | Ref |
| 6381 | MG945556 | Unassigned | Unassigned | Unassigned | 1285_0 | Clustered | 25 | VC_1285_0 | 25 | 1 | 1 | 1 | 0 | 0 | 0 | 1 | Ref |
| 6382 | MG945557 | Unassigned | Unassigned | Unassigned | 1233_0 | Clustered | 16 | VC_1233_0 | 16 | 0.8971 | 1 | 0.8971 | 0 | 0 | 0 | 1 | Ref |
| 6383 | MG945559 | Unassigned | Unassigned | Unassigned | 1104_0 | Clustered | 154 | VC_1104_0 | 154 | 0.8484 | 1 | 0.8484 | 0 | 0 | 0 | 1 | Ref |
| 6384 | MG945560 | Unassigned | Unassigned | Unassigned | 1056_0 | Clustered | 4 | VC_1056_0 | 4 | 0.169 | 0.84110627 | 0.1421 | 0 | 0 | 0 | 1 | Ref |
| 6385 | MG945561 | Unassigned | Unassigned | Unassigned | 1139_0 | Clustered | 4 | VC_1139_0 | 4 | 1 | 1 | 1 | 0 | 0 | 0 | 1 | Ref |
| 6386 | MG945562 | Unassigned | Unassigned | Unassigned | 1285_0 | Clustered | 25 | VC_1285_0 | 25 | 1 | 1 | 1 | 0 | 0 | 0 | 1 | Ref |
| 6387 | MG945563 | Unassigned | Unassigned | Unassigned | 1277_0 | Clustered | 14 | VC_1277_0 | 14 | 0.8218 | 1 | 0.8218 | 0 | 0 | 0 | 1 | Ref |
| 6388 | MG945564 | Unassigned | Unassigned | Unassigned | 1211_0 | Clustered | 25 | VC_1211_0 | 25 | 1 | 1 | 1 | 0 | 0 | 0 | 1 | Ref |
| 6389 | MG945565 | Unassigned | Unassigned | Unassigned | 1104_0 | Clustered | 154 | VC_1104_0 | 154 | 0.8484 | 1 | 0.8484 | 0 | 0 | 0 | 1 | Ref |
| 6390 | MG945566 | Unassigned | Unassigned | Unassigned | 1104_0 | Clustered | 154 | VC_1104_0 | 154 | 0.8484 | 1 | 0.8484 | 0 | 0 | 0 | 1 | Ref |
| 6391 | MG945567 | Unassigned | Unassigned | Unassigned | 1274_0 | Clustered | 2 | VC_1274_0 | 2 | 0.6386 | 0.72985431 | 0.4661 | 0 | 0 | 0 | 1 | Ref |
| 6392 | MG945568 | Unassigned | Unassigned | Unassigned | 1311_0 | Clustered | 5 | VC_1311_0 | 5 | 0.8316 | 0.99879053 | 0.8306 | 0 | 0 | 0 | 1 | Ref |
| 6393 | MG945569 | Unassigned | Unassigned | Unassigned | 1122_0 | Clustered | 10 | VC_1122_0 | 10 | 0.9643 | 0.99999697 | 0.9643 | 0 | 0 | 0 | 1 | Ref |
| 6394 | MG945570 | Unassigned | Unassigned | Unassigned | 1104_0 | Clustered | 154 | VC_1104_0 | 154 | 0.8484 | 1 | 0.8484 | 0 | 0 | 0 | 1 | Ref |
| 6395 | MG945571 | Unassigned | Unassigned | Unassigned | 198_0 | Clustered | 28 | VC_198_0 | 28 | 0.2429 | 0.99971175 | 0.2429 | 0 | 0 | 0 | 1 | Ref |
| 6396 | MG945572 | Unassigned | Unassigned | Unassigned | 1276_0 | Clustered | 7 | VC_1276_0 | 7 | 0.8695 | 0.99982476 | 0.8693 | 0 | 0 | 0 | 1 | Ref |
| 6397 | MG945573 | Unassigned | Unassigned | Unassigned | 1143_0 | Clustered | 9 | VC_1143_0 | 9 | 0.7268 | 1 | 0.7268 | 0 | 0 | 0 | 1 | Ref |
| 6398 | MG945574 | Unassigned | Unassigned | Unassigned | 1302_0 | Clustered | 3 | VC_1302_0 | 3 | 1 | 1 | 1 | 0 | 0 | 0 | 1 | Ref |
| 6399 | MG945575 | Unassigned | Unassigned | Unassigned | 1319_0 | Clustered | 2 | VC_1319_0 | 2 | 1 | 1 | 1 | 0 | 0 | 0 | 1 | Ref |
| 6400 | MG945576 | Unassigned | Unassigned | Unassigned | 1294_0 | Clustered | 6 | VC_1294_0 | 6 | 0.6626 | 0.99999272 | 0.6626 | 0 | 0 | 0 | 1 | Ref |
| 6401 | MG945578 | Unassigned | Unassigned | Unassigned | 1253_0 | Clustered | 6 | VC_1253_0 | 6 | 0.6097 | 1 | 0.6097 | 0 | 0 | 0 | 1 | Ref |
| 6402 | MG945579 | Unassigned | Unassigned | Unassigned | 1285_0 | Clustered | 25 | VC_1285_0 | 25 | 1 | 1 | 1 | 0 | 0 | 0 | 1 | Ref |
| 6403 | MG945580 | Unassigned | Unassigned | Unassigned | 1258_0 | Clustered | 3 | VC_1258_0 | 3 | 0.2422 | 0.85925819 | 0.2081 | 0 | 0 | 0 | 1 | Ref |
| 6404 | MG945582 | Unassigned | Unassigned | Unassigned | 1268_0 | Clustered | 3 | VC_1268_0 | 3 | 0.0807 | 0.6862532 | 0.0554 | 0 | 0 | 0 | 1 | Ref |
| 6405 | MG945584 | Unassigned | Unassigned | Unassigned | 1275_0 | Clustered | 4 | VC_1275_0 | 4 | 1 | 1 | 1 | 0 | 0 | 0 | 1 | Ref |
| 6406 | MG945585 | Unassigned | Unassigned | Unassigned | 1277_0 | Clustered | 14 | VC_1277_0 | 14 | 0.8218 | 1 | 0.8218 | 0 | 0 | 0 | 1 | Ref |
| 6407 | MG945586 | Unassigned | Unassigned | Unassigned | 1104_0 | Clustered | 154 | VC_1104_0 | 154 | 0.8484 | 1 | 0.8484 | 0 | 0 | 0 | 1 | Ref |
| 6408 | MG945587 | Unassigned | Unassigned | Unassigned | 1104_0 | Clustered | 154 | VC_1104_0 | 154 | 0.8484 | 1 | 0.8484 | 0 | 0 | 0 | 1 | Ref |
| 6409 | MG945589 | Unassigned | Unassigned | Unassigned | 1104_0 | Clustered | 154 | VC_1104_0 | 154 | 0.8484 | 1 | 0.8484 | 0 | 0 | 0 | 1 | Ref |
| 6410 | MG945591 | Unassigned | Unassigned | Unassigned | 1113_0 | Clustered/Singleton | 1 | VC_1113_0 | 1 | 0 | 1 | 0 | 0 | 0 | 0 | 1 | Ref |
| 6411 | MG945594 | Unassigned | Unassigned | Unassigned | 1053_0 | Clustered/Singleton | 1 | VC_1053_0 | 1 | 0 | 1 | 0 | 0 | 0 | 0 | 1 | Ref |
| 6412 | MG945596 | Unassigned | Unassigned | Unassigned | 1104_0 | Clustered | 154 | VC_1104_0 | 154 | 0.8484 | 1 | 0.8484 | 0 | 0 | 0 | 1 | Ref |
| 6413 | MG945597 | Unassigned | Unassigned | Unassigned | 1105_0 | Clustered | 3 | VC_1105_0 | 3 | 0.0755 | 0.99466875 | 0.0751 | 0 | 0 | 0 | 1 | Ref |
| 6414 | MG945602 | Unassigned | Unassigned | Unassigned | 1104_0 | Clustered | 154 | VC_1104_0 | 154 | 0.8484 | 1 | 0.8484 | 0 | 0 | 0 | 1 | Ref |
| 6415 | MG945605 | Unassigned | Unassigned | Unassigned | 1281_0 | Clustered | 7 | VC_1281_0 | 7 | 0.9212 | 0.99995294 | 0.9212 | 0 | 0 | 0 | 1 | Ref |
| 6416 | MG945609 | Unassigned | Unassigned | Unassigned | 1148_0 | Clustered | 16 | VC_1148_0 | 16 | 0.8821 | 1 | 0.8821 | 0 | 0 | 0 | 1 | Ref |
| 6417 | MG945611 | Unassigned | Unassigned | Unassigned | 1253_0 | Clustered | 6 | VC_1253_0 | 6 | 0.6097 | 1 | 0.6097 | 0 | 0 | 0 | 1 | Ref |
| 6418 | MG945612 | Unassigned | Unassigned | Unassigned | 1285_0 | Clustered | 25 | VC_1285_0 | 25 | 1 | 1 | 1 | 0 | 0 | 0 | 1 | Ref |
| 6419 | MG945613 | Unassigned | Unassigned | Unassigned | 1119_0 | Clustered | 20 | VC_1119_0 | 20 | 0.6751 | 0.99997892 | 0.6751 | 0 | 0 | 0 | 1 | Ref |
| 6420 | MG945614 | Unassigned | Unassigned | Unassigned | 1295_0 | Clustered | 3 | VC_1295_0 | 3 | 1 | 0 | 0 | 0 | 0 | 0 | 1 | Ref |
| 6421 | MG945615 | Unassigned | Unassigned | Unassigned | 1322_0 | Clustered | 2 | VC_1322_0 | 2 | 1 | 1 | 1 | 0 | 0 | 0 | 1 | Ref |
| 6422 | MG945616 | Unassigned | Unassigned | Unassigned | 1197_0 | Clustered | 15 | VC_1197_0 | 15 | 0.8991 | 0.76608394 | 0.6888 | 0 | 0 | 0 | 1 | Ref |
| 6423 | MG945617 | Unassigned | Unassigned | Unassigned | 1310_0 | Clustered | 2 | VC_1310_0 | 2 | 1 | 1 | 1 | 0 | 0 | 0 | 1 | Ref |
| 6424 | MG945618 | Unassigned | Unassigned | Unassigned | 1243_0 | Clustered | 52 | VC_1243_0 | 52 | 0.9126 | 0.92708311 | 0.8461 | 0 | 0 | 0 | 1 | Ref |
| 6425 | MG945619 | Unassigned | Unassigned | Unassigned | 1324_0 | Clustered | 4 | VC_1324_0 | 4 | 0.6008 | 0.54407361 | 0.3269 | 0 | 0 | 0 | 1 | Ref |
| 6426 | MG945620 | Unassigned | Unassigned | Unassigned | 1137_0 | Clustered | 3 | VC_1137_0 | 3 | 0.9176 | 0.93067968 | 0.854 | 0 | 0 | 0 | 1 | Ref |
| 6427 | MG945621 | Unassigned | Unassigned | Unassigned | 1308_0 | Clustered | 12 | VC_1308_0 | 12 | 0.8184 | 0.9810731 | 0.8029 | 0 | 0 | 0 | 1 | Ref |
| 6428 | MG945622 | Unassigned | Unassigned | Unassigned | 1255_0 | Clustered | 4 | VC_1255_0 | 4 | 0.248 | 0.99828153 | 0.2476 | 0 | 0 | 0 | 1 | Ref |
| 6429 | MG945623 | Unassigned | Unassigned | Unassigned | 1194_0 | Clustered | 5 | VC_1194_0 | 5 | 0.5236 | 0.99999899 | 0.5236 | 0 | 0 | 0 | 1 | Ref |
| 6430 | MG945625 | Unassigned | Unassigned | Unassigned | 1250_0 | Clustered | 5 | VC_1250_0 | 5 | 0.6384 | 0.99918945 | 0.6379 | 0 | 0 | 0 | 1 | Ref |
| 6431 | MG945628 | Unassigned | Unassigned | Unassigned | 1303_0 | Clustered | 6 | VC_1303_0 | 6 | 1 | 1 | 1 | 0 | 0 | 0 | 1 | Ref |
| 6432 | MG945629 | Unassigned | Unassigned | Unassigned | 1250_0 | Clustered | 5 | VC_1250_0 | 5 | 0.6384 | 0.99918945 | 0.6379 | 0 | 0 | 0 | 1 | Ref |
| 6433 | MG945631 | Unassigned | Unassigned | Unassigned | 1274_0 | Clustered | 2 | VC_1274_0 | 2 | 0.6386 | 0.72985431 | 0.4661 | 0 | 0 | 0 | 1 | Ref |
| 6434 | MG945632 | Unassigned | Unassigned | Unassigned | 1250_0 | Clustered | 5 | VC_1250_0 | 5 | 0.6384 | 0.99918945 | 0.6379 | 0 | 0 | 0 | 1 | Ref |
| 6435 | MG945633 | Unassigned | Unassigned | Unassigned | 1250_0 | Clustered | 5 | VC_1250_0 | 5 | 0.6384 | 0.99918945 | 0.6379 | 0 | 0 | 0 | 1 | Ref |
| 6436 | MG945634 | Unassigned | Unassigned | Unassigned | 1243_0 | Clustered | 52 | VC_1243_0 | 52 | 0.9126 | 0.92708311 | 0.8461 | 0 | 0 | 0 | 1 | Ref |
| 6437 | MG945635 | Unassigned | Unassigned | Unassigned | 1125_0 | Clustered | 20 | VC_1125_0 | 20 | 0.5599 | 1 | 0.5599 | 0 | 0 | 0 | 1 | Ref |
| 6438 | MG945637 | Unassigned | Unassigned | Unassigned | 1294_0 | Clustered | 6 | VC_1294_0 | 6 | 0.6626 | 0.99999272 | 0.6626 | 0 | 0 | 0 | 1 | Ref |
| 6439 | MG945639 | Unassigned | Unassigned | Unassigned | 1125_0 | Clustered | 20 | VC_1125_0 | 20 | 0.5599 | 1 | 0.5599 | 0 | 0 | 0 | 1 | Ref |
| 6440 | MG945640 | Unassigned | Unassigned | Unassigned | 1285_0 | Clustered | 25 | VC_1285_0 | 25 | 1 | 1 | 1 | 0 | 0 | 0 | 1 | Ref |
| 6441 | MG945641 | Unassigned | Unassigned | Unassigned | 1243_0 | Clustered | 52 | VC_1243_0 | 52 | 0.9126 | 0.92708311 | 0.8461 | 0 | 0 | 0 | 1 | Ref |
| 6442 | MG945642 | Unassigned | Unassigned | Unassigned | 1127_0 | Clustered | 19 | VC_1127_0 | 19 | 0.7611 | 1 | 0.7611 | 0 | 0 | 0 | 1 | Ref |
| 6443 | MG945643 | Unassigned | Unassigned | Unassigned | 1256_0 | Clustered | 4 | VC_1256_0 | 4 | 0.581 | 0.94751743 | 0.5505 | 0 | 0 | 0 | 1 | Ref |
| 6444 | MG945644 | Unassigned | Unassigned | Unassigned | 1140_0 | Clustered | 4 | VC_1140_0 | 4 | 0.1325 | 0.66207199 | 0.0877 | 0 | 0 | 0 | 1 | Ref |
| 6445 | MG945645 | Unassigned | Unassigned | Unassigned | 1116_0 | Clustered | 14 | VC_1116_0 | 14 | 0.7927 | 1 | 0.7927 | 0 | 0 | 0 | 1 | Ref |
| 6446 | MG945646 | Unassigned | Unassigned | Unassigned | 1104_0 | Clustered | 154 | VC_1104_0 | 154 | 0.8484 | 1 | 0.8484 | 0 | 0 | 0 | 1 | Ref |
| 6447 | MG945647 | Unassigned | Unassigned | Unassigned | 1243_0 | Clustered | 52 | VC_1243_0 | 52 | 0.9126 | 0.92708311 | 0.8461 | 0 | 0 | 0 | 1 | Ref |
| 6448 | MG945662 | Unassigned | Unassigned | Unassigned | 1257_0 | Clustered | 6 | VC_1257_0 | 6 | 0.6517 | 0.99999969 | 0.6517 | 0 | 0 | 0 | 1 | Ref |
| 6449 | MG945663 | Unassigned | Unassigned | Unassigned | 995_0 | Clustered | 69 | VC_995_0 | 69 | 0.8547 | 1 | 0.8547 | 0 | 0 | 0 | 1 | Ref |
| 6450 | MG945664 | Unassigned | Unassigned | Unassigned | 1211_0 | Clustered | 25 | VC_1211_0 | 25 | 1 | 1 | 1 | 0 | 0 | 0 | 1 | Ref |
| 6451 | MG945665 | Unassigned | Unassigned | Unassigned | 1287_0 | Clustered | 7 | VC_1287_0 | 7 | 1 | 1 | 1 | 0 | 0 | 0 | 1 | Ref |
| 6452 | MG945667 | Unassigned | Unassigned | Unassigned | 1232_0 | Clustered | 2 | VC_1232_0 | 2 | 0.4709 | 0.85557782 | 0.4029 | 0 | 0 | 0 | 1 | Ref |
| 6453 | MG945668 | Unassigned | Unassigned | Unassigned | 1285_0 | Clustered | 25 | VC_1285_0 | 25 | 1 | 1 | 1 | 0 | 0 | 0 | 1 | Ref |
| 6454 | MG945669 | Unassigned | Unassigned | Unassigned | 198_0 | Clustered | 28 | VC_198_0 | 28 | 0.2429 | 0.99971175 | 0.2429 | 0 | 0 | 0 | 1 | Ref |
| 6455 | MG945670 | Unassigned | Unassigned | Unassigned | 1077_0 | Clustered | 48 | VC_1077_0 | 48 | 0.8525 | 1 | 0.8525 | 0 | 0 | 0 | 1 | Ref |
| 6456 | MG945671 | Unassigned | Unassigned | Unassigned | 1104_0 | Clustered | 154 | VC_1104_0 | 154 | 0.8484 | 1 | 0.8484 | 0 | 0 | 0 | 1 | Ref |
| 6457 | MG945673 | Unassigned | Unassigned | Unassigned | 1104_0 | Clustered | 154 | VC_1104_0 | 154 | 0.8484 | 1 | 0.8484 | 0 | 0 | 0 | 1 | Ref |
| 6458 | MG945674 | Unassigned | Unassigned | Unassigned | 1104_0 | Clustered | 154 | VC_1104_0 | 154 | 0.8484 | 1 | 0.8484 | 0 | 0 | 0 | 1 | Ref |
| 6459 | MG945676 | Unassigned | Unassigned | Unassigned | 1148_0 | Clustered | 16 | VC_1148_0 | 16 | 0.8821 | 1 | 0.8821 | 0 | 0 | 0 | 1 | Ref |
| 6460 | MG945677 | Unassigned | Unassigned | Unassigned | 1230_0 | Clustered | 3 | VC_1230_0 | 3 | 0.4802 | 0.82711071 | 0.3971 | 0 | 0 | 0 | 1 | Ref |
| 6461 | MG945678 | Unassigned | Unassigned | Unassigned | 1155_0 | Clustered | 2 | VC_1155_0 | 2 | 1 | 1 | 1 | 0 | 0 | 0 | 1 | Ref |
| 6462 | MG945680 | Unassigned | Unassigned | Unassigned | 1211_0 | Clustered | 25 | VC_1211_0 | 25 | 1 | 1 | 1 | 0 | 0 | 0 | 1 | Ref |
| 6463 | MG945681 | Unassigned | Unassigned | Unassigned | 1104_0 | Clustered | 154 | VC_1104_0 | 154 | 0.8484 | 1 | 0.8484 | 0 | 0 | 0 | 1 | Ref |
| 6464 | MG945682 | Unassigned | Unassigned | Unassigned | 398_0 | Clustered | 48 | VC_398_0 | 48 | 0.4813 | 1 | 0.4813 | 0 | 0 | 0 | 1 | Ref |
| 6465 | MG945683 | Unassigned | Unassigned | Unassigned | 1211_0 | Clustered | 25 | VC_1211_0 | 25 | 1 | 1 | 1 | 0 | 0 | 0 | 1 | Ref |
| 6466 | MG945685 | Unassigned | Unassigned | Unassigned | 1251_0 | Clustered | 15 | VC_1251_0 | 15 | 0.9082 | 1 | 0.9082 | 0 | 0 | 0 | 1 | Ref |
| 6467 | MG945686 | Unassigned | Unassigned | Unassigned | 672_0 | Clustered | 12 | VC_672_0 | 12 | 1 | 1 | 1 | 0 | 0 | 0 | 1 | Ref |
| 6468 | MG945687 | Unassigned | Unassigned | Unassigned | 1259_0 | Clustered | 8 | VC_1259_0 | 8 | 0.8225 | 1 | 0.8225 | 0 | 0 | 0 | 1 | Ref |
| 6469 | MG945689 | Unassigned | Unassigned | Unassigned | 1122_0 | Clustered | 10 | VC_1122_0 | 10 | 0.9643 | 0.99999697 | 0.9643 | 0 | 0 | 0 | 1 | Ref |
| 6470 | MG945690 | Unassigned | Unassigned | Unassigned | 1326_0 | Clustered | 2 | VC_1326_0 | 2 | 1 | 1 | 1 | 0 | 0 | 0 | 1 | Ref |
| 6471 | MG945691 | Unassigned | Unassigned | Unassigned | 1104_0 | Clustered | 154 | VC_1104_0 | 154 | 0.8484 | 1 | 0.8484 | 0 | 0 | 0 | 1 | Ref |
| 6472 | MG945692 | Unassigned | Unassigned | Unassigned | 1247_0 | Clustered | 7 | VC_1247_0 | 7 | 0.9086 | 0.99998959 | 0.9086 | 0 | 0 | 0 | 1 | Ref |
| 6473 | MG945693 | Unassigned | Unassigned | Unassigned | 1138_0 | Clustered | 4 | VC_1138_0 | 4 | 0.9437 | 0.98964986 | 0.934 | 0 | 0 | 0 | 1 | Ref |
| 6474 | MG945694 | Unassigned | Unassigned | Unassigned | 1127_0 | Clustered | 19 | VC_1127_0 | 19 | 0.7611 | 1 | 0.7611 | 0 | 0 | 0 | 1 | Ref |
| 6475 | MG945696 | Unassigned | Unassigned | Unassigned | 1297_0 | Clustered | 7 | VC_1297_0 | 7 | 1 | 1 | 1 | 0 | 0 | 0 | 1 | Ref |
| 6476 | MG945697 | Unassigned | Unassigned | Unassigned | 1247_0 | Clustered | 7 | VC_1247_0 | 7 | 0.9086 | 0.99998959 | 0.9086 | 0 | 0 | 0 | 1 | Ref |
| 6477 | MG945698 | Unassigned | Unassigned | Unassigned | 1247_0 | Clustered | 7 | VC_1247_0 | 7 | 0.9086 | 0.99998959 | 0.9086 | 0 | 0 | 0 | 1 | Ref |
| 6478 | MG945699 | Unassigned | Unassigned | Unassigned | 1104_0 | Clustered | 154 | VC_1104_0 | 154 | 0.8484 | 1 | 0.8484 | 0 | 0 | 0 | 1 | Ref |

| | | | | | | | | | | | | | | | | | |
|---|---|---|---|---|---|---|---|---|---|---|---|---|---|---|---|---|---|
| 6479 | MG945702 | Unassigned | Unassigned | Unassigned | 1292_0 | Clustered | 3 | VC_1292_0 | 3 | 1 | 1 | 1 | 0 | 0 | 0 | 1 | Ref |
| 6480 | MG945703 | Unassigned | Unassigned | Unassigned | 1054_0 | Clustered | 12 | VC_1054_0 | 12 | 0.2863 | 0.99746644 | 0.2855 | 0 | 0 | 0 | 1 | Ref |
| 6481 | MG945704 | Unassigned | Unassigned | Unassigned | 1120_0 | Clustered | 7 | VC_1120_0 | 7 | 0.2688 | 0.9951868 | 0.2675 | 0 | 0 | 0 | 1 | Ref |
| 6482 | MG945705 | Unassigned | Unassigned | Unassigned | 1104_0 | Clustered | 154 | VC_1104_0 | 154 | 0.8484 | 1 | 0.8484 | 0 | 0 | 0 | 1 | Ref |
| 6483 | MG945706 | Unassigned | Unassigned | Unassigned | 1148_0 | Clustered | 16 | VC_1148_0 | 16 | 0.8821 | 1 | 0.8821 | 0 | 0 | 0 | 1 | Ref |
| 6484 | MG945708 | Unassigned | Unassigned | Unassigned | 1104_0 | Clustered | 154 | VC_1104_0 | 154 | 0.8484 | 1 | 0.8484 | 0 | 0 | 0 | 1 | Ref |
| 6485 | MG945710 | Unassigned | Unassigned | Unassigned | 1104_0 | Clustered | 154 | VC_1104_0 | 154 | 0.8484 | 1 | 0.8484 | 0 | 0 | 0 | 1 | Ref |
| 6486 | MG945712 | Unassigned | Unassigned | Unassigned | 1285_0 | Clustered | 25 | VC_1285_0 | 25 | 1 | 1 | 1 | 0 | 0 | 0 | 1 | Ref |
| 6487 | MG945713 | Unassigned | Unassigned | Unassigned | 1211_0 | Clustered | 25 | VC_1211_0 | 25 | 1 | 1 | 1 | 0 | 0 | 0 | 1 | Ref |
| 6488 | MG945716 | Unassigned | Unassigned | Unassigned | 1211_0 | Clustered | 25 | VC_1211_0 | 25 | 1 | 1 | 1 | 0 | 0 | 0 | 1 | Ref |
| 6489 | MG945717 | Unassigned | Unassigned | Unassigned | 1311_0 | Clustered | 5 | VC_1311_0 | 5 | 0.8316 | 0.99879053 | 0.8306 | 0 | 0 | 0 | 1 | Ref |
| 6490 | MG945718 | Unassigned | Unassigned | Unassigned | 1110_0 | Clustered | 8 | VC_1110_0 | 8 | 0.6915 | 1 | 0.6915 | 0 | 0 | 0 | 1 | Ref |
| 6491 | MG945719 | Unassigned | Unassigned | Unassigned | 1144_0 | Clustered | 7 | VC_1144_0 | 7 | 0.7076 | 1 | 0.7076 | 0 | 0 | 0 | 1 | Ref |
| 6492 | MG945721 | Unassigned | Unassigned | Unassigned | 1139_0 | Clustered | 4 | VC_1139_0 | 4 | 1 | 1 | 1 | 0 | 0 | 0 | 1 | Ref |
| 6493 | MG945723 | Unassigned | Unassigned | Unassigned | 1252_0 | Clustered | 3 | VC_1252_0 | 3 | 0.3237 | 0.99790393 | 0.323 | 0 | 0 | 0 | 1 | Ref |
| 6494 | MG945724 | Unassigned | Unassigned | Unassigned | 1327_0 | Clustered | 4 | VC_1327_0 | 4 | 1 | 1 | 1 | 0 | 0 | 0 | 1 | Ref |
| 6495 | MG945725 | Unassigned | Unassigned | Unassigned | 1233_0 | Clustered | 16 | VC_1233_0 | 16 | 0.8971 | 1 | 0.8971 | 0 | 0 | 0 | 1 | Ref |
| 6496 | MG945726 | Unassigned | Unassigned | Unassigned | 1151_0 | Clustered | 9 | VC_1151_0 | 9 | 0.8548 | 1 | 0.8548 | 0 | 0 | 0 | 1 | Ref |
| 6497 | MG945727 | Unassigned | Unassigned | Unassigned | 1152_0 | Clustered | 4 | VC_1152_0 | 4 | 0.7206 | 0.99971165 | 0.7204 | 0 | 0 | 0 | 1 | Ref |
| 6498 | MG945728 | Unassigned | Unassigned | Unassigned | 1104_0 | Clustered | 154 | VC_1104_0 | 154 | 0.8484 | 1 | 0.8484 | 0 | 0 | 0 | 1 | Ref |
| 6499 | MG945729 | Unassigned | Unassigned | Unassigned | 1287_0 | Clustered | 7 | VC_1287_0 | 7 | 1 | 1 | 1 | 0 | 0 | 0 | 1 | Ref |
| 6500 | MG945730 | Unassigned | Unassigned | Unassigned | 1211_0 | Clustered | 25 | VC_1211_0 | 25 | 1 | 1 | 1 | 0 | 0 | 0 | 1 | Ref |
| 6501 | MG945731 | Unassigned | Unassigned | Unassigned | 1104_0 | Clustered | 154 | VC_1104_0 | 154 | 0.8484 | 1 | 0.8484 | 0 | 0 | 0 | 1 | Ref |
| 6502 | MG945733 | Unassigned | Unassigned | Unassigned | 1285_0 | Clustered | 25 | VC_1285_0 | 25 | 1 | 1 | 1 | 0 | 0 | 0 | 1 | Ref |
| 6503 | MG945734 | Unassigned | Unassigned | Unassigned | 1247_0 | Clustered | 7 | VC_1247_0 | 7 | 0.9086 | 0.99998959 | 0.9086 | 0 | 0 | 0 | 1 | Ref |
| 6504 | MG945735 | Unassigned | Unassigned | Unassigned | 1193_0 | Clustered | 3 | VC_1193_0 | 3 | 0.5473 | 0.99523779 | 0.5447 | 0 | 0 | 0 | 1 | Ref |
| 6505 | MG945736 | Unassigned | Unassigned | Unassigned | 1116_0 | Clustered | 14 | VC_1116_0 | 14 | 0.7927 | 1 | 0.7927 | 0 | 0 | 0 | 1 | Ref |
| 6506 | MG945738 | Unassigned | Unassigned | Unassigned | 1132_0 | Clustered | 2 | VC_1132_0 | 2 | 0.0978 | 0.83194774 | 0.0814 | 0 | 0 | 0 | 1 | Ref |
| 6507 | MG945739 | Unassigned | Unassigned | Unassigned | 1311_0 | Clustered | 5 | VC_1311_0 | 5 | 0.8316 | 0.99879053 | 0.8306 | 0 | 0 | 0 | 1 | Ref |
| 6508 | MG945740 | Unassigned | Unassigned | Unassigned | 1116_0 | Clustered | 14 | VC_1116_0 | 14 | 0.7927 | 1 | 0.7927 | 0 | 0 | 0 | 1 | Ref |
| 6509 | MG945741 | Unassigned | Unassigned | Unassigned | 1137_0 | Clustered | 3 | VC_1137_0 | 3 | 0.9176 | 0.93067968 | 0.854 | 0 | 0 | 0 | 1 | Ref |
| 6510 | MG945742 | Unassigned | Unassigned | Unassigned | 1114_0 | Clustered | 2 | VC_1114_0 | 2 | 0.0522 | 0.94760452 | 0.0495 | 0 | 0 | 0 | 1 | Ref |
| 6511 | MG945743 | Unassigned | Unassigned | Unassigned | 1243_0 | Clustered | 52 | VC_1243_0 | 52 | 0.9126 | 0.92708311 | 0.8461 | 0 | 0 | 0 | 1 | Ref |
| 6512 | MG945744 | Unassigned | Unassigned | Unassigned | 1104_0 | Clustered | 154 | VC_1104_0 | 154 | 0.8484 | 1 | 0.8484 | 0 | 0 | 0 | 1 | Ref |
| 6513 | MG945745 | Unassigned | Unassigned | Unassigned | 1251_0 | Clustered | 15 | VC_1251_0 | 15 | 0.9082 | 1 | 0.9082 | 0 | 0 | 0 | 1 | Ref |
| 6514 | MG945746 | Unassigned | Unassigned | Unassigned | 1125_0 | Clustered | 20 | VC_1125_0 | 20 | 0.5599 | 1 | 0.5599 | 0 | 0 | 0 | 1 | Ref |
| 6515 | MG945747 | Unassigned | Unassigned | Unassigned | 1154_0 | Clustered | 5 | VC_1154_0 | 5 | 1 | 1 | 1 | 0 | 0 | 0 | 1 | Ref |
| 6516 | MG945748 | Unassigned | Unassigned | Unassigned | 1196_0 | Clustered | 3 | VC_1196_0 | 3 | 0.3378 | 0.99743975 | 0.3369 | 0 | 0 | 0 | 1 | Ref |
| 6517 | MG945750 | Unassigned | Unassigned | Unassigned | 1267_0 | Clustered | 7 | VC_1267_0 | 7 | 1 | 1 | 1 | 0 | 0 | 0 | 1 | Ref |
| 6518 | MG945751 | Unassigned | Unassigned | Unassigned | 1281_0 | Clustered | 7 | VC_1281_0 | 7 | 0.9212 | 0.99995294 | 0.9212 | 0 | 0 | 0 | 1 | Ref |
| 6519 | MG945752 | Unassigned | Unassigned | Unassigned | 1193_0 | Clustered | 3 | VC_1193_0 | 3 | 0.5473 | 0.99523779 | 0.5447 | 0 | 0 | 0 | 1 | Ref |
| 6520 | MG945753 | Unassigned | Unassigned | Unassigned | 1127_0 | Clustered | 19 | VC_1127_0 | 19 | 0.7611 | 1 | 0.7611 | 0 | 0 | 0 | 1 | Ref |
| 6521 | MG945754 | Unassigned | Unassigned | Unassigned | 1151_0 | Clustered | 9 | VC_1151_0 | 9 | 0.8548 | 1 | 0.8548 | 0 | 0 | 0 | 1 | Ref |
| 6522 | MG945755 | Unassigned | Unassigned | Unassigned | 1293_0 | Clustered | 4 | VC_1293_0 | 4 | 0.7847 | 0.99289298 | 0.7791 | 0 | 0 | 0 | 1 | Ref |
| 6523 | MG945756 | Unassigned | Unassigned | Unassigned | 672_0 | Clustered | 12 | VC_672_0 | 12 | 1 | 1 | 1 | 0 | 0 | 0 | 1 | Ref |
| 6524 | MG945757 | Unassigned | Unassigned | Unassigned | 1233_0 | Clustered | 16 | VC_1233_0 | 16 | 0.8971 | 1 | 0.8971 | 0 | 0 | 0 | 1 | Ref |
| 6525 | MG945758 | Unassigned | Unassigned | Unassigned | 1104_0 | Clustered | 154 | VC_1104_0 | 154 | 0.8484 | 1 | 0.8484 | 0 | 0 | 0 | 1 | Ref |
| 6526 | MG945759 | Unassigned | Unassigned | Unassigned | 1143_0 | Clustered | 9 | VC_1143_0 | 9 | 0.7268 | 1 | 0.7268 | 0 | 0 | 0 | 1 | Ref |
| 6527 | MG945760 | Unassigned | Unassigned | Unassigned | 1143_0 | Clustered | 9 | VC_1143_0 | 9 | 0.7268 | 1 | 0.7268 | 0 | 0 | 0 | 1 | Ref |
| 6528 | MG945761 | Unassigned | Unassigned | Unassigned | 1104_0 | Clustered | 154 | VC_1104_0 | 154 | 0.8484 | 1 | 0.8484 | 0 | 0 | 0 | 1 | Ref |
| 6529 | MG945764 | Unassigned | Unassigned | Unassigned | 1148_0 | Clustered | 16 | VC_1148_0 | 16 | 0.8821 | 1 | 0.8821 | 0 | 0 | 0 | 1 | Ref |
| 6530 | MG945765 | Unassigned | Unassigned | Unassigned | 1104_0 | Clustered | 154 | VC_1104_0 | 154 | 0.8484 | 1 | 0.8484 | 0 | 0 | 0 | 1 | Ref |
| 6531 | MG945766 | Unassigned | Unassigned | Unassigned | 1328_0 | Clustered | 2 | VC_1328_0 | 2 | 1 | 1 | 1 | 0 | 0 | 0 | 1 | Ref |
| 6532 | MG945767 | Unassigned | Unassigned | Unassigned | 1125_0 | Clustered | 20 | VC_1125_0 | 20 | 0.5599 | 1 | 0.5599 | 0 | 0 | 0 | 1 | Ref |
| 6533 | MG945768 | Unassigned | Unassigned | Unassigned | 1104_0 | Clustered | 154 | VC_1104_0 | 154 | 0.8484 | 1 | 0.8484 | 0 | 0 | 0 | 1 | Ref |
| 6534 | MG945769 | Unassigned | Unassigned | Unassigned | 1329_0 | Clustered | 2 | VC_1329_0 | 2 | 1 | 1 | 1 | 0 | 0 | 0 | 1 | Ref |
| 6535 | MG945770 | Unassigned | Unassigned | Unassigned | 1104_0 | Clustered | 154 | VC_1104_0 | 154 | 0.8484 | 1 | 0.8484 | 0 | 0 | 0 | 1 | Ref |
| 6536 | MG945771 | Unassigned | Unassigned | Unassigned | 1116_0 | Clustered | 14 | VC_1116_0 | 14 | 0.7927 | 1 | 0.7927 | 0 | 0 | 0 | 1 | Ref |
| 6537 | MG945774 | Unassigned | Unassigned | Unassigned | 1251_0 | Clustered | 15 | VC_1251_0 | 15 | 0.9082 | 1 | 0.9082 | 0 | 0 | 0 | 1 | Ref |
| 6538 | MG945775 | Unassigned | Unassigned | Unassigned | 1108_0 | Clustered | 4 | VC_1108_0 | 4 | 0.0485 | 0.99999021 | 0.0485 | 0 | 0 | 0 | 1 | Ref |
| 6539 | MG945776 | Unassigned | Unassigned | Unassigned | 1267_0 | Clustered | 7 | VC_1267_0 | 7 | 1 | 1 | 1 | 0 | 0 | 0 | 1 | Ref |
| 6540 | MG945778 | Unassigned | Unassigned | Unassigned | 1288_0 | Clustered | 3 | VC_1288_0 | 3 | 1 | 1 | 1 | 0 | 0 | 0 | 1 | Ref |
| 6541 | MG945780 | Unassigned | Unassigned | Unassigned | 1126_0 | Clustered | 8 | VC_1126_0 | 8 | 0.2864 | 1 | 0.2864 | 0 | 0 | 0 | 1 | Ref |
| 6542 | MG945781 | Unassigned | Unassigned | Unassigned | 1127_0 | Clustered | 19 | VC_1127_0 | 19 | 0.7611 | 1 | 0.7611 | 0 | 0 | 0 | 1 | Ref |
| 6543 | MG945782 | Unassigned | Unassigned | Unassigned | 1287_0 | Clustered | 7 | VC_1287_0 | 7 | 1 | 1 | 1 | 0 | 0 | 0 | 1 | Ref |
| 6544 | MG945783 | Unassigned | Unassigned | Unassigned | 1110_0 | Clustered | 8 | VC_1110_0 | 8 | 0.6915 | 1 | 0.6915 | 0 | 0 | 0 | 1 | Ref |
| 6545 | MG945784 | Unassigned | Unassigned | Unassigned | 1211_0 | Clustered | 25 | VC_1211_0 | 25 | 1 | 1 | 1 | 0 | 0 | 0 | 1 | Ref |
| 6546 | MG945785 | Unassigned | Unassigned | Unassigned | 1152_0 | Clustered | 4 | VC_1152_0 | 4 | 0.7206 | 0.99971165 | 0.7204 | 0 | 0 | 0 | 1 | Ref |
| 6547 | MG945786 | Unassigned | Unassigned | Unassigned | 1104_0 | Clustered | 154 | VC_1104_0 | 154 | 0.8484 | 1 | 0.8484 | 0 | 0 | 0 | 1 | Ref |
| 6548 | MG945787 | Unassigned | Unassigned | Unassigned | 1104_0 | Clustered | 154 | VC_1104_0 | 154 | 0.8484 | 1 | 0.8484 | 0 | 0 | 0 | 1 | Ref |
| 6549 | MG945788 | Unassigned | Unassigned | Unassigned | 1330_0 | Clustered | 2 | VC_1330_0 | 2 | 1 | 1 | 1 | 0 | 0 | 0 | 1 | Ref |
| 6550 | MG945789 | Unassigned | Unassigned | Unassigned | 1278_0 | Clustered | 3 | VC_1278_0 | 3 | 0.3961 | 0.99705286 | 0.395 | 0 | 0 | 0 | 1 | Ref |
| 6551 | MG945790 | Unassigned | Unassigned | Unassigned | 1211_0 | Clustered | 25 | VC_1211_0 | 25 | 1 | 1 | 1 | 0 | 0 | 0 | 1 | Ref |
| 6552 | MG945791 | Unassigned | Unassigned | Unassigned | 1267_0 | Clustered | 7 | VC_1267_0 | 7 | 1 | 1 | 1 | 0 | 0 | 0 | 1 | Ref |
| 6553 | MG945792 | Unassigned | Unassigned | Unassigned | 1329_0 | Clustered | 2 | VC_1329_0 | 2 | 1 | 1 | 1 | 0 | 0 | 0 | 1 | Ref |
| 6554 | MG945793 | Unassigned | Unassigned | Unassigned | 1104_0 | Clustered | 154 | VC_1104_0 | 154 | 0.8484 | 1 | 0.8484 | 0 | 0 | 0 | 1 | Ref |
| 6555 | MG945794 | Unassigned | Unassigned | Unassigned | 1197_0 | Clustered | 15 | VC_1197_0 | 15 | 0.8991 | 0.76608394 | 0.6888 | 0 | 0 | 0 | 1 | Ref |
| 6556 | MG945795 | Unassigned | Unassigned | Unassigned | 1330_0 | Clustered | 2 | VC_1330_0 | 2 | 1 | 1 | 1 | 0 | 0 | 0 | 1 | Ref |
| 6557 | MG945796 | Unassigned | Unassigned | Unassigned | 1104_0 | Clustered | 154 | VC_1104_0 | 154 | 0.8484 | 1 | 0.8484 | 0 | 0 | 0 | 1 | Ref |
| 6558 | MG945797 | Unassigned | Unassigned | Unassigned | 1122_0 | Clustered | 10 | VC_1122_0 | 10 | 0.9643 | 0.99999697 | 0.9643 | 0 | 0 | 0 | 1 | Ref |
| 6559 | MG945798 | Unassigned | Unassigned | Unassigned | 1331_0 | Clustered | 4 | VC_1331_0 | 4 | 0.7343 | 0.99899459 | 0.7335 | 0 | 0 | 0 | 1 | Ref |
| 6560 | MG945799 | Unassigned | Unassigned | Unassigned | 1196_0 | Clustered | 3 | VC_1196_0 | 3 | 0.3378 | 0.99743975 | 0.3369 | 0 | 0 | 0 | 1 | Ref |
| 6561 | MG945800 | Unassigned | Unassigned | Unassigned | 1196_0 | Clustered | 3 | VC_1196_0 | 3 | 0.3378 | 0.99743975 | 0.3369 | 0 | 0 | 0 | 1 | Ref |
| 6562 | MG945801 | Unassigned | Unassigned | Unassigned | 1104_0 | Clustered | 154 | VC_1104_0 | 154 | 0.8484 | 1 | 0.8484 | 0 | 0 | 0 | 1 | Ref |
| 6563 | MG945802 | Unassigned | Unassigned | Unassigned | 1104_0 | Clustered | 154 | VC_1104_0 | 154 | 0.8484 | 1 | 0.8484 | 0 | 0 | 0 | 1 | Ref |
| 6564 | MG945803 | Unassigned | Unassigned | Unassigned | 1251_0 | Clustered | 15 | VC_1251_0 | 15 | 0.9082 | 1 | 0.9082 | 0 | 0 | 0 | 1 | Ref |
| 6565 | MG945804 | Unassigned | Unassigned | Unassigned | 1258_0 | Clustered | 3 | VC_1258_0 | 3 | 0.2422 | 0.85925819 | 0.2081 | 0 | 0 | 0 | 1 | Ref |
| 6566 | MG945805 | Unassigned | Unassigned | Unassigned | 1144_0 | Clustered | 7 | VC_1144_0 | 7 | 0.7076 | 1 | 0.7076 | 0 | 0 | 0 | 1 | Ref |
| 6567 | MG945806 | Unassigned | Unassigned | Unassigned | 1285_0 | Clustered | 25 | VC_1285_0 | 25 | 1 | 1 | 1 | 0 | 0 | 0 | 1 | Ref |
| 6568 | MG945807 | Unassigned | Unassigned | Unassigned | 1117_0 | Clustered | 5 | VC_1117_0 | 5 | 0.4688 | 0.99999974 | 0.4688 | 0 | 0 | 0 | 1 | Ref |
| 6569 | MG945808 | Unassigned | Unassigned | Unassigned | 1333_0 | Clustered | 2 | VC_1333_0 | 2 | 1 | 1 | 1 | 0 | 0 | 0 | 1 | Ref |
| 6570 | MG945809 | Unassigned | Unassigned | Unassigned | 1138_0 | Clustered | 4 | VC_1138_0 | 4 | 0.9437 | 0.98964986 | 0.934 | 0 | 0 | 0 | 1 | Ref |
| 6571 | MG945810 | Unassigned | Unassigned | Unassigned | 1152_0 | Clustered | 4 | VC_1152_0 | 4 | 0.7206 | 0.99971165 | 0.7204 | 0 | 0 | 0 | 1 | Ref |
| 6572 | MG945811 | Unassigned | Unassigned | Unassigned | 1143_0 | Clustered | 9 | VC_1143_0 | 9 | 0.7268 | 1 | 0.7268 | 0 | 0 | 0 | 1 | Ref |
| 6573 | MG945812 | Unassigned | Unassigned | Unassigned | 1281_0 | Clustered | 7 | VC_1281_0 | 7 | 0.9212 | 0.99995294 | 0.9212 | 0 | 0 | 0 | 1 | Ref |
| 6574 | MG945813 | Unassigned | Unassigned | Unassigned | 1211_0 | Clustered | 25 | VC_1211_0 | 25 | 1 | 1 | 1 | 0 | 0 | 0 | 1 | Ref |
| 6575 | MG945814 | Unassigned | Unassigned | Unassigned | 1285_0 | Clustered | 25 | VC_1285_0 | 25 | 1 | 1 | 1 | 0 | 0 | 0 | 1 | Ref |
| 6576 | MG945815 | Unassigned | Unassigned | Unassigned | 1257_0 | Clustered | 6 | VC_1257_0 | 6 | 0.6517 | 0.99999969 | 0.6517 | 0 | 0 | 0 | 1 | Ref |
| 6577 | MG945816 | Unassigned | Unassigned | Unassigned | 1151_0 | Clustered | 9 | VC_1151_0 | 9 | 0.8548 | 1 | 0.8548 | 0 | 0 | 0 | 1 | Ref |
| 6578 | MG945817 | Unassigned | Unassigned | Unassigned | 1148_0 | Clustered | 16 | VC_1148_0 | 16 | 0.8821 | 1 | 0.8821 | 0 | 0 | 0 | 1 | Ref |
| 6579 | MG945818 | Unassigned | Unassigned | Unassigned | 1291_0 | Clustered | 2 | VC_1291_0 | 2 | 1 | 1 | 1 | 0 | 0 | 0 | 1 | Ref |
| 6580 | MG945819 | Unassigned | Unassigned | Unassigned | 1116_0 | Clustered | 14 | VC_1116_0 | 14 | 0.7927 | 1 | 0.7927 | 0 | 0 | 0 | 1 | Ref |
| 6581 | MG945820 | Unassigned | Unassigned | Unassigned | 1104_0 | Clustered | 154 | VC_1104_0 | 154 | 0.8484 | 1 | 0.8484 | 0 | 0 | 0 | 1 | Ref |
| 6582 | MG945821 | Unassigned | Unassigned | Unassigned | 1334_0 | Clustered/Singleton | 1 | VC_1334_0 | 1 | 0 | 1 | 0 | 0 | 0 | 0 | 1 | Ref |
| 6583 | MG945822 | Unassigned | Unassigned | Unassigned | 1273_0 | Clustered | 6 | VC_1273_0 | 6 | 0.7385 | 0.9999804 | 0.7385 | 0 | 0 | 0 | 1 | Ref |
| 6584 | MG945823 | Unassigned | Unassigned | Unassigned | 1328_0 | Clustered | 2 | VC_1328_0 | 2 | 1 | 1 | 1 | 0 | 0 | 0 | 1 | Ref |
| 6585 | MG945824 | Unassigned | Unassigned | Unassigned | 1305_0 | Clustered | 5 | VC_1305_0 | 5 | 0.7701 | 0.66985041 | 0.5159 | 0 | 0 | 0 | 1 | Ref |
| 6586 | MG945825 | Unassigned | Unassigned | Unassigned | 1281_0 | Clustered | 7 | VC_1281_0 | 7 | 0.9212 | 0.99995294 | 0.9212 | 0 | 0 | 0 | 1 | Ref |
| 6587 | MG945826 | Unassigned | Unassigned | Unassigned | 1119_0 | Clustered | 20 | VC_1119_0 | 20 | 0.6751 | 0.99997892 | 0.6751 | 0 | 0 | 0 | 1 | Ref |
| 6588 | MG945827 | Unassigned | Unassigned | Unassigned | 1146_0 | Clustered | 3 | VC_1146_0 | 3 | 0.3641 | 0.99867077 | 0.3636 | 0 | 0 | 0 | 1 | Ref |
| 6589 | MG945828 | Unassigned | Unassigned | Unassigned | 1300_0 | Clustered | 2 | VC_1300_0 | 2 | 0.664 | 0.72985431 | 0.4846 | 0 | 0 | 0 | 1 | Ref |
| 6590 | MG945829 | Unassigned | Unassigned | Unassigned | 1251_0 | Clustered | 15 | VC_1251_0 | 15 | 0.9082 | 1 | 0.9082 | 0 | 0 | 0 | 1 | Ref |
| 6591 | MG945830 | Unassigned | Unassigned | Unassigned | 1255_0 | Clustered | 4 | VC_1255_0 | 4 | 0.248 | 0.99828153 | 0.2476 | 0 | 0 | 0 | 1 | Ref |
| 6592 | MG945831 | Unassigned | Unassigned | Unassigned | 1335_0 | Clustered | 7 | VC_1335_0 | 7 | 0.8346 | 0.99999844 | 0.8346 | 0 | 0 | 0 | 1 | Ref |
| 6593 | MG945832 | Unassigned | Unassigned | Unassigned | 1337_0 | Clustered | 2 | VC_1337_0 | 2 | 1 | 1 | 1 | 0 | 0 | 0 | 1 | Ref |
| 6594 | MG945835 | Unassigned | Unassigned | Unassigned | 1059_0 | Clustered | 3 | VC_1059_0 | 3 | 0.3928 | 0.9839946 | 0.3865 | 0 | 0 | 0 | 1 | Ref |
| 6595 | MG945836 | Unassigned | Unassigned | Unassigned | 1077_0 | Clustered | 48 | VC_1077_0 | 48 | 0.8525 | 1 | 0.8525 | 0 | 0 | 0 | 1 | Ref |
| 6596 | MG945837 | Unassigned | Unassigned | Unassigned | 398_0 | Clustered | 48 | VC_398_0 | 48 | 0.4813 | 1 | 0.4813 | 0 | 0 | 0 | 1 | Ref |
| 6597 | MG945838 | Unassigned | Unassigned | Unassigned | 995_0 | Clustered | 69 | VC_995_0 | 69 | 0.8547 | 1 | 0.8547 | 0 | 0 | 0 | 1 | Ref |
| 6598 | MG945839 | Unassigned | Unassigned | Unassigned | 1338_0 | Clustered | 4 | VC_1338_0 | 4 | 1 | 1 | 1 | 0 | 0 | 0 | 1 | Ref |
| 6599 | MG945840 | Unassigned | Unassigned | Unassigned | 1077_0 | Clustered | 48 | VC_1077_0 | 48 | 0.8525 | 1 | 0.8525 | 0 | 0 | 0 | 1 | Ref |
| 6600 | MG945842 | Unassigned | Unassigned | Unassigned | 1339_0 | Clustered | 7 | VC_1339_0 | 7 | 0.9177 | 0.99772305 | 0.9156 | 0 | 0 | 0 | 1 | Ref |
| 6601 | MG945843 | Unassigned | Unassigned | Unassigned | 995_0 | Clustered | 69 | VC_995_0 | 69 | 0.8547 | 1 | 0.8547 | 0 | 0 | 0 | 1 | Ref |
| 6602 | MG945845 | Unassigned | Unassigned | Unassigned | 1211_0 | Clustered | 25 | VC_1211_0 | 25 | 1 | 1 | 1 | 0 | 0 | 0 | 1 | Ref |
| 6603 | MG945846 | Unassigned | Unassigned | Unassigned | 1077_0 | Clustered | 48 | VC_1077_0 | 48 | 0.8525 | 1 | 0.8525 | 0 | 0 | 0 | 1 | Ref |
| 6604 | MG945848 | Unassigned | Unassigned | Unassigned | 1266_0 | Clustered | 5 | VC_1266_0 | 5 | 1 | 1 | 1 | 0 | 0 | 0 | 1 | Ref |
| 6605 | MG945849 | Unassigned | Unassigned | Unassigned | 1333_0 | Clustered | 2 | VC_1333_0 | 2 | 1 | 1 | 1 | 0 | 0 | 0 | 1 | Ref |
| 6606 | MG945850 | Unassigned | Unassigned | Unassigned | 1340_0 | Clustered | 5 | VC_1340_0 | 5 | 1 | 1 | 1 | 0 | 0 | 0 | 1 | Ref |
| 6607 | MG945852 | Unassigned | Unassigned | Unassigned | 1266_0 | Clustered | 5 | VC_1266_0 | 5 | 1 | 1 | 1 | 0 | 0 | 0 | 1 | Ref |
| 6608 | MG945853 | Unassigned | Unassigned | Unassigned | 1104_0 | Clustered | 154 | VC_1104_0 | 154 | 0.8484 | 1 | 0.8484 | 0 | 0 | 0 | 1 | Ref |
| 6609 | MG945855 | Unassigned | Unassigned | Unassigned | 1104_0 | Clustered | 154 | VC_1104_0 | 154 | 0.8484 | 1 | 0.8484 | 0 | 0 | 0 | 1 | Ref |
| 6610 | MG945857 | Unassigned | Unassigned | Unassigned | 1342_0 | Clustered | 3 | VC_1342_0 | 3 | 1 | 1 | 1 | 0 | 0 | 0 | 1 | Ref |
| 6611 | MG945858 | Unassigned | Unassigned | Unassigned | 1125_0 | Clustered | 20 | VC_1125_0 | 20 | 0.5599 | 1 | 0.5599 | 0 | 0 | 0 | 1 | Ref |
| 6612 | MG945859 | Unassigned | Unassigned | Unassigned | 1342_0 | Clustered | 3 | VC_1342_0 | 3 | 1 | 1 | 1 | 0 | 0 | 0 | 1 | Ref |
| 6613 | MG945862 | Unassigned | Unassigned | Unassigned | 1104_0 | Clustered | 154 | VC_1104_0 | 154 | 0.8484 | 1 | 0.8484 | 0 | 0 | 0 | 1 | Ref |
| 6614 | MG945864 | Unassigned | Unassigned | Unassigned | 1104_0 | Clustered | 154 | VC_1104_0 | 154 | 0.8484 | 1 | 0.8484 | 0 | 0 | 0 | 1 | Ref |
| 6615 | MG945866 | Unassigned | Unassigned | Unassigned | 1342_0 | Clustered | 3 | VC_1342_0 | 3 | 1 | 1 | 1 | 0 | 0 | 0 | 1 | Ref |
| 6616 | MG945867 | Unassigned | Unassigned | Unassigned | 1124_0 | Clustered | 10 | VC_1124_0 | 10 | 0.6268 | 1 | 0.6268 | 0 | 0 | 0 | 1 | Ref |
| 6617 | MG945869 | Unassigned | Unassigned | Unassigned | 1104_0 | Clustered | 154 | VC_1104_0 | 154 | 0.8484 | 1 | 0.8484 | 0 | 0 | 0 | 1 | Ref |
| 6618 | MG945870 | Unassigned | Unassigned | Unassigned | 1122_0 | Clustered | 10 | VC_1122_0 | 10 | 0.9643 | 0.99999697 | 0.9643 | 0 | 0 | 0 | 1 | Ref |
| 6619 | MG945909 | Unassigned | Unassigned | Unassigned | 1258_0 | Clustered | 3 | VC_1258_0 | 3 | 0.2422 | 0.85925819 | 0.2081 | 0 | 0 | 0 | 1 | Ref |
| 6620 | MG957431 | Unassigned | Unassigned | Unassigned | 82_0 | Clustered | 24 | VC_82_0 | 24 | 0.7351 | 1 | 0.7351 | 0 | 0 | 0 | 1 | Ref |
| 6621 | MG962360 | Unassigned | Unassigned | Unassigned | 1240_0 | Clustered | 82 | VC_1240_0 | 82 | 0.7945 | 1 | 0.7945 | 0 | 0 | 0 | 1 | Ref |
| 6622 | MG962361 | Unassigned | Unassigned | Unassigned | 520_64 | Clustered/Singleton | 1 | VC_520_64 | 1 | 0 | 1 | 0 | 0 | 0 | 0 | 1 | Ref |
| 6623 | MG962362 | Unassigned | Unassigned | Unassigned | 371_3 | Clustered | 22 | VC_371_3 | 22 | 0.0787 | 1 | 0.0787 | 0 | 0 | 0 | 0.9048 | Ref |
| 6624 | MG962363 | Unassigned | Unassigned | Unassigned | 371_13 | Clustered | 33 | VC_371_13 | 33 | 0.179 | 1 | 0.179 | 0 | 0 | 0 | 0.8769 | Ref |
| 6625 | MG962364 | Unassigned | Unassigned | Unassigned | 236_0 | Clustered | 72 | VC_236_0 | 72 | 0.8104 | 1 | 0.8104 | 0 | 0 | 0 | 1 | Ref |
| 6626 | MG962365 | Unassigned | Unassigned | Unassigned | 371_16 | Clustered | 42 | VC_371_16 | 42 | 0.1341 | 1 | 0.1341 | 0 | 0 | 0 | 0.7282 | Ref |
| 6627 | MG962367 | Unassigned | Unassigned | Unassigned | 1240_0 | Clustered | 82 | VC_1240_0 | 82 | 0.7945 | 1 | 0.7945 | 0 | 0 | 0 | 1 | Ref |
| 6628 | MG962368 | Unassigned | Unassigned | Unassigned | 799_1 | Clustered | 27 | VC_799_1 | 27 | 0.8611 | 1 | 0.8611 | 0 | 0 | 0 | 1 | Ref |
| 6629 | MG962369 | Unassigned | Unassigned | Unassigned | 799_1 | Clustered | 27 | VC_799_1 | 27 | 0.8611 | 1 | 0.8611 | 0 | 0 | 0 | 1 | Ref |
| 6630 | MG962370 | Unassigned | Unassigned | Unassigned | 325_47 | Clustered | 2 | VC_325_47 | 2 | 0.0045 | 0.9575092 | 0.0043 | 0 | 0 | 0 | 1 | Ref |
| 6631 | MG962371 | Unassigned | Unassigned | Unassigned | 236_0 | Clustered | 72 | VC_236_0 | 72 | 0.8104 | 1 | 0.8104 | 0 | 0 | 0 | 1 | Ref |
| 6632 | MG962372 | Unassigned | Unassigned | Unassigned | 325_5 | Clustered | 11 | VC_325_5 | 11 | 0.047 | 1 | 0.047 | 0 | 0 | 0 | 0.7636 | Ref |
| 6633 | MG962373 | Unassigned | Unassigned | Unassigned | 517_1 | Clustered | 108 | VC_517_1 | 108 | 0.8483 | 1 | 0.8483 | 0 | 0 | 0 | 0.9955 | Ref |
| 6634 | MG962374 | Unassigned | Unassigned | Unassigned | 238_0 | Clustered | 17 | VC_238_0 | 17 | 0.6109 | 1 | 0.6109 | 0 | 0 | 0 | 1 | Ref |
| 6635 | MG962375 | Unassigned | Unassigned | Unassigned | 371_2 | Clustered | 8 | VC_371_2 | 8 | 0.0232 | 1 | 0.0232 | 0 | 0 | 0 | 0.8571 | Ref |
| 6636 | MG962376 | Unassigned | Unassigned | Unassigned | 567_0 | Clustered | 8 | VC_567_0 | 8 | 0.4935 | 1 | 0.4935 | 0 | 0 | 0 | 1 | Ref |
| 6637 | MG963916 | Unassigned | Unassigned | Unassigned | 772_16 | Clustered | 10 | VC_772_16 | 10 | 0.0563 | 0.9434928 | 0.0531 | 0 | 0 | 0 | 0.8667 | Ref |
| 6638 | MG966531 | Unassigned | Unassigned | Unassigned | 392_0 | Clustered | 126 | VC_392_0 | 126 | 0.9621 | 1 | 0.9621 | 0 | 0 | 0 | 1 | Ref |
| 6639 | MG966533 | Unassigned | Unassigned | Unassigned | 277_0 | Clustered | 3 | VC_277_0 | 3 | 1 | 1 | 1 | 0 | 0 | 0 | 1 | Ref |
| 6640 | MG967616 | Unassigned | Unassigned | Unassigned | 1006_0 | Clustered | 8 | VC_1006_0 | 8 | 0.7407 | 1 | 0.7407 | 0 | 0 | 0 | 1 | Ref |

| | | | | | | | | | | | | | | | | | |
|---|---|---|---|---|---|---|---|---|---|---|---|---|---|---|---|---|---|
| 6641 | MG967617 | Unassigned | Unassigned | Unassigned | 1006_0 | Clustered | 8 | VC_1006_0 | 8 | 0.7407 | 1 | 0.7407 | 0 | 0 | 0 | 1 | Ref |
| 6642 | MG967618 | Unassigned | Unassigned | Unassigned | 1006_0 | Clustered | 8 | VC_1006_0 | 8 | 0.7407 | 1 | 0.7407 | 0 | 0 | 0 | 1 | Ref |
| 6643 | MG969404 | Unassigned | Unassigned | Unassigned | 329_14 | Clustered/Singleton | 1 | VC_329_14 | 1 | 0 | 1 | 0 | 0 | 0 | 0 | 1 | Ref |
| 6644 | MG969405 | Unassigned | Unassigned | Unassigned | 329_1 | Clustered | 17 | VC_329_1 | 17 | 0.0867 | 1 | 0.0867 | 0 | 0 | 0 | 0.8235 | Ref |
| 6645 | MG969406 | Unassigned | Unassigned | Unassigned | 329_1 | Clustered | 17 | VC_329_1 | 17 | 0.0867 | 1 | 0.0867 | 0 | 0 | 0 | 0.8235 | Ref |
| 6646 | MG969408 | Unassigned | Unassigned | Unassigned | 637_0 | Clustered | 115 | VC_637_0 | 115 | 0.731 | 1 | 0.731 | 0 | 0 | 0 | 0.8764 | Ref |
| 6647 | MG969409 | Unassigned | Unassigned | Unassigned | 637_0 | Clustered | 115 | VC_637_0 | 115 | 0.731 | 1 | 0.731 | 0 | 0 | 0 | 0.8764 | Ref |
| 6648 | MG969410 | Unassigned | Unassigned | Unassigned | 637_0 | Clustered | 115 | VC_637_0 | 115 | 0.731 | 1 | 0.731 | 0 | 0 | 0 | 0.8764 | Ref |
| 6649 | MG969411 | Unassigned | Unassigned | Unassigned | 582_6 | Clustered/Singleton | 1 | VC_582_6 | 1 | 0 | 1 | 0 | 0 | 0 | 0 | 1 | Ref |
| 6650 | MG969412 | Unassigned | Unassigned | Unassigned | 190_40 | Clustered | 2 | VC_190_40 | 2 | 0.0083 | 0.95556395 | 0.0079 | 0 | 0 | 0 | 1 | Ref |
| 6651 | MG969415 | Unassigned | Unassigned | Unassigned | 28_1 | Clustered | 53 | VC_28_1 | 53 | 0.7164 | 1 | 0.7164 | 0 | 0 | 0 | 0.9325 | Ref |
| 6652 | MG972768 | Unassigned | Unassigned | Unassigned | 111_0 | Clustered | 91 | VC_111_0 | 91 | 0.9587 | 1 | 0.9587 | 0 | 0 | 0 | 1 | Ref |
| 6653 | MG976803 | Unassigned | Unassigned | Unassigned | 813_1 | Clustered | 24 | VC_813_1 | 24 | 0.535 | 1 | 0.535 | 0 | 0 | 0 | 1 | Ref |
| 6654 | MG983742 | Unassigned | Unassigned | Unassigned | 100_0 | Clustered | 15 | VC_100_0 | 15 | 0.6536 | 1 | 0.6536 | 0 | 0 | 0 | 1 | Ref |
| 6655 | MG983743 | Unassigned | Unassigned | Unassigned | 779_3 | Clustered | 7 | VC_779_3 | 7 | 0.0766 | 1 | 0.0766 | 0 | 0 | 0 | 0.7143 | Ref |
| 6656 | MG983840 | Unassigned | Unassigned | Unassigned | 813_1 | Clustered | 24 | VC_813_1 | 24 | 0.535 | 1 | 0.535 | 0 | 0 | 0 | 1 | Ref |
| 6657 | MH000602 | Unassigned | Unassigned | Unassigned | 252_0 | Clustered | 121 | VC_252_0 | 121 | 0.7376 | 1 | 0.7376 | 0 | 0 | 0 | 0.9999 | Ref |
| 6658 | MH000603 | Unassigned | Unassigned | Unassigned | 252_0 | Clustered | 121 | VC_252_0 | 121 | 0.7376 | 1 | 0.7376 | 0 | 0 | 0 | 0.9999 | Ref |
| 6659 | MH000604 | Unassigned | Unassigned | Unassigned | 252_0 | Clustered | 121 | VC_252_0 | 121 | 0.7376 | 1 | 0.7376 | 0 | 0 | 0 | 0.9999 | Ref |
| 6660 | MH000606 | Unassigned | Unassigned | Unassigned | 325_38 | Clustered | 6 | VC_325_38 | 6 | 0.0267 | 1 | 0.0267 | 0 | 0 | 0 | 0.8 | Ref |
| 6661 | MH000607 | Unassigned | Unassigned | Unassigned | 551_7 | Clustered | 28 | VC_551_7 | 28 | 0.1661 | 1 | 0.1661 | 0 | 0 | 0 | 0.8519 | Ref |
| 6662 | MH001446 | Unassigned | Unassigned | Unassigned | 507_0 | Clustered | 8 | VC_507_0 | 8 | 0.4466 | 1 | 0.4466 | 0 | 0 | 0 | 1 | Ref |
| 6663 | MH001447 | Unassigned | Unassigned | Unassigned | 568_0 | Clustered | 8 | VC_568_0 | 8 | 0.5738 | 1 | 0.5738 | 0 | 0 | 0 | 1 | Ref |
| 6664 | MH001448 | Unassigned | Unassigned | Unassigned | 520_59 | Clustered | 5 | VC_520_59 | 5 | 0.0459 | 0.99999997 | 0.0459 | 0 | 0 | 0 | 1 | Ref |
| 6665 | MH001450 | Unassigned | Unassigned | Unassigned | 507_0 | Clustered | 8 | VC_507_0 | 8 | 0.4466 | 1 | 0.4466 | 0 | 0 | 0 | 1 | Ref |
| 6666 | MH001451 | Unassigned | Unassigned | Unassigned | 1046_0 | Clustered | 7 | VC_1046_0 | 7 | 0.9413 | 1 | 0.9413 | 0 | 0 | 0 | 1 | Ref |
| 6667 | MH001452 | Unassigned | Unassigned | Unassigned | 508_0 | Clustered | 32 | VC_508_0 | 32 | 0.906 | 1 | 0.906 | 0 | 0 | 0 | 1 | Ref |
| 6668 | MH001453 | Unassigned | Unassigned | Unassigned | 236_0 | Clustered | 72 | VC_236_0 | 72 | 0.8104 | 1 | 0.8104 | 0 | 0 | 0 | 1 | Ref |
| 6669 | MH001454 | Unassigned | Unassigned | Unassigned | 224_0 | Clustered | 19 | VC_224_0 | 19 | 0.9416 | 1 | 0.9416 | 0 | 0 | 0 | 1 | Ref |
| 6670 | MH001455 | Unassigned | Unassigned | Unassigned | 761_0 | Clustered | 67 | VC_761_0 | 67 | 0.8947 | 1 | 0.8947 | 0 | 0 | 0 | 1 | Ref |
| 6671 | MH001456 | Unassigned | Unassigned | Unassigned | 761_0 | Clustered | 67 | VC_761_0 | 67 | 0.8947 | 1 | 0.8947 | 0 | 0 | 0 | 1 | Ref |
| 6672 | MH001457 | Unassigned | Unassigned | Unassigned | 375_0 | Clustered | 3 | VC_375_0 | 3 | 0.2479 | 0.99807286 | 0.2474 | 0 | 0 | 0 | 1 | Ref |
| 6673 | MH001458 | Unassigned | Unassigned | Unassigned | 325_11 | Clustered/Singleton | 1 | VC_325_11 | 1 | 0 | 1 | 0 | 0 | 0 | 0 | 1 | Ref |
| 6674 | MH001459 | Unassigned | Unassigned | Unassigned | 515_0 | Clustered | 16 | VC_515_0 | 16 | 0.3793 | 1 | 0.3793 | 0 | 0 | 0 | 1 | Ref |
| 6675 | MH001460 | Unassigned | Unassigned | Unassigned | 981_1 | Clustered | 26 | VC_981_1 | 26 | 0.7009 | 1 | 0.7009 | 0 | 0 | 0 | 1 | Ref |
| 6676 | MH015249 | Unassigned | Unassigned | Unassigned | 593_0 | Clustered | 5 | VC_593_0 | 5 | 0.4608 | 0.99999995 | 0.4608 | 0 | 0 | 0 | 1 | Ref |
| 6677 | MH015250 | Unassigned | Unassigned | Unassigned | 833_0 | Clustered | 16 | VC_833_0 | 16 | 0.9351 | 1 | 0.9351 | 0 | 0 | 0 | 1 | Ref |
| 6678 | MH015251 | Unassigned | Unassigned | Unassigned | 1212_0 | Clustered | 2 | VC_1212_0 | 2 | 1 | 1 | 1 | 0 | 0 | 0 | 1 | Ref |
| 6679 | MH015252 | Unassigned | Unassigned | Unassigned | 593_0 | Clustered | 5 | VC_593_0 | 5 | 0.4608 | 0.99999995 | 0.4608 | 0 | 0 | 0 | 1 | Ref |
| 6680 | MH015253 | Unassigned | Unassigned | Unassigned | 833_0 | Clustered | 16 | VC_833_0 | 16 | 0.9351 | 1 | 0.9351 | 0 | 0 | 0 | 1 | Ref |
| 6681 | MH015254 | Unassigned | Unassigned | Unassigned | 593_0 | Clustered | 5 | VC_593_0 | 5 | 0.4608 | 0.99999995 | 0.4608 | 0 | 0 | 0 | 1 | Ref |
| 6682 | MH015255 | Unassigned | Unassigned | Unassigned | 977_0 | Clustered | 2 | VC_977_0 | 2 | 0.0806 | 0.93920319 | 0.0757 | 0 | 0 | 0 | 1 | Ref |
| 6683 | MH015256 | Unassigned | Unassigned | Unassigned | 833_0 | Clustered | 16 | VC_833_0 | 16 | 0.9351 | 1 | 0.9351 | 0 | 0 | 0 | 1 | Ref |
| 6684 | MH015257 | Unassigned | Unassigned | Unassigned | 833_0 | Clustered | 16 | VC_833_0 | 16 | 0.9351 | 1 | 0.9351 | 0 | 0 | 0 | 1 | Ref |
| 6685 | MH015258 | Unassigned | Unassigned | Unassigned | 593_0 | Clustered | 5 | VC_593_0 | 5 | 0.4608 | 0.99999995 | 0.4608 | 0 | 0 | 0 | 1 | Ref |
| 6686 | MH015259 | Unassigned | Unassigned | Unassigned | 833_0 | Clustered | 16 | VC_833_0 | 16 | 0.9351 | 1 | 0.9351 | 0 | 0 | 0 | 1 | Ref |
| 6687 | MH019215 | Unassigned | Unassigned | Unassigned | 859_0 | Clustered | 9 | VC_859_0 | 9 | 0.9426 | 1 | 0.9426 | 0 | 0 | 0 | 1 | Ref |
| 6688 | MH019216 | Unassigned | Unassigned | Unassigned | 859_0 | Clustered | 9 | VC_859_0 | 9 | 0.9426 | 1 | 0.9426 | 0 | 0 | 0 | 1 | Ref |
| 6689 | MH020235 | Unassigned | Unassigned | Unassigned | 520_31 | Clustered | 11 | VC_520_31 | 11 | 0.0897 | 1 | 0.0897 | 0 | 0 | 0 | 0.6 | Ref |
| 6690 | MH020236 | Unassigned | Unassigned | Unassigned | 325_53 | Clustered | 7 | VC_325_53 | 7 | 0.0242 | 1 | 0.0242 | 0 | 0 | 0 | 0.9048 | Ref |
| 6691 | MH020237 | Unassigned | Unassigned | Unassigned | 215_2 | Clustered | 5 | VC_215_2 | 5 | 0.0572 | 0.99999997 | 0.0572 | 0 | 0 | 0 | 1 | Ref |
| 6692 | MH020238 | Unassigned | Unassigned | Unassigned | 514_0 | Clustered | 75 | VC_514_0 | 75 | 0.9402 | 1 | 0.9402 | 0 | 0 | 0 | 1 | Ref |
| 6693 | MH020239 | Unassigned | Unassigned | Unassigned | 940_0 | Clustered | 34 | VC_940_0 | 34 | 0.7959 | 1 | 0.7959 | 0 | 0 | 0 | 1 | Ref |
| 6694 | MH020240 | Unassigned | Unassigned | Unassigned | 223_0 | Clustered | 38 | VC_223_0 | 38 | 0.9095 | 1 | 0.9095 | 0 | 0 | 0 | 1 | Ref |
| 6695 | MH020241 | Unassigned | Unassigned | Unassigned | 1176_0 | Clustered | 41 | VC_1176_0 | 41 | 0.9558 | 1 | 0.9558 | 0 | 0 | 0 | 0.9976 | Ref |
| 6696 | MH020244 | Unassigned | Unassigned | Unassigned | 325_23 | Clustered | 17 | VC_325_23 | 17 | 0.0716 | 1 | 0.0716 | 0 | 0 | 0 | 0.6397 | Ref |
| 6697 | MH020246 | Unassigned | Unassigned | Unassigned | 556_0 | Clustered | 21 | VC_556_0 | 21 | 0.9912 | 1 | 0.9912 | 0 | 0 | 0 | 1 | Ref |
| 6698 | MH020247 | Unassigned | Unassigned | Unassigned | 551_8 | Clustered | 19 | VC_551_8 | 19 | 0.1039 | 1 | 0.1039 | 0 | 0 | 0 | 0.8713 | Ref |
| 6699 | MH020248 | Unassigned | Unassigned | Unassigned | 513_0 | Clustered | 32 | VC_513_0 | 32 | 0.6881 | 1 | 0.6881 | 0 | 0 | 0 | 1 | Ref |
| 6700 | MH020249 | Unassigned | Unassigned | Unassigned | 517_1 | Clustered | 108 | VC_517_1 | 108 | 0.8483 | 1 | 0.8483 | 0 | 0 | 0 | 0.9955 | Ref |
| 6701 | MH023293 | Unassigned | Unassigned | Unassigned | 461_0 | Clustered | 96 | VC_461_0 | 96 | 0.9307 | 1 | 0.9307 | 0 | 0 | 0 | 0.9575 | Ref |
| 6702 | MH025888 | Unassigned | Unassigned | Unassigned | 565_0 | Clustered | 20 | VC_565_0 | 20 | 0.8398 | 1 | 0.8398 | 0 | 0 | 0 | 1 | Ref |
| 6703 | MH025889 | Unassigned | Unassigned | Unassigned | 866_1 | Clustered | 20 | VC_866_1 | 20 | 0.4814 | 1 | 0.4814 | 0 | 0 | 0 | 1 | Ref |
| 6704 | MH025890 | Unassigned | Unassigned | Unassigned | 570_0 | Clustered | 9 | VC_570_0 | 9 | 0.499 | 1 | 0.499 | 0 | 0 | 0 | 1 | Ref |
| 6705 | MH031343 | Unassigned | Unassigned | Unassigned | 386_0 | Clustered | 49 | VC_386_0 | 49 | 0.7941 | 1 | 0.7941 | 0 | 0 | 0 | 1 | Ref |
| 6706 | MH042230 | Unassigned | Unassigned | Unassigned | 699_0 | Clustered | 71 | VC_699_0 | 71 | 0.9352 | 1 | 0.9352 | 0 | 0 | 0 | 1 | Ref |
| 6707 | MH045554 | Unassigned | Unassigned | Unassigned | 1240_0 | Clustered | 82 | VC_1240_0 | 82 | 0.7945 | 1 | 0.7945 | 0 | 0 | 0 | 1 | Ref |
| 6708 | MH045557 | Unassigned | Unassigned | Unassigned | 1344_0 | Clustered | 78 | VC_1344_0 | 78 | 0.9732 | 1 | 0.9732 | 0 | 0 | 0 | 1 | Ref |
| 6709 | MH045558 | Unassigned | Unassigned | Unassigned | 1240_0 | Clustered | 82 | VC_1240_0 | 82 | 0.7945 | 1 | 0.7945 | 0 | 0 | 0 | 1 | Ref |
| 6710 | MH045560 | Unassigned | Unassigned | Unassigned | 1240_0 | Clustered | 82 | VC_1240_0 | 82 | 0.7945 | 1 | 0.7945 | 0 | 0 | 0 | 1 | Ref |
| 6711 | MH045561 | Unassigned | Unassigned | Unassigned | 1344_0 | Clustered | 78 | VC_1344_0 | 78 | 0.9732 | 1 | 0.9732 | 0 | 0 | 0 | 1 | Ref |
| 6712 | MH045562 | Unassigned | Unassigned | Unassigned | 1240_0 | Clustered | 82 | VC_1240_0 | 82 | 0.7945 | 1 | 0.7945 | 0 | 0 | 0 | 1 | Ref |
| 6713 | MH045563 | Unassigned | Unassigned | Unassigned | 1240_0 | Clustered | 82 | VC_1240_0 | 82 | 0.7945 | 1 | 0.7945 | 0 | 0 | 0 | 1 | Ref |
| 6714 | MH045564 | Unassigned | Unassigned | Unassigned | 1240_0 | Clustered | 82 | VC_1240_0 | 82 | 0.7945 | 1 | 0.7945 | 0 | 0 | 0 | 1 | Ref |
| 6715 | MH045565 | Unassigned | Unassigned | Unassigned | 1240_0 | Clustered | 82 | VC_1240_0 | 82 | 0.7945 | 1 | 0.7945 | 0 | 0 | 0 | 1 | Ref |
| 6716 | MH045566 | Unassigned | Unassigned | Unassigned | 1234_0 | Clustered | 44 | VC_1234_0 | 44 | 1 | 1 | 1 | 0 | 0 | 0 | 1 | Ref |
| 6717 | MH045567 | Unassigned | Unassigned | Unassigned | 1234_0 | Clustered | 44 | VC_1234_0 | 44 | 1 | 1 | 1 | 0 | 0 | 0 | 1 | Ref |
| 6718 | MH045568 | Unassigned | Unassigned | Unassigned | 1234_0 | Clustered | 44 | VC_1234_0 | 44 | 1 | 1 | 1 | 0 | 0 | 0 | 1 | Ref |
| 6719 | MH045569 | Unassigned | Unassigned | Unassigned | 761_0 | Clustered | 67 | VC_761_0 | 67 | 0.8947 | 1 | 0.8947 | 0 | 0 | 0 | 1 | Ref |
| 6720 | MH047632 | Unassigned | Unassigned | Unassigned | 513_0 | Clustered | 32 | VC_513_0 | 32 | 0.6881 | 1 | 0.6881 | 0 | 0 | 0 | 1 | Ref |
| 6721 | MH047633 | Unassigned | Unassigned | Unassigned | 513_0 | Clustered | 32 | VC_513_0 | 32 | 0.6881 | 1 | 0.6881 | 0 | 0 | 0 | 1 | Ref |
| 6722 | MH051246 | Unassigned | Unassigned | Unassigned | 236_0 | Clustered | 72 | VC_236_0 | 72 | 0.8104 | 1 | 0.8104 | 0 | 0 | 0 | 1 | Ref |
| 6723 | MH051247 | Unassigned | Unassigned | Unassigned | 517_1 | Clustered | 108 | VC_517_1 | 108 | 0.8483 | 1 | 0.8483 | 0 | 0 | 0 | 0.9955 | Ref |
| 6724 | MH051249 | Unassigned | Unassigned | Unassigned | 508_0 | Clustered | 32 | VC_508_0 | 32 | 0.906 | 1 | 0.906 | 0 | 0 | 0 | 1 | Ref |
| 6725 | MH051250 | Unassigned | Unassigned | Unassigned | 940_0 | Clustered | 34 | VC_940_0 | 34 | 0.7959 | 1 | 0.7959 | 0 | 0 | 0 | 1 | Ref |
| 6726 | MH051251 | Unassigned | Unassigned | Unassigned | 371_13 | Clustered | 33 | VC_371_13 | 33 | 0.179 | 1 | 0.179 | 0 | 0 | 0 | 0.8769 | Ref |
| 6727 | MH051252 | Unassigned | Unassigned | Unassigned | 216_6 | Clustered | 42 | VC_216_6 | 42 | 0.473 | 1 | 0.473 | 0 | 0 | 0 | 0.8351 | Ref |
| 6728 | MH051253 | Unassigned | Unassigned | Unassigned | 223_0 | Clustered | 38 | VC_223_0 | 38 | 0.9095 | 1 | 0.9095 | 0 | 0 | 0 | 1 | Ref |
| 6729 | MH051254 | Unassigned | Unassigned | Unassigned | 940_0 | Clustered | 34 | VC_940_0 | 34 | 0.7959 | 1 | 0.7959 | 0 | 0 | 0 | 1 | Ref |
| 6730 | MH051255 | Unassigned | Unassigned | Unassigned | 238_0 | Clustered | 17 | VC_238_0 | 17 | 0.6109 | 1 | 0.6109 | 0 | 0 | 0 | 1 | Ref |
| 6731 | MH051256 | Unassigned | Unassigned | Unassigned | 940_0 | Clustered | 34 | VC_940_0 | 34 | 0.7959 | 1 | 0.7959 | 0 | 0 | 0 | 1 | Ref |
| 6732 | MH051257 | Unassigned | Unassigned | Unassigned | 520_51 | Clustered/Singleton | 1 | VC_520_51 | 1 | 0 | 1 | 0 | 0 | 0 | 0 | 1 | Ref |
| 6733 | MH051258 | Unassigned | Unassigned | Unassigned | 234_0 | Clustered | 21 | VC_234_0 | 21 | 0.8355 | 1 | 0.8355 | 0 | 0 | 0 | 1 | Ref |
| 6734 | MH051259 | Unassigned | Unassigned | Unassigned | 513_0 | Clustered | 32 | VC_513_0 | 32 | 0.6881 | 1 | 0.6881 | 0 | 0 | 0 | 1 | Ref |
| 6735 | MH051260 | Unassigned | Unassigned | Unassigned | 558_61 | Clustered | 2 | VC_558_61 | 2 | 0.0032 | 0.66165479 | 0.0021 | 0 | 0 | 0 | 1 | Ref |
| 6736 | MH051261 | Unassigned | Unassigned | Unassigned | 558_7 | Clustered/Singleton | 1 | VC_558_7 | 1 | 0 | 1 | 0 | 0 | 0 | 0 | 1 | Ref |
| 6737 | MH051264 | Unassigned | Unassigned | Unassigned | 371_5 | Clustered | 2 | VC_371_5 | 2 | 0.0035 | 0.95771071 | 0.0033 | 0 | 0 | 0 | 1 | Ref |
| 6738 | MH051265 | Unassigned | Unassigned | Unassigned | 509_0 | Clustered | 37 | VC_509_0 | 37 | 0.9689 | 1 | 0.9689 | 0 | 0 | 0 | 1 | Ref |
| 6739 | MH051266 | Unassigned | Unassigned | Unassigned | 558_50 | Clustered | 15 | VC_558_50 | 15 | 0.0468 | 1 | 0.0468 | 0 | 0 | 0 | 0.9238 | Ref |
| 6740 | MH051333 | Unassigned | Unassigned | Unassigned | 847_1 | Clustered | 26 | VC_847_1 | 26 | 0.6323 | 1 | 0.6323 | 0 | 0 | 0 | 0.9815 | Ref |
| 6741 | MH051334 | Unassigned | Unassigned | Unassigned | 638_0 | Clustered | 45 | VC_638_0 | 45 | 0.7741 | 1 | 0.7741 | 0 | 0 | 0 | 1 | Ref |
| 6742 | MH051335 | Unassigned | Unassigned | Unassigned | 618_0 | Clustered | 13 | VC_618_0 | 13 | 0.6252 | 1 | 0.6252 | 0 | 0 | 0 | 1 | Ref |
| 6743 | MH051911 | Unassigned | Unassigned | Unassigned | 575_0 | Clustered | 35 | VC_575_0 | 35 | 0.8815 | 1 | 0.8815 | 0 | 0 | 0 | 1 | Ref |
| 6744 | MH051912 | Unassigned | Unassigned | Unassigned | 575_0 | Clustered | 35 | VC_575_0 | 35 | 0.8815 | 1 | 0.8815 | 0 | 0 | 0 | 1 | Ref |
| 6745 | MH051913 | Unassigned | Unassigned | Unassigned | 290_6 | Clustered | 12 | VC_290_6 | 12 | 0.1149 | 1 | 0.1149 | 0 | 0 | 0 | 0.8182 | Ref |
| 6746 | MH051914 | Unassigned | Unassigned | Unassigned | 329_21 | Clustered/Singleton | 1 | VC_329_21 | 1 | 0 | 1 | 0 | 0 | 0 | 0 | 1 | Ref |
| 6747 | MH051915 | Unassigned | Unassigned | Unassigned | 283_151 | Clustered/Singleton | 1 | VC_283_151 | 1 | 0 | 1 | 0 | 0 | 0 | 0 | 1 | Ref |
| 6748 | MH051916 | Unassigned | Unassigned | Unassigned | 283_170 | Clustered/Singleton | 1 | VC_283_170 | 1 | 0 | 1 | 0 | 0 | 0 | 0 | 1 | Ref |
| 6749 | MH051917 | Unassigned | Unassigned | Unassigned | 290_5 | Clustered | 14 | VC_290_5 | 14 | 0.1422 | 1 | 0.1422 | 0 | 0 | 0 | 0.9231 | Ref |
| 6750 | MH051918 | Unassigned | Unassigned | Unassigned | 576_0 | Clustered | 4 | VC_576_0 | 4 | 0.0606 | 0.99998766 | 0.0606 | 0 | 0 | 0 | 1 | Ref |
| 6751 | MH059632 | Unassigned | Unassigned | Unassigned | 458_0 | Clustered | 5 | VC_458_0 | 5 | 0.1938 | 0.99999997 | 0.1938 | 0 | 0 | 0 | 1 | Ref |
| 6752 | MH059635 | Unassigned | Unassigned | Unassigned | 117_0 | Clustered | 9 | VC_117_0 | 9 | 0.9977 | 0.98988637 | 0.9876 | 0 | 0 | 0 | 1 | Ref |
| 6753 | MH059636 | Unassigned | Unassigned | Unassigned | 490_5 | Clustered/Singleton | 1 | VC_490_5 | 1 | 0 | 1 | 0 | 0 | 0 | 0 | 1 | Ref |
| 6754 | MH059638 | Unassigned | Unassigned | Unassigned | 841_0 | Clustered | 10 | VC_841_0 | 10 | 0.5997 | 1 | 0.5997 | 0 | 0 | 0 | 1 | Ref |
| 6755 | MH059639 | Unassigned | Unassigned | Unassigned | 393_0 | Clustered | 7 | VC_393_0 | 7 | 0.1775 | 1 | 0.1775 | 0 | 0 | 0 | 1 | Ref |
| 6756 | MH077576 | Unassigned | Unassigned | Unassigned | 216_6 | Clustered | 42 | VC_216_6 | 42 | 0.473 | 1 | 0.473 | 0 | 0 | 0 | 0.8351 | Ref |
| 6757 | MH077577 | Unassigned | Unassigned | Unassigned | 236_0 | Clustered | 72 | VC_236_0 | 72 | 0.8104 | 1 | 0.8104 | 0 | 0 | 0 | 1 | Ref |
| 6758 | MH077578 | Unassigned | Unassigned | Unassigned | 520_34 | Clustered/Singleton | 1 | VC_520_34 | 1 | 0 | 1 | 0 | 0 | 0 | 0 | 1 | Ref |
| 6759 | MH077579 | Unassigned | Unassigned | Unassigned | 546_4 | Clustered | 16 | VC_546_4 | 16 | 0.2517 | 0.9999998 | 0.2517 | 0 | 0 | 0 | 0.9417 | Ref |
| 6760 | MH077580 | Unassigned | Unassigned | Unassigned | 520_31 | Clustered | 11 | VC_520_31 | 11 | 0.0897 | 1 | 0.0897 | 0 | 0 | 0 | 0.6 | Ref |
| 6761 | MH077581 | Unassigned | Unassigned | Unassigned | 517_1 | Clustered | 108 | VC_517_1 | 108 | 0.8483 | 1 | 0.8483 | 0 | 0 | 0 | 0.9955 | Ref |
| 6762 | MH077582 | Unassigned | Unassigned | Unassigned | 371_2 | Clustered | 8 | VC_371_2 | 8 | 0.0232 | 1 | 0.0232 | 0 | 0 | 0 | 0.8571 | Ref |
| 6763 | MH077583 | Unassigned | Unassigned | Unassigned | 514_0 | Clustered | 75 | VC_514_0 | 75 | 0.9402 | 1 | 0.9402 | 0 | 0 | 0 | 1 | Ref |
| 6764 | MH077584 | Unassigned | Unassigned | Unassigned | 761_0 | Clustered | 67 | VC_761_0 | 67 | 0.8947 | 1 | 0.8947 | 0 | 0 | 0 | 1 | Ref |
| 6765 | MH102284 | Unassigned | Unassigned | Unassigned | 584_0 | Clustered | 28 | VC_584_0 | 28 | 0.7704 | 1 | 0.7704 | 0 | 0 | 0 | 1 | Ref |
| 6766 | MH102285 | Unassigned | Unassigned | Unassigned | 190_23 | Clustered | 10 | VC_190_23 | 10 | 0.0704 | 1 | 0.0704 | 0 | 0 | 0 | 0.8444 | Ref |
| 6767 | MH105773 | Unassigned | Unassigned | Unassigned | 607_0 | Clustered | 16 | VC_607_0 | 16 | 0.9179 | 1 | 0.9179 | 0 | 0 | 0 | 1 | Ref |
| 6768 | MH107028 | Unassigned | Unassigned | Unassigned | 844_0 | Clustered | 27 | VC_844_0 | 27 | 0.9385 | 1 | 0.9385 | 0 | 0 | 0 | 1 | Ref |
| 6769 | MH107118 | Unassigned | Unassigned | Unassigned | 96_0 | Clustered | 30 | VC_96_0 | 30 | 0.718 | 1 | 0.718 | 0 | 0 | 0 | 1 | Ref |
| 6770 | MH107769 | Unassigned | Unassigned | Unassigned | 149_12 | Clustered | 2 | VC_149_12 | 2 | 0.0044 | 0.83156708 | 0.0037 | 0 | 0 | 0 | 1 | Ref |
| 6771 | MH107770 | Unassigned | Unassigned | Unassigned | 423_0 | Clustered | 45 | VC_423_0 | 45 | 0.9484 | 1 | 0.9484 | 0 | 0 | 0 | 1 | Ref |
| 6772 | MH113812 | Unassigned | Unassigned | Unassigned | 1027_0 | Clustered | 20 | VC_1027_0 | 20 | 0.7114 | 1 | 0.7114 | 0 | 0 | 0 | 1 | Ref |
| 6773 | MH113813 | Unassigned | Unassigned | Unassigned | 1027_0 | Clustered | 20 | VC_1027_0 | 20 | 0.7114 | 1 | 0.7114 | 0 | 0 | 0 | 1 | Ref |
| 6774 | MH113814 | Unassigned | Unassigned | Unassigned | 1027_0 | Clustered | 20 | VC_1027_0 | 20 | 0.7114 | 1 | 0.7114 | 0 | 0 | 0 | 1 | Ref |
| 6775 | MH113815 | Unassigned | Unassigned | Unassigned | 1027_0 | Clustered | 20 | VC_1027_0 | 20 | 0.7114 | 1 | 0.7114 | 0 | 0 | 0 | 1 | Ref |
| 6776 | MH115576 | Unassigned | Unassigned | Unassigned | 696_1 | Clustered | 8 | VC_696_1 | 8 | 0.5452 | 1 | 0.5452 | 0 | 0 | 0 | 0.9286 | Ref |
| 6777 | MH128984 | Unassigned | Unassigned | Unassigned | 279_0 | Clustered | 2 | VC_279_0 | 2 | 0.1888 | 0.95124665 | 0.1796 | 0 | 0 | 0 | 1 | Ref |
| 6778 | MH133207 | Unassigned | Unassigned | Unassigned | 701_1 | Clustered | 2 | VC_701_1 | 2 | 0.1304 | 0.95114164 | 0.124 | 0 | 0 | 0 | 1 | Ref |
| 6779 | MH136584 | Unassigned | Unassigned | Unassigned | 149_8 | Clustered | 66 | VC_149_8 | 66 | 0.4565 | 1 | 0.4565 | 0 | 0 | 0 | 0.8182 | Ref |
| 6780 | MH142219 | Unassigned | Unassigned | Unassigned | 24_0 | Clustered | 3 | VC_24_0 | 3 | 0.3455 | 0.99844825 | 0.345 | 0 | 0 | 0 | 1 | Ref |
| 6781 | MH142220 | Unassigned | Unassigned | Unassigned | 24_0 | Clustered | 3 | VC_24_0 | 3 | 0.3455 | 0.99844825 | 0.345 | 0 | 0 | 0 | 1 | Ref |
| 6782 | MH153799 | Unassigned | Unassigned | Unassigned | 1348_0 | Clustered | 5 | VC_1348_0 | 5 | 0.5462 | 0.99999985 | 0.5462 | 0 | 0 | 0 | 1 | Ref |
| 6783 | MH153800 | Unassigned | Unassigned | Unassigned | 803_0 | Clustered | 3 | VC_803_0 | 3 | 0.0513 | 0.99857736 | 0.0512 | 0 | 0 | 0 | 0.6667 | Ref |
| 6784 | MH153801 | Unassigned | Unassigned | Unassigned | 803_0 | Clustered | 3 | VC_803_0 | 3 | 0.0513 | 0.99857736 | 0.0512 | 0 | 0 | 0 | 0.6667 | Ref |
| 6785 | MH153802 | Unassigned | Unassigned | Unassigned | 1240_0 | Clustered | 82 | VC_1240_0 | 82 | 0.7945 | 1 | 0.7945 | 0 | 0 | 0 | 1 | Ref |
| 6786 | MH153803 | Unassigned | Unassigned | Unassigned | 1171_0 | Clustered | 21 | VC_1171_0 | 21 | 0.7125 | 1 | 0.7125 | 0 | 0 | 0 | 1 | Ref |
| 6787 | MH153804 | Unassigned | Unassigned | Unassigned | 726_0 | Clustered | 3 | VC_726_0 | 3 | 0.0381 | 0.88955608 | 0.0339 | 0 | 0 | 0 | 1 | Ref |
| 6788 | MH153805 | Unassigned | Unassigned | Unassigned | 1240_0 | Clustered | 82 | VC_1240_0 | 82 | 0.7945 | 1 | 0.7945 | 0 | 0 | 0 | 1 | Ref |
| 6789 | MH153806 | Unassigned | Unassigned | Unassigned | 1240_0 | Clustered | 82 | VC_1240_0 | 82 | 0.7945 | 1 | 0.7945 | 0 | 0 | 0 | 1 | Ref |
| 6790 | MH153807 | Unassigned | Unassigned | Unassigned | 1070_0 | Clustered | 2 | VC_1070_0 | 2 | 0.1299 | 0.95375267 | 0.1239 | 0 | 0 | 0 | 1 | Ref |
| 6791 | MH153808 | Unassigned | Unassigned | Unassigned | 1176_0 | Clustered | 41 | VC_1176_0 | 41 | 0.9558 | 1 | 0.9558 | 0 | 0 | 0 | 0.9976 | Ref |
| 6792 | MH153809 | Unassigned | Unassigned | Unassigned | 1093_0 | Clustered | 40 | VC_1093_0 | 40 | 0.8754 | 1 | 0.8754 | 0 | 0 | 0 | 1 | Ref |
| 6793 | MH153810 | Unassigned | Unassigned | Unassigned | 510_0 | Clustered | 12 | VC_510_0 | 12 | 0.3963 | 1 | 0.3963 | 0 | 0 | 0 | 0.9697 | Ref |
| 6794 | MH153811 | Unassigned | Unassigned | Unassigned | 1240_0 | Clustered | 82 | VC_1240_0 | 82 | 0.7945 | 1 | 0.7945 | 0 | 0 | 0 | 1 | Ref |
| 6795 | MH153812 | Unassigned | Unassigned | Unassigned | 1070_0 | Clustered | 2 | VC_1070_0 | 2 | 0.1299 | 0.95375267 | 0.1239 | 0 | 0 | 0 | 1 | Ref |
| 6796 | MH153813 | Unassigned | Unassigned | Unassigned | 1171_0 | Clustered | 21 | VC_1171_0 | 21 | 0.7125 | 1 | 0.7125 | 0 | 0 | 0 | 1 | Ref |
| 6797 | MH155864 | Unassigned | Unassigned | Unassigned | 514_0 | Clustered | 75 | VC_514_0 | 75 | 0.9402 | 1 | 0.9402 | 0 | 0 | 0 | 1 | Ref |
| 6798 | MH155865 | Unassigned | Unassigned | Unassigned | 520_38 | Clustered | 6 | VC_520_38 | 6 | 0.0519 | 1 | 0.0519 | 0 | 0 | 0 | 0.8667 | Ref |
| 6799 | MH155866 | Unassigned | Unassigned | Unassigned | 520_4 | Clustered/Singleton | 1 | VC_520_4 | 1 | 0 | 1 | 0 | 0 | 0 | 0 | 1 | Ref |
| 6800 | MH155868 | Unassigned | Unassigned | Unassigned | 1224_0 | Clustered | 19 | VC_1224_0 | 19 | 1 | 1 | 1 | 0 | 0 | 0 | 1 | Ref |
| 6801 | MH155870 | Unassigned | Unassigned | Unassigned | 1351_0 | Clustered | 2 | VC_1351_0 | 2 | 1 | 1 | 1 | 0 | 0 | 0 | 1 | Ref |
| 6802 | MH155871 | Unassigned | Unassigned | Unassigned | 520_38 | Clustered | 6 | VC_520_38 | 6 | 0.0519 | 1 | 0.0519 | 0 | 0 | 0 | 0.8667 | Ref |

| | | | | | | | | | | | | | | | | | |
|---|---|---|---|---|---|---|---|---|---|---|---|---|---|---|---|---|---|
| 6803 | MH155873 | Unassigned | Unassigned | Unassigned | 1352_0 | Clustered | 31 | VC_1352_0 | 31 | 0.849 | 1 | 0.849 | 0 | 0 | 0 | 1 | Ref |
| 6804 | MH155874 | Unassigned | Unassigned | Unassigned | 371_13 | Clustered | 33 | VC_371_13 | 33 | 0.179 | 1 | 0.179 | 0 | 0 | 0 | 0.8769 | Ref |
| 6805 | MH155875 | Unassigned | Unassigned | Unassigned | 517_1 | Clustered | 108 | VC_517_1 | 108 | 0.8483 | 1 | 0.8483 | 0 | 0 | 0 | 0.9955 | Ref |
| 6806 | MH155876 | Unassigned | Unassigned | Unassigned | 223_0 | Clustered | 38 | VC_223_0 | 38 | 0.9095 | 1 | 0.9095 | 0 | 0 | 0 | 1 | Ref |
| 6807 | MH155877 | Unassigned | Unassigned | Unassigned | 872_0 | Clustered | 31 | VC_872_0 | 31 | 0.8151 | 1 | 0.8151 | 0 | 0 | 0 | 0.8194 | Ref |
| 6808 | MH155879 | Unassigned | Unassigned | Unassigned | 517_1 | Clustered | 108 | VC_517_1 | 108 | 0.8483 | 1 | 0.8483 | 0 | 0 | 0 | 0.9955 | Ref |
| 6809 | MH155880 | Unassigned | Unassigned | Unassigned | 872_0 | Clustered | 31 | VC_872_0 | 31 | 0.8151 | 1 | 0.8151 | 0 | 0 | 0 | 0.8194 | Ref |
| 6810 | MH159197 | Unassigned | Unassigned | Unassigned | 149_15 | Clustered | 16 | VC_149_15 | 16 | 0.0788 | 1 | 0.0788 | 0 | 0 | 0 | 0.8083 | Ref |
| 6811 | MH160392 | Unassigned | Unassigned | Unassigned | 779_2 | Clustered | 5 | VC_779_2 | 5 | 0.0876 | 0.99999997 | 0.0876 | 0 | 0 | 0 | 1 | Ref |
| 6812 | MH160766 | Unassigned | Unassigned | Unassigned | 847_1 | Clustered | 26 | VC_847_1 | 26 | 0.6323 | 1 | 0.6323 | 0 | 0 | 0 | 0.9815 | Ref |
| 6813 | MH160767 | Unassigned | Unassigned | Unassigned | 247_0 | Clustered | 14 | VC_247_0 | 14 | 0.9119 | 1 | 0.9119 | 0 | 0 | 0 | 0.967 | Ref |
| 6814 | MH165274 | Unassigned | Unassigned | Unassigned | 492_1 | Clustered | 20 | VC_492_1 | 20 | 0.4176 | 1 | 0.4176 | 0 | 0 | 0 | 1 | Ref |
| 6815 | MH171093 | Unassigned | Unassigned | Unassigned | 982_0 | Clustered | 62 | VC_982_0 | 62 | 0.8646 | 1 | 0.8646 | 0 | 0 | 0 | 0.743 | Ref |
| 6816 | MH171094 | Unassigned | Unassigned | Unassigned | 982_0 | Clustered | 62 | VC_982_0 | 62 | 0.8646 | 1 | 0.8646 | 0 | 0 | 0 | 0.743 | Ref |
| 6817 | MH171095 | Unassigned | Unassigned | Unassigned | 982_0 | Clustered | 62 | VC_982_0 | 62 | 0.8646 | 1 | 0.8646 | 0 | 0 | 0 | 0.743 | Ref |
| 6818 | MH171096 | Unassigned | Unassigned | Unassigned | 982_0 | Clustered | 62 | VC_982_0 | 62 | 0.8646 | 1 | 0.8646 | 0 | 0 | 0 | 0.743 | Ref |
| 6819 | MH171097 | Unassigned | Unassigned | Unassigned | 982_0 | Clustered | 62 | VC_982_0 | 62 | 0.8646 | 1 | 0.8646 | 0 | 0 | 0 | 0.743 | Ref |
| 6820 | MH171098 | Unassigned | Unassigned | Unassigned | 982_0 | Clustered | 62 | VC_982_0 | 62 | 0.8646 | 1 | 0.8646 | 0 | 0 | 0 | 0.743 | Ref |
| 6821 | MH172261 | Unassigned | Unassigned | Unassigned | 823_0 | Clustered | 124 | VC_823_0 | 124 | 0.9492 | 1 | 0.9492 | 0 | 0 | 0 | 1 | Ref |
| 6822 | MH172262 | Unassigned | Unassigned | Unassigned | 823_0 | Clustered | 124 | VC_823_0 | 124 | 0.9492 | 1 | 0.9492 | 0 | 0 | 0 | 1 | Ref |
| 6823 | MH172263 | Unassigned | Unassigned | Unassigned | 823_0 | Clustered | 124 | VC_823_0 | 124 | 0.9492 | 1 | 0.9492 | 0 | 0 | 0 | 1 | Ref |
| 6824 | MH172264 | Unassigned | Unassigned | Unassigned | 823_0 | Clustered | 124 | VC_823_0 | 124 | 0.9492 | 1 | 0.9492 | 0 | 0 | 0 | 1 | Ref |
| 6825 | MH178096 | Unassigned | Unassigned | Unassigned | 33_0 | Clustered | 4 | VC_33_0 | 4 | 0.06 | 0.99994826 | 0.06 | 0 | 0 | 0 | 1 | Ref |
| 6826 | MH178381 | Unassigned | Unassigned | Unassigned | 975_0 | Clustered | 11 | VC_975_0 | 11 | 0.9687 | 1 | 0.9687 | 0 | 0 | 0 | 1 | Ref |
| 6827 | MH178382 | Unassigned | Unassigned | Unassigned | 975_0 | Clustered | 11 | VC_975_0 | 11 | 0.9687 | 1 | 0.9687 | 0 | 0 | 0 | 1 | Ref |
| 6828 | MH179470 | Unassigned | Unassigned | Unassigned | 1189_0 | Clustered | 9 | VC_1189_0 | 9 | 0.968 | 1 | 0.968 | 0 | 0 | 0 | 1 | Ref |
| 6829 | MH179471 | Unassigned | Unassigned | Unassigned | 1189_0 | Clustered | 9 | VC_1189_0 | 9 | 0.968 | 1 | 0.968 | 0 | 0 | 0 | 1 | Ref |
| 6830 | MH179472 | Unassigned | Unassigned | Unassigned | 130_0 | Clustered | 19 | VC_130_0 | 19 | 0.6068 | 1 | 0.6068 | 0 | 0 | 0 | 1 | Ref |
| 6831 | MH179473 | Unassigned | Unassigned | Unassigned | 878_0 | Clustered | 32 | VC_878_0 | 32 | 0.9965 | 1 | 0.9965 | 0 | 0 | 0 | 1 | Ref |
| 6832 | MH179474 | Unassigned | Unassigned | Unassigned | 369_0 | Clustered | 6 | VC_369_0 | 6 | 0.5924 | 1 | 0.5924 | 0 | 0 | 0 | 1 | Ref |
| 6833 | MH179475 | Unassigned | Unassigned | Unassigned | 130_0 | Clustered | 19 | VC_130_0 | 19 | 0.6068 | 1 | 0.6068 | 0 | 0 | 0 | 1 | Ref |
| 6834 | MH179476 | Unassigned | Unassigned | Unassigned | 600_2 | Clustered | 10 | VC_600_2 | 10 | 0.3041 | 1 | 0.3041 | 0 | 0 | 0 | 1 | Ref |
| 6835 | MH179477 | Unassigned | Unassigned | Unassigned | 600_2 | Clustered | 10 | VC_600_2 | 10 | 0.3041 | 1 | 0.3041 | 0 | 0 | 0 | 1 | Ref |
| 6836 | MH179478 | Unassigned | Unassigned | Unassigned | 130_0 | Clustered | 19 | VC_130_0 | 19 | 0.6068 | 1 | 0.6068 | 0 | 0 | 0 | 1 | Ref |
| 6837 | MH179479 | Unassigned | Unassigned | Unassigned | 1189_0 | Clustered | 9 | VC_1189_0 | 9 | 0.968 | 1 | 0.968 | 0 | 0 | 0 | 1 | Ref |
| 6838 | MH179480 | Unassigned | Unassigned | Unassigned | 842_0 | Clustered | 2 | VC_842_0 | 2 | 0.0559 | 0.95564851 | 0.0534 | 0 | 0 | 0 | 1 | Ref |
| 6839 | MH181876 | Unassigned | Unassigned | Unassigned | 428_0 | Clustered | 33 | VC_428_0 | 33 | 0.9915 | 1 | 0.9915 | 0 | 0 | 0 | 1 | Ref |
| 6840 | MH183162 | Unassigned | Unassigned | Unassigned | 1345_1 | Clustered | 2 | VC_1345_1 | 2 | 0.0956 | 0.9561938 | 0.0914 | 0 | 0 | 0 | 1 | Ref |
| 6841 | MH191398 | Unassigned | Unassigned | Unassigned | 792_0 | Clustered | 35 | VC_792_0 | 35 | 0.9828 | 1 | 0.9828 | 0 | 0 | 0 | 1 | Ref |
| 6842 | MH193369 | Unassigned | Unassigned | Unassigned | 695_0 | Clustered | 48 | VC_695_0 | 48 | 0.8162 | 1 | 0.8162 | 0 | 0 | 0 | 1 | Ref |
| 6843 | MH203051 | Unassigned | Unassigned | Unassigned | 190_22 | Clustered | 3 | VC_190_22 | 3 | 0.0186 | 0.9985967 | 0.0185 | 0 | 0 | 0 | 1 | Ref |
| 6844 | MH203383 | Unassigned | Unassigned | Unassigned | 695_0 | Clustered | 48 | VC_695_0 | 48 | 0.8162 | 1 | 0.8162 | 0 | 0 | 0 | 1 | Ref |
| 6845 | MH203384 | Unassigned | Unassigned | Unassigned | 695_0 | Clustered | 48 | VC_695_0 | 48 | 0.8162 | 1 | 0.8162 | 0 | 0 | 0 | 1 | Ref |
| 6846 | MH206183 | Unassigned | Unassigned | Unassigned | 178_0 | Clustered | 5 | VC_178_0 | 5 | 0.7741 | 0.99998991 | 0.7741 | 0 | 0 | 0 | 1 | Ref |
| 6847 | MH206184 | Unassigned | Unassigned | Unassigned | 178_0 | Clustered | 5 | VC_178_0 | 5 | 0.7741 | 0.99998991 | 0.7741 | 0 | 0 | 0 | 1 | Ref |
| 6848 | MH218848 | Unassigned | Unassigned | Unassigned | 178_0 | Clustered | 5 | VC_178_0 | 5 | 0.7741 | 0.99998991 | 0.7741 | 0 | 0 | 0 | 1 | Ref |
| 6849 | MH220877 | Unassigned | Unassigned | Unassigned | 274_0 | Clustered | 2 | VC_274_0 | 2 | 0.0799 | 0.953709 | 0.0762 | 0 | 0 | 0 | 1 | Ref |
| 6850 | MH221128 | Unassigned | Unassigned | Unassigned | 959_6 | Clustered/Singleton | 1 | VC_959_6 | 1 | 0 | 1 | 0 | 0 | 0 | 0 | 1 | Ref |
| 6851 | MH221129 | Unassigned | Unassigned | Unassigned | 959_5 | Clustered | 19 | VC_959_5 | 19 | 0.2626 | 1 | 0.2626 | 0 | 0 | 0 | 1 | Ref |
| 6852 | MH229862 | Unassigned | Unassigned | Unassigned | 1353_0 | Clustered | 10 | VC_1353_0 | 10 | 1 | 1 | 1 | 0 | 0 | 0 | 1 | Ref |
| 6853 | MH229863 | Unassigned | Unassigned | Unassigned | 1353_0 | Clustered | 10 | VC_1353_0 | 10 | 1 | 1 | 1 | 0 | 0 | 0 | 1 | Ref |
| 6854 | MH229864 | Unassigned | Unassigned | Unassigned | 1353_0 | Clustered | 10 | VC_1353_0 | 10 | 1 | 1 | 1 | 0 | 0 | 0 | 1 | Ref |
| 6855 | MH229865 | Unassigned | Unassigned | Unassigned | 1353_0 | Clustered | 10 | VC_1353_0 | 10 | 1 | 1 | 1 | 0 | 0 | 0 | 1 | Ref |
| 6856 | MH230177 | Unassigned | Unassigned | Unassigned | 795_0 | Clustered | 19 | VC_795_0 | 19 | 0.6848 | 1 | 0.6848 | 0 | 0 | 0 | 1 | Ref |
| 6857 | MH230874 | Unassigned | Unassigned | Unassigned | 371_11 | Clustered | 31 | VC_371_11 | 31 | 0.0927 | 1 | 0.0927 | 0 | 0 | 0 | 0.7011 | Ref |
| 6858 | MH230875 | Unassigned | Unassigned | Unassigned | 371_3 | Clustered | 22 | VC_371_3 | 22 | 0.0787 | 1 | 0.0787 | 0 | 0 | 0 | 0.9048 | Ref |
| 6859 | MH230876 | Unassigned | Unassigned | Unassigned | 325_24 | Clustered | 3 | VC_325_24 | 3 | 0.009 | 0.99853949 | 0.009 | 0 | 0 | 0 | 1 | Ref |
| 6860 | MH230877 | Unassigned | Unassigned | Unassigned | 517_1 | Clustered | 108 | VC_517_1 | 108 | 0.8483 | 1 | 0.8483 | 0 | 0 | 0 | 0.9955 | Ref |
| 6861 | MH230878 | Unassigned | Unassigned | Unassigned | 325_52 | Clustered | 5 | VC_325_52 | 5 | 0.0147 | 0.99999073 | 0.0147 | 0 | 0 | 0 | 0.7 | Ref |
| 6862 | MH243438 | Unassigned | Unassigned | Unassigned | 286_1 | Clustered | 21 | VC_286_1 | 21 | 0.1696 | 1 | 0.1696 | 0 | 0 | 0 | 0.9048 | Ref |
| 6863 | MH243439 | Unassigned | Unassigned | Unassigned | 283_181 | Clustered/Singleton | 1 | VC_283_181 | 1 | 0 | 1 | 0 | 0 | 0 | 0 | 1 | Ref |
| 6864 | MH248069 | Unassigned | Unassigned | Unassigned | 898_0 | Clustered | 10 | VC_898_0 | 10 | 0.7284 | 1 | 0.7284 | 0 | 0 | 0 | 1 | Ref |
| 6865 | MH248138 | Unassigned | Unassigned | Unassigned | 67_3 | Clustered | 20 | VC_67_3 | 20 | 0.3542 | 1 | 0.3542 | 0 | 0 | 0 | 1 | Ref |
| 6866 | MH248947 | Unassigned | Unassigned | Unassigned | 982_0 | Clustered | 62 | VC_982_0 | 62 | 0.8646 | 1 | 0.8646 | 0 | 0 | 0 | 0.743 | Ref |
| 6867 | MH252123 | Unassigned | Unassigned | Unassigned | 847_1 | Clustered | 26 | VC_847_1 | 26 | 0.6323 | 1 | 0.6323 | 0 | 0 | 0 | 0.9815 | Ref |
| 6868 | MH252365 | Unassigned | Unassigned | Unassigned | 125_0 | Clustered | 17 | VC_125_0 | 17 | 0.9244 | 1 | 0.9244 | 0 | 0 | 0 | 1 | Ref |
| 6869 | MH268168 | Unassigned | Unassigned | Unassigned | 130_0 | Clustered | 19 | VC_130_0 | 19 | 0.6068 | 1 | 0.6068 | 0 | 0 | 0 | 1 | Ref |
| 6870 | MH271291 | Unassigned | Unassigned | Unassigned | 956_0 | Clustered | 39 | VC_956_0 | 39 | 1 | 1 | 1 | 0 | 0 | 0 | 1 | Ref |
| 6871 | MH271292 | Unassigned | Unassigned | Unassigned | 1343_0 | Clustered | 21 | VC_1343_0 | 21 | 0.9843 | 1 | 0.9843 | 0 | 0 | 0 | 1 | Ref |
| 6872 | MH271293 | Unassigned | Unassigned | Unassigned | 956_0 | Clustered | 39 | VC_956_0 | 39 | 1 | 1 | 1 | 0 | 0 | 0 | 1 | Ref |
| 6873 | MH271294 | Unassigned | Unassigned | Unassigned | 956_0 | Clustered | 39 | VC_956_0 | 39 | 1 | 1 | 1 | 0 | 0 | 0 | 1 | Ref |
| 6874 | MH271295 | Unassigned | Unassigned | Unassigned | 1175_0 | Clustered | 22 | VC_1175_0 | 22 | 0.4562 | 1 | 0.4562 | 0 | 0 | 0 | 1 | Ref |
| 6875 | MH271296 | Unassigned | Unassigned | Unassigned | 949_0 | Clustered | 10 | VC_949_0 | 10 | 0.5977 | 1 | 0.5977 | 0 | 0 | 0 | 1 | Ref |
| 6876 | MH271297 | Unassigned | Unassigned | Unassigned | 956_0 | Clustered | 39 | VC_956_0 | 39 | 1 | 1 | 1 | 0 | 0 | 0 | 1 | Ref |
| 6877 | MH271298 | Unassigned | Unassigned | Unassigned | 1206_0 | Clustered | 8 | VC_1206_0 | 8 | 0.1913 | 1 | 0.1913 | 0 | 0 | 0 | 1 | Ref |
| 6878 | MH271299 | Unassigned | Unassigned | Unassigned | 956_0 | Clustered | 39 | VC_956_0 | 39 | 1 | 1 | 1 | 0 | 0 | 0 | 1 | Ref |
| 6879 | MH271300 | Unassigned | Unassigned | Unassigned | 856_1 | Clustered | 13 | VC_856_1 | 13 | 0.5848 | 1 | 0.5848 | 0 | 0 | 0 | 1 | Ref |
| 6880 | MH271301 | Unassigned | Unassigned | Unassigned | 1343_0 | Clustered | 21 | VC_1343_0 | 21 | 0.9843 | 1 | 0.9843 | 0 | 0 | 0 | 1 | Ref |
| 6881 | MH271302 | Unassigned | Unassigned | Unassigned | 919_0 | Clustered | 39 | VC_919_0 | 39 | 0.9069 | 1 | 0.9069 | 0 | 0 | 0 | 1 | Ref |
| 6882 | MH271303 | Unassigned | Unassigned | Unassigned | 1349_0 | Clustered | 7 | VC_1349_0 | 7 | 0.2495 | 1 | 0.2495 | 0 | 0 | 0 | 1 | Ref |
| 6883 | MH271304 | Unassigned | Unassigned | Unassigned | 1352_0 | Clustered | 31 | VC_1352_0 | 31 | 0.849 | 1 | 0.849 | 0 | 0 | 0 | 1 | Ref |
| 6884 | MH271305 | Unassigned | Unassigned | Unassigned | 410_0 | Clustered | 7 | VC_410_0 | 7 | 0.7905 | 1 | 0.7905 | 0 | 0 | 0 | 1 | Ref |
| 6885 | MH271306 | Unassigned | Unassigned | Unassigned | 956_0 | Clustered | 39 | VC_956_0 | 39 | 1 | 1 | 1 | 0 | 0 | 0 | 1 | Ref |
| 6886 | MH271307 | Unassigned | Unassigned | Unassigned | 856_1 | Clustered | 13 | VC_856_1 | 13 | 0.5848 | 1 | 0.5848 | 0 | 0 | 0 | 1 | Ref |
| 6887 | MH271308 | Unassigned | Unassigned | Unassigned | 1206_0 | Clustered | 8 | VC_1206_0 | 8 | 0.1913 | 1 | 0.1913 | 0 | 0 | 0 | 1 | Ref |
| 6888 | MH271309 | Unassigned | Unassigned | Unassigned | 956_0 | Clustered | 39 | VC_956_0 | 39 | 1 | 1 | 1 | 0 | 0 | 0 | 1 | Ref |
| 6889 | MH271310 | Unassigned | Unassigned | Unassigned | 856_1 | Clustered | 13 | VC_856_1 | 13 | 0.5848 | 1 | 0.5848 | 0 | 0 | 0 | 1 | Ref |
| 6890 | MH271311 | Unassigned | Unassigned | Unassigned | 956_0 | Clustered | 39 | VC_956_0 | 39 | 1 | 1 | 1 | 0 | 0 | 0 | 1 | Ref |
| 6891 | MH271312 | Unassigned | Unassigned | Unassigned | 1352_0 | Clustered | 31 | VC_1352_0 | 31 | 0.849 | 1 | 0.849 | 0 | 0 | 0 | 1 | Ref |
| 6892 | MH271313 | Unassigned | Unassigned | Unassigned | 570_0 | Clustered | 9 | VC_570_0 | 9 | 0.499 | 1 | 0.499 | 0 | 0 | 0 | 1 | Ref |
| 6893 | MH271314 | Unassigned | Unassigned | Unassigned | 956_0 | Clustered | 39 | VC_956_0 | 39 | 1 | 1 | 1 | 0 | 0 | 0 | 1 | Ref |
| 6894 | MH271315 | Unassigned | Unassigned | Unassigned | 956_0 | Clustered | 39 | VC_956_0 | 39 | 1 | 1 | 1 | 0 | 0 | 0 | 1 | Ref |
| 6895 | MH271316 | Unassigned | Unassigned | Unassigned | 856_1 | Clustered | 13 | VC_856_1 | 13 | 0.5848 | 1 | 0.5848 | 0 | 0 | 0 | 1 | Ref |
| 6896 | MH271317 | Unassigned | Unassigned | Unassigned | 1200_0 | Clustered | 4 | VC_1200_0 | 4 | 0.8691 | 0.9999992 | 0.8691 | 0 | 0 | 0 | 1 | Ref |
| 6897 | MH271318 | Unassigned | Unassigned | Unassigned | 410_0 | Clustered | 7 | VC_410_0 | 7 | 0.7905 | 1 | 0.7905 | 0 | 0 | 0 | 1 | Ref |
| 6898 | MH271319 | Unassigned | Unassigned | Unassigned | 956_0 | Clustered | 39 | VC_956_0 | 39 | 1 | 1 | 1 | 0 | 0 | 0 | 1 | Ref |
| 6899 | MH271320 | Unassigned | Unassigned | Unassigned | 1206_0 | Clustered | 8 | VC_1206_0 | 8 | 0.1913 | 1 | 0.1913 | 0 | 0 | 0 | 1 | Ref |
| 6900 | MH271321 | Unassigned | Unassigned | Unassigned | 1352_0 | Clustered | 31 | VC_1352_0 | 31 | 0.849 | 1 | 0.849 | 0 | 0 | 0 | 1 | Ref |
| 6901 | MH285980 | Unassigned | Unassigned | Unassigned | 575_0 | Clustered | 35 | VC_575_0 | 35 | 0.8815 | 1 | 0.8815 | 0 | 0 | 0 | 1 | Ref |
| 6902 | MH299806 | Unassigned | Unassigned | Unassigned | 980_0 | Clustered | 10 | VC_980_0 | 10 | 0.9996 | 1 | 0.9996 | 0 | 0 | 0 | 1 | Ref |
| 6903 | MH310933 | Unassigned | Unassigned | Unassigned | 495_0 | Clustered | 39 | VC_495_0 | 39 | 0.308 | 1 | 0.308 | 0 | 0 | 0 | 1 | Ref |
| 6904 | MH310934 | Unassigned | Unassigned | Unassigned | 495_1 | Clustered | 41 | VC_495_1 | 41 | 0.328 | 1 | 0.328 | 0 | 0 | 0 | 0.911 | Ref |
| 6905 | MH310935 | Unassigned | Unassigned | Unassigned | 495_0 | Clustered | 39 | VC_495_0 | 39 | 0.308 | 1 | 0.308 | 0 | 0 | 0 | 1 | Ref |
| 6906 | MH310936 | Unassigned | Unassigned | Unassigned | 495_1 | Clustered | 41 | VC_495_1 | 41 | 0.328 | 1 | 0.328 | 0 | 0 | 0 | 0.911 | Ref |
| 6907 | MH316561 | Unassigned | Unassigned | Unassigned | 558_14 | Clustered | 8 | VC_558_14 | 8 | 0.0228 | 0.99945246 | 0.0228 | 0 | 0 | 0 | 0.8214 | Ref |
| 6908 | MH316562 | Unassigned | Unassigned | Unassigned | 565_0 | Clustered | 20 | VC_565_0 | 20 | 0.8398 | 1 | 0.8398 | 0 | 0 | 0 | 1 | Ref |
| 6909 | MH316563 | Unassigned | Unassigned | Unassigned | 513_0 | Clustered | 32 | VC_513_0 | 32 | 0.6881 | 1 | 0.6881 | 0 | 0 | 0 | 1 | Ref |
| 6910 | MH316564 | Unassigned | Unassigned | Unassigned | 520_45 | Clustered | 7 | VC_520_45 | 7 | 0.0635 | 1 | 0.0635 | 0 | 0 | 0 | 0.8095 | Ref |
| 6911 | MH316565 | Unassigned | Unassigned | Unassigned | 509_0 | Clustered | 37 | VC_509_0 | 37 | 0.9689 | 1 | 0.9689 | 0 | 0 | 0 | 1 | Ref |
| 6912 | MH316566 | Unassigned | Unassigned | Unassigned | 224_0 | Clustered | 19 | VC_224_0 | 19 | 0.9416 | 1 | 0.9416 | 0 | 0 | 0 | 1 | Ref |
| 6913 | MH316567 | Unassigned | Unassigned | Unassigned | 558_50 | Clustered | 15 | VC_558_50 | 15 | 0.0468 | 1 | 0.0468 | 0 | 0 | 0 | 0.9238 | Ref |
| 6914 | MH316568 | Unassigned | Unassigned | Unassigned | 371_13 | Clustered | 33 | VC_371_13 | 33 | 0.179 | 1 | 0.179 | 0 | 0 | 0 | 0.8769 | Ref |
| 6915 | MH316569 | Unassigned | Unassigned | Unassigned | 956_0 | Clustered | 39 | VC_956_0 | 39 | 1 | 1 | 1 | 0 | 0 | 0 | 1 | Ref |
| 6916 | MH316570 | Unassigned | Unassigned | Unassigned | 521_0 | Clustered | 37 | VC_521_0 | 37 | 0.912 | 1 | 0.912 | 0 | 0 | 0 | 1 | Ref |
| 6917 | MH319720 | Unassigned | Unassigned | Unassigned | 438_0 | Clustered | 6 | VC_438_0 | 6 | 0.0917 | 0.99994271 | 0.0917 | 0 | 0 | 0 | 1 | Ref |
| 6918 | MH319733 | Unassigned | Unassigned | Unassigned | 438_0 | Clustered | 6 | VC_438_0 | 6 | 0.0917 | 0.99994271 | 0.0917 | 0 | 0 | 0 | 1 | Ref |
| 6919 | MH319737 | Unassigned | Unassigned | Unassigned | 659_0 | Clustered | 2 | VC_659_0 | 2 | 0.0254 | 0.54598917 | 0.0139 | 0 | 0 | 0 | 1 | Ref |
| 6920 | MH319742 | Unassigned | Unassigned | Unassigned | 438_0 | Clustered | 6 | VC_438_0 | 6 | 0.0917 | 0.99994271 | 0.0917 | 0 | 0 | 0 | 1 | Ref |
| 6921 | MH319746 | Unassigned | Unassigned | Unassigned | 659_0 | Clustered | 2 | VC_659_0 | 2 | 0.0254 | 0.54598917 | 0.0139 | 0 | 0 | 0 | 1 | Ref |
| 6922 | MH319749 | Unassigned | Unassigned | Unassigned | 432_6 | Clustered/Singleton | 1 | VC_432_6 | 1 | 0 | 1 | 0 | 0 | 0 | 0 | 1 | Ref |
| 6923 | MH319750 | Unassigned | Unassigned | Unassigned | 439_0 | Clustered | 2 | VC_439_0 | 2 | 0.0291 | 0.90533848 | 0.0263 | 0 | 0 | 0 | 1 | Ref |
| 6924 | MH319753 | Unassigned | Unassigned | Unassigned | 438_0 | Clustered | 6 | VC_438_0 | 6 | 0.0917 | 0.99994271 | 0.0917 | 0 | 0 | 0 | 1 | Ref |
| 6925 | MH319754 | Unassigned | Unassigned | Unassigned | 432_7 | Clustered/Singleton | 1 | VC_432_7 | 1 | 0 | 1 | 0 | 0 | 0 | 0 | 1 | Ref |
| 6926 | MH319758 | Unassigned | Unassigned | Unassigned | 438_0 | Clustered | 6 | VC_438_0 | 6 | 0.0917 | 0.99994271 | 0.0917 | 0 | 0 | 0 | 1 | Ref |
| 6927 | MH319759 | Unassigned | Unassigned | Unassigned | 439_0 | Clustered | 2 | VC_439_0 | 2 | 0.0291 | 0.90533848 | 0.0263 | 0 | 0 | 0 | 1 | Ref |
| 6928 | MH321490 | Unassigned | Unassigned | Unassigned | 150_0 | Clustered | 13 | VC_150_0 | 13 | 0.2417 | 1 | 0.2417 | 0 | 0 | 0 | 0.9872 | Ref |
| 6929 | MH321491 | Unassigned | Unassigned | Unassigned | 150_0 | Clustered | 13 | VC_150_0 | 13 | 0.2417 | 1 | 0.2417 | 0 | 0 | 0 | 0.9872 | Ref |
| 6930 | MH321492 | Unassigned | Unassigned | Unassigned | 9_0 | Clustered | 38 | VC_9_0 | 38 | 0.4895 | 1 | 0.4895 | 0 | 0 | 0 | 1 | Ref |
| 6931 | MH321493 | Unassigned | Unassigned | Unassigned | 461_0 | Clustered | 96 | VC_461_0 | 96 | 0.9307 | 1 | 0.9307 | 0 | 0 | 0 | 0.9575 | Ref |
| 6932 | MH327487 | Unassigned | Unassigned | Unassigned | 438_0 | Clustered | 6 | VC_438_0 | 6 | 0.0917 | 0.99994271 | 0.0917 | 0 | 0 | 0 | 1 | Ref |
| 6933 | MH333064 | Unassigned | Unassigned | Unassigned | 491_0 | Clustered | 36 | VC_491_0 | 36 | 0.5336 | 1 | 0.5336 | 0 | 0 | 0 | 0.9714 | Ref |
| 6934 | MH338234 | Unassigned | Unassigned | Unassigned | 216_6 | Clustered | 42 | VC_216_6 | 42 | 0.473 | 1 | 0.473 | 0 | 0 | 0 | 0.8351 | Ref |
| 6935 | MH338235 | Unassigned | Unassigned | Unassigned | 940_0 | Clustered | 34 | VC_940_0 | 34 | 0.7959 | 1 | 0.7959 | 0 | 0 | 0 | 1 | Ref |
| 6936 | MH338236 | Unassigned | Unassigned | Unassigned | 221_0 | Clustered | 22 | VC_221_0 | 22 | 0.7447 | 1 | 0.7447 | 0 | 0 | 0 | 1 | Ref |
| 6937 | MH338237 | Unassigned | Unassigned | Unassigned | 221_0 | Clustered | 22 | VC_221_0 | 22 | 0.7447 | 1 | 0.7447 | 0 | 0 | 0 | 1 | Ref |
| 6938 | MH338238 | Unassigned | Unassigned | Unassigned | 325_19 | Clustered | 3 | VC_325_19 | 3 | 0.0108 | 0.99859221 | 0.0107 | 0 | 0 | 0 | 1 | Ref |
| 6939 | MH338239 | Unassigned | Unassigned | Unassigned | 325_60 | Clustered | 2 | VC_325_60 | 2 | 0.0038 | 0.957513 | 0.0036 | 0 | 0 | 0 | 1 | Ref |
| 6940 | MH338240 | Unassigned | Unassigned | Unassigned | 514_0 | Clustered | 75 | VC_514_0 | 75 | 0.9402 | 1 | 0.9402 | 0 | 0 | 0 | 1 | Ref |
| 6941 | MH338241 | Unassigned | Unassigned | Unassigned | 940_0 | Clustered | 34 | VC_940_0 | 34 | 0.7959 | 1 | 0.7959 | 0 | 0 | 0 | 1 | Ref |
| 6942 | MH341451 | Unassigned | Unassigned | Unassigned | 380_0 | Clustered | 3 | VC_380_0 | 3 | 0.1631 | 0.99675097 | 0.1625 | 0 | 0 | 0 | 1 | Ref |
| 6943 | MH341452 | Unassigned | Unassigned | Unassigned | 378_0 | Clustered | 9 | VC_378_0 | 9 | 0.7603 | 1 | 0.7603 | 0 | 0 | 0 | 1 | Ref |
| 6944 | MH341453 | Unassigned | Unassigned | Unassigned | 140_0 | Clustered | 2 | VC_140_0 | 2 | 0.4111 | 0.94824791 | 0.3898 | 0 | 0 | 0 | 1 | Ref |
| 6945 | MH355583 | Unassigned | Unassigned | Unassigned | 695_0 | Clustered | 48 | VC_695_0 | 48 | 0.8162 | 1 | 0.8162 | 0 | 0 | 0 | 1 | Ref |
| 6946 | MH355584 | Unassigned | Unassigned | Unassigned | 283_45 | Clustered/Singleton | 1 | VC_283_45 | 1 | 0 | 1 | 0 | 0 | 0 | 0 | 1 | Ref |
| 6947 | MH356729 | Unassigned | Unassigned | Unassigned | 838_0 | Clustered | 29 | VC_838_0 | 29 | 0.8611 | 1 | 0.8611 | 0 | 0 | 0 | 1 | Ref |
| 6948 | MH356730 | Unassigned | Unassigned | Unassigned | 667_1 | Clustered/Singleton | 1 | VC_667_1 | 1 | 0 | 1 | 0 | 0 | 0 | 0 | 1 | Ref |
| 6949 | MH358359 | Unassigned | Unassigned | Unassigned | 637_0 | Clustered | 115 | VC_637_0 | 115 | 0.731 | 1 | 0.731 | 0 | 0 | 0 | 0.8764 | Ref |
| 6950 | MH358458 | Unassigned | Unassigned | Unassigned | 779_0 | Clustered | 37 | VC_779_0 | 37 | 0.5682 | 1 | 0.5682 | 0 | 0 | 0 | 1 | Ref |
| 6951 | MH359124 | Unassigned | Unassigned | Unassigned | 286_11 | Clustered | 3 | VC_286_11 | 3 | 0.0146 | 0.93460284 | 0.0137 | 0 | 0 | 0 | 1 | Ref |
| 6952 | MH363700 | Unassigned | Unassigned | Unassigned | 134_0 | Clustered | 3 | VC_134_0 | 3 | 0.0956 | 0.99858494 | 0.0955 | 0 | 0 | 0 | 1 | Ref |
| 6953 | MH363708 | Unassigned | Unassigned | Unassigned | 705_0 | Clustered | 11 | VC_705_0 | 11 | 0.8861 | 1 | 0.8861 | 0 | 0 | 0 | 1 | Ref |
| 6954 | MH370358 | Unassigned | Unassigned | Unassigned | 637_0 | Clustered | 115 | VC_637_0 | 115 | 0.731 | 1 | 0.731 | 0 | 0 | 0 | 0.8764 | Ref |
| 6955 | MH370359 | Unassigned | Unassigned | Unassigned | 637_0 | Clustered | 115 | VC_637_0 | 115 | 0.731 | 1 | 0.731 | 0 | 0 | 0 | 0.8764 | Ref |
| 6956 | MH370360 | Unassigned | Unassigned | Unassigned | 637_0 | Clustered | 115 | VC_637_0 | 115 | 0.731 | 1 | 0.731 | 0 | 0 | 0 | 0.8764 | Ref |
| 6957 | MH370361 | Unassigned | Unassigned | Unassigned | 637_0 | Clustered | 115 | VC_637_0 | 115 | 0.731 | 1 | 0.731 | 0 | 0 | 0 | 0.8764 | Ref |
| 6958 | MH370362 | Unassigned | Unassigned | Unassigned | 637_0 | Clustered | 115 | VC_637_0 | 115 | 0.731 | 1 | 0.731 | 0 | 0 | 0 | 0.8764 | Ref |
| 6959 | MH370363 | Unassigned | Unassigned | Unassigned | 637_0 | Clustered | 115 | VC_637_0 | 115 | 0.731 | 1 | 0.731 | 0 | 0 | 0 | 0.8764 | Ref |
| 6960 | MH370365 | Unassigned | Unassigned | Unassigned | 637_0 | Clustered | 115 | VC_637_0 | 115 | 0.731 | 1 | 0.731 | 0 | 0 | 0 | 0.8764 | Ref |
| 6961 | MH370366 | Unassigned | Unassigned | Unassigned | 191_4 | Clustered | 2 | VC_191_4 | 2 | 0.0055 | 0.88936514 | 0.0049 | 0 | 0 | 0 | 1 | Ref |
| 6962 | MH370367 | Unassigned | Unassigned | Unassigned | 191_9 | Clustered | 5 | VC_191_9 | 5 | 0.0228 | 0.99999662 | 0.0228 | 0 | 0 | 0 | 0.7 | Ref |
| 6963 | MH370368 | Unassigned | Unassigned | Unassigned | 133_3 | Clustered | 3 | VC_133_3 | 3 | 0.0105 | 0.99119575 | 0.0104 | 0 | 0 | 0 | 1 | Ref |
| 6964 | MH370369 | Unassigned | Unassigned | Unassigned | 191_16 | Clustered | 12 | VC_191_16 | 12 | 0.0643 | 1 | 0.0643 | 0 | 0 | 0 | 0.9394 | Ref |

| | | | | | | | | | | | | | | | | | |
|---|---|---|---|---|---|---|---|---|---|---|---|---|---|---|---|---|---|
| 6965 | MH370370 | Unassigned | Unassigned | Unassigned | 133_31 | Clustered | 6 | VC_133_31 | 6 | 0.0263 | 0.99999694 | 0.0263 | 0 | 0 | 0 | 1 | Ref |
| 6966 | MH370371 | Unassigned | Unassigned | Unassigned | 133_3 | Clustered | 3 | VC_133_3 | 3 | 0.0105 | 0.99119575 | 0.0104 | 0 | 0 | 0 | 1 | Ref |
| 6967 | MH370372 | Unassigned | Unassigned | Unassigned | 637_0 | Clustered | 115 | VC_637_0 | 115 | 0.731 | 1 | 0.731 | 0 | 0 | 0 | 0.8764 | Ref |
| 6968 | MH370373 | Unassigned | Unassigned | Unassigned | 637_0 | Clustered | 115 | VC_637_0 | 115 | 0.731 | 1 | 0.731 | 0 | 0 | 0 | 0.8764 | Ref |
| 6969 | MH370374 | Unassigned | Unassigned | Unassigned | 637_0 | Clustered | 115 | VC_637_0 | 115 | 0.731 | 1 | 0.731 | 0 | 0 | 0 | 0.8764 | Ref |
| 6970 | MH370375 | Unassigned | Unassigned | Unassigned | 191_29 | Clustered/Singleton | 1 | VC_191_29 | 1 | 0 | 1 | 0 | 0 | 0 | 0 | 1 | Ref |
| 6971 | MH370376 | Unassigned | Unassigned | Unassigned | 191_8 | Clustered | 2 | VC_191_8 | 2 | 0.0055 | 0.91831352 | 0.0051 | 0 | 0 | 0 | 1 | Ref |
| 6972 | MH370377 | Unassigned | Unassigned | Unassigned | 190_13 | Clustered | 4 | VC_190_13 | 4 | 0.0216 | 0.99997041 | 0.0216 | 0 | 0 | 0 | 0.6667 | Ref |
| 6973 | MH370378 | Unassigned | Unassigned | Unassigned | 190_13 | Clustered | 4 | VC_190_13 | 4 | 0.0216 | 0.99997041 | 0.0216 | 0 | 0 | 0 | 0.6667 | Ref |
| 6974 | MH370379 | Unassigned | Unassigned | Unassigned | 191_23 | Clustered/Singleton | 1 | VC_191_23 | 1 | 0 | 1 | 0 | 0 | 0 | 0 | 1 | Ref |
| 6975 | MH370380 | Unassigned | Unassigned | Unassigned | 191_9 | Clustered | 5 | VC_191_9 | 5 | 0.0228 | 0.99999662 | 0.0228 | 0 | 0 | 0 | 0.7 | Ref |
| 6976 | MH370381 | Unassigned | Unassigned | Unassigned | 637_0 | Clustered | 115 | VC_637_0 | 115 | 0.731 | 1 | 0.731 | 0 | 0 | 0 | 0.8764 | Ref |
| 6977 | MH370384 | Unassigned | Unassigned | Unassigned | 637_0 | Clustered | 115 | VC_637_0 | 115 | 0.731 | 1 | 0.731 | 0 | 0 | 0 | 0.8764 | Ref |
| 6978 | MH370385 | Unassigned | Unassigned | Unassigned | 637_0 | Clustered | 115 | VC_637_0 | 115 | 0.731 | 1 | 0.731 | 0 | 0 | 0 | 0.8764 | Ref |
| 6979 | MH370386 | Unassigned | Unassigned | Unassigned | 191_5 | Clustered | 6 | VC_191_5 | 6 | 0.0285 | 0.99999996 | 0.0285 | 0 | 0 | 0 | 1 | Ref |
| 6980 | MH370387 | Unassigned | Unassigned | Unassigned | 28_1 | Clustered | 53 | VC_28_1 | 53 | 0.7164 | 1 | 0.7164 | 0 | 0 | 0 | 0.9325 | Ref |
| 6981 | MH370477 | Unassigned | Unassigned | Unassigned | 394_0 | Clustered | 2 | VC_394_0 | 2 | 0.0211 | 0.957922 | 0.0202 | 0 | 0 | 0 | 1 | Ref |
| 6982 | MH370478 | Unassigned | Unassigned | Unassigned | 76_0 | Clustered | 37 | VC_76_0 | 37 | 0.9403 | 1 | 0.9403 | 0 | 0 | 0 | 1 | Ref |
| 6983 | MH371107 | Unassigned | Unassigned | Unassigned | 371_13 | Clustered | 33 | VC_371_13 | 33 | 0.179 | 1 | 0.179 | 0 | 0 | 0 | 0.8769 | Ref |
| 6984 | MH371108 | Unassigned | Unassigned | Unassigned | 856_0 | Clustered | 7 | VC_856_0 | 7 | 0.3463 | 1 | 0.3463 | 0 | 0 | 0 | 1 | Ref |
| 6985 | MH371109 | Unassigned | Unassigned | Unassigned | 856_0 | Clustered | 7 | VC_856_0 | 7 | 0.3463 | 1 | 0.3463 | 0 | 0 | 0 | 1 | Ref |
| 6986 | MH371110 | Unassigned | Unassigned | Unassigned | 325_36 | Clustered | 2 | VC_325_36 | 2 | 0.0043 | 0.95751115 | 0.0041 | 0 | 0 | 0 | 1 | Ref |
| 6987 | MH371111 | Unassigned | Unassigned | Unassigned | 1344_0 | Clustered | 78 | VC_1344_0 | 78 | 0.9732 | 1 | 0.9732 | 0 | 0 | 0 | 1 | Ref |
| 6988 | MH371112 | Unassigned | Unassigned | Unassigned | 551_6 | Clustered | 22 | VC_551_6 | 22 | 0.1214 | 1 | 0.1214 | 0 | 0 | 0 | 0.8095 | Ref |
| 6989 | MH371113 | Unassigned | Unassigned | Unassigned | 236_0 | Clustered | 72 | VC_236_0 | 72 | 0.8104 | 1 | 0.8104 | 0 | 0 | 0 | 1 | Ref |
| 6990 | MH371114 | Unassigned | Unassigned | Unassigned | 371_16 | Clustered | 42 | VC_371_16 | 42 | 0.1341 | 1 | 0.1341 | 0 | 0 | 0 | 0.7282 | Ref |
| 6991 | MH371115 | Unassigned | Unassigned | Unassigned | 520_40 | Clustered | 4 | VC_520_40 | 4 | 0.0396 | 0.99998776 | 0.0396 | 0 | 0 | 0 | 1 | Ref |
| 6992 | MH371116 | Unassigned | Unassigned | Unassigned | 761_0 | Clustered | 67 | VC_761_0 | 67 | 0.8947 | 1 | 0.8947 | 0 | 0 | 0 | 1 | Ref |
| 6993 | MH371117 | Unassigned | Unassigned | Unassigned | 371_16 | Clustered | 42 | VC_371_16 | 42 | 0.1341 | 1 | 0.1341 | 0 | 0 | 0 | 0.7282 | Ref |
| 6994 | MH371118 | Unassigned | Unassigned | Unassigned | 1344_0 | Clustered | 78 | VC_1344_0 | 78 | 0.9732 | 1 | 0.9732 | 0 | 0 | 0 | 1 | Ref |
| 6995 | MH371119 | Unassigned | Unassigned | Unassigned | 761_0 | Clustered | 67 | VC_761_0 | 67 | 0.8947 | 1 | 0.8947 | 0 | 0 | 0 | 1 | Ref |
| 6996 | MH371120 | Unassigned | Unassigned | Unassigned | 520_39 | Clustered/Singleton | 1 | VC_520_39 | 1 | 0 | 1 | 0 | 0 | 0 | 0 | 1 | Ref |
| 6997 | MH371121 | Unassigned | Unassigned | Unassigned | 514_0 | Clustered | 75 | VC_514_0 | 75 | 0.9402 | 1 | 0.9402 | 0 | 0 | 0 | 1 | Ref |
| 6998 | MH371122 | Unassigned | Unassigned | Unassigned | 225_0 | Clustered | 32 | VC_225_0 | 32 | 0.848 | 1 | 0.848 | 0 | 0 | 0 | 0.996 | Ref |
| 6999 | MH371123 | Unassigned | Unassigned | Unassigned | 520_33 | Clustered | 2 | VC_520_33 | 2 | 0.0087 | 0.95761571 | 0.0083 | 0 | 0 | 0 | 1 | Ref |
| 7000 | MH371124 | Unassigned | Unassigned | Unassigned | 1344_0 | Clustered | 78 | VC_1344_0 | 78 | 0.9732 | 1 | 0.9732 | 0 | 0 | 0 | 1 | Ref |
| 7001 | MH371125 | Unassigned | Unassigned | Unassigned | 371_16 | Clustered | 42 | VC_371_16 | 42 | 0.1341 | 1 | 0.1341 | 0 | 0 | 0 | 0.7282 | Ref |
| 7002 | MH375074 | Unassigned | Unassigned | Unassigned | 695_0 | Clustered | 48 | VC_695_0 | 48 | 0.8162 | 1 | 0.8162 | 0 | 0 | 0 | 1 | Ref |
| 7003 | MH375593 | Unassigned | Unassigned | Unassigned | 254_0 | Clustered | 46 | VC_254_0 | 46 | 0.5159 | 1 | 0.5159 | 0 | 0 | 0 | 1 | Ref |
| 7004 | MH375594 | Unassigned | Unassigned | Unassigned | 252_0 | Clustered | 121 | VC_252_0 | 121 | 0.7376 | 1 | 0.7376 | 0 | 0 | 0 | 0.9999 | Ref |
| 7005 | MH375595 | Unassigned | Unassigned | Unassigned | 252_0 | Clustered | 121 | VC_252_0 | 121 | 0.7376 | 1 | 0.7376 | 0 | 0 | 0 | 0.9999 | Ref |
| 7006 | MH375596 | Unassigned | Unassigned | Unassigned | 252_0 | Clustered | 121 | VC_252_0 | 121 | 0.7376 | 1 | 0.7376 | 0 | 0 | 0 | 0.9999 | Ref |
| 7007 | MH375597 | Unassigned | Unassigned | Unassigned | 252_0 | Clustered | 121 | VC_252_0 | 121 | 0.7376 | 1 | 0.7376 | 0 | 0 | 0 | 0.9999 | Ref |
| 7008 | MH375598 | Unassigned | Unassigned | Unassigned | 252_0 | Clustered | 121 | VC_252_0 | 121 | 0.7376 | 1 | 0.7376 | 0 | 0 | 0 | 0.9999 | Ref |
| 7009 | MH375599 | Unassigned | Unassigned | Unassigned | 252_0 | Clustered | 121 | VC_252_0 | 121 | 0.7376 | 1 | 0.7376 | 0 | 0 | 0 | 0.9999 | Ref |
| 7010 | MH375600 | Unassigned | Unassigned | Unassigned | 252_0 | Clustered | 121 | VC_252_0 | 121 | 0.7376 | 1 | 0.7376 | 0 | 0 | 0 | 0.9999 | Ref |
| 7011 | MH375644 | Unassigned | Unassigned | Unassigned | 136_0 | Clustered | 2 | VC_136_0 | 2 | 0.0412 | 0.95741268 | 0.0394 | 0 | 0 | 0 | 1 | Ref |
| 7012 | MH378441 | Unassigned | Unassigned | Unassigned | 303_0 | Clustered | 155 | VC_303_0 | 155 | 0.9979 | 1 | 0.9979 | 0 | 0 | 0 | 1 | Ref |
| 7013 | MH378442 | Unassigned | Unassigned | Unassigned | 303_0 | Clustered | 155 | VC_303_0 | 155 | 0.9979 | 1 | 0.9979 | 0 | 0 | 0 | 1 | Ref |
| 7014 | MH378443 | Unassigned | Unassigned | Unassigned | 303_0 | Clustered | 155 | VC_303_0 | 155 | 0.9979 | 1 | 0.9979 | 0 | 0 | 0 | 1 | Ref |
| 7015 | MH382198 | Unassigned | Unassigned | Unassigned | 392_0 | Clustered | 126 | VC_392_0 | 126 | 0.9621 | 1 | 0.9621 | 0 | 0 | 0 | 1 | Ref |
| 7016 | MH383160 | Unassigned | Unassigned | Unassigned | 164_11 | Clustered | 10 | VC_164_11 | 10 | 0.1507 | 1 | 0.1507 | 0 | 0 | 0 | 0.9111 | Ref |
| 7017 | MH384259 | Unassigned | Unassigned | Unassigned | 10_0 | Clustered | 25 | VC_10_0 | 25 | 0.3467 | 1 | 0.3467 | 0 | 0 | 0 | 0.9733 | Ref |
| 7018 | MH384260 | Unassigned | Unassigned | Unassigned | 7_0 | Clustered | 28 | VC_7_0 | 28 | 0.3916 | 1 | 0.3916 | 0 | 0 | 0 | 0.9947 | Ref |
| 7019 | MH384261 | Unassigned | Unassigned | Unassigned | 8_0 | Clustered | 58 | VC_8_0 | 58 | 0.8306 | 1 | 0.8306 | 0 | 0 | 0 | 0.9946 | Ref |
| 7020 | MH389777 | Unassigned | Unassigned | Unassigned | 67_4 | Clustered | 4 | VC_67_4 | 4 | 0.0431 | 0.90943035 | 0.0392 | 0 | 0 | 0 | 1 | Ref |
| 7021 | MH393889 | Unassigned | Unassigned | Unassigned | 899_0 | Clustered | 4 | VC_899_0 | 4 | 0.2655 | 0.99997104 | 0.2655 | 0 | 0 | 0 | 1 | Ref |
| 7022 | MH399771 | Unassigned | Unassigned | Unassigned | 520_42 | Clustered/Singleton | 1 | VC_520_42 | 1 | 0 | 1 | 0 | 0 | 0 | 0 | 1 | Ref |
| 7023 | MH399772 | Unassigned | Unassigned | Unassigned | 546_0 | Clustered | 2 | VC_546_0 | 2 | 0.0136 | 0.63748598 | 0.0087 | 0 | 0 | 0 | 1 | Ref |
| 7024 | MH399773 | Unassigned | Unassigned | Unassigned | 371_16 | Clustered | 42 | VC_371_16 | 42 | 0.1341 | 1 | 0.1341 | 0 | 0 | 0 | 0.7282 | Ref |
| 7025 | MH399774 | Unassigned | Unassigned | Unassigned | 551_0 | Clustered | 16 | VC_551_0 | 16 | 0.0915 | 1 | 0.0915 | 0 | 0 | 0 | 0.6667 | Ref |
| 7026 | MH399775 | Unassigned | Unassigned | Unassigned | 371_15 | Clustered | 44 | VC_371_15 | 44 | 0.1391 | 1 | 0.1391 | 0 | 0 | 0 | 0.7474 | Ref |
| 7027 | MH399776 | Unassigned | Unassigned | Unassigned | 514_0 | Clustered | 75 | VC_514_0 | 75 | 0.9402 | 1 | 0.9402 | 0 | 0 | 0 | 1 | Ref |
| 7028 | MH399777 | Unassigned | Unassigned | Unassigned | 517_1 | Clustered | 108 | VC_517_1 | 108 | 0.8483 | 1 | 0.8483 | 0 | 0 | 0 | 0.9955 | Ref |
| 7029 | MH399778 | Unassigned | Unassigned | Unassigned | 551_0 | Clustered | 16 | VC_551_0 | 16 | 0.0915 | 1 | 0.0915 | 0 | 0 | 0 | 0.6667 | Ref |
| 7030 | MH399779 | Unassigned | Unassigned | Unassigned | 856_1 | Clustered | 13 | VC_856_1 | 13 | 0.5848 | 1 | 0.5848 | 0 | 0 | 0 | 1 | Ref |
| 7031 | MH399780 | Unassigned | Unassigned | Unassigned | 371_16 | Clustered | 42 | VC_371_16 | 42 | 0.1341 | 1 | 0.1341 | 0 | 0 | 0 | 0.7282 | Ref |
| 7032 | MH399781 | Unassigned | Unassigned | Unassigned | 954_0 | Clustered | 13 | VC_954_0 | 13 | 0.863 | 1 | 0.863 | 0 | 0 | 0 | 1 | Ref |
| 7033 | MH399782 | Unassigned | Unassigned | Unassigned | 325_53 | Clustered | 7 | VC_325_53 | 7 | 0.0242 | 1 | 0.0242 | 0 | 0 | 0 | 0.9048 | Ref |
| 7034 | MH399783 | Unassigned | Unassigned | Unassigned | 1344_0 | Clustered | 78 | VC_1344_0 | 78 | 0.9732 | 1 | 0.9732 | 0 | 0 | 0 | 1 | Ref |
| 7035 | MH399784 | Unassigned | Unassigned | Unassigned | 520_45 | Clustered | 7 | VC_520_45 | 7 | 0.0635 | 1 | 0.0635 | 0 | 0 | 0 | 0.8095 | Ref |
| 7036 | MH399785 | Unassigned | Unassigned | Unassigned | 517_1 | Clustered | 108 | VC_517_1 | 108 | 0.8483 | 1 | 0.8483 | 0 | 0 | 0 | 0.9955 | Ref |
| 7037 | MH399786 | Unassigned | Unassigned | Unassigned | 371_15 | Clustered | 44 | VC_371_15 | 44 | 0.1391 | 1 | 0.1391 | 0 | 0 | 0 | 0.7474 | Ref |
| 7038 | MH399787 | Unassigned | Unassigned | Unassigned | 521_0 | Clustered | 37 | VC_521_0 | 37 | 0.912 | 1 | 0.912 | 0 | 0 | 0 | 1 | Ref |
| 7039 | MH399788 | Unassigned | Unassigned | Unassigned | 565_0 | Clustered | 20 | VC_565_0 | 20 | 0.8398 | 1 | 0.8398 | 0 | 0 | 0 | 1 | Ref |
| 7040 | MH399789 | Unassigned | Unassigned | Unassigned | 1164_0 | Clustered | 25 | VC_1164_0 | 25 | 0.7312 | 1 | 0.7312 | 0 | 0 | 0 | 1 | Ref |
| 7041 | MH400309 | Unassigned | Unassigned | Unassigned | 451_0 | Clustered | 80 | VC_451_0 | 80 | 0.9991 | 1 | 0.9991 | 0 | 0 | 0 | 1 | Ref |
| 7042 | MH401414 | Unassigned | Unassigned | Unassigned | 7_0 | Clustered | 28 | VC_7_0 | 28 | 0.3916 | 1 | 0.3916 | 0 | 0 | 0 | 0.9947 | Ref |
| 7043 | MH401415 | Unassigned | Unassigned | Unassigned | 7_0 | Clustered | 28 | VC_7_0 | 28 | 0.3916 | 1 | 0.3916 | 0 | 0 | 0 | 0.9947 | Ref |
| 7044 | MH401416 | Unassigned | Unassigned | Unassigned | 10_0 | Clustered | 25 | VC_10_0 | 25 | 0.3467 | 1 | 0.3467 | 0 | 0 | 0 | 0.9733 | Ref |
| 7045 | MH412654 | Unassigned | Unassigned | Unassigned | 432_1 | Clustered/Singleton | 1 | VC_432_1 | 1 | 0 | 1 | 0 | 0 | 0 | 0 | 1 | Ref |
| 7046 | MH422554 | Unassigned | Unassigned | Unassigned | 247_0 | Clustered | 14 | VC_247_0 | 14 | 0.9119 | 1 | 0.9119 | 0 | 0 | 0 | 0.967 | Ref |
| 7047 | MH424443 | Unassigned | Unassigned | Unassigned | 637_0 | Clustered | 115 | VC_637_0 | 115 | 0.731 | 1 | 0.731 | 0 | 0 | 0 | 0.8764 | Ref |
| 7048 | MH424444 | Unassigned | Unassigned | Unassigned | 540_0 | Clustered | 15 | VC_540_0 | 15 | 0.2866 | 1 | 0.2866 | 0 | 0 | 0 | 1 | Ref |
| 7049 | MH424445 | Unassigned | Unassigned | Unassigned | 637_0 | Clustered | 115 | VC_637_0 | 115 | 0.731 | 1 | 0.731 | 0 | 0 | 0 | 0.8764 | Ref |
| 7050 | MH424446 | Unassigned | Unassigned | Unassigned | 461_0 | Clustered | 96 | VC_461_0 | 96 | 0.9307 | 1 | 0.9307 | 0 | 0 | 0 | 0.9575 | Ref |
| 7051 | MH426724 | Unassigned | Unassigned | Unassigned | 959_3 | Clustered | 3 | VC_959_3 | 3 | 0.0237 | 0.75911548 | 0.018 | 0 | 0 | 0 | 1 | Ref |
| 7052 | MH426725 | Unassigned | Unassigned | Unassigned | 328_0 | Clustered | 11 | VC_328_0 | 11 | 0.4707 | 1 | 0.4707 | 0 | 0 | 0 | 1 | Ref |
| 7053 | MH426726 | Unassigned | Unassigned | Unassigned | 359_0 | Clustered | 7 | VC_359_0 | 7 | 0.3703 | 1 | 0.3703 | 0 | 0 | 0 | 1 | Ref |
| 7054 | MH427377 | Unassigned | Unassigned | Unassigned | 133_22 | Clustered | 13 | VC_133_22 | 13 | 0.0804 | 1 | 0.0804 | 0 | 0 | 0 | 0.8205 | Ref |
| 7055 | MH431931 | Unassigned | Unassigned | Unassigned | 708_0 | Clustered | 47 | VC_708_0 | 47 | 0.8654 | 1 | 0.8654 | 0 | 0 | 0 | 1 | Ref |
| 7056 | MH431932 | Unassigned | Unassigned | Unassigned | 708_0 | Clustered | 47 | VC_708_0 | 47 | 0.8654 | 1 | 0.8654 | 0 | 0 | 0 | 1 | Ref |
| 7057 | MH431933 | Unassigned | Unassigned | Unassigned | 708_0 | Clustered | 47 | VC_708_0 | 47 | 0.8654 | 1 | 0.8654 | 0 | 0 | 0 | 1 | Ref |
| 7058 | MH431934 | Unassigned | Unassigned | Unassigned | 708_0 | Clustered | 47 | VC_708_0 | 47 | 0.8654 | 1 | 0.8654 | 0 | 0 | 0 | 1 | Ref |
| 7059 | MH431935 | Unassigned | Unassigned | Unassigned | 708_0 | Clustered | 47 | VC_708_0 | 47 | 0.8654 | 1 | 0.8654 | 0 | 0 | 0 | 1 | Ref |
| 7060 | MH431936 | Unassigned | Unassigned | Unassigned | 708_0 | Clustered | 47 | VC_708_0 | 47 | 0.8654 | 1 | 0.8654 | 0 | 0 | 0 | 1 | Ref |
| 7061 | MH431937 | Unassigned | Unassigned | Unassigned | 708_0 | Clustered | 47 | VC_708_0 | 47 | 0.8654 | 1 | 0.8654 | 0 | 0 | 0 | 1 | Ref |
| 7062 | MH431938 | Unassigned | Unassigned | Unassigned | 1156_0 | Clustered | 8 | VC_1156_0 | 8 | 0.8137 | 1 | 0.8137 | 0 | 0 | 0 | 1 | Ref |
| 7063 | MH443100 | Unassigned | Unassigned | Unassigned | 32_0 | Clustered/Singleton | 1 | VC_32_0 | 1 | 0 | 1 | 0 | 0 | 0 | 0 | 1 | Ref |
| 7064 | MH444512 | Unassigned | Unassigned | Unassigned | 851_0 | Clustered | 2 | VC_851_0 | 2 | 0.4472 | 0.9410422 | 0.4209 | 0 | 0 | 0 | 1 | Ref |
| 7065 | MH445380 | Unassigned | Unassigned | Unassigned | 247_0 | Clustered | 14 | VC_247_0 | 14 | 0.9119 | 1 | 0.9119 | 0 | 0 | 0 | 0.967 | Ref |
| 7066 | MH445381 | Unassigned | Unassigned | Unassigned | 247_0 | Clustered | 14 | VC_247_0 | 14 | 0.9119 | 1 | 0.9119 | 0 | 0 | 0 | 0.967 | Ref |
| 7067 | MH445453 | Unassigned | Unassigned | Unassigned | 938_0 | Clustered | 99 | VC_938_0 | 99 | 0.9119 | 1 | 0.9119 | 0 | 0 | 0 | 0.9928 | Ref |
| 7068 | MH447526 | Unassigned | Unassigned | Unassigned | 1354_0 | Clustered | 3 | VC_1354_0 | 3 | 0.983 | 0.9595722 | 0.9432 | 0 | 0 | 0 | 1 | Ref |
| 7069 | MH450113 | Unassigned | Unassigned | Unassigned | 325_23 | Clustered | 17 | VC_325_23 | 17 | 0.0716 | 1 | 0.0716 | 0 | 0 | 0 | 0.6397 | Ref |
| 7070 | MH450114 | Unassigned | Unassigned | Unassigned | 1167_0 | Clustered | 6 | VC_1167_0 | 6 | 0.8642 | 0.99999946 | 0.8642 | 0 | 0 | 0 | 1 | Ref |
| 7071 | MH450115 | Unassigned | Unassigned | Unassigned | 1164_0 | Clustered | 25 | VC_1164_0 | 25 | 0.7312 | 1 | 0.7312 | 0 | 0 | 0 | 1 | Ref |
| 7072 | MH450116 | Unassigned | Unassigned | Unassigned | 371_16 | Clustered | 42 | VC_371_16 | 42 | 0.1341 | 1 | 0.1341 | 0 | 0 | 0 | 0.7282 | Ref |
| 7073 | MH450117 | Unassigned | Unassigned | Unassigned | 1160_0 | Clustered | 73 | VC_1160_0 | 73 | 0.9835 | 1 | 0.9835 | 0 | 0 | 0 | 1 | Ref |
| 7074 | MH450118 | Unassigned | Unassigned | Unassigned | 1160_0 | Clustered | 73 | VC_1160_0 | 73 | 0.9835 | 1 | 0.9835 | 0 | 0 | 0 | 1 | Ref |
| 7075 | MH450119 | Unassigned | Unassigned | Unassigned | 514_0 | Clustered | 75 | VC_514_0 | 75 | 0.9402 | 1 | 0.9402 | 0 | 0 | 0 | 1 | Ref |
| 7076 | MH450120 | Unassigned | Unassigned | Unassigned | 802_0 | Clustered | 17 | VC_802_0 | 17 | 0.8025 | 1 | 0.8025 | 0 | 0 | 0 | 1 | Ref |
| 7077 | MH450121 | Unassigned | Unassigned | Unassigned | 1160_0 | Clustered | 73 | VC_1160_0 | 73 | 0.9835 | 1 | 0.9835 | 0 | 0 | 0 | 1 | Ref |
| 7078 | MH450122 | Unassigned | Unassigned | Unassigned | 371_16 | Clustered | 42 | VC_371_16 | 42 | 0.1341 | 1 | 0.1341 | 0 | 0 | 0 | 0.7282 | Ref |
| 7079 | MH450124 | Unassigned | Unassigned | Unassigned | 514_0 | Clustered | 75 | VC_514_0 | 75 | 0.9402 | 1 | 0.9402 | 0 | 0 | 0 | 1 | Ref |
| 7080 | MH450125 | Unassigned | Unassigned | Unassigned | 1164_0 | Clustered | 25 | VC_1164_0 | 25 | 0.7312 | 1 | 0.7312 | 0 | 0 | 0 | 1 | Ref |
| 7081 | MH450126 | Unassigned | Unassigned | Unassigned | 514_0 | Clustered | 75 | VC_514_0 | 75 | 0.9402 | 1 | 0.9402 | 0 | 0 | 0 | 1 | Ref |
| 7082 | MH450127 | Unassigned | Unassigned | Unassigned | 761_0 | Clustered | 67 | VC_761_0 | 67 | 0.8947 | 1 | 0.8947 | 0 | 0 | 0 | 1 | Ref |
| 7083 | MH450128 | Unassigned | Unassigned | Unassigned | 514_0 | Clustered | 75 | VC_514_0 | 75 | 0.9402 | 1 | 0.9402 | 0 | 0 | 0 | 1 | Ref |
| 7084 | MH450129 | Unassigned | Unassigned | Unassigned | 1093_0 | Clustered | 40 | VC_1093_0 | 40 | 0.8754 | 1 | 0.8754 | 0 | 0 | 0 | 1 | Ref |
| 7085 | MH450130 | Unassigned | Unassigned | Unassigned | 325_23 | Clustered | 17 | VC_325_23 | 17 | 0.0716 | 1 | 0.0716 | 0 | 0 | 0 | 0.6397 | Ref |
| 7086 | MH450131 | Unassigned | Unassigned | Unassigned | 1160_0 | Clustered | 73 | VC_1160_0 | 73 | 0.9835 | 1 | 0.9835 | 0 | 0 | 0 | 1 | Ref |
| 7087 | MH450132 | Unassigned | Unassigned | Unassigned | 919_0 | Clustered | 39 | VC_919_0 | 39 | 0.9069 | 1 | 0.9069 | 0 | 0 | 0 | 1 | Ref |
| 7088 | MH450133 | Unassigned | Unassigned | Unassigned | 325_61 | Clustered | 8 | VC_325_61 | 8 | 0.0395 | 1 | 0.0395 | 0 | 0 | 0 | 0.7857 | Ref |
| 7089 | MH454076 | Unassigned | Unassigned | Unassigned | 1156_0 | Clustered | 8 | VC_1156_0 | 8 | 0.8137 | 1 | 0.8137 | 0 | 0 | 0 | 1 | Ref |
| 7090 | MH454077 | Unassigned | Unassigned | Unassigned | 708_0 | Clustered | 47 | VC_708_0 | 47 | 0.8654 | 1 | 0.8654 | 0 | 0 | 0 | 1 | Ref |
| 7091 | MH454078 | Unassigned | Unassigned | Unassigned | 708_0 | Clustered | 47 | VC_708_0 | 47 | 0.8654 | 1 | 0.8654 | 0 | 0 | 0 | 1 | Ref |
| 7092 | MH454079 | Unassigned | Unassigned | Unassigned | 708_0 | Clustered | 47 | VC_708_0 | 47 | 0.8654 | 1 | 0.8654 | 0 | 0 | 0 | 1 | Ref |
| 7093 | MH454080 | Unassigned | Unassigned | Unassigned | 1156_0 | Clustered | 8 | VC_1156_0 | 8 | 0.8137 | 1 | 0.8137 | 0 | 0 | 0 | 1 | Ref |
| 7094 | MH454082 | Unassigned | Unassigned | Unassigned | 708_0 | Clustered | 47 | VC_708_0 | 47 | 0.8654 | 1 | 0.8654 | 0 | 0 | 0 | 1 | Ref |
| 7095 | MH454083 | Unassigned | Unassigned | Unassigned | 708_0 | Clustered | 47 | VC_708_0 | 47 | 0.8654 | 1 | 0.8654 | 0 | 0 | 0 | 1 | Ref |
| 7096 | MH454084 | Unassigned | Unassigned | Unassigned | 708_0 | Clustered | 47 | VC_708_0 | 47 | 0.8654 | 1 | 0.8654 | 0 | 0 | 0 | 1 | Ref |
| 7097 | MH460459 | Unassigned | Unassigned | Unassigned | 393_0 | Clustered | 7 | VC_393_0 | 7 | 0.1775 | 1 | 0.1775 | 0 | 0 | 0 | 1 | Ref |
| 7098 | MH460460 | Unassigned | Unassigned | Unassigned | 67_4 | Clustered | 4 | VC_67_4 | 4 | 0.0431 | 0.90943035 | 0.0392 | 0 | 0 | 0 | 1 | Ref |
| 7099 | MH460461 | Unassigned | Unassigned | Unassigned | 67_4 | Clustered | 4 | VC_67_4 | 4 | 0.0431 | 0.90943035 | 0.0392 | 0 | 0 | 0 | 1 | Ref |
| 7100 | MH460462 | Unassigned | Unassigned | Unassigned | 67_4 | Clustered | 4 | VC_67_4 | 4 | 0.0431 | 0.90943035 | 0.0392 | 0 | 0 | 0 | 1 | Ref |
| 7101 | MH460463 | Unassigned | Unassigned | Unassigned | 67_3 | Clustered | 20 | VC_67_3 | 20 | 0.3542 | 1 | 0.3542 | 0 | 0 | 0 | 1 | Ref |
| 7102 | MH460824 | Unassigned | Unassigned | Unassigned | 1156_0 | Clustered | 8 | VC_1156_0 | 8 | 0.8137 | 1 | 0.8137 | 0 | 0 | 0 | 1 | Ref |
| 7103 | MH460825 | Unassigned | Unassigned | Unassigned | 1156_0 | Clustered | 8 | VC_1156_0 | 8 | 0.8137 | 1 | 0.8137 | 0 | 0 | 0 | 1 | Ref |
| 7104 | MH460826 | Unassigned | Unassigned | Unassigned | 1156_0 | Clustered | 8 | VC_1156_0 | 8 | 0.8137 | 1 | 0.8137 | 0 | 0 | 0 | 1 | Ref |
| 7105 | MH460827 | Unassigned | Unassigned | Unassigned | 1156_0 | Clustered | 8 | VC_1156_0 | 8 | 0.8137 | 1 | 0.8137 | 0 | 0 | 0 | 1 | Ref |
| 7106 | MH460829 | Unassigned | Unassigned | Unassigned | 492_1 | Clustered | 20 | VC_492_1 | 20 | 0.4176 | 1 | 0.4176 | 0 | 0 | 0 | 1 | Ref |
| 7107 | MH464253 | Unassigned | Unassigned | Unassigned | 451_0 | Clustered | 80 | VC_451_0 | 80 | 0.9991 | 1 | 0.9991 | 0 | 0 | 0 | 1 | Ref |
| 7108 | MH479909 | Unassigned | Unassigned | Unassigned | 517_1 | Clustered | 108 | VC_517_1 | 108 | 0.8483 | 1 | 0.8483 | 0 | 0 | 0 | 0.9955 | Ref |
| 7109 | MH479910 | Unassigned | Unassigned | Unassigned | 1178_0 | Clustered | 24 | VC_1178_0 | 24 | 0.9524 | 1 | 0.9524 | 0 | 0 | 0 | 1 | Ref |
| 7110 | MH479911 | Unassigned | Unassigned | Unassigned | 517_1 | Clustered | 108 | VC_517_1 | 108 | 0.8483 | 1 | 0.8483 | 0 | 0 | 0 | 0.9955 | Ref |
| 7111 | MH479912 | Unassigned | Unassigned | Unassigned | 525_0 | Clustered | 25 | VC_525_0 | 25 | 0.4597 | 1 | 0.4597 | 0 | 0 | 0 | 1 | Ref |
| 7112 | MH479913 | Unassigned | Unassigned | Unassigned | 1183_0 | Clustered | 6 | VC_1183_0 | 6 | 0.995 | 0.99996974 | 0.995 | 0 | 0 | 0 | 1 | Ref |
| 7113 | MH479914 | Unassigned | Unassigned | Unassigned | 371_16 | Clustered | 42 | VC_371_16 | 42 | 0.1341 | 1 | 0.1341 | 0 | 0 | 0 | 0.7282 | Ref |
| 7114 | MH479915 | Unassigned | Unassigned | Unassigned | 1177_0 | Clustered | 14 | VC_1177_0 | 14 | 0.7847 | 1 | 0.7847 | 0 | 0 | 0 | 0.989 | Ref |
| 7115 | MH479916 | Unassigned | Unassigned | Unassigned | 371_13 | Clustered | 33 | VC_371_13 | 33 | 0.179 | 1 | 0.179 | 0 | 0 | 0 | 0.8769 | Ref |
| 7116 | MH479917 | Unassigned | Unassigned | Unassigned | 1178_0 | Clustered | 24 | VC_1178_0 | 24 | 0.9524 | 1 | 0.9524 | 0 | 0 | 0 | 1 | Ref |
| 7117 | MH479918 | Unassigned | Unassigned | Unassigned | 371_13 | Clustered | 33 | VC_371_13 | 33 | 0.179 | 1 | 0.179 | 0 | 0 | 0 | 0.8769 | Ref |
| 7118 | MH479919 | Unassigned | Unassigned | Unassigned | 325_61 | Clustered | 8 | VC_325_61 | 8 | 0.0395 | 1 | 0.0395 | 0 | 0 | 0 | 0.7857 | Ref |
| 7119 | MH479920 | Unassigned | Unassigned | Unassigned | 761_0 | Clustered | 67 | VC_761_0 | 67 | 0.8947 | 1 | 0.8947 | 0 | 0 | 0 | 1 | Ref |
| 7120 | MH479921 | Unassigned | Unassigned | Unassigned | 371_16 | Clustered | 42 | VC_371_16 | 42 | 0.1341 | 1 | 0.1341 | 0 | 0 | 0 | 0.7282 | Ref |
| 7121 | MH479923 | Unassigned | Unassigned | Unassigned | 870_0 | Clustered | 5 | VC_870_0 | 5 | 0.424 | 0.99999997 | 0.424 | 0 | 0 | 0 | 1 | Ref |
| 7122 | MH479924 | Unassigned | Unassigned | Unassigned | 1093_0 | Clustered | 40 | VC_1093_0 | 40 | 0.8754 | 1 | 0.8754 | 0 | 0 | 0 | 1 | Ref |
| 7123 | MH479925 | Unassigned | Unassigned | Unassigned | 1183_0 | Clustered | 6 | VC_1183_0 | 6 | 0.995 | 0.99996974 | 0.995 | 0 | 0 | 0 | 1 | Ref |
| 7124 | MH479926 | Unassigned | Unassigned | Unassigned | 1164_0 | Clustered | 25 | VC_1164_0 | 25 | 0.7312 | 1 | 0.7312 | 0 | 0 | 0 | 1 | Ref |
| 7125 | MH479927 | Unassigned | Unassigned | Unassigned | 525_0 | Clustered | 25 | VC_525_0 | 25 | 0.4597 | 1 | 0.4597 | 0 | 0 | 0 | 1 | Ref |
| 7126 | MH487649 | Unassigned | Unassigned | Unassigned | 187_1 | Clustered | 7 | VC_187_1 | 7 | 0.1423 | 1 | 0.1423 | 0 | 0 | 0 | 1 | Ref |

| | | | | | | | | | | | | | | | | | |
|---|---|---|---|---|---|---|---|---|---|---|---|---|---|---|---|---|---|
| 7127 | MH491167 | Unassigned | Unassigned | Unassigned | 451_0 | Clustered | 80 | VC_451_0 | 80 | 0.9991 | 1 | 0.9991 | 0 | 0 | 0 | 1 | Ref |
| 7128 | MH491968 | Unassigned | Unassigned | Unassigned | 584_0 | Clustered | 28 | VC_584_0 | 28 | 0.7704 | 1 | 0.7704 | 0 | 0 | 0 | 1 | Ref |
| 7129 | MH491969 | Unassigned | Unassigned | Unassigned | 772_11 | Clustered | 6 | VC_772_11 | 6 | 0.0305 | 0.84253803 | 0.0257 | 0 | 0 | 0 | 0.8 | Ref |
| 7130 | MH494197 | Unassigned | Unassigned | Unassigned | 164_2 | Clustered/Singleton | 1 | VC_164_2 | 1 | 0 | 1 | 0 | 0 | 0 | 0 | 1 | Ref |
| 7131 | MH509443 | Unassigned | Unassigned | Unassigned | 517_1 | Clustered | 108 | VC_517_1 | 108 | 0.8483 | 1 | 0.8483 | 0 | 0 | 0 | 0.9955 | Ref |
| 7132 | MH509444 | Unassigned | Unassigned | Unassigned | 236_0 | Clustered | 72 | VC_236_0 | 72 | 0.8104 | 1 | 0.8104 | 0 | 0 | 0 | 1 | Ref |
| 7133 | MH509445 | Unassigned | Unassigned | Unassigned | 567_0 | Clustered | 8 | VC_567_0 | 8 | 0.4935 | 1 | 0.4935 | 0 | 0 | 0 | 1 | Ref |
| 7134 | MH509446 | Unassigned | Unassigned | Unassigned | 954_0 | Clustered | 13 | VC_954_0 | 13 | 0.863 | 1 | 0.863 | 0 | 0 | 0 | 1 | Ref |
| 7135 | MH509447 | Unassigned | Unassigned | Unassigned | 1234_0 | Clustered | 44 | VC_1234_0 | 44 | 1 | 1 | 1 | 0 | 0 | 0 | 1 | Ref |
| 7136 | MH512890 | Unassigned | Unassigned | Unassigned | 655_0 | Clustered | 2 | VC_655_0 | 2 | 0.0111 | 0.95704495 | 0.0106 | 0 | 0 | 0 | 1 | Ref |
| 7137 | MH513970 | Unassigned | Unassigned | Unassigned | 372_0 | Clustered | 24 | VC_372_0 | 24 | 0.5539 | 1 | 0.5539 | 0 | 0 | 0 | 1 | Ref |
| 7138 | MH513971 | Unassigned | Unassigned | Unassigned | 325_0 | Clustered | 2 | VC_325_0 | 2 | 0.0041 | 0.95574439 | 0.0039 | 0 | 0 | 0 | 1 | Ref |
| 7139 | MH513972 | Unassigned | Unassigned | Unassigned | 551_2 | Clustered | 17 | VC_551_2 | 17 | 0.0973 | 1 | 0.0973 | 0 | 0 | 0 | 0.9779 | Ref |
| 7140 | MH513973 | Unassigned | Unassigned | Unassigned | 371_13 | Clustered | 33 | VC_371_13 | 33 | 0.179 | 1 | 0.179 | 0 | 0 | 0 | 0.8769 | Ref |
| 7141 | MH513974 | Unassigned | Unassigned | Unassigned | 224_0 | Clustered | 19 | VC_224_0 | 19 | 0.9416 | 1 | 0.9416 | 0 | 0 | 0 | 1 | Ref |
| 7142 | MH513975 | Unassigned | Unassigned | Unassigned | 514_0 | Clustered | 75 | VC_514_0 | 75 | 0.9402 | 1 | 0.9402 | 0 | 0 | 0 | 1 | Ref |
| 7143 | MH513976 | Unassigned | Unassigned | Unassigned | 509_0 | Clustered | 37 | VC_509_0 | 37 | 0.9689 | 1 | 0.9689 | 0 | 0 | 0 | 1 | Ref |
| 7144 | MH513977 | Unassigned | Unassigned | Unassigned | 236_0 | Clustered | 72 | VC_236_0 | 72 | 0.8104 | 1 | 0.8104 | 0 | 0 | 0 | 1 | Ref |
| 7145 | MH513978 | Unassigned | Unassigned | Unassigned | 551_2 | Clustered | 17 | VC_551_2 | 17 | 0.0973 | 1 | 0.0973 | 0 | 0 | 0 | 0.9779 | Ref |
| 7146 | MH513979 | Unassigned | Unassigned | Unassigned | 685_0 | Clustered | 16 | VC_685_0 | 16 | 0.2972 | 1 | 0.2972 | 0 | 0 | 0 | 0.9167 | Ref |
| 7147 | MH513980 | Unassigned | Unassigned | Unassigned | 371_11 | Clustered | 31 | VC_371_11 | 31 | 0.0927 | 1 | 0.0927 | 0 | 0 | 0 | 0.7011 | Ref |
| 7148 | MH513981 | Unassigned | Unassigned | Unassigned | 1240_0 | Clustered | 82 | VC_1240_0 | 82 | 0.7945 | 1 | 0.7945 | 0 | 0 | 0 | 1 | Ref |
| 7149 | MH513982 | Unassigned | Unassigned | Unassigned | 1163_0 | Clustered | 28 | VC_1163_0 | 28 | 0.4923 | 1 | 0.4923 | 0 | 0 | 0 | 1 | Ref |
| 7150 | MH513983 | Unassigned | Unassigned | Unassigned | 551_6 | Clustered | 22 | VC_551_6 | 22 | 0.1214 | 1 | 0.1214 | 0 | 0 | 0 | 0.8095 | Ref |
| 7151 | MH513984 | Unassigned | Unassigned | Unassigned | 222_0 | Clustered | 5 | VC_222_0 | 5 | 0.0565 | 0.99999997 | 0.0565 | 0 | 0 | 0 | 1 | Ref |
| 7152 | MH513985 | Unassigned | Unassigned | Unassigned | 215_1 | Clustered | 19 | VC_215_1 | 19 | 0.4188 | 1 | 0.4188 | 0 | 0 | 0 | 1 | Ref |
| 7153 | MH523359 | Unassigned | Unassigned | Unassigned | 133_40 | Clustered/Singleton | 1 | VC_133_40 | 1 | 0 | 1 | 0 | 0 | 0 | 0 | 1 | Ref |
| 7154 | MH533020 | Unassigned | Unassigned | Unassigned | 849_0 | Clustered | 3 | VC_849_0 | 3 | 0.069 | 0.9984794 | 0.0689 | 0 | 0 | 0 | 1 | Ref |
| 7155 | MH536736 | Unassigned | Unassigned | Unassigned | 80_0 | Clustered | 41 | VC_80_0 | 41 | 0.5837 | 1 | 0.5837 | 0 | 0 | 0 | 0.9805 | Ref |
| 7156 | MH536811 | Unassigned | Unassigned | Unassigned | 1072_4 | Clustered | 4 | VC_1072_4 | 4 | 0.0736 | 0.99956559 | 0.0736 | 0 | 0 | 0 | 1 | Ref |
| 7157 | MH536812 | Unassigned | Unassigned | Unassigned | 557_2 | Clustered/Singleton | 1 | VC_557_2 | 1 | 0 | 1 | 0 | 0 | 0 | 0 | 1 | Ref |
| 7158 | MH536813 | Unassigned | Unassigned | Unassigned | 514_0 | Clustered | 75 | VC_514_0 | 75 | 0.9402 | 1 | 0.9402 | 0 | 0 | 0 | 1 | Ref |
| 7159 | MH536814 | Unassigned | Unassigned | Unassigned | 1072_4 | Clustered | 4 | VC_1072_4 | 4 | 0.0736 | 0.99956559 | 0.0736 | 0 | 0 | 0 | 1 | Ref |
| 7160 | MH536815 | Unassigned | Unassigned | Unassigned | 1353_0 | Clustered | 10 | VC_1353_0 | 10 | 1 | 1 | 1 | 0 | 0 | 0 | 1 | Ref |
| 7161 | MH536816 | Unassigned | Unassigned | Unassigned | 325_27 | Clustered | 2 | VC_325_27 | 2 | 0.0044 | 0.95750955 | 0.0042 | 0 | 0 | 0 | 1 | Ref |
| 7162 | MH536817 | Unassigned | Unassigned | Unassigned | 517_1 | Clustered | 108 | VC_517_1 | 108 | 0.8483 | 1 | 0.8483 | 0 | 0 | 0 | 0.9955 | Ref |
| 7163 | MH536818 | Unassigned | Unassigned | Unassigned | 1176_0 | Clustered | 41 | VC_1176_0 | 41 | 0.9558 | 1 | 0.9558 | 0 | 0 | 0 | 0.9976 | Ref |
| 7164 | MH536819 | Unassigned | Unassigned | Unassigned | 767_0 | Clustered | 34 | VC_767_0 | 34 | 0.9173 | 1 | 0.9173 | 0 | 0 | 0 | 1 | Ref |
| 7165 | MH536820 | Unassigned | Unassigned | Unassigned | 551_0 | Clustered | 16 | VC_551_0 | 16 | 0.0915 | 1 | 0.0915 | 0 | 0 | 0 | 0.6667 | Ref |
| 7166 | MH536821 | Unassigned | Unassigned | Unassigned | 236_0 | Clustered | 72 | VC_236_0 | 72 | 0.8104 | 1 | 0.8104 | 0 | 0 | 0 | 1 | Ref |
| 7167 | MH536822 | Unassigned | Unassigned | Unassigned | 236_0 | Clustered | 72 | VC_236_0 | 72 | 0.8104 | 1 | 0.8104 | 0 | 0 | 0 | 1 | Ref |
| 7168 | MH536823 | Unassigned | Unassigned | Unassigned | 856_0 | Clustered | 7 | VC_856_0 | 7 | 0.3463 | 1 | 0.3463 | 0 | 0 | 0 | 1 | Ref |
| 7169 | MH536824 | Unassigned | Unassigned | Unassigned | 767_0 | Clustered | 34 | VC_767_0 | 34 | 0.9173 | 1 | 0.9173 | 0 | 0 | 0 | 1 | Ref |
| 7170 | MH536825 | Unassigned | Unassigned | Unassigned | 514_0 | Clustered | 75 | VC_514_0 | 75 | 0.9402 | 1 | 0.9402 | 0 | 0 | 0 | 1 | Ref |
| 7171 | MH536827 | Unassigned | Unassigned | Unassigned | 551_7 | Clustered | 28 | VC_551_7 | 28 | 0.1661 | 1 | 0.1661 | 0 | 0 | 0 | 0.8519 | Ref |
| 7172 | MH536828 | Unassigned | Unassigned | Unassigned | 221_0 | Clustered | 22 | VC_221_0 | 22 | 0.7447 | 1 | 0.7447 | 0 | 0 | 0 | 1 | Ref |
| 7173 | MH536829 | Unassigned | Unassigned | Unassigned | 551_6 | Clustered | 22 | VC_551_6 | 22 | 0.1214 | 1 | 0.1214 | 0 | 0 | 0 | 0.8095 | Ref |
| 7174 | MH538193 | Unassigned | Unassigned | Unassigned | 884_0 | Clustered | 28 | VC_884_0 | 28 | 0.6211 | 1 | 0.6211 | 0 | 0 | 0 | 0.9841 | Ref |
| 7175 | MH540083 | Unassigned | Unassigned | Unassigned | 340_0 | Clustered | 31 | VC_340_0 | 31 | 0.9549 | 1 | 0.9549 | 0 | 0 | 0 | 1 | Ref |
| 7176 | MH542234 | Unassigned | Unassigned | Unassigned | 150_0 | Clustered | 13 | VC_150_0 | 13 | 0.2417 | 1 | 0.2417 | 0 | 0 | 0 | 0.9872 | Ref |
| 7177 | MH547045 | Unassigned | Unassigned | Unassigned | 43_0 | Clustered | 38 | VC_43_0 | 38 | 0.5885 | 1 | 0.5885 | 0 | 0 | 0 | 0.9659 | Ref |
| 7178 | MH550421 | Unassigned | Unassigned | Unassigned | 283_88 | Clustered | 2 | VC_283_88 | 2 | 0.0026 | 0.95596547 | 0.0024 | 0 | 0 | 0 | 1 | Ref |
| 7179 | MH552500 | Unassigned | Unassigned | Unassigned | 673_0 | Clustered | 15 | VC_673_0 | 15 | 0.8234 | 1 | 0.8234 | 0 | 0 | 0 | 0.7619 | Ref |
| 7180 | MH553563 | Unassigned | Unassigned | Unassigned | 283_52 | Clustered/Singleton | 1 | VC_283_52 | 1 | 0 | 1 | 0 | 0 | 0 | 0 | 1 | Ref |
| 7181 | MH560358 | Unassigned | Unassigned | Unassigned | 590_0 | Clustered | 27 | VC_590_0 | 27 | 0.9947 | 1 | 0.9947 | 0 | 0 | 0 | 1 | Ref |
| 7182 | MH571750 | Unassigned | Unassigned | Unassigned | 329_7 | Clustered | 18 | VC_329_7 | 18 | 0.0938 | 1 | 0.0938 | 0 | 0 | 0 | 0.7778 | Ref |
| 7183 | MH572269 | Unassigned | Unassigned | Unassigned | 398_0 | Clustered | 48 | VC_398_0 | 48 | 0.4813 | 1 | 0.4813 | 0 | 0 | 0 | 1 | Ref |
| 7184 | MH572274 | Unassigned | Unassigned | Unassigned | 1355_0 | Clustered | 3 | VC_1355_0 | 3 | 1 | 1 | 1 | 0 | 0 | 0 | 1 | Ref |
| 7185 | MH572275 | Unassigned | Unassigned | Unassigned | 1355_0 | Clustered | 3 | VC_1355_0 | 3 | 1 | 1 | 1 | 0 | 0 | 0 | 1 | Ref |
| 7186 | MH572276 | Unassigned | Unassigned | Unassigned | 1355_0 | Clustered | 3 | VC_1355_0 | 3 | 1 | 1 | 1 | 0 | 0 | 0 | 1 | Ref |
| 7187 | MH572277 | Unassigned | Unassigned | Unassigned | 198_0 | Clustered | 28 | VC_198_0 | 28 | 0.2429 | 0.99971175 | 0.2429 | 0 | 0 | 0 | 1 | Ref |
| 7188 | MH572279 | Unassigned | Unassigned | Unassigned | 404_0 | Clustered | 25 | VC_404_0 | 25 | 0.5649 | 1 | 0.5649 | 0 | 0 | 0 | 1 | Ref |
| 7189 | MH572280 | Unassigned | Unassigned | Unassigned | 198_0 | Clustered | 28 | VC_198_0 | 28 | 0.2429 | 0.99971175 | 0.2429 | 0 | 0 | 0 | 1 | Ref |
| 7190 | MH572282 | Unassigned | Unassigned | Unassigned | 198_0 | Clustered | 28 | VC_198_0 | 28 | 0.2429 | 0.99971175 | 0.2429 | 0 | 0 | 0 | 1 | Ref |
| 7191 | MH572283 | Unassigned | Unassigned | Unassigned | 995_0 | Clustered | 69 | VC_995_0 | 69 | 0.8547 | 1 | 0.8547 | 0 | 0 | 0 | 1 | Ref |
| 7192 | MH572284 | Unassigned | Unassigned | Unassigned | 995_0 | Clustered | 69 | VC_995_0 | 69 | 0.8547 | 1 | 0.8547 | 0 | 0 | 0 | 1 | Ref |
| 7193 | MH572285 | Unassigned | Unassigned | Unassigned | 995_0 | Clustered | 69 | VC_995_0 | 69 | 0.8547 | 1 | 0.8547 | 0 | 0 | 0 | 1 | Ref |
| 7194 | MH572286 | Unassigned | Unassigned | Unassigned | 995_0 | Clustered | 69 | VC_995_0 | 69 | 0.8547 | 1 | 0.8547 | 0 | 0 | 0 | 1 | Ref |
| 7195 | MH572287 | Unassigned | Unassigned | Unassigned | 1084_0 | Clustered | 9 | VC_1084_0 | 9 | 0.6558 | 1 | 0.6558 | 0 | 0 | 0 | 1 | Ref |
| 7196 | MH572288 | Unassigned | Unassigned | Unassigned | 1084_0 | Clustered | 9 | VC_1084_0 | 9 | 0.6558 | 1 | 0.6558 | 0 | 0 | 0 | 1 | Ref |
| 7197 | MH572290 | Unassigned | Unassigned | Unassigned | 198_0 | Clustered | 28 | VC_198_0 | 28 | 0.2429 | 0.99971175 | 0.2429 | 0 | 0 | 0 | 1 | Ref |
| 7198 | MH572291 | Unassigned | Unassigned | Unassigned | 995_0 | Clustered | 69 | VC_995_0 | 69 | 0.8547 | 1 | 0.8547 | 0 | 0 | 0 | 1 | Ref |
| 7199 | MH572292 | Unassigned | Unassigned | Unassigned | 995_0 | Clustered | 69 | VC_995_0 | 69 | 0.8547 | 1 | 0.8547 | 0 | 0 | 0 | 1 | Ref |
| 7200 | MH572293 | Unassigned | Unassigned | Unassigned | 995_0 | Clustered | 69 | VC_995_0 | 69 | 0.8547 | 1 | 0.8547 | 0 | 0 | 0 | 1 | Ref |
| 7201 | MH572294 | Unassigned | Unassigned | Unassigned | 403_0 | Clustered | 20 | VC_403_0 | 20 | 0.6598 | 1 | 0.6598 | 0 | 0 | 0 | 1 | Ref |
| 7202 | MH572295 | Unassigned | Unassigned | Unassigned | 403_0 | Clustered | 20 | VC_403_0 | 20 | 0.6598 | 1 | 0.6598 | 0 | 0 | 0 | 1 | Ref |
| 7203 | MH572296 | Unassigned | Unassigned | Unassigned | 403_0 | Clustered | 20 | VC_403_0 | 20 | 0.6598 | 1 | 0.6598 | 0 | 0 | 0 | 1 | Ref |
| 7204 | MH572299 | Unassigned | Unassigned | Unassigned | 998_0 | Clustered | 2 | VC_998_0 | 2 | 0.0515 | 0.93901809 | 0.0484 | 0 | 0 | 0 | 1 | Ref |
| 7205 | MH572300 | Unassigned | Unassigned | Unassigned | 404_0 | Clustered | 25 | VC_404_0 | 25 | 0.5649 | 1 | 0.5649 | 0 | 0 | 0 | 1 | Ref |
| 7206 | MH572301 | Unassigned | Unassigned | Unassigned | 404_0 | Clustered | 25 | VC_404_0 | 25 | 0.5649 | 1 | 0.5649 | 0 | 0 | 0 | 1 | Ref |
| 7207 | MH572302 | Unassigned | Unassigned | Unassigned | 404_0 | Clustered | 25 | VC_404_0 | 25 | 0.5649 | 1 | 0.5649 | 0 | 0 | 0 | 1 | Ref |
| 7208 | MH572303 | Unassigned | Unassigned | Unassigned | 404_0 | Clustered | 25 | VC_404_0 | 25 | 0.5649 | 1 | 0.5649 | 0 | 0 | 0 | 1 | Ref |
| 7209 | MH572305 | Unassigned | Unassigned | Unassigned | 404_0 | Clustered | 25 | VC_404_0 | 25 | 0.5649 | 1 | 0.5649 | 0 | 0 | 0 | 1 | Ref |
| 7210 | MH572306 | Unassigned | Unassigned | Unassigned | 404_0 | Clustered | 25 | VC_404_0 | 25 | 0.5649 | 1 | 0.5649 | 0 | 0 | 0 | 1 | Ref |
| 7211 | MH572307 | Unassigned | Unassigned | Unassigned | 404_0 | Clustered | 25 | VC_404_0 | 25 | 0.5649 | 1 | 0.5649 | 0 | 0 | 0 | 1 | Ref |
| 7212 | MH572308 | Unassigned | Unassigned | Unassigned | 404_0 | Clustered | 25 | VC_404_0 | 25 | 0.5649 | 1 | 0.5649 | 0 | 0 | 0 | 1 | Ref |
| 7213 | MH572309 | Unassigned | Unassigned | Unassigned | 404_0 | Clustered | 25 | VC_404_0 | 25 | 0.5649 | 1 | 0.5649 | 0 | 0 | 0 | 1 | Ref |
| 7214 | MH572310 | Unassigned | Unassigned | Unassigned | 404_0 | Clustered | 25 | VC_404_0 | 25 | 0.5649 | 1 | 0.5649 | 0 | 0 | 0 | 1 | Ref |
| 7215 | MH572311 | Unassigned | Unassigned | Unassigned | 404_0 | Clustered | 25 | VC_404_0 | 25 | 0.5649 | 1 | 0.5649 | 0 | 0 | 0 | 1 | Ref |
| 7216 | MH572312 | Unassigned | Unassigned | Unassigned | 404_0 | Clustered | 25 | VC_404_0 | 25 | 0.5649 | 1 | 0.5649 | 0 | 0 | 0 | 1 | Ref |
| 7217 | MH572313 | Unassigned | Unassigned | Unassigned | 404_0 | Clustered | 25 | VC_404_0 | 25 | 0.5649 | 1 | 0.5649 | 0 | 0 | 0 | 1 | Ref |
| 7218 | MH572314 | Unassigned | Unassigned | Unassigned | 398_0 | Clustered | 48 | VC_398_0 | 48 | 0.4813 | 1 | 0.4813 | 0 | 0 | 0 | 1 | Ref |
| 7219 | MH572315 | Unassigned | Unassigned | Unassigned | 398_0 | Clustered | 48 | VC_398_0 | 48 | 0.4813 | 1 | 0.4813 | 0 | 0 | 0 | 1 | Ref |
| 7220 | MH572316 | Unassigned | Unassigned | Unassigned | 398_0 | Clustered | 48 | VC_398_0 | 48 | 0.4813 | 1 | 0.4813 | 0 | 0 | 0 | 1 | Ref |
| 7221 | MH572317 | Unassigned | Unassigned | Unassigned | 398_0 | Clustered | 48 | VC_398_0 | 48 | 0.4813 | 1 | 0.4813 | 0 | 0 | 0 | 1 | Ref |
| 7222 | MH572318 | Unassigned | Unassigned | Unassigned | 398_0 | Clustered | 48 | VC_398_0 | 48 | 0.4813 | 1 | 0.4813 | 0 | 0 | 0 | 1 | Ref |
| 7223 | MH572319 | Unassigned | Unassigned | Unassigned | 398_0 | Clustered | 48 | VC_398_0 | 48 | 0.4813 | 1 | 0.4813 | 0 | 0 | 0 | 1 | Ref |
| 7224 | MH572320 | Unassigned | Unassigned | Unassigned | 398_0 | Clustered | 48 | VC_398_0 | 48 | 0.4813 | 1 | 0.4813 | 0 | 0 | 0 | 1 | Ref |
| 7225 | MH572321 | Unassigned | Unassigned | Unassigned | 398_0 | Clustered | 48 | VC_398_0 | 48 | 0.4813 | 1 | 0.4813 | 0 | 0 | 0 | 1 | Ref |
| 7226 | MH572325 | Unassigned | Unassigned | Unassigned | 398_0 | Clustered | 48 | VC_398_0 | 48 | 0.4813 | 1 | 0.4813 | 0 | 0 | 0 | 1 | Ref |
| 7227 | MH572326 | Unassigned | Unassigned | Unassigned | 398_0 | Clustered | 48 | VC_398_0 | 48 | 0.4813 | 1 | 0.4813 | 0 | 0 | 0 | 1 | Ref |
| 7228 | MH572328 | Unassigned | Unassigned | Unassigned | 398_0 | Clustered | 48 | VC_398_0 | 48 | 0.4813 | 1 | 0.4813 | 0 | 0 | 0 | 1 | Ref |
| 7229 | MH572329 | Unassigned | Unassigned | Unassigned | 398_0 | Clustered | 48 | VC_398_0 | 48 | 0.4813 | 1 | 0.4813 | 0 | 0 | 0 | 1 | Ref |
| 7230 | MH572330 | Unassigned | Unassigned | Unassigned | 198_0 | Clustered | 28 | VC_198_0 | 28 | 0.2429 | 0.99971175 | 0.2429 | 0 | 0 | 0 | 1 | Ref |
| 7231 | MH572334 | Unassigned | Unassigned | Unassigned | 198_0 | Clustered | 28 | VC_198_0 | 28 | 0.2429 | 0.99971175 | 0.2429 | 0 | 0 | 0 | 1 | Ref |
| 7232 | MH572336 | Unassigned | Unassigned | Unassigned | 398_0 | Clustered | 48 | VC_398_0 | 48 | 0.4813 | 1 | 0.4813 | 0 | 0 | 0 | 1 | Ref |
| 7233 | MH572340 | Unassigned | Unassigned | Unassigned | 400_0 | Clustered/Singleton | 1 | VC_400_0 | 1 | 0 | 1 | 0 | 0 | 0 | 0 | 1 | Ref |
| 7234 | MH572341 | Unassigned | Unassigned | Unassigned | 199_0 | Clustered | 12 | VC_199_0 | 12 | 0.448 | 1 | 0.448 | 0 | 0 | 0 | 1 | Ref |
| 7235 | MH572342 | Unassigned | Unassigned | Unassigned | 199_0 | Clustered | 12 | VC_199_0 | 12 | 0.448 | 1 | 0.448 | 0 | 0 | 0 | 1 | Ref |
| 7236 | MH572343 | Unassigned | Unassigned | Unassigned | 199_0 | Clustered | 12 | VC_199_0 | 12 | 0.448 | 1 | 0.448 | 0 | 0 | 0 | 1 | Ref |
| 7237 | MH572345 | Unassigned | Unassigned | Unassigned | 404_0 | Clustered | 25 | VC_404_0 | 25 | 0.5649 | 1 | 0.5649 | 0 | 0 | 0 | 1 | Ref |
| 7238 | MH572347 | Unassigned | Unassigned | Unassigned | 992_0 | Clustered | 8 | VC_992_0 | 8 | 0.3873 | 1 | 0.3873 | 0 | 0 | 0 | 1 | Ref |
| 7239 | MH572348 | Unassigned | Unassigned | Unassigned | 992_0 | Clustered | 8 | VC_992_0 | 8 | 0.3873 | 1 | 0.3873 | 0 | 0 | 0 | 1 | Ref |
| 7240 | MH572349 | Unassigned | Unassigned | Unassigned | 992_0 | Clustered | 8 | VC_992_0 | 8 | 0.3873 | 1 | 0.3873 | 0 | 0 | 0 | 1 | Ref |
| 7241 | MH572351 | Unassigned | Unassigned | Unassigned | 398_0 | Clustered | 48 | VC_398_0 | 48 | 0.4813 | 1 | 0.4813 | 0 | 0 | 0 | 1 | Ref |
| 7242 | MH572353 | Unassigned | Unassigned | Unassigned | 1356_0 | Clustered | 8 | VC_1356_0 | 8 | 0.8729 | 0.99998222 | 0.8729 | 0 | 0 | 0 | 1 | Ref |
| 7243 | MH572354 | Unassigned | Unassigned | Unassigned | 1356_0 | Clustered | 8 | VC_1356_0 | 8 | 0.8729 | 0.99998222 | 0.8729 | 0 | 0 | 0 | 1 | Ref |
| 7244 | MH572355 | Unassigned | Unassigned | Unassigned | 1356_0 | Clustered | 8 | VC_1356_0 | 8 | 0.8729 | 0.99998222 | 0.8729 | 0 | 0 | 0 | 1 | Ref |
| 7245 | MH572357 | Unassigned | Unassigned | Unassigned | 1356_0 | Clustered | 8 | VC_1356_0 | 8 | 0.8729 | 0.99998222 | 0.8729 | 0 | 0 | 0 | 1 | Ref |
| 7246 | MH572358 | Unassigned | Unassigned | Unassigned | 1356_0 | Clustered | 8 | VC_1356_0 | 8 | 0.8729 | 0.99998222 | 0.8729 | 0 | 0 | 0 | 1 | Ref |
| 7247 | MH572360 | Unassigned | Unassigned | Unassigned | 1079_0 | Clustered | 6 | VC_1079_0 | 6 | 0.9301 | 0.89096579 | 0.8287 | 0 | 0 | 0 | 1 | Ref |
| 7248 | MH572361 | Unassigned | Unassigned | Unassigned | 1079_0 | Clustered | 6 | VC_1079_0 | 6 | 0.9301 | 0.89096579 | 0.8287 | 0 | 0 | 0 | 1 | Ref |
| 7249 | MH572362 | Unassigned | Unassigned | Unassigned | 1077_0 | Clustered | 48 | VC_1077_0 | 48 | 0.8525 | 1 | 0.8525 | 0 | 0 | 0 | 1 | Ref |
| 7250 | MH572364 | Unassigned | Unassigned | Unassigned | 1088_0 | Clustered | 3 | VC_1088_0 | 3 | 0.7739 | 0.82711071 | 0.6401 | 0 | 0 | 0 | 1 | Ref |
| 7251 | MH572366 | Unassigned | Unassigned | Unassigned | 1082_0 | Clustered | 3 | VC_1082_0 | 3 | 0.6925 | 0.61954664 | 0.429 | 0 | 0 | 0 | 1 | Ref |
| 7252 | MH572367 | Unassigned | Unassigned | Unassigned | 1062_0 | Clustered | 45 | VC_1062_0 | 45 | 0.9734 | 0.50949847 | 0.4959 | 0 | 0 | 0 | 1 | Ref |
| 7253 | MH572368 | Unassigned | Unassigned | Unassigned | 1062_0 | Clustered | 45 | VC_1062_0 | 45 | 0.9734 | 0.50949847 | 0.4959 | 0 | 0 | 0 | 1 | Ref |
| 7254 | MH572369 | Unassigned | Unassigned | Unassigned | 1308_0 | Clustered | 12 | VC_1308_0 | 12 | 0.8184 | 0.9810731 | 0.8029 | 0 | 0 | 0 | 1 | Ref |
| 7255 | MH572371 | Unassigned | Unassigned | Unassigned | 1087_0 | Clustered | 5 | VC_1087_0 | 5 | 0.8109 | 0.98759579 | 0.8008 | 0 | 0 | 0 | 1 | Ref |
| 7256 | MH572373 | Unassigned | Unassigned | Unassigned | 1087_0 | Clustered | 5 | VC_1087_0 | 5 | 0.8109 | 0.98759579 | 0.8008 | 0 | 0 | 0 | 1 | Ref |
| 7257 | MH572374 | Unassigned | Unassigned | Unassigned | 1323_0 | Clustered | 6 | VC_1323_0 | 6 | 0.598 | 0.99460389 | 0.5948 | 0 | 0 | 0 | 1 | Ref |
| 7258 | MH572375 | Unassigned | Unassigned | Unassigned | 1323_0 | Clustered | 6 | VC_1323_0 | 6 | 0.598 | 0.99460389 | 0.5948 | 0 | 0 | 0 | 1 | Ref |
| 7259 | MH572376 | Unassigned | Unassigned | Unassigned | 1323_0 | Clustered | 6 | VC_1323_0 | 6 | 0.598 | 0.99460389 | 0.5948 | 0 | 0 | 0 | 1 | Ref |
| 7260 | MH572377 | Unassigned | Unassigned | Unassigned | 1062_0 | Clustered | 45 | VC_1062_0 | 45 | 0.9734 | 0.50949847 | 0.4959 | 0 | 0 | 0 | 1 | Ref |
| 7261 | MH572378 | Unassigned | Unassigned | Unassigned | 1062_0 | Clustered | 45 | VC_1062_0 | 45 | 0.9734 | 0.50949847 | 0.4959 | 0 | 0 | 0 | 1 | Ref |
| 7262 | MH572379 | Unassigned | Unassigned | Unassigned | 1062_0 | Clustered | 45 | VC_1062_0 | 45 | 0.9734 | 0.50949847 | 0.4959 | 0 | 0 | 0 | 1 | Ref |
| 7263 | MH572380 | Unassigned | Unassigned | Unassigned | 1087_0 | Clustered | 5 | VC_1087_0 | 5 | 0.8109 | 0.98759579 | 0.8008 | 0 | 0 | 0 | 1 | Ref |
| 7264 | MH572382 | Unassigned | Unassigned | Unassigned | 995_0 | Clustered | 69 | VC_995_0 | 69 | 0.8547 | 1 | 0.8547 | 0 | 0 | 0 | 1 | Ref |
| 7265 | MH572383 | Unassigned | Unassigned | Unassigned | 995_0 | Clustered | 69 | VC_995_0 | 69 | 0.8547 | 1 | 0.8547 | 0 | 0 | 0 | 1 | Ref |
| 7266 | MH572384 | Unassigned | Unassigned | Unassigned | 403_0 | Clustered | 20 | VC_403_0 | 20 | 0.6598 | 1 | 0.6598 | 0 | 0 | 0 | 1 | Ref |
| 7267 | MH572385 | Unassigned | Unassigned | Unassigned | 403_0 | Clustered | 20 | VC_403_0 | 20 | 0.6598 | 1 | 0.6598 | 0 | 0 | 0 | 1 | Ref |
| 7268 | MH572386 | Unassigned | Unassigned | Unassigned | 403_0 | Clustered | 20 | VC_403_0 | 20 | 0.6598 | 1 | 0.6598 | 0 | 0 | 0 | 1 | Ref |
| 7269 | MH572387 | Unassigned | Unassigned | Unassigned | 403_0 | Clustered | 20 | VC_403_0 | 20 | 0.6598 | 1 | 0.6598 | 0 | 0 | 0 | 1 | Ref |
| 7270 | MH572388 | Unassigned | Unassigned | Unassigned | 403_0 | Clustered | 20 | VC_403_0 | 20 | 0.6598 | 1 | 0.6598 | 0 | 0 | 0 | 1 | Ref |
| 7271 | MH572389 | Unassigned | Unassigned | Unassigned | 403_0 | Clustered | 20 | VC_403_0 | 20 | 0.6598 | 1 | 0.6598 | 0 | 0 | 0 | 1 | Ref |
| 7272 | MH572390 | Unassigned | Unassigned | Unassigned | 403_0 | Clustered | 20 | VC_403_0 | 20 | 0.6598 | 1 | 0.6598 | 0 | 0 | 0 | 1 | Ref |
| 7273 | MH572391 | Unassigned | Unassigned | Unassigned | 403_0 | Clustered | 20 | VC_403_0 | 20 | 0.6598 | 1 | 0.6598 | 0 | 0 | 0 | 1 | Ref |
| 7274 | MH572392 | Unassigned | Unassigned | Unassigned | 404_0 | Clustered | 25 | VC_404_0 | 25 | 0.5649 | 1 | 0.5649 | 0 | 0 | 0 | 1 | Ref |
| 7275 | MH572393 | Unassigned | Unassigned | Unassigned | 404_0 | Clustered | 25 | VC_404_0 | 25 | 0.5649 | 1 | 0.5649 | 0 | 0 | 0 | 1 | Ref |
| 7276 | MH572394 | Unassigned | Unassigned | Unassigned | 398_0 | Clustered | 48 | VC_398_0 | 48 | 0.4813 | 1 | 0.4813 | 0 | 0 | 0 | 1 | Ref |
| 7277 | MH572395 | Unassigned | Unassigned | Unassigned | 398_0 | Clustered | 48 | VC_398_0 | 48 | 0.4813 | 1 | 0.4813 | 0 | 0 | 0 | 1 | Ref |
| 7278 | MH572396 | Unassigned | Unassigned | Unassigned | 398_0 | Clustered | 48 | VC_398_0 | 48 | 0.4813 | 1 | 0.4813 | 0 | 0 | 0 | 1 | Ref |
| 7279 | MH572397 | Unassigned | Unassigned | Unassigned | 398_0 | Clustered | 48 | VC_398_0 | 48 | 0.4813 | 1 | 0.4813 | 0 | 0 | 0 | 1 | Ref |
| 7280 | MH572399 | Unassigned | Unassigned | Unassigned | 199_0 | Clustered | 12 | VC_199_0 | 12 | 0.448 | 1 | 0.448 | 0 | 0 | 0 | 1 | Ref |
| 7281 | MH572400 | Unassigned | Unassigned | Unassigned | 199_0 | Clustered | 12 | VC_199_0 | 12 | 0.448 | 1 | 0.448 | 0 | 0 | 0 | 1 | Ref |
| 7282 | MH572401 | Unassigned | Unassigned | Unassigned | 199_0 | Clustered | 12 | VC_199_0 | 12 | 0.448 | 1 | 0.448 | 0 | 0 | 0 | 1 | Ref |
| 7283 | MH572402 | Unassigned | Unassigned | Unassigned | 199_0 | Clustered | 12 | VC_199_0 | 12 | 0.448 | 1 | 0.448 | 0 | 0 | 0 | 1 | Ref |
| 7284 | MH572403 | Unassigned | Unassigned | Unassigned | 199_0 | Clustered | 12 | VC_199_0 | 12 | 0.448 | 1 | 0.448 | 0 | 0 | 0 | 1 | Ref |
| 7285 | MH572404 | Unassigned | Unassigned | Unassigned | 398_0 | Clustered | 48 | VC_398_0 | 48 | 0.4813 | 1 | 0.4813 | 0 | 0 | 0 | 1 | Ref |
| 7286 | MH572405 | Unassigned | Unassigned | Unassigned | 992_0 | Clustered | 8 | VC_992_0 | 8 | 0.3873 | 1 | 0.3873 | 0 | 0 | 0 | 1 | Ref |
| 7287 | MH572406 | Unassigned | Unassigned | Unassigned | 404_0 | Clustered | 25 | VC_404_0 | 25 | 0.5649 | 1 | 0.5649 | 0 | 0 | 0 | 1 | Ref |
| 7288 | MH572407 | Unassigned | Unassigned | Unassigned | 1356_0 | Clustered | 8 | VC_1356_0 | 8 | 0.8729 | 0.99998222 | 0.8729 | 0 | 0 | 0 | 1 | Ref |

| | | | | | | | | | | | | | | | | | |
|---|---|---|---|---|---|---|---|---|---|---|---|---|---|---|---|---|---|
| 7289 | MH572408 | Unassigned | Unassigned | Unassigned | 1356_0 | Clustered | 8 | VC_1356_0 | 8 | 0.8729 | 0.99998222 | 0.8729 | 0 | 0 | 0 | 1 | Ref |
| 7290 | MH572409 | Unassigned | Unassigned | Unassigned | 1079_0 | Clustered | 6 | VC_1079_0 | 6 | 0.9301 | 0.89096579 | 0.8287 | 0 | 0 | 0 | 1 | Ref |
| 7291 | MH572410 | Unassigned | Unassigned | Unassigned | 1077_0 | Clustered | 48 | VC_1077_0 | 48 | 0.8525 | 1 | 0.8525 | 0 | 0 | 0 | 1 | Ref |
| 7292 | MH572411 | Unassigned | Unassigned | Unassigned | 1077_0 | Clustered | 48 | VC_1077_0 | 48 | 0.8525 | 1 | 0.8525 | 0 | 0 | 0 | 1 | Ref |
| 7293 | MH572412 | Unassigned | Unassigned | Unassigned | 1077_0 | Clustered | 48 | VC_1077_0 | 48 | 0.8525 | 1 | 0.8525 | 0 | 0 | 0 | 1 | Ref |
| 7294 | MH572414 | Unassigned | Unassigned | Unassigned | 1323_0 | Clustered | 6 | VC_1323_0 | 6 | 0.598 | 0.99460389 | 0.5948 | 0 | 0 | 0 | 1 | Ref |
| 7295 | MH572418 | Unassigned | Unassigned | Unassigned | 992_0 | Clustered | 8 | VC_992_0 | 8 | 0.3873 | 1 | 0.3873 | 0 | 0 | 0 | 1 | Ref |
| 7296 | MH572419 | Unassigned | Unassigned | Unassigned | 398_0 | Clustered | 48 | VC_398_0 | 48 | 0.4813 | 1 | 0.4813 | 0 | 0 | 0 | 1 | Ref |
| 7297 | MH572422 | Unassigned | Unassigned | Unassigned | 198_0 | Clustered | 28 | VC_198_0 | 28 | 0.2429 | 0.99971175 | 0.2429 | 0 | 0 | 0 | 1 | Ref |
| 7298 | MH572423 | Unassigned | Unassigned | Unassigned | 995_0 | Clustered | 69 | VC_995_0 | 69 | 0.8547 | 1 | 0.8547 | 0 | 0 | 0 | 1 | Ref |
| 7299 | MH572424 | Unassigned | Unassigned | Unassigned | 995_0 | Clustered | 69 | VC_995_0 | 69 | 0.8547 | 1 | 0.8547 | 0 | 0 | 0 | 1 | Ref |
| 7300 | MH572425 | Unassigned | Unassigned | Unassigned | 998_0 | Clustered | 2 | VC_998_0 | 2 | 0.0515 | 0.93901809 | 0.0484 | 0 | 0 | 0 | 1 | Ref |
| 7301 | MH572426 | Unassigned | Unassigned | Unassigned | 999_0 | Clustered | 2 | VC_999_0 | 2 | 0.0273 | 0.73989325 | 0.0202 | 0 | 0 | 0 | 1 | Ref |
| 7302 | MH572427 | Unassigned | Unassigned | Unassigned | 995_0 | Clustered | 69 | VC_995_0 | 69 | 0.8547 | 1 | 0.8547 | 0 | 0 | 0 | 1 | Ref |
| 7303 | MH572428 | Unassigned | Unassigned | Unassigned | 995_0 | Clustered | 69 | VC_995_0 | 69 | 0.8547 | 1 | 0.8547 | 0 | 0 | 0 | 1 | Ref |
| 7304 | MH572430 | Unassigned | Unassigned | Unassigned | 403_0 | Clustered | 20 | VC_403_0 | 20 | 0.6598 | 1 | 0.6598 | 0 | 0 | 0 | 1 | Ref |
| 7305 | MH572431 | Unassigned | Unassigned | Unassigned | 398_0 | Clustered | 48 | VC_398_0 | 48 | 0.4813 | 1 | 0.4813 | 0 | 0 | 0 | 1 | Ref |
| 7306 | MH572434 | Unassigned | Unassigned | Unassigned | 201_0 | Clustered | 3 | VC_201_0 | 3 | 0.0998 | 0.69690761 | 0.0695 | 0 | 0 | 0 | 1 | Ref |
| 7307 | MH572435 | Unassigned | Unassigned | Unassigned | 404_0 | Clustered | 25 | VC_404_0 | 25 | 0.5649 | 1 | 0.5649 | 0 | 0 | 0 | 1 | Ref |
| 7308 | MH572436 | Unassigned | Unassigned | Unassigned | 404_0 | Clustered | 25 | VC_404_0 | 25 | 0.5649 | 1 | 0.5649 | 0 | 0 | 0 | 1 | Ref |
| 7309 | MH572437 | Unassigned | Unassigned | Unassigned | 404_0 | Clustered | 25 | VC_404_0 | 25 | 0.5649 | 1 | 0.5649 | 0 | 0 | 0 | 1 | Ref |
| 7310 | MH572438 | Unassigned | Unassigned | Unassigned | 198_0 | Clustered | 28 | VC_198_0 | 28 | 0.2429 | 0.99971175 | 0.2429 | 0 | 0 | 0 | 1 | Ref |
| 7311 | MH572439 | Unassigned | Unassigned | Unassigned | 398_0 | Clustered | 48 | VC_398_0 | 48 | 0.4813 | 1 | 0.4813 | 0 | 0 | 0 | 1 | Ref |
| 7312 | MH572441 | Unassigned | Unassigned | Unassigned | 398_0 | Clustered | 48 | VC_398_0 | 48 | 0.4813 | 1 | 0.4813 | 0 | 0 | 0 | 1 | Ref |
| 7313 | MH572443 | Unassigned | Unassigned | Unassigned | 398_0 | Clustered | 48 | VC_398_0 | 48 | 0.4813 | 1 | 0.4813 | 0 | 0 | 0 | 1 | Ref |
| 7314 | MH572444 | Unassigned | Unassigned | Unassigned | 198_0 | Clustered | 28 | VC_198_0 | 28 | 0.2429 | 0.99971175 | 0.2429 | 0 | 0 | 0 | 1 | Ref |
| 7315 | MH572445 | Unassigned | Unassigned | Unassigned | 398_0 | Clustered | 48 | VC_398_0 | 48 | 0.4813 | 1 | 0.4813 | 0 | 0 | 0 | 1 | Ref |
| 7316 | MH572446 | Unassigned | Unassigned | Unassigned | 398_0 | Clustered | 48 | VC_398_0 | 48 | 0.4813 | 1 | 0.4813 | 0 | 0 | 0 | 1 | Ref |
| 7317 | MH572448 | Unassigned | Unassigned | Unassigned | 198_0 | Clustered | 28 | VC_198_0 | 28 | 0.2429 | 0.99971175 | 0.2429 | 0 | 0 | 0 | 1 | Ref |
| 7318 | MH572451 | Unassigned | Unassigned | Unassigned | 398_0 | Clustered | 48 | VC_398_0 | 48 | 0.4813 | 1 | 0.4813 | 0 | 0 | 0 | 1 | Ref |
| 7319 | MH572452 | Unassigned | Unassigned | Unassigned | 198_0 | Clustered | 28 | VC_198_0 | 28 | 0.2429 | 0.99971175 | 0.2429 | 0 | 0 | 0 | 1 | Ref |
| 7320 | MH572453 | Unassigned | Unassigned | Unassigned | 199_0 | Clustered | 12 | VC_199_0 | 12 | 0.448 | 1 | 0.448 | 0 | 0 | 0 | 1 | Ref |
| 7321 | MH572454 | Unassigned | Unassigned | Unassigned | 198_0 | Clustered | 28 | VC_198_0 | 28 | 0.2429 | 0.99971175 | 0.2429 | 0 | 0 | 0 | 1 | Ref |
| 7322 | MH572455 | Unassigned | Unassigned | Unassigned | 992_0 | Clustered | 8 | VC_992_0 | 8 | 0.3873 | 1 | 0.3873 | 0 | 0 | 0 | 1 | Ref |
| 7323 | MH572457 | Unassigned | Unassigned | Unassigned | 1079_0 | Clustered | 6 | VC_1079_0 | 6 | 0.9301 | 0.89096579 | 0.8287 | 0 | 0 | 0 | 1 | Ref |
| 7324 | MH572458 | Unassigned | Unassigned | Unassigned | 1088_0 | Clustered | 3 | VC_1088_0 | 3 | 0.7739 | 0.82711071 | 0.6401 | 0 | 0 | 0 | 1 | Ref |
| 7325 | MH572459 | Unassigned | Unassigned | Unassigned | 1062_0 | Clustered | 45 | VC_1062_0 | 45 | 0.9734 | 0.50949847 | 0.4959 | 0 | 0 | 0 | 1 | Ref |
| 7326 | MH572460 | Unassigned | Unassigned | Unassigned | 1062_0 | Clustered | 45 | VC_1062_0 | 45 | 0.9734 | 0.50949847 | 0.4959 | 0 | 0 | 0 | 1 | Ref |
| 7327 | MH572461 | Unassigned | Unassigned | Unassigned | 1062_0 | Clustered | 45 | VC_1062_0 | 45 | 0.9734 | 0.50949847 | 0.4959 | 0 | 0 | 0 | 1 | Ref |
| 7328 | MH572462 | Unassigned | Unassigned | Unassigned | 1062_0 | Clustered | 45 | VC_1062_0 | 45 | 0.9734 | 0.50949847 | 0.4959 | 0 | 0 | 0 | 1 | Ref |
| 7329 | MH572463 | Unassigned | Unassigned | Unassigned | 1062_0 | Clustered | 45 | VC_1062_0 | 45 | 0.9734 | 0.50949847 | 0.4959 | 0 | 0 | 0 | 1 | Ref |
| 7330 | MH572465 | Unassigned | Unassigned | Unassigned | 1062_0 | Clustered | 45 | VC_1062_0 | 45 | 0.9734 | 0.50949847 | 0.4959 | 0 | 0 | 0 | 1 | Ref |
| 7331 | MH572466 | Unassigned | Unassigned | Unassigned | 995_0 | Clustered | 69 | VC_995_0 | 69 | 0.8547 | 1 | 0.8547 | 0 | 0 | 0 | 1 | Ref |
| 7332 | MH572467 | Unassigned | Unassigned | Unassigned | 1084_0 | Clustered | 9 | VC_1084_0 | 9 | 0.6558 | 1 | 0.6558 | 0 | 0 | 0 | 1 | Ref |
| 7333 | MH572468 | Unassigned | Unassigned | Unassigned | 1084_0 | Clustered | 9 | VC_1084_0 | 9 | 0.6558 | 1 | 0.6558 | 0 | 0 | 0 | 1 | Ref |
| 7334 | MH572469 | Unassigned | Unassigned | Unassigned | 404_0 | Clustered | 25 | VC_404_0 | 25 | 0.5649 | 1 | 0.5649 | 0 | 0 | 0 | 1 | Ref |
| 7335 | MH572470 | Unassigned | Unassigned | Unassigned | 403_0 | Clustered | 20 | VC_403_0 | 20 | 0.6598 | 1 | 0.6598 | 0 | 0 | 0 | 1 | Ref |
| 7336 | MH572471 | Unassigned | Unassigned | Unassigned | 403_0 | Clustered | 20 | VC_403_0 | 20 | 0.6598 | 1 | 0.6598 | 0 | 0 | 0 | 1 | Ref |
| 7337 | MH572472 | Unassigned | Unassigned | Unassigned | 403_0 | Clustered | 20 | VC_403_0 | 20 | 0.6598 | 1 | 0.6598 | 0 | 0 | 0 | 1 | Ref |
| 7338 | MH572473 | Unassigned | Unassigned | Unassigned | 398_0 | Clustered | 48 | VC_398_0 | 48 | 0.4813 | 1 | 0.4813 | 0 | 0 | 0 | 1 | Ref |
| 7339 | MH572475 | Unassigned | Unassigned | Unassigned | 201_0 | Clustered | 3 | VC_201_0 | 3 | 0.0998 | 0.69690761 | 0.0695 | 0 | 0 | 0 | 1 | Ref |
| 7340 | MH572479 | Unassigned | Unassigned | Unassigned | 401_0 | Clustered/Singleton | 1 | VC_401_0 | 1 | 0 | 1 | 0 | 0 | 0 | 0 | 1 | Ref |
| 7341 | MH572480 | Unassigned | Unassigned | Unassigned | 404_0 | Clustered | 25 | VC_404_0 | 25 | 0.5649 | 1 | 0.5649 | 0 | 0 | 0 | 1 | Ref |
| 7342 | MH572482 | Unassigned | Unassigned | Unassigned | 1079_0 | Clustered | 6 | VC_1079_0 | 6 | 0.9301 | 0.89096579 | 0.8287 | 0 | 0 | 0 | 1 | Ref |
| 7343 | MH572483 | Unassigned | Unassigned | Unassigned | 1079_0 | Clustered | 6 | VC_1079_0 | 6 | 0.9301 | 0.89096579 | 0.8287 | 0 | 0 | 0 | 1 | Ref |
| 7344 | MH572484 | Unassigned | Unassigned | Unassigned | 1338_0 | Clustered | 4 | VC_1338_0 | 4 | 1 | 1 | 1 | 0 | 0 | 0 | 1 | Ref |
| 7345 | MH572485 | Unassigned | Unassigned | Unassigned | 1323_0 | Clustered | 6 | VC_1323_0 | 6 | 0.598 | 0.99460389 | 0.5948 | 0 | 0 | 0 | 1 | Ref |
| 7346 | MH572486 | Unassigned | Unassigned | Unassigned | 1077_0 | Clustered | 48 | VC_1077_0 | 48 | 0.8525 | 1 | 0.8525 | 0 | 0 | 0 | 1 | Ref |
| 7347 | MH572487 | Unassigned | Unassigned | Unassigned | 398_0 | Clustered | 48 | VC_398_0 | 48 | 0.4813 | 1 | 0.4813 | 0 | 0 | 0 | 1 | Ref |
| 7348 | MH572489 | Unassigned | Unassigned | Unassigned | 1089_0 | Clustered | 2 | VC_1089_0 | 2 | 0.546 | 0.5 | 0.273 | 0 | 0 | 0 | 1 | Ref |
| 7349 | MH572490 | Unassigned | Unassigned | Unassigned | 1357_0 | Clustered | 6 | VC_1357_0 | 6 | 1 | 1 | 1 | 0 | 0 | 0 | 1 | Ref |
| 7350 | MH572491 | Unassigned | Unassigned | Unassigned | 1357_0 | Clustered | 6 | VC_1357_0 | 6 | 1 | 1 | 1 | 0 | 0 | 0 | 1 | Ref |
| 7351 | MH572492 | Unassigned | Unassigned | Unassigned | 398_0 | Clustered | 48 | VC_398_0 | 48 | 0.4813 | 1 | 0.4813 | 0 | 0 | 0 | 1 | Ref |
| 7352 | MH572493 | Unassigned | Unassigned | Unassigned | 398_0 | Clustered | 48 | VC_398_0 | 48 | 0.4813 | 1 | 0.4813 | 0 | 0 | 0 | 1 | Ref |
| 7353 | MH572494 | Unassigned | Unassigned | Unassigned | 398_0 | Clustered | 48 | VC_398_0 | 48 | 0.4813 | 1 | 0.4813 | 0 | 0 | 0 | 1 | Ref |
| 7354 | MH572495 | Unassigned | Unassigned | Unassigned | 995_0 | Clustered | 69 | VC_995_0 | 69 | 0.8547 | 1 | 0.8547 | 0 | 0 | 0 | 1 | Ref |
| 7355 | MH572496 | Unassigned | Unassigned | Unassigned | 1084_0 | Clustered | 9 | VC_1084_0 | 9 | 0.6558 | 1 | 0.6558 | 0 | 0 | 0 | 1 | Ref |
| 7356 | MH572497 | Unassigned | Unassigned | Unassigned | 1084_0 | Clustered | 9 | VC_1084_0 | 9 | 0.6558 | 1 | 0.6558 | 0 | 0 | 0 | 1 | Ref |
| 7357 | MH572498 | Unassigned | Unassigned | Unassigned | 1084_0 | Clustered | 9 | VC_1084_0 | 9 | 0.6558 | 1 | 0.6558 | 0 | 0 | 0 | 1 | Ref |
| 7358 | MH572499 | Unassigned | Unassigned | Unassigned | 1084_0 | Clustered | 9 | VC_1084_0 | 9 | 0.6558 | 1 | 0.6558 | 0 | 0 | 0 | 1 | Ref |
| 7359 | MH572500 | Unassigned | Unassigned | Unassigned | 1084_0 | Clustered | 9 | VC_1084_0 | 9 | 0.6558 | 1 | 0.6558 | 0 | 0 | 0 | 1 | Ref |
| 7360 | MH572503 | Unassigned | Unassigned | Unassigned | 398_0 | Clustered | 48 | VC_398_0 | 48 | 0.4813 | 1 | 0.4813 | 0 | 0 | 0 | 1 | Ref |
| 7361 | MH572504 | Unassigned | Unassigned | Unassigned | 995_0 | Clustered | 69 | VC_995_0 | 69 | 0.8547 | 1 | 0.8547 | 0 | 0 | 0 | 1 | Ref |
| 7362 | MH572505 | Unassigned | Unassigned | Unassigned | 403_0 | Clustered | 20 | VC_403_0 | 20 | 0.6598 | 1 | 0.6598 | 0 | 0 | 0 | 1 | Ref |
| 7363 | MH572506 | Unassigned | Unassigned | Unassigned | 403_0 | Clustered | 20 | VC_403_0 | 20 | 0.6598 | 1 | 0.6598 | 0 | 0 | 0 | 1 | Ref |
| 7364 | MH572507 | Unassigned | Unassigned | Unassigned | 403_0 | Clustered | 20 | VC_403_0 | 20 | 0.6598 | 1 | 0.6598 | 0 | 0 | 0 | 1 | Ref |
| 7365 | MH572508 | Unassigned | Unassigned | Unassigned | 404_0 | Clustered | 25 | VC_404_0 | 25 | 0.5649 | 1 | 0.5649 | 0 | 0 | 0 | 1 | Ref |
| 7366 | MH572509 | Unassigned | Unassigned | Unassigned | 404_0 | Clustered | 25 | VC_404_0 | 25 | 0.5649 | 1 | 0.5649 | 0 | 0 | 0 | 1 | Ref |
| 7367 | MH572510 | Unassigned | Unassigned | Unassigned | 398_0 | Clustered | 48 | VC_398_0 | 48 | 0.4813 | 1 | 0.4813 | 0 | 0 | 0 | 1 | Ref |
| 7368 | MH572511 | Unassigned | Unassigned | Unassigned | 398_0 | Clustered | 48 | VC_398_0 | 48 | 0.4813 | 1 | 0.4813 | 0 | 0 | 0 | 1 | Ref |
| 7369 | MH572512 | Unassigned | Unassigned | Unassigned | 402_0 | Clustered/Singleton | 1 | VC_402_0 | 1 | 0 | 1 | 0 | 0 | 0 | 0 | 1 | Ref |
| 7370 | MH572513 | Unassigned | Unassigned | Unassigned | 199_0 | Clustered | 12 | VC_199_0 | 12 | 0.448 | 1 | 0.448 | 0 | 0 | 0 | 1 | Ref |
| 7371 | MH572514 | Unassigned | Unassigned | Unassigned | 199_0 | Clustered | 12 | VC_199_0 | 12 | 0.448 | 1 | 0.448 | 0 | 0 | 0 | 1 | Ref |
| 7372 | MH572515 | Unassigned | Unassigned | Unassigned | 992_0 | Clustered | 8 | VC_992_0 | 8 | 0.3873 | 1 | 0.3873 | 0 | 0 | 0 | 1 | Ref |
| 7373 | MH572516 | Unassigned | Unassigned | Unassigned | 1356_0 | Clustered | 8 | VC_1356_0 | 8 | 0.8729 | 0.99998222 | 0.8729 | 0 | 0 | 0 | 1 | Ref |
| 7374 | MH572517 | Unassigned | Unassigned | Unassigned | 1357_0 | Clustered | 6 | VC_1357_0 | 6 | 1 | 1 | 1 | 0 | 0 | 0 | 1 | Ref |
| 7375 | MH572518 | Unassigned | Unassigned | Unassigned | 1357_0 | Clustered | 6 | VC_1357_0 | 6 | 1 | 1 | 1 | 0 | 0 | 0 | 1 | Ref |
| 7376 | MH572519 | Unassigned | Unassigned | Unassigned | 1357_0 | Clustered | 6 | VC_1357_0 | 6 | 1 | 1 | 1 | 0 | 0 | 0 | 1 | Ref |
| 7377 | MH572520 | Unassigned | Unassigned | Unassigned | 1357_0 | Clustered | 6 | VC_1357_0 | 6 | 1 | 1 | 1 | 0 | 0 | 0 | 1 | Ref |
| 7378 | MH572521 | Unassigned | Unassigned | Unassigned | 1077_0 | Clustered | 48 | VC_1077_0 | 48 | 0.8525 | 1 | 0.8525 | 0 | 0 | 0 | 1 | Ref |
| 7379 | MH572522 | Unassigned | Unassigned | Unassigned | 1323_0 | Clustered | 6 | VC_1323_0 | 6 | 0.598 | 0.99460389 | 0.5948 | 0 | 0 | 0 | 1 | Ref |
| 7380 | MH572524 | Unassigned | Unassigned | Unassigned | 1324_0 | Clustered | 4 | VC_1324_0 | 4 | 0.6008 | 0.54407361 | 0.3269 | 0 | 0 | 0 | 1 | Ref |
| 7381 | MH572525 | Unassigned | Unassigned | Unassigned | 1243_0 | Clustered | 52 | VC_1243_0 | 52 | 0.9126 | 0.92708311 | 0.8461 | 0 | 0 | 0 | 1 | Ref |
| 7382 | MH572526 | Unassigned | Unassigned | Unassigned | 1243_0 | Clustered | 52 | VC_1243_0 | 52 | 0.9126 | 0.92708311 | 0.8461 | 0 | 0 | 0 | 1 | Ref |
| 7383 | MH576948 | Unassigned | Unassigned | Unassigned | 950_0 | Clustered | 38 | VC_950_0 | 38 | 0.9443 | 1 | 0.9443 | 0 | 0 | 0 | 1 | Ref |
| 7384 | MH576949 | Unassigned | Unassigned | Unassigned | 514_0 | Clustered | 75 | VC_514_0 | 75 | 0.9402 | 1 | 0.9402 | 0 | 0 | 0 | 1 | Ref |
| 7385 | MH576950 | Unassigned | Unassigned | Unassigned | 950_0 | Clustered | 38 | VC_950_0 | 38 | 0.9443 | 1 | 0.9443 | 0 | 0 | 0 | 1 | Ref |
| 7386 | MH576951 | Unassigned | Unassigned | Unassigned | 950_0 | Clustered | 38 | VC_950_0 | 38 | 0.9443 | 1 | 0.9443 | 0 | 0 | 0 | 1 | Ref |
| 7387 | MH576952 | Unassigned | Unassigned | Unassigned | 950_0 | Clustered | 38 | VC_950_0 | 38 | 0.9443 | 1 | 0.9443 | 0 | 0 | 0 | 1 | Ref |
| 7388 | MH576953 | Unassigned | Unassigned | Unassigned | 551_7 | Clustered | 28 | VC_551_7 | 28 | 0.1661 | 1 | 0.1661 | 0 | 0 | 0 | 0.8519 | Ref |
| 7389 | MH576954 | Unassigned | Unassigned | Unassigned | 371_4 | Clustered | 5 | VC_371_4 | 5 | 0.0135 | 0.99999971 | 0.0135 | 0 | 0 | 0 | 1 | Ref |
| 7390 | MH576955 | Unassigned | Unassigned | Unassigned | 950_0 | Clustered | 38 | VC_950_0 | 38 | 0.9443 | 1 | 0.9443 | 0 | 0 | 0 | 1 | Ref |
| 7391 | MH576956 | Unassigned | Unassigned | Unassigned | 551_1 | Clustered | 4 | VC_551_1 | 4 | 0.0159 | 0.99968337 | 0.0159 | 0 | 0 | 0 | 0.6667 | Ref |
| 7392 | MH576957 | Unassigned | Unassigned | Unassigned | 950_0 | Clustered | 38 | VC_950_0 | 38 | 0.9443 | 1 | 0.9443 | 0 | 0 | 0 | 1 | Ref |
| 7393 | MH576958 | Unassigned | Unassigned | Unassigned | 371_13 | Clustered | 33 | VC_371_13 | 33 | 0.179 | 1 | 0.179 | 0 | 0 | 0 | 0.8769 | Ref |
| 7394 | MH576959 | Unassigned | Unassigned | Unassigned | 325_23 | Clustered | 17 | VC_325_23 | 17 | 0.0716 | 1 | 0.0716 | 0 | 0 | 0 | 0.6397 | Ref |
| 7395 | MH576960 | Unassigned | Unassigned | Unassigned | 767_0 | Clustered | 34 | VC_767_0 | 34 | 0.9173 | 1 | 0.9173 | 0 | 0 | 0 | 1 | Ref |
| 7396 | MH576961 | Unassigned | Unassigned | Unassigned | 950_0 | Clustered | 38 | VC_950_0 | 38 | 0.9443 | 1 | 0.9443 | 0 | 0 | 0 | 1 | Ref |
| 7397 | MH576962 | Unassigned | Unassigned | Unassigned | 1158_0 | Clustered | 3 | VC_1158_0 | 3 | 0.3179 | 0.99335558 | 0.3158 | 0 | 0 | 0 | 1 | Ref |
| 7398 | MH576963 | Unassigned | Unassigned | Unassigned | 1344_0 | Clustered | 78 | VC_1344_0 | 78 | 0.9732 | 1 | 0.9732 | 0 | 0 | 0 | 1 | Ref |
| 7399 | MH576964 | Unassigned | Unassigned | Unassigned | 1071_0 | Clustered | 20 | VC_1071_0 | 20 | 0.3268 | 1 | 0.3268 | 0 | 0 | 0 | 0.9842 | Ref |
| 7400 | MH576965 | Unassigned | Unassigned | Unassigned | 1071_0 | Clustered | 20 | VC_1071_0 | 20 | 0.3268 | 1 | 0.3268 | 0 | 0 | 0 | 0.9842 | Ref |
| 7401 | MH576966 | Unassigned | Unassigned | Unassigned | 238_0 | Clustered | 17 | VC_238_0 | 17 | 0.6109 | 1 | 0.6109 | 0 | 0 | 0 | 1 | Ref |
| 7402 | MH576967 | Unassigned | Unassigned | Unassigned | 371_4 | Clustered | 5 | VC_371_4 | 5 | 0.0135 | 0.99999971 | 0.0135 | 0 | 0 | 0 | 1 | Ref |
| 7403 | MH576968 | Unassigned | Unassigned | Unassigned | 1071_0 | Clustered | 20 | VC_1071_0 | 20 | 0.3268 | 1 | 0.3268 | 0 | 0 | 0 | 0.9842 | Ref |
| 7404 | MH576969 | Unassigned | Unassigned | Unassigned | 1176_0 | Clustered | 41 | VC_1176_0 | 41 | 0.9558 | 1 | 0.9558 | 0 | 0 | 0 | 0.9976 | Ref |
| 7405 | MH576970 | Unassigned | Unassigned | Unassigned | 371_15 | Clustered | 44 | VC_371_15 | 44 | 0.1391 | 1 | 0.1391 | 0 | 0 | 0 | 0.7474 | Ref |
| 7406 | MH576971 | Unassigned | Unassigned | Unassigned | 325_23 | Clustered | 17 | VC_325_23 | 17 | 0.0716 | 1 | 0.0716 | 0 | 0 | 0 | 0.6397 | Ref |
| 7407 | MH576972 | Unassigned | Unassigned | Unassigned | 570_0 | Clustered | 9 | VC_570_0 | 9 | 0.499 | 1 | 0.499 | 0 | 0 | 0 | 1 | Ref |
| 7408 | MH576973 | Unassigned | Unassigned | Unassigned | 888_1 | Clustered | 3 | VC_888_1 | 3 | 0.0289 | 0.99848448 | 0.0289 | 0 | 0 | 0 | 1 | Ref |
| 7409 | MH576974 | Unassigned | Unassigned | Unassigned | 325_23 | Clustered | 17 | VC_325_23 | 17 | 0.0716 | 1 | 0.0716 | 0 | 0 | 0 | 0.6397 | Ref |
| 7410 | MH576975 | Unassigned | Unassigned | Unassigned | 325_46 | Clustered | 2 | VC_325_46 | 2 | 0.004 | 0.95751617 | 0.0039 | 0 | 0 | 0 | 1 | Ref |
| 7411 | MH576976 | Unassigned | Unassigned | Unassigned | 869_0 | Clustered | 9 | VC_869_0 | 9 | 0.2624 | 1 | 0.2624 | 0 | 0 | 0 | 1 | Ref |
| 7412 | MH576977 | Unassigned | Unassigned | Unassigned | 509_0 | Clustered | 37 | VC_509_0 | 37 | 0.9689 | 1 | 0.9689 | 0 | 0 | 0 | 1 | Ref |
| 7413 | MH579717 | Unassigned | Unassigned | Unassigned | 480_0 | Clustered | 19 | VC_480_0 | 19 | 0.5445 | 1 | 0.5445 | 0 | 0 | 0 | 1 | Ref |
| 7414 | MH586730 | Unassigned | Unassigned | Unassigned | 1049_0 | Clustered | 8 | VC_1049_0 | 8 | 0.6413 | 1 | 0.6413 | 0 | 0 | 0 | 1 | Ref |
| 7415 | MH586731 | Unassigned | Unassigned | Unassigned | 329_0 | Clustered | 2 | VC_329_0 | 2 | 0.0051 | 0.95464582 | 0.0048 | 0 | 0 | 0 | 1 | Ref |
| 7416 | MH587638 | Unassigned | Unassigned | Unassigned | 823_0 | Clustered | 124 | VC_823_0 | 124 | 0.9492 | 1 | 0.9492 | 0 | 0 | 0 | 1 | Ref |
| 7417 | MH588544 | Unassigned | Unassigned | Unassigned | 976_3 | Clustered | 2 | VC_976_3 | 2 | 0.0258 | 0.88402892 | 0.0228 | 0 | 0 | 0 | 1 | Ref |
| 7418 | MH588545 | Unassigned | Unassigned | Unassigned | 976_2 | Clustered | 2 | VC_976_2 | 2 | 0.0278 | 0.95410794 | 0.0266 | 0 | 0 | 0 | 1 | Ref |
| 7419 | MH590587 | Unassigned | Unassigned | Unassigned | 551_8 | Clustered | 19 | VC_551_8 | 19 | 0.1039 | 1 | 0.1039 | 0 | 0 | 0 | 0.8713 | Ref |
| 7420 | MH590588 | Unassigned | Unassigned | Unassigned | 371_11 | Clustered | 31 | VC_371_11 | 31 | 0.0927 | 1 | 0.0927 | 0 | 0 | 0 | 0.7011 | Ref |
| 7421 | MH590589 | Unassigned | Unassigned | Unassigned | 1072_2 | Clustered | 5 | VC_1072_2 | 5 | 0.1003 | 0.99999315 | 0.1003 | 0 | 0 | 0 | 1 | Ref |
| 7422 | MH590591 | Unassigned | Unassigned | Unassigned | 508_0 | Clustered | 32 | VC_508_0 | 32 | 0.906 | 1 | 0.906 | 0 | 0 | 0 | 1 | Ref |
| 7423 | MH590592 | Unassigned | Unassigned | Unassigned | 556_0 | Clustered | 21 | VC_556_0 | 21 | 0.9912 | 1 | 0.9912 | 0 | 0 | 0 | 1 | Ref |
| 7424 | MH590593 | Unassigned | Unassigned | Unassigned | 558_22 | Clustered | 4 | VC_558_22 | 4 | 0.01 | 0.99312819 | 0.0099 | 0 | 0 | 0 | 0.8333 | Ref |
| 7425 | MH590594 | Unassigned | Unassigned | Unassigned | 371_10 | Clustered/Singleton | 1 | VC_371_10 | 1 | 0 | 1 | 0 | 0 | 0 | 0 | 1 | Ref |
| 7426 | MH590595 | Unassigned | Unassigned | Unassigned | 325_53 | Clustered | 7 | VC_325_53 | 7 | 0.0242 | 1 | 0.0242 | 0 | 0 | 0 | 0.9048 | Ref |
| 7427 | MH590596 | Unassigned | Unassigned | Unassigned | 520_78 | Clustered | 2 | VC_520_78 | 2 | 0.008 | 0.95761153 | 0.0076 | 0 | 0 | 0 | 1 | Ref |
| 7428 | MH590597 | Unassigned | Unassigned | Unassigned | 1071_0 | Clustered | 20 | VC_1071_0 | 20 | 0.3268 | 1 | 0.3268 | 0 | 0 | 0 | 0.9842 | Ref |
| 7429 | MH590598 | Unassigned | Unassigned | Unassigned | 520_25 | Clustered/Singleton | 1 | VC_520_25 | 1 | 0 | 1 | 0 | 0 | 0 | 0 | 1 | Ref |
| 7430 | MH590599 | Unassigned | Unassigned | Unassigned | 1071_0 | Clustered | 20 | VC_1071_0 | 20 | 0.3268 | 1 | 0.3268 | 0 | 0 | 0 | 0.9842 | Ref |
| 7431 | MH590600 | Unassigned | Unassigned | Unassigned | 1352_0 | Clustered | 31 | VC_1352_0 | 31 | 0.849 | 1 | 0.849 | 0 | 0 | 0 | 1 | Ref |
| 7432 | MH590601 | Unassigned | Unassigned | Unassigned | 981_1 | Clustered | 26 | VC_981_1 | 26 | 0.7009 | 1 | 0.7009 | 0 | 0 | 0 | 1 | Ref |
| 7433 | MH590602 | Unassigned | Unassigned | Unassigned | 520_38 | Clustered | 6 | VC_520_38 | 6 | 0.0519 | 1 | 0.0519 | 0 | 0 | 0 | 0.8667 | Ref |
| 7434 | MH590603 | Unassigned | Unassigned | Unassigned | 1182_1 | Clustered/Singleton | 1 | VC_1182_1 | 1 | 0 | 1 | 0 | 0 | 0 | 0 | 1 | Ref |
| 7435 | MH590604 | Unassigned | Unassigned | Unassigned | 1163_0 | Clustered | 28 | VC_1163_0 | 28 | 0.4923 | 1 | 0.4923 | 0 | 0 | 0 | 1 | Ref |
| 7436 | MH590605 | Unassigned | Unassigned | Unassigned | 761_0 | Clustered | 67 | VC_761_0 | 67 | 0.8947 | 1 | 0.8947 | 0 | 0 | 0 | 1 | Ref |
| 7437 | MH590606 | Unassigned | Unassigned | Unassigned | 1163_0 | Clustered | 28 | VC_1163_0 | 28 | 0.4923 | 1 | 0.4923 | 0 | 0 | 0 | 1 | Ref |
| 7438 | MH595538 | Unassigned | Unassigned | Unassigned | 346_3 | Clustered/Singleton | 1 | VC_346_3 | 1 | 0 | 1 | 0 | 0 | 0 | 0 | 1 | Ref |
| 7439 | MH598512 | Unassigned | Unassigned | Unassigned | 828_0 | Clustered | 3 | VC_828_0 | 3 | 0.4748 | 0.99843555 | 0.4741 | 0 | 0 | 0 | 1 | Ref |
| 7440 | MH598798 | Unassigned | Unassigned | Unassigned | 480_0 | Clustered | 19 | VC_480_0 | 19 | 0.5445 | 1 | 0.5445 | 0 | 0 | 0 | 1 | Ref |
| 7441 | MH598800 | Unassigned | Unassigned | Unassigned | 480_0 | Clustered | 19 | VC_480_0 | 19 | 0.5445 | 1 | 0.5445 | 0 | 0 | 0 | 1 | Ref |
| 7442 | MH598802 | Unassigned | Unassigned | Unassigned | 480_0 | Clustered | 19 | VC_480_0 | 19 | 0.5445 | 1 | 0.5445 | 0 | 0 | 0 | 1 | Ref |
| 7443 | MH598803 | Unassigned | Unassigned | Unassigned | 480_0 | Clustered | 19 | VC_480_0 | 19 | 0.5445 | 1 | 0.5445 | 0 | 0 | 0 | 1 | Ref |
| 7444 | MH598804 | Unassigned | Unassigned | Unassigned | 480_0 | Clustered | 19 | VC_480_0 | 19 | 0.5445 | 1 | 0.5445 | 0 | 0 | 0 | 1 | Ref |
| 7445 | MH598805 | Unassigned | Unassigned | Unassigned | 480_0 | Clustered | 19 | VC_480_0 | 19 | 0.5445 | 1 | 0.5445 | 0 | 0 | 0 | 1 | Ref |
| 7446 | MH598807 | Unassigned | Unassigned | Unassigned | 480_0 | Clustered | 19 | VC_480_0 | 19 | 0.5445 | 1 | 0.5445 | 0 | 0 | 0 | 1 | Ref |
| 7447 | MH599087 | Unassigned | Unassigned | Unassigned | 882_0 | Clustered | 20 | VC_882_0 | 20 | 0.9917 | 1 | 0.9917 | 0 | 0 | 0 | 1 | Ref |
| 7448 | MH606185 | Unassigned | Unassigned | Unassigned | 85_5 | Clustered/Singleton | 1 | VC_85_5 | 1 | 0 | 1 | 0 | 0 | 0 | 0 | 1 | Ref |
| 7449 | MH616963 | Unassigned | Unassigned | Unassigned | 673_0 | Clustered | 15 | VC_673_0 | 15 | 0.8234 | 1 | 0.8234 | 0 | 0 | 0 | 0.7619 | Ref |
| 7450 | MH618488 | Unassigned | Unassigned | Unassigned | 108_0 | Clustered | 33 | VC_108_0 | 33 | 0.949 | 1 | 0.949 | 0 | 0 | 0 | 1 | Ref |

| | | | | | | | | | | | | | | | | | |
|---|---|---|---|---|---|---|---|---|---|---|---|---|---|---|---|---|---|
| 7451 | MH629685 | Unassigned | Unassigned | Unassigned | 432_16 | Clustered/Singleton | 1 | VC_432_16 | 1 | 0 | 1 | 0 | 0 | 0 | 0 | 1 | Ref |
| 7452 | MH631453 | Unassigned | Unassigned | Unassigned | 592_0 | Clustered | 50 | VC_592_0 | 50 | 0.7314 | 1 | 0.7314 | 0 | 0 | 0 | 1 | Ref |
| 7453 | MH631454 | Unassigned | Unassigned | Unassigned | 191_19 | Clustered | 12 | VC_191_19 | 12 | 0.0655 | 1 | 0.0655 | 0 | 0 | 0 | 0.8636 | Ref |
| 7454 | MH632117 | Unassigned | Unassigned | Unassigned | 325_49 | Clustered | 4 | VC_325_49 | 4 | 0.014 | 0.99995965 | 0.014 | 0 | 0 | 0 | 1 | Ref |
| 7455 | MH632118 | Unassigned | Unassigned | Unassigned | 325_27 | Clustered | 2 | VC_325_27 | 2 | 0.0044 | 0.95750955 | 0.0042 | 0 | 0 | 0 | 1 | Ref |
| 7456 | MH632119 | Unassigned | Unassigned | Unassigned | 520_93 | Clustered | 2 | VC_520_93 | 2 | 0.0104 | 0.95762079 | 0.0099 | 0 | 0 | 0 | 1 | Ref |
| 7457 | MH633484 | Unassigned | Unassigned | Unassigned | 938_0 | Clustered | 99 | VC_938_0 | 99 | 0.9119 | 1 | 0.9119 | 0 | 0 | 0 | 0.9928 | Ref |
| 7458 | MH633485 | Unassigned | Unassigned | Unassigned | 938_0 | Clustered | 99 | VC_938_0 | 99 | 0.9119 | 1 | 0.9119 | 0 | 0 | 0 | 0.9928 | Ref |
| 7459 | MH633486 | Unassigned | Unassigned | Unassigned | 938_0 | Clustered | 99 | VC_938_0 | 99 | 0.9119 | 1 | 0.9119 | 0 | 0 | 0 | 0.9928 | Ref |
| 7460 | MH633487 | Unassigned | Unassigned | Unassigned | 938_0 | Clustered | 99 | VC_938_0 | 99 | 0.9119 | 1 | 0.9119 | 0 | 0 | 0 | 0.9928 | Ref |
| 7461 | MH638294 | Unassigned | Unassigned | Unassigned | 162_0 | Clustered | 18 | VC_162_0 | 18 | 0.7403 | 1 | 0.7403 | 0 | 0 | 0 | 0.8627 | Ref |
| 7462 | MH638309 | Unassigned | Unassigned | Unassigned | 187_3 | Clustered | 9 | VC_187_3 | 9 | 0.1971 | 1 | 0.1971 | 0 | 0 | 0 | 0.8611 | Ref |
| 7463 | MH638310 | Unassigned | Unassigned | Unassigned | 883_0 | Clustered | 10 | VC_883_0 | 10 | 0.2015 | 1 | 0.2015 | 0 | 0 | 0 | 1 | Ref |
| 7464 | MH638311 | Unassigned | Unassigned | Unassigned | 884_0 | Clustered | 28 | VC_884_0 | 28 | 0.6211 | 1 | 0.6211 | 0 | 0 | 0 | 0.9841 | Ref |
| 7465 | MH638312 | Unassigned | Unassigned | Unassigned | 187_3 | Clustered | 9 | VC_187_3 | 9 | 0.1971 | 1 | 0.1971 | 0 | 0 | 0 | 0.8611 | Ref |
| 7466 | MH643777 | Unassigned | Unassigned | Unassigned | 179_0 | Clustered | 53 | VC_179_0 | 53 | 0.9137 | 1 | 0.9137 | 0 | 0 | 0 | 1 | Ref |
| 7467 | MH643778 | Unassigned | Unassigned | Unassigned | 179_0 | Clustered | 53 | VC_179_0 | 53 | 0.9137 | 1 | 0.9137 | 0 | 0 | 0 | 1 | Ref |
| 7468 | MH645904 | Unassigned | Unassigned | Unassigned | 972_0 | Clustered | 6 | VC_972_0 | 6 | 0.2892 | 1 | 0.2892 | 0 | 0 | 0 | 1 | Ref |
| 7469 | MH651166 | Unassigned | Unassigned | Unassigned | 1093_0 | Clustered | 40 | VC_1093_0 | 40 | 0.8754 | 1 | 0.8754 | 0 | 0 | 0 | 1 | Ref |
| 7470 | MH651167 | Unassigned | Unassigned | Unassigned | 685_0 | Clustered | 16 | VC_685_0 | 16 | 0.2972 | 1 | 0.2972 | 0 | 0 | 0 | 0.9167 | Ref |
| 7471 | MH651168 | Unassigned | Unassigned | Unassigned | 1093_0 | Clustered | 40 | VC_1093_0 | 40 | 0.8754 | 1 | 0.8754 | 0 | 0 | 0 | 1 | Ref |
| 7472 | MH651169 | Unassigned | Unassigned | Unassigned | 520_45 | Clustered | 7 | VC_520_45 | 7 | 0.0635 | 1 | 0.0635 | 0 | 0 | 0 | 0.8095 | Ref |
| 7473 | MH651170 | Unassigned | Unassigned | Unassigned | 517_1 | Clustered | 108 | VC_517_1 | 108 | 0.8483 | 1 | 0.8483 | 0 | 0 | 0 | 0.9955 | Ref |
| 7474 | MH651171 | Unassigned | Unassigned | Unassigned | 238_0 | Clustered | 17 | VC_238_0 | 17 | 0.6109 | 1 | 0.6109 | 0 | 0 | 0 | 1 | Ref |
| 7475 | MH651172 | Unassigned | Unassigned | Unassigned | 1072_2 | Clustered | 5 | VC_1072_2 | 5 | 0.1003 | 0.99999315 | 0.1003 | 0 | 0 | 0 | 1 | Ref |
| 7476 | MH651174 | Unassigned | Unassigned | Unassigned | 551_6 | Clustered | 22 | VC_551_6 | 22 | 0.1214 | 1 | 0.1214 | 0 | 0 | 0 | 0.8095 | Ref |
| 7477 | MH651175 | Unassigned | Unassigned | Unassigned | 371_16 | Clustered | 42 | VC_371_16 | 42 | 0.1341 | 1 | 0.1341 | 0 | 0 | 0 | 0.7282 | Ref |
| 7478 | MH651176 | Unassigned | Unassigned | Unassigned | 557_1 | Clustered | 11 | VC_557_1 | 11 | 0.4945 | 1 | 0.4945 | 0 | 0 | 0 | 0.7818 | Ref |
| 7479 | MH651177 | Unassigned | Unassigned | Unassigned | 371_4 | Clustered | 5 | VC_371_4 | 5 | 0.0135 | 0.99999971 | 0.0135 | 0 | 0 | 0 | 1 | Ref |
| 7480 | MH651178 | Unassigned | Unassigned | Unassigned | 520_32 | Clustered/Singleton | 1 | VC_520_32 | 1 | 0 | 1 | 0 | 0 | 0 | 0 | 1 | Ref |
| 7481 | MH651179 | Unassigned | Unassigned | Unassigned | 514_0 | Clustered | 75 | VC_514_0 | 75 | 0.9402 | 1 | 0.9402 | 0 | 0 | 0 | 1 | Ref |
| 7482 | MH651180 | Unassigned | Unassigned | Unassigned | 325_38 | Clustered | 6 | VC_325_38 | 6 | 0.0267 | 1 | 0.0267 | 0 | 0 | 0 | 0.8 | Ref |
| 7483 | MH651181 | Unassigned | Unassigned | Unassigned | 1344_0 | Clustered | 78 | VC_1344_0 | 78 | 0.9732 | 1 | 0.9732 | 0 | 0 | 0 | 1 | Ref |
| 7484 | MH651182 | Unassigned | Unassigned | Unassigned | 890_0 | Clustered | 3 | VC_890_0 | 3 | 0.3565 | 0.99830951 | 0.3559 | 0 | 0 | 0 | 1 | Ref |
| 7485 | MH651183 | Unassigned | Unassigned | Unassigned | 520_41 | Clustered | 2 | VC_520_41 | 2 | 0.0139 | 0.95761457 | 0.0134 | 0 | 0 | 0 | 1 | Ref |
| 7486 | MH651184 | Unassigned | Unassigned | Unassigned | 371_15 | Clustered | 44 | VC_371_15 | 44 | 0.1391 | 1 | 0.1391 | 0 | 0 | 0 | 0.7474 | Ref |
| 7487 | MH651185 | Unassigned | Unassigned | Unassigned | 557_1 | Clustered | 11 | VC_557_1 | 11 | 0.4945 | 1 | 0.4945 | 0 | 0 | 0 | 0.7818 | Ref |
| 7488 | MH651186 | Unassigned | Unassigned | Unassigned | 371_16 | Clustered | 42 | VC_371_16 | 42 | 0.1341 | 1 | 0.1341 | 0 | 0 | 0 | 0.7282 | Ref |
| 7489 | MH651188 | Unassigned | Unassigned | Unassigned | 520_80 | Clustered | 3 | VC_520_80 | 3 | 0.0295 | 0.99859904 | 0.0295 | 0 | 0 | 0 | 1 | Ref |
| 7490 | MH651189 | Unassigned | Unassigned | Unassigned | 406_0 | Clustered | 11 | VC_406_0 | 11 | 0.9449 | 1 | 0.9449 | 0 | 0 | 0 | 1 | Ref |
| 7491 | MH651190 | Unassigned | Unassigned | Unassigned | 981_1 | Clustered | 26 | VC_981_1 | 26 | 0.7009 | 1 | 0.7009 | 0 | 0 | 0 | 1 | Ref |
| 7492 | MH669000 | Unassigned | Unassigned | Unassigned | 1095_0 | Clustered | 16 | VC_1095_0 | 16 | 0.458 | 1 | 0.458 | 0 | 0 | 0 | 1 | Ref |
| 7493 | MH669001 | Unassigned | Unassigned | Unassigned | 520_21 | Clustered/Singleton | 1 | VC_520_21 | 1 | 0 | 1 | 0 | 0 | 0 | 0 | 1 | Ref |
| 7494 | MH669002 | Unassigned | Unassigned | Unassigned | 551_6 | Clustered | 22 | VC_551_6 | 22 | 0.1214 | 1 | 0.1214 | 0 | 0 | 0 | 0.8095 | Ref |
| 7495 | MH669003 | Unassigned | Unassigned | Unassigned | 520_80 | Clustered | 3 | VC_520_80 | 3 | 0.0295 | 0.99859904 | 0.0295 | 0 | 0 | 0 | 1 | Ref |
| 7496 | MH669004 | Unassigned | Unassigned | Unassigned | 981_1 | Clustered | 26 | VC_981_1 | 26 | 0.7009 | 1 | 0.7009 | 0 | 0 | 0 | 1 | Ref |
| 7497 | MH669006 | Unassigned | Unassigned | Unassigned | 872_0 | Clustered | 31 | VC_872_0 | 31 | 0.8151 | 1 | 0.8151 | 0 | 0 | 0 | 0.8194 | Ref |
| 7498 | MH669007 | Unassigned | Unassigned | Unassigned | 767_0 | Clustered | 34 | VC_767_0 | 34 | 0.9173 | 1 | 0.9173 | 0 | 0 | 0 | 1 | Ref |
| 7499 | MH669008 | Unassigned | Unassigned | Unassigned | 558_50 | Clustered | 15 | VC_558_50 | 15 | 0.0468 | 1 | 0.0468 | 0 | 0 | 0 | 0.9238 | Ref |
| 7500 | MH669011 | Unassigned | Unassigned | Unassigned | 325_44 | Clustered | 3 | VC_325_44 | 3 | 0.0157 | 0.99859171 | 0.0157 | 0 | 0 | 0 | 1 | Ref |
| 7501 | MH669012 | Unassigned | Unassigned | Unassigned | 520_73 | Clustered | 2 | VC_520_73 | 2 | 0.0107 | 0.95762743 | 0.0103 | 0 | 0 | 0 | 1 | Ref |
| 7502 | MH669013 | Unassigned | Unassigned | Unassigned | 767_0 | Clustered | 34 | VC_767_0 | 34 | 0.9173 | 1 | 0.9173 | 0 | 0 | 0 | 1 | Ref |
| 7503 | MH669014 | Unassigned | Unassigned | Unassigned | 520_69 | Clustered/Singleton | 1 | VC_520_69 | 1 | 0 | 1 | 0 | 0 | 0 | 0 | 1 | Ref |
| 7504 | MH669015 | Unassigned | Unassigned | Unassigned | 1178_0 | Clustered | 24 | VC_1178_0 | 24 | 0.9524 | 1 | 0.9524 | 0 | 0 | 0 | 1 | Ref |
| 7505 | MH669016 | Unassigned | Unassigned | Unassigned | 981_1 | Clustered | 26 | VC_981_1 | 26 | 0.7009 | 1 | 0.7009 | 0 | 0 | 0 | 1 | Ref |
| 7506 | MH669017 | Unassigned | Unassigned | Unassigned | 546_4 | Clustered | 16 | VC_546_4 | 16 | 0.2517 | 0.9999998 | 0.2517 | 0 | 0 | 0 | 0.9417 | Ref |
| 7507 | MH669274 | Unassigned | Unassigned | Unassigned | 779_0 | Clustered | 37 | VC_779_0 | 37 | 0.5682 | 1 | 0.5682 | 0 | 0 | 0 | 1 | Ref |
| 7508 | MH674343 | Unassigned | Unassigned | Unassigned | 233_0 | Clustered | 2 | VC_233_0 | 2 | 0.7179 | 0.72985431 | 0.5239 | 0 | 0 | 0 | 1 | Ref |
| 7509 | MH675552 | Unassigned | Unassigned | Unassigned | 1018_1 | Clustered | 10 | VC_1018_1 | 10 | 0.7518 | 1 | 0.7518 | 0 | 0 | 0 | 0.8 | Ref |
| 7510 | MH675927 | Unassigned | Unassigned | Unassigned | 848_0 | Clustered | 28 | VC_848_0 | 28 | 0.7927 | 1 | 0.7927 | 0 | 0 | 0 | 1 | Ref |
| 7511 | MH684921 | Unassigned | Unassigned | Unassigned | 70_0 | Clustered | 27 | VC_70_0 | 27 | 0.5981 | 1 | 0.5981 | 0 | 0 | 0 | 0.9886 | Ref |
| 7512 | MH688040 | Unassigned | Unassigned | Unassigned | 133_22 | Clustered | 13 | VC_133_22 | 13 | 0.0804 | 1 | 0.0804 | 0 | 0 | 0 | 0.8205 | Ref |
| 7513 | MH688453 | Unassigned | Unassigned | Unassigned | 938_0 | Clustered | 99 | VC_938_0 | 99 | 0.9119 | 1 | 0.9119 | 0 | 0 | 0 | 0.9928 | Ref |
| 7514 | MH697575 | Unassigned | Unassigned | Unassigned | 325_52 | Clustered | 5 | VC_325_52 | 5 | 0.0147 | 0.99999073 | 0.0147 | 0 | 0 | 0 | 0.7 | Ref |
| 7515 | MH697576 | Unassigned | Unassigned | Unassigned | 521_0 | Clustered | 37 | VC_521_0 | 37 | 0.912 | 1 | 0.912 | 0 | 0 | 0 | 1 | Ref |
| 7516 | MH697577 | Unassigned | Unassigned | Unassigned | 810_0 | Clustered | 18 | VC_810_0 | 18 | 0.9121 | 1 | 0.9121 | 0 | 0 | 0 | 1 | Ref |
| 7517 | MH697578 | Unassigned | Unassigned | Unassigned | 236_0 | Clustered | 72 | VC_236_0 | 72 | 0.8104 | 1 | 0.8104 | 0 | 0 | 0 | 1 | Ref |
| 7518 | MH697579 | Unassigned | Unassigned | Unassigned | 558_73 | Clustered | 2 | VC_558_73 | 2 | 0.0033 | 0.84374839 | 0.0028 | 0 | 0 | 0 | 1 | Ref |
| 7519 | MH697580 | Unassigned | Unassigned | Unassigned | 406_0 | Clustered | 11 | VC_406_0 | 11 | 0.9449 | 1 | 0.9449 | 0 | 0 | 0 | 1 | Ref |
| 7520 | MH697581 | Unassigned | Unassigned | Unassigned | 325_40 | Clustered | 2 | VC_325_40 | 2 | 0.004 | 0.95574341 | 0.0038 | 0 | 0 | 0 | 1 | Ref |
| 7521 | MH697582 | Unassigned | Unassigned | Unassigned | 546_4 | Clustered | 16 | VC_546_4 | 16 | 0.2517 | 0.9999998 | 0.2517 | 0 | 0 | 0 | 0.9417 | Ref |
| 7522 | MH697583 | Unassigned | Unassigned | Unassigned | 546_4 | Clustered | 16 | VC_546_4 | 16 | 0.2517 | 0.9999998 | 0.2517 | 0 | 0 | 0 | 0.9417 | Ref |
| 7523 | MH697584 | Unassigned | Unassigned | Unassigned | 508_0 | Clustered | 32 | VC_508_0 | 32 | 0.906 | 1 | 0.906 | 0 | 0 | 0 | 1 | Ref |
| 7524 | MH697585 | Unassigned | Unassigned | Unassigned | 521_0 | Clustered | 37 | VC_521_0 | 37 | 0.912 | 1 | 0.912 | 0 | 0 | 0 | 1 | Ref |
| 7525 | MH697586 | Unassigned | Unassigned | Unassigned | 558_20 | Clustered | 17 | VC_558_20 | 17 | 0.054 | 1 | 0.054 | 0 | 0 | 0 | 0.9926 | Ref |
| 7526 | MH697587 | Unassigned | Unassigned | Unassigned | 763_1 | Clustered | 9 | VC_763_1 | 9 | 0.3706 | 1 | 0.3706 | 0 | 0 | 0 | 1 | Ref |
| 7527 | MH697588 | Unassigned | Unassigned | Unassigned | 517_1 | Clustered | 108 | VC_517_1 | 108 | 0.8483 | 1 | 0.8483 | 0 | 0 | 0 | 0.9955 | Ref |
| 7528 | MH697590 | Unassigned | Unassigned | Unassigned | 558_20 | Clustered | 17 | VC_558_20 | 17 | 0.054 | 1 | 0.054 | 0 | 0 | 0 | 0.9926 | Ref |
| 7529 | MH697591 | Unassigned | Unassigned | Unassigned | 216_6 | Clustered | 42 | VC_216_6 | 42 | 0.473 | 1 | 0.473 | 0 | 0 | 0 | 0.8351 | Ref |
| 7530 | MH697592 | Unassigned | Unassigned | Unassigned | 238_0 | Clustered | 17 | VC_238_0 | 17 | 0.6109 | 1 | 0.6109 | 0 | 0 | 0 | 1 | Ref |
| 7531 | MH697593 | Unassigned | Unassigned | Unassigned | 521_0 | Clustered | 37 | VC_521_0 | 37 | 0.912 | 1 | 0.912 | 0 | 0 | 0 | 1 | Ref |
| 7532 | MH707430 | Unassigned | Unassigned | Unassigned | 1358_0 | Clustered | 5 | VC_1358_0 | 5 | 1 | 1 | 1 | 0 | 0 | 0 | 1 | Ref |
| 7533 | MH709120 | Unassigned | Unassigned | Unassigned | 133_17 | Clustered/Singleton | 1 | VC_133_17 | 1 | 0 | 1 | 0 | 0 | 0 | 0 | 1 | Ref |
| 7534 | MH709121 | Unassigned | Unassigned | Unassigned | 133_10 | Clustered | 10 | VC_133_10 | 10 | 0.0527 | 1 | 0.0527 | 0 | 0 | 0 | 1 | Ref |
| 7535 | MH713599 | Unassigned | Unassigned | Unassigned | 492_1 | Clustered | 20 | VC_492_1 | 20 | 0.4176 | 1 | 0.4176 | 0 | 0 | 0 | 1 | Ref |
| 7536 | MH717096 | Unassigned | Unassigned | Unassigned | 30_1 | Clustered | 16 | VC_30_1 | 16 | 0.1973 | 1 | 0.1973 | 0 | 0 | 0 | 0.8333 | Ref |
| 7537 | MH717097 | Unassigned | Unassigned | Unassigned | 461_0 | Clustered | 96 | VC_461_0 | 96 | 0.9307 | 1 | 0.9307 | 0 | 0 | 0 | 0.9575 | Ref |
| 7538 | MH717098 | Unassigned | Unassigned | Unassigned | 392_0 | Clustered | 126 | VC_392_0 | 126 | 0.9621 | 1 | 0.9621 | 0 | 0 | 0 | 1 | Ref |
| 7539 | MH717099 | Unassigned | Unassigned | Unassigned | 392_0 | Clustered | 126 | VC_392_0 | 126 | 0.9621 | 1 | 0.9621 | 0 | 0 | 0 | 1 | Ref |
| 7540 | MH717709 | Unassigned | Unassigned | Unassigned | 385_0 | Clustered | 51 | VC_385_0 | 51 | 0.8754 | 1 | 0.8754 | 0 | 0 | 0 | 1 | Ref |
| 7541 | MH719189 | Unassigned | Unassigned | Unassigned | 251_0 | Clustered | 22 | VC_251_0 | 22 | 0.6508 | 1 | 0.6508 | 0 | 0 | 0 | 1 | Ref |
| 7542 | MH719190 | Unassigned | Unassigned | Unassigned | 251_0 | Clustered | 22 | VC_251_0 | 22 | 0.6508 | 1 | 0.6508 | 0 | 0 | 0 | 1 | Ref |
| 7543 | MH719191 | Unassigned | Unassigned | Unassigned | 251_0 | Clustered | 22 | VC_251_0 | 22 | 0.6508 | 1 | 0.6508 | 0 | 0 | 0 | 1 | Ref |
| 7544 | MH719192 | Unassigned | Unassigned | Unassigned | 251_0 | Clustered | 22 | VC_251_0 | 22 | 0.6508 | 1 | 0.6508 | 0 | 0 | 0 | 1 | Ref |
| 7545 | MH719193 | Unassigned | Unassigned | Unassigned | 251_0 | Clustered | 22 | VC_251_0 | 22 | 0.6508 | 1 | 0.6508 | 0 | 0 | 0 | 1 | Ref |
| 7546 | MH719194 | Unassigned | Unassigned | Unassigned | 251_0 | Clustered | 22 | VC_251_0 | 22 | 0.6508 | 1 | 0.6508 | 0 | 0 | 0 | 1 | Ref |
| 7547 | MH719195 | Unassigned | Unassigned | Unassigned | 251_0 | Clustered | 22 | VC_251_0 | 22 | 0.6508 | 1 | 0.6508 | 0 | 0 | 0 | 1 | Ref |
| 7548 | MH725810 | Unassigned | Unassigned | Unassigned | 76_0 | Clustered | 37 | VC_76_0 | 37 | 0.9403 | 1 | 0.9403 | 0 | 0 | 0 | 1 | Ref |
| 7549 | MH727543 | Unassigned | Unassigned | Unassigned | 558_2 | Clustered | 2 | VC_558_2 | 2 | 0.0032 | 0.66859979 | 0.0021 | 0 | 0 | 0 | 1 | Ref |
| 7550 | MH727544 | Unassigned | Unassigned | Unassigned | 236_0 | Clustered | 72 | VC_236_0 | 72 | 0.8104 | 1 | 0.8104 | 0 | 0 | 0 | 1 | Ref |
| 7551 | MH727546 | Unassigned | Unassigned | Unassigned | 508_0 | Clustered | 32 | VC_508_0 | 32 | 0.906 | 1 | 0.906 | 0 | 0 | 0 | 1 | Ref |
| 7552 | MH727547 | Unassigned | Unassigned | Unassigned | 767_0 | Clustered | 34 | VC_767_0 | 34 | 0.9173 | 1 | 0.9173 | 0 | 0 | 0 | 1 | Ref |
| 7553 | MH727548 | Unassigned | Unassigned | Unassigned | 520_5 | Clustered/Singleton | 1 | VC_520_5 | 1 | 0 | 1 | 0 | 0 | 0 | 0 | 1 | Ref |
| 7554 | MH727549 | Unassigned | Unassigned | Unassigned | 810_0 | Clustered | 18 | VC_810_0 | 18 | 0.9121 | 1 | 0.9121 | 0 | 0 | 0 | 1 | Ref |
| 7555 | MH727551 | Unassigned | Unassigned | Unassigned | 546_4 | Clustered | 16 | VC_546_4 | 16 | 0.2517 | 0.9999998 | 0.2517 | 0 | 0 | 0 | 0.9417 | Ref |
| 7556 | MH727552 | Unassigned | Unassigned | Unassigned | 767_0 | Clustered | 34 | VC_767_0 | 34 | 0.9173 | 1 | 0.9173 | 0 | 0 | 0 | 1 | Ref |
| 7557 | MH727553 | Unassigned | Unassigned | Unassigned | 939_0 | Clustered | 15 | VC_939_0 | 15 | 1 | 0 | 0 | 0 | 0 | 0 | 1 | Ref |
| 7558 | MH727554 | Unassigned | Unassigned | Unassigned | 514_0 | Clustered | 75 | VC_514_0 | 75 | 0.9402 | 1 | 0.9402 | 0 | 0 | 0 | 1 | Ref |
| 7559 | MH727555 | Unassigned | Unassigned | Unassigned | 371_16 | Clustered | 42 | VC_371_16 | 42 | 0.1341 | 1 | 0.1341 | 0 | 0 | 0 | 0.7282 | Ref |
| 7560 | MH727557 | Unassigned | Unassigned | Unassigned | 551_6 | Clustered | 22 | VC_551_6 | 22 | 0.1214 | 1 | 0.1214 | 0 | 0 | 0 | 0.8095 | Ref |
| 7561 | MH727558 | Unassigned | Unassigned | Unassigned | 558_56 | Clustered/Singleton | 1 | VC_558_56 | 1 | 0 | 1 | 0 | 0 | 0 | 0 | 1 | Ref |
| 7562 | MH727559 | Unassigned | Unassigned | Unassigned | 889_0 | Clustered | 16 | VC_889_0 | 16 | 0.2969 | 1 | 0.2969 | 0 | 0 | 0 | 1 | Ref |
| 7563 | MH727560 | Unassigned | Unassigned | Unassigned | 1199_0 | Clustered | 8 | VC_1199_0 | 8 | 0.7928 | 1 | 0.7928 | 0 | 0 | 0 | 1 | Ref |
| 7564 | MH727561 | Unassigned | Unassigned | Unassigned | 372_1 | Clustered | 9 | VC_372_1 | 9 | 0.194 | 1 | 0.194 | 0 | 0 | 0 | 1 | Ref |
| 7565 | MH727562 | Unassigned | Unassigned | Unassigned | 939_0 | Clustered | 15 | VC_939_0 | 15 | 1 | 0 | 0 | 0 | 0 | 0 | 1 | Ref |
| 7566 | MH727563 | Unassigned | Unassigned | Unassigned | 517_1 | Clustered | 108 | VC_517_1 | 108 | 0.8483 | 1 | 0.8483 | 0 | 0 | 0 | 0.9955 | Ref |
| 7567 | MH727564 | Unassigned | Unassigned | Unassigned | 1071_0 | Clustered | 20 | VC_1071_0 | 20 | 0.3268 | 1 | 0.3268 | 0 | 0 | 0 | 0.9842 | Ref |
| 7568 | MH727593 | Unassigned | Unassigned | Unassigned | 184_0 | Clustered | 2 | VC_184_0 | 2 | 0.7964 | 0.93668477 | 0.746 | 0 | 0 | 0 | 1 | Ref |
| 7569 | MH729818 | Unassigned | Unassigned | Unassigned | 452_0 | Clustered | 66 | VC_452_0 | 66 | 0.9248 | 1 | 0.9248 | 0 | 0 | 0 | 1 | Ref |
| 7570 | MH729819 | Unassigned | Unassigned | Unassigned | 584_0 | Clustered | 28 | VC_584_0 | 28 | 0.7704 | 1 | 0.7704 | 0 | 0 | 0 | 1 | Ref |
| 7571 | MH729874 | Unassigned | Unassigned | Unassigned | 491_0 | Clustered | 36 | VC_491_0 | 36 | 0.5336 | 1 | 0.5336 | 0 | 0 | 0 | 0.9714 | Ref |
| 7572 | MH730557 | Unassigned | Unassigned | Unassigned | 1359_0 | Clustered | 11 | VC_1359_0 | 11 | 1 | 0 | 0 | 0 | 0 | 0 | 1 | Ref |
| 7573 | MH733496 | Unassigned | Unassigned | Unassigned | 779_0 | Clustered | 37 | VC_779_0 | 37 | 0.5682 | 1 | 0.5682 | 0 | 0 | 0 | 1 | Ref |
| 7574 | MH744414 | Unassigned | Unassigned | Unassigned | 325_10 | Clustered | 3 | VC_325_10 | 3 | 0.012 | 0.99859169 | 0.012 | 0 | 0 | 0 | 1 | Ref |
| 7575 | MH744415 | Unassigned | Unassigned | Unassigned | 520_36 | Clustered/Singleton | 1 | VC_520_36 | 1 | 0 | 1 | 0 | 0 | 0 | 0 | 1 | Ref |
| 7576 | MH744416 | Unassigned | Unassigned | Unassigned | 1344_0 | Clustered | 78 | VC_1344_0 | 78 | 0.9732 | 1 | 0.9732 | 0 | 0 | 0 | 1 | Ref |
| 7577 | MH744417 | Unassigned | Unassigned | Unassigned | 371_16 | Clustered | 42 | VC_371_16 | 42 | 0.1341 | 1 | 0.1341 | 0 | 0 | 0 | 0.7282 | Ref |
| 7578 | MH744418 | Unassigned | Unassigned | Unassigned | 1158_1 | Clustered | 2 | VC_1158_1 | 2 | 0.1372 | 0.89820314 | 0.1233 | 0 | 0 | 0 | 1 | Ref |
| 7579 | MH744419 | Unassigned | Unassigned | Unassigned | 1074_0 | Clustered | 19 | VC_1074_0 | 19 | 0.9198 | 1 | 0.9198 | 0 | 0 | 0 | 1 | Ref |
| 7580 | MH744420 | Unassigned | Unassigned | Unassigned | 588_0 | Clustered | 2 | VC_588_0 | 2 | 0.0832 | 0.94395095 | 0.0785 | 0 | 0 | 0 | 1 | Ref |
| 7581 | MH744421 | Unassigned | Unassigned | Unassigned | 1160_0 | Clustered | 73 | VC_1160_0 | 73 | 0.9835 | 1 | 0.9835 | 0 | 0 | 0 | 1 | Ref |
| 7582 | MH744422 | Unassigned | Unassigned | Unassigned | 558_52 | Clustered/Singleton | 1 | VC_558_52 | 1 | 0 | 1 | 0 | 0 | 0 | 0 | 1 | Ref |
| 7583 | MH744423 | Unassigned | Unassigned | Unassigned | 372_2 | Clustered | 5 | VC_372_2 | 5 | 0.072 | 0.99999997 | 0.072 | 0 | 0 | 0 | 0.9 | Ref |
| 7584 | MH744424 | Unassigned | Unassigned | Unassigned | 767_0 | Clustered | 34 | VC_767_0 | 34 | 0.9173 | 1 | 0.9173 | 0 | 0 | 0 | 1 | Ref |
| 7585 | MH744425 | Unassigned | Unassigned | Unassigned | 887_0 | Clustered | 18 | VC_887_0 | 18 | 0.3663 | 1 | 0.3663 | 0 | 0 | 0 | 1 | Ref |
| 7586 | MH746814 | Unassigned | Unassigned | Unassigned | 700_0 | Clustered | 10 | VC_700_0 | 10 | 0.8076 | 1 | 0.8076 | 0 | 0 | 0 | 1 | Ref |
| 7587 | MH746817 | Unassigned | Unassigned | Unassigned | 171_0 | Clustered | 8 | VC_171_0 | 8 | 0.5884 | 1 | 0.5884 | 0 | 0 | 0 | 1 | Ref |
| 7588 | MH751506 | Unassigned | Unassigned | Unassigned | 283_92 | Clustered | 4 | VC_283_92 | 4 | 0.0079 | 0.9999582 | 0.0079 | 0 | 0 | 0 | 1 | Ref |
| 7589 | MH752385 | Unassigned | Unassigned | Unassigned | 104_0 | Clustered | 6 | VC_104_0 | 6 | 0.8291 | 1 | 0.8291 | 0 | 0 | 0 | 1 | Ref |
| 7590 | MH752840 | Unassigned | Unassigned | Unassigned | 772_11 | Clustered | 6 | VC_772_11 | 6 | 0.0305 | 0.84253803 | 0.0257 | 0 | 0 | 0 | 0.8 | Ref |
| 7591 | MH763831 | Unassigned | Unassigned | Unassigned | 699_0 | Clustered | 71 | VC_699_0 | 71 | 0.9352 | 1 | 0.9352 | 0 | 0 | 0 | 1 | Ref |
| 7592 | MH779498 | Unassigned | Unassigned | Unassigned | 558_49 | Clustered | 7 | VC_558_49 | 7 | 0.0194 | 0.99510369 | 0.0193 | 0 | 0 | 0 | 0.8571 | Ref |
| 7593 | MH779499 | Unassigned | Unassigned | Unassigned | 767_0 | Clustered | 34 | VC_767_0 | 34 | 0.9173 | 1 | 0.9173 | 0 | 0 | 0 | 1 | Ref |
| 7594 | MH779500 | Unassigned | Unassigned | Unassigned | 371_3 | Clustered | 22 | VC_371_3 | 22 | 0.0787 | 1 | 0.0787 | 0 | 0 | 0 | 0.9048 | Ref |
| 7595 | MH779501 | Unassigned | Unassigned | Unassigned | 767_0 | Clustered | 34 | VC_767_0 | 34 | 0.9173 | 1 | 0.9173 | 0 | 0 | 0 | 1 | Ref |
| 7596 | MH779502 | Unassigned | Unassigned | Unassigned | 558_54 | Clustered/Singleton | 1 | VC_558_54 | 1 | 0 | 1 | 0 | 0 | 0 | 0 | 1 | Ref |
| 7597 | MH779503 | Unassigned | Unassigned | Unassigned | 551_6 | Clustered | 22 | VC_551_6 | 22 | 0.1214 | 1 | 0.1214 | 0 | 0 | 0 | 0.8095 | Ref |
| 7598 | MH779504 | Unassigned | Unassigned | Unassigned | 557_1 | Clustered | 11 | VC_557_1 | 11 | 0.4945 | 1 | 0.4945 | 0 | 0 | 0 | 0.7818 | Ref |
| 7599 | MH779505 | Unassigned | Unassigned | Unassigned | 761_0 | Clustered | 67 | VC_761_0 | 67 | 0.8947 | 1 | 0.8947 | 0 | 0 | 0 | 1 | Ref |
| 7600 | MH779506 | Unassigned | Unassigned | Unassigned | 950_0 | Clustered | 38 | VC_950_0 | 38 | 0.9443 | 1 | 0.9443 | 0 | 0 | 0 | 1 | Ref |
| 7601 | MH779507 | Unassigned | Unassigned | Unassigned | 761_0 | Clustered | 67 | VC_761_0 | 67 | 0.8947 | 1 | 0.8947 | 0 | 0 | 0 | 1 | Ref |
| 7602 | MH779508 | Unassigned | Unassigned | Unassigned | 767_0 | Clustered | 34 | VC_767_0 | 34 | 0.9173 | 1 | 0.9173 | 0 | 0 | 0 | 1 | Ref |
| 7603 | MH779509 | Unassigned | Unassigned | Unassigned | 761_0 | Clustered | 67 | VC_761_0 | 67 | 0.8947 | 1 | 0.8947 | 0 | 0 | 0 | 1 | Ref |
| 7604 | MH779510 | Unassigned | Unassigned | Unassigned | 767_0 | Clustered | 34 | VC_767_0 | 34 | 0.9173 | 1 | 0.9173 | 0 | 0 | 0 | 1 | Ref |
| 7605 | MH779511 | Unassigned | Unassigned | Unassigned | 888_0 | Clustered | 28 | VC_888_0 | 28 | 0.5497 | 1 | 0.5497 | 0 | 0 | 0 | 1 | Ref |
| 7606 | MH779512 | Unassigned | Unassigned | Unassigned | 1344_0 | Clustered | 78 | VC_1344_0 | 78 | 0.9732 | 1 | 0.9732 | 0 | 0 | 0 | 1 | Ref |
| 7607 | MH779513 | Unassigned | Unassigned | Unassigned | 761_0 | Clustered | 67 | VC_761_0 | 67 | 0.8947 | 1 | 0.8947 | 0 | 0 | 0 | 1 | Ref |
| 7608 | MH779514 | Unassigned | Unassigned | Unassigned | 761_0 | Clustered | 67 | VC_761_0 | 67 | 0.8947 | 1 | 0.8947 | 0 | 0 | 0 | 1 | Ref |
| 7609 | MH779515 | Unassigned | Unassigned | Unassigned | 761_0 | Clustered | 67 | VC_761_0 | 67 | 0.8947 | 1 | 0.8947 | 0 | 0 | 0 | 1 | Ref |
| 7610 | MH779516 | Unassigned | Unassigned | Unassigned | 371_16 | Clustered | 42 | VC_371_16 | 42 | 0.1341 | 1 | 0.1341 | 0 | 0 | 0 | 0.7282 | Ref |
| 7611 | MH779517 | Unassigned | Unassigned | Unassigned | 223_0 | Clustered | 38 | VC_223_0 | 38 | 0.9095 | 1 | 0.9095 | 0 | 0 | 0 | 1 | Ref |
| 7612 | MH779518 | Unassigned | Unassigned | Unassigned | 690_0 | Clustered | 50 | VC_690_0 | 50 | 0.9805 | 1 | 0.9805 | 0 | 0 | 0 | 1 | Ref |

| | | | | | | | | | | | | | | | | | |
|---|---|---|---|---|---|---|---|---|---|---|---|---|---|---|---|---|---|
| 7613 | MH779519 | Unassigned | Unassigned | Unassigned | 690_0 | Clustered | 50 | VC_690_0 | 50 | 0.9805 | 1 | 0.9805 | 0 | 0 | 0 | 1 | Ref |
| 7614 | MH779520 | Unassigned | Unassigned | Unassigned | 690_0 | Clustered | 50 | VC_690_0 | 50 | 0.9805 | 1 | 0.9805 | 0 | 0 | 0 | 1 | Ref |
| 7615 | MH779521 | Unassigned | Unassigned | Unassigned | 690_0 | Clustered | 50 | VC_690_0 | 50 | 0.9805 | 1 | 0.9805 | 0 | 0 | 0 | 1 | Ref |
| 7616 | MH779522 | Unassigned | Unassigned | Unassigned | 690_0 | Clustered | 50 | VC_690_0 | 50 | 0.9805 | 1 | 0.9805 | 0 | 0 | 0 | 1 | Ref |
| 7617 | MH779523 | Unassigned | Unassigned | Unassigned | 690_0 | Clustered | 50 | VC_690_0 | 50 | 0.9805 | 1 | 0.9805 | 0 | 0 | 0 | 1 | Ref |
| 7618 | MH779524 | Unassigned | Unassigned | Unassigned | 690_0 | Clustered | 50 | VC_690_0 | 50 | 0.9805 | 1 | 0.9805 | 0 | 0 | 0 | 1 | Ref |
| 7619 | MH779525 | Unassigned | Unassigned | Unassigned | 690_0 | Clustered | 50 | VC_690_0 | 50 | 0.9805 | 1 | 0.9805 | 0 | 0 | 0 | 1 | Ref |
| 7620 | MH779526 | Unassigned | Unassigned | Unassigned | 690_0 | Clustered | 50 | VC_690_0 | 50 | 0.9805 | 1 | 0.9805 | 0 | 0 | 0 | 1 | Ref |
| 7621 | MH779527 | Unassigned | Unassigned | Unassigned | 690_0 | Clustered | 50 | VC_690_0 | 50 | 0.9805 | 1 | 0.9805 | 0 | 0 | 0 | 1 | Ref |
| 7622 | MH779619 | Unassigned | Unassigned | Unassigned | 392_0 | Clustered | 126 | VC_392_0 | 126 | 0.9621 | 1 | 0.9621 | 0 | 0 | 0 | 1 | Ref |
| 7623 | MH791395 | Unassigned | Unassigned | Unassigned | 592_0 | Clustered | 50 | VC_592_0 | 50 | 0.7314 | 1 | 0.7314 | 0 | 0 | 0 | 1 | Ref |
| 7624 | MH791396 | Unassigned | Unassigned | Unassigned | 592_0 | Clustered | 50 | VC_592_0 | 50 | 0.7314 | 1 | 0.7314 | 0 | 0 | 0 | 1 | Ref |
| 7625 | MH791397 | Unassigned | Unassigned | Unassigned | 92_0 | Clustered | 22 | VC_92_0 | 22 | 0.5639 | 1 | 0.5639 | 0 | 0 | 0 | 1 | Ref |
| 7626 | MH791398 | Unassigned | Unassigned | Unassigned | 443_2 | Clustered | 10 | VC_443_2 | 10 | 0.3344 | 1 | 0.3344 | 0 | 0 | 0 | 0.9333 | Ref |
| 7627 | MH791399 | Unassigned | Unassigned | Unassigned | 76_0 | Clustered | 37 | VC_76_0 | 37 | 0.9403 | 1 | 0.9403 | 0 | 0 | 0 | 1 | Ref |
| 7628 | MH791400 | Unassigned | Unassigned | Unassigned | 330_0 | Clustered | 27 | VC_330_0 | 27 | 0.5088 | 1 | 0.5088 | 0 | 0 | 0 | 0.8661 | Ref |
| 7629 | MH791402 | Unassigned | Unassigned | Unassigned | 461_0 | Clustered | 96 | VC_461_0 | 96 | 0.9307 | 1 | 0.9307 | 0 | 0 | 0 | 0.9575 | Ref |
| 7630 | MH791403 | Unassigned | Unassigned | Unassigned | 166_0 | Clustered | 26 | VC_166_0 | 26 | 0.7923 | 1 | 0.7923 | 0 | 0 | 0 | 1 | Ref |
| 7631 | MH791404 | Unassigned | Unassigned | Unassigned | 443_2 | Clustered | 10 | VC_443_2 | 10 | 0.3344 | 1 | 0.3344 | 0 | 0 | 0 | 0.9333 | Ref |
| 7632 | MH791405 | Unassigned | Unassigned | Unassigned | 133_6 | Clustered/Singleton | 1 | VC_133_6 | 1 | 0 | 1 | 0 | 0 | 0 | 0 | 1 | Ref |
| 7633 | MH791406 | Unassigned | Unassigned | Unassigned | 166_0 | Clustered | 26 | VC_166_0 | 26 | 0.7923 | 1 | 0.7923 | 0 | 0 | 0 | 1 | Ref |
| 7634 | MH791408 | Unassigned | Unassigned | Unassigned | 76_0 | Clustered | 37 | VC_76_0 | 37 | 0.9403 | 1 | 0.9403 | 0 | 0 | 0 | 1 | Ref |
| 7635 | MH791409 | Unassigned | Unassigned | Unassigned | 290_6 | Clustered | 12 | VC_290_6 | 12 | 0.1149 | 1 | 0.1149 | 0 | 0 | 0 | 0.8182 | Ref |
| 7636 | MH791410 | Unassigned | Unassigned | Unassigned | 461_0 | Clustered | 96 | VC_461_0 | 96 | 0.9307 | 1 | 0.9307 | 0 | 0 | 0 | 0.9575 | Ref |
| 7637 | MH791411 | Unassigned | Unassigned | Unassigned | 164_11 | Clustered | 10 | VC_164_11 | 10 | 0.1507 | 1 | 0.1507 | 0 | 0 | 0 | 0.9111 | Ref |
| 7638 | MH791412 | Unassigned | Unassigned | Unassigned | 461_0 | Clustered | 96 | VC_461_0 | 96 | 0.9307 | 1 | 0.9307 | 0 | 0 | 0 | 0.9575 | Ref |
| 7639 | MH791413 | Unassigned | Unassigned | Unassigned | 330_0 | Clustered | 27 | VC_330_0 | 27 | 0.5088 | 1 | 0.5088 | 0 | 0 | 0 | 0.8661 | Ref |
| 7640 | MH791414 | Unassigned | Unassigned | Unassigned | 443_0 | Clustered | 2 | VC_443_0 | 2 | 0.029 | 0.87409603 | 0.0253 | 0 | 0 | 0 | 1 | Ref |
| 7641 | MH791415 | Unassigned | Unassigned | Unassigned | 108_0 | Clustered | 33 | VC_108_0 | 33 | 0.949 | 1 | 0.949 | 0 | 0 | 0 | 1 | Ref |
| 7642 | MH791416 | Unassigned | Unassigned | Unassigned | 76_0 | Clustered | 37 | VC_76_0 | 37 | 0.9403 | 1 | 0.9403 | 0 | 0 | 0 | 1 | Ref |
| 7643 | MH800198 | Unassigned | Unassigned | Unassigned | 700_0 | Clustered | 10 | VC_700_0 | 10 | 0.8076 | 1 | 0.8076 | 0 | 0 | 0 | 1 | Ref |
| 7644 | MH800199 | Unassigned | Unassigned | Unassigned | 700_0 | Clustered | 10 | VC_700_0 | 10 | 0.8076 | 1 | 0.8076 | 0 | 0 | 0 | 1 | Ref |
| 7645 | MH800200 | Unassigned | Unassigned | Unassigned | 699_0 | Clustered | 71 | VC_699_0 | 71 | 0.9352 | 1 | 0.9352 | 0 | 0 | 0 | 1 | Ref |
| 7646 | MH807810 | Unassigned | Unassigned | Unassigned | 458_0 | Clustered | 5 | VC_458_0 | 5 | 0.1938 | 0.99999997 | 0.1938 | 0 | 0 | 0 | 1 | Ref |
| 7647 | MH807811 | Unassigned | Unassigned | Unassigned | 984_0 | Clustered | 38 | VC_984_0 | 38 | 0.9758 | 1 | 0.9758 | 0 | 0 | 0 | 1 | Ref |
| 7648 | MH807812 | Unassigned | Unassigned | Unassigned | 131_0 | Clustered | 17 | VC_131_0 | 17 | 0.2671 | 1 | 0.2671 | 0 | 0 | 0 | 1 | Ref |
| 7649 | MH807813 | Unassigned | Unassigned | Unassigned | 458_0 | Clustered | 5 | VC_458_0 | 5 | 0.1938 | 0.99999997 | 0.1938 | 0 | 0 | 0 | 1 | Ref |
| 7650 | MH807814 | Unassigned | Unassigned | Unassigned | 984_0 | Clustered | 38 | VC_984_0 | 38 | 0.9758 | 1 | 0.9758 | 0 | 0 | 0 | 1 | Ref |
| 7651 | MH807815 | Unassigned | Unassigned | Unassigned | 458_0 | Clustered | 5 | VC_458_0 | 5 | 0.1938 | 0.99999997 | 0.1938 | 0 | 0 | 0 | 1 | Ref |
| 7652 | MH807816 | Unassigned | Unassigned | Unassigned | 984_0 | Clustered | 38 | VC_984_0 | 38 | 0.9758 | 1 | 0.9758 | 0 | 0 | 0 | 1 | Ref |
| 7653 | MH807817 | Unassigned | Unassigned | Unassigned | 458_0 | Clustered | 5 | VC_458_0 | 5 | 0.1938 | 0.99999997 | 0.1938 | 0 | 0 | 0 | 1 | Ref |
| 7654 | MH807818 | Unassigned | Unassigned | Unassigned | 984_0 | Clustered | 38 | VC_984_0 | 38 | 0.9758 | 1 | 0.9758 | 0 | 0 | 0 | 1 | Ref |
| 7655 | MH807819 | Unassigned | Unassigned | Unassigned | 984_0 | Clustered | 38 | VC_984_0 | 38 | 0.9758 | 1 | 0.9758 | 0 | 0 | 0 | 1 | Ref |
| 7656 | MH807820 | Unassigned | Unassigned | Unassigned | 131_0 | Clustered | 17 | VC_131_0 | 17 | 0.2671 | 1 | 0.2671 | 0 | 0 | 0 | 1 | Ref |
| 7657 | MH809528 | Unassigned | Unassigned | Unassigned | 577_0 | Clustered | 6 | VC_577_0 | 6 | 0.9267 | 0.99999993 | 0.9267 | 0 | 0 | 0 | 1 | Ref |
| 7658 | MH809529 | Unassigned | Unassigned | Unassigned | 526_0 | Clustered | 6 | VC_526_0 | 6 | 0.4386 | 1 | 0.4386 | 0 | 0 | 0 | 1 | Ref |
| 7659 | MH809530 | Unassigned | Unassigned | Unassigned | 526_0 | Clustered | 6 | VC_526_0 | 6 | 0.4386 | 1 | 0.4386 | 0 | 0 | 0 | 1 | Ref |
| 7660 | MH809531 | Unassigned | Unassigned | Unassigned | 526_0 | Clustered | 6 | VC_526_0 | 6 | 0.4386 | 1 | 0.4386 | 0 | 0 | 0 | 1 | Ref |
| 7661 | MH809534 | Unassigned | Unassigned | Unassigned | 386_0 | Clustered | 49 | VC_386_0 | 49 | 0.7941 | 1 | 0.7941 | 0 | 0 | 0 | 1 | Ref |
| 7662 | MH809535 | Unassigned | Unassigned | Unassigned | 283_34 | Clustered | 4 | VC_283_34 | 4 | 0.0082 | 0.9997481 | 0.0082 | 0 | 0 | 0 | 1 | Ref |
| 7663 | MH816848 | Unassigned | Unassigned | Unassigned | 848_0 | Clustered | 28 | VC_848_0 | 28 | 0.7927 | 1 | 0.7927 | 0 | 0 | 0 | 1 | Ref |
| 7664 | MH816966 | Unassigned | Unassigned | Unassigned | 618_0 | Clustered | 13 | VC_618_0 | 13 | 0.6252 | 1 | 0.6252 | 0 | 0 | 0 | 1 | Ref |
| 7665 | MH817022 | Unassigned | Unassigned | Unassigned | 818_0 | Clustered | 20 | VC_818_0 | 20 | 0.9601 | 1 | 0.9601 | 0 | 0 | 0 | 1 | Ref |
| 7666 | MH823906 | Unassigned | Unassigned | Unassigned | 669_0 | Clustered | 21 | VC_669_0 | 21 | 0.3623 | 1 | 0.3623 | 0 | 0 | 0 | 1 | Ref |
| 7667 | MH825697 | Unassigned | Unassigned | Unassigned | 558_12 | Clustered | 3 | VC_558_12 | 3 | 0.0064 | 0.87854253 | 0.0057 | 0 | 0 | 0 | 1 | Ref |
| 7668 | MH825698 | Unassigned | Unassigned | Unassigned | 1361_0 | Clustered | 6 | VC_1361_0 | 6 | 0.2371 | 1 | 0.2371 | 0 | 0 | 0 | 1 | Ref |
| 7669 | MH825699 | Unassigned | Unassigned | Unassigned | 727_0 | Clustered | 11 | VC_727_0 | 11 | 0.5054 | 1 | 0.5054 | 0 | 0 | 0 | 1 | Ref |
| 7670 | MH825700 | Unassigned | Unassigned | Unassigned | 1344_0 | Clustered | 78 | VC_1344_0 | 78 | 0.9732 | 1 | 0.9732 | 0 | 0 | 0 | 1 | Ref |
| 7671 | MH825701 | Unassigned | Unassigned | Unassigned | 216_4 | Clustered | 13 | VC_216_4 | 13 | 0.1657 | 1 | 0.1657 | 0 | 0 | 0 | 1 | Ref |
| 7672 | MH825702 | Unassigned | Unassigned | Unassigned | 371_15 | Clustered | 44 | VC_371_15 | 44 | 0.1391 | 1 | 0.1391 | 0 | 0 | 0 | 0.7474 | Ref |
| 7673 | MH825703 | Unassigned | Unassigned | Unassigned | 1073_0 | Clustered | 24 | VC_1073_0 | 24 | 0.8388 | 1 | 0.8388 | 0 | 0 | 0 | 1 | Ref |
| 7674 | MH825704 | Unassigned | Unassigned | Unassigned | 216_6 | Clustered | 42 | VC_216_6 | 42 | 0.473 | 1 | 0.473 | 0 | 0 | 0 | 0.8351 | Ref |
| 7675 | MH825705 | Unassigned | Unassigned | Unassigned | 371_15 | Clustered | 44 | VC_371_15 | 44 | 0.1391 | 1 | 0.1391 | 0 | 0 | 0 | 0.7474 | Ref |
| 7676 | MH825706 | Unassigned | Unassigned | Unassigned | 1353_0 | Clustered | 10 | VC_1353_0 | 10 | 1 | 1 | 1 | 0 | 0 | 0 | 1 | Ref |
| 7677 | MH825707 | Unassigned | Unassigned | Unassigned | 520_30 | Clustered | 2 | VC_520_30 | 2 | 0.0073 | 0.9544492 | 0.0069 | 0 | 0 | 0 | 1 | Ref |
| 7678 | MH825709 | Unassigned | Unassigned | Unassigned | 216_3 | Clustered | 8 | VC_216_3 | 8 | 0.0701 | 1 | 0.0701 | 0 | 0 | 0 | 0.8571 | Ref |
| 7679 | MH825710 | Unassigned | Unassigned | Unassigned | 517_1 | Clustered | 108 | VC_517_1 | 108 | 0.8483 | 1 | 0.8483 | 0 | 0 | 0 | 0.9955 | Ref |
| 7680 | MH825711 | Unassigned | Unassigned | Unassigned | 1164_0 | Clustered | 25 | VC_1164_0 | 25 | 0.7312 | 1 | 0.7312 | 0 | 0 | 0 | 1 | Ref |
| 7681 | MH825713 | Unassigned | Unassigned | Unassigned | 558_50 | Clustered | 15 | VC_558_50 | 15 | 0.0468 | 1 | 0.0468 | 0 | 0 | 0 | 0.9238 | Ref |
| 7682 | MH834594 | Unassigned | Unassigned | Unassigned | 951_0 | Clustered | 3 | VC_951_0 | 3 | 0.1049 | 0.99921864 | 0.1048 | 0 | 0 | 0 | 1 | Ref |
| 7683 | MH834595 | Unassigned | Unassigned | Unassigned | 1166_0 | Clustered | 20 | VC_1166_0 | 20 | 0.99 | 1 | 0.99 | 0 | 0 | 0 | 1 | Ref |
| 7684 | MH834596 | Unassigned | Unassigned | Unassigned | 1234_0 | Clustered | 44 | VC_1234_0 | 44 | 1 | 1 | 1 | 0 | 0 | 0 | 1 | Ref |
| 7685 | MH834597 | Unassigned | Unassigned | Unassigned | 951_0 | Clustered | 3 | VC_951_0 | 3 | 0.1049 | 0.99921864 | 0.1048 | 0 | 0 | 0 | 1 | Ref |
| 7686 | MH834598 | Unassigned | Unassigned | Unassigned | 1036_0 | Clustered | 11 | VC_1036_0 | 11 | 0.656 | 1 | 0.656 | 0 | 0 | 0 | 0.8727 | Ref |
| 7687 | MH834599 | Unassigned | Unassigned | Unassigned | 1234_0 | Clustered | 44 | VC_1234_0 | 44 | 1 | 1 | 1 | 0 | 0 | 0 | 1 | Ref |
| 7688 | MH834600 | Unassigned | Unassigned | Unassigned | 888_0 | Clustered | 28 | VC_888_0 | 28 | 0.5497 | 1 | 0.5497 | 0 | 0 | 0 | 1 | Ref |
| 7689 | MH834602 | Unassigned | Unassigned | Unassigned | 1234_0 | Clustered | 44 | VC_1234_0 | 44 | 1 | 1 | 1 | 0 | 0 | 0 | 1 | Ref |
| 7690 | MH834603 | Unassigned | Unassigned | Unassigned | 1035_0 | Clustered | 6 | VC_1035_0 | 6 | 0.4854 | 1 | 0.4854 | 0 | 0 | 0 | 1 | Ref |
| 7691 | MH834604 | Unassigned | Unassigned | Unassigned | 1234_0 | Clustered | 44 | VC_1234_0 | 44 | 1 | 1 | 1 | 0 | 0 | 0 | 1 | Ref |
| 7692 | MH834605 | Unassigned | Unassigned | Unassigned | 1035_0 | Clustered | 6 | VC_1035_0 | 6 | 0.4854 | 1 | 0.4854 | 0 | 0 | 0 | 1 | Ref |
| 7693 | MH834606 | Unassigned | Unassigned | Unassigned | 1166_0 | Clustered | 20 | VC_1166_0 | 20 | 0.99 | 1 | 0.99 | 0 | 0 | 0 | 1 | Ref |
| 7694 | MH834607 | Unassigned | Unassigned | Unassigned | 1362_0 | Clustered | 6 | VC_1362_0 | 6 | 1 | 1 | 1 | 0 | 0 | 0 | 1 | Ref |
| 7695 | MH834608 | Unassigned | Unassigned | Unassigned | 1166_0 | Clustered | 20 | VC_1166_0 | 20 | 0.99 | 1 | 0.99 | 0 | 0 | 0 | 1 | Ref |
| 7696 | MH834609 | Unassigned | Unassigned | Unassigned | 1166_0 | Clustered | 20 | VC_1166_0 | 20 | 0.99 | 1 | 0.99 | 0 | 0 | 0 | 1 | Ref |
| 7697 | MH834610 | Unassigned | Unassigned | Unassigned | 1207_0 | Clustered | 29 | VC_1207_0 | 29 | 0.8225 | 1 | 0.8225 | 0 | 0 | 0 | 1 | Ref |
| 7698 | MH834611 | Unassigned | Unassigned | Unassigned | 1035_0 | Clustered | 6 | VC_1035_0 | 6 | 0.4854 | 1 | 0.4854 | 0 | 0 | 0 | 1 | Ref |
| 7699 | MH834612 | Unassigned | Unassigned | Unassigned | 1036_0 | Clustered | 11 | VC_1036_0 | 11 | 0.656 | 1 | 0.656 | 0 | 0 | 0 | 0.8727 | Ref |
| 7700 | MH834613 | Unassigned | Unassigned | Unassigned | 571_0 | Clustered | 4 | VC_571_0 | 4 | 0.1921 | 0.99998591 | 0.1921 | 0 | 0 | 0 | 1 | Ref |
| 7701 | MH834614 | Unassigned | Unassigned | Unassigned | 1035_0 | Clustered | 6 | VC_1035_0 | 6 | 0.4854 | 1 | 0.4854 | 0 | 0 | 0 | 1 | Ref |
| 7702 | MH834615 | Unassigned | Unassigned | Unassigned | 565_0 | Clustered | 20 | VC_565_0 | 20 | 0.8398 | 1 | 0.8398 | 0 | 0 | 0 | 1 | Ref |
| 7703 | MH834616 | Unassigned | Unassigned | Unassigned | 1166_0 | Clustered | 20 | VC_1166_0 | 20 | 0.99 | 1 | 0.99 | 0 | 0 | 0 | 1 | Ref |
| 7704 | MH834617 | Unassigned | Unassigned | Unassigned | 520_45 | Clustered | 7 | VC_520_45 | 7 | 0.0635 | 1 | 0.0635 | 0 | 0 | 0 | 0.8095 | Ref |
| 7705 | MH834618 | Unassigned | Unassigned | Unassigned | 1166_0 | Clustered | 20 | VC_1166_0 | 20 | 0.99 | 1 | 0.99 | 0 | 0 | 0 | 1 | Ref |
| 7706 | MH834619 | Unassigned | Unassigned | Unassigned | 1207_0 | Clustered | 29 | VC_1207_0 | 29 | 0.8225 | 1 | 0.8225 | 0 | 0 | 0 | 1 | Ref |
| 7707 | MH834620 | Unassigned | Unassigned | Unassigned | 1166_0 | Clustered | 20 | VC_1166_0 | 20 | 0.99 | 1 | 0.99 | 0 | 0 | 0 | 1 | Ref |
| 7708 | MH834621 | Unassigned | Unassigned | Unassigned | 1037_0 | Clustered | 6 | VC_1037_0 | 6 | 0.6798 | 1 | 0.6798 | 0 | 0 | 0 | 1 | Ref |
| 7709 | MH834622 | Unassigned | Unassigned | Unassigned | 1362_0 | Clustered | 6 | VC_1362_0 | 6 | 1 | 1 | 1 | 0 | 0 | 0 | 1 | Ref |
| 7710 | MH834623 | Unassigned | Unassigned | Unassigned | 1035_0 | Clustered | 6 | VC_1035_0 | 6 | 0.4854 | 1 | 0.4854 | 0 | 0 | 0 | 1 | Ref |
| 7711 | MH834624 | Unassigned | Unassigned | Unassigned | 1166_0 | Clustered | 20 | VC_1166_0 | 20 | 0.99 | 1 | 0.99 | 0 | 0 | 0 | 1 | Ref |
| 7712 | MH834625 | Unassigned | Unassigned | Unassigned | 1036_0 | Clustered | 11 | VC_1036_0 | 11 | 0.656 | 1 | 0.656 | 0 | 0 | 0 | 0.8727 | Ref |
| 7713 | MH834626 | Unassigned | Unassigned | Unassigned | 1234_0 | Clustered | 44 | VC_1234_0 | 44 | 1 | 1 | 1 | 0 | 0 | 0 | 1 | Ref |
| 7714 | MH834627 | Unassigned | Unassigned | Unassigned | 1037_0 | Clustered | 6 | VC_1037_0 | 6 | 0.6798 | 1 | 0.6798 | 0 | 0 | 0 | 1 | Ref |
| 7715 | MH834628 | Unassigned | Unassigned | Unassigned | 951_0 | Clustered | 3 | VC_951_0 | 3 | 0.1049 | 0.99921864 | 0.1048 | 0 | 0 | 0 | 1 | Ref |
| 7716 | MH834629 | Unassigned | Unassigned | Unassigned | 1207_0 | Clustered | 29 | VC_1207_0 | 29 | 0.8225 | 1 | 0.8225 | 0 | 0 | 0 | 1 | Ref |
| 7717 | MH837542 | Unassigned | Unassigned | Unassigned | 302_0 | Clustered | 2 | VC_302_0 | 2 | 0.1917 | 0.94974203 | 0.182 | 0 | 0 | 0 | 1 | Ref |
| 7718 | MH837543 | Unassigned | Unassigned | Unassigned | 302_0 | Clustered | 2 | VC_302_0 | 2 | 0.1917 | 0.94974203 | 0.182 | 0 | 0 | 0 | 1 | Ref |
| 7719 | MH837626 | Unassigned | Unassigned | Unassigned | 283_159 | Clustered/Singleton | 1 | VC_283_159 | 1 | 0 | 1 | 0 | 0 | 0 | 0 | 1 | Ref |
| 7720 | MH844528 | Unassigned | Unassigned | Unassigned | 149_8 | Clustered | 66 | VC_149_8 | 66 | 0.4565 | 1 | 0.4565 | 0 | 0 | 0 | 0.8182 | Ref |
| 7721 | MH844529 | Unassigned | Unassigned | Unassigned | 149_8 | Clustered | 66 | VC_149_8 | 66 | 0.4565 | 1 | 0.4565 | 0 | 0 | 0 | 0.8182 | Ref |
| 7722 | MH844531 | Unassigned | Unassigned | Unassigned | 938_0 | Clustered | 99 | VC_938_0 | 99 | 0.9119 | 1 | 0.9119 | 0 | 0 | 0 | 0.9928 | Ref |
| 7723 | MH853786 | Unassigned | Unassigned | Unassigned | 698_0 | Clustered | 26 | VC_698_0 | 26 | 0.9366 | 1 | 0.9366 | 0 | 0 | 0 | 1 | Ref |
| 7724 | MH853787 | Unassigned | Unassigned | Unassigned | 698_0 | Clustered | 26 | VC_698_0 | 26 | 0.9366 | 1 | 0.9366 | 0 | 0 | 0 | 1 | Ref |
| 7725 | MH853788 | Unassigned | Unassigned | Unassigned | 698_0 | Clustered | 26 | VC_698_0 | 26 | 0.9366 | 1 | 0.9366 | 0 | 0 | 0 | 1 | Ref |
| 7726 | MH880817 | Unassigned | Unassigned | Unassigned | 92_0 | Clustered | 22 | VC_92_0 | 22 | 0.5639 | 1 | 0.5639 | 0 | 0 | 0 | 1 | Ref |
| 7727 | MH884508 | Unassigned | Unassigned | Unassigned | 475_4 | Clustered/Singleton | 1 | VC_475_4 | 1 | 0 | 1 | 0 | 0 | 0 | 0 | 1 | Ref |
| 7728 | MH884511 | Unassigned | Unassigned | Unassigned | 87_2 | Clustered/Singleton | 1 | VC_87_2 | 1 | 0 | 1 | 0 | 0 | 0 | 0 | 1 | Ref |
| 7729 | MH884512 | Unassigned | Unassigned | Unassigned | 622_0 | Clustered | 2 | VC_622_0 | 2 | 0.0507 | 0.6259393 | 0.0317 | 0 | 0 | 0 | 1 | Ref |
| 7730 | MH884513 | Unassigned | Unassigned | Unassigned | 100_0 | Clustered | 15 | VC_100_0 | 15 | 0.6536 | 1 | 0.6536 | 0 | 0 | 0 | 1 | Ref |
| 7731 | MH884514 | Unassigned | Unassigned | Unassigned | 101_0 | Clustered | 3 | VC_101_0 | 3 | 0.3621 | 0.99764884 | 0.3612 | 0 | 0 | 0 | 1 | Ref |
| 7732 | MH885472 | Unassigned | Unassigned | Unassigned | 111_0 | Clustered | 91 | VC_111_0 | 91 | 0.9587 | 1 | 0.9587 | 0 | 0 | 0 | 1 | Ref |
| 7733 | MH892350 | Unassigned | Unassigned | Unassigned | 256_0 | Clustered | 14 | VC_256_0 | 14 | 0.2368 | 1 | 0.2368 | 0 | 0 | 0 | 1 | Ref |
| 7734 | MH892351 | Unassigned | Unassigned | Unassigned | 252_0 | Clustered | 121 | VC_252_0 | 121 | 0.7376 | 1 | 0.7376 | 0 | 0 | 0 | 0.9999 | Ref |
| 7735 | MH892352 | Unassigned | Unassigned | Unassigned | 252_0 | Clustered | 121 | VC_252_0 | 121 | 0.7376 | 1 | 0.7376 | 0 | 0 | 0 | 0.9999 | Ref |
| 7736 | MH892353 | Unassigned | Unassigned | Unassigned | 252_0 | Clustered | 121 | VC_252_0 | 121 | 0.7376 | 1 | 0.7376 | 0 | 0 | 0 | 0.9999 | Ref |
| 7737 | MH892354 | Unassigned | Unassigned | Unassigned | 252_0 | Clustered | 121 | VC_252_0 | 121 | 0.7376 | 1 | 0.7376 | 0 | 0 | 0 | 0.9999 | Ref |
| 7738 | MH892355 | Unassigned | Unassigned | Unassigned | 255_0 | Clustered | 13 | VC_255_0 | 13 | 0.2048 | 1 | 0.2048 | 0 | 0 | 0 | 1 | Ref |
| 7739 | MH892356 | Unassigned | Unassigned | Unassigned | 252_0 | Clustered | 121 | VC_252_0 | 121 | 0.7376 | 1 | 0.7376 | 0 | 0 | 0 | 0.9999 | Ref |
| 7740 | MH892357 | Unassigned | Unassigned | Unassigned | 252_0 | Clustered | 121 | VC_252_0 | 121 | 0.7376 | 1 | 0.7376 | 0 | 0 | 0 | 0.9999 | Ref |
| 7741 | MH892358 | Unassigned | Unassigned | Unassigned | 252_0 | Clustered | 121 | VC_252_0 | 121 | 0.7376 | 1 | 0.7376 | 0 | 0 | 0 | 0.9999 | Ref |
| 7742 | MH892359 | Unassigned | Unassigned | Unassigned | 252_0 | Clustered | 121 | VC_252_0 | 121 | 0.7376 | 1 | 0.7376 | 0 | 0 | 0 | 0.9999 | Ref |
| 7743 | MH892360 | Unassigned | Unassigned | Unassigned | 252_0 | Clustered | 121 | VC_252_0 | 121 | 0.7376 | 1 | 0.7376 | 0 | 0 | 0 | 0.9999 | Ref |
| 7744 | MH892361 | Unassigned | Unassigned | Unassigned | 252_0 | Clustered | 121 | VC_252_0 | 121 | 0.7376 | 1 | 0.7376 | 0 | 0 | 0 | 0.9999 | Ref |
| 7745 | MH892362 | Unassigned | Unassigned | Unassigned | 254_0 | Clustered | 46 | VC_254_0 | 46 | 0.5159 | 1 | 0.5159 | 0 | 0 | 0 | 1 | Ref |
| 7746 | MH892363 | Unassigned | Unassigned | Unassigned | 254_0 | Clustered | 46 | VC_254_0 | 46 | 0.5159 | 1 | 0.5159 | 0 | 0 | 0 | 1 | Ref |
| 7747 | MH892364 | Unassigned | Unassigned | Unassigned | 254_0 | Clustered | 46 | VC_254_0 | 46 | 0.5159 | 1 | 0.5159 | 0 | 0 | 0 | 1 | Ref |
| 7748 | MH892365 | Unassigned | Unassigned | Unassigned | 256_0 | Clustered | 14 | VC_256_0 | 14 | 0.2368 | 1 | 0.2368 | 0 | 0 | 0 | 1 | Ref |
| 7749 | MH892366 | Unassigned | Unassigned | Unassigned | 254_0 | Clustered | 46 | VC_254_0 | 46 | 0.5159 | 1 | 0.5159 | 0 | 0 | 0 | 1 | Ref |
| 7750 | MH892367 | Unassigned | Unassigned | Unassigned | 255_0 | Clustered | 13 | VC_255_0 | 13 | 0.2048 | 1 | 0.2048 | 0 | 0 | 0 | 1 | Ref |
| 7751 | MH892368 | Unassigned | Unassigned | Unassigned | 256_0 | Clustered | 14 | VC_256_0 | 14 | 0.2368 | 1 | 0.2368 | 0 | 0 | 0 | 1 | Ref |
| 7752 | MH892369 | Unassigned | Unassigned | Unassigned | 256_0 | Clustered | 14 | VC_256_0 | 14 | 0.2368 | 1 | 0.2368 | 0 | 0 | 0 | 1 | Ref |
| 7753 | MH892370 | Unassigned | Unassigned | Unassigned | 256_0 | Clustered | 14 | VC_256_0 | 14 | 0.2368 | 1 | 0.2368 | 0 | 0 | 0 | 1 | Ref |
| 7754 | MH892371 | Unassigned | Unassigned | Unassigned | 256_0 | Clustered | 14 | VC_256_0 | 14 | 0.2368 | 1 | 0.2368 | 0 | 0 | 0 | 1 | Ref |
| 7755 | MH892372 | Unassigned | Unassigned | Unassigned | 256_0 | Clustered | 14 | VC_256_0 | 14 | 0.2368 | 1 | 0.2368 | 0 | 0 | 0 | 1 | Ref |
| 7756 | MH892373 | Unassigned | Unassigned | Unassigned | 255_0 | Clustered | 13 | VC_255_0 | 13 | 0.2048 | 1 | 0.2048 | 0 | 0 | 0 | 1 | Ref |
| 7757 | MH892374 | Unassigned | Unassigned | Unassigned | 256_0 | Clustered | 14 | VC_256_0 | 14 | 0.2368 | 1 | 0.2368 | 0 | 0 | 0 | 1 | Ref |
| 7758 | MH892375 | Unassigned | Unassigned | Unassigned | 252_0 | Clustered | 121 | VC_252_0 | 121 | 0.7376 | 1 | 0.7376 | 0 | 0 | 0 | 0.9999 | Ref |
| 7759 | MH892376 | Unassigned | Unassigned | Unassigned | 254_0 | Clustered | 46 | VC_254_0 | 46 | 0.5159 | 1 | 0.5159 | 0 | 0 | 0 | 1 | Ref |
| 7760 | MH892377 | Unassigned | Unassigned | Unassigned | 252_0 | Clustered | 121 | VC_252_0 | 121 | 0.7376 | 1 | 0.7376 | 0 | 0 | 0 | 0.9999 | Ref |
| 7761 | MH892378 | Unassigned | Unassigned | Unassigned | 252_0 | Clustered | 121 | VC_252_0 | 121 | 0.7376 | 1 | 0.7376 | 0 | 0 | 0 | 0.9999 | Ref |
| 7762 | MH892379 | Unassigned | Unassigned | Unassigned | 252_0 | Clustered | 121 | VC_252_0 | 121 | 0.7376 | 1 | 0.7376 | 0 | 0 | 0 | 0.9999 | Ref |
| 7763 | MH892380 | Unassigned | Unassigned | Unassigned | 252_0 | Clustered | 121 | VC_252_0 | 121 | 0.7376 | 1 | 0.7376 | 0 | 0 | 0 | 0.9999 | Ref |
| 7764 | MH892381 | Unassigned | Unassigned | Unassigned | 252_0 | Clustered | 121 | VC_252_0 | 121 | 0.7376 | 1 | 0.7376 | 0 | 0 | 0 | 0.9999 | Ref |
| 7765 | MH892382 | Unassigned | Unassigned | Unassigned | 252_0 | Clustered | 121 | VC_252_0 | 121 | 0.7376 | 1 | 0.7376 | 0 | 0 | 0 | 0.9999 | Ref |
| 7766 | MH892383 | Unassigned | Unassigned | Unassigned | 254_0 | Clustered | 46 | VC_254_0 | 46 | 0.5159 | 1 | 0.5159 | 0 | 0 | 0 | 1 | Ref |
| 7767 | MH892384 | Unassigned | Unassigned | Unassigned | 254_0 | Clustered | 46 | VC_254_0 | 46 | 0.5159 | 1 | 0.5159 | 0 | 0 | 0 | 1 | Ref |
| 7768 | MH892385 | Unassigned | Unassigned | Unassigned | 254_0 | Clustered | 46 | VC_254_0 | 46 | 0.5159 | 1 | 0.5159 | 0 | 0 | 0 | 1 | Ref |
| 7769 | MH899585 | Unassigned | Unassigned | Unassigned | 780_0 | Clustered | 8 | VC_780_0 | 8 | 0.5207 | 1 | 0.5207 | 0 | 0 | 0 | 1 | Ref |
| 7770 | MH910036 | Unassigned | Unassigned | Unassigned | 1036_0 | Clustered | 11 | VC_1036_0 | 11 | 0.656 | 1 | 0.656 | 0 | 0 | 0 | 0.8727 | Ref |
| 7771 | MH910037 | Unassigned | Unassigned | Unassigned | 1036_0 | Clustered | 11 | VC_1036_0 | 11 | 0.656 | 1 | 0.656 | 0 | 0 | 0 | 0.8727 | Ref |
| 7772 | MH910038 | Unassigned | Unassigned | Unassigned | 237_3 | Clustered | 8 | VC_237_3 | 8 | 0.1092 | 1 | 0.1092 | 0 | 0 | 0 | 1 | Ref |
| 7773 | MH910039 | Unassigned | Unassigned | Unassigned | 237_3 | Clustered | 8 | VC_237_3 | 8 | 0.1092 | 1 | 0.1092 | 0 | 0 | 0 | 1 | Ref |
| 7774 | MH910040 | Unassigned | Unassigned | Unassigned | 558_33 | Clustered/Singleton | 1 | VC_558_33 | 1 | 0 | 1 | 0 | 0 | 0 | 0 | 1 | Ref |

| | | | | | | | | | | | | | | | | | |
|---|---|---|---|---|---|---|---|---|---|---|---|---|---|---|---|---|---|
| 7775 | MH910041 | Unassigned | Unassigned | Unassigned | 1036_0 | Clustered | 11 | VC_1036_0 | 11 | 0.656 | 1 | 0.656 | 0 | 0 | 0 | 0.8727 | Ref |
| 7776 | MH910042 | Unassigned | Unassigned | Unassigned | 237_3 | Clustered | 8 | VC_237_3 | 8 | 0.1092 | 1 | 0.1092 | 0 | 0 | 0 | 1 | Ref |
| 7777 | MH917278 | Unassigned | Unassigned | Unassigned | 575_0 | Clustered | 35 | VC_575_0 | 35 | 0.8815 | 1 | 0.8815 | 0 | 0 | 0 | 1 | Ref |
| 7778 | MH920362 | Unassigned | Unassigned | Unassigned | 330_0 | Clustered | 27 | VC_330_0 | 27 | 0.5088 | 1 | 0.5088 | 0 | 0 | 0 | 0.8661 | Ref |
| 7779 | MH920639 | Unassigned | Unassigned | Unassigned | 655_7 | Clustered | 3 | VC_655_7 | 3 | 0.0166 | 0.99856978 | 0.0166 | 0 | 0 | 0 | 1 | Ref |
| 7780 | MH920640 | Unassigned | Unassigned | Unassigned | 655_7 | Clustered | 3 | VC_655_7 | 3 | 0.0166 | 0.99856978 | 0.0166 | 0 | 0 | 0 | 1 | Ref |
| 7781 | MH925090 | Unassigned | Unassigned | Unassigned | 882_0 | Clustered | 20 | VC_882_0 | 20 | 0.9917 | 1 | 0.9917 | 0 | 0 | 0 | 1 | Ref |
| 7782 | MH925091 | Unassigned | Unassigned | Unassigned | 904_2 | Clustered | 7 | VC_904_2 | 7 | 0.2857 | 1 | 0.2857 | 0 | 0 | 0 | 1 | Ref |
| 7783 | MH925092 | Unassigned | Unassigned | Unassigned | 1050_1 | Clustered | 29 | VC_1050_1 | 29 | 0.4861 | 1 | 0.4861 | 0 | 0 | 0 | 0.9581 | Ref |
| 7784 | MH925094 | Unassigned | Unassigned | Unassigned | 972_0 | Clustered | 6 | VC_972_0 | 6 | 0.2892 | 1 | 0.2892 | 0 | 0 | 0 | 1 | Ref |
| 7785 | MH926055 | Unassigned | Unassigned | Unassigned | 521_0 | Clustered | 37 | VC_521_0 | 37 | 0.912 | 1 | 0.912 | 0 | 0 | 0 | 1 | Ref |
| 7786 | MH926056 | Unassigned | Unassigned | Unassigned | 768_0 | Clustered | 8 | VC_768_0 | 8 | 0.7012 | 1 | 0.7012 | 0 | 0 | 0 | 1 | Ref |
| 7787 | MH926057 | Unassigned | Unassigned | Unassigned | 768_0 | Clustered | 8 | VC_768_0 | 8 | 0.7012 | 1 | 0.7012 | 0 | 0 | 0 | 1 | Ref |
| 7788 | MH926058 | Unassigned | Unassigned | Unassigned | 234_0 | Clustered | 21 | VC_234_0 | 21 | 0.8355 | 1 | 0.8355 | 0 | 0 | 0 | 1 | Ref |
| 7789 | MH926059 | Unassigned | Unassigned | Unassigned | 568_0 | Clustered | 8 | VC_568_0 | 8 | 0.5738 | 1 | 0.5738 | 0 | 0 | 0 | 1 | Ref |
| 7790 | MH926060 | Unassigned | Unassigned | Unassigned | 371_1 | Clustered | 11 | VC_371_1 | 11 | 0.0379 | 1 | 0.0379 | 0 | 0 | 0 | 0.8545 | Ref |
| 7791 | MH926061 | Unassigned | Unassigned | Unassigned | 1199_0 | Clustered | 8 | VC_1199_0 | 8 | 0.7928 | 1 | 0.7928 | 0 | 0 | 0 | 1 | Ref |
| 7792 | MH926062 | Unassigned | Unassigned | Unassigned | 556_0 | Clustered | 21 | VC_556_0 | 21 | 0.9912 | 1 | 0.9912 | 0 | 0 | 0 | 1 | Ref |
| 7793 | MH929319 | Unassigned | Unassigned | Unassigned | 132_0 | Clustered | 4 | VC_132_0 | 4 | 0.036 | 0.99998777 | 0.036 | 0 | 0 | 0 | 1 | Ref |
| 7794 | MH931003 | Unassigned | Unassigned | Unassigned | 1054_0 | Clustered | 12 | VC_1054_0 | 12 | 0.2863 | 0.99746644 | 0.2855 | 0 | 0 | 0 | 1 | Ref |
| 7795 | MH931004 | Unassigned | Unassigned | Unassigned | 1077_0 | Clustered | 48 | VC_1077_0 | 48 | 0.8525 | 1 | 0.8525 | 0 | 0 | 0 | 1 | Ref |
| 7796 | MH937457 | Unassigned | Unassigned | Unassigned | 254_0 | Clustered | 46 | VC_254_0 | 46 | 0.5159 | 1 | 0.5159 | 0 | 0 | 0 | 1 | Ref |
| 7797 | MH937458 | Unassigned | Unassigned | Unassigned | 252_0 | Clustered | 121 | VC_252_0 | 121 | 0.7376 | 1 | 0.7376 | 0 | 0 | 0 | 0.9999 | Ref |
| 7798 | MH937459 | Unassigned | Unassigned | Unassigned | 252_0 | Clustered | 121 | VC_252_0 | 121 | 0.7376 | 1 | 0.7376 | 0 | 0 | 0 | 0.9999 | Ref |
| 7799 | MH937460 | Unassigned | Unassigned | Unassigned | 254_0 | Clustered | 46 | VC_254_0 | 46 | 0.5159 | 1 | 0.5159 | 0 | 0 | 0 | 1 | Ref |
| 7800 | MH937461 | Unassigned | Unassigned | Unassigned | 252_0 | Clustered | 121 | VC_252_0 | 121 | 0.7376 | 1 | 0.7376 | 0 | 0 | 0 | 0.9999 | Ref |
| 7801 | MH937462 | Unassigned | Unassigned | Unassigned | 252_0 | Clustered | 121 | VC_252_0 | 121 | 0.7376 | 1 | 0.7376 | 0 | 0 | 0 | 0.9999 | Ref |
| 7802 | MH937463 | Unassigned | Unassigned | Unassigned | 254_0 | Clustered | 46 | VC_254_0 | 46 | 0.5159 | 1 | 0.5159 | 0 | 0 | 0 | 1 | Ref |
| 7803 | MH937464 | Unassigned | Unassigned | Unassigned | 254_0 | Clustered | 46 | VC_254_0 | 46 | 0.5159 | 1 | 0.5159 | 0 | 0 | 0 | 1 | Ref |
| 7804 | MH937465 | Unassigned | Unassigned | Unassigned | 252_0 | Clustered | 121 | VC_252_0 | 121 | 0.7376 | 1 | 0.7376 | 0 | 0 | 0 | 0.9999 | Ref |
| 7805 | MH937466 | Unassigned | Unassigned | Unassigned | 252_0 | Clustered | 121 | VC_252_0 | 121 | 0.7376 | 1 | 0.7376 | 0 | 0 | 0 | 0.9999 | Ref |
| 7806 | MH937467 | Unassigned | Unassigned | Unassigned | 252_0 | Clustered | 121 | VC_252_0 | 121 | 0.7376 | 1 | 0.7376 | 0 | 0 | 0 | 0.9999 | Ref |
| 7807 | MH937468 | Unassigned | Unassigned | Unassigned | 252_0 | Clustered | 121 | VC_252_0 | 121 | 0.7376 | 1 | 0.7376 | 0 | 0 | 0 | 0.9999 | Ref |
| 7808 | MH937469 | Unassigned | Unassigned | Unassigned | 252_0 | Clustered | 121 | VC_252_0 | 121 | 0.7376 | 1 | 0.7376 | 0 | 0 | 0 | 0.9999 | Ref |
| 7809 | MH937470 | Unassigned | Unassigned | Unassigned | 252_0 | Clustered | 121 | VC_252_0 | 121 | 0.7376 | 1 | 0.7376 | 0 | 0 | 0 | 0.9999 | Ref |
| 7810 | MH937471 | Unassigned | Unassigned | Unassigned | 252_0 | Clustered | 121 | VC_252_0 | 121 | 0.7376 | 1 | 0.7376 | 0 | 0 | 0 | 0.9999 | Ref |
| 7811 | MH937472 | Unassigned | Unassigned | Unassigned | 252_0 | Clustered | 121 | VC_252_0 | 121 | 0.7376 | 1 | 0.7376 | 0 | 0 | 0 | 0.9999 | Ref |
| 7812 | MH937473 | Unassigned | Unassigned | Unassigned | 254_0 | Clustered | 46 | VC_254_0 | 46 | 0.5159 | 1 | 0.5159 | 0 | 0 | 0 | 1 | Ref |
| 7813 | MH937474 | Unassigned | Unassigned | Unassigned | 252_0 | Clustered | 121 | VC_252_0 | 121 | 0.7376 | 1 | 0.7376 | 0 | 0 | 0 | 0.9999 | Ref |
| 7814 | MH937475 | Unassigned | Unassigned | Unassigned | 254_0 | Clustered | 46 | VC_254_0 | 46 | 0.5159 | 1 | 0.5159 | 0 | 0 | 0 | 1 | Ref |
| 7815 | MH937476 | Unassigned | Unassigned | Unassigned | 254_0 | Clustered | 46 | VC_254_0 | 46 | 0.5159 | 1 | 0.5159 | 0 | 0 | 0 | 1 | Ref |
| 7816 | MH937477 | Unassigned | Unassigned | Unassigned | 252_0 | Clustered | 121 | VC_252_0 | 121 | 0.7376 | 1 | 0.7376 | 0 | 0 | 0 | 0.9999 | Ref |
| 7817 | MH937478 | Unassigned | Unassigned | Unassigned | 254_0 | Clustered | 46 | VC_254_0 | 46 | 0.5159 | 1 | 0.5159 | 0 | 0 | 0 | 1 | Ref |
| 7818 | MH937479 | Unassigned | Unassigned | Unassigned | 254_0 | Clustered | 46 | VC_254_0 | 46 | 0.5159 | 1 | 0.5159 | 0 | 0 | 0 | 1 | Ref |
| 7819 | MH937480 | Unassigned | Unassigned | Unassigned | 252_0 | Clustered | 121 | VC_252_0 | 121 | 0.7376 | 1 | 0.7376 | 0 | 0 | 0 | 0.9999 | Ref |
| 7820 | MH937481 | Unassigned | Unassigned | Unassigned | 252_0 | Clustered | 121 | VC_252_0 | 121 | 0.7376 | 1 | 0.7376 | 0 | 0 | 0 | 0.9999 | Ref |
| 7821 | MH937482 | Unassigned | Unassigned | Unassigned | 252_0 | Clustered | 121 | VC_252_0 | 121 | 0.7376 | 1 | 0.7376 | 0 | 0 | 0 | 0.9999 | Ref |
| 7822 | MH937483 | Unassigned | Unassigned | Unassigned | 252_0 | Clustered | 121 | VC_252_0 | 121 | 0.7376 | 1 | 0.7376 | 0 | 0 | 0 | 0.9999 | Ref |
| 7823 | MH937484 | Unassigned | Unassigned | Unassigned | 254_0 | Clustered | 46 | VC_254_0 | 46 | 0.5159 | 1 | 0.5159 | 0 | 0 | 0 | 1 | Ref |
| 7824 | MH937485 | Unassigned | Unassigned | Unassigned | 252_0 | Clustered | 121 | VC_252_0 | 121 | 0.7376 | 1 | 0.7376 | 0 | 0 | 0 | 0.9999 | Ref |
| 7825 | MH937486 | Unassigned | Unassigned | Unassigned | 252_0 | Clustered | 121 | VC_252_0 | 121 | 0.7376 | 1 | 0.7376 | 0 | 0 | 0 | 0.9999 | Ref |
| 7826 | MH937487 | Unassigned | Unassigned | Unassigned | 252_0 | Clustered | 121 | VC_252_0 | 121 | 0.7376 | 1 | 0.7376 | 0 | 0 | 0 | 0.9999 | Ref |
| 7827 | MH937488 | Unassigned | Unassigned | Unassigned | 252_0 | Clustered | 121 | VC_252_0 | 121 | 0.7376 | 1 | 0.7376 | 0 | 0 | 0 | 0.9999 | Ref |
| 7828 | MH937489 | Unassigned | Unassigned | Unassigned | 252_0 | Clustered | 121 | VC_252_0 | 121 | 0.7376 | 1 | 0.7376 | 0 | 0 | 0 | 0.9999 | Ref |
| 7829 | MH937490 | Unassigned | Unassigned | Unassigned | 252_0 | Clustered | 121 | VC_252_0 | 121 | 0.7376 | 1 | 0.7376 | 0 | 0 | 0 | 0.9999 | Ref |
| 7830 | MH937491 | Unassigned | Unassigned | Unassigned | 252_0 | Clustered | 121 | VC_252_0 | 121 | 0.7376 | 1 | 0.7376 | 0 | 0 | 0 | 0.9999 | Ref |
| 7831 | MH937492 | Unassigned | Unassigned | Unassigned | 252_0 | Clustered | 121 | VC_252_0 | 121 | 0.7376 | 1 | 0.7376 | 0 | 0 | 0 | 0.9999 | Ref |
| 7832 | MH937493 | Unassigned | Unassigned | Unassigned | 252_0 | Clustered | 121 | VC_252_0 | 121 | 0.7376 | 1 | 0.7376 | 0 | 0 | 0 | 0.9999 | Ref |
| 7833 | MH937494 | Unassigned | Unassigned | Unassigned | 254_0 | Clustered | 46 | VC_254_0 | 46 | 0.5159 | 1 | 0.5159 | 0 | 0 | 0 | 1 | Ref |
| 7834 | MH937495 | Unassigned | Unassigned | Unassigned | 252_0 | Clustered | 121 | VC_252_0 | 121 | 0.7376 | 1 | 0.7376 | 0 | 0 | 0 | 0.9999 | Ref |
| 7835 | MH937496 | Unassigned | Unassigned | Unassigned | 252_0 | Clustered | 121 | VC_252_0 | 121 | 0.7376 | 1 | 0.7376 | 0 | 0 | 0 | 0.9999 | Ref |
| 7836 | MH937497 | Unassigned | Unassigned | Unassigned | 252_0 | Clustered | 121 | VC_252_0 | 121 | 0.7376 | 1 | 0.7376 | 0 | 0 | 0 | 0.9999 | Ref |
| 7837 | MH937498 | Unassigned | Unassigned | Unassigned | 254_0 | Clustered | 46 | VC_254_0 | 46 | 0.5159 | 1 | 0.5159 | 0 | 0 | 0 | 1 | Ref |
| 7838 | MH937499 | Unassigned | Unassigned | Unassigned | 252_0 | Clustered | 121 | VC_252_0 | 121 | 0.7376 | 1 | 0.7376 | 0 | 0 | 0 | 0.9999 | Ref |
| 7839 | MH937500 | Unassigned | Unassigned | Unassigned | 252_0 | Clustered | 121 | VC_252_0 | 121 | 0.7376 | 1 | 0.7376 | 0 | 0 | 0 | 0.9999 | Ref |
| 7840 | MH937501 | Unassigned | Unassigned | Unassigned | 252_0 | Clustered | 121 | VC_252_0 | 121 | 0.7376 | 1 | 0.7376 | 0 | 0 | 0 | 0.9999 | Ref |
| 7841 | MH937502 | Unassigned | Unassigned | Unassigned | 254_0 | Clustered | 46 | VC_254_0 | 46 | 0.5159 | 1 | 0.5159 | 0 | 0 | 0 | 1 | Ref |
| 7842 | MH937503 | Unassigned | Unassigned | Unassigned | 254_0 | Clustered | 46 | VC_254_0 | 46 | 0.5159 | 1 | 0.5159 | 0 | 0 | 0 | 1 | Ref |
| 7843 | MH937504 | Unassigned | Unassigned | Unassigned | 252_0 | Clustered | 121 | VC_252_0 | 121 | 0.7376 | 1 | 0.7376 | 0 | 0 | 0 | 0.9999 | Ref |
| 7844 | MH937505 | Unassigned | Unassigned | Unassigned | 254_0 | Clustered | 46 | VC_254_0 | 46 | 0.5159 | 1 | 0.5159 | 0 | 0 | 0 | 1 | Ref |
| 7845 | MH937506 | Unassigned | Unassigned | Unassigned | 252_0 | Clustered | 121 | VC_252_0 | 121 | 0.7376 | 1 | 0.7376 | 0 | 0 | 0 | 0.9999 | Ref |
| 7846 | MH937507 | Unassigned | Unassigned | Unassigned | 254_0 | Clustered | 46 | VC_254_0 | 46 | 0.5159 | 1 | 0.5159 | 0 | 0 | 0 | 1 | Ref |
| 7847 | MH937508 | Unassigned | Unassigned | Unassigned | 252_0 | Clustered | 121 | VC_252_0 | 121 | 0.7376 | 1 | 0.7376 | 0 | 0 | 0 | 0.9999 | Ref |
| 7848 | MH937509 | Unassigned | Unassigned | Unassigned | 254_0 | Clustered | 46 | VC_254_0 | 46 | 0.5159 | 1 | 0.5159 | 0 | 0 | 0 | 1 | Ref |
| 7849 | MH937510 | Unassigned | Unassigned | Unassigned | 254_0 | Clustered | 46 | VC_254_0 | 46 | 0.5159 | 1 | 0.5159 | 0 | 0 | 0 | 1 | Ref |
| 7850 | MH937511 | Unassigned | Unassigned | Unassigned | 254_0 | Clustered | 46 | VC_254_0 | 46 | 0.5159 | 1 | 0.5159 | 0 | 0 | 0 | 1 | Ref |
| 7851 | MH939157 | Unassigned | Unassigned | Unassigned | 1363_0 | Clustered | 2 | VC_1363_0 | 2 | 0.8326 | 0.72985431 | 0.6077 | 0 | 0 | 0 | 1 | Ref |
| 7852 | MH940212 | Unassigned | Unassigned | Unassigned | 133_22 | Clustered | 13 | VC_133_22 | 13 | 0.0804 | 1 | 0.0804 | 0 | 0 | 0 | 0.8205 | Ref |
| 7853 | MH972260 | Unassigned | Unassigned | Unassigned | 96_0 | Clustered | 30 | VC_96_0 | 30 | 0.718 | 1 | 0.718 | 0 | 0 | 0 | 1 | Ref |
| 7854 | MH973661 | Unassigned | Unassigned | Unassigned | 252_0 | Clustered | 121 | VC_252_0 | 121 | 0.7376 | 1 | 0.7376 | 0 | 0 | 0 | 0.9999 | Ref |
| 7855 | MH973662 | Unassigned | Unassigned | Unassigned | 252_0 | Clustered | 121 | VC_252_0 | 121 | 0.7376 | 1 | 0.7376 | 0 | 0 | 0 | 0.9999 | Ref |
| 7856 | MH973663 | Unassigned | Unassigned | Unassigned | 255_0 | Clustered | 13 | VC_255_0 | 13 | 0.2048 | 1 | 0.2048 | 0 | 0 | 0 | 1 | Ref |
| 7857 | MH973725 | Unassigned | Unassigned | Unassigned | 472_0 | Clustered | 47 | VC_472_0 | 47 | 0.9025 | 1 | 0.9025 | 0 | 0 | 0 | 1 | Ref |
| 7858 | MH976506 | Unassigned | Unassigned | Unassigned | 1093_0 | Clustered | 40 | VC_1093_0 | 40 | 0.8754 | 1 | 0.8754 | 0 | 0 | 0 | 1 | Ref |
| 7859 | MH976507 | Unassigned | Unassigned | Unassigned | 1093_0 | Clustered | 40 | VC_1093_0 | 40 | 0.8754 | 1 | 0.8754 | 0 | 0 | 0 | 1 | Ref |
| 7860 | MH976508 | Unassigned | Unassigned | Unassigned | 1093_0 | Clustered | 40 | VC_1093_0 | 40 | 0.8754 | 1 | 0.8754 | 0 | 0 | 0 | 1 | Ref |
| 7861 | MH976509 | Unassigned | Unassigned | Unassigned | 1093_0 | Clustered | 40 | VC_1093_0 | 40 | 0.8754 | 1 | 0.8754 | 0 | 0 | 0 | 1 | Ref |
| 7862 | MH976510 | Unassigned | Unassigned | Unassigned | 1093_0 | Clustered | 40 | VC_1093_0 | 40 | 0.8754 | 1 | 0.8754 | 0 | 0 | 0 | 1 | Ref |
| 7863 | MH976511 | Unassigned | Unassigned | Unassigned | 1093_0 | Clustered | 40 | VC_1093_0 | 40 | 0.8754 | 1 | 0.8754 | 0 | 0 | 0 | 1 | Ref |
| 7864 | MH976512 | Unassigned | Unassigned | Unassigned | 1093_0 | Clustered | 40 | VC_1093_0 | 40 | 0.8754 | 1 | 0.8754 | 0 | 0 | 0 | 1 | Ref |
| 7865 | MH976513 | Unassigned | Unassigned | Unassigned | 1093_0 | Clustered | 40 | VC_1093_0 | 40 | 0.8754 | 1 | 0.8754 | 0 | 0 | 0 | 1 | Ref |
| 7866 | MH976514 | Unassigned | Unassigned | Unassigned | 1093_0 | Clustered | 40 | VC_1093_0 | 40 | 0.8754 | 1 | 0.8754 | 0 | 0 | 0 | 1 | Ref |
| 7867 | MH976515 | Unassigned | Unassigned | Unassigned | 890_1 | Clustered | 4 | VC_890_1 | 4 | 0.4613 | 0.99998458 | 0.4613 | 0 | 0 | 0 | 1 | Ref |
| 7868 | MH976516 | Unassigned | Unassigned | Unassigned | 768_0 | Clustered | 8 | VC_768_0 | 8 | 0.7012 | 1 | 0.7012 | 0 | 0 | 0 | 1 | Ref |
| 7869 | MH976517 | Unassigned | Unassigned | Unassigned | 513_0 | Clustered | 32 | VC_513_0 | 32 | 0.6881 | 1 | 0.6881 | 0 | 0 | 0 | 1 | Ref |
| 7870 | MH976518 | Unassigned | Unassigned | Unassigned | 1093_0 | Clustered | 40 | VC_1093_0 | 40 | 0.8754 | 1 | 0.8754 | 0 | 0 | 0 | 1 | Ref |
| 7871 | MH976519 | Unassigned | Unassigned | Unassigned | 1093_0 | Clustered | 40 | VC_1093_0 | 40 | 0.8754 | 1 | 0.8754 | 0 | 0 | 0 | 1 | Ref |
| 7872 | MH979674 | Unassigned | Unassigned | Unassigned | 423_0 | Clustered | 45 | VC_423_0 | 45 | 0.9484 | 1 | 0.9484 | 0 | 0 | 0 | 1 | Ref |
| 7873 | MH983004 | Unassigned | Unassigned | Unassigned | 272_0 | Clustered | 11 | VC_272_0 | 11 | 0.4915 | 1 | 0.4915 | 0 | 0 | 0 | 0.8727 | Ref |
| 7874 | MH992121 | Unassigned | Unassigned | Unassigned | 283_188 | Clustered/Singleton | 1 | VC_283_188 | 1 | 0 | 1 | 0 | 0 | 0 | 0 | 1 | Ref |
| 7875 | MH992122 | Unassigned | Unassigned | Unassigned | 286_10 | Clustered | 9 | VC_286_10 | 9 | 0.0615 | 0.99999878 | 0.0615 | 0 | 0 | 0 | 0.6667 | Ref |
| 7876 | MH992123 | Unassigned | Unassigned | Unassigned | 779_0 | Clustered | 37 | VC_779_0 | 37 | 0.5682 | 1 | 0.5682 | 0 | 0 | 0 | 1 | Ref |
| 7877 | MH992131 | Unassigned | Unassigned | Unassigned | 805_0 | Clustered | 4 | VC_805_0 | 4 | 0.9081 | 0.99622688 | 0.9046 | 0 | 0 | 0 | 1 | Ref |
| 7878 | MH992154 | Unassigned | Unassigned | Unassigned | 1364_0 | Clustered | 3 | VC_1364_0 | 3 | 1 | 1 | 1 | 0 | 0 | 0 | 1 | Ref |
| 7879 | MH992155 | Unassigned | Unassigned | Unassigned | 1365_0 | Clustered | 3 | VC_1365_0 | 3 | 1 | 1 | 1 | 0 | 0 | 0 | 1 | Ref |
| 7880 | MH992156 | Unassigned | Unassigned | Unassigned | 995_0 | Clustered | 69 | VC_995_0 | 69 | 0.8547 | 1 | 0.8547 | 0 | 0 | 0 | 1 | Ref |
| 7881 | MH992157 | Unassigned | Unassigned | Unassigned | 198_0 | Clustered | 28 | VC_198_0 | 28 | 0.2429 | 0.99971175 | 0.2429 | 0 | 0 | 0 | 1 | Ref |
| 7882 | MH992158 | Unassigned | Unassigned | Unassigned | 327_0 | Clustered | 12 | VC_327_0 | 12 | 0.8925 | 1 | 0.8925 | 0 | 0 | 0 | 1 | Ref |
| 7883 | MH992159 | Unassigned | Unassigned | Unassigned | 1339_0 | Clustered | 7 | VC_1339_0 | 7 | 0.9177 | 0.99772305 | 0.9156 | 0 | 0 | 0 | 1 | Ref |
| 7884 | MH992160 | Unassigned | Unassigned | Unassigned | 327_0 | Clustered | 12 | VC_327_0 | 12 | 0.8925 | 1 | 0.8925 | 0 | 0 | 0 | 1 | Ref |
| 7885 | MH992161 | Unassigned | Unassigned | Unassigned | 995_0 | Clustered | 69 | VC_995_0 | 69 | 0.8547 | 1 | 0.8547 | 0 | 0 | 0 | 1 | Ref |
| 7886 | MH992163 | Unassigned | Unassigned | Unassigned | 1077_0 | Clustered | 48 | VC_1077_0 | 48 | 0.8525 | 1 | 0.8525 | 0 | 0 | 0 | 1 | Ref |
| 7887 | MH992164 | Unassigned | Unassigned | Unassigned | 1077_0 | Clustered | 48 | VC_1077_0 | 48 | 0.8525 | 1 | 0.8525 | 0 | 0 | 0 | 1 | Ref |
| 7888 | MH992165 | Unassigned | Unassigned | Unassigned | 1066_0 | Clustered | 52 | VC_1066_0 | 52 | 0.7682 | 1 | 0.7682 | 0 | 0 | 0 | 1 | Ref |
| 7889 | MH992167 | Unassigned | Unassigned | Unassigned | 1367_0 | Clustered | 2 | VC_1367_0 | 2 | 1 | 1 | 1 | 0 | 0 | 0 | 1 | Ref |
| 7890 | MH992169 | Unassigned | Unassigned | Unassigned | 997_0 | Clustered/Singleton | 1 | VC_997_0 | 1 | 0 | 1 | 0 | 0 | 0 | 0 | 1 | Ref |
| 7891 | MH992170 | Unassigned | Unassigned | Unassigned | 1059_0 | Clustered | 3 | VC_1059_0 | 3 | 0.3928 | 0.9839946 | 0.3865 | 0 | 0 | 0 | 1 | Ref |
| 7892 | MH992171 | Unassigned | Unassigned | Unassigned | 1368_0 | Clustered | 2 | VC_1368_0 | 2 | 1 | 1 | 1 | 0 | 0 | 0 | 1 | Ref |
| 7893 | MH992173 | Unassigned | Unassigned | Unassigned | 1324_0 | Clustered | 4 | VC_1324_0 | 4 | 0.6008 | 0.54407361 | 0.3269 | 0 | 0 | 0 | 1 | Ref |
| 7894 | MH992174 | Unassigned | Unassigned | Unassigned | 198_0 | Clustered | 28 | VC_198_0 | 28 | 0.2429 | 0.99971175 | 0.2429 | 0 | 0 | 0 | 1 | Ref |
| 7895 | MH992175 | Unassigned | Unassigned | Unassigned | 1339_0 | Clustered | 7 | VC_1339_0 | 7 | 0.9177 | 0.99772305 | 0.9156 | 0 | 0 | 0 | 1 | Ref |
| 7896 | MH992177 | Unassigned | Unassigned | Unassigned | 327_0 | Clustered | 12 | VC_327_0 | 12 | 0.8925 | 1 | 0.8925 | 0 | 0 | 0 | 1 | Ref |
| 7897 | MH992178 | Unassigned | Unassigned | Unassigned | 327_0 | Clustered | 12 | VC_327_0 | 12 | 0.8925 | 1 | 0.8925 | 0 | 0 | 0 | 1 | Ref |
| 7898 | MH992180 | Unassigned | Unassigned | Unassigned | 1062_0 | Clustered | 45 | VC_1062_0 | 45 | 0.9734 | 0.50949847 | 0.4959 | 0 | 0 | 0 | 1 | Ref |
| 7899 | MH992181 | Unassigned | Unassigned | Unassigned | 200_0 | Clustered/Singleton | 1 | VC_200_0 | 1 | 0 | 1 | 0 | 0 | 0 | 0 | 1 | Ref |
| 7900 | MH992182 | Unassigned | Unassigned | Unassigned | 198_0 | Clustered | 28 | VC_198_0 | 28 | 0.2429 | 0.99971175 | 0.2429 | 0 | 0 | 0 | 1 | Ref |
| 7901 | MH992184 | Unassigned | Unassigned | Unassigned | 995_0 | Clustered | 69 | VC_995_0 | 69 | 0.8547 | 1 | 0.8547 | 0 | 0 | 0 | 1 | Ref |
| 7902 | MH992185 | Unassigned | Unassigned | Unassigned | 995_0 | Clustered | 69 | VC_995_0 | 69 | 0.8547 | 1 | 0.8547 | 0 | 0 | 0 | 1 | Ref |
| 7903 | MH992189 | Unassigned | Unassigned | Unassigned | 327_0 | Clustered | 12 | VC_327_0 | 12 | 0.8925 | 1 | 0.8925 | 0 | 0 | 0 | 1 | Ref |
| 7904 | MH992191 | Unassigned | Unassigned | Unassigned | 327_0 | Clustered | 12 | VC_327_0 | 12 | 0.8925 | 1 | 0.8925 | 0 | 0 | 0 | 1 | Ref |
| 7905 | MH992192 | Unassigned | Unassigned | Unassigned | 1077_0 | Clustered | 48 | VC_1077_0 | 48 | 0.8525 | 1 | 0.8525 | 0 | 0 | 0 | 1 | Ref |
| 7906 | MH992193 | Unassigned | Unassigned | Unassigned | 1059_0 | Clustered | 3 | VC_1059_0 | 3 | 0.3928 | 0.9839946 | 0.3865 | 0 | 0 | 0 | 1 | Ref |
| 7907 | MH992194 | Unassigned | Unassigned | Unassigned | 327_0 | Clustered | 12 | VC_327_0 | 12 | 0.8925 | 1 | 0.8925 | 0 | 0 | 0 | 1 | Ref |
| 7908 | MH992195 | Unassigned | Unassigned | Unassigned | 398_0 | Clustered | 48 | VC_398_0 | 48 | 0.4813 | 1 | 0.4813 | 0 | 0 | 0 | 1 | Ref |
| 7909 | MH992196 | Unassigned | Unassigned | Unassigned | 1365_0 | Clustered | 3 | VC_1365_0 | 3 | 1 | 1 | 1 | 0 | 0 | 0 | 1 | Ref |
| 7910 | MH992197 | Unassigned | Unassigned | Unassigned | 327_0 | Clustered | 12 | VC_327_0 | 12 | 0.8925 | 1 | 0.8925 | 0 | 0 | 0 | 1 | Ref |
| 7911 | MH992199 | Unassigned | Unassigned | Unassigned | 1077_0 | Clustered | 48 | VC_1077_0 | 48 | 0.8525 | 1 | 0.8525 | 0 | 0 | 0 | 1 | Ref |
| 7912 | MH992200 | Unassigned | Unassigned | Unassigned | 1339_0 | Clustered | 7 | VC_1339_0 | 7 | 0.9177 | 0.99772305 | 0.9156 | 0 | 0 | 0 | 1 | Ref |
| 7913 | MH992202 | Unassigned | Unassigned | Unassigned | 1077_0 | Clustered | 48 | VC_1077_0 | 48 | 0.8525 | 1 | 0.8525 | 0 | 0 | 0 | 1 | Ref |
| 7914 | MH992205 | Unassigned | Unassigned | Unassigned | 1365_0 | Clustered | 3 | VC_1365_0 | 3 | 1 | 1 | 1 | 0 | 0 | 0 | 1 | Ref |
| 7915 | MH992206 | Unassigned | Unassigned | Unassigned | 1077_0 | Clustered | 48 | VC_1077_0 | 48 | 0.8525 | 1 | 0.8525 | 0 | 0 | 0 | 1 | Ref |
| 7916 | MH992207 | Unassigned | Unassigned | Unassigned | 327_0 | Clustered | 12 | VC_327_0 | 12 | 0.8925 | 1 | 0.8925 | 0 | 0 | 0 | 1 | Ref |
| 7917 | MH992208 | Unassigned | Unassigned | Unassigned | 1077_0 | Clustered | 48 | VC_1077_0 | 48 | 0.8525 | 1 | 0.8525 | 0 | 0 | 0 | 1 | Ref |
| 7918 | MH992210 | Unassigned | Unassigned | Unassigned | 198_0 | Clustered | 28 | VC_198_0 | 28 | 0.2429 | 0.99971175 | 0.2429 | 0 | 0 | 0 | 1 | Ref |
| 7919 | MH992211 | Unassigned | Unassigned | Unassigned | 398_0 | Clustered | 48 | VC_398_0 | 48 | 0.4813 | 1 | 0.4813 | 0 | 0 | 0 | 1 | Ref |
| 7920 | MH992213 | Unassigned | Unassigned | Unassigned | 995_0 | Clustered | 69 | VC_995_0 | 69 | 0.8547 | 1 | 0.8547 | 0 | 0 | 0 | 1 | Ref |
| 7921 | MH992214 | Unassigned | Unassigned | Unassigned | 1077_0 | Clustered | 48 | VC_1077_0 | 48 | 0.8525 | 1 | 0.8525 | 0 | 0 | 0 | 1 | Ref |
| 7922 | MH992215 | Unassigned | Unassigned | Unassigned | 1077_0 | Clustered | 48 | VC_1077_0 | 48 | 0.8525 | 1 | 0.8525 | 0 | 0 | 0 | 1 | Ref |
| 7923 | MH992216 | Unassigned | Unassigned | Unassigned | 1077_0 | Clustered | 48 | VC_1077_0 | 48 | 0.8525 | 1 | 0.8525 | 0 | 0 | 0 | 1 | Ref |
| 7924 | MH992217 | Unassigned | Unassigned | Unassigned | 1077_0 | Clustered | 48 | VC_1077_0 | 48 | 0.8525 | 1 | 0.8525 | 0 | 0 | 0 | 1 | Ref |
| 7925 | MH992218 | Unassigned | Unassigned | Unassigned | 327_0 | Clustered | 12 | VC_327_0 | 12 | 0.8925 | 1 | 0.8925 | 0 | 0 | 0 | 1 | Ref |
| 7926 | MH992219 | Unassigned | Unassigned | Unassigned | 1077_0 | Clustered | 48 | VC_1077_0 | 48 | 0.8525 | 1 | 0.8525 | 0 | 0 | 0 | 1 | Ref |
| 7927 | MH992220 | Unassigned | Unassigned | Unassigned | 1077_0 | Clustered | 48 | VC_1077_0 | 48 | 0.8525 | 1 | 0.8525 | 0 | 0 | 0 | 1 | Ref |
| 7928 | MH992222 | Unassigned | Unassigned | Unassigned | 1077_0 | Clustered | 48 | VC_1077_0 | 48 | 0.8525 | 1 | 0.8525 | 0 | 0 | 0 | 1 | Ref |
| 7929 | MH992509 | Unassigned | Unassigned | Unassigned | 637_0 | Clustered | 115 | VC_637_0 | 115 | 0.731 | 1 | 0.731 | 0 | 0 | 0 | 0.8764 | Ref |
| 7930 | MH992510 | Unassigned | Unassigned | Unassigned | 283_129 | Clustered/Singleton | 1 | VC_283_129 | 1 | 0 | 1 | 0 | 0 | 0 | 0 | 1 | Ref |
| 7931 | MH992513 | Unassigned | Unassigned | Unassigned | 878_0 | Clustered | 32 | VC_878_0 | 32 | 0.9965 | 1 | 0.9965 | 0 | 0 | 0 | 1 | Ref |
| 7932 | MH999279 | Unassigned | Unassigned | Unassigned | 897_0 | Clustered | 6 | VC_897_0 | 6 | 0.4156 | 1 | 0.4156 | 0 | 0 | 0 | 1 | Ref |
| 7933 | MH999280 | Unassigned | Unassigned | Unassigned | 815_0 | Clustered | 8 | VC_815_0 | 8 | 0.9801 | 1 | 0.9801 | 0 | 0 | 0 | 1 | Ref |
| 7934 | MK002701 | Unassigned | Unassigned | Unassigned | 352_0 | Clustered | 3 | VC_352_0 | 3 | 0.7306 | 0.98867275 | 0.7223 | 0 | 0 | 0 | 1 | Ref |
| 7935 | MK005300 | Unassigned | Unassigned | Unassigned | 191_20 | Clustered/Singleton | 1 | VC_191_20 | 1 | 0 | 1 | 0 | 0 | 0 | 0 | 1 | Ref |
| 7936 | MK016490 | Unassigned | Unassigned | Unassigned | 744_0 | Clustered | 10 | VC_744_0 | 10 | 0.7049 | 1 | 0.7049 | 0 | 0 | 0 | 1 | Ref |

| | | | | | | | | | | | | | | | | | |
|---|---|---|---|---|---|---|---|---|---|---|---|---|---|---|---|---|---|
| 7937 | MK016491 | Unassigned | Unassigned | Unassigned | 551_5 | Clustered | 2 | VC_551_5 | 2 | 0.0054 | 0.95548062 | 0.0052 | 0 | 0 | 0 | 1 | Ref |
| 7938 | MK016492 | Unassigned | Unassigned | Unassigned | 525_0 | Clustered | 25 | VC_525_0 | 25 | 0.4597 | 1 | 0.4597 | 0 | 0 | 0 | 1 | Ref |
| 7939 | MK016494 | Unassigned | Unassigned | Unassigned | 520_31 | Clustered | 11 | VC_520_31 | 11 | 0.0897 | 1 | 0.0897 | 0 | 0 | 0 | 0.6 | Ref |
| 7940 | MK016495 | Unassigned | Unassigned | Unassigned | 1162_0 | Clustered | 10 | VC_1162_0 | 10 | 0.5715 | 1 | 0.5715 | 0 | 0 | 0 | 1 | Ref |
| 7941 | MK016496 | Unassigned | Unassigned | Unassigned | 520_77 | Clustered | 3 | VC_520_77 | 3 | 0.0218 | 0.99859911 | 0.0218 | 0 | 0 | 0 | 1 | Ref |
| 7942 | MK016497 | Unassigned | Unassigned | Unassigned | 1162_0 | Clustered | 10 | VC_1162_0 | 10 | 0.5715 | 1 | 0.5715 | 0 | 0 | 0 | 1 | Ref |
| 7943 | MK016498 | Unassigned | Unassigned | Unassigned | 551_5 | Clustered | 2 | VC_551_5 | 2 | 0.0054 | 0.95548062 | 0.0052 | 0 | 0 | 0 | 1 | Ref |
| 7944 | MK016499 | Unassigned | Unassigned | Unassigned | 519_0 | Clustered | 39 | VC_519_0 | 39 | 0.8156 | 1 | 0.8156 | 0 | 0 | 0 | 1 | Ref |
| 7945 | MK016500 | Unassigned | Unassigned | Unassigned | 744_0 | Clustered | 10 | VC_744_0 | 10 | 0.7049 | 1 | 0.7049 | 0 | 0 | 0 | 1 | Ref |
| 7946 | MK016501 | Unassigned | Unassigned | Unassigned | 563_0 | Clustered | 2 | VC_563_0 | 2 | 0.0727 | 0.956426 | 0.0695 | 0 | 0 | 0 | 1 | Ref |
| 7947 | MK016502 | Unassigned | Unassigned | Unassigned | 551_0 | Clustered | 16 | VC_551_0 | 16 | 0.0915 | 1 | 0.0915 | 0 | 0 | 0 | 0.6667 | Ref |
| 7948 | MK016503 | Unassigned | Unassigned | Unassigned | 236_0 | Clustered | 72 | VC_236_0 | 72 | 0.8104 | 1 | 0.8104 | 0 | 0 | 0 | 1 | Ref |
| 7949 | MK016504 | Unassigned | Unassigned | Unassigned | 520_90 | Clustered/Singleton | 1 | VC_520_90 | 1 | 0 | 1 | 0 | 0 | 0 | 0 | 1 | Ref |
| 7950 | MK016662 | Unassigned | Unassigned | Unassigned | 340_0 | Clustered | 31 | VC_340_0 | 31 | 0.9549 | 1 | 0.9549 | 0 | 0 | 0 | 1 | Ref |
| 7951 | MK016664 | Unassigned | Unassigned | Unassigned | 435_1 | Clustered/Singleton | 1 | VC_435_1 | 1 | 0 | 1 | 0 | 0 | 0 | 0 | 1 | Ref |
| 7952 | MK017819 | Unassigned | Unassigned | Unassigned | 1022_0 | Clustered | 4 | VC_1022_0 | 4 | 0.8047 | 0.99996089 | 0.8046 | 0 | 0 | 0 | 1 | Ref |
| 7953 | MK017820 | Unassigned | Unassigned | Unassigned | 899_0 | Clustered | 4 | VC_899_0 | 4 | 0.2655 | 0.99997104 | 0.2655 | 0 | 0 | 0 | 1 | Ref |
| 7954 | MK024806 | Unassigned | Unassigned | Unassigned | 582_3 | Clustered | 12 | VC_582_3 | 12 | 0.2357 | 1 | 0.2357 | 0 | 0 | 0 | 0.8636 | Ref |
| 7955 | MK033136 | Unassigned | Unassigned | Unassigned | 451_0 | Clustered | 80 | VC_451_0 | 80 | 0.9991 | 1 | 0.9991 | 0 | 0 | 0 | 1 | Ref |
| 7956 | MK033137 | Unassigned | Unassigned | Unassigned | 130_0 | Clustered | 19 | VC_130_0 | 19 | 0.6068 | 1 | 0.6068 | 0 | 0 | 0 | 1 | Ref |
| 7957 | MK034952 | Unassigned | Unassigned | Unassigned | 2_0 | Clustered | 4 | VC_2_0 | 4 | 0.1311 | 0.99998692 | 0.1311 | 0 | 0 | 0 | 1 | Ref |
| 7958 | MK047717 | Unassigned | Unassigned | Unassigned | 286_15 | Clustered | 2 | VC_286_15 | 2 | 0.0073 | 0.75626308 | 0.0055 | 0 | 0 | 0 | 1 | Ref |
| 7959 | MK047718 | Unassigned | Unassigned | Unassigned | 286_17 | Clustered | 8 | VC_286_17 | 8 | 0.0535 | 0.99997829 | 0.0535 | 0 | 0 | 0 | 0.6071 | Ref |
| 7960 | MK050846 | Unassigned | Unassigned | Unassigned | 191_32 | Clustered | 12 | VC_191_32 | 12 | 0.0683 | 1 | 0.0683 | 0 | 0 | 0 | 0.7576 | Ref |
| 7961 | MK053931 | Unassigned | Unassigned | Unassigned | 878_0 | Clustered | 32 | VC_878_0 | 32 | 0.9965 | 1 | 0.9965 | 0 | 0 | 0 | 1 | Ref |
| 7962 | MK054204 | Unassigned | Unassigned | Unassigned | 599_0 | Clustered | 50 | VC_599_0 | 50 | 0.9989 | 1 | 0.9989 | 0 | 0 | 0 | 1 | Ref |
| 7963 | MK054205 | Unassigned | Unassigned | Unassigned | 599_0 | Clustered | 50 | VC_599_0 | 50 | 0.9989 | 1 | 0.9989 | 0 | 0 | 0 | 1 | Ref |
| 7964 | MK054206 | Unassigned | Unassigned | Unassigned | 599_0 | Clustered | 50 | VC_599_0 | 50 | 0.9989 | 1 | 0.9989 | 0 | 0 | 0 | 1 | Ref |
| 7965 | MK054207 | Unassigned | Unassigned | Unassigned | 599_0 | Clustered | 50 | VC_599_0 | 50 | 0.9989 | 1 | 0.9989 | 0 | 0 | 0 | 1 | Ref |
| 7966 | MK054208 | Unassigned | Unassigned | Unassigned | 599_0 | Clustered | 50 | VC_599_0 | 50 | 0.9989 | 1 | 0.9989 | 0 | 0 | 0 | 1 | Ref |
| 7967 | MK054209 | Unassigned | Unassigned | Unassigned | 599_0 | Clustered | 50 | VC_599_0 | 50 | 0.9989 | 1 | 0.9989 | 0 | 0 | 0 | 1 | Ref |
| 7968 | MK054210 | Unassigned | Unassigned | Unassigned | 599_0 | Clustered | 50 | VC_599_0 | 50 | 0.9989 | 1 | 0.9989 | 0 | 0 | 0 | 1 | Ref |
| 7969 | MK054211 | Unassigned | Unassigned | Unassigned | 599_0 | Clustered | 50 | VC_599_0 | 50 | 0.9989 | 1 | 0.9989 | 0 | 0 | 0 | 1 | Ref |
| 7970 | MK054212 | Unassigned | Unassigned | Unassigned | 599_0 | Clustered | 50 | VC_599_0 | 50 | 0.9989 | 1 | 0.9989 | 0 | 0 | 0 | 1 | Ref |
| 7971 | MK054213 | Unassigned | Unassigned | Unassigned | 599_0 | Clustered | 50 | VC_599_0 | 50 | 0.9989 | 1 | 0.9989 | 0 | 0 | 0 | 1 | Ref |
| 7972 | MK054214 | Unassigned | Unassigned | Unassigned | 599_0 | Clustered | 50 | VC_599_0 | 50 | 0.9989 | 1 | 0.9989 | 0 | 0 | 0 | 1 | Ref |
| 7973 | MK054215 | Unassigned | Unassigned | Unassigned | 599_0 | Clustered | 50 | VC_599_0 | 50 | 0.9989 | 1 | 0.9989 | 0 | 0 | 0 | 1 | Ref |
| 7974 | MK054216 | Unassigned | Unassigned | Unassigned | 599_0 | Clustered | 50 | VC_599_0 | 50 | 0.9989 | 1 | 0.9989 | 0 | 0 | 0 | 1 | Ref |
| 7975 | MK054217 | Unassigned | Unassigned | Unassigned | 599_0 | Clustered | 50 | VC_599_0 | 50 | 0.9989 | 1 | 0.9989 | 0 | 0 | 0 | 1 | Ref |
| 7976 | MK054218 | Unassigned | Unassigned | Unassigned | 599_0 | Clustered | 50 | VC_599_0 | 50 | 0.9989 | 1 | 0.9989 | 0 | 0 | 0 | 1 | Ref |
| 7977 | MK054219 | Unassigned | Unassigned | Unassigned | 599_0 | Clustered | 50 | VC_599_0 | 50 | 0.9989 | 1 | 0.9989 | 0 | 0 | 0 | 1 | Ref |
| 7978 | MK054220 | Unassigned | Unassigned | Unassigned | 599_0 | Clustered | 50 | VC_599_0 | 50 | 0.9989 | 1 | 0.9989 | 0 | 0 | 0 | 1 | Ref |
| 7979 | MK054221 | Unassigned | Unassigned | Unassigned | 599_0 | Clustered | 50 | VC_599_0 | 50 | 0.9989 | 1 | 0.9989 | 0 | 0 | 0 | 1 | Ref |
| 7980 | MK054222 | Unassigned | Unassigned | Unassigned | 599_0 | Clustered | 50 | VC_599_0 | 50 | 0.9989 | 1 | 0.9989 | 0 | 0 | 0 | 1 | Ref |
| 7981 | MK054223 | Unassigned | Unassigned | Unassigned | 599_0 | Clustered | 50 | VC_599_0 | 50 | 0.9989 | 1 | 0.9989 | 0 | 0 | 0 | 1 | Ref |
| 7982 | MK054224 | Unassigned | Unassigned | Unassigned | 599_0 | Clustered | 50 | VC_599_0 | 50 | 0.9989 | 1 | 0.9989 | 0 | 0 | 0 | 1 | Ref |
| 7983 | MK054225 | Unassigned | Unassigned | Unassigned | 599_0 | Clustered | 50 | VC_599_0 | 50 | 0.9989 | 1 | 0.9989 | 0 | 0 | 0 | 1 | Ref |
| 7984 | MK054226 | Unassigned | Unassigned | Unassigned | 599_0 | Clustered | 50 | VC_599_0 | 50 | 0.9989 | 1 | 0.9989 | 0 | 0 | 0 | 1 | Ref |
| 7985 | MK054227 | Unassigned | Unassigned | Unassigned | 599_0 | Clustered | 50 | VC_599_0 | 50 | 0.9989 | 1 | 0.9989 | 0 | 0 | 0 | 1 | Ref |
| 7986 | MK054228 | Unassigned | Unassigned | Unassigned | 599_0 | Clustered | 50 | VC_599_0 | 50 | 0.9989 | 1 | 0.9989 | 0 | 0 | 0 | 1 | Ref |
| 7987 | MK054229 | Unassigned | Unassigned | Unassigned | 599_0 | Clustered | 50 | VC_599_0 | 50 | 0.9989 | 1 | 0.9989 | 0 | 0 | 0 | 1 | Ref |
| 7988 | MK054230 | Unassigned | Unassigned | Unassigned | 599_0 | Clustered | 50 | VC_599_0 | 50 | 0.9989 | 1 | 0.9989 | 0 | 0 | 0 | 1 | Ref |
| 7989 | MK054231 | Unassigned | Unassigned | Unassigned | 599_0 | Clustered | 50 | VC_599_0 | 50 | 0.9989 | 1 | 0.9989 | 0 | 0 | 0 | 1 | Ref |
| 7990 | MK054232 | Unassigned | Unassigned | Unassigned | 599_0 | Clustered | 50 | VC_599_0 | 50 | 0.9989 | 1 | 0.9989 | 0 | 0 | 0 | 1 | Ref |
| 7991 | MK054233 | Unassigned | Unassigned | Unassigned | 599_0 | Clustered | 50 | VC_599_0 | 50 | 0.9989 | 1 | 0.9989 | 0 | 0 | 0 | 1 | Ref |
| 7992 | MK054234 | Unassigned | Unassigned | Unassigned | 599_0 | Clustered | 50 | VC_599_0 | 50 | 0.9989 | 1 | 0.9989 | 0 | 0 | 0 | 1 | Ref |
| 7993 | MK054235 | Unassigned | Unassigned | Unassigned | 599_0 | Clustered | 50 | VC_599_0 | 50 | 0.9989 | 1 | 0.9989 | 0 | 0 | 0 | 1 | Ref |
| 7994 | MK054236 | Unassigned | Unassigned | Unassigned | 599_0 | Clustered | 50 | VC_599_0 | 50 | 0.9989 | 1 | 0.9989 | 0 | 0 | 0 | 1 | Ref |
| 7995 | MK054237 | Unassigned | Unassigned | Unassigned | 599_0 | Clustered | 50 | VC_599_0 | 50 | 0.9989 | 1 | 0.9989 | 0 | 0 | 0 | 1 | Ref |
| 7996 | MK059749 | Unassigned | Unassigned | Unassigned | 372_2 | Clustered | 5 | VC_372_2 | 5 | 0.072 | 0.99999997 | 0.072 | 0 | 0 | 0 | 0.9 | Ref |
| 7997 | MK061407 | Unassigned | Unassigned | Unassigned | 939_0 | Clustered | 15 | VC_939_0 | 15 | 1 | 0 | 0 | 0 | 0 | 0 | 1 | Ref |
| 7998 | MK061408 | Unassigned | Unassigned | Unassigned | 216_4 | Clustered | 13 | VC_216_4 | 13 | 0.1657 | 1 | 0.1657 | 0 | 0 | 0 | 1 | Ref |
| 7999 | MK061409 | Unassigned | Unassigned | Unassigned | 225_0 | Clustered | 32 | VC_225_0 | 32 | 0.848 | 1 | 0.848 | 0 | 0 | 0 | 0.996 | Ref |
| 8000 | MK061410 | Unassigned | Unassigned | Unassigned | 1177_0 | Clustered | 14 | VC_1177_0 | 14 | 0.7847 | 1 | 0.7847 | 0 | 0 | 0 | 0.989 | Ref |
| 8001 | MK061411 | Unassigned | Unassigned | Unassigned | 508_0 | Clustered | 32 | VC_508_0 | 32 | 0.906 | 1 | 0.906 | 0 | 0 | 0 | 1 | Ref |
| 8002 | MK061412 | Unassigned | Unassigned | Unassigned | 1072_3 | Clustered | 6 | VC_1072_3 | 6 | 0.1304 | 0.99999994 | 0.1304 | 0 | 0 | 0 | 0.8667 | Ref |
| 8003 | MK061413 | Unassigned | Unassigned | Unassigned | 1207_0 | Clustered | 29 | VC_1207_0 | 29 | 0.8225 | 1 | 0.8225 | 0 | 0 | 0 | 1 | Ref |
| 8004 | MK061414 | Unassigned | Unassigned | Unassigned | 509_0 | Clustered | 37 | VC_509_0 | 37 | 0.9689 | 1 | 0.9689 | 0 | 0 | 0 | 1 | Ref |
| 8005 | MK061415 | Unassigned | Unassigned | Unassigned | 325_37 | Clustered | 3 | VC_325_37 | 3 | 0.008 | 0.99634936 | 0.008 | 0 | 0 | 0 | 0.6667 | Ref |
| 8006 | MK064562 | Unassigned | Unassigned | Unassigned | 1354_0 | Clustered | 3 | VC_1354_0 | 3 | 0.983 | 0.9595722 | 0.9432 | 0 | 0 | 0 | 1 | Ref |
| 8007 | MK064563 | Unassigned | Unassigned | Unassigned | 1354_0 | Clustered | 3 | VC_1354_0 | 3 | 0.983 | 0.9595722 | 0.9432 | 0 | 0 | 0 | 1 | Ref |
| 8008 | MK064564 | Unassigned | Unassigned | Unassigned | 350_0 | Clustered | 2 | VC_350_0 | 2 | 0.324 | 0.93865445 | 0.3041 | 0 | 0 | 0 | 1 | Ref |
| 8009 | MK064565 | Unassigned | Unassigned | Unassigned | 348_0 | Clustered | 22 | VC_348_0 | 22 | 0.9602 | 1 | 0.9602 | 0 | 0 | 0 | 1 | Ref |
| 8010 | MK069403 | Unassigned | Unassigned | Unassigned | 673_0 | Clustered | 15 | VC_673_0 | 15 | 0.8234 | 1 | 0.8234 | 0 | 0 | 0 | 0.7619 | Ref |
| 8011 | MK071653 | Unassigned | Unassigned | Unassigned | 1363_0 | Clustered | 2 | VC_1363_0 | 2 | 0.8326 | 0.72985431 | 0.6077 | 0 | 0 | 0 | 1 | Ref |
| 8012 | MK075001 | Unassigned | Unassigned | Unassigned | 15_1 | Clustered | 9 | VC_15_1 | 9 | 0.182 | 1 | 0.182 | 0 | 0 | 0 | 0.8889 | Ref |
| 8013 | MK075002 | Unassigned | Unassigned | Unassigned | 15_0 | Clustered | 3 | VC_15_0 | 3 | 0.0481 | 0.9985346 | 0.048 | 0 | 0 | 0 | 1 | Ref |
| 8014 | MK075003 | Unassigned | Unassigned | Unassigned | 11_0 | Clustered | 19 | VC_11_0 | 19 | 0.7562 | 1 | 0.7562 | 0 | 0 | 0 | 1 | Ref |
| 8015 | MK075004 | Unassigned | Unassigned | Unassigned | 15_1 | Clustered | 9 | VC_15_1 | 9 | 0.182 | 1 | 0.182 | 0 | 0 | 0 | 0.8889 | Ref |
| 8016 | MK075006 | Unassigned | Unassigned | Unassigned | 11_0 | Clustered | 19 | VC_11_0 | 19 | 0.7562 | 1 | 0.7562 | 0 | 0 | 0 | 1 | Ref |
| 8017 | MK080312 | Unassigned | Unassigned | Unassigned | 1218_0 | Clustered | 4 | VC_1218_0 | 4 | 0.8905 | 0.99530591 | 0.8863 | 0 | 0 | 0 | 1 | Ref |
| 8018 | MK084630 | Unassigned | Unassigned | Unassigned | 818_0 | Clustered | 20 | VC_818_0 | 20 | 0.9601 | 1 | 0.9601 | 0 | 0 | 0 | 1 | Ref |
| 8019 | MK085976 | Unassigned | Unassigned | Unassigned | 704_0 | Clustered | 16 | VC_704_0 | 16 | 0.7274 | 1 | 0.7274 | 0 | 0 | 0 | 1 | Ref |
| 8020 | MK088078 | Unassigned | Unassigned | Unassigned | 185_0 | Clustered | 3 | VC_185_0 | 3 | 0.4487 | 0.97148006 | 0.4359 | 0 | 0 | 0 | 1 | Ref |
| 8021 | MK089780 | Unassigned | Unassigned | Unassigned | 699_0 | Clustered | 71 | VC_699_0 | 71 | 0.9352 | 1 | 0.9352 | 0 | 0 | 0 | 1 | Ref |
| 8022 | MK095192 | Unassigned | Unassigned | Unassigned | 984_0 | Clustered | 38 | VC_984_0 | 38 | 0.9758 | 1 | 0.9758 | 0 | 0 | 0 | 1 | Ref |
| 8023 | MK095193 | Unassigned | Unassigned | Unassigned | 984_0 | Clustered | 38 | VC_984_0 | 38 | 0.9758 | 1 | 0.9758 | 0 | 0 | 0 | 1 | Ref |
| 8024 | MK095194 | Unassigned | Unassigned | Unassigned | 984_0 | Clustered | 38 | VC_984_0 | 38 | 0.9758 | 1 | 0.9758 | 0 | 0 | 0 | 1 | Ref |
| 8025 | MK095195 | Unassigned | Unassigned | Unassigned | 984_0 | Clustered | 38 | VC_984_0 | 38 | 0.9758 | 1 | 0.9758 | 0 | 0 | 0 | 1 | Ref |
| 8026 | MK095196 | Unassigned | Unassigned | Unassigned | 984_0 | Clustered | 38 | VC_984_0 | 38 | 0.9758 | 1 | 0.9758 | 0 | 0 | 0 | 1 | Ref |
| 8027 | MK095197 | Unassigned | Unassigned | Unassigned | 984_0 | Clustered | 38 | VC_984_0 | 38 | 0.9758 | 1 | 0.9758 | 0 | 0 | 0 | 1 | Ref |
| 8028 | MK095198 | Unassigned | Unassigned | Unassigned | 984_0 | Clustered | 38 | VC_984_0 | 38 | 0.9758 | 1 | 0.9758 | 0 | 0 | 0 | 1 | Ref |
| 8029 | MK095199 | Unassigned | Unassigned | Unassigned | 984_0 | Clustered | 38 | VC_984_0 | 38 | 0.9758 | 1 | 0.9758 | 0 | 0 | 0 | 1 | Ref |
| 8030 | MK095200 | Unassigned | Unassigned | Unassigned | 984_0 | Clustered | 38 | VC_984_0 | 38 | 0.9758 | 1 | 0.9758 | 0 | 0 | 0 | 1 | Ref |
| 8031 | MK095201 | Unassigned | Unassigned | Unassigned | 984_0 | Clustered | 38 | VC_984_0 | 38 | 0.9758 | 1 | 0.9758 | 0 | 0 | 0 | 1 | Ref |
| 8032 | MK095202 | Unassigned | Unassigned | Unassigned | 984_0 | Clustered | 38 | VC_984_0 | 38 | 0.9758 | 1 | 0.9758 | 0 | 0 | 0 | 1 | Ref |
| 8033 | MK095203 | Unassigned | Unassigned | Unassigned | 984_0 | Clustered | 38 | VC_984_0 | 38 | 0.9758 | 1 | 0.9758 | 0 | 0 | 0 | 1 | Ref |
| 8034 | MK095204 | Unassigned | Unassigned | Unassigned | 984_0 | Clustered | 38 | VC_984_0 | 38 | 0.9758 | 1 | 0.9758 | 0 | 0 | 0 | 1 | Ref |
| 8035 | MK095205 | Unassigned | Unassigned | Unassigned | 984_0 | Clustered | 38 | VC_984_0 | 38 | 0.9758 | 1 | 0.9758 | 0 | 0 | 0 | 1 | Ref |
| 8036 | MK095206 | Unassigned | Unassigned | Unassigned | 984_0 | Clustered | 38 | VC_984_0 | 38 | 0.9758 | 1 | 0.9758 | 0 | 0 | 0 | 1 | Ref |
| 8037 | MK095207 | Unassigned | Unassigned | Unassigned | 984_0 | Clustered | 38 | VC_984_0 | 38 | 0.9758 | 1 | 0.9758 | 0 | 0 | 0 | 1 | Ref |
| 8038 | MK095208 | Unassigned | Unassigned | Unassigned | 984_0 | Clustered | 38 | VC_984_0 | 38 | 0.9758 | 1 | 0.9758 | 0 | 0 | 0 | 1 | Ref |
| 8039 | MK095209 | Unassigned | Unassigned | Unassigned | 984_0 | Clustered | 38 | VC_984_0 | 38 | 0.9758 | 1 | 0.9758 | 0 | 0 | 0 | 1 | Ref |
| 8040 | MK095210 | Unassigned | Unassigned | Unassigned | 984_0 | Clustered | 38 | VC_984_0 | 38 | 0.9758 | 1 | 0.9758 | 0 | 0 | 0 | 1 | Ref |
| 8041 | MK095211 | Unassigned | Unassigned | Unassigned | 984_0 | Clustered | 38 | VC_984_0 | 38 | 0.9758 | 1 | 0.9758 | 0 | 0 | 0 | 1 | Ref |
| 8042 | MK097141 | Unassigned | Unassigned | Unassigned | 642_2 | Clustered | 6 | VC_642_2 | 6 | 0.1679 | 1 | 0.1679 | 0 | 0 | 0 | 1 | Ref |
| 8043 | MK105819 | Unassigned | Unassigned | Unassigned | 356_0 | Clustered | 2 | VC_356_0 | 2 | 0.1992 | 0.93211095 | 0.1857 | 0 | 0 | 0 | 1 | Ref |
| 8044 | MK105855 | Unassigned | Unassigned | Unassigned | 705_0 | Clustered | 11 | VC_705_0 | 11 | 0.8861 | 1 | 0.8861 | 0 | 0 | 0 | 1 | Ref |
| 8045 | MK112526 | Unassigned | Unassigned | Unassigned | 1160_0 | Clustered | 73 | VC_1160_0 | 73 | 0.9835 | 1 | 0.9835 | 0 | 0 | 0 | 1 | Ref |
| 8046 | MK112527 | Unassigned | Unassigned | Unassigned | 371_15 | Clustered | 44 | VC_371_15 | 44 | 0.1391 | 1 | 0.1391 | 0 | 0 | 0 | 0.7474 | Ref |
| 8047 | MK112528 | Unassigned | Unassigned | Unassigned | 1160_0 | Clustered | 73 | VC_1160_0 | 73 | 0.9835 | 1 | 0.9835 | 0 | 0 | 0 | 1 | Ref |
| 8048 | MK112529 | Unassigned | Unassigned | Unassigned | 1160_0 | Clustered | 73 | VC_1160_0 | 73 | 0.9835 | 1 | 0.9835 | 0 | 0 | 0 | 1 | Ref |
| 8049 | MK112530 | Unassigned | Unassigned | Unassigned | 325_41 | Clustered | 2 | VC_325_41 | 2 | 0.0036 | 0.94071129 | 0.0034 | 0 | 0 | 0 | 1 | Ref |
| 8050 | MK112531 | Unassigned | Unassigned | Unassigned | 1160_0 | Clustered | 73 | VC_1160_0 | 73 | 0.9835 | 1 | 0.9835 | 0 | 0 | 0 | 1 | Ref |
| 8051 | MK112532 | Unassigned | Unassigned | Unassigned | 325_33 | Clustered | 4 | VC_325_33 | 4 | 0.0149 | 0.99998565 | 0.0149 | 0 | 0 | 0 | 1 | Ref |
| 8052 | MK112533 | Unassigned | Unassigned | Unassigned | 558_51 | Clustered/Singleton | 1 | VC_558_51 | 1 | 0 | 1 | 0 | 0 | 0 | 0 | 1 | Ref |
| 8053 | MK112534 | Unassigned | Unassigned | Unassigned | 558_10 | Clustered/Singleton | 1 | VC_558_10 | 1 | 0 | 1 | 0 | 0 | 0 | 0 | 1 | Ref |
| 8054 | MK112535 | Unassigned | Unassigned | Unassigned | 1160_0 | Clustered | 73 | VC_1160_0 | 73 | 0.9835 | 1 | 0.9835 | 0 | 0 | 0 | 1 | Ref |
| 8055 | MK112536 | Unassigned | Unassigned | Unassigned | 371_15 | Clustered | 44 | VC_371_15 | 44 | 0.1391 | 1 | 0.1391 | 0 | 0 | 0 | 0.7474 | Ref |
| 8056 | MK112537 | Unassigned | Unassigned | Unassigned | 236_0 | Clustered | 72 | VC_236_0 | 72 | 0.8104 | 1 | 0.8104 | 0 | 0 | 0 | 1 | Ref |
| 8057 | MK112538 | Unassigned | Unassigned | Unassigned | 1160_0 | Clustered | 73 | VC_1160_0 | 73 | 0.9835 | 1 | 0.9835 | 0 | 0 | 0 | 1 | Ref |
| 8058 | MK112539 | Unassigned | Unassigned | Unassigned | 371_11 | Clustered | 31 | VC_371_11 | 31 | 0.0927 | 1 | 0.0927 | 0 | 0 | 0 | 0.7011 | Ref |
| 8059 | MK112540 | Unassigned | Unassigned | Unassigned | 325_48 | Clustered | 2 | VC_325_48 | 2 | 0.0039 | 0.95008941 | 0.0037 | 0 | 0 | 0 | 1 | Ref |
| 8060 | MK112541 | Unassigned | Unassigned | Unassigned | 558_47 | Clustered | 3 | VC_558_47 | 3 | 0.0064 | 0.80043943 | 0.0051 | 0 | 0 | 0 | 1 | Ref |
| 8061 | MK112542 | Unassigned | Unassigned | Unassigned | 371_16 | Clustered | 42 | VC_371_16 | 42 | 0.1341 | 1 | 0.1341 | 0 | 0 | 0 | 0.7282 | Ref |
| 8062 | MK112543 | Unassigned | Unassigned | Unassigned | 514_0 | Clustered | 75 | VC_514_0 | 75 | 0.9402 | 1 | 0.9402 | 0 | 0 | 0 | 1 | Ref |
| 8063 | MK112544 | Unassigned | Unassigned | Unassigned | 371_12 | Clustered | 4 | VC_371_12 | 4 | 0.0126 | 0.99998489 | 0.0126 | 0 | 0 | 0 | 0.8333 | Ref |
| 8064 | MK112545 | Unassigned | Unassigned | Unassigned | 558_47 | Clustered | 3 | VC_558_47 | 3 | 0.0064 | 0.80043943 | 0.0051 | 0 | 0 | 0 | 1 | Ref |
| 8065 | MK112546 | Unassigned | Unassigned | Unassigned | 371_11 | Clustered | 31 | VC_371_11 | 31 | 0.0927 | 1 | 0.0927 | 0 | 0 | 0 | 0.7011 | Ref |
| 8066 | MK112547 | Unassigned | Unassigned | Unassigned | 525_0 | Clustered | 25 | VC_525_0 | 25 | 0.4597 | 1 | 0.4597 | 0 | 0 | 0 | 1 | Ref |
| 8067 | MK112548 | Unassigned | Unassigned | Unassigned | 558_47 | Clustered | 3 | VC_558_47 | 3 | 0.0064 | 0.80043943 | 0.0051 | 0 | 0 | 0 | 1 | Ref |
| 8068 | MK112549 | Unassigned | Unassigned | Unassigned | 558_34 | Clustered/Singleton | 1 | VC_558_34 | 1 | 0 | 1 | 0 | 0 | 0 | 0 | 1 | Ref |
| 8069 | MK112550 | Unassigned | Unassigned | Unassigned | 1224_0 | Clustered | 19 | VC_1224_0 | 19 | 1 | 1 | 1 | 0 | 0 | 0 | 1 | Ref |
| 8070 | MK112551 | Unassigned | Unassigned | Unassigned | 371_11 | Clustered | 31 | VC_371_11 | 31 | 0.0927 | 1 | 0.0927 | 0 | 0 | 0 | 0.7011 | Ref |
| 8071 | MK112552 | Unassigned | Unassigned | Unassigned | 371_2 | Clustered | 8 | VC_371_2 | 8 | 0.0232 | 1 | 0.0232 | 0 | 0 | 0 | 0.8571 | Ref |
| 8072 | MK112553 | Unassigned | Unassigned | Unassigned | 371_16 | Clustered | 42 | VC_371_16 | 42 | 0.1341 | 1 | 0.1341 | 0 | 0 | 0 | 0.7282 | Ref |
| 8073 | MK112554 | Unassigned | Unassigned | Unassigned | 558_49 | Clustered | 7 | VC_558_49 | 7 | 0.0194 | 0.99510369 | 0.0193 | 0 | 0 | 0 | 0.8571 | Ref |
| 8074 | MK112555 | Unassigned | Unassigned | Unassigned | 371_15 | Clustered | 44 | VC_371_15 | 44 | 0.1391 | 1 | 0.1391 | 0 | 0 | 0 | 0.7474 | Ref |
| 8075 | MK112556 | Unassigned | Unassigned | Unassigned | 520_74 | Clustered | 2 | VC_520_74 | 2 | 0.0075 | 0.95760586 | 0.0072 | 0 | 0 | 0 | 1 | Ref |
| 8076 | MK112901 | Unassigned | Unassigned | Unassigned | 461_0 | Clustered | 96 | VC_461_0 | 96 | 0.9307 | 1 | 0.9307 | 0 | 0 | 0 | 0.9575 | Ref |
| 8077 | MK125140 | Unassigned | Unassigned | Unassigned | 695_0 | Clustered | 48 | VC_695_0 | 48 | 0.8162 | 1 | 0.8162 | 0 | 0 | 0 | 1 | Ref |
| 8078 | MK125141 | Unassigned | Unassigned | Unassigned | 705_0 | Clustered | 11 | VC_705_0 | 11 | 0.8861 | 1 | 0.8861 | 0 | 0 | 0 | 1 | Ref |
| 8079 | MK134560 | Unassigned | Unassigned | Unassigned | 823_0 | Clustered | 124 | VC_823_0 | 124 | 0.9492 | 1 | 0.9492 | 0 | 0 | 0 | 1 | Ref |
| 8080 | MK135468 | Unassigned | Unassigned | Unassigned | 111_0 | Clustered | 91 | VC_111_0 | 91 | 0.9587 | 1 | 0.9587 | 0 | 0 | 0 | 1 | Ref |
| 8081 | MK138526 | Unassigned | Unassigned | Unassigned | 76_0 | Clustered | 37 | VC_76_0 | 37 | 0.9403 | 1 | 0.9403 | 0 | 0 | 0 | 1 | Ref |
| 8082 | MK138527 | Unassigned | Unassigned | Unassigned | 835_0 | Clustered | 67 | VC_835_0 | 67 | 0.9624 | 1 | 0.9624 | 0 | 0 | 0 | 1 | Ref |
| 8083 | MK144667 | Unassigned | Unassigned | Unassigned | 179_0 | Clustered | 53 | VC_179_0 | 53 | 0.9137 | 1 | 0.9137 | 0 | 0 | 0 | 1 | Ref |
| 8084 | MK165087 | Unassigned | Unassigned | Unassigned | 722_0 | Clustered | 13 | VC_722_0 | 13 | 0.5104 | 1 | 0.5104 | 0 | 0 | 0 | 1 | Ref |
| 8085 | MK165657 | Unassigned | Unassigned | Unassigned | 749_0 | Clustered | 40 | VC_749_0 | 40 | 0.7517 | 1 | 0.7517 | 0 | 0 | 0 | 1 | Ref |
| 8086 | MK165658 | Unassigned | Unassigned | Unassigned | 749_0 | Clustered | 40 | VC_749_0 | 40 | 0.7517 | 1 | 0.7517 | 0 | 0 | 0 | 1 | Ref |
| 8087 | MK170160 | Unassigned | Unassigned | Unassigned | 700_0 | Clustered | 10 | VC_700_0 | 10 | 0.8076 | 1 | 0.8076 | 0 | 0 | 0 | 1 | Ref |
| 8088 | MK170446 | Unassigned | Unassigned | Unassigned | 938_0 | Clustered | 99 | VC_938_0 | 99 | 0.9119 | 1 | 0.9119 | 0 | 0 | 0 | 0.9928 | Ref |
| 8089 | MK202159 | Unassigned | Unassigned | Unassigned | 255_0 | Clustered | 13 | VC_255_0 | 13 | 0.2048 | 1 | 0.2048 | 0 | 0 | 0 | 1 | Ref |
| 8090 | MK202160 | Unassigned | Unassigned | Unassigned | 255_0 | Clustered | 13 | VC_255_0 | 13 | 0.2048 | 1 | 0.2048 | 0 | 0 | 0 | 1 | Ref |
| 8091 | MK202161 | Unassigned | Unassigned | Unassigned | 255_0 | Clustered | 13 | VC_255_0 | 13 | 0.2048 | 1 | 0.2048 | 0 | 0 | 0 | 1 | Ref |
| 8092 | MK203841 | Unassigned | Unassigned | Unassigned | 823_0 | Clustered | 124 | VC_823_0 | 124 | 0.9492 | 1 | 0.9492 | 0 | 0 | 0 | 1 | Ref |
| 8093 | MK211557 | Unassigned | Unassigned | Unassigned | 12_2 | Clustered | 19 | VC_12_2 | 19 | 0.2018 | 1 | 0.2018 | 0 | 0 | 0 | 1 | Ref |
| 8094 | MK213795 | Unassigned | Unassigned | Unassigned | 315_0 | Clustered | 12 | VC_315_0 | 12 | 0.8947 | 1 | 0.8947 | 0 | 0 | 0 | 1 | Ref |
| 8095 | MK213796 | Unassigned | Unassigned | Unassigned | 575_0 | Clustered | 35 | VC_575_0 | 35 | 0.8815 | 1 | 0.8815 | 0 | 0 | 0 | 1 | Ref |
| 8096 | MK214385 | Unassigned | Unassigned | Unassigned | 540_0 | Clustered | 15 | VC_540_0 | 15 | 0.2866 | 1 | 0.2866 | 0 | 0 | 0 | 1 | Ref |
| 8097 | MK215646 | Unassigned | Unassigned | Unassigned | 883_0 | Clustered | 10 | VC_883_0 | 10 | 0.2015 | 1 | 0.2015 | 0 | 0 | 0 | 1 | Ref |
| 8098 | MK224497 | Unassigned | Unassigned | Unassigned | 234_0 | Clustered | 21 | VC_234_0 | 21 | 0.8355 | 1 | 0.8355 | 0 | 0 | 0 | 1 | Ref |

| | | | | | | | | | | | | | | | | | |
|---|---|---|---|---|---|---|---|---|---|---|---|---|---|---|---|---|---|
| 8099 | MK224498 | Unassigned | Unassigned | Unassigned | 76_0 | Clustered | 37 | VC_76_0 | 37 | 0.9403 | 1 | 0.9403 | 0 | 0 | 0 | 1 | Ref |
| 8100 | MK224592 | Unassigned | Unassigned | Unassigned | 520_95 | Clustered/Singleton | 1 | VC_520_95 | 1 | 0 | 1 | 0 | 0 | 0 | 0 | 1 | Ref |
| 8101 | MK234886 | Unassigned | Unassigned | Unassigned | 290_5 | Clustered | 14 | VC_290_5 | 14 | 0.1422 | 1 | 0.1422 | 0 | 0 | 0 | 0.9231 | Ref |
| 8102 | MK238400 | Unassigned | Unassigned | Unassigned | 673_0 | Clustered | 15 | VC_673_0 | 15 | 0.8234 | 1 | 0.8234 | 0 | 0 | 0 | 0.7619 | Ref |
| 8103 | MK240351 | Unassigned | Unassigned | Unassigned | 492_0 | Clustered | 2 | VC_492_0 | 2 | 0.0158 | 0.6640397 | 0.0105 | 0 | 0 | 0 | 1 | Ref |
| 8104 | MK240575 | Unassigned | Unassigned | Unassigned | 1351_0 | Clustered | 2 | VC_1351_0 | 2 | 1 | 1 | 1 | 0 | 0 | 0 | 1 | Ref |
| 8105 | MK249126 | Unassigned | Unassigned | Unassigned | 637_0 | Clustered | 115 | VC_637_0 | 115 | 0.731 | 1 | 0.731 | 0 | 0 | 0 | 0.8764 | Ref |
| 8106 | MK249127 | Unassigned | Unassigned | Unassigned | 1066_0 | Clustered | 52 | VC_1066_0 | 52 | 0.7682 | 1 | 0.7682 | 0 | 0 | 0 | 1 | Ref |
| 8107 | MK249128 | Unassigned | Unassigned | Unassigned | 1066_0 | Clustered | 52 | VC_1066_0 | 52 | 0.7682 | 1 | 0.7682 | 0 | 0 | 0 | 1 | Ref |
| 8108 | MK249129 | Unassigned | Unassigned | Unassigned | 1066_0 | Clustered | 52 | VC_1066_0 | 52 | 0.7682 | 1 | 0.7682 | 0 | 0 | 0 | 1 | Ref |
| 8109 | MK249131 | Unassigned | Unassigned | Unassigned | 1000_0 | Clustered | 21 | VC_1000_0 | 21 | 0.8437 | 0.99999998 | 0.8437 | 0 | 0 | 0 | 1 | Ref |
| 8110 | MK249132 | Unassigned | Unassigned | Unassigned | 1067_0 | Clustered | 2 | VC_1067_0 | 2 | 0.012 | 0.85807772 | 0.0103 | 0 | 0 | 0 | 1 | Ref |
| 8111 | MK249133 | Unassigned | Unassigned | Unassigned | 1062_0 | Clustered | 45 | VC_1062_0 | 45 | 0.9734 | 0.50949847 | 0.4959 | 0 | 0 | 0 | 1 | Ref |
| 8112 | MK249135 | Unassigned | Unassigned | Unassigned | 994_0 | Clustered/Singleton | 1 | VC_994_0 | 1 | 0 | 1 | 0 | 0 | 0 | 0 | 1 | Ref |
| 8113 | MK249138 | Unassigned | Unassigned | Unassigned | 1062_0 | Clustered | 45 | VC_1062_0 | 45 | 0.9734 | 0.50949847 | 0.4959 | 0 | 0 | 0 | 1 | Ref |
| 8114 | MK249139 | Unassigned | Unassigned | Unassigned | 1066_0 | Clustered | 52 | VC_1066_0 | 52 | 0.7682 | 1 | 0.7682 | 0 | 0 | 0 | 1 | Ref |
| 8115 | MK249141 | Unassigned | Unassigned | Unassigned | 995_0 | Clustered | 69 | VC_995_0 | 69 | 0.8547 | 1 | 0.8547 | 0 | 0 | 0 | 1 | Ref |
| 8116 | MK249143 | Unassigned | Unassigned | Unassigned | 398_0 | Clustered | 48 | VC_398_0 | 48 | 0.4813 | 1 | 0.4813 | 0 | 0 | 0 | 1 | Ref |
| 8117 | MK249144 | Unassigned | Unassigned | Unassigned | 1000_0 | Clustered | 21 | VC_1000_0 | 21 | 0.8437 | 0.99999998 | 0.8437 | 0 | 0 | 0 | 1 | Ref |
| 8118 | MK249147 | Unassigned | Unassigned | Unassigned | 995_0 | Clustered | 69 | VC_995_0 | 69 | 0.8547 | 1 | 0.8547 | 0 | 0 | 0 | 1 | Ref |
| 8119 | MK249148 | Unassigned | Unassigned | Unassigned | 1264_0 | Clustered | 6 | VC_1264_0 | 6 | 0.7062 | 0.92055725 | 0.6501 | 0 | 0 | 0 | 1 | Ref |
| 8120 | MK249149 | Unassigned | Unassigned | Unassigned | 1066_0 | Clustered | 52 | VC_1066_0 | 52 | 0.7682 | 1 | 0.7682 | 0 | 0 | 0 | 1 | Ref |
| 8121 | MK249150 | Unassigned | Unassigned | Unassigned | 1066_0 | Clustered | 52 | VC_1066_0 | 52 | 0.7682 | 1 | 0.7682 | 0 | 0 | 0 | 1 | Ref |
| 8122 | MK249151 | Unassigned | Unassigned | Unassigned | 398_0 | Clustered | 48 | VC_398_0 | 48 | 0.4813 | 1 | 0.4813 | 0 | 0 | 0 | 1 | Ref |
| 8123 | MK249153 | Unassigned | Unassigned | Unassigned | 1066_0 | Clustered | 52 | VC_1066_0 | 52 | 0.7682 | 1 | 0.7682 | 0 | 0 | 0 | 1 | Ref |
| 8124 | MK249154 | Unassigned | Unassigned | Unassigned | 1066_0 | Clustered | 52 | VC_1066_0 | 52 | 0.7682 | 1 | 0.7682 | 0 | 0 | 0 | 1 | Ref |
| 8125 | MK249155 | Unassigned | Unassigned | Unassigned | 1066_0 | Clustered | 52 | VC_1066_0 | 52 | 0.7682 | 1 | 0.7682 | 0 | 0 | 0 | 1 | Ref |
| 8126 | MK249156 | Unassigned | Unassigned | Unassigned | 1000_0 | Clustered | 21 | VC_1000_0 | 21 | 0.8437 | 0.99999998 | 0.8437 | 0 | 0 | 0 | 1 | Ref |
| 8127 | MK249158 | Unassigned | Unassigned | Unassigned | 1066_0 | Clustered | 52 | VC_1066_0 | 52 | 0.7682 | 1 | 0.7682 | 0 | 0 | 0 | 1 | Ref |
| 8128 | MK249159 | Unassigned | Unassigned | Unassigned | 1000_0 | Clustered | 21 | VC_1000_0 | 21 | 0.8437 | 0.99999998 | 0.8437 | 0 | 0 | 0 | 1 | Ref |
| 8129 | MK249160 | Unassigned | Unassigned | Unassigned | 1000_0 | Clustered | 21 | VC_1000_0 | 21 | 0.8437 | 0.99999998 | 0.8437 | 0 | 0 | 0 | 1 | Ref |
| 8130 | MK249162 | Unassigned | Unassigned | Unassigned | 1066_0 | Clustered | 52 | VC_1066_0 | 52 | 0.7682 | 1 | 0.7682 | 0 | 0 | 0 | 1 | Ref |
| 8131 | MK249163 | Unassigned | Unassigned | Unassigned | 1066_0 | Clustered | 52 | VC_1066_0 | 52 | 0.7682 | 1 | 0.7682 | 0 | 0 | 0 | 1 | Ref |
| 8132 | MK249165 | Unassigned | Unassigned | Unassigned | 995_0 | Clustered | 69 | VC_995_0 | 69 | 0.8547 | 1 | 0.8547 | 0 | 0 | 0 | 1 | Ref |
| 8133 | MK249166 | Unassigned | Unassigned | Unassigned | 1066_0 | Clustered | 52 | VC_1066_0 | 52 | 0.7682 | 1 | 0.7682 | 0 | 0 | 0 | 1 | Ref |
| 8134 | MK249167 | Unassigned | Unassigned | Unassigned | 1000_0 | Clustered | 21 | VC_1000_0 | 21 | 0.8437 | 0.99999998 | 0.8437 | 0 | 0 | 0 | 1 | Ref |
| 8135 | MK249168 | Unassigned | Unassigned | Unassigned | 1066_0 | Clustered | 52 | VC_1066_0 | 52 | 0.7682 | 1 | 0.7682 | 0 | 0 | 0 | 1 | Ref |
| 8136 | MK249174 | Unassigned | Unassigned | Unassigned | 1066_0 | Clustered | 52 | VC_1066_0 | 52 | 0.7682 | 1 | 0.7682 | 0 | 0 | 0 | 1 | Ref |
| 8137 | MK249176 | Unassigned | Unassigned | Unassigned | 1066_0 | Clustered | 52 | VC_1066_0 | 52 | 0.7682 | 1 | 0.7682 | 0 | 0 | 0 | 1 | Ref |
| 8138 | MK249178 | Unassigned | Unassigned | Unassigned | 1066_0 | Clustered | 52 | VC_1066_0 | 52 | 0.7682 | 1 | 0.7682 | 0 | 0 | 0 | 1 | Ref |
| 8139 | MK249179 | Unassigned | Unassigned | Unassigned | 1000_0 | Clustered | 21 | VC_1000_0 | 21 | 0.8437 | 0.99999998 | 0.8437 | 0 | 0 | 0 | 1 | Ref |
| 8140 | MK249180 | Unassigned | Unassigned | Unassigned | 1066_0 | Clustered | 52 | VC_1066_0 | 52 | 0.7682 | 1 | 0.7682 | 0 | 0 | 0 | 1 | Ref |
| 8141 | MK249181 | Unassigned | Unassigned | Unassigned | 1000_0 | Clustered | 21 | VC_1000_0 | 21 | 0.8437 | 0.99999998 | 0.8437 | 0 | 0 | 0 | 1 | Ref |
| 8142 | MK249185 | Unassigned | Unassigned | Unassigned | 995_0 | Clustered | 69 | VC_995_0 | 69 | 0.8547 | 1 | 0.8547 | 0 | 0 | 0 | 1 | Ref |
| 8143 | MK249188 | Unassigned | Unassigned | Unassigned | 1066_0 | Clustered | 52 | VC_1066_0 | 52 | 0.7682 | 1 | 0.7682 | 0 | 0 | 0 | 1 | Ref |
| 8144 | MK249189 | Unassigned | Unassigned | Unassigned | 999_0 | Clustered | 2 | VC_999_0 | 2 | 0.0273 | 0.73989325 | 0.0202 | 0 | 0 | 0 | 1 | Ref |
| 8145 | MK249190 | Unassigned | Unassigned | Unassigned | 995_0 | Clustered | 69 | VC_995_0 | 69 | 0.8547 | 1 | 0.8547 | 0 | 0 | 0 | 1 | Ref |
| 8146 | MK249191 | Unassigned | Unassigned | Unassigned | 1062_0 | Clustered | 45 | VC_1062_0 | 45 | 0.9734 | 0.50949847 | 0.4959 | 0 | 0 | 0 | 1 | Ref |
| 8147 | MK249192 | Unassigned | Unassigned | Unassigned | 1066_0 | Clustered | 52 | VC_1066_0 | 52 | 0.7682 | 1 | 0.7682 | 0 | 0 | 0 | 1 | Ref |
| 8148 | MK249194 | Unassigned | Unassigned | Unassigned | 1066_0 | Clustered | 52 | VC_1066_0 | 52 | 0.7682 | 1 | 0.7682 | 0 | 0 | 0 | 1 | Ref |
| 8149 | MK249195 | Unassigned | Unassigned | Unassigned | 1066_0 | Clustered | 52 | VC_1066_0 | 52 | 0.7682 | 1 | 0.7682 | 0 | 0 | 0 | 1 | Ref |
| 8150 | MK249196 | Unassigned | Unassigned | Unassigned | 1066_0 | Clustered | 52 | VC_1066_0 | 52 | 0.7682 | 1 | 0.7682 | 0 | 0 | 0 | 1 | Ref |
| 8151 | MK249197 | Unassigned | Unassigned | Unassigned | 995_0 | Clustered | 69 | VC_995_0 | 69 | 0.8547 | 1 | 0.8547 | 0 | 0 | 0 | 1 | Ref |
| 8152 | MK249198 | Unassigned | Unassigned | Unassigned | 1066_0 | Clustered | 52 | VC_1066_0 | 52 | 0.7682 | 1 | 0.7682 | 0 | 0 | 0 | 1 | Ref |
| 8153 | MK249199 | Unassigned | Unassigned | Unassigned | 1062_0 | Clustered | 45 | VC_1062_0 | 45 | 0.9734 | 0.50949847 | 0.4959 | 0 | 0 | 0 | 1 | Ref |
| 8154 | MK249201 | Unassigned | Unassigned | Unassigned | 995_0 | Clustered | 69 | VC_995_0 | 69 | 0.8547 | 1 | 0.8547 | 0 | 0 | 0 | 1 | Ref |
| 8155 | MK249202 | Unassigned | Unassigned | Unassigned | 995_0 | Clustered | 69 | VC_995_0 | 69 | 0.8547 | 1 | 0.8547 | 0 | 0 | 0 | 1 | Ref |
| 8156 | MK249203 | Unassigned | Unassigned | Unassigned | 995_0 | Clustered | 69 | VC_995_0 | 69 | 0.8547 | 1 | 0.8547 | 0 | 0 | 0 | 1 | Ref |
| 8157 | MK249204 | Unassigned | Unassigned | Unassigned | 1066_0 | Clustered | 52 | VC_1066_0 | 52 | 0.7682 | 1 | 0.7682 | 0 | 0 | 0 | 1 | Ref |
| 8158 | MK249205 | Unassigned | Unassigned | Unassigned | 1067_0 | Clustered | 2 | VC_1067_0 | 2 | 0.012 | 0.85807772 | 0.0103 | 0 | 0 | 0 | 1 | Ref |
| 8159 | MK249207 | Unassigned | Unassigned | Unassigned | 1062_0 | Clustered | 45 | VC_1062_0 | 45 | 0.9734 | 0.50949847 | 0.4959 | 0 | 0 | 0 | 1 | Ref |
| 8160 | MK249208 | Unassigned | Unassigned | Unassigned | 1062_0 | Clustered | 45 | VC_1062_0 | 45 | 0.9734 | 0.50949847 | 0.4959 | 0 | 0 | 0 | 1 | Ref |
| 8161 | MK249211 | Unassigned | Unassigned | Unassigned | 995_0 | Clustered | 69 | VC_995_0 | 69 | 0.8547 | 1 | 0.8547 | 0 | 0 | 0 | 1 | Ref |
| 8162 | MK249212 | Unassigned | Unassigned | Unassigned | 1066_0 | Clustered | 52 | VC_1066_0 | 52 | 0.7682 | 1 | 0.7682 | 0 | 0 | 0 | 1 | Ref |
| 8163 | MK249213 | Unassigned | Unassigned | Unassigned | 995_0 | Clustered | 69 | VC_995_0 | 69 | 0.8547 | 1 | 0.8547 | 0 | 0 | 0 | 1 | Ref |
| 8164 | MK249214 | Unassigned | Unassigned | Unassigned | 1066_0 | Clustered | 52 | VC_1066_0 | 52 | 0.7682 | 1 | 0.7682 | 0 | 0 | 0 | 1 | Ref |
| 8165 | MK249215 | Unassigned | Unassigned | Unassigned | 1087_0 | Clustered | 5 | VC_1087_0 | 5 | 0.8109 | 0.98759579 | 0.8008 | 0 | 0 | 0 | 1 | Ref |
| 8166 | MK249216 | Unassigned | Unassigned | Unassigned | 1364_0 | Clustered | 3 | VC_1364_0 | 3 | 1 | 1 | 1 | 0 | 0 | 0 | 1 | Ref |
| 8167 | MK249217 | Unassigned | Unassigned | Unassigned | 1364_0 | Clustered | 3 | VC_1364_0 | 3 | 1 | 1 | 1 | 0 | 0 | 0 | 1 | Ref |
| 8168 | MK249218 | Unassigned | Unassigned | Unassigned | 1339_0 | Clustered | 7 | VC_1339_0 | 7 | 0.9177 | 0.99772305 | 0.9156 | 0 | 0 | 0 | 1 | Ref |
| 8169 | MK249219 | Unassigned | Unassigned | Unassigned | 1066_0 | Clustered | 52 | VC_1066_0 | 52 | 0.7682 | 1 | 0.7682 | 0 | 0 | 0 | 1 | Ref |
| 8170 | MK249221 | Unassigned | Unassigned | Unassigned | 995_0 | Clustered | 69 | VC_995_0 | 69 | 0.8547 | 1 | 0.8547 | 0 | 0 | 0 | 1 | Ref |
| 8171 | MK249222 | Unassigned | Unassigned | Unassigned | 1068_0 | Clustered | 5 | VC_1068_0 | 5 | 1 | 1 | 1 | 0 | 0 | 0 | 1 | Ref |
| 8172 | MK249223 | Unassigned | Unassigned | Unassigned | 398_0 | Clustered | 48 | VC_398_0 | 48 | 0.4813 | 1 | 0.4813 | 0 | 0 | 0 | 1 | Ref |
| 8173 | MK249228 | Unassigned | Unassigned | Unassigned | 1062_0 | Clustered | 45 | VC_1062_0 | 45 | 0.9734 | 0.50949847 | 0.4959 | 0 | 0 | 0 | 1 | Ref |
| 8174 | MK249229 | Unassigned | Unassigned | Unassigned | 1339_0 | Clustered | 7 | VC_1339_0 | 7 | 0.9177 | 0.99772305 | 0.9156 | 0 | 0 | 0 | 1 | Ref |
| 8175 | MK249231 | Unassigned | Unassigned | Unassigned | 398_0 | Clustered | 48 | VC_398_0 | 48 | 0.4813 | 1 | 0.4813 | 0 | 0 | 0 | 1 | Ref |
| 8176 | MK249232 | Unassigned | Unassigned | Unassigned | 1062_0 | Clustered | 45 | VC_1062_0 | 45 | 0.9734 | 0.50949847 | 0.4959 | 0 | 0 | 0 | 1 | Ref |
| 8177 | MK249233 | Unassigned | Unassigned | Unassigned | 1339_0 | Clustered | 7 | VC_1339_0 | 7 | 0.9177 | 0.99772305 | 0.9156 | 0 | 0 | 0 | 1 | Ref |
| 8178 | MK249308 | Unassigned | Unassigned | Unassigned | 592_0 | Clustered | 50 | VC_592_0 | 50 | 0.7314 | 1 | 0.7314 | 0 | 0 | 0 | 1 | Ref |
| 8179 | MK249872 | Unassigned | Unassigned | Unassigned | 643_0 | Clustered | 2 | VC_643_0 | 2 | 0.1092 | 0.95085347 | 0.1038 | 0 | 0 | 0 | 1 | Ref |
| 8180 | MK250015 | Unassigned | Unassigned | Unassigned | 1210_2 | Clustered | 4 | VC_1210_2 | 4 | 0.1183 | 0.92062211 | 0.109 | 0 | 0 | 0 | 1 | Ref |
| 8181 | MK250016 | Unassigned | Unassigned | Unassigned | 1210_2 | Clustered | 4 | VC_1210_2 | 4 | 0.1183 | 0.92062211 | 0.109 | 0 | 0 | 0 | 1 | Ref |
| 8182 | MK250017 | Unassigned | Unassigned | Unassigned | 1210_2 | Clustered | 4 | VC_1210_2 | 4 | 0.1183 | 0.92062211 | 0.109 | 0 | 0 | 0 | 1 | Ref |
| 8183 | MK250018 | Unassigned | Unassigned | Unassigned | 1210_2 | Clustered | 4 | VC_1210_2 | 4 | 0.1183 | 0.92062211 | 0.109 | 0 | 0 | 0 | 1 | Ref |
| 8184 | MK250019 | Unassigned | Unassigned | Unassigned | 1210_3 | Clustered | 2 | VC_1210_3 | 2 | 0.0366 | 0.68120968 | 0.0249 | 0 | 0 | 0 | 1 | Ref |
| 8185 | MK250020 | Unassigned | Unassigned | Unassigned | 1210_1 | Clustered | 7 | VC_1210_1 | 7 | 0.2693 | 0.99874141 | 0.2689 | 0 | 0 | 0 | 1 | Ref |
| 8186 | MK250021 | Unassigned | Unassigned | Unassigned | 1210_0 | Clustered | 2 | VC_1210_0 | 2 | 0.0367 | 0.68120968 | 0.025 | 0 | 0 | 0 | 1 | Ref |
| 8187 | MK250022 | Unassigned | Unassigned | Unassigned | 1210_1 | Clustered | 7 | VC_1210_1 | 7 | 0.2693 | 0.99874141 | 0.2689 | 0 | 0 | 0 | 1 | Ref |
| 8188 | MK250023 | Unassigned | Unassigned | Unassigned | 1210_0 | Clustered | 2 | VC_1210_0 | 2 | 0.0367 | 0.68120968 | 0.025 | 0 | 0 | 0 | 1 | Ref |
| 8189 | MK250024 | Unassigned | Unassigned | Unassigned | 1210_1 | Clustered | 7 | VC_1210_1 | 7 | 0.2693 | 0.99874141 | 0.2689 | 0 | 0 | 0 | 1 | Ref |
| 8190 | MK250025 | Unassigned | Unassigned | Unassigned | 1210_1 | Clustered | 7 | VC_1210_1 | 7 | 0.2693 | 0.99874141 | 0.2689 | 0 | 0 | 0 | 1 | Ref |
| 8191 | MK250026 | Unassigned | Unassigned | Unassigned | 1210_1 | Clustered | 7 | VC_1210_1 | 7 | 0.2693 | 0.99874141 | 0.2689 | 0 | 0 | 0 | 1 | Ref |
| 8192 | MK250027 | Unassigned | Unassigned | Unassigned | 1210_1 | Clustered | 7 | VC_1210_1 | 7 | 0.2693 | 0.99874141 | 0.2689 | 0 | 0 | 0 | 1 | Ref |
| 8193 | MK250028 | Unassigned | Unassigned | Unassigned | 1210_1 | Clustered | 7 | VC_1210_1 | 7 | 0.2693 | 0.99874141 | 0.2689 | 0 | 0 | 0 | 1 | Ref |
| 8194 | MK250029 | Unassigned | Unassigned | Unassigned | 1210_3 | Clustered | 2 | VC_1210_3 | 2 | 0.0366 | 0.68120968 | 0.0249 | 0 | 0 | 0 | 1 | Ref |
| 8195 | MK250904 | Unassigned | Unassigned | Unassigned | 149_15 | Clustered | 16 | VC_149_15 | 16 | 0.0788 | 1 | 0.0788 | 0 | 0 | 0 | 0.8083 | Ref |
| 8196 | MK257719 | Unassigned | Unassigned | Unassigned | 699_0 | Clustered | 71 | VC_699_0 | 71 | 0.9352 | 1 | 0.9352 | 0 | 0 | 0 | 1 | Ref |
| 8197 | MK257720 | Unassigned | Unassigned | Unassigned | 699_0 | Clustered | 71 | VC_699_0 | 71 | 0.9352 | 1 | 0.9352 | 0 | 0 | 0 | 1 | Ref |
| 8198 | MK257721 | Unassigned | Unassigned | Unassigned | 699_0 | Clustered | 71 | VC_699_0 | 71 | 0.9352 | 1 | 0.9352 | 0 | 0 | 0 | 1 | Ref |
| 8199 | MK257722 | Unassigned | Unassigned | Unassigned | 699_0 | Clustered | 71 | VC_699_0 | 71 | 0.9352 | 1 | 0.9352 | 0 | 0 | 0 | 1 | Ref |
| 8200 | MK257723 | Unassigned | Unassigned | Unassigned | 699_0 | Clustered | 71 | VC_699_0 | 71 | 0.9352 | 1 | 0.9352 | 0 | 0 | 0 | 1 | Ref |
| 8201 | MK257744 | Unassigned | Unassigned | Unassigned | 477_0 | Clustered | 4 | VC_477_0 | 4 | 0.9836 | 0.99748973 | 0.9811 | 0 | 0 | 0 | 1 | Ref |
| 8202 | MK268344 | Unassigned | Unassigned | Unassigned | 164_0 | Clustered | 3 | VC_164_0 | 3 | 0.0298 | 0.82983726 | 0.0247 | 0 | 0 | 0 | 1 | Ref |
| 8203 | MK268686 | Unassigned | Unassigned | Unassigned | 91_0 | Clustered | 22 | VC_91_0 | 22 | 0.5318 | 1 | 0.5318 | 0 | 0 | 0 | 1 | Ref |
| 8204 | MK278859 | Unassigned | Unassigned | Unassigned | 699_0 | Clustered | 71 | VC_699_0 | 71 | 0.9352 | 1 | 0.9352 | 0 | 0 | 0 | 1 | Ref |
| 8205 | MK278860 | Unassigned | Unassigned | Unassigned | 492_2 | Clustered | 8 | VC_492_2 | 8 | 0.1221 | 0.98786574 | 0.1206 | 0 | 0 | 0 | 1 | Ref |
| 8206 | MK278861 | Unassigned | Unassigned | Unassigned | 938_0 | Clustered | 99 | VC_938_0 | 99 | 0.9119 | 1 | 0.9119 | 0 | 0 | 0 | 0.9928 | Ref |
| 8207 | MK279840 | Unassigned | Unassigned | Unassigned | 574_0 | Clustered | 7 | VC_574_0 | 7 | 0.2734 | 1 | 0.2734 | 0 | 0 | 0 | 1 | Ref |
| 8208 | MK279841 | Unassigned | Unassigned | Unassigned | 728_0 | Clustered | 3 | VC_728_0 | 3 | 0.4017 | 0.99850259 | 0.4011 | 0 | 0 | 0 | 1 | Ref |
| 8209 | MK279842 | Unassigned | Unassigned | Unassigned | 939_0 | Clustered | 15 | VC_939_0 | 15 | 1 | 0 | 0 | 0 | 0 | 0 | 1 | Ref |
| 8210 | MK279843 | Unassigned | Unassigned | Unassigned | 371_11 | Clustered | 31 | VC_371_11 | 31 | 0.0927 | 1 | 0.0927 | 0 | 0 | 0 | 0.7011 | Ref |
| 8211 | MK279844 | Unassigned | Unassigned | Unassigned | 1160_0 | Clustered | 73 | VC_1160_0 | 73 | 0.9835 | 1 | 0.9835 | 0 | 0 | 0 | 1 | Ref |
| 8212 | MK279845 | Unassigned | Unassigned | Unassigned | 1160_0 | Clustered | 73 | VC_1160_0 | 73 | 0.9835 | 1 | 0.9835 | 0 | 0 | 0 | 1 | Ref |
| 8213 | MK279846 | Unassigned | Unassigned | Unassigned | 371_16 | Clustered | 42 | VC_371_16 | 42 | 0.1341 | 1 | 0.1341 | 0 | 0 | 0 | 0.7282 | Ref |
| 8214 | MK279847 | Unassigned | Unassigned | Unassigned | 1160_0 | Clustered | 73 | VC_1160_0 | 73 | 0.9835 | 1 | 0.9835 | 0 | 0 | 0 | 1 | Ref |
| 8215 | MK279849 | Unassigned | Unassigned | Unassigned | 546_7 | Clustered | 4 | VC_546_7 | 4 | 0.041 | 0.9963251 | 0.0408 | 0 | 0 | 0 | 1 | Ref |
| 8216 | MK279850 | Unassigned | Unassigned | Unassigned | 371_13 | Clustered | 33 | VC_371_13 | 33 | 0.179 | 1 | 0.179 | 0 | 0 | 0 | 0.8769 | Ref |
| 8217 | MK279851 | Unassigned | Unassigned | Unassigned | 1160_0 | Clustered | 73 | VC_1160_0 | 73 | 0.9835 | 1 | 0.9835 | 0 | 0 | 0 | 1 | Ref |
| 8218 | MK279852 | Unassigned | Unassigned | Unassigned | 371_16 | Clustered | 42 | VC_371_16 | 42 | 0.1341 | 1 | 0.1341 | 0 | 0 | 0 | 0.7282 | Ref |
| 8219 | MK279853 | Unassigned | Unassigned | Unassigned | 1096_0 | Clustered | 5 | VC_1096_0 | 5 | 0.8566 | 0.99869977 | 0.8555 | 0 | 0 | 0 | 1 | Ref |
| 8220 | MK279854 | Unassigned | Unassigned | Unassigned | 1160_0 | Clustered | 73 | VC_1160_0 | 73 | 0.9835 | 1 | 0.9835 | 0 | 0 | 0 | 1 | Ref |
| 8221 | MK279855 | Unassigned | Unassigned | Unassigned | 371_15 | Clustered | 44 | VC_371_15 | 44 | 0.1391 | 1 | 0.1391 | 0 | 0 | 0 | 0.7474 | Ref |
| 8222 | MK279856 | Unassigned | Unassigned | Unassigned | 1160_0 | Clustered | 73 | VC_1160_0 | 73 | 0.9835 | 1 | 0.9835 | 0 | 0 | 0 | 1 | Ref |
| 8223 | MK279857 | Unassigned | Unassigned | Unassigned | 217_0 | Clustered | 5 | VC_217_0 | 5 | 0.069 | 0.99999998 | 0.069 | 0 | 0 | 0 | 1 | Ref |
| 8224 | MK279858 | Unassigned | Unassigned | Unassigned | 371_13 | Clustered | 33 | VC_371_13 | 33 | 0.179 | 1 | 0.179 | 0 | 0 | 0 | 0.8769 | Ref |
| 8225 | MK279859 | Unassigned | Unassigned | Unassigned | 371_15 | Clustered | 44 | VC_371_15 | 44 | 0.1391 | 1 | 0.1391 | 0 | 0 | 0 | 0.7474 | Ref |
| 8226 | MK279860 | Unassigned | Unassigned | Unassigned | 1160_0 | Clustered | 73 | VC_1160_0 | 73 | 0.9835 | 1 | 0.9835 | 0 | 0 | 0 | 1 | Ref |
| 8227 | MK279861 | Unassigned | Unassigned | Unassigned | 371_15 | Clustered | 44 | VC_371_15 | 44 | 0.1391 | 1 | 0.1391 | 0 | 0 | 0 | 0.7474 | Ref |
| 8228 | MK279862 | Unassigned | Unassigned | Unassigned | 1160_0 | Clustered | 73 | VC_1160_0 | 73 | 0.9835 | 1 | 0.9835 | 0 | 0 | 0 | 1 | Ref |
| 8229 | MK279863 | Unassigned | Unassigned | Unassigned | 1160_0 | Clustered | 73 | VC_1160_0 | 73 | 0.9835 | 1 | 0.9835 | 0 | 0 | 0 | 1 | Ref |
| 8230 | MK279864 | Unassigned | Unassigned | Unassigned | 371_13 | Clustered | 33 | VC_371_13 | 33 | 0.179 | 1 | 0.179 | 0 | 0 | 0 | 0.8769 | Ref |
| 8231 | MK279865 | Unassigned | Unassigned | Unassigned | 371_11 | Clustered | 31 | VC_371_11 | 31 | 0.0927 | 1 | 0.0927 | 0 | 0 | 0 | 0.7011 | Ref |
| 8232 | MK279866 | Unassigned | Unassigned | Unassigned | 371_15 | Clustered | 44 | VC_371_15 | 44 | 0.1391 | 1 | 0.1391 | 0 | 0 | 0 | 0.7474 | Ref |
| 8233 | MK279867 | Unassigned | Unassigned | Unassigned | 1160_0 | Clustered | 73 | VC_1160_0 | 73 | 0.9835 | 1 | 0.9835 | 0 | 0 | 0 | 1 | Ref |
| 8234 | MK279868 | Unassigned | Unassigned | Unassigned | 1160_0 | Clustered | 73 | VC_1160_0 | 73 | 0.9835 | 1 | 0.9835 | 0 | 0 | 0 | 1 | Ref |
| 8235 | MK279869 | Unassigned | Unassigned | Unassigned | 1160_0 | Clustered | 73 | VC_1160_0 | 73 | 0.9835 | 1 | 0.9835 | 0 | 0 | 0 | 1 | Ref |
| 8236 | MK279870 | Unassigned | Unassigned | Unassigned | 371_11 | Clustered | 31 | VC_371_11 | 31 | 0.0927 | 1 | 0.0927 | 0 | 0 | 0 | 0.7011 | Ref |
| 8237 | MK279871 | Unassigned | Unassigned | Unassigned | 371_16 | Clustered | 42 | VC_371_16 | 42 | 0.1341 | 1 | 0.1341 | 0 | 0 | 0 | 0.7282 | Ref |
| 8238 | MK279872 | Unassigned | Unassigned | Unassigned | 1160_0 | Clustered | 73 | VC_1160_0 | 73 | 0.9835 | 1 | 0.9835 | 0 | 0 | 0 | 1 | Ref |
| 8239 | MK279873 | Unassigned | Unassigned | Unassigned | 371_11 | Clustered | 31 | VC_371_11 | 31 | 0.0927 | 1 | 0.0927 | 0 | 0 | 0 | 0.7011 | Ref |
| 8240 | MK279874 | Unassigned | Unassigned | Unassigned | 1160_0 | Clustered | 73 | VC_1160_0 | 73 | 0.9835 | 1 | 0.9835 | 0 | 0 | 0 | 1 | Ref |
| 8241 | MK279875 | Unassigned | Unassigned | Unassigned | 1160_0 | Clustered | 73 | VC_1160_0 | 73 | 0.9835 | 1 | 0.9835 | 0 | 0 | 0 | 1 | Ref |
| 8242 | MK279876 | Unassigned | Unassigned | Unassigned | 1160_0 | Clustered | 73 | VC_1160_0 | 73 | 0.9835 | 1 | 0.9835 | 0 | 0 | 0 | 1 | Ref |
| 8243 | MK279877 | Unassigned | Unassigned | Unassigned | 371_3 | Clustered | 22 | VC_371_3 | 22 | 0.0787 | 1 | 0.0787 | 0 | 0 | 0 | 0.9048 | Ref |
| 8244 | MK279878 | Unassigned | Unassigned | Unassigned | 371_11 | Clustered | 31 | VC_371_11 | 31 | 0.0927 | 1 | 0.0927 | 0 | 0 | 0 | 0.7011 | Ref |
| 8245 | MK279879 | Unassigned | Unassigned | Unassigned | 371_15 | Clustered | 44 | VC_371_15 | 44 | 0.1391 | 1 | 0.1391 | 0 | 0 | 0 | 0.7474 | Ref |
| 8246 | MK279880 | Unassigned | Unassigned | Unassigned | 1160_0 | Clustered | 73 | VC_1160_0 | 73 | 0.9835 | 1 | 0.9835 | 0 | 0 | 0 | 1 | Ref |
| 8247 | MK279881 | Unassigned | Unassigned | Unassigned | 371_15 | Clustered | 44 | VC_371_15 | 44 | 0.1391 | 1 | 0.1391 | 0 | 0 | 0 | 0.7474 | Ref |
| 8248 | MK279882 | Unassigned | Unassigned | Unassigned | 371_15 | Clustered | 44 | VC_371_15 | 44 | 0.1391 | 1 | 0.1391 | 0 | 0 | 0 | 0.7474 | Ref |
| 8249 | MK279883 | Unassigned | Unassigned | Unassigned | 371_13 | Clustered | 33 | VC_371_13 | 33 | 0.179 | 1 | 0.179 | 0 | 0 | 0 | 0.8769 | Ref |
| 8250 | MK279884 | Unassigned | Unassigned | Unassigned | 1160_0 | Clustered | 73 | VC_1160_0 | 73 | 0.9835 | 1 | 0.9835 | 0 | 0 | 0 | 1 | Ref |
| 8251 | MK279885 | Unassigned | Unassigned | Unassigned | 371_15 | Clustered | 44 | VC_371_15 | 44 | 0.1391 | 1 | 0.1391 | 0 | 0 | 0 | 0.7474 | Ref |
| 8252 | MK279886 | Unassigned | Unassigned | Unassigned | 1160_0 | Clustered | 73 | VC_1160_0 | 73 | 0.9835 | 1 | 0.9835 | 0 | 0 | 0 | 1 | Ref |
| 8253 | MK279887 | Unassigned | Unassigned | Unassigned | 371_15 | Clustered | 44 | VC_371_15 | 44 | 0.1391 | 1 | 0.1391 | 0 | 0 | 0 | 0.7474 | Ref |
| 8254 | MK279888 | Unassigned | Unassigned | Unassigned | 371_15 | Clustered | 44 | VC_371_15 | 44 | 0.1391 | 1 | 0.1391 | 0 | 0 | 0 | 0.7474 | Ref |
| 8255 | MK279889 | Unassigned | Unassigned | Unassigned | 371_2 | Clustered | 8 | VC_371_2 | 8 | 0.0232 | 1 | 0.0232 | 0 | 0 | 0 | 0.8571 | Ref |
| 8256 | MK279890 | Unassigned | Unassigned | Unassigned | 371_13 | Clustered | 33 | VC_371_13 | 33 | 0.179 | 1 | 0.179 | 0 | 0 | 0 | 0.8769 | Ref |
| 8257 | MK279891 | Unassigned | Unassigned | Unassigned | 371_13 | Clustered | 33 | VC_371_13 | 33 | 0.179 | 1 | 0.179 | 0 | 0 | 0 | 0.8769 | Ref |
| 8258 | MK279892 | Unassigned | Unassigned | Unassigned | 1160_0 | Clustered | 73 | VC_1160_0 | 73 | 0.9835 | 1 | 0.9835 | 0 | 0 | 0 | 1 | Ref |
| 8259 | MK279893 | Unassigned | Unassigned | Unassigned | 954_0 | Clustered | 13 | VC_954_0 | 13 | 0.863 | 1 | 0.863 | 0 | 0 | 0 | 1 | Ref |
| 8260 | MK279894 | Unassigned | Unassigned | Unassigned | 371_11 | Clustered | 31 | VC_371_11 | 31 | 0.0927 | 1 | 0.0927 | 0 | 0 | 0 | 0.7011 | Ref |

| | | | | | | | | | | | | | | | | | |
|---|---|---|---|---|---|---|---|---|---|---|---|---|---|---|---|---|---|
| 8261 | MK279895 | Unassigned | Unassigned | Unassigned | 371_11 | Clustered | 31 | VC_371_11 | 31 | 0.0927 | 1 | 0.0927 | 0 | 0 | 0 | 0.7011 | Ref |
| 8262 | MK279896 | Unassigned | Unassigned | Unassigned | 1160_0 | Clustered | 73 | VC_1160_0 | 73 | 0.9835 | 1 | 0.9835 | 0 | 0 | 0 | 1 | Ref |
| 8263 | MK279897 | Unassigned | Unassigned | Unassigned | 371_11 | Clustered | 31 | VC_371_11 | 31 | 0.0927 | 1 | 0.0927 | 0 | 0 | 0 | 0.7011 | Ref |
| 8264 | MK279898 | Unassigned | Unassigned | Unassigned | 517_0 | Clustered | 18 | VC_517_0 | 18 | 0.1599 | 1 | 0.1599 | 0 | 0 | 0 | 1 | Ref |
| 8265 | MK279900 | Unassigned | Unassigned | Unassigned | 919_0 | Clustered | 39 | VC_919_0 | 39 | 0.9069 | 1 | 0.9069 | 0 | 0 | 0 | 1 | Ref |
| 8266 | MK279901 | Unassigned | Unassigned | Unassigned | 1160_0 | Clustered | 73 | VC_1160_0 | 73 | 0.9835 | 1 | 0.9835 | 0 | 0 | 0 | 1 | Ref |
| 8267 | MK279902 | Unassigned | Unassigned | Unassigned | 517_1 | Clustered | 108 | VC_517_1 | 108 | 0.8483 | 1 | 0.8483 | 0 | 0 | 0 | 0.9955 | Ref |
| 8268 | MK279903 | Unassigned | Unassigned | Unassigned | 514_0 | Clustered | 75 | VC_514_0 | 75 | 0.9402 | 1 | 0.9402 | 0 | 0 | 0 | 1 | Ref |
| 8269 | MK279904 | Unassigned | Unassigned | Unassigned | 371_4 | Clustered | 5 | VC_371_4 | 5 | 0.0135 | 0.99999971 | 0.0135 | 0 | 0 | 0 | 1 | Ref |
| 8270 | MK279905 | Unassigned | Unassigned | Unassigned | 887_0 | Clustered | 18 | VC_887_0 | 18 | 0.3663 | 1 | 0.3663 | 0 | 0 | 0 | 1 | Ref |
| 8271 | MK279906 | Unassigned | Unassigned | Unassigned | 557_1 | Clustered | 11 | VC_557_1 | 11 | 0.4945 | 1 | 0.4945 | 0 | 0 | 0 | 0.7818 | Ref |
| 8272 | MK279907 | Unassigned | Unassigned | Unassigned | 1160_0 | Clustered | 73 | VC_1160_0 | 73 | 0.9835 | 1 | 0.9835 | 0 | 0 | 0 | 1 | Ref |
| 8273 | MK279908 | Unassigned | Unassigned | Unassigned | 371_3 | Clustered | 22 | VC_371_3 | 22 | 0.0787 | 1 | 0.0787 | 0 | 0 | 0 | 0.9048 | Ref |
| 8274 | MK279909 | Unassigned | Unassigned | Unassigned | 566_0 | Clustered | 2 | VC_566_0 | 2 | 0.0989 | 0.95622526 | 0.0945 | 0 | 0 | 0 | 1 | Ref |
| 8275 | MK279910 | Unassigned | Unassigned | Unassigned | 371_1 | Clustered | 11 | VC_371_1 | 11 | 0.0379 | 1 | 0.0379 | 0 | 0 | 0 | 0.8545 | Ref |
| 8276 | MK279911 | Unassigned | Unassigned | Unassigned | 1175_0 | Clustered | 22 | VC_1175_0 | 22 | 0.4562 | 1 | 0.4562 | 0 | 0 | 0 | 1 | Ref |
| 8277 | MK279912 | Unassigned | Unassigned | Unassigned | 1175_0 | Clustered | 22 | VC_1175_0 | 22 | 0.4562 | 1 | 0.4562 | 0 | 0 | 0 | 1 | Ref |
| 8278 | MK279913 | Unassigned | Unassigned | Unassigned | 514_0 | Clustered | 75 | VC_514_0 | 75 | 0.9402 | 1 | 0.9402 | 0 | 0 | 0 | 1 | Ref |
| 8279 | MK284518 | Unassigned | Unassigned | Unassigned | 514_0 | Clustered | 75 | VC_514_0 | 75 | 0.9402 | 1 | 0.9402 | 0 | 0 | 0 | 1 | Ref |
| 8280 | MK284519 | Unassigned | Unassigned | Unassigned | 513_0 | Clustered | 32 | VC_513_0 | 32 | 0.6881 | 1 | 0.6881 | 0 | 0 | 0 | 1 | Ref |
| 8281 | MK284520 | Unassigned | Unassigned | Unassigned | 685_0 | Clustered | 16 | VC_685_0 | 16 | 0.2972 | 1 | 0.2972 | 0 | 0 | 0 | 0.9167 | Ref |
| 8282 | MK284521 | Unassigned | Unassigned | Unassigned | 1177_0 | Clustered | 14 | VC_1177_0 | 14 | 0.7847 | 1 | 0.7847 | 0 | 0 | 0 | 0.989 | Ref |
| 8283 | MK284522 | Unassigned | Unassigned | Unassigned | 216_4 | Clustered | 13 | VC_216_4 | 13 | 0.1657 | 1 | 0.1657 | 0 | 0 | 0 | 1 | Ref |
| 8284 | MK284526 | Unassigned | Unassigned | Unassigned | 818_0 | Clustered | 20 | VC_818_0 | 20 | 0.9601 | 1 | 0.9601 | 0 | 0 | 0 | 1 | Ref |
| 8285 | MK284527 | Unassigned | Unassigned | Unassigned | 818_0 | Clustered | 20 | VC_818_0 | 20 | 0.9601 | 1 | 0.9601 | 0 | 0 | 0 | 1 | Ref |
| 8286 | MK284528 | Unassigned | Unassigned | Unassigned | 818_0 | Clustered | 20 | VC_818_0 | 20 | 0.9601 | 1 | 0.9601 | 0 | 0 | 0 | 1 | Ref |
| 8287 | MK286578 | Unassigned | Unassigned | Unassigned | 592_0 | Clustered | 50 | VC_592_0 | 50 | 0.7314 | 1 | 0.7314 | 0 | 0 | 0 | 1 | Ref |
| 8288 | MK288021 | Unassigned | Unassigned | Unassigned | 475_8 | Clustered | 2 | VC_475_8 | 2 | 0.0291 | 0.90184699 | 0.0262 | 0 | 0 | 0 | 1 | Ref |
| 8289 | MK288022 | Unassigned | Unassigned | Unassigned | 1006_0 | Clustered | 8 | VC_1006_0 | 8 | 0.7407 | 1 | 0.7407 | 0 | 0 | 0 | 1 | Ref |
| 8290 | MK290737 | Unassigned | Unassigned | Unassigned | 331_0 | Clustered | 4 | VC_331_0 | 4 | 0.0884 | 0.99998775 | 0.0884 | 0 | 0 | 0 | 1 | Ref |
| 8291 | MK290738 | Unassigned | Unassigned | Unassigned | 331_0 | Clustered | 4 | VC_331_0 | 4 | 0.0884 | 0.99998775 | 0.0884 | 0 | 0 | 0 | 1 | Ref |
| 8292 | MK290764 | Unassigned | Unassigned | Unassigned | 12_0 | Clustered | 21 | VC_12_0 | 21 | 0.2274 | 1 | 0.2274 | 0 | 0 | 0 | 0.9571 | Ref |
| 8293 | MK291441 | Unassigned | Unassigned | Unassigned | 922_0 | Clustered | 2 | VC_922_0 | 2 | 0.42 | 0.90478487 | 0.38 | 0 | 0 | 0 | 1 | Ref |
| 8294 | MK291443 | Unassigned | Unassigned | Unassigned | 922_0 | Clustered | 2 | VC_922_0 | 2 | 0.42 | 0.90478487 | 0.38 | 0 | 0 | 0 | 1 | Ref |
| 8295 | MK295203 | Unassigned | Unassigned | Unassigned | 290_7 | Clustered/Singleton | 1 | VC_290_7 | 1 | 0 | 1 | 0 | 0 | 0 | 0 | 1 | Ref |
| 8296 | MK295204 | Unassigned | Unassigned | Unassigned | 288_2 | Clustered | 2 | VC_288_2 | 2 | 0.0228 | 0.95538559 | 0.0218 | 0 | 0 | 0 | 1 | Ref |
| 8297 | MK301437 | Unassigned | Unassigned | Unassigned | 202_0 | Clustered | 102 | VC_202_0 | 102 | 0.4049 | 1 | 0.4049 | 0 | 0 | 0 | 0.7876 | Ref |
| 8298 | MK301442 | Unassigned | Unassigned | Unassigned | 690_0 | Clustered | 50 | VC_690_0 | 50 | 0.9805 | 1 | 0.9805 | 0 | 0 | 0 | 1 | Ref |
| 8299 | MK301443 | Unassigned | Unassigned | Unassigned | 690_0 | Clustered | 50 | VC_690_0 | 50 | 0.9805 | 1 | 0.9805 | 0 | 0 | 0 | 1 | Ref |
| 8300 | MK301532 | Unassigned | Unassigned | Unassigned | 451_0 | Clustered | 80 | VC_451_0 | 80 | 0.9991 | 1 | 0.9991 | 0 | 0 | 0 | 1 | Ref |
| 8301 | MK305884 | Unassigned | Unassigned | Unassigned | 761_0 | Clustered | 67 | VC_761_0 | 67 | 0.8947 | 1 | 0.8947 | 0 | 0 | 0 | 1 | Ref |
| 8302 | MK305885 | Unassigned | Unassigned | Unassigned | 954_0 | Clustered | 13 | VC_954_0 | 13 | 0.863 | 1 | 0.863 | 0 | 0 | 0 | 1 | Ref |
| 8303 | MK305887 | Unassigned | Unassigned | Unassigned | 940_0 | Clustered | 34 | VC_940_0 | 34 | 0.7959 | 1 | 0.7959 | 0 | 0 | 0 | 1 | Ref |
| 8304 | MK305888 | Unassigned | Unassigned | Unassigned | 727_0 | Clustered | 11 | VC_727_0 | 11 | 0.5054 | 1 | 0.5054 | 0 | 0 | 0 | 1 | Ref |
| 8305 | MK305889 | Unassigned | Unassigned | Unassigned | 1178_0 | Clustered | 24 | VC_1178_0 | 24 | 0.9524 | 1 | 0.9524 | 0 | 0 | 0 | 1 | Ref |
| 8306 | MK305890 | Unassigned | Unassigned | Unassigned | 1369_0 | Clustered | 3 | VC_1369_0 | 3 | 0.9809 | 0.9595722 | 0.9412 | 0 | 0 | 0 | 1 | Ref |
| 8307 | MK305891 | Unassigned | Unassigned | Unassigned | 859_0 | Clustered | 9 | VC_859_0 | 9 | 0.9426 | 1 | 0.9426 | 0 | 0 | 0 | 1 | Ref |
| 8308 | MK305892 | Unassigned | Unassigned | Unassigned | 939_0 | Clustered | 15 | VC_939_0 | 15 | 1 | 0 | 0 | 0 | 0 | 0 | 1 | Ref |
| 8309 | MK305893 | Unassigned | Unassigned | Unassigned | 325_55 | Clustered | 2 | VC_325_55 | 2 | 0.0061 | 0.95750995 | 0.0058 | 0 | 0 | 0 | 1 | Ref |
| 8310 | MK308637 | Unassigned | Unassigned | Unassigned | 1163_0 | Clustered | 28 | VC_1163_0 | 28 | 0.4923 | 1 | 0.4923 | 0 | 0 | 0 | 1 | Ref |
| 8311 | MK308638 | Unassigned | Unassigned | Unassigned | 1360_0 | Clustered | 31 | VC_1360_0 | 31 | 0.8837 | 1 | 0.8837 | 0 | 0 | 0 | 1 | Ref |
| 8312 | MK308674 | Unassigned | Unassigned | Unassigned | 346_0 | Clustered | 4 | VC_346_0 | 4 | 0.4606 | 0.99997905 | 0.4606 | 0 | 0 | 0 | 1 | Ref |
| 8313 | MK308675 | Unassigned | Unassigned | Unassigned | 346_0 | Clustered | 4 | VC_346_0 | 4 | 0.4606 | 0.99997905 | 0.4606 | 0 | 0 | 0 | 1 | Ref |
| 8314 | MK308676 | Unassigned | Unassigned | Unassigned | 346_0 | Clustered | 4 | VC_346_0 | 4 | 0.4606 | 0.99997905 | 0.4606 | 0 | 0 | 0 | 1 | Ref |
| 8315 | MK308677 | Unassigned | Unassigned | Unassigned | 346_0 | Clustered | 4 | VC_346_0 | 4 | 0.4606 | 0.99997905 | 0.4606 | 0 | 0 | 0 | 1 | Ref |
| 8316 | MK310138 | Unassigned | Unassigned | Unassigned | 237_0 | Clustered | 4 | VC_237_0 | 4 | 0.0429 | 0.99998759 | 0.0429 | 0 | 0 | 0 | 1 | Ref |
| 8317 | MK310139 | Unassigned | Unassigned | Unassigned | 371_5 | Clustered | 2 | VC_371_5 | 2 | 0.0035 | 0.95771071 | 0.0033 | 0 | 0 | 0 | 1 | Ref |
| 8318 | MK310140 | Unassigned | Unassigned | Unassigned | 514_0 | Clustered | 75 | VC_514_0 | 75 | 0.9402 | 1 | 0.9402 | 0 | 0 | 0 | 1 | Ref |
| 8319 | MK310141 | Unassigned | Unassigned | Unassigned | 325_16 | Clustered | 2 | VC_325_16 | 2 | 0.0095 | 0.95751393 | 0.0091 | 0 | 0 | 0 | 1 | Ref |
| 8320 | MK310142 | Unassigned | Unassigned | Unassigned | 325_61 | Clustered | 8 | VC_325_61 | 8 | 0.0395 | 1 | 0.0395 | 0 | 0 | 0 | 0.7857 | Ref |
| 8321 | MK310143 | Unassigned | Unassigned | Unassigned | 558_32 | Clustered/Singleton | 1 | VC_558_32 | 1 | 0 | 1 | 0 | 0 | 0 | 0 | 1 | Ref |
| 8322 | MK310144 | Unassigned | Unassigned | Unassigned | 887_0 | Clustered | 18 | VC_887_0 | 18 | 0.3663 | 1 | 0.3663 | 0 | 0 | 0 | 1 | Ref |
| 8323 | MK310145 | Unassigned | Unassigned | Unassigned | 517_1 | Clustered | 108 | VC_517_1 | 108 | 0.8483 | 1 | 0.8483 | 0 | 0 | 0 | 0.9955 | Ref |
| 8324 | MK310146 | Unassigned | Unassigned | Unassigned | 761_0 | Clustered | 67 | VC_761_0 | 67 | 0.8947 | 1 | 0.8947 | 0 | 0 | 0 | 1 | Ref |
| 8325 | MK310182 | Unassigned | Unassigned | Unassigned | 392_0 | Clustered | 126 | VC_392_0 | 126 | 0.9621 | 1 | 0.9621 | 0 | 0 | 0 | 1 | Ref |
| 8326 | MK310183 | Unassigned | Unassigned | Unassigned | 813_0 | Clustered | 12 | VC_813_0 | 12 | 0.2347 | 1 | 0.2347 | 0 | 0 | 0 | 1 | Ref |
| 8327 | MK310184 | Unassigned | Unassigned | Unassigned | 290_6 | Clustered | 12 | VC_290_6 | 12 | 0.1149 | 1 | 0.1149 | 0 | 0 | 0 | 0.8182 | Ref |
| 8328 | MK317959 | Unassigned | Unassigned | Unassigned | 80_0 | Clustered | 41 | VC_80_0 | 41 | 0.5837 | 1 | 0.5837 | 0 | 0 | 0 | 0.9805 | Ref |
| 8329 | MK318076 | Unassigned | Unassigned | Unassigned | 79_7 | Clustered | 33 | VC_79_7 | 33 | 0.216 | 1 | 0.216 | 0 | 0 | 0 | 0.8409 | Ref |
| 8330 | MK318083 | Unassigned | Unassigned | Unassigned | 195_1 | Clustered/Singleton | 1 | VC_195_1 | 1 | 0 | 1 | 0 | 0 | 0 | 0 | 1 | Ref |
| 8331 | MK327140 | Unassigned | Unassigned | Unassigned | 460_0 | Clustered | 19 | VC_460_0 | 19 | 0.3744 | 1 | 0.3744 | 0 | 0 | 0 | 1 | Ref |
| 8332 | MK327928 | Unassigned | Unassigned | Unassigned | 283_18 | Clustered/Singleton | 1 | VC_283_18 | 1 | 0 | 1 | 0 | 0 | 0 | 0 | 1 | Ref |
| 8333 | MK327929 | Unassigned | Unassigned | Unassigned | 283_127 | Clustered/Singleton | 1 | VC_283_127 | 1 | 0 | 1 | 0 | 0 | 0 | 0 | 1 | Ref |
| 8334 | MK327930 | Unassigned | Unassigned | Unassigned | 772_19 | Clustered | 16 | VC_772_19 | 16 | 0.0975 | 0.9970466 | 0.0972 | 0 | 0 | 0 | 0.8083 | Ref |
| 8335 | MK327931 | Unassigned | Unassigned | Unassigned | 164_11 | Clustered | 10 | VC_164_11 | 10 | 0.1507 | 1 | 0.1507 | 0 | 0 | 0 | 0.9111 | Ref |
| 8336 | MK327932 | Unassigned | Unassigned | Unassigned | 590_0 | Clustered | 27 | VC_590_0 | 27 | 0.9947 | 1 | 0.9947 | 0 | 0 | 0 | 1 | Ref |
| 8337 | MK327933 | Unassigned | Unassigned | Unassigned | 286_4 | Clustered/Singleton | 1 | VC_286_4 | 1 | 0 | 1 | 0 | 0 | 0 | 0 | 1 | Ref |
| 8338 | MK327934 | Unassigned | Unassigned | Unassigned | 286_1 | Clustered | 21 | VC_286_1 | 21 | 0.1696 | 1 | 0.1696 | 0 | 0 | 0 | 0.9048 | Ref |
| 8339 | MK327935 | Unassigned | Unassigned | Unassigned | 590_0 | Clustered | 27 | VC_590_0 | 27 | 0.9947 | 1 | 0.9947 | 0 | 0 | 0 | 1 | Ref |
| 8340 | MK327936 | Unassigned | Unassigned | Unassigned | 283_133 | Clustered/Singleton | 1 | VC_283_133 | 1 | 0 | 1 | 0 | 0 | 0 | 0 | 1 | Ref |
| 8341 | MK327937 | Unassigned | Unassigned | Unassigned | 283_16 | Clustered/Singleton | 1 | VC_283_16 | 1 | 0 | 1 | 0 | 0 | 0 | 0 | 1 | Ref |
| 8342 | MK327939 | Unassigned | Unassigned | Unassigned | 283_119 | Clustered/Singleton | 1 | VC_283_119 | 1 | 0 | 1 | 0 | 0 | 0 | 0 | 1 | Ref |
| 8343 | MK327940 | Unassigned | Unassigned | Unassigned | 283_121 | Clustered | 2 | VC_283_121 | 2 | 0.0026 | 0.90700595 | 0.0023 | 0 | 0 | 0 | 1 | Ref |
| 8344 | MK327941 | Unassigned | Unassigned | Unassigned | 590_0 | Clustered | 27 | VC_590_0 | 27 | 0.9947 | 1 | 0.9947 | 0 | 0 | 0 | 1 | Ref |
| 8345 | MK327942 | Unassigned | Unassigned | Unassigned | 283_156 | Clustered/Singleton | 1 | VC_283_156 | 1 | 0 | 1 | 0 | 0 | 0 | 0 | 1 | Ref |
| 8346 | MK327943 | Unassigned | Unassigned | Unassigned | 286_1 | Clustered | 21 | VC_286_1 | 21 | 0.1696 | 1 | 0.1696 | 0 | 0 | 0 | 0.9048 | Ref |
| 8347 | MK327944 | Unassigned | Unassigned | Unassigned | 283_134 | Clustered/Singleton | 1 | VC_283_134 | 1 | 0 | 1 | 0 | 0 | 0 | 0 | 1 | Ref |
| 8348 | MK327945 | Unassigned | Unassigned | Unassigned | 283_121 | Clustered | 2 | VC_283_121 | 2 | 0.0026 | 0.90700595 | 0.0023 | 0 | 0 | 0 | 1 | Ref |
| 8349 | MK327946 | Unassigned | Unassigned | Unassigned | 283_17 | Clustered/Singleton | 1 | VC_283_17 | 1 | 0 | 1 | 0 | 0 | 0 | 0 | 1 | Ref |
| 8350 | MK327947 | Unassigned | Unassigned | Unassigned | 590_0 | Clustered | 27 | VC_590_0 | 27 | 0.9947 | 1 | 0.9947 | 0 | 0 | 0 | 1 | Ref |
| 8351 | MK330684 | Unassigned | Unassigned | Unassigned | 878_0 | Clustered | 32 | VC_878_0 | 32 | 0.9965 | 1 | 0.9965 | 0 | 0 | 0 | 1 | Ref |
| 8352 | MK331930 | Unassigned | Unassigned | Unassigned | 149_8 | Clustered | 66 | VC_149_8 | 66 | 0.4565 | 1 | 0.4565 | 0 | 0 | 0 | 0.8182 | Ref |
| 8353 | MK331931 | Unassigned | Unassigned | Unassigned | 149_8 | Clustered | 66 | VC_149_8 | 66 | 0.4565 | 1 | 0.4565 | 0 | 0 | 0 | 0.8182 | Ref |
| 8354 | MK335533 | Unassigned | Unassigned | Unassigned | 575_0 | Clustered | 35 | VC_575_0 | 35 | 0.8815 | 1 | 0.8815 | 0 | 0 | 0 | 1 | Ref |
| 8355 | MK340760 | Unassigned | Unassigned | Unassigned | 79_9 | Clustered | 2 | VC_79_9 | 2 | 0.0061 | 0.94860001 | 0.0058 | 0 | 0 | 0 | 1 | Ref |
| 8356 | MK340761 | Unassigned | Unassigned | Unassigned | 76_0 | Clustered | 37 | VC_76_0 | 37 | 0.9403 | 1 | 0.9403 | 0 | 0 | 0 | 1 | Ref |
| 8357 | MK340941 | Unassigned | Unassigned | Unassigned | 242_0 | Clustered | 2 | VC_242_0 | 2 | 0.0867 | 0.95379254 | 0.0827 | 0 | 0 | 0 | 1 | Ref |
| 8358 | MK348510 | Unassigned | Unassigned | Unassigned | 8_0 | Clustered | 58 | VC_8_0 | 58 | 0.8306 | 1 | 0.8306 | 0 | 0 | 0 | 0.9946 | Ref |
| 8359 | MK359299 | Unassigned | Unassigned | Unassigned | 325_23 | Clustered | 17 | VC_325_23 | 17 | 0.0716 | 1 | 0.0716 | 0 | 0 | 0 | 0.6397 | Ref |
| 8360 | MK359300 | Unassigned | Unassigned | Unassigned | 325_53 | Clustered | 7 | VC_325_53 | 7 | 0.0242 | 1 | 0.0242 | 0 | 0 | 0 | 0.9048 | Ref |
| 8361 | MK359301 | Unassigned | Unassigned | Unassigned | 517_1 | Clustered | 108 | VC_517_1 | 108 | 0.8483 | 1 | 0.8483 | 0 | 0 | 0 | 0.9955 | Ref |
| 8362 | MK359302 | Unassigned | Unassigned | Unassigned | 868_0 | Clustered | 25 | VC_868_0 | 25 | 0.5791 | 1 | 0.5791 | 0 | 0 | 0 | 1 | Ref |
| 8363 | MK359303 | Unassigned | Unassigned | Unassigned | 558_14 | Clustered | 8 | VC_558_14 | 8 | 0.0228 | 0.99945246 | 0.0228 | 0 | 0 | 0 | 0.8214 | Ref |
| 8364 | MK359304 | Unassigned | Unassigned | Unassigned | 325_53 | Clustered | 7 | VC_325_53 | 7 | 0.0242 | 1 | 0.0242 | 0 | 0 | 0 | 0.9048 | Ref |
| 8365 | MK359305 | Unassigned | Unassigned | Unassigned | 558_62 | Clustered/Singleton | 1 | VC_558_62 | 1 | 0 | 1 | 0 | 0 | 0 | 0 | 1 | Ref |
| 8366 | MK359306 | Unassigned | Unassigned | Unassigned | 558_53 | Clustered/Singleton | 1 | VC_558_53 | 1 | 0 | 1 | 0 | 0 | 0 | 0 | 1 | Ref |
| 8367 | MK359307 | Unassigned | Unassigned | Unassigned | 520_92 | Clustered | 5 | VC_520_92 | 5 | 0.0663 | 0.99999997 | 0.0663 | 0 | 0 | 0 | 1 | Ref |
| 8368 | MK359308 | Unassigned | Unassigned | Unassigned | 558_77 | Clustered | 3 | VC_558_77 | 3 | 0.0065 | 0.90877939 | 0.0059 | 0 | 0 | 0 | 0.6667 | Ref |
| 8369 | MK359309 | Unassigned | Unassigned | Unassigned | 551_7 | Clustered | 28 | VC_551_7 | 28 | 0.1661 | 1 | 0.1661 | 0 | 0 | 0 | 0.8519 | Ref |
| 8370 | MK359310 | Unassigned | Unassigned | Unassigned | 517_0 | Clustered | 18 | VC_517_0 | 18 | 0.1599 | 1 | 0.1599 | 0 | 0 | 0 | 1 | Ref |
| 8371 | MK359311 | Unassigned | Unassigned | Unassigned | 558_14 | Clustered | 8 | VC_558_14 | 8 | 0.0228 | 0.99945246 | 0.0228 | 0 | 0 | 0 | 0.8214 | Ref |
| 8372 | MK359312 | Unassigned | Unassigned | Unassigned | 325_35 | Clustered | 17 | VC_325_35 | 17 | 0.0732 | 1 | 0.0732 | 0 | 0 | 0 | 0.7721 | Ref |
| 8373 | MK359313 | Unassigned | Unassigned | Unassigned | 1175_0 | Clustered | 22 | VC_1175_0 | 22 | 0.4562 | 1 | 0.4562 | 0 | 0 | 0 | 1 | Ref |
| 8374 | MK359314 | Unassigned | Unassigned | Unassigned | 558_50 | Clustered | 15 | VC_558_50 | 15 | 0.0468 | 1 | 0.0468 | 0 | 0 | 0 | 0.9238 | Ref |
| 8375 | MK359315 | Unassigned | Unassigned | Unassigned | 520_80 | Clustered | 3 | VC_520_80 | 3 | 0.0295 | 0.99859904 | 0.0295 | 0 | 0 | 0 | 1 | Ref |
| 8376 | MK359316 | Unassigned | Unassigned | Unassigned | 325_33 | Clustered | 4 | VC_325_33 | 4 | 0.0149 | 0.99998565 | 0.0149 | 0 | 0 | 0 | 1 | Ref |
| 8377 | MK359317 | Unassigned | Unassigned | Unassigned | 558_59 | Clustered/Singleton | 1 | VC_558_59 | 1 | 0 | 1 | 0 | 0 | 0 | 0 | 1 | Ref |
| 8378 | MK359318 | Unassigned | Unassigned | Unassigned | 558_61 | Clustered | 2 | VC_558_61 | 2 | 0.0032 | 0.66165479 | 0.0021 | 0 | 0 | 0 | 1 | Ref |
| 8379 | MK359319 | Unassigned | Unassigned | Unassigned | 954_0 | Clustered | 13 | VC_954_0 | 13 | 0.863 | 1 | 0.863 | 0 | 0 | 0 | 1 | Ref |
| 8380 | MK359320 | Unassigned | Unassigned | Unassigned | 551_0 | Clustered | 16 | VC_551_0 | 16 | 0.0915 | 1 | 0.0915 | 0 | 0 | 0 | 0.6667 | Ref |
| 8381 | MK359321 | Unassigned | Unassigned | Unassigned | 558_13 | Clustered/Singleton | 1 | VC_558_13 | 1 | 0 | 1 | 0 | 0 | 0 | 0 | 1 | Ref |
| 8382 | MK359322 | Unassigned | Unassigned | Unassigned | 517_0 | Clustered | 18 | VC_517_0 | 18 | 0.1599 | 1 | 0.1599 | 0 | 0 | 0 | 1 | Ref |
| 8383 | MK359323 | Unassigned | Unassigned | Unassigned | 517_1 | Clustered | 108 | VC_517_1 | 108 | 0.8483 | 1 | 0.8483 | 0 | 0 | 0 | 0.9955 | Ref |
| 8384 | MK359324 | Unassigned | Unassigned | Unassigned | 558_9 | Clustered | 3 | VC_558_9 | 3 | 0.0064 | 0.82911529 | 0.0053 | 0 | 0 | 0 | 0.6667 | Ref |
| 8385 | MK359325 | Unassigned | Unassigned | Unassigned | 517_1 | Clustered | 108 | VC_517_1 | 108 | 0.8483 | 1 | 0.8483 | 0 | 0 | 0 | 0.9955 | Ref |
| 8386 | MK359326 | Unassigned | Unassigned | Unassigned | 558_20 | Clustered | 17 | VC_558_20 | 17 | 0.054 | 1 | 0.054 | 0 | 0 | 0 | 0.9926 | Ref |
| 8387 | MK359327 | Unassigned | Unassigned | Unassigned | 558_45 | Clustered/Singleton | 1 | VC_558_45 | 1 | 0 | 1 | 0 | 0 | 0 | 0 | 1 | Ref |
| 8388 | MK359328 | Unassigned | Unassigned | Unassigned | 558_17 | Clustered/Singleton | 1 | VC_558_17 | 1 | 0 | 1 | 0 | 0 | 0 | 0 | 1 | Ref |
| 8389 | MK359329 | Unassigned | Unassigned | Unassigned | 558_69 | Clustered/Singleton | 1 | VC_558_69 | 1 | 0 | 1 | 0 | 0 | 0 | 0 | 1 | Ref |
| 8390 | MK359330 | Unassigned | Unassigned | Unassigned | 558_66 | Clustered/Singleton | 1 | VC_558_66 | 1 | 0 | 1 | 0 | 0 | 0 | 0 | 1 | Ref |
| 8391 | MK359331 | Unassigned | Unassigned | Unassigned | 325_10 | Clustered | 3 | VC_325_10 | 3 | 0.012 | 0.99859169 | 0.012 | 0 | 0 | 0 | 1 | Ref |
| 8392 | MK359332 | Unassigned | Unassigned | Unassigned | 1071_0 | Clustered | 20 | VC_1071_0 | 20 | 0.3268 | 1 | 0.3268 | 0 | 0 | 0 | 0.9842 | Ref |
| 8393 | MK359333 | Unassigned | Unassigned | Unassigned | 558_28 | Clustered/Singleton | 1 | VC_558_28 | 1 | 0 | 1 | 0 | 0 | 0 | 0 | 1 | Ref |
| 8394 | MK359334 | Unassigned | Unassigned | Unassigned | 558_49 | Clustered | 7 | VC_558_49 | 7 | 0.0194 | 0.99510369 | 0.0193 | 0 | 0 | 0 | 0.8571 | Ref |
| 8395 | MK359335 | Unassigned | Unassigned | Unassigned | 558_20 | Clustered | 17 | VC_558_20 | 17 | 0.054 | 1 | 0.054 | 0 | 0 | 0 | 0.9926 | Ref |
| 8396 | MK359336 | Unassigned | Unassigned | Unassigned | 558_36 | Clustered | 4 | VC_558_36 | 4 | 0.0096 | 0.85913852 | 0.0082 | 0 | 0 | 0 | 1 | Ref |
| 8397 | MK359337 | Unassigned | Unassigned | Unassigned | 558_43 | Clustered/Singleton | 1 | VC_558_43 | 1 | 0 | 1 | 0 | 0 | 0 | 0 | 1 | Ref |
| 8398 | MK359338 | Unassigned | Unassigned | Unassigned | 558_20 | Clustered | 17 | VC_558_20 | 17 | 0.054 | 1 | 0.054 | 0 | 0 | 0 | 0.9926 | Ref |
| 8399 | MK359339 | Unassigned | Unassigned | Unassigned | 558_20 | Clustered | 17 | VC_558_20 | 17 | 0.054 | 1 | 0.054 | 0 | 0 | 0 | 0.9926 | Ref |
| 8400 | MK359340 | Unassigned | Unassigned | Unassigned | 517_1 | Clustered | 108 | VC_517_1 | 108 | 0.8483 | 1 | 0.8483 | 0 | 0 | 0 | 0.9955 | Ref |
| 8401 | MK359341 | Unassigned | Unassigned | Unassigned | 514_0 | Clustered | 75 | VC_514_0 | 75 | 0.9402 | 1 | 0.9402 | 0 | 0 | 0 | 1 | Ref |
| 8402 | MK359342 | Unassigned | Unassigned | Unassigned | 558_15 | Clustered/Singleton | 1 | VC_558_15 | 1 | 0 | 1 | 0 | 0 | 0 | 0 | 1 | Ref |
| 8403 | MK359343 | Unassigned | Unassigned | Unassigned | 520_57 | Clustered/Singleton | 1 | VC_520_57 | 1 | 0 | 1 | 0 | 0 | 0 | 0 | 1 | Ref |
| 8404 | MK359344 | Unassigned | Unassigned | Unassigned | 558_30 | Clustered | 3 | VC_558_30 | 3 | 0.0064 | 0.76574053 | 0.0049 | 0 | 0 | 0 | 1 | Ref |
| 8405 | MK359345 | Unassigned | Unassigned | Unassigned | 558_44 | Clustered/Singleton | 1 | VC_558_44 | 1 | 0 | 1 | 0 | 0 | 0 | 0 | 1 | Ref |
| 8406 | MK359346 | Unassigned | Unassigned | Unassigned | 558_71 | Clustered/Singleton | 1 | VC_558_71 | 1 | 0 | 1 | 0 | 0 | 0 | 0 | 1 | Ref |
| 8407 | MK359347 | Unassigned | Unassigned | Unassigned | 558_20 | Clustered | 17 | VC_558_20 | 17 | 0.054 | 1 | 0.054 | 0 | 0 | 0 | 0.9926 | Ref |
| 8408 | MK359348 | Unassigned | Unassigned | Unassigned | 558_12 | Clustered | 3 | VC_558_12 | 3 | 0.0064 | 0.87854253 | 0.0057 | 0 | 0 | 0 | 1 | Ref |
| 8409 | MK359349 | Unassigned | Unassigned | Unassigned | 558_46 | Clustered/Singleton | 1 | VC_558_46 | 1 | 0 | 1 | 0 | 0 | 0 | 0 | 1 | Ref |
| 8410 | MK359350 | Unassigned | Unassigned | Unassigned | 558_64 | Clustered/Singleton | 1 | VC_558_64 | 1 | 0 | 1 | 0 | 0 | 0 | 0 | 1 | Ref |
| 8411 | MK359351 | Unassigned | Unassigned | Unassigned | 1071_0 | Clustered | 20 | VC_1071_0 | 20 | 0.3268 | 1 | 0.3268 | 0 | 0 | 0 | 0.9842 | Ref |
| 8412 | MK359352 | Unassigned | Unassigned | Unassigned | 223_0 | Clustered | 38 | VC_223_0 | 38 | 0.9095 | 1 | 0.9095 | 0 | 0 | 0 | 1 | Ref |
| 8413 | MK359353 | Unassigned | Unassigned | Unassigned | 558_4 | Clustered/Singleton | 1 | VC_558_4 | 1 | 0 | 1 | 0 | 0 | 0 | 0 | 1 | Ref |
| 8414 | MK359354 | Unassigned | Unassigned | Unassigned | 325_5 | Clustered | 11 | VC_325_5 | 11 | 0.047 | 1 | 0.047 | 0 | 0 | 0 | 0.7636 | Ref |
| 8415 | MK359355 | Unassigned | Unassigned | Unassigned | 558_3 | Clustered/Singleton | 1 | VC_558_3 | 1 | 0 | 1 | 0 | 0 | 0 | 0 | 1 | Ref |
| 8416 | MK359356 | Unassigned | Unassigned | Unassigned | 517_1 | Clustered | 108 | VC_517_1 | 108 | 0.8483 | 1 | 0.8483 | 0 | 0 | 0 | 0.9955 | Ref |
| 8417 | MK359357 | Unassigned | Unassigned | Unassigned | 509_0 | Clustered | 37 | VC_509_0 | 37 | 0.9689 | 1 | 0.9689 | 0 | 0 | 0 | 1 | Ref |
| 8418 | MK359358 | Unassigned | Unassigned | Unassigned | 558_14 | Clustered | 8 | VC_558_14 | 8 | 0.0228 | 0.99945246 | 0.0228 | 0 | 0 | 0 | 0.8214 | Ref |
| 8419 | MK359359 | Unassigned | Unassigned | Unassigned | 558_73 | Clustered | 2 | VC_558_73 | 2 | 0.0033 | 0.84374839 | 0.0028 | 0 | 0 | 0 | 1 | Ref |
| 8420 | MK359360 | Unassigned | Unassigned | Unassigned | 223_0 | Clustered | 38 | VC_223_0 | 38 | 0.9095 | 1 | 0.9095 | 0 | 0 | 0 | 1 | Ref |
| 8421 | MK359361 | Unassigned | Unassigned | Unassigned | 558_27 | Clustered | 2 | VC_558_27 | 2 | 0.0032 | 0.64135461 | 0.002 | 0 | 0 | 0 | 1 | Ref |
| 8422 | MK359362 | Unassigned | Unassigned | Unassigned | 558_20 | Clustered | 17 | VC_558_20 | 17 | 0.054 | 1 | 0.054 | 0 | 0 | 0 | 0.9926 | Ref |

| | | | | | | | | | | | | | | | | | |
|---|---|---|---|---|---|---|---|---|---|---|---|---|---|---|---|---|---|
| 8423 | MK359363 | Unassigned | Unassigned | Unassigned | 558_36 | Clustered | 4 | VC_558_36 | 4 | 0.0096 | 0.85913852 | 0.0082 | 0 | 0 | 0 | 1 | Ref |
| 8424 | MK359364 | Unassigned | Unassigned | Unassigned | 558_39 | Clustered | 3 | VC_558_39 | 3 | 0.0063 | 0.73849574 | 0.0047 | 0 | 0 | 0 | 0.6667 | Ref |
| 8425 | MK359365 | Unassigned | Unassigned | Unassigned | 558_50 | Clustered | 15 | VC_558_50 | 15 | 0.0468 | 1 | 0.0468 | 0 | 0 | 0 | 0.9238 | Ref |
| 8426 | MK359366 | Unassigned | Unassigned | Unassigned | 558_36 | Clustered | 4 | VC_558_36 | 4 | 0.0096 | 0.85913852 | 0.0082 | 0 | 0 | 0 | 1 | Ref |
| 8427 | MK359990 | Unassigned | Unassigned | Unassigned | 632_0 | Clustered | 37 | VC_632_0 | 37 | 0.581 | 1 | 0.581 | 0 | 0 | 0 | 0.9234 | Ref |
| 8428 | MK360024 | Unassigned | Unassigned | Unassigned | 695_0 | Clustered | 48 | VC_695_0 | 48 | 0.8162 | 1 | 0.8162 | 0 | 0 | 0 | 1 | Ref |
| 8429 | MK360025 | Unassigned | Unassigned | Unassigned | 968_0 | Clustered | 4 | VC_968_0 | 4 | 0.2959 | 0.99997642 | 0.2959 | 0 | 0 | 0 | 1 | Ref |
| 8430 | MK370036 | Unassigned | Unassigned | Unassigned | 191_1 | Clustered/Singleton | 1 | VC_191_1 | 1 | 0 | 1 | 0 | 0 | 0 | 0 | 1 | Ref |
| 8431 | MK372342 | Unassigned | Unassigned | Unassigned | 456_0 | Clustered | 62 | VC_456_0 | 62 | 0.9026 | 1 | 0.9026 | 0 | 0 | 0 | 1 | Ref |
| 8432 | MK373770 | Unassigned | Unassigned | Unassigned | 813_1 | Clustered | 24 | VC_813_1 | 24 | 0.535 | 1 | 0.535 | 0 | 0 | 0 | 1 | Ref |
| 8433 | MK373771 | Unassigned | Unassigned | Unassigned | 452_0 | Clustered | 66 | VC_452_0 | 66 | 0.9248 | 1 | 0.9248 | 0 | 0 | 0 | 1 | Ref |
| 8434 | MK373772 | Unassigned | Unassigned | Unassigned | 1228_0 | Clustered | 2 | VC_1228_0 | 2 | 0.9902 | 0.72985431 | 0.7227 | 0 | 0 | 0 | 1 | Ref |
| 8435 | MK373773 | Unassigned | Unassigned | Unassigned | 452_0 | Clustered | 66 | VC_452_0 | 66 | 0.9248 | 1 | 0.9248 | 0 | 0 | 0 | 1 | Ref |
| 8436 | MK373774 | Unassigned | Unassigned | Unassigned | 286_3 | Clustered | 2 | VC_286_3 | 2 | 0.0073 | 0.74885804 | 0.0055 | 0 | 0 | 0 | 1 | Ref |
| 8437 | MK373775 | Unassigned | Unassigned | Unassigned | 286_3 | Clustered | 2 | VC_286_3 | 2 | 0.0073 | 0.74885804 | 0.0055 | 0 | 0 | 0 | 1 | Ref |
| 8438 | MK373776 | Unassigned | Unassigned | Unassigned | 618_0 | Clustered | 13 | VC_618_0 | 13 | 0.6252 | 1 | 0.6252 | 0 | 0 | 0 | 1 | Ref |
| 8439 | MK373777 | Unassigned | Unassigned | Unassigned | 618_0 | Clustered | 13 | VC_618_0 | 13 | 0.6252 | 1 | 0.6252 | 0 | 0 | 0 | 1 | Ref |
| 8440 | MK373778 | Unassigned | Unassigned | Unassigned | 286_1 | Clustered | 21 | VC_286_1 | 21 | 0.1696 | 1 | 0.1696 | 0 | 0 | 0 | 0.9048 | Ref |
| 8441 | MK373779 | Unassigned | Unassigned | Unassigned | 283_124 | Clustered/Singleton | 1 | VC_283_124 | 1 | 0 | 1 | 0 | 0 | 0 | 0 | 1 | Ref |
| 8442 | MK373780 | Unassigned | Unassigned | Unassigned | 772_19 | Clustered | 16 | VC_772_19 | 16 | 0.0975 | 0.9970466 | 0.0972 | 0 | 0 | 0 | 0.8083 | Ref |
| 8443 | MK373781 | Unassigned | Unassigned | Unassigned | 283_53 | Clustered/Singleton | 1 | VC_283_53 | 1 | 0 | 1 | 0 | 0 | 0 | 0 | 1 | Ref |
| 8444 | MK373782 | Unassigned | Unassigned | Unassigned | 286_1 | Clustered | 21 | VC_286_1 | 21 | 0.1696 | 1 | 0.1696 | 0 | 0 | 0 | 0.9048 | Ref |
| 8445 | MK373783 | Unassigned | Unassigned | Unassigned | 132_1 | Clustered | 17 | VC_132_1 | 17 | 0.1738 | 1 | 0.1738 | 0 | 0 | 0 | 0.9853 | Ref |
| 8446 | MK373784 | Unassigned | Unassigned | Unassigned | 286_1 | Clustered | 21 | VC_286_1 | 21 | 0.1696 | 1 | 0.1696 | 0 | 0 | 0 | 0.9048 | Ref |
| 8447 | MK373785 | Unassigned | Unassigned | Unassigned | 283_12 | Clustered/Singleton | 1 | VC_283_12 | 1 | 0 | 1 | 0 | 0 | 0 | 0 | 1 | Ref |
| 8448 | MK373786 | Unassigned | Unassigned | Unassigned | 283_43 | Clustered/Singleton | 1 | VC_283_43 | 1 | 0 | 1 | 0 | 0 | 0 | 0 | 1 | Ref |
| 8449 | MK373787 | Unassigned | Unassigned | Unassigned | 283_76 | Clustered/Singleton | 1 | VC_283_76 | 1 | 0 | 1 | 0 | 0 | 0 | 0 | 1 | Ref |
| 8450 | MK373788 | Unassigned | Unassigned | Unassigned | 847_1 | Clustered | 26 | VC_847_1 | 26 | 0.6323 | 1 | 0.6323 | 0 | 0 | 0 | 0.9815 | Ref |
| 8451 | MK373789 | Unassigned | Unassigned | Unassigned | 835_0 | Clustered | 67 | VC_835_0 | 67 | 0.9624 | 1 | 0.9624 | 0 | 0 | 0 | 1 | Ref |
| 8452 | MK373790 | Unassigned | Unassigned | Unassigned | 835_0 | Clustered | 67 | VC_835_0 | 67 | 0.9624 | 1 | 0.9624 | 0 | 0 | 0 | 1 | Ref |
| 8453 | MK373791 | Unassigned | Unassigned | Unassigned | 835_0 | Clustered | 67 | VC_835_0 | 67 | 0.9624 | 1 | 0.9624 | 0 | 0 | 0 | 1 | Ref |
| 8454 | MK373792 | Unassigned | Unassigned | Unassigned | 190_1 | Clustered/Singleton | 1 | VC_190_1 | 1 | 0 | 1 | 0 | 0 | 0 | 0 | 1 | Ref |
| 8455 | MK373793 | Unassigned | Unassigned | Unassigned | 456_0 | Clustered | 62 | VC_456_0 | 62 | 0.9026 | 1 | 0.9026 | 0 | 0 | 0 | 1 | Ref |
| 8456 | MK373794 | Unassigned | Unassigned | Unassigned | 722_0 | Clustered | 13 | VC_722_0 | 13 | 0.5104 | 1 | 0.5104 | 0 | 0 | 0 | 1 | Ref |
| 8457 | MK373795 | Unassigned | Unassigned | Unassigned | 722_0 | Clustered | 13 | VC_722_0 | 13 | 0.5104 | 1 | 0.5104 | 0 | 0 | 0 | 1 | Ref |
| 8458 | MK373796 | Unassigned | Unassigned | Unassigned | 190_9 | Clustered/Singleton | 1 | VC_190_9 | 1 | 0 | 1 | 0 | 0 | 0 | 0 | 1 | Ref |
| 8459 | MK373797 | Unassigned | Unassigned | Unassigned | 190_2 | Clustered | 2 | VC_190_2 | 2 | 0.0073 | 0.95531062 | 0.007 | 0 | 0 | 0 | 1 | Ref |
| 8460 | MK373798 | Unassigned | Unassigned | Unassigned | 941_0 | Clustered | 40 | VC_941_0 | 40 | 0.6903 | 1 | 0.6903 | 0 | 0 | 0 | 1 | Ref |
| 8461 | MK373799 | Unassigned | Unassigned | Unassigned | 190_2 | Clustered | 2 | VC_190_2 | 2 | 0.0073 | 0.95531062 | 0.007 | 0 | 0 | 0 | 1 | Ref |
| 8462 | MK376953 | Unassigned | Unassigned | Unassigned | 799_1 | Clustered | 27 | VC_799_1 | 27 | 0.8611 | 1 | 0.8611 | 0 | 0 | 0 | 1 | Ref |
| 8463 | MK376954 | Unassigned | Unassigned | Unassigned | 767_0 | Clustered | 34 | VC_767_0 | 34 | 0.9173 | 1 | 0.9173 | 0 | 0 | 0 | 1 | Ref |
| 8464 | MK376955 | Unassigned | Unassigned | Unassigned | 237_3 | Clustered | 8 | VC_237_3 | 8 | 0.1092 | 1 | 0.1092 | 0 | 0 | 0 | 1 | Ref |
| 8465 | MK376956 | Unassigned | Unassigned | Unassigned | 1181_1 | Clustered | 10 | VC_1181_1 | 10 | 0.5828 | 1 | 0.5828 | 0 | 0 | 0 | 1 | Ref |
| 8466 | MK376957 | Unassigned | Unassigned | Unassigned | 799_1 | Clustered | 27 | VC_799_1 | 27 | 0.8611 | 1 | 0.8611 | 0 | 0 | 0 | 1 | Ref |
| 8467 | MK376958 | Unassigned | Unassigned | Unassigned | 954_0 | Clustered | 13 | VC_954_0 | 13 | 0.863 | 1 | 0.863 | 0 | 0 | 0 | 1 | Ref |
| 8468 | MK376959 | Unassigned | Unassigned | Unassigned | 1181_1 | Clustered | 10 | VC_1181_1 | 10 | 0.5828 | 1 | 0.5828 | 0 | 0 | 0 | 1 | Ref |
| 8469 | MK376960 | Unassigned | Unassigned | Unassigned | 767_0 | Clustered | 34 | VC_767_0 | 34 | 0.9173 | 1 | 0.9173 | 0 | 0 | 0 | 1 | Ref |
| 8470 | MK376961 | Unassigned | Unassigned | Unassigned | 557_1 | Clustered | 11 | VC_557_1 | 11 | 0.4945 | 1 | 0.4945 | 0 | 0 | 0 | 0.7818 | Ref |
| 8471 | MK376962 | Unassigned | Unassigned | Unassigned | 767_0 | Clustered | 34 | VC_767_0 | 34 | 0.9173 | 1 | 0.9173 | 0 | 0 | 0 | 1 | Ref |
| 8472 | MK376963 | Unassigned | Unassigned | Unassigned | 856_1 | Clustered | 13 | VC_856_1 | 13 | 0.5848 | 1 | 0.5848 | 0 | 0 | 0 | 1 | Ref |
| 8473 | MK380014 | Unassigned | Unassigned | Unassigned | 452_0 | Clustered | 66 | VC_452_0 | 66 | 0.9248 | 1 | 0.9248 | 0 | 0 | 0 | 1 | Ref |
| 8474 | MK380015 | Unassigned | Unassigned | Unassigned | 823_0 | Clustered | 124 | VC_823_0 | 124 | 0.9492 | 1 | 0.9492 | 0 | 0 | 0 | 1 | Ref |
| 8475 | MK380016 | Unassigned | Unassigned | Unassigned | 823_0 | Clustered | 124 | VC_823_0 | 124 | 0.9492 | 1 | 0.9492 | 0 | 0 | 0 | 1 | Ref |
| 8476 | MK387309 | Unassigned | Unassigned | Unassigned | 644_0 | Clustered | 5 | VC_644_0 | 5 | 0.6649 | 0.99999975 | 0.6649 | 0 | 0 | 0 | 1 | Ref |
| 8477 | MK387337 | Unassigned | Unassigned | Unassigned | 845_0 | Clustered | 3 | VC_845_0 | 3 | 0.3348 | 0.99831497 | 0.3343 | 0 | 0 | 0 | 1 | Ref |
| 8478 | MK387869 | Unassigned | Unassigned | Unassigned | 112_0 | Clustered | 8 | VC_112_0 | 8 | 0.2537 | 1 | 0.2537 | 0 | 0 | 0 | 1 | Ref |
| 8479 | MK388689 | Unassigned | Unassigned | Unassigned | 846_0 | Clustered | 2 | VC_846_0 | 2 | 0.5455 | 0.92357512 | 0.5038 | 0 | 0 | 0 | 1 | Ref |
| 8480 | MK388859 | Unassigned | Unassigned | Unassigned | 181_0 | Clustered | 5 | VC_181_0 | 5 | 0.1168 | 0.99999995 | 0.1168 | 0 | 0 | 0 | 1 | Ref |
| 8481 | MK392363 | Unassigned | Unassigned | Unassigned | 981_1 | Clustered | 26 | VC_981_1 | 26 | 0.7009 | 1 | 0.7009 | 0 | 0 | 0 | 1 | Ref |
| 8482 | MK392364 | Unassigned | Unassigned | Unassigned | 981_1 | Clustered | 26 | VC_981_1 | 26 | 0.7009 | 1 | 0.7009 | 0 | 0 | 0 | 1 | Ref |
| 8483 | MK392365 | Unassigned | Unassigned | Unassigned | 981_1 | Clustered | 26 | VC_981_1 | 26 | 0.7009 | 1 | 0.7009 | 0 | 0 | 0 | 1 | Ref |
| 8484 | MK392366 | Unassigned | Unassigned | Unassigned | 981_1 | Clustered | 26 | VC_981_1 | 26 | 0.7009 | 1 | 0.7009 | 0 | 0 | 0 | 1 | Ref |
| 8485 | MK392367 | Unassigned | Unassigned | Unassigned | 521_0 | Clustered | 37 | VC_521_0 | 37 | 0.912 | 1 | 0.912 | 0 | 0 | 0 | 1 | Ref |
| 8486 | MK392368 | Unassigned | Unassigned | Unassigned | 1037_0 | Clustered | 6 | VC_1037_0 | 6 | 0.6798 | 1 | 0.6798 | 0 | 0 | 0 | 1 | Ref |
| 8487 | MK393882 | Unassigned | Unassigned | Unassigned | 191_16 | Clustered | 12 | VC_191_16 | 12 | 0.0643 | 1 | 0.0643 | 0 | 0 | 0 | 0.9394 | Ref |
| 8488 | MK408758 | Unassigned | Unassigned | Unassigned | 843_0 | Clustered | 5 | VC_843_0 | 5 | 0.2843 | 0.99999994 | 0.2843 | 0 | 0 | 0 | 1 | Ref |
| 8489 | MK411746 | Unassigned | Unassigned | Unassigned | 1370_0 | Clustered | 2 | VC_1370_0 | 2 | 0.8214 | 0.91237788 | 0.7494 | 0 | 0 | 0 | 1 | Ref |
| 8490 | MK416007 | Unassigned | Unassigned | Unassigned | 242_1 | Clustered | 20 | VC_242_1 | 20 | 0.684 | 1 | 0.684 | 0 | 0 | 0 | 0.8368 | Ref |
| 8491 | MK416008 | Unassigned | Unassigned | Unassigned | 35_0 | Clustered | 6 | VC_35_0 | 6 | 0.2254 | 1 | 0.2254 | 0 | 0 | 0 | 0.9333 | Ref |
| 8492 | MK416009 | Unassigned | Unassigned | Unassigned | 6_0 | Clustered | 44 | VC_6_0 | 44 | 0.908 | 1 | 0.908 | 0 | 0 | 0 | 1 | Ref |
| 8493 | MK416010 | Unassigned | Unassigned | Unassigned | 476_0 | Clustered | 19 | VC_476_0 | 19 | 0.7814 | 1 | 0.7814 | 0 | 0 | 0 | 1 | Ref |
| 8494 | MK416011 | Unassigned | Unassigned | Unassigned | 6_0 | Clustered | 44 | VC_6_0 | 44 | 0.908 | 1 | 0.908 | 0 | 0 | 0 | 1 | Ref |
| 8495 | MK416012 | Unassigned | Unassigned | Unassigned | 6_0 | Clustered | 44 | VC_6_0 | 44 | 0.908 | 1 | 0.908 | 0 | 0 | 0 | 1 | Ref |
| 8496 | MK416013 | Unassigned | Unassigned | Unassigned | 35_0 | Clustered | 6 | VC_35_0 | 6 | 0.2254 | 1 | 0.2254 | 0 | 0 | 0 | 0.9333 | Ref |
| 8497 | MK416014 | Unassigned | Unassigned | Unassigned | 243_0 | Clustered | 2 | VC_243_0 | 2 | 0.0637 | 0.95609892 | 0.0609 | 0 | 0 | 0 | 1 | Ref |
| 8498 | MK416015 | Unassigned | Unassigned | Unassigned | 229_0 | Clustered | 32 | VC_229_0 | 32 | 0.9119 | 1 | 0.9119 | 0 | 0 | 0 | 1 | Ref |
| 8499 | MK416017 | Unassigned | Unassigned | Unassigned | 244_0 | Clustered | 12 | VC_244_0 | 12 | 0.4039 | 1 | 0.4039 | 0 | 0 | 0 | 1 | Ref |
| 8500 | MK416018 | Unassigned | Unassigned | Unassigned | 228_0 | Clustered | 60 | VC_228_0 | 60 | 0.942 | 1 | 0.942 | 0 | 0 | 0 | 1 | Ref |
| 8501 | MK416019 | Unassigned | Unassigned | Unassigned | 242_1 | Clustered | 20 | VC_242_1 | 20 | 0.684 | 1 | 0.684 | 0 | 0 | 0 | 0.8368 | Ref |
| 8502 | MK416020 | Unassigned | Unassigned | Unassigned | 6_0 | Clustered | 44 | VC_6_0 | 44 | 0.908 | 1 | 0.908 | 0 | 0 | 0 | 1 | Ref |
| 8503 | MK416022 | Unassigned | Unassigned | Unassigned | 35_0 | Clustered | 6 | VC_35_0 | 6 | 0.2254 | 1 | 0.2254 | 0 | 0 | 0 | 0.9333 | Ref |
| 8504 | MK417514 | Unassigned | Unassigned | Unassigned | 149_8 | Clustered | 66 | VC_149_8 | 66 | 0.4565 | 1 | 0.4565 | 0 | 0 | 0 | 0.8182 | Ref |
| 8505 | MK417515 | Unassigned | Unassigned | Unassigned | 149_8 | Clustered | 66 | VC_149_8 | 66 | 0.4565 | 1 | 0.4565 | 0 | 0 | 0 | 0.8182 | Ref |
| 8506 | MK417516 | Unassigned | Unassigned | Unassigned | 149_8 | Clustered | 66 | VC_149_8 | 66 | 0.4565 | 1 | 0.4565 | 0 | 0 | 0 | 0.8182 | Ref |
| 8507 | MK421971 | Unassigned | Unassigned | Unassigned | 491_0 | Clustered | 36 | VC_491_0 | 36 | 0.5336 | 1 | 0.5336 | 0 | 0 | 0 | 0.9714 | Ref |
| 8508 | MK422448 | Unassigned | Unassigned | Unassigned | 476_0 | Clustered | 19 | VC_476_0 | 19 | 0.7814 | 1 | 0.7814 | 0 | 0 | 0 | 1 | Ref |
| 8509 | MK422449 | Unassigned | Unassigned | Unassigned | 6_0 | Clustered | 44 | VC_6_0 | 44 | 0.908 | 1 | 0.908 | 0 | 0 | 0 | 1 | Ref |
| 8510 | MK422451 | Unassigned | Unassigned | Unassigned | 723_0 | Clustered | 9 | VC_723_0 | 9 | 0.7138 | 1 | 0.7138 | 0 | 0 | 0 | 1 | Ref |
| 8511 | MK422453 | Unassigned | Unassigned | Unassigned | 6_0 | Clustered | 44 | VC_6_0 | 44 | 0.908 | 1 | 0.908 | 0 | 0 | 0 | 1 | Ref |
| 8512 | MK422454 | Unassigned | Unassigned | Unassigned | 6_0 | Clustered | 44 | VC_6_0 | 44 | 0.908 | 1 | 0.908 | 0 | 0 | 0 | 1 | Ref |
| 8513 | MK422455 | Unassigned | Unassigned | Unassigned | 242_1 | Clustered | 20 | VC_242_1 | 20 | 0.684 | 1 | 0.684 | 0 | 0 | 0 | 0.8368 | Ref |
| 8514 | MK424903 | Unassigned | Unassigned | Unassigned | 277_0 | Clustered | 3 | VC_277_0 | 3 | 1 | 1 | 1 | 0 | 0 | 0 | 1 | Ref |
| 8515 | MK433259 | Unassigned | Unassigned | Unassigned | 872_0 | Clustered | 31 | VC_872_0 | 31 | 0.8151 | 1 | 0.8151 | 0 | 0 | 0 | 0.8194 | Ref |
| 8516 | MK433260 | Unassigned | Unassigned | Unassigned | 872_0 | Clustered | 31 | VC_872_0 | 31 | 0.8151 | 1 | 0.8151 | 0 | 0 | 0 | 0.8194 | Ref |
| 8517 | MK433261 | Unassigned | Unassigned | Unassigned | 868_0 | Clustered | 25 | VC_868_0 | 25 | 0.5791 | 1 | 0.5791 | 0 | 0 | 0 | 1 | Ref |
| 8518 | MK433262 | Unassigned | Unassigned | Unassigned | 551_8 | Clustered | 19 | VC_551_8 | 19 | 0.1039 | 1 | 0.1039 | 0 | 0 | 0 | 0.8713 | Ref |
| 8519 | MK433263 | Unassigned | Unassigned | Unassigned | 868_0 | Clustered | 25 | VC_868_0 | 25 | 0.5791 | 1 | 0.5791 | 0 | 0 | 0 | 1 | Ref |
| 8520 | MK433265 | Unassigned | Unassigned | Unassigned | 868_0 | Clustered | 25 | VC_868_0 | 25 | 0.5791 | 1 | 0.5791 | 0 | 0 | 0 | 1 | Ref |
| 8521 | MK433266 | Unassigned | Unassigned | Unassigned | 373_0 | Clustered | 10 | VC_373_0 | 10 | 0.7471 | 1 | 0.7471 | 0 | 0 | 0 | 1 | Ref |
| 8522 | MK433267 | Unassigned | Unassigned | Unassigned | 868_0 | Clustered | 25 | VC_868_0 | 25 | 0.5791 | 1 | 0.5791 | 0 | 0 | 0 | 1 | Ref |
| 8523 | MK433268 | Unassigned | Unassigned | Unassigned | 872_0 | Clustered | 31 | VC_872_0 | 31 | 0.8151 | 1 | 0.8151 | 0 | 0 | 0 | 0.8194 | Ref |
| 8524 | MK433269 | Unassigned | Unassigned | Unassigned | 872_0 | Clustered | 31 | VC_872_0 | 31 | 0.8151 | 1 | 0.8151 | 0 | 0 | 0 | 0.8194 | Ref |
| 8525 | MK433270 | Unassigned | Unassigned | Unassigned | 982_0 | Clustered | 62 | VC_982_0 | 62 | 0.8646 | 1 | 0.8646 | 0 | 0 | 0 | 0.743 | Ref |
| 8526 | MK433271 | Unassigned | Unassigned | Unassigned | 982_0 | Clustered | 62 | VC_982_0 | 62 | 0.8646 | 1 | 0.8646 | 0 | 0 | 0 | 0.743 | Ref |
| 8527 | MK433272 | Unassigned | Unassigned | Unassigned | 767_0 | Clustered | 34 | VC_767_0 | 34 | 0.9173 | 1 | 0.9173 | 0 | 0 | 0 | 1 | Ref |
| 8528 | MK433273 | Unassigned | Unassigned | Unassigned | 514_0 | Clustered | 75 | VC_514_0 | 75 | 0.9402 | 1 | 0.9402 | 0 | 0 | 0 | 1 | Ref |
| 8529 | MK433274 | Unassigned | Unassigned | Unassigned | 685_0 | Clustered | 16 | VC_685_0 | 16 | 0.2972 | 1 | 0.2972 | 0 | 0 | 0 | 0.9167 | Ref |
| 8530 | MK433275 | Unassigned | Unassigned | Unassigned | 685_0 | Clustered | 16 | VC_685_0 | 16 | 0.2972 | 1 | 0.2972 | 0 | 0 | 0 | 0.9167 | Ref |
| 8531 | MK433276 | Unassigned | Unassigned | Unassigned | 982_0 | Clustered | 62 | VC_982_0 | 62 | 0.8646 | 1 | 0.8646 | 0 | 0 | 0 | 0.743 | Ref |
| 8532 | MK433277 | Unassigned | Unassigned | Unassigned | 761_0 | Clustered | 67 | VC_761_0 | 67 | 0.8947 | 1 | 0.8947 | 0 | 0 | 0 | 1 | Ref |
| 8533 | MK433278 | Unassigned | Unassigned | Unassigned | 982_0 | Clustered | 62 | VC_982_0 | 62 | 0.8646 | 1 | 0.8646 | 0 | 0 | 0 | 0.743 | Ref |
| 8534 | MK433279 | Unassigned | Unassigned | Unassigned | 761_0 | Clustered | 67 | VC_761_0 | 67 | 0.8947 | 1 | 0.8947 | 0 | 0 | 0 | 1 | Ref |
| 8535 | MK433577 | Unassigned | Unassigned | Unassigned | 6_0 | Clustered | 44 | VC_6_0 | 44 | 0.908 | 1 | 0.908 | 0 | 0 | 0 | 1 | Ref |
| 8536 | MK433578 | Unassigned | Unassigned | Unassigned | 6_0 | Clustered | 44 | VC_6_0 | 44 | 0.908 | 1 | 0.908 | 0 | 0 | 0 | 1 | Ref |
| 8537 | MK433579 | Unassigned | Unassigned | Unassigned | 242_1 | Clustered | 20 | VC_242_1 | 20 | 0.684 | 1 | 0.684 | 0 | 0 | 0 | 0.8368 | Ref |
| 8538 | MK433580 | Unassigned | Unassigned | Unassigned | 6_0 | Clustered | 44 | VC_6_0 | 44 | 0.908 | 1 | 0.908 | 0 | 0 | 0 | 1 | Ref |
| 8539 | MK433581 | Unassigned | Unassigned | Unassigned | 6_0 | Clustered | 44 | VC_6_0 | 44 | 0.908 | 1 | 0.908 | 0 | 0 | 0 | 1 | Ref |
| 8540 | MK433582 | Unassigned | Unassigned | Unassigned | 6_0 | Clustered | 44 | VC_6_0 | 44 | 0.908 | 1 | 0.908 | 0 | 0 | 0 | 1 | Ref |
| 8541 | MK443264 | Unassigned | Unassigned | Unassigned | 76_0 | Clustered | 37 | VC_76_0 | 37 | 0.9403 | 1 | 0.9403 | 0 | 0 | 0 | 1 | Ref |
| 8542 | MK443265 | Unassigned | Unassigned | Unassigned | 76_1 | Clustered/Singleton | 1 | VC_76_1 | 1 | 0 | 1 | 0 | 0 | 0 | 0 | 1 | Ref |
| 8543 | MK443266 | Unassigned | Unassigned | Unassigned | 167_0 | Clustered | 3 | VC_167_0 | 3 | 0.6952 | 0.99826575 | 0.694 | 0 | 0 | 0 | 1 | Ref |
| 8544 | MK448230 | Unassigned | Unassigned | Unassigned | 242_1 | Clustered | 20 | VC_242_1 | 20 | 0.684 | 1 | 0.684 | 0 | 0 | 0 | 0.8368 | Ref |
| 8545 | MK448231 | Unassigned | Unassigned | Unassigned | 31_0 | Clustered | 9 | VC_31_0 | 9 | 0.3435 | 1 | 0.3435 | 0 | 0 | 0 | 0.6944 | Ref |
| 8546 | MK448233 | Unassigned | Unassigned | Unassigned | 31_0 | Clustered | 9 | VC_31_0 | 9 | 0.3435 | 1 | 0.3435 | 0 | 0 | 0 | 0.6944 | Ref |
| 8547 | MK448234 | Unassigned | Unassigned | Unassigned | 35_0 | Clustered | 6 | VC_35_0 | 6 | 0.2254 | 1 | 0.2254 | 0 | 0 | 0 | 0.9333 | Ref |
| 8548 | MK448235 | Unassigned | Unassigned | Unassigned | 31_0 | Clustered | 9 | VC_31_0 | 9 | 0.3435 | 1 | 0.3435 | 0 | 0 | 0 | 0.6944 | Ref |
| 8549 | MK448237 | Unassigned | Unassigned | Unassigned | 35_0 | Clustered | 6 | VC_35_0 | 6 | 0.2254 | 1 | 0.2254 | 0 | 0 | 0 | 0.9333 | Ref |
| 8550 | MK448667 | Unassigned | Unassigned | Unassigned | 626_0 | Clustered | 23 | VC_626_0 | 23 | 0.66 | 1 | 0.66 | 0 | 0 | 0 | 0.8379 | Ref |
| 8551 | MK448671 | Unassigned | Unassigned | Unassigned | 412_0 | Clustered | 39 | VC_412_0 | 39 | 0.845 | 1 | 0.845 | 0 | 0 | 0 | 1 | Ref |
| 8552 | MK448673 | Unassigned | Unassigned | Unassigned | 775_1 | Clustered | 14 | VC_775_1 | 14 | 0.2797 | 1 | 0.2797 | 0 | 0 | 0 | 0.7582 | Ref |
| 8553 | MK448674 | Unassigned | Unassigned | Unassigned | 632_0 | Clustered | 37 | VC_632_0 | 37 | 0.581 | 1 | 0.581 | 0 | 0 | 0 | 0.9234 | Ref |
| 8554 | MK448675 | Unassigned | Unassigned | Unassigned | 293_3 | Clustered | 19 | VC_293_3 | 19 | 0.2627 | 1 | 0.2627 | 0 | 0 | 0 | 0.8187 | Ref |
| 8555 | MK448676 | Unassigned | Unassigned | Unassigned | 293_3 | Clustered | 19 | VC_293_3 | 19 | 0.2627 | 1 | 0.2627 | 0 | 0 | 0 | 0.8187 | Ref |
| 8556 | MK448678 | Unassigned | Unassigned | Unassigned | 773_0 | Clustered | 23 | VC_773_0 | 23 | 0.5514 | 1 | 0.5514 | 0 | 0 | 0 | 1 | Ref |
| 8557 | MK448680 | Unassigned | Unassigned | Unassigned | 419_0 | Clustered | 5 | VC_419_0 | 5 | 0.2498 | 0.99999997 | 0.2498 | 0 | 0 | 0 | 1 | Ref |
| 8558 | MK448681 | Unassigned | Unassigned | Unassigned | 636_0 | Clustered | 9 | VC_636_0 | 9 | 0.2422 | 1 | 0.2422 | 0 | 0 | 0 | 0.9722 | Ref |
| 8559 | MK448683 | Unassigned | Unassigned | Unassigned | 636_0 | Clustered | 9 | VC_636_0 | 9 | 0.2422 | 1 | 0.2422 | 0 | 0 | 0 | 0.9722 | Ref |
| 8560 | MK448684 | Unassigned | Unassigned | Unassigned | 419_0 | Clustered | 5 | VC_419_0 | 5 | 0.2498 | 0.99999997 | 0.2498 | 0 | 0 | 0 | 1 | Ref |
| 8561 | MK448685 | Unassigned | Unassigned | Unassigned | 317_0 | Clustered | 18 | VC_317_0 | 18 | 0.3304 | 1 | 0.3304 | 0 | 0 | 0 | 1 | Ref |
| 8562 | MK448686 | Unassigned | Unassigned | Unassigned | 416_0 | Clustered | 14 | VC_416_0 | 14 | 0.2592 | 1 | 0.2592 | 0 | 0 | 0 | 0.989 | Ref |
| 8563 | MK448688 | Unassigned | Unassigned | Unassigned | 293_3 | Clustered | 19 | VC_293_3 | 19 | 0.2627 | 1 | 0.2627 | 0 | 0 | 0 | 0.8187 | Ref |
| 8564 | MK448689 | Unassigned | Unassigned | Unassigned | 773_0 | Clustered | 23 | VC_773_0 | 23 | 0.5514 | 1 | 0.5514 | 0 | 0 | 0 | 1 | Ref |
| 8565 | MK448690 | Unassigned | Unassigned | Unassigned | 293_3 | Clustered | 19 | VC_293_3 | 19 | 0.2627 | 1 | 0.2627 | 0 | 0 | 0 | 0.8187 | Ref |
| 8566 | MK448692 | Unassigned | Unassigned | Unassigned | 431_0 | Clustered | 9 | VC_431_0 | 9 | 0.1673 | 1 | 0.1673 | 0 | 0 | 0 | 1 | Ref |
| 8567 | MK448693 | Unassigned | Unassigned | Unassigned | 773_0 | Clustered | 23 | VC_773_0 | 23 | 0.5514 | 1 | 0.5514 | 0 | 0 | 0 | 1 | Ref |
| 8568 | MK448694 | Unassigned | Unassigned | Unassigned | 293_3 | Clustered | 19 | VC_293_3 | 19 | 0.2627 | 1 | 0.2627 | 0 | 0 | 0 | 0.8187 | Ref |
| 8569 | MK448696 | Unassigned | Unassigned | Unassigned | 773_0 | Clustered | 23 | VC_773_0 | 23 | 0.5514 | 1 | 0.5514 | 0 | 0 | 0 | 1 | Ref |
| 8570 | MK448697 | Unassigned | Unassigned | Unassigned | 773_0 | Clustered | 23 | VC_773_0 | 23 | 0.5514 | 1 | 0.5514 | 0 | 0 | 0 | 1 | Ref |
| 8571 | MK448698 | Unassigned | Unassigned | Unassigned | 419_0 | Clustered | 5 | VC_419_0 | 5 | 0.2498 | 0.99999997 | 0.2498 | 0 | 0 | 0 | 1 | Ref |
| 8572 | MK448699 | Unassigned | Unassigned | Unassigned | 773_0 | Clustered | 23 | VC_773_0 | 23 | 0.5514 | 1 | 0.5514 | 0 | 0 | 0 | 1 | Ref |
| 8573 | MK448700 | Unassigned | Unassigned | Unassigned | 626_0 | Clustered | 23 | VC_626_0 | 23 | 0.66 | 1 | 0.66 | 0 | 0 | 0 | 0.8379 | Ref |
| 8574 | MK448702 | Unassigned | Unassigned | Unassigned | 431_0 | Clustered | 9 | VC_431_0 | 9 | 0.1673 | 1 | 0.1673 | 0 | 0 | 0 | 1 | Ref |
| 8575 | MK448704 | Unassigned | Unassigned | Unassigned | 292_0 | Clustered | 4 | VC_292_0 | 4 | 0.1677 | 0.99998671 | 0.1677 | 0 | 0 | 0 | 1 | Ref |
| 8576 | MK448706 | Unassigned | Unassigned | Unassigned | 317_1 | Clustered | 2 | VC_317_1 | 2 | 0.0186 | 0.95657151 | 0.0178 | 0 | 0 | 0 | 1 | Ref |
| 8577 | MK448707 | Unassigned | Unassigned | Unassigned | 292_0 | Clustered | 4 | VC_292_0 | 4 | 0.1677 | 0.99998671 | 0.1677 | 0 | 0 | 0 | 1 | Ref |
| 8578 | MK448708 | Unassigned | Unassigned | Unassigned | 412_0 | Clustered | 39 | VC_412_0 | 39 | 0.845 | 1 | 0.845 | 0 | 0 | 0 | 1 | Ref |
| 8579 | MK448709 | Unassigned | Unassigned | Unassigned | 292_0 | Clustered | 4 | VC_292_0 | 4 | 0.1677 | 0.99998671 | 0.1677 | 0 | 0 | 0 | 1 | Ref |
| 8580 | MK448710 | Unassigned | Unassigned | Unassigned | 292_0 | Clustered | 4 | VC_292_0 | 4 | 0.1677 | 0.99998671 | 0.1677 | 0 | 0 | 0 | 1 | Ref |
| 8581 | MK448711 | Unassigned | Unassigned | Unassigned | 431_0 | Clustered | 9 | VC_431_0 | 9 | 0.1673 | 1 | 0.1673 | 0 | 0 | 0 | 1 | Ref |
| 8582 | MK448714 | Unassigned | Unassigned | Unassigned | 412_0 | Clustered | 39 | VC_412_0 | 39 | 0.845 | 1 | 0.845 | 0 | 0 | 0 | 1 | Ref |
| 8583 | MK448716 | Unassigned | Unassigned | Unassigned | 431_0 | Clustered | 9 | VC_431_0 | 9 | 0.1673 | 1 | 0.1673 | 0 | 0 | 0 | 1 | Ref |
| 8584 | MK448720 | Unassigned | Unassigned | Unassigned | 626_0 | Clustered | 23 | VC_626_0 | 23 | 0.66 | 1 | 0.66 | 0 | 0 | 0 | 0.8379 | Ref |

| | | | | | | | | | | | | | | | | | |
|---|---|---|---|---|---|---|---|---|---|---|---|---|---|---|---|---|---|
| 8585 | MK448721 | Unassigned | Unassigned | Unassigned | 317_1 | Clustered | 2 | VC_317_1 | 2 | 0.0186 | 0.95657151 | 0.0178 | 0 | 0 | 0 | 1 | Ref |
| 8586 | MK448724 | Unassigned | Unassigned | Unassigned | 431_0 | Clustered | 9 | VC_431_0 | 9 | 0.1673 | 1 | 0.1673 | 0 | 0 | 0 | 1 | Ref |
| 8587 | MK448730 | Unassigned | Unassigned | Unassigned | 431_0 | Clustered | 9 | VC_431_0 | 9 | 0.1673 | 1 | 0.1673 | 0 | 0 | 0 | 1 | Ref |
| 8588 | MK448732 | Unassigned | Unassigned | Unassigned | 333_0 | Clustered | 25 | VC_333_0 | 25 | 0.4681 | 1 | 0.4681 | 0 | 0 | 0 | 1 | Ref |
| 8589 | MK448733 | Unassigned | Unassigned | Unassigned | 413_0 | Clustered | 3 | VC_413_0 | 3 | 0.0415 | 0.99841754 | 0.0415 | 0 | 0 | 0 | 1 | Ref |
| 8590 | MK448735 | Unassigned | Unassigned | Unassigned | 323_1 | Clustered | 9 | VC_323_1 | 9 | 0.1798 | 1 | 0.1798 | 0 | 0 | 0 | 0.75 | Ref |
| 8591 | MK448739 | Unassigned | Unassigned | Unassigned | 333_0 | Clustered | 25 | VC_333_0 | 25 | 0.4681 | 1 | 0.4681 | 0 | 0 | 0 | 1 | Ref |
| 8592 | MK448740 | Unassigned | Unassigned | Unassigned | 293_2 | Clustered | 2 | VC_293_2 | 2 | 0.0311 | 0.95710449 | 0.0298 | 0 | 0 | 0 | 1 | Ref |
| 8593 | MK448741 | Unassigned | Unassigned | Unassigned | 632_0 | Clustered | 37 | VC_632_0 | 37 | 0.581 | 1 | 0.581 | 0 | 0 | 0 | 0.9234 | Ref |
| 8594 | MK448744 | Unassigned | Unassigned | Unassigned | 431_0 | Clustered | 9 | VC_431_0 | 9 | 0.1673 | 1 | 0.1673 | 0 | 0 | 0 | 1 | Ref |
| 8595 | MK448745 | Unassigned | Unassigned | Unassigned | 431_0 | Clustered | 9 | VC_431_0 | 9 | 0.1673 | 1 | 0.1673 | 0 | 0 | 0 | 1 | Ref |
| 8596 | MK448750 | Unassigned | Unassigned | Unassigned | 293_0 | Clustered/Singleton | 1 | VC_293_0 | 1 | 0 | 1 | 0 | 0 | 0 | 0 | 1 | Ref |
| 8597 | MK448752 | Unassigned | Unassigned | Unassigned | 632_0 | Clustered | 37 | VC_632_0 | 37 | 0.581 | 1 | 0.581 | 0 | 0 | 0 | 0.9234 | Ref |
| 8598 | MK448757 | Unassigned | Unassigned | Unassigned | 323_1 | Clustered | 9 | VC_323_1 | 9 | 0.1798 | 1 | 0.1798 | 0 | 0 | 0 | 0.75 | Ref |
| 8599 | MK448758 | Unassigned | Unassigned | Unassigned | 323_1 | Clustered | 9 | VC_323_1 | 9 | 0.1798 | 1 | 0.1798 | 0 | 0 | 0 | 0.75 | Ref |
| 8600 | MK448759 | Unassigned | Unassigned | Unassigned | 323_1 | Clustered | 9 | VC_323_1 | 9 | 0.1798 | 1 | 0.1798 | 0 | 0 | 0 | 0.75 | Ref |
| 8601 | MK448762 | Unassigned | Unassigned | Unassigned | 416_0 | Clustered | 14 | VC_416_0 | 14 | 0.2592 | 1 | 0.2592 | 0 | 0 | 0 | 0.989 | Ref |
| 8602 | MK448763 | Unassigned | Unassigned | Unassigned | 317_0 | Clustered | 18 | VC_317_0 | 18 | 0.3304 | 1 | 0.3304 | 0 | 0 | 0 | 1 | Ref |
| 8603 | MK448765 | Unassigned | Unassigned | Unassigned | 775_1 | Clustered | 14 | VC_775_1 | 14 | 0.2797 | 1 | 0.2797 | 0 | 0 | 0 | 0.7582 | Ref |
| 8604 | MK448766 | Unassigned | Unassigned | Unassigned | 773_0 | Clustered | 23 | VC_773_0 | 23 | 0.5514 | 1 | 0.5514 | 0 | 0 | 0 | 1 | Ref |
| 8605 | MK448767 | Unassigned | Unassigned | Unassigned | 317_0 | Clustered | 18 | VC_317_0 | 18 | 0.3304 | 1 | 0.3304 | 0 | 0 | 0 | 1 | Ref |
| 8606 | MK448769 | Unassigned | Unassigned | Unassigned | 419_0 | Clustered | 5 | VC_419_0 | 5 | 0.2498 | 0.99999997 | 0.2498 | 0 | 0 | 0 | 1 | Ref |
| 8607 | MK448771 | Unassigned | Unassigned | Unassigned | 317_0 | Clustered | 18 | VC_317_0 | 18 | 0.3304 | 1 | 0.3304 | 0 | 0 | 0 | 1 | Ref |
| 8608 | MK448772 | Unassigned | Unassigned | Unassigned | 293_3 | Clustered | 19 | VC_293_3 | 19 | 0.2627 | 1 | 0.2627 | 0 | 0 | 0 | 0.8187 | Ref |
| 8609 | MK448773 | Unassigned | Unassigned | Unassigned | 317_0 | Clustered | 18 | VC_317_0 | 18 | 0.3304 | 1 | 0.3304 | 0 | 0 | 0 | 1 | Ref |
| 8610 | MK448774 | Unassigned | Unassigned | Unassigned | 317_0 | Clustered | 18 | VC_317_0 | 18 | 0.3304 | 1 | 0.3304 | 0 | 0 | 0 | 1 | Ref |
| 8611 | MK448775 | Unassigned | Unassigned | Unassigned | 775_1 | Clustered | 14 | VC_775_1 | 14 | 0.2797 | 1 | 0.2797 | 0 | 0 | 0 | 0.7582 | Ref |
| 8612 | MK448777 | Unassigned | Unassigned | Unassigned | 317_0 | Clustered | 18 | VC_317_0 | 18 | 0.3304 | 1 | 0.3304 | 0 | 0 | 0 | 1 | Ref |
| 8613 | MK448778 | Unassigned | Unassigned | Unassigned | 293_3 | Clustered | 19 | VC_293_3 | 19 | 0.2627 | 1 | 0.2627 | 0 | 0 | 0 | 0.8187 | Ref |
| 8614 | MK448779 | Unassigned | Unassigned | Unassigned | 773_0 | Clustered | 23 | VC_773_0 | 23 | 0.5514 | 1 | 0.5514 | 0 | 0 | 0 | 1 | Ref |
| 8615 | MK448782 | Unassigned | Unassigned | Unassigned | 293_3 | Clustered | 19 | VC_293_3 | 19 | 0.2627 | 1 | 0.2627 | 0 | 0 | 0 | 0.8187 | Ref |
| 8616 | MK448783 | Unassigned | Unassigned | Unassigned | 636_0 | Clustered | 9 | VC_636_0 | 9 | 0.2422 | 1 | 0.2422 | 0 | 0 | 0 | 0.9722 | Ref |
| 8617 | MK448784 | Unassigned | Unassigned | Unassigned | 773_0 | Clustered | 23 | VC_773_0 | 23 | 0.5514 | 1 | 0.5514 | 0 | 0 | 0 | 1 | Ref |
| 8618 | MK448786 | Unassigned | Unassigned | Unassigned | 317_0 | Clustered | 18 | VC_317_0 | 18 | 0.3304 | 1 | 0.3304 | 0 | 0 | 0 | 1 | Ref |
| 8619 | MK448788 | Unassigned | Unassigned | Unassigned | 416_0 | Clustered | 14 | VC_416_0 | 14 | 0.2592 | 1 | 0.2592 | 0 | 0 | 0 | 0.989 | Ref |
| 8620 | MK448789 | Unassigned | Unassigned | Unassigned | 773_0 | Clustered | 23 | VC_773_0 | 23 | 0.5514 | 1 | 0.5514 | 0 | 0 | 0 | 1 | Ref |
| 8621 | MK448790 | Unassigned | Unassigned | Unassigned | 636_0 | Clustered | 9 | VC_636_0 | 9 | 0.2422 | 1 | 0.2422 | 0 | 0 | 0 | 0.9722 | Ref |
| 8622 | MK448791 | Unassigned | Unassigned | Unassigned | 416_0 | Clustered | 14 | VC_416_0 | 14 | 0.2592 | 1 | 0.2592 | 0 | 0 | 0 | 0.989 | Ref |
| 8623 | MK448793 | Unassigned | Unassigned | Unassigned | 416_0 | Clustered | 14 | VC_416_0 | 14 | 0.2592 | 1 | 0.2592 | 0 | 0 | 0 | 0.989 | Ref |
| 8624 | MK448794 | Unassigned | Unassigned | Unassigned | 775_1 | Clustered | 14 | VC_775_1 | 14 | 0.2797 | 1 | 0.2797 | 0 | 0 | 0 | 0.7582 | Ref |
| 8625 | MK448795 | Unassigned | Unassigned | Unassigned | 775_1 | Clustered | 14 | VC_775_1 | 14 | 0.2797 | 1 | 0.2797 | 0 | 0 | 0 | 0.7582 | Ref |
| 8626 | MK448797 | Unassigned | Unassigned | Unassigned | 773_0 | Clustered | 23 | VC_773_0 | 23 | 0.5514 | 1 | 0.5514 | 0 | 0 | 0 | 1 | Ref |
| 8627 | MK448798 | Unassigned | Unassigned | Unassigned | 636_0 | Clustered | 9 | VC_636_0 | 9 | 0.2422 | 1 | 0.2422 | 0 | 0 | 0 | 0.9722 | Ref |
| 8628 | MK448804 | Unassigned | Unassigned | Unassigned | 335_0 | Clustered | 6 | VC_335_0 | 6 | 0.3678 | 1 | 0.3678 | 0 | 0 | 0 | 1 | Ref |
| 8629 | MK448805 | Unassigned | Unassigned | Unassigned | 335_0 | Clustered | 6 | VC_335_0 | 6 | 0.3678 | 1 | 0.3678 | 0 | 0 | 0 | 1 | Ref |
| 8630 | MK448807 | Unassigned | Unassigned | Unassigned | 335_0 | Clustered | 6 | VC_335_0 | 6 | 0.3678 | 1 | 0.3678 | 0 | 0 | 0 | 1 | Ref |
| 8631 | MK448808 | Unassigned | Unassigned | Unassigned | 335_0 | Clustered | 6 | VC_335_0 | 6 | 0.3678 | 1 | 0.3678 | 0 | 0 | 0 | 1 | Ref |
| 8632 | MK448811 | Unassigned | Unassigned | Unassigned | 632_0 | Clustered | 37 | VC_632_0 | 37 | 0.581 | 1 | 0.581 | 0 | 0 | 0 | 0.9234 | Ref |
| 8633 | MK448814 | Unassigned | Unassigned | Unassigned | 317_2 | Clustered | 20 | VC_317_2 | 20 | 0.1976 | 1 | 0.1976 | 0 | 0 | 0 | 0.6842 | Ref |
| 8634 | MK448815 | Unassigned | Unassigned | Unassigned | 335_0 | Clustered | 6 | VC_335_0 | 6 | 0.3678 | 1 | 0.3678 | 0 | 0 | 0 | 1 | Ref |
| 8635 | MK448818 | Unassigned | Unassigned | Unassigned | 623_0 | Clustered/Singleton | 1 | VC_623_0 | 1 | 0 | 1 | 0 | 0 | 0 | 0 | 1 | Ref |
| 8636 | MK448819 | Unassigned | Unassigned | Unassigned | 632_0 | Clustered | 37 | VC_632_0 | 37 | 0.581 | 1 | 0.581 | 0 | 0 | 0 | 0.9234 | Ref |
| 8637 | MK448821 | Unassigned | Unassigned | Unassigned | 323_0 | Clustered | 2 | VC_323_0 | 2 | 0.0281 | 0.95656566 | 0.0269 | 0 | 0 | 0 | 1 | Ref |
| 8638 | MK448822 | Unassigned | Unassigned | Unassigned | 323_0 | Clustered | 2 | VC_323_0 | 2 | 0.0281 | 0.95656566 | 0.0269 | 0 | 0 | 0 | 1 | Ref |
| 8639 | MK448824 | Unassigned | Unassigned | Unassigned | 259_0 | Clustered | 3 | VC_259_0 | 3 | 0.1148 | 0.99858602 | 0.1146 | 0 | 0 | 0 | 1 | Ref |
| 8640 | MK448828 | Unassigned | Unassigned | Unassigned | 632_0 | Clustered | 37 | VC_632_0 | 37 | 0.581 | 1 | 0.581 | 0 | 0 | 0 | 0.9234 | Ref |
| 8641 | MK448831 | Unassigned | Unassigned | Unassigned | 412_0 | Clustered | 39 | VC_412_0 | 39 | 0.845 | 1 | 0.845 | 0 | 0 | 0 | 1 | Ref |
| 8642 | MK448832 | Unassigned | Unassigned | Unassigned | 294_0 | Clustered | 2 | VC_294_0 | 2 | 0.0836 | 0.95731776 | 0.0801 | 0 | 0 | 0 | 1 | Ref |
| 8643 | MK448833 | Unassigned | Unassigned | Unassigned | 636_0 | Clustered | 9 | VC_636_0 | 9 | 0.2422 | 1 | 0.2422 | 0 | 0 | 0 | 0.9722 | Ref |
| 8644 | MK448834 | Unassigned | Unassigned | Unassigned | 412_0 | Clustered | 39 | VC_412_0 | 39 | 0.845 | 1 | 0.845 | 0 | 0 | 0 | 1 | Ref |
| 8645 | MK448839 | Unassigned | Unassigned | Unassigned | 626_0 | Clustered | 23 | VC_626_0 | 23 | 0.66 | 1 | 0.66 | 0 | 0 | 0 | 0.8379 | Ref |
| 8646 | MK448841 | Unassigned | Unassigned | Unassigned | 626_0 | Clustered | 23 | VC_626_0 | 23 | 0.66 | 1 | 0.66 | 0 | 0 | 0 | 0.8379 | Ref |
| 8647 | MK448843 | Unassigned | Unassigned | Unassigned | 626_0 | Clustered | 23 | VC_626_0 | 23 | 0.66 | 1 | 0.66 | 0 | 0 | 0 | 0.8379 | Ref |
| 8648 | MK448845 | Unassigned | Unassigned | Unassigned | 317_0 | Clustered | 18 | VC_317_0 | 18 | 0.3304 | 1 | 0.3304 | 0 | 0 | 0 | 1 | Ref |
| 8649 | MK448850 | Unassigned | Unassigned | Unassigned | 293_3 | Clustered | 19 | VC_293_3 | 19 | 0.2627 | 1 | 0.2627 | 0 | 0 | 0 | 0.8187 | Ref |
| 8650 | MK448852 | Unassigned | Unassigned | Unassigned | 773_0 | Clustered | 23 | VC_773_0 | 23 | 0.5514 | 1 | 0.5514 | 0 | 0 | 0 | 1 | Ref |
| 8651 | MK448853 | Unassigned | Unassigned | Unassigned | 317_0 | Clustered | 18 | VC_317_0 | 18 | 0.3304 | 1 | 0.3304 | 0 | 0 | 0 | 1 | Ref |
| 8652 | MK448854 | Unassigned | Unassigned | Unassigned | 775_0 | Clustered | 3 | VC_775_0 | 3 | 0.0364 | 0.99850073 | 0.0364 | 0 | 0 | 0 | 1 | Ref |
| 8653 | MK448855 | Unassigned | Unassigned | Unassigned | 317_0 | Clustered | 18 | VC_317_0 | 18 | 0.3304 | 1 | 0.3304 | 0 | 0 | 0 | 1 | Ref |
| 8654 | MK448858 | Unassigned | Unassigned | Unassigned | 775_1 | Clustered | 14 | VC_775_1 | 14 | 0.2797 | 1 | 0.2797 | 0 | 0 | 0 | 0.7582 | Ref |
| 8655 | MK448859 | Unassigned | Unassigned | Unassigned | 293_3 | Clustered | 19 | VC_293_3 | 19 | 0.2627 | 1 | 0.2627 | 0 | 0 | 0 | 0.8187 | Ref |
| 8656 | MK448861 | Unassigned | Unassigned | Unassigned | 412_0 | Clustered | 39 | VC_412_0 | 39 | 0.845 | 1 | 0.845 | 0 | 0 | 0 | 1 | Ref |
| 8657 | MK448862 | Unassigned | Unassigned | Unassigned | 416_0 | Clustered | 14 | VC_416_0 | 14 | 0.2592 | 1 | 0.2592 | 0 | 0 | 0 | 0.989 | Ref |
| 8658 | MK448864 | Unassigned | Unassigned | Unassigned | 775_1 | Clustered | 14 | VC_775_1 | 14 | 0.2797 | 1 | 0.2797 | 0 | 0 | 0 | 0.7582 | Ref |
| 8659 | MK448866 | Unassigned | Unassigned | Unassigned | 773_0 | Clustered | 23 | VC_773_0 | 23 | 0.5514 | 1 | 0.5514 | 0 | 0 | 0 | 1 | Ref |
| 8660 | MK448867 | Unassigned | Unassigned | Unassigned | 773_0 | Clustered | 23 | VC_773_0 | 23 | 0.5514 | 1 | 0.5514 | 0 | 0 | 0 | 1 | Ref |
| 8661 | MK448868 | Unassigned | Unassigned | Unassigned | 773_0 | Clustered | 23 | VC_773_0 | 23 | 0.5514 | 1 | 0.5514 | 0 | 0 | 0 | 1 | Ref |
| 8662 | MK448871 | Unassigned | Unassigned | Unassigned | 773_0 | Clustered | 23 | VC_773_0 | 23 | 0.5514 | 1 | 0.5514 | 0 | 0 | 0 | 1 | Ref |
| 8663 | MK448874 | Unassigned | Unassigned | Unassigned | 412_0 | Clustered | 39 | VC_412_0 | 39 | 0.845 | 1 | 0.845 | 0 | 0 | 0 | 1 | Ref |
| 8664 | MK448875 | Unassigned | Unassigned | Unassigned | 412_0 | Clustered | 39 | VC_412_0 | 39 | 0.845 | 1 | 0.845 | 0 | 0 | 0 | 1 | Ref |
| 8665 | MK448877 | Unassigned | Unassigned | Unassigned | 293_3 | Clustered | 19 | VC_293_3 | 19 | 0.2627 | 1 | 0.2627 | 0 | 0 | 0 | 0.8187 | Ref |
| 8666 | MK448881 | Unassigned | Unassigned | Unassigned | 317_2 | Clustered | 20 | VC_317_2 | 20 | 0.1976 | 1 | 0.1976 | 0 | 0 | 0 | 0.6842 | Ref |
| 8667 | MK448882 | Unassigned | Unassigned | Unassigned | 317_2 | Clustered | 20 | VC_317_2 | 20 | 0.1976 | 1 | 0.1976 | 0 | 0 | 0 | 0.6842 | Ref |
| 8668 | MK448886 | Unassigned | Unassigned | Unassigned | 412_0 | Clustered | 39 | VC_412_0 | 39 | 0.845 | 1 | 0.845 | 0 | 0 | 0 | 1 | Ref |
| 8669 | MK448887 | Unassigned | Unassigned | Unassigned | 412_0 | Clustered | 39 | VC_412_0 | 39 | 0.845 | 1 | 0.845 | 0 | 0 | 0 | 1 | Ref |
| 8670 | MK448890 | Unassigned | Unassigned | Unassigned | 625_0 | Clustered | 2 | VC_625_0 | 2 | 0.0259 | 0.92480562 | 0.0239 | 0 | 0 | 0 | 1 | Ref |
| 8671 | MK448893 | Unassigned | Unassigned | Unassigned | 323_1 | Clustered | 9 | VC_323_1 | 9 | 0.1798 | 1 | 0.1798 | 0 | 0 | 0 | 0.75 | Ref |
| 8672 | MK448895 | Unassigned | Unassigned | Unassigned | 419_0 | Clustered | 5 | VC_419_0 | 5 | 0.2498 | 0.99999997 | 0.2498 | 0 | 0 | 0 | 1 | Ref |
| 8673 | MK448896 | Unassigned | Unassigned | Unassigned | 317_2 | Clustered | 20 | VC_317_2 | 20 | 0.1976 | 1 | 0.1976 | 0 | 0 | 0 | 0.6842 | Ref |
| 8674 | MK448897 | Unassigned | Unassigned | Unassigned | 412_0 | Clustered | 39 | VC_412_0 | 39 | 0.845 | 1 | 0.845 | 0 | 0 | 0 | 1 | Ref |
| 8675 | MK448898 | Unassigned | Unassigned | Unassigned | 412_0 | Clustered | 39 | VC_412_0 | 39 | 0.845 | 1 | 0.845 | 0 | 0 | 0 | 1 | Ref |
| 8676 | MK448899 | Unassigned | Unassigned | Unassigned | 253_0 | Clustered/Singleton | 1 | VC_253_0 | 1 | 0 | 1 | 0 | 0 | 0 | 0 | 1 | Ref |
| 8677 | MK448900 | Unassigned | Unassigned | Unassigned | 317_4 | Clustered/Singleton | 1 | VC_317_4 | 1 | 0 | 1 | 0 | 0 | 0 | 0 | 1 | Ref |
| 8678 | MK448901 | Unassigned | Unassigned | Unassigned | 333_0 | Clustered | 25 | VC_333_0 | 25 | 0.4681 | 1 | 0.4681 | 0 | 0 | 0 | 1 | Ref |
| 8679 | MK448902 | Unassigned | Unassigned | Unassigned | 137_1 | Clustered/Singleton | 1 | VC_137_1 | 1 | 0 | 1 | 0 | 0 | 0 | 0 | 1 | Ref |
| 8680 | MK448904 | Unassigned | Unassigned | Unassigned | 413_0 | Clustered | 3 | VC_413_0 | 3 | 0.0415 | 0.99841754 | 0.0415 | 0 | 0 | 0 | 1 | Ref |
| 8681 | MK448905 | Unassigned | Unassigned | Unassigned | 412_0 | Clustered | 39 | VC_412_0 | 39 | 0.845 | 1 | 0.845 | 0 | 0 | 0 | 1 | Ref |
| 8682 | MK448909 | Unassigned | Unassigned | Unassigned | 317_2 | Clustered | 20 | VC_317_2 | 20 | 0.1976 | 1 | 0.1976 | 0 | 0 | 0 | 0.6842 | Ref |
| 8683 | MK448910 | Unassigned | Unassigned | Unassigned | 137_0 | Clustered | 3 | VC_137_0 | 3 | 0.1129 | 0.998547 | 0.1128 | 0 | 0 | 0 | 1 | Ref |
| 8684 | MK448912 | Unassigned | Unassigned | Unassigned | 333_0 | Clustered | 25 | VC_333_0 | 25 | 0.4681 | 1 | 0.4681 | 0 | 0 | 0 | 1 | Ref |
| 8685 | MK448914 | Unassigned | Unassigned | Unassigned | 333_0 | Clustered | 25 | VC_333_0 | 25 | 0.4681 | 1 | 0.4681 | 0 | 0 | 0 | 1 | Ref |
| 8686 | MK448915 | Unassigned | Unassigned | Unassigned | 323_1 | Clustered | 9 | VC_323_1 | 9 | 0.1798 | 1 | 0.1798 | 0 | 0 | 0 | 0.75 | Ref |
| 8687 | MK448917 | Unassigned | Unassigned | Unassigned | 293_2 | Clustered | 2 | VC_293_2 | 2 | 0.0311 | 0.95710449 | 0.0298 | 0 | 0 | 0 | 1 | Ref |
| 8688 | MK448918 | Unassigned | Unassigned | Unassigned | 333_0 | Clustered | 25 | VC_333_0 | 25 | 0.4681 | 1 | 0.4681 | 0 | 0 | 0 | 1 | Ref |
| 8689 | MK448920 | Unassigned | Unassigned | Unassigned | 333_0 | Clustered | 25 | VC_333_0 | 25 | 0.4681 | 1 | 0.4681 | 0 | 0 | 0 | 1 | Ref |
| 8690 | MK448923 | Unassigned | Unassigned | Unassigned | 431_0 | Clustered | 9 | VC_431_0 | 9 | 0.1673 | 1 | 0.1673 | 0 | 0 | 0 | 1 | Ref |
| 8691 | MK448925 | Unassigned | Unassigned | Unassigned | 412_0 | Clustered | 39 | VC_412_0 | 39 | 0.845 | 1 | 0.845 | 0 | 0 | 0 | 1 | Ref |
| 8692 | MK448927 | Unassigned | Unassigned | Unassigned | 412_0 | Clustered | 39 | VC_412_0 | 39 | 0.845 | 1 | 0.845 | 0 | 0 | 0 | 1 | Ref |
| 8693 | MK448930 | Unassigned | Unassigned | Unassigned | 625_0 | Clustered | 2 | VC_625_0 | 2 | 0.0259 | 0.92480562 | 0.0239 | 0 | 0 | 0 | 1 | Ref |
| 8694 | MK448931 | Unassigned | Unassigned | Unassigned | 293_3 | Clustered | 19 | VC_293_3 | 19 | 0.2627 | 1 | 0.2627 | 0 | 0 | 0 | 0.8187 | Ref |
| 8695 | MK448933 | Unassigned | Unassigned | Unassigned | 775_0 | Clustered | 3 | VC_775_0 | 3 | 0.0364 | 0.99850073 | 0.0364 | 0 | 0 | 0 | 1 | Ref |
| 8696 | MK448934 | Unassigned | Unassigned | Unassigned | 626_0 | Clustered | 23 | VC_626_0 | 23 | 0.66 | 1 | 0.66 | 0 | 0 | 0 | 0.8379 | Ref |
| 8697 | MK448939 | Unassigned | Unassigned | Unassigned | 323_1 | Clustered | 9 | VC_323_1 | 9 | 0.1798 | 1 | 0.1798 | 0 | 0 | 0 | 0.75 | Ref |
| 8698 | MK448941 | Unassigned | Unassigned | Unassigned | 775_1 | Clustered | 14 | VC_775_1 | 14 | 0.2797 | 1 | 0.2797 | 0 | 0 | 0 | 0.7582 | Ref |
| 8699 | MK448942 | Unassigned | Unassigned | Unassigned | 317_0 | Clustered | 18 | VC_317_0 | 18 | 0.3304 | 1 | 0.3304 | 0 | 0 | 0 | 1 | Ref |
| 8700 | MK448943 | Unassigned | Unassigned | Unassigned | 317_0 | Clustered | 18 | VC_317_0 | 18 | 0.3304 | 1 | 0.3304 | 0 | 0 | 0 | 1 | Ref |
| 8701 | MK448944 | Unassigned | Unassigned | Unassigned | 775_1 | Clustered | 14 | VC_775_1 | 14 | 0.2797 | 1 | 0.2797 | 0 | 0 | 0 | 0.7582 | Ref |
| 8702 | MK448945 | Unassigned | Unassigned | Unassigned | 775_1 | Clustered | 14 | VC_775_1 | 14 | 0.2797 | 1 | 0.2797 | 0 | 0 | 0 | 0.7582 | Ref |
| 8703 | MK448946 | Unassigned | Unassigned | Unassigned | 775_1 | Clustered | 14 | VC_775_1 | 14 | 0.2797 | 1 | 0.2797 | 0 | 0 | 0 | 0.7582 | Ref |
| 8704 | MK448947 | Unassigned | Unassigned | Unassigned | 773_0 | Clustered | 23 | VC_773_0 | 23 | 0.5514 | 1 | 0.5514 | 0 | 0 | 0 | 1 | Ref |
| 8705 | MK448949 | Unassigned | Unassigned | Unassigned | 416_0 | Clustered | 14 | VC_416_0 | 14 | 0.2592 | 1 | 0.2592 | 0 | 0 | 0 | 0.989 | Ref |
| 8706 | MK448950 | Unassigned | Unassigned | Unassigned | 775_0 | Clustered | 3 | VC_775_0 | 3 | 0.0364 | 0.99850073 | 0.0364 | 0 | 0 | 0 | 1 | Ref |
| 8707 | MK448951 | Unassigned | Unassigned | Unassigned | 416_0 | Clustered | 14 | VC_416_0 | 14 | 0.2592 | 1 | 0.2592 | 0 | 0 | 0 | 0.989 | Ref |
| 8708 | MK448952 | Unassigned | Unassigned | Unassigned | 773_0 | Clustered | 23 | VC_773_0 | 23 | 0.5514 | 1 | 0.5514 | 0 | 0 | 0 | 1 | Ref |
| 8709 | MK448953 | Unassigned | Unassigned | Unassigned | 775_1 | Clustered | 14 | VC_775_1 | 14 | 0.2797 | 1 | 0.2797 | 0 | 0 | 0 | 0.7582 | Ref |
| 8710 | MK448954 | Unassigned | Unassigned | Unassigned | 416_0 | Clustered | 14 | VC_416_0 | 14 | 0.2592 | 1 | 0.2592 | 0 | 0 | 0 | 0.989 | Ref |
| 8711 | MK448955 | Unassigned | Unassigned | Unassigned | 293_3 | Clustered | 19 | VC_293_3 | 19 | 0.2627 | 1 | 0.2627 | 0 | 0 | 0 | 0.8187 | Ref |
| 8712 | MK448957 | Unassigned | Unassigned | Unassigned | 775_1 | Clustered | 14 | VC_775_1 | 14 | 0.2797 | 1 | 0.2797 | 0 | 0 | 0 | 0.7582 | Ref |
| 8713 | MK448958 | Unassigned | Unassigned | Unassigned | 416_0 | Clustered | 14 | VC_416_0 | 14 | 0.2592 | 1 | 0.2592 | 0 | 0 | 0 | 0.989 | Ref |
| 8714 | MK448959 | Unassigned | Unassigned | Unassigned | 416_0 | Clustered | 14 | VC_416_0 | 14 | 0.2592 | 1 | 0.2592 | 0 | 0 | 0 | 0.989 | Ref |
| 8715 | MK448960 | Unassigned | Unassigned | Unassigned | 416_0 | Clustered | 14 | VC_416_0 | 14 | 0.2592 | 1 | 0.2592 | 0 | 0 | 0 | 0.989 | Ref |
| 8716 | MK448961 | Unassigned | Unassigned | Unassigned | 773_0 | Clustered | 23 | VC_773_0 | 23 | 0.5514 | 1 | 0.5514 | 0 | 0 | 0 | 1 | Ref |
| 8717 | MK448962 | Unassigned | Unassigned | Unassigned | 293_3 | Clustered | 19 | VC_293_3 | 19 | 0.2627 | 1 | 0.2627 | 0 | 0 | 0 | 0.8187 | Ref |
| 8718 | MK448963 | Unassigned | Unassigned | Unassigned | 317_0 | Clustered | 18 | VC_317_0 | 18 | 0.3304 | 1 | 0.3304 | 0 | 0 | 0 | 1 | Ref |
| 8719 | MK448964 | Unassigned | Unassigned | Unassigned | 317_0 | Clustered | 18 | VC_317_0 | 18 | 0.3304 | 1 | 0.3304 | 0 | 0 | 0 | 1 | Ref |
| 8720 | MK448965 | Unassigned | Unassigned | Unassigned | 416_0 | Clustered | 14 | VC_416_0 | 14 | 0.2592 | 1 | 0.2592 | 0 | 0 | 0 | 0.989 | Ref |
| 8721 | MK448966 | Unassigned | Unassigned | Unassigned | 293_3 | Clustered | 19 | VC_293_3 | 19 | 0.2627 | 1 | 0.2627 | 0 | 0 | 0 | 0.8187 | Ref |
| 8722 | MK448967 | Unassigned | Unassigned | Unassigned | 636_0 | Clustered | 9 | VC_636_0 | 9 | 0.2422 | 1 | 0.2422 | 0 | 0 | 0 | 0.9722 | Ref |
| 8723 | MK448968 | Unassigned | Unassigned | Unassigned | 773_0 | Clustered | 23 | VC_773_0 | 23 | 0.5514 | 1 | 0.5514 | 0 | 0 | 0 | 1 | Ref |
| 8724 | MK448969 | Unassigned | Unassigned | Unassigned | 317_0 | Clustered | 18 | VC_317_0 | 18 | 0.3304 | 1 | 0.3304 | 0 | 0 | 0 | 1 | Ref |
| 8725 | MK448970 | Unassigned | Unassigned | Unassigned | 775_1 | Clustered | 14 | VC_775_1 | 14 | 0.2797 | 1 | 0.2797 | 0 | 0 | 0 | 0.7582 | Ref |
| 8726 | MK448972 | Unassigned | Unassigned | Unassigned | 317_0 | Clustered | 18 | VC_317_0 | 18 | 0.3304 | 1 | 0.3304 | 0 | 0 | 0 | 1 | Ref |
| 8727 | MK448973 | Unassigned | Unassigned | Unassigned | 293_3 | Clustered | 19 | VC_293_3 | 19 | 0.2627 | 1 | 0.2627 | 0 | 0 | 0 | 0.8187 | Ref |
| 8728 | MK448974 | Unassigned | Unassigned | Unassigned | 773_0 | Clustered | 23 | VC_773_0 | 23 | 0.5514 | 1 | 0.5514 | 0 | 0 | 0 | 1 | Ref |
| 8729 | MK448975 | Unassigned | Unassigned | Unassigned | 293_3 | Clustered | 19 | VC_293_3 | 19 | 0.2627 | 1 | 0.2627 | 0 | 0 | 0 | 0.8187 | Ref |
| 8730 | MK448976 | Unassigned | Unassigned | Unassigned | 773_0 | Clustered | 23 | VC_773_0 | 23 | 0.5514 | 1 | 0.5514 | 0 | 0 | 0 | 1 | Ref |
| 8731 | MK448977 | Unassigned | Unassigned | Unassigned | 416_0 | Clustered | 14 | VC_416_0 | 14 | 0.2592 | 1 | 0.2592 | 0 | 0 | 0 | 0.989 | Ref |
| 8732 | MK448982 | Unassigned | Unassigned | Unassigned | 318_0 | Clustered | 3 | VC_318_0 | 3 | 0.0414 | 0.98579852 | 0.0408 | 0 | 0 | 0 | 1 | Ref |
| 8733 | MK448983 | Unassigned | Unassigned | Unassigned | 632_0 | Clustered | 37 | VC_632_0 | 37 | 0.581 | 1 | 0.581 | 0 | 0 | 0 | 0.9234 | Ref |
| 8734 | MK448988 | Unassigned | Unassigned | Unassigned | 317_2 | Clustered | 20 | VC_317_2 | 20 | 0.1976 | 1 | 0.1976 | 0 | 0 | 0 | 0.6842 | Ref |
| 8735 | MK448989 | Unassigned | Unassigned | Unassigned | 317_2 | Clustered | 20 | VC_317_2 | 20 | 0.1976 | 1 | 0.1976 | 0 | 0 | 0 | 0.6842 | Ref |
| 8736 | MK448991 | Unassigned | Unassigned | Unassigned | 335_0 | Clustered | 6 | VC_335_0 | 6 | 0.3678 | 1 | 0.3678 | 0 | 0 | 0 | 1 | Ref |
| 8737 | MK448995 | Unassigned | Unassigned | Unassigned | 293_1 | Clustered/Singleton | 1 | VC_293_1 | 1 | 0 | 1 | 0 | 0 | 0 | 0 | 1 | Ref |
| 8738 | MK448997 | Unassigned | Unassigned | Unassigned | 626_0 | Clustered | 23 | VC_626_0 | 23 | 0.66 | 1 | 0.66 | 0 | 0 | 0 | 0.8379 | Ref |
| 8739 | MK449000 | Unassigned | Unassigned | Unassigned | 334_0 | Clustered/Singleton | 1 | VC_334_0 | 1 | 0 | 1 | 0 | 0 | 0 | 0 | 1 | Ref |
| 8740 | MK449002 | Unassigned | Unassigned | Unassigned | 259_0 | Clustered | 3 | VC_259_0 | 3 | 0.1148 | 0.99858602 | 0.1146 | 0 | 0 | 0 | 1 | Ref |
| 8741 | MK449003 | Unassigned | Unassigned | Unassigned | 259_0 | Clustered | 3 | VC_259_0 | 3 | 0.1148 | 0.99858602 | 0.1146 | 0 | 0 | 0 | 1 | Ref |
| 8742 | MK449004 | Unassigned | Unassigned | Unassigned | 293_3 | Clustered | 19 | VC_293_3 | 19 | 0.2627 | 1 | 0.2627 | 0 | 0 | 0 | 0.8187 | Ref |
| 8743 | MK449006 | Unassigned | Unassigned | Unassigned | 412_0 | Clustered | 39 | VC_412_0 | 39 | 0.845 | 1 | 0.845 | 0 | 0 | 0 | 1 | Ref |
| 8744 | MK449008 | Unassigned | Unassigned | Unassigned | 294_0 | Clustered | 2 | VC_294_0 | 2 | 0.0836 | 0.95731776 | 0.0801 | 0 | 0 | 0 | 1 | Ref |
| 8745 | MK449009 | Unassigned | Unassigned | Unassigned | 412_0 | Clustered | 39 | VC_412_0 | 39 | 0.845 | 1 | 0.845 | 0 | 0 | 0 | 1 | Ref |
| 8746 | MK449010 | Unassigned | Unassigned | Unassigned | 636_0 | Clustered | 9 | VC_636_0 | 9 | 0.2422 | 1 | 0.2422 | 0 | 0 | 0 | 0.9722 | Ref |

| | | | | | | | | | | | | | | | | | |
|---|---|---|---|---|---|---|---|---|---|---|---|---|---|---|---|---|---|
| 8747 | MK449012 | Unassigned | Unassigned | Unassigned | 317_0 | Clustered | 18 | VC_317_0 | 18 | 0.3304 | 1 | 0.3304 | 0 | 0 | 0 | 1 | Ref |
| 8748 | MK450421 | Unassigned | Unassigned | Unassigned | 373_0 | Clustered | 10 | VC_373_0 | 10 | 0.7471 | 1 | 0.7471 | 0 | 0 | 0 | 1 | Ref |
| 8749 | MK450422 | Unassigned | Unassigned | Unassigned | 558_65 | Clustered | 2 | VC_558_65 | 2 | 0.0032 | 0.61174182 | 0.0019 | 0 | 0 | 0 | 1 | Ref |
| 8750 | MK450423 | Unassigned | Unassigned | Unassigned | 558_22 | Clustered | 4 | VC_558_22 | 4 | 0.01 | 0.99312819 | 0.0099 | 0 | 0 | 0 | 0.8333 | Ref |
| 8751 | MK450424 | Unassigned | Unassigned | Unassigned | 558_41 | Clustered/Singleton | 1 | VC_558_41 | 1 | 0 | 1 | 0 | 0 | 0 | 0 | 1 | Ref |
| 8752 | MK450425 | Unassigned | Unassigned | Unassigned | 558_57 | Clustered/Singleton | 1 | VC_558_57 | 1 | 0 | 1 | 0 | 0 | 0 | 0 | 1 | Ref |
| 8753 | MK450426 | Unassigned | Unassigned | Unassigned | 373_0 | Clustered | 10 | VC_373_0 | 10 | 0.7471 | 1 | 0.7471 | 0 | 0 | 0 | 1 | Ref |
| 8754 | MK450427 | Unassigned | Unassigned | Unassigned | 558_14 | Clustered | 8 | VC_558_14 | 8 | 0.0228 | 0.99945246 | 0.0228 | 0 | 0 | 0 | 0.8214 | Ref |
| 8755 | MK450428 | Unassigned | Unassigned | Unassigned | 866_1 | Clustered | 20 | VC_866_1 | 20 | 0.4814 | 1 | 0.4814 | 0 | 0 | 0 | 1 | Ref |
| 8756 | MK450429 | Unassigned | Unassigned | Unassigned | 558_50 | Clustered | 15 | VC_558_50 | 15 | 0.0468 | 1 | 0.0468 | 0 | 0 | 0 | 0.9238 | Ref |
| 8757 | MK450430 | Unassigned | Unassigned | Unassigned | 866_1 | Clustered | 20 | VC_866_1 | 20 | 0.4814 | 1 | 0.4814 | 0 | 0 | 0 | 1 | Ref |
| 8758 | MK450431 | Unassigned | Unassigned | Unassigned | 373_0 | Clustered | 10 | VC_373_0 | 10 | 0.7471 | 1 | 0.7471 | 0 | 0 | 0 | 1 | Ref |
| 8759 | MK450432 | Unassigned | Unassigned | Unassigned | 558_20 | Clustered | 17 | VC_558_20 | 17 | 0.054 | 1 | 0.054 | 0 | 0 | 0 | 0.9926 | Ref |
| 8760 | MK450433 | Unassigned | Unassigned | Unassigned | 373_0 | Clustered | 10 | VC_373_0 | 10 | 0.7471 | 1 | 0.7471 | 0 | 0 | 0 | 1 | Ref |
| 8761 | MK450434 | Unassigned | Unassigned | Unassigned | 558_50 | Clustered | 15 | VC_558_50 | 15 | 0.0468 | 1 | 0.0468 | 0 | 0 | 0 | 0.9238 | Ref |
| 8762 | MK450435 | Unassigned | Unassigned | Unassigned | 866_1 | Clustered | 20 | VC_866_1 | 20 | 0.4814 | 1 | 0.4814 | 0 | 0 | 0 | 1 | Ref |
| 8763 | MK450436 | Unassigned | Unassigned | Unassigned | 866_1 | Clustered | 20 | VC_866_1 | 20 | 0.4814 | 1 | 0.4814 | 0 | 0 | 0 | 1 | Ref |
| 8764 | MK450538 | Unassigned | Unassigned | Unassigned | 15_1 | Clustered | 9 | VC_15_1 | 9 | 0.182 | 1 | 0.182 | 0 | 0 | 0 | 0.8889 | Ref |
| 8765 | MK450543 | Unassigned | Unassigned | Unassigned | 352_0 | Clustered | 3 | VC_352_0 | 3 | 0.7306 | 0.98867275 | 0.7223 | 0 | 0 | 0 | 1 | Ref |
| 8766 | MK455769 | Unassigned | Unassigned | Unassigned | 1027_0 | Clustered | 20 | VC_1027_0 | 20 | 0.7114 | 1 | 0.7114 | 0 | 0 | 0 | 1 | Ref |
| 8767 | MK455770 | Unassigned | Unassigned | Unassigned | 1027_0 | Clustered | 20 | VC_1027_0 | 20 | 0.7114 | 1 | 0.7114 | 0 | 0 | 0 | 1 | Ref |
| 8768 | MK460245 | Unassigned | Unassigned | Unassigned | 981_1 | Clustered | 26 | VC_981_1 | 26 | 0.7009 | 1 | 0.7009 | 0 | 0 | 0 | 1 | Ref |
| 8769 | MK460246 | Unassigned | Unassigned | Unassigned | 237_3 | Clustered | 8 | VC_237_3 | 8 | 0.1092 | 1 | 0.1092 | 0 | 0 | 0 | 1 | Ref |
| 8770 | MK460247 | Unassigned | Unassigned | Unassigned | 236_0 | Clustered | 72 | VC_236_0 | 72 | 0.8104 | 1 | 0.8104 | 0 | 0 | 0 | 1 | Ref |
| 8771 | MK460248 | Unassigned | Unassigned | Unassigned | 1071_3 | Clustered | 16 | VC_1071_3 | 16 | 0.2354 | 1 | 0.2354 | 0 | 0 | 0 | 0.9667 | Ref |
| 8772 | MK468591 | Unassigned | Unassigned | Unassigned | 225_0 | Clustered | 32 | VC_225_0 | 32 | 0.848 | 1 | 0.848 | 0 | 0 | 0 | 0.996 | Ref |
| 8773 | MK473373 | Unassigned | Unassigned | Unassigned | 1030_0 | Clustered | 3 | VC_1030_0 | 3 | 0.7895 | 0.99542416 | 0.7859 | 0 | 0 | 0 | 1 | Ref |
| 8774 | MK473382 | Unassigned | Unassigned | Unassigned | 647_0 | Clustered | 11 | VC_647_0 | 11 | 0.598 | 1 | 0.598 | 0 | 0 | 0 | 1 | Ref |
| 8775 | MK474470 | Unassigned | Unassigned | Unassigned | 837_0 | Clustered | 29 | VC_837_0 | 29 | 0.9955 | 1 | 0.9955 | 0 | 0 | 0 | 1 | Ref |
| 8776 | MK479295 | Unassigned | Unassigned | Unassigned | 540_0 | Clustered | 15 | VC_540_0 | 15 | 0.2866 | 1 | 0.2866 | 0 | 0 | 0 | 1 | Ref |
| 8777 | MK482688 | Unassigned | Unassigned | Unassigned | 329_7 | Clustered | 18 | VC_329_7 | 18 | 0.0938 | 1 | 0.0938 | 0 | 0 | 0 | 0.7778 | Ref |
| 8778 | MK482689 | Unassigned | Unassigned | Unassigned | 329_7 | Clustered | 18 | VC_329_7 | 18 | 0.0938 | 1 | 0.0938 | 0 | 0 | 0 | 0.7778 | Ref |
| 8779 | MK482690 | Unassigned | Unassigned | Unassigned | 329_7 | Clustered | 18 | VC_329_7 | 18 | 0.0938 | 1 | 0.0938 | 0 | 0 | 0 | 0.7778 | Ref |
| 8780 | MK483107 | Unassigned | Unassigned | Unassigned | 530_0 | Clustered | 9 | VC_530_0 | 9 | 0.8027 | 1 | 0.8027 | 0 | 0 | 0 | 1 | Ref |
| 8781 | MK494087 | Unassigned | Unassigned | Unassigned | 215_1 | Clustered | 19 | VC_215_1 | 19 | 0.4188 | 1 | 0.4188 | 0 | 0 | 0 | 1 | Ref |
| 8782 | MK494088 | Unassigned | Unassigned | Unassigned | 517_1 | Clustered | 108 | VC_517_1 | 108 | 0.8483 | 1 | 0.8483 | 0 | 0 | 0 | 0.9955 | Ref |
| 8783 | MK494089 | Unassigned | Unassigned | Unassigned | 1046_0 | Clustered | 7 | VC_1046_0 | 7 | 0.9413 | 1 | 0.9413 | 0 | 0 | 0 | 1 | Ref |
| 8784 | MK494090 | Unassigned | Unassigned | Unassigned | 223_0 | Clustered | 38 | VC_223_0 | 38 | 0.9095 | 1 | 0.9095 | 0 | 0 | 0 | 1 | Ref |
| 8785 | MK494091 | Unassigned | Unassigned | Unassigned | 940_0 | Clustered | 34 | VC_940_0 | 34 | 0.7959 | 1 | 0.7959 | 0 | 0 | 0 | 1 | Ref |
| 8786 | MK494092 | Unassigned | Unassigned | Unassigned | 887_0 | Clustered | 18 | VC_887_0 | 18 | 0.3663 | 1 | 0.3663 | 0 | 0 | 0 | 1 | Ref |
| 8787 | MK494093 | Unassigned | Unassigned | Unassigned | 517_1 | Clustered | 108 | VC_517_1 | 108 | 0.8483 | 1 | 0.8483 | 0 | 0 | 0 | 0.9955 | Ref |
| 8788 | MK494094 | Unassigned | Unassigned | Unassigned | 517_1 | Clustered | 108 | VC_517_1 | 108 | 0.8483 | 1 | 0.8483 | 0 | 0 | 0 | 0.9955 | Ref |
| 8789 | MK494095 | Unassigned | Unassigned | Unassigned | 810_0 | Clustered | 18 | VC_810_0 | 18 | 0.9121 | 1 | 0.9121 | 0 | 0 | 0 | 1 | Ref |
| 8790 | MK494096 | Unassigned | Unassigned | Unassigned | 514_0 | Clustered | 75 | VC_514_0 | 75 | 0.9402 | 1 | 0.9402 | 0 | 0 | 0 | 1 | Ref |
| 8791 | MK494097 | Unassigned | Unassigned | Unassigned | 517_1 | Clustered | 108 | VC_517_1 | 108 | 0.8483 | 1 | 0.8483 | 0 | 0 | 0 | 0.9955 | Ref |
| 8792 | MK494098 | Unassigned | Unassigned | Unassigned | 869_0 | Clustered | 9 | VC_869_0 | 9 | 0.2624 | 1 | 0.2624 | 0 | 0 | 0 | 1 | Ref |
| 8793 | MK494099 | Unassigned | Unassigned | Unassigned | 686_0 | Clustered | 2 | VC_686_0 | 2 | 0.2147 | 0.9539194 | 0.2048 | 0 | 0 | 0 | 1 | Ref |
| 8794 | MK494100 | Unassigned | Unassigned | Unassigned | 887_0 | Clustered | 18 | VC_887_0 | 18 | 0.3663 | 1 | 0.3663 | 0 | 0 | 0 | 1 | Ref |
| 8795 | MK494101 | Unassigned | Unassigned | Unassigned | 517_1 | Clustered | 108 | VC_517_1 | 108 | 0.8483 | 1 | 0.8483 | 0 | 0 | 0 | 0.9955 | Ref |
| 8796 | MK494102 | Unassigned | Unassigned | Unassigned | 565_0 | Clustered | 20 | VC_565_0 | 20 | 0.8398 | 1 | 0.8398 | 0 | 0 | 0 | 1 | Ref |
| 8797 | MK494103 | Unassigned | Unassigned | Unassigned | 950_0 | Clustered | 38 | VC_950_0 | 38 | 0.9443 | 1 | 0.9443 | 0 | 0 | 0 | 1 | Ref |
| 8798 | MK494104 | Unassigned | Unassigned | Unassigned | 371_16 | Clustered | 42 | VC_371_16 | 42 | 0.1341 | 1 | 0.1341 | 0 | 0 | 0 | 0.7282 | Ref |
| 8799 | MK494105 | Unassigned | Unassigned | Unassigned | 762_0 | Clustered | 9 | VC_762_0 | 9 | 0.0885 | 1 | 0.0885 | 0 | 0 | 0 | 1 | Ref |
| 8800 | MK494106 | Unassigned | Unassigned | Unassigned | 866_1 | Clustered | 20 | VC_866_1 | 20 | 0.4814 | 1 | 0.4814 | 0 | 0 | 0 | 1 | Ref |
| 8801 | MK494107 | Unassigned | Unassigned | Unassigned | 1074_0 | Clustered | 19 | VC_1074_0 | 19 | 0.9198 | 1 | 0.9198 | 0 | 0 | 0 | 1 | Ref |
| 8802 | MK494108 | Unassigned | Unassigned | Unassigned | 225_0 | Clustered | 32 | VC_225_0 | 32 | 0.848 | 1 | 0.848 | 0 | 0 | 0 | 0.996 | Ref |
| 8803 | MK494109 | Unassigned | Unassigned | Unassigned | 866_1 | Clustered | 20 | VC_866_1 | 20 | 0.4814 | 1 | 0.4814 | 0 | 0 | 0 | 1 | Ref |
| 8804 | MK494110 | Unassigned | Unassigned | Unassigned | 520_45 | Clustered | 7 | VC_520_45 | 7 | 0.0635 | 1 | 0.0635 | 0 | 0 | 0 | 0.8095 | Ref |
| 8805 | MK494111 | Unassigned | Unassigned | Unassigned | 950_0 | Clustered | 38 | VC_950_0 | 38 | 0.9443 | 1 | 0.9443 | 0 | 0 | 0 | 1 | Ref |
| 8806 | MK494112 | Unassigned | Unassigned | Unassigned | 1071_3 | Clustered | 16 | VC_1071_3 | 16 | 0.2354 | 1 | 0.2354 | 0 | 0 | 0 | 0.9667 | Ref |
| 8807 | MK494113 | Unassigned | Unassigned | Unassigned | 222_0 | Clustered | 5 | VC_222_0 | 5 | 0.0565 | 0.99999997 | 0.0565 | 0 | 0 | 0 | 1 | Ref |
| 8808 | MK494114 | Unassigned | Unassigned | Unassigned | 507_0 | Clustered | 8 | VC_507_0 | 8 | 0.4466 | 1 | 0.4466 | 0 | 0 | 0 | 1 | Ref |
| 8809 | MK494115 | Unassigned | Unassigned | Unassigned | 517_0 | Clustered | 18 | VC_517_0 | 18 | 0.1599 | 1 | 0.1599 | 0 | 0 | 0 | 1 | Ref |
| 8810 | MK494116 | Unassigned | Unassigned | Unassigned | 325_30 | Clustered | 4 | VC_325_30 | 4 | 0.0144 | 0.99998616 | 0.0144 | 0 | 0 | 0 | 0.8333 | Ref |
| 8811 | MK494117 | Unassigned | Unassigned | Unassigned | 517_0 | Clustered | 18 | VC_517_0 | 18 | 0.1599 | 1 | 0.1599 | 0 | 0 | 0 | 1 | Ref |
| 8812 | MK494118 | Unassigned | Unassigned | Unassigned | 520_72 | Clustered/Singleton | 1 | VC_520_72 | 1 | 0 | 1 | 0 | 0 | 0 | 0 | 1 | Ref |
| 8813 | MK494119 | Unassigned | Unassigned | Unassigned | 237_0 | Clustered | 4 | VC_237_0 | 4 | 0.0429 | 0.99998759 | 0.0429 | 0 | 0 | 0 | 1 | Ref |
| 8814 | MK494121 | Unassigned | Unassigned | Unassigned | 866_1 | Clustered | 20 | VC_866_1 | 20 | 0.4814 | 1 | 0.4814 | 0 | 0 | 0 | 1 | Ref |
| 8815 | MK494122 | Unassigned | Unassigned | Unassigned | 519_0 | Clustered | 39 | VC_519_0 | 39 | 0.8156 | 1 | 0.8156 | 0 | 0 | 0 | 1 | Ref |
| 8816 | MK494123 | Unassigned | Unassigned | Unassigned | 939_0 | Clustered | 15 | VC_939_0 | 15 | 1 | 0 | 0 | 0 | 0 | 0 | 1 | Ref |
| 8817 | MK494124 | Unassigned | Unassigned | Unassigned | 515_0 | Clustered | 16 | VC_515_0 | 16 | 0.3793 | 1 | 0.3793 | 0 | 0 | 0 | 1 | Ref |
| 8818 | MK494125 | Unassigned | Unassigned | Unassigned | 517_1 | Clustered | 108 | VC_517_1 | 108 | 0.8483 | 1 | 0.8483 | 0 | 0 | 0 | 0.9955 | Ref |
| 8819 | MK494126 | Unassigned | Unassigned | Unassigned | 517_1 | Clustered | 108 | VC_517_1 | 108 | 0.8483 | 1 | 0.8483 | 0 | 0 | 0 | 0.9955 | Ref |
| 8820 | MK494127 | Unassigned | Unassigned | Unassigned | 513_0 | Clustered | 32 | VC_513_0 | 32 | 0.6881 | 1 | 0.6881 | 0 | 0 | 0 | 1 | Ref |
| 8821 | MK494128 | Unassigned | Unassigned | Unassigned | 514_0 | Clustered | 75 | VC_514_0 | 75 | 0.9402 | 1 | 0.9402 | 0 | 0 | 0 | 1 | Ref |
| 8822 | MK494129 | Unassigned | Unassigned | Unassigned | 517_0 | Clustered | 18 | VC_517_0 | 18 | 0.1599 | 1 | 0.1599 | 0 | 0 | 0 | 1 | Ref |
| 8823 | MK494130 | Unassigned | Unassigned | Unassigned | 221_0 | Clustered | 22 | VC_221_0 | 22 | 0.7447 | 1 | 0.7447 | 0 | 0 | 0 | 1 | Ref |
| 8824 | MK494131 | Unassigned | Unassigned | Unassigned | 507_0 | Clustered | 8 | VC_507_0 | 8 | 0.4466 | 1 | 0.4466 | 0 | 0 | 0 | 1 | Ref |
| 8825 | MK494132 | Unassigned | Unassigned | Unassigned | 507_0 | Clustered | 8 | VC_507_0 | 8 | 0.4466 | 1 | 0.4466 | 0 | 0 | 0 | 1 | Ref |
| 8826 | MK496679 | Unassigned | Unassigned | Unassigned | 1371_0 | Clustered | 5 | VC_1371_0 | 5 | 0.7004 | 0.52188382 | 0.3656 | 0 | 0 | 0 | 1 | Ref |
| 8827 | MK496680 | Unassigned | Unassigned | Unassigned | 1371_0 | Clustered | 5 | VC_1371_0 | 5 | 0.7004 | 0.52188382 | 0.3656 | 0 | 0 | 0 | 1 | Ref |
| 8828 | MK496681 | Unassigned | Unassigned | Unassigned | 1373_0 | Clustered | 2 | VC_1373_0 | 2 | 0.6485 | 0.86822376 | 0.563 | 0 | 0 | 0 | 1 | Ref |
| 8829 | MK496682 | Unassigned | Unassigned | Unassigned | 1367_0 | Clustered | 2 | VC_1367_0 | 2 | 1 | 1 | 1 | 0 | 0 | 0 | 1 | Ref |
| 8830 | MK496683 | Unassigned | Unassigned | Unassigned | 1374_0 | Clustered | 2 | VC_1374_0 | 2 | 1 | 1 | 1 | 0 | 0 | 0 | 1 | Ref |
| 8831 | MK496685 | Unassigned | Unassigned | Unassigned | 1264_0 | Clustered | 6 | VC_1264_0 | 6 | 0.7062 | 0.92055725 | 0.6501 | 0 | 0 | 0 | 1 | Ref |
| 8832 | MK496686 | Unassigned | Unassigned | Unassigned | 1375_0 | Clustered | 2 | VC_1375_0 | 2 | 1 | 1 | 1 | 0 | 0 | 0 | 1 | Ref |
| 8833 | MK496688 | Unassigned | Unassigned | Unassigned | 1376_0 | Clustered | 3 | VC_1376_0 | 3 | 1 | 1 | 1 | 0 | 0 | 0 | 1 | Ref |
| 8834 | MK496689 | Unassigned | Unassigned | Unassigned | 1377_0 | Clustered | 3 | VC_1377_0 | 3 | 1 | 0 | 0 | 0 | 0 | 0 | 1 | Ref |
| 8835 | MK496690 | Unassigned | Unassigned | Unassigned | 1376_0 | Clustered | 3 | VC_1376_0 | 3 | 1 | 1 | 1 | 0 | 0 | 0 | 1 | Ref |
| 8836 | MK496692 | Unassigned | Unassigned | Unassigned | 1326_0 | Clustered | 2 | VC_1326_0 | 2 | 1 | 1 | 1 | 0 | 0 | 0 | 1 | Ref |
| 8837 | MK496693 | Unassigned | Unassigned | Unassigned | 1197_0 | Clustered | 15 | VC_1197_0 | 15 | 0.8991 | 0.76608394 | 0.6888 | 0 | 0 | 0 | 1 | Ref |
| 8838 | MK496694 | Unassigned | Unassigned | Unassigned | 1243_0 | Clustered | 52 | VC_1243_0 | 52 | 0.9126 | 0.92708311 | 0.8461 | 0 | 0 | 0 | 1 | Ref |
| 8839 | MK496696 | Unassigned | Unassigned | Unassigned | 1297_0 | Clustered | 7 | VC_1297_0 | 7 | 1 | 1 | 1 | 0 | 0 | 0 | 1 | Ref |
| 8840 | MK496697 | Unassigned | Unassigned | Unassigned | 1378_0 | Clustered | 2 | VC_1378_0 | 2 | 1 | 1 | 1 | 0 | 0 | 0 | 1 | Ref |
| 8841 | MK496698 | Unassigned | Unassigned | Unassigned | 1372_0 | Clustered | 2 | VC_1372_0 | 2 | 0.1774 | 0.73645537 | 0.1307 | 0 | 0 | 0 | 1 | Ref |
| 8842 | MK496699 | Unassigned | Unassigned | Unassigned | 1379_0 | Clustered | 2 | VC_1379_0 | 2 | 1 | 1 | 1 | 0 | 0 | 0 | 1 | Ref |
| 8843 | MK496700 | Unassigned | Unassigned | Unassigned | 1377_0 | Clustered | 3 | VC_1377_0 | 3 | 1 | 0 | 0 | 0 | 0 | 0 | 1 | Ref |
| 8844 | MK496701 | Unassigned | Unassigned | Unassigned | 1380_0 | Clustered | 2 | VC_1380_0 | 2 | 1 | 1 | 1 | 0 | 0 | 0 | 1 | Ref |
| 8845 | MK496702 | Unassigned | Unassigned | Unassigned | 1380_0 | Clustered | 2 | VC_1380_0 | 2 | 1 | 1 | 1 | 0 | 0 | 0 | 1 | Ref |
| 8846 | MK496704 | Unassigned | Unassigned | Unassigned | 1296_0 | Clustered | 4 | VC_1296_0 | 4 | 0.8684 | 0.91151676 | 0.7915 | 0 | 0 | 0 | 1 | Ref |
| 8847 | MK496710 | Unassigned | Unassigned | Unassigned | 1381_0 | Clustered | 3 | VC_1381_0 | 3 | 1 | 1 | 1 | 0 | 0 | 0 | 1 | Ref |
| 8848 | MK496711 | Unassigned | Unassigned | Unassigned | 1297_0 | Clustered | 7 | VC_1297_0 | 7 | 1 | 1 | 1 | 0 | 0 | 0 | 1 | Ref |
| 8849 | MK496712 | Unassigned | Unassigned | Unassigned | 1382_0 | Clustered | 2 | VC_1382_0 | 2 | 1 | 1 | 1 | 0 | 0 | 0 | 1 | Ref |
| 8850 | MK496714 | Unassigned | Unassigned | Unassigned | 1134_0 | Clustered | 5 | VC_1134_0 | 5 | 0.129 | 0.7637089 | 0.0985 | 0 | 0 | 0 | 1 | Ref |
| 8851 | MK496716 | Unassigned | Unassigned | Unassigned | 1383_0 | Clustered | 2 | VC_1383_0 | 2 | 0.6415 | 0.72985431 | 0.4682 | 0 | 0 | 0 | 1 | Ref |
| 8852 | MK496717 | Unassigned | Unassigned | Unassigned | 1384_0 | Clustered | 2 | VC_1384_0 | 2 | 1 | 1 | 1 | 0 | 0 | 0 | 1 | Ref |
| 8853 | MK496719 | Unassigned | Unassigned | Unassigned | 1112_0 | Clustered | 2 | VC_1112_0 | 2 | 0.2212 | 0.72377355 | 0.1601 | 0 | 0 | 0 | 1 | Ref |
| 8854 | MK496720 | Unassigned | Unassigned | Unassigned | 1385_0 | Clustered | 2 | VC_1385_0 | 2 | 1 | 1 | 1 | 0 | 0 | 0 | 1 | Ref |
| 8855 | MK496721 | Unassigned | Unassigned | Unassigned | 1386_0 | Clustered | 3 | VC_1386_0 | 3 | 1 | 0 | 0 | 0 | 0 | 0 | 1 | Ref |
| 8856 | MK496723 | Unassigned | Unassigned | Unassigned | 1054_0 | Clustered | 12 | VC_1054_0 | 12 | 0.2863 | 0.99746644 | 0.2855 | 0 | 0 | 0 | 1 | Ref |
| 8857 | MK496725 | Unassigned | Unassigned | Unassigned | 1387_0 | Clustered | 3 | VC_1387_0 | 3 | 0.7373 | 0.96173875 | 0.709 | 0 | 0 | 0 | 1 | Ref |
| 8858 | MK496729 | Unassigned | Unassigned | Unassigned | 1296_0 | Clustered | 4 | VC_1296_0 | 4 | 0.8684 | 0.91151676 | 0.7915 | 0 | 0 | 0 | 1 | Ref |
| 8859 | MK496732 | Unassigned | Unassigned | Unassigned | 1292_0 | Clustered | 3 | VC_1292_0 | 3 | 1 | 1 | 1 | 0 | 0 | 0 | 1 | Ref |
| 8860 | MK496738 | Unassigned | Unassigned | Unassigned | 1112_0 | Clustered | 2 | VC_1112_0 | 2 | 0.2212 | 0.72377355 | 0.1601 | 0 | 0 | 0 | 1 | Ref |
| 8861 | MK496743 | Unassigned | Unassigned | Unassigned | 1105_0 | Clustered | 3 | VC_1105_0 | 3 | 0.0755 | 0.99466875 | 0.0751 | 0 | 0 | 0 | 1 | Ref |
| 8862 | MK496744 | Unassigned | Unassigned | Unassigned | 1105_0 | Clustered | 3 | VC_1105_0 | 3 | 0.0755 | 0.99466875 | 0.0751 | 0 | 0 | 0 | 1 | Ref |
| 8863 | MK496745 | Unassigned | Unassigned | Unassigned | 1147_0 | Clustered | 3 | VC_1147_0 | 3 | 0.4334 | 0.65335909 | 0.2832 | 0 | 0 | 0 | 1 | Ref |
| 8864 | MK496746 | Unassigned | Unassigned | Unassigned | 1387_0 | Clustered | 3 | VC_1387_0 | 3 | 0.7373 | 0.96173875 | 0.709 | 0 | 0 | 0 | 1 | Ref |
| 8865 | MK496748 | Unassigned | Unassigned | Unassigned | 1388_0 | Clustered | 2 | VC_1388_0 | 2 | 1 | 1 | 1 | 0 | 0 | 0 | 1 | Ref |
| 8866 | MK496750 | Unassigned | Unassigned | Unassigned | 1389_0 | Clustered | 2 | VC_1389_0 | 2 | 0.6339 | 0.72985431 | 0.4627 | 0 | 0 | 0 | 1 | Ref |
| 8867 | MK496753 | Unassigned | Unassigned | Unassigned | 1390_0 | Clustered | 2 | VC_1390_0 | 2 | 1 | 1 | 1 | 0 | 0 | 0 | 1 | Ref |
| 8868 | MK496754 | Unassigned | Unassigned | Unassigned | 1119_0 | Clustered | 20 | VC_1119_0 | 20 | 0.6751 | 0.99997892 | 0.6751 | 0 | 0 | 0 | 1 | Ref |
| 8869 | MK496756 | Unassigned | Unassigned | Unassigned | 1391_0 | Clustered | 2 | VC_1391_0 | 2 | 1 | 1 | 1 | 0 | 0 | 0 | 1 | Ref |
| 8870 | MK496757 | Unassigned | Unassigned | Unassigned | 1303_0 | Clustered | 6 | VC_1303_0 | 6 | 1 | 1 | 1 | 0 | 0 | 0 | 1 | Ref |
| 8871 | MK496758 | Unassigned | Unassigned | Unassigned | 1392_0 | Clustered | 2 | VC_1392_0 | 2 | 1 | 1 | 1 | 0 | 0 | 0 | 1 | Ref |
| 8872 | MK496760 | Unassigned | Unassigned | Unassigned | 1373_0 | Clustered | 2 | VC_1373_0 | 2 | 0.6485 | 0.86822376 | 0.563 | 0 | 0 | 0 | 1 | Ref |
| 8873 | MK496761 | Unassigned | Unassigned | Unassigned | 1374_0 | Clustered | 2 | VC_1374_0 | 2 | 1 | 1 | 1 | 0 | 0 | 0 | 1 | Ref |
| 8874 | MK496764 | Unassigned | Unassigned | Unassigned | 1375_0 | Clustered | 2 | VC_1375_0 | 2 | 1 | 1 | 1 | 0 | 0 | 0 | 1 | Ref |
| 8875 | MK496765 | Unassigned | Unassigned | Unassigned | 1294_0 | Clustered | 6 | VC_1294_0 | 6 | 0.6626 | 0.99999272 | 0.6626 | 0 | 0 | 0 | 1 | Ref |
| 8876 | MK496766 | Unassigned | Unassigned | Unassigned | 1376_0 | Clustered | 3 | VC_1376_0 | 3 | 1 | 1 | 1 | 0 | 0 | 0 | 1 | Ref |
| 8877 | MK496767 | Unassigned | Unassigned | Unassigned | 1381_0 | Clustered | 3 | VC_1381_0 | 3 | 1 | 1 | 1 | 0 | 0 | 0 | 1 | Ref |
| 8878 | MK496768 | Unassigned | Unassigned | Unassigned | 1381_0 | Clustered | 3 | VC_1381_0 | 3 | 1 | 1 | 1 | 0 | 0 | 0 | 1 | Ref |
| 8879 | MK496770 | Unassigned | Unassigned | Unassigned | 1393_0 | Clustered | 2 | VC_1393_0 | 2 | 1 | 1 | 1 | 0 | 0 | 0 | 1 | Ref |
| 8880 | MK496771 | Unassigned | Unassigned | Unassigned | 1393_0 | Clustered | 2 | VC_1393_0 | 2 | 1 | 1 | 1 | 0 | 0 | 0 | 1 | Ref |
| 8881 | MK496772 | Unassigned | Unassigned | Unassigned | 1372_0 | Clustered | 2 | VC_1372_0 | 2 | 0.1774 | 0.73645537 | 0.1307 | 0 | 0 | 0 | 1 | Ref |
| 8882 | MK496774 | Unassigned | Unassigned | Unassigned | 1197_0 | Clustered | 15 | VC_1197_0 | 15 | 0.8991 | 0.76608394 | 0.6888 | 0 | 0 | 0 | 1 | Ref |
| 8883 | MK496775 | Unassigned | Unassigned | Unassigned | 1243_0 | Clustered | 52 | VC_1243_0 | 52 | 0.9126 | 0.92708311 | 0.8461 | 0 | 0 | 0 | 1 | Ref |
| 8884 | MK496776 | Unassigned | Unassigned | Unassigned | 1371_0 | Clustered | 5 | VC_1371_0 | 5 | 0.7004 | 0.52188382 | 0.3656 | 0 | 0 | 0 | 1 | Ref |
| 8885 | MK496779 | Unassigned | Unassigned | Unassigned | 1134_0 | Clustered | 5 | VC_1134_0 | 5 | 0.129 | 0.7637089 | 0.0985 | 0 | 0 | 0 | 1 | Ref |
| 8886 | MK496780 | Unassigned | Unassigned | Unassigned | 1378_0 | Clustered | 2 | VC_1378_0 | 2 | 1 | 1 | 1 | 0 | 0 | 0 | 1 | Ref |
| 8887 | MK496781 | Unassigned | Unassigned | Unassigned | 1386_0 | Clustered | 3 | VC_1386_0 | 3 | 1 | 0 | 0 | 0 | 0 | 0 | 1 | Ref |
| 8888 | MK496782 | Unassigned | Unassigned | Unassigned | 1296_0 | Clustered | 4 | VC_1296_0 | 4 | 0.8684 | 0.91151676 | 0.7915 | 0 | 0 | 0 | 1 | Ref |
| 8889 | MK496784 | Unassigned | Unassigned | Unassigned | 1377_0 | Clustered | 3 | VC_1377_0 | 3 | 1 | 0 | 0 | 0 | 0 | 0 | 1 | Ref |
| 8890 | MK496785 | Unassigned | Unassigned | Unassigned | 1379_0 | Clustered | 2 | VC_1379_0 | 2 | 1 | 1 | 1 | 0 | 0 | 0 | 1 | Ref |
| 8891 | MK496786 | Unassigned | Unassigned | Unassigned | 1386_0 | Clustered | 3 | VC_1386_0 | 3 | 1 | 0 | 0 | 0 | 0 | 0 | 1 | Ref |
| 8892 | MK496787 | Unassigned | Unassigned | Unassigned | 1382_0 | Clustered | 2 | VC_1382_0 | 2 | 1 | 1 | 1 | 0 | 0 | 0 | 1 | Ref |
| 8893 | MK496790 | Unassigned | Unassigned | Unassigned | 1383_0 | Clustered | 2 | VC_1383_0 | 2 | 0.6415 | 0.72985431 | 0.4682 | 0 | 0 | 0 | 1 | Ref |
| 8894 | MK496792 | Unassigned | Unassigned | Unassigned | 1134_0 | Clustered | 5 | VC_1134_0 | 5 | 0.129 | 0.7637089 | 0.0985 | 0 | 0 | 0 | 1 | Ref |
| 8895 | MK496796 | Unassigned | Unassigned | Unassigned | 1385_0 | Clustered | 2 | VC_1385_0 | 2 | 1 | 1 | 1 | 0 | 0 | 0 | 1 | Ref |
| 8896 | MK496804 | Unassigned | Unassigned | Unassigned | 1384_0 | Clustered | 2 | VC_1384_0 | 2 | 1 | 1 | 1 | 0 | 0 | 0 | 1 | Ref |
| 8897 | MK496805 | Unassigned | Unassigned | Unassigned | 1119_0 | Clustered | 20 | VC_1119_0 | 20 | 0.6751 | 0.99997892 | 0.6751 | 0 | 0 | 0 | 1 | Ref |
| 8898 | MK496808 | Unassigned | Unassigned | Unassigned | 1392_0 | Clustered | 2 | VC_1392_0 | 2 | 1 | 1 | 1 | 0 | 0 | 0 | 1 | Ref |
| 8899 | MK496811 | Unassigned | Unassigned | Unassigned | 1387_0 | Clustered | 3 | VC_1387_0 | 3 | 0.7373 | 0.96173875 | 0.709 | 0 | 0 | 0 | 1 | Ref |
| 8900 | MK496812 | Unassigned | Unassigned | Unassigned | 1119_0 | Clustered | 20 | VC_1119_0 | 20 | 0.6751 | 0.99997892 | 0.6751 | 0 | 0 | 0 | 1 | Ref |
| 8901 | MK496813 | Unassigned | Unassigned | Unassigned | 1119_0 | Clustered | 20 | VC_1119_0 | 20 | 0.6751 | 0.99997892 | 0.6751 | 0 | 0 | 0 | 1 | Ref |
| 8902 | MK496815 | Unassigned | Unassigned | Unassigned | 1388_0 | Clustered | 2 | VC_1388_0 | 2 | 1 | 1 | 1 | 0 | 0 | 0 | 1 | Ref |
| 8903 | MK496816 | Unassigned | Unassigned | Unassigned | 1389_0 | Clustered | 2 | VC_1389_0 | 2 | 0.6339 | 0.72985431 | 0.4627 | 0 | 0 | 0 | 1 | Ref |
| 8904 | MK496818 | Unassigned | Unassigned | Unassigned | 1390_0 | Clustered | 2 | VC_1390_0 | 2 | 1 | 1 | 1 | 0 | 0 | 0 | 1 | Ref |
| 8905 | MK496820 | Unassigned | Unassigned | Unassigned | 1394_0 | Clustered | 2 | VC_1394_0 | 2 | 1 | 1 | 1 | 0 | 0 | 0 | 1 | Ref |
| 8906 | MK496824 | Unassigned | Unassigned | Unassigned | 1391_0 | Clustered | 2 | VC_1391_0 | 2 | 1 | 1 | 1 | 0 | 0 | 0 | 1 | Ref |
| 8907 | MK496825 | Unassigned | Unassigned | Unassigned | 995_0 | Clustered | 69 | VC_995_0 | 69 | 0.8547 | 1 | 0.8547 | 0 | 0 | 0 | 1 | Ref |
| 8908 | MK501729 | Unassigned | Unassigned | Unassigned | 1176_0 | Clustered | 41 | VC_1176_0 | 41 | 0.9558 | 1 | 0.9558 | 0 | 0 | 0 | 0.9976 | Ref |

| | | | | | | | | | | | | | | | | | |
|---|---|---|---|---|---|---|---|---|---|---|---|---|---|---|---|---|---|
| 8909 | MK501730 | Unassigned | Unassigned | Unassigned | 1176_0 | Clustered | 41 | VC_1176_0 | 41 | 0.9558 | 1 | 0.9558 | 0 | 0 | 0 | 0.9976 | Ref |
| 8910 | MK504442 | Unassigned | Unassigned | Unassigned | 578_0 | Clustered | 2 | VC_578_0 | 2 | 0.1712 | 0.93973606 | 0.1608 | 0 | 0 | 0 | 1 | Ref |
| 8911 | MK504443 | Unassigned | Unassigned | Unassigned | 95_0 | Clustered | 3 | VC_95_0 | 3 | 0.2092 | 0.99852577 | 0.2089 | 0 | 0 | 0 | 1 | Ref |
| 8912 | MK504444 | Unassigned | Unassigned | Unassigned | 526_3 | Clustered/Singleton | 1 | VC_526_3 | 1 | 0 | 1 | 0 | 0 | 0 | 0 | 1 | Ref |
| 8913 | MK504446 | Unassigned | Unassigned | Unassigned | 95_0 | Clustered | 3 | VC_95_0 | 3 | 0.2092 | 0.99852577 | 0.2089 | 0 | 0 | 0 | 1 | Ref |
| 8914 | MK509462 | Unassigned | Unassigned | Unassigned | 131_2 | Clustered | 7 | VC_131_2 | 7 | 0.0685 | 1 | 0.0685 | 0 | 0 | 0 | 0.9048 | Ref |
| 8915 | MK510962 | Unassigned | Unassigned | Unassigned | 747_0 | Clustered | 23 | VC_747_0 | 23 | 0.8202 | 1 | 0.8202 | 0 | 0 | 0 | 1 | Ref |
| 8916 | MK510963 | Unassigned | Unassigned | Unassigned | 747_0 | Clustered | 23 | VC_747_0 | 23 | 0.8202 | 1 | 0.8202 | 0 | 0 | 0 | 1 | Ref |
| 8917 | MK510964 | Unassigned | Unassigned | Unassigned | 747_0 | Clustered | 23 | VC_747_0 | 23 | 0.8202 | 1 | 0.8202 | 0 | 0 | 0 | 1 | Ref |
| 8918 | MK510965 | Unassigned | Unassigned | Unassigned | 747_0 | Clustered | 23 | VC_747_0 | 23 | 0.8202 | 1 | 0.8202 | 0 | 0 | 0 | 1 | Ref |
| 8919 | MK510966 | Unassigned | Unassigned | Unassigned | 748_0 | Clustered | 12 | VC_748_0 | 12 | 0.6879 | 1 | 0.6879 | 0 | 0 | 0 | 1 | Ref |
| 8920 | MK510967 | Unassigned | Unassigned | Unassigned | 748_0 | Clustered | 12 | VC_748_0 | 12 | 0.6879 | 1 | 0.6879 | 0 | 0 | 0 | 1 | Ref |
| 8921 | MK510968 | Unassigned | Unassigned | Unassigned | 747_0 | Clustered | 23 | VC_747_0 | 23 | 0.8202 | 1 | 0.8202 | 0 | 0 | 0 | 1 | Ref |
| 8922 | MK510969 | Unassigned | Unassigned | Unassigned | 747_0 | Clustered | 23 | VC_747_0 | 23 | 0.8202 | 1 | 0.8202 | 0 | 0 | 0 | 1 | Ref |
| 8923 | MK510970 | Unassigned | Unassigned | Unassigned | 747_0 | Clustered | 23 | VC_747_0 | 23 | 0.8202 | 1 | 0.8202 | 0 | 0 | 0 | 1 | Ref |
| 8924 | MK510971 | Unassigned | Unassigned | Unassigned | 747_0 | Clustered | 23 | VC_747_0 | 23 | 0.8202 | 1 | 0.8202 | 0 | 0 | 0 | 1 | Ref |
| 8925 | MK510972 | Unassigned | Unassigned | Unassigned | 748_0 | Clustered | 12 | VC_748_0 | 12 | 0.6879 | 1 | 0.6879 | 0 | 0 | 0 | 1 | Ref |
| 8926 | MK510973 | Unassigned | Unassigned | Unassigned | 748_0 | Clustered | 12 | VC_748_0 | 12 | 0.6879 | 1 | 0.6879 | 0 | 0 | 0 | 1 | Ref |
| 8927 | MK510974 | Unassigned | Unassigned | Unassigned | 747_0 | Clustered | 23 | VC_747_0 | 23 | 0.8202 | 1 | 0.8202 | 0 | 0 | 0 | 1 | Ref |
| 8928 | MK510975 | Unassigned | Unassigned | Unassigned | 747_0 | Clustered | 23 | VC_747_0 | 23 | 0.8202 | 1 | 0.8202 | 0 | 0 | 0 | 1 | Ref |
| 8929 | MK510976 | Unassigned | Unassigned | Unassigned | 747_0 | Clustered | 23 | VC_747_0 | 23 | 0.8202 | 1 | 0.8202 | 0 | 0 | 0 | 1 | Ref |
| 8930 | MK510977 | Unassigned | Unassigned | Unassigned | 748_0 | Clustered | 12 | VC_748_0 | 12 | 0.6879 | 1 | 0.6879 | 0 | 0 | 0 | 1 | Ref |
| 8931 | MK510978 | Unassigned | Unassigned | Unassigned | 747_0 | Clustered | 23 | VC_747_0 | 23 | 0.8202 | 1 | 0.8202 | 0 | 0 | 0 | 1 | Ref |
| 8932 | MK510979 | Unassigned | Unassigned | Unassigned | 748_0 | Clustered | 12 | VC_748_0 | 12 | 0.6879 | 1 | 0.6879 | 0 | 0 | 0 | 1 | Ref |
| 8933 | MK510980 | Unassigned | Unassigned | Unassigned | 748_0 | Clustered | 12 | VC_748_0 | 12 | 0.6879 | 1 | 0.6879 | 0 | 0 | 0 | 1 | Ref |
| 8934 | MK510981 | Unassigned | Unassigned | Unassigned | 748_0 | Clustered | 12 | VC_748_0 | 12 | 0.6879 | 1 | 0.6879 | 0 | 0 | 0 | 1 | Ref |
| 8935 | MK510982 | Unassigned | Unassigned | Unassigned | 748_0 | Clustered | 12 | VC_748_0 | 12 | 0.6879 | 1 | 0.6879 | 0 | 0 | 0 | 1 | Ref |
| 8936 | MK510983 | Unassigned | Unassigned | Unassigned | 748_0 | Clustered | 12 | VC_748_0 | 12 | 0.6879 | 1 | 0.6879 | 0 | 0 | 0 | 1 | Ref |
| 8937 | MK510984 | Unassigned | Unassigned | Unassigned | 747_0 | Clustered | 23 | VC_747_0 | 23 | 0.8202 | 1 | 0.8202 | 0 | 0 | 0 | 1 | Ref |
| 8938 | MK510985 | Unassigned | Unassigned | Unassigned | 747_0 | Clustered | 23 | VC_747_0 | 23 | 0.8202 | 1 | 0.8202 | 0 | 0 | 0 | 1 | Ref |
| 8939 | MK510986 | Unassigned | Unassigned | Unassigned | 747_0 | Clustered | 23 | VC_747_0 | 23 | 0.8202 | 1 | 0.8202 | 0 | 0 | 0 | 1 | Ref |
| 8940 | MK510987 | Unassigned | Unassigned | Unassigned | 747_0 | Clustered | 23 | VC_747_0 | 23 | 0.8202 | 1 | 0.8202 | 0 | 0 | 0 | 1 | Ref |
| 8941 | MK510988 | Unassigned | Unassigned | Unassigned | 747_0 | Clustered | 23 | VC_747_0 | 23 | 0.8202 | 1 | 0.8202 | 0 | 0 | 0 | 1 | Ref |
| 8942 | MK510989 | Unassigned | Unassigned | Unassigned | 748_0 | Clustered | 12 | VC_748_0 | 12 | 0.6879 | 1 | 0.6879 | 0 | 0 | 0 | 1 | Ref |
| 8943 | MK510990 | Unassigned | Unassigned | Unassigned | 747_0 | Clustered | 23 | VC_747_0 | 23 | 0.8202 | 1 | 0.8202 | 0 | 0 | 0 | 1 | Ref |
| 8944 | MK510991 | Unassigned | Unassigned | Unassigned | 748_0 | Clustered | 12 | VC_748_0 | 12 | 0.6879 | 1 | 0.6879 | 0 | 0 | 0 | 1 | Ref |
| 8945 | MK510992 | Unassigned | Unassigned | Unassigned | 747_0 | Clustered | 23 | VC_747_0 | 23 | 0.8202 | 1 | 0.8202 | 0 | 0 | 0 | 1 | Ref |
| 8946 | MK510993 | Unassigned | Unassigned | Unassigned | 747_0 | Clustered | 23 | VC_747_0 | 23 | 0.8202 | 1 | 0.8202 | 0 | 0 | 0 | 1 | Ref |
| 8947 | MK511001 | Unassigned | Unassigned | Unassigned | 297_1 | Clustered | 20 | VC_297_1 | 20 | 0.5653 | 1 | 0.5653 | 0 | 0 | 0 | 0.7684 | Ref |
| 8948 | MK511002 | Unassigned | Unassigned | Unassigned | 297_1 | Clustered | 20 | VC_297_1 | 20 | 0.5653 | 1 | 0.5653 | 0 | 0 | 0 | 0.7684 | Ref |
| 8949 | MK511003 | Unassigned | Unassigned | Unassigned | 297_1 | Clustered | 20 | VC_297_1 | 20 | 0.5653 | 1 | 0.5653 | 0 | 0 | 0 | 0.7684 | Ref |
| 8950 | MK511004 | Unassigned | Unassigned | Unassigned | 297_1 | Clustered | 20 | VC_297_1 | 20 | 0.5653 | 1 | 0.5653 | 0 | 0 | 0 | 0.7684 | Ref |
| 8951 | MK511005 | Unassigned | Unassigned | Unassigned | 297_1 | Clustered | 20 | VC_297_1 | 20 | 0.5653 | 1 | 0.5653 | 0 | 0 | 0 | 0.7684 | Ref |
| 8952 | MK511006 | Unassigned | Unassigned | Unassigned | 297_1 | Clustered | 20 | VC_297_1 | 20 | 0.5653 | 1 | 0.5653 | 0 | 0 | 0 | 0.7684 | Ref |
| 8953 | MK511007 | Unassigned | Unassigned | Unassigned | 297_1 | Clustered | 20 | VC_297_1 | 20 | 0.5653 | 1 | 0.5653 | 0 | 0 | 0 | 0.7684 | Ref |
| 8954 | MK511008 | Unassigned | Unassigned | Unassigned | 297_1 | Clustered | 20 | VC_297_1 | 20 | 0.5653 | 1 | 0.5653 | 0 | 0 | 0 | 0.7684 | Ref |
| 8955 | MK511009 | Unassigned | Unassigned | Unassigned | 297_1 | Clustered | 20 | VC_297_1 | 20 | 0.5653 | 1 | 0.5653 | 0 | 0 | 0 | 0.7684 | Ref |
| 8956 | MK511010 | Unassigned | Unassigned | Unassigned | 297_1 | Clustered | 20 | VC_297_1 | 20 | 0.5653 | 1 | 0.5653 | 0 | 0 | 0 | 0.7684 | Ref |
| 8957 | MK511011 | Unassigned | Unassigned | Unassigned | 297_1 | Clustered | 20 | VC_297_1 | 20 | 0.5653 | 1 | 0.5653 | 0 | 0 | 0 | 0.7684 | Ref |
| 8958 | MK511012 | Unassigned | Unassigned | Unassigned | 297_1 | Clustered | 20 | VC_297_1 | 20 | 0.5653 | 1 | 0.5653 | 0 | 0 | 0 | 0.7684 | Ref |
| 8959 | MK511013 | Unassigned | Unassigned | Unassigned | 297_1 | Clustered | 20 | VC_297_1 | 20 | 0.5653 | 1 | 0.5653 | 0 | 0 | 0 | 0.7684 | Ref |
| 8960 | MK511014 | Unassigned | Unassigned | Unassigned | 297_1 | Clustered | 20 | VC_297_1 | 20 | 0.5653 | 1 | 0.5653 | 0 | 0 | 0 | 0.7684 | Ref |
| 8961 | MK511015 | Unassigned | Unassigned | Unassigned | 297_1 | Clustered | 20 | VC_297_1 | 20 | 0.5653 | 1 | 0.5653 | 0 | 0 | 0 | 0.7684 | Ref |
| 8962 | MK511016 | Unassigned | Unassigned | Unassigned | 297_1 | Clustered | 20 | VC_297_1 | 20 | 0.5653 | 1 | 0.5653 | 0 | 0 | 0 | 0.7684 | Ref |
| 8963 | MK511017 | Unassigned | Unassigned | Unassigned | 179_0 | Clustered | 53 | VC_179_0 | 53 | 0.9137 | 1 | 0.9137 | 0 | 0 | 0 | 1 | Ref |
| 8964 | MK511018 | Unassigned | Unassigned | Unassigned | 179_0 | Clustered | 53 | VC_179_0 | 53 | 0.9137 | 1 | 0.9137 | 0 | 0 | 0 | 1 | Ref |
| 8965 | MK511019 | Unassigned | Unassigned | Unassigned | 179_0 | Clustered | 53 | VC_179_0 | 53 | 0.9137 | 1 | 0.9137 | 0 | 0 | 0 | 1 | Ref |
| 8966 | MK511020 | Unassigned | Unassigned | Unassigned | 179_0 | Clustered | 53 | VC_179_0 | 53 | 0.9137 | 1 | 0.9137 | 0 | 0 | 0 | 1 | Ref |
| 8967 | MK511021 | Unassigned | Unassigned | Unassigned | 179_0 | Clustered | 53 | VC_179_0 | 53 | 0.9137 | 1 | 0.9137 | 0 | 0 | 0 | 1 | Ref |
| 8968 | MK511022 | Unassigned | Unassigned | Unassigned | 179_0 | Clustered | 53 | VC_179_0 | 53 | 0.9137 | 1 | 0.9137 | 0 | 0 | 0 | 1 | Ref |
| 8969 | MK511023 | Unassigned | Unassigned | Unassigned | 179_0 | Clustered | 53 | VC_179_0 | 53 | 0.9137 | 1 | 0.9137 | 0 | 0 | 0 | 1 | Ref |
| 8970 | MK511024 | Unassigned | Unassigned | Unassigned | 179_0 | Clustered | 53 | VC_179_0 | 53 | 0.9137 | 1 | 0.9137 | 0 | 0 | 0 | 1 | Ref |
| 8971 | MK511025 | Unassigned | Unassigned | Unassigned | 179_0 | Clustered | 53 | VC_179_0 | 53 | 0.9137 | 1 | 0.9137 | 0 | 0 | 0 | 1 | Ref |
| 8972 | MK511026 | Unassigned | Unassigned | Unassigned | 179_0 | Clustered | 53 | VC_179_0 | 53 | 0.9137 | 1 | 0.9137 | 0 | 0 | 0 | 1 | Ref |
| 8973 | MK511027 | Unassigned | Unassigned | Unassigned | 179_0 | Clustered | 53 | VC_179_0 | 53 | 0.9137 | 1 | 0.9137 | 0 | 0 | 0 | 1 | Ref |
| 8974 | MK511028 | Unassigned | Unassigned | Unassigned | 179_0 | Clustered | 53 | VC_179_0 | 53 | 0.9137 | 1 | 0.9137 | 0 | 0 | 0 | 1 | Ref |
| 8975 | MK511029 | Unassigned | Unassigned | Unassigned | 179_0 | Clustered | 53 | VC_179_0 | 53 | 0.9137 | 1 | 0.9137 | 0 | 0 | 0 | 1 | Ref |
| 8976 | MK511030 | Unassigned | Unassigned | Unassigned | 179_0 | Clustered | 53 | VC_179_0 | 53 | 0.9137 | 1 | 0.9137 | 0 | 0 | 0 | 1 | Ref |
| 8977 | MK511031 | Unassigned | Unassigned | Unassigned | 179_0 | Clustered | 53 | VC_179_0 | 53 | 0.9137 | 1 | 0.9137 | 0 | 0 | 0 | 1 | Ref |
| 8978 | MK511032 | Unassigned | Unassigned | Unassigned | 179_0 | Clustered | 53 | VC_179_0 | 53 | 0.9137 | 1 | 0.9137 | 0 | 0 | 0 | 1 | Ref |
| 8979 | MK511033 | Unassigned | Unassigned | Unassigned | 179_0 | Clustered | 53 | VC_179_0 | 53 | 0.9137 | 1 | 0.9137 | 0 | 0 | 0 | 1 | Ref |
| 8980 | MK511034 | Unassigned | Unassigned | Unassigned | 179_0 | Clustered | 53 | VC_179_0 | 53 | 0.9137 | 1 | 0.9137 | 0 | 0 | 0 | 1 | Ref |
| 8981 | MK511035 | Unassigned | Unassigned | Unassigned | 179_0 | Clustered | 53 | VC_179_0 | 53 | 0.9137 | 1 | 0.9137 | 0 | 0 | 0 | 1 | Ref |
| 8982 | MK511036 | Unassigned | Unassigned | Unassigned | 179_0 | Clustered | 53 | VC_179_0 | 53 | 0.9137 | 1 | 0.9137 | 0 | 0 | 0 | 1 | Ref |
| 8983 | MK511037 | Unassigned | Unassigned | Unassigned | 179_0 | Clustered | 53 | VC_179_0 | 53 | 0.9137 | 1 | 0.9137 | 0 | 0 | 0 | 1 | Ref |
| 8984 | MK511038 | Unassigned | Unassigned | Unassigned | 179_0 | Clustered | 53 | VC_179_0 | 53 | 0.9137 | 1 | 0.9137 | 0 | 0 | 0 | 1 | Ref |
| 8985 | MK511039 | Unassigned | Unassigned | Unassigned | 630_0 | Clustered | 20 | VC_630_0 | 20 | 0.8513 | 1 | 0.8513 | 0 | 0 | 0 | 1 | Ref |
| 8986 | MK511040 | Unassigned | Unassigned | Unassigned | 630_0 | Clustered | 20 | VC_630_0 | 20 | 0.8513 | 1 | 0.8513 | 0 | 0 | 0 | 1 | Ref |
| 8987 | MK511041 | Unassigned | Unassigned | Unassigned | 630_0 | Clustered | 20 | VC_630_0 | 20 | 0.8513 | 1 | 0.8513 | 0 | 0 | 0 | 1 | Ref |
| 8988 | MK511042 | Unassigned | Unassigned | Unassigned | 630_0 | Clustered | 20 | VC_630_0 | 20 | 0.8513 | 1 | 0.8513 | 0 | 0 | 0 | 1 | Ref |
| 8989 | MK511043 | Unassigned | Unassigned | Unassigned | 630_0 | Clustered | 20 | VC_630_0 | 20 | 0.8513 | 1 | 0.8513 | 0 | 0 | 0 | 1 | Ref |
| 8990 | MK511044 | Unassigned | Unassigned | Unassigned | 630_0 | Clustered | 20 | VC_630_0 | 20 | 0.8513 | 1 | 0.8513 | 0 | 0 | 0 | 1 | Ref |
| 8991 | MK511045 | Unassigned | Unassigned | Unassigned | 630_0 | Clustered | 20 | VC_630_0 | 20 | 0.8513 | 1 | 0.8513 | 0 | 0 | 0 | 1 | Ref |
| 8992 | MK511046 | Unassigned | Unassigned | Unassigned | 630_0 | Clustered | 20 | VC_630_0 | 20 | 0.8513 | 1 | 0.8513 | 0 | 0 | 0 | 1 | Ref |
| 8993 | MK511047 | Unassigned | Unassigned | Unassigned | 630_0 | Clustered | 20 | VC_630_0 | 20 | 0.8513 | 1 | 0.8513 | 0 | 0 | 0 | 1 | Ref |
| 8994 | MK511048 | Unassigned | Unassigned | Unassigned | 630_0 | Clustered | 20 | VC_630_0 | 20 | 0.8513 | 1 | 0.8513 | 0 | 0 | 0 | 1 | Ref |
| 8995 | MK511049 | Unassigned | Unassigned | Unassigned | 630_0 | Clustered | 20 | VC_630_0 | 20 | 0.8513 | 1 | 0.8513 | 0 | 0 | 0 | 1 | Ref |
| 8996 | MK511050 | Unassigned | Unassigned | Unassigned | 630_0 | Clustered | 20 | VC_630_0 | 20 | 0.8513 | 1 | 0.8513 | 0 | 0 | 0 | 1 | Ref |
| 8997 | MK511051 | Unassigned | Unassigned | Unassigned | 630_0 | Clustered | 20 | VC_630_0 | 20 | 0.8513 | 1 | 0.8513 | 0 | 0 | 0 | 1 | Ref |
| 8998 | MK511057 | Unassigned | Unassigned | Unassigned | 251_0 | Clustered | 22 | VC_251_0 | 22 | 0.6508 | 1 | 0.6508 | 0 | 0 | 0 | 1 | Ref |
| 8999 | MK511058 | Unassigned | Unassigned | Unassigned | 251_0 | Clustered | 22 | VC_251_0 | 22 | 0.6508 | 1 | 0.6508 | 0 | 0 | 0 | 1 | Ref |
| 9000 | MK511059 | Unassigned | Unassigned | Unassigned | 251_0 | Clustered | 22 | VC_251_0 | 22 | 0.6508 | 1 | 0.6508 | 0 | 0 | 0 | 1 | Ref |
| 9001 | MK511060 | Unassigned | Unassigned | Unassigned | 251_0 | Clustered | 22 | VC_251_0 | 22 | 0.6508 | 1 | 0.6508 | 0 | 0 | 0 | 1 | Ref |
| 9002 | MK511061 | Unassigned | Unassigned | Unassigned | 251_0 | Clustered | 22 | VC_251_0 | 22 | 0.6508 | 1 | 0.6508 | 0 | 0 | 0 | 1 | Ref |
| 9003 | MK511063 | Unassigned | Unassigned | Unassigned | 251_0 | Clustered | 22 | VC_251_0 | 22 | 0.6508 | 1 | 0.6508 | 0 | 0 | 0 | 1 | Ref |
| 9004 | MK511064 | Unassigned | Unassigned | Unassigned | 251_0 | Clustered | 22 | VC_251_0 | 22 | 0.6508 | 1 | 0.6508 | 0 | 0 | 0 | 1 | Ref |
| 9005 | MK511065 | Unassigned | Unassigned | Unassigned | 251_0 | Clustered | 22 | VC_251_0 | 22 | 0.6508 | 1 | 0.6508 | 0 | 0 | 0 | 1 | Ref |
| 9006 | MK511066 | Unassigned | Unassigned | Unassigned | 2_0 | Clustered | 4 | VC_2_0 | 4 | 0.1311 | 0.99998692 | 0.1311 | 0 | 0 | 0 | 1 | Ref |
| 9007 | MK512531 | Unassigned | Unassigned | Unassigned | 178_0 | Clustered | 5 | VC_178_0 | 5 | 0.7741 | 0.99998991 | 0.7741 | 0 | 0 | 0 | 1 | Ref |
| 9008 | MK514281 | Unassigned | Unassigned | Unassigned | 489_1 | Clustered | 11 | VC_489_1 | 11 | 0.4621 | 1 | 0.4621 | 0 | 0 | 0 | 1 | Ref |
| 9009 | MK514282 | Unassigned | Unassigned | Unassigned | 959_0 | Clustered | 5 | VC_959_0 | 5 | 0.0499 | 0.99999231 | 0.0499 | 0 | 0 | 0 | 1 | Ref |
| 9010 | MK514283 | Unassigned | Unassigned | Unassigned | 612_0 | Clustered | 9 | VC_612_0 | 9 | 0.9545 | 1 | 0.9545 | 0 | 0 | 0 | 1 | Ref |
| 9011 | MK521904 | Unassigned | Unassigned | Unassigned | 938_0 | Clustered | 99 | VC_938_0 | 99 | 0.9119 | 1 | 0.9119 | 0 | 0 | 0 | 0.9928 | Ref |
| 9012 | MK521905 | Unassigned | Unassigned | Unassigned | 823_0 | Clustered | 124 | VC_823_0 | 124 | 0.9492 | 1 | 0.9492 | 0 | 0 | 0 | 1 | Ref |
| 9013 | MK521906 | Unassigned | Unassigned | Unassigned | 123_0 | Clustered | 23 | VC_123_0 | 23 | 0.817 | 1 | 0.817 | 0 | 0 | 0 | 1 | Ref |
| 9014 | MK521907 | Unassigned | Unassigned | Unassigned | 192_0 | Clustered | 15 | VC_192_0 | 15 | 0.5071 | 1 | 0.5071 | 0 | 0 | 0 | 1 | Ref |
| 9015 | MK524176 | Unassigned | Unassigned | Unassigned | 329_18 | Clustered | 7 | VC_329_18 | 7 | 0.0328 | 1 | 0.0328 | 0 | 0 | 0 | 0.8095 | Ref |
| 9016 | MK524177 | Unassigned | Unassigned | Unassigned | 230_0 | Clustered | 3 | VC_230_0 | 3 | 0.0739 | 0.9934951 | 0.0734 | 0 | 0 | 0 | 0.6667 | Ref |
| 9017 | MK524178 | Unassigned | Unassigned | Unassigned | 286_11 | Clustered | 3 | VC_286_11 | 3 | 0.0146 | 0.93460284 | 0.0137 | 0 | 0 | 0 | 1 | Ref |
| 9018 | MK524485 | Unassigned | Unassigned | Unassigned | 234_0 | Clustered | 21 | VC_234_0 | 21 | 0.8355 | 1 | 0.8355 | 0 | 0 | 0 | 1 | Ref |
| 9019 | MK524486 | Unassigned | Unassigned | Unassigned | 372_0 | Clustered | 24 | VC_372_0 | 24 | 0.5539 | 1 | 0.5539 | 0 | 0 | 0 | 1 | Ref |
| 9020 | MK524487 | Unassigned | Unassigned | Unassigned | 956_0 | Clustered | 39 | VC_956_0 | 39 | 1 | 1 | 1 | 0 | 0 | 0 | 1 | Ref |
| 9021 | MK524488 | Unassigned | Unassigned | Unassigned | 234_0 | Clustered | 21 | VC_234_0 | 21 | 0.8355 | 1 | 0.8355 | 0 | 0 | 0 | 1 | Ref |
| 9022 | MK524489 | Unassigned | Unassigned | Unassigned | 225_0 | Clustered | 32 | VC_225_0 | 32 | 0.848 | 1 | 0.848 | 0 | 0 | 0 | 0.996 | Ref |
| 9023 | MK524490 | Unassigned | Unassigned | Unassigned | 372_1 | Clustered | 9 | VC_372_1 | 9 | 0.194 | 1 | 0.194 | 0 | 0 | 0 | 1 | Ref |
| 9024 | MK524491 | Unassigned | Unassigned | Unassigned | 216_6 | Clustered | 42 | VC_216_6 | 42 | 0.473 | 1 | 0.473 | 0 | 0 | 0 | 0.8351 | Ref |
| 9025 | MK524493 | Unassigned | Unassigned | Unassigned | 761_0 | Clustered | 67 | VC_761_0 | 67 | 0.8947 | 1 | 0.8947 | 0 | 0 | 0 | 1 | Ref |
| 9026 | MK524494 | Unassigned | Unassigned | Unassigned | 761_0 | Clustered | 67 | VC_761_0 | 67 | 0.8947 | 1 | 0.8947 | 0 | 0 | 0 | 1 | Ref |
| 9027 | MK524495 | Unassigned | Unassigned | Unassigned | 956_0 | Clustered | 39 | VC_956_0 | 39 | 1 | 1 | 1 | 0 | 0 | 0 | 1 | Ref |
| 9028 | MK524496 | Unassigned | Unassigned | Unassigned | 513_0 | Clustered | 32 | VC_513_0 | 32 | 0.6881 | 1 | 0.6881 | 0 | 0 | 0 | 1 | Ref |
| 9029 | MK524497 | Unassigned | Unassigned | Unassigned | 325_21 | Clustered | 5 | VC_325_21 | 5 | 0.0209 | 0.99999976 | 0.0209 | 0 | 0 | 0 | 0.8 | Ref |
| 9030 | MK524498 | Unassigned | Unassigned | Unassigned | 565_0 | Clustered | 20 | VC_565_0 | 20 | 0.8398 | 1 | 0.8398 | 0 | 0 | 0 | 1 | Ref |
| 9031 | MK524499 | Unassigned | Unassigned | Unassigned | 325_22 | Clustered/Singleton | 1 | VC_325_22 | 1 | 0 | 1 | 0 | 0 | 0 | 0 | 1 | Ref |
| 9032 | MK524500 | Unassigned | Unassigned | Unassigned | 521_0 | Clustered | 37 | VC_521_0 | 37 | 0.912 | 1 | 0.912 | 0 | 0 | 0 | 1 | Ref |
| 9033 | MK524501 | Unassigned | Unassigned | Unassigned | 1181_0 | Clustered | 2 | VC_1181_0 | 2 | 0.0461 | 0.95126432 | 0.0439 | 0 | 0 | 0 | 1 | Ref |
| 9034 | MK524502 | Unassigned | Unassigned | Unassigned | 1344_0 | Clustered | 78 | VC_1344_0 | 78 | 0.9732 | 1 | 0.9732 | 0 | 0 | 0 | 1 | Ref |
| 9035 | MK524503 | Unassigned | Unassigned | Unassigned | 520_87 | Clustered/Singleton | 1 | VC_520_87 | 1 | 0 | 1 | 0 | 0 | 0 | 0 | 1 | Ref |
| 9036 | MK524504 | Unassigned | Unassigned | Unassigned | 546_4 | Clustered | 16 | VC_546_4 | 16 | 0.2517 | 0.9999998 | 0.2517 | 0 | 0 | 0 | 0.9417 | Ref |
| 9037 | MK524505 | Unassigned | Unassigned | Unassigned | 1184_0 | Clustered | 5 | VC_1184_0 | 5 | 0.6591 | 0.99999982 | 0.6591 | 0 | 0 | 0 | 1 | Ref |
| 9038 | MK524506 | Unassigned | Unassigned | Unassigned | 325_35 | Clustered | 17 | VC_325_35 | 17 | 0.0732 | 1 | 0.0732 | 0 | 0 | 0 | 0.7721 | Ref |
| 9039 | MK524507 | Unassigned | Unassigned | Unassigned | 221_0 | Clustered | 22 | VC_221_0 | 22 | 0.7447 | 1 | 0.7447 | 0 | 0 | 0 | 1 | Ref |
| 9040 | MK524508 | Unassigned | Unassigned | Unassigned | 519_0 | Clustered | 39 | VC_519_0 | 39 | 0.8156 | 1 | 0.8156 | 0 | 0 | 0 | 1 | Ref |
| 9041 | MK524509 | Unassigned | Unassigned | Unassigned | 521_0 | Clustered | 37 | VC_521_0 | 37 | 0.912 | 1 | 0.912 | 0 | 0 | 0 | 1 | Ref |
| 9042 | MK524510 | Unassigned | Unassigned | Unassigned | 1352_0 | Clustered | 31 | VC_1352_0 | 31 | 0.849 | 1 | 0.849 | 0 | 0 | 0 | 1 | Ref |
| 9043 | MK524511 | Unassigned | Unassigned | Unassigned | 221_0 | Clustered | 22 | VC_221_0 | 22 | 0.7447 | 1 | 0.7447 | 0 | 0 | 0 | 1 | Ref |
| 9044 | MK524512 | Unassigned | Unassigned | Unassigned | 371_1 | Clustered | 11 | VC_371_1 | 11 | 0.0379 | 1 | 0.0379 | 0 | 0 | 0 | 0.8545 | Ref |
| 9045 | MK524513 | Unassigned | Unassigned | Unassigned | 221_0 | Clustered | 22 | VC_221_0 | 22 | 0.7447 | 1 | 0.7447 | 0 | 0 | 0 | 1 | Ref |
| 9046 | MK524514 | Unassigned | Unassigned | Unassigned | 221_0 | Clustered | 22 | VC_221_0 | 22 | 0.7447 | 1 | 0.7447 | 0 | 0 | 0 | 1 | Ref |
| 9047 | MK524515 | Unassigned | Unassigned | Unassigned | 521_0 | Clustered | 37 | VC_521_0 | 37 | 0.912 | 1 | 0.912 | 0 | 0 | 0 | 1 | Ref |
| 9048 | MK524516 | Unassigned | Unassigned | Unassigned | 546_9 | Clustered/Singleton | 1 | VC_546_9 | 1 | 0 | 1 | 0 | 0 | 0 | 0 | 1 | Ref |
| 9049 | MK524517 | Unassigned | Unassigned | Unassigned | 520_38 | Clustered | 6 | VC_520_38 | 6 | 0.0519 | 1 | 0.0519 | 0 | 0 | 0 | 0.8667 | Ref |
| 9050 | MK524518 | Unassigned | Unassigned | Unassigned | 521_0 | Clustered | 37 | VC_521_0 | 37 | 0.912 | 1 | 0.912 | 0 | 0 | 0 | 1 | Ref |
| 9051 | MK524519 | Unassigned | Unassigned | Unassigned | 221_0 | Clustered | 22 | VC_221_0 | 22 | 0.7447 | 1 | 0.7447 | 0 | 0 | 0 | 1 | Ref |
| 9052 | MK524520 | Unassigned | Unassigned | Unassigned | 521_0 | Clustered | 37 | VC_521_0 | 37 | 0.912 | 1 | 0.912 | 0 | 0 | 0 | 1 | Ref |
| 9053 | MK524521 | Unassigned | Unassigned | Unassigned | 546_4 | Clustered | 16 | VC_546_4 | 16 | 0.2517 | 0.9999998 | 0.2517 | 0 | 0 | 0 | 0.9417 | Ref |
| 9054 | MK524522 | Unassigned | Unassigned | Unassigned | 521_0 | Clustered | 37 | VC_521_0 | 37 | 0.912 | 1 | 0.912 | 0 | 0 | 0 | 1 | Ref |
| 9055 | MK524523 | Unassigned | Unassigned | Unassigned | 325_16 | Clustered | 2 | VC_325_16 | 2 | 0.0095 | 0.95751393 | 0.0091 | 0 | 0 | 0 | 1 | Ref |
| 9056 | MK524524 | Unassigned | Unassigned | Unassigned | 981_1 | Clustered | 26 | VC_981_1 | 26 | 0.7009 | 1 | 0.7009 | 0 | 0 | 0 | 1 | Ref |
| 9057 | MK524525 | Unassigned | Unassigned | Unassigned | 325_45 | Clustered/Singleton | 1 | VC_325_45 | 1 | 0 | 1 | 0 | 0 | 0 | 0 | 1 | Ref |
| 9058 | MK524526 | Unassigned | Unassigned | Unassigned | 371_1 | Clustered | 11 | VC_371_1 | 11 | 0.0379 | 1 | 0.0379 | 0 | 0 | 0 | 0.8545 | Ref |
| 9059 | MK524527 | Unassigned | Unassigned | Unassigned | 546_4 | Clustered | 16 | VC_546_4 | 16 | 0.2517 | 0.9999998 | 0.2517 | 0 | 0 | 0 | 0.9417 | Ref |
| 9060 | MK524528 | Unassigned | Unassigned | Unassigned | 521_0 | Clustered | 37 | VC_521_0 | 37 | 0.912 | 1 | 0.912 | 0 | 0 | 0 | 1 | Ref |
| 9061 | MK524529 | Unassigned | Unassigned | Unassigned | 546_4 | Clustered | 16 | VC_546_4 | 16 | 0.2517 | 0.9999998 | 0.2517 | 0 | 0 | 0 | 0.9417 | Ref |
| 9062 | MK524530 | Unassigned | Unassigned | Unassigned | 222_0 | Clustered | 5 | VC_222_0 | 5 | 0.0565 | 0.99999997 | 0.0565 | 0 | 0 | 0 | 1 | Ref |
| 9063 | MK524531 | Unassigned | Unassigned | Unassigned | 325_35 | Clustered | 17 | VC_325_35 | 17 | 0.0732 | 1 | 0.0732 | 0 | 0 | 0 | 0.7721 | Ref |
| 9064 | MK542015 | Unassigned | Unassigned | Unassigned | 638_0 | Clustered | 45 | VC_638_0 | 45 | 0.7741 | 1 | 0.7741 | 0 | 0 | 0 | 1 | Ref |
| 9065 | MK542821 | Unassigned | Unassigned | Unassigned | 392_0 | Clustered | 126 | VC_392_0 | 126 | 0.9621 | 1 | 0.9621 | 0 | 0 | 0 | 1 | Ref |
| 9066 | MK552140 | Unassigned | Unassigned | Unassigned | 66_0 | Clustered | 2 | VC_66_0 | 2 | 0.1995 | 0.95487418 | 0.1905 | 0 | 0 | 0 | 1 | Ref |
| 9067 | MK552141 | Unassigned | Unassigned | Unassigned | 66_0 | Clustered | 2 | VC_66_0 | 2 | 0.1995 | 0.95487418 | 0.1905 | 0 | 0 | 0 | 1 | Ref |
| 9068 | MK552327 | Unassigned | Unassigned | Unassigned | 344_3 | Clustered/Singleton | 1 | VC_344_3 | 1 | 0 | 1 | 0 | 0 | 0 | 0 | 1 | Ref |
| 9069 | MK559427 | Unassigned | Unassigned | Unassigned | 565_0 | Clustered | 20 | VC_565_0 | 20 | 0.8398 | 1 | 0.8398 | 0 | 0 | 0 | 1 | Ref |
| 9070 | MK559428 | Unassigned | Unassigned | Unassigned | 225_0 | Clustered | 32 | VC_225_0 | 32 | 0.848 | 1 | 0.848 | 0 | 0 | 0 | 0.996 | Ref |

| | | | | | | | | | | | | | | | | | |
|---|---|---|---|---|---|---|---|---|---|---|---|---|---|---|---|---|---|
| 9071 | MK559429 | Unassigned | Unassigned | Unassigned | 551_6 | Clustered | 22 | VC_551_6 | 22 | 0.1214 | 1 | 0.1214 | 0 | 0 | 0 | 0.8095 | Ref |
| 9072 | MK559459 | Unassigned | Unassigned | Unassigned | 904_0 | Clustered | 6 | VC_904_0 | 6 | 0.233 | 1 | 0.233 | 0 | 0 | 0 | 1 | Ref |
| 9073 | MK562502 | Unassigned | Unassigned | Unassigned | 330_0 | Clustered | 27 | VC_330_0 | 27 | 0.5088 | 1 | 0.5088 | 0 | 0 | 0 | 0.8661 | Ref |
| 9074 | MK562503 | Unassigned | Unassigned | Unassigned | 111_0 | Clustered | 91 | VC_111_0 | 91 | 0.9587 | 1 | 0.9587 | 0 | 0 | 0 | 1 | Ref |
| 9075 | MK562504 | Unassigned | Unassigned | Unassigned | 330_0 | Clustered | 27 | VC_330_0 | 27 | 0.5088 | 1 | 0.5088 | 0 | 0 | 0 | 0.8661 | Ref |
| 9076 | MK562505 | Unassigned | Unassigned | Unassigned | 330_0 | Clustered | 27 | VC_330_0 | 27 | 0.5088 | 1 | 0.5088 | 0 | 0 | 0 | 0.8661 | Ref |
| 9077 | MK568062 | Unassigned | Unassigned | Unassigned | 428_0 | Clustered | 33 | VC_428_0 | 33 | 0.9915 | 1 | 0.9915 | 0 | 0 | 0 | 1 | Ref |
| 9078 | MK568540 | Unassigned | Unassigned | Unassigned | 531_0 | Clustered | 9 | VC_531_0 | 9 | 0.26 | 1 | 0.26 | 0 | 0 | 0 | 1 | Ref |
| 9079 | MK570053 | Unassigned | Unassigned | Unassigned | 1395_0 | Clustered | 3 | VC_1395_0 | 3 | 0.7761 | 0.9639009 | 0.7481 | 0 | 0 | 0 | 1 | Ref |
| 9080 | MK570054 | Unassigned | Unassigned | Unassigned | 1395_0 | Clustered | 3 | VC_1395_0 | 3 | 0.7761 | 0.9639009 | 0.7481 | 0 | 0 | 0 | 1 | Ref |
| 9081 | MK570055 | Unassigned | Unassigned | Unassigned | 1395_0 | Clustered | 3 | VC_1395_0 | 3 | 0.7761 | 0.9639009 | 0.7481 | 0 | 0 | 0 | 1 | Ref |
| 9082 | MK570225 | Unassigned | Unassigned | Unassigned | 108_0 | Clustered | 33 | VC_108_0 | 33 | 0.949 | 1 | 0.949 | 0 | 0 | 0 | 1 | Ref |
| 9083 | MK573636 | Unassigned | Unassigned | Unassigned | 590_0 | Clustered | 27 | VC_590_0 | 27 | 0.9947 | 1 | 0.9947 | 0 | 0 | 0 | 1 | Ref |
| 9084 | MK574011 | Unassigned | Unassigned | Unassigned | 117_0 | Clustered | 9 | VC_117_0 | 9 | 0.9977 | 0.98988637 | 0.9876 | 0 | 0 | 0 | 1 | Ref |
| 9085 | MK574078 | Unassigned | Unassigned | Unassigned | 749_0 | Clustered | 40 | VC_749_0 | 40 | 0.7517 | 1 | 0.7517 | 0 | 0 | 0 | 1 | Ref |
| 9086 | MK577502 | Unassigned | Unassigned | Unassigned | 600_0 | Clustered | 3 | VC_600_0 | 3 | 0.0541 | 0.97110069 | 0.0526 | 0 | 0 | 0 | 1 | Ref |
| 9087 | MK578530 | Unassigned | Unassigned | Unassigned | 637_1 | Clustered | 27 | VC_637_1 | 27 | 0.2578 | 1 | 0.2578 | 0 | 0 | 0 | 0.9858 | Ref |
| 9088 | MK580972 | Unassigned | Unassigned | Unassigned | 506_0 | Clustered | 8 | VC_506_0 | 8 | 0.7376 | 1 | 0.7376 | 0 | 0 | 0 | 1 | Ref |
| 9089 | MK584893 | Unassigned | Unassigned | Unassigned | 149_8 | Clustered | 66 | VC_149_8 | 66 | 0.4565 | 1 | 0.4565 | 0 | 0 | 0 | 0.8182 | Ref |
| 9090 | MK598851 | Unassigned | Unassigned | Unassigned | 111_0 | Clustered | 91 | VC_111_0 | 91 | 0.9587 | 1 | 0.9587 | 0 | 0 | 0 | 1 | Ref |
| 9091 | MK599315 | Unassigned | Unassigned | Unassigned | 344_1 | Clustered | 3 | VC_344_1 | 3 | 0.0359 | 0.93348877 | 0.0335 | 0 | 0 | 0 | 1 | Ref |
| 9092 | MK599416 | Unassigned | Unassigned | Unassigned | 461_0 | Clustered | 96 | VC_461_0 | 96 | 0.9307 | 1 | 0.9307 | 0 | 0 | 0 | 0.9575 | Ref |
| 9093 | MK605242 | Unassigned | Unassigned | Unassigned | 1173_0 | Clustered | 6 | VC_1173_0 | 6 | 0.9951 | 0.99999296 | 0.9951 | 0 | 0 | 0 | 1 | Ref |
| 9094 | MK605243 | Unassigned | Unassigned | Unassigned | 1173_0 | Clustered | 6 | VC_1173_0 | 6 | 0.9951 | 0.99999296 | 0.9951 | 0 | 0 | 0 | 1 | Ref |
| 9095 | MK605244 | Unassigned | Unassigned | Unassigned | 1173_0 | Clustered | 6 | VC_1173_0 | 6 | 0.9951 | 0.99999296 | 0.9951 | 0 | 0 | 0 | 1 | Ref |
| 9096 | MK605245 | Unassigned | Unassigned | Unassigned | 1173_0 | Clustered | 6 | VC_1173_0 | 6 | 0.9951 | 0.99999296 | 0.9951 | 0 | 0 | 0 | 1 | Ref |
| 9097 | MK605246 | Unassigned | Unassigned | Unassigned | 1173_0 | Clustered | 6 | VC_1173_0 | 6 | 0.9951 | 0.99999296 | 0.9951 | 0 | 0 | 0 | 1 | Ref |
| 9098 | MK608335 | Unassigned | Unassigned | Unassigned | 385_0 | Clustered | 51 | VC_385_0 | 51 | 0.8754 | 1 | 0.8754 | 0 | 0 | 0 | 1 | Ref |
| 9099 | MK608336 | Unassigned | Unassigned | Unassigned | 966_0 | Clustered | 3 | VC_966_0 | 3 | 0.3549 | 0.99840333 | 0.3543 | 0 | 0 | 0 | 1 | Ref |
| 9100 | MK610268 | Unassigned | Unassigned | Unassigned | 451_0 | Clustered | 80 | VC_451_0 | 80 | 0.9991 | 1 | 0.9991 | 0 | 0 | 0 | 1 | Ref |
| 9101 | MK613343 | Unassigned | Unassigned | Unassigned | 305_0 | Clustered | 12 | VC_305_0 | 12 | 0.5745 | 1 | 0.5745 | 0 | 0 | 0 | 0.8485 | Ref |
| 9102 | MK613344 | Unassigned | Unassigned | Unassigned | 305_0 | Clustered | 12 | VC_305_0 | 12 | 0.5745 | 1 | 0.5745 | 0 | 0 | 0 | 0.8485 | Ref |
| 9103 | MK613345 | Unassigned | Unassigned | Unassigned | 305_0 | Clustered | 12 | VC_305_0 | 12 | 0.5745 | 1 | 0.5745 | 0 | 0 | 0 | 0.8485 | Ref |
| 9104 | MK613346 | Unassigned | Unassigned | Unassigned | 304_0 | Clustered | 11 | VC_304_0 | 11 | 0.7869 | 1 | 0.7869 | 0 | 0 | 0 | 1 | Ref |
| 9105 | MK618657 | Unassigned | Unassigned | Unassigned | 938_0 | Clustered | 99 | VC_938_0 | 99 | 0.9119 | 1 | 0.9119 | 0 | 0 | 0 | 0.9928 | Ref |
| 9106 | MK618658 | Unassigned | Unassigned | Unassigned | 823_0 | Clustered | 124 | VC_823_0 | 124 | 0.9492 | 1 | 0.9492 | 0 | 0 | 0 | 1 | Ref |
| 9107 | MK618715 | Unassigned | Unassigned | Unassigned | 359_0 | Clustered | 7 | VC_359_0 | 7 | 0.3703 | 1 | 0.3703 | 0 | 0 | 0 | 1 | Ref |
| 9108 | MK618716 | Unassigned | Unassigned | Unassigned | 12_0 | Clustered | 21 | VC_12_0 | 21 | 0.2274 | 1 | 0.2274 | 0 | 0 | 0 | 0.9571 | Ref |
| 9109 | MK620893 | Unassigned | Unassigned | Unassigned | 551_7 | Clustered | 28 | VC_551_7 | 28 | 0.1661 | 1 | 0.1661 | 0 | 0 | 0 | 0.8519 | Ref |
| 9110 | MK620894 | Unassigned | Unassigned | Unassigned | 1158_0 | Clustered | 3 | VC_1158_0 | 3 | 0.3179 | 0.99335558 | 0.3158 | 0 | 0 | 0 | 1 | Ref |
| 9111 | MK620895 | Unassigned | Unassigned | Unassigned | 1240_0 | Clustered | 82 | VC_1240_0 | 82 | 0.7945 | 1 | 0.7945 | 0 | 0 | 0 | 1 | Ref |
| 9112 | MK620896 | Unassigned | Unassigned | Unassigned | 1072_0 | Clustered | 2 | VC_1072_0 | 2 | 0.0264 | 0.95117401 | 0.0251 | 0 | 0 | 0 | 1 | Ref |
| 9113 | MK620897 | Unassigned | Unassigned | Unassigned | 1344_0 | Clustered | 78 | VC_1344_0 | 78 | 0.9732 | 1 | 0.9732 | 0 | 0 | 0 | 1 | Ref |
| 9114 | MK620898 | Unassigned | Unassigned | Unassigned | 508_0 | Clustered | 32 | VC_508_0 | 32 | 0.906 | 1 | 0.906 | 0 | 0 | 0 | 1 | Ref |
| 9115 | MK620900 | Unassigned | Unassigned | Unassigned | 1369_0 | Clustered | 3 | VC_1369_0 | 3 | 0.9809 | 0.9595722 | 0.9412 | 0 | 0 | 0 | 1 | Ref |
| 9116 | MK620901 | Unassigned | Unassigned | Unassigned | 1163_0 | Clustered | 28 | VC_1163_0 | 28 | 0.4923 | 1 | 0.4923 | 0 | 0 | 0 | 1 | Ref |
| 9117 | MK620902 | Unassigned | Unassigned | Unassigned | 1240_0 | Clustered | 82 | VC_1240_0 | 82 | 0.7945 | 1 | 0.7945 | 0 | 0 | 0 | 1 | Ref |
| 9118 | MK620903 | Unassigned | Unassigned | Unassigned | 1240_0 | Clustered | 82 | VC_1240_0 | 82 | 0.7945 | 1 | 0.7945 | 0 | 0 | 0 | 1 | Ref |
| 9119 | MK620904 | Unassigned | Unassigned | Unassigned | 1163_0 | Clustered | 28 | VC_1163_0 | 28 | 0.4923 | 1 | 0.4923 | 0 | 0 | 0 | 1 | Ref |
| 9120 | MK620905 | Unassigned | Unassigned | Unassigned | 1240_0 | Clustered | 82 | VC_1240_0 | 82 | 0.7945 | 1 | 0.7945 | 0 | 0 | 0 | 1 | Ref |
| 9121 | MK629527 | Unassigned | Unassigned | Unassigned | 743_0 | Clustered | 15 | VC_743_0 | 15 | 1 | 1 | 1 | 0 | 0 | 0 | 1 | Ref |
| 9122 | MK629528 | Unassigned | Unassigned | Unassigned | 156_0 | Clustered | 12 | VC_156_0 | 12 | 0.9062 | 1 | 0.9062 | 0 | 0 | 0 | 1 | Ref |
| 9123 | MK630230 | Unassigned | Unassigned | Unassigned | 192_0 | Clustered | 15 | VC_192_0 | 15 | 0.5071 | 1 | 0.5071 | 0 | 0 | 0 | 1 | Ref |
| 9124 | MK637516 | Unassigned | Unassigned | Unassigned | 969_0 | Clustered | 3 | VC_969_0 | 3 | 0.8314 | 0.99762851 | 0.8295 | 0 | 0 | 0 | 1 | Ref |
| 9125 | MK639187 | Unassigned | Unassigned | Unassigned | 286_13 | Clustered | 10 | VC_286_13 | 10 | 0.0698 | 0.99999929 | 0.0698 | 0 | 0 | 0 | 0.9556 | Ref |
| 9126 | MK651787 | Unassigned | Unassigned | Unassigned | 705_0 | Clustered | 11 | VC_705_0 | 11 | 0.8861 | 1 | 0.8861 | 0 | 0 | 0 | 1 | Ref |
| 9127 | MK656892 | Unassigned | Unassigned | Unassigned | 149_8 | Clustered | 66 | VC_149_8 | 66 | 0.4565 | 1 | 0.4565 | 0 | 0 | 0 | 0.8182 | Ref |
| 9128 | MK658834 | Unassigned | Unassigned | Unassigned | 9_0 | Clustered | 38 | VC_9_0 | 38 | 0.4895 | 1 | 0.4895 | 0 | 0 | 0 | 1 | Ref |
| 9129 | MK660712 | Unassigned | Unassigned | Unassigned | 949_0 | Clustered | 10 | VC_949_0 | 10 | 0.5977 | 1 | 0.5977 | 0 | 0 | 0 | 1 | Ref |
| 9130 | MK660714 | Unassigned | Unassigned | Unassigned | 1160_0 | Clustered | 73 | VC_1160_0 | 73 | 0.9835 | 1 | 0.9835 | 0 | 0 | 0 | 1 | Ref |
| 9131 | MK672798 | Unassigned | Unassigned | Unassigned | 795_0 | Clustered | 19 | VC_795_0 | 19 | 0.6848 | 1 | 0.6848 | 0 | 0 | 0 | 1 | Ref |
| 9132 | MK672799 | Unassigned | Unassigned | Unassigned | 299_0 | Clustered | 24 | VC_299_0 | 24 | 0.9905 | 1 | 0.9905 | 0 | 0 | 0 | 1 | Ref |
| 9133 | MK672800 | Unassigned | Unassigned | Unassigned | 299_0 | Clustered | 24 | VC_299_0 | 24 | 0.9905 | 1 | 0.9905 | 0 | 0 | 0 | 1 | Ref |
| 9134 | MK672801 | Unassigned | Unassigned | Unassigned | 299_0 | Clustered | 24 | VC_299_0 | 24 | 0.9905 | 1 | 0.9905 | 0 | 0 | 0 | 1 | Ref |
| 9135 | MK672803 | Unassigned | Unassigned | Unassigned | 299_0 | Clustered | 24 | VC_299_0 | 24 | 0.9905 | 1 | 0.9905 | 0 | 0 | 0 | 1 | Ref |
| 9136 | MK672804 | Unassigned | Unassigned | Unassigned | 299_0 | Clustered | 24 | VC_299_0 | 24 | 0.9905 | 1 | 0.9905 | 0 | 0 | 0 | 1 | Ref |
| 9137 | MK673511 | Unassigned | Unassigned | Unassigned | 428_0 | Clustered | 33 | VC_428_0 | 33 | 0.9915 | 1 | 0.9915 | 0 | 0 | 0 | 1 | Ref |
| 9138 | MK685667 | Unassigned | Unassigned | Unassigned | 123_0 | Clustered | 23 | VC_123_0 | 23 | 0.817 | 1 | 0.817 | 0 | 0 | 0 | 1 | Ref |
| 9139 | MK685668 | Unassigned | Unassigned | Unassigned | 386_0 | Clustered | 49 | VC_386_0 | 49 | 0.7941 | 1 | 0.7941 | 0 | 0 | 0 | 1 | Ref |
| 9140 | MK686068 | Unassigned | Unassigned | Unassigned | 373_0 | Clustered | 10 | VC_373_0 | 10 | 0.7471 | 1 | 0.7471 | 0 | 0 | 0 | 1 | Ref |
| 9141 | MK686069 | Unassigned | Unassigned | Unassigned | 373_0 | Clustered | 10 | VC_373_0 | 10 | 0.7471 | 1 | 0.7471 | 0 | 0 | 0 | 1 | Ref |
| 9142 | MK686071 | Unassigned | Unassigned | Unassigned | 975_0 | Clustered | 11 | VC_975_0 | 11 | 0.9687 | 1 | 0.9687 | 0 | 0 | 0 | 1 | Ref |
| 9143 | MK689364 | Unassigned | Unassigned | Unassigned | 442_2 | Clustered | 4 | VC_442_2 | 4 | 0.0844 | 0.99988129 | 0.0844 | 0 | 0 | 0 | 0.8333 | Ref |
| 9144 | MK693005 | Unassigned | Unassigned | Unassigned | 111_0 | Clustered | 91 | VC_111_0 | 91 | 0.9587 | 1 | 0.9587 | 0 | 0 | 0 | 1 | Ref |
| 9145 | MK693030 | Unassigned | Unassigned | Unassigned | 91_0 | Clustered | 22 | VC_91_0 | 22 | 0.5318 | 1 | 0.5318 | 0 | 0 | 0 | 1 | Ref |
| 9146 | MK714353 | Unassigned | Unassigned | Unassigned | 31_0 | Clustered | 9 | VC_31_0 | 9 | 0.3435 | 1 | 0.3435 | 0 | 0 | 0 | 0.6944 | Ref |
| 9147 | MK719701 | Unassigned | Unassigned | Unassigned | 1396_0 | Clustered | 110 | VC_1396_0 | 110 | 1 | 1 | 1 | 0 | 0 | 0 | 0.9746 | Ref |
| 9148 | MK719702 | Unassigned | Unassigned | Unassigned | 1396_0 | Clustered | 110 | VC_1396_0 | 110 | 1 | 1 | 1 | 0 | 0 | 0 | 0.9746 | Ref |
| 9149 | MK719703 | Unassigned | Unassigned | Unassigned | 1396_0 | Clustered | 110 | VC_1396_0 | 110 | 1 | 1 | 1 | 0 | 0 | 0 | 0.9746 | Ref |
| 9150 | MK719704 | Unassigned | Unassigned | Unassigned | 1396_0 | Clustered | 110 | VC_1396_0 | 110 | 1 | 1 | 1 | 0 | 0 | 0 | 0.9746 | Ref |
| 9151 | MK719705 | Unassigned | Unassigned | Unassigned | 1396_0 | Clustered | 110 | VC_1396_0 | 110 | 1 | 1 | 1 | 0 | 0 | 0 | 0.9746 | Ref |
| 9152 | MK719706 | Unassigned | Unassigned | Unassigned | 1396_0 | Clustered | 110 | VC_1396_0 | 110 | 1 | 1 | 1 | 0 | 0 | 0 | 0.9746 | Ref |
| 9153 | MK719707 | Unassigned | Unassigned | Unassigned | 1396_0 | Clustered | 110 | VC_1396_0 | 110 | 1 | 1 | 1 | 0 | 0 | 0 | 0.9746 | Ref |
| 9154 | MK719708 | Unassigned | Unassigned | Unassigned | 1396_0 | Clustered | 110 | VC_1396_0 | 110 | 1 | 1 | 1 | 0 | 0 | 0 | 0.9746 | Ref |
| 9155 | MK719709 | Unassigned | Unassigned | Unassigned | 1396_0 | Clustered | 110 | VC_1396_0 | 110 | 1 | 1 | 1 | 0 | 0 | 0 | 0.9746 | Ref |
| 9156 | MK719710 | Unassigned | Unassigned | Unassigned | 1396_0 | Clustered | 110 | VC_1396_0 | 110 | 1 | 1 | 1 | 0 | 0 | 0 | 0.9746 | Ref |
| 9157 | MK719711 | Unassigned | Unassigned | Unassigned | 1396_0 | Clustered | 110 | VC_1396_0 | 110 | 1 | 1 | 1 | 0 | 0 | 0 | 0.9746 | Ref |
| 9158 | MK719712 | Unassigned | Unassigned | Unassigned | 1396_0 | Clustered | 110 | VC_1396_0 | 110 | 1 | 1 | 1 | 0 | 0 | 0 | 0.9746 | Ref |
| 9159 | MK719713 | Unassigned | Unassigned | Unassigned | 1396_0 | Clustered | 110 | VC_1396_0 | 110 | 1 | 1 | 1 | 0 | 0 | 0 | 0.9746 | Ref |
| 9160 | MK719714 | Unassigned | Unassigned | Unassigned | 1396_0 | Clustered | 110 | VC_1396_0 | 110 | 1 | 1 | 1 | 0 | 0 | 0 | 0.9746 | Ref |
| 9161 | MK719715 | Unassigned | Unassigned | Unassigned | 1396_0 | Clustered | 110 | VC_1396_0 | 110 | 1 | 1 | 1 | 0 | 0 | 0 | 0.9746 | Ref |
| 9162 | MK719716 | Unassigned | Unassigned | Unassigned | 1396_0 | Clustered | 110 | VC_1396_0 | 110 | 1 | 1 | 1 | 0 | 0 | 0 | 0.9746 | Ref |
| 9163 | MK719717 | Unassigned | Unassigned | Unassigned | 1396_0 | Clustered | 110 | VC_1396_0 | 110 | 1 | 1 | 1 | 0 | 0 | 0 | 0.9746 | Ref |
| 9164 | MK719718 | Unassigned | Unassigned | Unassigned | 1396_0 | Clustered | 110 | VC_1396_0 | 110 | 1 | 1 | 1 | 0 | 0 | 0 | 0.9746 | Ref |
| 9165 | MK719719 | Unassigned | Unassigned | Unassigned | 1396_0 | Clustered | 110 | VC_1396_0 | 110 | 1 | 1 | 1 | 0 | 0 | 0 | 0.9746 | Ref |
| 9166 | MK719720 | Unassigned | Unassigned | Unassigned | 1396_0 | Clustered | 110 | VC_1396_0 | 110 | 1 | 1 | 1 | 0 | 0 | 0 | 0.9746 | Ref |
| 9167 | MK719721 | Unassigned | Unassigned | Unassigned | 1396_0 | Clustered | 110 | VC_1396_0 | 110 | 1 | 1 | 1 | 0 | 0 | 0 | 0.9746 | Ref |
| 9168 | MK719722 | Unassigned | Unassigned | Unassigned | 1396_0 | Clustered | 110 | VC_1396_0 | 110 | 1 | 1 | 1 | 0 | 0 | 0 | 0.9746 | Ref |
| 9169 | MK719723 | Unassigned | Unassigned | Unassigned | 1396_0 | Clustered | 110 | VC_1396_0 | 110 | 1 | 1 | 1 | 0 | 0 | 0 | 0.9746 | Ref |
| 9170 | MK719724 | Unassigned | Unassigned | Unassigned | 1396_0 | Clustered | 110 | VC_1396_0 | 110 | 1 | 1 | 1 | 0 | 0 | 0 | 0.9746 | Ref |
| 9171 | MK719725 | Unassigned | Unassigned | Unassigned | 1396_0 | Clustered | 110 | VC_1396_0 | 110 | 1 | 1 | 1 | 0 | 0 | 0 | 0.9746 | Ref |
| 9172 | MK719726 | Unassigned | Unassigned | Unassigned | 1396_0 | Clustered | 110 | VC_1396_0 | 110 | 1 | 1 | 1 | 0 | 0 | 0 | 0.9746 | Ref |
| 9173 | MK719727 | Unassigned | Unassigned | Unassigned | 1396_0 | Clustered | 110 | VC_1396_0 | 110 | 1 | 1 | 1 | 0 | 0 | 0 | 0.9746 | Ref |
| 9174 | MK719728 | Unassigned | Unassigned | Unassigned | 1396_0 | Clustered | 110 | VC_1396_0 | 110 | 1 | 1 | 1 | 0 | 0 | 0 | 0.9746 | Ref |
| 9175 | MK719729 | Unassigned | Unassigned | Unassigned | 1396_0 | Clustered | 110 | VC_1396_0 | 110 | 1 | 1 | 1 | 0 | 0 | 0 | 0.9746 | Ref |
| 9176 | MK719730 | Unassigned | Unassigned | Unassigned | 1396_0 | Clustered | 110 | VC_1396_0 | 110 | 1 | 1 | 1 | 0 | 0 | 0 | 0.9746 | Ref |
| 9177 | MK719731 | Unassigned | Unassigned | Unassigned | 1396_0 | Clustered | 110 | VC_1396_0 | 110 | 1 | 1 | 1 | 0 | 0 | 0 | 0.9746 | Ref |
| 9178 | MK719732 | Unassigned | Unassigned | Unassigned | 1396_0 | Clustered | 110 | VC_1396_0 | 110 | 1 | 1 | 1 | 0 | 0 | 0 | 0.9746 | Ref |
| 9179 | MK719733 | Unassigned | Unassigned | Unassigned | 1396_0 | Clustered | 110 | VC_1396_0 | 110 | 1 | 1 | 1 | 0 | 0 | 0 | 0.9746 | Ref |
| 9180 | MK719734 | Unassigned | Unassigned | Unassigned | 1396_0 | Clustered | 110 | VC_1396_0 | 110 | 1 | 1 | 1 | 0 | 0 | 0 | 0.9746 | Ref |
| 9181 | MK719735 | Unassigned | Unassigned | Unassigned | 1396_0 | Clustered | 110 | VC_1396_0 | 110 | 1 | 1 | 1 | 0 | 0 | 0 | 0.9746 | Ref |
| 9182 | MK719736 | Unassigned | Unassigned | Unassigned | 1396_0 | Clustered | 110 | VC_1396_0 | 110 | 1 | 1 | 1 | 0 | 0 | 0 | 0.9746 | Ref |
| 9183 | MK719737 | Unassigned | Unassigned | Unassigned | 1396_0 | Clustered | 110 | VC_1396_0 | 110 | 1 | 1 | 1 | 0 | 0 | 0 | 0.9746 | Ref |
| 9184 | MK719738 | Unassigned | Unassigned | Unassigned | 1396_0 | Clustered | 110 | VC_1396_0 | 110 | 1 | 1 | 1 | 0 | 0 | 0 | 0.9746 | Ref |
| 9185 | MK719739 | Unassigned | Unassigned | Unassigned | 1396_0 | Clustered | 110 | VC_1396_0 | 110 | 1 | 1 | 1 | 0 | 0 | 0 | 0.9746 | Ref |
| 9186 | MK719740 | Unassigned | Unassigned | Unassigned | 1396_0 | Clustered | 110 | VC_1396_0 | 110 | 1 | 1 | 1 | 0 | 0 | 0 | 0.9746 | Ref |
| 9187 | MK719741 | Unassigned | Unassigned | Unassigned | 1396_0 | Clustered | 110 | VC_1396_0 | 110 | 1 | 1 | 1 | 0 | 0 | 0 | 0.9746 | Ref |
| 9188 | MK719742 | Unassigned | Unassigned | Unassigned | 1396_0 | Clustered | 110 | VC_1396_0 | 110 | 1 | 1 | 1 | 0 | 0 | 0 | 0.9746 | Ref |
| 9189 | MK719743 | Unassigned | Unassigned | Unassigned | 1396_0 | Clustered | 110 | VC_1396_0 | 110 | 1 | 1 | 1 | 0 | 0 | 0 | 0.9746 | Ref |
| 9190 | MK719744 | Unassigned | Unassigned | Unassigned | 1396_0 | Clustered | 110 | VC_1396_0 | 110 | 1 | 1 | 1 | 0 | 0 | 0 | 0.9746 | Ref |
| 9191 | MK719745 | Unassigned | Unassigned | Unassigned | 1396_0 | Clustered | 110 | VC_1396_0 | 110 | 1 | 1 | 1 | 0 | 0 | 0 | 0.9746 | Ref |
| 9192 | MK719746 | Unassigned | Unassigned | Unassigned | 1396_0 | Clustered | 110 | VC_1396_0 | 110 | 1 | 1 | 1 | 0 | 0 | 0 | 0.9746 | Ref |
| 9193 | MK719747 | Unassigned | Unassigned | Unassigned | 1396_0 | Clustered | 110 | VC_1396_0 | 110 | 1 | 1 | 1 | 0 | 0 | 0 | 0.9746 | Ref |
| 9194 | MK719748 | Unassigned | Unassigned | Unassigned | 1396_0 | Clustered | 110 | VC_1396_0 | 110 | 1 | 1 | 1 | 0 | 0 | 0 | 0.9746 | Ref |
| 9195 | MK719749 | Unassigned | Unassigned | Unassigned | 1396_0 | Clustered | 110 | VC_1396_0 | 110 | 1 | 1 | 1 | 0 | 0 | 0 | 0.9746 | Ref |
| 9196 | MK719750 | Unassigned | Unassigned | Unassigned | 1396_0 | Clustered | 110 | VC_1396_0 | 110 | 1 | 1 | 1 | 0 | 0 | 0 | 0.9746 | Ref |
| 9197 | MK719751 | Unassigned | Unassigned | Unassigned | 1396_0 | Clustered | 110 | VC_1396_0 | 110 | 1 | 1 | 1 | 0 | 0 | 0 | 0.9746 | Ref |
| 9198 | MK719752 | Unassigned | Unassigned | Unassigned | 1396_0 | Clustered | 110 | VC_1396_0 | 110 | 1 | 1 | 1 | 0 | 0 | 0 | 0.9746 | Ref |
| 9199 | MK719753 | Unassigned | Unassigned | Unassigned | 1396_0 | Clustered | 110 | VC_1396_0 | 110 | 1 | 1 | 1 | 0 | 0 | 0 | 0.9746 | Ref |
| 9200 | MK719754 | Unassigned | Unassigned | Unassigned | 1396_0 | Clustered | 110 | VC_1396_0 | 110 | 1 | 1 | 1 | 0 | 0 | 0 | 0.9746 | Ref |
| 9201 | MK721183 | Unassigned | Unassigned | Unassigned | 967_0 | Clustered | 16 | VC_967_0 | 16 | 0.8971 | 1 | 0.8971 | 0 | 0 | 0 | 1 | Ref |
| 9202 | MK721184 | Unassigned | Unassigned | Unassigned | 967_0 | Clustered | 16 | VC_967_0 | 16 | 0.8971 | 1 | 0.8971 | 0 | 0 | 0 | 1 | Ref |
| 9203 | MK721185 | Unassigned | Unassigned | Unassigned | 967_0 | Clustered | 16 | VC_967_0 | 16 | 0.8971 | 1 | 0.8971 | 0 | 0 | 0 | 1 | Ref |
| 9204 | MK721186 | Unassigned | Unassigned | Unassigned | 695_0 | Clustered | 48 | VC_695_0 | 48 | 0.8162 | 1 | 0.8162 | 0 | 0 | 0 | 1 | Ref |
| 9205 | MK721187 | Unassigned | Unassigned | Unassigned | 695_0 | Clustered | 48 | VC_695_0 | 48 | 0.8162 | 1 | 0.8162 | 0 | 0 | 0 | 1 | Ref |
| 9206 | MK721188 | Unassigned | Unassigned | Unassigned | 967_0 | Clustered | 16 | VC_967_0 | 16 | 0.8971 | 1 | 0.8971 | 0 | 0 | 0 | 1 | Ref |
| 9207 | MK721189 | Unassigned | Unassigned | Unassigned | 108_0 | Clustered | 33 | VC_108_0 | 33 | 0.949 | 1 | 0.949 | 0 | 0 | 0 | 1 | Ref |
| 9208 | MK721190 | Unassigned | Unassigned | Unassigned | 695_0 | Clustered | 48 | VC_695_0 | 48 | 0.8162 | 1 | 0.8162 | 0 | 0 | 0 | 1 | Ref |
| 9209 | MK721191 | Unassigned | Unassigned | Unassigned | 695_0 | Clustered | 48 | VC_695_0 | 48 | 0.8162 | 1 | 0.8162 | 0 | 0 | 0 | 1 | Ref |
| 9210 | MK721192 | Unassigned | Unassigned | Unassigned | 91_0 | Clustered | 22 | VC_91_0 | 22 | 0.5318 | 1 | 0.5318 | 0 | 0 | 0 | 1 | Ref |
| 9211 | MK721193 | Unassigned | Unassigned | Unassigned | 967_0 | Clustered | 16 | VC_967_0 | 16 | 0.8971 | 1 | 0.8971 | 0 | 0 | 0 | 1 | Ref |
| 9212 | MK721194 | Unassigned | Unassigned | Unassigned | 108_0 | Clustered | 33 | VC_108_0 | 33 | 0.949 | 1 | 0.949 | 0 | 0 | 0 | 1 | Ref |
| 9213 | MK721195 | Unassigned | Unassigned | Unassigned | 967_0 | Clustered | 16 | VC_967_0 | 16 | 0.8971 | 1 | 0.8971 | 0 | 0 | 0 | 1 | Ref |
| 9214 | MK721196 | Unassigned | Unassigned | Unassigned | 967_0 | Clustered | 16 | VC_967_0 | 16 | 0.8971 | 1 | 0.8971 | 0 | 0 | 0 | 1 | Ref |
| 9215 | MK721197 | Unassigned | Unassigned | Unassigned | 967_0 | Clustered | 16 | VC_967_0 | 16 | 0.8971 | 1 | 0.8971 | 0 | 0 | 0 | 1 | Ref |
| 9216 | MK721198 | Unassigned | Unassigned | Unassigned | 967_0 | Clustered | 16 | VC_967_0 | 16 | 0.8971 | 1 | 0.8971 | 0 | 0 | 0 | 1 | Ref |
| 9217 | MK721199 | Unassigned | Unassigned | Unassigned | 695_0 | Clustered | 48 | VC_695_0 | 48 | 0.8162 | 1 | 0.8162 | 0 | 0 | 0 | 1 | Ref |
| 9218 | MK721200 | Unassigned | Unassigned | Unassigned | 695_0 | Clustered | 48 | VC_695_0 | 48 | 0.8162 | 1 | 0.8162 | 0 | 0 | 0 | 1 | Ref |
| 9219 | MK728541 | Unassigned | Unassigned | Unassigned | 835_0 | Clustered | 67 | VC_835_0 | 67 | 0.9624 | 1 | 0.9624 | 0 | 0 | 0 | 1 | Ref |
| 9220 | MK728824 | Unassigned | Unassigned | Unassigned | 191_19 | Clustered | 12 | VC_191_19 | 12 | 0.0655 | 1 | 0.0655 | 0 | 0 | 0 | 0.8636 | Ref |
| 9221 | MK728825 | Unassigned | Unassigned | Unassigned | 191_19 | Clustered | 12 | VC_191_19 | 12 | 0.0655 | 1 | 0.0655 | 0 | 0 | 0 | 0.8636 | Ref |
| 9222 | MK733259 | Unassigned | Unassigned | Unassigned | 45_0 | Clustered | 12 | VC_45_0 | 12 | 0.35 | 1 | 0.35 | 0 | 0 | 0 | 1 | Ref |
| 9223 | MK733260 | Unassigned | Unassigned | Unassigned | 45_0 | Clustered | 12 | VC_45_0 | 12 | 0.35 | 1 | 0.35 | 0 | 0 | 0 | 1 | Ref |
| 9224 | MK733261 | Unassigned | Unassigned | Unassigned | 45_0 | Clustered | 12 | VC_45_0 | 12 | 0.35 | 1 | 0.35 | 0 | 0 | 0 | 1 | Ref |
| 9225 | MK733262 | Unassigned | Unassigned | Unassigned | 281_0 | Clustered | 3 | VC_281_0 | 3 | 0.3561 | 0.99828319 | 0.3555 | 0 | 0 | 0 | 1 | Ref |
| 9226 | MK733263 | Unassigned | Unassigned | Unassigned | 281_0 | Clustered | 3 | VC_281_0 | 3 | 0.3561 | 0.99828319 | 0.3555 | 0 | 0 | 0 | 1 | Ref |
| 9227 | MK733264 | Unassigned | Unassigned | Unassigned | 281_0 | Clustered | 3 | VC_281_0 | 3 | 0.3561 | 0.99828319 | 0.3555 | 0 | 0 | 0 | 1 | Ref |
| 9228 | MK737937 | Unassigned | Unassigned | Unassigned | 638_0 | Clustered | 45 | VC_638_0 | 45 | 0.7741 | 1 | 0.7741 | 0 | 0 | 0 | 1 | Ref |
| 9229 | MK737941 | Unassigned | Unassigned | Unassigned | 1343_0 | Clustered | 21 | VC_1343_0 | 21 | 0.9843 | 1 | 0.9843 | 0 | 0 | 0 | 1 | Ref |
| 9230 | MK737942 | Unassigned | Unassigned | Unassigned | 1162_0 | Clustered | 10 | VC_1162_0 | 10 | 0.5715 | 1 | 0.5715 | 0 | 0 | 0 | 1 | Ref |
| 9231 | MK737943 | Unassigned | Unassigned | Unassigned | 950_0 | Clustered | 38 | VC_950_0 | 38 | 0.9443 | 1 | 0.9443 | 0 | 0 | 0 | 1 | Ref |
| 9232 | MK737944 | Unassigned | Unassigned | Unassigned | 1348_0 | Clustered | 5 | VC_1348_0 | 5 | 0.5462 | 0.99999985 | 0.5462 | 0 | 0 | 0 | 1 | Ref |

| | | | | | | | | | | | | | | | | | |
|---|---|---|---|---|---|---|---|---|---|---|---|---|---|---|---|---|---|
| 9233 | MK756083 | Unassigned | Unassigned | Unassigned | 912_0 | Clustered | 76 | VC_912_0 | 76 | 0.979 | 1 | 0.979 | 0 | 0 | 0 | 1 | Ref |
| 9234 | MK756084 | Unassigned | Unassigned | Unassigned | 912_0 | Clustered | 76 | VC_912_0 | 76 | 0.979 | 1 | 0.979 | 0 | 0 | 0 | 1 | Ref |
| 9235 | MK756085 | Unassigned | Unassigned | Unassigned | 912_0 | Clustered | 76 | VC_912_0 | 76 | 0.979 | 1 | 0.979 | 0 | 0 | 0 | 1 | Ref |
| 9236 | MK756086 | Unassigned | Unassigned | Unassigned | 912_0 | Clustered | 76 | VC_912_0 | 76 | 0.979 | 1 | 0.979 | 0 | 0 | 0 | 1 | Ref |
| 9237 | MK756087 | Unassigned | Unassigned | Unassigned | 912_0 | Clustered | 76 | VC_912_0 | 76 | 0.979 | 1 | 0.979 | 0 | 0 | 0 | 1 | Ref |
| 9238 | MK756088 | Unassigned | Unassigned | Unassigned | 912_0 | Clustered | 76 | VC_912_0 | 76 | 0.979 | 1 | 0.979 | 0 | 0 | 0 | 1 | Ref |
| 9239 | MK756089 | Unassigned | Unassigned | Unassigned | 912_0 | Clustered | 76 | VC_912_0 | 76 | 0.979 | 1 | 0.979 | 0 | 0 | 0 | 1 | Ref |
| 9240 | MK756090 | Unassigned | Unassigned | Unassigned | 912_0 | Clustered | 76 | VC_912_0 | 76 | 0.979 | 1 | 0.979 | 0 | 0 | 0 | 1 | Ref |
| 9241 | MK756091 | Unassigned | Unassigned | Unassigned | 912_0 | Clustered | 76 | VC_912_0 | 76 | 0.979 | 1 | 0.979 | 0 | 0 | 0 | 1 | Ref |
| 9242 | MK756092 | Unassigned | Unassigned | Unassigned | 912_0 | Clustered | 76 | VC_912_0 | 76 | 0.979 | 1 | 0.979 | 0 | 0 | 0 | 1 | Ref |
| 9243 | MK756093 | Unassigned | Unassigned | Unassigned | 912_0 | Clustered | 76 | VC_912_0 | 76 | 0.979 | 1 | 0.979 | 0 | 0 | 0 | 1 | Ref |
| 9244 | MK756094 | Unassigned | Unassigned | Unassigned | 912_0 | Clustered | 76 | VC_912_0 | 76 | 0.979 | 1 | 0.979 | 0 | 0 | 0 | 1 | Ref |
| 9245 | MK756095 | Unassigned | Unassigned | Unassigned | 912_0 | Clustered | 76 | VC_912_0 | 76 | 0.979 | 1 | 0.979 | 0 | 0 | 0 | 1 | Ref |
| 9246 | MK756096 | Unassigned | Unassigned | Unassigned | 912_0 | Clustered | 76 | VC_912_0 | 76 | 0.979 | 1 | 0.979 | 0 | 0 | 0 | 1 | Ref |
| 9247 | MK757445 | Unassigned | Unassigned | Unassigned | 551_7 | Clustered | 28 | VC_551_7 | 28 | 0.1661 | 1 | 0.1661 | 0 | 0 | 0 | 0.8519 | Ref |
| 9248 | MK757446 | Unassigned | Unassigned | Unassigned | 568_0 | Clustered | 8 | VC_568_0 | 8 | 0.5738 | 1 | 0.5738 | 0 | 0 | 0 | 1 | Ref |
| 9249 | MK757447 | Unassigned | Unassigned | Unassigned | 1240_0 | Clustered | 82 | VC_1240_0 | 82 | 0.7945 | 1 | 0.7945 | 0 | 0 | 0 | 1 | Ref |
| 9250 | MK757448 | Unassigned | Unassigned | Unassigned | 1362_0 | Clustered | 6 | VC_1362_0 | 6 | 1 | 1 | 1 | 0 | 0 | 0 | 1 | Ref |
| 9251 | MK757449 | Unassigned | Unassigned | Unassigned | 1360_0 | Clustered | 31 | VC_1360_0 | 31 | 0.8837 | 1 | 0.8837 | 0 | 0 | 0 | 1 | Ref |
| 9252 | MK759854 | Unassigned | Unassigned | Unassigned | 461_0 | Clustered | 96 | VC_461_0 | 96 | 0.9307 | 1 | 0.9307 | 0 | 0 | 0 | 0.9575 | Ref |
| 9253 | MK759883 | Unassigned | Unassigned | Unassigned | 637_1 | Clustered | 27 | VC_637_1 | 27 | 0.2578 | 1 | 0.2578 | 0 | 0 | 0 | 0.9858 | Ref |
| 9254 | MK759884 | Unassigned | Unassigned | Unassigned | 328_0 | Clustered | 11 | VC_328_0 | 11 | 0.4707 | 1 | 0.4707 | 0 | 0 | 0 | 1 | Ref |
| 9255 | MK759918 | Unassigned | Unassigned | Unassigned | 731_0 | Clustered | 2 | VC_731_0 | 2 | 0.3697 | 0.94979793 | 0.3512 | 0 | 0 | 0 | 1 | Ref |
| 9256 | MK761195 | Unassigned | Unassigned | Unassigned | 637_0 | Clustered | 115 | VC_637_0 | 115 | 0.731 | 1 | 0.731 | 0 | 0 | 0 | 0.8764 | Ref |
| 9257 | MK761196 | Unassigned | Unassigned | Unassigned | 637_0 | Clustered | 115 | VC_637_0 | 115 | 0.731 | 1 | 0.731 | 0 | 0 | 0 | 0.8764 | Ref |
| 9258 | MK761197 | Unassigned | Unassigned | Unassigned | 637_0 | Clustered | 115 | VC_637_0 | 115 | 0.731 | 1 | 0.731 | 0 | 0 | 0 | 0.8764 | Ref |
| 9259 | MK761198 | Unassigned | Unassigned | Unassigned | 637_0 | Clustered | 115 | VC_637_0 | 115 | 0.731 | 1 | 0.731 | 0 | 0 | 0 | 0.8764 | Ref |
| 9260 | MK761199 | Unassigned | Unassigned | Unassigned | 637_0 | Clustered | 115 | VC_637_0 | 115 | 0.731 | 1 | 0.731 | 0 | 0 | 0 | 0.8764 | Ref |
| 9261 | MK764384 | Unassigned | Unassigned | Unassigned | 96_0 | Clustered | 30 | VC_96_0 | 30 | 0.718 | 1 | 0.718 | 0 | 0 | 0 | 1 | Ref |
| 9262 | MK764436 | Unassigned | Unassigned | Unassigned | 1040_0 | Clustered | 23 | VC_1040_0 | 23 | 0.9605 | 1 | 0.9605 | 0 | 0 | 0 | 1 | Ref |
| 9263 | MK764437 | Unassigned | Unassigned | Unassigned | 1023_0 | Clustered | 8 | VC_1023_0 | 8 | 0.8574 | 1 | 0.8574 | 0 | 0 | 0 | 1 | Ref |
| 9264 | MK764438 | Unassigned | Unassigned | Unassigned | 1040_0 | Clustered | 23 | VC_1040_0 | 23 | 0.9605 | 1 | 0.9605 | 0 | 0 | 0 | 1 | Ref |
| 9265 | MK764439 | Unassigned | Unassigned | Unassigned | 1040_0 | Clustered | 23 | VC_1040_0 | 23 | 0.9605 | 1 | 0.9605 | 0 | 0 | 0 | 1 | Ref |
| 9266 | MK764440 | Unassigned | Unassigned | Unassigned | 1023_0 | Clustered | 8 | VC_1023_0 | 8 | 0.8574 | 1 | 0.8574 | 0 | 0 | 0 | 1 | Ref |
| 9267 | MK764441 | Unassigned | Unassigned | Unassigned | 1157_0 | Clustered | 24 | VC_1157_0 | 24 | 1 | 1 | 1 | 0 | 0 | 0 | 1 | Ref |
| 9268 | MK764442 | Unassigned | Unassigned | Unassigned | 1021_0 | Clustered | 10 | VC_1021_0 | 10 | 0.8876 | 1 | 0.8876 | 0 | 0 | 0 | 1 | Ref |
| 9269 | MK764443 | Unassigned | Unassigned | Unassigned | 1040_0 | Clustered | 23 | VC_1040_0 | 23 | 0.9605 | 1 | 0.9605 | 0 | 0 | 0 | 1 | Ref |
| 9270 | MK764444 | Unassigned | Unassigned | Unassigned | 1040_0 | Clustered | 23 | VC_1040_0 | 23 | 0.9605 | 1 | 0.9605 | 0 | 0 | 0 | 1 | Ref |
| 9271 | MK764445 | Unassigned | Unassigned | Unassigned | 1021_0 | Clustered | 10 | VC_1021_0 | 10 | 0.8876 | 1 | 0.8876 | 0 | 0 | 0 | 1 | Ref |
| 9272 | MK764446 | Unassigned | Unassigned | Unassigned | 1040_0 | Clustered | 23 | VC_1040_0 | 23 | 0.9605 | 1 | 0.9605 | 0 | 0 | 0 | 1 | Ref |
| 9273 | MK764447 | Unassigned | Unassigned | Unassigned | 1040_0 | Clustered | 23 | VC_1040_0 | 23 | 0.9605 | 1 | 0.9605 | 0 | 0 | 0 | 1 | Ref |
| 9274 | MK764448 | Unassigned | Unassigned | Unassigned | 1021_0 | Clustered | 10 | VC_1021_0 | 10 | 0.8876 | 1 | 0.8876 | 0 | 0 | 0 | 1 | Ref |
| 9275 | MK764449 | Unassigned | Unassigned | Unassigned | 1157_0 | Clustered | 24 | VC_1157_0 | 24 | 1 | 1 | 1 | 0 | 0 | 0 | 1 | Ref |
| 9276 | MK764450 | Unassigned | Unassigned | Unassigned | 1023_0 | Clustered | 8 | VC_1023_0 | 8 | 0.8574 | 1 | 0.8574 | 0 | 0 | 0 | 1 | Ref |
| 9277 | MK765551 | Unassigned | Unassigned | Unassigned | 1398_0 | Clustered | 2 | VC_1398_0 | 2 | 1 | 1 | 1 | 0 | 0 | 0 | 1 | Ref |
| 9278 | MK765552 | Unassigned | Unassigned | Unassigned | 1283_0 | Clustered | 26 | VC_1283_0 | 26 | 0.5446 | 1 | 0.5446 | 0 | 0 | 0 | 1 | Ref |
| 9279 | MK765553 | Unassigned | Unassigned | Unassigned | 1283_0 | Clustered | 26 | VC_1283_0 | 26 | 0.5446 | 1 | 0.5446 | 0 | 0 | 0 | 1 | Ref |
| 9280 | MK765555 | Unassigned | Unassigned | Unassigned | 1325_0 | Clustered | 7 | VC_1325_0 | 7 | 0.9166 | 0.99999981 | 0.9166 | 0 | 0 | 0 | 1 | Ref |
| 9281 | MK765556 | Unassigned | Unassigned | Unassigned | 1283_0 | Clustered | 26 | VC_1283_0 | 26 | 0.5446 | 1 | 0.5446 | 0 | 0 | 0 | 1 | Ref |
| 9282 | MK765557 | Unassigned | Unassigned | Unassigned | 1211_0 | Clustered | 25 | VC_1211_0 | 25 | 1 | 1 | 1 | 0 | 0 | 0 | 1 | Ref |
| 9283 | MK765558 | Unassigned | Unassigned | Unassigned | 1399_0 | Clustered | 9 | VC_1399_0 | 9 | 1 | 0 | 0 | 0 | 0 | 0 | 1 | Ref |
| 9284 | MK765559 | Unassigned | Unassigned | Unassigned | 1119_0 | Clustered | 20 | VC_1119_0 | 20 | 0.6751 | 0.99997892 | 0.6751 | 0 | 0 | 0 | 1 | Ref |
| 9285 | MK765560 | Unassigned | Unassigned | Unassigned | 1283_0 | Clustered | 26 | VC_1283_0 | 26 | 0.5446 | 1 | 0.5446 | 0 | 0 | 0 | 1 | Ref |
| 9286 | MK765561 | Unassigned | Unassigned | Unassigned | 1243_0 | Clustered | 52 | VC_1243_0 | 52 | 0.9126 | 0.92708311 | 0.8461 | 0 | 0 | 0 | 1 | Ref |
| 9287 | MK765562 | Unassigned | Unassigned | Unassigned | 1283_0 | Clustered | 26 | VC_1283_0 | 26 | 0.5446 | 1 | 0.5446 | 0 | 0 | 0 | 1 | Ref |
| 9288 | MK765563 | Unassigned | Unassigned | Unassigned | 1243_0 | Clustered | 52 | VC_1243_0 | 52 | 0.9126 | 0.92708311 | 0.8461 | 0 | 0 | 0 | 1 | Ref |
| 9289 | MK765564 | Unassigned | Unassigned | Unassigned | 1243_0 | Clustered | 52 | VC_1243_0 | 52 | 0.9126 | 0.92708311 | 0.8461 | 0 | 0 | 0 | 1 | Ref |
| 9290 | MK765565 | Unassigned | Unassigned | Unassigned | 1283_0 | Clustered | 26 | VC_1283_0 | 26 | 0.5446 | 1 | 0.5446 | 0 | 0 | 0 | 1 | Ref |
| 9291 | MK765567 | Unassigned | Unassigned | Unassigned | 1325_0 | Clustered | 7 | VC_1325_0 | 7 | 0.9166 | 0.99999981 | 0.9166 | 0 | 0 | 0 | 1 | Ref |
| 9292 | MK765568 | Unassigned | Unassigned | Unassigned | 1243_0 | Clustered | 52 | VC_1243_0 | 52 | 0.9126 | 0.92708311 | 0.8461 | 0 | 0 | 0 | 1 | Ref |
| 9293 | MK765569 | Unassigned | Unassigned | Unassigned | 1243_0 | Clustered | 52 | VC_1243_0 | 52 | 0.9126 | 0.92708311 | 0.8461 | 0 | 0 | 0 | 1 | Ref |
| 9294 | MK765571 | Unassigned | Unassigned | Unassigned | 1285_0 | Clustered | 25 | VC_1285_0 | 25 | 1 | 1 | 1 | 0 | 0 | 0 | 1 | Ref |
| 9295 | MK765572 | Unassigned | Unassigned | Unassigned | 1280_0 | Clustered | 2 | VC_1280_0 | 2 | 0.0695 | 0.94944661 | 0.066 | 0 | 0 | 0 | 1 | Ref |
| 9296 | MK765573 | Unassigned | Unassigned | Unassigned | 1400_0 | Clustered | 2 | VC_1400_0 | 2 | 1 | 1 | 1 | 0 | 0 | 0 | 1 | Ref |
| 9297 | MK765574 | Unassigned | Unassigned | Unassigned | 1285_0 | Clustered | 25 | VC_1285_0 | 25 | 1 | 1 | 1 | 0 | 0 | 0 | 1 | Ref |
| 9298 | MK765575 | Unassigned | Unassigned | Unassigned | 1401_0 | Clustered | 2 | VC_1401_0 | 2 | 1 | 1 | 1 | 0 | 0 | 0 | 1 | Ref |
| 9299 | MK765576 | Unassigned | Unassigned | Unassigned | 1285_0 | Clustered | 25 | VC_1285_0 | 25 | 1 | 1 | 1 | 0 | 0 | 0 | 1 | Ref |
| 9300 | MK765577 | Unassigned | Unassigned | Unassigned | 1285_0 | Clustered | 25 | VC_1285_0 | 25 | 1 | 1 | 1 | 0 | 0 | 0 | 1 | Ref |
| 9301 | MK765578 | Unassigned | Unassigned | Unassigned | 1401_0 | Clustered | 2 | VC_1401_0 | 2 | 1 | 1 | 1 | 0 | 0 | 0 | 1 | Ref |
| 9302 | MK765579 | Unassigned | Unassigned | Unassigned | 1062_0 | Clustered | 45 | VC_1062_0 | 45 | 0.9734 | 0.50949847 | 0.4959 | 0 | 0 | 0 | 1 | Ref |
| 9303 | MK765580 | Unassigned | Unassigned | Unassigned | 1285_0 | Clustered | 25 | VC_1285_0 | 25 | 1 | 1 | 1 | 0 | 0 | 0 | 1 | Ref |
| 9304 | MK765581 | Unassigned | Unassigned | Unassigned | 1399_0 | Clustered | 9 | VC_1399_0 | 9 | 1 | 0 | 0 | 0 | 0 | 0 | 1 | Ref |
| 9305 | MK765582 | Unassigned | Unassigned | Unassigned | 1062_0 | Clustered | 45 | VC_1062_0 | 45 | 0.9734 | 0.50949847 | 0.4959 | 0 | 0 | 0 | 1 | Ref |
| 9306 | MK765583 | Unassigned | Unassigned | Unassigned | 1308_0 | Clustered | 12 | VC_1308_0 | 12 | 0.8184 | 0.9810731 | 0.8029 | 0 | 0 | 0 | 1 | Ref |
| 9307 | MK765587 | Unassigned | Unassigned | Unassigned | 1402_0 | Clustered | 14 | VC_1402_0 | 14 | 0.1673 | 0.51390354 | 0.086 | 0 | 0 | 0 | 1 | Ref |
| 9308 | MK765588 | Unassigned | Unassigned | Unassigned | 1402_0 | Clustered | 14 | VC_1402_0 | 14 | 0.1673 | 0.51390354 | 0.086 | 0 | 0 | 0 | 1 | Ref |
| 9309 | MK765589 | Unassigned | Unassigned | Unassigned | 1403_0 | Clustered | 28 | VC_1403_0 | 28 | 0.753 | 1 | 0.753 | 0 | 0 | 0 | 1 | Ref |
| 9310 | MK765590 | Unassigned | Unassigned | Unassigned | 1277_0 | Clustered | 14 | VC_1277_0 | 14 | 0.8218 | 1 | 0.8218 | 0 | 0 | 0 | 1 | Ref |
| 9311 | MK765591 | Unassigned | Unassigned | Unassigned | 1283_0 | Clustered | 26 | VC_1283_0 | 26 | 0.5446 | 1 | 0.5446 | 0 | 0 | 0 | 1 | Ref |
| 9312 | MK765592 | Unassigned | Unassigned | Unassigned | 1283_0 | Clustered | 26 | VC_1283_0 | 26 | 0.5446 | 1 | 0.5446 | 0 | 0 | 0 | 1 | Ref |
| 9313 | MK765593 | Unassigned | Unassigned | Unassigned | 1283_0 | Clustered | 26 | VC_1283_0 | 26 | 0.5446 | 1 | 0.5446 | 0 | 0 | 0 | 1 | Ref |
| 9314 | MK765594 | Unassigned | Unassigned | Unassigned | 1243_0 | Clustered | 52 | VC_1243_0 | 52 | 0.9126 | 0.92708311 | 0.8461 | 0 | 0 | 0 | 1 | Ref |
| 9315 | MK765596 | Unassigned | Unassigned | Unassigned | 1283_0 | Clustered | 26 | VC_1283_0 | 26 | 0.5446 | 1 | 0.5446 | 0 | 0 | 0 | 1 | Ref |
| 9316 | MK765598 | Unassigned | Unassigned | Unassigned | 1402_0 | Clustered | 14 | VC_1402_0 | 14 | 0.1673 | 0.51390354 | 0.086 | 0 | 0 | 0 | 1 | Ref |
| 9317 | MK765599 | Unassigned | Unassigned | Unassigned | 1402_0 | Clustered | 14 | VC_1402_0 | 14 | 0.1673 | 0.51390354 | 0.086 | 0 | 0 | 0 | 1 | Ref |
| 9318 | MK765600 | Unassigned | Unassigned | Unassigned | 1325_0 | Clustered | 7 | VC_1325_0 | 7 | 0.9166 | 0.99999981 | 0.9166 | 0 | 0 | 0 | 1 | Ref |
| 9319 | MK765601 | Unassigned | Unassigned | Unassigned | 1303_0 | Clustered | 6 | VC_1303_0 | 6 | 1 | 1 | 1 | 0 | 0 | 0 | 1 | Ref |
| 9320 | MK765602 | Unassigned | Unassigned | Unassigned | 1140_0 | Clustered | 4 | VC_1140_0 | 4 | 0.1325 | 0.66207199 | 0.0877 | 0 | 0 | 0 | 1 | Ref |
| 9321 | MK765603 | Unassigned | Unassigned | Unassigned | 1119_0 | Clustered | 20 | VC_1119_0 | 20 | 0.6751 | 0.99997892 | 0.6751 | 0 | 0 | 0 | 1 | Ref |
| 9322 | MK765604 | Unassigned | Unassigned | Unassigned | 1399_0 | Clustered | 9 | VC_1399_0 | 9 | 1 | 0 | 0 | 0 | 0 | 0 | 1 | Ref |
| 9323 | MK765605 | Unassigned | Unassigned | Unassigned | 1406_0 | Clustered | 2 | VC_1406_0 | 2 | 1 | 1 | 1 | 0 | 0 | 0 | 1 | Ref |
| 9324 | MK765606 | Unassigned | Unassigned | Unassigned | 1283_0 | Clustered | 26 | VC_1283_0 | 26 | 0.5446 | 1 | 0.5446 | 0 | 0 | 0 | 1 | Ref |
| 9325 | MK765607 | Unassigned | Unassigned | Unassigned | 1283_0 | Clustered | 26 | VC_1283_0 | 26 | 0.5446 | 1 | 0.5446 | 0 | 0 | 0 | 1 | Ref |
| 9326 | MK765609 | Unassigned | Unassigned | Unassigned | 1283_0 | Clustered | 26 | VC_1283_0 | 26 | 0.5446 | 1 | 0.5446 | 0 | 0 | 0 | 1 | Ref |
| 9327 | MK765610 | Unassigned | Unassigned | Unassigned | 1325_0 | Clustered | 7 | VC_1325_0 | 7 | 0.9166 | 0.99999981 | 0.9166 | 0 | 0 | 0 | 1 | Ref |
| 9328 | MK765611 | Unassigned | Unassigned | Unassigned | 1283_0 | Clustered | 26 | VC_1283_0 | 26 | 0.5446 | 1 | 0.5446 | 0 | 0 | 0 | 1 | Ref |
| 9329 | MK765612 | Unassigned | Unassigned | Unassigned | 1283_0 | Clustered | 26 | VC_1283_0 | 26 | 0.5446 | 1 | 0.5446 | 0 | 0 | 0 | 1 | Ref |
| 9330 | MK765613 | Unassigned | Unassigned | Unassigned | 1283_0 | Clustered | 26 | VC_1283_0 | 26 | 0.5446 | 1 | 0.5446 | 0 | 0 | 0 | 1 | Ref |
| 9331 | MK765615 | Unassigned | Unassigned | Unassigned | 1283_0 | Clustered | 26 | VC_1283_0 | 26 | 0.5446 | 1 | 0.5446 | 0 | 0 | 0 | 1 | Ref |
| 9332 | MK765617 | Unassigned | Unassigned | Unassigned | 1325_0 | Clustered | 7 | VC_1325_0 | 7 | 0.9166 | 0.99999981 | 0.9166 | 0 | 0 | 0 | 1 | Ref |
| 9333 | MK765620 | Unassigned | Unassigned | Unassigned | 1104_0 | Clustered | 154 | VC_1104_0 | 154 | 0.8484 | 1 | 0.8484 | 0 | 0 | 0 | 1 | Ref |
| 9334 | MK765621 | Unassigned | Unassigned | Unassigned | 1119_0 | Clustered | 20 | VC_1119_0 | 20 | 0.6751 | 0.99997892 | 0.6751 | 0 | 0 | 0 | 1 | Ref |
| 9335 | MK765622 | Unassigned | Unassigned | Unassigned | 1399_0 | Clustered | 9 | VC_1399_0 | 9 | 1 | 0 | 0 | 0 | 0 | 0 | 1 | Ref |
| 9336 | MK765623 | Unassigned | Unassigned | Unassigned | 1402_0 | Clustered | 14 | VC_1402_0 | 14 | 0.1673 | 0.51390354 | 0.086 | 0 | 0 | 0 | 1 | Ref |
| 9337 | MK765625 | Unassigned | Unassigned | Unassigned | 1402_0 | Clustered | 14 | VC_1402_0 | 14 | 0.1673 | 0.51390354 | 0.086 | 0 | 0 | 0 | 1 | Ref |
| 9338 | MK765626 | Unassigned | Unassigned | Unassigned | 1404_0 | Clustered/Singleton | 1 | VC_1404_0 | 1 | 0 | 1 | 0 | 0 | 0 | 0 | 1 | Ref |
| 9339 | MK765628 | Unassigned | Unassigned | Unassigned | 1402_0 | Clustered | 14 | VC_1402_0 | 14 | 0.1673 | 0.51390354 | 0.086 | 0 | 0 | 0 | 1 | Ref |
| 9340 | MK765630 | Unassigned | Unassigned | Unassigned | 1402_0 | Clustered | 14 | VC_1402_0 | 14 | 0.1673 | 0.51390354 | 0.086 | 0 | 0 | 0 | 1 | Ref |
| 9341 | MK765631 | Unassigned | Unassigned | Unassigned | 1402_0 | Clustered | 14 | VC_1402_0 | 14 | 0.1673 | 0.51390354 | 0.086 | 0 | 0 | 0 | 1 | Ref |
| 9342 | MK765633 | Unassigned | Unassigned | Unassigned | 1283_0 | Clustered | 26 | VC_1283_0 | 26 | 0.5446 | 1 | 0.5446 | 0 | 0 | 0 | 1 | Ref |
| 9343 | MK765634 | Unassigned | Unassigned | Unassigned | 1402_0 | Clustered | 14 | VC_1402_0 | 14 | 0.1673 | 0.51390354 | 0.086 | 0 | 0 | 0 | 1 | Ref |
| 9344 | MK765638 | Unassigned | Unassigned | Unassigned | 1243_0 | Clustered | 52 | VC_1243_0 | 52 | 0.9126 | 0.92708311 | 0.8461 | 0 | 0 | 0 | 1 | Ref |
| 9345 | MK765639 | Unassigned | Unassigned | Unassigned | 1062_0 | Clustered | 45 | VC_1062_0 | 45 | 0.9734 | 0.50949847 | 0.4959 | 0 | 0 | 0 | 1 | Ref |
| 9346 | MK765640 | Unassigned | Unassigned | Unassigned | 1399_0 | Clustered | 9 | VC_1399_0 | 9 | 1 | 0 | 0 | 0 | 0 | 0 | 1 | Ref |
| 9347 | MK765641 | Unassigned | Unassigned | Unassigned | 1406_0 | Clustered | 2 | VC_1406_0 | 2 | 1 | 1 | 1 | 0 | 0 | 0 | 1 | Ref |
| 9348 | MK765642 | Unassigned | Unassigned | Unassigned | 1062_0 | Clustered | 45 | VC_1062_0 | 45 | 0.9734 | 0.50949847 | 0.4959 | 0 | 0 | 0 | 1 | Ref |
| 9349 | MK765643 | Unassigned | Unassigned | Unassigned | 1062_0 | Clustered | 45 | VC_1062_0 | 45 | 0.9734 | 0.50949847 | 0.4959 | 0 | 0 | 0 | 1 | Ref |
| 9350 | MK765644 | Unassigned | Unassigned | Unassigned | 1062_0 | Clustered | 45 | VC_1062_0 | 45 | 0.9734 | 0.50949847 | 0.4959 | 0 | 0 | 0 | 1 | Ref |
| 9351 | MK765645 | Unassigned | Unassigned | Unassigned | 1077_0 | Clustered | 48 | VC_1077_0 | 48 | 0.8525 | 1 | 0.8525 | 0 | 0 | 0 | 1 | Ref |
| 9352 | MK765646 | Unassigned | Unassigned | Unassigned | 1062_0 | Clustered | 45 | VC_1062_0 | 45 | 0.9734 | 0.50949847 | 0.4959 | 0 | 0 | 0 | 1 | Ref |
| 9353 | MK765648 | Unassigned | Unassigned | Unassigned | 1283_0 | Clustered | 26 | VC_1283_0 | 26 | 0.5446 | 1 | 0.5446 | 0 | 0 | 0 | 1 | Ref |
| 9354 | MK765649 | Unassigned | Unassigned | Unassigned | 1282_0 | Clustered | 3 | VC_1282_0 | 3 | 1 | 1 | 1 | 0 | 0 | 0 | 1 | Ref |
| 9355 | MK765650 | Unassigned | Unassigned | Unassigned | 1398_0 | Clustered | 2 | VC_1398_0 | 2 | 1 | 1 | 1 | 0 | 0 | 0 | 1 | Ref |
| 9356 | MK765651 | Unassigned | Unassigned | Unassigned | 1283_0 | Clustered | 26 | VC_1283_0 | 26 | 0.5446 | 1 | 0.5446 | 0 | 0 | 0 | 1 | Ref |
| 9357 | MK765653 | Unassigned | Unassigned | Unassigned | 1331_0 | Clustered | 4 | VC_1331_0 | 4 | 0.7343 | 0.99899459 | 0.7335 | 0 | 0 | 0 | 1 | Ref |
| 9358 | MK765654 | Unassigned | Unassigned | Unassigned | 1399_0 | Clustered | 9 | VC_1399_0 | 9 | 1 | 0 | 0 | 0 | 0 | 0 | 1 | Ref |
| 9359 | MK765655 | Unassigned | Unassigned | Unassigned | 1211_0 | Clustered | 25 | VC_1211_0 | 25 | 1 | 1 | 1 | 0 | 0 | 0 | 1 | Ref |
| 9360 | MK765656 | Unassigned | Unassigned | Unassigned | 1399_0 | Clustered | 9 | VC_1399_0 | 9 | 1 | 0 | 0 | 0 | 0 | 0 | 1 | Ref |
| 9361 | MK765657 | Unassigned | Unassigned | Unassigned | 1283_0 | Clustered | 26 | VC_1283_0 | 26 | 0.5446 | 1 | 0.5446 | 0 | 0 | 0 | 1 | Ref |
| 9362 | MK765658 | Unassigned | Unassigned | Unassigned | 1283_0 | Clustered | 26 | VC_1283_0 | 26 | 0.5446 | 1 | 0.5446 | 0 | 0 | 0 | 1 | Ref |
| 9363 | MK765659 | Unassigned | Unassigned | Unassigned | 1325_0 | Clustered | 7 | VC_1325_0 | 7 | 0.9166 | 0.99999981 | 0.9166 | 0 | 0 | 0 | 1 | Ref |
| 9364 | MK765660 | Unassigned | Unassigned | Unassigned | 1283_0 | Clustered | 26 | VC_1283_0 | 26 | 0.5446 | 1 | 0.5446 | 0 | 0 | 0 | 1 | Ref |
| 9365 | MK765661 | Unassigned | Unassigned | Unassigned | 1211_0 | Clustered | 25 | VC_1211_0 | 25 | 1 | 1 | 1 | 0 | 0 | 0 | 1 | Ref |
| 9366 | MK765662 | Unassigned | Unassigned | Unassigned | 1400_0 | Clustered | 2 | VC_1400_0 | 2 | 1 | 1 | 1 | 0 | 0 | 0 | 1 | Ref |
| 9367 | MK765663 | Unassigned | Unassigned | Unassigned | 1399_0 | Clustered | 9 | VC_1399_0 | 9 | 1 | 0 | 0 | 0 | 0 | 0 | 1 | Ref |
| 9368 | MK765664 | Unassigned | Unassigned | Unassigned | 1325_0 | Clustered | 7 | VC_1325_0 | 7 | 0.9166 | 0.99999981 | 0.9166 | 0 | 0 | 0 | 1 | Ref |
| 9369 | MK765666 | Unassigned | Unassigned | Unassigned | 1283_0 | Clustered | 26 | VC_1283_0 | 26 | 0.5446 | 1 | 0.5446 | 0 | 0 | 0 | 1 | Ref |
| 9370 | MK765667 | Unassigned | Unassigned | Unassigned | 1211_0 | Clustered | 25 | VC_1211_0 | 25 | 1 | 1 | 1 | 0 | 0 | 0 | 1 | Ref |
| 9371 | MK765668 | Unassigned | Unassigned | Unassigned | 1399_0 | Clustered | 9 | VC_1399_0 | 9 | 1 | 0 | 0 | 0 | 0 | 0 | 1 | Ref |
| 9372 | MK765669 | Unassigned | Unassigned | Unassigned | 1119_0 | Clustered | 20 | VC_1119_0 | 20 | 0.6751 | 0.99997892 | 0.6751 | 0 | 0 | 0 | 1 | Ref |
| 9373 | MK770409 | Unassigned | Unassigned | Unassigned | 637_0 | Clustered | 115 | VC_637_0 | 115 | 0.731 | 1 | 0.731 | 0 | 0 | 0 | 0.8764 | Ref |
| 9374 | MK770410 | Unassigned | Unassigned | Unassigned | 191_31 | Clustered | 2 | VC_191_31 | 2 | 0.0057 | 0.92424379 | 0.0052 | 0 | 0 | 0 | 1 | Ref |
| 9375 | MK770411 | Unassigned | Unassigned | Unassigned | 428_0 | Clustered | 33 | VC_428_0 | 33 | 0.9915 | 1 | 0.9915 | 0 | 0 | 0 | 1 | Ref |
| 9376 | MK770412 | Unassigned | Unassigned | Unassigned | 328_0 | Clustered | 11 | VC_328_0 | 11 | 0.4707 | 1 | 0.4707 | 0 | 0 | 0 | 1 | Ref |
| 9377 | MK770413 | Unassigned | Unassigned | Unassigned | 428_0 | Clustered | 33 | VC_428_0 | 33 | 0.9915 | 1 | 0.9915 | 0 | 0 | 0 | 1 | Ref |
| 9378 | MK770414 | Unassigned | Unassigned | Unassigned | 28_1 | Clustered | 53 | VC_28_1 | 53 | 0.7164 | 1 | 0.7164 | 0 | 0 | 0 | 0.9325 | Ref |
| 9379 | MK770415 | Unassigned | Unassigned | Unassigned | 28_1 | Clustered | 53 | VC_28_1 | 53 | 0.7164 | 1 | 0.7164 | 0 | 0 | 0 | 0.9325 | Ref |
| 9380 | MK773491 | Unassigned | Unassigned | Unassigned | 164_0 | Clustered | 3 | VC_164_0 | 3 | 0.0298 | 0.82983726 | 0.0247 | 0 | 0 | 0 | 1 | Ref |
| 9381 | MK774614 | Unassigned | Unassigned | Unassigned | 442_4 | Clustered | 2 | VC_442_4 | 2 | 0.0254 | 0.91859498 | 0.0233 | 0 | 0 | 0 | 1 | Ref |
| 9382 | MK778457 | Unassigned | Unassigned | Unassigned | 585_1 | Clustered | 3 | VC_585_1 | 3 | 0.0313 | 0.99857668 | 0.0312 | 0 | 0 | 0 | 1 | Ref |
| 9383 | MK779875 | Unassigned | Unassigned | Unassigned | 800_0 | Clustered | 3 | VC_800_0 | 3 | 0.4591 | 0.9984737 | 0.4584 | 0 | 0 | 0 | 1 | Ref |
| 9384 | MK792752 | Unassigned | Unassigned | Unassigned | 739_0 | Clustered | 26 | VC_739_0 | 26 | 0.9571 | 1 | 0.9571 | 0 | 0 | 0 | 1 | Ref |
| 9385 | MK795384 | Unassigned | Unassigned | Unassigned | 134_0 | Clustered | 3 | VC_134_0 | 3 | 0.0956 | 0.99858494 | 0.0955 | 0 | 0 | 0 | 1 | Ref |
| 9386 | MK795385 | Unassigned | Unassigned | Unassigned | 823_0 | Clustered | 124 | VC_823_0 | 124 | 0.9492 | 1 | 0.9492 | 0 | 0 | 0 | 1 | Ref |
| 9387 | MK796244 | Unassigned | Unassigned | Unassigned | 971_1 | Clustered | 24 | VC_971_1 | 24 | 0.7393 | 1 | 0.7393 | 0 | 0 | 0 | 0.8913 | Ref |
| 9388 | MK796797 | Unassigned | Unassigned | Unassigned | 807_0 | Clustered | 17 | VC_807_0 | 17 | 0.907 | 1 | 0.907 | 0 | 0 | 0 | 1 | Ref |
| 9389 | MK797984 | Unassigned | Unassigned | Unassigned | 165_0 | Clustered | 4 | VC_165_0 | 4 | 0.2247 | 0.99998328 | 0.2247 | 0 | 0 | 0 | 1 | Ref |
| 9390 | MK798142 | Unassigned | Unassigned | Unassigned | 123_0 | Clustered | 23 | VC_123_0 | 23 | 0.817 | 1 | 0.817 | 0 | 0 | 0 | 1 | Ref |
| 9391 | MK798143 | Unassigned | Unassigned | Unassigned | 64_0 | Clustered | 3 | VC_64_0 | 3 | 0.5503 | 0.99785685 | 0.5491 | 0 | 0 | 0 | 1 | Ref |
| 9392 | MK798144 | Unassigned | Unassigned | Unassigned | 64_0 | Clustered | 3 | VC_64_0 | 3 | 0.5503 | 0.99785685 | 0.5491 | 0 | 0 | 0 | 1 | Ref |
| 9393 | MK799650 | Unassigned | Unassigned | Unassigned | 77_0 | Clustered | 20 | VC_77_0 | 20 | 0.9657 | 1 | 0.9657 | 0 | 0 | 0 | 1 | Ref |
| 9394 | MK799832 | Unassigned | Unassigned | Unassigned | 432_13 | Clustered/Singleton | 1 | VC_432_13 | 1 | 0 | 1 | 0 | 0 | 0 | 0 | 1 | Ref |

| | | | | | | | | | | | | | | | | | |
|---|---|---|---|---|---|---|---|---|---|---|---|---|---|---|---|---|---|
| 9395 | MK800154 | Unassigned | Unassigned | Unassigned | 108_0 | Clustered | 33 | VC_108_0 | 33 | 0.949 | 1 | 0.949 | 0 | 0 | 0 | 1 | Ref |
| 9396 | MK801680 | Unassigned | Unassigned | Unassigned | 8_0 | Clustered | 58 | VC_8_0 | 58 | 0.8306 | 1 | 0.8306 | 0 | 0 | 0 | 0.9946 | Ref |
| 9397 | MK801681 | Unassigned | Unassigned | Unassigned | 8_0 | Clustered | 58 | VC_8_0 | 58 | 0.8306 | 1 | 0.8306 | 0 | 0 | 0 | 0.9946 | Ref |
| 9398 | MK801682 | Unassigned | Unassigned | Unassigned | 9_0 | Clustered | 38 | VC_9_0 | 38 | 0.4895 | 1 | 0.4895 | 0 | 0 | 0 | 1 | Ref |
| 9399 | MK801683 | Unassigned | Unassigned | Unassigned | 9_0 | Clustered | 38 | VC_9_0 | 38 | 0.4895 | 1 | 0.4895 | 0 | 0 | 0 | 1 | Ref |
| 9400 | MK801721 | Unassigned | Unassigned | Unassigned | 1176_0 | Clustered | 41 | VC_1176_0 | 41 | 0.9558 | 1 | 0.9558 | 0 | 0 | 0 | 0.9976 | Ref |
| 9401 | MK801722 | Unassigned | Unassigned | Unassigned | 1071_0 | Clustered | 20 | VC_1071_0 | 20 | 0.3268 | 1 | 0.3268 | 0 | 0 | 0 | 0.9842 | Ref |
| 9402 | MK801723 | Unassigned | Unassigned | Unassigned | 949_0 | Clustered | 10 | VC_949_0 | 10 | 0.5977 | 1 | 0.5977 | 0 | 0 | 0 | 1 | Ref |
| 9403 | MK801724 | Unassigned | Unassigned | Unassigned | 1176_0 | Clustered | 41 | VC_1176_0 | 41 | 0.9558 | 1 | 0.9558 | 0 | 0 | 0 | 0.9976 | Ref |
| 9404 | MK801725 | Unassigned | Unassigned | Unassigned | 510_0 | Clustered | 12 | VC_510_0 | 12 | 0.3963 | 1 | 0.3963 | 0 | 0 | 0 | 0.9697 | Ref |
| 9405 | MK801726 | Unassigned | Unassigned | Unassigned | 551_0 | Clustered | 16 | VC_551_0 | 16 | 0.0915 | 1 | 0.0915 | 0 | 0 | 0 | 0.6667 | Ref |
| 9406 | MK801727 | Unassigned | Unassigned | Unassigned | 525_0 | Clustered | 25 | VC_525_0 | 25 | 0.4597 | 1 | 0.4597 | 0 | 0 | 0 | 1 | Ref |
| 9407 | MK801728 | Unassigned | Unassigned | Unassigned | 868_0 | Clustered | 25 | VC_868_0 | 25 | 0.5791 | 1 | 0.5791 | 0 | 0 | 0 | 1 | Ref |
| 9408 | MK801730 | Unassigned | Unassigned | Unassigned | 1344_0 | Clustered | 78 | VC_1344_0 | 78 | 0.9732 | 1 | 0.9732 | 0 | 0 | 0 | 1 | Ref |
| 9409 | MK801731 | Unassigned | Unassigned | Unassigned | 1343_0 | Clustered | 21 | VC_1343_0 | 21 | 0.9843 | 1 | 0.9843 | 0 | 0 | 0 | 1 | Ref |
| 9410 | MK801732 | Unassigned | Unassigned | Unassigned | 1343_0 | Clustered | 21 | VC_1343_0 | 21 | 0.9843 | 1 | 0.9843 | 0 | 0 | 0 | 1 | Ref |
| 9411 | MK801733 | Unassigned | Unassigned | Unassigned | 868_0 | Clustered | 25 | VC_868_0 | 25 | 0.5791 | 1 | 0.5791 | 0 | 0 | 0 | 1 | Ref |
| 9412 | MK801734 | Unassigned | Unassigned | Unassigned | 1175_0 | Clustered | 22 | VC_1175_0 | 22 | 0.4562 | 1 | 0.4562 | 0 | 0 | 0 | 1 | Ref |
| 9413 | MK801735 | Unassigned | Unassigned | Unassigned | 216_0 | Clustered | 4 | VC_216_0 | 4 | 0.048 | 0.9999878 | 0.048 | 0 | 0 | 0 | 1 | Ref |
| 9414 | MK803322 | Unassigned | Unassigned | Unassigned | 167_0 | Clustered | 3 | VC_167_0 | 3 | 0.6952 | 0.99826575 | 0.694 | 0 | 0 | 0 | 1 | Ref |
| 9415 | MK804891 | Unassigned | Unassigned | Unassigned | 369_0 | Clustered | 6 | VC_369_0 | 6 | 0.5924 | 1 | 0.5924 | 0 | 0 | 0 | 1 | Ref |
| 9416 | MK804892 | Unassigned | Unassigned | Unassigned | 369_0 | Clustered | 6 | VC_369_0 | 6 | 0.5924 | 1 | 0.5924 | 0 | 0 | 0 | 1 | Ref |
| 9417 | MK804893 | Unassigned | Unassigned | Unassigned | 865_0 | Clustered | 2 | VC_865_0 | 2 | 0.0766 | 0.9388444 | 0.0719 | 0 | 0 | 0 | 1 | Ref |
| 9418 | MK809530 | Unassigned | Unassigned | Unassigned | 191_32 | Clustered | 12 | VC_191_32 | 12 | 0.0683 | 1 | 0.0683 | 0 | 0 | 0 | 0.7576 | Ref |
| 9419 | MK813938 | Unassigned | Unassigned | Unassigned | 865_1 | Clustered/Singleton | 1 | VC_865_1 | 1 | 0 | 1 | 0 | 0 | 0 | 0 | 1 | Ref |
| 9420 | MK813939 | Unassigned | Unassigned | Unassigned | 865_0 | Clustered | 2 | VC_865_0 | 2 | 0.0766 | 0.9388444 | 0.0719 | 0 | 0 | 0 | 1 | Ref |
| 9421 | MK813940 | Unassigned | Unassigned | Unassigned | 639_0 | Clustered | 2 | VC_639_0 | 2 | 0.2791 | 0.95526917 | 0.2666 | 0 | 0 | 0 | 1 | Ref |
| 9422 | MK813941 | Unassigned | Unassigned | Unassigned | 865_2 | Clustered/Singleton | 1 | VC_865_2 | 1 | 0 | 1 | 0 | 0 | 0 | 0 | 1 | Ref |
| 9423 | MK814748 | Unassigned | Unassigned | Unassigned | 216_1 | Clustered/Singleton | 1 | VC_216_1 | 1 | 0 | 1 | 0 | 0 | 0 | 0 | 1 | Ref |
| 9424 | MK814749 | Unassigned | Unassigned | Unassigned | 1160_0 | Clustered | 73 | VC_1160_0 | 73 | 0.9835 | 1 | 0.9835 | 0 | 0 | 0 | 1 | Ref |
| 9425 | MK814750 | Unassigned | Unassigned | Unassigned | 1184_0 | Clustered | 5 | VC_1184_0 | 5 | 0.6591 | 0.99999982 | 0.6591 | 0 | 0 | 0 | 1 | Ref |
| 9426 | MK814751 | Unassigned | Unassigned | Unassigned | 508_0 | Clustered | 32 | VC_508_0 | 32 | 0.906 | 1 | 0.906 | 0 | 0 | 0 | 1 | Ref |
| 9427 | MK814752 | Unassigned | Unassigned | Unassigned | 557_1 | Clustered | 11 | VC_557_1 | 11 | 0.4945 | 1 | 0.4945 | 0 | 0 | 0 | 0.7818 | Ref |
| 9428 | MK814753 | Unassigned | Unassigned | Unassigned | 1234_0 | Clustered | 44 | VC_1234_0 | 44 | 1 | 1 | 1 | 0 | 0 | 0 | 1 | Ref |
| 9429 | MK814754 | Unassigned | Unassigned | Unassigned | 325_31 | Clustered/Singleton | 1 | VC_325_31 | 1 | 0 | 1 | 0 | 0 | 0 | 0 | 1 | Ref |
| 9430 | MK814755 | Unassigned | Unassigned | Unassigned | 525_0 | Clustered | 25 | VC_525_0 | 25 | 0.4597 | 1 | 0.4597 | 0 | 0 | 0 | 1 | Ref |
| 9431 | MK814756 | Unassigned | Unassigned | Unassigned | 803_0 | Clustered | 3 | VC_803_0 | 3 | 0.0513 | 0.99857736 | 0.0512 | 0 | 0 | 0 | 0.6667 | Ref |
| 9432 | MK814757 | Unassigned | Unassigned | Unassigned | 1176_0 | Clustered | 41 | VC_1176_0 | 41 | 0.9558 | 1 | 0.9558 | 0 | 0 | 0 | 0.9976 | Ref |
| 9433 | MK814758 | Unassigned | Unassigned | Unassigned | 519_0 | Clustered | 39 | VC_519_0 | 39 | 0.8156 | 1 | 0.8156 | 0 | 0 | 0 | 1 | Ref |
| 9434 | MK814759 | Unassigned | Unassigned | Unassigned | 1180_0 | Clustered | 3 | VC_1180_0 | 3 | 0.5161 | 0.9981786 | 0.5151 | 0 | 0 | 0 | 1 | Ref |
| 9435 | MK814760 | Unassigned | Unassigned | Unassigned | 1098_0 | Clustered | 4 | VC_1098_0 | 4 | 0.5465 | 0.99996462 | 0.5465 | 0 | 0 | 0 | 1 | Ref |
| 9436 | MK814761 | Unassigned | Unassigned | Unassigned | 1178_0 | Clustered | 24 | VC_1178_0 | 24 | 0.9524 | 1 | 0.9524 | 0 | 0 | 0 | 1 | Ref |
| 9437 | MK814762 | Unassigned | Unassigned | Unassigned | 1163_0 | Clustered | 28 | VC_1163_0 | 28 | 0.4923 | 1 | 0.4923 | 0 | 0 | 0 | 1 | Ref |
| 9438 | MK816297 | Unassigned | Unassigned | Unassigned | 796_0 | Clustered | 5 | VC_796_0 | 5 | 0.3352 | 0.99999997 | 0.3352 | 0 | 0 | 0 | 1 | Ref |
| 9439 | MK817115 | Unassigned | Unassigned | Unassigned | 164_11 | Clustered | 10 | VC_164_11 | 10 | 0.1507 | 1 | 0.1507 | 0 | 0 | 0 | 0.9111 | Ref |
| 9440 | MK819239 | Unassigned | Unassigned | Unassigned | 295_0 | Clustered | 2 | VC_295_0 | 2 | 0.0336 | 0.94464231 | 0.0317 | 0 | 0 | 0 | 1 | Ref |
| 9441 | MK820013 | Unassigned | Unassigned | Unassigned | 1027_0 | Clustered | 20 | VC_1027_0 | 20 | 0.7114 | 1 | 0.7114 | 0 | 0 | 0 | 1 | Ref |
| 9442 | MK820638 | Unassigned | Unassigned | Unassigned | 325_23 | Clustered | 17 | VC_325_23 | 17 | 0.0716 | 1 | 0.0716 | 0 | 0 | 0 | 0.6397 | Ref |
| 9443 | MK820639 | Unassigned | Unassigned | Unassigned | 519_0 | Clustered | 39 | VC_519_0 | 39 | 0.8156 | 1 | 0.8156 | 0 | 0 | 0 | 1 | Ref |
| 9444 | MK820640 | Unassigned | Unassigned | Unassigned | 950_0 | Clustered | 38 | VC_950_0 | 38 | 0.9443 | 1 | 0.9443 | 0 | 0 | 0 | 1 | Ref |
| 9445 | MK820641 | Unassigned | Unassigned | Unassigned | 685_0 | Clustered | 16 | VC_685_0 | 16 | 0.2972 | 1 | 0.2972 | 0 | 0 | 0 | 0.9167 | Ref |
| 9446 | MK820642 | Unassigned | Unassigned | Unassigned | 525_0 | Clustered | 25 | VC_525_0 | 25 | 0.4597 | 1 | 0.4597 | 0 | 0 | 0 | 1 | Ref |
| 9447 | MK837009 | Unassigned | Unassigned | Unassigned | 79_5 | Clustered | 2 | VC_79_5 | 2 | 0.0046 | 0.92781966 | 0.0043 | 0 | 0 | 0 | 1 | Ref |
| 9448 | MK837010 | Unassigned | Unassigned | Unassigned | 79_6 | Clustered | 11 | VC_79_6 | 11 | 0.0528 | 1 | 0.0528 | 0 | 0 | 0 | 0.7455 | Ref |
| 9449 | MK837011 | Unassigned | Unassigned | Unassigned | 472_0 | Clustered | 47 | VC_472_0 | 47 | 0.9025 | 1 | 0.9025 | 0 | 0 | 0 | 1 | Ref |
| 9450 | MK837012 | Unassigned | Unassigned | Unassigned | 472_0 | Clustered | 47 | VC_472_0 | 47 | 0.9025 | 1 | 0.9025 | 0 | 0 | 0 | 1 | Ref |
| 9451 | MK838107 | Unassigned | Unassigned | Unassigned | 878_0 | Clustered | 32 | VC_878_0 | 32 | 0.9965 | 1 | 0.9965 | 0 | 0 | 0 | 1 | Ref |
| 9452 | MK838108 | Unassigned | Unassigned | Unassigned | 878_0 | Clustered | 32 | VC_878_0 | 32 | 0.9965 | 1 | 0.9965 | 0 | 0 | 0 | 1 | Ref |
| 9453 | MK838109 | Unassigned | Unassigned | Unassigned | 878_0 | Clustered | 32 | VC_878_0 | 32 | 0.9965 | 1 | 0.9965 | 0 | 0 | 0 | 1 | Ref |
| 9454 | MK838110 | Unassigned | Unassigned | Unassigned | 878_0 | Clustered | 32 | VC_878_0 | 32 | 0.9965 | 1 | 0.9965 | 0 | 0 | 0 | 1 | Ref |
| 9455 | MK838111 | Unassigned | Unassigned | Unassigned | 878_0 | Clustered | 32 | VC_878_0 | 32 | 0.9965 | 1 | 0.9965 | 0 | 0 | 0 | 1 | Ref |
| 9456 | MK838113 | Unassigned | Unassigned | Unassigned | 597_0 | Clustered | 6 | VC_597_0 | 6 | 0.1801 | 1 | 0.1801 | 0 | 0 | 0 | 1 | Ref |
| 9457 | MK838116 | Unassigned | Unassigned | Unassigned | 442_0 | Clustered | 4 | VC_442_0 | 4 | 0.0842 | 0.99996321 | 0.0842 | 0 | 0 | 0 | 1 | Ref |
| 9458 | MK863032 | Unassigned | Unassigned | Unassigned | 779_3 | Clustered | 7 | VC_779_3 | 7 | 0.0766 | 1 | 0.0766 | 0 | 0 | 0 | 0.7143 | Ref |
| 9459 | MK864263 | Unassigned | Unassigned | Unassigned | 1176_0 | Clustered | 41 | VC_1176_0 | 41 | 0.9558 | 1 | 0.9558 | 0 | 0 | 0 | 0.9976 | Ref |
| 9460 | MK864264 | Unassigned | Unassigned | Unassigned | 1178_0 | Clustered | 24 | VC_1178_0 | 24 | 0.9524 | 1 | 0.9524 | 0 | 0 | 0 | 1 | Ref |
| 9461 | MK864265 | Unassigned | Unassigned | Unassigned | 1178_0 | Clustered | 24 | VC_1178_0 | 24 | 0.9524 | 1 | 0.9524 | 0 | 0 | 0 | 1 | Ref |
| 9462 | MK864266 | Unassigned | Unassigned | Unassigned | 1178_0 | Clustered | 24 | VC_1178_0 | 24 | 0.9524 | 1 | 0.9524 | 0 | 0 | 0 | 1 | Ref |
| 9463 | MK864267 | Unassigned | Unassigned | Unassigned | 1178_0 | Clustered | 24 | VC_1178_0 | 24 | 0.9524 | 1 | 0.9524 | 0 | 0 | 0 | 1 | Ref |
| 9464 | MK867354 | Unassigned | Unassigned | Unassigned | 432_5 | Clustered/Singleton | 1 | VC_432_5 | 1 | 0 | 1 | 0 | 0 | 0 | 0 | 1 | Ref |
| 9465 | MK867835 | Unassigned | Unassigned | Unassigned | 191_27 | Clustered/Singleton | 1 | VC_191_27 | 1 | 0 | 1 | 0 | 0 | 0 | 0 | 1 | Ref |
| 9466 | MK875791 | Unassigned | Unassigned | Unassigned | 919_0 | Clustered | 39 | VC_919_0 | 39 | 0.9069 | 1 | 0.9069 | 0 | 0 | 0 | 1 | Ref |
| 9467 | MK875793 | Unassigned | Unassigned | Unassigned | 525_0 | Clustered | 25 | VC_525_0 | 25 | 0.4597 | 1 | 0.4597 | 0 | 0 | 0 | 1 | Ref |
| 9468 | MK875795 | Unassigned | Unassigned | Unassigned | 919_0 | Clustered | 39 | VC_919_0 | 39 | 0.9069 | 1 | 0.9069 | 0 | 0 | 0 | 1 | Ref |
| 9469 | MK875796 | Unassigned | Unassigned | Unassigned | 525_0 | Clustered | 25 | VC_525_0 | 25 | 0.4597 | 1 | 0.4597 | 0 | 0 | 0 | 1 | Ref |
| 9470 | MK875797 | Unassigned | Unassigned | Unassigned | 1175_0 | Clustered | 22 | VC_1175_0 | 22 | 0.4562 | 1 | 0.4562 | 0 | 0 | 0 | 1 | Ref |
| 9471 | MK875798 | Unassigned | Unassigned | Unassigned | 1352_0 | Clustered | 31 | VC_1352_0 | 31 | 0.849 | 1 | 0.849 | 0 | 0 | 0 | 1 | Ref |
| 9472 | MK875799 | Unassigned | Unassigned | Unassigned | 517_0 | Clustered | 18 | VC_517_0 | 18 | 0.1599 | 1 | 0.1599 | 0 | 0 | 0 | 1 | Ref |
| 9473 | MK875800 | Unassigned | Unassigned | Unassigned | 514_0 | Clustered | 75 | VC_514_0 | 75 | 0.9402 | 1 | 0.9402 | 0 | 0 | 0 | 1 | Ref |
| 9474 | MK878895 | Unassigned | Unassigned | Unassigned | 517_1 | Clustered | 108 | VC_517_1 | 108 | 0.8483 | 1 | 0.8483 | 0 | 0 | 0 | 0.9955 | Ref |
| 9475 | MK878896 | Unassigned | Unassigned | Unassigned | 1176_0 | Clustered | 41 | VC_1176_0 | 41 | 0.9558 | 1 | 0.9558 | 0 | 0 | 0 | 0.9976 | Ref |
| 9476 | MK878897 | Unassigned | Unassigned | Unassigned | 558_38 | Clustered | 2 | VC_558_38 | 2 | 0.0032 | 0.64328629 | 0.002 | 0 | 0 | 0 | 1 | Ref |
| 9477 | MK878898 | Unassigned | Unassigned | Unassigned | 525_0 | Clustered | 25 | VC_525_0 | 25 | 0.4597 | 1 | 0.4597 | 0 | 0 | 0 | 1 | Ref |
| 9478 | MK878899 | Unassigned | Unassigned | Unassigned | 1344_0 | Clustered | 78 | VC_1344_0 | 78 | 0.9732 | 1 | 0.9732 | 0 | 0 | 0 | 1 | Ref |
| 9479 | MK878900 | Unassigned | Unassigned | Unassigned | 889_0 | Clustered | 16 | VC_889_0 | 16 | 0.2969 | 1 | 0.2969 | 0 | 0 | 0 | 1 | Ref |
| 9480 | MK878901 | Unassigned | Unassigned | Unassigned | 1177_0 | Clustered | 14 | VC_1177_0 | 14 | 0.7847 | 1 | 0.7847 | 0 | 0 | 0 | 0.989 | Ref |
| 9481 | MK878903 | Unassigned | Unassigned | Unassigned | 866_1 | Clustered | 20 | VC_866_1 | 20 | 0.4814 | 1 | 0.4814 | 0 | 0 | 0 | 1 | Ref |
| 9482 | MK878904 | Unassigned | Unassigned | Unassigned | 1360_0 | Clustered | 31 | VC_1360_0 | 31 | 0.8837 | 1 | 0.8837 | 0 | 0 | 0 | 1 | Ref |
| 9483 | MK878905 | Unassigned | Unassigned | Unassigned | 517_1 | Clustered | 108 | VC_517_1 | 108 | 0.8483 | 1 | 0.8483 | 0 | 0 | 0 | 0.9955 | Ref |
| 9484 | MK880124 | Unassigned | Unassigned | Unassigned | 1206_0 | Clustered | 8 | VC_1206_0 | 8 | 0.1913 | 1 | 0.1913 | 0 | 0 | 0 | 1 | Ref |
| 9485 | MK883717 | Unassigned | Unassigned | Unassigned | 772_4 | Clustered | 4 | VC_772_4 | 4 | 0.0181 | 0.73018303 | 0.0132 | 0 | 0 | 0 | 1 | Ref |
| 9486 | MK883718 | Unassigned | Unassigned | Unassigned | 772_4 | Clustered | 4 | VC_772_4 | 4 | 0.0181 | 0.73018303 | 0.0132 | 0 | 0 | 0 | 1 | Ref |
| 9487 | MK886800 | Unassigned | Unassigned | Unassigned | 283_164 | Clustered | 2 | VC_283_164 | 2 | 0.003 | 0.93475788 | 0.0028 | 0 | 0 | 0 | 1 | Ref |
| 9488 | MK893987 | Unassigned | Unassigned | Unassigned | 667_2 | Clustered/Singleton | 1 | VC_667_2 | 1 | 0 | 1 | 0 | 0 | 0 | 0 | 1 | Ref |
| 9489 | MK894432 | Unassigned | Unassigned | Unassigned | 1163_0 | Clustered | 28 | VC_1163_0 | 28 | 0.4923 | 1 | 0.4923 | 0 | 0 | 0 | 1 | Ref |
| 9490 | MK894433 | Unassigned | Unassigned | Unassigned | 223_0 | Clustered | 38 | VC_223_0 | 38 | 0.9095 | 1 | 0.9095 | 0 | 0 | 0 | 1 | Ref |
| 9491 | MK894434 | Unassigned | Unassigned | Unassigned | 1344_0 | Clustered | 78 | VC_1344_0 | 78 | 0.9732 | 1 | 0.9732 | 0 | 0 | 0 | 1 | Ref |
| 9492 | MK894435 | Unassigned | Unassigned | Unassigned | 1360_0 | Clustered | 31 | VC_1360_0 | 31 | 0.8837 | 1 | 0.8837 | 0 | 0 | 0 | 1 | Ref |
| 9493 | MK894436 | Unassigned | Unassigned | Unassigned | 1098_0 | Clustered | 4 | VC_1098_0 | 4 | 0.5465 | 0.99996462 | 0.5465 | 0 | 0 | 0 | 1 | Ref |
| 9494 | MK894438 | Unassigned | Unassigned | Unassigned | 1360_0 | Clustered | 31 | VC_1360_0 | 31 | 0.8837 | 1 | 0.8837 | 0 | 0 | 0 | 1 | Ref |
| 9495 | MK895508 | Unassigned | Unassigned | Unassigned | 971_1 | Clustered | 24 | VC_971_1 | 24 | 0.7393 | 1 | 0.7393 | 0 | 0 | 0 | 0.8913 | Ref |
| 9496 | MK903033 | Unassigned | Unassigned | Unassigned | 96_0 | Clustered | 30 | VC_96_0 | 30 | 0.718 | 1 | 0.718 | 0 | 0 | 0 | 1 | Ref |
| 9497 | MK903277 | Unassigned | Unassigned | Unassigned | 451_0 | Clustered | 80 | VC_451_0 | 80 | 0.9991 | 1 | 0.9991 | 0 | 0 | 0 | 1 | Ref |
| 9498 | MK903278 | Unassigned | Unassigned | Unassigned | 74_0 | Clustered | 15 | VC_74_0 | 15 | 0.5204 | 1 | 0.5204 | 0 | 0 | 0 | 1 | Ref |
| 9499 | MK903279 | Unassigned | Unassigned | Unassigned | 451_0 | Clustered | 80 | VC_451_0 | 80 | 0.9991 | 1 | 0.9991 | 0 | 0 | 0 | 1 | Ref |
| 9500 | MK903281 | Unassigned | Unassigned | Unassigned | 451_0 | Clustered | 80 | VC_451_0 | 80 | 0.9991 | 1 | 0.9991 | 0 | 0 | 0 | 1 | Ref |
| 9501 | MK903282 | Unassigned | Unassigned | Unassigned | 618_0 | Clustered | 13 | VC_618_0 | 13 | 0.6252 | 1 | 0.6252 | 0 | 0 | 0 | 1 | Ref |
| 9502 | MK903728 | Unassigned | Unassigned | Unassigned | 823_0 | Clustered | 124 | VC_823_0 | 124 | 0.9492 | 1 | 0.9492 | 0 | 0 | 0 | 1 | Ref |
| 9503 | MK907226 | Unassigned | Unassigned | Unassigned | 456_0 | Clustered | 62 | VC_456_0 | 62 | 0.9026 | 1 | 0.9026 | 0 | 0 | 0 | 1 | Ref |
| 9504 | MK907227 | Unassigned | Unassigned | Unassigned | 456_0 | Clustered | 62 | VC_456_0 | 62 | 0.9026 | 1 | 0.9026 | 0 | 0 | 0 | 1 | Ref |
| 9505 | MK907228 | Unassigned | Unassigned | Unassigned | 456_0 | Clustered | 62 | VC_456_0 | 62 | 0.9026 | 1 | 0.9026 | 0 | 0 | 0 | 1 | Ref |
| 9506 | MK907229 | Unassigned | Unassigned | Unassigned | 456_0 | Clustered | 62 | VC_456_0 | 62 | 0.9026 | 1 | 0.9026 | 0 | 0 | 0 | 1 | Ref |
| 9507 | MK907230 | Unassigned | Unassigned | Unassigned | 456_0 | Clustered | 62 | VC_456_0 | 62 | 0.9026 | 1 | 0.9026 | 0 | 0 | 0 | 1 | Ref |
| 9508 | MK907231 | Unassigned | Unassigned | Unassigned | 456_0 | Clustered | 62 | VC_456_0 | 62 | 0.9026 | 1 | 0.9026 | 0 | 0 | 0 | 1 | Ref |
| 9509 | MK907232 | Unassigned | Unassigned | Unassigned | 456_0 | Clustered | 62 | VC_456_0 | 62 | 0.9026 | 1 | 0.9026 | 0 | 0 | 0 | 1 | Ref |
| 9510 | MK907233 | Unassigned | Unassigned | Unassigned | 456_0 | Clustered | 62 | VC_456_0 | 62 | 0.9026 | 1 | 0.9026 | 0 | 0 | 0 | 1 | Ref |
| 9511 | MK907234 | Unassigned | Unassigned | Unassigned | 456_0 | Clustered | 62 | VC_456_0 | 62 | 0.9026 | 1 | 0.9026 | 0 | 0 | 0 | 1 | Ref |
| 9512 | MK907235 | Unassigned | Unassigned | Unassigned | 456_0 | Clustered | 62 | VC_456_0 | 62 | 0.9026 | 1 | 0.9026 | 0 | 0 | 0 | 1 | Ref |
| 9513 | MK907236 | Unassigned | Unassigned | Unassigned | 228_0 | Clustered | 60 | VC_228_0 | 60 | 0.942 | 1 | 0.942 | 0 | 0 | 0 | 1 | Ref |
| 9514 | MK907237 | Unassigned | Unassigned | Unassigned | 228_0 | Clustered | 60 | VC_228_0 | 60 | 0.942 | 1 | 0.942 | 0 | 0 | 0 | 1 | Ref |
| 9515 | MK907238 | Unassigned | Unassigned | Unassigned | 228_0 | Clustered | 60 | VC_228_0 | 60 | 0.942 | 1 | 0.942 | 0 | 0 | 0 | 1 | Ref |
| 9516 | MK907239 | Unassigned | Unassigned | Unassigned | 228_0 | Clustered | 60 | VC_228_0 | 60 | 0.942 | 1 | 0.942 | 0 | 0 | 0 | 1 | Ref |
| 9517 | MK907240 | Unassigned | Unassigned | Unassigned | 456_0 | Clustered | 62 | VC_456_0 | 62 | 0.9026 | 1 | 0.9026 | 0 | 0 | 0 | 1 | Ref |
| 9518 | MK907241 | Unassigned | Unassigned | Unassigned | 456_0 | Clustered | 62 | VC_456_0 | 62 | 0.9026 | 1 | 0.9026 | 0 | 0 | 0 | 1 | Ref |
| 9519 | MK907242 | Unassigned | Unassigned | Unassigned | 456_0 | Clustered | 62 | VC_456_0 | 62 | 0.9026 | 1 | 0.9026 | 0 | 0 | 0 | 1 | Ref |
| 9520 | MK907243 | Unassigned | Unassigned | Unassigned | 456_0 | Clustered | 62 | VC_456_0 | 62 | 0.9026 | 1 | 0.9026 | 0 | 0 | 0 | 1 | Ref |
| 9521 | MK907244 | Unassigned | Unassigned | Unassigned | 456_0 | Clustered | 62 | VC_456_0 | 62 | 0.9026 | 1 | 0.9026 | 0 | 0 | 0 | 1 | Ref |
| 9522 | MK907245 | Unassigned | Unassigned | Unassigned | 456_0 | Clustered | 62 | VC_456_0 | 62 | 0.9026 | 1 | 0.9026 | 0 | 0 | 0 | 1 | Ref |
| 9523 | MK907246 | Unassigned | Unassigned | Unassigned | 456_0 | Clustered | 62 | VC_456_0 | 62 | 0.9026 | 1 | 0.9026 | 0 | 0 | 0 | 1 | Ref |
| 9524 | MK907247 | Unassigned | Unassigned | Unassigned | 456_0 | Clustered | 62 | VC_456_0 | 62 | 0.9026 | 1 | 0.9026 | 0 | 0 | 0 | 1 | Ref |
| 9525 | MK907248 | Unassigned | Unassigned | Unassigned | 456_0 | Clustered | 62 | VC_456_0 | 62 | 0.9026 | 1 | 0.9026 | 0 | 0 | 0 | 1 | Ref |
| 9526 | MK907249 | Unassigned | Unassigned | Unassigned | 456_0 | Clustered | 62 | VC_456_0 | 62 | 0.9026 | 1 | 0.9026 | 0 | 0 | 0 | 1 | Ref |
| 9527 | MK907250 | Unassigned | Unassigned | Unassigned | 456_0 | Clustered | 62 | VC_456_0 | 62 | 0.9026 | 1 | 0.9026 | 0 | 0 | 0 | 1 | Ref |
| 9528 | MK907251 | Unassigned | Unassigned | Unassigned | 456_0 | Clustered | 62 | VC_456_0 | 62 | 0.9026 | 1 | 0.9026 | 0 | 0 | 0 | 1 | Ref |
| 9529 | MK907252 | Unassigned | Unassigned | Unassigned | 456_0 | Clustered | 62 | VC_456_0 | 62 | 0.9026 | 1 | 0.9026 | 0 | 0 | 0 | 1 | Ref |
| 9530 | MK907253 | Unassigned | Unassigned | Unassigned | 456_0 | Clustered | 62 | VC_456_0 | 62 | 0.9026 | 1 | 0.9026 | 0 | 0 | 0 | 1 | Ref |
| 9531 | MK907254 | Unassigned | Unassigned | Unassigned | 456_0 | Clustered | 62 | VC_456_0 | 62 | 0.9026 | 1 | 0.9026 | 0 | 0 | 0 | 1 | Ref |
| 9532 | MK907255 | Unassigned | Unassigned | Unassigned | 456_0 | Clustered | 62 | VC_456_0 | 62 | 0.9026 | 1 | 0.9026 | 0 | 0 | 0 | 1 | Ref |
| 9533 | MK907256 | Unassigned | Unassigned | Unassigned | 456_0 | Clustered | 62 | VC_456_0 | 62 | 0.9026 | 1 | 0.9026 | 0 | 0 | 0 | 1 | Ref |
| 9534 | MK907257 | Unassigned | Unassigned | Unassigned | 456_0 | Clustered | 62 | VC_456_0 | 62 | 0.9026 | 1 | 0.9026 | 0 | 0 | 0 | 1 | Ref |
| 9535 | MK907258 | Unassigned | Unassigned | Unassigned | 456_0 | Clustered | 62 | VC_456_0 | 62 | 0.9026 | 1 | 0.9026 | 0 | 0 | 0 | 1 | Ref |
| 9536 | MK907259 | Unassigned | Unassigned | Unassigned | 456_0 | Clustered | 62 | VC_456_0 | 62 | 0.9026 | 1 | 0.9026 | 0 | 0 | 0 | 1 | Ref |
| 9537 | MK907260 | Unassigned | Unassigned | Unassigned | 456_0 | Clustered | 62 | VC_456_0 | 62 | 0.9026 | 1 | 0.9026 | 0 | 0 | 0 | 1 | Ref |
| 9538 | MK907261 | Unassigned | Unassigned | Unassigned | 456_0 | Clustered | 62 | VC_456_0 | 62 | 0.9026 | 1 | 0.9026 | 0 | 0 | 0 | 1 | Ref |
| 9539 | MK907262 | Unassigned | Unassigned | Unassigned | 456_0 | Clustered | 62 | VC_456_0 | 62 | 0.9026 | 1 | 0.9026 | 0 | 0 | 0 | 1 | Ref |
| 9540 | MK907263 | Unassigned | Unassigned | Unassigned | 456_0 | Clustered | 62 | VC_456_0 | 62 | 0.9026 | 1 | 0.9026 | 0 | 0 | 0 | 1 | Ref |
| 9541 | MK907264 | Unassigned | Unassigned | Unassigned | 456_0 | Clustered | 62 | VC_456_0 | 62 | 0.9026 | 1 | 0.9026 | 0 | 0 | 0 | 1 | Ref |
| 9542 | MK907265 | Unassigned | Unassigned | Unassigned | 456_0 | Clustered | 62 | VC_456_0 | 62 | 0.9026 | 1 | 0.9026 | 0 | 0 | 0 | 1 | Ref |
| 9543 | MK907266 | Unassigned | Unassigned | Unassigned | 456_0 | Clustered | 62 | VC_456_0 | 62 | 0.9026 | 1 | 0.9026 | 0 | 0 | 0 | 1 | Ref |
| 9544 | MK907267 | Unassigned | Unassigned | Unassigned | 191_13 | Clustered | 5 | VC_191_13 | 5 | 0.0233 | 0.99999927 | 0.0233 | 0 | 0 | 0 | 1 | Ref |
| 9545 | MK907268 | Unassigned | Unassigned | Unassigned | 191_13 | Clustered | 5 | VC_191_13 | 5 | 0.0233 | 0.99999927 | 0.0233 | 0 | 0 | 0 | 1 | Ref |
| 9546 | MK907269 | Unassigned | Unassigned | Unassigned | 191_13 | Clustered | 5 | VC_191_13 | 5 | 0.0233 | 0.99999927 | 0.0233 | 0 | 0 | 0 | 1 | Ref |
| 9547 | MK907270 | Unassigned | Unassigned | Unassigned | 191_13 | Clustered | 5 | VC_191_13 | 5 | 0.0233 | 0.99999927 | 0.0233 | 0 | 0 | 0 | 1 | Ref |
| 9548 | MK907271 | Unassigned | Unassigned | Unassigned | 191_13 | Clustered | 5 | VC_191_13 | 5 | 0.0233 | 0.99999927 | 0.0233 | 0 | 0 | 0 | 1 | Ref |
| 9549 | MK907272 | Unassigned | Unassigned | Unassigned | 456_0 | Clustered | 62 | VC_456_0 | 62 | 0.9026 | 1 | 0.9026 | 0 | 0 | 0 | 1 | Ref |
| 9550 | MK907273 | Unassigned | Unassigned | Unassigned | 456_0 | Clustered | 62 | VC_456_0 | 62 | 0.9026 | 1 | 0.9026 | 0 | 0 | 0 | 1 | Ref |
| 9551 | MK907274 | Unassigned | Unassigned | Unassigned | 456_0 | Clustered | 62 | VC_456_0 | 62 | 0.9026 | 1 | 0.9026 | 0 | 0 | 0 | 1 | Ref |
| 9552 | MK907275 | Unassigned | Unassigned | Unassigned | 456_0 | Clustered | 62 | VC_456_0 | 62 | 0.9026 | 1 | 0.9026 | 0 | 0 | 0 | 1 | Ref |
| 9553 | MK907276 | Unassigned | Unassigned | Unassigned | 456_0 | Clustered | 62 | VC_456_0 | 62 | 0.9026 | 1 | 0.9026 | 0 | 0 | 0 | 1 | Ref |
| 9554 | MK907285 | Unassigned | Unassigned | Unassigned | 386_0 | Clustered | 49 | VC_386_0 | 49 | 0.7941 | 1 | 0.7941 | 0 | 0 | 0 | 1 | Ref |
| 9555 | MK907780 | Unassigned | Unassigned | Unassigned | 971_1 | Clustered | 24 | VC_971_1 | 24 | 0.7393 | 1 | 0.7393 | 0 | 0 | 0 | 0.8913 | Ref |
| 9556 | MK919469 | Unassigned | Unassigned | Unassigned | 525_1 | Clustered | 5 | VC_525_1 | 5 | 0.0485 | 0.99999947 | 0.0485 | 0 | 0 | 0 | 0.9 | Ref |

| | | | | | | | | | | | | | | | | | |
|---|---|---|---|---|---|---|---|---|---|---|---|---|---|---|---|---|---|
| 9557 | MK919470 | Unassigned | Unassigned | Unassigned | 1176_0 | Clustered | 41 | VC_1176_0 | 41 | 0.9558 | 1 | 0.9558 | 0 | 0 | 0 | 0.9976 | Ref |
| 9558 | MK919471 | Unassigned | Unassigned | Unassigned | 1240_0 | Clustered | 82 | VC_1240_0 | 82 | 0.7945 | 1 | 0.7945 | 0 | 0 | 0 | 1 | Ref |
| 9559 | MK919472 | Unassigned | Unassigned | Unassigned | 216_6 | Clustered | 42 | VC_216_6 | 42 | 0.473 | 1 | 0.473 | 0 | 0 | 0 | 0.8351 | Ref |
| 9560 | MK919473 | Unassigned | Unassigned | Unassigned | 802_0 | Clustered | 17 | VC_802_0 | 17 | 0.8025 | 1 | 0.8025 | 0 | 0 | 0 | 1 | Ref |
| 9561 | MK919474 | Unassigned | Unassigned | Unassigned | 856_1 | Clustered | 13 | VC_856_1 | 13 | 0.5848 | 1 | 0.5848 | 0 | 0 | 0 | 1 | Ref |
| 9562 | MK919475 | Unassigned | Unassigned | Unassigned | 761_0 | Clustered | 67 | VC_761_0 | 67 | 0.8947 | 1 | 0.8947 | 0 | 0 | 0 | 1 | Ref |
| 9563 | MK919476 | Unassigned | Unassigned | Unassigned | 1160_0 | Clustered | 73 | VC_1160_0 | 73 | 0.9835 | 1 | 0.9835 | 0 | 0 | 0 | 1 | Ref |
| 9564 | MK919477 | Unassigned | Unassigned | Unassigned | 866_1 | Clustered | 20 | VC_866_1 | 20 | 0.4814 | 1 | 0.4814 | 0 | 0 | 0 | 1 | Ref |
| 9565 | MK919478 | Unassigned | Unassigned | Unassigned | 1183_0 | Clustered | 6 | VC_1183_0 | 6 | 0.995 | 0.99996974 | 0.995 | 0 | 0 | 0 | 1 | Ref |
| 9566 | MK919479 | Unassigned | Unassigned | Unassigned | 1073_0 | Clustered | 24 | VC_1073_0 | 24 | 0.8388 | 1 | 0.8388 | 0 | 0 | 0 | 1 | Ref |
| 9567 | MK919480 | Unassigned | Unassigned | Unassigned | 519_0 | Clustered | 39 | VC_519_0 | 39 | 0.8156 | 1 | 0.8156 | 0 | 0 | 0 | 1 | Ref |
| 9568 | MK919481 | Unassigned | Unassigned | Unassigned | 866_1 | Clustered | 20 | VC_866_1 | 20 | 0.4814 | 1 | 0.4814 | 0 | 0 | 0 | 1 | Ref |
| 9569 | MK919482 | Unassigned | Unassigned | Unassigned | 810_0 | Clustered | 18 | VC_810_0 | 18 | 0.9121 | 1 | 0.9121 | 0 | 0 | 0 | 1 | Ref |
| 9570 | MK919483 | Unassigned | Unassigned | Unassigned | 525_0 | Clustered | 25 | VC_525_0 | 25 | 0.4597 | 1 | 0.4597 | 0 | 0 | 0 | 1 | Ref |
| 9571 | MK919484 | Unassigned | Unassigned | Unassigned | 1160_0 | Clustered | 73 | VC_1160_0 | 73 | 0.9835 | 1 | 0.9835 | 0 | 0 | 0 | 1 | Ref |
| 9572 | MK922546 | Unassigned | Unassigned | Unassigned | 96_0 | Clustered | 30 | VC_96_0 | 30 | 0.718 | 1 | 0.718 | 0 | 0 | 0 | 1 | Ref |
| 9573 | MK922547 | Unassigned | Unassigned | Unassigned | 96_0 | Clustered | 30 | VC_96_0 | 30 | 0.718 | 1 | 0.718 | 0 | 0 | 0 | 1 | Ref |
| 9574 | MK922548 | Unassigned | Unassigned | Unassigned | 96_0 | Clustered | 30 | VC_96_0 | 30 | 0.718 | 1 | 0.718 | 0 | 0 | 0 | 1 | Ref |
| 9575 | MK929790 | Unassigned | Unassigned | Unassigned | 1407_0 | Clustered | 2 | VC_1407_0 | 2 | 0.8176 | 0.81445332 | 0.6659 | 0 | 0 | 0 | 1 | Ref |
| 9576 | MK931297 | Unassigned | Unassigned | Unassigned | 779_0 | Clustered | 37 | VC_779_0 | 37 | 0.5682 | 1 | 0.5682 | 0 | 0 | 0 | 1 | Ref |
| 9577 | MK931438 | Unassigned | Unassigned | Unassigned | 638_0 | Clustered | 45 | VC_638_0 | 45 | 0.7741 | 1 | 0.7741 | 0 | 0 | 0 | 1 | Ref |
| 9578 | MK931439 | Unassigned | Unassigned | Unassigned | 638_0 | Clustered | 45 | VC_638_0 | 45 | 0.7741 | 1 | 0.7741 | 0 | 0 | 0 | 1 | Ref |
| 9579 | MK931440 | Unassigned | Unassigned | Unassigned | 638_0 | Clustered | 45 | VC_638_0 | 45 | 0.7741 | 1 | 0.7741 | 0 | 0 | 0 | 1 | Ref |
| 9580 | MK931441 | Unassigned | Unassigned | Unassigned | 638_0 | Clustered | 45 | VC_638_0 | 45 | 0.7741 | 1 | 0.7741 | 0 | 0 | 0 | 1 | Ref |
| 9581 | MK931442 | Unassigned | Unassigned | Unassigned | 938_0 | Clustered | 99 | VC_938_0 | 99 | 0.9119 | 1 | 0.9119 | 0 | 0 | 0 | 0.9928 | Ref |
| 9582 | MK931443 | Unassigned | Unassigned | Unassigned | 938_0 | Clustered | 99 | VC_938_0 | 99 | 0.9119 | 1 | 0.9119 | 0 | 0 | 0 | 0.9928 | Ref |
| 9583 | MK931444 | Unassigned | Unassigned | Unassigned | 938_0 | Clustered | 99 | VC_938_0 | 99 | 0.9119 | 1 | 0.9119 | 0 | 0 | 0 | 0.9928 | Ref |
| 9584 | MK931445 | Unassigned | Unassigned | Unassigned | 938_0 | Clustered | 99 | VC_938_0 | 99 | 0.9119 | 1 | 0.9119 | 0 | 0 | 0 | 0.9928 | Ref |
| 9585 | MK931446 | Unassigned | Unassigned | Unassigned | 637_1 | Clustered | 27 | VC_637_1 | 27 | 0.2578 | 1 | 0.2578 | 0 | 0 | 0 | 0.9858 | Ref |
| 9586 | MK932884 | Unassigned | Unassigned | Unassigned | 851_0 | Clustered | 2 | VC_851_0 | 2 | 0.4472 | 0.9410422 | 0.4209 | 0 | 0 | 0 | 1 | Ref |
| 9587 | MK934841 | Unassigned | Unassigned | Unassigned | 251_0 | Clustered | 22 | VC_251_0 | 22 | 0.6508 | 1 | 0.6508 | 0 | 0 | 0 | 1 | Ref |
| 9588 | MK936475 | Unassigned | Unassigned | Unassigned | 96_0 | Clustered | 30 | VC_96_0 | 30 | 0.718 | 1 | 0.718 | 0 | 0 | 0 | 1 | Ref |
| 9589 | MK936476 | Unassigned | Unassigned | Unassigned | 96_0 | Clustered | 30 | VC_96_0 | 30 | 0.718 | 1 | 0.718 | 0 | 0 | 0 | 1 | Ref |
| 9590 | MK937591 | Unassigned | Unassigned | Unassigned | 1349_0 | Clustered | 7 | VC_1349_0 | 7 | 0.2495 | 1 | 0.2495 | 0 | 0 | 0 | 1 | Ref |
| 9591 | MK937593 | Unassigned | Unassigned | Unassigned | 551_0 | Clustered | 16 | VC_551_0 | 16 | 0.0915 | 1 | 0.0915 | 0 | 0 | 0 | 0.6667 | Ref |
| 9592 | MK937594 | Unassigned | Unassigned | Unassigned | 1093_0 | Clustered | 40 | VC_1093_0 | 40 | 0.8754 | 1 | 0.8754 | 0 | 0 | 0 | 1 | Ref |
| 9593 | MK937595 | Unassigned | Unassigned | Unassigned | 374_0 | Clustered | 2 | VC_374_0 | 2 | 0.1258 | 0.95578317 | 0.1203 | 0 | 0 | 0 | 1 | Ref |
| 9594 | MK937596 | Unassigned | Unassigned | Unassigned | 856_1 | Clustered | 13 | VC_856_1 | 13 | 0.5848 | 1 | 0.5848 | 0 | 0 | 0 | 1 | Ref |
| 9595 | MK937597 | Unassigned | Unassigned | Unassigned | 557_0 | Clustered | 3 | VC_557_0 | 3 | 0.0814 | 0.99853279 | 0.0812 | 0 | 0 | 0 | 0.6667 | Ref |
| 9596 | MK937598 | Unassigned | Unassigned | Unassigned | 1234_0 | Clustered | 44 | VC_1234_0 | 44 | 1 | 1 | 1 | 0 | 0 | 0 | 1 | Ref |
| 9597 | MK937599 | Unassigned | Unassigned | Unassigned | 1177_0 | Clustered | 14 | VC_1177_0 | 14 | 0.7847 | 1 | 0.7847 | 0 | 0 | 0 | 0.989 | Ref |
| 9598 | MK937600 | Unassigned | Unassigned | Unassigned | 888_0 | Clustered | 28 | VC_888_0 | 28 | 0.5497 | 1 | 0.5497 | 0 | 0 | 0 | 1 | Ref |
| 9599 | MK937601 | Unassigned | Unassigned | Unassigned | 1093_0 | Clustered | 40 | VC_1093_0 | 40 | 0.8754 | 1 | 0.8754 | 0 | 0 | 0 | 1 | Ref |
| 9600 | MK937602 | Unassigned | Unassigned | Unassigned | 1163_0 | Clustered | 28 | VC_1163_0 | 28 | 0.4923 | 1 | 0.4923 | 0 | 0 | 0 | 1 | Ref |
| 9601 | MK937603 | Unassigned | Unassigned | Unassigned | 1178_0 | Clustered | 24 | VC_1178_0 | 24 | 0.9524 | 1 | 0.9524 | 0 | 0 | 0 | 1 | Ref |
| 9602 | MK937604 | Unassigned | Unassigned | Unassigned | 519_0 | Clustered | 39 | VC_519_0 | 39 | 0.8156 | 1 | 0.8156 | 0 | 0 | 0 | 1 | Ref |
| 9603 | MK937606 | Unassigned | Unassigned | Unassigned | 1349_0 | Clustered | 7 | VC_1349_0 | 7 | 0.2495 | 1 | 0.2495 | 0 | 0 | 0 | 1 | Ref |
| 9604 | MK937607 | Unassigned | Unassigned | Unassigned | 1095_0 | Clustered | 16 | VC_1095_0 | 16 | 0.458 | 1 | 0.458 | 0 | 0 | 0 | 1 | Ref |
| 9605 | MK937608 | Unassigned | Unassigned | Unassigned | 1349_0 | Clustered | 7 | VC_1349_0 | 7 | 0.2495 | 1 | 0.2495 | 0 | 0 | 0 | 1 | Ref |
| 9606 | MK937610 | Unassigned | Unassigned | Unassigned | 1234_0 | Clustered | 44 | VC_1234_0 | 44 | 1 | 1 | 1 | 0 | 0 | 0 | 1 | Ref |
| 9607 | MK937611 | Unassigned | Unassigned | Unassigned | 1095_0 | Clustered | 16 | VC_1095_0 | 16 | 0.458 | 1 | 0.458 | 0 | 0 | 0 | 1 | Ref |
| 9608 | MK937612 | Unassigned | Unassigned | Unassigned | 763_1 | Clustered | 9 | VC_763_1 | 9 | 0.3706 | 1 | 0.3706 | 0 | 0 | 0 | 1 | Ref |
| 9609 | MK937613 | Unassigned | Unassigned | Unassigned | 1199_0 | Clustered | 8 | VC_1199_0 | 8 | 0.7928 | 1 | 0.7928 | 0 | 0 | 0 | 1 | Ref |
| 9610 | MK940809 | Unassigned | Unassigned | Unassigned | 8_0 | Clustered | 58 | VC_8_0 | 58 | 0.8306 | 1 | 0.8306 | 0 | 0 | 0 | 0.9946 | Ref |
| 9611 | MK947458 | Unassigned | Unassigned | Unassigned | 191_16 | Clustered | 12 | VC_191_16 | 12 | 0.0643 | 1 | 0.0643 | 0 | 0 | 0 | 0.9394 | Ref |
| 9612 | MK947459 | Unassigned | Unassigned | Unassigned | 191_17 | Clustered | 4 | VC_191_17 | 4 | 0.0166 | 0.99930063 | 0.0166 | 0 | 0 | 0 | 0.8333 | Ref |
| 9613 | MK947460 | Unassigned | Unassigned | Unassigned | 461_0 | Clustered | 96 | VC_461_0 | 96 | 0.9307 | 1 | 0.9307 | 0 | 0 | 0 | 0.9575 | Ref |
| 9614 | MK962626 | Unassigned | Unassigned | Unassigned | 779_1 | Clustered | 11 | VC_779_1 | 11 | 0.1587 | 1 | 0.1587 | 0 | 0 | 0 | 1 | Ref |
| 9615 | MK962627 | Unassigned | Unassigned | Unassigned | 171_0 | Clustered | 8 | VC_171_0 | 8 | 0.5884 | 1 | 0.5884 | 0 | 0 | 0 | 1 | Ref |
| 9616 | MK962628 | Unassigned | Unassigned | Unassigned | 157_0 | Clustered | 3 | VC_157_0 | 3 | 0.3732 | 0.99808097 | 0.3725 | 0 | 0 | 0 | 1 | Ref |
| 9617 | MK962629 | Unassigned | Unassigned | Unassigned | 779_1 | Clustered | 11 | VC_779_1 | 11 | 0.1587 | 1 | 0.1587 | 0 | 0 | 0 | 1 | Ref |
| 9618 | MK962630 | Unassigned | Unassigned | Unassigned | 779_1 | Clustered | 11 | VC_779_1 | 11 | 0.1587 | 1 | 0.1587 | 0 | 0 | 0 | 1 | Ref |
| 9619 | MK962631 | Unassigned | Unassigned | Unassigned | 779_1 | Clustered | 11 | VC_779_1 | 11 | 0.1587 | 1 | 0.1587 | 0 | 0 | 0 | 1 | Ref |
| 9620 | MK962632 | Unassigned | Unassigned | Unassigned | 779_1 | Clustered | 11 | VC_779_1 | 11 | 0.1587 | 1 | 0.1587 | 0 | 0 | 0 | 1 | Ref |
| 9621 | MK962633 | Unassigned | Unassigned | Unassigned | 171_0 | Clustered | 8 | VC_171_0 | 8 | 0.5884 | 1 | 0.5884 | 0 | 0 | 0 | 1 | Ref |
| 9622 | MK962634 | Unassigned | Unassigned | Unassigned | 171_0 | Clustered | 8 | VC_171_0 | 8 | 0.5884 | 1 | 0.5884 | 0 | 0 | 0 | 1 | Ref |
| 9623 | MK962635 | Unassigned | Unassigned | Unassigned | 751_0 | Clustered | 4 | VC_751_0 | 4 | 0.1763 | 0.99998526 | 0.1763 | 0 | 0 | 0 | 1 | Ref |
| 9624 | MK962636 | Unassigned | Unassigned | Unassigned | 751_0 | Clustered | 4 | VC_751_0 | 4 | 0.1763 | 0.99998526 | 0.1763 | 0 | 0 | 0 | 1 | Ref |
| 9625 | MK962637 | Unassigned | Unassigned | Unassigned | 751_0 | Clustered | 4 | VC_751_0 | 4 | 0.1763 | 0.99998526 | 0.1763 | 0 | 0 | 0 | 1 | Ref |
| 9626 | MK962638 | Unassigned | Unassigned | Unassigned | 157_0 | Clustered | 3 | VC_157_0 | 3 | 0.3732 | 0.99808097 | 0.3725 | 0 | 0 | 0 | 1 | Ref |
| 9627 | MK962639 | Unassigned | Unassigned | Unassigned | 779_1 | Clustered | 11 | VC_779_1 | 11 | 0.1587 | 1 | 0.1587 | 0 | 0 | 0 | 1 | Ref |
| 9628 | MK962640 | Unassigned | Unassigned | Unassigned | 157_0 | Clustered | 3 | VC_157_0 | 3 | 0.3732 | 0.99808097 | 0.3725 | 0 | 0 | 0 | 1 | Ref |
| 9629 | MK962641 | Unassigned | Unassigned | Unassigned | 779_1 | Clustered | 11 | VC_779_1 | 11 | 0.1587 | 1 | 0.1587 | 0 | 0 | 0 | 1 | Ref |
| 9630 | MK962749 | Unassigned | Unassigned | Unassigned | 772_1 | Clustered | 2 | VC_772_1 | 2 | 0.006 | 0.57682361 | 0.0034 | 0 | 0 | 0 | 1 | Ref |
| 9631 | MK962750 | Unassigned | Unassigned | Unassigned | 283_169 | Clustered/Singleton | 1 | VC_283_169 | 1 | 0 | 1 | 0 | 0 | 0 | 0 | 1 | Ref |
| 9632 | MK962751 | Unassigned | Unassigned | Unassigned | 942_0 | Clustered | 36 | VC_942_0 | 36 | 0.7848 | 1 | 0.7848 | 0 | 0 | 0 | 1 | Ref |
| 9633 | MK962752 | Unassigned | Unassigned | Unassigned | 283_120 | Clustered/Singleton | 1 | VC_283_120 | 1 | 0 | 1 | 0 | 0 | 0 | 0 | 1 | Ref |
| 9634 | MK962753 | Unassigned | Unassigned | Unassigned | 590_0 | Clustered | 27 | VC_590_0 | 27 | 0.9947 | 1 | 0.9947 | 0 | 0 | 0 | 1 | Ref |
| 9635 | MK962754 | Unassigned | Unassigned | Unassigned | 286_1 | Clustered | 21 | VC_286_1 | 21 | 0.1696 | 1 | 0.1696 | 0 | 0 | 0 | 0.9048 | Ref |
| 9636 | MK962755 | Unassigned | Unassigned | Unassigned | 283_35 | Clustered/Singleton | 1 | VC_283_35 | 1 | 0 | 1 | 0 | 0 | 0 | 0 | 1 | Ref |
| 9637 | MK962756 | Unassigned | Unassigned | Unassigned | 286_1 | Clustered | 21 | VC_286_1 | 21 | 0.1696 | 1 | 0.1696 | 0 | 0 | 0 | 0.9048 | Ref |
| 9638 | MK962757 | Unassigned | Unassigned | Unassigned | 286_1 | Clustered | 21 | VC_286_1 | 21 | 0.1696 | 1 | 0.1696 | 0 | 0 | 0 | 0.9048 | Ref |
| 9639 | MK962758 | Unassigned | Unassigned | Unassigned | 329_3 | Clustered | 24 | VC_329_3 | 24 | 0.1206 | 1 | 0.1206 | 0 | 0 | 0 | 0.7283 | Ref |
| 9640 | MK965969 | Unassigned | Unassigned | Unassigned | 329_1 | Clustered | 17 | VC_329_1 | 17 | 0.0867 | 1 | 0.0867 | 0 | 0 | 0 | 0.8235 | Ref |
| 9641 | MK965970 | Unassigned | Unassigned | Unassigned | 329_1 | Clustered | 17 | VC_329_1 | 17 | 0.0867 | 1 | 0.0867 | 0 | 0 | 0 | 0.8235 | Ref |
| 9642 | MK967377 | Unassigned | Unassigned | Unassigned | 1177_0 | Clustered | 14 | VC_1177_0 | 14 | 0.7847 | 1 | 0.7847 | 0 | 0 | 0 | 0.989 | Ref |
| 9643 | MK967378 | Unassigned | Unassigned | Unassigned | 890_0 | Clustered | 3 | VC_890_0 | 3 | 0.3565 | 0.99830951 | 0.3559 | 0 | 0 | 0 | 1 | Ref |
| 9644 | MK967379 | Unassigned | Unassigned | Unassigned | 546_4 | Clustered | 16 | VC_546_4 | 16 | 0.2517 | 0.9999998 | 0.2517 | 0 | 0 | 0 | 0.9417 | Ref |
| 9645 | MK967381 | Unassigned | Unassigned | Unassigned | 1178_0 | Clustered | 24 | VC_1178_0 | 24 | 0.9524 | 1 | 0.9524 | 0 | 0 | 0 | 1 | Ref |
| 9646 | MK967382 | Unassigned | Unassigned | Unassigned | 224_0 | Clustered | 19 | VC_224_0 | 19 | 0.9416 | 1 | 0.9416 | 0 | 0 | 0 | 1 | Ref |
| 9647 | MK967383 | Unassigned | Unassigned | Unassigned | 919_0 | Clustered | 39 | VC_919_0 | 39 | 0.9069 | 1 | 0.9069 | 0 | 0 | 0 | 1 | Ref |
| 9648 | MK967384 | Unassigned | Unassigned | Unassigned | 525_1 | Clustered | 5 | VC_525_1 | 5 | 0.0485 | 0.99999947 | 0.0485 | 0 | 0 | 0 | 0.9 | Ref |
| 9649 | MK967385 | Unassigned | Unassigned | Unassigned | 919_0 | Clustered | 39 | VC_919_0 | 39 | 0.9069 | 1 | 0.9069 | 0 | 0 | 0 | 1 | Ref |
| 9650 | MK967386 | Unassigned | Unassigned | Unassigned | 872_0 | Clustered | 31 | VC_872_0 | 31 | 0.8151 | 1 | 0.8151 | 0 | 0 | 0 | 0.8194 | Ref |
| 9651 | MK967387 | Unassigned | Unassigned | Unassigned | 325_19 | Clustered | 3 | VC_325_19 | 3 | 0.0108 | 0.99859221 | 0.0107 | 0 | 0 | 0 | 1 | Ref |
| 9652 | MK967388 | Unassigned | Unassigned | Unassigned | 1240_0 | Clustered | 82 | VC_1240_0 | 82 | 0.7945 | 1 | 0.7945 | 0 | 0 | 0 | 1 | Ref |
| 9653 | MK967389 | Unassigned | Unassigned | Unassigned | 1176_0 | Clustered | 41 | VC_1176_0 | 41 | 0.9558 | 1 | 0.9558 | 0 | 0 | 0 | 0.9976 | Ref |
| 9654 | MK967390 | Unassigned | Unassigned | Unassigned | 868_0 | Clustered | 25 | VC_868_0 | 25 | 0.5791 | 1 | 0.5791 | 0 | 0 | 0 | 1 | Ref |
| 9655 | MK967391 | Unassigned | Unassigned | Unassigned | 1240_0 | Clustered | 82 | VC_1240_0 | 82 | 0.7945 | 1 | 0.7945 | 0 | 0 | 0 | 1 | Ref |
| 9656 | MK967392 | Unassigned | Unassigned | Unassigned | 954_0 | Clustered | 13 | VC_954_0 | 13 | 0.863 | 1 | 0.863 | 0 | 0 | 0 | 1 | Ref |
| 9657 | MK967393 | Unassigned | Unassigned | Unassigned | 953_0 | Clustered | 2 | VC_953_0 | 2 | 0.2116 | 0.94820857 | 0.2007 | 0 | 0 | 0 | 1 | Ref |
| 9658 | MK967394 | Unassigned | Unassigned | Unassigned | 1240_0 | Clustered | 82 | VC_1240_0 | 82 | 0.7945 | 1 | 0.7945 | 0 | 0 | 0 | 1 | Ref |
| 9659 | MK967395 | Unassigned | Unassigned | Unassigned | 216_4 | Clustered | 13 | VC_216_4 | 13 | 0.1657 | 1 | 0.1657 | 0 | 0 | 0 | 1 | Ref |
| 9660 | MK967396 | Unassigned | Unassigned | Unassigned | 1093_0 | Clustered | 40 | VC_1093_0 | 40 | 0.8754 | 1 | 0.8754 | 0 | 0 | 0 | 1 | Ref |
| 9661 | MK967397 | Unassigned | Unassigned | Unassigned | 520_94 | Clustered/Singleton | 1 | VC_520_94 | 1 | 0 | 1 | 0 | 0 | 0 | 0 | 1 | Ref |
| 9662 | MK967398 | Unassigned | Unassigned | Unassigned | 802_0 | Clustered | 17 | VC_802_0 | 17 | 0.8025 | 1 | 0.8025 | 0 | 0 | 0 | 1 | Ref |
| 9663 | MK967399 | Unassigned | Unassigned | Unassigned | 514_0 | Clustered | 75 | VC_514_0 | 75 | 0.9402 | 1 | 0.9402 | 0 | 0 | 0 | 1 | Ref |
| 9664 | MK967400 | Unassigned | Unassigned | Unassigned | 1240_0 | Clustered | 82 | VC_1240_0 | 82 | 0.7945 | 1 | 0.7945 | 0 | 0 | 0 | 1 | Ref |
| 9665 | MK967401 | Unassigned | Unassigned | Unassigned | 325_46 | Clustered | 2 | VC_325_46 | 2 | 0.004 | 0.95751617 | 0.0039 | 0 | 0 | 0 | 1 | Ref |
| 9666 | MK967402 | Unassigned | Unassigned | Unassigned | 546_4 | Clustered | 16 | VC_546_4 | 16 | 0.2517 | 0.9999998 | 0.2517 | 0 | 0 | 0 | 0.9417 | Ref |
| 9667 | MK972685 | Unassigned | Unassigned | Unassigned | 28_1 | Clustered | 53 | VC_28_1 | 53 | 0.7164 | 1 | 0.7164 | 0 | 0 | 0 | 0.9325 | Ref |
| 9668 | MK972686 | Unassigned | Unassigned | Unassigned | 28_1 | Clustered | 53 | VC_28_1 | 53 | 0.7164 | 1 | 0.7164 | 0 | 0 | 0 | 0.9325 | Ref |
| 9669 | MK972687 | Unassigned | Unassigned | Unassigned | 28_1 | Clustered | 53 | VC_28_1 | 53 | 0.7164 | 1 | 0.7164 | 0 | 0 | 0 | 0.9325 | Ref |
| 9670 | MK972690 | Unassigned | Unassigned | Unassigned | 133_21 | Clustered/Singleton | 1 | VC_133_21 | 1 | 0 | 1 | 0 | 0 | 0 | 0 | 1 | Ref |
| 9671 | MK972691 | Unassigned | Unassigned | Unassigned | 637_0 | Clustered | 115 | VC_637_0 | 115 | 0.731 | 1 | 0.731 | 0 | 0 | 0 | 0.8764 | Ref |
| 9672 | MK972692 | Unassigned | Unassigned | Unassigned | 28_1 | Clustered | 53 | VC_28_1 | 53 | 0.7164 | 1 | 0.7164 | 0 | 0 | 0 | 0.9325 | Ref |
| 9673 | MK972693 | Unassigned | Unassigned | Unassigned | 28_1 | Clustered | 53 | VC_28_1 | 53 | 0.7164 | 1 | 0.7164 | 0 | 0 | 0 | 0.9325 | Ref |
| 9674 | MK972694 | Unassigned | Unassigned | Unassigned | 28_1 | Clustered | 53 | VC_28_1 | 53 | 0.7164 | 1 | 0.7164 | 0 | 0 | 0 | 0.9325 | Ref |
| 9675 | MK972698 | Unassigned | Unassigned | Unassigned | 133_48 | Clustered | 2 | VC_133_48 | 2 | 0.0052 | 0.93639952 | 0.0048 | 0 | 0 | 0 | 1 | Ref |
| 9676 | MK972699 | Unassigned | Unassigned | Unassigned | 133_48 | Clustered | 2 | VC_133_48 | 2 | 0.0052 | 0.93639952 | 0.0048 | 0 | 0 | 0 | 1 | Ref |
| 9677 | MK972700 | Unassigned | Unassigned | Unassigned | 637_0 | Clustered | 115 | VC_637_0 | 115 | 0.731 | 1 | 0.731 | 0 | 0 | 0 | 0.8764 | Ref |
| 9678 | MK972701 | Unassigned | Unassigned | Unassigned | 637_0 | Clustered | 115 | VC_637_0 | 115 | 0.731 | 1 | 0.731 | 0 | 0 | 0 | 0.8764 | Ref |
| 9679 | MK972702 | Unassigned | Unassigned | Unassigned | 637_0 | Clustered | 115 | VC_637_0 | 115 | 0.731 | 1 | 0.731 | 0 | 0 | 0 | 0.8764 | Ref |
| 9680 | MK972703 | Unassigned | Unassigned | Unassigned | 637_0 | Clustered | 115 | VC_637_0 | 115 | 0.731 | 1 | 0.731 | 0 | 0 | 0 | 0.8764 | Ref |
| 9681 | MK972704 | Unassigned | Unassigned | Unassigned | 191_3 | Clustered | 5 | VC_191_3 | 5 | 0.0231 | 0.99999849 | 0.0231 | 0 | 0 | 0 | 0.8 | Ref |
| 9682 | MK972705 | Unassigned | Unassigned | Unassigned | 637_0 | Clustered | 115 | VC_637_0 | 115 | 0.731 | 1 | 0.731 | 0 | 0 | 0 | 0.8764 | Ref |
| 9683 | MK972706 | Unassigned | Unassigned | Unassigned | 637_0 | Clustered | 115 | VC_637_0 | 115 | 0.731 | 1 | 0.731 | 0 | 0 | 0 | 0.8764 | Ref |
| 9684 | MK972707 | Unassigned | Unassigned | Unassigned | 191_3 | Clustered | 5 | VC_191_3 | 5 | 0.0231 | 0.99999849 | 0.0231 | 0 | 0 | 0 | 0.8 | Ref |
| 9685 | MK972708 | Unassigned | Unassigned | Unassigned | 191_31 | Clustered | 2 | VC_191_31 | 2 | 0.0057 | 0.92424379 | 0.0052 | 0 | 0 | 0 | 1 | Ref |
| 9686 | MK972709 | Unassigned | Unassigned | Unassigned | 191_3 | Clustered | 5 | VC_191_3 | 5 | 0.0231 | 0.99999849 | 0.0231 | 0 | 0 | 0 | 0.8 | Ref |
| 9687 | MK972710 | Unassigned | Unassigned | Unassigned | 228_0 | Clustered | 60 | VC_228_0 | 60 | 0.942 | 1 | 0.942 | 0 | 0 | 0 | 1 | Ref |
| 9688 | MK972711 | Unassigned | Unassigned | Unassigned | 228_0 | Clustered | 60 | VC_228_0 | 60 | 0.942 | 1 | 0.942 | 0 | 0 | 0 | 1 | Ref |
| 9689 | MK972712 | Unassigned | Unassigned | Unassigned | 5_0 | Clustered | 30 | VC_5_0 | 30 | 0.9647 | 1 | 0.9647 | 0 | 0 | 0 | 1 | Ref |
| 9690 | MK972713 | Unassigned | Unassigned | Unassigned | 5_0 | Clustered | 30 | VC_5_0 | 30 | 0.9647 | 1 | 0.9647 | 0 | 0 | 0 | 1 | Ref |
| 9691 | MK972714 | Unassigned | Unassigned | Unassigned | 5_0 | Clustered | 30 | VC_5_0 | 30 | 0.9647 | 1 | 0.9647 | 0 | 0 | 0 | 1 | Ref |
| 9692 | MK972715 | Unassigned | Unassigned | Unassigned | 190_20 | Clustered | 4 | VC_190_20 | 4 | 0.0229 | 0.99997885 | 0.0229 | 0 | 0 | 0 | 1 | Ref |
| 9693 | MK972716 | Unassigned | Unassigned | Unassigned | 190_20 | Clustered | 4 | VC_190_20 | 4 | 0.0229 | 0.99997885 | 0.0229 | 0 | 0 | 0 | 1 | Ref |
| 9694 | MK972717 | Unassigned | Unassigned | Unassigned | 191_16 | Clustered | 12 | VC_191_16 | 12 | 0.0643 | 1 | 0.0643 | 0 | 0 | 0 | 0.9394 | Ref |
| 9695 | MK972831 | Unassigned | Unassigned | Unassigned | 575_0 | Clustered | 35 | VC_575_0 | 35 | 0.8815 | 1 | 0.8815 | 0 | 0 | 0 | 1 | Ref |
| 9696 | MK976036 | Unassigned | Unassigned | Unassigned | 22_0 | Clustered | 11 | VC_22_0 | 11 | 0.5163 | 1 | 0.5163 | 0 | 0 | 0 | 1 | Ref |
| 9697 | MK977694 | Unassigned | Unassigned | Unassigned | 283_190 | Clustered/Singleton | 1 | VC_283_190 | 1 | 0 | 1 | 0 | 0 | 0 | 0 | 1 | Ref |
| 9698 | MK977697 | Unassigned | Unassigned | Unassigned | 1240_0 | Clustered | 82 | VC_1240_0 | 82 | 0.7945 | 1 | 0.7945 | 0 | 0 | 0 | 1 | Ref |
| 9699 | MK977698 | Unassigned | Unassigned | Unassigned | 1352_0 | Clustered | 31 | VC_1352_0 | 31 | 0.849 | 1 | 0.849 | 0 | 0 | 0 | 1 | Ref |
| 9700 | MK977699 | Unassigned | Unassigned | Unassigned | 767_0 | Clustered | 34 | VC_767_0 | 34 | 0.9173 | 1 | 0.9173 | 0 | 0 | 0 | 1 | Ref |
| 9701 | MK977700 | Unassigned | Unassigned | Unassigned | 768_0 | Clustered | 8 | VC_768_0 | 8 | 0.7012 | 1 | 0.7012 | 0 | 0 | 0 | 1 | Ref |
| 9702 | MK977701 | Unassigned | Unassigned | Unassigned | 686_1 | Clustered | 2 | VC_686_1 | 2 | 0.2204 | 0.91327915 | 0.2013 | 0 | 0 | 0 | 1 | Ref |
| 9703 | MK977702 | Unassigned | Unassigned | Unassigned | 1093_0 | Clustered | 40 | VC_1093_0 | 40 | 0.8754 | 1 | 0.8754 | 0 | 0 | 0 | 1 | Ref |
| 9704 | MK977705 | Unassigned | Unassigned | Unassigned | 1182_0 | Clustered | 4 | VC_1182_0 | 4 | 0.2845 | 0.99997676 | 0.2845 | 0 | 0 | 0 | 1 | Ref |
| 9705 | MK977706 | Unassigned | Unassigned | Unassigned | 1199_0 | Clustered | 8 | VC_1199_0 | 8 | 0.7928 | 1 | 0.7928 | 0 | 0 | 0 | 1 | Ref |
| 9706 | MK977708 | Unassigned | Unassigned | Unassigned | 521_0 | Clustered | 37 | VC_521_0 | 37 | 0.912 | 1 | 0.912 | 0 | 0 | 0 | 1 | Ref |
| 9707 | MK977709 | Unassigned | Unassigned | Unassigned | 523_0 | Clustered/Singleton | 1 | VC_523_0 | 1 | 0 | 1 | 0 | 0 | 0 | 0 | 1 | Ref |
| 9708 | MK977710 | Unassigned | Unassigned | Unassigned | 1199_0 | Clustered | 8 | VC_1199_0 | 8 | 0.7928 | 1 | 0.7928 | 0 | 0 | 0 | 1 | Ref |
| 9709 | MK977711 | Unassigned | Unassigned | Unassigned | 1071_3 | Clustered | 16 | VC_1071_3 | 16 | 0.2354 | 1 | 0.2354 | 0 | 0 | 0 | 0.9667 | Ref |
| 9710 | MK977712 | Unassigned | Unassigned | Unassigned | 1199_0 | Clustered | 8 | VC_1199_0 | 8 | 0.7928 | 1 | 0.7928 | 0 | 0 | 0 | 1 | Ref |
| 9711 | MK977713 | Unassigned | Unassigned | Unassigned | 225_0 | Clustered | 32 | VC_225_0 | 32 | 0.848 | 1 | 0.848 | 0 | 0 | 0 | 0.996 | Ref |
| 9712 | MK977714 | Unassigned | Unassigned | Unassigned | 1199_0 | Clustered | 8 | VC_1199_0 | 8 | 0.7928 | 1 | 0.7928 | 0 | 0 | 0 | 1 | Ref |
| 9713 | MK977715 | Unassigned | Unassigned | Unassigned | 1199_0 | Clustered | 8 | VC_1199_0 | 8 | 0.7928 | 1 | 0.7928 | 0 | 0 | 0 | 1 | Ref |
| 9714 | MK982307 | Unassigned | Unassigned | Unassigned | 695_0 | Clustered | 48 | VC_695_0 | 48 | 0.8162 | 1 | 0.8162 | 0 | 0 | 0 | 1 | Ref |
| 9715 | MK994515 | Unassigned | Unassigned | Unassigned | 488_1 | Clustered | 8 | VC_488_1 | 8 | 0.2927 | 1 | 0.2927 | 0 | 0 | 0 | 1 | Ref |
| 9716 | MN010755 | Unassigned | Unassigned | Unassigned | 1344_0 | Clustered | 78 | VC_1344_0 | 78 | 0.9732 | 1 | 0.9732 | 0 | 0 | 0 | 1 | Ref |
| 9717 | MN010756 | Unassigned | Unassigned | Unassigned | 567_0 | Clustered | 8 | VC_567_0 | 8 | 0.4935 | 1 | 0.4935 | 0 | 0 | 0 | 1 | Ref |
| 9718 | MN010757 | Unassigned | Unassigned | Unassigned | 519_0 | Clustered | 39 | VC_519_0 | 39 | 0.8156 | 1 | 0.8156 | 0 | 0 | 0 | 1 | Ref |

| | | | | | | | | | | | | | | | | | |
|---|---|---|---|---|---|---|---|---|---|---|---|---|---|---|---|---|---|
| 9719 | MN010758 | Unassigned | Unassigned | Unassigned | 406_0 | Clustered | 11 | VC_406_0 | 11 | 0.9449 | 1 | 0.9449 | 0 | 0 | 0 | 1 | Ref |
| 9720 | MN010759 | Unassigned | Unassigned | Unassigned | 1344_0 | Clustered | 78 | VC_1344_0 | 78 | 0.9732 | 1 | 0.9732 | 0 | 0 | 0 | 1 | Ref |
| 9721 | MN010760 | Unassigned | Unassigned | Unassigned | 1178_0 | Clustered | 24 | VC_1178_0 | 24 | 0.9524 | 1 | 0.9524 | 0 | 0 | 0 | 1 | Ref |
| 9722 | MN010761 | Unassigned | Unassigned | Unassigned | 799_1 | Clustered | 27 | VC_799_1 | 27 | 0.8611 | 1 | 0.8611 | 0 | 0 | 0 | 1 | Ref |
| 9723 | MN010762 | Unassigned | Unassigned | Unassigned | 1176_0 | Clustered | 41 | VC_1176_0 | 41 | 0.9558 | 1 | 0.9558 | 0 | 0 | 0 | 0.9976 | Ref |
| 9724 | MN013074 | Unassigned | Unassigned | Unassigned | 385_0 | Clustered | 51 | VC_385_0 | 51 | 0.8754 | 1 | 0.8754 | 0 | 0 | 0 | 1 | Ref |
| 9725 | MN013075 | Unassigned | Unassigned | Unassigned | 937_0 | Clustered | 40 | VC_937_0 | 40 | 0.6809 | 1 | 0.6809 | 0 | 0 | 0 | 0.9359 | Ref |
| 9726 | MN013076 | Unassigned | Unassigned | Unassigned | 937_0 | Clustered | 40 | VC_937_0 | 40 | 0.6809 | 1 | 0.6809 | 0 | 0 | 0 | 0.9359 | Ref |
| 9727 | MN013077 | Unassigned | Unassigned | Unassigned | 849_6 | Clustered | 7 | VC_849_6 | 7 | 0.164 | 0.99999999 | 0.164 | 0 | 0 | 0 | 0.9524 | Ref |
| 9728 | MN013078 | Unassigned | Unassigned | Unassigned | 937_0 | Clustered | 40 | VC_937_0 | 40 | 0.6809 | 1 | 0.6809 | 0 | 0 | 0 | 0.9359 | Ref |
| 9729 | MN013079 | Unassigned | Unassigned | Unassigned | 937_0 | Clustered | 40 | VC_937_0 | 40 | 0.6809 | 1 | 0.6809 | 0 | 0 | 0 | 0.9359 | Ref |
| 9730 | MN013080 | Unassigned | Unassigned | Unassigned | 937_0 | Clustered | 40 | VC_937_0 | 40 | 0.6809 | 1 | 0.6809 | 0 | 0 | 0 | 0.9359 | Ref |
| 9731 | MN013082 | Unassigned | Unassigned | Unassigned | 393_0 | Clustered | 7 | VC_393_0 | 7 | 0.1775 | 1 | 0.1775 | 0 | 0 | 0 | 1 | Ref |
| 9732 | MN013083 | Unassigned | Unassigned | Unassigned | 937_0 | Clustered | 40 | VC_937_0 | 40 | 0.6809 | 1 | 0.6809 | 0 | 0 | 0 | 0.9359 | Ref |
| 9733 | MN013085 | Unassigned | Unassigned | Unassigned | 393_0 | Clustered | 7 | VC_393_0 | 7 | 0.1775 | 1 | 0.1775 | 0 | 0 | 0 | 1 | Ref |
| 9734 | MN013086 | Unassigned | Unassigned | Unassigned | 37_1 | Clustered | 3 | VC_37_1 | 3 | 0.0833 | 0.9484635 | 0.079 | 0 | 0 | 0 | 1 | Ref |
| 9735 | MN013087 | Unassigned | Unassigned | Unassigned | 937_0 | Clustered | 40 | VC_937_0 | 40 | 0.6809 | 1 | 0.6809 | 0 | 0 | 0 | 0.9359 | Ref |
| 9736 | MN013088 | Unassigned | Unassigned | Unassigned | 104_0 | Clustered | 6 | VC_104_0 | 6 | 0.8291 | 1 | 0.8291 | 0 | 0 | 0 | 1 | Ref |
| 9737 | MN013089 | Unassigned | Unassigned | Unassigned | 187_1 | Clustered | 7 | VC_187_1 | 7 | 0.1423 | 1 | 0.1423 | 0 | 0 | 0 | 1 | Ref |
| 9738 | MN013356 | Unassigned | Unassigned | Unassigned | 76_0 | Clustered | 37 | VC_76_0 | 37 | 0.9403 | 1 | 0.9403 | 0 | 0 | 0 | 1 | Ref |
| 9739 | MN016939 | Unassigned | Unassigned | Unassigned | 228_0 | Clustered | 60 | VC_228_0 | 60 | 0.942 | 1 | 0.942 | 0 | 0 | 0 | 1 | Ref |
| 9740 | MN018232 | Unassigned | Unassigned | Unassigned | 432_14 | Clustered/Singleton | 1 | VC_432_14 | 1 | 0 | 1 | 0 | 0 | 0 | 0 | 1 | Ref |
| 9741 | MN019128 | Unassigned | Unassigned | Unassigned | 585_0 | Clustered | 4 | VC_585_0 | 4 | 0.0533 | 0.99998755 | 0.0533 | 0 | 0 | 0 | 1 | Ref |
| 9742 | MN022785 | Unassigned | Unassigned | Unassigned | 283_130 | Clustered/Singleton | 1 | VC_283_130 | 1 | 0 | 1 | 0 | 0 | 0 | 0 | 1 | Ref |
| 9743 | MN022787 | Unassigned | Unassigned | Unassigned | 190_14 | Clustered | 3 | VC_190_14 | 3 | 0.0144 | 0.99696852 | 0.0143 | 0 | 0 | 0 | 0.6667 | Ref |
| 9744 | MN023176 | Unassigned | Unassigned | Unassigned | 860_0 | Clustered | 16 | VC_860_0 | 16 | 0.9964 | 1 | 0.9964 | 0 | 0 | 0 | 1 | Ref |
| 9745 | MN023177 | Unassigned | Unassigned | Unassigned | 860_0 | Clustered | 16 | VC_860_0 | 16 | 0.9964 | 1 | 0.9964 | 0 | 0 | 0 | 1 | Ref |
| 9746 | MN023178 | Unassigned | Unassigned | Unassigned | 860_0 | Clustered | 16 | VC_860_0 | 16 | 0.9964 | 1 | 0.9964 | 0 | 0 | 0 | 1 | Ref |
| 9747 | MN023179 | Unassigned | Unassigned | Unassigned | 860_0 | Clustered | 16 | VC_860_0 | 16 | 0.9964 | 1 | 0.9964 | 0 | 0 | 0 | 1 | Ref |
| 9748 | MN023180 | Unassigned | Unassigned | Unassigned | 860_0 | Clustered | 16 | VC_860_0 | 16 | 0.9964 | 1 | 0.9964 | 0 | 0 | 0 | 1 | Ref |
| 9749 | MN023181 | Unassigned | Unassigned | Unassigned | 860_0 | Clustered | 16 | VC_860_0 | 16 | 0.9964 | 1 | 0.9964 | 0 | 0 | 0 | 1 | Ref |
| 9750 | MN023182 | Unassigned | Unassigned | Unassigned | 860_0 | Clustered | 16 | VC_860_0 | 16 | 0.9964 | 1 | 0.9964 | 0 | 0 | 0 | 1 | Ref |
| 9751 | MN023183 | Unassigned | Unassigned | Unassigned | 860_0 | Clustered | 16 | VC_860_0 | 16 | 0.9964 | 1 | 0.9964 | 0 | 0 | 0 | 1 | Ref |
| 9752 | MN023184 | Unassigned | Unassigned | Unassigned | 860_0 | Clustered | 16 | VC_860_0 | 16 | 0.9964 | 1 | 0.9964 | 0 | 0 | 0 | 1 | Ref |
| 9753 | MN023185 | Unassigned | Unassigned | Unassigned | 860_0 | Clustered | 16 | VC_860_0 | 16 | 0.9964 | 1 | 0.9964 | 0 | 0 | 0 | 1 | Ref |
| 9754 | MN023186 | Unassigned | Unassigned | Unassigned | 860_0 | Clustered | 16 | VC_860_0 | 16 | 0.9964 | 1 | 0.9964 | 0 | 0 | 0 | 1 | Ref |
| 9755 | MN023187 | Unassigned | Unassigned | Unassigned | 860_0 | Clustered | 16 | VC_860_0 | 16 | 0.9964 | 1 | 0.9964 | 0 | 0 | 0 | 1 | Ref |
| 9756 | MN023188 | Unassigned | Unassigned | Unassigned | 860_0 | Clustered | 16 | VC_860_0 | 16 | 0.9964 | 1 | 0.9964 | 0 | 0 | 0 | 1 | Ref |
| 9757 | MN023189 | Unassigned | Unassigned | Unassigned | 860_0 | Clustered | 16 | VC_860_0 | 16 | 0.9964 | 1 | 0.9964 | 0 | 0 | 0 | 1 | Ref |
| 9758 | MN023190 | Unassigned | Unassigned | Unassigned | 860_0 | Clustered | 16 | VC_860_0 | 16 | 0.9964 | 1 | 0.9964 | 0 | 0 | 0 | 1 | Ref |
| 9759 | MN023191 | Unassigned | Unassigned | Unassigned | 860_0 | Clustered | 16 | VC_860_0 | 16 | 0.9964 | 1 | 0.9964 | 0 | 0 | 0 | 1 | Ref |
| 9760 | MN026740 | Unassigned | Unassigned | Unassigned | 392_0 | Clustered | 126 | VC_392_0 | 126 | 0.9621 | 1 | 0.9621 | 0 | 0 | 0 | 1 | Ref |
| 9761 | MN026741 | Unassigned | Unassigned | Unassigned | 190_23 | Clustered | 10 | VC_190_23 | 10 | 0.0704 | 1 | 0.0704 | 0 | 0 | 0 | 0.8444 | Ref |
| 9762 | MN027503 | Unassigned | Unassigned | Unassigned | 92_0 | Clustered | 22 | VC_92_0 | 22 | 0.5639 | 1 | 0.5639 | 0 | 0 | 0 | 1 | Ref |
| 9763 | MN028536 | Unassigned | Unassigned | Unassigned | 11_0 | Clustered | 19 | VC_11_0 | 19 | 0.7562 | 1 | 0.7562 | 0 | 0 | 0 | 1 | Ref |
| 9764 | MN029011 | Unassigned | Unassigned | Unassigned | 796_0 | Clustered | 5 | VC_796_0 | 5 | 0.3352 | 0.99999997 | 0.3352 | 0 | 0 | 0 | 1 | Ref |
| 9765 | MN032614 | Unassigned | Unassigned | Unassigned | 442_3 | Clustered | 3 | VC_442_3 | 3 | 0.0529 | 0.99675181 | 0.0527 | 0 | 0 | 0 | 1 | Ref |
| 9766 | MN038175 | Unassigned | Unassigned | Unassigned | 669_3 | Clustered/Singleton | 1 | VC_669_3 | 1 | 0 | 1 | 0 | 0 | 0 | 0 | 1 | Ref |
| 9767 | MN038176 | Unassigned | Unassigned | Unassigned | 884_0 | Clustered | 28 | VC_884_0 | 28 | 0.6211 | 1 | 0.6211 | 0 | 0 | 0 | 0.9841 | Ref |
| 9768 | MN038177 | Unassigned | Unassigned | Unassigned | 618_0 | Clustered | 13 | VC_618_0 | 13 | 0.6252 | 1 | 0.6252 | 0 | 0 | 0 | 1 | Ref |
| 9769 | MN038178 | Unassigned | Unassigned | Unassigned | 883_1 | Clustered | 12 | VC_883_1 | 12 | 0.2441 | 1 | 0.2441 | 0 | 0 | 0 | 1 | Ref |
| 9770 | MN038179 | Unassigned | Unassigned | Unassigned | 884_0 | Clustered | 28 | VC_884_0 | 28 | 0.6211 | 1 | 0.6211 | 0 | 0 | 0 | 0.9841 | Ref |
| 9771 | MN043729 | Unassigned | Unassigned | Unassigned | 85_10 | Clustered/Singleton | 1 | VC_85_10 | 1 | 0 | 1 | 0 | 0 | 0 | 0 | 1 | Ref |
| 9772 | MN043730 | Unassigned | Unassigned | Unassigned | 85_4 | Clustered | 3 | VC_85_4 | 3 | 0.0294 | 0.98125255 | 0.0288 | 0 | 0 | 0 | 1 | Ref |
| 9773 | MN045228 | Unassigned | Unassigned | Unassigned | 149_12 | Clustered | 2 | VC_149_12 | 2 | 0.0044 | 0.83156708 | 0.0037 | 0 | 0 | 0 | 1 | Ref |
| 9774 | MN045229 | Unassigned | Unassigned | Unassigned | 792_0 | Clustered | 35 | VC_792_0 | 35 | 0.9828 | 1 | 0.9828 | 0 | 0 | 0 | 1 | Ref |
| 9775 | MN045230 | Unassigned | Unassigned | Unassigned | 132_1 | Clustered | 17 | VC_132_1 | 17 | 0.1738 | 1 | 0.1738 | 0 | 0 | 0 | 0.9853 | Ref |
| 9776 | MN045231 | Unassigned | Unassigned | Unassigned | 813_1 | Clustered | 24 | VC_813_1 | 24 | 0.535 | 1 | 0.535 | 0 | 0 | 0 | 1 | Ref |
| 9777 | MN047438 | Unassigned | Unassigned | Unassigned | 149_8 | Clustered | 66 | VC_149_8 | 66 | 0.4565 | 1 | 0.4565 | 0 | 0 | 0 | 0.8182 | Ref |
| 9778 | MN047793 | Unassigned | Unassigned | Unassigned | 68_1 | Clustered/Singleton | 1 | VC_68_1 | 1 | 0 | 1 | 0 | 0 | 0 | 0 | 1 | Ref |
| 9779 | MN052874 | Unassigned | Unassigned | Unassigned | 823_0 | Clustered | 124 | VC_823_0 | 124 | 0.9492 | 1 | 0.9492 | 0 | 0 | 0 | 1 | Ref |
| 9780 | MN055691 | Unassigned | Unassigned | Unassigned | 941_0 | Clustered | 40 | VC_941_0 | 40 | 0.6903 | 1 | 0.6903 | 0 | 0 | 0 | 1 | Ref |
| 9781 | MN061582 | Unassigned | Unassigned | Unassigned | 699_0 | Clustered | 71 | VC_699_0 | 71 | 0.9352 | 1 | 0.9352 | 0 | 0 | 0 | 1 | Ref |
| 9782 | MN062185 | Unassigned | Unassigned | Unassigned | 530_0 | Clustered | 9 | VC_530_0 | 9 | 0.8027 | 1 | 0.8027 | 0 | 0 | 0 | 1 | Ref |
| 9783 | MN062186 | Unassigned | Unassigned | Unassigned | 779_3 | Clustered | 7 | VC_779_3 | 7 | 0.0766 | 1 | 0.0766 | 0 | 0 | 0 | 0.7143 | Ref |
| 9784 | MN062187 | Unassigned | Unassigned | Unassigned | 749_0 | Clustered | 40 | VC_749_0 | 40 | 0.7517 | 1 | 0.7517 | 0 | 0 | 0 | 1 | Ref |
| 9785 | MN062188 | Unassigned | Unassigned | Unassigned | 591_0 | Clustered | 14 | VC_591_0 | 14 | 0.5184 | 1 | 0.5184 | 0 | 0 | 0 | 1 | Ref |
| 9786 | MN062701 | Unassigned | Unassigned | Unassigned | 546_4 | Clustered | 16 | VC_546_4 | 16 | 0.2517 | 0.9999998 | 0.2517 | 0 | 0 | 0 | 0.9417 | Ref |
| 9787 | MN062702 | Unassigned | Unassigned | Unassigned | 1240_0 | Clustered | 82 | VC_1240_0 | 82 | 0.7945 | 1 | 0.7945 | 0 | 0 | 0 | 1 | Ref |
| 9788 | MN062704 | Unassigned | Unassigned | Unassigned | 1176_0 | Clustered | 41 | VC_1176_0 | 41 | 0.9558 | 1 | 0.9558 | 0 | 0 | 0 | 0.9976 | Ref |
| 9789 | MN062706 | Unassigned | Unassigned | Unassigned | 513_0 | Clustered | 32 | VC_513_0 | 32 | 0.6881 | 1 | 0.6881 | 0 | 0 | 0 | 1 | Ref |
| 9790 | MN062707 | Unassigned | Unassigned | Unassigned | 558_77 | Clustered | 3 | VC_558_77 | 3 | 0.0065 | 0.90877939 | 0.0059 | 0 | 0 | 0 | 0.6667 | Ref |
| 9791 | MN062708 | Unassigned | Unassigned | Unassigned | 217_1 | Clustered/Singleton | 1 | VC_217_1 | 1 | 0 | 1 | 0 | 0 | 0 | 0 | 1 | Ref |
| 9792 | MN062709 | Unassigned | Unassigned | Unassigned | 1240_0 | Clustered | 82 | VC_1240_0 | 82 | 0.7945 | 1 | 0.7945 | 0 | 0 | 0 | 1 | Ref |
| 9793 | MN062710 | Unassigned | Unassigned | Unassigned | 325_52 | Clustered | 5 | VC_325_52 | 5 | 0.0147 | 0.99999073 | 0.0147 | 0 | 0 | 0 | 0.7 | Ref |
| 9794 | MN062711 | Unassigned | Unassigned | Unassigned | 1361_0 | Clustered | 6 | VC_1361_0 | 6 | 0.2371 | 1 | 0.2371 | 0 | 0 | 0 | 1 | Ref |
| 9795 | MN062712 | Unassigned | Unassigned | Unassigned | 513_0 | Clustered | 32 | VC_513_0 | 32 | 0.6881 | 1 | 0.6881 | 0 | 0 | 0 | 1 | Ref |
| 9796 | MN062713 | Unassigned | Unassigned | Unassigned | 1240_0 | Clustered | 82 | VC_1240_0 | 82 | 0.7945 | 1 | 0.7945 | 0 | 0 | 0 | 1 | Ref |
| 9797 | MN062714 | Unassigned | Unassigned | Unassigned | 1234_0 | Clustered | 44 | VC_1234_0 | 44 | 1 | 1 | 1 | 0 | 0 | 0 | 1 | Ref |
| 9798 | MN062715 | Unassigned | Unassigned | Unassigned | 685_0 | Clustered | 16 | VC_685_0 | 16 | 0.2972 | 1 | 0.2972 | 0 | 0 | 0 | 0.9167 | Ref |
| 9799 | MN062716 | Unassigned | Unassigned | Unassigned | 919_0 | Clustered | 39 | VC_919_0 | 39 | 0.9069 | 1 | 0.9069 | 0 | 0 | 0 | 1 | Ref |
| 9800 | MN062717 | Unassigned | Unassigned | Unassigned | 1234_0 | Clustered | 44 | VC_1234_0 | 44 | 1 | 1 | 1 | 0 | 0 | 0 | 1 | Ref |
| 9801 | MN062718 | Unassigned | Unassigned | Unassigned | 514_0 | Clustered | 75 | VC_514_0 | 75 | 0.9402 | 1 | 0.9402 | 0 | 0 | 0 | 1 | Ref |
| 9802 | MN062719 | Unassigned | Unassigned | Unassigned | 1352_0 | Clustered | 31 | VC_1352_0 | 31 | 0.849 | 1 | 0.849 | 0 | 0 | 0 | 1 | Ref |
| 9803 | MN065183 | Unassigned | Unassigned | Unassigned | 20_0 | Clustered | 5 | VC_20_0 | 5 | 0.4387 | 0.99999934 | 0.4387 | 0 | 0 | 0 | 1 | Ref |
| 9804 | MN066127 | Unassigned | Unassigned | Unassigned | 133_22 | Clustered | 13 | VC_133_22 | 13 | 0.0804 | 1 | 0.0804 | 0 | 0 | 0 | 0.8205 | Ref |
| 9805 | MN067333 | Unassigned | Unassigned | Unassigned | 42_0 | Clustered | 49 | VC_42_0 | 49 | 0.4517 | 1 | 0.4517 | 0 | 0 | 0 | 0.8597 | Ref |
| 9806 | MN067430 | Unassigned | Unassigned | Unassigned | 451_0 | Clustered | 80 | VC_451_0 | 80 | 0.9991 | 1 | 0.9991 | 0 | 0 | 0 | 1 | Ref |
| 9807 | MN070121 | Unassigned | Unassigned | Unassigned | 540_0 | Clustered | 15 | VC_540_0 | 15 | 0.2866 | 1 | 0.2866 | 0 | 0 | 0 | 1 | Ref |
| 9808 | MN078104 | Unassigned | Unassigned | Unassigned | 830_0 | Clustered | 6 | VC_830_0 | 6 | 0.6536 | 1 | 0.6536 | 0 | 0 | 0 | 1 | Ref |
| 9809 | MN080711 | Unassigned | Unassigned | Unassigned | 386_0 | Clustered | 49 | VC_386_0 | 49 | 0.7941 | 1 | 0.7941 | 0 | 0 | 0 | 1 | Ref |
| 9810 | MN082624 | Unassigned | Unassigned | Unassigned | 818_0 | Clustered | 20 | VC_818_0 | 20 | 0.9601 | 1 | 0.9601 | 0 | 0 | 0 | 1 | Ref |
| 9811 | MN087708 | Unassigned | Unassigned | Unassigned | 490_1 | Clustered | 5 | VC_490_1 | 5 | 0.0447 | 0.99999996 | 0.0447 | 0 | 0 | 0 | 1 | Ref |
| 9812 | MN090155 | Unassigned | Unassigned | Unassigned | 585_1 | Clustered | 3 | VC_585_1 | 3 | 0.0313 | 0.99857668 | 0.0312 | 0 | 0 | 0 | 1 | Ref |
| 9813 | MN091625 | Unassigned | Unassigned | Unassigned | 149_5 | Clustered/Singleton | 1 | VC_149_5 | 1 | 0 | 1 | 0 | 0 | 0 | 0 | 1 | Ref |
| 9814 | MN091626 | Unassigned | Unassigned | Unassigned | 449_0 | Clustered | 7 | VC_449_0 | 7 | 0.6015 | 1 | 0.6015 | 0 | 0 | 0 | 1 | Ref |
| 9815 | MN094788 | Unassigned | Unassigned | Unassigned | 66_1 | Clustered/Singleton | 1 | VC_66_1 | 1 | 0 | 1 | 0 | 0 | 0 | 0 | 1 | Ref |
| 9816 | MN095770 | Unassigned | Unassigned | Unassigned | 196_0 | Clustered | 2 | VC_196_0 | 2 | 0.0218 | 0.95762696 | 0.0209 | 0 | 0 | 0 | 1 | Ref |
| 9817 | MN095771 | Unassigned | Unassigned | Unassigned | 284_0 | Clustered | 3 | VC_284_0 | 3 | 0.0495 | 0.99860966 | 0.0494 | 0 | 0 | 0 | 1 | Ref |
| 9818 | MN095772 | Unassigned | Unassigned | Unassigned | 506_0 | Clustered | 8 | VC_506_0 | 8 | 0.7376 | 1 | 0.7376 | 0 | 0 | 0 | 1 | Ref |
| 9819 | MN096356 | Unassigned | Unassigned | Unassigned | 519_0 | Clustered | 39 | VC_519_0 | 39 | 0.8156 | 1 | 0.8156 | 0 | 0 | 0 | 1 | Ref |
| 9820 | MN096357 | Unassigned | Unassigned | Unassigned | 1031_0 | Clustered | 4 | VC_1031_0 | 4 | 0.7884 | 0.99997621 | 0.7884 | 0 | 0 | 0 | 1 | Ref |
| 9821 | MN096358 | Unassigned | Unassigned | Unassigned | 1031_0 | Clustered | 4 | VC_1031_0 | 4 | 0.7884 | 0.99997621 | 0.7884 | 0 | 0 | 0 | 1 | Ref |
| 9822 | MN096359 | Unassigned | Unassigned | Unassigned | 509_0 | Clustered | 37 | VC_509_0 | 37 | 0.9689 | 1 | 0.9689 | 0 | 0 | 0 | 1 | Ref |
| 9823 | MN096360 | Unassigned | Unassigned | Unassigned | 509_0 | Clustered | 37 | VC_509_0 | 37 | 0.9689 | 1 | 0.9689 | 0 | 0 | 0 | 1 | Ref |
| 9824 | MN096361 | Unassigned | Unassigned | Unassigned | 551_0 | Clustered | 16 | VC_551_0 | 16 | 0.0915 | 1 | 0.0915 | 0 | 0 | 0 | 0.6667 | Ref |
| 9825 | MN096363 | Unassigned | Unassigned | Unassigned | 870_0 | Clustered | 5 | VC_870_0 | 5 | 0.424 | 0.99999997 | 0.424 | 0 | 0 | 0 | 1 | Ref |
| 9826 | MN096364 | Unassigned | Unassigned | Unassigned | 551_8 | Clustered | 19 | VC_551_8 | 19 | 0.1039 | 1 | 0.1039 | 0 | 0 | 0 | 0.8713 | Ref |
| 9827 | MN096365 | Unassigned | Unassigned | Unassigned | 1176_0 | Clustered | 41 | VC_1176_0 | 41 | 0.9558 | 1 | 0.9558 | 0 | 0 | 0 | 0.9976 | Ref |
| 9828 | MN096366 | Unassigned | Unassigned | Unassigned | 371_2 | Clustered | 8 | VC_371_2 | 8 | 0.0232 | 1 | 0.0232 | 0 | 0 | 0 | 0.8571 | Ref |
| 9829 | MN096367 | Unassigned | Unassigned | Unassigned | 1163_0 | Clustered | 28 | VC_1163_0 | 28 | 0.4923 | 1 | 0.4923 | 0 | 0 | 0 | 1 | Ref |
| 9830 | MN096368 | Unassigned | Unassigned | Unassigned | 514_0 | Clustered | 75 | VC_514_0 | 75 | 0.9402 | 1 | 0.9402 | 0 | 0 | 0 | 1 | Ref |
| 9831 | MN096370 | Unassigned | Unassigned | Unassigned | 557_0 | Clustered | 3 | VC_557_0 | 3 | 0.0814 | 0.99853279 | 0.0812 | 0 | 0 | 0 | 0.6667 | Ref |
| 9832 | MN096371 | Unassigned | Unassigned | Unassigned | 1071_3 | Clustered | 16 | VC_1071_3 | 16 | 0.2354 | 1 | 0.2354 | 0 | 0 | 0 | 0.9667 | Ref |
| 9833 | MN096372 | Unassigned | Unassigned | Unassigned | 940_0 | Clustered | 34 | VC_940_0 | 34 | 0.7959 | 1 | 0.7959 | 0 | 0 | 0 | 1 | Ref |
| 9834 | MN096373 | Unassigned | Unassigned | Unassigned | 982_0 | Clustered | 62 | VC_982_0 | 62 | 0.8646 | 1 | 0.8646 | 0 | 0 | 0 | 0.743 | Ref |
| 9835 | MN096374 | Unassigned | Unassigned | Unassigned | 856_1 | Clustered | 13 | VC_856_1 | 13 | 0.5848 | 1 | 0.5848 | 0 | 0 | 0 | 1 | Ref |
| 9836 | MN096375 | Unassigned | Unassigned | Unassigned | 982_0 | Clustered | 62 | VC_982_0 | 62 | 0.8646 | 1 | 0.8646 | 0 | 0 | 0 | 0.743 | Ref |
| 9837 | MN096376 | Unassigned | Unassigned | Unassigned | 218_0 | Clustered | 3 | VC_218_0 | 3 | 0.0481 | 0.99860979 | 0.0481 | 0 | 0 | 0 | 1 | Ref |
| 9838 | MN096377 | Unassigned | Unassigned | Unassigned | 889_0 | Clustered | 16 | VC_889_0 | 16 | 0.2969 | 1 | 0.2969 | 0 | 0 | 0 | 1 | Ref |
| 9839 | MN096378 | Unassigned | Unassigned | Unassigned | 1163_0 | Clustered | 28 | VC_1163_0 | 28 | 0.4923 | 1 | 0.4923 | 0 | 0 | 0 | 1 | Ref |
| 9840 | MN096379 | Unassigned | Unassigned | Unassigned | 982_0 | Clustered | 62 | VC_982_0 | 62 | 0.8646 | 1 | 0.8646 | 0 | 0 | 0 | 0.743 | Ref |
| 9841 | MN096380 | Unassigned | Unassigned | Unassigned | 888_0 | Clustered | 28 | VC_888_0 | 28 | 0.5497 | 1 | 0.5497 | 0 | 0 | 0 | 1 | Ref |
| 9842 | MN096381 | Unassigned | Unassigned | Unassigned | 982_0 | Clustered | 62 | VC_982_0 | 62 | 0.8646 | 1 | 0.8646 | 0 | 0 | 0 | 0.743 | Ref |
| 9843 | MN096382 | Unassigned | Unassigned | Unassigned | 887_0 | Clustered | 18 | VC_887_0 | 18 | 0.3663 | 1 | 0.3663 | 0 | 0 | 0 | 1 | Ref |
| 9844 | MN098325 | Unassigned | Unassigned | Unassigned | 96_0 | Clustered | 30 | VC_96_0 | 30 | 0.718 | 1 | 0.718 | 0 | 0 | 0 | 1 | Ref |
| 9845 | MN098326 | Unassigned | Unassigned | Unassigned | 792_0 | Clustered | 35 | VC_792_0 | 35 | 0.9828 | 1 | 0.9828 | 0 | 0 | 0 | 1 | Ref |
| 9846 | MN098327 | Unassigned | Unassigned | Unassigned | 242_1 | Clustered | 20 | VC_242_1 | 20 | 0.684 | 1 | 0.684 | 0 | 0 | 0 | 0.8368 | Ref |
| 9847 | MN098328 | Unassigned | Unassigned | Unassigned | 506_0 | Clustered | 8 | VC_506_0 | 8 | 0.7376 | 1 | 0.7376 | 0 | 0 | 0 | 1 | Ref |
| 9848 | MN098329 | Unassigned | Unassigned | Unassigned | 392_0 | Clustered | 126 | VC_392_0 | 126 | 0.9621 | 1 | 0.9621 | 0 | 0 | 0 | 1 | Ref |
| 9849 | MN101216 | Unassigned | Unassigned | Unassigned | 823_0 | Clustered | 124 | VC_823_0 | 124 | 0.9492 | 1 | 0.9492 | 0 | 0 | 0 | 1 | Ref |
| 9850 | MN101217 | Unassigned | Unassigned | Unassigned | 111_0 | Clustered | 91 | VC_111_0 | 91 | 0.9587 | 1 | 0.9587 | 0 | 0 | 0 | 1 | Ref |
| 9851 | MN101218 | Unassigned | Unassigned | Unassigned | 823_0 | Clustered | 124 | VC_823_0 | 124 | 0.9492 | 1 | 0.9492 | 0 | 0 | 0 | 1 | Ref |
| 9852 | MN101219 | Unassigned | Unassigned | Unassigned | 111_0 | Clustered | 91 | VC_111_0 | 91 | 0.9587 | 1 | 0.9587 | 0 | 0 | 0 | 1 | Ref |
| 9853 | MN101220 | Unassigned | Unassigned | Unassigned | 111_0 | Clustered | 91 | VC_111_0 | 91 | 0.9587 | 1 | 0.9587 | 0 | 0 | 0 | 1 | Ref |
| 9854 | MN101221 | Unassigned | Unassigned | Unassigned | 668_2 | Clustered/Singleton | 1 | VC_668_2 | 1 | 0 | 1 | 0 | 0 | 0 | 0 | 1 | Ref |
| 9855 | MN101222 | Unassigned | Unassigned | Unassigned | 586_0 | Clustered | 3 | VC_586_0 | 3 | 0.0495 | 0.99859933 | 0.0495 | 0 | 0 | 0 | 1 | Ref |
| 9856 | MN101223 | Unassigned | Unassigned | Unassigned | 668_0 | Clustered | 25 | VC_668_0 | 25 | 0.5539 | 1 | 0.5539 | 0 | 0 | 0 | 0.96 | Ref |
| 9857 | MN101224 | Unassigned | Unassigned | Unassigned | 668_3 | Clustered/Singleton | 1 | VC_668_3 | 1 | 0 | 1 | 0 | 0 | 0 | 0 | 1 | Ref |
| 9858 | MN101225 | Unassigned | Unassigned | Unassigned | 491_1 | Clustered/Singleton | 1 | VC_491_1 | 1 | 0 | 1 | 0 | 0 | 0 | 0 | 1 | Ref |
| 9859 | MN101226 | Unassigned | Unassigned | Unassigned | 491_0 | Clustered | 36 | VC_491_0 | 36 | 0.5336 | 1 | 0.5336 | 0 | 0 | 0 | 0.9714 | Ref |
| 9860 | MN101227 | Unassigned | Unassigned | Unassigned | 823_0 | Clustered | 124 | VC_823_0 | 124 | 0.9492 | 1 | 0.9492 | 0 | 0 | 0 | 1 | Ref |
| 9861 | MN101228 | Unassigned | Unassigned | Unassigned | 192_0 | Clustered | 15 | VC_192_0 | 15 | 0.5071 | 1 | 0.5071 | 0 | 0 | 0 | 1 | Ref |
| 9862 | MN101229 | Unassigned | Unassigned | Unassigned | 491_0 | Clustered | 36 | VC_491_0 | 36 | 0.5336 | 1 | 0.5336 | 0 | 0 | 0 | 0.9714 | Ref |
| 9863 | MN101230 | Unassigned | Unassigned | Unassigned | 491_0 | Clustered | 36 | VC_491_0 | 36 | 0.5336 | 1 | 0.5336 | 0 | 0 | 0 | 0.9714 | Ref |
| 9864 | MN102098 | Unassigned | Unassigned | Unassigned | 442_0 | Clustered | 4 | VC_442_0 | 4 | 0.0842 | 0.99996321 | 0.0842 | 0 | 0 | 0 | 1 | Ref |
| 9865 | MN103542 | Unassigned | Unassigned | Unassigned | 108_0 | Clustered | 33 | VC_108_0 | 33 | 0.949 | 1 | 0.949 | 0 | 0 | 0 | 1 | Ref |
| 9866 | MN103543 | Unassigned | Unassigned | Unassigned | 344_1 | Clustered | 3 | VC_344_1 | 3 | 0.0359 | 0.93348877 | 0.0335 | 0 | 0 | 0 | 1 | Ref |
| 9867 | MN114082 | Unassigned | Unassigned | Unassigned | 980_0 | Clustered | 10 | VC_980_0 | 10 | 0.9996 | 1 | 0.9996 | 0 | 0 | 0 | 1 | Ref |
| 9868 | MN114083 | Unassigned | Unassigned | Unassigned | 980_0 | Clustered | 10 | VC_980_0 | 10 | 0.9996 | 1 | 0.9996 | 0 | 0 | 0 | 1 | Ref |
| 9869 | MN116494 | Unassigned | Unassigned | Unassigned | 111_0 | Clustered | 91 | VC_111_0 | 91 | 0.9587 | 1 | 0.9587 | 0 | 0 | 0 | 1 | Ref |
| 9870 | MN117721 | Unassigned | Unassigned | Unassigned | 792_0 | Clustered | 35 | VC_792_0 | 35 | 0.9828 | 1 | 0.9828 | 0 | 0 | 0 | 1 | Ref |
| 9871 | MN119376 | Unassigned | Unassigned | Unassigned | 1224_0 | Clustered | 19 | VC_1224_0 | 19 | 1 | 1 | 1 | 0 | 0 | 0 | 1 | Ref |
| 9872 | MN119377 | Unassigned | Unassigned | Unassigned | 325_42 | Clustered | 4 | VC_325_42 | 4 | 0.0112 | 0.99980442 | 0.0112 | 0 | 0 | 0 | 0.6667 | Ref |
| 9873 | MN119378 | Unassigned | Unassigned | Unassigned | 565_0 | Clustered | 20 | VC_565_0 | 20 | 0.8398 | 1 | 0.8398 | 0 | 0 | 0 | 1 | Ref |
| 9874 | MN119379 | Unassigned | Unassigned | Unassigned | 325_10 | Clustered | 3 | VC_325_10 | 3 | 0.012 | 0.99859169 | 0.012 | 0 | 0 | 0 | 1 | Ref |
| 9875 | MN122072 | Unassigned | Unassigned | Unassigned | 792_0 | Clustered | 35 | VC_792_0 | 35 | 0.9828 | 1 | 0.9828 | 0 | 0 | 0 | 1 | Ref |
| 9876 | MN128593 | Unassigned | Unassigned | Unassigned | 980_0 | Clustered | 10 | VC_980_0 | 10 | 0.9996 | 1 | 0.9996 | 0 | 0 | 0 | 1 | Ref |
| 9877 | MN128594 | Unassigned | Unassigned | Unassigned | 739_0 | Clustered | 26 | VC_739_0 | 26 | 0.9571 | 1 | 0.9571 | 0 | 0 | 0 | 1 | Ref |
| 9878 | MN131137 | Unassigned | Unassigned | Unassigned | 442_0 | Clustered | 4 | VC_442_0 | 4 | 0.0842 | 0.99996321 | 0.0842 | 0 | 0 | 0 | 1 | Ref |
| 9879 | MN131141 | Unassigned | Unassigned | Unassigned | 79_7 | Clustered | 33 | VC_79_7 | 33 | 0.216 | 1 | 0.216 | 0 | 0 | 0 | 0.8409 | Ref |
| 9880 | MN131142 | Unassigned | Unassigned | Unassigned | 79_7 | Clustered | 33 | VC_79_7 | 33 | 0.216 | 1 | 0.216 | 0 | 0 | 0 | 0.8409 | Ref |

| | | | | | | | | | | | | | | | | | |
|---|---|---|---|---|---|---|---|---|---|---|---|---|---|---|---|---|---|
| 9881 | MN131143 | Unassigned | Unassigned | Unassigned | 79_7 | Clustered | 33 | VC_79_7 | 33 | 0.216 | 1 | 0.216 | 0 | 0 | 0 | 0.8409 | Ref |
| 9882 | MN136198 | Unassigned | Unassigned | Unassigned | 792_0 | Clustered | 35 | VC_792_0 | 35 | 0.9828 | 1 | 0.9828 | 0 | 0 | 0 | 1 | Ref |
| 9883 | MN148435 | Unassigned | Unassigned | Unassigned | 813_1 | Clustered | 24 | VC_813_1 | 24 | 0.535 | 1 | 0.535 | 0 | 0 | 0 | 1 | Ref |
| 9884 | MN149903 | Unassigned | Unassigned | Unassigned | 823_0 | Clustered | 124 | VC_823_0 | 124 | 0.9492 | 1 | 0.9492 | 0 | 0 | 0 | 1 | Ref |
| 9885 | MN149904 | Unassigned | Unassigned | Unassigned | 385_0 | Clustered | 51 | VC_385_0 | 51 | 0.8754 | 1 | 0.8754 | 0 | 0 | 0 | 1 | Ref |
| 9886 | MN150686 | Unassigned | Unassigned | Unassigned | 187_0 | Clustered | 4 | VC_187_0 | 4 | 0.0661 | 0.99994098 | 0.0661 | 0 | 0 | 0 | 1 | Ref |
| 9887 | MN150710 | Unassigned | Unassigned | Unassigned | 96_0 | Clustered | 30 | VC_96_0 | 30 | 0.718 | 1 | 0.718 | 0 | 0 | 0 | 1 | Ref |
| 9888 | MN153503 | Unassigned | Unassigned | Unassigned | 883_4 | Clustered/Singleton | 1 | VC_883_4 | 1 | 0 | 1 | 0 | 0 | 0 | 0 | 1 | Ref |
| 9889 | MN153797 | Unassigned | Unassigned | Unassigned | 575_0 | Clustered | 35 | VC_575_0 | 35 | 0.8815 | 1 | 0.8815 | 0 | 0 | 0 | 1 | Ref |
| 9890 | MN159079 | Unassigned | Unassigned | Unassigned | 442_0 | Clustered | 4 | VC_442_0 | 4 | 0.0842 | 0.99996321 | 0.0842 | 0 | 0 | 0 | 1 | Ref |
| 9891 | MN161573 | Unassigned | Unassigned | Unassigned | 28_1 | Clustered | 53 | VC_28_1 | 53 | 0.7164 | 1 | 0.7164 | 0 | 0 | 0 | 0.9325 | Ref |
| 9892 | MN163280 | Unassigned | Unassigned | Unassigned | 192_0 | Clustered | 15 | VC_192_0 | 15 | 0.5071 | 1 | 0.5071 | 0 | 0 | 0 | 1 | Ref |
| 9893 | MN163281 | Unassigned | Unassigned | Unassigned | 780_0 | Clustered | 8 | VC_780_0 | 8 | 0.5207 | 1 | 0.5207 | 0 | 0 | 0 | 1 | Ref |
| 9894 | MN164484 | Unassigned | Unassigned | Unassigned | 456_0 | Clustered | 62 | VC_456_0 | 62 | 0.9026 | 1 | 0.9026 | 0 | 0 | 0 | 1 | Ref |
| 9895 | MN166083 | Unassigned | Unassigned | Unassigned | 698_0 | Clustered | 26 | VC_698_0 | 26 | 0.9366 | 1 | 0.9366 | 0 | 0 | 0 | 1 | Ref |
| 9896 | MN166823 | Unassigned | Unassigned | Unassigned | 242_1 | Clustered | 20 | VC_242_1 | 20 | 0.684 | 1 | 0.684 | 0 | 0 | 0 | 0.8368 | Ref |
| 9897 | MN172529 | Unassigned | Unassigned | Unassigned | 739_0 | Clustered | 26 | VC_739_0 | 26 | 0.9571 | 1 | 0.9571 | 0 | 0 | 0 | 1 | Ref |
| 9898 | MN175603 | Unassigned | Unassigned | Unassigned | 1352_0 | Clustered | 31 | VC_1352_0 | 31 | 0.849 | 1 | 0.849 | 0 | 0 | 0 | 1 | Ref |
| 9899 | MN175604 | Unassigned | Unassigned | Unassigned | 522_0 | Clustered | 3 | VC_522_0 | 3 | 0.0836 | 0.9985266 | 0.0835 | 0 | 0 | 0 | 1 | Ref |
| 9900 | MN176217 | Unassigned | Unassigned | Unassigned | 618_0 | Clustered | 13 | VC_618_0 | 13 | 0.6252 | 1 | 0.6252 | 0 | 0 | 0 | 1 | Ref |
| 9901 | MN176219 | Unassigned | Unassigned | Unassigned | 104_0 | Clustered | 6 | VC_104_0 | 6 | 0.8291 | 1 | 0.8291 | 0 | 0 | 0 | 1 | Ref |
| 9902 | MN176222 | Unassigned | Unassigned | Unassigned | 103_0 | Clustered | 6 | VC_103_0 | 6 | 0.7337 | 1 | 0.7337 | 0 | 0 | 0 | 1 | Ref |
| 9903 | MN176223 | Unassigned | Unassigned | Unassigned | 103_0 | Clustered | 6 | VC_103_0 | 6 | 0.7337 | 1 | 0.7337 | 0 | 0 | 0 | 1 | Ref |
| 9904 | MN176224 | Unassigned | Unassigned | Unassigned | 103_0 | Clustered | 6 | VC_103_0 | 6 | 0.7337 | 1 | 0.7337 | 0 | 0 | 0 | 1 | Ref |
| 9905 | MN176225 | Unassigned | Unassigned | Unassigned | 103_0 | Clustered | 6 | VC_103_0 | 6 | 0.7337 | 1 | 0.7337 | 0 | 0 | 0 | 1 | Ref |
| 9906 | MN176226 | Unassigned | Unassigned | Unassigned | 103_0 | Clustered | 6 | VC_103_0 | 6 | 0.7337 | 1 | 0.7337 | 0 | 0 | 0 | 1 | Ref |
| 9907 | MN176227 | Unassigned | Unassigned | Unassigned | 104_0 | Clustered | 6 | VC_104_0 | 6 | 0.8291 | 1 | 0.8291 | 0 | 0 | 0 | 1 | Ref |
| 9908 | MN176228 | Unassigned | Unassigned | Unassigned | 104_0 | Clustered | 6 | VC_104_0 | 6 | 0.8291 | 1 | 0.8291 | 0 | 0 | 0 | 1 | Ref |
| 9909 | MN176229 | Unassigned | Unassigned | Unassigned | 103_0 | Clustered | 6 | VC_103_0 | 6 | 0.7337 | 1 | 0.7337 | 0 | 0 | 0 | 1 | Ref |
| 9910 | MN176572 | Unassigned | Unassigned | Unassigned | 111_0 | Clustered | 91 | VC_111_0 | 91 | 0.9587 | 1 | 0.9587 | 0 | 0 | 0 | 1 | Ref |
| 9911 | MN176573 | Unassigned | Unassigned | Unassigned | 111_0 | Clustered | 91 | VC_111_0 | 91 | 0.9587 | 1 | 0.9587 | 0 | 0 | 0 | 1 | Ref |
| 9912 | MN176574 | Unassigned | Unassigned | Unassigned | 823_0 | Clustered | 124 | VC_823_0 | 124 | 0.9492 | 1 | 0.9492 | 0 | 0 | 0 | 1 | Ref |
| 9913 | MN180248 | Unassigned | Unassigned | Unassigned | 22_0 | Clustered | 11 | VC_22_0 | 11 | 0.5163 | 1 | 0.5163 | 0 | 0 | 0 | 1 | Ref |
| 9914 | MN180249 | Unassigned | Unassigned | Unassigned | 22_0 | Clustered | 11 | VC_22_0 | 11 | 0.5163 | 1 | 0.5163 | 0 | 0 | 0 | 1 | Ref |
| 9915 | MN180250 | Unassigned | Unassigned | Unassigned | 22_0 | Clustered | 11 | VC_22_0 | 11 | 0.5163 | 1 | 0.5163 | 0 | 0 | 0 | 1 | Ref |
| 9916 | MN180251 | Unassigned | Unassigned | Unassigned | 22_0 | Clustered | 11 | VC_22_0 | 11 | 0.5163 | 1 | 0.5163 | 0 | 0 | 0 | 1 | Ref |
| 9917 | MN180252 | Unassigned | Unassigned | Unassigned | 22_0 | Clustered | 11 | VC_22_0 | 11 | 0.5163 | 1 | 0.5163 | 0 | 0 | 0 | 1 | Ref |
| 9918 | MN183279 | Unassigned | Unassigned | Unassigned | 1073_0 | Clustered | 24 | VC_1073_0 | 24 | 0.8388 | 1 | 0.8388 | 0 | 0 | 0 | 1 | Ref |
| 9919 | MN183280 | Unassigned | Unassigned | Unassigned | 558_60 | Clustered/Singleton | 1 | VC_558_60 | 1 | 0 | 1 | 0 | 0 | 0 | 0 | 1 | Ref |
| 9920 | MN183281 | Unassigned | Unassigned | Unassigned | 1234_0 | Clustered | 44 | VC_1234_0 | 44 | 1 | 1 | 1 | 0 | 0 | 0 | 1 | Ref |
| 9921 | MN183282 | Unassigned | Unassigned | Unassigned | 1043_0 | Clustered | 2 | VC_1043_0 | 2 | 0.0576 | 0.95654247 | 0.0551 | 0 | 0 | 0 | 1 | Ref |
| 9922 | MN183283 | Unassigned | Unassigned | Unassigned | 1073_0 | Clustered | 24 | VC_1073_0 | 24 | 0.8388 | 1 | 0.8388 | 0 | 0 | 0 | 1 | Ref |
| 9923 | MN183284 | Unassigned | Unassigned | Unassigned | 1171_0 | Clustered | 21 | VC_1171_0 | 21 | 0.7125 | 1 | 0.7125 | 0 | 0 | 0 | 1 | Ref |
| 9924 | MN183285 | Unassigned | Unassigned | Unassigned | 558_18 | Clustered/Singleton | 1 | VC_558_18 | 1 | 0 | 1 | 0 | 0 | 0 | 0 | 1 | Ref |
| 9925 | MN183286 | Unassigned | Unassigned | Unassigned | 546_4 | Clustered | 16 | VC_546_4 | 16 | 0.2517 | 0.9999998 | 0.2517 | 0 | 0 | 0 | 0.9417 | Ref |
| 9926 | MN184885 | Unassigned | Unassigned | Unassigned | 395_0 | Clustered | 5 | VC_395_0 | 5 | 0.1021 | 0.99999998 | 0.1021 | 0 | 0 | 0 | 1 | Ref |
| 9927 | MN184887 | Unassigned | Unassigned | Unassigned | 582_0 | Clustered | 4 | VC_582_0 | 4 | 0.0589 | 0.99996986 | 0.0589 | 0 | 0 | 0 | 1 | Ref |
| 9928 | MN187550 | Unassigned | Unassigned | Unassigned | 476_0 | Clustered | 19 | VC_476_0 | 19 | 0.7814 | 1 | 0.7814 | 0 | 0 | 0 | 1 | Ref |
| 9929 | MN187969 | Unassigned | Unassigned | Unassigned | 191_19 | Clustered | 12 | VC_191_19 | 12 | 0.0655 | 1 | 0.0655 | 0 | 0 | 0 | 0.8636 | Ref |
| 9930 | MN191861 | Unassigned | Unassigned | Unassigned | 70_0 | Clustered | 27 | VC_70_0 | 27 | 0.5981 | 1 | 0.5981 | 0 | 0 | 0 | 0.9886 | Ref |
| 9931 | MN197465 | Unassigned | Unassigned | Unassigned | 191_10 | Clustered/Singleton | 1 | VC_191_10 | 1 | 0 | 1 | 0 | 0 | 0 | 0 | 1 | Ref |
| 9932 | MN200777 | Unassigned | Unassigned | Unassigned | 26_0 | Clustered | 4 | VC_26_0 | 4 | 0.7433 | 0.99936153 | 0.7428 | 0 | 0 | 0 | 1 | Ref |
| 9933 | MN200778 | Unassigned | Unassigned | Unassigned | 26_0 | Clustered | 4 | VC_26_0 | 4 | 0.7433 | 0.99936153 | 0.7428 | 0 | 0 | 0 | 1 | Ref |
| 9934 | MN204492 | Unassigned | Unassigned | Unassigned | 872_0 | Clustered | 31 | VC_872_0 | 31 | 0.8151 | 1 | 0.8151 | 0 | 0 | 0 | 0.8194 | Ref |
| 9935 | MN204493 | Unassigned | Unassigned | Unassigned | 920_0 | Clustered | 2 | VC_920_0 | 2 | 0.1671 | 0.95556296 | 0.1597 | 0 | 0 | 0 | 1 | Ref |
| 9936 | MN204494 | Unassigned | Unassigned | Unassigned | 872_0 | Clustered | 31 | VC_872_0 | 31 | 0.8151 | 1 | 0.8151 | 0 | 0 | 0 | 0.8194 | Ref |
| 9937 | MN204495 | Unassigned | Unassigned | Unassigned | 872_0 | Clustered | 31 | VC_872_0 | 31 | 0.8151 | 1 | 0.8151 | 0 | 0 | 0 | 0.8194 | Ref |
| 9938 | MN204496 | Unassigned | Unassigned | Unassigned | 872_0 | Clustered | 31 | VC_872_0 | 31 | 0.8151 | 1 | 0.8151 | 0 | 0 | 0 | 0.8194 | Ref |
| 9939 | MN204497 | Unassigned | Unassigned | Unassigned | 872_0 | Clustered | 31 | VC_872_0 | 31 | 0.8151 | 1 | 0.8151 | 0 | 0 | 0 | 0.8194 | Ref |
| 9940 | MN204498 | Unassigned | Unassigned | Unassigned | 982_0 | Clustered | 62 | VC_982_0 | 62 | 0.8646 | 1 | 0.8646 | 0 | 0 | 0 | 0.743 | Ref |
| 9941 | MN204499 | Unassigned | Unassigned | Unassigned | 1074_0 | Clustered | 19 | VC_1074_0 | 19 | 0.9198 | 1 | 0.9198 | 0 | 0 | 0 | 1 | Ref |
| 9942 | MN204500 | Unassigned | Unassigned | Unassigned | 872_0 | Clustered | 31 | VC_872_0 | 31 | 0.8151 | 1 | 0.8151 | 0 | 0 | 0 | 0.8194 | Ref |
| 9943 | MN204501 | Unassigned | Unassigned | Unassigned | 799_1 | Clustered | 27 | VC_799_1 | 27 | 0.8611 | 1 | 0.8611 | 0 | 0 | 0 | 1 | Ref |
| 9944 | MN204503 | Unassigned | Unassigned | Unassigned | 1074_0 | Clustered | 19 | VC_1074_0 | 19 | 0.9198 | 1 | 0.9198 | 0 | 0 | 0 | 1 | Ref |
| 9945 | MN204504 | Unassigned | Unassigned | Unassigned | 872_0 | Clustered | 31 | VC_872_0 | 31 | 0.8151 | 1 | 0.8151 | 0 | 0 | 0 | 0.8194 | Ref |
| 9946 | MN212906 | Unassigned | Unassigned | Unassigned | 699_0 | Clustered | 71 | VC_699_0 | 71 | 0.9352 | 1 | 0.9352 | 0 | 0 | 0 | 1 | Ref |
| 9947 | MN215888 | Unassigned | Unassigned | Unassigned | 882_0 | Clustered | 20 | VC_882_0 | 20 | 0.9917 | 1 | 0.9917 | 0 | 0 | 0 | 1 | Ref |
| 9948 | MN218190 | Unassigned | Unassigned | Unassigned | 191_16 | Clustered | 12 | VC_191_16 | 12 | 0.0643 | 1 | 0.0643 | 0 | 0 | 0 | 0.9394 | Ref |
| 9949 | MN218191 | Unassigned | Unassigned | Unassigned | 191_16 | Clustered | 12 | VC_191_16 | 12 | 0.0643 | 1 | 0.0643 | 0 | 0 | 0 | 0.9394 | Ref |
| 9950 | MN218775 | Unassigned | Unassigned | Unassigned | 451_0 | Clustered | 80 | VC_451_0 | 80 | 0.9991 | 1 | 0.9991 | 0 | 0 | 0 | 1 | Ref |
| 9951 | MN224564 | Unassigned | Unassigned | Unassigned | 1353_0 | Clustered | 10 | VC_1353_0 | 10 | 1 | 1 | 1 | 0 | 0 | 0 | 1 | Ref |
| 9952 | MN224565 | Unassigned | Unassigned | Unassigned | 1353_0 | Clustered | 10 | VC_1353_0 | 10 | 1 | 1 | 1 | 0 | 0 | 0 | 1 | Ref |
| 9953 | MN227145 | Unassigned | Unassigned | Unassigned | 329_11 | Clustered | 15 | VC_329_11 | 15 | 0.0791 | 1 | 0.0791 | 0 | 0 | 0 | 0.9333 | Ref |
| 9954 | MN228696 | Unassigned | Unassigned | Unassigned | 783_0 | Clustered | 3 | VC_783_0 | 3 | 0.0639 | 0.99854307 | 0.0638 | 0 | 0 | 0 | 1 | Ref |
| 9955 | MN234161 | Unassigned | Unassigned | Unassigned | 1176_0 | Clustered | 41 | VC_1176_0 | 41 | 0.9558 | 1 | 0.9558 | 0 | 0 | 0 | 0.9976 | Ref |
| 9956 | MN234162 | Unassigned | Unassigned | Unassigned | 572_0 | Clustered | 16 | VC_572_0 | 16 | 0.7286 | 1 | 0.7286 | 0 | 0 | 0 | 1 | Ref |
| 9957 | MN234163 | Unassigned | Unassigned | Unassigned | 1240_0 | Clustered | 82 | VC_1240_0 | 82 | 0.7945 | 1 | 0.7945 | 0 | 0 | 0 | 1 | Ref |
| 9958 | MN234164 | Unassigned | Unassigned | Unassigned | 857_0 | Clustered | 11 | VC_857_0 | 11 | 0.9712 | 1 | 0.9712 | 0 | 0 | 0 | 1 | Ref |
| 9959 | MN234165 | Unassigned | Unassigned | Unassigned | 236_1 | Clustered | 5 | VC_236_1 | 5 | 0.0358 | 0.99999997 | 0.0358 | 0 | 0 | 0 | 1 | Ref |
| 9960 | MN234166 | Unassigned | Unassigned | Unassigned | 869_0 | Clustered | 9 | VC_869_0 | 9 | 0.2624 | 1 | 0.2624 | 0 | 0 | 0 | 1 | Ref |
| 9961 | MN234167 | Unassigned | Unassigned | Unassigned | 225_0 | Clustered | 32 | VC_225_0 | 32 | 0.848 | 1 | 0.848 | 0 | 0 | 0 | 0.996 | Ref |
| 9962 | MN234168 | Unassigned | Unassigned | Unassigned | 1164_0 | Clustered | 25 | VC_1164_0 | 25 | 0.7312 | 1 | 0.7312 | 0 | 0 | 0 | 1 | Ref |
| 9963 | MN234169 | Unassigned | Unassigned | Unassigned | 215_1 | Clustered | 19 | VC_215_1 | 19 | 0.4188 | 1 | 0.4188 | 0 | 0 | 0 | 1 | Ref |
| 9964 | MN234171 | Unassigned | Unassigned | Unassigned | 372_0 | Clustered | 24 | VC_372_0 | 24 | 0.5539 | 1 | 0.5539 | 0 | 0 | 0 | 1 | Ref |
| 9965 | MN234174 | Unassigned | Unassigned | Unassigned | 236_1 | Clustered | 5 | VC_236_1 | 5 | 0.0358 | 0.99999997 | 0.0358 | 0 | 0 | 0 | 1 | Ref |
| 9966 | MN234176 | Unassigned | Unassigned | Unassigned | 237_2 | Clustered | 11 | VC_237_2 | 11 | 0.1409 | 1 | 0.1409 | 0 | 0 | 0 | 0.9818 | Ref |
| 9967 | MN234177 | Unassigned | Unassigned | Unassigned | 1240_0 | Clustered | 82 | VC_1240_0 | 82 | 0.7945 | 1 | 0.7945 | 0 | 0 | 0 | 1 | Ref |
| 9968 | MN234179 | Unassigned | Unassigned | Unassigned | 1176_0 | Clustered | 41 | VC_1176_0 | 41 | 0.9558 | 1 | 0.9558 | 0 | 0 | 0 | 0.9976 | Ref |
| 9969 | MN234181 | Unassigned | Unassigned | Unassigned | 1240_0 | Clustered | 82 | VC_1240_0 | 82 | 0.7945 | 1 | 0.7945 | 0 | 0 | 0 | 1 | Ref |
| 9970 | MN234182 | Unassigned | Unassigned | Unassigned | 236_0 | Clustered | 72 | VC_236_0 | 72 | 0.8104 | 1 | 0.8104 | 0 | 0 | 0 | 1 | Ref |
| 9971 | MN234183 | Unassigned | Unassigned | Unassigned | 762_0 | Clustered | 9 | VC_762_0 | 9 | 0.0885 | 1 | 0.0885 | 0 | 0 | 0 | 1 | Ref |
| 9972 | MN234185 | Unassigned | Unassigned | Unassigned | 762_0 | Clustered | 9 | VC_762_0 | 9 | 0.0885 | 1 | 0.0885 | 0 | 0 | 0 | 1 | Ref |
| 9973 | MN234187 | Unassigned | Unassigned | Unassigned | 888_1 | Clustered | 3 | VC_888_1 | 3 | 0.0289 | 0.99848448 | 0.0289 | 0 | 0 | 0 | 1 | Ref |
| 9974 | MN234190 | Unassigned | Unassigned | Unassigned | 572_0 | Clustered | 16 | VC_572_0 | 16 | 0.7286 | 1 | 0.7286 | 0 | 0 | 0 | 1 | Ref |
| 9975 | MN234192 | Unassigned | Unassigned | Unassigned | 981_1 | Clustered | 26 | VC_981_1 | 26 | 0.7009 | 1 | 0.7009 | 0 | 0 | 0 | 1 | Ref |
| 9976 | MN234193 | Unassigned | Unassigned | Unassigned | 558_36 | Clustered | 4 | VC_558_36 | 4 | 0.0096 | 0.85913852 | 0.0082 | 0 | 0 | 0 | 1 | Ref |
| 9977 | MN234194 | Unassigned | Unassigned | Unassigned | 1164_0 | Clustered | 25 | VC_1164_0 | 25 | 0.7312 | 1 | 0.7312 | 0 | 0 | 0 | 1 | Ref |
| 9978 | MN234195 | Unassigned | Unassigned | Unassigned | 518_0 | Clustered | 6 | VC_518_0 | 6 | 0.1295 | 1 | 0.1295 | 0 | 0 | 0 | 1 | Ref |
| 9979 | MN234196 | Unassigned | Unassigned | Unassigned | 510_0 | Clustered | 12 | VC_510_0 | 12 | 0.3963 | 1 | 0.3963 | 0 | 0 | 0 | 0.9697 | Ref |
| 9980 | MN234197 | Unassigned | Unassigned | Unassigned | 236_0 | Clustered | 72 | VC_236_0 | 72 | 0.8104 | 1 | 0.8104 | 0 | 0 | 0 | 1 | Ref |
| 9981 | MN234198 | Unassigned | Unassigned | Unassigned | 236_0 | Clustered | 72 | VC_236_0 | 72 | 0.8104 | 1 | 0.8104 | 0 | 0 | 0 | 1 | Ref |
| 9982 | MN234199 | Unassigned | Unassigned | Unassigned | 237_1 | Clustered | 13 | VC_237_1 | 13 | 0.1773 | 1 | 0.1773 | 0 | 0 | 0 | 0.7308 | Ref |
| 9983 | MN234200 | Unassigned | Unassigned | Unassigned | 225_0 | Clustered | 32 | VC_225_0 | 32 | 0.848 | 1 | 0.848 | 0 | 0 | 0 | 0.996 | Ref |
| 9984 | MN234201 | Unassigned | Unassigned | Unassigned | 236_1 | Clustered | 5 | VC_236_1 | 5 | 0.0358 | 0.99999997 | 0.0358 | 0 | 0 | 0 | 1 | Ref |
| 9985 | MN234203 | Unassigned | Unassigned | Unassigned | 216_6 | Clustered | 42 | VC_216_6 | 42 | 0.473 | 1 | 0.473 | 0 | 0 | 0 | 0.8351 | Ref |
| 9986 | MN234204 | Unassigned | Unassigned | Unassigned | 1240_0 | Clustered | 82 | VC_1240_0 | 82 | 0.7945 | 1 | 0.7945 | 0 | 0 | 0 | 1 | Ref |
| 9987 | MN234205 | Unassigned | Unassigned | Unassigned | 236_0 | Clustered | 72 | VC_236_0 | 72 | 0.8104 | 1 | 0.8104 | 0 | 0 | 0 | 1 | Ref |
| 9988 | MN234206 | Unassigned | Unassigned | Unassigned | 1099_0 | Clustered | 2 | VC_1099_0 | 2 | 0.2612 | 0.94673154 | 0.2473 | 0 | 0 | 0 | 1 | Ref |
| 9989 | MN234207 | Unassigned | Unassigned | Unassigned | 572_0 | Clustered | 16 | VC_572_0 | 16 | 0.7286 | 1 | 0.7286 | 0 | 0 | 0 | 1 | Ref |
| 9990 | MN234208 | Unassigned | Unassigned | Unassigned | 981_1 | Clustered | 26 | VC_981_1 | 26 | 0.7009 | 1 | 0.7009 | 0 | 0 | 0 | 1 | Ref |
| 9991 | MN234209 | Unassigned | Unassigned | Unassigned | 225_0 | Clustered | 32 | VC_225_0 | 32 | 0.848 | 1 | 0.848 | 0 | 0 | 0 | 0.996 | Ref |
| 9992 | MN234211 | Unassigned | Unassigned | Unassigned | 1099_0 | Clustered | 2 | VC_1099_0 | 2 | 0.2612 | 0.94673154 | 0.2473 | 0 | 0 | 0 | 1 | Ref |
| 9993 | MN234212 | Unassigned | Unassigned | Unassigned | 325_52 | Clustered | 5 | VC_325_52 | 5 | 0.0147 | 0.99999073 | 0.0147 | 0 | 0 | 0 | 0.7 | Ref |
| 9994 | MN234213 | Unassigned | Unassigned | Unassigned | 949_0 | Clustered | 10 | VC_949_0 | 10 | 0.5977 | 1 | 0.5977 | 0 | 0 | 0 | 1 | Ref |
| 9995 | MN234214 | Unassigned | Unassigned | Unassigned | 1166_0 | Clustered | 20 | VC_1166_0 | 20 | 0.99 | 1 | 0.99 | 0 | 0 | 0 | 1 | Ref |
| 9996 | MN234215 | Unassigned | Unassigned | Unassigned | 919_0 | Clustered | 39 | VC_919_0 | 39 | 0.9069 | 1 | 0.9069 | 0 | 0 | 0 | 1 | Ref |
| 9997 | MN234216 | Unassigned | Unassigned | Unassigned | 864_0 | Clustered | 2 | VC_864_0 | 2 | 1 | 1 | 1 | 0 | 0 | 0 | 1 | Ref |
| 9998 | MN234217 | Unassigned | Unassigned | Unassigned | 857_0 | Clustered | 11 | VC_857_0 | 11 | 0.9712 | 1 | 0.9712 | 0 | 0 | 0 | 1 | Ref |
| 9999 | MN234219 | Unassigned | Unassigned | Unassigned | 762_0 | Clustered | 9 | VC_762_0 | 9 | 0.0885 | 1 | 0.0885 | 0 | 0 | 0 | 1 | Ref |
| 10000 | MN234220 | Unassigned | Unassigned | Unassigned | 572_0 | Clustered | 16 | VC_572_0 | 16 | 0.7286 | 1 | 0.7286 | 0 | 0 | 0 | 1 | Ref |
| 10001 | MN234221 | Unassigned | Unassigned | Unassigned | 1164_0 | Clustered | 25 | VC_1164_0 | 25 | 0.7312 | 1 | 0.7312 | 0 | 0 | 0 | 1 | Ref |
| 10002 | MN234222 | Unassigned | Unassigned | Unassigned | 1240_0 | Clustered | 82 | VC_1240_0 | 82 | 0.7945 | 1 | 0.7945 | 0 | 0 | 0 | 1 | Ref |
| 10003 | MN234223 | Unassigned | Unassigned | Unassigned | 509_0 | Clustered | 37 | VC_509_0 | 37 | 0.9689 | 1 | 0.9689 | 0 | 0 | 0 | 1 | Ref |
| 10004 | MN234224 | Unassigned | Unassigned | Unassigned | 857_0 | Clustered | 11 | VC_857_0 | 11 | 0.9712 | 1 | 0.9712 | 0 | 0 | 0 | 1 | Ref |
| 10005 | MN234225 | Unassigned | Unassigned | Unassigned | 508_0 | Clustered | 32 | VC_508_0 | 32 | 0.906 | 1 | 0.906 | 0 | 0 | 0 | 1 | Ref |
| 10006 | MN234226 | Unassigned | Unassigned | Unassigned | 236_1 | Clustered | 5 | VC_236_1 | 5 | 0.0358 | 0.99999997 | 0.0358 | 0 | 0 | 0 | 1 | Ref |
| 10007 | MN234227 | Unassigned | Unassigned | Unassigned | 565_0 | Clustered | 20 | VC_565_0 | 20 | 0.8398 | 1 | 0.8398 | 0 | 0 | 0 | 1 | Ref |
| 10008 | MN234228 | Unassigned | Unassigned | Unassigned | 888_0 | Clustered | 28 | VC_888_0 | 28 | 0.5497 | 1 | 0.5497 | 0 | 0 | 0 | 1 | Ref |
| 10009 | MN234230 | Unassigned | Unassigned | Unassigned | 236_0 | Clustered | 72 | VC_236_0 | 72 | 0.8104 | 1 | 0.8104 | 0 | 0 | 0 | 1 | Ref |
| 10010 | MN234231 | Unassigned | Unassigned | Unassigned | 236_0 | Clustered | 72 | VC_236_0 | 72 | 0.8104 | 1 | 0.8104 | 0 | 0 | 0 | 1 | Ref |
| 10011 | MN234232 | Unassigned | Unassigned | Unassigned | 572_0 | Clustered | 16 | VC_572_0 | 16 | 0.7286 | 1 | 0.7286 | 0 | 0 | 0 | 1 | Ref |
| 10012 | MN234233 | Unassigned | Unassigned | Unassigned | 1200_0 | Clustered | 4 | VC_1200_0 | 4 | 0.8691 | 0.9999992 | 0.8691 | 0 | 0 | 0 | 1 | Ref |
| 10013 | MN234234 | Unassigned | Unassigned | Unassigned | 1074_0 | Clustered | 19 | VC_1074_0 | 19 | 0.9198 | 1 | 0.9198 | 0 | 0 | 0 | 1 | Ref |
| 10014 | MN234235 | Unassigned | Unassigned | Unassigned | 225_0 | Clustered | 32 | VC_225_0 | 32 | 0.848 | 1 | 0.848 | 0 | 0 | 0 | 0.996 | Ref |
| 10015 | MN234236 | Unassigned | Unassigned | Unassigned | 518_0 | Clustered | 6 | VC_518_0 | 6 | 0.1295 | 1 | 0.1295 | 0 | 0 | 0 | 1 | Ref |
| 10016 | MN241318 | Unassigned | Unassigned | Unassigned | 92_0 | Clustered | 22 | VC_92_0 | 22 | 0.5639 | 1 | 0.5639 | 0 | 0 | 0 | 1 | Ref |
| 10017 | MN252582 | Unassigned | Unassigned | Unassigned | 386_0 | Clustered | 49 | VC_386_0 | 49 | 0.7941 | 1 | 0.7941 | 0 | 0 | 0 | 1 | Ref |
| 10018 | MN263053 | Unassigned | Unassigned | Unassigned | 178_0 | Clustered | 5 | VC_178_0 | 5 | 0.7741 | 0.99998991 | 0.7741 | 0 | 0 | 0 | 1 | Ref |
| 10019 | MN270258 | Unassigned | Unassigned | Unassigned | 632_0 | Clustered | 37 | VC_632_0 | 37 | 0.581 | 1 | 0.581 | 0 | 0 | 0 | 0.9234 | Ref |
| 10020 | MN270259 | Unassigned | Unassigned | Unassigned | 632_0 | Clustered | 37 | VC_632_0 | 37 | 0.581 | 1 | 0.581 | 0 | 0 | 0 | 0.9234 | Ref |
| 10021 | MN270260 | Unassigned | Unassigned | Unassigned | 632_0 | Clustered | 37 | VC_632_0 | 37 | 0.581 | 1 | 0.581 | 0 | 0 | 0 | 0.9234 | Ref |
| 10022 | MN270261 | Unassigned | Unassigned | Unassigned | 632_0 | Clustered | 37 | VC_632_0 | 37 | 0.581 | 1 | 0.581 | 0 | 0 | 0 | 0.9234 | Ref |
| 10023 | MN270262 | Unassigned | Unassigned | Unassigned | 632_0 | Clustered | 37 | VC_632_0 | 37 | 0.581 | 1 | 0.581 | 0 | 0 | 0 | 0.9234 | Ref |
| 10024 | MN270263 | Unassigned | Unassigned | Unassigned | 632_0 | Clustered | 37 | VC_632_0 | 37 | 0.581 | 1 | 0.581 | 0 | 0 | 0 | 0.9234 | Ref |
| 10025 | MN270264 | Unassigned | Unassigned | Unassigned | 632_0 | Clustered | 37 | VC_632_0 | 37 | 0.581 | 1 | 0.581 | 0 | 0 | 0 | 0.9234 | Ref |
| 10026 | MN270265 | Unassigned | Unassigned | Unassigned | 632_0 | Clustered | 37 | VC_632_0 | 37 | 0.581 | 1 | 0.581 | 0 | 0 | 0 | 0.9234 | Ref |
| 10027 | MN270266 | Unassigned | Unassigned | Unassigned | 632_0 | Clustered | 37 | VC_632_0 | 37 | 0.581 | 1 | 0.581 | 0 | 0 | 0 | 0.9234 | Ref |
| 10028 | MN270267 | Unassigned | Unassigned | Unassigned | 632_0 | Clustered | 37 | VC_632_0 | 37 | 0.581 | 1 | 0.581 | 0 | 0 | 0 | 0.9234 | Ref |
| 10029 | MN270268 | Unassigned | Unassigned | Unassigned | 632_0 | Clustered | 37 | VC_632_0 | 37 | 0.581 | 1 | 0.581 | 0 | 0 | 0 | 0.9234 | Ref |
| 10030 | MN270269 | Unassigned | Unassigned | Unassigned | 632_0 | Clustered | 37 | VC_632_0 | 37 | 0.581 | 1 | 0.581 | 0 | 0 | 0 | 0.9234 | Ref |
| 10031 | MN270270 | Unassigned | Unassigned | Unassigned | 632_0 | Clustered | 37 | VC_632_0 | 37 | 0.581 | 1 | 0.581 | 0 | 0 | 0 | 0.9234 | Ref |
| 10032 | MN270271 | Unassigned | Unassigned | Unassigned | 632_0 | Clustered | 37 | VC_632_0 | 37 | 0.581 | 1 | 0.581 | 0 | 0 | 0 | 0.9234 | Ref |
| 10033 | MN270272 | Unassigned | Unassigned | Unassigned | 632_0 | Clustered | 37 | VC_632_0 | 37 | 0.581 | 1 | 0.581 | 0 | 0 | 0 | 0.9234 | Ref |
| 10034 | MN270273 | Unassigned | Unassigned | Unassigned | 632_0 | Clustered | 37 | VC_632_0 | 37 | 0.581 | 1 | 0.581 | 0 | 0 | 0 | 0.9234 | Ref |
| 10035 | MN270274 | Unassigned | Unassigned | Unassigned | 632_0 | Clustered | 37 | VC_632_0 | 37 | 0.581 | 1 | 0.581 | 0 | 0 | 0 | 0.9234 | Ref |
| 10036 | MN270275 | Unassigned | Unassigned | Unassigned | 632_0 | Clustered | 37 | VC_632_0 | 37 | 0.581 | 1 | 0.581 | 0 | 0 | 0 | 0.9234 | Ref |
| 10037 | MN270276 | Unassigned | Unassigned | Unassigned | 632_0 | Clustered | 37 | VC_632_0 | 37 | 0.581 | 1 | 0.581 | 0 | 0 | 0 | 0.9234 | Ref |
| 10038 | MN270277 | Unassigned | Unassigned | Unassigned | 632_0 | Clustered | 37 | VC_632_0 | 37 | 0.581 | 1 | 0.581 | 0 | 0 | 0 | 0.9234 | Ref |
| 10039 | MN270278 | Unassigned | Unassigned | Unassigned | 632_0 | Clustered | 37 | VC_632_0 | 37 | 0.581 | 1 | 0.581 | 0 | 0 | 0 | 0.9234 | Ref |
| 10040 | MN270279 | Unassigned | Unassigned | Unassigned | 632_0 | Clustered | 37 | VC_632_0 | 37 | 0.581 | 1 | 0.581 | 0 | 0 | 0 | 0.9234 | Ref |
| 10041 | MN270887 | Unassigned | Unassigned | Unassigned | 984_0 | Clustered | 38 | VC_984_0 | 38 | 0.9758 | 1 | 0.9758 | 0 | 0 | 0 | 1 | Ref |
| 10042 | MN270888 | Unassigned | Unassigned | Unassigned | 984_0 | Clustered | 38 | VC_984_0 | 38 | 0.9758 | 1 | 0.9758 | 0 | 0 | 0 | 1 | Ref |

| | | | | | | | | | | | | | | | | | |
|---|---|---|---|---|---|---|---|---|---|---|---|---|---|---|---|---|---|
| 10043 | MN270891 | Unassigned | Unassigned | Unassigned | 331_0 | Clustered | 4 | VC_331_0 | 4 | 0.0884 | 0.99998775 | 0.0884 | 0 | 0 | 0 | 1 | Ref |
| 10044 | MN270892 | Unassigned | Unassigned | Unassigned | 331_0 | Clustered | 4 | VC_331_0 | 4 | 0.0884 | 0.99998775 | 0.0884 | 0 | 0 | 0 | 1 | Ref |
| 10045 | MN271656 | Unassigned | Unassigned | Unassigned | 984_0 | Clustered | 38 | VC_984_0 | 38 | 0.9758 | 1 | 0.9758 | 0 | 0 | 0 | 1 | Ref |
| 10046 | MN276049 | Unassigned | Unassigned | Unassigned | 701_0 | Clustered | 2 | VC_701_0 | 2 | 0.1487 | 0.95249915 | 0.1417 | 0 | 0 | 0 | 1 | Ref |
| 10047 | MN276192 | Unassigned | Unassigned | Unassigned | 558_40 | Clustered/Singleton | 1 | VC_558_40 | 1 | 0 | 1 | 0 | 0 | 0 | 0 | 1 | Ref |
| 10048 | MN284892 | Unassigned | Unassigned | Unassigned | 1344_0 | Clustered | 78 | VC_1344_0 | 78 | 0.9732 | 1 | 0.9732 | 0 | 0 | 0 | 1 | Ref |
| 10049 | MN284893 | Unassigned | Unassigned | Unassigned | 372_1 | Clustered | 9 | VC_372_1 | 9 | 0.194 | 1 | 0.194 | 0 | 0 | 0 | 1 | Ref |
| 10050 | MN284894 | Unassigned | Unassigned | Unassigned | 1224_0 | Clustered | 19 | VC_1224_0 | 19 | 1 | 1 | 1 | 0 | 0 | 0 | 1 | Ref |
| 10051 | MN284895 | Unassigned | Unassigned | Unassigned | 237_3 | Clustered | 8 | VC_237_3 | 8 | 0.1092 | 1 | 0.1092 | 0 | 0 | 0 | 1 | Ref |
| 10052 | MN284896 | Unassigned | Unassigned | Unassigned | 799_1 | Clustered | 27 | VC_799_1 | 27 | 0.8611 | 1 | 0.8611 | 0 | 0 | 0 | 1 | Ref |
| 10053 | MN284897 | Unassigned | Unassigned | Unassigned | 802_0 | Clustered | 17 | VC_802_0 | 17 | 0.8025 | 1 | 0.8025 | 0 | 0 | 0 | 1 | Ref |
| 10054 | MN284898 | Unassigned | Unassigned | Unassigned | 224_0 | Clustered | 19 | VC_224_0 | 19 | 0.9416 | 1 | 0.9416 | 0 | 0 | 0 | 1 | Ref |
| 10055 | MN284899 | Unassigned | Unassigned | Unassigned | 982_0 | Clustered | 62 | VC_982_0 | 62 | 0.8646 | 1 | 0.8646 | 0 | 0 | 0 | 0.743 | Ref |
| 10056 | MN284900 | Unassigned | Unassigned | Unassigned | 514_0 | Clustered | 75 | VC_514_0 | 75 | 0.9402 | 1 | 0.9402 | 0 | 0 | 0 | 1 | Ref |
| 10057 | MN284901 | Unassigned | Unassigned | Unassigned | 1163_0 | Clustered | 28 | VC_1163_0 | 28 | 0.4923 | 1 | 0.4923 | 0 | 0 | 0 | 1 | Ref |
| 10058 | MN284902 | Unassigned | Unassigned | Unassigned | 1164_0 | Clustered | 25 | VC_1164_0 | 25 | 0.7312 | 1 | 0.7312 | 0 | 0 | 0 | 1 | Ref |
| 10059 | MN284904 | Unassigned | Unassigned | Unassigned | 799_1 | Clustered | 27 | VC_799_1 | 27 | 0.8611 | 1 | 0.8611 | 0 | 0 | 0 | 1 | Ref |
| 10060 | MN284905 | Unassigned | Unassigned | Unassigned | 517_0 | Clustered | 18 | VC_517_0 | 18 | 0.1599 | 1 | 0.1599 | 0 | 0 | 0 | 1 | Ref |
| 10061 | MN284906 | Unassigned | Unassigned | Unassigned | 919_0 | Clustered | 39 | VC_919_0 | 39 | 0.9069 | 1 | 0.9069 | 0 | 0 | 0 | 1 | Ref |
| 10062 | MN284907 | Unassigned | Unassigned | Unassigned | 1178_0 | Clustered | 24 | VC_1178_0 | 24 | 0.9524 | 1 | 0.9524 | 0 | 0 | 0 | 1 | Ref |
| 10063 | MN284908 | Unassigned | Unassigned | Unassigned | 565_0 | Clustered | 20 | VC_565_0 | 20 | 0.8398 | 1 | 0.8398 | 0 | 0 | 0 | 1 | Ref |
| 10064 | MN294712 | Unassigned | Unassigned | Unassigned | 699_0 | Clustered | 71 | VC_699_0 | 71 | 0.9352 | 1 | 0.9352 | 0 | 0 | 0 | 1 | Ref |
| 10065 | MN296515 | Unassigned | Unassigned | Unassigned | 575_0 | Clustered | 35 | VC_575_0 | 35 | 0.8815 | 1 | 0.8815 | 0 | 0 | 0 | 1 | Ref |
| 10066 | MN304941 | Unassigned | Unassigned | Unassigned | 149_15 | Clustered | 16 | VC_149_15 | 16 | 0.0788 | 1 | 0.0788 | 0 | 0 | 0 | 0.8083 | Ref |
| 10067 | MN310541 | Unassigned | Unassigned | Unassigned | 220_0 | Clustered | 4 | VC_220_0 | 4 | 0.0615 | 0.99998802 | 0.0615 | 0 | 0 | 0 | 1 | Ref |
| 10068 | MN310542 | Unassigned | Unassigned | Unassigned | 225_0 | Clustered | 32 | VC_225_0 | 32 | 0.848 | 1 | 0.848 | 0 | 0 | 0 | 0.996 | Ref |
| 10069 | MN310543 | Unassigned | Unassigned | Unassigned | 1163_0 | Clustered | 28 | VC_1163_0 | 28 | 0.4923 | 1 | 0.4923 | 0 | 0 | 0 | 1 | Ref |
| 10070 | MN310544 | Unassigned | Unassigned | Unassigned | 520_75 | Clustered/Singleton | 1 | VC_520_75 | 1 | 0 | 1 | 0 | 0 | 0 | 0 | 1 | Ref |
| 10071 | MN310545 | Unassigned | Unassigned | Unassigned | 520_82 | Clustered/Singleton | 1 | VC_520_82 | 1 | 0 | 1 | 0 | 0 | 0 | 0 | 1 | Ref |
| 10072 | MN310546 | Unassigned | Unassigned | Unassigned | 1360_0 | Clustered | 31 | VC_1360_0 | 31 | 0.8837 | 1 | 0.8837 | 0 | 0 | 0 | 1 | Ref |
| 10073 | MN310547 | Unassigned | Unassigned | Unassigned | 221_0 | Clustered | 22 | VC_221_0 | 22 | 0.7447 | 1 | 0.7447 | 0 | 0 | 0 | 1 | Ref |
| 10074 | MN310548 | Unassigned | Unassigned | Unassigned | 520_7 | Clustered/Singleton | 1 | VC_520_7 | 1 | 0 | 1 | 0 | 0 | 0 | 0 | 1 | Ref |
| 10075 | MN310549 | Unassigned | Unassigned | Unassigned | 919_0 | Clustered | 39 | VC_919_0 | 39 | 0.9069 | 1 | 0.9069 | 0 | 0 | 0 | 1 | Ref |
| 10076 | MN311487 | Unassigned | Unassigned | Unassigned | 1262_0 | Clustered | 2 | VC_1262_0 | 2 | 0.1193 | 0.95613564 | 0.1141 | 0 | 0 | 0 | 1 | Ref |
| 10077 | MN311488 | Unassigned | Unassigned | Unassigned | 1062_0 | Clustered | 45 | VC_1062_0 | 45 | 0.9734 | 0.50949847 | 0.4959 | 0 | 0 | 0 | 1 | Ref |
| 10078 | MN311489 | Unassigned | Unassigned | Unassigned | 1062_0 | Clustered | 45 | VC_1062_0 | 45 | 0.9734 | 0.50949847 | 0.4959 | 0 | 0 | 0 | 1 | Ref |
| 10079 | MN311490 | Unassigned | Unassigned | Unassigned | 1062_0 | Clustered | 45 | VC_1062_0 | 45 | 0.9734 | 0.50949847 | 0.4959 | 0 | 0 | 0 | 1 | Ref |
| 10080 | MN311491 | Unassigned | Unassigned | Unassigned | 1062_0 | Clustered | 45 | VC_1062_0 | 45 | 0.9734 | 0.50949847 | 0.4959 | 0 | 0 | 0 | 1 | Ref |
| 10081 | MN311492 | Unassigned | Unassigned | Unassigned | 1262_0 | Clustered | 2 | VC_1262_0 | 2 | 0.1193 | 0.95613564 | 0.1141 | 0 | 0 | 0 | 1 | Ref |
| 10082 | MN311493 | Unassigned | Unassigned | Unassigned | 1243_0 | Clustered | 52 | VC_1243_0 | 52 | 0.9126 | 0.92708311 | 0.8461 | 0 | 0 | 0 | 1 | Ref |
| 10083 | MN313256 | Unassigned | Unassigned | Unassigned | 644_0 | Clustered | 5 | VC_644_0 | 5 | 0.6649 | 0.99999975 | 0.6649 | 0 | 0 | 0 | 1 | Ref |
| 10084 | MN316588 | Unassigned | Unassigned | Unassigned | 190_14 | Clustered | 3 | VC_190_14 | 3 | 0.0144 | 0.99696852 | 0.0143 | 0 | 0 | 0 | 0.6667 | Ref |
| 10085 | MN317029 | Unassigned | Unassigned | Unassigned | 597_0 | Clustered | 6 | VC_597_0 | 6 | 0.1801 | 1 | 0.1801 | 0 | 0 | 0 | 1 | Ref |
| 10086 | MN329673 | Unassigned | Unassigned | Unassigned | 1093_0 | Clustered | 40 | VC_1093_0 | 40 | 0.8754 | 1 | 0.8754 | 0 | 0 | 0 | 1 | Ref |
| 10087 | MN329674 | Unassigned | Unassigned | Unassigned | 939_0 | Clustered | 15 | VC_939_0 | 15 | 1 | 0 | 0 | 0 | 0 | 0 | 1 | Ref |
| 10088 | MN329675 | Unassigned | Unassigned | Unassigned | 1234_0 | Clustered | 44 | VC_1234_0 | 44 | 1 | 1 | 1 | 0 | 0 | 0 | 1 | Ref |
| 10089 | MN329676 | Unassigned | Unassigned | Unassigned | 939_0 | Clustered | 15 | VC_939_0 | 15 | 1 | 0 | 0 | 0 | 0 | 0 | 1 | Ref |
| 10090 | MN329677 | Unassigned | Unassigned | Unassigned | 1093_0 | Clustered | 40 | VC_1093_0 | 40 | 0.8754 | 1 | 0.8754 | 0 | 0 | 0 | 1 | Ref |
| 10091 | MN329678 | Unassigned | Unassigned | Unassigned | 765_0 | Clustered | 132 | VC_765_0 | 132 | 1 | 1 | 1 | 0 | 0 | 0 | 1 | Ref |
| 10092 | MN329679 | Unassigned | Unassigned | Unassigned | 1344_0 | Clustered | 78 | VC_1344_0 | 78 | 0.9732 | 1 | 0.9732 | 0 | 0 | 0 | 1 | Ref |
| 10093 | MN334766 | Unassigned | Unassigned | Unassigned | 488_2 | Clustered | 2 | VC_488_2 | 2 | 0.0325 | 0.95511029 | 0.0311 | 0 | 0 | 0 | 1 | Ref |
| 10094 | MN335248 | Unassigned | Unassigned | Unassigned | 177_0 | Clustered | 5 | VC_177_0 | 5 | 0.7468 | 0.99885646 | 0.746 | 0 | 0 | 0 | 1 | Ref |
| 10095 | MN336261 | Unassigned | Unassigned | Unassigned | 149_8 | Clustered | 66 | VC_149_8 | 66 | 0.4565 | 1 | 0.4565 | 0 | 0 | 0 | 0.8182 | Ref |
| 10096 | MN336262 | Unassigned | Unassigned | Unassigned | 149_8 | Clustered | 66 | VC_149_8 | 66 | 0.4565 | 1 | 0.4565 | 0 | 0 | 0 | 0.8182 | Ref |
| 10097 | MN336263 | Unassigned | Unassigned | Unassigned | 149_8 | Clustered | 66 | VC_149_8 | 66 | 0.4565 | 1 | 0.4565 | 0 | 0 | 0 | 0.8182 | Ref |
| 10098 | MN336264 | Unassigned | Unassigned | Unassigned | 637_0 | Clustered | 115 | VC_637_0 | 115 | 0.731 | 1 | 0.731 | 0 | 0 | 0 | 0.8764 | Ref |
| 10099 | MN336265 | Unassigned | Unassigned | Unassigned | 637_0 | Clustered | 115 | VC_637_0 | 115 | 0.731 | 1 | 0.731 | 0 | 0 | 0 | 0.8764 | Ref |
| 10100 | MN336266 | Unassigned | Unassigned | Unassigned | 637_0 | Clustered | 115 | VC_637_0 | 115 | 0.731 | 1 | 0.731 | 0 | 0 | 0 | 0.8764 | Ref |
| 10101 | MN337573 | Unassigned | Unassigned | Unassigned | 575_0 | Clustered | 35 | VC_575_0 | 35 | 0.8815 | 1 | 0.8815 | 0 | 0 | 0 | 1 | Ref |
| 10102 | MN337887 | Unassigned | Unassigned | Unassigned | 690_0 | Clustered | 50 | VC_690_0 | 50 | 0.9805 | 1 | 0.9805 | 0 | 0 | 0 | 1 | Ref |
| 10103 | MN340231 | Unassigned | Unassigned | Unassigned | 30_1 | Clustered | 16 | VC_30_1 | 16 | 0.1973 | 1 | 0.1973 | 0 | 0 | 0 | 0.8333 | Ref |
| 10104 | MN342150 | Unassigned | Unassigned | Unassigned | 133_22 | Clustered | 13 | VC_133_22 | 13 | 0.0804 | 1 | 0.0804 | 0 | 0 | 0 | 0.8205 | Ref |
| 10105 | MN342151 | Unassigned | Unassigned | Unassigned | 330_0 | Clustered | 27 | VC_330_0 | 27 | 0.5088 | 1 | 0.5088 | 0 | 0 | 0 | 0.8661 | Ref |
| 10106 | MN342247 | Unassigned | Unassigned | Unassigned | 575_0 | Clustered | 35 | VC_575_0 | 35 | 0.8815 | 1 | 0.8815 | 0 | 0 | 0 | 1 | Ref |
| 10107 | MN364664 | Unassigned | Unassigned | Unassigned | 187_4 | Clustered/Singleton | 1 | VC_187_4 | 1 | 0 | 1 | 0 | 0 | 0 | 0 | 1 | Ref |
| 10108 | MN369737 | Unassigned | Unassigned | Unassigned | 1240_0 | Clustered | 82 | VC_1240_0 | 82 | 0.7945 | 1 | 0.7945 | 0 | 0 | 0 | 1 | Ref |
| 10109 | MN369738 | Unassigned | Unassigned | Unassigned | 371_14 | Clustered/Singleton | 1 | VC_371_14 | 1 | 0 | 1 | 0 | 0 | 0 | 0 | 1 | Ref |
| 10110 | MN369739 | Unassigned | Unassigned | Unassigned | 520_17 | Clustered/Singleton | 1 | VC_520_17 | 1 | 0 | 1 | 0 | 0 | 0 | 0 | 1 | Ref |
| 10111 | MN369740 | Unassigned | Unassigned | Unassigned | 1176_0 | Clustered | 41 | VC_1176_0 | 41 | 0.9558 | 1 | 0.9558 | 0 | 0 | 0 | 0.9976 | Ref |
| 10112 | MN369741 | Unassigned | Unassigned | Unassigned | 225_0 | Clustered | 32 | VC_225_0 | 32 | 0.848 | 1 | 0.848 | 0 | 0 | 0 | 0.996 | Ref |
| 10113 | MN369742 | Unassigned | Unassigned | Unassigned | 236_0 | Clustered | 72 | VC_236_0 | 72 | 0.8104 | 1 | 0.8104 | 0 | 0 | 0 | 1 | Ref |
| 10114 | MN369743 | Unassigned | Unassigned | Unassigned | 1071_3 | Clustered | 16 | VC_1071_3 | 16 | 0.2354 | 1 | 0.2354 | 0 | 0 | 0 | 0.9667 | Ref |
| 10115 | MN369744 | Unassigned | Unassigned | Unassigned | 371_11 | Clustered | 31 | VC_371_11 | 31 | 0.0927 | 1 | 0.0927 | 0 | 0 | 0 | 0.7011 | Ref |
| 10116 | MN369745 | Unassigned | Unassigned | Unassigned | 1344_0 | Clustered | 78 | VC_1344_0 | 78 | 0.9732 | 1 | 0.9732 | 0 | 0 | 0 | 1 | Ref |
| 10117 | MN369746 | Unassigned | Unassigned | Unassigned | 236_0 | Clustered | 72 | VC_236_0 | 72 | 0.8104 | 1 | 0.8104 | 0 | 0 | 0 | 1 | Ref |
| 10118 | MN369747 | Unassigned | Unassigned | Unassigned | 1360_0 | Clustered | 31 | VC_1360_0 | 31 | 0.8837 | 1 | 0.8837 | 0 | 0 | 0 | 1 | Ref |
| 10119 | MN369748 | Unassigned | Unassigned | Unassigned | 216_0 | Clustered | 4 | VC_216_0 | 4 | 0.048 | 0.9999878 | 0.048 | 0 | 0 | 0 | 1 | Ref |
| 10120 | MN369749 | Unassigned | Unassigned | Unassigned | 520_19 | Clustered | 2 | VC_520_19 | 2 | 0.0098 | 0.95761552 | 0.0094 | 0 | 0 | 0 | 1 | Ref |
| 10121 | MN369750 | Unassigned | Unassigned | Unassigned | 1071_0 | Clustered | 20 | VC_1071_0 | 20 | 0.3268 | 1 | 0.3268 | 0 | 0 | 0 | 0.9842 | Ref |
| 10122 | MN369751 | Unassigned | Unassigned | Unassigned | 520_85 | Clustered | 2 | VC_520_85 | 2 | 0.0148 | 0.95762405 | 0.0142 | 0 | 0 | 0 | 1 | Ref |
| 10123 | MN369752 | Unassigned | Unassigned | Unassigned | 520_10 | Clustered/Singleton | 1 | VC_520_10 | 1 | 0 | 1 | 0 | 0 | 0 | 0 | 1 | Ref |
| 10124 | MN369753 | Unassigned | Unassigned | Unassigned | 767_0 | Clustered | 34 | VC_767_0 | 34 | 0.9173 | 1 | 0.9173 | 0 | 0 | 0 | 1 | Ref |
| 10125 | MN369754 | Unassigned | Unassigned | Unassigned | 1072_4 | Clustered | 4 | VC_1072_4 | 4 | 0.0736 | 0.99956559 | 0.0736 | 0 | 0 | 0 | 1 | Ref |
| 10126 | MN369755 | Unassigned | Unassigned | Unassigned | 520_33 | Clustered | 2 | VC_520_33 | 2 | 0.0087 | 0.95761571 | 0.0083 | 0 | 0 | 0 | 1 | Ref |
| 10127 | MN369756 | Unassigned | Unassigned | Unassigned | 224_0 | Clustered | 19 | VC_224_0 | 19 | 0.9416 | 1 | 0.9416 | 0 | 0 | 0 | 1 | Ref |
| 10128 | MN369757 | Unassigned | Unassigned | Unassigned | 1071_0 | Clustered | 20 | VC_1071_0 | 20 | 0.3268 | 1 | 0.3268 | 0 | 0 | 0 | 0.9842 | Ref |
| 10129 | MN369758 | Unassigned | Unassigned | Unassigned | 510_0 | Clustered | 12 | VC_510_0 | 12 | 0.3963 | 1 | 0.3963 | 0 | 0 | 0 | 0.9697 | Ref |
| 10130 | MN369759 | Unassigned | Unassigned | Unassigned | 372_0 | Clustered | 24 | VC_372_0 | 24 | 0.5539 | 1 | 0.5539 | 0 | 0 | 0 | 1 | Ref |
| 10131 | MN369760 | Unassigned | Unassigned | Unassigned | 325_36 | Clustered | 2 | VC_325_36 | 2 | 0.0043 | 0.95751115 | 0.0041 | 0 | 0 | 0 | 1 | Ref |
| 10132 | MN369761 | Unassigned | Unassigned | Unassigned | 234_0 | Clustered | 21 | VC_234_0 | 21 | 0.8355 | 1 | 0.8355 | 0 | 0 | 0 | 1 | Ref |
| 10133 | MN369762 | Unassigned | Unassigned | Unassigned | 237_2 | Clustered | 11 | VC_237_2 | 11 | 0.1409 | 1 | 0.1409 | 0 | 0 | 0 | 0.9818 | Ref |
| 10134 | MN369763 | Unassigned | Unassigned | Unassigned | 225_0 | Clustered | 32 | VC_225_0 | 32 | 0.848 | 1 | 0.848 | 0 | 0 | 0 | 0.996 | Ref |
| 10135 | MN369764 | Unassigned | Unassigned | Unassigned | 225_0 | Clustered | 32 | VC_225_0 | 32 | 0.848 | 1 | 0.848 | 0 | 0 | 0 | 0.996 | Ref |
| 10136 | MN369765 | Unassigned | Unassigned | Unassigned | 1162_0 | Clustered | 10 | VC_1162_0 | 10 | 0.5715 | 1 | 0.5715 | 0 | 0 | 0 | 1 | Ref |
| 10137 | MN369766 | Unassigned | Unassigned | Unassigned | 767_0 | Clustered | 34 | VC_767_0 | 34 | 0.9173 | 1 | 0.9173 | 0 | 0 | 0 | 1 | Ref |
| 10138 | MN379460 | Unassigned | Unassigned | Unassigned | 121_0 | Clustered | 4 | VC_121_0 | 4 | 0.1814 | 0.99998531 | 0.1814 | 0 | 0 | 0 | 1 | Ref |
| 10139 | MN379629 | Unassigned | Unassigned | Unassigned | 1409_0 | Clustered | 2 | VC_1409_0 | 2 | 1 | 1 | 1 | 0 | 0 | 0 | 1 | Ref |
| 10140 | MN379630 | Unassigned | Unassigned | Unassigned | 1122_0 | Clustered | 10 | VC_1122_0 | 10 | 0.9643 | 0.99999697 | 0.9643 | 0 | 0 | 0 | 1 | Ref |
| 10141 | MN379632 | Unassigned | Unassigned | Unassigned | 1081_0 | Clustered/Singleton | 1 | VC_1081_0 | 1 | 0 | 1 | 0 | 0 | 0 | 0 | 1 | Ref |
| 10142 | MN379633 | Unassigned | Unassigned | Unassigned | 1054_0 | Clustered | 12 | VC_1054_0 | 12 | 0.2863 | 0.99746644 | 0.2855 | 0 | 0 | 0 | 1 | Ref |
| 10143 | MN379636 | Unassigned | Unassigned | Unassigned | 1409_0 | Clustered | 2 | VC_1409_0 | 2 | 1 | 1 | 1 | 0 | 0 | 0 | 1 | Ref |
| 10144 | MN379637 | Unassigned | Unassigned | Unassigned | 1233_0 | Clustered | 16 | VC_1233_0 | 16 | 0.8971 | 1 | 0.8971 | 0 | 0 | 0 | 1 | Ref |
| 10145 | MN379638 | Unassigned | Unassigned | Unassigned | 1277_0 | Clustered | 14 | VC_1277_0 | 14 | 0.8218 | 1 | 0.8218 | 0 | 0 | 0 | 1 | Ref |
| 10146 | MN379639 | Unassigned | Unassigned | Unassigned | 1410_0 | Clustered | 2 | VC_1410_0 | 2 | 1 | 1 | 1 | 0 | 0 | 0 | 1 | Ref |
| 10147 | MN379640 | Unassigned | Unassigned | Unassigned | 1122_0 | Clustered | 10 | VC_1122_0 | 10 | 0.9643 | 0.99999697 | 0.9643 | 0 | 0 | 0 | 1 | Ref |
| 10148 | MN379641 | Unassigned | Unassigned | Unassigned | 1153_0 | Clustered | 2 | VC_1153_0 | 2 | 0.073 | 0.80562642 | 0.0588 | 0 | 0 | 0 | 1 | Ref |
| 10149 | MN379643 | Unassigned | Unassigned | Unassigned | 1411_0 | Clustered | 2 | VC_1411_0 | 2 | 1 | 1 | 1 | 0 | 0 | 0 | 1 | Ref |
| 10150 | MN379645 | Unassigned | Unassigned | Unassigned | 1275_0 | Clustered | 4 | VC_1275_0 | 4 | 1 | 1 | 1 | 0 | 0 | 0 | 1 | Ref |
| 10151 | MN379646 | Unassigned | Unassigned | Unassigned | 1410_0 | Clustered | 2 | VC_1410_0 | 2 | 1 | 1 | 1 | 0 | 0 | 0 | 1 | Ref |
| 10152 | MN379647 | Unassigned | Unassigned | Unassigned | 1233_0 | Clustered | 16 | VC_1233_0 | 16 | 0.8971 | 1 | 0.8971 | 0 | 0 | 0 | 1 | Ref |
| 10153 | MN379648 | Unassigned | Unassigned | Unassigned | 1277_0 | Clustered | 14 | VC_1277_0 | 14 | 0.8218 | 1 | 0.8218 | 0 | 0 | 0 | 1 | Ref |
| 10154 | MN379650 | Unassigned | Unassigned | Unassigned | 1411_0 | Clustered | 2 | VC_1411_0 | 2 | 1 | 1 | 1 | 0 | 0 | 0 | 1 | Ref |
| 10155 | MN379651 | Unassigned | Unassigned | Unassigned | 1054_0 | Clustered | 12 | VC_1054_0 | 12 | 0.2863 | 0.99746644 | 0.2855 | 0 | 0 | 0 | 1 | Ref |
| 10156 | MN379739 | Unassigned | Unassigned | Unassigned | 191_16 | Clustered | 12 | VC_191_16 | 12 | 0.0643 | 1 | 0.0643 | 0 | 0 | 0 | 0.9394 | Ref |
| 10157 | MN379740 | Unassigned | Unassigned | Unassigned | 428_0 | Clustered | 33 | VC_428_0 | 33 | 0.9915 | 1 | 0.9915 | 0 | 0 | 0 | 1 | Ref |
| 10158 | MN379832 | Unassigned | Unassigned | Unassigned | 938_0 | Clustered | 99 | VC_938_0 | 99 | 0.9119 | 1 | 0.9119 | 0 | 0 | 0 | 0.9928 | Ref |
| 10159 | MN380459 | Unassigned | Unassigned | Unassigned | 111_0 | Clustered | 91 | VC_111_0 | 91 | 0.9587 | 1 | 0.9587 | 0 | 0 | 0 | 1 | Ref |
| 10160 | MN384978 | Unassigned | Unassigned | Unassigned | 452_0 | Clustered | 66 | VC_452_0 | 66 | 0.9248 | 1 | 0.9248 | 0 | 0 | 0 | 1 | Ref |
| 10161 | MN384979 | Unassigned | Unassigned | Unassigned | 133_9 | Clustered | 3 | VC_133_9 | 3 | 0.0099 | 0.98457109 | 0.0098 | 0 | 0 | 0 | 1 | Ref |
| 10162 | MN385565 | Unassigned | Unassigned | Unassigned | 303_0 | Clustered | 155 | VC_303_0 | 155 | 0.9979 | 1 | 0.9979 | 0 | 0 | 0 | 1 | Ref |
| 10163 | MN393078 | Unassigned | Unassigned | Unassigned | 637_0 | Clustered | 115 | VC_637_0 | 115 | 0.731 | 1 | 0.731 | 0 | 0 | 0 | 0.8764 | Ref |
| 10164 | MN393079 | Unassigned | Unassigned | Unassigned | 540_0 | Clustered | 15 | VC_540_0 | 15 | 0.2866 | 1 | 0.2866 | 0 | 0 | 0 | 1 | Ref |
| 10165 | MN393473 | Unassigned | Unassigned | Unassigned | 575_0 | Clustered | 35 | VC_575_0 | 35 | 0.8815 | 1 | 0.8815 | 0 | 0 | 0 | 1 | Ref |
| 10166 | MN395285 | Unassigned | Unassigned | Unassigned | 938_0 | Clustered | 99 | VC_938_0 | 99 | 0.9119 | 1 | 0.9119 | 0 | 0 | 0 | 0.9928 | Ref |
| 10167 | MN395286 | Unassigned | Unassigned | Unassigned | 938_0 | Clustered | 99 | VC_938_0 | 99 | 0.9119 | 1 | 0.9119 | 0 | 0 | 0 | 0.9928 | Ref |
| 10168 | MN395291 | Unassigned | Unassigned | Unassigned | 699_0 | Clustered | 71 | VC_699_0 | 71 | 0.9352 | 1 | 0.9352 | 0 | 0 | 0 | 1 | Ref |
| 10169 | MN402506 | Unassigned | Unassigned | Unassigned | 495_1 | Clustered | 41 | VC_495_1 | 41 | 0.328 | 1 | 0.328 | 0 | 0 | 0 | 0.911 | Ref |
| 10170 | MN414250 | Unassigned | Unassigned | Unassigned | 451_0 | Clustered | 80 | VC_451_0 | 80 | 0.9991 | 1 | 0.9991 | 0 | 0 | 0 | 1 | Ref |
| 10171 | MN417334 | Unassigned | Unassigned | Unassigned | 1022_0 | Clustered | 4 | VC_1022_0 | 4 | 0.8047 | 0.99996089 | 0.8046 | 0 | 0 | 0 | 1 | Ref |
| 10172 | MN419153 | Unassigned | Unassigned | Unassigned | 495_0 | Clustered | 39 | VC_495_0 | 39 | 0.308 | 1 | 0.308 | 0 | 0 | 0 | 1 | Ref |
| 10173 | MN428049 | Unassigned | Unassigned | Unassigned | 508_0 | Clustered | 32 | VC_508_0 | 32 | 0.906 | 1 | 0.906 | 0 | 0 | 0 | 1 | Ref |
| 10174 | MN428050 | Unassigned | Unassigned | Unassigned | 372_0 | Clustered | 24 | VC_372_0 | 24 | 0.5539 | 1 | 0.5539 | 0 | 0 | 0 | 1 | Ref |
| 10175 | MN428051 | Unassigned | Unassigned | Unassigned | 520_92 | Clustered | 5 | VC_520_92 | 5 | 0.0663 | 0.99999997 | 0.0663 | 0 | 0 | 0 | 1 | Ref |
| 10176 | MN428052 | Unassigned | Unassigned | Unassigned | 556_0 | Clustered | 21 | VC_556_0 | 21 | 0.9912 | 1 | 0.9912 | 0 | 0 | 0 | 1 | Ref |
| 10177 | MN428053 | Unassigned | Unassigned | Unassigned | 522_1 | Clustered | 5 | VC_522_1 | 5 | 0.1017 | 0.99999997 | 0.1017 | 0 | 0 | 0 | 0.8 | Ref |
| 10178 | MN428054 | Unassigned | Unassigned | Unassigned | 325_35 | Clustered | 17 | VC_325_35 | 17 | 0.0732 | 1 | 0.0732 | 0 | 0 | 0 | 0.7721 | Ref |
| 10179 | MN428056 | Unassigned | Unassigned | Unassigned | 939_0 | Clustered | 15 | VC_939_0 | 15 | 1 | 0 | 0 | 0 | 0 | 0 | 1 | Ref |
| 10180 | MN428058 | Unassigned | Unassigned | Unassigned | 556_0 | Clustered | 21 | VC_556_0 | 21 | 0.9912 | 1 | 0.9912 | 0 | 0 | 0 | 1 | Ref |
| 10181 | MN428059 | Unassigned | Unassigned | Unassigned | 551_6 | Clustered | 22 | VC_551_6 | 22 | 0.1214 | 1 | 0.1214 | 0 | 0 | 0 | 0.8095 | Ref |
| 10182 | MN428060 | Unassigned | Unassigned | Unassigned | 1071_0 | Clustered | 20 | VC_1071_0 | 20 | 0.3268 | 1 | 0.3268 | 0 | 0 | 0 | 0.9842 | Ref |
| 10183 | MN428061 | Unassigned | Unassigned | Unassigned | 939_0 | Clustered | 15 | VC_939_0 | 15 | 1 | 0 | 0 | 0 | 0 | 0 | 1 | Ref |
| 10184 | MN428062 | Unassigned | Unassigned | Unassigned | 1344_0 | Clustered | 78 | VC_1344_0 | 78 | 0.9732 | 1 | 0.9732 | 0 | 0 | 0 | 1 | Ref |
| 10185 | MN428063 | Unassigned | Unassigned | Unassigned | 767_0 | Clustered | 34 | VC_767_0 | 34 | 0.9173 | 1 | 0.9173 | 0 | 0 | 0 | 1 | Ref |
| 10186 | MN428064 | Unassigned | Unassigned | Unassigned | 1344_0 | Clustered | 78 | VC_1344_0 | 78 | 0.9732 | 1 | 0.9732 | 0 | 0 | 0 | 1 | Ref |
| 10187 | MN428065 | Unassigned | Unassigned | Unassigned | 558_38 | Clustered | 2 | VC_558_38 | 2 | 0.0032 | 0.64328629 | 0.002 | 0 | 0 | 0 | 1 | Ref |
| 10188 | MN428066 | Unassigned | Unassigned | Unassigned | 558_20 | Clustered | 17 | VC_558_20 | 17 | 0.054 | 1 | 0.054 | 0 | 0 | 0 | 0.9926 | Ref |
| 10189 | MN432485 | Unassigned | Unassigned | Unassigned | 941_0 | Clustered | 40 | VC_941_0 | 40 | 0.6903 | 1 | 0.6903 | 0 | 0 | 0 | 1 | Ref |
| 10190 | MN433707 | Unassigned | Unassigned | Unassigned | 699_0 | Clustered | 71 | VC_699_0 | 71 | 0.9352 | 1 | 0.9352 | 0 | 0 | 0 | 1 | Ref |
| 10191 | MN434093 | Unassigned | Unassigned | Unassigned | 823_0 | Clustered | 124 | VC_823_0 | 124 | 0.9492 | 1 | 0.9492 | 0 | 0 | 0 | 1 | Ref |
| 10192 | MN434094 | Unassigned | Unassigned | Unassigned | 192_0 | Clustered | 15 | VC_192_0 | 15 | 0.5071 | 1 | 0.5071 | 0 | 0 | 0 | 1 | Ref |
| 10193 | MN434095 | Unassigned | Unassigned | Unassigned | 491_0 | Clustered | 36 | VC_491_0 | 36 | 0.5336 | 1 | 0.5336 | 0 | 0 | 0 | 0.9714 | Ref |
| 10194 | MN434096 | Unassigned | Unassigned | Unassigned | 192_0 | Clustered | 15 | VC_192_0 | 15 | 0.5071 | 1 | 0.5071 | 0 | 0 | 0 | 1 | Ref |
| 10195 | MN444868 | Unassigned | Unassigned | Unassigned | 371_3 | Clustered | 22 | VC_371_3 | 22 | 0.0787 | 1 | 0.0787 | 0 | 0 | 0 | 0.9048 | Ref |
| 10196 | MN444869 | Unassigned | Unassigned | Unassigned | 325_6 | Clustered/Singleton | 1 | VC_325_6 | 1 | 0 | 1 | 0 | 0 | 0 | 0 | 1 | Ref |
| 10197 | MN444870 | Unassigned | Unassigned | Unassigned | 890_1 | Clustered | 4 | VC_890_1 | 4 | 0.4613 | 0.99998458 | 0.4613 | 0 | 0 | 0 | 1 | Ref |
| 10198 | MN444871 | Unassigned | Unassigned | Unassigned | 919_0 | Clustered | 39 | VC_919_0 | 39 | 0.9069 | 1 | 0.9069 | 0 | 0 | 0 | 1 | Ref |
| 10199 | MN444872 | Unassigned | Unassigned | Unassigned | 509_0 | Clustered | 37 | VC_509_0 | 37 | 0.9689 | 1 | 0.9689 | 0 | 0 | 0 | 1 | Ref |
| 10200 | MN444873 | Unassigned | Unassigned | Unassigned | 509_0 | Clustered | 37 | VC_509_0 | 37 | 0.9689 | 1 | 0.9689 | 0 | 0 | 0 | 1 | Ref |
| 10201 | MN444874 | Unassigned | Unassigned | Unassigned | 868_0 | Clustered | 25 | VC_868_0 | 25 | 0.5791 | 1 | 0.5791 | 0 | 0 | 0 | 1 | Ref |
| 10202 | MN444875 | Unassigned | Unassigned | Unassigned | 761_0 | Clustered | 67 | VC_761_0 | 67 | 0.8947 | 1 | 0.8947 | 0 | 0 | 0 | 1 | Ref |
| 10203 | MN444876 | Unassigned | Unassigned | Unassigned | 1071_4 | Clustered/Singleton | 1 | VC_1071_4 | 1 | 0 | 1 | 0 | 0 | 0 | 0 | 1 | Ref |
| 10204 | MN444877 | Unassigned | Unassigned | Unassigned | 1352_0 | Clustered | 31 | VC_1352_0 | 31 | 0.849 | 1 | 0.849 | 0 | 0 | 0 | 1 | Ref |

| | | | | | | | | | | | | | | | | | |
|---|---|---|---|---|---|---|---|---|---|---|---|---|---|---|---|---|---|
| 10205 | MN444878 | Unassigned | Unassigned | Unassigned | 919_0 | Clustered | 39 | VC_919_0 | 39 | 0.9069 | 1 | 0.9069 | 0 | 0 | 0 | 1 | Ref |
| 10206 | MN445182 | Unassigned | Unassigned | Unassigned | 190_34 | Clustered/Singleton | 1 | VC_190_34 | 1 | 0 | 1 | 0 | 0 | 0 | 0 | 1 | Ref |
| 10207 | MN445183 | Unassigned | Unassigned | Unassigned | 489_0 | Clustered | 3 | VC_489_0 | 3 | 0.0677 | 0.98259158 | 0.0666 | 0 | 0 | 0 | 1 | Ref |
| 10208 | MN445184 | Unassigned | Unassigned | Unassigned | 133_42 | Clustered/Singleton | 1 | VC_133_42 | 1 | 0 | 1 | 0 | 0 | 0 | 0 | 1 | Ref |
| 10209 | MN445185 | Unassigned | Unassigned | Unassigned | 959_5 | Clustered | 19 | VC_959_5 | 19 | 0.2626 | 1 | 0.2626 | 0 | 0 | 0 | 1 | Ref |
| 10210 | MN450150 | Unassigned | Unassigned | Unassigned | 387_0 | Clustered | 3 | VC_387_0 | 3 | 0.0424 | 0.99861589 | 0.0423 | 0 | 0 | 0 | 1 | Ref |
| 10211 | MN453779 | Unassigned | Unassigned | Unassigned | 442_3 | Clustered | 3 | VC_442_3 | 3 | 0.0529 | 0.99675181 | 0.0527 | 0 | 0 | 0 | 1 | Ref |
| 10212 | MN461279 | Unassigned | Unassigned | Unassigned | 364_0 | Clustered | 16 | VC_364_0 | 16 | 0.6491 | 1 | 0.6491 | 0 | 0 | 0 | 1 | Ref |
| 10213 | MN478375 | Unassigned | Unassigned | Unassigned | 74_0 | Clustered | 15 | VC_74_0 | 15 | 0.5204 | 1 | 0.5204 | 0 | 0 | 0 | 1 | Ref |
| 10214 | MN478377 | Unassigned | Unassigned | Unassigned | 192_2 | Clustered/Singleton | 1 | VC_192_2 | 1 | 0 | 1 | 0 | 0 | 0 | 0 | 1 | Ref |
| 10215 | MN478483 | Unassigned | Unassigned | Unassigned | 132_1 | Clustered | 17 | VC_132_1 | 17 | 0.1738 | 1 | 0.1738 | 0 | 0 | 0 | 0.9853 | Ref |
| 10216 | MN481365 | Unassigned | Unassigned | Unassigned | 392_0 | Clustered | 126 | VC_392_0 | 126 | 0.9621 | 1 | 0.9621 | 0 | 0 | 0 | 1 | Ref |
| 10217 | MN481366 | Unassigned | Unassigned | Unassigned | 392_0 | Clustered | 126 | VC_392_0 | 126 | 0.9621 | 1 | 0.9621 | 0 | 0 | 0 | 1 | Ref |
| 10218 | MN481367 | Unassigned | Unassigned | Unassigned | 329_11 | Clustered | 15 | VC_329_11 | 15 | 0.0791 | 1 | 0.0791 | 0 | 0 | 0 | 0.9333 | Ref |
| 10219 | MN484599 | Unassigned | Unassigned | Unassigned | 1071_0 | Clustered | 20 | VC_1071_0 | 20 | 0.3268 | 1 | 0.3268 | 0 | 0 | 0 | 0.9842 | Ref |
| 10220 | MN484600 | Unassigned | Unassigned | Unassigned | 1166_0 | Clustered | 20 | VC_1166_0 | 20 | 0.99 | 1 | 0.99 | 0 | 0 | 0 | 1 | Ref |
| 10221 | MN484601 | Unassigned | Unassigned | Unassigned | 1098_0 | Clustered | 4 | VC_1098_0 | 4 | 0.5465 | 0.99996462 | 0.5465 | 0 | 0 | 0 | 1 | Ref |
| 10222 | MN496305 | Unassigned | Unassigned | Unassigned | 599_0 | Clustered | 50 | VC_599_0 | 50 | 0.9989 | 1 | 0.9989 | 0 | 0 | 0 | 1 | Ref |
| 10223 | MN496306 | Unassigned | Unassigned | Unassigned | 599_0 | Clustered | 50 | VC_599_0 | 50 | 0.9989 | 1 | 0.9989 | 0 | 0 | 0 | 1 | Ref |
| 10224 | MN496307 | Unassigned | Unassigned | Unassigned | 599_0 | Clustered | 50 | VC_599_0 | 50 | 0.9989 | 1 | 0.9989 | 0 | 0 | 0 | 1 | Ref |
| 10225 | MN496308 | Unassigned | Unassigned | Unassigned | 599_0 | Clustered | 50 | VC_599_0 | 50 | 0.9989 | 1 | 0.9989 | 0 | 0 | 0 | 1 | Ref |
| 10226 | MN497953 | Unassigned | Unassigned | Unassigned | 514_0 | Clustered | 75 | VC_514_0 | 75 | 0.9402 | 1 | 0.9402 | 0 | 0 | 0 | 1 | Ref |
| 10227 | MN497954 | Unassigned | Unassigned | Unassigned | 1343_0 | Clustered | 21 | VC_1343_0 | 21 | 0.9843 | 1 | 0.9843 | 0 | 0 | 0 | 1 | Ref |
| 10228 | MN504636 | Unassigned | Unassigned | Unassigned | 465_0 | Clustered | 7 | VC_465_0 | 7 | 0.3322 | 1 | 0.3322 | 0 | 0 | 0 | 1 | Ref |
| 10229 | MN508614 | Unassigned | Unassigned | Unassigned | 283_0 | Clustered | 2 | VC_283_0 | 2 | 0.0025 | 0.94936553 | 0.0024 | 0 | 0 | 0 | 1 | Ref |
| 10230 | MN508615 | Unassigned | Unassigned | Unassigned | 813_1 | Clustered | 24 | VC_813_1 | 24 | 0.535 | 1 | 0.535 | 0 | 0 | 0 | 1 | Ref |
| 10231 | MN508616 | Unassigned | Unassigned | Unassigned | 283_110 | Clustered | 2 | VC_283_110 | 2 | 0.0027 | 0.90910416 | 0.0024 | 0 | 0 | 0 | 1 | Ref |
| 10232 | MN508617 | Unassigned | Unassigned | Unassigned | 283_110 | Clustered | 2 | VC_283_110 | 2 | 0.0027 | 0.90910416 | 0.0024 | 0 | 0 | 0 | 1 | Ref |
| 10233 | MN508618 | Unassigned | Unassigned | Unassigned | 283_0 | Clustered | 2 | VC_283_0 | 2 | 0.0025 | 0.94936553 | 0.0024 | 0 | 0 | 0 | 1 | Ref |
| 10234 | MN508619 | Unassigned | Unassigned | Unassigned | 472_0 | Clustered | 47 | VC_472_0 | 47 | 0.9025 | 1 | 0.9025 | 0 | 0 | 0 | 1 | Ref |
| 10235 | MN508620 | Unassigned | Unassigned | Unassigned | 80_0 | Clustered | 41 | VC_80_0 | 41 | 0.5837 | 1 | 0.5837 | 0 | 0 | 0 | 0.9805 | Ref |
| 10236 | MN508621 | Unassigned | Unassigned | Unassigned | 669_0 | Clustered | 21 | VC_669_0 | 21 | 0.3623 | 1 | 0.3623 | 0 | 0 | 0 | 1 | Ref |
| 10237 | MN508622 | Unassigned | Unassigned | Unassigned | 669_0 | Clustered | 21 | VC_669_0 | 21 | 0.3623 | 1 | 0.3623 | 0 | 0 | 0 | 1 | Ref |
| 10238 | MN508623 | Unassigned | Unassigned | Unassigned | 669_0 | Clustered | 21 | VC_669_0 | 21 | 0.3623 | 1 | 0.3623 | 0 | 0 | 0 | 1 | Ref |
| 10239 | MN508624 | Unassigned | Unassigned | Unassigned | 669_0 | Clustered | 21 | VC_669_0 | 21 | 0.3623 | 1 | 0.3623 | 0 | 0 | 0 | 1 | Ref |
| 10240 | MN508817 | Unassigned | Unassigned | Unassigned | 669_2 | Clustered/Singleton | 1 | VC_669_2 | 1 | 0 | 1 | 0 | 0 | 0 | 0 | 1 | Ref |
| 10241 | MN509793 | Unassigned | Unassigned | Unassigned | 878_0 | Clustered | 32 | VC_878_0 | 32 | 0.9965 | 1 | 0.9965 | 0 | 0 | 0 | 1 | Ref |
| 10242 | MN510331 | Unassigned | Unassigned | Unassigned | 451_0 | Clustered | 80 | VC_451_0 | 80 | 0.9991 | 1 | 0.9991 | 0 | 0 | 0 | 1 | Ref |
| 10243 | MN512538 | Unassigned | Unassigned | Unassigned | 354_0 | Clustered | 4 | VC_354_0 | 4 | 0.1749 | 0.99998684 | 0.1749 | 0 | 0 | 0 | 0.8333 | Ref |
| 10244 | MN516421 | Unassigned | Unassigned | Unassigned | 698_0 | Clustered | 26 | VC_698_0 | 26 | 0.9366 | 1 | 0.9366 | 0 | 0 | 0 | 1 | Ref |
| 10245 | MN516422 | Unassigned | Unassigned | Unassigned | 698_0 | Clustered | 26 | VC_698_0 | 26 | 0.9366 | 1 | 0.9366 | 0 | 0 | 0 | 1 | Ref |
| 10246 | MN518893 | Unassigned | Unassigned | Unassigned | 451_0 | Clustered | 80 | VC_451_0 | 80 | 0.9991 | 1 | 0.9991 | 0 | 0 | 0 | 1 | Ref |
| 10247 | MN518894 | Unassigned | Unassigned | Unassigned | 451_0 | Clustered | 80 | VC_451_0 | 80 | 0.9991 | 1 | 0.9991 | 0 | 0 | 0 | 1 | Ref |
| 10248 | MN528768 | Unassigned | Unassigned | Unassigned | 534_0 | Clustered | 9 | VC_534_0 | 9 | 0.99 | 1 | 0.99 | 0 | 0 | 0 | 1 | Ref |
| 10249 | MN534316 | Unassigned | Unassigned | Unassigned | 260_0 | Clustered | 13 | VC_260_0 | 13 | 0.3705 | 1 | 0.3705 | 0 | 0 | 0 | 1 | Ref |
| 10250 | MN534317 | Unassigned | Unassigned | Unassigned | 260_0 | Clustered | 13 | VC_260_0 | 13 | 0.3705 | 1 | 0.3705 | 0 | 0 | 0 | 1 | Ref |
| 10251 | MN534320 | Unassigned | Unassigned | Unassigned | 262_0 | Clustered | 5 | VC_262_0 | 5 | 0.0724 | 0.99999989 | 0.0724 | 0 | 0 | 0 | 1 | Ref |
| 10252 | MN536023 | Unassigned | Unassigned | Unassigned | 16_0 | Clustered | 12 | VC_16_0 | 12 | 0.6615 | 1 | 0.6615 | 0 | 0 | 0 | 1 | Ref |
| 10253 | MN536027 | Unassigned | Unassigned | Unassigned | 251_0 | Clustered | 22 | VC_251_0 | 22 | 0.6508 | 1 | 0.6508 | 0 | 0 | 0 | 1 | Ref |
| 10254 | MN539736 | Unassigned | Unassigned | Unassigned | 149_8 | Clustered | 66 | VC_149_8 | 66 | 0.4565 | 1 | 0.4565 | 0 | 0 | 0 | 0.8182 | Ref |
| 10255 | MN539737 | Unassigned | Unassigned | Unassigned | 149_8 | Clustered | 66 | VC_149_8 | 66 | 0.4565 | 1 | 0.4565 | 0 | 0 | 0 | 0.8182 | Ref |
| 10256 | MN539738 | Unassigned | Unassigned | Unassigned | 149_8 | Clustered | 66 | VC_149_8 | 66 | 0.4565 | 1 | 0.4565 | 0 | 0 | 0 | 0.8182 | Ref |
| 10257 | MN549360 | Unassigned | Unassigned | Unassigned | 1052_3 | Clustered | 5 | VC_1052_3 | 5 | 0.185 | 0.98656048 | 0.1825 | 0 | 0 | 0 | 1 | Ref |
| 10258 | MN549361 | Unassigned | Unassigned | Unassigned | 1052_3 | Clustered | 5 | VC_1052_3 | 5 | 0.185 | 0.98656048 | 0.1825 | 0 | 0 | 0 | 1 | Ref |
| 10259 | MN552143 | Unassigned | Unassigned | Unassigned | 800_0 | Clustered | 3 | VC_800_0 | 3 | 0.4591 | 0.9984737 | 0.4584 | 0 | 0 | 0 | 1 | Ref |
| 10260 | MN552144 | Unassigned | Unassigned | Unassigned | 268_0 | Clustered | 7 | VC_268_0 | 7 | 0.3427 | 1 | 0.3427 | 0 | 0 | 0 | 1 | Ref |
| 10261 | MN552145 | Unassigned | Unassigned | Unassigned | 474_0 | Clustered | 14 | VC_474_0 | 14 | 0.8466 | 1 | 0.8466 | 0 | 0 | 0 | 1 | Ref |
| 10262 | MN552146 | Unassigned | Unassigned | Unassigned | 861_0 | Clustered | 9 | VC_861_0 | 9 | 0.5869 | 1 | 0.5869 | 0 | 0 | 0 | 1 | Ref |
| 10263 | MN552147 | Unassigned | Unassigned | Unassigned | 862_0 | Clustered | 11 | VC_862_0 | 11 | 0.6745 | 1 | 0.6745 | 0 | 0 | 0 | 1 | Ref |
| 10264 | MN552148 | Unassigned | Unassigned | Unassigned | 202_0 | Clustered | 102 | VC_202_0 | 102 | 0.4049 | 1 | 0.4049 | 0 | 0 | 0 | 0.7876 | Ref |
| 10265 | MN552149 | Unassigned | Unassigned | Unassigned | 202_0 | Clustered | 102 | VC_202_0 | 102 | 0.4049 | 1 | 0.4049 | 0 | 0 | 0 | 0.7876 | Ref |
| 10266 | MN552150 | Unassigned | Unassigned | Unassigned | 202_0 | Clustered | 102 | VC_202_0 | 102 | 0.4049 | 1 | 0.4049 | 0 | 0 | 0 | 0.7876 | Ref |
| 10267 | MN553583 | Unassigned | Unassigned | Unassigned | 838_0 | Clustered | 29 | VC_838_0 | 29 | 0.8611 | 1 | 0.8611 | 0 | 0 | 0 | 1 | Ref |
| 10268 | MN553584 | Unassigned | Unassigned | Unassigned | 838_0 | Clustered | 29 | VC_838_0 | 29 | 0.8611 | 1 | 0.8611 | 0 | 0 | 0 | 1 | Ref |
| 10269 | MN553585 | Unassigned | Unassigned | Unassigned | 423_0 | Clustered | 45 | VC_423_0 | 45 | 0.9484 | 1 | 0.9484 | 0 | 0 | 0 | 1 | Ref |
| 10270 | MN553586 | Unassigned | Unassigned | Unassigned | 472_0 | Clustered | 47 | VC_472_0 | 47 | 0.9025 | 1 | 0.9025 | 0 | 0 | 0 | 1 | Ref |
| 10271 | MN553587 | Unassigned | Unassigned | Unassigned | 423_0 | Clustered | 45 | VC_423_0 | 45 | 0.9484 | 1 | 0.9484 | 0 | 0 | 0 | 1 | Ref |
| 10272 | MN553588 | Unassigned | Unassigned | Unassigned | 423_0 | Clustered | 45 | VC_423_0 | 45 | 0.9484 | 1 | 0.9484 | 0 | 0 | 0 | 1 | Ref |
| 10273 | MN553589 | Unassigned | Unassigned | Unassigned | 423_0 | Clustered | 45 | VC_423_0 | 45 | 0.9484 | 1 | 0.9484 | 0 | 0 | 0 | 1 | Ref |
| 10274 | MN562749 | Unassigned | Unassigned | Unassigned | 472_0 | Clustered | 47 | VC_472_0 | 47 | 0.9025 | 1 | 0.9025 | 0 | 0 | 0 | 1 | Ref |
| 10275 | MN563783 | Unassigned | Unassigned | Unassigned | 80_0 | Clustered | 41 | VC_80_0 | 41 | 0.5837 | 1 | 0.5837 | 0 | 0 | 0 | 0.9805 | Ref |
| 10276 | MN563784 | Unassigned | Unassigned | Unassigned | 344_0 | Clustered | 21 | VC_344_0 | 21 | 0.5436 | 1 | 0.5436 | 0 | 0 | 0 | 1 | Ref |
| 10277 | MN563785 | Unassigned | Unassigned | Unassigned | 76_0 | Clustered | 37 | VC_76_0 | 37 | 0.9403 | 1 | 0.9403 | 0 | 0 | 0 | 1 | Ref |
| 10278 | MN563793 | Unassigned | Unassigned | Unassigned | 5_0 | Clustered | 30 | VC_5_0 | 30 | 0.9647 | 1 | 0.9647 | 0 | 0 | 0 | 1 | Ref |
| 10279 | MN564818 | Unassigned | Unassigned | Unassigned | 80_0 | Clustered | 41 | VC_80_0 | 41 | 0.5837 | 1 | 0.5837 | 0 | 0 | 0 | 0.9805 | Ref |
| 10280 | MN580668 | Unassigned | Unassigned | Unassigned | 283_179 | Clustered/Singleton | 1 | VC_283_179 | 1 | 0 | 1 | 0 | 0 | 0 | 0 | 1 | Ref |
| 10281 | MN585130 | Unassigned | Unassigned | Unassigned | 823_0 | Clustered | 124 | VC_823_0 | 124 | 0.9492 | 1 | 0.9492 | 0 | 0 | 0 | 1 | Ref |
| 10282 | MN585963 | Unassigned | Unassigned | Unassigned | 1176_0 | Clustered | 41 | VC_1176_0 | 41 | 0.9558 | 1 | 0.9558 | 0 | 0 | 0 | 0.9976 | Ref |
| 10283 | MN585964 | Unassigned | Unassigned | Unassigned | 556_0 | Clustered | 21 | VC_556_0 | 21 | 0.9912 | 1 | 0.9912 | 0 | 0 | 0 | 1 | Ref |
| 10284 | MN585965 | Unassigned | Unassigned | Unassigned | 371_13 | Clustered | 33 | VC_371_13 | 33 | 0.179 | 1 | 0.179 | 0 | 0 | 0 | 0.8769 | Ref |
| 10285 | MN585966 | Unassigned | Unassigned | Unassigned | 522_1 | Clustered | 5 | VC_522_1 | 5 | 0.1017 | 0.99999997 | 0.1017 | 0 | 0 | 0 | 0.8 | Ref |
| 10286 | MN585967 | Unassigned | Unassigned | Unassigned | 371_1 | Clustered | 11 | VC_371_1 | 11 | 0.0379 | 1 | 0.0379 | 0 | 0 | 0 | 0.8545 | Ref |
| 10287 | MN585968 | Unassigned | Unassigned | Unassigned | 216_2 | Clustered | 2 | VC_216_2 | 2 | 0.0139 | 0.95744997 | 0.0133 | 0 | 0 | 0 | 1 | Ref |
| 10288 | MN585969 | Unassigned | Unassigned | Unassigned | 514_0 | Clustered | 75 | VC_514_0 | 75 | 0.9402 | 1 | 0.9402 | 0 | 0 | 0 | 1 | Ref |
| 10289 | MN585970 | Unassigned | Unassigned | Unassigned | 325_20 | Clustered/Singleton | 1 | VC_325_20 | 1 | 0 | 1 | 0 | 0 | 0 | 0 | 1 | Ref |
| 10290 | MN585971 | Unassigned | Unassigned | Unassigned | 799_1 | Clustered | 27 | VC_799_1 | 27 | 0.8611 | 1 | 0.8611 | 0 | 0 | 0 | 1 | Ref |
| 10291 | MN585972 | Unassigned | Unassigned | Unassigned | 1169_0 | Clustered | 7 | VC_1169_0 | 7 | 0.9811 | 0.99995613 | 0.981 | 0 | 0 | 0 | 1 | Ref |
| 10292 | MN585973 | Unassigned | Unassigned | Unassigned | 520_31 | Clustered | 11 | VC_520_31 | 11 | 0.0897 | 1 | 0.0897 | 0 | 0 | 0 | 0.6 | Ref |
| 10293 | MN585974 | Unassigned | Unassigned | Unassigned | 225_0 | Clustered | 32 | VC_225_0 | 32 | 0.848 | 1 | 0.848 | 0 | 0 | 0 | 0.996 | Ref |
| 10294 | MN585975 | Unassigned | Unassigned | Unassigned | 520_89 | Clustered/Singleton | 1 | VC_520_89 | 1 | 0 | 1 | 0 | 0 | 0 | 0 | 1 | Ref |
| 10295 | MN585976 | Unassigned | Unassigned | Unassigned | 325_24 | Clustered | 3 | VC_325_24 | 3 | 0.009 | 0.99853949 | 0.009 | 0 | 0 | 0 | 1 | Ref |
| 10296 | MN585977 | Unassigned | Unassigned | Unassigned | 519_0 | Clustered | 39 | VC_519_0 | 39 | 0.8156 | 1 | 0.8156 | 0 | 0 | 0 | 1 | Ref |
| 10297 | MN585978 | Unassigned | Unassigned | Unassigned | 225_0 | Clustered | 32 | VC_225_0 | 32 | 0.848 | 1 | 0.848 | 0 | 0 | 0 | 0.996 | Ref |
| 10298 | MN585979 | Unassigned | Unassigned | Unassigned | 225_0 | Clustered | 32 | VC_225_0 | 32 | 0.848 | 1 | 0.848 | 0 | 0 | 0 | 0.996 | Ref |
| 10299 | MN585980 | Unassigned | Unassigned | Unassigned | 1352_0 | Clustered | 31 | VC_1352_0 | 31 | 0.849 | 1 | 0.849 | 0 | 0 | 0 | 1 | Ref |
| 10300 | MN585981 | Unassigned | Unassigned | Unassigned | 216_6 | Clustered | 42 | VC_216_6 | 42 | 0.473 | 1 | 0.473 | 0 | 0 | 0 | 0.8351 | Ref |
| 10301 | MN585982 | Unassigned | Unassigned | Unassigned | 799_1 | Clustered | 27 | VC_799_1 | 27 | 0.8611 | 1 | 0.8611 | 0 | 0 | 0 | 1 | Ref |
| 10302 | MN585983 | Unassigned | Unassigned | Unassigned | 216_6 | Clustered | 42 | VC_216_6 | 42 | 0.473 | 1 | 0.473 | 0 | 0 | 0 | 0.8351 | Ref |
| 10303 | MN585984 | Unassigned | Unassigned | Unassigned | 514_0 | Clustered | 75 | VC_514_0 | 75 | 0.9402 | 1 | 0.9402 | 0 | 0 | 0 | 1 | Ref |
| 10304 | MN585985 | Unassigned | Unassigned | Unassigned | 325_37 | Clustered | 3 | VC_325_37 | 3 | 0.008 | 0.99634936 | 0.008 | 0 | 0 | 0 | 0.6667 | Ref |
| 10305 | MN585986 | Unassigned | Unassigned | Unassigned | 1352_0 | Clustered | 31 | VC_1352_0 | 31 | 0.849 | 1 | 0.849 | 0 | 0 | 0 | 1 | Ref |
| 10306 | MN585987 | Unassigned | Unassigned | Unassigned | 520_59 | Clustered | 5 | VC_520_59 | 5 | 0.0459 | 0.99999997 | 0.0459 | 0 | 0 | 0 | 1 | Ref |
| 10307 | MN585988 | Unassigned | Unassigned | Unassigned | 799_1 | Clustered | 27 | VC_799_1 | 27 | 0.8611 | 1 | 0.8611 | 0 | 0 | 0 | 1 | Ref |
| 10308 | MN585989 | Unassigned | Unassigned | Unassigned | 215_0 | Clustered | 5 | VC_215_0 | 5 | 0.0724 | 0.99999997 | 0.0724 | 0 | 0 | 0 | 1 | Ref |
| 10309 | MN585990 | Unassigned | Unassigned | Unassigned | 1344_0 | Clustered | 78 | VC_1344_0 | 78 | 0.9732 | 1 | 0.9732 | 0 | 0 | 0 | 1 | Ref |
| 10310 | MN585991 | Unassigned | Unassigned | Unassigned | 325_51 | Clustered | 2 | VC_325_51 | 2 | 0.0039 | 0.95663826 | 0.0037 | 0 | 0 | 0 | 1 | Ref |
| 10311 | MN585992 | Unassigned | Unassigned | Unassigned | 764_0 | Clustered | 4 | VC_764_0 | 4 | 0.3556 | 0.99997469 | 0.3556 | 0 | 0 | 0 | 1 | Ref |
| 10312 | MN585993 | Unassigned | Unassigned | Unassigned | 571_0 | Clustered | 4 | VC_571_0 | 4 | 0.1921 | 0.99998591 | 0.1921 | 0 | 0 | 0 | 1 | Ref |
| 10313 | MN585994 | Unassigned | Unassigned | Unassigned | 225_0 | Clustered | 32 | VC_225_0 | 32 | 0.848 | 1 | 0.848 | 0 | 0 | 0 | 0.996 | Ref |
| 10314 | MN585995 | Unassigned | Unassigned | Unassigned | 520_11 | Clustered/Singleton | 1 | VC_520_11 | 1 | 0 | 1 | 0 | 0 | 0 | 0 | 1 | Ref |
| 10315 | MN585996 | Unassigned | Unassigned | Unassigned | 799_1 | Clustered | 27 | VC_799_1 | 27 | 0.8611 | 1 | 0.8611 | 0 | 0 | 0 | 1 | Ref |
| 10316 | MN585997 | Unassigned | Unassigned | Unassigned | 325_56 | Clustered | 2 | VC_325_56 | 2 | 0.0036 | 0.94604798 | 0.0034 | 0 | 0 | 0 | 0 | Ref |
| 10317 | MN585998 | Unassigned | Unassigned | Unassigned | 216_6 | Clustered | 42 | VC_216_6 | 42 | 0.473 | 1 | 0.473 | 0 | 0 | 0 | 0.8351 | Ref |
| 10318 | MN585999 | Unassigned | Unassigned | Unassigned | 325_5 | Clustered | 11 | VC_325_5 | 11 | 0.047 | 1 | 0.047 | 0 | 0 | 0 | 0.7636 | Ref |
| 10319 | MN586000 | Unassigned | Unassigned | Unassigned | 325_35 | Clustered | 17 | VC_325_35 | 17 | 0.0732 | 1 | 0.0732 | 0 | 0 | 0 | 0.7721 | Ref |
| 10320 | MN586001 | Unassigned | Unassigned | Unassigned | 520_18 | Clustered/Singleton | 1 | VC_520_18 | 1 | 0 | 1 | 0 | 0 | 0 | 0 | 1 | Ref |
| 10321 | MN586002 | Unassigned | Unassigned | Unassigned | 216_2 | Clustered | 2 | VC_216_2 | 2 | 0.0139 | 0.95744997 | 0.0133 | 0 | 0 | 0 | 1 | Ref |
| 10322 | MN586003 | Unassigned | Unassigned | Unassigned | 572_0 | Clustered | 16 | VC_572_0 | 16 | 0.7286 | 1 | 0.7286 | 0 | 0 | 0 | 1 | Ref |
| 10323 | MN586004 | Unassigned | Unassigned | Unassigned | 520_6 | Clustered | 3 | VC_520_6 | 3 | 0.0276 | 0.99859841 | 0.0276 | 0 | 0 | 0 | 1 | Ref |
| 10324 | MN586005 | Unassigned | Unassigned | Unassigned | 520_59 | Clustered | 5 | VC_520_59 | 5 | 0.0459 | 0.99999997 | 0.0459 | 0 | 0 | 0 | 1 | Ref |
| 10325 | MN586006 | Unassigned | Unassigned | Unassigned | 221_0 | Clustered | 22 | VC_221_0 | 22 | 0.7447 | 1 | 0.7447 | 0 | 0 | 0 | 1 | Ref |
| 10326 | MN586007 | Unassigned | Unassigned | Unassigned | 1352_0 | Clustered | 31 | VC_1352_0 | 31 | 0.849 | 1 | 0.849 | 0 | 0 | 0 | 1 | Ref |
| 10327 | MN586008 | Unassigned | Unassigned | Unassigned | 1344_0 | Clustered | 78 | VC_1344_0 | 78 | 0.9732 | 1 | 0.9732 | 0 | 0 | 0 | 1 | Ref |
| 10328 | MN586009 | Unassigned | Unassigned | Unassigned | 982_0 | Clustered | 62 | VC_982_0 | 62 | 0.8646 | 1 | 0.8646 | 0 | 0 | 0 | 0.743 | Ref |
| 10329 | MN586010 | Unassigned | Unassigned | Unassigned | 1344_0 | Clustered | 78 | VC_1344_0 | 78 | 0.9732 | 1 | 0.9732 | 0 | 0 | 0 | 1 | Ref |
| 10330 | MN586011 | Unassigned | Unassigned | Unassigned | 520_41 | Clustered | 2 | VC_520_41 | 2 | 0.0139 | 0.95761457 | 0.0134 | 0 | 0 | 0 | 1 | Ref |
| 10331 | MN586012 | Unassigned | Unassigned | Unassigned | 520_66 | Clustered/Singleton | 1 | VC_520_66 | 1 | 0 | 1 | 0 | 0 | 0 | 0 | 1 | Ref |
| 10332 | MN586013 | Unassigned | Unassigned | Unassigned | 551_0 | Clustered | 16 | VC_551_0 | 16 | 0.0915 | 1 | 0.0915 | 0 | 0 | 0 | 0.6667 | Ref |
| 10333 | MN586014 | Unassigned | Unassigned | Unassigned | 1183_0 | Clustered | 6 | VC_1183_0 | 6 | 0.995 | 0.99996974 | 0.995 | 0 | 0 | 0 | 1 | Ref |
| 10334 | MN586015 | Unassigned | Unassigned | Unassigned | 919_0 | Clustered | 39 | VC_919_0 | 39 | 0.9069 | 1 | 0.9069 | 0 | 0 | 0 | 1 | Ref |
| 10335 | MN586016 | Unassigned | Unassigned | Unassigned | 940_0 | Clustered | 34 | VC_940_0 | 34 | 0.7959 | 1 | 0.7959 | 0 | 0 | 0 | 1 | Ref |
| 10336 | MN586017 | Unassigned | Unassigned | Unassigned | 551_7 | Clustered | 28 | VC_551_7 | 28 | 0.1661 | 1 | 0.1661 | 0 | 0 | 0 | 0.8519 | Ref |
| 10337 | MN586018 | Unassigned | Unassigned | Unassigned | 887_0 | Clustered | 18 | VC_887_0 | 18 | 0.3663 | 1 | 0.3663 | 0 | 0 | 0 | 1 | Ref |
| 10338 | MN586019 | Unassigned | Unassigned | Unassigned | 551_7 | Clustered | 28 | VC_551_7 | 28 | 0.1661 | 1 | 0.1661 | 0 | 0 | 0 | 0.8519 | Ref |
| 10339 | MN586021 | Unassigned | Unassigned | Unassigned | 1170_0 | Clustered | 3 | VC_1170_0 | 3 | 0.3275 | 0.99834213 | 0.327 | 0 | 0 | 0 | 1 | Ref |
| 10340 | MN586022 | Unassigned | Unassigned | Unassigned | 1096_0 | Clustered | 5 | VC_1096_0 | 5 | 0.8566 | 0.99869977 | 0.8555 | 0 | 0 | 0 | 1 | Ref |
| 10341 | MN586023 | Unassigned | Unassigned | Unassigned | 887_0 | Clustered | 18 | VC_887_0 | 18 | 0.3663 | 1 | 0.3663 | 0 | 0 | 0 | 1 | Ref |
| 10342 | MN586024 | Unassigned | Unassigned | Unassigned | 763_1 | Clustered | 9 | VC_763_1 | 9 | 0.3706 | 1 | 0.3706 | 0 | 0 | 0 | 1 | Ref |
| 10343 | MN586026 | Unassigned | Unassigned | Unassigned | 1095_0 | Clustered | 16 | VC_1095_0 | 16 | 0.458 | 1 | 0.458 | 0 | 0 | 0 | 1 | Ref |
| 10344 | MN586027 | Unassigned | Unassigned | Unassigned | 1370_0 | Clustered | 2 | VC_1370_0 | 2 | 0.8214 | 0.91237788 | 0.7494 | 0 | 0 | 0 | 1 | Ref |
| 10345 | MN586028 | Unassigned | Unassigned | Unassigned | 1095_0 | Clustered | 16 | VC_1095_0 | 16 | 0.458 | 1 | 0.458 | 0 | 0 | 0 | 1 | Ref |
| 10346 | MN586029 | Unassigned | Unassigned | Unassigned | 546_1 | Clustered | 2 | VC_546_1 | 2 | 0.0136 | 0.68405151 | 0.0093 | 0 | 0 | 0 | 1 | Ref |
| 10347 | MN586030 | Unassigned | Unassigned | Unassigned | 888_0 | Clustered | 28 | VC_888_0 | 28 | 0.5497 | 1 | 0.5497 | 0 | 0 | 0 | 1 | Ref |
| 10348 | MN586031 | Unassigned | Unassigned | Unassigned | 1176_0 | Clustered | 41 | VC_1176_0 | 41 | 0.9558 | 1 | 0.9558 | 0 | 0 | 0 | 0.9976 | Ref |
| 10349 | MN586032 | Unassigned | Unassigned | Unassigned | 551_2 | Clustered | 17 | VC_551_2 | 17 | 0.0973 | 1 | 0.0973 | 0 | 0 | 0 | 0.9779 | Ref |
| 10350 | MN586034 | Unassigned | Unassigned | Unassigned | 551_6 | Clustered | 22 | VC_551_6 | 22 | 0.1214 | 1 | 0.1214 | 0 | 0 | 0 | 0.8095 | Ref |
| 10351 | MN586035 | Unassigned | Unassigned | Unassigned | 551_7 | Clustered | 28 | VC_551_7 | 28 | 0.1661 | 1 | 0.1661 | 0 | 0 | 0 | 0.8519 | Ref |
| 10352 | MN586036 | Unassigned | Unassigned | Unassigned | 919_0 | Clustered | 39 | VC_919_0 | 39 | 0.9069 | 1 | 0.9069 | 0 | 0 | 0 | 1 | Ref |
| 10353 | MN586037 | Unassigned | Unassigned | Unassigned | 551_6 | Clustered | 22 | VC_551_6 | 22 | 0.1214 | 1 | 0.1214 | 0 | 0 | 0 | 0.8095 | Ref |
| 10354 | MN586038 | Unassigned | Unassigned | Unassigned | 515_0 | Clustered | 16 | VC_515_0 | 16 | 0.3793 | 1 | 0.3793 | 0 | 0 | 0 | 1 | Ref |
| 10355 | MN586039 | Unassigned | Unassigned | Unassigned | 223_0 | Clustered | 38 | VC_223_0 | 38 | 0.9095 | 1 | 0.9095 | 0 | 0 | 0 | 1 | Ref |
| 10356 | MN586041 | Unassigned | Unassigned | Unassigned | 551_7 | Clustered | 28 | VC_551_7 | 28 | 0.1661 | 1 | 0.1661 | 0 | 0 | 0 | 0.8519 | Ref |
| 10357 | MN586042 | Unassigned | Unassigned | Unassigned | 1352_0 | Clustered | 31 | VC_1352_0 | 31 | 0.849 | 1 | 0.849 | 0 | 0 | 0 | 1 | Ref |
| 10358 | MN586043 | Unassigned | Unassigned | Unassigned | 551_1 | Clustered | 4 | VC_551_1 | 4 | 0.0159 | 0.99968337 | 0.0159 | 0 | 0 | 0 | 0.6667 | Ref |
| 10359 | MN586044 | Unassigned | Unassigned | Unassigned | 551_1 | Clustered | 4 | VC_551_1 | 4 | 0.0159 | 0.99968337 | 0.0159 | 0 | 0 | 0 | 0.6667 | Ref |
| 10360 | MN586045 | Unassigned | Unassigned | Unassigned | 508_0 | Clustered | 32 | VC_508_0 | 32 | 0.906 | 1 | 0.906 | 0 | 0 | 0 | 1 | Ref |
| 10361 | MN586046 | Unassigned | Unassigned | Unassigned | 508_0 | Clustered | 32 | VC_508_0 | 32 | 0.906 | 1 | 0.906 | 0 | 0 | 0 | 1 | Ref |
| 10362 | MN586047 | Unassigned | Unassigned | Unassigned | 1095_0 | Clustered | 16 | VC_1095_0 | 16 | 0.458 | 1 | 0.458 | 0 | 0 | 0 | 1 | Ref |
| 10363 | MN586048 | Unassigned | Unassigned | Unassigned | 514_0 | Clustered | 75 | VC_514_0 | 75 | 0.9402 | 1 | 0.9402 | 0 | 0 | 0 | 1 | Ref |
| 10364 | MN586049 | Unassigned | Unassigned | Unassigned | 919_0 | Clustered | 39 | VC_919_0 | 39 | 0.9069 | 1 | 0.9069 | 0 | 0 | 0 | 1 | Ref |
| 10365 | MN586050 | Unassigned | Unassigned | Unassigned | 551_7 | Clustered | 28 | VC_551_7 | 28 | 0.1661 | 1 | 0.1661 | 0 | 0 | 0 | 0.8519 | Ref |
| 10366 | MN586051 | Unassigned | Unassigned | Unassigned | 551_2 | Clustered | 17 | VC_551_2 | 17 | 0.0973 | 1 | 0.0973 | 0 | 0 | 0 | 0.9779 | Ref |

| | | | | | | | | | | | | | | | | | |
|---|---|---|---|---|---|---|---|---|---|---|---|---|---|---|---|---|---|
| 10367 | MN586052 | Unassigned | Unassigned | Unassigned | 888_0 | Clustered | 28 | VC_888_0 | 28 | 0.5497 | 1 | 0.5497 | 0 | 0 | 0 | 1 | Ref |
| 10368 | MN586053 | Unassigned | Unassigned | Unassigned | 1179_0 | Clustered | 3 | VC_1179_0 | 3 | 0.7588 | 0.96819872 | 0.7347 | 0 | 0 | 0 | 1 | Ref |
| 10369 | MN586054 | Unassigned | Unassigned | Unassigned | 872_0 | Clustered | 31 | VC_872_0 | 31 | 0.8151 | 1 | 0.8151 | 0 | 0 | 0 | 0.8194 | Ref |
| 10370 | MN586055 | Unassigned | Unassigned | Unassigned | 1175_0 | Clustered | 22 | VC_1175_0 | 22 | 0.4562 | 1 | 0.4562 | 0 | 0 | 0 | 1 | Ref |
| 10371 | MN586056 | Unassigned | Unassigned | Unassigned | 872_0 | Clustered | 31 | VC_872_0 | 31 | 0.8151 | 1 | 0.8151 | 0 | 0 | 0 | 0.8194 | Ref |
| 10372 | MN586057 | Unassigned | Unassigned | Unassigned | 919_0 | Clustered | 39 | VC_919_0 | 39 | 0.9069 | 1 | 0.9069 | 0 | 0 | 0 | 1 | Ref |
| 10373 | MN586058 | Unassigned | Unassigned | Unassigned | 371_1 | Clustered | 11 | VC_371_1 | 11 | 0.0379 | 1 | 0.0379 | 0 | 0 | 0 | 0.8545 | Ref |
| 10374 | MN586059 | Unassigned | Unassigned | Unassigned | 1352_0 | Clustered | 31 | VC_1352_0 | 31 | 0.849 | 1 | 0.849 | 0 | 0 | 0 | 1 | Ref |
| 10375 | MN586060 | Unassigned | Unassigned | Unassigned | 940_0 | Clustered | 34 | VC_940_0 | 34 | 0.7959 | 1 | 0.7959 | 0 | 0 | 0 | 1 | Ref |
| 10376 | MN586061 | Unassigned | Unassigned | Unassigned | 1160_0 | Clustered | 73 | VC_1160_0 | 73 | 0.9835 | 1 | 0.9835 | 0 | 0 | 0 | 1 | Ref |
| 10377 | MN586062 | Unassigned | Unassigned | Unassigned | 940_0 | Clustered | 34 | VC_940_0 | 34 | 0.7959 | 1 | 0.7959 | 0 | 0 | 0 | 1 | Ref |
| 10378 | MN586063 | Unassigned | Unassigned | Unassigned | 513_0 | Clustered | 32 | VC_513_0 | 32 | 0.6881 | 1 | 0.6881 | 0 | 0 | 0 | 1 | Ref |
| 10379 | MN592896 | Unassigned | Unassigned | Unassigned | 925_0 | Clustered/Singleton | 1 | VC_925_0 | 1 | 0 | 1 | 0 | 0 | 0 | 0 | 1 | Ref |
| 10380 | MN592897 | Unassigned | Unassigned | Unassigned | 296_0 | Clustered | 6 | VC_296_0 | 6 | 0.5315 | 1 | 0.5315 | 0 | 0 | 0 | 1 | Ref |
| 10381 | MN602045 | Unassigned | Unassigned | Unassigned | 423_0 | Clustered | 45 | VC_423_0 | 45 | 0.9484 | 1 | 0.9484 | 0 | 0 | 0 | 1 | Ref |
| 10382 | MN602266 | Unassigned | Unassigned | Unassigned | 936_0 | Clustered | 2 | VC_936_0 | 2 | 0.9284 | 0.81445332 | 0.7561 | 0 | 0 | 0 | 1 | Ref |
| 10383 | MN602267 | Unassigned | Unassigned | Unassigned | 936_0 | Clustered | 2 | VC_936_0 | 2 | 0.9284 | 0.81445332 | 0.7561 | 0 | 0 | 0 | 1 | Ref |
| 10384 | MN602268 | Unassigned | Unassigned | Unassigned | 977_1 | Clustered | 2 | VC_977_1 | 2 | 0.079 | 0.93977186 | 0.0743 | 0 | 0 | 0 | 1 | Ref |
| 10385 | MN604053 | Unassigned | Unassigned | Unassigned | 591_0 | Clustered | 14 | VC_591_0 | 14 | 0.5184 | 1 | 0.5184 | 0 | 0 | 0 | 1 | Ref |
| 10386 | MN604230 | Unassigned | Unassigned | Unassigned | 144_0 | Clustered | 2 | VC_144_0 | 2 | 0.4237 | 0.94768242 | 0.4015 | 0 | 0 | 0 | 1 | Ref |
| 10387 | MN604238 | Unassigned | Unassigned | Unassigned | 699_0 | Clustered | 71 | VC_699_0 | 71 | 0.9352 | 1 | 0.9352 | 0 | 0 | 0 | 1 | Ref |
| 10388 | MN604239 | Unassigned | Unassigned | Unassigned | 699_0 | Clustered | 71 | VC_699_0 | 71 | 0.9352 | 1 | 0.9352 | 0 | 0 | 0 | 1 | Ref |
| 10389 | MN604407 | Unassigned | Unassigned | Unassigned | 328_0 | Clustered | 11 | VC_328_0 | 11 | 0.4707 | 1 | 0.4707 | 0 | 0 | 0 | 1 | Ref |
| 10390 | MN604698 | Unassigned | Unassigned | Unassigned | 144_0 | Clustered | 2 | VC_144_0 | 2 | 0.4237 | 0.94768242 | 0.4015 | 0 | 0 | 0 | 1 | Ref |
| 10391 | MN614471 | Unassigned | Unassigned | Unassigned | 699_0 | Clustered | 71 | VC_699_0 | 71 | 0.9352 | 1 | 0.9352 | 0 | 0 | 0 | 1 | Ref |
| 10392 | MN615698 | Unassigned | Unassigned | Unassigned | 423_0 | Clustered | 45 | VC_423_0 | 45 | 0.9484 | 1 | 0.9484 | 0 | 0 | 0 | 1 | Ref |
| 10393 | MN615699 | Unassigned | Unassigned | Unassigned | 423_0 | Clustered | 45 | VC_423_0 | 45 | 0.9484 | 1 | 0.9484 | 0 | 0 | 0 | 1 | Ref |
| 10394 | MN615700 | Unassigned | Unassigned | Unassigned | 79_7 | Clustered | 33 | VC_79_7 | 33 | 0.216 | 1 | 0.216 | 0 | 0 | 0 | 0.8409 | Ref |
| 10395 | MN615701 | Unassigned | Unassigned | Unassigned | 79_7 | Clustered | 33 | VC_79_7 | 33 | 0.216 | 1 | 0.216 | 0 | 0 | 0 | 0.8409 | Ref |
| 10396 | MN615702 | Unassigned | Unassigned | Unassigned | 79_7 | Clustered | 33 | VC_79_7 | 33 | 0.216 | 1 | 0.216 | 0 | 0 | 0 | 0.8409 | Ref |
| 10397 | MN617773 | Unassigned | Unassigned | Unassigned | 450_0 | Clustered | 16 | VC_450_0 | 16 | 0.9564 | 1 | 0.9564 | 0 | 0 | 0 | 1 | Ref |
| 10398 | MN617840 | Unassigned | Unassigned | Unassigned | 1240_0 | Clustered | 82 | VC_1240_0 | 82 | 0.7945 | 1 | 0.7945 | 0 | 0 | 0 | 1 | Ref |
| 10399 | MN617841 | Unassigned | Unassigned | Unassigned | 887_0 | Clustered | 18 | VC_887_0 | 18 | 0.3663 | 1 | 0.3663 | 0 | 0 | 0 | 1 | Ref |
| 10400 | MN617842 | Unassigned | Unassigned | Unassigned | 950_0 | Clustered | 38 | VC_950_0 | 38 | 0.9443 | 1 | 0.9443 | 0 | 0 | 0 | 1 | Ref |
| 10401 | MN617843 | Unassigned | Unassigned | Unassigned | 372_2 | Clustered | 5 | VC_372_2 | 5 | 0.072 | 0.99999997 | 0.072 | 0 | 0 | 0 | 0.9 | Ref |
| 10402 | MN617844 | Unassigned | Unassigned | Unassigned | 940_0 | Clustered | 34 | VC_940_0 | 34 | 0.7959 | 1 | 0.7959 | 0 | 0 | 0 | 1 | Ref |
| 10403 | MN617845 | Unassigned | Unassigned | Unassigned | 325_1 | Clustered | 2 | VC_325_1 | 2 | 0.0043 | 0.95663348 | 0.0041 | 0 | 0 | 0 | 1 | Ref |
| 10404 | MN642089 | Unassigned | Unassigned | Unassigned | 442_1 | Clustered | 3 | VC_442_1 | 3 | 0.0538 | 0.99419552 | 0.0535 | 0 | 0 | 0 | 1 | Ref |
| 10405 | MN648195 | Unassigned | Unassigned | Unassigned | 492_1 | Clustered | 20 | VC_492_1 | 20 | 0.4176 | 1 | 0.4176 | 0 | 0 | 0 | 1 | Ref |
| 10406 | MN648445 | Unassigned | Unassigned | Unassigned | 286_17 | Clustered | 8 | VC_286_17 | 8 | 0.0535 | 0.99997829 | 0.0535 | 0 | 0 | 0 | 0.6071 | Ref |
| 10407 | MN651570 | Unassigned | Unassigned | Unassigned | 699_0 | Clustered | 71 | VC_699_0 | 71 | 0.9352 | 1 | 0.9352 | 0 | 0 | 0 | 1 | Ref |
| 10408 | MN655998 | Unassigned | Unassigned | Unassigned | 590_0 | Clustered | 27 | VC_590_0 | 27 | 0.9947 | 1 | 0.9947 | 0 | 0 | 0 | 1 | Ref |
| 10409 | MN655999 | Unassigned | Unassigned | Unassigned | 329_11 | Clustered | 15 | VC_329_11 | 15 | 0.0791 | 1 | 0.0791 | 0 | 0 | 0 | 0.9333 | Ref |
| 10410 | MN656993 | Unassigned | Unassigned | Unassigned | 836_0 | Clustered | 9 | VC_836_0 | 9 | 0.7733 | 1 | 0.7733 | 0 | 0 | 0 | 1 | Ref |
| 10411 | MN662249 | Unassigned | Unassigned | Unassigned | 492_1 | Clustered | 20 | VC_492_1 | 20 | 0.4176 | 1 | 0.4176 | 0 | 0 | 0 | 1 | Ref |
| 10412 | MN688132 | Unassigned | Unassigned | Unassigned | 33_0 | Clustered | 4 | VC_33_0 | 4 | 0.06 | 0.99994826 | 0.06 | 0 | 0 | 0 | 1 | Ref |
| 10413 | MN689503 | Unassigned | Unassigned | Unassigned | 690_0 | Clustered | 50 | VC_690_0 | 50 | 0.9805 | 1 | 0.9805 | 0 | 0 | 0 | 1 | Ref |
| 10414 | MN689504 | Unassigned | Unassigned | Unassigned | 690_0 | Clustered | 50 | VC_690_0 | 50 | 0.9805 | 1 | 0.9805 | 0 | 0 | 0 | 1 | Ref |
| 10415 | MN689505 | Unassigned | Unassigned | Unassigned | 690_0 | Clustered | 50 | VC_690_0 | 50 | 0.9805 | 1 | 0.9805 | 0 | 0 | 0 | 1 | Ref |
| 10416 | MN689506 | Unassigned | Unassigned | Unassigned | 690_0 | Clustered | 50 | VC_690_0 | 50 | 0.9805 | 1 | 0.9805 | 0 | 0 | 0 | 1 | Ref |
| 10417 | MN689507 | Unassigned | Unassigned | Unassigned | 690_0 | Clustered | 50 | VC_690_0 | 50 | 0.9805 | 1 | 0.9805 | 0 | 0 | 0 | 1 | Ref |
| 10418 | MN689508 | Unassigned | Unassigned | Unassigned | 690_0 | Clustered | 50 | VC_690_0 | 50 | 0.9805 | 1 | 0.9805 | 0 | 0 | 0 | 1 | Ref |
| 10419 | MN689509 | Unassigned | Unassigned | Unassigned | 260_0 | Clustered | 13 | VC_260_0 | 13 | 0.3705 | 1 | 0.3705 | 0 | 0 | 0 | 1 | Ref |
| 10420 | MN689510 | Unassigned | Unassigned | Unassigned | 690_0 | Clustered | 50 | VC_690_0 | 50 | 0.9805 | 1 | 0.9805 | 0 | 0 | 0 | 1 | Ref |
| 10421 | MN689511 | Unassigned | Unassigned | Unassigned | 690_0 | Clustered | 50 | VC_690_0 | 50 | 0.9805 | 1 | 0.9805 | 0 | 0 | 0 | 1 | Ref |
| 10422 | MN689512 | Unassigned | Unassigned | Unassigned | 690_0 | Clustered | 50 | VC_690_0 | 50 | 0.9805 | 1 | 0.9805 | 0 | 0 | 0 | 1 | Ref |
| 10423 | MN689513 | Unassigned | Unassigned | Unassigned | 690_0 | Clustered | 50 | VC_690_0 | 50 | 0.9805 | 1 | 0.9805 | 0 | 0 | 0 | 1 | Ref |
| 10424 | MN689514 | Unassigned | Unassigned | Unassigned | 202_0 | Clustered | 102 | VC_202_0 | 102 | 0.4049 | 1 | 0.4049 | 0 | 0 | 0 | 0.7876 | Ref |
| 10425 | MN689515 | Unassigned | Unassigned | Unassigned | 690_0 | Clustered | 50 | VC_690_0 | 50 | 0.9805 | 1 | 0.9805 | 0 | 0 | 0 | 1 | Ref |
| 10426 | MN689516 | Unassigned | Unassigned | Unassigned | 260_0 | Clustered | 13 | VC_260_0 | 13 | 0.3705 | 1 | 0.3705 | 0 | 0 | 0 | 1 | Ref |
| 10427 | MN689517 | Unassigned | Unassigned | Unassigned | 202_0 | Clustered | 102 | VC_202_0 | 102 | 0.4049 | 1 | 0.4049 | 0 | 0 | 0 | 0.7876 | Ref |
| 10428 | MN689518 | Unassigned | Unassigned | Unassigned | 202_0 | Clustered | 102 | VC_202_0 | 102 | 0.4049 | 1 | 0.4049 | 0 | 0 | 0 | 0.7876 | Ref |
| 10429 | MN689519 | Unassigned | Unassigned | Unassigned | 202_0 | Clustered | 102 | VC_202_0 | 102 | 0.4049 | 1 | 0.4049 | 0 | 0 | 0 | 0.7876 | Ref |
| 10430 | MN689520 | Unassigned | Unassigned | Unassigned | 202_0 | Clustered | 102 | VC_202_0 | 102 | 0.4049 | 1 | 0.4049 | 0 | 0 | 0 | 0.7876 | Ref |
| 10431 | MN689521 | Unassigned | Unassigned | Unassigned | 260_0 | Clustered | 13 | VC_260_0 | 13 | 0.3705 | 1 | 0.3705 | 0 | 0 | 0 | 1 | Ref |
| 10432 | MN689522 | Unassigned | Unassigned | Unassigned | 202_0 | Clustered | 102 | VC_202_0 | 102 | 0.4049 | 1 | 0.4049 | 0 | 0 | 0 | 0.7876 | Ref |
| 10433 | MN689523 | Unassigned | Unassigned | Unassigned | 202_0 | Clustered | 102 | VC_202_0 | 102 | 0.4049 | 1 | 0.4049 | 0 | 0 | 0 | 0.7876 | Ref |
| 10434 | MN689524 | Unassigned | Unassigned | Unassigned | 202_0 | Clustered | 102 | VC_202_0 | 102 | 0.4049 | 1 | 0.4049 | 0 | 0 | 0 | 0.7876 | Ref |
| 10435 | MN689525 | Unassigned | Unassigned | Unassigned | 202_0 | Clustered | 102 | VC_202_0 | 102 | 0.4049 | 1 | 0.4049 | 0 | 0 | 0 | 0.7876 | Ref |
| 10436 | MN689526 | Unassigned | Unassigned | Unassigned | 690_0 | Clustered | 50 | VC_690_0 | 50 | 0.9805 | 1 | 0.9805 | 0 | 0 | 0 | 1 | Ref |
| 10437 | MN689527 | Unassigned | Unassigned | Unassigned | 690_0 | Clustered | 50 | VC_690_0 | 50 | 0.9805 | 1 | 0.9805 | 0 | 0 | 0 | 1 | Ref |
| 10438 | MN689528 | Unassigned | Unassigned | Unassigned | 690_0 | Clustered | 50 | VC_690_0 | 50 | 0.9805 | 1 | 0.9805 | 0 | 0 | 0 | 1 | Ref |
| 10439 | MN689529 | Unassigned | Unassigned | Unassigned | 690_0 | Clustered | 50 | VC_690_0 | 50 | 0.9805 | 1 | 0.9805 | 0 | 0 | 0 | 1 | Ref |
| 10440 | MN689530 | Unassigned | Unassigned | Unassigned | 260_0 | Clustered | 13 | VC_260_0 | 13 | 0.3705 | 1 | 0.3705 | 0 | 0 | 0 | 1 | Ref |
| 10441 | MN689531 | Unassigned | Unassigned | Unassigned | 690_0 | Clustered | 50 | VC_690_0 | 50 | 0.9805 | 1 | 0.9805 | 0 | 0 | 0 | 1 | Ref |
| 10442 | MN689532 | Unassigned | Unassigned | Unassigned | 690_0 | Clustered | 50 | VC_690_0 | 50 | 0.9805 | 1 | 0.9805 | 0 | 0 | 0 | 1 | Ref |
| 10443 | MN689533 | Unassigned | Unassigned | Unassigned | 690_0 | Clustered | 50 | VC_690_0 | 50 | 0.9805 | 1 | 0.9805 | 0 | 0 | 0 | 1 | Ref |
| 10444 | MN689778 | Unassigned | Unassigned | Unassigned | 123_0 | Clustered | 23 | VC_123_0 | 23 | 0.817 | 1 | 0.817 | 0 | 0 | 0 | 1 | Ref |
| 10445 | MN689779 | Unassigned | Unassigned | Unassigned | 938_0 | Clustered | 99 | VC_938_0 | 99 | 0.9119 | 1 | 0.9119 | 0 | 0 | 0 | 0.9928 | Ref |
| 10446 | MN690600 | Unassigned | Unassigned | Unassigned | 16_0 | Clustered | 12 | VC_16_0 | 12 | 0.6615 | 1 | 0.6615 | 0 | 0 | 0 | 1 | Ref |
| 10447 | MN692672 | Unassigned | Unassigned | Unassigned | 423_0 | Clustered | 45 | VC_423_0 | 45 | 0.9484 | 1 | 0.9484 | 0 | 0 | 0 | 1 | Ref |
| 10448 | MN692673 | Unassigned | Unassigned | Unassigned | 461_0 | Clustered | 96 | VC_461_0 | 96 | 0.9307 | 1 | 0.9307 | 0 | 0 | 0 | 0.9575 | Ref |
| 10449 | MN695334 | Unassigned | Unassigned | Unassigned | 432_0 | Clustered | 2 | VC_432_0 | 2 | 0.0362 | 0.95680396 | 0.0346 | 0 | 0 | 0 | 1 | Ref |
| 10450 | MN695335 | Unassigned | Unassigned | Unassigned | 432_0 | Clustered | 2 | VC_432_0 | 2 | 0.0362 | 0.95680396 | 0.0346 | 0 | 0 | 0 | 1 | Ref |
| 10451 | MN698239 | Unassigned | Unassigned | Unassigned | 470_0 | Clustered | 3 | VC_470_0 | 3 | 0.5552 | 0.99823354 | 0.5543 | 0 | 0 | 0 | 1 | Ref |
| 10452 | MN698240 | Unassigned | Unassigned | Unassigned | 1003_0 | Clustered | 4 | VC_1003_0 | 4 | 0.8718 | 0.99983781 | 0.8716 | 0 | 0 | 0 | 1 | Ref |
| 10453 | MN698241 | Unassigned | Unassigned | Unassigned | 470_0 | Clustered | 3 | VC_470_0 | 3 | 0.5552 | 0.99823354 | 0.5543 | 0 | 0 | 0 | 1 | Ref |
| 10454 | MN698242 | Unassigned | Unassigned | Unassigned | 485_0 | Clustered | 3 | VC_485_0 | 3 | 0.2389 | 0.99844364 | 0.2385 | 0 | 0 | 0 | 1 | Ref |
| 10455 | MN698243 | Unassigned | Unassigned | Unassigned | 485_0 | Clustered | 3 | VC_485_0 | 3 | 0.2389 | 0.99844364 | 0.2385 | 0 | 0 | 0 | 1 | Ref |
| 10456 | MN698245 | Unassigned | Unassigned | Unassigned | 1003_0 | Clustered | 4 | VC_1003_0 | 4 | 0.8718 | 0.99983781 | 0.8716 | 0 | 0 | 0 | 1 | Ref |
| 10457 | MN698246 | Unassigned | Unassigned | Unassigned | 1003_0 | Clustered | 4 | VC_1003_0 | 4 | 0.8718 | 0.99983781 | 0.8716 | 0 | 0 | 0 | 1 | Ref |
| 10458 | MN698247 | Unassigned | Unassigned | Unassigned | 485_0 | Clustered | 3 | VC_485_0 | 3 | 0.2389 | 0.99844364 | 0.2385 | 0 | 0 | 0 | 1 | Ref |
| 10459 | MN698248 | Unassigned | Unassigned | Unassigned | 1003_0 | Clustered | 4 | VC_1003_0 | 4 | 0.8718 | 0.99983781 | 0.8716 | 0 | 0 | 0 | 1 | Ref |
| 10460 | MN703405 | Unassigned | Unassigned | Unassigned | 221_0 | Clustered | 22 | VC_221_0 | 22 | 0.7447 | 1 | 0.7447 | 0 | 0 | 0 | 1 | Ref |
| 10461 | MN703406 | Unassigned | Unassigned | Unassigned | 888_0 | Clustered | 28 | VC_888_0 | 28 | 0.5497 | 1 | 0.5497 | 0 | 0 | 0 | 1 | Ref |
| 10462 | MN703407 | Unassigned | Unassigned | Unassigned | 1345_0 | Clustered | 2 | VC_1345_0 | 2 | 0.1133 | 0.95579173 | 0.1083 | 0 | 0 | 0 | 1 | Ref |
| 10463 | MN703408 | Unassigned | Unassigned | Unassigned | 217_0 | Clustered | 5 | VC_217_0 | 5 | 0.069 | 0.99999998 | 0.069 | 0 | 0 | 0 | 1 | Ref |
| 10464 | MN703409 | Unassigned | Unassigned | Unassigned | 1166_0 | Clustered | 20 | VC_1166_0 | 20 | 0.99 | 1 | 0.99 | 0 | 0 | 0 | 1 | Ref |
| 10465 | MN703410 | Unassigned | Unassigned | Unassigned | 888_0 | Clustered | 28 | VC_888_0 | 28 | 0.5497 | 1 | 0.5497 | 0 | 0 | 0 | 1 | Ref |
| 10466 | MN703411 | Unassigned | Unassigned | Unassigned | 1074_0 | Clustered | 19 | VC_1074_0 | 19 | 0.9198 | 1 | 0.9198 | 0 | 0 | 0 | 1 | Ref |
| 10467 | MN703412 | Unassigned | Unassigned | Unassigned | 1164_0 | Clustered | 25 | VC_1164_0 | 25 | 0.7312 | 1 | 0.7312 | 0 | 0 | 0 | 1 | Ref |
| 10468 | MN703413 | Unassigned | Unassigned | Unassigned | 1207_0 | Clustered | 29 | VC_1207_0 | 29 | 0.8225 | 1 | 0.8225 | 0 | 0 | 0 | 1 | Ref |
| 10469 | MN703414 | Unassigned | Unassigned | Unassigned | 1345_0 | Clustered | 2 | VC_1345_0 | 2 | 0.1133 | 0.95579173 | 0.1083 | 0 | 0 | 0 | 1 | Ref |
| 10470 | MN703415 | Unassigned | Unassigned | Unassigned | 371_20 | Clustered/Singleton | 1 | VC_371_20 | 1 | 0 | 1 | 0 | 0 | 0 | 0 | 1 | Ref |
| 10471 | MN709127 | Unassigned | Unassigned | Unassigned | 590_0 | Clustered | 27 | VC_590_0 | 27 | 0.9947 | 1 | 0.9947 | 0 | 0 | 0 | 1 | Ref |
| 10472 | MN709128 | Unassigned | Unassigned | Unassigned | 492_1 | Clustered | 20 | VC_492_1 | 20 | 0.4176 | 1 | 0.4176 | 0 | 0 | 0 | 1 | Ref |
| 10473 | MN710383 | Unassigned | Unassigned | Unassigned | 589_0 | Clustered | 2 | VC_589_0 | 2 | 1 | 1 | 1 | 0 | 0 | 0 | 1 | Ref |
| 10474 | MN715150 | Unassigned | Unassigned | Unassigned | 451_0 | Clustered | 80 | VC_451_0 | 80 | 0.9991 | 1 | 0.9991 | 0 | 0 | 0 | 1 | Ref |
| 10475 | MN715356 | Unassigned | Unassigned | Unassigned | 813_1 | Clustered | 24 | VC_813_1 | 24 | 0.535 | 1 | 0.535 | 0 | 0 | 0 | 1 | Ref |
| 10476 | MN716856 | Unassigned | Unassigned | Unassigned | 283_131 | Clustered/Singleton | 1 | VC_283_131 | 1 | 0 | 1 | 0 | 0 | 0 | 0 | 1 | Ref |
| 10477 | MN718463 | Unassigned | Unassigned | Unassigned | 766_0 | Clustered | 2 | VC_766_0 | 2 | 0.2674 | 0.94916107 | 0.2538 | 0 | 0 | 0 | 1 | Ref |
| 10478 | MN719123 | Unassigned | Unassigned | Unassigned | 17_0 | Clustered | 9 | VC_17_0 | 9 | 0.6969 | 1 | 0.6969 | 0 | 0 | 0 | 1 | Ref |
| 10479 | MN722429 | Unassigned | Unassigned | Unassigned | 637_0 | Clustered | 115 | VC_637_0 | 115 | 0.731 | 1 | 0.731 | 0 | 0 | 0 | 0.8764 | Ref |
| 10480 | MN723850 | Unassigned | Unassigned | Unassigned | 492_1 | Clustered | 20 | VC_492_1 | 20 | 0.4176 | 1 | 0.4176 | 0 | 0 | 0 | 1 | Ref |
| 10481 | MN727882 | Unassigned | Unassigned | Unassigned | 100_0 | Clustered | 15 | VC_100_0 | 15 | 0.6536 | 1 | 0.6536 | 0 | 0 | 0 | 1 | Ref |
| 10482 | MN728683 | Unassigned | Unassigned | Unassigned | 223_0 | Clustered | 38 | VC_223_0 | 38 | 0.9095 | 1 | 0.9095 | 0 | 0 | 0 | 1 | Ref |
| 10483 | MN728684 | Unassigned | Unassigned | Unassigned | 517_1 | Clustered | 108 | VC_517_1 | 108 | 0.8483 | 1 | 0.8483 | 0 | 0 | 0 | 0.9955 | Ref |
| 10484 | MN729595 | Unassigned | Unassigned | Unassigned | 129_0 | Clustered | 43 | VC_129_0 | 43 | 0.886 | 1 | 0.886 | 0 | 0 | 0 | 1 | Ref |
| 10485 | MN729596 | Unassigned | Unassigned | Unassigned | 129_0 | Clustered | 43 | VC_129_0 | 43 | 0.886 | 1 | 0.886 | 0 | 0 | 0 | 1 | Ref |
| 10486 | MN729597 | Unassigned | Unassigned | Unassigned | 129_0 | Clustered | 43 | VC_129_0 | 43 | 0.886 | 1 | 0.886 | 0 | 0 | 0 | 1 | Ref |
| 10487 | MN729598 | Unassigned | Unassigned | Unassigned | 129_0 | Clustered | 43 | VC_129_0 | 43 | 0.886 | 1 | 0.886 | 0 | 0 | 0 | 1 | Ref |
| 10488 | MN729599 | Unassigned | Unassigned | Unassigned | 129_0 | Clustered | 43 | VC_129_0 | 43 | 0.886 | 1 | 0.886 | 0 | 0 | 0 | 1 | Ref |
| 10489 | MN729600 | Unassigned | Unassigned | Unassigned | 129_0 | Clustered | 43 | VC_129_0 | 43 | 0.886 | 1 | 0.886 | 0 | 0 | 0 | 1 | Ref |
| 10490 | MN732867 | Unassigned | Unassigned | Unassigned | 582_0 | Clustered | 4 | VC_582_0 | 4 | 0.0589 | 0.99996986 | 0.0589 | 0 | 0 | 0 | 1 | Ref |
| 10491 | MN732883 | Unassigned | Unassigned | Unassigned | 492_1 | Clustered | 20 | VC_492_1 | 20 | 0.4176 | 1 | 0.4176 | 0 | 0 | 0 | 1 | Ref |
| 10492 | MN732896 | Unassigned | Unassigned | Unassigned | 1010_0 | Clustered | 2 | VC_1010_0 | 2 | 0.2036 | 0.77337265 | 0.1574 | 0 | 0 | 0 | 1 | Ref |
| 10493 | MN734437 | Unassigned | Unassigned | Unassigned | 752_0 | Clustered | 3 | VC_752_0 | 3 | 0.0788 | 0.9984602 | 0.0787 | 0 | 0 | 0 | 1 | Ref |
| 10494 | MN734438 | Unassigned | Unassigned | Unassigned | 752_0 | Clustered | 3 | VC_752_0 | 3 | 0.0788 | 0.9984602 | 0.0787 | 0 | 0 | 0 | 1 | Ref |
| 10495 | MN734439 | Unassigned | Unassigned | Unassigned | 752_0 | Clustered | 3 | VC_752_0 | 3 | 0.0788 | 0.9984602 | 0.0787 | 0 | 0 | 0 | 1 | Ref |
| 10496 | MN735429 | Unassigned | Unassigned | Unassigned | 1160_0 | Clustered | 73 | VC_1160_0 | 73 | 0.9835 | 1 | 0.9835 | 0 | 0 | 0 | 1 | Ref |
| 10497 | MN735430 | Unassigned | Unassigned | Unassigned | 1240_0 | Clustered | 82 | VC_1240_0 | 82 | 0.7945 | 1 | 0.7945 | 0 | 0 | 0 | 1 | Ref |
| 10498 | MN735431 | Unassigned | Unassigned | Unassigned | 520_12 | Clustered/Singleton | 1 | VC_520_12 | 1 | 0 | 1 | 0 | 0 | 0 | 0 | 1 | Ref |
| 10499 | MN735432 | Unassigned | Unassigned | Unassigned | 1046_0 | Clustered | 7 | VC_1046_0 | 7 | 0.9413 | 1 | 0.9413 | 0 | 0 | 0 | 1 | Ref |
| 10500 | MN735434 | Unassigned | Unassigned | Unassigned | 558_79 | Clustered/Singleton | 1 | VC_558_79 | 1 | 0 | 1 | 0 | 0 | 0 | 0 | 1 | Ref |
| 10501 | MN735435 | Unassigned | Unassigned | Unassigned | 1240_0 | Clustered | 82 | VC_1240_0 | 82 | 0.7945 | 1 | 0.7945 | 0 | 0 | 0 | 1 | Ref |
| 10502 | MN737551 | Unassigned | Unassigned | Unassigned | 461_0 | Clustered | 96 | VC_461_0 | 96 | 0.9307 | 1 | 0.9307 | 0 | 0 | 0 | 0.9575 | Ref |
| 10503 | MN737622 | Unassigned | Unassigned | Unassigned | 233_0 | Clustered | 2 | VC_233_0 | 2 | 0.7179 | 0.72985431 | 0.5239 | 0 | 0 | 0 | 1 | Ref |
| 10504 | MN781580 | Unassigned | Unassigned | Unassigned | 283_21 | Clustered/Singleton | 1 | VC_283_21 | 1 | 0 | 1 | 0 | 0 | 0 | 0 | 1 | Ref |
| 10505 | MN781674 | Unassigned | Unassigned | Unassigned | 942_0 | Clustered | 36 | VC_942_0 | 36 | 0.7848 | 1 | 0.7848 | 0 | 0 | 0 | 1 | Ref |
| 10506 | MN782535 | Unassigned | Unassigned | Unassigned | 492_1 | Clustered | 20 | VC_492_1 | 20 | 0.4176 | 1 | 0.4176 | 0 | 0 | 0 | 1 | Ref |
| 10507 | MN783016 | Unassigned | Unassigned | Unassigned | 938_0 | Clustered | 99 | VC_938_0 | 99 | 0.9119 | 1 | 0.9119 | 0 | 0 | 0 | 0.9928 | Ref |
| 10508 | MN788369 | Unassigned | Unassigned | Unassigned | 131_0 | Clustered | 17 | VC_131_0 | 17 | 0.2671 | 1 | 0.2671 | 0 | 0 | 0 | 1 | Ref |
| 10509 | MN794000 | Unassigned | Unassigned | Unassigned | 111_0 | Clustered | 91 | VC_111_0 | 91 | 0.9587 | 1 | 0.9587 | 0 | 0 | 0 | 1 | Ref |
| 10510 | MN794001 | Unassigned | Unassigned | Unassigned | 111_0 | Clustered | 91 | VC_111_0 | 91 | 0.9587 | 1 | 0.9587 | 0 | 0 | 0 | 1 | Ref |
| 10511 | MN794002 | Unassigned | Unassigned | Unassigned | 111_0 | Clustered | 91 | VC_111_0 | 91 | 0.9587 | 1 | 0.9587 | 0 | 0 | 0 | 1 | Ref |
| 10512 | MN794003 | Unassigned | Unassigned | Unassigned | 111_0 | Clustered | 91 | VC_111_0 | 91 | 0.9587 | 1 | 0.9587 | 0 | 0 | 0 | 1 | Ref |
| 10513 | MN794232 | Unassigned | Unassigned | Unassigned | 531_1 | Clustered | 6 | VC_531_1 | 6 | 0.1481 | 0.99978665 | 0.148 | 0 | 0 | 0 | 1 | Ref |
| 10514 | MN807240 | Unassigned | Unassigned | Unassigned | 1027_0 | Clustered | 20 | VC_1027_0 | 20 | 0.7114 | 1 | 0.7114 | 0 | 0 | 0 | 1 | Ref |
| 10515 | MN807246 | Unassigned | Unassigned | Unassigned | 1240_0 | Clustered | 82 | VC_1240_0 | 82 | 0.7945 | 1 | 0.7945 | 0 | 0 | 0 | 1 | Ref |
| 10516 | MN807247 | Unassigned | Unassigned | Unassigned | 515_0 | Clustered | 16 | VC_515_0 | 16 | 0.3793 | 1 | 0.3793 | 0 | 0 | 0 | 1 | Ref |
| 10517 | MN807248 | Unassigned | Unassigned | Unassigned | 519_0 | Clustered | 39 | VC_519_0 | 39 | 0.8156 | 1 | 0.8156 | 0 | 0 | 0 | 1 | Ref |
| 10518 | MN807249 | Unassigned | Unassigned | Unassigned | 519_0 | Clustered | 39 | VC_519_0 | 39 | 0.8156 | 1 | 0.8156 | 0 | 0 | 0 | 1 | Ref |
| 10519 | MN807250 | Unassigned | Unassigned | Unassigned | 519_0 | Clustered | 39 | VC_519_0 | 39 | 0.8156 | 1 | 0.8156 | 0 | 0 | 0 | 1 | Ref |
| 10520 | MN807295 | Unassigned | Unassigned | Unassigned | 699_0 | Clustered | 71 | VC_699_0 | 71 | 0.9352 | 1 | 0.9352 | 0 | 0 | 0 | 1 | Ref |
| 10521 | MN812203 | Unassigned | Unassigned | Unassigned | 1021_0 | Clustered | 10 | VC_1021_0 | 10 | 0.8876 | 1 | 0.8876 | 0 | 0 | 0 | 1 | Ref |
| 10522 | MN812204 | Unassigned | Unassigned | Unassigned | 1021_0 | Clustered | 10 | VC_1021_0 | 10 | 0.8876 | 1 | 0.8876 | 0 | 0 | 0 | 1 | Ref |
| 10523 | MN812205 | Unassigned | Unassigned | Unassigned | 1021_0 | Clustered | 10 | VC_1021_0 | 10 | 0.8876 | 1 | 0.8876 | 0 | 0 | 0 | 1 | Ref |
| 10524 | MN812206 | Unassigned | Unassigned | Unassigned | 1013_0 | Clustered | 34 | VC_1013_0 | 34 | 0.9939 | 1 | 0.9939 | 0 | 0 | 0 | 1 | Ref |
| 10525 | MN812207 | Unassigned | Unassigned | Unassigned | 1013_0 | Clustered | 34 | VC_1013_0 | 34 | 0.9939 | 1 | 0.9939 | 0 | 0 | 0 | 1 | Ref |
| 10526 | MN812208 | Unassigned | Unassigned | Unassigned | 1013_0 | Clustered | 34 | VC_1013_0 | 34 | 0.9939 | 1 | 0.9939 | 0 | 0 | 0 | 1 | Ref |
| 10527 | MN812209 | Unassigned | Unassigned | Unassigned | 1013_0 | Clustered | 34 | VC_1013_0 | 34 | 0.9939 | 1 | 0.9939 | 0 | 0 | 0 | 1 | Ref |
| 10528 | MN812210 | Unassigned | Unassigned | Unassigned | 1013_0 | Clustered | 34 | VC_1013_0 | 34 | 0.9939 | 1 | 0.9939 | 0 | 0 | 0 | 1 | Ref |

| | | | | | | | | | | | | | | | | | |
|---|---|---|---|---|---|---|---|---|---|---|---|---|---|---|---|---|---|
| 10529 | MN812212 | Unassigned | Unassigned | Unassigned | 1013_0 | Clustered | 34 | VC_1013_0 | 34 | 0.9939 | 1 | 0.9939 | 0 | 0 | 0 | 1 | Ref |
| 10530 | MN812213 | Unassigned | Unassigned | Unassigned | 1013_0 | Clustered | 34 | VC_1013_0 | 34 | 0.9939 | 1 | 0.9939 | 0 | 0 | 0 | 1 | Ref |
| 10531 | MN812214 | Unassigned | Unassigned | Unassigned | 1013_0 | Clustered | 34 | VC_1013_0 | 34 | 0.9939 | 1 | 0.9939 | 0 | 0 | 0 | 1 | Ref |
| 10532 | MN812215 | Unassigned | Unassigned | Unassigned | 1013_0 | Clustered | 34 | VC_1013_0 | 34 | 0.9939 | 1 | 0.9939 | 0 | 0 | 0 | 1 | Ref |
| 10533 | MN812216 | Unassigned | Unassigned | Unassigned | 1013_0 | Clustered | 34 | VC_1013_0 | 34 | 0.9939 | 1 | 0.9939 | 0 | 0 | 0 | 1 | Ref |
| 10534 | MN812217 | Unassigned | Unassigned | Unassigned | 1013_0 | Clustered | 34 | VC_1013_0 | 34 | 0.9939 | 1 | 0.9939 | 0 | 0 | 0 | 1 | Ref |
| 10535 | MN812218 | Unassigned | Unassigned | Unassigned | 1013_0 | Clustered | 34 | VC_1013_0 | 34 | 0.9939 | 1 | 0.9939 | 0 | 0 | 0 | 1 | Ref |
| 10536 | MN812219 | Unassigned | Unassigned | Unassigned | 1013_0 | Clustered | 34 | VC_1013_0 | 34 | 0.9939 | 1 | 0.9939 | 0 | 0 | 0 | 1 | Ref |
| 10537 | MN812220 | Unassigned | Unassigned | Unassigned | 1013_0 | Clustered | 34 | VC_1013_0 | 34 | 0.9939 | 1 | 0.9939 | 0 | 0 | 0 | 1 | Ref |
| 10538 | MN812221 | Unassigned | Unassigned | Unassigned | 1013_0 | Clustered | 34 | VC_1013_0 | 34 | 0.9939 | 1 | 0.9939 | 0 | 0 | 0 | 1 | Ref |
| 10539 | MN812222 | Unassigned | Unassigned | Unassigned | 1013_0 | Clustered | 34 | VC_1013_0 | 34 | 0.9939 | 1 | 0.9939 | 0 | 0 | 0 | 1 | Ref |
| 10540 | MN812223 | Unassigned | Unassigned | Unassigned | 1013_0 | Clustered | 34 | VC_1013_0 | 34 | 0.9939 | 1 | 0.9939 | 0 | 0 | 0 | 1 | Ref |
| 10541 | MN812224 | Unassigned | Unassigned | Unassigned | 1013_0 | Clustered | 34 | VC_1013_0 | 34 | 0.9939 | 1 | 0.9939 | 0 | 0 | 0 | 1 | Ref |
| 10542 | MN812225 | Unassigned | Unassigned | Unassigned | 1013_0 | Clustered | 34 | VC_1013_0 | 34 | 0.9939 | 1 | 0.9939 | 0 | 0 | 0 | 1 | Ref |
| 10543 | MN812226 | Unassigned | Unassigned | Unassigned | 1013_0 | Clustered | 34 | VC_1013_0 | 34 | 0.9939 | 1 | 0.9939 | 0 | 0 | 0 | 1 | Ref |
| 10544 | MN812227 | Unassigned | Unassigned | Unassigned | 1013_0 | Clustered | 34 | VC_1013_0 | 34 | 0.9939 | 1 | 0.9939 | 0 | 0 | 0 | 1 | Ref |
| 10545 | MN812228 | Unassigned | Unassigned | Unassigned | 1013_0 | Clustered | 34 | VC_1013_0 | 34 | 0.9939 | 1 | 0.9939 | 0 | 0 | 0 | 1 | Ref |
| 10546 | MN812229 | Unassigned | Unassigned | Unassigned | 1013_0 | Clustered | 34 | VC_1013_0 | 34 | 0.9939 | 1 | 0.9939 | 0 | 0 | 0 | 1 | Ref |
| 10547 | MN812230 | Unassigned | Unassigned | Unassigned | 1013_0 | Clustered | 34 | VC_1013_0 | 34 | 0.9939 | 1 | 0.9939 | 0 | 0 | 0 | 1 | Ref |
| 10548 | MN812231 | Unassigned | Unassigned | Unassigned | 1013_0 | Clustered | 34 | VC_1013_0 | 34 | 0.9939 | 1 | 0.9939 | 0 | 0 | 0 | 1 | Ref |
| 10549 | MN812232 | Unassigned | Unassigned | Unassigned | 1013_0 | Clustered | 34 | VC_1013_0 | 34 | 0.9939 | 1 | 0.9939 | 0 | 0 | 0 | 1 | Ref |
| 10550 | MN812233 | Unassigned | Unassigned | Unassigned | 1013_0 | Clustered | 34 | VC_1013_0 | 34 | 0.9939 | 1 | 0.9939 | 0 | 0 | 0 | 1 | Ref |
| 10551 | MN812234 | Unassigned | Unassigned | Unassigned | 1013_0 | Clustered | 34 | VC_1013_0 | 34 | 0.9939 | 1 | 0.9939 | 0 | 0 | 0 | 1 | Ref |
| 10552 | MN812235 | Unassigned | Unassigned | Unassigned | 1013_0 | Clustered | 34 | VC_1013_0 | 34 | 0.9939 | 1 | 0.9939 | 0 | 0 | 0 | 1 | Ref |
| 10553 | MN812236 | Unassigned | Unassigned | Unassigned | 1013_0 | Clustered | 34 | VC_1013_0 | 34 | 0.9939 | 1 | 0.9939 | 0 | 0 | 0 | 1 | Ref |
| 10554 | MN812237 | Unassigned | Unassigned | Unassigned | 1013_0 | Clustered | 34 | VC_1013_0 | 34 | 0.9939 | 1 | 0.9939 | 0 | 0 | 0 | 1 | Ref |
| 10555 | MN812238 | Unassigned | Unassigned | Unassigned | 1013_0 | Clustered | 34 | VC_1013_0 | 34 | 0.9939 | 1 | 0.9939 | 0 | 0 | 0 | 1 | Ref |
| 10556 | MN812240 | Unassigned | Unassigned | Unassigned | 1013_0 | Clustered | 34 | VC_1013_0 | 34 | 0.9939 | 1 | 0.9939 | 0 | 0 | 0 | 1 | Ref |
| 10557 | MN812241 | Unassigned | Unassigned | Unassigned | 1013_0 | Clustered | 34 | VC_1013_0 | 34 | 0.9939 | 1 | 0.9939 | 0 | 0 | 0 | 1 | Ref |
| 10558 | MN812685 | Unassigned | Unassigned | Unassigned | 841_0 | Clustered | 10 | VC_841_0 | 10 | 0.5997 | 1 | 0.5997 | 0 | 0 | 0 | 1 | Ref |
| 10559 | MN812686 | Unassigned | Unassigned | Unassigned | 451_0 | Clustered | 80 | VC_451_0 | 80 | 0.9991 | 1 | 0.9991 | 0 | 0 | 0 | 1 | Ref |
| 10560 | MN812687 | Unassigned | Unassigned | Unassigned | 841_0 | Clustered | 10 | VC_841_0 | 10 | 0.5997 | 1 | 0.5997 | 0 | 0 | 0 | 1 | Ref |
| 10561 | MN812688 | Unassigned | Unassigned | Unassigned | 792_0 | Clustered | 35 | VC_792_0 | 35 | 0.9828 | 1 | 0.9828 | 0 | 0 | 0 | 1 | Ref |
| 10562 | MN812690 | Unassigned | Unassigned | Unassigned | 984_0 | Clustered | 38 | VC_984_0 | 38 | 0.9758 | 1 | 0.9758 | 0 | 0 | 0 | 1 | Ref |
| 10563 | MN812691 | Unassigned | Unassigned | Unassigned | 841_0 | Clustered | 10 | VC_841_0 | 10 | 0.5997 | 1 | 0.5997 | 0 | 0 | 0 | 1 | Ref |
| 10564 | MN812692 | Unassigned | Unassigned | Unassigned | 984_0 | Clustered | 38 | VC_984_0 | 38 | 0.9758 | 1 | 0.9758 | 0 | 0 | 0 | 1 | Ref |
| 10565 | MN812693 | Unassigned | Unassigned | Unassigned | 393_0 | Clustered | 7 | VC_393_0 | 7 | 0.1775 | 1 | 0.1775 | 0 | 0 | 0 | 1 | Ref |
| 10566 | MN812694 | Unassigned | Unassigned | Unassigned | 78_4 | Clustered/Singleton | 1 | VC_78_4 | 1 | 0 | 1 | 0 | 0 | 0 | 0 | 1 | Ref |
| 10567 | MN812722 | Unassigned | Unassigned | Unassigned | 354_0 | Clustered | 4 | VC_354_0 | 4 | 0.1749 | 0.99998684 | 0.1749 | 0 | 0 | 0 | 0.8333 | Ref |
| 10568 | MN813048 | Unassigned | Unassigned | Unassigned | 131_0 | Clustered | 17 | VC_131_0 | 17 | 0.2671 | 1 | 0.2671 | 0 | 0 | 0 | 1 | Ref |
| 10569 | MN813049 | Unassigned | Unassigned | Unassigned | 131_0 | Clustered | 17 | VC_131_0 | 17 | 0.2671 | 1 | 0.2671 | 0 | 0 | 0 | 1 | Ref |
| 10570 | MN813050 | Unassigned | Unassigned | Unassigned | 131_0 | Clustered | 17 | VC_131_0 | 17 | 0.2671 | 1 | 0.2671 | 0 | 0 | 0 | 1 | Ref |
| 10571 | MN813051 | Unassigned | Unassigned | Unassigned | 131_0 | Clustered | 17 | VC_131_0 | 17 | 0.2671 | 1 | 0.2671 | 0 | 0 | 0 | 1 | Ref |
| 10572 | MN813052 | Unassigned | Unassigned | Unassigned | 131_0 | Clustered | 17 | VC_131_0 | 17 | 0.2671 | 1 | 0.2671 | 0 | 0 | 0 | 1 | Ref |
| 10573 | MN813053 | Unassigned | Unassigned | Unassigned | 131_0 | Clustered | 17 | VC_131_0 | 17 | 0.2671 | 1 | 0.2671 | 0 | 0 | 0 | 1 | Ref |
| 10574 | MN813675 | Unassigned | Unassigned | Unassigned | 765_0 | Clustered | 132 | VC_765_0 | 132 | 1 | 1 | 1 | 0 | 0 | 0 | 1 | Ref |
| 10575 | MN813676 | Unassigned | Unassigned | Unassigned | 1207_0 | Clustered | 29 | VC_1207_0 | 29 | 0.8225 | 1 | 0.8225 | 0 | 0 | 0 | 1 | Ref |
| 10576 | MN813677 | Unassigned | Unassigned | Unassigned | 765_0 | Clustered | 132 | VC_765_0 | 132 | 1 | 1 | 1 | 0 | 0 | 0 | 1 | Ref |
| 10577 | MN813678 | Unassigned | Unassigned | Unassigned | 508_0 | Clustered | 32 | VC_508_0 | 32 | 0.906 | 1 | 0.906 | 0 | 0 | 0 | 1 | Ref |
| 10578 | MN813679 | Unassigned | Unassigned | Unassigned | 765_0 | Clustered | 132 | VC_765_0 | 132 | 1 | 1 | 1 | 0 | 0 | 0 | 1 | Ref |
| 10579 | MN813680 | Unassigned | Unassigned | Unassigned | 1160_0 | Clustered | 73 | VC_1160_0 | 73 | 0.9835 | 1 | 0.9835 | 0 | 0 | 0 | 1 | Ref |
| 10580 | MN813682 | Unassigned | Unassigned | Unassigned | 1349_0 | Clustered | 7 | VC_1349_0 | 7 | 0.2495 | 1 | 0.2495 | 0 | 0 | 0 | 1 | Ref |
| 10581 | MN813683 | Unassigned | Unassigned | Unassigned | 765_0 | Clustered | 132 | VC_765_0 | 132 | 1 | 1 | 1 | 0 | 0 | 0 | 1 | Ref |
| 10582 | MN813684 | Unassigned | Unassigned | Unassigned | 1349_0 | Clustered | 7 | VC_1349_0 | 7 | 0.2495 | 1 | 0.2495 | 0 | 0 | 0 | 1 | Ref |
| 10583 | MN813687 | Unassigned | Unassigned | Unassigned | 573_0 | Clustered | 2 | VC_573_0 | 2 | 0.11 | 0.95639318 | 0.1052 | 0 | 0 | 0 | 1 | Ref |
| 10584 | MN813688 | Unassigned | Unassigned | Unassigned | 765_0 | Clustered | 132 | VC_765_0 | 132 | 1 | 1 | 1 | 0 | 0 | 0 | 1 | Ref |
| 10585 | MN813689 | Unassigned | Unassigned | Unassigned | 765_0 | Clustered | 132 | VC_765_0 | 132 | 1 | 1 | 1 | 0 | 0 | 0 | 1 | Ref |
| 10586 | MN813690 | Unassigned | Unassigned | Unassigned | 765_0 | Clustered | 132 | VC_765_0 | 132 | 1 | 1 | 1 | 0 | 0 | 0 | 1 | Ref |
| 10587 | MN813691 | Unassigned | Unassigned | Unassigned | 765_0 | Clustered | 132 | VC_765_0 | 132 | 1 | 1 | 1 | 0 | 0 | 0 | 1 | Ref |
| 10588 | MN813692 | Unassigned | Unassigned | Unassigned | 765_0 | Clustered | 132 | VC_765_0 | 132 | 1 | 1 | 1 | 0 | 0 | 0 | 1 | Ref |
| 10589 | MN813693 | Unassigned | Unassigned | Unassigned | 372_4 | Clustered | 3 | VC_372_4 | 3 | 0.033 | 0.99853257 | 0.0329 | 0 | 0 | 0 | 1 | Ref |
| 10590 | MN813694 | Unassigned | Unassigned | Unassigned | 1177_0 | Clustered | 14 | VC_1177_0 | 14 | 0.7847 | 1 | 0.7847 | 0 | 0 | 0 | 0.989 | Ref |
| 10591 | MN813695 | Unassigned | Unassigned | Unassigned | 950_0 | Clustered | 38 | VC_950_0 | 38 | 0.9443 | 1 | 0.9443 | 0 | 0 | 0 | 1 | Ref |
| 10592 | MN813696 | Unassigned | Unassigned | Unassigned | 517_1 | Clustered | 108 | VC_517_1 | 108 | 0.8483 | 1 | 0.8483 | 0 | 0 | 0 | 0.9955 | Ref |
| 10593 | MN813697 | Unassigned | Unassigned | Unassigned | 517_0 | Clustered | 18 | VC_517_0 | 18 | 0.1599 | 1 | 0.1599 | 0 | 0 | 0 | 1 | Ref |
| 10594 | MN813698 | Unassigned | Unassigned | Unassigned | 1183_0 | Clustered | 6 | VC_1183_0 | 6 | 0.995 | 0.99996974 | 0.995 | 0 | 0 | 0 | 1 | Ref |
| 10595 | MN813699 | Unassigned | Unassigned | Unassigned | 514_0 | Clustered | 75 | VC_514_0 | 75 | 0.9402 | 1 | 0.9402 | 0 | 0 | 0 | 1 | Ref |
| 10596 | MN813700 | Unassigned | Unassigned | Unassigned | 325_12 | Clustered/Singleton | 1 | VC_325_12 | 1 | 0 | 1 | 0 | 0 | 0 | 0 | 1 | Ref |
| 10597 | MN830254 | Unassigned | Unassigned | Unassigned | 877_0 | Clustered | 2 | VC_877_0 | 2 | 0.7762 | 0.86822376 | 0.6739 | 0 | 0 | 0 | 1 | Ref |
| 10598 | MN830255 | Unassigned | Unassigned | Unassigned | 877_0 | Clustered | 2 | VC_877_0 | 2 | 0.7762 | 0.86822376 | 0.6739 | 0 | 0 | 0 | 1 | Ref |
| 10599 | MN830256 | Unassigned | Unassigned | Unassigned | 309_0 | Clustered | 3 | VC_309_0 | 3 | 0.1265 | 0.99498896 | 0.1258 | 0 | 0 | 0 | 1 | Ref |
| 10600 | MN830258 | Unassigned | Unassigned | Unassigned | 272_0 | Clustered | 11 | VC_272_0 | 11 | 0.4915 | 1 | 0.4915 | 0 | 0 | 0 | 0.8727 | Ref |
| 10601 | MN840485 | Unassigned | Unassigned | Unassigned | 456_0 | Clustered | 62 | VC_456_0 | 62 | 0.9026 | 1 | 0.9026 | 0 | 0 | 0 | 1 | Ref |
| 10602 | MN840486 | Unassigned | Unassigned | Unassigned | 591_0 | Clustered | 14 | VC_591_0 | 14 | 0.5184 | 1 | 0.5184 | 0 | 0 | 0 | 1 | Ref |
| 10603 | MN844877 | Unassigned | Unassigned | Unassigned | 154_0 | Clustered | 4 | VC_154_0 | 4 | 0.1766 | 0.99995106 | 0.1766 | 0 | 0 | 0 | 1 | Ref |
| 10604 | MN850564 | Unassigned | Unassigned | Unassigned | 329_3 | Clustered | 24 | VC_329_3 | 24 | 0.1206 | 1 | 0.1206 | 0 | 0 | 0 | 0.7283 | Ref |
| 10605 | MN850565 | Unassigned | Unassigned | Unassigned | 847_1 | Clustered | 26 | VC_847_1 | 26 | 0.6323 | 1 | 0.6323 | 0 | 0 | 0 | 0.9815 | Ref |
| 10606 | MN850566 | Unassigned | Unassigned | Unassigned | 329_3 | Clustered | 24 | VC_329_3 | 24 | 0.1206 | 1 | 0.1206 | 0 | 0 | 0 | 0.7283 | Ref |
| 10607 | MN850567 | Unassigned | Unassigned | Unassigned | 941_0 | Clustered | 40 | VC_941_0 | 40 | 0.6903 | 1 | 0.6903 | 0 | 0 | 0 | 1 | Ref |
| 10608 | MN850568 | Unassigned | Unassigned | Unassigned | 156_0 | Clustered | 12 | VC_156_0 | 12 | 0.9062 | 1 | 0.9062 | 0 | 0 | 0 | 1 | Ref |
| 10609 | MN850569 | Unassigned | Unassigned | Unassigned | 941_0 | Clustered | 40 | VC_941_0 | 40 | 0.6903 | 1 | 0.6903 | 0 | 0 | 0 | 1 | Ref |
| 10610 | MN850570 | Unassigned | Unassigned | Unassigned | 156_0 | Clustered | 12 | VC_156_0 | 12 | 0.9062 | 1 | 0.9062 | 0 | 0 | 0 | 1 | Ref |
| 10611 | MN850571 | Unassigned | Unassigned | Unassigned | 847_1 | Clustered | 26 | VC_847_1 | 26 | 0.6323 | 1 | 0.6323 | 0 | 0 | 0 | 0.9815 | Ref |
| 10612 | MN850572 | Unassigned | Unassigned | Unassigned | 941_0 | Clustered | 40 | VC_941_0 | 40 | 0.6903 | 1 | 0.6903 | 0 | 0 | 0 | 1 | Ref |
| 10613 | MN850573 | Unassigned | Unassigned | Unassigned | 848_0 | Clustered | 28 | VC_848_0 | 28 | 0.7927 | 1 | 0.7927 | 0 | 0 | 0 | 1 | Ref |
| 10614 | MN850574 | Unassigned | Unassigned | Unassigned | 590_0 | Clustered | 27 | VC_590_0 | 27 | 0.9947 | 1 | 0.9947 | 0 | 0 | 0 | 1 | Ref |
| 10615 | MN850575 | Unassigned | Unassigned | Unassigned | 835_0 | Clustered | 67 | VC_835_0 | 67 | 0.9624 | 1 | 0.9624 | 0 | 0 | 0 | 1 | Ref |
| 10616 | MN850576 | Unassigned | Unassigned | Unassigned | 772_19 | Clustered | 16 | VC_772_19 | 16 | 0.0975 | 0.9970466 | 0.0972 | 0 | 0 | 0 | 0.8083 | Ref |
| 10617 | MN850577 | Unassigned | Unassigned | Unassigned | 329_3 | Clustered | 24 | VC_329_3 | 24 | 0.1206 | 1 | 0.1206 | 0 | 0 | 0 | 0.7283 | Ref |
| 10618 | MN850578 | Unassigned | Unassigned | Unassigned | 772_19 | Clustered | 16 | VC_772_19 | 16 | 0.0975 | 0.9970466 | 0.0972 | 0 | 0 | 0 | 0.8083 | Ref |
| 10619 | MN850579 | Unassigned | Unassigned | Unassigned | 286_1 | Clustered | 21 | VC_286_1 | 21 | 0.1696 | 1 | 0.1696 | 0 | 0 | 0 | 0.9048 | Ref |
| 10620 | MN850580 | Unassigned | Unassigned | Unassigned | 329_3 | Clustered | 24 | VC_329_3 | 24 | 0.1206 | 1 | 0.1206 | 0 | 0 | 0 | 0.7283 | Ref |
| 10621 | MN850581 | Unassigned | Unassigned | Unassigned | 705_0 | Clustered | 11 | VC_705_0 | 11 | 0.8861 | 1 | 0.8861 | 0 | 0 | 0 | 1 | Ref |
| 10622 | MN850582 | Unassigned | Unassigned | Unassigned | 942_0 | Clustered | 36 | VC_942_0 | 36 | 0.7848 | 1 | 0.7848 | 0 | 0 | 0 | 1 | Ref |
| 10623 | MN850583 | Unassigned | Unassigned | Unassigned | 156_0 | Clustered | 12 | VC_156_0 | 12 | 0.9062 | 1 | 0.9062 | 0 | 0 | 0 | 1 | Ref |
| 10624 | MN850584 | Unassigned | Unassigned | Unassigned | 848_0 | Clustered | 28 | VC_848_0 | 28 | 0.7927 | 1 | 0.7927 | 0 | 0 | 0 | 1 | Ref |
| 10625 | MN850585 | Unassigned | Unassigned | Unassigned | 329_3 | Clustered | 24 | VC_329_3 | 24 | 0.1206 | 1 | 0.1206 | 0 | 0 | 0 | 0.7283 | Ref |
| 10626 | MN850586 | Unassigned | Unassigned | Unassigned | 942_0 | Clustered | 36 | VC_942_0 | 36 | 0.7848 | 1 | 0.7848 | 0 | 0 | 0 | 1 | Ref |
| 10627 | MN850587 | Unassigned | Unassigned | Unassigned | 330_0 | Clustered | 27 | VC_330_0 | 27 | 0.5088 | 1 | 0.5088 | 0 | 0 | 0 | 0.8661 | Ref |
| 10628 | MN850588 | Unassigned | Unassigned | Unassigned | 705_0 | Clustered | 11 | VC_705_0 | 11 | 0.8861 | 1 | 0.8861 | 0 | 0 | 0 | 1 | Ref |
| 10629 | MN850589 | Unassigned | Unassigned | Unassigned | 835_0 | Clustered | 67 | VC_835_0 | 67 | 0.9624 | 1 | 0.9624 | 0 | 0 | 0 | 1 | Ref |
| 10630 | MN850590 | Unassigned | Unassigned | Unassigned | 286_1 | Clustered | 21 | VC_286_1 | 21 | 0.1696 | 1 | 0.1696 | 0 | 0 | 0 | 0.9048 | Ref |
| 10631 | MN850591 | Unassigned | Unassigned | Unassigned | 941_0 | Clustered | 40 | VC_941_0 | 40 | 0.6903 | 1 | 0.6903 | 0 | 0 | 0 | 1 | Ref |
| 10632 | MN850592 | Unassigned | Unassigned | Unassigned | 156_0 | Clustered | 12 | VC_156_0 | 12 | 0.9062 | 1 | 0.9062 | 0 | 0 | 0 | 1 | Ref |
| 10633 | MN850593 | Unassigned | Unassigned | Unassigned | 772_20 | Clustered | 2 | VC_772_20 | 2 | 0.006 | 0.58868587 | 0.0035 | 0 | 0 | 0 | 1 | Ref |
| 10634 | MN850594 | Unassigned | Unassigned | Unassigned | 792_0 | Clustered | 35 | VC_792_0 | 35 | 0.9828 | 1 | 0.9828 | 0 | 0 | 0 | 1 | Ref |
| 10635 | MN850595 | Unassigned | Unassigned | Unassigned | 772_19 | Clustered | 16 | VC_772_19 | 16 | 0.0975 | 0.9970466 | 0.0972 | 0 | 0 | 0 | 0.8083 | Ref |
| 10636 | MN850596 | Unassigned | Unassigned | Unassigned | 942_0 | Clustered | 36 | VC_942_0 | 36 | 0.7848 | 1 | 0.7848 | 0 | 0 | 0 | 1 | Ref |
| 10637 | MN850597 | Unassigned | Unassigned | Unassigned | 772_19 | Clustered | 16 | VC_772_19 | 16 | 0.0975 | 0.9970466 | 0.0972 | 0 | 0 | 0 | 0.8083 | Ref |
| 10638 | MN850598 | Unassigned | Unassigned | Unassigned | 847_1 | Clustered | 26 | VC_847_1 | 26 | 0.6323 | 1 | 0.6323 | 0 | 0 | 0 | 0.9815 | Ref |
| 10639 | MN850600 | Unassigned | Unassigned | Unassigned | 941_0 | Clustered | 40 | VC_941_0 | 40 | 0.6903 | 1 | 0.6903 | 0 | 0 | 0 | 1 | Ref |
| 10640 | MN850601 | Unassigned | Unassigned | Unassigned | 848_0 | Clustered | 28 | VC_848_0 | 28 | 0.7927 | 1 | 0.7927 | 0 | 0 | 0 | 1 | Ref |
| 10641 | MN850602 | Unassigned | Unassigned | Unassigned | 941_0 | Clustered | 40 | VC_941_0 | 40 | 0.6903 | 1 | 0.6903 | 0 | 0 | 0 | 1 | Ref |
| 10642 | MN850603 | Unassigned | Unassigned | Unassigned | 329_3 | Clustered | 24 | VC_329_3 | 24 | 0.1206 | 1 | 0.1206 | 0 | 0 | 0 | 0.7283 | Ref |
| 10643 | MN850604 | Unassigned | Unassigned | Unassigned | 942_0 | Clustered | 36 | VC_942_0 | 36 | 0.7848 | 1 | 0.7848 | 0 | 0 | 0 | 1 | Ref |
| 10644 | MN850605 | Unassigned | Unassigned | Unassigned | 329_3 | Clustered | 24 | VC_329_3 | 24 | 0.1206 | 1 | 0.1206 | 0 | 0 | 0 | 0.7283 | Ref |
| 10645 | MN850606 | Unassigned | Unassigned | Unassigned | 942_0 | Clustered | 36 | VC_942_0 | 36 | 0.7848 | 1 | 0.7848 | 0 | 0 | 0 | 1 | Ref |
| 10646 | MN850607 | Unassigned | Unassigned | Unassigned | 941_0 | Clustered | 40 | VC_941_0 | 40 | 0.6903 | 1 | 0.6903 | 0 | 0 | 0 | 1 | Ref |
| 10647 | MN850608 | Unassigned | Unassigned | Unassigned | 156_0 | Clustered | 12 | VC_156_0 | 12 | 0.9062 | 1 | 0.9062 | 0 | 0 | 0 | 1 | Ref |
| 10648 | MN850609 | Unassigned | Unassigned | Unassigned | 290_6 | Clustered | 12 | VC_290_6 | 12 | 0.1149 | 1 | 0.1149 | 0 | 0 | 0 | 0.8182 | Ref |
| 10649 | MN850610 | Unassigned | Unassigned | Unassigned | 329_3 | Clustered | 24 | VC_329_3 | 24 | 0.1206 | 1 | 0.1206 | 0 | 0 | 0 | 0.7283 | Ref |
| 10650 | MN850611 | Unassigned | Unassigned | Unassigned | 329_3 | Clustered | 24 | VC_329_3 | 24 | 0.1206 | 1 | 0.1206 | 0 | 0 | 0 | 0.7283 | Ref |
| 10651 | MN850612 | Unassigned | Unassigned | Unassigned | 772_19 | Clustered | 16 | VC_772_19 | 16 | 0.0975 | 0.9970466 | 0.0972 | 0 | 0 | 0 | 0.8083 | Ref |
| 10652 | MN850613 | Unassigned | Unassigned | Unassigned | 942_0 | Clustered | 36 | VC_942_0 | 36 | 0.7848 | 1 | 0.7848 | 0 | 0 | 0 | 1 | Ref |
| 10653 | MN850614 | Unassigned | Unassigned | Unassigned | 329_3 | Clustered | 24 | VC_329_3 | 24 | 0.1206 | 1 | 0.1206 | 0 | 0 | 0 | 0.7283 | Ref |
| 10654 | MN850615 | Unassigned | Unassigned | Unassigned | 590_0 | Clustered | 27 | VC_590_0 | 27 | 0.9947 | 1 | 0.9947 | 0 | 0 | 0 | 1 | Ref |
| 10655 | MN850616 | Unassigned | Unassigned | Unassigned | 637_0 | Clustered | 115 | VC_637_0 | 115 | 0.731 | 1 | 0.731 | 0 | 0 | 0 | 0.8764 | Ref |
| 10656 | MN850617 | Unassigned | Unassigned | Unassigned | 156_0 | Clustered | 12 | VC_156_0 | 12 | 0.9062 | 1 | 0.9062 | 0 | 0 | 0 | 1 | Ref |
| 10657 | MN850618 | Unassigned | Unassigned | Unassigned | 847_1 | Clustered | 26 | VC_847_1 | 26 | 0.6323 | 1 | 0.6323 | 0 | 0 | 0 | 0.9815 | Ref |
| 10658 | MN850619 | Unassigned | Unassigned | Unassigned | 329_3 | Clustered | 24 | VC_329_3 | 24 | 0.1206 | 1 | 0.1206 | 0 | 0 | 0 | 0.7283 | Ref |
| 10659 | MN850620 | Unassigned | Unassigned | Unassigned | 942_0 | Clustered | 36 | VC_942_0 | 36 | 0.7848 | 1 | 0.7848 | 0 | 0 | 0 | 1 | Ref |
| 10660 | MN850621 | Unassigned | Unassigned | Unassigned | 290_6 | Clustered | 12 | VC_290_6 | 12 | 0.1149 | 1 | 0.1149 | 0 | 0 | 0 | 0.8182 | Ref |
| 10661 | MN850622 | Unassigned | Unassigned | Unassigned | 286_17 | Clustered | 8 | VC_286_17 | 8 | 0.0535 | 0.99997829 | 0.0535 | 0 | 0 | 0 | 0.6071 | Ref |
| 10662 | MN850623 | Unassigned | Unassigned | Unassigned | 705_0 | Clustered | 11 | VC_705_0 | 11 | 0.8861 | 1 | 0.8861 | 0 | 0 | 0 | 1 | Ref |
| 10663 | MN850624 | Unassigned | Unassigned | Unassigned | 156_0 | Clustered | 12 | VC_156_0 | 12 | 0.9062 | 1 | 0.9062 | 0 | 0 | 0 | 1 | Ref |
| 10664 | MN850625 | Unassigned | Unassigned | Unassigned | 156_0 | Clustered | 12 | VC_156_0 | 12 | 0.9062 | 1 | 0.9062 | 0 | 0 | 0 | 1 | Ref |
| 10665 | MN850626 | Unassigned | Unassigned | Unassigned | 772_19 | Clustered | 16 | VC_772_19 | 16 | 0.0975 | 0.9970466 | 0.0972 | 0 | 0 | 0 | 0.8083 | Ref |
| 10666 | MN850627 | Unassigned | Unassigned | Unassigned | 772_19 | Clustered | 16 | VC_772_19 | 16 | 0.0975 | 0.9970466 | 0.0972 | 0 | 0 | 0 | 0.8083 | Ref |
| 10667 | MN850628 | Unassigned | Unassigned | Unassigned | 835_0 | Clustered | 67 | VC_835_0 | 67 | 0.9624 | 1 | 0.9624 | 0 | 0 | 0 | 1 | Ref |
| 10668 | MN850629 | Unassigned | Unassigned | Unassigned | 835_0 | Clustered | 67 | VC_835_0 | 67 | 0.9624 | 1 | 0.9624 | 0 | 0 | 0 | 1 | Ref |
| 10669 | MN850630 | Unassigned | Unassigned | Unassigned | 772_10 | Clustered | 8 | VC_772_10 | 8 | 0.0433 | 0.9088183 | 0.0394 | 0 | 0 | 0 | 0.9286 | Ref |
| 10670 | MN850631 | Unassigned | Unassigned | Unassigned | 329_2 | Clustered | 2 | VC_329_2 | 2 | 0.0049 | 0.95560085 | 0.0047 | 0 | 0 | 0 | 1 | Ref |
| 10671 | MN850632 | Unassigned | Unassigned | Unassigned | 848_0 | Clustered | 28 | VC_848_0 | 28 | 0.7927 | 1 | 0.7927 | 0 | 0 | 0 | 1 | Ref |
| 10672 | MN850633 | Unassigned | Unassigned | Unassigned | 329_3 | Clustered | 24 | VC_329_3 | 24 | 0.1206 | 1 | 0.1206 | 0 | 0 | 0 | 0.7283 | Ref |
| 10673 | MN850634 | Unassigned | Unassigned | Unassigned | 942_0 | Clustered | 36 | VC_942_0 | 36 | 0.7848 | 1 | 0.7848 | 0 | 0 | 0 | 1 | Ref |
| 10674 | MN850635 | Unassigned | Unassigned | Unassigned | 329_3 | Clustered | 24 | VC_329_3 | 24 | 0.1206 | 1 | 0.1206 | 0 | 0 | 0 | 0.7283 | Ref |
| 10675 | MN850636 | Unassigned | Unassigned | Unassigned | 329_3 | Clustered | 24 | VC_329_3 | 24 | 0.1206 | 1 | 0.1206 | 0 | 0 | 0 | 0.7283 | Ref |
| 10676 | MN850637 | Unassigned | Unassigned | Unassigned | 329_2 | Clustered | 2 | VC_329_2 | 2 | 0.0049 | 0.95560085 | 0.0047 | 0 | 0 | 0 | 1 | Ref |
| 10677 | MN850638 | Unassigned | Unassigned | Unassigned | 942_0 | Clustered | 36 | VC_942_0 | 36 | 0.7848 | 1 | 0.7848 | 0 | 0 | 0 | 1 | Ref |
| 10678 | MN850639 | Unassigned | Unassigned | Unassigned | 329_3 | Clustered | 24 | VC_329_3 | 24 | 0.1206 | 1 | 0.1206 | 0 | 0 | 0 | 0.7283 | Ref |
| 10679 | MN850640 | Unassigned | Unassigned | Unassigned | 941_0 | Clustered | 40 | VC_941_0 | 40 | 0.6903 | 1 | 0.6903 | 0 | 0 | 0 | 1 | Ref |
| 10680 | MN850641 | Unassigned | Unassigned | Unassigned | 942_0 | Clustered | 36 | VC_942_0 | 36 | 0.7848 | 1 | 0.7848 | 0 | 0 | 0 | 1 | Ref |
| 10681 | MN850642 | Unassigned | Unassigned | Unassigned | 772_16 | Clustered | 10 | VC_772_16 | 10 | 0.0563 | 0.9434928 | 0.0531 | 0 | 0 | 0 | 0.8667 | Ref |
| 10682 | MN850643 | Unassigned | Unassigned | Unassigned | 942_0 | Clustered | 36 | VC_942_0 | 36 | 0.7848 | 1 | 0.7848 | 0 | 0 | 0 | 1 | Ref |
| 10683 | MN850644 | Unassigned | Unassigned | Unassigned | 329_3 | Clustered | 24 | VC_329_3 | 24 | 0.1206 | 1 | 0.1206 | 0 | 0 | 0 | 0.7283 | Ref |
| 10684 | MN850645 | Unassigned | Unassigned | Unassigned | 848_0 | Clustered | 28 | VC_848_0 | 28 | 0.7927 | 1 | 0.7927 | 0 | 0 | 0 | 1 | Ref |
| 10685 | MN850646 | Unassigned | Unassigned | Unassigned | 772_6 | Clustered | 3 | VC_772_6 | 3 | 0.012 | 0.63979507 | 0.0077 | 0 | 0 | 0 | 1 | Ref |
| 10686 | MN850647 | Unassigned | Unassigned | Unassigned | 329_3 | Clustered | 24 | VC_329_3 | 24 | 0.1206 | 1 | 0.1206 | 0 | 0 | 0 | 0.7283 | Ref |
| 10687 | MN850648 | Unassigned | Unassigned | Unassigned | 847_1 | Clustered | 26 | VC_847_1 | 26 | 0.6323 | 1 | 0.6323 | 0 | 0 | 0 | 0.9815 | Ref |
| 10688 | MN850649 | Unassigned | Unassigned | Unassigned | 772_6 | Clustered | 3 | VC_772_6 | 3 | 0.012 | 0.63979507 | 0.0077 | 0 | 0 | 0 | 1 | Ref |
| 10689 | MN850650 | Unassigned | Unassigned | Unassigned | 835_0 | Clustered | 67 | VC_835_0 | 67 | 0.9624 | 1 | 0.9624 | 0 | 0 | 0 | 1 | Ref |
| 10690 | MN850651 | Unassigned | Unassigned | Unassigned | 286_2 | Clustered/Singleton | 1 | VC_286_2 | 1 | 0 | 1 | 0 | 0 | 0 | 0 | 1 | Ref |

| | | | | | | | | | | | | | | | | | |
|---|---|---|---|---|---|---|---|---|---|---|---|---|---|---|---|---|---|
| 10691 | MN854830 | Unassigned | Unassigned | Unassigned | 91_0 | Clustered | 22 | VC_91_0 | 22 | 0.5318 | 1 | 0.5318 | 0 | 0 | 0 | 1 | Ref |
| 10692 | MN862068 | Unassigned | Unassigned | Unassigned | 467_0 | Clustered | 5 | VC_467_0 | 5 | 0.6874 | 0.99999996 | 0.6874 | 0 | 0 | 0 | 1 | Ref |
| 10693 | MN862333 | Unassigned | Unassigned | Unassigned | 995_0 | Clustered | 69 | VC_995_0 | 69 | 0.8547 | 1 | 0.8547 | 0 | 0 | 0 | 1 | Ref |
| 10694 | MN862335 | Unassigned | Unassigned | Unassigned | 995_0 | Clustered | 69 | VC_995_0 | 69 | 0.8547 | 1 | 0.8547 | 0 | 0 | 0 | 1 | Ref |
| 10695 | MN862338 | Unassigned | Unassigned | Unassigned | 1066_0 | Clustered | 52 | VC_1066_0 | 52 | 0.7682 | 1 | 0.7682 | 0 | 0 | 0 | 1 | Ref |
| 10696 | MN862339 | Unassigned | Unassigned | Unassigned | 996_0 | Clustered | 2 | VC_996_0 | 2 | 0.0422 | 0.59038153 | 0.0249 | 0 | 0 | 0 | 1 | Ref |
| 10697 | MN862340 | Unassigned | Unassigned | Unassigned | 996_0 | Clustered | 2 | VC_996_0 | 2 | 0.0422 | 0.59038153 | 0.0249 | 0 | 0 | 0 | 1 | Ref |
| 10698 | MN862341 | Unassigned | Unassigned | Unassigned | 995_0 | Clustered | 69 | VC_995_0 | 69 | 0.8547 | 1 | 0.8547 | 0 | 0 | 0 | 1 | Ref |
| 10699 | MN862343 | Unassigned | Unassigned | Unassigned | 198_0 | Clustered | 28 | VC_198_0 | 28 | 0.2429 | 0.99971175 | 0.2429 | 0 | 0 | 0 | 1 | Ref |
| 10700 | MN862345 | Unassigned | Unassigned | Unassigned | 197_0 | Clustered | 5 | VC_197_0 | 5 | 0.1324 | 0.84698401 | 0.1122 | 0 | 0 | 0 | 1 | Ref |
| 10701 | MN862346 | Unassigned | Unassigned | Unassigned | 1077_0 | Clustered | 48 | VC_1077_0 | 48 | 0.8525 | 1 | 0.8525 | 0 | 0 | 0 | 1 | Ref |
| 10702 | MN864145 | Unassigned | Unassigned | Unassigned | 286_6 | Clustered/Singleton | 1 | VC_286_6 | 1 | 0 | 1 | 0 | 0 | 0 | 0 | 1 | Ref |
| 10703 | MN864865 | Unassigned | Unassigned | Unassigned | 492_2 | Clustered | 8 | VC_492_2 | 8 | 0.1221 | 0.98786574 | 0.1206 | 0 | 0 | 0 | 1 | Ref |
| 10704 | MN871441 | Unassigned | Unassigned | Unassigned | 443_2 | Clustered | 10 | VC_443_2 | 10 | 0.3344 | 1 | 0.3344 | 0 | 0 | 0 | 0.9333 | Ref |
| 10705 | MN871442 | Unassigned | Unassigned | Unassigned | 443_2 | Clustered | 10 | VC_443_2 | 10 | 0.3344 | 1 | 0.3344 | 0 | 0 | 0 | 0.9333 | Ref |
| 10706 | MN871443 | Unassigned | Unassigned | Unassigned | 108_0 | Clustered | 33 | VC_108_0 | 33 | 0.949 | 1 | 0.949 | 0 | 0 | 0 | 1 | Ref |
| 10707 | MN871444 | Unassigned | Unassigned | Unassigned | 347_0 | Clustered | 3 | VC_347_0 | 3 | 0.1545 | 0.9984446 | 0.1543 | 0 | 0 | 0 | 1 | Ref |
| 10708 | MN871445 | Unassigned | Unassigned | Unassigned | 347_0 | Clustered | 3 | VC_347_0 | 3 | 0.1545 | 0.9984446 | 0.1543 | 0 | 0 | 0 | 1 | Ref |
| 10709 | MN871446 | Unassigned | Unassigned | Unassigned | 488_0 | Clustered | 8 | VC_488_0 | 8 | 0.2938 | 1 | 0.2938 | 0 | 0 | 0 | 1 | Ref |
| 10710 | MN871447 | Unassigned | Unassigned | Unassigned | 488_0 | Clustered | 8 | VC_488_0 | 8 | 0.2938 | 1 | 0.2938 | 0 | 0 | 0 | 1 | Ref |
| 10711 | MN871448 | Unassigned | Unassigned | Unassigned | 488_0 | Clustered | 8 | VC_488_0 | 8 | 0.2938 | 1 | 0.2938 | 0 | 0 | 0 | 1 | Ref |
| 10712 | MN871449 | Unassigned | Unassigned | Unassigned | 488_0 | Clustered | 8 | VC_488_0 | 8 | 0.2938 | 1 | 0.2938 | 0 | 0 | 0 | 1 | Ref |
| 10713 | MN871450 | Unassigned | Unassigned | Unassigned | 592_0 | Clustered | 50 | VC_592_0 | 50 | 0.7314 | 1 | 0.7314 | 0 | 0 | 0 | 1 | Ref |
| 10714 | MN871451 | Unassigned | Unassigned | Unassigned | 488_0 | Clustered | 8 | VC_488_0 | 8 | 0.2938 | 1 | 0.2938 | 0 | 0 | 0 | 1 | Ref |
| 10715 | MN871452 | Unassigned | Unassigned | Unassigned | 488_0 | Clustered | 8 | VC_488_0 | 8 | 0.2938 | 1 | 0.2938 | 0 | 0 | 0 | 1 | Ref |
| 10716 | MN871453 | Unassigned | Unassigned | Unassigned | 488_0 | Clustered | 8 | VC_488_0 | 8 | 0.2938 | 1 | 0.2938 | 0 | 0 | 0 | 1 | Ref |
| 10717 | MN871454 | Unassigned | Unassigned | Unassigned | 80_0 | Clustered | 41 | VC_80_0 | 41 | 0.5837 | 1 | 0.5837 | 0 | 0 | 0 | 0.9805 | Ref |
| 10718 | MN871455 | Unassigned | Unassigned | Unassigned | 423_0 | Clustered | 45 | VC_423_0 | 45 | 0.9484 | 1 | 0.9484 | 0 | 0 | 0 | 1 | Ref |
| 10719 | MN871456 | Unassigned | Unassigned | Unassigned | 166_0 | Clustered | 26 | VC_166_0 | 26 | 0.7923 | 1 | 0.7923 | 0 | 0 | 0 | 1 | Ref |
| 10720 | MN871457 | Unassigned | Unassigned | Unassigned | 79_6 | Clustered | 11 | VC_79_6 | 11 | 0.0528 | 1 | 0.0528 | 0 | 0 | 0 | 0.7455 | Ref |
| 10721 | MN871458 | Unassigned | Unassigned | Unassigned | 79_0 | Clustered | 64 | VC_79_0 | 64 | 0.4185 | 1 | 0.4185 | 0 | 0 | 0 | 0.7649 | Ref |
| 10722 | MN871459 | Unassigned | Unassigned | Unassigned | 79_0 | Clustered | 64 | VC_79_0 | 64 | 0.4185 | 1 | 0.4185 | 0 | 0 | 0 | 0.7649 | Ref |
| 10723 | MN871460 | Unassigned | Unassigned | Unassigned | 79_6 | Clustered | 11 | VC_79_6 | 11 | 0.0528 | 1 | 0.0528 | 0 | 0 | 0 | 0.7455 | Ref |
| 10724 | MN871461 | Unassigned | Unassigned | Unassigned | 79_0 | Clustered | 64 | VC_79_0 | 64 | 0.4185 | 1 | 0.4185 | 0 | 0 | 0 | 0.7649 | Ref |
| 10725 | MN871462 | Unassigned | Unassigned | Unassigned | 79_0 | Clustered | 64 | VC_79_0 | 64 | 0.4185 | 1 | 0.4185 | 0 | 0 | 0 | 0.7649 | Ref |
| 10726 | MN871463 | Unassigned | Unassigned | Unassigned | 166_0 | Clustered | 26 | VC_166_0 | 26 | 0.7923 | 1 | 0.7923 | 0 | 0 | 0 | 1 | Ref |
| 10727 | MN871464 | Unassigned | Unassigned | Unassigned | 79_0 | Clustered | 64 | VC_79_0 | 64 | 0.4185 | 1 | 0.4185 | 0 | 0 | 0 | 0.7649 | Ref |
| 10728 | MN871465 | Unassigned | Unassigned | Unassigned | 79_0 | Clustered | 64 | VC_79_0 | 64 | 0.4185 | 1 | 0.4185 | 0 | 0 | 0 | 0.7649 | Ref |
| 10729 | MN871466 | Unassigned | Unassigned | Unassigned | 79_0 | Clustered | 64 | VC_79_0 | 64 | 0.4185 | 1 | 0.4185 | 0 | 0 | 0 | 0.7649 | Ref |
| 10730 | MN871467 | Unassigned | Unassigned | Unassigned | 79_6 | Clustered | 11 | VC_79_6 | 11 | 0.0528 | 1 | 0.0528 | 0 | 0 | 0 | 0.7455 | Ref |
| 10731 | MN871468 | Unassigned | Unassigned | Unassigned | 79_9 | Clustered | 2 | VC_79_9 | 2 | 0.0061 | 0.94860001 | 0.0058 | 0 | 0 | 0 | 1 | Ref |
| 10732 | MN871469 | Unassigned | Unassigned | Unassigned | 166_0 | Clustered | 26 | VC_166_0 | 26 | 0.7923 | 1 | 0.7923 | 0 | 0 | 0 | 1 | Ref |
| 10733 | MN871470 | Unassigned | Unassigned | Unassigned | 76_0 | Clustered | 37 | VC_76_0 | 37 | 0.9403 | 1 | 0.9403 | 0 | 0 | 0 | 1 | Ref |
| 10734 | MN871471 | Unassigned | Unassigned | Unassigned | 796_0 | Clustered | 5 | VC_796_0 | 5 | 0.3352 | 0.99999997 | 0.3352 | 0 | 0 | 0 | 1 | Ref |
| 10735 | MN871472 | Unassigned | Unassigned | Unassigned | 796_0 | Clustered | 5 | VC_796_0 | 5 | 0.3352 | 0.99999997 | 0.3352 | 0 | 0 | 0 | 1 | Ref |
| 10736 | MN871473 | Unassigned | Unassigned | Unassigned | 79_0 | Clustered | 64 | VC_79_0 | 64 | 0.4185 | 1 | 0.4185 | 0 | 0 | 0 | 0.7649 | Ref |
| 10737 | MN871474 | Unassigned | Unassigned | Unassigned | 796_0 | Clustered | 5 | VC_796_0 | 5 | 0.3352 | 0.99999997 | 0.3352 | 0 | 0 | 0 | 1 | Ref |
| 10738 | MN871475 | Unassigned | Unassigned | Unassigned | 133_29 | Clustered | 2 | VC_133_29 | 2 | 0.0052 | 0.8586516 | 0.0044 | 0 | 0 | 0 | 1 | Ref |
| 10739 | MN871476 | Unassigned | Unassigned | Unassigned | 472_0 | Clustered | 47 | VC_472_0 | 47 | 0.9025 | 1 | 0.9025 | 0 | 0 | 0 | 1 | Ref |
| 10740 | MN871478 | Unassigned | Unassigned | Unassigned | 166_0 | Clustered | 26 | VC_166_0 | 26 | 0.7923 | 1 | 0.7923 | 0 | 0 | 0 | 1 | Ref |
| 10741 | MN871479 | Unassigned | Unassigned | Unassigned | 79_0 | Clustered | 64 | VC_79_0 | 64 | 0.4185 | 1 | 0.4185 | 0 | 0 | 0 | 0.7649 | Ref |
| 10742 | MN871480 | Unassigned | Unassigned | Unassigned | 344_0 | Clustered | 21 | VC_344_0 | 21 | 0.5436 | 1 | 0.5436 | 0 | 0 | 0 | 1 | Ref |
| 10743 | MN871481 | Unassigned | Unassigned | Unassigned | 76_0 | Clustered | 37 | VC_76_0 | 37 | 0.9403 | 1 | 0.9403 | 0 | 0 | 0 | 1 | Ref |
| 10744 | MN871482 | Unassigned | Unassigned | Unassigned | 166_0 | Clustered | 26 | VC_166_0 | 26 | 0.7923 | 1 | 0.7923 | 0 | 0 | 0 | 1 | Ref |
| 10745 | MN871483 | Unassigned | Unassigned | Unassigned | 423_0 | Clustered | 45 | VC_423_0 | 45 | 0.9484 | 1 | 0.9484 | 0 | 0 | 0 | 1 | Ref |
| 10746 | MN871484 | Unassigned | Unassigned | Unassigned | 472_0 | Clustered | 47 | VC_472_0 | 47 | 0.9025 | 1 | 0.9025 | 0 | 0 | 0 | 1 | Ref |
| 10747 | MN871485 | Unassigned | Unassigned | Unassigned | 79_0 | Clustered | 64 | VC_79_0 | 64 | 0.4185 | 1 | 0.4185 | 0 | 0 | 0 | 0.7649 | Ref |
| 10748 | MN871486 | Unassigned | Unassigned | Unassigned | 166_0 | Clustered | 26 | VC_166_0 | 26 | 0.7923 | 1 | 0.7923 | 0 | 0 | 0 | 1 | Ref |
| 10749 | MN871487 | Unassigned | Unassigned | Unassigned | 79_0 | Clustered | 64 | VC_79_0 | 64 | 0.4185 | 1 | 0.4185 | 0 | 0 | 0 | 0.7649 | Ref |
| 10750 | MN871488 | Unassigned | Unassigned | Unassigned | 76_0 | Clustered | 37 | VC_76_0 | 37 | 0.9403 | 1 | 0.9403 | 0 | 0 | 0 | 1 | Ref |
| 10751 | MN871489 | Unassigned | Unassigned | Unassigned | 344_0 | Clustered | 21 | VC_344_0 | 21 | 0.5436 | 1 | 0.5436 | 0 | 0 | 0 | 1 | Ref |
| 10752 | MN871490 | Unassigned | Unassigned | Unassigned | 461_0 | Clustered | 96 | VC_461_0 | 96 | 0.9307 | 1 | 0.9307 | 0 | 0 | 0 | 0.9575 | Ref |
| 10753 | MN871491 | Unassigned | Unassigned | Unassigned | 133_36 | Clustered | 2 | VC_133_36 | 2 | 0.0052 | 0.86421148 | 0.0045 | 0 | 0 | 0 | 1 | Ref |
| 10754 | MN871492 | Unassigned | Unassigned | Unassigned | 133_36 | Clustered | 2 | VC_133_36 | 2 | 0.0052 | 0.86421148 | 0.0045 | 0 | 0 | 0 | 1 | Ref |
| 10755 | MN871493 | Unassigned | Unassigned | Unassigned | 133_37 | Clustered/Singleton | 1 | VC_133_37 | 1 | 0 | 1 | 0 | 0 | 0 | 0 | 1 | Ref |
| 10756 | MN871494 | Unassigned | Unassigned | Unassigned | 133_12 | Clustered | 2 | VC_133_12 | 2 | 0.0051 | 0.8672083 | 0.0044 | 0 | 0 | 0 | 1 | Ref |
| 10757 | MN871495 | Unassigned | Unassigned | Unassigned | 133_12 | Clustered | 2 | VC_133_12 | 2 | 0.0051 | 0.8672083 | 0.0044 | 0 | 0 | 0 | 1 | Ref |
| 10758 | MN871496 | Unassigned | Unassigned | Unassigned | 133_43 | Clustered | 4 | VC_133_43 | 4 | 0.015 | 0.99995682 | 0.015 | 0 | 0 | 0 | 1 | Ref |
| 10759 | MN871497 | Unassigned | Unassigned | Unassigned | 133_43 | Clustered | 4 | VC_133_43 | 4 | 0.015 | 0.99995682 | 0.015 | 0 | 0 | 0 | 1 | Ref |
| 10760 | MN871498 | Unassigned | Unassigned | Unassigned | 133_43 | Clustered | 4 | VC_133_43 | 4 | 0.015 | 0.99995682 | 0.015 | 0 | 0 | 0 | 1 | Ref |
| 10761 | MN871499 | Unassigned | Unassigned | Unassigned | 133_43 | Clustered | 4 | VC_133_43 | 4 | 0.015 | 0.99995682 | 0.015 | 0 | 0 | 0 | 1 | Ref |
| 10762 | MN871500 | Unassigned | Unassigned | Unassigned | 133_19 | Clustered | 2 | VC_133_19 | 2 | 0.0051 | 0.8696643 | 0.0044 | 0 | 0 | 0 | 1 | Ref |
| 10763 | MN871501 | Unassigned | Unassigned | Unassigned | 133_29 | Clustered | 2 | VC_133_29 | 2 | 0.0052 | 0.8586516 | 0.0044 | 0 | 0 | 0 | 1 | Ref |
| 10764 | MN871502 | Unassigned | Unassigned | Unassigned | 133_38 | Clustered/Singleton | 1 | VC_133_38 | 1 | 0 | 1 | 0 | 0 | 0 | 0 | 1 | Ref |
| 10765 | MN871503 | Unassigned | Unassigned | Unassigned | 461_0 | Clustered | 96 | VC_461_0 | 96 | 0.9307 | 1 | 0.9307 | 0 | 0 | 0 | 0.9575 | Ref |
| 10766 | MN871504 | Unassigned | Unassigned | Unassigned | 133_10 | Clustered | 10 | VC_133_10 | 10 | 0.0527 | 1 | 0.0527 | 0 | 0 | 0 | 1 | Ref |
| 10767 | MN871505 | Unassigned | Unassigned | Unassigned | 972_0 | Clustered | 6 | VC_972_0 | 6 | 0.2892 | 1 | 0.2892 | 0 | 0 | 0 | 1 | Ref |
| 10768 | MN871506 | Unassigned | Unassigned | Unassigned | 904_2 | Clustered | 7 | VC_904_2 | 7 | 0.2857 | 1 | 0.2857 | 0 | 0 | 0 | 1 | Ref |
| 10769 | MN871507 | Unassigned | Unassigned | Unassigned | 600_0 | Clustered | 3 | VC_600_0 | 3 | 0.0541 | 0.97110069 | 0.0526 | 0 | 0 | 0 | 1 | Ref |
| 10770 | MN871508 | Unassigned | Unassigned | Unassigned | 79_0 | Clustered | 64 | VC_79_0 | 64 | 0.4185 | 1 | 0.4185 | 0 | 0 | 0 | 0.7649 | Ref |
| 10771 | MN871509 | Unassigned | Unassigned | Unassigned | 79_0 | Clustered | 64 | VC_79_0 | 64 | 0.4185 | 1 | 0.4185 | 0 | 0 | 0 | 0.7649 | Ref |
| 10772 | MN871510 | Unassigned | Unassigned | Unassigned | 972_0 | Clustered | 6 | VC_972_0 | 6 | 0.2892 | 1 | 0.2892 | 0 | 0 | 0 | 1 | Ref |
| 10773 | MN871511 | Unassigned | Unassigned | Unassigned | 972_0 | Clustered | 6 | VC_972_0 | 6 | 0.2892 | 1 | 0.2892 | 0 | 0 | 0 | 1 | Ref |
| 10774 | MN876841 | Unassigned | Unassigned | Unassigned | 348_0 | Clustered | 22 | VC_348_0 | 22 | 0.9602 | 1 | 0.9602 | 0 | 0 | 0 | 1 | Ref |
| 10775 | MN876842 | Unassigned | Unassigned | Unassigned | 348_0 | Clustered | 22 | VC_348_0 | 22 | 0.9602 | 1 | 0.9602 | 0 | 0 | 0 | 1 | Ref |
| 10776 | MN876843 | Unassigned | Unassigned | Unassigned | 348_0 | Clustered | 22 | VC_348_0 | 22 | 0.9602 | 1 | 0.9602 | 0 | 0 | 0 | 1 | Ref |
| 10777 | MN876845 | Unassigned | Unassigned | Unassigned | 440_0 | Clustered | 4 | VC_440_0 | 4 | 1 | 1 | 1 | 0 | 0 | 0 | 1 | Ref |
| 10778 | MN882349 | Unassigned | Unassigned | Unassigned | 590_0 | Clustered | 27 | VC_590_0 | 27 | 0.9947 | 1 | 0.9947 | 0 | 0 | 0 | 1 | Ref |
| 10779 | MN882542 | Unassigned | Unassigned | Unassigned | 329_18 | Clustered | 7 | VC_329_18 | 7 | 0.0328 | 1 | 0.0328 | 0 | 0 | 0 | 0.8095 | Ref |
| 10780 | MN882551 | Unassigned | Unassigned | Unassigned | 1413_0 | Clustered | 2 | VC_1413_0 | 2 | 0.6796 | 0.89435023 | 0.6078 | 0 | 0 | 0 | 1 | Ref |
| 10781 | MN882552 | Unassigned | Unassigned | Unassigned | 1413_0 | Clustered | 2 | VC_1413_0 | 2 | 0.6796 | 0.89435023 | 0.6078 | 0 | 0 | 0 | 1 | Ref |
| 10782 | MN882610 | Unassigned | Unassigned | Unassigned | 490_1 | Clustered | 5 | VC_490_1 | 5 | 0.0447 | 0.99999996 | 0.0447 | 0 | 0 | 0 | 1 | Ref |
| 10783 | MN890136 | Unassigned | Unassigned | Unassigned | 721_0 | Clustered | 9 | VC_721_0 | 9 | 1 | 1 | 1 | 0 | 0 | 0 | 1 | Ref |
| 10784 | MN890137 | Unassigned | Unassigned | Unassigned | 721_0 | Clustered | 9 | VC_721_0 | 9 | 1 | 1 | 1 | 0 | 0 | 0 | 1 | Ref |
| 10785 | MN890138 | Unassigned | Unassigned | Unassigned | 721_0 | Clustered | 9 | VC_721_0 | 9 | 1 | 1 | 1 | 0 | 0 | 0 | 1 | Ref |
| 10786 | MN890139 | Unassigned | Unassigned | Unassigned | 721_0 | Clustered | 9 | VC_721_0 | 9 | 1 | 1 | 1 | 0 | 0 | 0 | 1 | Ref |
| 10787 | MN890140 | Unassigned | Unassigned | Unassigned | 721_0 | Clustered | 9 | VC_721_0 | 9 | 1 | 1 | 1 | 0 | 0 | 0 | 1 | Ref |
| 10788 | MN890141 | Unassigned | Unassigned | Unassigned | 721_0 | Clustered | 9 | VC_721_0 | 9 | 1 | 1 | 1 | 0 | 0 | 0 | 1 | Ref |
| 10789 | MN890142 | Unassigned | Unassigned | Unassigned | 721_0 | Clustered | 9 | VC_721_0 | 9 | 1 | 1 | 1 | 0 | 0 | 0 | 1 | Ref |
| 10790 | MN890143 | Unassigned | Unassigned | Unassigned | 721_0 | Clustered | 9 | VC_721_0 | 9 | 1 | 1 | 1 | 0 | 0 | 0 | 1 | Ref |
| 10791 | MN892483 | Unassigned | Unassigned | Unassigned | 1240_0 | Clustered | 82 | VC_1240_0 | 82 | 0.7945 | 1 | 0.7945 | 0 | 0 | 0 | 1 | Ref |
| 10792 | MN892484 | Unassigned | Unassigned | Unassigned | 519_0 | Clustered | 39 | VC_519_0 | 39 | 0.8156 | 1 | 0.8156 | 0 | 0 | 0 | 1 | Ref |
| 10793 | MN892486 | Unassigned | Unassigned | Unassigned | 519_0 | Clustered | 39 | VC_519_0 | 39 | 0.8156 | 1 | 0.8156 | 0 | 0 | 0 | 1 | Ref |
| 10794 | MN892487 | Unassigned | Unassigned | Unassigned | 221_0 | Clustered | 22 | VC_221_0 | 22 | 0.7447 | 1 | 0.7447 | 0 | 0 | 0 | 1 | Ref |
| 10795 | MN894885 | Unassigned | Unassigned | Unassigned | 590_0 | Clustered | 27 | VC_590_0 | 27 | 0.9947 | 1 | 0.9947 | 0 | 0 | 0 | 1 | Ref |
| 10796 | MN895434 | Unassigned | Unassigned | Unassigned | 283_39 | Clustered/Singleton | 1 | VC_283_39 | 1 | 0 | 1 | 0 | 0 | 0 | 0 | 1 | Ref |
| 10797 | MN895435 | Unassigned | Unassigned | Unassigned | 283_51 | Clustered/Singleton | 1 | VC_283_51 | 1 | 0 | 1 | 0 | 0 | 0 | 0 | 1 | Ref |
| 10798 | MN895436 | Unassigned | Unassigned | Unassigned | 283_48 | Clustered/Singleton | 1 | VC_283_48 | 1 | 0 | 1 | 0 | 0 | 0 | 0 | 1 | Ref |
| 10799 | MN895437 | Unassigned | Unassigned | Unassigned | 283_31 | Clustered/Singleton | 1 | VC_283_31 | 1 | 0 | 1 | 0 | 0 | 0 | 0 | 1 | Ref |
| 10800 | MN895438 | Unassigned | Unassigned | Unassigned | 283_44 | Clustered/Singleton | 1 | VC_283_44 | 1 | 0 | 1 | 0 | 0 | 0 | 0 | 1 | Ref |
| 10801 | MN901520 | Unassigned | Unassigned | Unassigned | 310_0 | Clustered | 33 | VC_310_0 | 33 | 0.8771 | 1 | 0.8771 | 0 | 0 | 0 | 1 | Ref |
| 10802 | MN901521 | Unassigned | Unassigned | Unassigned | 310_0 | Clustered | 33 | VC_310_0 | 33 | 0.8771 | 1 | 0.8771 | 0 | 0 | 0 | 1 | Ref |
| 10803 | MN901924 | Unassigned | Unassigned | Unassigned | 423_0 | Clustered | 45 | VC_423_0 | 45 | 0.9484 | 1 | 0.9484 | 0 | 0 | 0 | 1 | Ref |
| 10804 | MN904502 | Unassigned | Unassigned | Unassigned | 980_0 | Clustered | 10 | VC_980_0 | 10 | 0.9996 | 1 | 0.9996 | 0 | 0 | 0 | 1 | Ref |
| 10805 | MN904510 | Unassigned | Unassigned | Unassigned | 12_2 | Clustered | 19 | VC_12_2 | 19 | 0.2018 | 1 | 0.2018 | 0 | 0 | 0 | 1 | Ref |
| 10806 | MN905160 | Unassigned | Unassigned | Unassigned | 78_3 | Clustered | 11 | VC_78_3 | 11 | 0.2572 | 1 | 0.2572 | 0 | 0 | 0 | 1 | Ref |
| 10807 | MN906996 | Unassigned | Unassigned | Unassigned | 838_0 | Clustered | 29 | VC_838_0 | 29 | 0.8611 | 1 | 0.8611 | 0 | 0 | 0 | 1 | Ref |
| 10808 | MN908683 | Unassigned | Unassigned | Unassigned | 519_0 | Clustered | 39 | VC_519_0 | 39 | 0.8156 | 1 | 0.8156 | 0 | 0 | 0 | 1 | Ref |
| 10809 | MN908685 | Unassigned | Unassigned | Unassigned | 1100_0 | Clustered | 2 | VC_1100_0 | 2 | 0.1285 | 0.94673154 | 0.1217 | 0 | 0 | 0 | 1 | Ref |
| 10810 | MN908689 | Unassigned | Unassigned | Unassigned | 1207_0 | Clustered | 29 | VC_1207_0 | 29 | 0.8225 | 1 | 0.8225 | 0 | 0 | 0 | 1 | Ref |
| 10811 | MN908690 | Unassigned | Unassigned | Unassigned | 856_1 | Clustered | 13 | VC_856_1 | 13 | 0.5848 | 1 | 0.5848 | 0 | 0 | 0 | 1 | Ref |
| 10812 | MN908691 | Unassigned | Unassigned | Unassigned | 1182_0 | Clustered | 4 | VC_1182_0 | 4 | 0.2845 | 0.99997676 | 0.2845 | 0 | 0 | 0 | 1 | Ref |
| 10813 | MN908692 | Unassigned | Unassigned | Unassigned | 236_0 | Clustered | 72 | VC_236_0 | 72 | 0.8104 | 1 | 0.8104 | 0 | 0 | 0 | 1 | Ref |
| 10814 | MN908693 | Unassigned | Unassigned | Unassigned | 510_0 | Clustered | 12 | VC_510_0 | 12 | 0.3963 | 1 | 0.3963 | 0 | 0 | 0 | 0.9697 | Ref |
| 10815 | MN927225 | Unassigned | Unassigned | Unassigned | 638_0 | Clustered | 45 | VC_638_0 | 45 | 0.7741 | 1 | 0.7741 | 0 | 0 | 0 | 1 | Ref |
| 10816 | MN927226 | Unassigned | Unassigned | Unassigned | 638_0 | Clustered | 45 | VC_638_0 | 45 | 0.7741 | 1 | 0.7741 | 0 | 0 | 0 | 1 | Ref |
| 10817 | MN929097 | Unassigned | Unassigned | Unassigned | 831_0 | Clustered | 5 | VC_831_0 | 5 | 0.4369 | 0.99999952 | 0.4369 | 0 | 0 | 0 | 1 | Ref |
| 10818 | MN935200 | Unassigned | Unassigned | Unassigned | 149_28 | Clustered/Singleton | 1 | VC_149_28 | 1 | 0 | 1 | 0 | 0 | 0 | 0 | 1 | Ref |
| 10819 | MN937349 | Unassigned | Unassigned | Unassigned | 671_0 | Clustered | 5 | VC_671_0 | 5 | 0.5289 | 0.99999996 | 0.5289 | 0 | 0 | 0 | 1 | Ref |
| 10820 | MN939408 | Unassigned | Unassigned | Unassigned | 108_0 | Clustered | 33 | VC_108_0 | 33 | 0.949 | 1 | 0.949 | 0 | 0 | 0 | 1 | Ref |
| 10821 | MN939539 | Unassigned | Unassigned | Unassigned | 739_0 | Clustered | 26 | VC_739_0 | 26 | 0.9571 | 1 | 0.9571 | 0 | 0 | 0 | 1 | Ref |
| 10822 | MN939540 | Unassigned | Unassigned | Unassigned | 739_0 | Clustered | 26 | VC_739_0 | 26 | 0.9571 | 1 | 0.9571 | 0 | 0 | 0 | 1 | Ref |
| 10823 | MN940411 | Unassigned | Unassigned | Unassigned | 85_11 | Clustered/Singleton | 1 | VC_85_11 | 1 | 0 | 1 | 0 | 0 | 0 | 0 | 1 | Ref |
| 10824 | MN945899 | Unassigned | Unassigned | Unassigned | 371_16 | Clustered | 42 | VC_371_16 | 42 | 0.1341 | 1 | 0.1341 | 0 | 0 | 0 | 0.7282 | Ref |
| 10825 | MN945901 | Unassigned | Unassigned | Unassigned | 237_2 | Clustered | 11 | VC_237_2 | 11 | 0.1409 | 1 | 0.1409 | 0 | 0 | 0 | 0.9818 | Ref |
| 10826 | MN945902 | Unassigned | Unassigned | Unassigned | 956_0 | Clustered | 39 | VC_956_0 | 39 | 1 | 1 | 1 | 0 | 0 | 0 | 1 | Ref |
| 10827 | MN945903 | Unassigned | Unassigned | Unassigned | 371_13 | Clustered | 33 | VC_371_13 | 33 | 0.179 | 1 | 0.179 | 0 | 0 | 0 | 0.8769 | Ref |
| 10828 | MN945904 | Unassigned | Unassigned | Unassigned | 234_0 | Clustered | 21 | VC_234_0 | 21 | 0.8355 | 1 | 0.8355 | 0 | 0 | 0 | 1 | Ref |
| 10829 | MN945905 | Unassigned | Unassigned | Unassigned | 868_0 | Clustered | 25 | VC_868_0 | 25 | 0.5791 | 1 | 0.5791 | 0 | 0 | 0 | 1 | Ref |
| 10830 | MN953776 | Unassigned | Unassigned | Unassigned | 133_14 | Clustered | 3 | VC_133_14 | 3 | 0.0102 | 0.96327256 | 0.0098 | 0 | 0 | 0 | 1 | Ref |
| 10831 | MN954399 | Unassigned | Unassigned | Unassigned | 971_1 | Clustered | 24 | VC_971_1 | 24 | 0.7393 | 1 | 0.7393 | 0 | 0 | 0 | 0.8913 | Ref |
| 10832 | MN956514 | Unassigned | Unassigned | Unassigned | 190_25 | Clustered/Singleton | 1 | VC_190_25 | 1 | 0 | 1 | 0 | 0 | 0 | 0 | 1 | Ref |
| 10833 | MN958086 | Unassigned | Unassigned | Unassigned | 971_1 | Clustered | 24 | VC_971_1 | 24 | 0.7393 | 1 | 0.7393 | 0 | 0 | 0 | 0.8913 | Ref |
| 10834 | MN966730 | Unassigned | Unassigned | Unassigned | 971_1 | Clustered | 24 | VC_971_1 | 24 | 0.7393 | 1 | 0.7393 | 0 | 0 | 0 | 0.8913 | Ref |
| 10835 | MN966731 | Unassigned | Unassigned | Unassigned | 971_1 | Clustered | 24 | VC_971_1 | 24 | 0.7393 | 1 | 0.7393 | 0 | 0 | 0 | 0.8913 | Ref |
| 10836 | MN966732 | Unassigned | Unassigned | Unassigned | 971_1 | Clustered | 24 | VC_971_1 | 24 | 0.7393 | 1 | 0.7393 | 0 | 0 | 0 | 0.8913 | Ref |
| 10837 | MN970211 | Unassigned | Unassigned | Unassigned | 190_13 | Clustered | 4 | VC_190_13 | 4 | 0.0216 | 0.99997041 | 0.0216 | 0 | 0 | 0 | 0.6667 | Ref |
| 10838 | MN974282 | Unassigned | Unassigned | Unassigned | 837_0 | Clustered | 29 | VC_837_0 | 29 | 0.9955 | 1 | 0.9955 | 0 | 0 | 0 | 1 | Ref |
| 10839 | MN988459 | Unassigned | Unassigned | Unassigned | 886_0 | Clustered | 2 | VC_886_0 | 2 | 0.1375 | 0.95352335 | 0.1311 | 0 | 0 | 0 | 1 | Ref |
| 10840 | MN988460 | Unassigned | Unassigned | Unassigned | 1403_0 | Clustered | 28 | VC_1403_0 | 28 | 0.753 | 1 | 0.753 | 0 | 0 | 0 | 1 | Ref |
| 10841 | MN988461 | Unassigned | Unassigned | Unassigned | 1414_0 | Clustered | 4 | VC_1414_0 | 4 | 1 | 0 | 0 | 0 | 0 | 0 | 1 | Ref |
| 10842 | MN988462 | Unassigned | Unassigned | Unassigned | 1414_0 | Clustered | 4 | VC_1414_0 | 4 | 1 | 0 | 0 | 0 | 0 | 0 | 1 | Ref |
| 10843 | MN988463 | Unassigned | Unassigned | Unassigned | 1414_0 | Clustered | 4 | VC_1414_0 | 4 | 1 | 0 | 0 | 0 | 0 | 0 | 1 | Ref |
| 10844 | MN988465 | Unassigned | Unassigned | Unassigned | 468_0 | Clustered | 4 | VC_468_0 | 4 | 0.143 | 0.99998478 | 0.143 | 0 | 0 | 0 | 1 | Ref |
| 10845 | MN988466 | Unassigned | Unassigned | Unassigned | 987_0 | Clustered | 22 | VC_987_0 | 22 | 0.8955 | 1 | 0.8955 | 0 | 0 | 0 | 1 | Ref |
| 10846 | MN988467 | Unassigned | Unassigned | Unassigned | 1403_0 | Clustered | 28 | VC_1403_0 | 28 | 0.753 | 1 | 0.753 | 0 | 0 | 0 | 1 | Ref |
| 10847 | MN988468 | Unassigned | Unassigned | Unassigned | 1045_0 | Clustered | 2 | VC_1045_0 | 2 | 0.4318 | 0.94398033 | 0.4076 | 0 | 0 | 0 | 1 | Ref |
| 10848 | MN988469 | Unassigned | Unassigned | Unassigned | 1403_0 | Clustered | 28 | VC_1403_0 | 28 | 0.753 | 1 | 0.753 | 0 | 0 | 0 | 1 | Ref |
| 10849 | MN988470 | Unassigned | Unassigned | Unassigned | 970_0 | Clustered | 9 | VC_970_0 | 9 | 0.9685 | 1 | 0.9685 | 0 | 0 | 0 | 1 | Ref |
| 10850 | MN988471 | Unassigned | Unassigned | Unassigned | 1403_0 | Clustered | 28 | VC_1403_0 | 28 | 0.753 | 1 | 0.753 | 0 | 0 | 0 | 1 | Ref |
| 10851 | MN988472 | Unassigned | Unassigned | Unassigned | 1414_0 | Clustered | 4 | VC_1414_0 | 4 | 1 | 0 | 0 | 0 | 0 | 0 | 1 | Ref |
| 10852 | MN988473 | Unassigned | Unassigned | Unassigned | 1403_0 | Clustered | 28 | VC_1403_0 | 28 | 0.753 | 1 | 0.753 | 0 | 0 | 0 | 1 | Ref |

| | | | | | | | | | | | | | | | | | |
|---|---|---|---|---|---|---|---|---|---|---|---|---|---|---|---|---|---|
| 10853 | MN988474 | Unassigned | Unassigned | Unassigned | 1403_0 | Clustered | 28 | VC_1403_0 | 28 | 0.753 | 1 | 0.753 | 0 | 0 | 0 | 1 | Ref |
| 10854 | MN988475 | Unassigned | Unassigned | Unassigned | 1403_0 | Clustered | 28 | VC_1403_0 | 28 | 0.753 | 1 | 0.753 | 0 | 0 | 0 | 1 | Ref |
| 10855 | MN988478 | Unassigned | Unassigned | Unassigned | 1403_0 | Clustered | 28 | VC_1403_0 | 28 | 0.753 | 1 | 0.753 | 0 | 0 | 0 | 1 | Ref |
| 10856 | MN988479 | Unassigned | Unassigned | Unassigned | 1403_0 | Clustered | 28 | VC_1403_0 | 28 | 0.753 | 1 | 0.753 | 0 | 0 | 0 | 1 | Ref |
| 10857 | MN988480 | Unassigned | Unassigned | Unassigned | 1403_0 | Clustered | 28 | VC_1403_0 | 28 | 0.753 | 1 | 0.753 | 0 | 0 | 0 | 1 | Ref |
| 10858 | MN988481 | Unassigned | Unassigned | Unassigned | 1403_0 | Clustered | 28 | VC_1403_0 | 28 | 0.753 | 1 | 0.753 | 0 | 0 | 0 | 1 | Ref |
| 10859 | MN988482 | Unassigned | Unassigned | Unassigned | 946_0 | Clustered | 11 | VC_946_0 | 11 | 0.7352 | 1 | 0.7352 | 0 | 0 | 0 | 1 | Ref |
| 10860 | MN988484 | Unassigned | Unassigned | Unassigned | 970_0 | Clustered | 9 | VC_970_0 | 9 | 0.9685 | 1 | 0.9685 | 0 | 0 | 0 | 1 | Ref |
| 10861 | MN988485 | Unassigned | Unassigned | Unassigned | 1415_0 | Clustered | 2 | VC_1415_0 | 2 | 1 | 1 | 1 | 0 | 0 | 0 | 1 | Ref |
| 10862 | MN988486 | Unassigned | Unassigned | Unassigned | 1052_4 | Clustered | 6 | VC_1052_4 | 6 | 0.2437 | 0.99795925 | 0.2432 | 0 | 0 | 0 | 1 | Ref |
| 10863 | MN988488 | Unassigned | Unassigned | Unassigned | 987_0 | Clustered | 22 | VC_987_0 | 22 | 0.8955 | 1 | 0.8955 | 0 | 0 | 0 | 1 | Ref |
| 10864 | MN988489 | Unassigned | Unassigned | Unassigned | 1403_0 | Clustered | 28 | VC_1403_0 | 28 | 0.753 | 1 | 0.753 | 0 | 0 | 0 | 1 | Ref |
| 10865 | MN988490 | Unassigned | Unassigned | Unassigned | 771_0 | Clustered | 2 | VC_771_0 | 2 | 0.762 | 0.91600907 | 0.698 | 0 | 0 | 0 | 1 | Ref |
| 10866 | MN988491 | Unassigned | Unassigned | Unassigned | 1403_0 | Clustered | 28 | VC_1403_0 | 28 | 0.753 | 1 | 0.753 | 0 | 0 | 0 | 1 | Ref |
| 10867 | MN988492 | Unassigned | Unassigned | Unassigned | 987_0 | Clustered | 22 | VC_987_0 | 22 | 0.8955 | 1 | 0.8955 | 0 | 0 | 0 | 1 | Ref |
| 10868 | MN988493 | Unassigned | Unassigned | Unassigned | 987_0 | Clustered | 22 | VC_987_0 | 22 | 0.8955 | 1 | 0.8955 | 0 | 0 | 0 | 1 | Ref |
| 10869 | MN988494 | Unassigned | Unassigned | Unassigned | 1403_0 | Clustered | 28 | VC_1403_0 | 28 | 0.753 | 1 | 0.753 | 0 | 0 | 0 | 1 | Ref |
| 10870 | MN988495 | Unassigned | Unassigned | Unassigned | 970_0 | Clustered | 9 | VC_970_0 | 9 | 0.9685 | 1 | 0.9685 | 0 | 0 | 0 | 1 | Ref |
| 10871 | MN988496 | Unassigned | Unassigned | Unassigned | 770_0 | Clustered | 5 | VC_770_0 | 5 | 0.8394 | 0.99999405 | 0.8394 | 0 | 0 | 0 | 1 | Ref |
| 10872 | MN988498 | Unassigned | Unassigned | Unassigned | 1403_0 | Clustered | 28 | VC_1403_0 | 28 | 0.753 | 1 | 0.753 | 0 | 0 | 0 | 1 | Ref |
| 10873 | MN988499 | Unassigned | Unassigned | Unassigned | 1403_0 | Clustered | 28 | VC_1403_0 | 28 | 0.753 | 1 | 0.753 | 0 | 0 | 0 | 1 | Ref |
| 10874 | MN988500 | Unassigned | Unassigned | Unassigned | 946_0 | Clustered | 11 | VC_946_0 | 11 | 0.7352 | 1 | 0.7352 | 0 | 0 | 0 | 1 | Ref |
| 10875 | MN988501 | Unassigned | Unassigned | Unassigned | 946_0 | Clustered | 11 | VC_946_0 | 11 | 0.7352 | 1 | 0.7352 | 0 | 0 | 0 | 1 | Ref |
| 10876 | MN988502 | Unassigned | Unassigned | Unassigned | 771_0 | Clustered | 2 | VC_771_0 | 2 | 0.762 | 0.91600907 | 0.698 | 0 | 0 | 0 | 1 | Ref |
| 10877 | MN988503 | Unassigned | Unassigned | Unassigned | 1403_0 | Clustered | 28 | VC_1403_0 | 28 | 0.753 | 1 | 0.753 | 0 | 0 | 0 | 1 | Ref |
| 10878 | MN988504 | Unassigned | Unassigned | Unassigned | 1403_0 | Clustered | 28 | VC_1403_0 | 28 | 0.753 | 1 | 0.753 | 0 | 0 | 0 | 1 | Ref |
| 10879 | MN988505 | Unassigned | Unassigned | Unassigned | 724_1 | Clustered | 6 | VC_724_1 | 6 | 0.4379 | 1 | 0.4379 | 0 | 0 | 0 | 1 | Ref |
| 10880 | MN988506 | Unassigned | Unassigned | Unassigned | 724_1 | Clustered | 6 | VC_724_1 | 6 | 0.4379 | 1 | 0.4379 | 0 | 0 | 0 | 1 | Ref |
| 10881 | MN988507 | Unassigned | Unassigned | Unassigned | 759_0 | Clustered | 3 | VC_759_0 | 3 | 0.4748 | 0.99813327 | 0.4739 | 0 | 0 | 0 | 1 | Ref |
| 10882 | MN988508 | Unassigned | Unassigned | Unassigned | 946_0 | Clustered | 11 | VC_946_0 | 11 | 0.7352 | 1 | 0.7352 | 0 | 0 | 0 | 1 | Ref |
| 10883 | MN988509 | Unassigned | Unassigned | Unassigned | 1052_4 | Clustered | 6 | VC_1052_4 | 6 | 0.2437 | 0.99795925 | 0.2432 | 0 | 0 | 0 | 1 | Ref |
| 10884 | MN988510 | Unassigned | Unassigned | Unassigned | 759_0 | Clustered | 3 | VC_759_0 | 3 | 0.4748 | 0.99813327 | 0.4739 | 0 | 0 | 0 | 1 | Ref |
| 10885 | MN988511 | Unassigned | Unassigned | Unassigned | 1403_0 | Clustered | 28 | VC_1403_0 | 28 | 0.753 | 1 | 0.753 | 0 | 0 | 0 | 1 | Ref |
| 10886 | MN988512 | Unassigned | Unassigned | Unassigned | 987_0 | Clustered | 22 | VC_987_0 | 22 | 0.8955 | 1 | 0.8955 | 0 | 0 | 0 | 1 | Ref |
| 10887 | MN988513 | Unassigned | Unassigned | Unassigned | 987_0 | Clustered | 22 | VC_987_0 | 22 | 0.8955 | 1 | 0.8955 | 0 | 0 | 0 | 1 | Ref |
| 10888 | MN988514 | Unassigned | Unassigned | Unassigned | 987_0 | Clustered | 22 | VC_987_0 | 22 | 0.8955 | 1 | 0.8955 | 0 | 0 | 0 | 1 | Ref |
| 10889 | MN988515 | Unassigned | Unassigned | Unassigned | 1416_0 | Clustered | 5 | VC_1416_0 | 5 | 0.9955 | 0.99988353 | 0.9954 | 0 | 0 | 0 | 1 | Ref |
| 10890 | MN988516 | Unassigned | Unassigned | Unassigned | 1045_0 | Clustered | 2 | VC_1045_0 | 2 | 0.4318 | 0.94398033 | 0.4076 | 0 | 0 | 0 | 1 | Ref |
| 10891 | MN988518 | Unassigned | Unassigned | Unassigned | 1403_0 | Clustered | 28 | VC_1403_0 | 28 | 0.753 | 1 | 0.753 | 0 | 0 | 0 | 1 | Ref |
| 10892 | MN988520 | Unassigned | Unassigned | Unassigned | 1052_4 | Clustered | 6 | VC_1052_4 | 6 | 0.2437 | 0.99795925 | 0.2432 | 0 | 0 | 0 | 1 | Ref |
| 10893 | MN988521 | Unassigned | Unassigned | Unassigned | 1203_0 | Clustered | 7 | VC_1203_0 | 7 | 0.6515 | 1 | 0.6515 | 0 | 0 | 0 | 1 | Ref |
| 10894 | MN988522 | Unassigned | Unassigned | Unassigned | 1203_0 | Clustered | 7 | VC_1203_0 | 7 | 0.6515 | 1 | 0.6515 | 0 | 0 | 0 | 1 | Ref |
| 10895 | MN988523 | Unassigned | Unassigned | Unassigned | 1203_0 | Clustered | 7 | VC_1203_0 | 7 | 0.6515 | 1 | 0.6515 | 0 | 0 | 0 | 1 | Ref |
| 10896 | MN988524 | Unassigned | Unassigned | Unassigned | 1203_0 | Clustered | 7 | VC_1203_0 | 7 | 0.6515 | 1 | 0.6515 | 0 | 0 | 0 | 1 | Ref |
| 10897 | MN988525 | Unassigned | Unassigned | Unassigned | 1203_0 | Clustered | 7 | VC_1203_0 | 7 | 0.6515 | 1 | 0.6515 | 0 | 0 | 0 | 1 | Ref |
| 10898 | MN988526 | Unassigned | Unassigned | Unassigned | 342_0 | Clustered | 2 | VC_342_0 | 2 | 0.1817 | 0.93411407 | 0.1698 | 0 | 0 | 0 | 1 | Ref |
| 10899 | MN988527 | Unassigned | Unassigned | Unassigned | 759_0 | Clustered | 3 | VC_759_0 | 3 | 0.4748 | 0.99813327 | 0.4739 | 0 | 0 | 0 | 1 | Ref |
| 10900 | MN988528 | Unassigned | Unassigned | Unassigned | 987_0 | Clustered | 22 | VC_987_0 | 22 | 0.8955 | 1 | 0.8955 | 0 | 0 | 0 | 1 | Ref |
| 10901 | MN988529 | Unassigned | Unassigned | Unassigned | 987_0 | Clustered | 22 | VC_987_0 | 22 | 0.8955 | 1 | 0.8955 | 0 | 0 | 0 | 1 | Ref |
| 10902 | MN988530 | Unassigned | Unassigned | Unassigned | 946_0 | Clustered | 11 | VC_946_0 | 11 | 0.7352 | 1 | 0.7352 | 0 | 0 | 0 | 1 | Ref |
| 10903 | MN988531 | Unassigned | Unassigned | Unassigned | 970_0 | Clustered | 9 | VC_970_0 | 9 | 0.9685 | 1 | 0.9685 | 0 | 0 | 0 | 1 | Ref |
| 10904 | MN988532 | Unassigned | Unassigned | Unassigned | 1052_4 | Clustered | 6 | VC_1052_4 | 6 | 0.2437 | 0.99795925 | 0.2432 | 0 | 0 | 0 | 1 | Ref |
| 10905 | MN988533 | Unassigned | Unassigned | Unassigned | 908_0 | Clustered | 3 | VC_908_0 | 3 | 0.5083 | 0.9972453 | 0.5069 | 0 | 0 | 0 | 1 | Ref |
| 10906 | MN988534 | Unassigned | Unassigned | Unassigned | 1052_4 | Clustered | 6 | VC_1052_4 | 6 | 0.2437 | 0.99795925 | 0.2432 | 0 | 0 | 0 | 1 | Ref |
| 10907 | MN988535 | Unassigned | Unassigned | Unassigned | 1416_0 | Clustered | 5 | VC_1416_0 | 5 | 0.9955 | 0.99988353 | 0.9954 | 0 | 0 | 0 | 1 | Ref |
| 10908 | MN988536 | Unassigned | Unassigned | Unassigned | 935_0 | Clustered | 2 | VC_935_0 | 2 | 0.7079 | 0.72985431 | 0.5167 | 0 | 0 | 0 | 1 | Ref |
| 10909 | MN988537 | Unassigned | Unassigned | Unassigned | 970_0 | Clustered | 9 | VC_970_0 | 9 | 0.9685 | 1 | 0.9685 | 0 | 0 | 0 | 1 | Ref |
| 10910 | MN988538 | Unassigned | Unassigned | Unassigned | 908_0 | Clustered | 3 | VC_908_0 | 3 | 0.5083 | 0.9972453 | 0.5069 | 0 | 0 | 0 | 1 | Ref |
| 10911 | MN988540 | Unassigned | Unassigned | Unassigned | 1416_0 | Clustered | 5 | VC_1416_0 | 5 | 0.9955 | 0.99988353 | 0.9954 | 0 | 0 | 0 | 1 | Ref |
| 10912 | MN988541 | Unassigned | Unassigned | Unassigned | 468_0 | Clustered | 4 | VC_468_0 | 4 | 0.143 | 0.99998478 | 0.143 | 0 | 0 | 0 | 1 | Ref |
| 10913 | MN988543 | Unassigned | Unassigned | Unassigned | 724_1 | Clustered | 6 | VC_724_1 | 6 | 0.4379 | 1 | 0.4379 | 0 | 0 | 0 | 1 | Ref |
| 10914 | MN988544 | Unassigned | Unassigned | Unassigned | 970_0 | Clustered | 9 | VC_970_0 | 9 | 0.9685 | 1 | 0.9685 | 0 | 0 | 0 | 1 | Ref |
| 10915 | MN988545 | Unassigned | Unassigned | Unassigned | 946_0 | Clustered | 11 | VC_946_0 | 11 | 0.7352 | 1 | 0.7352 | 0 | 0 | 0 | 1 | Ref |
| 10916 | MN988546 | Unassigned | Unassigned | Unassigned | 970_0 | Clustered | 9 | VC_970_0 | 9 | 0.9685 | 1 | 0.9685 | 0 | 0 | 0 | 1 | Ref |
| 10917 | MN988547 | Unassigned | Unassigned | Unassigned | 724_1 | Clustered | 6 | VC_724_1 | 6 | 0.4379 | 1 | 0.4379 | 0 | 0 | 0 | 1 | Ref |
| 10918 | MN988548 | Unassigned | Unassigned | Unassigned | 724_1 | Clustered | 6 | VC_724_1 | 6 | 0.4379 | 1 | 0.4379 | 0 | 0 | 0 | 1 | Ref |
| 10919 | MN988549 | Unassigned | Unassigned | Unassigned | 946_0 | Clustered | 11 | VC_946_0 | 11 | 0.7352 | 1 | 0.7352 | 0 | 0 | 0 | 1 | Ref |
| 10920 | MN988550 | Unassigned | Unassigned | Unassigned | 1415_0 | Clustered | 2 | VC_1415_0 | 2 | 1 | 1 | 1 | 0 | 0 | 0 | 1 | Ref |
| 10921 | MN988551 | Unassigned | Unassigned | Unassigned | 744_0 | Clustered | 10 | VC_744_0 | 10 | 0.7049 | 1 | 0.7049 | 0 | 0 | 0 | 1 | Ref |
| 10922 | MN988552 | Unassigned | Unassigned | Unassigned | 990_0 | Clustered | 3 | VC_990_0 | 3 | 0.6363 | 0.99819562 | 0.6351 | 0 | 0 | 0 | 1 | Ref |
| 10923 | MN988553 | Unassigned | Unassigned | Unassigned | 1052_4 | Clustered | 6 | VC_1052_4 | 6 | 0.2437 | 0.99795925 | 0.2432 | 0 | 0 | 0 | 1 | Ref |
| 10924 | MN988554 | Unassigned | Unassigned | Unassigned | 468_0 | Clustered | 4 | VC_468_0 | 4 | 0.143 | 0.99998478 | 0.143 | 0 | 0 | 0 | 1 | Ref |
| 10925 | MN988556 | Unassigned | Unassigned | Unassigned | 342_0 | Clustered | 2 | VC_342_0 | 2 | 0.1817 | 0.93411407 | 0.1698 | 0 | 0 | 0 | 1 | Ref |
| 10926 | MN988557 | Unassigned | Unassigned | Unassigned | 970_0 | Clustered | 9 | VC_970_0 | 9 | 0.9685 | 1 | 0.9685 | 0 | 0 | 0 | 1 | Ref |
| 10927 | MN988558 | Unassigned | Unassigned | Unassigned | 886_0 | Clustered | 2 | VC_886_0 | 2 | 0.1375 | 0.95352335 | 0.1311 | 0 | 0 | 0 | 1 | Ref |
| 10928 | MN988559 | Unassigned | Unassigned | Unassigned | 1203_0 | Clustered | 7 | VC_1203_0 | 7 | 0.6515 | 1 | 0.6515 | 0 | 0 | 0 | 1 | Ref |
| 10929 | MN988560 | Unassigned | Unassigned | Unassigned | 1203_1 | Clustered/Singleton | 1 | VC_1203_1 | 1 | 0 | 1 | 0 | 0 | 0 | 0 | 1 | Ref |
| 10930 | MN988561 | Unassigned | Unassigned | Unassigned | 1203_0 | Clustered | 7 | VC_1203_0 | 7 | 0.6515 | 1 | 0.6515 | 0 | 0 | 0 | 1 | Ref |
| 10931 | MN994275 | Unassigned | Unassigned | Unassigned | 1224_0 | Clustered | 19 | VC_1224_0 | 19 | 1 | 1 | 1 | 0 | 0 | 0 | 1 | Ref |
| 10932 | MN994496 | Unassigned | Unassigned | Unassigned | 190_15 | Clustered | 3 | VC_190_15 | 3 | 0.0146 | 0.99759465 | 0.0145 | 0 | 0 | 0 | 0.6667 | Ref |
| 10933 | MN994497 | Unassigned | Unassigned | Unassigned | 283_185 | Clustered/Singleton | 1 | VC_283_185 | 1 | 0 | 1 | 0 | 0 | 0 | 0 | 1 | Ref |
| 10934 | MN994498 | Unassigned | Unassigned | Unassigned | 329_10 | Clustered/Singleton | 1 | VC_329_10 | 1 | 0 | 1 | 0 | 0 | 0 | 0 | 1 | Ref |
| 10935 | MN994499 | Unassigned | Unassigned | Unassigned | 329_5 | Clustered/Singleton | 1 | VC_329_5 | 1 | 0 | 1 | 0 | 0 | 0 | 0 | 1 | Ref |
| 10936 | MN994500 | Unassigned | Unassigned | Unassigned | 584_0 | Clustered | 28 | VC_584_0 | 28 | 0.7704 | 1 | 0.7704 | 0 | 0 | 0 | 1 | Ref |
| 10937 | MN994501 | Unassigned | Unassigned | Unassigned | 191_16 | Clustered | 12 | VC_191_16 | 12 | 0.0643 | 1 | 0.0643 | 0 | 0 | 0 | 0.9394 | Ref |
| 10938 | MN994502 | Unassigned | Unassigned | Unassigned | 190_13 | Clustered | 4 | VC_190_13 | 4 | 0.0216 | 0.99997041 | 0.0216 | 0 | 0 | 0 | 0.6667 | Ref |
| 10939 | MN994503 | Unassigned | Unassigned | Unassigned | 329_8 | Clustered | 2 | VC_329_8 | 2 | 0.0048 | 0.95654014 | 0.0046 | 0 | 0 | 0 | 1 | Ref |
| 10940 | MN994504 | Unassigned | Unassigned | Unassigned | 190_3 | Clustered/Singleton | 1 | VC_190_3 | 1 | 0 | 1 | 0 | 0 | 0 | 0 | 1 | Ref |
| 10941 | MN996301 | Unassigned | Unassigned | Unassigned | 160_0 | Clustered | 2 | VC_160_0 | 2 | 0.4956 | 0.94363853 | 0.4677 | 0 | 0 | 0 | 1 | Ref |
| 10942 | MPU46938 | Unassigned | Unassigned | Unassigned | 554_0 | Clustered | 2 | VC_554_0 | 2 | 0.2292 | 0.95193785 | 0.2181 | 0 | 0 | 0 | 1 | Ref |
| 10943 | MT001829 | Unassigned | Unassigned | Unassigned | 164_0 | Clustered | 3 | VC_164_0 | 3 | 0.0298 | 0.82983726 | 0.0247 | 0 | 0 | 0 | 1 | Ref |
| 10944 | MT002875 | Unassigned | Unassigned | Unassigned | 896_0 | Clustered | 5 | VC_896_0 | 5 | 0.4981 | 0.99999977 | 0.4981 | 0 | 0 | 0 | 1 | Ref |
| 10945 | MT002975 | Unassigned | Unassigned | Unassigned | 879_0 | Clustered | 36 | VC_879_0 | 36 | 0.994 | 1 | 0.994 | 0 | 0 | 0 | 1 | Ref |
| 10946 | MT004791 | Unassigned | Unassigned | Unassigned | 191_5 | Clustered | 6 | VC_191_5 | 6 | 0.0285 | 0.99999996 | 0.0285 | 0 | 0 | 0 | 1 | Ref |
| 10947 | MT006214 | Unassigned | Unassigned | Unassigned | 673_0 | Clustered | 15 | VC_673_0 | 15 | 0.8234 | 1 | 0.8234 | 0 | 0 | 0 | 0.7619 | Ref |
| 10948 | MT006233 | Unassigned | Unassigned | Unassigned | 745_0 | Clustered | 4 | VC_745_0 | 4 | 0.5585 | 0.99995802 | 0.5585 | 0 | 0 | 0 | 1 | Ref |
| 10949 | MT006234 | Unassigned | Unassigned | Unassigned | 1418_0 | Clustered | 2 | VC_1418_0 | 2 | 1 | 1 | 1 | 0 | 0 | 0 | 1 | Ref |
| 10950 | MT006235 | Unassigned | Unassigned | Unassigned | 1418_0 | Clustered | 2 | VC_1418_0 | 2 | 1 | 1 | 1 | 0 | 0 | 0 | 1 | Ref |
| 10951 | MT006237 | Unassigned | Unassigned | Unassigned | 1198_0 | Clustered | 3 | VC_1198_0 | 3 | 0.2794 | 0.95148141 | 0.2658 | 0 | 0 | 0 | 1 | Ref |
| 10952 | MT006238 | Unassigned | Unassigned | Unassigned | 745_0 | Clustered | 4 | VC_745_0 | 4 | 0.5585 | 0.99995802 | 0.5585 | 0 | 0 | 0 | 1 | Ref |
| 10953 | MT006239 | Unassigned | Unassigned | Unassigned | 745_0 | Clustered | 4 | VC_745_0 | 4 | 0.5585 | 0.99995802 | 0.5585 | 0 | 0 | 0 | 1 | Ref |
| 10954 | MT006240 | Unassigned | Unassigned | Unassigned | 745_0 | Clustered | 4 | VC_745_0 | 4 | 0.5585 | 0.99995802 | 0.5585 | 0 | 0 | 0 | 1 | Ref |
| 10955 | MT012302 | Unassigned | Unassigned | Unassigned | 125_0 | Clustered | 17 | VC_125_0 | 17 | 0.9244 | 1 | 0.9244 | 0 | 0 | 0 | 1 | Ref |
| 10956 | MT012729 | Unassigned | Unassigned | Unassigned | 540_0 | Clustered | 15 | VC_540_0 | 15 | 0.2866 | 1 | 0.2866 | 0 | 0 | 0 | 1 | Ref |
| 10957 | MT012730 | Unassigned | Unassigned | Unassigned | 428_0 | Clustered | 33 | VC_428_0 | 33 | 0.9915 | 1 | 0.9915 | 0 | 0 | 0 | 1 | Ref |
| 10958 | MT013111 | Unassigned | Unassigned | Unassigned | 9_0 | Clustered | 38 | VC_9_0 | 38 | 0.4895 | 1 | 0.4895 | 0 | 0 | 0 | 1 | Ref |
| 10959 | MT023084 | Unassigned | Unassigned | Unassigned | 576_0 | Clustered | 4 | VC_576_0 | 4 | 0.0606 | 0.99998766 | 0.0606 | 0 | 0 | 0 | 1 | Ref |
| 10960 | MT024860 | Unassigned | Unassigned | Unassigned | 1234_0 | Clustered | 44 | VC_1234_0 | 44 | 1 | 1 | 1 | 0 | 0 | 0 | 1 | Ref |
| 10961 | MT024861 | Unassigned | Unassigned | Unassigned | 223_0 | Clustered | 38 | VC_223_0 | 38 | 0.9095 | 1 | 0.9095 | 0 | 0 | 0 | 1 | Ref |
| 10962 | MT024863 | Unassigned | Unassigned | Unassigned | 1361_0 | Clustered | 6 | VC_1361_0 | 6 | 0.2371 | 1 | 0.2371 | 0 | 0 | 0 | 1 | Ref |
| 10963 | MT024864 | Unassigned | Unassigned | Unassigned | 1240_0 | Clustered | 82 | VC_1240_0 | 82 | 0.7945 | 1 | 0.7945 | 0 | 0 | 0 | 1 | Ref |
| 10964 | MT024865 | Unassigned | Unassigned | Unassigned | 1072_1 | Clustered | 2 | VC_1072_1 | 2 | 0.0259 | 0.95128042 | 0.0246 | 0 | 0 | 0 | 1 | Ref |
| 10965 | MT024866 | Unassigned | Unassigned | Unassigned | 519_0 | Clustered | 39 | VC_519_0 | 39 | 0.8156 | 1 | 0.8156 | 0 | 0 | 0 | 1 | Ref |
| 10966 | MT024867 | Unassigned | Unassigned | Unassigned | 1362_0 | Clustered | 6 | VC_1362_0 | 6 | 1 | 1 | 1 | 0 | 0 | 0 | 1 | Ref |
| 10967 | MT024868 | Unassigned | Unassigned | Unassigned | 1073_0 | Clustered | 24 | VC_1073_0 | 24 | 0.8388 | 1 | 0.8388 | 0 | 0 | 0 | 1 | Ref |
| 10968 | MT024869 | Unassigned | Unassigned | Unassigned | 1207_0 | Clustered | 29 | VC_1207_0 | 29 | 0.8225 | 1 | 0.8225 | 0 | 0 | 0 | 1 | Ref |
| 10969 | MT024870 | Unassigned | Unassigned | Unassigned | 1362_0 | Clustered | 6 | VC_1362_0 | 6 | 1 | 1 | 1 | 0 | 0 | 0 | 1 | Ref |
| 10970 | MT024871 | Unassigned | Unassigned | Unassigned | 1207_0 | Clustered | 29 | VC_1207_0 | 29 | 0.8225 | 1 | 0.8225 | 0 | 0 | 0 | 1 | Ref |
| 10971 | MT024872 | Unassigned | Unassigned | Unassigned | 1072_1 | Clustered | 2 | VC_1072_1 | 2 | 0.0259 | 0.95128042 | 0.0246 | 0 | 0 | 0 | 1 | Ref |
| 10972 | MT028297 | Unassigned | Unassigned | Unassigned | 812_1 | Clustered | 4 | VC_812_1 | 4 | 0.2266 | 0.99998426 | 0.2266 | 0 | 0 | 0 | 1 | Ref |
| 10973 | MT047590 | Unassigned | Unassigned | Unassigned | 440_0 | Clustered | 4 | VC_440_0 | 4 | 1 | 1 | 1 | 0 | 0 | 0 | 1 | Ref |
| 10974 | MT066160 | Unassigned | Unassigned | Unassigned | 754_0 | Clustered | 8 | VC_754_0 | 8 | 0.752 | 1 | 0.752 | 0 | 0 | 0 | 1 | Ref |
| 10975 | MT074134 | Unassigned | Unassigned | Unassigned | 1007_0 | Clustered | 19 | VC_1007_0 | 19 | 0.86 | 1 | 0.86 | 0 | 0 | 0 | 1 | Ref |
| 10976 | MT074135 | Unassigned | Unassigned | Unassigned | 1007_0 | Clustered | 19 | VC_1007_0 | 19 | 0.86 | 1 | 0.86 | 0 | 0 | 0 | 1 | Ref |
| 10977 | MT074136 | Unassigned | Unassigned | Unassigned | 1018_1 | Clustered | 10 | VC_1018_1 | 10 | 0.7518 | 1 | 0.7518 | 0 | 0 | 0 | 0.8 | Ref |
| 10978 | MT074137 | Unassigned | Unassigned | Unassigned | 1419_0 | Clustered | 6 | VC_1419_0 | 6 | 0.9543 | 0.99999304 | 0.9543 | 0 | 0 | 0 | 1 | Ref |
| 10979 | MT074138 | Unassigned | Unassigned | Unassigned | 1018_1 | Clustered | 10 | VC_1018_1 | 10 | 0.7518 | 1 | 0.7518 | 0 | 0 | 0 | 0.8 | Ref |
| 10980 | MT074139 | Unassigned | Unassigned | Unassigned | 1419_0 | Clustered | 6 | VC_1419_0 | 6 | 0.9543 | 0.99999304 | 0.9543 | 0 | 0 | 0 | 1 | Ref |
| 10981 | MT074140 | Unassigned | Unassigned | Unassigned | 1419_0 | Clustered | 6 | VC_1419_0 | 6 | 0.9543 | 0.99999304 | 0.9543 | 0 | 0 | 0 | 1 | Ref |
| 10982 | MT074141 | Unassigned | Unassigned | Unassigned | 1419_0 | Clustered | 6 | VC_1419_0 | 6 | 0.9543 | 0.99999304 | 0.9543 | 0 | 0 | 0 | 1 | Ref |
| 10983 | MT074142 | Unassigned | Unassigned | Unassigned | 1419_0 | Clustered | 6 | VC_1419_0 | 6 | 0.9543 | 0.99999304 | 0.9543 | 0 | 0 | 0 | 1 | Ref |
| 10984 | MT074143 | Unassigned | Unassigned | Unassigned | 1007_0 | Clustered | 19 | VC_1007_0 | 19 | 0.86 | 1 | 0.86 | 0 | 0 | 0 | 1 | Ref |
| 10985 | MT074144 | Unassigned | Unassigned | Unassigned | 1007_0 | Clustered | 19 | VC_1007_0 | 19 | 0.86 | 1 | 0.86 | 0 | 0 | 0 | 1 | Ref |
| 10986 | MT074145 | Unassigned | Unassigned | Unassigned | 1007_0 | Clustered | 19 | VC_1007_0 | 19 | 0.86 | 1 | 0.86 | 0 | 0 | 0 | 1 | Ref |
| 10987 | MT074146 | Unassigned | Unassigned | Unassigned | 1419_0 | Clustered | 6 | VC_1419_0 | 6 | 0.9543 | 0.99999304 | 0.9543 | 0 | 0 | 0 | 1 | Ref |
| 10988 | MT074147 | Unassigned | Unassigned | Unassigned | 1007_0 | Clustered | 19 | VC_1007_0 | 19 | 0.86 | 1 | 0.86 | 0 | 0 | 0 | 1 | Ref |
| 10989 | MT074148 | Unassigned | Unassigned | Unassigned | 1007_0 | Clustered | 19 | VC_1007_0 | 19 | 0.86 | 1 | 0.86 | 0 | 0 | 0 | 1 | Ref |
| 10990 | MT074149 | Unassigned | Unassigned | Unassigned | 1007_0 | Clustered | 19 | VC_1007_0 | 19 | 0.86 | 1 | 0.86 | 0 | 0 | 0 | 1 | Ref |
| 10991 | MT074150 | Unassigned | Unassigned | Unassigned | 1007_0 | Clustered | 19 | VC_1007_0 | 19 | 0.86 | 1 | 0.86 | 0 | 0 | 0 | 1 | Ref |
| 10992 | MT074151 | Unassigned | Unassigned | Unassigned | 1007_0 | Clustered | 19 | VC_1007_0 | 19 | 0.86 | 1 | 0.86 | 0 | 0 | 0 | 1 | Ref |
| 10993 | MT074152 | Unassigned | Unassigned | Unassigned | 1007_0 | Clustered | 19 | VC_1007_0 | 19 | 0.86 | 1 | 0.86 | 0 | 0 | 0 | 1 | Ref |
| 10994 | MT074153 | Unassigned | Unassigned | Unassigned | 1007_0 | Clustered | 19 | VC_1007_0 | 19 | 0.86 | 1 | 0.86 | 0 | 0 | 0 | 1 | Ref |
| 10995 | MT074154 | Unassigned | Unassigned | Unassigned | 1007_0 | Clustered | 19 | VC_1007_0 | 19 | 0.86 | 1 | 0.86 | 0 | 0 | 0 | 1 | Ref |
| 10996 | MT074155 | Unassigned | Unassigned | Unassigned | 1007_0 | Clustered | 19 | VC_1007_0 | 19 | 0.86 | 1 | 0.86 | 0 | 0 | 0 | 1 | Ref |
| 10997 | MT074156 | Unassigned | Unassigned | Unassigned | 1007_0 | Clustered | 19 | VC_1007_0 | 19 | 0.86 | 1 | 0.86 | 0 | 0 | 0 | 1 | Ref |
| 10998 | MT074157 | Unassigned | Unassigned | Unassigned | 1007_0 | Clustered | 19 | VC_1007_0 | 19 | 0.86 | 1 | 0.86 | 0 | 0 | 0 | 1 | Ref |
| 10999 | MT074158 | Unassigned | Unassigned | Unassigned | 1007_0 | Clustered | 19 | VC_1007_0 | 19 | 0.86 | 1 | 0.86 | 0 | 0 | 0 | 1 | Ref |
| 11000 | MT074159 | Unassigned | Unassigned | Unassigned | 1007_0 | Clustered | 19 | VC_1007_0 | 19 | 0.86 | 1 | 0.86 | 0 | 0 | 0 | 1 | Ref |
| 11001 | MT074160 | Unassigned | Unassigned | Unassigned | 1007_0 | Clustered | 19 | VC_1007_0 | 19 | 0.86 | 1 | 0.86 | 0 | 0 | 0 | 1 | Ref |
| 11002 | MT074429 | Unassigned | Unassigned | Unassigned | 1218_0 | Clustered | 4 | VC_1218_0 | 4 | 0.8905 | 0.99530591 | 0.8863 | 0 | 0 | 0 | 1 | Ref |
| 11003 | MT074430 | Unassigned | Unassigned | Unassigned | 637_1 | Clustered | 27 | VC_637_1 | 27 | 0.2578 | 1 | 0.2578 | 0 | 0 | 0 | 0.9858 | Ref |
| 11004 | MT074431 | Unassigned | Unassigned | Unassigned | 637_0 | Clustered | 115 | VC_637_0 | 115 | 0.731 | 1 | 0.731 | 0 | 0 | 0 | 0.8764 | Ref |
| 11005 | MT074432 | Unassigned | Unassigned | Unassigned | 637_0 | Clustered | 115 | VC_637_0 | 115 | 0.731 | 1 | 0.731 | 0 | 0 | 0 | 0.8764 | Ref |
| 11006 | MT074433 | Unassigned | Unassigned | Unassigned | 941_0 | Clustered | 40 | VC_941_0 | 40 | 0.6903 | 1 | 0.6903 | 0 | 0 | 0 | 1 | Ref |
| 11007 | MT074434 | Unassigned | Unassigned | Unassigned | 428_0 | Clustered | 33 | VC_428_0 | 33 | 0.9915 | 1 | 0.9915 | 0 | 0 | 0 | 1 | Ref |
| 11008 | MT074435 | Unassigned | Unassigned | Unassigned | 428_0 | Clustered | 33 | VC_428_0 | 33 | 0.9915 | 1 | 0.9915 | 0 | 0 | 0 | 1 | Ref |
| 11009 | MT074436 | Unassigned | Unassigned | Unassigned | 428_0 | Clustered | 33 | VC_428_0 | 33 | 0.9915 | 1 | 0.9915 | 0 | 0 | 0 | 1 | Ref |
| 11010 | MT074437 | Unassigned | Unassigned | Unassigned | 428_0 | Clustered | 33 | VC_428_0 | 33 | 0.9915 | 1 | 0.9915 | 0 | 0 | 0 | 1 | Ref |
| 11011 | MT074438 | Unassigned | Unassigned | Unassigned | 428_0 | Clustered | 33 | VC_428_0 | 33 | 0.9915 | 1 | 0.9915 | 0 | 0 | 0 | 1 | Ref |
| 11012 | MT074439 | Unassigned | Unassigned | Unassigned | 428_0 | Clustered | 33 | VC_428_0 | 33 | 0.9915 | 1 | 0.9915 | 0 | 0 | 0 | 1 | Ref |
| 11013 | MT074440 | Unassigned | Unassigned | Unassigned | 1218_0 | Clustered | 4 | VC_1218_0 | 4 | 0.8905 | 0.99530591 | 0.8863 | 0 | 0 | 0 | 1 | Ref |
| 11014 | MT074441 | Unassigned | Unassigned | Unassigned | 428_0 | Clustered | 33 | VC_428_0 | 33 | 0.9915 | 1 | 0.9915 | 0 | 0 | 0 | 1 | Ref |

| | | | | | | | | | | | | | | | | | |
|---|---|---|---|---|---|---|---|---|---|---|---|---|---|---|---|---|---|
| 11015 | MT074442 | Unassigned | Unassigned | Unassigned | 428_0 | Clustered | 33 | VC_428_0 | 33 | 0.9915 | 1 | 0.9915 | 0 | 0 | 0 | 1 | Ref |
| 11016 | MT074443 | Unassigned | Unassigned | Unassigned | 428_0 | Clustered | 33 | VC_428_0 | 33 | 0.9915 | 1 | 0.9915 | 0 | 0 | 0 | 1 | Ref |
| 11017 | MT074444 | Unassigned | Unassigned | Unassigned | 428_0 | Clustered | 33 | VC_428_0 | 33 | 0.9915 | 1 | 0.9915 | 0 | 0 | 0 | 1 | Ref |
| 11018 | MT074445 | Unassigned | Unassigned | Unassigned | 428_0 | Clustered | 33 | VC_428_0 | 33 | 0.9915 | 1 | 0.9915 | 0 | 0 | 0 | 1 | Ref |
| 11019 | MT074446 | Unassigned | Unassigned | Unassigned | 428_0 | Clustered | 33 | VC_428_0 | 33 | 0.9915 | 1 | 0.9915 | 0 | 0 | 0 | 1 | Ref |
| 11020 | MT074447 | Unassigned | Unassigned | Unassigned | 191_30 | Clustered | 2 | VC_191_30 | 2 | 0.0061 | 0.95531751 | 0.0058 | 0 | 0 | 0 | 1 | Ref |
| 11021 | MT074448 | Unassigned | Unassigned | Unassigned | 190_7 | Clustered | 7 | VC_190_7 | 7 | 0.0476 | 1 | 0.0476 | 0 | 0 | 0 | 1 | Ref |
| 11022 | MT074449 | Unassigned | Unassigned | Unassigned | 191_32 | Clustered | 12 | VC_191_32 | 12 | 0.0683 | 1 | 0.0683 | 0 | 0 | 0 | 0.7576 | Ref |
| 11023 | MT074450 | Unassigned | Unassigned | Unassigned | 191_16 | Clustered | 12 | VC_191_16 | 12 | 0.0643 | 1 | 0.0643 | 0 | 0 | 0 | 0.9394 | Ref |
| 11024 | MT074451 | Unassigned | Unassigned | Unassigned | 637_0 | Clustered | 115 | VC_637_0 | 115 | 0.731 | 1 | 0.731 | 0 | 0 | 0 | 0.8764 | Ref |
| 11025 | MT074452 | Unassigned | Unassigned | Unassigned | 191_5 | Clustered | 6 | VC_191_5 | 6 | 0.0285 | 0.99999996 | 0.0285 | 0 | 0 | 0 | 1 | Ref |
| 11026 | MT074453 | Unassigned | Unassigned | Unassigned | 191_3 | Clustered | 5 | VC_191_3 | 5 | 0.0231 | 0.99999849 | 0.0231 | 0 | 0 | 0 | 0.8 | Ref |
| 11027 | MT074454 | Unassigned | Unassigned | Unassigned | 191_8 | Clustered | 2 | VC_191_8 | 2 | 0.0055 | 0.91831352 | 0.0051 | 0 | 0 | 0 | 1 | Ref |
| 11028 | MT074455 | Unassigned | Unassigned | Unassigned | 191_4 | Clustered | 2 | VC_191_4 | 2 | 0.0055 | 0.88936514 | 0.0049 | 0 | 0 | 0 | 1 | Ref |
| 11029 | MT074456 | Unassigned | Unassigned | Unassigned | 191_5 | Clustered | 6 | VC_191_5 | 6 | 0.0285 | 0.99999996 | 0.0285 | 0 | 0 | 0 | 1 | Ref |
| 11030 | MT074457 | Unassigned | Unassigned | Unassigned | 191_3 | Clustered | 5 | VC_191_3 | 5 | 0.0231 | 0.99999849 | 0.0231 | 0 | 0 | 0 | 0.8 | Ref |
| 11031 | MT074458 | Unassigned | Unassigned | Unassigned | 191_16 | Clustered | 12 | VC_191_16 | 12 | 0.0643 | 1 | 0.0643 | 0 | 0 | 0 | 0.9394 | Ref |
| 11032 | MT074459 | Unassigned | Unassigned | Unassigned | 191_9 | Clustered | 5 | VC_191_9 | 5 | 0.0228 | 0.99999662 | 0.0228 | 0 | 0 | 0 | 0.7 | Ref |
| 11033 | MT074460 | Unassigned | Unassigned | Unassigned | 191_5 | Clustered | 6 | VC_191_5 | 6 | 0.0285 | 0.99999996 | 0.0285 | 0 | 0 | 0 | 1 | Ref |
| 11034 | MT074461 | Unassigned | Unassigned | Unassigned | 191_30 | Clustered | 2 | VC_191_30 | 2 | 0.0061 | 0.95531751 | 0.0058 | 0 | 0 | 0 | 1 | Ref |
| 11035 | MT074462 | Unassigned | Unassigned | Unassigned | 637_0 | Clustered | 115 | VC_637_0 | 115 | 0.731 | 1 | 0.731 | 0 | 0 | 0 | 0.8764 | Ref |
| 11036 | MT074463 | Unassigned | Unassigned | Unassigned | 191_5 | Clustered | 6 | VC_191_5 | 6 | 0.0285 | 0.99999996 | 0.0285 | 0 | 0 | 0 | 1 | Ref |
| 11037 | MT074464 | Unassigned | Unassigned | Unassigned | 191_12 | Clustered/Singleton | 1 | VC_191_12 | 1 | 0 | 1 | 0 | 0 | 0 | 0 | 1 | Ref |
| 11038 | MT074465 | Unassigned | Unassigned | Unassigned | 191_32 | Clustered | 12 | VC_191_32 | 12 | 0.0683 | 1 | 0.0683 | 0 | 0 | 0 | 0.7576 | Ref |
| 11039 | MT074466 | Unassigned | Unassigned | Unassigned | 191_9 | Clustered | 5 | VC_191_9 | 5 | 0.0228 | 0.99999662 | 0.0228 | 0 | 0 | 0 | 0.7 | Ref |
| 11040 | MT074467 | Unassigned | Unassigned | Unassigned | 191_7 | Clustered/Singleton | 1 | VC_191_7 | 1 | 0 | 1 | 0 | 0 | 0 | 0 | 1 | Ref |
| 11041 | MT074468 | Unassigned | Unassigned | Unassigned | 191_6 | Clustered/Singleton | 1 | VC_191_6 | 1 | 0 | 1 | 0 | 0 | 0 | 0 | 1 | Ref |
| 11042 | MT074469 | Unassigned | Unassigned | Unassigned | 133_3 | Clustered | 3 | VC_133_3 | 3 | 0.0105 | 0.99119575 | 0.0104 | 0 | 0 | 0 | 1 | Ref |
| 11043 | MT074470 | Unassigned | Unassigned | Unassigned | 133_23 | Clustered/Singleton | 1 | VC_133_23 | 1 | 0 | 1 | 0 | 0 | 0 | 0 | 1 | Ref |
| 11044 | MT074471 | Unassigned | Unassigned | Unassigned | 133_45 | Clustered | 2 | VC_133_45 | 2 | 0.0052 | 0.9489868 | 0.0049 | 0 | 0 | 0 | 1 | Ref |
| 11045 | MT074472 | Unassigned | Unassigned | Unassigned | 133_2 | Clustered/Singleton | 1 | VC_133_2 | 1 | 0 | 1 | 0 | 0 | 0 | 0 | 1 | Ref |
| 11046 | MT074473 | Unassigned | Unassigned | Unassigned | 637_0 | Clustered | 115 | VC_637_0 | 115 | 0.731 | 1 | 0.731 | 0 | 0 | 0 | 0.8764 | Ref |
| 11047 | MT074474 | Unassigned | Unassigned | Unassigned | 133_41 | Clustered/Singleton | 1 | VC_133_41 | 1 | 0 | 1 | 0 | 0 | 0 | 0 | 1 | Ref |
| 11048 | MT074475 | Unassigned | Unassigned | Unassigned | 133_19 | Clustered | 2 | VC_133_19 | 2 | 0.0051 | 0.8696643 | 0.0044 | 0 | 0 | 0 | 1 | Ref |
| 11049 | MT074476 | Unassigned | Unassigned | Unassigned | 133_31 | Clustered | 6 | VC_133_31 | 6 | 0.0263 | 0.99999694 | 0.0263 | 0 | 0 | 0 | 1 | Ref |
| 11050 | MT074477 | Unassigned | Unassigned | Unassigned | 133_8 | Clustered/Singleton | 1 | VC_133_8 | 1 | 0 | 1 | 0 | 0 | 0 | 0 | 1 | Ref |
| 11051 | MT074478 | Unassigned | Unassigned | Unassigned | 133_39 | Clustered/Singleton | 1 | VC_133_39 | 1 | 0 | 1 | 0 | 0 | 0 | 0 | 1 | Ref |
| 11052 | MT074479 | Unassigned | Unassigned | Unassigned | 133_47 | Clustered/Singleton | 1 | VC_133_47 | 1 | 0 | 1 | 0 | 0 | 0 | 0 | 1 | Ref |
| 11053 | MT074480 | Unassigned | Unassigned | Unassigned | 133_45 | Clustered | 2 | VC_133_45 | 2 | 0.0052 | 0.9489868 | 0.0049 | 0 | 0 | 0 | 1 | Ref |
| 11054 | MT074481 | Unassigned | Unassigned | Unassigned | 637_0 | Clustered | 115 | VC_637_0 | 115 | 0.731 | 1 | 0.731 | 0 | 0 | 0 | 0.8764 | Ref |
| 11055 | MT074482 | Unassigned | Unassigned | Unassigned | 637_0 | Clustered | 115 | VC_637_0 | 115 | 0.731 | 1 | 0.731 | 0 | 0 | 0 | 0.8764 | Ref |
| 11056 | MT074483 | Unassigned | Unassigned | Unassigned | 637_0 | Clustered | 115 | VC_637_0 | 115 | 0.731 | 1 | 0.731 | 0 | 0 | 0 | 0.8764 | Ref |
| 11057 | MT074484 | Unassigned | Unassigned | Unassigned | 637_0 | Clustered | 115 | VC_637_0 | 115 | 0.731 | 1 | 0.731 | 0 | 0 | 0 | 0.8764 | Ref |
| 11058 | MT075871 | Unassigned | Unassigned | Unassigned | 836_0 | Clustered | 9 | VC_836_0 | 9 | 0.7733 | 1 | 0.7733 | 0 | 0 | 0 | 1 | Ref |
| 11059 | MT078988 | Unassigned | Unassigned | Unassigned | 131_2 | Clustered | 7 | VC_131_2 | 7 | 0.0685 | 1 | 0.0685 | 0 | 0 | 0 | 0.9048 | Ref |
| 11060 | MT080102 | Unassigned | Unassigned | Unassigned | 838_0 | Clustered | 29 | VC_838_0 | 29 | 0.8611 | 1 | 0.8611 | 0 | 0 | 0 | 1 | Ref |
| 11061 | MT080103 | Unassigned | Unassigned | Unassigned | 190_29 | Clustered/Singleton | 1 | VC_190_29 | 1 | 0 | 1 | 0 | 0 | 0 | 0 | 1 | Ref |
| 11062 | MT080582 | Unassigned | Unassigned | Unassigned | 149_11 | Clustered | 4 | VC_149_11 | 4 | 0.0203 | 0.9998357 | 0.0203 | 0 | 0 | 0 | 1 | Ref |
| 11063 | MT080583 | Unassigned | Unassigned | Unassigned | 149_11 | Clustered | 4 | VC_149_11 | 4 | 0.0203 | 0.9998357 | 0.0203 | 0 | 0 | 0 | 1 | Ref |
| 11064 | MT080584 | Unassigned | Unassigned | Unassigned | 153_0 | Clustered | 25 | VC_153_0 | 25 | 0.7729 | 1 | 0.7729 | 0 | 0 | 0 | 1 | Ref |
| 11065 | MT080585 | Unassigned | Unassigned | Unassigned | 149_17 | Clustered | 4 | VC_149_17 | 4 | 0.013 | 0.99951815 | 0.013 | 0 | 0 | 0 | 1 | Ref |
| 11066 | MT080586 | Unassigned | Unassigned | Unassigned | 149_11 | Clustered | 4 | VC_149_11 | 4 | 0.0203 | 0.9998357 | 0.0203 | 0 | 0 | 0 | 1 | Ref |
| 11067 | MT080587 | Unassigned | Unassigned | Unassigned | 153_0 | Clustered | 25 | VC_153_0 | 25 | 0.7729 | 1 | 0.7729 | 0 | 0 | 0 | 1 | Ref |
| 11068 | MT080588 | Unassigned | Unassigned | Unassigned | 153_0 | Clustered | 25 | VC_153_0 | 25 | 0.7729 | 1 | 0.7729 | 0 | 0 | 0 | 1 | Ref |
| 11069 | MT080589 | Unassigned | Unassigned | Unassigned | 149_17 | Clustered | 4 | VC_149_17 | 4 | 0.013 | 0.99951815 | 0.013 | 0 | 0 | 0 | 1 | Ref |
| 11070 | MT080590 | Unassigned | Unassigned | Unassigned | 153_0 | Clustered | 25 | VC_153_0 | 25 | 0.7729 | 1 | 0.7729 | 0 | 0 | 0 | 1 | Ref |
| 11071 | MT080591 | Unassigned | Unassigned | Unassigned | 149_17 | Clustered | 4 | VC_149_17 | 4 | 0.013 | 0.99951815 | 0.013 | 0 | 0 | 0 | 1 | Ref |
| 11072 | MT080592 | Unassigned | Unassigned | Unassigned | 149_0 | Clustered | 5 | VC_149_0 | 5 | 0.0281 | 0.99999897 | 0.0281 | 0 | 0 | 0 | 1 | Ref |
| 11073 | MT080593 | Unassigned | Unassigned | Unassigned | 149_22 | Clustered | 2 | VC_149_22 | 2 | 0.0062 | 0.92916562 | 0.0058 | 0 | 0 | 0 | 1 | Ref |
| 11074 | MT080594 | Unassigned | Unassigned | Unassigned | 149_11 | Clustered | 4 | VC_149_11 | 4 | 0.0203 | 0.9998357 | 0.0203 | 0 | 0 | 0 | 1 | Ref |
| 11075 | MT080595 | Unassigned | Unassigned | Unassigned | 153_0 | Clustered | 25 | VC_153_0 | 25 | 0.7729 | 1 | 0.7729 | 0 | 0 | 0 | 1 | Ref |
| 11076 | MT080596 | Unassigned | Unassigned | Unassigned | 149_22 | Clustered | 2 | VC_149_22 | 2 | 0.0062 | 0.92916562 | 0.0058 | 0 | 0 | 0 | 1 | Ref |
| 11077 | MT080597 | Unassigned | Unassigned | Unassigned | 149_0 | Clustered | 5 | VC_149_0 | 5 | 0.0281 | 0.99999897 | 0.0281 | 0 | 0 | 0 | 1 | Ref |
| 11078 | MT080598 | Unassigned | Unassigned | Unassigned | 149_0 | Clustered | 5 | VC_149_0 | 5 | 0.0281 | 0.99999897 | 0.0281 | 0 | 0 | 0 | 1 | Ref |
| 11079 | MT080599 | Unassigned | Unassigned | Unassigned | 149_0 | Clustered | 5 | VC_149_0 | 5 | 0.0281 | 0.99999897 | 0.0281 | 0 | 0 | 0 | 1 | Ref |
| 11080 | MT080600 | Unassigned | Unassigned | Unassigned | 149_8 | Clustered | 66 | VC_149_8 | 66 | 0.4565 | 1 | 0.4565 | 0 | 0 | 0 | 0.8182 | Ref |
| 11081 | MT080601 | Unassigned | Unassigned | Unassigned | 149_17 | Clustered | 4 | VC_149_17 | 4 | 0.013 | 0.99951815 | 0.013 | 0 | 0 | 0 | 1 | Ref |
| 11082 | MT080602 | Unassigned | Unassigned | Unassigned | 149_8 | Clustered | 66 | VC_149_8 | 66 | 0.4565 | 1 | 0.4565 | 0 | 0 | 0 | 0.8182 | Ref |
| 11083 | MT080603 | Unassigned | Unassigned | Unassigned | 149_0 | Clustered | 5 | VC_149_0 | 5 | 0.0281 | 0.99999897 | 0.0281 | 0 | 0 | 0 | 1 | Ref |
| 11084 | MT084350 | Unassigned | Unassigned | Unassigned | 280_0 | Clustered | 2 | VC_280_0 | 2 | 0.34 | 0.95158328 | 0.3235 | 0 | 0 | 0 | 1 | Ref |
| 11085 | MT090077 | Unassigned | Unassigned | Unassigned | 111_0 | Clustered | 91 | VC_111_0 | 91 | 0.9587 | 1 | 0.9587 | 0 | 0 | 0 | 1 | Ref |
| 11086 | MT094431 | Unassigned | Unassigned | Unassigned | 130_0 | Clustered | 19 | VC_130_0 | 19 | 0.6068 | 1 | 0.6068 | 0 | 0 | 0 | 1 | Ref |
| 11087 | MT104465 | Unassigned | Unassigned | Unassigned | 129_0 | Clustered | 43 | VC_129_0 | 43 | 0.886 | 1 | 0.886 | 0 | 0 | 0 | 1 | Ref |
| 11088 | MT104466 | Unassigned | Unassigned | Unassigned | 129_0 | Clustered | 43 | VC_129_0 | 43 | 0.886 | 1 | 0.886 | 0 | 0 | 0 | 1 | Ref |
| 11089 | MT104468 | Unassigned | Unassigned | Unassigned | 155_0 | Clustered | 14 | VC_155_0 | 14 | 0.6095 | 1 | 0.6095 | 0 | 0 | 0 | 1 | Ref |
| 11090 | MT104469 | Unassigned | Unassigned | Unassigned | 155_0 | Clustered | 14 | VC_155_0 | 14 | 0.6095 | 1 | 0.6095 | 0 | 0 | 0 | 1 | Ref |
| 11091 | MT104470 | Unassigned | Unassigned | Unassigned | 155_0 | Clustered | 14 | VC_155_0 | 14 | 0.6095 | 1 | 0.6095 | 0 | 0 | 0 | 1 | Ref |
| 11092 | MT104471 | Unassigned | Unassigned | Unassigned | 155_0 | Clustered | 14 | VC_155_0 | 14 | 0.6095 | 1 | 0.6095 | 0 | 0 | 0 | 1 | Ref |
| 11093 | MT104472 | Unassigned | Unassigned | Unassigned | 155_0 | Clustered | 14 | VC_155_0 | 14 | 0.6095 | 1 | 0.6095 | 0 | 0 | 0 | 1 | Ref |
| 11094 | MT104473 | Unassigned | Unassigned | Unassigned | 697_0 | Clustered | 5 | VC_697_0 | 5 | 0.8303 | 0.99999971 | 0.8303 | 0 | 0 | 0 | 1 | Ref |
| 11095 | MT104475 | Unassigned | Unassigned | Unassigned | 697_0 | Clustered | 5 | VC_697_0 | 5 | 0.8303 | 0.99999971 | 0.8303 | 0 | 0 | 0 | 1 | Ref |
| 11096 | MT104476 | Unassigned | Unassigned | Unassigned | 155_0 | Clustered | 14 | VC_155_0 | 14 | 0.6095 | 1 | 0.6095 | 0 | 0 | 0 | 1 | Ref |
| 11097 | MT104477 | Unassigned | Unassigned | Unassigned | 155_0 | Clustered | 14 | VC_155_0 | 14 | 0.6095 | 1 | 0.6095 | 0 | 0 | 0 | 1 | Ref |
| 11098 | MT108723 | Unassigned | Unassigned | Unassigned | 472_0 | Clustered | 47 | VC_472_0 | 47 | 0.9025 | 1 | 0.9025 | 0 | 0 | 0 | 1 | Ref |
| 11099 | MT108724 | Unassigned | Unassigned | Unassigned | 472_0 | Clustered | 47 | VC_472_0 | 47 | 0.9025 | 1 | 0.9025 | 0 | 0 | 0 | 1 | Ref |
| 11100 | MT108725 | Unassigned | Unassigned | Unassigned | 464_0 | Clustered | 8 | VC_464_0 | 8 | 0.2629 | 1 | 0.2629 | 0 | 0 | 0 | 1 | Ref |
| 11101 | MT108726 | Unassigned | Unassigned | Unassigned | 80_0 | Clustered | 41 | VC_80_0 | 41 | 0.5837 | 1 | 0.5837 | 0 | 0 | 0 | 0.9805 | Ref |
| 11102 | MT108727 | Unassigned | Unassigned | Unassigned | 79_7 | Clustered | 33 | VC_79_7 | 33 | 0.216 | 1 | 0.216 | 0 | 0 | 0 | 0.8409 | Ref |
| 11103 | MT108728 | Unassigned | Unassigned | Unassigned | 77_0 | Clustered | 20 | VC_77_0 | 20 | 0.9657 | 1 | 0.9657 | 0 | 0 | 0 | 1 | Ref |
| 11104 | MT108729 | Unassigned | Unassigned | Unassigned | 79_6 | Clustered | 11 | VC_79_6 | 11 | 0.0528 | 1 | 0.0528 | 0 | 0 | 0 | 0.7455 | Ref |
| 11105 | MT108730 | Unassigned | Unassigned | Unassigned | 77_0 | Clustered | 20 | VC_77_0 | 20 | 0.9657 | 1 | 0.9657 | 0 | 0 | 0 | 1 | Ref |
| 11106 | MT108731 | Unassigned | Unassigned | Unassigned | 464_0 | Clustered | 8 | VC_464_0 | 8 | 0.2629 | 1 | 0.2629 | 0 | 0 | 0 | 1 | Ref |
| 11107 | MT110073 | Unassigned | Unassigned | Unassigned | 905_0 | Clustered | 3 | VC_905_0 | 3 | 0.3424 | 0.99823775 | 0.3417 | 0 | 0 | 0 | 1 | Ref |
| 11108 | MT114158 | Unassigned | Unassigned | Unassigned | 517_1 | Clustered | 108 | VC_517_1 | 108 | 0.8483 | 1 | 0.8483 | 0 | 0 | 0 | 0.9955 | Ref |
| 11109 | MT114159 | Unassigned | Unassigned | Unassigned | 810_0 | Clustered | 18 | VC_810_0 | 18 | 0.9121 | 1 | 0.9121 | 0 | 0 | 0 | 1 | Ref |
| 11110 | MT114160 | Unassigned | Unassigned | Unassigned | 868_0 | Clustered | 25 | VC_868_0 | 25 | 0.5791 | 1 | 0.5791 | 0 | 0 | 0 | 1 | Ref |
| 11111 | MT114161 | Unassigned | Unassigned | Unassigned | 1073_0 | Clustered | 24 | VC_1073_0 | 24 | 0.8388 | 1 | 0.8388 | 0 | 0 | 0 | 1 | Ref |
| 11112 | MT114162 | Unassigned | Unassigned | Unassigned | 1072_3 | Clustered | 6 | VC_1072_3 | 6 | 0.1304 | 0.99999994 | 0.1304 | 0 | 0 | 0 | 0.8667 | Ref |
| 11113 | MT114164 | Unassigned | Unassigned | Unassigned | 520_76 | Clustered | 2 | VC_520_76 | 2 | 0.0092 | 0.95761376 | 0.0088 | 0 | 0 | 0 | 1 | Ref |
| 11114 | MT114165 | Unassigned | Unassigned | Unassigned | 551_2 | Clustered | 17 | VC_551_2 | 17 | 0.0973 | 1 | 0.0973 | 0 | 0 | 0 | 0.9779 | Ref |
| 11115 | MT114166 | Unassigned | Unassigned | Unassigned | 1171_0 | Clustered | 21 | VC_1171_0 | 21 | 0.7125 | 1 | 0.7125 | 0 | 0 | 0 | 1 | Ref |
| 11116 | MT114167 | Unassigned | Unassigned | Unassigned | 520_28 | Clustered | 2 | VC_520_28 | 2 | 0.0137 | 0.95761531 | 0.0131 | 0 | 0 | 0 | 1 | Ref |
| 11117 | MT118288 | Unassigned | Unassigned | Unassigned | 472_0 | Clustered | 47 | VC_472_0 | 47 | 0.9025 | 1 | 0.9025 | 0 | 0 | 0 | 1 | Ref |
| 11118 | MT118289 | Unassigned | Unassigned | Unassigned | 79_4 | Clustered | 3 | VC_79_4 | 3 | 0.0106 | 0.99632092 | 0.0105 | 0 | 0 | 0 | 0.6667 | Ref |
| 11119 | MT118290 | Unassigned | Unassigned | Unassigned | 79_7 | Clustered | 33 | VC_79_7 | 33 | 0.216 | 1 | 0.216 | 0 | 0 | 0 | 0.8409 | Ref |
| 11120 | MT118291 | Unassigned | Unassigned | Unassigned | 79_0 | Clustered | 64 | VC_79_0 | 64 | 0.4185 | 1 | 0.4185 | 0 | 0 | 0 | 0.7649 | Ref |
| 11121 | MT118292 | Unassigned | Unassigned | Unassigned | 79_4 | Clustered | 3 | VC_79_4 | 3 | 0.0106 | 0.99632092 | 0.0105 | 0 | 0 | 0 | 0.6667 | Ref |
| 11122 | MT118293 | Unassigned | Unassigned | Unassigned | 80_0 | Clustered | 41 | VC_80_0 | 41 | 0.5837 | 1 | 0.5837 | 0 | 0 | 0 | 0.9805 | Ref |
| 11123 | MT118294 | Unassigned | Unassigned | Unassigned | 77_0 | Clustered | 20 | VC_77_0 | 20 | 0.9657 | 1 | 0.9657 | 0 | 0 | 0 | 1 | Ref |
| 11124 | MT118295 | Unassigned | Unassigned | Unassigned | 77_0 | Clustered | 20 | VC_77_0 | 20 | 0.9657 | 1 | 0.9657 | 0 | 0 | 0 | 1 | Ref |
| 11125 | MT118297 | Unassigned | Unassigned | Unassigned | 79_0 | Clustered | 64 | VC_79_0 | 64 | 0.4185 | 1 | 0.4185 | 0 | 0 | 0 | 0.7649 | Ref |
| 11126 | MT118298 | Unassigned | Unassigned | Unassigned | 79_0 | Clustered | 64 | VC_79_0 | 64 | 0.4185 | 1 | 0.4185 | 0 | 0 | 0 | 0.7649 | Ref |
| 11127 | MT118299 | Unassigned | Unassigned | Unassigned | 79_0 | Clustered | 64 | VC_79_0 | 64 | 0.4185 | 1 | 0.4185 | 0 | 0 | 0 | 0.7649 | Ref |
| 11128 | MT118300 | Unassigned | Unassigned | Unassigned | 77_0 | Clustered | 20 | VC_77_0 | 20 | 0.9657 | 1 | 0.9657 | 0 | 0 | 0 | 1 | Ref |
| 11129 | MT118301 | Unassigned | Unassigned | Unassigned | 630_0 | Clustered | 20 | VC_630_0 | 20 | 0.8513 | 1 | 0.8513 | 0 | 0 | 0 | 1 | Ref |
| 11130 | MT118302 | Unassigned | Unassigned | Unassigned | 166_0 | Clustered | 26 | VC_166_0 | 26 | 0.7923 | 1 | 0.7923 | 0 | 0 | 0 | 1 | Ref |
| 11131 | MT118303 | Unassigned | Unassigned | Unassigned | 80_0 | Clustered | 41 | VC_80_0 | 41 | 0.5837 | 1 | 0.5837 | 0 | 0 | 0 | 0.9805 | Ref |
| 11132 | MT118304 | Unassigned | Unassigned | Unassigned | 749_0 | Clustered | 40 | VC_749_0 | 40 | 0.7517 | 1 | 0.7517 | 0 | 0 | 0 | 1 | Ref |
| 11133 | MT118305 | Unassigned | Unassigned | Unassigned | 749_0 | Clustered | 40 | VC_749_0 | 40 | 0.7517 | 1 | 0.7517 | 0 | 0 | 0 | 1 | Ref |
| 11134 | MT119359 | Unassigned | Unassigned | Unassigned | 695_0 | Clustered | 48 | VC_695_0 | 48 | 0.8162 | 1 | 0.8162 | 0 | 0 | 0 | 1 | Ref |
| 11135 | MT119360 | Unassigned | Unassigned | Unassigned | 691_1 | Clustered | 5 | VC_691_1 | 5 | 0.4358 | 0.99999993 | 0.4358 | 0 | 0 | 0 | 1 | Ref |
| 11136 | MT119361 | Unassigned | Unassigned | Unassigned | 691_1 | Clustered | 5 | VC_691_1 | 5 | 0.4358 | 0.99999993 | 0.4358 | 0 | 0 | 0 | 1 | Ref |
| 11137 | MT119362 | Unassigned | Unassigned | Unassigned | 472_0 | Clustered | 47 | VC_472_0 | 47 | 0.9025 | 1 | 0.9025 | 0 | 0 | 0 | 1 | Ref |
| 11138 | MT119363 | Unassigned | Unassigned | Unassigned | 79_0 | Clustered | 64 | VC_79_0 | 64 | 0.4185 | 1 | 0.4185 | 0 | 0 | 0 | 0.7649 | Ref |
| 11139 | MT119364 | Unassigned | Unassigned | Unassigned | 80_0 | Clustered | 41 | VC_80_0 | 41 | 0.5837 | 1 | 0.5837 | 0 | 0 | 0 | 0.9805 | Ref |
| 11140 | MT119365 | Unassigned | Unassigned | Unassigned | 79_0 | Clustered | 64 | VC_79_0 | 64 | 0.4185 | 1 | 0.4185 | 0 | 0 | 0 | 0.7649 | Ref |
| 11141 | MT119366 | Unassigned | Unassigned | Unassigned | 79_7 | Clustered | 33 | VC_79_7 | 33 | 0.216 | 1 | 0.216 | 0 | 0 | 0 | 0.8409 | Ref |
| 11142 | MT119367 | Unassigned | Unassigned | Unassigned | 80_0 | Clustered | 41 | VC_80_0 | 41 | 0.5837 | 1 | 0.5837 | 0 | 0 | 0 | 0.9805 | Ref |
| 11143 | MT119368 | Unassigned | Unassigned | Unassigned | 79_0 | Clustered | 64 | VC_79_0 | 64 | 0.4185 | 1 | 0.4185 | 0 | 0 | 0 | 0.7649 | Ref |
| 11144 | MT119369 | Unassigned | Unassigned | Unassigned | 79_7 | Clustered | 33 | VC_79_7 | 33 | 0.216 | 1 | 0.216 | 0 | 0 | 0 | 0.8409 | Ref |
| 11145 | MT119370 | Unassigned | Unassigned | Unassigned | 80_0 | Clustered | 41 | VC_80_0 | 41 | 0.5837 | 1 | 0.5837 | 0 | 0 | 0 | 0.9805 | Ref |
| 11146 | MT119371 | Unassigned | Unassigned | Unassigned | 472_0 | Clustered | 47 | VC_472_0 | 47 | 0.9025 | 1 | 0.9025 | 0 | 0 | 0 | 1 | Ref |
| 11147 | MT119372 | Unassigned | Unassigned | Unassigned | 80_0 | Clustered | 41 | VC_80_0 | 41 | 0.5837 | 1 | 0.5837 | 0 | 0 | 0 | 0.9805 | Ref |
| 11148 | MT119373 | Unassigned | Unassigned | Unassigned | 472_0 | Clustered | 47 | VC_472_0 | 47 | 0.9025 | 1 | 0.9025 | 0 | 0 | 0 | 1 | Ref |
| 11149 | MT119374 | Unassigned | Unassigned | Unassigned | 80_0 | Clustered | 41 | VC_80_0 | 41 | 0.5837 | 1 | 0.5837 | 0 | 0 | 0 | 0.9805 | Ref |
| 11150 | MT119375 | Unassigned | Unassigned | Unassigned | 80_0 | Clustered | 41 | VC_80_0 | 41 | 0.5837 | 1 | 0.5837 | 0 | 0 | 0 | 0.9805 | Ref |
| 11151 | MT119376 | Unassigned | Unassigned | Unassigned | 79_0 | Clustered | 64 | VC_79_0 | 64 | 0.4185 | 1 | 0.4185 | 0 | 0 | 0 | 0.7649 | Ref |
| 11152 | MT119377 | Unassigned | Unassigned | Unassigned | 80_0 | Clustered | 41 | VC_80_0 | 41 | 0.5837 | 1 | 0.5837 | 0 | 0 | 0 | 0.9805 | Ref |
| 11153 | MT119378 | Unassigned | Unassigned | Unassigned | 80_0 | Clustered | 41 | VC_80_0 | 41 | 0.5837 | 1 | 0.5837 | 0 | 0 | 0 | 0.9805 | Ref |
| 11154 | MT119766 | Unassigned | Unassigned | Unassigned | 934_0 | Clustered | 2 | VC_934_0 | 2 | 0.8613 | 0.92499224 | 0.7967 | 0 | 0 | 0 | 1 | Ref |
| 11155 | MT121960 | Unassigned | Unassigned | Unassigned | 1420_0 | Clustered | 5 | VC_1420_0 | 5 | 1 | 1 | 1 | 0 | 0 | 0 | 1 | Ref |
| 11156 | MT121964 | Unassigned | Unassigned | Unassigned | 1420_0 | Clustered | 5 | VC_1420_0 | 5 | 1 | 1 | 1 | 0 | 0 | 0 | 1 | Ref |
| 11157 | MT121966 | Unassigned | Unassigned | Unassigned | 241_0 | Clustered | 2 | VC_241_0 | 2 | 0.072 | 0.95433582 | 0.0687 | 0 | 0 | 0 | 1 | Ref |
| 11158 | MT121967 | Unassigned | Unassigned | Unassigned | 355_0 | Clustered | 2 | VC_355_0 | 2 | 0.0513 | 0.93418388 | 0.0479 | 0 | 0 | 0 | 1 | Ref |
| 11159 | MT127619 | Unassigned | Unassigned | Unassigned | 941_0 | Clustered | 40 | VC_941_0 | 40 | 0.6903 | 1 | 0.6903 | 0 | 0 | 0 | 1 | Ref |
| 11160 | MT127620 | Unassigned | Unassigned | Unassigned | 813_1 | Clustered | 24 | VC_813_1 | 24 | 0.535 | 1 | 0.535 | 0 | 0 | 0 | 1 | Ref |
| 11161 | MT129652 | Unassigned | Unassigned | Unassigned | 290_0 | Clustered | 10 | VC_290_0 | 10 | 0.0981 | 1 | 0.0981 | 0 | 0 | 0 | 1 | Ref |
| 11162 | MT129653 | Unassigned | Unassigned | Unassigned | 813_1 | Clustered | 24 | VC_813_1 | 24 | 0.535 | 1 | 0.535 | 0 | 0 | 0 | 1 | Ref |
| 11163 | MT129654 | Unassigned | Unassigned | Unassigned | 290_6 | Clustered | 12 | VC_290_6 | 12 | 0.1149 | 1 | 0.1149 | 0 | 0 | 0 | 0.8182 | Ref |
| 11164 | MT129655 | Unassigned | Unassigned | Unassigned | 813_1 | Clustered | 24 | VC_813_1 | 24 | 0.535 | 1 | 0.535 | 0 | 0 | 0 | 1 | Ref |
| 11165 | MT133560 | Unassigned | Unassigned | Unassigned | 344_0 | Clustered | 21 | VC_344_0 | 21 | 0.5436 | 1 | 0.5436 | 0 | 0 | 0 | 1 | Ref |
| 11166 | MT133561 | Unassigned | Unassigned | Unassigned | 79_0 | Clustered | 64 | VC_79_0 | 64 | 0.4185 | 1 | 0.4185 | 0 | 0 | 0 | 0.7649 | Ref |
| 11167 | MT133562 | Unassigned | Unassigned | Unassigned | 79_0 | Clustered | 64 | VC_79_0 | 64 | 0.4185 | 1 | 0.4185 | 0 | 0 | 0 | 0.7649 | Ref |
| 11168 | MT133563 | Unassigned | Unassigned | Unassigned | 80_0 | Clustered | 41 | VC_80_0 | 41 | 0.5837 | 1 | 0.5837 | 0 | 0 | 0 | 0.9805 | Ref |
| 11169 | MT135024 | Unassigned | Unassigned | Unassigned | 531_6 | Clustered/Singleton | 1 | VC_531_6 | 1 | 0 | 1 | 0 | 0 | 0 | 0 | 1 | Ref |
| 11170 | MT135025 | Unassigned | Unassigned | Unassigned | 531_5 | Clustered/Singleton | 1 | VC_531_5 | 1 | 0 | 1 | 0 | 0 | 0 | 0 | 1 | Ref |
| 11171 | MT135026 | Unassigned | Unassigned | Unassigned | 531_1 | Clustered | 6 | VC_531_1 | 6 | 0.1481 | 0.99978665 | 0.148 | 0 | 0 | 0 | 1 | Ref |
| 11172 | MT135176 | Unassigned | Unassigned | Unassigned | 242_1 | Clustered | 20 | VC_242_1 | 20 | 0.684 | 1 | 0.684 | 0 | 0 | 0 | 0.8368 | Ref |
| 11173 | MT135255 | Unassigned | Unassigned | Unassigned | 1149_0 | Clustered | 5 | VC_1149_0 | 5 | 0.4768 | 0.9999998 | 0.4768 | 0 | 0 | 0 | 1 | Ref |
| 11174 | MT135256 | Unassigned | Unassigned | Unassigned | 1117_0 | Clustered | 5 | VC_1117_0 | 5 | 0.4688 | 0.99999974 | 0.4688 | 0 | 0 | 0 | 1 | Ref |
| 11175 | MT135257 | Unassigned | Unassigned | Unassigned | 1104_0 | Clustered | 154 | VC_1104_0 | 154 | 0.8484 | 1 | 0.8484 | 0 | 0 | 0 | 1 | Ref |
| 11176 | MT135258 | Unassigned | Unassigned | Unassigned | 1124_0 | Clustered | 10 | VC_1124_0 | 10 | 0.6268 | 1 | 0.6268 | 0 | 0 | 0 | 1 | Ref |

| | | | | | | | | | | | | | | | | | |
|---|---|---|---|---|---|---|---|---|---|---|---|---|---|---|---|---|---|
| 11177 | MT135259 | Unassigned | Unassigned | Unassigned | 1126_0 | Clustered | 8 | VC_1126_0 | 8 | 0.2864 | 1 | 0.2864 | 0 | 0 | 0 | 1 | Ref |
| 11178 | MT135261 | Unassigned | Unassigned | Unassigned | 1110_0 | Clustered | 8 | VC_1110_0 | 8 | 0.6915 | 1 | 0.6915 | 0 | 0 | 0 | 1 | Ref |
| 11179 | MT135262 | Unassigned | Unassigned | Unassigned | 1421_0 | Clustered | 4 | VC_1421_0 | 4 | 1 | 1 | 1 | 0 | 0 | 0 | 1 | Ref |
| 11180 | MT135263 | Unassigned | Unassigned | Unassigned | 1104_0 | Clustered | 154 | VC_1104_0 | 154 | 0.8484 | 1 | 0.8484 | 0 | 0 | 0 | 1 | Ref |
| 11181 | MT135264 | Unassigned | Unassigned | Unassigned | 1104_0 | Clustered | 154 | VC_1104_0 | 154 | 0.8484 | 1 | 0.8484 | 0 | 0 | 0 | 1 | Ref |
| 11182 | MT135265 | Unassigned | Unassigned | Unassigned | 1311_0 | Clustered | 5 | VC_1311_0 | 5 | 0.8316 | 0.99879053 | 0.8306 | 0 | 0 | 0 | 1 | Ref |
| 11183 | MT135266 | Unassigned | Unassigned | Unassigned | 1117_0 | Clustered | 5 | VC_1117_0 | 5 | 0.4688 | 0.99999974 | 0.4688 | 0 | 0 | 0 | 1 | Ref |
| 11184 | MT135267 | Unassigned | Unassigned | Unassigned | 1148_0 | Clustered | 16 | VC_1148_0 | 16 | 0.8821 | 1 | 0.8821 | 0 | 0 | 0 | 1 | Ref |
| 11185 | MT135269 | Unassigned | Unassigned | Unassigned | 1341_0 | Clustered | 2 | VC_1341_0 | 2 | 0.7733 | 0.5 | 0.3867 | 0 | 0 | 0 | 1 | Ref |
| 11186 | MT135271 | Unassigned | Unassigned | Unassigned | 1148_0 | Clustered | 16 | VC_1148_0 | 16 | 0.8821 | 1 | 0.8821 | 0 | 0 | 0 | 1 | Ref |
| 11187 | MT135272 | Unassigned | Unassigned | Unassigned | 1104_0 | Clustered | 154 | VC_1104_0 | 154 | 0.8484 | 1 | 0.8484 | 0 | 0 | 0 | 1 | Ref |
| 11188 | MT135273 | Unassigned | Unassigned | Unassigned | 1104_0 | Clustered | 154 | VC_1104_0 | 154 | 0.8484 | 1 | 0.8484 | 0 | 0 | 0 | 1 | Ref |
| 11189 | MT135274 | Unassigned | Unassigned | Unassigned | 1421_0 | Clustered | 4 | VC_1421_0 | 4 | 1 | 1 | 1 | 0 | 0 | 0 | 1 | Ref |
| 11190 | MT135275 | Unassigned | Unassigned | Unassigned | 1149_0 | Clustered | 5 | VC_1149_0 | 5 | 0.4768 | 0.9999998 | 0.4768 | 0 | 0 | 0 | 1 | Ref |
| 11191 | MT135276 | Unassigned | Unassigned | Unassigned | 1283_0 | Clustered | 26 | VC_1283_0 | 26 | 0.5446 | 1 | 0.5446 | 0 | 0 | 0 | 1 | Ref |
| 11192 | MT135277 | Unassigned | Unassigned | Unassigned | 1127_0 | Clustered | 19 | VC_1127_0 | 19 | 0.7611 | 1 | 0.7611 | 0 | 0 | 0 | 1 | Ref |
| 11193 | MT135278 | Unassigned | Unassigned | Unassigned | 1104_0 | Clustered | 154 | VC_1104_0 | 154 | 0.8484 | 1 | 0.8484 | 0 | 0 | 0 | 1 | Ref |
| 11194 | MT135281 | Unassigned | Unassigned | Unassigned | 1148_0 | Clustered | 16 | VC_1148_0 | 16 | 0.8821 | 1 | 0.8821 | 0 | 0 | 0 | 1 | Ref |
| 11195 | MT135282 | Unassigned | Unassigned | Unassigned | 1111_0 | Clustered | 2 | VC_1111_0 | 2 | 0.2635 | 0.89847992 | 0.2367 | 0 | 0 | 0 | 1 | Ref |
| 11196 | MT135284 | Unassigned | Unassigned | Unassigned | 1422_0 | Clustered | 2 | VC_1422_0 | 2 | 1 | 1 | 1 | 0 | 0 | 0 | 1 | Ref |
| 11197 | MT135285 | Unassigned | Unassigned | Unassigned | 1423_0 | Clustered | 8 | VC_1423_0 | 8 | 0.896 | 0.99983932 | 0.8959 | 0 | 0 | 0 | 1 | Ref |
| 11198 | MT135286 | Unassigned | Unassigned | Unassigned | 1423_0 | Clustered | 8 | VC_1423_0 | 8 | 0.896 | 0.99983932 | 0.8959 | 0 | 0 | 0 | 1 | Ref |
| 11199 | MT135287 | Unassigned | Unassigned | Unassigned | 1425_0 | Clustered | 7 | VC_1425_0 | 7 | 0.6976 | 0.99999999 | 0.6976 | 0 | 0 | 0 | 1 | Ref |
| 11200 | MT135288 | Unassigned | Unassigned | Unassigned | 1424_0 | Clustered | 8 | VC_1424_0 | 8 | 0.7842 | 1 | 0.7842 | 0 | 0 | 0 | 1 | Ref |
| 11201 | MT135289 | Unassigned | Unassigned | Unassigned | 1423_0 | Clustered | 8 | VC_1423_0 | 8 | 0.896 | 0.99983932 | 0.8959 | 0 | 0 | 0 | 1 | Ref |
| 11202 | MT135290 | Unassigned | Unassigned | Unassigned | 1424_0 | Clustered | 8 | VC_1424_0 | 8 | 0.7842 | 1 | 0.7842 | 0 | 0 | 0 | 1 | Ref |
| 11203 | MT135291 | Unassigned | Unassigned | Unassigned | 1424_0 | Clustered | 8 | VC_1424_0 | 8 | 0.7842 | 1 | 0.7842 | 0 | 0 | 0 | 1 | Ref |
| 11204 | MT135292 | Unassigned | Unassigned | Unassigned | 1426_0 | Clustered | 4 | VC_1426_0 | 4 | 1 | 1 | 1 | 0 | 0 | 0 | 1 | Ref |
| 11205 | MT135293 | Unassigned | Unassigned | Unassigned | 1426_0 | Clustered | 4 | VC_1426_0 | 4 | 1 | 1 | 1 | 0 | 0 | 0 | 1 | Ref |
| 11206 | MT135294 | Unassigned | Unassigned | Unassigned | 1425_0 | Clustered | 7 | VC_1425_0 | 7 | 0.6976 | 0.99999999 | 0.6976 | 0 | 0 | 0 | 1 | Ref |
| 11207 | MT135296 | Unassigned | Unassigned | Unassigned | 1426_0 | Clustered | 4 | VC_1426_0 | 4 | 1 | 1 | 1 | 0 | 0 | 0 | 1 | Ref |
| 11208 | MT135297 | Unassigned | Unassigned | Unassigned | 1423_0 | Clustered | 8 | VC_1423_0 | 8 | 0.896 | 0.99983932 | 0.8959 | 0 | 0 | 0 | 1 | Ref |
| 11209 | MT135298 | Unassigned | Unassigned | Unassigned | 1425_0 | Clustered | 7 | VC_1425_0 | 7 | 0.6976 | 0.99999999 | 0.6976 | 0 | 0 | 0 | 1 | Ref |
| 11210 | MT135299 | Unassigned | Unassigned | Unassigned | 1425_0 | Clustered | 7 | VC_1425_0 | 7 | 0.6976 | 0.99999999 | 0.6976 | 0 | 0 | 0 | 1 | Ref |
| 11211 | MT135301 | Unassigned | Unassigned | Unassigned | 1424_0 | Clustered | 8 | VC_1424_0 | 8 | 0.7842 | 1 | 0.7842 | 0 | 0 | 0 | 1 | Ref |
| 11212 | MT135302 | Unassigned | Unassigned | Unassigned | 1423_0 | Clustered | 8 | VC_1423_0 | 8 | 0.896 | 0.99983932 | 0.8959 | 0 | 0 | 0 | 1 | Ref |
| 11213 | MT135303 | Unassigned | Unassigned | Unassigned | 1424_0 | Clustered | 8 | VC_1424_0 | 8 | 0.7842 | 1 | 0.7842 | 0 | 0 | 0 | 1 | Ref |
| 11214 | MT135304 | Unassigned | Unassigned | Unassigned | 1425_0 | Clustered | 7 | VC_1425_0 | 7 | 0.6976 | 0.99999999 | 0.6976 | 0 | 0 | 0 | 1 | Ref |
| 11215 | MT135305 | Unassigned | Unassigned | Unassigned | 1426_0 | Clustered | 4 | VC_1426_0 | 4 | 1 | 1 | 1 | 0 | 0 | 0 | 1 | Ref |
| 11216 | MT135306 | Unassigned | Unassigned | Unassigned | 1425_0 | Clustered | 7 | VC_1425_0 | 7 | 0.6976 | 0.99999999 | 0.6976 | 0 | 0 | 0 | 1 | Ref |
| 11217 | MT135307 | Unassigned | Unassigned | Unassigned | 1424_0 | Clustered | 8 | VC_1424_0 | 8 | 0.7842 | 1 | 0.7842 | 0 | 0 | 0 | 1 | Ref |
| 11218 | MT135308 | Unassigned | Unassigned | Unassigned | 1423_0 | Clustered | 8 | VC_1423_0 | 8 | 0.896 | 0.99983932 | 0.8959 | 0 | 0 | 0 | 1 | Ref |
| 11219 | MT135309 | Unassigned | Unassigned | Unassigned | 1424_0 | Clustered | 8 | VC_1424_0 | 8 | 0.7842 | 1 | 0.7842 | 0 | 0 | 0 | 1 | Ref |
| 11220 | MT135310 | Unassigned | Unassigned | Unassigned | 1425_0 | Clustered | 7 | VC_1425_0 | 7 | 0.6976 | 0.99999999 | 0.6976 | 0 | 0 | 0 | 1 | Ref |
| 11221 | MT135311 | Unassigned | Unassigned | Unassigned | 1424_0 | Clustered | 8 | VC_1424_0 | 8 | 0.7842 | 1 | 0.7842 | 0 | 0 | 0 | 1 | Ref |
| 11222 | MT135314 | Unassigned | Unassigned | Unassigned | 1423_0 | Clustered | 8 | VC_1423_0 | 8 | 0.896 | 0.99983932 | 0.8959 | 0 | 0 | 0 | 1 | Ref |
| 11223 | MT135326 | Unassigned | Unassigned | Unassigned | 1308_0 | Clustered | 12 | VC_1308_0 | 12 | 0.8184 | 0.9810731 | 0.8029 | 0 | 0 | 0 | 1 | Ref |
| 11224 | MT135328 | Unassigned | Unassigned | Unassigned | 1309_0 | Clustered | 2 | VC_1309_0 | 2 | 0.309 | 0.72985431 | 0.2255 | 0 | 0 | 0 | 1 | Ref |
| 11225 | MT135329 | Unassigned | Unassigned | Unassigned | 1259_0 | Clustered | 8 | VC_1259_0 | 8 | 0.8225 | 1 | 0.8225 | 0 | 0 | 0 | 1 | Ref |
| 11226 | MT135330 | Unassigned | Unassigned | Unassigned | 1285_0 | Clustered | 25 | VC_1285_0 | 25 | 1 | 1 | 1 | 0 | 0 | 0 | 1 | Ref |
| 11227 | MT135331 | Unassigned | Unassigned | Unassigned | 1298_0 | Clustered | 3 | VC_1298_0 | 3 | 0.4889 | 0.98167473 | 0.4799 | 0 | 0 | 0 | 1 | Ref |
| 11228 | MT135332 | Unassigned | Unassigned | Unassigned | 1427_0 | Clustered | 2 | VC_1427_0 | 2 | 1 | 1 | 1 | 0 | 0 | 0 | 1 | Ref |
| 11229 | MT135333 | Unassigned | Unassigned | Unassigned | 1277_0 | Clustered | 14 | VC_1277_0 | 14 | 0.8218 | 1 | 0.8218 | 0 | 0 | 0 | 1 | Ref |
| 11230 | MT135334 | Unassigned | Unassigned | Unassigned | 1110_0 | Clustered | 8 | VC_1110_0 | 8 | 0.6915 | 1 | 0.6915 | 0 | 0 | 0 | 1 | Ref |
| 11231 | MT135335 | Unassigned | Unassigned | Unassigned | 1340_0 | Clustered | 5 | VC_1340_0 | 5 | 1 | 1 | 1 | 0 | 0 | 0 | 1 | Ref |
| 11232 | MT135336 | Unassigned | Unassigned | Unassigned | 1251_0 | Clustered | 15 | VC_1251_0 | 15 | 0.9082 | 1 | 0.9082 | 0 | 0 | 0 | 1 | Ref |
| 11233 | MT135337 | Unassigned | Unassigned | Unassigned | 1144_0 | Clustered | 7 | VC_1144_0 | 7 | 0.7076 | 1 | 0.7076 | 0 | 0 | 0 | 1 | Ref |
| 11234 | MT135338 | Unassigned | Unassigned | Unassigned | 1104_0 | Clustered | 154 | VC_1104_0 | 154 | 0.8484 | 1 | 0.8484 | 0 | 0 | 0 | 1 | Ref |
| 11235 | MT135339 | Unassigned | Unassigned | Unassigned | 1233_0 | Clustered | 16 | VC_1233_0 | 16 | 0.8971 | 1 | 0.8971 | 0 | 0 | 0 | 1 | Ref |
| 11236 | MT135340 | Unassigned | Unassigned | Unassigned | 1340_0 | Clustered | 5 | VC_1340_0 | 5 | 1 | 1 | 1 | 0 | 0 | 0 | 1 | Ref |
| 11237 | MT135342 | Unassigned | Unassigned | Unassigned | 1211_0 | Clustered | 25 | VC_1211_0 | 25 | 1 | 1 | 1 | 0 | 0 | 0 | 1 | Ref |
| 11238 | MT135343 | Unassigned | Unassigned | Unassigned | 1312_0 | Clustered | 3 | VC_1312_0 | 3 | 1 | 1 | 1 | 0 | 0 | 0 | 1 | Ref |
| 11239 | MT135344 | Unassigned | Unassigned | Unassigned | 672_0 | Clustered | 12 | VC_672_0 | 12 | 1 | 1 | 1 | 0 | 0 | 0 | 1 | Ref |
| 11240 | MT135345 | Unassigned | Unassigned | Unassigned | 1268_0 | Clustered | 3 | VC_1268_0 | 3 | 0.0807 | 0.6862532 | 0.0554 | 0 | 0 | 0 | 1 | Ref |
| 11241 | MT135346 | Unassigned | Unassigned | Unassigned | 1428_0 | Clustered | 2 | VC_1428_0 | 2 | 1 | 1 | 1 | 0 | 0 | 0 | 1 | Ref |
| 11242 | MT135347 | Unassigned | Unassigned | Unassigned | 1243_0 | Clustered | 52 | VC_1243_0 | 52 | 0.9126 | 0.92708311 | 0.8461 | 0 | 0 | 0 | 1 | Ref |
| 11243 | MT135348 | Unassigned | Unassigned | Unassigned | 1233_0 | Clustered | 16 | VC_1233_0 | 16 | 0.8971 | 1 | 0.8971 | 0 | 0 | 0 | 1 | Ref |
| 11244 | MT135349 | Unassigned | Unassigned | Unassigned | 1315_0 | Clustered | 4 | VC_1315_0 | 4 | 0.8005 | 0.98875328 | 0.7915 | 0 | 0 | 0 | 1 | Ref |
| 11245 | MT135350 | Unassigned | Unassigned | Unassigned | 1331_0 | Clustered | 4 | VC_1331_0 | 4 | 0.7343 | 0.99899459 | 0.7335 | 0 | 0 | 0 | 1 | Ref |
| 11246 | MT135351 | Unassigned | Unassigned | Unassigned | 1122_0 | Clustered | 10 | VC_1122_0 | 10 | 0.9643 | 0.99999697 | 0.9643 | 0 | 0 | 0 | 1 | Ref |
| 11247 | MT135352 | Unassigned | Unassigned | Unassigned | 1243_0 | Clustered | 52 | VC_1243_0 | 52 | 0.9126 | 0.92708311 | 0.8461 | 0 | 0 | 0 | 1 | Ref |
| 11248 | MT135353 | Unassigned | Unassigned | Unassigned | 1305_0 | Clustered | 5 | VC_1305_0 | 5 | 0.7701 | 0.66985041 | 0.5159 | 0 | 0 | 0 | 1 | Ref |
| 11249 | MT135354 | Unassigned | Unassigned | Unassigned | 1260_0 | Clustered | 2 | VC_1260_0 | 2 | 1 | 1 | 1 | 0 | 0 | 0 | 1 | Ref |
| 11250 | MT135355 | Unassigned | Unassigned | Unassigned | 1243_0 | Clustered | 52 | VC_1243_0 | 52 | 0.9126 | 0.92708311 | 0.8461 | 0 | 0 | 0 | 1 | Ref |
| 11251 | MT135357 | Unassigned | Unassigned | Unassigned | 1271_0 | Clustered | 2 | VC_1271_0 | 2 | 1 | 1 | 1 | 0 | 0 | 0 | 1 | Ref |
| 11252 | MT135358 | Unassigned | Unassigned | Unassigned | 1273_0 | Clustered | 6 | VC_1273_0 | 6 | 0.7385 | 0.9999804 | 0.7385 | 0 | 0 | 0 | 1 | Ref |
| 11253 | MT135359 | Unassigned | Unassigned | Unassigned | 1429_0 | Clustered | 8 | VC_1429_0 | 8 | 0.9714 | 0.94517081 | 0.9182 | 0 | 0 | 0 | 1 | Ref |
| 11254 | MT135361 | Unassigned | Unassigned | Unassigned | 1233_0 | Clustered | 16 | VC_1233_0 | 16 | 0.8971 | 1 | 0.8971 | 0 | 0 | 0 | 1 | Ref |
| 11255 | MT135363 | Unassigned | Unassigned | Unassigned | 1315_0 | Clustered | 4 | VC_1315_0 | 4 | 0.8005 | 0.98875328 | 0.7915 | 0 | 0 | 0 | 1 | Ref |
| 11256 | MT135365 | Unassigned | Unassigned | Unassigned | 1289_0 | Clustered | 2 | VC_1289_0 | 2 | 0.4661 | 0.85557782 | 0.3987 | 0 | 0 | 0 | 1 | Ref |
| 11257 | MT135366 | Unassigned | Unassigned | Unassigned | 1243_0 | Clustered | 52 | VC_1243_0 | 52 | 0.9126 | 0.92708311 | 0.8461 | 0 | 0 | 0 | 1 | Ref |
| 11258 | MT135367 | Unassigned | Unassigned | Unassigned | 1253_0 | Clustered | 6 | VC_1253_0 | 6 | 0.6097 | 1 | 0.6097 | 0 | 0 | 0 | 1 | Ref |
| 11259 | MT135368 | Unassigned | Unassigned | Unassigned | 1429_0 | Clustered | 8 | VC_1429_0 | 8 | 0.9714 | 0.94517081 | 0.9182 | 0 | 0 | 0 | 1 | Ref |
| 11260 | MT135369 | Unassigned | Unassigned | Unassigned | 1430_0 | Clustered | 4 | VC_1430_0 | 4 | 1 | 1 | 1 | 0 | 0 | 0 | 1 | Ref |
| 11261 | MT135370 | Unassigned | Unassigned | Unassigned | 1335_0 | Clustered | 7 | VC_1335_0 | 7 | 0.8346 | 0.99999844 | 0.8346 | 0 | 0 | 0 | 1 | Ref |
| 11262 | MT135371 | Unassigned | Unassigned | Unassigned | 1192_0 | Clustered | 3 | VC_1192_0 | 3 | 0.407 | 0.99427327 | 0.4046 | 0 | 0 | 0 | 1 | Ref |
| 11263 | MT135372 | Unassigned | Unassigned | Unassigned | 1322_0 | Clustered | 2 | VC_1322_0 | 2 | 1 | 1 | 1 | 0 | 0 | 0 | 1 | Ref |
| 11264 | MT135373 | Unassigned | Unassigned | Unassigned | 1335_0 | Clustered | 7 | VC_1335_0 | 7 | 0.8346 | 0.99999844 | 0.8346 | 0 | 0 | 0 | 1 | Ref |
| 11265 | MT135374 | Unassigned | Unassigned | Unassigned | 1429_0 | Clustered | 8 | VC_1429_0 | 8 | 0.9714 | 0.94517081 | 0.9182 | 0 | 0 | 0 | 1 | Ref |
| 11266 | MT135375 | Unassigned | Unassigned | Unassigned | 1431_0 | Clustered | 3 | VC_1431_0 | 3 | 1 | 1 | 1 | 0 | 0 | 0 | 1 | Ref |
| 11267 | MT135376 | Unassigned | Unassigned | Unassigned | 1340_0 | Clustered | 5 | VC_1340_0 | 5 | 1 | 1 | 1 | 0 | 0 | 0 | 1 | Ref |
| 11268 | MT135377 | Unassigned | Unassigned | Unassigned | 1273_0 | Clustered | 6 | VC_1273_0 | 6 | 0.7385 | 0.9999804 | 0.7385 | 0 | 0 | 0 | 1 | Ref |
| 11269 | MT135378 | Unassigned | Unassigned | Unassigned | 1429_0 | Clustered | 8 | VC_1429_0 | 8 | 0.9714 | 0.94517081 | 0.9182 | 0 | 0 | 0 | 1 | Ref |
| 11270 | MT135379 | Unassigned | Unassigned | Unassigned | 1277_0 | Clustered | 14 | VC_1277_0 | 14 | 0.8218 | 1 | 0.8218 | 0 | 0 | 0 | 1 | Ref |
| 11271 | MT135380 | Unassigned | Unassigned | Unassigned | 1270_0 | Clustered | 3 | VC_1270_0 | 3 | 0.6808 | 0.98688028 | 0.6719 | 0 | 0 | 0 | 1 | Ref |
| 11272 | MT135381 | Unassigned | Unassigned | Unassigned | 1243_0 | Clustered | 52 | VC_1243_0 | 52 | 0.9126 | 0.92708311 | 0.8461 | 0 | 0 | 0 | 1 | Ref |
| 11273 | MT135382 | Unassigned | Unassigned | Unassigned | 1243_0 | Clustered | 52 | VC_1243_0 | 52 | 0.9126 | 0.92708311 | 0.8461 | 0 | 0 | 0 | 1 | Ref |
| 11274 | MT135383 | Unassigned | Unassigned | Unassigned | 1327_0 | Clustered | 4 | VC_1327_0 | 4 | 1 | 1 | 1 | 0 | 0 | 0 | 1 | Ref |
| 11275 | MT135384 | Unassigned | Unassigned | Unassigned | 1233_0 | Clustered | 16 | VC_1233_0 | 16 | 0.8971 | 1 | 0.8971 | 0 | 0 | 0 | 1 | Ref |
| 11276 | MT135385 | Unassigned | Unassigned | Unassigned | 1288_0 | Clustered | 3 | VC_1288_0 | 3 | 1 | 1 | 1 | 0 | 0 | 0 | 1 | Ref |
| 11277 | MT135387 | Unassigned | Unassigned | Unassigned | 1243_0 | Clustered | 52 | VC_1243_0 | 52 | 0.9126 | 0.92708311 | 0.8461 | 0 | 0 | 0 | 1 | Ref |
| 11278 | MT135388 | Unassigned | Unassigned | Unassigned | 1272_0 | Clustered | 5 | VC_1272_0 | 5 | 1 | 1 | 1 | 0 | 0 | 0 | 1 | Ref |
| 11279 | MT135389 | Unassigned | Unassigned | Unassigned | 1421_0 | Clustered | 4 | VC_1421_0 | 4 | 1 | 1 | 1 | 0 | 0 | 0 | 1 | Ref |
| 11280 | MT135391 | Unassigned | Unassigned | Unassigned | 1277_0 | Clustered | 14 | VC_1277_0 | 14 | 0.8218 | 1 | 0.8218 | 0 | 0 | 0 | 1 | Ref |
| 11281 | MT135392 | Unassigned | Unassigned | Unassigned | 1315_0 | Clustered | 4 | VC_1315_0 | 4 | 0.8005 | 0.98875328 | 0.7915 | 0 | 0 | 0 | 1 | Ref |
| 11282 | MT135393 | Unassigned | Unassigned | Unassigned | 1340_0 | Clustered | 5 | VC_1340_0 | 5 | 1 | 1 | 1 | 0 | 0 | 0 | 1 | Ref |
| 11283 | MT135394 | Unassigned | Unassigned | Unassigned | 1246_0 | Clustered | 2 | VC_1246_0 | 2 | 1 | 1 | 1 | 0 | 0 | 0 | 1 | Ref |
| 11284 | MT135395 | Unassigned | Unassigned | Unassigned | 1428_0 | Clustered | 2 | VC_1428_0 | 2 | 1 | 1 | 1 | 0 | 0 | 0 | 1 | Ref |
| 11285 | MT135396 | Unassigned | Unassigned | Unassigned | 1430_0 | Clustered | 4 | VC_1430_0 | 4 | 1 | 1 | 1 | 0 | 0 | 0 | 1 | Ref |
| 11286 | MT135397 | Unassigned | Unassigned | Unassigned | 1307_0 | Clustered | 2 | VC_1307_0 | 2 | 0.522 | 0.72985431 | 0.381 | 0 | 0 | 0 | 1 | Ref |
| 11287 | MT135399 | Unassigned | Unassigned | Unassigned | 1429_0 | Clustered | 8 | VC_1429_0 | 8 | 0.9714 | 0.94517081 | 0.9182 | 0 | 0 | 0 | 1 | Ref |
| 11288 | MT135400 | Unassigned | Unassigned | Unassigned | 1305_0 | Clustered | 5 | VC_1305_0 | 5 | 0.7701 | 0.66985041 | 0.5159 | 0 | 0 | 0 | 1 | Ref |
| 11289 | MT135401 | Unassigned | Unassigned | Unassigned | 1277_0 | Clustered | 14 | VC_1277_0 | 14 | 0.8218 | 1 | 0.8218 | 0 | 0 | 0 | 1 | Ref |
| 11290 | MT135402 | Unassigned | Unassigned | Unassigned | 1305_0 | Clustered | 5 | VC_1305_0 | 5 | 0.7701 | 0.66985041 | 0.5159 | 0 | 0 | 0 | 1 | Ref |
| 11291 | MT135403 | Unassigned | Unassigned | Unassigned | 1327_0 | Clustered | 4 | VC_1327_0 | 4 | 1 | 1 | 1 | 0 | 0 | 0 | 1 | Ref |
| 11292 | MT135404 | Unassigned | Unassigned | Unassigned | 672_0 | Clustered | 12 | VC_672_0 | 12 | 1 | 1 | 1 | 0 | 0 | 0 | 1 | Ref |
| 11293 | MT135405 | Unassigned | Unassigned | Unassigned | 1069_0 | Clustered | 4 | VC_1069_0 | 4 | 1 | 1 | 1 | 0 | 0 | 0 | 1 | Ref |
| 11294 | MT135406 | Unassigned | Unassigned | Unassigned | 1285_0 | Clustered | 25 | VC_1285_0 | 25 | 1 | 1 | 1 | 0 | 0 | 0 | 1 | Ref |
| 11295 | MT135407 | Unassigned | Unassigned | Unassigned | 1307_0 | Clustered | 2 | VC_1307_0 | 2 | 0.522 | 0.72985431 | 0.381 | 0 | 0 | 0 | 1 | Ref |
| 11296 | MT135408 | Unassigned | Unassigned | Unassigned | 1315_0 | Clustered | 4 | VC_1315_0 | 4 | 0.8005 | 0.98875328 | 0.7915 | 0 | 0 | 0 | 1 | Ref |
| 11297 | MT135409 | Unassigned | Unassigned | Unassigned | 1233_0 | Clustered | 16 | VC_1233_0 | 16 | 0.8971 | 1 | 0.8971 | 0 | 0 | 0 | 1 | Ref |
| 11298 | MT135410 | Unassigned | Unassigned | Unassigned | 1290_0 | Clustered | 2 | VC_1290_0 | 2 | 1 | 1 | 1 | 0 | 0 | 0 | 1 | Ref |
| 11299 | MT135411 | Unassigned | Unassigned | Unassigned | 1302_0 | Clustered | 3 | VC_1302_0 | 3 | 1 | 1 | 1 | 0 | 0 | 0 | 1 | Ref |
| 11300 | MT135412 | Unassigned | Unassigned | Unassigned | 672_0 | Clustered | 12 | VC_672_0 | 12 | 1 | 1 | 1 | 0 | 0 | 0 | 1 | Ref |
| 11301 | MT135413 | Unassigned | Unassigned | Unassigned | 1421_0 | Clustered | 4 | VC_1421_0 | 4 | 1 | 1 | 1 | 0 | 0 | 0 | 1 | Ref |
| 11302 | MT135414 | Unassigned | Unassigned | Unassigned | 1429_0 | Clustered | 8 | VC_1429_0 | 8 | 0.9714 | 0.94517081 | 0.9182 | 0 | 0 | 0 | 1 | Ref |
| 11303 | MT135415 | Unassigned | Unassigned | Unassigned | 1278_0 | Clustered | 3 | VC_1278_0 | 3 | 0.3961 | 0.99705286 | 0.395 | 0 | 0 | 0 | 1 | Ref |
| 11304 | MT135416 | Unassigned | Unassigned | Unassigned | 1312_0 | Clustered | 3 | VC_1312_0 | 3 | 1 | 1 | 1 | 0 | 0 | 0 | 1 | Ref |
| 11305 | MT135418 | Unassigned | Unassigned | Unassigned | 1335_0 | Clustered | 7 | VC_1335_0 | 7 | 0.8346 | 0.99999844 | 0.8346 | 0 | 0 | 0 | 1 | Ref |
| 11306 | MT135419 | Unassigned | Unassigned | Unassigned | 1429_0 | Clustered | 8 | VC_1429_0 | 8 | 0.9714 | 0.94517081 | 0.9182 | 0 | 0 | 0 | 1 | Ref |
| 11307 | MT135420 | Unassigned | Unassigned | Unassigned | 1306_0 | Clustered | 2 | VC_1306_0 | 2 | 1 | 1 | 1 | 0 | 0 | 0 | 1 | Ref |
| 11308 | MT135421 | Unassigned | Unassigned | Unassigned | 1337_0 | Clustered | 2 | VC_1337_0 | 2 | 1 | 1 | 1 | 0 | 0 | 0 | 1 | Ref |
| 11309 | MT135422 | Unassigned | Unassigned | Unassigned | 1192_0 | Clustered | 3 | VC_1192_0 | 3 | 0.407 | 0.99427327 | 0.4046 | 0 | 0 | 0 | 1 | Ref |
| 11310 | MT135423 | Unassigned | Unassigned | Unassigned | 1270_0 | Clustered | 3 | VC_1270_0 | 3 | 0.6808 | 0.98688028 | 0.6719 | 0 | 0 | 0 | 1 | Ref |
| 11311 | MT135424 | Unassigned | Unassigned | Unassigned | 1144_0 | Clustered | 7 | VC_1144_0 | 7 | 0.7076 | 1 | 0.7076 | 0 | 0 | 0 | 1 | Ref |
| 11312 | MT135425 | Unassigned | Unassigned | Unassigned | 1283_0 | Clustered | 26 | VC_1283_0 | 26 | 0.5446 | 1 | 0.5446 | 0 | 0 | 0 | 1 | Ref |
| 11313 | MT135426 | Unassigned | Unassigned | Unassigned | 1273_0 | Clustered | 6 | VC_1273_0 | 6 | 0.7385 | 0.9999804 | 0.7385 | 0 | 0 | 0 | 1 | Ref |
| 11314 | MT135427 | Unassigned | Unassigned | Unassigned | 1273_0 | Clustered | 6 | VC_1273_0 | 6 | 0.7385 | 0.9999804 | 0.7385 | 0 | 0 | 0 | 1 | Ref |
| 11315 | MT135429 | Unassigned | Unassigned | Unassigned | 1341_0 | Clustered | 2 | VC_1341_0 | 2 | 0.7733 | 0.5 | 0.3867 | 0 | 0 | 0 | 1 | Ref |
| 11316 | MT135430 | Unassigned | Unassigned | Unassigned | 1335_0 | Clustered | 7 | VC_1335_0 | 7 | 0.8346 | 0.99999844 | 0.8346 | 0 | 0 | 0 | 1 | Ref |
| 11317 | MT135431 | Unassigned | Unassigned | Unassigned | 1285_0 | Clustered | 25 | VC_1285_0 | 25 | 1 | 1 | 1 | 0 | 0 | 0 | 1 | Ref |
| 11318 | MT135432 | Unassigned | Unassigned | Unassigned | 1192_0 | Clustered | 3 | VC_1192_0 | 3 | 0.407 | 0.99427327 | 0.4046 | 0 | 0 | 0 | 1 | Ref |
| 11319 | MT135433 | Unassigned | Unassigned | Unassigned | 1253_0 | Clustered | 6 | VC_1253_0 | 6 | 0.6097 | 1 | 0.6097 | 0 | 0 | 0 | 1 | Ref |
| 11320 | MT135434 | Unassigned | Unassigned | Unassigned | 1279_0 | Clustered | 4 | VC_1279_0 | 4 | 0.6058 | 0.76972092 | 0.4663 | 0 | 0 | 0 | 1 | Ref |
| 11321 | MT135435 | Unassigned | Unassigned | Unassigned | 1233_0 | Clustered | 16 | VC_1233_0 | 16 | 0.8971 | 1 | 0.8971 | 0 | 0 | 0 | 1 | Ref |
| 11322 | MT135436 | Unassigned | Unassigned | Unassigned | 1427_0 | Clustered | 2 | VC_1427_0 | 2 | 1 | 1 | 1 | 0 | 0 | 0 | 1 | Ref |
| 11323 | MT135437 | Unassigned | Unassigned | Unassigned | 1277_0 | Clustered | 14 | VC_1277_0 | 14 | 0.8218 | 1 | 0.8218 | 0 | 0 | 0 | 1 | Ref |
| 11324 | MT135438 | Unassigned | Unassigned | Unassigned | 1431_0 | Clustered | 3 | VC_1431_0 | 3 | 1 | 1 | 1 | 0 | 0 | 0 | 1 | Ref |
| 11325 | MT135439 | Unassigned | Unassigned | Unassigned | 1252_0 | Clustered | 3 | VC_1252_0 | 3 | 0.3237 | 0.99790393 | 0.323 | 0 | 0 | 0 | 1 | Ref |
| 11326 | MT135440 | Unassigned | Unassigned | Unassigned | 1194_0 | Clustered | 5 | VC_1194_0 | 5 | 0.5236 | 0.99999899 | 0.5236 | 0 | 0 | 0 | 1 | Ref |
| 11327 | MT135441 | Unassigned | Unassigned | Unassigned | 1251_0 | Clustered | 15 | VC_1251_0 | 15 | 0.9082 | 1 | 0.9082 | 0 | 0 | 0 | 1 | Ref |
| 11328 | MT135443 | Unassigned | Unassigned | Unassigned | 1272_0 | Clustered | 5 | VC_1272_0 | 5 | 1 | 1 | 1 | 0 | 0 | 0 | 1 | Ref |
| 11329 | MT135444 | Unassigned | Unassigned | Unassigned | 1243_0 | Clustered | 52 | VC_1243_0 | 52 | 0.9126 | 0.92708311 | 0.8461 | 0 | 0 | 0 | 1 | Ref |
| 11330 | MT135445 | Unassigned | Unassigned | Unassigned | 1277_0 | Clustered | 14 | VC_1277_0 | 14 | 0.8218 | 1 | 0.8218 | 0 | 0 | 0 | 1 | Ref |
| 11331 | MT135446 | Unassigned | Unassigned | Unassigned | 1194_0 | Clustered | 5 | VC_1194_0 | 5 | 0.5236 | 0.99999899 | 0.5236 | 0 | 0 | 0 | 1 | Ref |
| 11332 | MT135447 | Unassigned | Unassigned | Unassigned | 1293_0 | Clustered | 4 | VC_1293_0 | 4 | 0.7847 | 0.99289298 | 0.7791 | 0 | 0 | 0 | 1 | Ref |
| 11333 | MT135451 | Unassigned | Unassigned | Unassigned | 1279_0 | Clustered | 4 | VC_1279_0 | 4 | 0.6058 | 0.76972092 | 0.4663 | 0 | 0 | 0 | 1 | Ref |
| 11334 | MT135453 | Unassigned | Unassigned | Unassigned | 1431_0 | Clustered | 3 | VC_1431_0 | 3 | 1 | 1 | 1 | 0 | 0 | 0 | 1 | Ref |
| 11335 | MT135454 | Unassigned | Unassigned | Unassigned | 1430_0 | Clustered | 4 | VC_1430_0 | 4 | 1 | 1 | 1 | 0 | 0 | 0 | 1 | Ref |
| 11336 | MT135455 | Unassigned | Unassigned | Unassigned | 1335_0 | Clustered | 7 | VC_1335_0 | 7 | 0.8346 | 0.99999844 | 0.8346 | 0 | 0 | 0 | 1 | Ref |
| 11337 | MT135456 | Unassigned | Unassigned | Unassigned | 1327_0 | Clustered | 4 | VC_1327_0 | 4 | 1 | 1 | 1 | 0 | 0 | 0 | 1 | Ref |
| 11338 | MT135457 | Unassigned | Unassigned | Unassigned | 1331_0 | Clustered | 4 | VC_1331_0 | 4 | 0.7343 | 0.99899459 | 0.7335 | 0 | 0 | 0 | 1 | Ref |

| | | | | | | | | | | | | | | | | | |
|---|---|---|---|---|---|---|---|---|---|---|---|---|---|---|---|---|---|
| 11339 | MT135458 | Unassigned | Unassigned | Unassigned | 1285_0 | Clustered | 25 | VC_1285_0 | 25 | 1 | 1 | 1 | 0 | 0 | 0 | 1 | Ref |
| 11340 | MT135459 | Unassigned | Unassigned | Unassigned | 1335_0 | Clustered | 7 | VC_1335_0 | 7 | 0.8346 | 0.99999844 | 0.8346 | 0 | 0 | 0 | 1 | Ref |
| 11341 | MT135460 | Unassigned | Unassigned | Unassigned | 1430_0 | Clustered | 4 | VC_1430_0 | 4 | 1 | 1 | 1 | 0 | 0 | 0 | 1 | Ref |
| 11342 | MT135461 | Unassigned | Unassigned | Unassigned | 1298_0 | Clustered | 3 | VC_1298_0 | 3 | 0.4889 | 0.98167473 | 0.4799 | 0 | 0 | 0 | 1 | Ref |
| 11343 | MT135462 | Unassigned | Unassigned | Unassigned | 1272_0 | Clustered | 5 | VC_1272_0 | 5 | 1 | 1 | 1 | 0 | 0 | 0 | 1 | Ref |
| 11344 | MT135463 | Unassigned | Unassigned | Unassigned | 1429_0 | Clustered | 8 | VC_1429_0 | 8 | 0.9714 | 0.94517081 | 0.9182 | 0 | 0 | 0 | 1 | Ref |
| 11345 | MT136606 | Unassigned | Unassigned | Unassigned | 829_0 | Clustered | 5 | VC_829_0 | 5 | 0.8821 | 0.99999832 | 0.8821 | 0 | 0 | 0 | 1 | Ref |
| 11346 | MT150133 | Unassigned | Unassigned | Unassigned | 290_5 | Clustered | 14 | VC_290_5 | 14 | 0.1422 | 1 | 0.1422 | 0 | 0 | 0 | 0.9231 | Ref |
| 11347 | MT151604 | Unassigned | Unassigned | Unassigned | 85_12 | Clustered/Singleton | 1 | VC_85_12 | 1 | 0 | 1 | 0 | 0 | 0 | 0 | 1 | Ref |
| 11348 | MT152150 | Unassigned | Unassigned | Unassigned | 631_0 | Clustered | 3 | VC_631_0 | 3 | 0.1213 | 0.99819199 | 0.1211 | 0 | 0 | 0 | 1 | Ref |
| 11349 | MT152698 | Unassigned | Unassigned | Unassigned | 1002_0 | Clustered | 2 | VC_1002_0 | 2 | 0.1809 | 0.72090768 | 0.1304 | 0 | 0 | 0 | 1 | Ref |
| 11350 | MT157285 | Unassigned | Unassigned | Unassigned | 668_0 | Clustered | 25 | VC_668_0 | 25 | 0.5539 | 1 | 0.5539 | 0 | 0 | 0 | 0.96 | Ref |
| 11351 | MT161381 | Unassigned | Unassigned | Unassigned | 125_0 | Clustered | 17 | VC_125_0 | 17 | 0.9244 | 1 | 0.9244 | 0 | 0 | 0 | 1 | Ref |
| 11352 | MT161382 | Unassigned | Unassigned | Unassigned | 125_0 | Clustered | 17 | VC_125_0 | 17 | 0.9244 | 1 | 0.9244 | 0 | 0 | 0 | 1 | Ref |
| 11353 | MT161383 | Unassigned | Unassigned | Unassigned | 125_0 | Clustered | 17 | VC_125_0 | 17 | 0.9244 | 1 | 0.9244 | 0 | 0 | 0 | 1 | Ref |
| 11354 | MT161384 | Unassigned | Unassigned | Unassigned | 125_0 | Clustered | 17 | VC_125_0 | 17 | 0.9244 | 1 | 0.9244 | 0 | 0 | 0 | 1 | Ref |
| 11355 | MT162466 | Unassigned | Unassigned | Unassigned | 435_0 | Clustered | 2 | VC_435_0 | 2 | 0.0423 | 0.95727719 | 0.0405 | 0 | 0 | 0 | 1 | Ref |
| 11356 | MT162467 | Unassigned | Unassigned | Unassigned | 435_0 | Clustered | 2 | VC_435_0 | 2 | 0.0423 | 0.95727719 | 0.0405 | 0 | 0 | 0 | 1 | Ref |
| 11357 | MT176424 | Unassigned | Unassigned | Unassigned | 283_27 | Clustered/Singleton | 1 | VC_283_27 | 1 | 0 | 1 | 0 | 0 | 0 | 0 | 1 | Ref |
| 11358 | MT176425 | Unassigned | Unassigned | Unassigned | 283_49 | Clustered/Singleton | 1 | VC_283_49 | 1 | 0 | 1 | 0 | 0 | 0 | 0 | 1 | Ref |
| 11359 | MT176426 | Unassigned | Unassigned | Unassigned | 283_28 | Clustered/Singleton | 1 | VC_283_28 | 1 | 0 | 1 | 0 | 0 | 0 | 0 | 1 | Ref |
| 11360 | MT178275 | Unassigned | Unassigned | Unassigned | 582_3 | Clustered | 12 | VC_582_3 | 12 | 0.2357 | 1 | 0.2357 | 0 | 0 | 0 | 0.8636 | Ref |
| 11361 | MT178448 | Unassigned | Unassigned | Unassigned | 837_0 | Clustered | 29 | VC_837_0 | 29 | 0.9955 | 1 | 0.9955 | 0 | 0 | 0 | 1 | Ref |
| 11362 | MT178766 | Unassigned | Unassigned | Unassigned | 739_0 | Clustered | 26 | VC_739_0 | 26 | 0.9571 | 1 | 0.9571 | 0 | 0 | 0 | 1 | Ref |
| 11363 | MT178767 | Unassigned | Unassigned | Unassigned | 739_0 | Clustered | 26 | VC_739_0 | 26 | 0.9571 | 1 | 0.9571 | 0 | 0 | 0 | 1 | Ref |
| 11364 | MT179807 | Unassigned | Unassigned | Unassigned | 283_161 | Clustered/Singleton | 1 | VC_283_161 | 1 | 0 | 1 | 0 | 0 | 0 | 0 | 1 | Ref |
| 11365 | MT188223 | Unassigned | Unassigned | Unassigned | 700_0 | Clustered | 10 | VC_700_0 | 10 | 0.8076 | 1 | 0.8076 | 0 | 0 | 0 | 1 | Ref |
| 11366 | MT188662 | Unassigned | Unassigned | Unassigned | 17_0 | Clustered | 9 | VC_17_0 | 9 | 0.6969 | 1 | 0.6969 | 0 | 0 | 0 | 1 | Ref |
| 11367 | MT188663 | Unassigned | Unassigned | Unassigned | 17_0 | Clustered | 9 | VC_17_0 | 9 | 0.6969 | 1 | 0.6969 | 0 | 0 | 0 | 1 | Ref |
| 11368 | MT188664 | Unassigned | Unassigned | Unassigned | 17_0 | Clustered | 9 | VC_17_0 | 9 | 0.6969 | 1 | 0.6969 | 0 | 0 | 0 | 1 | Ref |
| 11369 | MT188665 | Unassigned | Unassigned | Unassigned | 17_0 | Clustered | 9 | VC_17_0 | 9 | 0.6969 | 1 | 0.6969 | 0 | 0 | 0 | 1 | Ref |
| 11370 | MT188666 | Unassigned | Unassigned | Unassigned | 17_0 | Clustered | 9 | VC_17_0 | 9 | 0.6969 | 1 | 0.6969 | 0 | 0 | 0 | 1 | Ref |
| 11371 | MT193276 | Unassigned | Unassigned | Unassigned | 647_0 | Clustered | 11 | VC_647_0 | 11 | 0.598 | 1 | 0.598 | 0 | 0 | 0 | 1 | Ref |
| 11372 | MT193890 | Unassigned | Unassigned | Unassigned | 17_0 | Clustered | 9 | VC_17_0 | 9 | 0.6969 | 1 | 0.6969 | 0 | 0 | 0 | 1 | Ref |
| 11373 | MT197175 | Unassigned | Unassigned | Unassigned | 111_0 | Clustered | 91 | VC_111_0 | 91 | 0.9587 | 1 | 0.9587 | 0 | 0 | 0 | 1 | Ref |
| 11374 | MT197176 | Unassigned | Unassigned | Unassigned | 111_0 | Clustered | 91 | VC_111_0 | 91 | 0.9587 | 1 | 0.9587 | 0 | 0 | 0 | 1 | Ref |
| 11375 | MT210154 | Unassigned | Unassigned | Unassigned | 934_0 | Clustered | 2 | VC_934_0 | 2 | 0.8613 | 0.92499224 | 0.7967 | 0 | 0 | 0 | 1 | Ref |
| 11376 | MT215167 | Unassigned | Unassigned | Unassigned | 82_0 | Clustered | 24 | VC_82_0 | 24 | 0.7351 | 1 | 0.7351 | 0 | 0 | 0 | 1 | Ref |
| 11377 | MT226657 | Unassigned | Unassigned | Unassigned | 149_8 | Clustered | 66 | VC_149_8 | 66 | 0.4565 | 1 | 0.4565 | 0 | 0 | 0 | 0.8182 | Ref |
| 11378 | MT227924 | Unassigned | Unassigned | Unassigned | 354_0 | Clustered | 4 | VC_354_0 | 4 | 0.1749 | 0.99998684 | 0.1749 | 0 | 0 | 0 | 0.8333 | Ref |
| 11379 | MT227925 | Unassigned | Unassigned | Unassigned | 989_0 | Clustered | 2 | VC_989_0 | 2 | 0.1205 | 0.95251219 | 0.1148 | 0 | 0 | 0 | 1 | Ref |
| 11380 | MT230534 | Unassigned | Unassigned | Unassigned | 792_0 | Clustered | 35 | VC_792_0 | 35 | 0.9828 | 1 | 0.9828 | 0 | 0 | 0 | 1 | Ref |
| 11381 | MT233524 | Unassigned | Unassigned | Unassigned | 190_39 | Clustered/Singleton | 1 | VC_190_39 | 1 | 0 | 1 | 0 | 0 | 0 | 0 | 1 | Ref |
| 11382 | MT234338 | Unassigned | Unassigned | Unassigned | 786_0 | Clustered | 3 | VC_786_0 | 3 | 0.0814 | 0.99854097 | 0.0813 | 0 | 0 | 0 | 1 | Ref |
| 11383 | MT234339 | Unassigned | Unassigned | Unassigned | 786_0 | Clustered | 3 | VC_786_0 | 3 | 0.0814 | 0.99854097 | 0.0813 | 0 | 0 | 0 | 1 | Ref |
| 11384 | MT234340 | Unassigned | Unassigned | Unassigned | 786_0 | Clustered | 3 | VC_786_0 | 3 | 0.0814 | 0.99854097 | 0.0813 | 0 | 0 | 0 | 1 | Ref |
| 11385 | MT234341 | Unassigned | Unassigned | Unassigned | 969_0 | Clustered | 3 | VC_969_0 | 3 | 0.8314 | 0.99762851 | 0.8295 | 0 | 0 | 0 | 1 | Ref |
| 11386 | MT234342 | Unassigned | Unassigned | Unassigned | 1052_0 | Clustered | 2 | VC_1052_0 | 2 | 0.0778 | 0.94600647 | 0.0736 | 0 | 0 | 0 | 1 | Ref |
| 11387 | MT234343 | Unassigned | Unassigned | Unassigned | 1052_0 | Clustered | 2 | VC_1052_0 | 2 | 0.0778 | 0.94600647 | 0.0736 | 0 | 0 | 0 | 1 | Ref |
| 11388 | MT234670 | Unassigned | Unassigned | Unassigned | 1433_0 | Clustered | 2 | VC_1433_0 | 2 | 1 | 1 | 1 | 0 | 0 | 0 | 1 | Ref |
| 11389 | MT239398 | Unassigned | Unassigned | Unassigned | 228_0 | Clustered | 60 | VC_228_0 | 60 | 0.942 | 1 | 0.942 | 0 | 0 | 0 | 1 | Ref |
| 11390 | MT241605 | Unassigned | Unassigned | Unassigned | 171_0 | Clustered | 8 | VC_171_0 | 8 | 0.5884 | 1 | 0.5884 | 0 | 0 | 0 | 1 | Ref |
| 11391 | MT241607 | Unassigned | Unassigned | Unassigned | 171_0 | Clustered | 8 | VC_171_0 | 8 | 0.5884 | 1 | 0.5884 | 0 | 0 | 0 | 1 | Ref |
| 11392 | MT251347 | Unassigned | Unassigned | Unassigned | 359_0 | Clustered | 7 | VC_359_0 | 7 | 0.3703 | 1 | 0.3703 | 0 | 0 | 0 | 1 | Ref |
| 11393 | MT251348 | Unassigned | Unassigned | Unassigned | 359_0 | Clustered | 7 | VC_359_0 | 7 | 0.3703 | 1 | 0.3703 | 0 | 0 | 0 | 1 | Ref |
| 11394 | MT254578 | Unassigned | Unassigned | Unassigned | 475_2 | Clustered | 3 | VC_475_2 | 3 | 0.0585 | 0.98883022 | 0.0578 | 0 | 0 | 0 | 1 | Ref |
| 11395 | MT261384 | Unassigned | Unassigned | Unassigned | 476_0 | Clustered | 19 | VC_476_0 | 19 | 0.7814 | 1 | 0.7814 | 0 | 0 | 0 | 1 | Ref |
| 11396 | MT263719 | Unassigned | Unassigned | Unassigned | 699_0 | Clustered | 71 | VC_699_0 | 71 | 0.9352 | 1 | 0.9352 | 0 | 0 | 0 | 1 | Ref |
| 11397 | MT270409 | Unassigned | Unassigned | Unassigned | 977_1 | Clustered | 2 | VC_977_1 | 2 | 0.079 | 0.93977186 | 0.0743 | 0 | 0 | 0 | 1 | Ref |
| 11398 | MT276994 | Unassigned | Unassigned | Unassigned | 1189_0 | Clustered | 9 | VC_1189_0 | 9 | 0.968 | 1 | 0.968 | 0 | 0 | 0 | 1 | Ref |
| 11399 | MT310850 | Unassigned | Unassigned | Unassigned | 799_1 | Clustered | 27 | VC_799_1 | 27 | 0.8611 | 1 | 0.8611 | 0 | 0 | 0 | 1 | Ref |
| 11400 | MT310851 | Unassigned | Unassigned | Unassigned | 1240_0 | Clustered | 82 | VC_1240_0 | 82 | 0.7945 | 1 | 0.7945 | 0 | 0 | 0 | 1 | Ref |
| 11401 | MT310852 | Unassigned | Unassigned | Unassigned | 872_0 | Clustered | 31 | VC_872_0 | 31 | 0.8151 | 1 | 0.8151 | 0 | 0 | 0 | 0.8194 | Ref |
| 11402 | MT310853 | Unassigned | Unassigned | Unassigned | 1234_0 | Clustered | 44 | VC_1234_0 | 44 | 1 | 1 | 1 | 0 | 0 | 0 | 1 | Ref |
| 11403 | MT310854 | Unassigned | Unassigned | Unassigned | 872_0 | Clustered | 31 | VC_872_0 | 31 | 0.8151 | 1 | 0.8151 | 0 | 0 | 0 | 0.8194 | Ref |
| 11404 | MT310855 | Unassigned | Unassigned | Unassigned | 1162_0 | Clustered | 10 | VC_1162_0 | 10 | 0.5715 | 1 | 0.5715 | 0 | 0 | 0 | 1 | Ref |
| 11405 | MT310856 | Unassigned | Unassigned | Unassigned | 1240_0 | Clustered | 82 | VC_1240_0 | 82 | 0.7945 | 1 | 0.7945 | 0 | 0 | 0 | 1 | Ref |
| 11406 | MT310858 | Unassigned | Unassigned | Unassigned | 216_7 | Clustered/Singleton | 1 | VC_216_7 | 1 | 0 | 1 | 0 | 0 | 0 | 0 | 1 | Ref |
| 11407 | MT310859 | Unassigned | Unassigned | Unassigned | 237_3 | Clustered | 8 | VC_237_3 | 8 | 0.1092 | 1 | 0.1092 | 0 | 0 | 0 | 1 | Ref |
| 11408 | MT310860 | Unassigned | Unassigned | Unassigned | 237_3 | Clustered | 8 | VC_237_3 | 8 | 0.1092 | 1 | 0.1092 | 0 | 0 | 0 | 1 | Ref |
| 11409 | MT310861 | Unassigned | Unassigned | Unassigned | 1240_0 | Clustered | 82 | VC_1240_0 | 82 | 0.7945 | 1 | 0.7945 | 0 | 0 | 0 | 1 | Ref |
| 11410 | MT310863 | Unassigned | Unassigned | Unassigned | 981_1 | Clustered | 26 | VC_981_1 | 26 | 0.7009 | 1 | 0.7009 | 0 | 0 | 0 | 1 | Ref |
| 11411 | MT310864 | Unassigned | Unassigned | Unassigned | 1344_0 | Clustered | 78 | VC_1344_0 | 78 | 0.9732 | 1 | 0.9732 | 0 | 0 | 0 | 1 | Ref |
| 11412 | MT310865 | Unassigned | Unassigned | Unassigned | 1071_2 | Clustered | 5 | VC_1071_2 | 5 | 0.0534 | 0.99997362 | 0.0534 | 0 | 0 | 0 | 0.9 | Ref |
| 11413 | MT310866 | Unassigned | Unassigned | Unassigned | 1240_0 | Clustered | 82 | VC_1240_0 | 82 | 0.7945 | 1 | 0.7945 | 0 | 0 | 0 | 1 | Ref |
| 11414 | MT310867 | Unassigned | Unassigned | Unassigned | 371_12 | Clustered | 4 | VC_371_12 | 4 | 0.0126 | 0.99998489 | 0.0126 | 0 | 0 | 0 | 0.8333 | Ref |
| 11415 | MT310868 | Unassigned | Unassigned | Unassigned | 371_11 | Clustered | 31 | VC_371_11 | 31 | 0.0927 | 1 | 0.0927 | 0 | 0 | 0 | 0.7011 | Ref |
| 11416 | MT310869 | Unassigned | Unassigned | Unassigned | 1360_0 | Clustered | 31 | VC_1360_0 | 31 | 0.8837 | 1 | 0.8837 | 0 | 0 | 0 | 1 | Ref |
| 11417 | MT310870 | Unassigned | Unassigned | Unassigned | 372_0 | Clustered | 24 | VC_372_0 | 24 | 0.5539 | 1 | 0.5539 | 0 | 0 | 0 | 1 | Ref |
| 11418 | MT310871 | Unassigned | Unassigned | Unassigned | 509_0 | Clustered | 37 | VC_509_0 | 37 | 0.9689 | 1 | 0.9689 | 0 | 0 | 0 | 1 | Ref |
| 11419 | MT310872 | Unassigned | Unassigned | Unassigned | 1178_0 | Clustered | 24 | VC_1178_0 | 24 | 0.9524 | 1 | 0.9524 | 0 | 0 | 0 | 1 | Ref |
| 11420 | MT310873 | Unassigned | Unassigned | Unassigned | 1240_0 | Clustered | 82 | VC_1240_0 | 82 | 0.7945 | 1 | 0.7945 | 0 | 0 | 0 | 1 | Ref |
| 11421 | MT310874 | Unassigned | Unassigned | Unassigned | 857_0 | Clustered | 11 | VC_857_0 | 11 | 0.9712 | 1 | 0.9712 | 0 | 0 | 0 | 1 | Ref |
| 11422 | MT310875 | Unassigned | Unassigned | Unassigned | 1360_0 | Clustered | 31 | VC_1360_0 | 31 | 0.8837 | 1 | 0.8837 | 0 | 0 | 0 | 1 | Ref |
| 11423 | MT310876 | Unassigned | Unassigned | Unassigned | 215_1 | Clustered | 19 | VC_215_1 | 19 | 0.4188 | 1 | 0.4188 | 0 | 0 | 0 | 1 | Ref |
| 11424 | MT310877 | Unassigned | Unassigned | Unassigned | 216_6 | Clustered | 42 | VC_216_6 | 42 | 0.473 | 1 | 0.473 | 0 | 0 | 0 | 0.8351 | Ref |
| 11425 | MT310878 | Unassigned | Unassigned | Unassigned | 513_0 | Clustered | 32 | VC_513_0 | 32 | 0.6881 | 1 | 0.6881 | 0 | 0 | 0 | 1 | Ref |
| 11426 | MT310879 | Unassigned | Unassigned | Unassigned | 513_0 | Clustered | 32 | VC_513_0 | 32 | 0.6881 | 1 | 0.6881 | 0 | 0 | 0 | 1 | Ref |
| 11427 | MT310880 | Unassigned | Unassigned | Unassigned | 513_0 | Clustered | 32 | VC_513_0 | 32 | 0.6881 | 1 | 0.6881 | 0 | 0 | 0 | 1 | Ref |
| 11428 | MT310881 | Unassigned | Unassigned | Unassigned | 513_0 | Clustered | 32 | VC_513_0 | 32 | 0.6881 | 1 | 0.6881 | 0 | 0 | 0 | 1 | Ref |
| 11429 | MT310882 | Unassigned | Unassigned | Unassigned | 234_0 | Clustered | 21 | VC_234_0 | 21 | 0.8355 | 1 | 0.8355 | 0 | 0 | 0 | 1 | Ref |
| 11430 | MT310883 | Unassigned | Unassigned | Unassigned | 215_1 | Clustered | 19 | VC_215_1 | 19 | 0.4188 | 1 | 0.4188 | 0 | 0 | 0 | 1 | Ref |
| 11431 | MT310884 | Unassigned | Unassigned | Unassigned | 215_1 | Clustered | 19 | VC_215_1 | 19 | 0.4188 | 1 | 0.4188 | 0 | 0 | 0 | 1 | Ref |
| 11432 | MT310885 | Unassigned | Unassigned | Unassigned | 215_1 | Clustered | 19 | VC_215_1 | 19 | 0.4188 | 1 | 0.4188 | 0 | 0 | 0 | 1 | Ref |
| 11433 | MT310886 | Unassigned | Unassigned | Unassigned | 216_6 | Clustered | 42 | VC_216_6 | 42 | 0.473 | 1 | 0.473 | 0 | 0 | 0 | 0.8351 | Ref |
| 11434 | MT310887 | Unassigned | Unassigned | Unassigned | 1360_0 | Clustered | 31 | VC_1360_0 | 31 | 0.8837 | 1 | 0.8837 | 0 | 0 | 0 | 1 | Ref |
| 11435 | MT310888 | Unassigned | Unassigned | Unassigned | 1360_0 | Clustered | 31 | VC_1360_0 | 31 | 0.8837 | 1 | 0.8837 | 0 | 0 | 0 | 1 | Ref |
| 11436 | MT310889 | Unassigned | Unassigned | Unassigned | 1360_0 | Clustered | 31 | VC_1360_0 | 31 | 0.8837 | 1 | 0.8837 | 0 | 0 | 0 | 1 | Ref |
| 11437 | MT310890 | Unassigned | Unassigned | Unassigned | 1360_0 | Clustered | 31 | VC_1360_0 | 31 | 0.8837 | 1 | 0.8837 | 0 | 0 | 0 | 1 | Ref |
| 11438 | MT310891 | Unassigned | Unassigned | Unassigned | 1360_0 | Clustered | 31 | VC_1360_0 | 31 | 0.8837 | 1 | 0.8837 | 0 | 0 | 0 | 1 | Ref |
| 11439 | MT310892 | Unassigned | Unassigned | Unassigned | 509_0 | Clustered | 37 | VC_509_0 | 37 | 0.9689 | 1 | 0.9689 | 0 | 0 | 0 | 1 | Ref |
| 11440 | MT310893 | Unassigned | Unassigned | Unassigned | 520_38 | Clustered | 6 | VC_520_38 | 6 | 0.0519 | 1 | 0.0519 | 0 | 0 | 0 | 0.8667 | Ref |
| 11441 | MT310895 | Unassigned | Unassigned | Unassigned | 1073_0 | Clustered | 24 | VC_1073_0 | 24 | 0.8388 | 1 | 0.8388 | 0 | 0 | 0 | 1 | Ref |
| 11442 | MT310896 | Unassigned | Unassigned | Unassigned | 1234_0 | Clustered | 44 | VC_1234_0 | 44 | 1 | 1 | 1 | 0 | 0 | 0 | 1 | Ref |
| 11443 | MT310897 | Unassigned | Unassigned | Unassigned | 325_5 | Clustered | 11 | VC_325_5 | 11 | 0.047 | 1 | 0.047 | 0 | 0 | 0 | 0.7636 | Ref |
| 11444 | MT310898 | Unassigned | Unassigned | Unassigned | 1158_1 | Clustered | 2 | VC_1158_1 | 2 | 0.1372 | 0.89820314 | 0.1233 | 0 | 0 | 0 | 1 | Ref |
| 11445 | MT311645 | Unassigned | Unassigned | Unassigned | 190_23 | Clustered | 10 | VC_190_23 | 10 | 0.0704 | 1 | 0.0704 | 0 | 0 | 0 | 0.8444 | Ref |
| 11446 | MT316458 | Unassigned | Unassigned | Unassigned | 1344_0 | Clustered | 78 | VC_1344_0 | 78 | 0.9732 | 1 | 0.9732 | 0 | 0 | 0 | 1 | Ref |
| 11447 | MT316459 | Unassigned | Unassigned | Unassigned | 1344_0 | Clustered | 78 | VC_1344_0 | 78 | 0.9732 | 1 | 0.9732 | 0 | 0 | 0 | 1 | Ref |
| 11448 | MT316460 | Unassigned | Unassigned | Unassigned | 371_11 | Clustered | 31 | VC_371_11 | 31 | 0.0927 | 1 | 0.0927 | 0 | 0 | 0 | 0.7011 | Ref |
| 11449 | MT316461 | Unassigned | Unassigned | Unassigned | 728_0 | Clustered | 3 | VC_728_0 | 3 | 0.4017 | 0.99850259 | 0.4011 | 0 | 0 | 0 | 1 | Ref |
| 11450 | MT316462 | Unassigned | Unassigned | Unassigned | 1344_0 | Clustered | 78 | VC_1344_0 | 78 | 0.9732 | 1 | 0.9732 | 0 | 0 | 0 | 1 | Ref |
| 11451 | MT316463 | Unassigned | Unassigned | Unassigned | 371_11 | Clustered | 31 | VC_371_11 | 31 | 0.0927 | 1 | 0.0927 | 0 | 0 | 0 | 0.7011 | Ref |
| 11452 | MT322327 | Unassigned | Unassigned | Unassigned | 329_17 | Clustered | 4 | VC_329_17 | 4 | 0.0163 | 0.99996247 | 0.0163 | 0 | 0 | 0 | 1 | Ref |
| 11453 | MT325768 | Unassigned | Unassigned | Unassigned | 211_3 | Clustered/Singleton | 1 | VC_211_3 | 1 | 0 | 1 | 0 | 0 | 0 | 0 | 1 | Ref |
| 11454 | MT331608 | Unassigned | Unassigned | Unassigned | 835_0 | Clustered | 67 | VC_835_0 | 67 | 0.9624 | 1 | 0.9624 | 0 | 0 | 0 | 1 | Ref |
| 11455 | MT334653 | Unassigned | Unassigned | Unassigned | 669_1 | Clustered | 10 | VC_669_1 | 10 | 0.1268 | 1 | 0.1268 | 0 | 0 | 0 | 0.9778 | Ref |
| 11456 | MT338525 | Unassigned | Unassigned | Unassigned | 149_8 | Clustered | 66 | VC_149_8 | 66 | 0.4565 | 1 | 0.4565 | 0 | 0 | 0 | 0.8182 | Ref |
| 11457 | MT341500 | Unassigned | Unassigned | Unassigned | 669_0 | Clustered | 21 | VC_669_0 | 21 | 0.3623 | 1 | 0.3623 | 0 | 0 | 0 | 1 | Ref |
| 11458 | MT344103 | Unassigned | Unassigned | Unassigned | 1188_0 | Clustered | 3 | VC_1188_0 | 3 | 0.9001 | 0.9944138 | 0.8951 | 0 | 0 | 0 | 1 | Ref |
| 11459 | MT344104 | Unassigned | Unassigned | Unassigned | 1188_0 | Clustered | 3 | VC_1188_0 | 3 | 0.9001 | 0.9944138 | 0.8951 | 0 | 0 | 0 | 1 | Ref |
| 11460 | MT344105 | Unassigned | Unassigned | Unassigned | 1188_0 | Clustered | 3 | VC_1188_0 | 3 | 0.9001 | 0.9944138 | 0.8951 | 0 | 0 | 0 | 1 | Ref |
| 11461 | MT349888 | Unassigned | Unassigned | Unassigned | 79_0 | Clustered | 64 | VC_79_0 | 64 | 0.4185 | 1 | 0.4185 | 0 | 0 | 0 | 0.7649 | Ref |
| 11462 | MT350292 | Unassigned | Unassigned | Unassigned | 451_0 | Clustered | 80 | VC_451_0 | 80 | 0.9991 | 1 | 0.9991 | 0 | 0 | 0 | 1 | Ref |
| 11463 | MT350293 | Unassigned | Unassigned | Unassigned | 392_0 | Clustered | 126 | VC_392_0 | 126 | 0.9621 | 1 | 0.9621 | 0 | 0 | 0 | 1 | Ref |
| 11464 | MT354569 | Unassigned | Unassigned | Unassigned | 725_0 | Clustered | 4 | VC_725_0 | 4 | 0.4268 | 0.99997355 | 0.4268 | 0 | 0 | 0 | 1 | Ref |
| 11465 | MT354570 | Unassigned | Unassigned | Unassigned | 155_0 | Clustered | 14 | VC_155_0 | 14 | 0.6095 | 1 | 0.6095 | 0 | 0 | 0 | 1 | Ref |
| 11466 | MT360680 | Unassigned | Unassigned | Unassigned | 111_0 | Clustered | 91 | VC_111_0 | 91 | 0.9587 | 1 | 0.9587 | 0 | 0 | 0 | 1 | Ref |
| 11467 | MT360681 | Unassigned | Unassigned | Unassigned | 941_0 | Clustered | 40 | VC_941_0 | 40 | 0.6903 | 1 | 0.6903 | 0 | 0 | 0 | 1 | Ref |
| 11468 | MT361768 | Unassigned | Unassigned | Unassigned | 162_0 | Clustered | 18 | VC_162_0 | 18 | 0.7403 | 1 | 0.7403 | 0 | 0 | 0 | 0.8627 | Ref |
| 11469 | MT361972 | Unassigned | Unassigned | Unassigned | 696_1 | Clustered | 8 | VC_696_1 | 8 | 0.5452 | 1 | 0.5452 | 0 | 0 | 0 | 0.9286 | Ref |
| 11470 | MT366568 | Unassigned | Unassigned | Unassigned | 8_0 | Clustered | 58 | VC_8_0 | 58 | 0.8306 | 1 | 0.8306 | 0 | 0 | 0 | 0.9946 | Ref |
| 11471 | MT366580 | Unassigned | Unassigned | Unassigned | 231_0 | Clustered | 3 | VC_231_0 | 3 | 0.236 | 0.99824503 | 0.2356 | 0 | 0 | 0 | 1 | Ref |
| 11472 | MT366760 | Unassigned | Unassigned | Unassigned | 65_0 | Clustered | 2 | VC_65_0 | 2 | 0.1825 | 0.9555882 | 0.1744 | 0 | 0 | 0 | 1 | Ref |
| 11473 | MT366761 | Unassigned | Unassigned | Unassigned | 65_0 | Clustered | 2 | VC_65_0 | 2 | 0.1825 | 0.9555882 | 0.1744 | 0 | 0 | 0 | 1 | Ref |
| 11474 | MT366762 | Unassigned | Unassigned | Unassigned | 136_0 | Clustered | 2 | VC_136_0 | 2 | 0.0412 | 0.95741268 | 0.0394 | 0 | 0 | 0 | 1 | Ref |
| 11475 | MT366931 | Unassigned | Unassigned | Unassigned | 450_0 | Clustered | 16 | VC_450_0 | 16 | 0.9564 | 1 | 0.9564 | 0 | 0 | 0 | 1 | Ref |
| 11476 | MT366932 | Unassigned | Unassigned | Unassigned | 450_0 | Clustered | 16 | VC_450_0 | 16 | 0.9564 | 1 | 0.9564 | 0 | 0 | 0 | 1 | Ref |
| 11477 | MT366933 | Unassigned | Unassigned | Unassigned | 450_0 | Clustered | 16 | VC_450_0 | 16 | 0.9564 | 1 | 0.9564 | 0 | 0 | 0 | 1 | Ref |
| 11478 | MT366934 | Unassigned | Unassigned | Unassigned | 450_0 | Clustered | 16 | VC_450_0 | 16 | 0.9564 | 1 | 0.9564 | 0 | 0 | 0 | 1 | Ref |
| 11479 | MT366935 | Unassigned | Unassigned | Unassigned | 450_0 | Clustered | 16 | VC_450_0 | 16 | 0.9564 | 1 | 0.9564 | 0 | 0 | 0 | 1 | Ref |
| 11480 | MT366936 | Unassigned | Unassigned | Unassigned | 450_0 | Clustered | 16 | VC_450_0 | 16 | 0.9564 | 1 | 0.9564 | 0 | 0 | 0 | 1 | Ref |
| 11481 | MT366937 | Unassigned | Unassigned | Unassigned | 450_0 | Clustered | 16 | VC_450_0 | 16 | 0.9564 | 1 | 0.9564 | 0 | 0 | 0 | 1 | Ref |
| 11482 | MT366938 | Unassigned | Unassigned | Unassigned | 450_0 | Clustered | 16 | VC_450_0 | 16 | 0.9564 | 1 | 0.9564 | 0 | 0 | 0 | 1 | Ref |
| 11483 | MT366939 | Unassigned | Unassigned | Unassigned | 450_0 | Clustered | 16 | VC_450_0 | 16 | 0.9564 | 1 | 0.9564 | 0 | 0 | 0 | 1 | Ref |
| 11484 | MT366940 | Unassigned | Unassigned | Unassigned | 450_0 | Clustered | 16 | VC_450_0 | 16 | 0.9564 | 1 | 0.9564 | 0 | 0 | 0 | 1 | Ref |
| 11485 | MT366941 | Unassigned | Unassigned | Unassigned | 450_0 | Clustered | 16 | VC_450_0 | 16 | 0.9564 | 1 | 0.9564 | 0 | 0 | 0 | 1 | Ref |
| 11486 | MT366942 | Unassigned | Unassigned | Unassigned | 450_0 | Clustered | 16 | VC_450_0 | 16 | 0.9564 | 1 | 0.9564 | 0 | 0 | 0 | 1 | Ref |
| 11487 | MT366943 | Unassigned | Unassigned | Unassigned | 450_0 | Clustered | 16 | VC_450_0 | 16 | 0.9564 | 1 | 0.9564 | 0 | 0 | 0 | 1 | Ref |
| 11488 | MT366944 | Unassigned | Unassigned | Unassigned | 450_0 | Clustered | 16 | VC_450_0 | 16 | 0.9564 | 1 | 0.9564 | 0 | 0 | 0 | 1 | Ref |
| 11489 | MT366945 | Unassigned | Unassigned | Unassigned | 89_2 | Clustered | 5 | VC_89_2 | 5 | 0.2033 | 0.99999643 | 0.2033 | 0 | 0 | 0 | 1 | Ref |
| 11490 | MT366946 | Unassigned | Unassigned | Unassigned | 89_2 | Clustered | 5 | VC_89_2 | 5 | 0.2033 | 0.99999643 | 0.2033 | 0 | 0 | 0 | 1 | Ref |
| 11491 | MT366947 | Unassigned | Unassigned | Unassigned | 89_2 | Clustered | 5 | VC_89_2 | 5 | 0.2033 | 0.99999643 | 0.2033 | 0 | 0 | 0 | 1 | Ref |
| 11492 | MT366948 | Unassigned | Unassigned | Unassigned | 89_2 | Clustered | 5 | VC_89_2 | 5 | 0.2033 | 0.99999643 | 0.2033 | 0 | 0 | 0 | 1 | Ref |
| 11493 | MT374852 | Unassigned | Unassigned | Unassigned | 50_0 | Clustered | 9 | VC_50_0 | 9 | 0.5531 | 1 | 0.5531 | 0 | 0 | 0 | 1 | Ref |
| 11494 | MT374853 | Unassigned | Unassigned | Unassigned | 50_0 | Clustered | 9 | VC_50_0 | 9 | 0.5531 | 1 | 0.5531 | 0 | 0 | 0 | 1 | Ref |
| 11495 | MT374854 | Unassigned | Unassigned | Unassigned | 50_0 | Clustered | 9 | VC_50_0 | 9 | 0.5531 | 1 | 0.5531 | 0 | 0 | 0 | 1 | Ref |
| 11496 | MT374855 | Unassigned | Unassigned | Unassigned | 228_0 | Clustered | 60 | VC_228_0 | 60 | 0.942 | 1 | 0.942 | 0 | 0 | 0 | 1 | Ref |
| 11497 | MT374856 | Unassigned | Unassigned | Unassigned | 50_0 | Clustered | 9 | VC_50_0 | 9 | 0.5531 | 1 | 0.5531 | 0 | 0 | 0 | 1 | Ref |
| 11498 | MT374857 | Unassigned | Unassigned | Unassigned | 228_0 | Clustered | 60 | VC_228_0 | 60 | 0.942 | 1 | 0.942 | 0 | 0 | 0 | 1 | Ref |
| 11499 | MT374858 | Unassigned | Unassigned | Unassigned | 228_0 | Clustered | 60 | VC_228_0 | 60 | 0.942 | 1 | 0.942 | 0 | 0 | 0 | 1 | Ref |
| 11500 | MT374859 | Unassigned | Unassigned | Unassigned | 386_0 | Clustered | 49 | VC_386_0 | 49 | 0.7941 | 1 | 0.7941 | 0 | 0 | 0 | 1 | Ref |

| | | | | | | | | | | | | | | | | | |
|---|---|---|---|---|---|---|---|---|---|---|---|---|---|---|---|---|---|
| 11501 | MT375042 | Unassigned | Unassigned | Unassigned | 125_0 | Clustered | 17 | VC_125_0 | 17 | 0.9244 | 1 | 0.9244 | 0 | 0 | 0 | 1 | Ref |
| 11502 | MT375519 | Unassigned | Unassigned | Unassigned | 487_0 | Clustered | 2 | VC_487_0 | 2 | 0.2529 | 0.94924918 | 0.2401 | 0 | 0 | 0 | 1 | Ref |
| 11503 | MT375520 | Unassigned | Unassigned | Unassigned | 480_0 | Clustered | 19 | VC_480_0 | 19 | 0.5445 | 1 | 0.5445 | 0 | 0 | 0 | 1 | Ref |
| 11504 | MT375521 | Unassigned | Unassigned | Unassigned | 480_0 | Clustered | 19 | VC_480_0 | 19 | 0.5445 | 1 | 0.5445 | 0 | 0 | 0 | 1 | Ref |
| 11505 | MT375525 | Unassigned | Unassigned | Unassigned | 486_0 | Clustered | 18 | VC_486_0 | 18 | 0.887 | 1 | 0.887 | 0 | 0 | 0 | 1 | Ref |
| 11506 | MT375526 | Unassigned | Unassigned | Unassigned | 480_0 | Clustered | 19 | VC_480_0 | 19 | 0.5445 | 1 | 0.5445 | 0 | 0 | 0 | 1 | Ref |
| 11507 | MT375527 | Unassigned | Unassigned | Unassigned | 480_0 | Clustered | 19 | VC_480_0 | 19 | 0.5445 | 1 | 0.5445 | 0 | 0 | 0 | 1 | Ref |
| 11508 | MT375528 | Unassigned | Unassigned | Unassigned | 480_0 | Clustered | 19 | VC_480_0 | 19 | 0.5445 | 1 | 0.5445 | 0 | 0 | 0 | 1 | Ref |
| 11509 | MT375529 | Unassigned | Unassigned | Unassigned | 487_0 | Clustered | 2 | VC_487_0 | 2 | 0.2529 | 0.94924918 | 0.2401 | 0 | 0 | 0 | 1 | Ref |
| 11510 | MT375530 | Unassigned | Unassigned | Unassigned | 480_0 | Clustered | 19 | VC_480_0 | 19 | 0.5445 | 1 | 0.5445 | 0 | 0 | 0 | 1 | Ref |
| 11511 | MT375531 | Unassigned | Unassigned | Unassigned | 480_0 | Clustered | 19 | VC_480_0 | 19 | 0.5445 | 1 | 0.5445 | 0 | 0 | 0 | 1 | Ref |
| 11512 | MT380195 | Unassigned | Unassigned | Unassigned | 192_0 | Clustered | 15 | VC_192_0 | 15 | 0.5071 | 1 | 0.5071 | 0 | 0 | 0 | 1 | Ref |
| 11513 | MT385367 | Unassigned | Unassigned | Unassigned | 492_1 | Clustered | 20 | VC_492_1 | 20 | 0.4176 | 1 | 0.4176 | 0 | 0 | 0 | 1 | Ref |
| 11514 | MT408532 | Unassigned | Unassigned | Unassigned | 432_4 | Clustered/Singleton | 1 | VC_432_4 | 1 | 0 | 1 | 0 | 0 | 0 | 0 | 1 | Ref |
| 11515 | MT409116 | Unassigned | Unassigned | Unassigned | 492_1 | Clustered | 20 | VC_492_1 | 20 | 0.4176 | 1 | 0.4176 | 0 | 0 | 0 | 1 | Ref |
| 11516 | MT409176 | Unassigned | Unassigned | Unassigned | 329_16 | Clustered | 4 | VC_329_16 | 4 | 0.0158 | 0.9999807 | 0.0158 | 0 | 0 | 0 | 1 | Ref |
| 11517 | MT411892 | Unassigned | Unassigned | Unassigned | 149_9 | Clustered | 3 | VC_149_9 | 3 | 0.0084 | 0.97854556 | 0.0082 | 0 | 0 | 0 | 1 | Ref |
| 11518 | MT413450 | Unassigned | Unassigned | Unassigned | 79_0 | Clustered | 64 | VC_79_0 | 64 | 0.4185 | 1 | 0.4185 | 0 | 0 | 0 | 0.7649 | Ref |
| 11519 | MT416090 | Unassigned | Unassigned | Unassigned | 839_0 | Clustered | 5 | VC_839_0 | 5 | 0.165 | 0.99999995 | 0.165 | 0 | 0 | 0 | 1 | Ref |
| 11520 | MT419366 | Unassigned | Unassigned | Unassigned | 385_0 | Clustered | 51 | VC_385_0 | 51 | 0.8754 | 1 | 0.8754 | 0 | 0 | 0 | 1 | Ref |
| 11521 | MT422786 | Unassigned | Unassigned | Unassigned | 100_0 | Clustered | 15 | VC_100_0 | 15 | 0.6536 | 1 | 0.6536 | 0 | 0 | 0 | 1 | Ref |
| 11522 | MT423823 | Unassigned | Unassigned | Unassigned | 11_0 | Clustered | 19 | VC_11_0 | 19 | 0.7562 | 1 | 0.7562 | 0 | 0 | 0 | 1 | Ref |
| 11523 | MT423824 | Unassigned | Unassigned | Unassigned | 11_0 | Clustered | 19 | VC_11_0 | 19 | 0.7562 | 1 | 0.7562 | 0 | 0 | 0 | 1 | Ref |
| 11524 | MT424636 | Unassigned | Unassigned | Unassigned | 340_0 | Clustered | 31 | VC_340_0 | 31 | 0.9549 | 1 | 0.9549 | 0 | 0 | 0 | 1 | Ref |
| 11525 | MT425185 | Unassigned | Unassigned | Unassigned | 823_0 | Clustered | 124 | VC_823_0 | 124 | 0.9492 | 1 | 0.9492 | 0 | 0 | 0 | 1 | Ref |
| 11526 | MT426122 | Unassigned | Unassigned | Unassigned | 1174_0 | Clustered | 4 | VC_1174_0 | 4 | 1 | 1 | 1 | 0 | 0 | 0 | 1 | Ref |
| 11527 | MT427400 | Unassigned | Unassigned | Unassigned | 575_0 | Clustered | 35 | VC_575_0 | 35 | 0.8815 | 1 | 0.8815 | 0 | 0 | 0 | 1 | Ref |
| 11528 | MT430910 | Unassigned | Unassigned | Unassigned | 463_0 | Clustered | 4 | VC_463_0 | 4 | 0.7906 | 0.99997156 | 0.7906 | 0 | 0 | 0 | 1 | Ref |
| 11529 | MT431535 | Unassigned | Unassigned | Unassigned | 912_0 | Clustered | 76 | VC_912_0 | 76 | 0.979 | 1 | 0.979 | 0 | 0 | 0 | 1 | Ref |
| 11530 | MT438396 | Unassigned | Unassigned | Unassigned | 582_3 | Clustered | 12 | VC_582_3 | 12 | 0.2357 | 1 | 0.2357 | 0 | 0 | 0 | 0.8636 | Ref |
| 11531 | MT438761 | Unassigned | Unassigned | Unassigned | 739_0 | Clustered | 26 | VC_739_0 | 26 | 0.9571 | 1 | 0.9571 | 0 | 0 | 0 | 1 | Ref |
| 11532 | MT442039 | Unassigned | Unassigned | Unassigned | 972_0 | Clustered | 6 | VC_972_0 | 6 | 0.2892 | 1 | 0.2892 | 0 | 0 | 0 | 1 | Ref |
| 11533 | MT446386 | Unassigned | Unassigned | Unassigned | 813_1 | Clustered | 24 | VC_813_1 | 24 | 0.535 | 1 | 0.535 | 0 | 0 | 0 | 1 | Ref |
| 11534 | MT446387 | Unassigned | Unassigned | Unassigned | 283_61 | Clustered/Singleton | 1 | VC_283_61 | 1 | 0 | 1 | 0 | 0 | 0 | 0 | 1 | Ref |
| 11535 | MT446388 | Unassigned | Unassigned | Unassigned | 329_7 | Clustered | 18 | VC_329_7 | 18 | 0.0938 | 1 | 0.0938 | 0 | 0 | 0 | 0.7778 | Ref |
| 11536 | MT446389 | Unassigned | Unassigned | Unassigned | 835_0 | Clustered | 67 | VC_835_0 | 67 | 0.9624 | 1 | 0.9624 | 0 | 0 | 0 | 1 | Ref |
| 11537 | MT446390 | Unassigned | Unassigned | Unassigned | 286_10 | Clustered | 9 | VC_286_10 | 9 | 0.0615 | 0.99999878 | 0.0615 | 0 | 0 | 0 | 0.6667 | Ref |
| 11538 | MT446391 | Unassigned | Unassigned | Unassigned | 835_0 | Clustered | 67 | VC_835_0 | 67 | 0.9624 | 1 | 0.9624 | 0 | 0 | 0 | 1 | Ref |
| 11539 | MT446392 | Unassigned | Unassigned | Unassigned | 290_3 | Clustered | 2 | VC_290_3 | 2 | 0.0102 | 0.92273843 | 0.0094 | 0 | 0 | 0 | 1 | Ref |
| 11540 | MT446393 | Unassigned | Unassigned | Unassigned | 451_0 | Clustered | 80 | VC_451_0 | 80 | 0.9991 | 1 | 0.9991 | 0 | 0 | 0 | 1 | Ref |
| 11541 | MT446394 | Unassigned | Unassigned | Unassigned | 451_0 | Clustered | 80 | VC_451_0 | 80 | 0.9991 | 1 | 0.9991 | 0 | 0 | 0 | 1 | Ref |
| 11542 | MT446396 | Unassigned | Unassigned | Unassigned | 290_0 | Clustered | 10 | VC_290_0 | 10 | 0.0981 | 1 | 0.0981 | 0 | 0 | 0 | 1 | Ref |
| 11543 | MT446397 | Unassigned | Unassigned | Unassigned | 792_0 | Clustered | 35 | VC_792_0 | 35 | 0.9828 | 1 | 0.9828 | 0 | 0 | 0 | 1 | Ref |
| 11544 | MT446398 | Unassigned | Unassigned | Unassigned | 792_0 | Clustered | 35 | VC_792_0 | 35 | 0.9828 | 1 | 0.9828 | 0 | 0 | 0 | 1 | Ref |
| 11545 | MT446399 | Unassigned | Unassigned | Unassigned | 792_0 | Clustered | 35 | VC_792_0 | 35 | 0.9828 | 1 | 0.9828 | 0 | 0 | 0 | 1 | Ref |
| 11546 | MT446400 | Unassigned | Unassigned | Unassigned | 792_0 | Clustered | 35 | VC_792_0 | 35 | 0.9828 | 1 | 0.9828 | 0 | 0 | 0 | 1 | Ref |
| 11547 | MT446402 | Unassigned | Unassigned | Unassigned | 792_0 | Clustered | 35 | VC_792_0 | 35 | 0.9828 | 1 | 0.9828 | 0 | 0 | 0 | 1 | Ref |
| 11548 | MT446403 | Unassigned | Unassigned | Unassigned | 792_0 | Clustered | 35 | VC_792_0 | 35 | 0.9828 | 1 | 0.9828 | 0 | 0 | 0 | 1 | Ref |
| 11549 | MT446405 | Unassigned | Unassigned | Unassigned | 618_0 | Clustered | 13 | VC_618_0 | 13 | 0.6252 | 1 | 0.6252 | 0 | 0 | 0 | 1 | Ref |
| 11550 | MT446406 | Unassigned | Unassigned | Unassigned | 792_0 | Clustered | 35 | VC_792_0 | 35 | 0.9828 | 1 | 0.9828 | 0 | 0 | 0 | 1 | Ref |
| 11551 | MT446407 | Unassigned | Unassigned | Unassigned | 813_0 | Clustered | 12 | VC_813_0 | 12 | 0.2347 | 1 | 0.2347 | 0 | 0 | 0 | 1 | Ref |
| 11552 | MT446408 | Unassigned | Unassigned | Unassigned | 618_0 | Clustered | 13 | VC_618_0 | 13 | 0.6252 | 1 | 0.6252 | 0 | 0 | 0 | 1 | Ref |
| 11553 | MT446409 | Unassigned | Unassigned | Unassigned | 792_0 | Clustered | 35 | VC_792_0 | 35 | 0.9828 | 1 | 0.9828 | 0 | 0 | 0 | 1 | Ref |
| 11554 | MT446410 | Unassigned | Unassigned | Unassigned | 813_1 | Clustered | 24 | VC_813_1 | 24 | 0.535 | 1 | 0.535 | 0 | 0 | 0 | 1 | Ref |
| 11555 | MT446411 | Unassigned | Unassigned | Unassigned | 290_0 | Clustered | 10 | VC_290_0 | 10 | 0.0981 | 1 | 0.0981 | 0 | 0 | 0 | 1 | Ref |
| 11556 | MT446412 | Unassigned | Unassigned | Unassigned | 290_0 | Clustered | 10 | VC_290_0 | 10 | 0.0981 | 1 | 0.0981 | 0 | 0 | 0 | 1 | Ref |
| 11557 | MT446413 | Unassigned | Unassigned | Unassigned | 813_0 | Clustered | 12 | VC_813_0 | 12 | 0.2347 | 1 | 0.2347 | 0 | 0 | 0 | 1 | Ref |
| 11558 | MT446414 | Unassigned | Unassigned | Unassigned | 813_0 | Clustered | 12 | VC_813_0 | 12 | 0.2347 | 1 | 0.2347 | 0 | 0 | 0 | 1 | Ref |
| 11559 | MT446415 | Unassigned | Unassigned | Unassigned | 290_0 | Clustered | 10 | VC_290_0 | 10 | 0.0981 | 1 | 0.0981 | 0 | 0 | 0 | 1 | Ref |
| 11560 | MT446416 | Unassigned | Unassigned | Unassigned | 164_1 | Clustered/Singleton | 1 | VC_164_1 | 1 | 0 | 1 | 0 | 0 | 0 | 0 | 1 | Ref |
| 11561 | MT446417 | Unassigned | Unassigned | Unassigned | 823_0 | Clustered | 124 | VC_823_0 | 124 | 0.9492 | 1 | 0.9492 | 0 | 0 | 0 | 1 | Ref |
| 11562 | MT446418 | Unassigned | Unassigned | Unassigned | 823_0 | Clustered | 124 | VC_823_0 | 124 | 0.9492 | 1 | 0.9492 | 0 | 0 | 0 | 1 | Ref |
| 11563 | MT446419 | Unassigned | Unassigned | Unassigned | 792_0 | Clustered | 35 | VC_792_0 | 35 | 0.9828 | 1 | 0.9828 | 0 | 0 | 0 | 1 | Ref |
| 11564 | MT446420 | Unassigned | Unassigned | Unassigned | 290_1 | Clustered | 8 | VC_290_1 | 8 | 0.0757 | 1 | 0.0757 | 0 | 0 | 0 | 0.8571 | Ref |
| 11565 | MT446421 | Unassigned | Unassigned | Unassigned | 290_0 | Clustered | 10 | VC_290_0 | 10 | 0.0981 | 1 | 0.0981 | 0 | 0 | 0 | 1 | Ref |
| 11566 | MT446422 | Unassigned | Unassigned | Unassigned | 290_1 | Clustered | 8 | VC_290_1 | 8 | 0.0757 | 1 | 0.0757 | 0 | 0 | 0 | 0.8571 | Ref |
| 11567 | MT446423 | Unassigned | Unassigned | Unassigned | 286_5 | Clustered/Singleton | 1 | VC_286_5 | 1 | 0 | 1 | 0 | 0 | 0 | 0 | 1 | Ref |
| 11568 | MT448616 | Unassigned | Unassigned | Unassigned | 971_1 | Clustered | 24 | VC_971_1 | 24 | 0.7393 | 1 | 0.7393 | 0 | 0 | 0 | 0.8913 | Ref |
| 11569 | MT448617 | Unassigned | Unassigned | Unassigned | 971_1 | Clustered | 24 | VC_971_1 | 24 | 0.7393 | 1 | 0.7393 | 0 | 0 | 0 | 0.8913 | Ref |
| 11570 | MT451873 | Unassigned | Unassigned | Unassigned | 904_1 | Clustered | 4 | VC_904_1 | 4 | 0.1486 | 0.99997723 | 0.1486 | 0 | 0 | 0 | 1 | Ref |
| 11571 | MT451985 | Unassigned | Unassigned | Unassigned | 1240_0 | Clustered | 82 | VC_1240_0 | 82 | 0.7945 | 1 | 0.7945 | 0 | 0 | 0 | 1 | Ref |
| 11572 | MT457475 | Unassigned | Unassigned | Unassigned | 777_0 | Clustered | 3 | VC_777_0 | 3 | 0.4763 | 0.9961812 | 0.4745 | 0 | 0 | 0 | 1 | Ref |
| 11573 | MT457552 | Unassigned | Unassigned | Unassigned | 287_0 | Clustered | 2 | VC_287_0 | 2 | 0.0279 | 0.95771461 | 0.0267 | 0 | 0 | 0 | 1 | Ref |
| 11574 | MT457553 | Unassigned | Unassigned | Unassigned | 287_0 | Clustered | 2 | VC_287_0 | 2 | 0.0279 | 0.95771461 | 0.0267 | 0 | 0 | 0 | 1 | Ref |
| 11575 | MT459144 | Unassigned | Unassigned | Unassigned | 971_1 | Clustered | 24 | VC_971_1 | 24 | 0.7393 | 1 | 0.7393 | 0 | 0 | 0 | 0.8913 | Ref |
| 11576 | MT459794 | Unassigned | Unassigned | Unassigned | 817_0 | Clustered | 14 | VC_817_0 | 14 | 0.6342 | 1 | 0.6342 | 0 | 0 | 0 | 1 | Ref |
| 11577 | MT460514 | Unassigned | Unassigned | Unassigned | 971_1 | Clustered | 24 | VC_971_1 | 24 | 0.7393 | 1 | 0.7393 | 0 | 0 | 0 | 0.8913 | Ref |
| 11578 | MT460515 | Unassigned | Unassigned | Unassigned | 971_1 | Clustered | 24 | VC_971_1 | 24 | 0.7393 | 1 | 0.7393 | 0 | 0 | 0 | 0.8913 | Ref |
| 11579 | MT460516 | Unassigned | Unassigned | Unassigned | 971_1 | Clustered | 24 | VC_971_1 | 24 | 0.7393 | 1 | 0.7393 | 0 | 0 | 0 | 0.8913 | Ref |
| 11580 | MT460517 | Unassigned | Unassigned | Unassigned | 971_1 | Clustered | 24 | VC_971_1 | 24 | 0.7393 | 1 | 0.7393 | 0 | 0 | 0 | 0.8913 | Ref |
| 11581 | MT460518 | Unassigned | Unassigned | Unassigned | 971_1 | Clustered | 24 | VC_971_1 | 24 | 0.7393 | 1 | 0.7393 | 0 | 0 | 0 | 0.8913 | Ref |
| 11582 | MT465573 | Unassigned | Unassigned | Unassigned | 394_0 | Clustered | 2 | VC_394_0 | 2 | 0.0211 | 0.957922 | 0.0202 | 0 | 0 | 0 | 1 | Ref |
| 11583 | MT478991 | Unassigned | Unassigned | Unassigned | 590_0 | Clustered | 27 | VC_590_0 | 27 | 0.9947 | 1 | 0.9947 | 0 | 0 | 0 | 1 | Ref |
| 11584 | MT478992 | Unassigned | Unassigned | Unassigned | 584_0 | Clustered | 28 | VC_584_0 | 28 | 0.7704 | 1 | 0.7704 | 0 | 0 | 0 | 1 | Ref |
| 11585 | MT478993 | Unassigned | Unassigned | Unassigned | 835_0 | Clustered | 67 | VC_835_0 | 67 | 0.9624 | 1 | 0.9624 | 0 | 0 | 0 | 1 | Ref |
| 11586 | MT490303 | Unassigned | Unassigned | Unassigned | 928_0 | Clustered | 3 | VC_928_0 | 3 | 0.7844 | 0.9944138 | 0.78 | 0 | 0 | 0 | 1 | Ref |
| 11587 | MT490373 | Unassigned | Unassigned | Unassigned | 517_1 | Clustered | 108 | VC_517_1 | 108 | 0.8483 | 1 | 0.8483 | 0 | 0 | 0 | 0.9955 | Ref |
| 11588 | MT491204 | Unassigned | Unassigned | Unassigned | 79_6 | Clustered | 11 | VC_79_6 | 11 | 0.0528 | 1 | 0.0528 | 0 | 0 | 0 | 0.7455 | Ref |
| 11589 | MT491205 | Unassigned | Unassigned | Unassigned | 79_6 | Clustered | 11 | VC_79_6 | 11 | 0.0528 | 1 | 0.0528 | 0 | 0 | 0 | 0.7455 | Ref |
| 11590 | MT491206 | Unassigned | Unassigned | Unassigned | 79_6 | Clustered | 11 | VC_79_6 | 11 | 0.0528 | 1 | 0.0528 | 0 | 0 | 0 | 0.7455 | Ref |
| 11591 | MT491207 | Unassigned | Unassigned | Unassigned | 631_0 | Clustered | 3 | VC_631_0 | 3 | 0.1213 | 0.99819199 | 0.1211 | 0 | 0 | 0 | 1 | Ref |
| 11592 | MT491208 | Unassigned | Unassigned | Unassigned | 631_0 | Clustered | 3 | VC_631_0 | 3 | 0.1213 | 0.99819199 | 0.1211 | 0 | 0 | 0 | 1 | Ref |
| 11593 | MT495635 | Unassigned | Unassigned | Unassigned | 54_0 | Clustered | 6 | VC_54_0 | 6 | 0.6725 | 0.9999999 | 0.6725 | 0 | 0 | 0 | 1 | Ref |
| 11594 | MT496969 | Unassigned | Unassigned | Unassigned | 451_0 | Clustered | 80 | VC_451_0 | 80 | 0.9991 | 1 | 0.9991 | 0 | 0 | 0 | 1 | Ref |
| 11595 | MT496970 | Unassigned | Unassigned | Unassigned | 847_1 | Clustered | 26 | VC_847_1 | 26 | 0.6323 | 1 | 0.6323 | 0 | 0 | 0 | 0.9815 | Ref |
| 11596 | MT496971 | Unassigned | Unassigned | Unassigned | 452_0 | Clustered | 66 | VC_452_0 | 66 | 0.9248 | 1 | 0.9248 | 0 | 0 | 0 | 1 | Ref |
| 11597 | MT497017 | Unassigned | Unassigned | Unassigned | 1015_0 | Clustered | 62 | VC_1015_0 | 62 | 0.9947 | 1 | 0.9947 | 0 | 0 | 0 | 1 | Ref |
| 11598 | MT497065 | Unassigned | Unassigned | Unassigned | 1015_0 | Clustered | 62 | VC_1015_0 | 62 | 0.9947 | 1 | 0.9947 | 0 | 0 | 0 | 1 | Ref |
| 11599 | MT497066 | Unassigned | Unassigned | Unassigned | 1015_0 | Clustered | 62 | VC_1015_0 | 62 | 0.9947 | 1 | 0.9947 | 0 | 0 | 0 | 1 | Ref |
| 11600 | MT497067 | Unassigned | Unassigned | Unassigned | 1015_0 | Clustered | 62 | VC_1015_0 | 62 | 0.9947 | 1 | 0.9947 | 0 | 0 | 0 | 1 | Ref |
| 11601 | MT497068 | Unassigned | Unassigned | Unassigned | 1015_0 | Clustered | 62 | VC_1015_0 | 62 | 0.9947 | 1 | 0.9947 | 0 | 0 | 0 | 1 | Ref |
| 11602 | MT497069 | Unassigned | Unassigned | Unassigned | 1015_0 | Clustered | 62 | VC_1015_0 | 62 | 0.9947 | 1 | 0.9947 | 0 | 0 | 0 | 1 | Ref |
| 11603 | MT497070 | Unassigned | Unassigned | Unassigned | 1015_0 | Clustered | 62 | VC_1015_0 | 62 | 0.9947 | 1 | 0.9947 | 0 | 0 | 0 | 1 | Ref |
| 11604 | MT497071 | Unassigned | Unassigned | Unassigned | 1015_0 | Clustered | 62 | VC_1015_0 | 62 | 0.9947 | 1 | 0.9947 | 0 | 0 | 0 | 1 | Ref |
| 11605 | MT497072 | Unassigned | Unassigned | Unassigned | 1015_0 | Clustered | 62 | VC_1015_0 | 62 | 0.9947 | 1 | 0.9947 | 0 | 0 | 0 | 1 | Ref |
| 11606 | MT497073 | Unassigned | Unassigned | Unassigned | 1015_0 | Clustered | 62 | VC_1015_0 | 62 | 0.9947 | 1 | 0.9947 | 0 | 0 | 0 | 1 | Ref |
| 11607 | MT497074 | Unassigned | Unassigned | Unassigned | 1015_0 | Clustered | 62 | VC_1015_0 | 62 | 0.9947 | 1 | 0.9947 | 0 | 0 | 0 | 1 | Ref |
| 11608 | MT497075 | Unassigned | Unassigned | Unassigned | 1015_0 | Clustered | 62 | VC_1015_0 | 62 | 0.9947 | 1 | 0.9947 | 0 | 0 | 0 | 1 | Ref |
| 11609 | MT497076 | Unassigned | Unassigned | Unassigned | 1015_0 | Clustered | 62 | VC_1015_0 | 62 | 0.9947 | 1 | 0.9947 | 0 | 0 | 0 | 1 | Ref |
| 11610 | MT497077 | Unassigned | Unassigned | Unassigned | 1015_0 | Clustered | 62 | VC_1015_0 | 62 | 0.9947 | 1 | 0.9947 | 0 | 0 | 0 | 1 | Ref |
| 11611 | MT497078 | Unassigned | Unassigned | Unassigned | 1015_0 | Clustered | 62 | VC_1015_0 | 62 | 0.9947 | 1 | 0.9947 | 0 | 0 | 0 | 1 | Ref |
| 11612 | MT497079 | Unassigned | Unassigned | Unassigned | 1015_0 | Clustered | 62 | VC_1015_0 | 62 | 0.9947 | 1 | 0.9947 | 0 | 0 | 0 | 1 | Ref |
| 11613 | MT497080 | Unassigned | Unassigned | Unassigned | 1015_0 | Clustered | 62 | VC_1015_0 | 62 | 0.9947 | 1 | 0.9947 | 0 | 0 | 0 | 1 | Ref |
| 11614 | MT497081 | Unassigned | Unassigned | Unassigned | 1015_0 | Clustered | 62 | VC_1015_0 | 62 | 0.9947 | 1 | 0.9947 | 0 | 0 | 0 | 1 | Ref |
| 11615 | MT497082 | Unassigned | Unassigned | Unassigned | 1015_0 | Clustered | 62 | VC_1015_0 | 62 | 0.9947 | 1 | 0.9947 | 0 | 0 | 0 | 1 | Ref |
| 11616 | MT497083 | Unassigned | Unassigned | Unassigned | 1015_0 | Clustered | 62 | VC_1015_0 | 62 | 0.9947 | 1 | 0.9947 | 0 | 0 | 0 | 1 | Ref |
| 11617 | MT497084 | Unassigned | Unassigned | Unassigned | 1015_0 | Clustered | 62 | VC_1015_0 | 62 | 0.9947 | 1 | 0.9947 | 0 | 0 | 0 | 1 | Ref |
| 11618 | MT497085 | Unassigned | Unassigned | Unassigned | 1015_0 | Clustered | 62 | VC_1015_0 | 62 | 0.9947 | 1 | 0.9947 | 0 | 0 | 0 | 1 | Ref |
| 11619 | MT497086 | Unassigned | Unassigned | Unassigned | 1015_0 | Clustered | 62 | VC_1015_0 | 62 | 0.9947 | 1 | 0.9947 | 0 | 0 | 0 | 1 | Ref |
| 11620 | MT497087 | Unassigned | Unassigned | Unassigned | 1015_0 | Clustered | 62 | VC_1015_0 | 62 | 0.9947 | 1 | 0.9947 | 0 | 0 | 0 | 1 | Ref |
| 11621 | MT497088 | Unassigned | Unassigned | Unassigned | 1015_0 | Clustered | 62 | VC_1015_0 | 62 | 0.9947 | 1 | 0.9947 | 0 | 0 | 0 | 1 | Ref |
| 11622 | MT497089 | Unassigned | Unassigned | Unassigned | 1015_0 | Clustered | 62 | VC_1015_0 | 62 | 0.9947 | 1 | 0.9947 | 0 | 0 | 0 | 1 | Ref |
| 11623 | MT497090 | Unassigned | Unassigned | Unassigned | 1015_0 | Clustered | 62 | VC_1015_0 | 62 | 0.9947 | 1 | 0.9947 | 0 | 0 | 0 | 1 | Ref |
| 11624 | MT497091 | Unassigned | Unassigned | Unassigned | 1015_0 | Clustered | 62 | VC_1015_0 | 62 | 0.9947 | 1 | 0.9947 | 0 | 0 | 0 | 1 | Ref |
| 11625 | MT497092 | Unassigned | Unassigned | Unassigned | 1015_0 | Clustered | 62 | VC_1015_0 | 62 | 0.9947 | 1 | 0.9947 | 0 | 0 | 0 | 1 | Ref |
| 11626 | MT497093 | Unassigned | Unassigned | Unassigned | 1015_0 | Clustered | 62 | VC_1015_0 | 62 | 0.9947 | 1 | 0.9947 | 0 | 0 | 0 | 1 | Ref |
| 11627 | MT497094 | Unassigned | Unassigned | Unassigned | 1015_0 | Clustered | 62 | VC_1015_0 | 62 | 0.9947 | 1 | 0.9947 | 0 | 0 | 0 | 1 | Ref |
| 11628 | MT497095 | Unassigned | Unassigned | Unassigned | 1015_0 | Clustered | 62 | VC_1015_0 | 62 | 0.9947 | 1 | 0.9947 | 0 | 0 | 0 | 1 | Ref |
| 11629 | MT497096 | Unassigned | Unassigned | Unassigned | 1015_0 | Clustered | 62 | VC_1015_0 | 62 | 0.9947 | 1 | 0.9947 | 0 | 0 | 0 | 1 | Ref |
| 11630 | MT497097 | Unassigned | Unassigned | Unassigned | 1015_0 | Clustered | 62 | VC_1015_0 | 62 | 0.9947 | 1 | 0.9947 | 0 | 0 | 0 | 1 | Ref |
| 11631 | MT497098 | Unassigned | Unassigned | Unassigned | 1015_0 | Clustered | 62 | VC_1015_0 | 62 | 0.9947 | 1 | 0.9947 | 0 | 0 | 0 | 1 | Ref |
| 11632 | MT497099 | Unassigned | Unassigned | Unassigned | 1015_0 | Clustered | 62 | VC_1015_0 | 62 | 0.9947 | 1 | 0.9947 | 0 | 0 | 0 | 1 | Ref |
| 11633 | MT497100 | Unassigned | Unassigned | Unassigned | 1015_0 | Clustered | 62 | VC_1015_0 | 62 | 0.9947 | 1 | 0.9947 | 0 | 0 | 0 | 1 | Ref |
| 11634 | MT497101 | Unassigned | Unassigned | Unassigned | 1015_0 | Clustered | 62 | VC_1015_0 | 62 | 0.9947 | 1 | 0.9947 | 0 | 0 | 0 | 1 | Ref |
| 11635 | MT497102 | Unassigned | Unassigned | Unassigned | 1015_0 | Clustered | 62 | VC_1015_0 | 62 | 0.9947 | 1 | 0.9947 | 0 | 0 | 0 | 1 | Ref |
| 11636 | MT497103 | Unassigned | Unassigned | Unassigned | 1015_0 | Clustered | 62 | VC_1015_0 | 62 | 0.9947 | 1 | 0.9947 | 0 | 0 | 0 | 1 | Ref |
| 11637 | MT497104 | Unassigned | Unassigned | Unassigned | 1015_0 | Clustered | 62 | VC_1015_0 | 62 | 0.9947 | 1 | 0.9947 | 0 | 0 | 0 | 1 | Ref |
| 11638 | MT497105 | Unassigned | Unassigned | Unassigned | 1015_0 | Clustered | 62 | VC_1015_0 | 62 | 0.9947 | 1 | 0.9947 | 0 | 0 | 0 | 1 | Ref |
| 11639 | MT497106 | Unassigned | Unassigned | Unassigned | 1015_0 | Clustered | 62 | VC_1015_0 | 62 | 0.9947 | 1 | 0.9947 | 0 | 0 | 0 | 1 | Ref |
| 11640 | MT497107 | Unassigned | Unassigned | Unassigned | 1015_0 | Clustered | 62 | VC_1015_0 | 62 | 0.9947 | 1 | 0.9947 | 0 | 0 | 0 | 1 | Ref |
| 11641 | MT497108 | Unassigned | Unassigned | Unassigned | 1015_0 | Clustered | 62 | VC_1015_0 | 62 | 0.9947 | 1 | 0.9947 | 0 | 0 | 0 | 1 | Ref |
| 11642 | MT497109 | Unassigned | Unassigned | Unassigned | 1015_0 | Clustered | 62 | VC_1015_0 | 62 | 0.9947 | 1 | 0.9947 | 0 | 0 | 0 | 1 | Ref |
| 11643 | MT497110 | Unassigned | Unassigned | Unassigned | 1015_0 | Clustered | 62 | VC_1015_0 | 62 | 0.9947 | 1 | 0.9947 | 0 | 0 | 0 | 1 | Ref |
| 11644 | MT497111 | Unassigned | Unassigned | Unassigned | 1015_0 | Clustered | 62 | VC_1015_0 | 62 | 0.9947 | 1 | 0.9947 | 0 | 0 | 0 | 1 | Ref |
| 11645 | MT497112 | Unassigned | Unassigned | Unassigned | 1015_0 | Clustered | 62 | VC_1015_0 | 62 | 0.9947 | 1 | 0.9947 | 0 | 0 | 0 | 1 | Ref |
| 11646 | MT497113 | Unassigned | Unassigned | Unassigned | 1015_0 | Clustered | 62 | VC_1015_0 | 62 | 0.9947 | 1 | 0.9947 | 0 | 0 | 0 | 1 | Ref |
| 11647 | MT497114 | Unassigned | Unassigned | Unassigned | 1015_0 | Clustered | 62 | VC_1015_0 | 62 | 0.9947 | 1 | 0.9947 | 0 | 0 | 0 | 1 | Ref |
| 11648 | MT497115 | Unassigned | Unassigned | Unassigned | 1015_0 | Clustered | 62 | VC_1015_0 | 62 | 0.9947 | 1 | 0.9947 | 0 | 0 | 0 | 1 | Ref |
| 11649 | MT497116 | Unassigned | Unassigned | Unassigned | 1015_0 | Clustered | 62 | VC_1015_0 | 62 | 0.9947 | 1 | 0.9947 | 0 | 0 | 0 | 1 | Ref |
| 11650 | MT497117 | Unassigned | Unassigned | Unassigned | 1015_0 | Clustered | 62 | VC_1015_0 | 62 | 0.9947 | 1 | 0.9947 | 0 | 0 | 0 | 1 | Ref |
| 11651 | MT497118 | Unassigned | Unassigned | Unassigned | 1015_0 | Clustered | 62 | VC_1015_0 | 62 | 0.9947 | 1 | 0.9947 | 0 | 0 | 0 | 1 | Ref |
| 11652 | MT497119 | Unassigned | Unassigned | Unassigned | 1015_0 | Clustered | 62 | VC_1015_0 | 62 | 0.9947 | 1 | 0.9947 | 0 | 0 | 0 | 1 | Ref |
| 11653 | MT497120 | Unassigned | Unassigned | Unassigned | 1015_0 | Clustered | 62 | VC_1015_0 | 62 | 0.9947 | 1 | 0.9947 | 0 | 0 | 0 | 1 | Ref |
| 11654 | MT497121 | Unassigned | Unassigned | Unassigned | 1015_0 | Clustered | 62 | VC_1015_0 | 62 | 0.9947 | 1 | 0.9947 | 0 | 0 | 0 | 1 | Ref |
| 11655 | MT497122 | Unassigned | Unassigned | Unassigned | 1015_0 | Clustered | 62 | VC_1015_0 | 62 | 0.9947 | 1 | 0.9947 | 0 | 0 | 0 | 1 | Ref |
| 11656 | MT497123 | Unassigned | Unassigned | Unassigned | 1015_0 | Clustered | 62 | VC_1015_0 | 62 | 0.9947 | 1 | 0.9947 | 0 | 0 | 0 | 1 | Ref |
| 11657 | MT497124 | Unassigned | Unassigned | Unassigned | 1015_0 | Clustered | 62 | VC_1015_0 | 62 | 0.9947 | 1 | 0.9947 | 0 | 0 | 0 | 1 | Ref |
| 11658 | MT497125 | Unassigned | Unassigned | Unassigned | 1015_0 | Clustered | 62 | VC_1015_0 | 62 | 0.9947 | 1 | 0.9947 | 0 | 0 | 0 | 1 | Ref |
| 11659 | MT497224 | Unassigned | Unassigned | Unassigned | 1396_0 | Clustered | 110 | VC_1396_0 | 110 | 1 | 1 | 1 | 0 | 0 | 0 | 0.9746 | Ref |
| 11660 | MT497225 | Unassigned | Unassigned | Unassigned | 1396_0 | Clustered | 110 | VC_1396_0 | 110 | 1 | 1 | 1 | 0 | 0 | 0 | 0.9746 | Ref |
| 11661 | MT497226 | Unassigned | Unassigned | Unassigned | 1396_0 | Clustered | 110 | VC_1396_0 | 110 | 1 | 1 | 1 | 0 | 0 | 0 | 0.9746 | Ref |
| 11662 | MT497227 | Unassigned | Unassigned | Unassigned | 1396_0 | Clustered | 110 | VC_1396_0 | 110 | 1 | 1 | 1 | 0 | 0 | 0 | 0.9746 | Ref |

| | | | | | | | | | | | | | | | | | |
|---|---|---|---|---|---|---|---|---|---|---|---|---|---|---|---|---|---|
| 11663 | MT497228 | Unassigned | Unassigned | Unassigned | 1396_0 | Clustered | 110 | VC_1396_0 | 110 | 1 | 1 | 1 | 0 | 0 | 0 | 0.9746 | Ref |
| 11664 | MT497229 | Unassigned | Unassigned | Unassigned | 1396_0 | Clustered | 110 | VC_1396_0 | 110 | 1 | 1 | 1 | 0 | 0 | 0 | 0.9746 | Ref |
| 11665 | MT497230 | Unassigned | Unassigned | Unassigned | 1396_0 | Clustered | 110 | VC_1396_0 | 110 | 1 | 1 | 1 | 0 | 0 | 0 | 0.9746 | Ref |
| 11666 | MT497231 | Unassigned | Unassigned | Unassigned | 1396_0 | Clustered | 110 | VC_1396_0 | 110 | 1 | 1 | 1 | 0 | 0 | 0 | 0.9746 | Ref |
| 11667 | MT497232 | Unassigned | Unassigned | Unassigned | 1396_0 | Clustered | 110 | VC_1396_0 | 110 | 1 | 1 | 1 | 0 | 0 | 0 | 0.9746 | Ref |
| 11668 | MT497233 | Unassigned | Unassigned | Unassigned | 1396_0 | Clustered | 110 | VC_1396_0 | 110 | 1 | 1 | 1 | 0 | 0 | 0 | 0.9746 | Ref |
| 11669 | MT497234 | Unassigned | Unassigned | Unassigned | 1396_0 | Clustered | 110 | VC_1396_0 | 110 | 1 | 1 | 1 | 0 | 0 | 0 | 0.9746 | Ref |
| 11670 | MT497235 | Unassigned | Unassigned | Unassigned | 1396_0 | Clustered | 110 | VC_1396_0 | 110 | 1 | 1 | 1 | 0 | 0 | 0 | 0.9746 | Ref |
| 11671 | MT497236 | Unassigned | Unassigned | Unassigned | 1396_0 | Clustered | 110 | VC_1396_0 | 110 | 1 | 1 | 1 | 0 | 0 | 0 | 0.9746 | Ref |
| 11672 | MT497237 | Unassigned | Unassigned | Unassigned | 1396_0 | Clustered | 110 | VC_1396_0 | 110 | 1 | 1 | 1 | 0 | 0 | 0 | 0.9746 | Ref |
| 11673 | MT497238 | Unassigned | Unassigned | Unassigned | 1396_0 | Clustered | 110 | VC_1396_0 | 110 | 1 | 1 | 1 | 0 | 0 | 0 | 0.9746 | Ref |
| 11674 | MT497239 | Unassigned | Unassigned | Unassigned | 1396_0 | Clustered | 110 | VC_1396_0 | 110 | 1 | 1 | 1 | 0 | 0 | 0 | 0.9746 | Ref |
| 11675 | MT497240 | Unassigned | Unassigned | Unassigned | 1396_0 | Clustered | 110 | VC_1396_0 | 110 | 1 | 1 | 1 | 0 | 0 | 0 | 0.9746 | Ref |
| 11676 | MT497241 | Unassigned | Unassigned | Unassigned | 1396_0 | Clustered | 110 | VC_1396_0 | 110 | 1 | 1 | 1 | 0 | 0 | 0 | 0.9746 | Ref |
| 11677 | MT497242 | Unassigned | Unassigned | Unassigned | 1396_0 | Clustered | 110 | VC_1396_0 | 110 | 1 | 1 | 1 | 0 | 0 | 0 | 0.9746 | Ref |
| 11678 | MT497243 | Unassigned | Unassigned | Unassigned | 1396_0 | Clustered | 110 | VC_1396_0 | 110 | 1 | 1 | 1 | 0 | 0 | 0 | 0.9746 | Ref |
| 11679 | MT497244 | Unassigned | Unassigned | Unassigned | 1396_0 | Clustered | 110 | VC_1396_0 | 110 | 1 | 1 | 1 | 0 | 0 | 0 | 0.9746 | Ref |
| 11680 | MT497245 | Unassigned | Unassigned | Unassigned | 1396_0 | Clustered | 110 | VC_1396_0 | 110 | 1 | 1 | 1 | 0 | 0 | 0 | 0.9746 | Ref |
| 11681 | MT497246 | Unassigned | Unassigned | Unassigned | 1396_0 | Clustered | 110 | VC_1396_0 | 110 | 1 | 1 | 1 | 0 | 0 | 0 | 0.9746 | Ref |
| 11682 | MT497247 | Unassigned | Unassigned | Unassigned | 1396_0 | Clustered | 110 | VC_1396_0 | 110 | 1 | 1 | 1 | 0 | 0 | 0 | 0.9746 | Ref |
| 11683 | MT497248 | Unassigned | Unassigned | Unassigned | 1396_0 | Clustered | 110 | VC_1396_0 | 110 | 1 | 1 | 1 | 0 | 0 | 0 | 0.9746 | Ref |
| 11684 | MT497249 | Unassigned | Unassigned | Unassigned | 1396_0 | Clustered | 110 | VC_1396_0 | 110 | 1 | 1 | 1 | 0 | 0 | 0 | 0.9746 | Ref |
| 11685 | MT497250 | Unassigned | Unassigned | Unassigned | 1396_0 | Clustered | 110 | VC_1396_0 | 110 | 1 | 1 | 1 | 0 | 0 | 0 | 0.9746 | Ref |
| 11686 | MT497251 | Unassigned | Unassigned | Unassigned | 1396_0 | Clustered | 110 | VC_1396_0 | 110 | 1 | 1 | 1 | 0 | 0 | 0 | 0.9746 | Ref |
| 11687 | MT497252 | Unassigned | Unassigned | Unassigned | 1396_0 | Clustered | 110 | VC_1396_0 | 110 | 1 | 1 | 1 | 0 | 0 | 0 | 0.9746 | Ref |
| 11688 | MT497253 | Unassigned | Unassigned | Unassigned | 1396_0 | Clustered | 110 | VC_1396_0 | 110 | 1 | 1 | 1 | 0 | 0 | 0 | 0.9746 | Ref |
| 11689 | MT497254 | Unassigned | Unassigned | Unassigned | 1396_0 | Clustered | 110 | VC_1396_0 | 110 | 1 | 1 | 1 | 0 | 0 | 0 | 0.9746 | Ref |
| 11690 | MT497255 | Unassigned | Unassigned | Unassigned | 1396_0 | Clustered | 110 | VC_1396_0 | 110 | 1 | 1 | 1 | 0 | 0 | 0 | 0.9746 | Ref |
| 11691 | MT497256 | Unassigned | Unassigned | Unassigned | 1396_0 | Clustered | 110 | VC_1396_0 | 110 | 1 | 1 | 1 | 0 | 0 | 0 | 0.9746 | Ref |
| 11692 | MT497257 | Unassigned | Unassigned | Unassigned | 1396_0 | Clustered | 110 | VC_1396_0 | 110 | 1 | 1 | 1 | 0 | 0 | 0 | 0.9746 | Ref |
| 11693 | MT497258 | Unassigned | Unassigned | Unassigned | 1396_0 | Clustered | 110 | VC_1396_0 | 110 | 1 | 1 | 1 | 0 | 0 | 0 | 0.9746 | Ref |
| 11694 | MT497259 | Unassigned | Unassigned | Unassigned | 1396_0 | Clustered | 110 | VC_1396_0 | 110 | 1 | 1 | 1 | 0 | 0 | 0 | 0.9746 | Ref |
| 11695 | MT497260 | Unassigned | Unassigned | Unassigned | 1396_0 | Clustered | 110 | VC_1396_0 | 110 | 1 | 1 | 1 | 0 | 0 | 0 | 0.9746 | Ref |
| 11696 | MT497261 | Unassigned | Unassigned | Unassigned | 1396_0 | Clustered | 110 | VC_1396_0 | 110 | 1 | 1 | 1 | 0 | 0 | 0 | 0.9746 | Ref |
| 11697 | MT497262 | Unassigned | Unassigned | Unassigned | 1396_0 | Clustered | 110 | VC_1396_0 | 110 | 1 | 1 | 1 | 0 | 0 | 0 | 0.9746 | Ref |
| 11698 | MT497263 | Unassigned | Unassigned | Unassigned | 1396_0 | Clustered | 110 | VC_1396_0 | 110 | 1 | 1 | 1 | 0 | 0 | 0 | 0.9746 | Ref |
| 11699 | MT497264 | Unassigned | Unassigned | Unassigned | 1396_0 | Clustered | 110 | VC_1396_0 | 110 | 1 | 1 | 1 | 0 | 0 | 0 | 0.9746 | Ref |
| 11700 | MT497265 | Unassigned | Unassigned | Unassigned | 1396_0 | Clustered | 110 | VC_1396_0 | 110 | 1 | 1 | 1 | 0 | 0 | 0 | 0.9746 | Ref |
| 11701 | MT497266 | Unassigned | Unassigned | Unassigned | 1396_0 | Clustered | 110 | VC_1396_0 | 110 | 1 | 1 | 1 | 0 | 0 | 0 | 0.9746 | Ref |
| 11702 | MT497267 | Unassigned | Unassigned | Unassigned | 1396_0 | Clustered | 110 | VC_1396_0 | 110 | 1 | 1 | 1 | 0 | 0 | 0 | 0.9746 | Ref |
| 11703 | MT497268 | Unassigned | Unassigned | Unassigned | 1396_0 | Clustered | 110 | VC_1396_0 | 110 | 1 | 1 | 1 | 0 | 0 | 0 | 0.9746 | Ref |
| 11704 | MT497269 | Unassigned | Unassigned | Unassigned | 1396_0 | Clustered | 110 | VC_1396_0 | 110 | 1 | 1 | 1 | 0 | 0 | 0 | 0.9746 | Ref |
| 11705 | MT497270 | Unassigned | Unassigned | Unassigned | 1396_0 | Clustered | 110 | VC_1396_0 | 110 | 1 | 1 | 1 | 0 | 0 | 0 | 0.9746 | Ref |
| 11706 | MT497271 | Unassigned | Unassigned | Unassigned | 1396_0 | Clustered | 110 | VC_1396_0 | 110 | 1 | 1 | 1 | 0 | 0 | 0 | 0.9746 | Ref |
| 11707 | MT497272 | Unassigned | Unassigned | Unassigned | 1396_0 | Clustered | 110 | VC_1396_0 | 110 | 1 | 1 | 1 | 0 | 0 | 0 | 0.9746 | Ref |
| 11708 | MT497273 | Unassigned | Unassigned | Unassigned | 1396_0 | Clustered | 110 | VC_1396_0 | 110 | 1 | 1 | 1 | 0 | 0 | 0 | 0.9746 | Ref |
| 11709 | MT497274 | Unassigned | Unassigned | Unassigned | 1396_0 | Clustered | 110 | VC_1396_0 | 110 | 1 | 1 | 1 | 0 | 0 | 0 | 0.9746 | Ref |
| 11710 | MT497275 | Unassigned | Unassigned | Unassigned | 1396_0 | Clustered | 110 | VC_1396_0 | 110 | 1 | 1 | 1 | 0 | 0 | 0 | 0.9746 | Ref |
| 11711 | MT497276 | Unassigned | Unassigned | Unassigned | 1396_0 | Clustered | 110 | VC_1396_0 | 110 | 1 | 1 | 1 | 0 | 0 | 0 | 0.9746 | Ref |
| 11712 | MT497277 | Unassigned | Unassigned | Unassigned | 1396_0 | Clustered | 110 | VC_1396_0 | 110 | 1 | 1 | 1 | 0 | 0 | 0 | 0.9746 | Ref |
| 11713 | MT497278 | Unassigned | Unassigned | Unassigned | 1396_0 | Clustered | 110 | VC_1396_0 | 110 | 1 | 1 | 1 | 0 | 0 | 0 | 0.9746 | Ref |
| 11714 | MT497279 | Unassigned | Unassigned | Unassigned | 1396_0 | Clustered | 110 | VC_1396_0 | 110 | 1 | 1 | 1 | 0 | 0 | 0 | 0.9746 | Ref |
| 11715 | MT498035 | Unassigned | Unassigned | Unassigned | 1093_0 | Clustered | 40 | VC_1093_0 | 40 | 0.8754 | 1 | 0.8754 | 0 | 0 | 0 | 1 | Ref |
| 11716 | MT498036 | Unassigned | Unassigned | Unassigned | 1095_0 | Clustered | 16 | VC_1095_0 | 16 | 0.458 | 1 | 0.458 | 0 | 0 | 0 | 1 | Ref |
| 11717 | MT498037 | Unassigned | Unassigned | Unassigned | 859_0 | Clustered | 9 | VC_859_0 | 9 | 0.9426 | 1 | 0.9426 | 0 | 0 | 0 | 1 | Ref |
| 11718 | MT498038 | Unassigned | Unassigned | Unassigned | 1344_0 | Clustered | 78 | VC_1344_0 | 78 | 0.9732 | 1 | 0.9732 | 0 | 0 | 0 | 1 | Ref |
| 11719 | MT498039 | Unassigned | Unassigned | Unassigned | 799_1 | Clustered | 27 | VC_799_1 | 27 | 0.8611 | 1 | 0.8611 | 0 | 0 | 0 | 1 | Ref |
| 11720 | MT498040 | Unassigned | Unassigned | Unassigned | 1344_0 | Clustered | 78 | VC_1344_0 | 78 | 0.9732 | 1 | 0.9732 | 0 | 0 | 0 | 1 | Ref |
| 11721 | MT498041 | Unassigned | Unassigned | Unassigned | 216_6 | Clustered | 42 | VC_216_6 | 42 | 0.473 | 1 | 0.473 | 0 | 0 | 0 | 0.8351 | Ref |
| 11722 | MT498042 | Unassigned | Unassigned | Unassigned | 521_0 | Clustered | 37 | VC_521_0 | 37 | 0.912 | 1 | 0.912 | 0 | 0 | 0 | 1 | Ref |
| 11723 | MT498044 | Unassigned | Unassigned | Unassigned | 521_0 | Clustered | 37 | VC_521_0 | 37 | 0.912 | 1 | 0.912 | 0 | 0 | 0 | 1 | Ref |
| 11724 | MT498045 | Unassigned | Unassigned | Unassigned | 1160_0 | Clustered | 73 | VC_1160_0 | 73 | 0.9835 | 1 | 0.9835 | 0 | 0 | 0 | 1 | Ref |
| 11725 | MT498046 | Unassigned | Unassigned | Unassigned | 1344_0 | Clustered | 78 | VC_1344_0 | 78 | 0.9732 | 1 | 0.9732 | 0 | 0 | 0 | 1 | Ref |
| 11726 | MT498047 | Unassigned | Unassigned | Unassigned | 1344_0 | Clustered | 78 | VC_1344_0 | 78 | 0.9732 | 1 | 0.9732 | 0 | 0 | 0 | 1 | Ref |
| 11727 | MT498048 | Unassigned | Unassigned | Unassigned | 521_0 | Clustered | 37 | VC_521_0 | 37 | 0.912 | 1 | 0.912 | 0 | 0 | 0 | 1 | Ref |
| 11728 | MT498049 | Unassigned | Unassigned | Unassigned | 1240_0 | Clustered | 82 | VC_1240_0 | 82 | 0.7945 | 1 | 0.7945 | 0 | 0 | 0 | 1 | Ref |
| 11729 | MT498050 | Unassigned | Unassigned | Unassigned | 1160_0 | Clustered | 73 | VC_1160_0 | 73 | 0.9835 | 1 | 0.9835 | 0 | 0 | 0 | 1 | Ref |
| 11730 | MT498051 | Unassigned | Unassigned | Unassigned | 1344_0 | Clustered | 78 | VC_1344_0 | 78 | 0.9732 | 1 | 0.9732 | 0 | 0 | 0 | 1 | Ref |
| 11731 | MT498052 | Unassigned | Unassigned | Unassigned | 799_1 | Clustered | 27 | VC_799_1 | 27 | 0.8611 | 1 | 0.8611 | 0 | 0 | 0 | 1 | Ref |
| 11732 | MT498053 | Unassigned | Unassigned | Unassigned | 859_0 | Clustered | 9 | VC_859_0 | 9 | 0.9426 | 1 | 0.9426 | 0 | 0 | 0 | 1 | Ref |
| 11733 | MT498054 | Unassigned | Unassigned | Unassigned | 1181_1 | Clustered | 10 | VC_1181_1 | 10 | 0.5828 | 1 | 0.5828 | 0 | 0 | 0 | 1 | Ref |
| 11734 | MT498055 | Unassigned | Unassigned | Unassigned | 1075_0 | Clustered | 4 | VC_1075_0 | 4 | 0.1483 | 0.99998427 | 0.1483 | 0 | 0 | 0 | 1 | Ref |
| 11735 | MT498056 | Unassigned | Unassigned | Unassigned | 517_1 | Clustered | 108 | VC_517_1 | 108 | 0.8483 | 1 | 0.8483 | 0 | 0 | 0 | 0.9955 | Ref |
| 11736 | MT498057 | Unassigned | Unassigned | Unassigned | 1162_0 | Clustered | 10 | VC_1162_0 | 10 | 0.5715 | 1 | 0.5715 | 0 | 0 | 0 | 1 | Ref |
| 11737 | MT498059 | Unassigned | Unassigned | Unassigned | 1177_0 | Clustered | 14 | VC_1177_0 | 14 | 0.7847 | 1 | 0.7847 | 0 | 0 | 0 | 0.989 | Ref |
| 11738 | MT498060 | Unassigned | Unassigned | Unassigned | 216_6 | Clustered | 42 | VC_216_6 | 42 | 0.473 | 1 | 0.473 | 0 | 0 | 0 | 0.8351 | Ref |
| 11739 | MT498061 | Unassigned | Unassigned | Unassigned | 519_0 | Clustered | 39 | VC_519_0 | 39 | 0.8156 | 1 | 0.8156 | 0 | 0 | 0 | 1 | Ref |
| 11740 | MT498064 | Unassigned | Unassigned | Unassigned | 1360_0 | Clustered | 31 | VC_1360_0 | 31 | 0.8837 | 1 | 0.8837 | 0 | 0 | 0 | 1 | Ref |
| 11741 | MT498065 | Unassigned | Unassigned | Unassigned | 1344_0 | Clustered | 78 | VC_1344_0 | 78 | 0.9732 | 1 | 0.9732 | 0 | 0 | 0 | 1 | Ref |
| 11742 | MT498066 | Unassigned | Unassigned | Unassigned | 216_6 | Clustered | 42 | VC_216_6 | 42 | 0.473 | 1 | 0.473 | 0 | 0 | 0 | 0.8351 | Ref |
| 11743 | MT498067 | Unassigned | Unassigned | Unassigned | 1344_0 | Clustered | 78 | VC_1344_0 | 78 | 0.9732 | 1 | 0.9732 | 0 | 0 | 0 | 1 | Ref |
| 11744 | MT499896 | Unassigned | Unassigned | Unassigned | 133_33 | Clustered/Singleton | 1 | VC_133_33 | 1 | 0 | 1 | 0 | 0 | 0 | 0 | 1 | Ref |
| 11745 | MT499897 | Unassigned | Unassigned | Unassigned | 592_0 | Clustered | 50 | VC_592_0 | 50 | 0.7314 | 1 | 0.7314 | 0 | 0 | 0 | 1 | Ref |
| 11746 | MT499898 | Unassigned | Unassigned | Unassigned | 592_0 | Clustered | 50 | VC_592_0 | 50 | 0.7314 | 1 | 0.7314 | 0 | 0 | 0 | 1 | Ref |
| 11747 | MT500540 | Unassigned | Unassigned | Unassigned | 379_0 | Clustered | 5 | VC_379_0 | 5 | 0.4779 | 0.99999992 | 0.4779 | 0 | 0 | 0 | 0.9 | Ref |
| 11748 | MT501516 | Unassigned | Unassigned | Unassigned | 837_0 | Clustered | 29 | VC_837_0 | 29 | 0.9955 | 1 | 0.9955 | 0 | 0 | 0 | 1 | Ref |
| 11749 | MT511058 | Unassigned | Unassigned | Unassigned | 456_0 | Clustered | 62 | VC_456_0 | 62 | 0.9026 | 1 | 0.9026 | 0 | 0 | 0 | 1 | Ref |
| 11750 | MT514532 | Unassigned | Unassigned | Unassigned | 100_0 | Clustered | 15 | VC_100_0 | 15 | 0.6536 | 1 | 0.6536 | 0 | 0 | 0 | 1 | Ref |
| 11751 | MT515751 | Unassigned | Unassigned | Unassigned | 283_34 | Clustered | 4 | VC_283_34 | 4 | 0.0082 | 0.9997481 | 0.0082 | 0 | 0 | 0 | 1 | Ref |
| 11752 | MT515752 | Unassigned | Unassigned | Unassigned | 283_100 | Clustered | 2 | VC_283_100 | 2 | 0.0027 | 0.90891788 | 0.0024 | 0 | 0 | 0 | 1 | Ref |
| 11753 | MT515753 | Unassigned | Unassigned | Unassigned | 283_136 | Clustered | 2 | VC_283_136 | 2 | 0.0026 | 0.94116137 | 0.0024 | 0 | 0 | 0 | 1 | Ref |
| 11754 | MT515754 | Unassigned | Unassigned | Unassigned | 283_67 | Clustered | 2 | VC_283_67 | 2 | 0.0027 | 0.90726003 | 0.0025 | 0 | 0 | 0 | 1 | Ref |
| 11755 | MT515755 | Unassigned | Unassigned | Unassigned | 283_136 | Clustered | 2 | VC_283_136 | 2 | 0.0026 | 0.94116137 | 0.0024 | 0 | 0 | 0 | 1 | Ref |
| 11756 | MT515756 | Unassigned | Unassigned | Unassigned | 283_180 | Clustered/Singleton | 1 | VC_283_180 | 1 | 0 | 1 | 0 | 0 | 0 | 0 | 1 | Ref |
| 11757 | MT515757 | Unassigned | Unassigned | Unassigned | 386_0 | Clustered | 49 | VC_386_0 | 49 | 0.7941 | 1 | 0.7941 | 0 | 0 | 0 | 1 | Ref |
| 11758 | MT520979 | Unassigned | Unassigned | Unassigned | 695_0 | Clustered | 48 | VC_695_0 | 48 | 0.8162 | 1 | 0.8162 | 0 | 0 | 0 | 1 | Ref |
| 11759 | MT521990 | Unassigned | Unassigned | Unassigned | 1344_0 | Clustered | 78 | VC_1344_0 | 78 | 0.9732 | 1 | 0.9732 | 0 | 0 | 0 | 1 | Ref |
| 11760 | MT521994 | Unassigned | Unassigned | Unassigned | 799_1 | Clustered | 27 | VC_799_1 | 27 | 0.8611 | 1 | 0.8611 | 0 | 0 | 0 | 1 | Ref |
| 11761 | MT521995 | Unassigned | Unassigned | Unassigned | 522_1 | Clustered | 5 | VC_522_1 | 5 | 0.1017 | 0.99999997 | 0.1017 | 0 | 0 | 0 | 0.8 | Ref |
| 11762 | MT521996 | Unassigned | Unassigned | Unassigned | 567_0 | Clustered | 8 | VC_567_0 | 8 | 0.4935 | 1 | 0.4935 | 0 | 0 | 0 | 1 | Ref |
| 11763 | MT521997 | Unassigned | Unassigned | Unassigned | 410_0 | Clustered | 7 | VC_410_0 | 7 | 0.7905 | 1 | 0.7905 | 0 | 0 | 0 | 1 | Ref |
| 11764 | MT521998 | Unassigned | Unassigned | Unassigned | 1178_0 | Clustered | 24 | VC_1178_0 | 24 | 0.9524 | 1 | 0.9524 | 0 | 0 | 0 | 1 | Ref |
| 11765 | MT521999 | Unassigned | Unassigned | Unassigned | 685_0 | Clustered | 16 | VC_685_0 | 16 | 0.2972 | 1 | 0.2972 | 0 | 0 | 0 | 0.9167 | Ref |
| 11766 | MT522001 | Unassigned | Unassigned | Unassigned | 1343_0 | Clustered | 21 | VC_1343_0 | 21 | 0.9843 | 1 | 0.9843 | 0 | 0 | 0 | 1 | Ref |
| 11767 | MT522002 | Unassigned | Unassigned | Unassigned | 515_0 | Clustered | 16 | VC_515_0 | 16 | 0.3793 | 1 | 0.3793 | 0 | 0 | 0 | 1 | Ref |
| 11768 | MT522003 | Unassigned | Unassigned | Unassigned | 1343_0 | Clustered | 21 | VC_1343_0 | 21 | 0.9843 | 1 | 0.9843 | 0 | 0 | 0 | 1 | Ref |
| 11769 | MT522004 | Unassigned | Unassigned | Unassigned | 220_0 | Clustered | 4 | VC_220_0 | 4 | 0.0615 | 0.99998802 | 0.0615 | 0 | 0 | 0 | 1 | Ref |
| 11770 | MT522005 | Unassigned | Unassigned | Unassigned | 551_8 | Clustered | 19 | VC_551_8 | 19 | 0.1039 | 1 | 0.1039 | 0 | 0 | 0 | 0.8713 | Ref |
| 11771 | MT522006 | Unassigned | Unassigned | Unassigned | 1177_0 | Clustered | 14 | VC_1177_0 | 14 | 0.7847 | 1 | 0.7847 | 0 | 0 | 0 | 0.989 | Ref |
| 11772 | MT522007 | Unassigned | Unassigned | Unassigned | 224_0 | Clustered | 19 | VC_224_0 | 19 | 0.9416 | 1 | 0.9416 | 0 | 0 | 0 | 1 | Ref |
| 11773 | MT526905 | Unassigned | Unassigned | Unassigned | 283_100 | Clustered | 2 | VC_283_100 | 2 | 0.0027 | 0.90891788 | 0.0024 | 0 | 0 | 0 | 1 | Ref |
| 11774 | MT533174 | Unassigned | Unassigned | Unassigned | 290_1 | Clustered | 8 | VC_290_1 | 8 | 0.0757 | 1 | 0.0757 | 0 | 0 | 0 | 0.8571 | Ref |
| 11775 | MT536174 | Unassigned | Unassigned | Unassigned | 469_0 | Clustered | 2 | VC_469_0 | 2 | 0.1492 | 0.95658144 | 0.1427 | 0 | 0 | 0 | 1 | Ref |
| 11776 | MT542512 | Unassigned | Unassigned | Unassigned | 386_0 | Clustered | 49 | VC_386_0 | 49 | 0.7941 | 1 | 0.7941 | 0 | 0 | 0 | 1 | Ref |
| 11777 | MT542697 | Unassigned | Unassigned | Unassigned | 823_0 | Clustered | 124 | VC_823_0 | 124 | 0.9492 | 1 | 0.9492 | 0 | 0 | 0 | 1 | Ref |
| 11778 | MT543041 | Unassigned | Unassigned | Unassigned | 392_0 | Clustered | 126 | VC_392_0 | 126 | 0.9621 | 1 | 0.9621 | 0 | 0 | 0 | 1 | Ref |
| 11779 | MT552976 | Unassigned | Unassigned | Unassigned | 849_4 | Clustered | 2 | VC_849_4 | 2 | 0.0239 | 0.86481074 | 0.0206 | 0 | 0 | 0 | 1 | Ref |
| 11780 | MT553335 | Unassigned | Unassigned | Unassigned | 558_1 | Clustered | 2 | VC_558_1 | 2 | 0.0032 | 0.78405032 | 0.0025 | 0 | 0 | 0 | 0 | Ref |
| 11781 | MT553336 | Unassigned | Unassigned | Unassigned | 1176_0 | Clustered | 41 | VC_1176_0 | 41 | 0.9558 | 1 | 0.9558 | 0 | 0 | 0 | 0.9976 | Ref |
| 11782 | MT553337 | Unassigned | Unassigned | Unassigned | 219_0 | Clustered | 5 | VC_219_0 | 5 | 0.08 | 0.99999998 | 0.08 | 0 | 0 | 0 | 1 | Ref |
| 11783 | MT553338 | Unassigned | Unassigned | Unassigned | 517_1 | Clustered | 108 | VC_517_1 | 108 | 0.8483 | 1 | 0.8483 | 0 | 0 | 0 | 0.9955 | Ref |
| 11784 | MT553339 | Unassigned | Unassigned | Unassigned | 1240_0 | Clustered | 82 | VC_1240_0 | 82 | 0.7945 | 1 | 0.7945 | 0 | 0 | 0 | 1 | Ref |
| 11785 | MT553340 | Unassigned | Unassigned | Unassigned | 1163_0 | Clustered | 28 | VC_1163_0 | 28 | 0.4923 | 1 | 0.4923 | 0 | 0 | 0 | 1 | Ref |
| 11786 | MT553341 | Unassigned | Unassigned | Unassigned | 1240_0 | Clustered | 82 | VC_1240_0 | 82 | 0.7945 | 1 | 0.7945 | 0 | 0 | 0 | 1 | Ref |
| 11787 | MT553343 | Unassigned | Unassigned | Unassigned | 1240_0 | Clustered | 82 | VC_1240_0 | 82 | 0.7945 | 1 | 0.7945 | 0 | 0 | 0 | 1 | Ref |
| 11788 | MT553344 | Unassigned | Unassigned | Unassigned | 406_0 | Clustered | 11 | VC_406_0 | 11 | 0.9449 | 1 | 0.9449 | 0 | 0 | 0 | 1 | Ref |
| 11789 | MT553345 | Unassigned | Unassigned | Unassigned | 520_40 | Clustered | 4 | VC_520_40 | 4 | 0.0396 | 0.99998776 | 0.0396 | 0 | 0 | 0 | 1 | Ref |
| 11790 | MT553346 | Unassigned | Unassigned | Unassigned | 517_1 | Clustered | 108 | VC_517_1 | 108 | 0.8483 | 1 | 0.8483 | 0 | 0 | 0 | 0.9955 | Ref |
| 11791 | MT554104 | Unassigned | Unassigned | Unassigned | 149_18 | Clustered/Singleton | 1 | VC_149_18 | 1 | 0 | 1 | 0 | 0 | 0 | 0 | 1 | Ref |
| 11792 | MT559526 | Unassigned | Unassigned | Unassigned | 111_0 | Clustered | 91 | VC_111_0 | 91 | 0.9587 | 1 | 0.9587 | 0 | 0 | 0 | 1 | Ref |
| 11793 | MT559527 | Unassigned | Unassigned | Unassigned | 111_0 | Clustered | 91 | VC_111_0 | 91 | 0.9587 | 1 | 0.9587 | 0 | 0 | 0 | 1 | Ref |
| 11794 | MT559528 | Unassigned | Unassigned | Unassigned | 123_0 | Clustered | 23 | VC_123_0 | 23 | 0.817 | 1 | 0.817 | 0 | 0 | 0 | 1 | Ref |
| 11795 | MT560058 | Unassigned | Unassigned | Unassigned | 669_5 | Clustered/Singleton | 1 | VC_669_5 | 1 | 0 | 1 | 0 | 0 | 0 | 0 | 1 | Ref |
| 11796 | MT560059 | Unassigned | Unassigned | Unassigned | 849_4 | Clustered | 2 | VC_849_4 | 2 | 0.0239 | 0.86481074 | 0.0206 | 0 | 0 | 0 | 1 | Ref |
| 11797 | MT577844 | Unassigned | Unassigned | Unassigned | 190_4 | Clustered/Singleton | 1 | VC_190_4 | 1 | 0 | 1 | 0 | 0 | 0 | 0 | 1 | Ref |
| 11798 | MT580116 | Unassigned | Unassigned | Unassigned | 476_0 | Clustered | 19 | VC_476_0 | 19 | 0.7814 | 1 | 0.7814 | 0 | 0 | 0 | 1 | Ref |
| 11799 | MT580117 | Unassigned | Unassigned | Unassigned | 476_0 | Clustered | 19 | VC_476_0 | 19 | 0.7814 | 1 | 0.7814 | 0 | 0 | 0 | 1 | Ref |
| 11800 | MT580897 | Unassigned | Unassigned | Unassigned | 491_0 | Clustered | 36 | VC_491_0 | 36 | 0.5336 | 1 | 0.5336 | 0 | 0 | 0 | 0.9714 | Ref |
| 11801 | MT584805 | Unassigned | Unassigned | Unassigned | 884_0 | Clustered | 28 | VC_884_0 | 28 | 0.6211 | 1 | 0.6211 | 0 | 0 | 0 | 0.9841 | Ref |
| 11802 | MT585273 | Unassigned | Unassigned | Unassigned | 912_0 | Clustered | 76 | VC_912_0 | 76 | 0.979 | 1 | 0.979 | 0 | 0 | 0 | 1 | Ref |
| 11803 | MT585274 | Unassigned | Unassigned | Unassigned | 912_0 | Clustered | 76 | VC_912_0 | 76 | 0.979 | 1 | 0.979 | 0 | 0 | 0 | 1 | Ref |
| 11804 | MT585275 | Unassigned | Unassigned | Unassigned | 912_0 | Clustered | 76 | VC_912_0 | 76 | 0.979 | 1 | 0.979 | 0 | 0 | 0 | 1 | Ref |
| 11805 | MT585276 | Unassigned | Unassigned | Unassigned | 912_0 | Clustered | 76 | VC_912_0 | 76 | 0.979 | 1 | 0.979 | 0 | 0 | 0 | 1 | Ref |
| 11806 | MT585277 | Unassigned | Unassigned | Unassigned | 912_0 | Clustered | 76 | VC_912_0 | 76 | 0.979 | 1 | 0.979 | 0 | 0 | 0 | 1 | Ref |
| 11807 | MT585278 | Unassigned | Unassigned | Unassigned | 912_0 | Clustered | 76 | VC_912_0 | 76 | 0.979 | 1 | 0.979 | 0 | 0 | 0 | 1 | Ref |
| 11808 | MT585279 | Unassigned | Unassigned | Unassigned | 912_0 | Clustered | 76 | VC_912_0 | 76 | 0.979 | 1 | 0.979 | 0 | 0 | 0 | 1 | Ref |
| 11809 | MT585280 | Unassigned | Unassigned | Unassigned | 912_0 | Clustered | 76 | VC_912_0 | 76 | 0.979 | 1 | 0.979 | 0 | 0 | 0 | 1 | Ref |
| 11810 | MT585281 | Unassigned | Unassigned | Unassigned | 912_0 | Clustered | 76 | VC_912_0 | 76 | 0.979 | 1 | 0.979 | 0 | 0 | 0 | 1 | Ref |
| 11811 | MT585282 | Unassigned | Unassigned | Unassigned | 912_0 | Clustered | 76 | VC_912_0 | 76 | 0.979 | 1 | 0.979 | 0 | 0 | 0 | 1 | Ref |
| 11812 | MT585283 | Unassigned | Unassigned | Unassigned | 912_0 | Clustered | 76 | VC_912_0 | 76 | 0.979 | 1 | 0.979 | 0 | 0 | 0 | 1 | Ref |
| 11813 | MT585284 | Unassigned | Unassigned | Unassigned | 912_0 | Clustered | 76 | VC_912_0 | 76 | 0.979 | 1 | 0.979 | 0 | 0 | 0 | 1 | Ref |
| 11814 | MT585285 | Unassigned | Unassigned | Unassigned | 912_0 | Clustered | 76 | VC_912_0 | 76 | 0.979 | 1 | 0.979 | 0 | 0 | 0 | 1 | Ref |
| 11815 | MT585286 | Unassigned | Unassigned | Unassigned | 912_0 | Clustered | 76 | VC_912_0 | 76 | 0.979 | 1 | 0.979 | 0 | 0 | 0 | 1 | Ref |
| 11816 | MT585287 | Unassigned | Unassigned | Unassigned | 912_0 | Clustered | 76 | VC_912_0 | 76 | 0.979 | 1 | 0.979 | 0 | 0 | 0 | 1 | Ref |
| 11817 | MT585288 | Unassigned | Unassigned | Unassigned | 912_0 | Clustered | 76 | VC_912_0 | 76 | 0.979 | 1 | 0.979 | 0 | 0 | 0 | 1 | Ref |
| 11818 | MT585289 | Unassigned | Unassigned | Unassigned | 912_0 | Clustered | 76 | VC_912_0 | 76 | 0.979 | 1 | 0.979 | 0 | 0 | 0 | 1 | Ref |
| 11819 | MT585290 | Unassigned | Unassigned | Unassigned | 912_0 | Clustered | 76 | VC_912_0 | 76 | 0.979 | 1 | 0.979 | 0 | 0 | 0 | 1 | Ref |
| 11820 | MT585291 | Unassigned | Unassigned | Unassigned | 912_0 | Clustered | 76 | VC_912_0 | 76 | 0.979 | 1 | 0.979 | 0 | 0 | 0 | 1 | Ref |
| 11821 | MT585292 | Unassigned | Unassigned | Unassigned | 912_0 | Clustered | 76 | VC_912_0 | 76 | 0.979 | 1 | 0.979 | 0 | 0 | 0 | 1 | Ref |
| 11822 | MT585293 | Unassigned | Unassigned | Unassigned | 912_0 | Clustered | 76 | VC_912_0 | 76 | 0.979 | 1 | 0.979 | 0 | 0 | 0 | 1 | Ref |
| 11823 | MT585294 | Unassigned | Unassigned | Unassigned | 912_0 | Clustered | 76 | VC_912_0 | 76 | 0.979 | 1 | 0.979 | 0 | 0 | 0 | 1 | Ref |
| 11824 | MT585295 | Unassigned | Unassigned | Unassigned | 912_0 | Clustered | 76 | VC_912_0 | 76 | 0.979 | 1 | 0.979 | 0 | 0 | 0 | 1 | Ref |

| | | | | | | | | | | | | | | | | | |
|---|---|---|---|---|---|---|---|---|---|---|---|---|---|---|---|---|---|
| 11825 | MT585296 | Unassigned | Unassigned | Unassigned | 912_0 | Clustered | 76 | VC_912_0 | 76 | 0.979 | 1 | 0.979 | 0 | 0 | 0 | 1 | Ref |
| 11826 | MT585297 | Unassigned | Unassigned | Unassigned | 912_0 | Clustered | 76 | VC_912_0 | 76 | 0.979 | 1 | 0.979 | 0 | 0 | 0 | 1 | Ref |
| 11827 | MT585298 | Unassigned | Unassigned | Unassigned | 912_0 | Clustered | 76 | VC_912_0 | 76 | 0.979 | 1 | 0.979 | 0 | 0 | 0 | 1 | Ref |
| 11828 | MT585299 | Unassigned | Unassigned | Unassigned | 912_0 | Clustered | 76 | VC_912_0 | 76 | 0.979 | 1 | 0.979 | 0 | 0 | 0 | 1 | Ref |
| 11829 | MT585300 | Unassigned | Unassigned | Unassigned | 912_0 | Clustered | 76 | VC_912_0 | 76 | 0.979 | 1 | 0.979 | 0 | 0 | 0 | 1 | Ref |
| 11830 | MT585301 | Unassigned | Unassigned | Unassigned | 912_0 | Clustered | 76 | VC_912_0 | 76 | 0.979 | 1 | 0.979 | 0 | 0 | 0 | 1 | Ref |
| 11831 | MT585302 | Unassigned | Unassigned | Unassigned | 912_0 | Clustered | 76 | VC_912_0 | 76 | 0.979 | 1 | 0.979 | 0 | 0 | 0 | 1 | Ref |
| 11832 | MT585303 | Unassigned | Unassigned | Unassigned | 912_0 | Clustered | 76 | VC_912_0 | 76 | 0.979 | 1 | 0.979 | 0 | 0 | 0 | 1 | Ref |
| 11833 | MT585304 | Unassigned | Unassigned | Unassigned | 912_0 | Clustered | 76 | VC_912_0 | 76 | 0.979 | 1 | 0.979 | 0 | 0 | 0 | 1 | Ref |
| 11834 | MT585305 | Unassigned | Unassigned | Unassigned | 912_0 | Clustered | 76 | VC_912_0 | 76 | 0.979 | 1 | 0.979 | 0 | 0 | 0 | 1 | Ref |
| 11835 | MT585306 | Unassigned | Unassigned | Unassigned | 912_0 | Clustered | 76 | VC_912_0 | 76 | 0.979 | 1 | 0.979 | 0 | 0 | 0 | 1 | Ref |
| 11836 | MT585307 | Unassigned | Unassigned | Unassigned | 912_0 | Clustered | 76 | VC_912_0 | 76 | 0.979 | 1 | 0.979 | 0 | 0 | 0 | 1 | Ref |
| 11837 | MT585308 | Unassigned | Unassigned | Unassigned | 912_0 | Clustered | 76 | VC_912_0 | 76 | 0.979 | 1 | 0.979 | 0 | 0 | 0 | 1 | Ref |
| 11838 | MT585309 | Unassigned | Unassigned | Unassigned | 912_0 | Clustered | 76 | VC_912_0 | 76 | 0.979 | 1 | 0.979 | 0 | 0 | 0 | 1 | Ref |
| 11839 | MT585310 | Unassigned | Unassigned | Unassigned | 912_0 | Clustered | 76 | VC_912_0 | 76 | 0.979 | 1 | 0.979 | 0 | 0 | 0 | 1 | Ref |
| 11840 | MT585311 | Unassigned | Unassigned | Unassigned | 912_0 | Clustered | 76 | VC_912_0 | 76 | 0.979 | 1 | 0.979 | 0 | 0 | 0 | 1 | Ref |
| 11841 | MT585312 | Unassigned | Unassigned | Unassigned | 912_0 | Clustered | 76 | VC_912_0 | 76 | 0.979 | 1 | 0.979 | 0 | 0 | 0 | 1 | Ref |
| 11842 | MT585313 | Unassigned | Unassigned | Unassigned | 912_0 | Clustered | 76 | VC_912_0 | 76 | 0.979 | 1 | 0.979 | 0 | 0 | 0 | 1 | Ref |
| 11843 | MT585314 | Unassigned | Unassigned | Unassigned | 912_0 | Clustered | 76 | VC_912_0 | 76 | 0.979 | 1 | 0.979 | 0 | 0 | 0 | 1 | Ref |
| 11844 | MT585315 | Unassigned | Unassigned | Unassigned | 912_0 | Clustered | 76 | VC_912_0 | 76 | 0.979 | 1 | 0.979 | 0 | 0 | 0 | 1 | Ref |
| 11845 | MT586120 | Unassigned | Unassigned | Unassigned | 657_0 | Clustered | 3 | VC_657_0 | 3 | 0.0217 | 0.92528126 | 0.0201 | 0 | 0 | 0 | 1 | Ref |
| 11846 | MT588083 | Unassigned | Unassigned | Unassigned | 637_0 | Clustered | 115 | VC_637_0 | 115 | 0.731 | 1 | 0.731 | 0 | 0 | 0 | 0.8764 | Ref |
| 11847 | MT591577 | Unassigned | Unassigned | Unassigned | 50_0 | Clustered | 9 | VC_50_0 | 9 | 0.5531 | 1 | 0.5531 | 0 | 0 | 0 | 1 | Ref |
| 11848 | MT596499 | Unassigned | Unassigned | Unassigned | 667_0 | Clustered | 7 | VC_667_0 | 7 | 0.4101 | 1 | 0.4101 | 0 | 0 | 0 | 1 | Ref |
| 11849 | MT596501 | Unassigned | Unassigned | Unassigned | 150_0 | Clustered | 13 | VC_150_0 | 13 | 0.2417 | 1 | 0.2417 | 0 | 0 | 0 | 0.9872 | Ref |
| 11850 | MT596502 | Unassigned | Unassigned | Unassigned | 150_0 | Clustered | 13 | VC_150_0 | 13 | 0.2417 | 1 | 0.2417 | 0 | 0 | 0 | 0.9872 | Ref |
| 11851 | MT596503 | Unassigned | Unassigned | Unassigned | 150_0 | Clustered | 13 | VC_150_0 | 13 | 0.2417 | 1 | 0.2417 | 0 | 0 | 0 | 0.9872 | Ref |
| 11852 | MT596504 | Unassigned | Unassigned | Unassigned | 150_0 | Clustered | 13 | VC_150_0 | 13 | 0.2417 | 1 | 0.2417 | 0 | 0 | 0 | 0.9872 | Ref |
| 11853 | MT596505 | Unassigned | Unassigned | Unassigned | 150_0 | Clustered | 13 | VC_150_0 | 13 | 0.2417 | 1 | 0.2417 | 0 | 0 | 0 | 0.9872 | Ref |
| 11854 | MT596506 | Unassigned | Unassigned | Unassigned | 150_0 | Clustered | 13 | VC_150_0 | 13 | 0.2417 | 1 | 0.2417 | 0 | 0 | 0 | 0.9872 | Ref |
| 11855 | MT601270 | Unassigned | Unassigned | Unassigned | 87_0 | Clustered | 8 | VC_87_0 | 8 | 0.5545 | 1 | 0.5545 | 0 | 0 | 0 | 1 | Ref |
| 11856 | MT601271 | Unassigned | Unassigned | Unassigned | 87_0 | Clustered | 8 | VC_87_0 | 8 | 0.5545 | 1 | 0.5545 | 0 | 0 | 0 | 1 | Ref |
| 11857 | MT601272 | Unassigned | Unassigned | Unassigned | 89_1 | Clustered | 2 | VC_89_1 | 2 | 0.0447 | 0.9464689 | 0.0423 | 0 | 0 | 0 | 1 | Ref |
| 11858 | MT601273 | Unassigned | Unassigned | Unassigned | 89_0 | Clustered | 2 | VC_89_0 | 2 | 0.0451 | 0.92849471 | 0.0419 | 0 | 0 | 0 | 1 | Ref |
| 11859 | MT601274 | Unassigned | Unassigned | Unassigned | 89_0 | Clustered | 2 | VC_89_0 | 2 | 0.0451 | 0.92849471 | 0.0419 | 0 | 0 | 0 | 1 | Ref |
| 11860 | MT611523 | Unassigned | Unassigned | Unassigned | 290_0 | Clustered | 10 | VC_290_0 | 10 | 0.0981 | 1 | 0.0981 | 0 | 0 | 0 | 1 | Ref |
| 11861 | MT612988 | Unassigned | Unassigned | Unassigned | 531_0 | Clustered | 9 | VC_531_0 | 9 | 0.26 | 1 | 0.26 | 0 | 0 | 0 | 1 | Ref |
| 11862 | MT612989 | Unassigned | Unassigned | Unassigned | 531_0 | Clustered | 9 | VC_531_0 | 9 | 0.26 | 1 | 0.26 | 0 | 0 | 0 | 1 | Ref |
| 11863 | MT613935 | Unassigned | Unassigned | Unassigned | 125_0 | Clustered | 17 | VC_125_0 | 17 | 0.9244 | 1 | 0.9244 | 0 | 0 | 0 | 1 | Ref |
| 11864 | MT623546 | Unassigned | Unassigned | Unassigned | 700_0 | Clustered | 10 | VC_700_0 | 10 | 0.8076 | 1 | 0.8076 | 0 | 0 | 0 | 1 | Ref |
| 11865 | MT623578 | Unassigned | Unassigned | Unassigned | 793_0 | Clustered | 12 | VC_793_0 | 12 | 0.8531 | 1 | 0.8531 | 0 | 0 | 0 | 1 | Ref |
| 11866 | MT625440 | Unassigned | Unassigned | Unassigned | 386_0 | Clustered | 49 | VC_386_0 | 49 | 0.7941 | 1 | 0.7941 | 0 | 0 | 0 | 1 | Ref |
| 11867 | MT627482 | Unassigned | Unassigned | Unassigned | 695_0 | Clustered | 48 | VC_695_0 | 48 | 0.8162 | 1 | 0.8162 | 0 | 0 | 0 | 1 | Ref |
| 11868 | MT630408 | Unassigned | Unassigned | Unassigned | 283_155 | Clustered | 2 | VC_283_155 | 2 | 0.0027 | 0.9528577 | 0.0026 | 0 | 0 | 0 | 1 | Ref |
| 11869 | MT630433 | Unassigned | Unassigned | Unassigned | 830_0 | Clustered | 6 | VC_830_0 | 6 | 0.6536 | 1 | 0.6536 | 0 | 0 | 0 | 1 | Ref |
| 11870 | MT632017 | Unassigned | Unassigned | Unassigned | 637_0 | Clustered | 115 | VC_637_0 | 115 | 0.731 | 1 | 0.731 | 0 | 0 | 0 | 0.8764 | Ref |
| 11871 | MT632018 | Unassigned | Unassigned | Unassigned | 637_0 | Clustered | 115 | VC_637_0 | 115 | 0.731 | 1 | 0.731 | 0 | 0 | 0 | 0.8764 | Ref |
| 11872 | MT632019 | Unassigned | Unassigned | Unassigned | 637_0 | Clustered | 115 | VC_637_0 | 115 | 0.731 | 1 | 0.731 | 0 | 0 | 0 | 0.8764 | Ref |
| 11873 | MT632020 | Unassigned | Unassigned | Unassigned | 637_0 | Clustered | 115 | VC_637_0 | 115 | 0.731 | 1 | 0.731 | 0 | 0 | 0 | 0.8764 | Ref |
| 11874 | MT633129 | Unassigned | Unassigned | Unassigned | 699_0 | Clustered | 71 | VC_699_0 | 71 | 0.9352 | 1 | 0.9352 | 0 | 0 | 0 | 1 | Ref |
| 11875 | MT635598 | Unassigned | Unassigned | Unassigned | 1008_0 | Clustered | 4 | VC_1008_0 | 4 | 0.6602 | 0.99993542 | 0.6601 | 0 | 0 | 0 | 1 | Ref |
| 11876 | MT639639 | Unassigned | Unassigned | Unassigned | 570_0 | Clustered | 9 | VC_570_0 | 9 | 0.499 | 1 | 0.499 | 0 | 0 | 0 | 1 | Ref |
| 11877 | MT639640 | Unassigned | Unassigned | Unassigned | 1093_0 | Clustered | 40 | VC_1093_0 | 40 | 0.8754 | 1 | 0.8754 | 0 | 0 | 0 | 1 | Ref |
| 11878 | MT639641 | Unassigned | Unassigned | Unassigned | 1171_0 | Clustered | 21 | VC_1171_0 | 21 | 0.7125 | 1 | 0.7125 | 0 | 0 | 0 | 1 | Ref |
| 11879 | MT639642 | Unassigned | Unassigned | Unassigned | 1344_0 | Clustered | 78 | VC_1344_0 | 78 | 0.9732 | 1 | 0.9732 | 0 | 0 | 0 | 1 | Ref |
| 11880 | MT639643 | Unassigned | Unassigned | Unassigned | 1343_0 | Clustered | 21 | VC_1343_0 | 21 | 0.9843 | 1 | 0.9843 | 0 | 0 | 0 | 1 | Ref |
| 11881 | MT639644 | Unassigned | Unassigned | Unassigned | 325_61 | Clustered | 8 | VC_325_61 | 8 | 0.0395 | 1 | 0.0395 | 0 | 0 | 0 | 0.7857 | Ref |
| 11882 | MT639645 | Unassigned | Unassigned | Unassigned | 218_0 | Clustered | 3 | VC_218_0 | 3 | 0.0481 | 0.99860979 | 0.0481 | 0 | 0 | 0 | 1 | Ref |
| 11883 | MT639646 | Unassigned | Unassigned | Unassigned | 1345_1 | Clustered | 2 | VC_1345_1 | 2 | 0.0956 | 0.9561938 | 0.0914 | 0 | 0 | 0 | 1 | Ref |
| 11884 | MT639647 | Unassigned | Unassigned | Unassigned | 1344_0 | Clustered | 78 | VC_1344_0 | 78 | 0.9732 | 1 | 0.9732 | 0 | 0 | 0 | 1 | Ref |
| 11885 | MT639648 | Unassigned | Unassigned | Unassigned | 372_0 | Clustered | 24 | VC_372_0 | 24 | 0.5539 | 1 | 0.5539 | 0 | 0 | 0 | 1 | Ref |
| 11886 | MT639649 | Unassigned | Unassigned | Unassigned | 1234_0 | Clustered | 44 | VC_1234_0 | 44 | 1 | 1 | 1 | 0 | 0 | 0 | 1 | Ref |
| 11887 | MT639650 | Unassigned | Unassigned | Unassigned | 525_1 | Clustered | 5 | VC_525_1 | 5 | 0.0485 | 0.99999947 | 0.0485 | 0 | 0 | 0 | 0.9 | Ref |
| 11888 | MT639651 | Unassigned | Unassigned | Unassigned | 1178_0 | Clustered | 24 | VC_1178_0 | 24 | 0.9524 | 1 | 0.9524 | 0 | 0 | 0 | 1 | Ref |
| 11889 | MT639652 | Unassigned | Unassigned | Unassigned | 508_0 | Clustered | 32 | VC_508_0 | 32 | 0.906 | 1 | 0.906 | 0 | 0 | 0 | 1 | Ref |
| 11890 | MT639653 | Unassigned | Unassigned | Unassigned | 1207_0 | Clustered | 29 | VC_1207_0 | 29 | 0.8225 | 1 | 0.8225 | 0 | 0 | 0 | 1 | Ref |
| 11891 | MT639654 | Unassigned | Unassigned | Unassigned | 325_23 | Clustered | 17 | VC_325_23 | 17 | 0.0716 | 1 | 0.0716 | 0 | 0 | 0 | 0.6397 | Ref |
| 11892 | MT647605 | Unassigned | Unassigned | Unassigned | 486_0 | Clustered | 18 | VC_486_0 | 18 | 0.887 | 1 | 0.887 | 0 | 0 | 0 | 1 | Ref |
| 11893 | MT647607 | Unassigned | Unassigned | Unassigned | 765_0 | Clustered | 132 | VC_765_0 | 132 | 1 | 1 | 1 | 0 | 0 | 0 | 1 | Ref |
| 11894 | MT653137 | Unassigned | Unassigned | Unassigned | 637_0 | Clustered | 115 | VC_637_0 | 115 | 0.731 | 1 | 0.731 | 0 | 0 | 0 | 0.8764 | Ref |
| 11895 | MT653143 | Unassigned | Unassigned | Unassigned | 190_36 | Clustered/Singleton | 1 | VC_190_36 | 1 | 0 | 1 | 0 | 0 | 0 | 0 | 1 | Ref |
| 11896 | MT653144 | Unassigned | Unassigned | Unassigned | 637_0 | Clustered | 115 | VC_637_0 | 115 | 0.731 | 1 | 0.731 | 0 | 0 | 0 | 0.8764 | Ref |
| 11897 | MT653145 | Unassigned | Unassigned | Unassigned | 191_32 | Clustered | 12 | VC_191_32 | 12 | 0.0683 | 1 | 0.0683 | 0 | 0 | 0 | 0.7576 | Ref |
| 11898 | MT653146 | Unassigned | Unassigned | Unassigned | 190_37 | Clustered/Singleton | 1 | VC_190_37 | 1 | 0 | 1 | 0 | 0 | 0 | 0 | 1 | Ref |
| 11899 | MT657330 | Unassigned | Unassigned | Unassigned | 810_0 | Clustered | 18 | VC_810_0 | 18 | 0.9121 | 1 | 0.9121 | 0 | 0 | 0 | 1 | Ref |
| 11900 | MT657331 | Unassigned | Unassigned | Unassigned | 514_0 | Clustered | 75 | VC_514_0 | 75 | 0.9402 | 1 | 0.9402 | 0 | 0 | 0 | 1 | Ref |
| 11901 | MT657332 | Unassigned | Unassigned | Unassigned | 1344_0 | Clustered | 78 | VC_1344_0 | 78 | 0.9732 | 1 | 0.9732 | 0 | 0 | 0 | 1 | Ref |
| 11902 | MT657333 | Unassigned | Unassigned | Unassigned | 1171_0 | Clustered | 21 | VC_1171_0 | 21 | 0.7125 | 1 | 0.7125 | 0 | 0 | 0 | 1 | Ref |
| 11903 | MT657334 | Unassigned | Unassigned | Unassigned | 1360_0 | Clustered | 31 | VC_1360_0 | 31 | 0.8837 | 1 | 0.8837 | 0 | 0 | 0 | 1 | Ref |
| 11904 | MT657335 | Unassigned | Unassigned | Unassigned | 510_0 | Clustered | 12 | VC_510_0 | 12 | 0.3963 | 1 | 0.3963 | 0 | 0 | 0 | 0.9697 | Ref |
| 11905 | MT657336 | Unassigned | Unassigned | Unassigned | 1352_0 | Clustered | 31 | VC_1352_0 | 31 | 0.849 | 1 | 0.849 | 0 | 0 | 0 | 1 | Ref |
| 11906 | MT657337 | Unassigned | Unassigned | Unassigned | 1360_0 | Clustered | 31 | VC_1360_0 | 31 | 0.8837 | 1 | 0.8837 | 0 | 0 | 0 | 1 | Ref |
| 11907 | MT657338 | Unassigned | Unassigned | Unassigned | 223_0 | Clustered | 38 | VC_223_0 | 38 | 0.9095 | 1 | 0.9095 | 0 | 0 | 0 | 1 | Ref |
| 11908 | MT657339 | Unassigned | Unassigned | Unassigned | 1182_0 | Clustered | 4 | VC_1182_0 | 4 | 0.2845 | 0.99997676 | 0.2845 | 0 | 0 | 0 | 1 | Ref |
| 11909 | MT657340 | Unassigned | Unassigned | Unassigned | 872_0 | Clustered | 31 | VC_872_0 | 31 | 0.8151 | 1 | 0.8151 | 0 | 0 | 0 | 0.8194 | Ref |
| 11910 | MT657341 | Unassigned | Unassigned | Unassigned | 763_2 | Clustered | 7 | VC_763_2 | 7 | 0.2612 | 1 | 0.2612 | 0 | 0 | 0 | 1 | Ref |
| 11911 | MT657342 | Unassigned | Unassigned | Unassigned | 325_35 | Clustered | 17 | VC_325_35 | 17 | 0.0732 | 1 | 0.0732 | 0 | 0 | 0 | 0.7721 | Ref |
| 11912 | MT657343 | Unassigned | Unassigned | Unassigned | 1163_0 | Clustered | 28 | VC_1163_0 | 28 | 0.4923 | 1 | 0.4923 | 0 | 0 | 0 | 1 | Ref |
| 11913 | MT657344 | Unassigned | Unassigned | Unassigned | 1163_0 | Clustered | 28 | VC_1163_0 | 28 | 0.4923 | 1 | 0.4923 | 0 | 0 | 0 | 1 | Ref |
| 11914 | MT658802 | Unassigned | Unassigned | Unassigned | 763_2 | Clustered | 7 | VC_763_2 | 7 | 0.2612 | 1 | 0.2612 | 0 | 0 | 0 | 1 | Ref |
| 11915 | MT658803 | Unassigned | Unassigned | Unassigned | 763_1 | Clustered | 9 | VC_763_1 | 9 | 0.3706 | 1 | 0.3706 | 0 | 0 | 0 | 1 | Ref |
| 11916 | MT658804 | Unassigned | Unassigned | Unassigned | 216_6 | Clustered | 42 | VC_216_6 | 42 | 0.473 | 1 | 0.473 | 0 | 0 | 0 | 0.8351 | Ref |
| 11917 | MT658805 | Unassigned | Unassigned | Unassigned | 890_0 | Clustered | 3 | VC_890_0 | 3 | 0.3565 | 0.99830951 | 0.3559 | 0 | 0 | 0 | 1 | Ref |
| 11918 | MT661596 | Unassigned | Unassigned | Unassigned | 489_2 | Clustered/Singleton | 1 | VC_489_2 | 1 | 0 | 1 | 0 | 0 | 0 | 0 | 1 | Ref |
| 11919 | MT661597 | Unassigned | Unassigned | Unassigned | 695_0 | Clustered | 48 | VC_695_0 | 48 | 0.8162 | 1 | 0.8162 | 0 | 0 | 0 | 1 | Ref |
| 11920 | MT661598 | Unassigned | Unassigned | Unassigned | 695_0 | Clustered | 48 | VC_695_0 | 48 | 0.8162 | 1 | 0.8162 | 0 | 0 | 0 | 1 | Ref |
| 11921 | MT662115 | Unassigned | Unassigned | Unassigned | 5_0 | Clustered | 30 | VC_5_0 | 30 | 0.9647 | 1 | 0.9647 | 0 | 0 | 0 | 1 | Ref |
| 11922 | MT663719 | Unassigned | Unassigned | Unassigned | 428_0 | Clustered | 33 | VC_428_0 | 33 | 0.9915 | 1 | 0.9915 | 0 | 0 | 0 | 1 | Ref |
| 11923 | MT664722 | Unassigned | Unassigned | Unassigned | 812_1 | Clustered | 4 | VC_812_1 | 4 | 0.2266 | 0.99998426 | 0.2266 | 0 | 0 | 0 | 1 | Ref |
| 11924 | MT664984 | Unassigned | Unassigned | Unassigned | 169_0 | Clustered | 20 | VC_169_0 | 20 | 0.7529 | 1 | 0.7529 | 0 | 0 | 0 | 1 | Ref |
| 11925 | MT668623 | Unassigned | Unassigned | Unassigned | 739_0 | Clustered | 26 | VC_739_0 | 26 | 0.9571 | 1 | 0.9571 | 0 | 0 | 0 | 1 | Ref |
| 11926 | MT668624 | Unassigned | Unassigned | Unassigned | 739_0 | Clustered | 26 | VC_739_0 | 26 | 0.9571 | 1 | 0.9571 | 0 | 0 | 0 | 1 | Ref |
| 11927 | MT670417 | Unassigned | Unassigned | Unassigned | 78_3 | Clustered | 11 | VC_78_3 | 11 | 0.2572 | 1 | 0.2572 | 0 | 0 | 0 | 1 | Ref |
| 11928 | MT670418 | Unassigned | Unassigned | Unassigned | 78_3 | Clustered | 11 | VC_78_3 | 11 | 0.2572 | 1 | 0.2572 | 0 | 0 | 0 | 1 | Ref |
| 11929 | MT670419 | Unassigned | Unassigned | Unassigned | 78_3 | Clustered | 11 | VC_78_3 | 11 | 0.2572 | 1 | 0.2572 | 0 | 0 | 0 | 1 | Ref |
| 11930 | MT670420 | Unassigned | Unassigned | Unassigned | 78_3 | Clustered | 11 | VC_78_3 | 11 | 0.2572 | 1 | 0.2572 | 0 | 0 | 0 | 1 | Ref |
| 11931 | MT670421 | Unassigned | Unassigned | Unassigned | 78_3 | Clustered | 11 | VC_78_3 | 11 | 0.2572 | 1 | 0.2572 | 0 | 0 | 0 | 1 | Ref |
| 11932 | MT670422 | Unassigned | Unassigned | Unassigned | 78_3 | Clustered | 11 | VC_78_3 | 11 | 0.2572 | 1 | 0.2572 | 0 | 0 | 0 | 1 | Ref |
| 11933 | MT675124 | Unassigned | Unassigned | Unassigned | 592_0 | Clustered | 50 | VC_592_0 | 50 | 0.7314 | 1 | 0.7314 | 0 | 0 | 0 | 1 | Ref |
| 11934 | MT675125 | Unassigned | Unassigned | Unassigned | 592_0 | Clustered | 50 | VC_592_0 | 50 | 0.7314 | 1 | 0.7314 | 0 | 0 | 0 | 1 | Ref |
| 11935 | MT675126 | Unassigned | Unassigned | Unassigned | 592_0 | Clustered | 50 | VC_592_0 | 50 | 0.7314 | 1 | 0.7314 | 0 | 0 | 0 | 1 | Ref |
| 11936 | MT677933 | Unassigned | Unassigned | Unassigned | 637_1 | Clustered | 27 | VC_637_1 | 27 | 0.2578 | 1 | 0.2578 | 0 | 0 | 0 | 0.9858 | Ref |
| 11937 | MT677934 | Unassigned | Unassigned | Unassigned | 637_0 | Clustered | 115 | VC_637_0 | 115 | 0.731 | 1 | 0.731 | 0 | 0 | 0 | 0.8764 | Ref |
| 11938 | MT679221 | Unassigned | Unassigned | Unassigned | 959_5 | Clustered | 19 | VC_959_5 | 19 | 0.2626 | 1 | 0.2626 | 0 | 0 | 0 | 1 | Ref |
| 11939 | MT680615 | Unassigned | Unassigned | Unassigned | 452_0 | Clustered | 66 | VC_452_0 | 66 | 0.9248 | 1 | 0.9248 | 0 | 0 | 0 | 1 | Ref |
| 11940 | MT681669 | Unassigned | Unassigned | Unassigned | 392_0 | Clustered | 126 | VC_392_0 | 126 | 0.9621 | 1 | 0.9621 | 0 | 0 | 0 | 1 | Ref |
| 11941 | MT682064 | Unassigned | Unassigned | Unassigned | 460_0 | Clustered | 19 | VC_460_0 | 19 | 0.3744 | 1 | 0.3744 | 0 | 0 | 0 | 1 | Ref |
| 11942 | MT682065 | Unassigned | Unassigned | Unassigned | 111_0 | Clustered | 91 | VC_111_0 | 91 | 0.9587 | 1 | 0.9587 | 0 | 0 | 0 | 1 | Ref |
| 11943 | MT682386 | Unassigned | Unassigned | Unassigned | 878_0 | Clustered | 32 | VC_878_0 | 32 | 0.9965 | 1 | 0.9965 | 0 | 0 | 0 | 1 | Ref |
| 11944 | MT682387 | Unassigned | Unassigned | Unassigned | 878_0 | Clustered | 32 | VC_878_0 | 32 | 0.9965 | 1 | 0.9965 | 0 | 0 | 0 | 1 | Ref |
| 11945 | MT682388 | Unassigned | Unassigned | Unassigned | 878_0 | Clustered | 32 | VC_878_0 | 32 | 0.9965 | 1 | 0.9965 | 0 | 0 | 0 | 1 | Ref |
| 11946 | MT682389 | Unassigned | Unassigned | Unassigned | 878_0 | Clustered | 32 | VC_878_0 | 32 | 0.9965 | 1 | 0.9965 | 0 | 0 | 0 | 1 | Ref |
| 11947 | MT682390 | Unassigned | Unassigned | Unassigned | 878_0 | Clustered | 32 | VC_878_0 | 32 | 0.9965 | 1 | 0.9965 | 0 | 0 | 0 | 1 | Ref |
| 11948 | MT682391 | Unassigned | Unassigned | Unassigned | 878_0 | Clustered | 32 | VC_878_0 | 32 | 0.9965 | 1 | 0.9965 | 0 | 0 | 0 | 1 | Ref |
| 11949 | MT682705 | Unassigned | Unassigned | Unassigned | 722_0 | Clustered | 13 | VC_722_0 | 13 | 0.5104 | 1 | 0.5104 | 0 | 0 | 0 | 1 | Ref |
| 11950 | MT682706 | Unassigned | Unassigned | Unassigned | 456_0 | Clustered | 62 | VC_456_0 | 62 | 0.9026 | 1 | 0.9026 | 0 | 0 | 0 | 1 | Ref |
| 11951 | MT682707 | Unassigned | Unassigned | Unassigned | 451_0 | Clustered | 80 | VC_451_0 | 80 | 0.9991 | 1 | 0.9991 | 0 | 0 | 0 | 1 | Ref |
| 11952 | MT682708 | Unassigned | Unassigned | Unassigned | 779_0 | Clustered | 37 | VC_779_0 | 37 | 0.5682 | 1 | 0.5682 | 0 | 0 | 0 | 1 | Ref |
| 11953 | MT682709 | Unassigned | Unassigned | Unassigned | 283_171 | Clustered/Singleton | 1 | VC_283_171 | 1 | 0 | 1 | 0 | 0 | 0 | 0 | 1 | Ref |
| 11954 | MT682710 | Unassigned | Unassigned | Unassigned | 283_72 | Clustered/Singleton | 1 | VC_283_72 | 1 | 0 | 1 | 0 | 0 | 0 | 0 | 1 | Ref |
| 11955 | MT682711 | Unassigned | Unassigned | Unassigned | 290_5 | Clustered | 14 | VC_290_5 | 14 | 0.1422 | 1 | 0.1422 | 0 | 0 | 0 | 0.9231 | Ref |
| 11956 | MT682712 | Unassigned | Unassigned | Unassigned | 283_77 | Clustered | 2 | VC_283_77 | 2 | 0.0025 | 0.95407419 | 0.0024 | 0 | 0 | 0 | 1 | Ref |
| 11957 | MT682713 | Unassigned | Unassigned | Unassigned | 283_54 | Clustered/Singleton | 1 | VC_283_54 | 1 | 0 | 1 | 0 | 0 | 0 | 0 | 1 | Ref |
| 11958 | MT682714 | Unassigned | Unassigned | Unassigned | 283_74 | Clustered/Singleton | 1 | VC_283_74 | 1 | 0 | 1 | 0 | 0 | 0 | 0 | 1 | Ref |
| 11959 | MT682715 | Unassigned | Unassigned | Unassigned | 575_0 | Clustered | 35 | VC_575_0 | 35 | 0.8815 | 1 | 0.8815 | 0 | 0 | 0 | 1 | Ref |
| 11960 | MT682716 | Unassigned | Unassigned | Unassigned | 772_20 | Clustered | 2 | VC_772_20 | 2 | 0.006 | 0.58868587 | 0.0035 | 0 | 0 | 0 | 1 | Ref |
| 11961 | MT684586 | Unassigned | Unassigned | Unassigned | 520_47 | Clustered/Singleton | 1 | VC_520_47 | 1 | 0 | 1 | 0 | 0 | 0 | 0 | 1 | Ref |
| 11962 | MT684587 | Unassigned | Unassigned | Unassigned | 1360_0 | Clustered | 31 | VC_1360_0 | 31 | 0.8837 | 1 | 0.8837 | 0 | 0 | 0 | 1 | Ref |
| 11963 | MT684588 | Unassigned | Unassigned | Unassigned | 521_0 | Clustered | 37 | VC_521_0 | 37 | 0.912 | 1 | 0.912 | 0 | 0 | 0 | 1 | Ref |
| 11964 | MT684589 | Unassigned | Unassigned | Unassigned | 872_0 | Clustered | 31 | VC_872_0 | 31 | 0.8151 | 1 | 0.8151 | 0 | 0 | 0 | 0.8194 | Ref |
| 11965 | MT684590 | Unassigned | Unassigned | Unassigned | 1071_2 | Clustered | 5 | VC_1071_2 | 5 | 0.0534 | 0.99997362 | 0.0534 | 0 | 0 | 0 | 0.9 | Ref |
| 11966 | MT684591 | Unassigned | Unassigned | Unassigned | 1343_0 | Clustered | 21 | VC_1343_0 | 21 | 0.9843 | 1 | 0.9843 | 0 | 0 | 0 | 1 | Ref |
| 11967 | MT684592 | Unassigned | Unassigned | Unassigned | 1343_0 | Clustered | 21 | VC_1343_0 | 21 | 0.9843 | 1 | 0.9843 | 0 | 0 | 0 | 1 | Ref |
| 11968 | MT684593 | Unassigned | Unassigned | Unassigned | 520_15 | Clustered | 2 | VC_520_15 | 2 | 0.0078 | 0.957604 | 0.0075 | 0 | 0 | 0 | 0 | Ref |
| 11969 | MT684594 | Unassigned | Unassigned | Unassigned | 810_0 | Clustered | 18 | VC_810_0 | 18 | 0.9121 | 1 | 0.9121 | 0 | 0 | 0 | 1 | Ref |
| 11970 | MT684595 | Unassigned | Unassigned | Unassigned | 520_14 | Clustered/Singleton | 1 | VC_520_14 | 1 | 0 | 1 | 0 | 0 | 0 | 0 | 1 | Ref |
| 11971 | MT684596 | Unassigned | Unassigned | Unassigned | 1224_0 | Clustered | 19 | VC_1224_0 | 19 | 1 | 1 | 1 | 0 | 0 | 0 | 1 | Ref |
| 11972 | MT684597 | Unassigned | Unassigned | Unassigned | 520_6 | Clustered | 3 | VC_520_6 | 3 | 0.0276 | 0.99859841 | 0.0276 | 0 | 0 | 0 | 1 | Ref |
| 11973 | MT684598 | Unassigned | Unassigned | Unassigned | 1072_5 | Clustered | 3 | VC_1072_5 | 3 | 0.0475 | 0.98997321 | 0.047 | 0 | 0 | 0 | 1 | Ref |
| 11974 | MT684599 | Unassigned | Unassigned | Unassigned | 890_1 | Clustered | 4 | VC_890_1 | 4 | 0.4613 | 0.99998458 | 0.4613 | 0 | 0 | 0 | 1 | Ref |
| 11975 | MT684600 | Unassigned | Unassigned | Unassigned | 872_0 | Clustered | 31 | VC_872_0 | 31 | 0.8151 | 1 | 0.8151 | 0 | 0 | 0 | 0.8194 | Ref |
| 11976 | MT684601 | Unassigned | Unassigned | Unassigned | 520_6 | Clustered | 3 | VC_520_6 | 3 | 0.0276 | 0.99859841 | 0.0276 | 0 | 0 | 0 | 1 | Ref |
| 11977 | MT684602 | Unassigned | Unassigned | Unassigned | 568_0 | Clustered | 8 | VC_568_0 | 8 | 0.5738 | 1 | 0.5738 | 0 | 0 | 0 | 1 | Ref |
| 11978 | MT701586 | Unassigned | Unassigned | Unassigned | 125_0 | Clustered | 17 | VC_125_0 | 17 | 0.9244 | 1 | 0.9244 | 0 | 0 | 0 | 1 | Ref |
| 11979 | MT701587 | Unassigned | Unassigned | Unassigned | 242_1 | Clustered | 20 | VC_242_1 | 20 | 0.684 | 1 | 0.684 | 0 | 0 | 0 | 0.8368 | Ref |
| 11980 | MT701588 | Unassigned | Unassigned | Unassigned | 491_0 | Clustered | 36 | VC_491_0 | 36 | 0.5336 | 1 | 0.5336 | 0 | 0 | 0 | 0.9714 | Ref |
| 11981 | MT701589 | Unassigned | Unassigned | Unassigned | 111_0 | Clustered | 91 | VC_111_0 | 91 | 0.9587 | 1 | 0.9587 | 0 | 0 | 0 | 1 | Ref |
| 11982 | MT701591 | Unassigned | Unassigned | Unassigned | 1_0 | Clustered | 54 | VC_1_0 | 54 | 0.8527 | 1 | 0.8527 | 0 | 0 | 0 | 1 | Ref |
| 11983 | MT701592 | Unassigned | Unassigned | Unassigned | 937_0 | Clustered | 40 | VC_937_0 | 40 | 0.6809 | 1 | 0.6809 | 0 | 0 | 0 | 0.9359 | Ref |
| 11984 | MT701593 | Unassigned | Unassigned | Unassigned | 376_0 | Clustered | 3 | VC_376_0 | 3 | 0.2389 | 0.99789919 | 0.2384 | 0 | 0 | 0 | 1 | Ref |
| 11985 | MT701594 | Unassigned | Unassigned | Unassigned | 981_1 | Clustered | 26 | VC_981_1 | 26 | 0.7009 | 1 | 0.7009 | 0 | 0 | 0 | 1 | Ref |
| 11986 | MT701595 | Unassigned | Unassigned | Unassigned | 376_0 | Clustered | 3 | VC_376_0 | 3 | 0.2389 | 0.99789919 | 0.2384 | 0 | 0 | 0 | 1 | Ref |

| | | | | | | | | | | | | | | | | | |
|---|---|---|---|---|---|---|---|---|---|---|---|---|---|---|---|---|---|
| 11987 | MT701596 | Unassigned | Unassigned | Unassigned | 376_0 | Clustered | 3 | VC_376_0 | 3 | 0.2389 | 0.99789919 | 0.2384 | 0 | 0 | 0 | 1 | Ref |
| 11988 | MT701597 | Unassigned | Unassigned | Unassigned | 981_1 | Clustered | 26 | VC_981_1 | 26 | 0.7009 | 1 | 0.7009 | 0 | 0 | 0 | 1 | Ref |
| 11989 | MT701598 | Unassigned | Unassigned | Unassigned | 982_0 | Clustered | 62 | VC_982_0 | 62 | 0.8646 | 1 | 0.8646 | 0 | 0 | 0 | 0.743 | Ref |
| 11990 | MT708544 | Unassigned | Unassigned | Unassigned | 987_0 | Clustered | 22 | VC_987_0 | 22 | 0.8955 | 1 | 0.8955 | 0 | 0 | 0 | 1 | Ref |
| 11991 | MT708545 | Unassigned | Unassigned | Unassigned | 481_0 | Clustered | 8 | VC_481_0 | 8 | 0.1348 | 1 | 0.1348 | 0 | 0 | 0 | 1 | Ref |
| 11992 | MT708546 | Unassigned | Unassigned | Unassigned | 987_0 | Clustered | 22 | VC_987_0 | 22 | 0.8955 | 1 | 0.8955 | 0 | 0 | 0 | 1 | Ref |
| 11993 | MT708547 | Unassigned | Unassigned | Unassigned | 164_10 | Clustered | 5 | VC_164_10 | 5 | 0.0615 | 0.96483582 | 0.0593 | 0 | 0 | 0 | 1 | Ref |
| 11994 | MT708548 | Unassigned | Unassigned | Unassigned | 981_1 | Clustered | 26 | VC_981_1 | 26 | 0.7009 | 1 | 0.7009 | 0 | 0 | 0 | 1 | Ref |
| 11995 | MT708549 | Unassigned | Unassigned | Unassigned | 536_0 | Clustered | 3 | VC_536_0 | 3 | 0.0878 | 0.9985359 | 0.0877 | 0 | 0 | 0 | 1 | Ref |
| 11996 | MT708550 | Unassigned | Unassigned | Unassigned | 125_0 | Clustered | 17 | VC_125_0 | 17 | 0.9244 | 1 | 0.9244 | 0 | 0 | 0 | 1 | Ref |
| 11997 | MT710307 | Unassigned | Unassigned | Unassigned | 959_1 | Clustered | 8 | VC_959_1 | 8 | 0.0878 | 0.98435099 | 0.0864 | 0 | 0 | 0 | 1 | Ref |
| 11998 | MT711523 | Unassigned | Unassigned | Unassigned | 638_0 | Clustered | 45 | VC_638_0 | 45 | 0.7741 | 1 | 0.7741 | 0 | 0 | 0 | 1 | Ref |
| 11999 | MT711524 | Unassigned | Unassigned | Unassigned | 638_0 | Clustered | 45 | VC_638_0 | 45 | 0.7741 | 1 | 0.7741 | 0 | 0 | 0 | 1 | Ref |
| 12000 | MT711525 | Unassigned | Unassigned | Unassigned | 638_0 | Clustered | 45 | VC_638_0 | 45 | 0.7741 | 1 | 0.7741 | 0 | 0 | 0 | 1 | Ref |
| 12001 | MT711526 | Unassigned | Unassigned | Unassigned | 638_0 | Clustered | 45 | VC_638_0 | 45 | 0.7741 | 1 | 0.7741 | 0 | 0 | 0 | 1 | Ref |
| 12002 | MT711527 | Unassigned | Unassigned | Unassigned | 638_0 | Clustered | 45 | VC_638_0 | 45 | 0.7741 | 1 | 0.7741 | 0 | 0 | 0 | 1 | Ref |
| 12003 | MT711528 | Unassigned | Unassigned | Unassigned | 638_0 | Clustered | 45 | VC_638_0 | 45 | 0.7741 | 1 | 0.7741 | 0 | 0 | 0 | 1 | Ref |
| 12004 | MT711887 | Unassigned | Unassigned | Unassigned | 130_0 | Clustered | 19 | VC_130_0 | 19 | 0.6068 | 1 | 0.6068 | 0 | 0 | 0 | 1 | Ref |
| 12005 | MT711888 | Unassigned | Unassigned | Unassigned | 127_2 | Clustered/Singleton | 1 | VC_127_2 | 1 | 0 | 1 | 0 | 0 | 0 | 0 | 1 | Ref |
| 12006 | MT711889 | Unassigned | Unassigned | Unassigned | 129_0 | Clustered | 43 | VC_129_0 | 43 | 0.886 | 1 | 0.886 | 0 | 0 | 0 | 1 | Ref |
| 12007 | MT711890 | Unassigned | Unassigned | Unassigned | 129_0 | Clustered | 43 | VC_129_0 | 43 | 0.886 | 1 | 0.886 | 0 | 0 | 0 | 1 | Ref |
| 12008 | MT711975 | Unassigned | Unassigned | Unassigned | 727_0 | Clustered | 11 | VC_727_0 | 11 | 0.5054 | 1 | 0.5054 | 0 | 0 | 0 | 1 | Ref |
| 12009 | MT711976 | Unassigned | Unassigned | Unassigned | 1071_5 | Clustered/Singleton | 1 | VC_1071_5 | 1 | 0 | 1 | 0 | 0 | 0 | 0 | 1 | Ref |
| 12010 | MT711978 | Unassigned | Unassigned | Unassigned | 982_0 | Clustered | 62 | VC_982_0 | 62 | 0.8646 | 1 | 0.8646 | 0 | 0 | 0 | 0.743 | Ref |
| 12011 | MT711979 | Unassigned | Unassigned | Unassigned | 982_0 | Clustered | 62 | VC_982_0 | 62 | 0.8646 | 1 | 0.8646 | 0 | 0 | 0 | 0.743 | Ref |
| 12012 | MT713136 | Unassigned | Unassigned | Unassigned | 443_2 | Clustered | 10 | VC_443_2 | 10 | 0.3344 | 1 | 0.3344 | 0 | 0 | 0 | 0.9333 | Ref |
| 12013 | MT720689 | Unassigned | Unassigned | Unassigned | 909_0 | Clustered | 5 | VC_909_0 | 5 | 0.7406 | 0.9999861 | 0.7406 | 0 | 0 | 0 | 1 | Ref |
| 12014 | MT723932 | Unassigned | Unassigned | Unassigned | 521_0 | Clustered | 37 | VC_521_0 | 37 | 0.912 | 1 | 0.912 | 0 | 0 | 0 | 1 | Ref |
| 12015 | MT723933 | Unassigned | Unassigned | Unassigned | 728_0 | Clustered | 3 | VC_728_0 | 3 | 0.4017 | 0.99850259 | 0.4011 | 0 | 0 | 0 | 1 | Ref |
| 12016 | MT723934 | Unassigned | Unassigned | Unassigned | 1093_0 | Clustered | 40 | VC_1093_0 | 40 | 0.8754 | 1 | 0.8754 | 0 | 0 | 0 | 1 | Ref |
| 12017 | MT723935 | Unassigned | Unassigned | Unassigned | 1176_0 | Clustered | 41 | VC_1176_0 | 41 | 0.9558 | 1 | 0.9558 | 0 | 0 | 0 | 0.9976 | Ref |
| 12018 | MT723936 | Unassigned | Unassigned | Unassigned | 1176_0 | Clustered | 41 | VC_1176_0 | 41 | 0.9558 | 1 | 0.9558 | 0 | 0 | 0 | 0.9976 | Ref |
| 12019 | MT723937 | Unassigned | Unassigned | Unassigned | 1093_0 | Clustered | 40 | VC_1093_0 | 40 | 0.8754 | 1 | 0.8754 | 0 | 0 | 0 | 1 | Ref |
| 12020 | MT723938 | Unassigned | Unassigned | Unassigned | 525_0 | Clustered | 25 | VC_525_0 | 25 | 0.4597 | 1 | 0.4597 | 0 | 0 | 0 | 1 | Ref |
| 12021 | MT723939 | Unassigned | Unassigned | Unassigned | 517_1 | Clustered | 108 | VC_517_1 | 108 | 0.8483 | 1 | 0.8483 | 0 | 0 | 0 | 0.9955 | Ref |
| 12022 | MT723940 | Unassigned | Unassigned | Unassigned | 237_1 | Clustered | 13 | VC_237_1 | 13 | 0.1773 | 1 | 0.1773 | 0 | 0 | 0 | 0.7308 | Ref |
| 12023 | MT723941 | Unassigned | Unassigned | Unassigned | 1093_0 | Clustered | 40 | VC_1093_0 | 40 | 0.8754 | 1 | 0.8754 | 0 | 0 | 0 | 1 | Ref |
| 12024 | MT723942 | Unassigned | Unassigned | Unassigned | 1176_0 | Clustered | 41 | VC_1176_0 | 41 | 0.9558 | 1 | 0.9558 | 0 | 0 | 0 | 0.9976 | Ref |
| 12025 | MT723943 | Unassigned | Unassigned | Unassigned | 551_0 | Clustered | 16 | VC_551_0 | 16 | 0.0915 | 1 | 0.0915 | 0 | 0 | 0 | 0.6667 | Ref |
| 12026 | MT723944 | Unassigned | Unassigned | Unassigned | 558_39 | Clustered | 3 | VC_558_39 | 3 | 0.0063 | 0.73849574 | 0.0047 | 0 | 0 | 0 | 0.6667 | Ref |
| 12027 | MT723945 | Unassigned | Unassigned | Unassigned | 558_21 | Clustered/Singleton | 1 | VC_558_21 | 1 | 0 | 1 | 0 | 0 | 0 | 0 | 1 | Ref |
| 12028 | MT732432 | Unassigned | Unassigned | Unassigned | 1019_0 | Clustered | 5 | VC_1019_0 | 5 | 0.866 | 0.99999999 | 0.866 | 0 | 0 | 0 | 1 | Ref |
| 12029 | MT732433 | Unassigned | Unassigned | Unassigned | 1019_0 | Clustered | 5 | VC_1019_0 | 5 | 0.866 | 0.99999999 | 0.866 | 0 | 0 | 0 | 1 | Ref |
| 12030 | MT732434 | Unassigned | Unassigned | Unassigned | 1019_0 | Clustered | 5 | VC_1019_0 | 5 | 0.866 | 0.99999999 | 0.866 | 0 | 0 | 0 | 1 | Ref |
| 12031 | MT732435 | Unassigned | Unassigned | Unassigned | 1016_0 | Clustered | 8 | VC_1016_0 | 8 | 0.9481 | 1 | 0.9481 | 0 | 0 | 0 | 1 | Ref |
| 12032 | MT732436 | Unassigned | Unassigned | Unassigned | 1016_0 | Clustered | 8 | VC_1016_0 | 8 | 0.9481 | 1 | 0.9481 | 0 | 0 | 0 | 1 | Ref |
| 12033 | MT732437 | Unassigned | Unassigned | Unassigned | 1016_0 | Clustered | 8 | VC_1016_0 | 8 | 0.9481 | 1 | 0.9481 | 0 | 0 | 0 | 1 | Ref |
| 12034 | MT732438 | Unassigned | Unassigned | Unassigned | 1016_0 | Clustered | 8 | VC_1016_0 | 8 | 0.9481 | 1 | 0.9481 | 0 | 0 | 0 | 1 | Ref |
| 12035 | MT732439 | Unassigned | Unassigned | Unassigned | 1016_0 | Clustered | 8 | VC_1016_0 | 8 | 0.9481 | 1 | 0.9481 | 0 | 0 | 0 | 1 | Ref |
| 12036 | MT732440 | Unassigned | Unassigned | Unassigned | 1016_0 | Clustered | 8 | VC_1016_0 | 8 | 0.9481 | 1 | 0.9481 | 0 | 0 | 0 | 1 | Ref |
| 12037 | MT732441 | Unassigned | Unassigned | Unassigned | 1016_0 | Clustered | 8 | VC_1016_0 | 8 | 0.9481 | 1 | 0.9481 | 0 | 0 | 0 | 1 | Ref |
| 12038 | MT732442 | Unassigned | Unassigned | Unassigned | 1016_0 | Clustered | 8 | VC_1016_0 | 8 | 0.9481 | 1 | 0.9481 | 0 | 0 | 0 | 1 | Ref |
| 12039 | MT732443 | Unassigned | Unassigned | Unassigned | 445_0 | Clustered | 2 | VC_445_0 | 2 | 0.3978 | 0.92263554 | 0.3671 | 0 | 0 | 0 | 1 | Ref |
| 12040 | MT732444 | Unassigned | Unassigned | Unassigned | 445_0 | Clustered | 2 | VC_445_0 | 2 | 0.3978 | 0.92263554 | 0.3671 | 0 | 0 | 0 | 1 | Ref |
| 12041 | MT732445 | Unassigned | Unassigned | Unassigned | 1011_0 | Clustered | 9 | VC_1011_0 | 9 | 1 | 1 | 1 | 0 | 0 | 0 | 1 | Ref |
| 12042 | MT732446 | Unassigned | Unassigned | Unassigned | 1011_0 | Clustered | 9 | VC_1011_0 | 9 | 1 | 1 | 1 | 0 | 0 | 0 | 1 | Ref |
| 12043 | MT732447 | Unassigned | Unassigned | Unassigned | 1011_0 | Clustered | 9 | VC_1011_0 | 9 | 1 | 1 | 1 | 0 | 0 | 0 | 1 | Ref |
| 12044 | MT732448 | Unassigned | Unassigned | Unassigned | 1011_0 | Clustered | 9 | VC_1011_0 | 9 | 1 | 1 | 1 | 0 | 0 | 0 | 1 | Ref |
| 12045 | MT732449 | Unassigned | Unassigned | Unassigned | 1011_0 | Clustered | 9 | VC_1011_0 | 9 | 1 | 1 | 1 | 0 | 0 | 0 | 1 | Ref |
| 12046 | MT732450 | Unassigned | Unassigned | Unassigned | 1438_0 | Clustered | 6 | VC_1438_0 | 6 | 1 | 0 | 0 | 0 | 0 | 0 | 1 | Ref |
| 12047 | MT732451 | Unassigned | Unassigned | Unassigned | 1438_0 | Clustered | 6 | VC_1438_0 | 6 | 1 | 0 | 0 | 0 | 0 | 0 | 1 | Ref |
| 12048 | MT732452 | Unassigned | Unassigned | Unassigned | 1438_0 | Clustered | 6 | VC_1438_0 | 6 | 1 | 0 | 0 | 0 | 0 | 0 | 1 | Ref |
| 12049 | MT732453 | Unassigned | Unassigned | Unassigned | 1438_0 | Clustered | 6 | VC_1438_0 | 6 | 1 | 0 | 0 | 0 | 0 | 0 | 1 | Ref |
| 12050 | MT732454 | Unassigned | Unassigned | Unassigned | 1438_0 | Clustered | 6 | VC_1438_0 | 6 | 1 | 0 | 0 | 0 | 0 | 0 | 1 | Ref |
| 12051 | MT732455 | Unassigned | Unassigned | Unassigned | 1438_0 | Clustered | 6 | VC_1438_0 | 6 | 1 | 0 | 0 | 0 | 0 | 0 | 1 | Ref |
| 12052 | MT732458 | Unassigned | Unassigned | Unassigned | 1417_0 | Clustered | 15 | VC_1417_0 | 15 | 0.9982 | 1 | 0.9982 | 0 | 0 | 0 | 1 | Ref |
| 12053 | MT732459 | Unassigned | Unassigned | Unassigned | 1417_0 | Clustered | 15 | VC_1417_0 | 15 | 0.9982 | 1 | 0.9982 | 0 | 0 | 0 | 1 | Ref |
| 12054 | MT732460 | Unassigned | Unassigned | Unassigned | 1417_0 | Clustered | 15 | VC_1417_0 | 15 | 0.9982 | 1 | 0.9982 | 0 | 0 | 0 | 1 | Ref |
| 12055 | MT732461 | Unassigned | Unassigned | Unassigned | 1417_0 | Clustered | 15 | VC_1417_0 | 15 | 0.9982 | 1 | 0.9982 | 0 | 0 | 0 | 1 | Ref |
| 12056 | MT732462 | Unassigned | Unassigned | Unassigned | 1417_0 | Clustered | 15 | VC_1417_0 | 15 | 0.9982 | 1 | 0.9982 | 0 | 0 | 0 | 1 | Ref |
| 12057 | MT732463 | Unassigned | Unassigned | Unassigned | 1417_0 | Clustered | 15 | VC_1417_0 | 15 | 0.9982 | 1 | 0.9982 | 0 | 0 | 0 | 1 | Ref |
| 12058 | MT732464 | Unassigned | Unassigned | Unassigned | 1417_0 | Clustered | 15 | VC_1417_0 | 15 | 0.9982 | 1 | 0.9982 | 0 | 0 | 0 | 1 | Ref |
| 12059 | MT732465 | Unassigned | Unassigned | Unassigned | 1417_0 | Clustered | 15 | VC_1417_0 | 15 | 0.9982 | 1 | 0.9982 | 0 | 0 | 0 | 1 | Ref |
| 12060 | MT732466 | Unassigned | Unassigned | Unassigned | 1417_0 | Clustered | 15 | VC_1417_0 | 15 | 0.9982 | 1 | 0.9982 | 0 | 0 | 0 | 1 | Ref |
| 12061 | MT732467 | Unassigned | Unassigned | Unassigned | 1417_0 | Clustered | 15 | VC_1417_0 | 15 | 0.9982 | 1 | 0.9982 | 0 | 0 | 0 | 1 | Ref |
| 12062 | MT732468 | Unassigned | Unassigned | Unassigned | 1417_0 | Clustered | 15 | VC_1417_0 | 15 | 0.9982 | 1 | 0.9982 | 0 | 0 | 0 | 1 | Ref |
| 12063 | MT732469 | Unassigned | Unassigned | Unassigned | 1417_0 | Clustered | 15 | VC_1417_0 | 15 | 0.9982 | 1 | 0.9982 | 0 | 0 | 0 | 1 | Ref |
| 12064 | MT732470 | Unassigned | Unassigned | Unassigned | 1417_0 | Clustered | 15 | VC_1417_0 | 15 | 0.9982 | 1 | 0.9982 | 0 | 0 | 0 | 1 | Ref |
| 12065 | MT732471 | Unassigned | Unassigned | Unassigned | 1417_0 | Clustered | 15 | VC_1417_0 | 15 | 0.9982 | 1 | 0.9982 | 0 | 0 | 0 | 1 | Ref |
| 12066 | MT732472 | Unassigned | Unassigned | Unassigned | 1417_0 | Clustered | 15 | VC_1417_0 | 15 | 0.9982 | 1 | 0.9982 | 0 | 0 | 0 | 1 | Ref |
| 12067 | MT735629 | Unassigned | Unassigned | Unassigned | 904_0 | Clustered | 6 | VC_904_0 | 6 | 0.233 | 1 | 0.233 | 0 | 0 | 0 | 1 | Ref |
| 12068 | MT740244 | Unassigned | Unassigned | Unassigned | 812_1 | Clustered | 4 | VC_812_1 | 4 | 0.2266 | 0.99998426 | 0.2266 | 0 | 0 | 0 | 1 | Ref |
| 12069 | MT740291 | Unassigned | Unassigned | Unassigned | 637_0 | Clustered | 115 | VC_637_0 | 115 | 0.731 | 1 | 0.731 | 0 | 0 | 0 | 0.8764 | Ref |
| 12070 | MT740314 | Unassigned | Unassigned | Unassigned | 772_5 | Clustered/Singleton | 1 | VC_772_5 | 1 | 0 | 1 | 0 | 0 | 0 | 0 | 1 | Ref |
| 12071 | MT740726 | Unassigned | Unassigned | Unassigned | 1205_0 | Clustered | 3 | VC_1205_0 | 3 | 0.9212 | 0.99501814 | 0.9166 | 0 | 0 | 0 | 1 | Ref |
| 12072 | MT740727 | Unassigned | Unassigned | Unassigned | 162_0 | Clustered | 18 | VC_162_0 | 18 | 0.7403 | 1 | 0.7403 | 0 | 0 | 0 | 0.8627 | Ref |
| 12073 | MT740728 | Unassigned | Unassigned | Unassigned | 70_0 | Clustered | 27 | VC_70_0 | 27 | 0.5981 | 1 | 0.5981 | 0 | 0 | 0 | 0.9886 | Ref |
| 12074 | MT740730 | Unassigned | Unassigned | Unassigned | 162_0 | Clustered | 18 | VC_162_0 | 18 | 0.7403 | 1 | 0.7403 | 0 | 0 | 0 | 0.8627 | Ref |
| 12075 | MT740731 | Unassigned | Unassigned | Unassigned | 162_0 | Clustered | 18 | VC_162_0 | 18 | 0.7403 | 1 | 0.7403 | 0 | 0 | 0 | 0.8627 | Ref |
| 12076 | MT740732 | Unassigned | Unassigned | Unassigned | 162_0 | Clustered | 18 | VC_162_0 | 18 | 0.7403 | 1 | 0.7403 | 0 | 0 | 0 | 0.8627 | Ref |
| 12077 | MT740733 | Unassigned | Unassigned | Unassigned | 162_0 | Clustered | 18 | VC_162_0 | 18 | 0.7403 | 1 | 0.7403 | 0 | 0 | 0 | 0.8627 | Ref |
| 12078 | MT740734 | Unassigned | Unassigned | Unassigned | 357_0 | Clustered | 53 | VC_357_0 | 53 | 0.8979 | 1 | 0.8979 | 0 | 0 | 0 | 1 | Ref |
| 12079 | MT740736 | Unassigned | Unassigned | Unassigned | 162_0 | Clustered | 18 | VC_162_0 | 18 | 0.7403 | 1 | 0.7403 | 0 | 0 | 0 | 0.8627 | Ref |
| 12080 | MT740737 | Unassigned | Unassigned | Unassigned | 161_0 | Clustered | 3 | VC_161_0 | 3 | 0.5777 | 0.99804396 | 0.5765 | 0 | 0 | 0 | 1 | Ref |
| 12081 | MT740738 | Unassigned | Unassigned | Unassigned | 162_0 | Clustered | 18 | VC_162_0 | 18 | 0.7403 | 1 | 0.7403 | 0 | 0 | 0 | 0.8627 | Ref |
| 12082 | MT740739 | Unassigned | Unassigned | Unassigned | 162_0 | Clustered | 18 | VC_162_0 | 18 | 0.7403 | 1 | 0.7403 | 0 | 0 | 0 | 0.8627 | Ref |
| 12083 | MT740740 | Unassigned | Unassigned | Unassigned | 162_0 | Clustered | 18 | VC_162_0 | 18 | 0.7403 | 1 | 0.7403 | 0 | 0 | 0 | 0.8627 | Ref |
| 12084 | MT740741 | Unassigned | Unassigned | Unassigned | 161_0 | Clustered | 3 | VC_161_0 | 3 | 0.5777 | 0.99804396 | 0.5765 | 0 | 0 | 0 | 1 | Ref |
| 12085 | MT740742 | Unassigned | Unassigned | Unassigned | 162_0 | Clustered | 18 | VC_162_0 | 18 | 0.7403 | 1 | 0.7403 | 0 | 0 | 0 | 0.8627 | Ref |
| 12086 | MT740743 | Unassigned | Unassigned | Unassigned | 1205_0 | Clustered | 3 | VC_1205_0 | 3 | 0.9212 | 0.99501814 | 0.9166 | 0 | 0 | 0 | 1 | Ref |
| 12087 | MT740745 | Unassigned | Unassigned | Unassigned | 1205_0 | Clustered | 3 | VC_1205_0 | 3 | 0.9212 | 0.99501814 | 0.9166 | 0 | 0 | 0 | 1 | Ref |
| 12088 | MT741943 | Unassigned | Unassigned | Unassigned | 492_1 | Clustered | 20 | VC_492_1 | 20 | 0.4176 | 1 | 0.4176 | 0 | 0 | 0 | 1 | Ref |
| 12089 | MT741944 | Unassigned | Unassigned | Unassigned | 699_0 | Clustered | 71 | VC_699_0 | 71 | 0.9352 | 1 | 0.9352 | 0 | 0 | 0 | 1 | Ref |
| 12090 | MT745954 | Unassigned | Unassigned | Unassigned | 704_0 | Clustered | 16 | VC_704_0 | 16 | 0.7274 | 1 | 0.7274 | 0 | 0 | 0 | 1 | Ref |
| 12091 | MT745955 | Unassigned | Unassigned | Unassigned | 704_0 | Clustered | 16 | VC_704_0 | 16 | 0.7274 | 1 | 0.7274 | 0 | 0 | 0 | 1 | Ref |
| 12092 | MT745956 | Unassigned | Unassigned | Unassigned | 704_0 | Clustered | 16 | VC_704_0 | 16 | 0.7274 | 1 | 0.7274 | 0 | 0 | 0 | 1 | Ref |
| 12093 | MT747434 | Unassigned | Unassigned | Unassigned | 967_0 | Clustered | 16 | VC_967_0 | 16 | 0.8971 | 1 | 0.8971 | 0 | 0 | 0 | 1 | Ref |
| 12094 | MT757392 | Unassigned | Unassigned | Unassigned | 123_0 | Clustered | 23 | VC_123_0 | 23 | 0.817 | 1 | 0.817 | 0 | 0 | 0 | 1 | Ref |
| 12095 | MT757399 | Unassigned | Unassigned | Unassigned | 967_0 | Clustered | 16 | VC_967_0 | 16 | 0.8971 | 1 | 0.8971 | 0 | 0 | 0 | 1 | Ref |
| 12096 | MT758688 | Unassigned | Unassigned | Unassigned | 888_0 | Clustered | 28 | VC_888_0 | 28 | 0.5497 | 1 | 0.5497 | 0 | 0 | 0 | 1 | Ref |
| 12097 | MT758689 | Unassigned | Unassigned | Unassigned | 93_0 | Clustered | 2 | VC_93_0 | 2 | 0.1063 | 0.95665672 | 0.1017 | 0 | 0 | 0 | 1 | Ref |
| 12098 | MT764206 | Unassigned | Unassigned | Unassigned | 283_80 | Clustered/Singleton | 1 | VC_283_80 | 1 | 0 | 1 | 0 | 0 | 0 | 0 | 1 | Ref |
| 12099 | MT764207 | Unassigned | Unassigned | Unassigned | 428_0 | Clustered | 33 | VC_428_0 | 33 | 0.9915 | 1 | 0.9915 | 0 | 0 | 0 | 1 | Ref |
| 12100 | MT764208 | Unassigned | Unassigned | Unassigned | 590_0 | Clustered | 27 | VC_590_0 | 27 | 0.9947 | 1 | 0.9947 | 0 | 0 | 0 | 1 | Ref |
| 12101 | MT767883 | Unassigned | Unassigned | Unassigned | 754_0 | Clustered | 8 | VC_754_0 | 8 | 0.752 | 1 | 0.752 | 0 | 0 | 0 | 1 | Ref |
| 12102 | MT768059 | Unassigned | Unassigned | Unassigned | 428_0 | Clustered | 33 | VC_428_0 | 33 | 0.9915 | 1 | 0.9915 | 0 | 0 | 0 | 1 | Ref |
| 12103 | MT768060 | Unassigned | Unassigned | Unassigned | 461_0 | Clustered | 96 | VC_461_0 | 96 | 0.9307 | 1 | 0.9307 | 0 | 0 | 0 | 0.9575 | Ref |
| 12104 | MT770738 | Unassigned | Unassigned | Unassigned | 724_0 | Clustered | 3 | VC_724_0 | 3 | 0.1586 | 0.99815158 | 0.1583 | 0 | 0 | 0 | 1 | Ref |
| 12105 | MT771339 | Unassigned | Unassigned | Unassigned | 1161_0 | Clustered | 2 | VC_1161_0 | 2 | 0.0296 | 0.9573719 | 0.0283 | 0 | 0 | 0 | 1 | Ref |
| 12106 | MT771340 | Unassigned | Unassigned | Unassigned | 325_59 | Clustered/Singleton | 1 | VC_325_59 | 1 | 0 | 1 | 0 | 0 | 0 | 0 | 1 | Ref |
| 12107 | MT771341 | Unassigned | Unassigned | Unassigned | 1177_0 | Clustered | 14 | VC_1177_0 | 14 | 0.7847 | 1 | 0.7847 | 0 | 0 | 0 | 0.989 | Ref |
| 12108 | MT771342 | Unassigned | Unassigned | Unassigned | 1175_0 | Clustered | 22 | VC_1175_0 | 22 | 0.4562 | 1 | 0.4562 | 0 | 0 | 0 | 1 | Ref |
| 12109 | MT771344 | Unassigned | Unassigned | Unassigned | 1360_0 | Clustered | 31 | VC_1360_0 | 31 | 0.8837 | 1 | 0.8837 | 0 | 0 | 0 | 1 | Ref |
| 12110 | MT771345 | Unassigned | Unassigned | Unassigned | 570_0 | Clustered | 9 | VC_570_0 | 9 | 0.499 | 1 | 0.499 | 0 | 0 | 0 | 1 | Ref |
| 12111 | MT771347 | Unassigned | Unassigned | Unassigned | 1043_0 | Clustered | 2 | VC_1043_0 | 2 | 0.0576 | 0.95654247 | 0.0551 | 0 | 0 | 0 | 1 | Ref |
| 12112 | MT771348 | Unassigned | Unassigned | Unassigned | 570_0 | Clustered | 9 | VC_570_0 | 9 | 0.499 | 1 | 0.499 | 0 | 0 | 0 | 1 | Ref |
| 12113 | MT771349 | Unassigned | Unassigned | Unassigned | 887_0 | Clustered | 18 | VC_887_0 | 18 | 0.3663 | 1 | 0.3663 | 0 | 0 | 0 | 1 | Ref |
| 12114 | MT771350 | Unassigned | Unassigned | Unassigned | 1240_0 | Clustered | 82 | VC_1240_0 | 82 | 0.7945 | 1 | 0.7945 | 0 | 0 | 0 | 1 | Ref |
| 12115 | MT774375 | Unassigned | Unassigned | Unassigned | 502_1 | Clustered | 6 | VC_502_1 | 6 | 0.234 | 1 | 0.234 | 0 | 0 | 0 | 1 | Ref |
| 12116 | MT774376 | Unassigned | Unassigned | Unassigned | 502_1 | Clustered | 6 | VC_502_1 | 6 | 0.234 | 1 | 0.234 | 0 | 0 | 0 | 1 | Ref |
| 12117 | MT774378 | Unassigned | Unassigned | Unassigned | 1018_1 | Clustered | 10 | VC_1018_1 | 10 | 0.7518 | 1 | 0.7518 | 0 | 0 | 0 | 0.8 | Ref |
| 12118 | MT774379 | Unassigned | Unassigned | Unassigned | 502_1 | Clustered | 6 | VC_502_1 | 6 | 0.234 | 1 | 0.234 | 0 | 0 | 0 | 1 | Ref |
| 12119 | MT774380 | Unassigned | Unassigned | Unassigned | 502_1 | Clustered | 6 | VC_502_1 | 6 | 0.234 | 1 | 0.234 | 0 | 0 | 0 | 1 | Ref |
| 12120 | MT774381 | Unassigned | Unassigned | Unassigned | 673_0 | Clustered | 15 | VC_673_0 | 15 | 0.8234 | 1 | 0.8234 | 0 | 0 | 0 | 0.7619 | Ref |
| 12121 | MT774382 | Unassigned | Unassigned | Unassigned | 502_1 | Clustered | 6 | VC_502_1 | 6 | 0.234 | 1 | 0.234 | 0 | 0 | 0 | 1 | Ref |
| 12122 | MT774383 | Unassigned | Unassigned | Unassigned | 673_0 | Clustered | 15 | VC_673_0 | 15 | 0.8234 | 1 | 0.8234 | 0 | 0 | 0 | 0.7619 | Ref |
| 12123 | MT774384 | Unassigned | Unassigned | Unassigned | 673_0 | Clustered | 15 | VC_673_0 | 15 | 0.8234 | 1 | 0.8234 | 0 | 0 | 0 | 0.7619 | Ref |
| 12124 | MT774385 | Unassigned | Unassigned | Unassigned | 1018_1 | Clustered | 10 | VC_1018_1 | 10 | 0.7518 | 1 | 0.7518 | 0 | 0 | 0 | 0.8 | Ref |
| 12125 | MT774386 | Unassigned | Unassigned | Unassigned | 673_0 | Clustered | 15 | VC_673_0 | 15 | 0.8234 | 1 | 0.8234 | 0 | 0 | 0 | 0.7619 | Ref |
| 12126 | MT774387 | Unassigned | Unassigned | Unassigned | 1018_1 | Clustered | 10 | VC_1018_1 | 10 | 0.7518 | 1 | 0.7518 | 0 | 0 | 0 | 0.8 | Ref |
| 12127 | MT774388 | Unassigned | Unassigned | Unassigned | 1018_1 | Clustered | 10 | VC_1018_1 | 10 | 0.7518 | 1 | 0.7518 | 0 | 0 | 0 | 0.8 | Ref |
| 12128 | MT774389 | Unassigned | Unassigned | Unassigned | 1018_0 | Clustered | 2 | VC_1018_0 | 2 | 0.0851 | 0.94401932 | 0.0804 | 0 | 0 | 0 | 1 | Ref |
| 12129 | MT774390 | Unassigned | Unassigned | Unassigned | 1018_1 | Clustered | 10 | VC_1018_1 | 10 | 0.7518 | 1 | 0.7518 | 0 | 0 | 0 | 0.8 | Ref |
| 12130 | MT774391 | Unassigned | Unassigned | Unassigned | 1018_1 | Clustered | 10 | VC_1018_1 | 10 | 0.7518 | 1 | 0.7518 | 0 | 0 | 0 | 0.8 | Ref |
| 12131 | MT774392 | Unassigned | Unassigned | Unassigned | 1018_1 | Clustered | 10 | VC_1018_1 | 10 | 0.7518 | 1 | 0.7518 | 0 | 0 | 0 | 0.8 | Ref |
| 12132 | MT774393 | Unassigned | Unassigned | Unassigned | 675_0 | Clustered | 2 | VC_675_0 | 2 | 0.081 | 0.95018582 | 0.077 | 0 | 0 | 0 | 1 | Ref |
| 12133 | MT774394 | Unassigned | Unassigned | Unassigned | 673_0 | Clustered | 15 | VC_673_0 | 15 | 0.8234 | 1 | 0.8234 | 0 | 0 | 0 | 0.7619 | Ref |
| 12134 | MT774395 | Unassigned | Unassigned | Unassigned | 502_4 | Clustered | 3 | VC_502_4 | 3 | 0.0997 | 0.99715347 | 0.0994 | 0 | 0 | 0 | 1 | Ref |
| 12135 | MT774396 | Unassigned | Unassigned | Unassigned | 502_5 | Clustered | 2 | VC_502_5 | 2 | 0.0539 | 0.93577964 | 0.0505 | 0 | 0 | 0 | 1 | Ref |
| 12136 | MT774397 | Unassigned | Unassigned | Unassigned | 502_4 | Clustered | 3 | VC_502_4 | 3 | 0.0997 | 0.99715347 | 0.0994 | 0 | 0 | 0 | 1 | Ref |
| 12137 | MT774398 | Unassigned | Unassigned | Unassigned | 502_4 | Clustered | 3 | VC_502_4 | 3 | 0.0997 | 0.99715347 | 0.0994 | 0 | 0 | 0 | 1 | Ref |
| 12138 | MT774399 | Unassigned | Unassigned | Unassigned | 502_3 | Clustered | 2 | VC_502_3 | 2 | 0.0724 | 0.95171362 | 0.0689 | 0 | 0 | 0 | 1 | Ref |
| 12139 | MT774400 | Unassigned | Unassigned | Unassigned | 502_7 | Clustered/Singleton | 1 | VC_502_7 | 1 | 0 | 1 | 0 | 0 | 0 | 0 | 1 | Ref |
| 12140 | MT774401 | Unassigned | Unassigned | Unassigned | 502_2 | Clustered/Singleton | 1 | VC_502_2 | 1 | 0 | 1 | 0 | 0 | 0 | 0 | 1 | Ref |
| 12141 | MT774403 | Unassigned | Unassigned | Unassigned | 502_3 | Clustered | 2 | VC_502_3 | 2 | 0.0724 | 0.95171362 | 0.0689 | 0 | 0 | 0 | 1 | Ref |
| 12142 | MT774404 | Unassigned | Unassigned | Unassigned | 502_6 | Clustered | 2 | VC_502_6 | 2 | 0.0804 | 0.95171362 | 0.0765 | 0 | 0 | 0 | 1 | Ref |
| 12143 | MT774405 | Unassigned | Unassigned | Unassigned | 502_5 | Clustered | 2 | VC_502_5 | 2 | 0.0539 | 0.93577964 | 0.0505 | 0 | 0 | 0 | 1 | Ref |
| 12144 | MT774407 | Unassigned | Unassigned | Unassigned | 502_6 | Clustered | 2 | VC_502_6 | 2 | 0.0804 | 0.95171362 | 0.0765 | 0 | 0 | 0 | 1 | Ref |
| 12145 | MT774487 | Unassigned | Unassigned | Unassigned | 28_1 | Clustered | 53 | VC_28_1 | 53 | 0.7164 | 1 | 0.7164 | 0 | 0 | 0 | 0.9325 | Ref |
| 12146 | MT776806 | Unassigned | Unassigned | Unassigned | 571_0 | Clustered | 4 | VC_571_0 | 4 | 0.1921 | 0.99998591 | 0.1921 | 0 | 0 | 0 | 1 | Ref |
| 12147 | MT776807 | Unassigned | Unassigned | Unassigned | 1344_0 | Clustered | 78 | VC_1344_0 | 78 | 0.9732 | 1 | 0.9732 | 0 | 0 | 0 | 1 | Ref |
| 12148 | MT776808 | Unassigned | Unassigned | Unassigned | 919_0 | Clustered | 39 | VC_919_0 | 39 | 0.9069 | 1 | 0.9069 | 0 | 0 | 0 | 1 | Ref |

| | | | | | | | | | | | | | | | | | |
|---|---|---|---|---|---|---|---|---|---|---|---|---|---|---|---|---|---|
| 12149 | MT776809 | Unassigned | Unassigned | Unassigned | 919_0 | Clustered | 39 | VC_919_0 | 39 | 0.9069 | 1 | 0.9069 | 0 | 0 | 0 | 1 | Ref |
| 12150 | MT776810 | Unassigned | Unassigned | Unassigned | 919_0 | Clustered | 39 | VC_919_0 | 39 | 0.9069 | 1 | 0.9069 | 0 | 0 | 0 | 1 | Ref |
| 12151 | MT776811 | Unassigned | Unassigned | Unassigned | 1073_0 | Clustered | 24 | VC_1073_0 | 24 | 0.8388 | 1 | 0.8388 | 0 | 0 | 0 | 1 | Ref |
| 12152 | MT776812 | Unassigned | Unassigned | Unassigned | 325_37 | Clustered | 3 | VC_325_37 | 3 | 0.008 | 0.99634936 | 0.008 | 0 | 0 | 0 | 0.6667 | Ref |
| 12153 | MT776813 | Unassigned | Unassigned | Unassigned | 371_9 | Clustered | 4 | VC_371_9 | 4 | 0.0109 | 0.9999766 | 0.0109 | 0 | 0 | 0 | 0.6667 | Ref |
| 12154 | MT777452 | Unassigned | Unassigned | Unassigned | 818_0 | Clustered | 20 | VC_818_0 | 20 | 0.9601 | 1 | 0.9601 | 0 | 0 | 0 | 1 | Ref |
| 12155 | MT778837 | Unassigned | Unassigned | Unassigned | 1052_3 | Clustered | 5 | VC_1052_3 | 5 | 0.185 | 0.98656048 | 0.1825 | 0 | 0 | 0 | 1 | Ref |
| 12156 | MT778838 | Unassigned | Unassigned | Unassigned | 724_0 | Clustered | 3 | VC_724_0 | 3 | 0.1586 | 0.99815158 | 0.1583 | 0 | 0 | 0 | 1 | Ref |
| 12157 | MT778839 | Unassigned | Unassigned | Unassigned | 1052_3 | Clustered | 5 | VC_1052_3 | 5 | 0.185 | 0.98656048 | 0.1825 | 0 | 0 | 0 | 1 | Ref |
| 12158 | MT778840 | Unassigned | Unassigned | Unassigned | 990_0 | Clustered | 3 | VC_990_0 | 3 | 0.6363 | 0.99819562 | 0.6351 | 0 | 0 | 0 | 1 | Ref |
| 12159 | MT782071 | Unassigned | Unassigned | Unassigned | 286_13 | Clustered | 10 | VC_286_13 | 10 | 0.0698 | 0.99999929 | 0.0698 | 0 | 0 | 0 | 0.9556 | Ref |
| 12160 | MT786458 | Unassigned | Unassigned | Unassigned | 149_15 | Clustered | 16 | VC_149_15 | 16 | 0.0788 | 1 | 0.0788 | 0 | 0 | 0 | 0.8083 | Ref |
| 12161 | MT787017 | Unassigned | Unassigned | Unassigned | 149_3 | Clustered/Singleton | 1 | VC_149_3 | 1 | 0 | 1 | 0 | 0 | 0 | 0 | 1 | Ref |
| 12162 | MT795651 | Unassigned | Unassigned | Unassigned | 614_0 | Clustered | 5 | VC_614_0 | 5 | 0.2395 | 0.99999997 | 0.2395 | 0 | 0 | 0 | 1 | Ref |
| 12163 | MT799840 | Unassigned | Unassigned | Unassigned | 959_5 | Clustered | 19 | VC_959_5 | 19 | 0.2626 | 1 | 0.2626 | 0 | 0 | 0 | 1 | Ref |
| 12164 | MT806185 | Unassigned | Unassigned | Unassigned | 1008_0 | Clustered | 4 | VC_1008_0 | 4 | 0.6602 | 0.99993542 | 0.6601 | 0 | 0 | 0 | 1 | Ref |
| 12165 | MT806186 | Unassigned | Unassigned | Unassigned | 1008_0 | Clustered | 4 | VC_1008_0 | 4 | 0.6602 | 0.99993542 | 0.6601 | 0 | 0 | 0 | 1 | Ref |
| 12166 | MT808983 | Unassigned | Unassigned | Unassigned | 451_0 | Clustered | 80 | VC_451_0 | 80 | 0.9991 | 1 | 0.9991 | 0 | 0 | 0 | 1 | Ref |
| 12167 | MT811961 | Unassigned | Unassigned | Unassigned | 346_1 | Clustered | 2 | VC_346_1 | 2 | 0.1238 | 0.95353448 | 0.1181 | 0 | 0 | 0 | 1 | Ref |
| 12168 | MT813140 | Unassigned | Unassigned | Unassigned | 823_0 | Clustered | 124 | VC_823_0 | 124 | 0.9492 | 1 | 0.9492 | 0 | 0 | 0 | 1 | Ref |
| 12169 | MT813141 | Unassigned | Unassigned | Unassigned | 823_0 | Clustered | 124 | VC_823_0 | 124 | 0.9492 | 1 | 0.9492 | 0 | 0 | 0 | 1 | Ref |
| 12170 | MT813142 | Unassigned | Unassigned | Unassigned | 385_0 | Clustered | 51 | VC_385_0 | 51 | 0.8754 | 1 | 0.8754 | 0 | 0 | 0 | 1 | Ref |
| 12171 | MT813197 | Unassigned | Unassigned | Unassigned | 30_1 | Clustered | 16 | VC_30_1 | 16 | 0.1973 | 1 | 0.1973 | 0 | 0 | 0 | 0.8333 | Ref |
| 12172 | MT818415 | Unassigned | Unassigned | Unassigned | 1352_0 | Clustered | 31 | VC_1352_0 | 31 | 0.849 | 1 | 0.849 | 0 | 0 | 0 | 1 | Ref |
| 12173 | MT818416 | Unassigned | Unassigned | Unassigned | 558_29 | Clustered/Singleton | 1 | VC_558_29 | 1 | 0 | 1 | 0 | 0 | 0 | 0 | 1 | Ref |
| 12174 | MT818417 | Unassigned | Unassigned | Unassigned | 517_1 | Clustered | 108 | VC_517_1 | 108 | 0.8483 | 1 | 0.8483 | 0 | 0 | 0 | 0.9955 | Ref |
| 12175 | MT818418 | Unassigned | Unassigned | Unassigned | 556_0 | Clustered | 21 | VC_556_0 | 21 | 0.9912 | 1 | 0.9912 | 0 | 0 | 0 | 1 | Ref |
| 12176 | MT818419 | Unassigned | Unassigned | Unassigned | 372_0 | Clustered | 24 | VC_372_0 | 24 | 0.5539 | 1 | 0.5539 | 0 | 0 | 0 | 1 | Ref |
| 12177 | MT818420 | Unassigned | Unassigned | Unassigned | 1343_0 | Clustered | 21 | VC_1343_0 | 21 | 0.9843 | 1 | 0.9843 | 0 | 0 | 0 | 1 | Ref |
| 12178 | MT818421 | Unassigned | Unassigned | Unassigned | 810_0 | Clustered | 18 | VC_810_0 | 18 | 0.9121 | 1 | 0.9121 | 0 | 0 | 0 | 1 | Ref |
| 12179 | MT818422 | Unassigned | Unassigned | Unassigned | 761_0 | Clustered | 67 | VC_761_0 | 67 | 0.8947 | 1 | 0.8947 | 0 | 0 | 0 | 1 | Ref |
| 12180 | MT818423 | Unassigned | Unassigned | Unassigned | 558_35 | Clustered/Singleton | 1 | VC_558_35 | 1 | 0 | 1 | 0 | 0 | 0 | 0 | 1 | Ref |
| 12181 | MT818424 | Unassigned | Unassigned | Unassigned | 1175_0 | Clustered | 22 | VC_1175_0 | 22 | 0.4562 | 1 | 0.4562 | 0 | 0 | 0 | 1 | Ref |
| 12182 | MT818425 | Unassigned | Unassigned | Unassigned | 556_0 | Clustered | 21 | VC_556_0 | 21 | 0.9912 | 1 | 0.9912 | 0 | 0 | 0 | 1 | Ref |
| 12183 | MT818426 | Unassigned | Unassigned | Unassigned | 1031_0 | Clustered | 4 | VC_1031_0 | 4 | 0.7884 | 0.99997621 | 0.7884 | 0 | 0 | 0 | 1 | Ref |
| 12184 | MT818427 | Unassigned | Unassigned | Unassigned | 1234_0 | Clustered | 44 | VC_1234_0 | 44 | 1 | 1 | 1 | 0 | 0 | 0 | 1 | Ref |
| 12185 | MT822284 | Unassigned | Unassigned | Unassigned | 395_0 | Clustered | 5 | VC_395_0 | 5 | 0.1021 | 0.99999998 | 0.1021 | 0 | 0 | 0 | 1 | Ref |
| 12186 | MT822285 | Unassigned | Unassigned | Unassigned | 395_0 | Clustered | 5 | VC_395_0 | 5 | 0.1021 | 0.99999998 | 0.1021 | 0 | 0 | 0 | 1 | Ref |
| 12187 | MT822286 | Unassigned | Unassigned | Unassigned | 395_0 | Clustered | 5 | VC_395_0 | 5 | 0.1021 | 0.99999998 | 0.1021 | 0 | 0 | 0 | 1 | Ref |
| 12188 | MT822287 | Unassigned | Unassigned | Unassigned | 395_0 | Clustered | 5 | VC_395_0 | 5 | 0.1021 | 0.99999998 | 0.1021 | 0 | 0 | 0 | 1 | Ref |
| 12189 | MT822288 | Unassigned | Unassigned | Unassigned | 393_0 | Clustered | 7 | VC_393_0 | 7 | 0.1775 | 1 | 0.1775 | 0 | 0 | 0 | 1 | Ref |
| 12190 | MT828551 | Unassigned | Unassigned | Unassigned | 283_34 | Clustered | 4 | VC_283_34 | 4 | 0.0082 | 0.9997481 | 0.0082 | 0 | 0 | 0 | 1 | Ref |
| 12191 | MT828552 | Unassigned | Unassigned | Unassigned | 283_23 | Clustered/Singleton | 1 | VC_283_23 | 1 | 0 | 1 | 0 | 0 | 0 | 0 | 1 | Ref |
| 12192 | MT828553 | Unassigned | Unassigned | Unassigned | 283_34 | Clustered | 4 | VC_283_34 | 4 | 0.0082 | 0.9997481 | 0.0082 | 0 | 0 | 0 | 1 | Ref |
| 12193 | MT829326 | Unassigned | Unassigned | Unassigned | 695_0 | Clustered | 48 | VC_695_0 | 48 | 0.8162 | 1 | 0.8162 | 0 | 0 | 0 | 1 | Ref |
| 12194 | MT830620 | Unassigned | Unassigned | Unassigned | 315_0 | Clustered | 12 | VC_315_0 | 12 | 0.8947 | 1 | 0.8947 | 0 | 0 | 0 | 1 | Ref |
| 12195 | MT833282 | Unassigned | Unassigned | Unassigned | 329_18 | Clustered | 7 | VC_329_18 | 7 | 0.0328 | 1 | 0.0328 | 0 | 0 | 0 | 0.8095 | Ref |
| 12196 | MT833283 | Unassigned | Unassigned | Unassigned | 191_18 | Clustered | 2 | VC_191_18 | 2 | 0.0057 | 0.91903589 | 0.0053 | 0 | 0 | 0 | 0 | Ref |
| 12197 | MT833387 | Unassigned | Unassigned | Unassigned | 30_1 | Clustered | 16 | VC_30_1 | 16 | 0.1973 | 1 | 0.1973 | 0 | 0 | 0 | 0.8333 | Ref |
| 12198 | MT843274 | Unassigned | Unassigned | Unassigned | 190_20 | Clustered | 4 | VC_190_20 | 4 | 0.0229 | 0.99997885 | 0.0229 | 0 | 0 | 0 | 1 | Ref |
| 12199 | MT843275 | Unassigned | Unassigned | Unassigned | 411_0 | Clustered | 3 | VC_411_0 | 3 | 0.1239 | 0.98986341 | 0.1227 | 0 | 0 | 0 | 1 | Ref |
| 12200 | MT856905 | Unassigned | Unassigned | Unassigned | 92_0 | Clustered | 22 | VC_92_0 | 22 | 0.5639 | 1 | 0.5639 | 0 | 0 | 0 | 1 | Ref |
| 12201 | MT857001 | Unassigned | Unassigned | Unassigned | 695_0 | Clustered | 48 | VC_695_0 | 48 | 0.8162 | 1 | 0.8162 | 0 | 0 | 0 | 1 | Ref |
| 12202 | MT857002 | Unassigned | Unassigned | Unassigned | 131_3 | Clustered/Singleton | 1 | VC_131_3 | 1 | 0 | 1 | 0 | 0 | 0 | 0 | 1 | Ref |
| 12203 | MT862763 | Unassigned | Unassigned | Unassigned | 451_0 | Clustered | 80 | VC_451_0 | 80 | 0.9991 | 1 | 0.9991 | 0 | 0 | 0 | 1 | Ref |
| 12204 | MT863714 | Unassigned | Unassigned | Unassigned | 844_0 | Clustered | 27 | VC_844_0 | 27 | 0.9385 | 1 | 0.9385 | 0 | 0 | 0 | 1 | Ref |
| 12205 | MT863715 | Unassigned | Unassigned | Unassigned | 844_0 | Clustered | 27 | VC_844_0 | 27 | 0.9385 | 1 | 0.9385 | 0 | 0 | 0 | 1 | Ref |
| 12206 | MT863716 | Unassigned | Unassigned | Unassigned | 844_0 | Clustered | 27 | VC_844_0 | 27 | 0.9385 | 1 | 0.9385 | 0 | 0 | 0 | 1 | Ref |
| 12207 | MT863717 | Unassigned | Unassigned | Unassigned | 844_0 | Clustered | 27 | VC_844_0 | 27 | 0.9385 | 1 | 0.9385 | 0 | 0 | 0 | 1 | Ref |
| 12208 | MT863718 | Unassigned | Unassigned | Unassigned | 844_0 | Clustered | 27 | VC_844_0 | 27 | 0.9385 | 1 | 0.9385 | 0 | 0 | 0 | 1 | Ref |
| 12209 | MT863719 | Unassigned | Unassigned | Unassigned | 844_0 | Clustered | 27 | VC_844_0 | 27 | 0.9385 | 1 | 0.9385 | 0 | 0 | 0 | 1 | Ref |
| 12210 | MT863720 | Unassigned | Unassigned | Unassigned | 844_0 | Clustered | 27 | VC_844_0 | 27 | 0.9385 | 1 | 0.9385 | 0 | 0 | 0 | 1 | Ref |
| 12211 | MT863721 | Unassigned | Unassigned | Unassigned | 844_0 | Clustered | 27 | VC_844_0 | 27 | 0.9385 | 1 | 0.9385 | 0 | 0 | 0 | 1 | Ref |
| 12212 | MT863722 | Unassigned | Unassigned | Unassigned | 844_0 | Clustered | 27 | VC_844_0 | 27 | 0.9385 | 1 | 0.9385 | 0 | 0 | 0 | 1 | Ref |
| 12213 | MT863723 | Unassigned | Unassigned | Unassigned | 844_0 | Clustered | 27 | VC_844_0 | 27 | 0.9385 | 1 | 0.9385 | 0 | 0 | 0 | 1 | Ref |
| 12214 | MT863724 | Unassigned | Unassigned | Unassigned | 844_0 | Clustered | 27 | VC_844_0 | 27 | 0.9385 | 1 | 0.9385 | 0 | 0 | 0 | 1 | Ref |
| 12215 | MT863725 | Unassigned | Unassigned | Unassigned | 844_0 | Clustered | 27 | VC_844_0 | 27 | 0.9385 | 1 | 0.9385 | 0 | 0 | 0 | 1 | Ref |
| 12216 | MT863726 | Unassigned | Unassigned | Unassigned | 844_0 | Clustered | 27 | VC_844_0 | 27 | 0.9385 | 1 | 0.9385 | 0 | 0 | 0 | 1 | Ref |
| 12217 | MT863727 | Unassigned | Unassigned | Unassigned | 844_0 | Clustered | 27 | VC_844_0 | 27 | 0.9385 | 1 | 0.9385 | 0 | 0 | 0 | 1 | Ref |
| 12218 | MT863728 | Unassigned | Unassigned | Unassigned | 844_0 | Clustered | 27 | VC_844_0 | 27 | 0.9385 | 1 | 0.9385 | 0 | 0 | 0 | 1 | Ref |
| 12219 | MT863729 | Unassigned | Unassigned | Unassigned | 844_0 | Clustered | 27 | VC_844_0 | 27 | 0.9385 | 1 | 0.9385 | 0 | 0 | 0 | 1 | Ref |
| 12220 | MT863730 | Unassigned | Unassigned | Unassigned | 844_0 | Clustered | 27 | VC_844_0 | 27 | 0.9385 | 1 | 0.9385 | 0 | 0 | 0 | 1 | Ref |
| 12221 | MT863731 | Unassigned | Unassigned | Unassigned | 844_0 | Clustered | 27 | VC_844_0 | 27 | 0.9385 | 1 | 0.9385 | 0 | 0 | 0 | 1 | Ref |
| 12222 | MT880872 | Unassigned | Unassigned | Unassigned | 213_0 | Clustered | 2 | VC_213_0 | 2 | 0.4293 | 0.95300227 | 0.4091 | 0 | 0 | 0 | 1 | Ref |
| 12223 | MT884006 | Unassigned | Unassigned | Unassigned | 283_184 | Clustered | 2 | VC_283_184 | 2 | 0.0025 | 0.93531224 | 0.0023 | 0 | 0 | 0 | 1 | Ref |
| 12224 | MT884007 | Unassigned | Unassigned | Unassigned | 283_184 | Clustered | 2 | VC_283_184 | 2 | 0.0025 | 0.93531224 | 0.0023 | 0 | 0 | 0 | 1 | Ref |
| 12225 | MT884008 | Unassigned | Unassigned | Unassigned | 772_10 | Clustered | 8 | VC_772_10 | 8 | 0.0433 | 0.9088183 | 0.0394 | 0 | 0 | 0 | 0.9286 | Ref |
| 12226 | MT884009 | Unassigned | Unassigned | Unassigned | 772_10 | Clustered | 8 | VC_772_10 | 8 | 0.0433 | 0.9088183 | 0.0394 | 0 | 0 | 0 | 0.9286 | Ref |
| 12227 | MT884010 | Unassigned | Unassigned | Unassigned | 772_10 | Clustered | 8 | VC_772_10 | 8 | 0.0433 | 0.9088183 | 0.0394 | 0 | 0 | 0 | 0.9286 | Ref |
| 12228 | MT884011 | Unassigned | Unassigned | Unassigned | 772_10 | Clustered | 8 | VC_772_10 | 8 | 0.0433 | 0.9088183 | 0.0394 | 0 | 0 | 0 | 0.9286 | Ref |
| 12229 | MT884012 | Unassigned | Unassigned | Unassigned | 835_0 | Clustered | 67 | VC_835_0 | 67 | 0.9624 | 1 | 0.9624 | 0 | 0 | 0 | 1 | Ref |
| 12230 | MT884013 | Unassigned | Unassigned | Unassigned | 835_0 | Clustered | 67 | VC_835_0 | 67 | 0.9624 | 1 | 0.9624 | 0 | 0 | 0 | 1 | Ref |
| 12231 | MT884014 | Unassigned | Unassigned | Unassigned | 835_0 | Clustered | 67 | VC_835_0 | 67 | 0.9624 | 1 | 0.9624 | 0 | 0 | 0 | 1 | Ref |
| 12232 | MT884015 | Unassigned | Unassigned | Unassigned | 835_0 | Clustered | 67 | VC_835_0 | 67 | 0.9624 | 1 | 0.9624 | 0 | 0 | 0 | 1 | Ref |
| 12233 | MT887289 | Unassigned | Unassigned | Unassigned | 329_18 | Clustered | 7 | VC_329_18 | 7 | 0.0328 | 1 | 0.0328 | 0 | 0 | 0 | 0.8095 | Ref |
| 12234 | MT889364 | Unassigned | Unassigned | Unassigned | 1362_0 | Clustered | 6 | VC_1362_0 | 6 | 1 | 1 | 1 | 0 | 0 | 0 | 1 | Ref |
| 12235 | MT889365 | Unassigned | Unassigned | Unassigned | 1234_0 | Clustered | 44 | VC_1234_0 | 44 | 1 | 1 | 1 | 0 | 0 | 0 | 1 | Ref |
| 12236 | MT889366 | Unassigned | Unassigned | Unassigned | 1207_0 | Clustered | 29 | VC_1207_0 | 29 | 0.8225 | 1 | 0.8225 | 0 | 0 | 0 | 1 | Ref |
| 12237 | MT889367 | Unassigned | Unassigned | Unassigned | 919_0 | Clustered | 39 | VC_919_0 | 39 | 0.9069 | 1 | 0.9069 | 0 | 0 | 0 | 1 | Ref |
| 12238 | MT889368 | Unassigned | Unassigned | Unassigned | 868_0 | Clustered | 25 | VC_868_0 | 25 | 0.5791 | 1 | 0.5791 | 0 | 0 | 0 | 1 | Ref |
| 12239 | MT889369 | Unassigned | Unassigned | Unassigned | 371_9 | Clustered | 4 | VC_371_9 | 4 | 0.0109 | 0.9999766 | 0.0109 | 0 | 0 | 0 | 0.6667 | Ref |
| 12240 | MT889370 | Unassigned | Unassigned | Unassigned | 868_0 | Clustered | 25 | VC_868_0 | 25 | 0.5791 | 1 | 0.5791 | 0 | 0 | 0 | 1 | Ref |
| 12241 | MT889371 | Unassigned | Unassigned | Unassigned | 1350_0 | Clustered | 2 | VC_1350_0 | 2 | 0.1631 | 0.95304852 | 0.1555 | 0 | 0 | 0 | 1 | Ref |
| 12242 | MT889372 | Unassigned | Unassigned | Unassigned | 1234_0 | Clustered | 44 | VC_1234_0 | 44 | 1 | 1 | 1 | 0 | 0 | 0 | 1 | Ref |
| 12243 | MT889373 | Unassigned | Unassigned | Unassigned | 1166_0 | Clustered | 20 | VC_1166_0 | 20 | 0.99 | 1 | 0.99 | 0 | 0 | 0 | 1 | Ref |
| 12244 | MT889374 | Unassigned | Unassigned | Unassigned | 520_1 | Clustered/Singleton | 1 | VC_520_1 | 1 | 0 | 1 | 0 | 0 | 0 | 0 | 1 | Ref |
| 12245 | MT889375 | Unassigned | Unassigned | Unassigned | 1207_0 | Clustered | 29 | VC_1207_0 | 29 | 0.8225 | 1 | 0.8225 | 0 | 0 | 0 | 1 | Ref |
| 12246 | MT889376 | Unassigned | Unassigned | Unassigned | 1207_0 | Clustered | 29 | VC_1207_0 | 29 | 0.8225 | 1 | 0.8225 | 0 | 0 | 0 | 1 | Ref |
| 12247 | MT889378 | Unassigned | Unassigned | Unassigned | 919_0 | Clustered | 39 | VC_919_0 | 39 | 0.9069 | 1 | 0.9069 | 0 | 0 | 0 | 1 | Ref |
| 12248 | MT889379 | Unassigned | Unassigned | Unassigned | 868_0 | Clustered | 25 | VC_868_0 | 25 | 0.5791 | 1 | 0.5791 | 0 | 0 | 0 | 1 | Ref |
| 12249 | MT889381 | Unassigned | Unassigned | Unassigned | 372_1 | Clustered | 9 | VC_372_1 | 9 | 0.194 | 1 | 0.194 | 0 | 0 | 0 | 1 | Ref |
| 12250 | MT889382 | Unassigned | Unassigned | Unassigned | 517_1 | Clustered | 108 | VC_517_1 | 108 | 0.8483 | 1 | 0.8483 | 0 | 0 | 0 | 0.9955 | Ref |
| 12251 | MT889383 | Unassigned | Unassigned | Unassigned | 889_0 | Clustered | 16 | VC_889_0 | 16 | 0.2969 | 1 | 0.2969 | 0 | 0 | 0 | 1 | Ref |
| 12252 | MT889384 | Unassigned | Unassigned | Unassigned | 325_14 | Clustered | 3 | VC_325_14 | 3 | 0.0094 | 0.99859159 | 0.0094 | 0 | 0 | 0 | 1 | Ref |
| 12253 | MT889385 | Unassigned | Unassigned | Unassigned | 565_0 | Clustered | 20 | VC_565_0 | 20 | 0.8398 | 1 | 0.8398 | 0 | 0 | 0 | 1 | Ref |
| 12254 | MT889386 | Unassigned | Unassigned | Unassigned | 517_1 | Clustered | 108 | VC_517_1 | 108 | 0.8483 | 1 | 0.8483 | 0 | 0 | 0 | 0.9955 | Ref |
| 12255 | MT889388 | Unassigned | Unassigned | Unassigned | 1073_0 | Clustered | 24 | VC_1073_0 | 24 | 0.8388 | 1 | 0.8388 | 0 | 0 | 0 | 1 | Ref |
| 12256 | MT889389 | Unassigned | Unassigned | Unassigned | 1234_0 | Clustered | 44 | VC_1234_0 | 44 | 1 | 1 | 1 | 0 | 0 | 0 | 1 | Ref |
| 12257 | MT889390 | Unassigned | Unassigned | Unassigned | 517_1 | Clustered | 108 | VC_517_1 | 108 | 0.8483 | 1 | 0.8483 | 0 | 0 | 0 | 0.9955 | Ref |
| 12258 | MT889391 | Unassigned | Unassigned | Unassigned | 889_0 | Clustered | 16 | VC_889_0 | 16 | 0.2969 | 1 | 0.2969 | 0 | 0 | 0 | 1 | Ref |
| 12259 | MT889392 | Unassigned | Unassigned | Unassigned | 1234_0 | Clustered | 44 | VC_1234_0 | 44 | 1 | 1 | 1 | 0 | 0 | 0 | 1 | Ref |
| 12260 | MT889393 | Unassigned | Unassigned | Unassigned | 767_0 | Clustered | 34 | VC_767_0 | 34 | 0.9173 | 1 | 0.9173 | 0 | 0 | 0 | 1 | Ref |
| 12261 | MT889394 | Unassigned | Unassigned | Unassigned | 1170_0 | Clustered | 3 | VC_1170_0 | 3 | 0.3275 | 0.99834213 | 0.327 | 0 | 0 | 0 | 1 | Ref |
| 12262 | MT889395 | Unassigned | Unassigned | Unassigned | 558_11 | Clustered | 2 | VC_558_11 | 2 | 0.0032 | 0.78528587 | 0.0025 | 0 | 0 | 0 | 1 | Ref |
| 12263 | MT889396 | Unassigned | Unassigned | Unassigned | 520_92 | Clustered | 5 | VC_520_92 | 5 | 0.0663 | 0.99999997 | 0.0663 | 0 | 0 | 0 | 1 | Ref |
| 12264 | MT889397 | Unassigned | Unassigned | Unassigned | 520_92 | Clustered | 5 | VC_520_92 | 5 | 0.0663 | 0.99999997 | 0.0663 | 0 | 0 | 0 | 1 | Ref |
| 12265 | MT889398 | Unassigned | Unassigned | Unassigned | 525_0 | Clustered | 25 | VC_525_0 | 25 | 0.4597 | 1 | 0.4597 | 0 | 0 | 0 | 1 | Ref |
| 12266 | MT894004 | Unassigned | Unassigned | Unassigned | 123_0 | Clustered | 23 | VC_123_0 | 23 | 0.817 | 1 | 0.817 | 0 | 0 | 0 | 1 | Ref |
| 12267 | MT894005 | Unassigned | Unassigned | Unassigned | 938_0 | Clustered | 99 | VC_938_0 | 99 | 0.9119 | 1 | 0.9119 | 0 | 0 | 0 | 0.9928 | Ref |
| 12268 | MT897901 | Unassigned | Unassigned | Unassigned | 371_18 | Clustered | 10 | VC_371_18 | 10 | 0.0302 | 1 | 0.0302 | 0 | 0 | 0 | 0.8 | Ref |
| 12269 | MT897902 | Unassigned | Unassigned | Unassigned | 371_3 | Clustered | 22 | VC_371_3 | 22 | 0.0787 | 1 | 0.0787 | 0 | 0 | 0 | 0.9048 | Ref |
| 12270 | MT897903 | Unassigned | Unassigned | Unassigned | 371_0 | Clustered | 2 | VC_371_0 | 2 | 0.0032 | 0.94731972 | 0.003 | 0 | 0 | 0 | 1 | Ref |
| 12271 | MT897904 | Unassigned | Unassigned | Unassigned | 520_50 | Clustered/Singleton | 1 | VC_520_50 | 1 | 0 | 1 | 0 | 0 | 0 | 0 | 1 | Ref |
| 12272 | MT897905 | Unassigned | Unassigned | Unassigned | 1071_2 | Clustered | 5 | VC_1071_2 | 5 | 0.0534 | 0.99997362 | 0.0534 | 0 | 0 | 0 | 0.9 | Ref |
| 12273 | MT897906 | Unassigned | Unassigned | Unassigned | 1207_0 | Clustered | 29 | VC_1207_0 | 29 | 0.8225 | 1 | 0.8225 | 0 | 0 | 0 | 1 | Ref |
| 12274 | MT897907 | Unassigned | Unassigned | Unassigned | 514_0 | Clustered | 75 | VC_514_0 | 75 | 0.9402 | 1 | 0.9402 | 0 | 0 | 0 | 1 | Ref |
| 12275 | MT897908 | Unassigned | Unassigned | Unassigned | 859_0 | Clustered | 9 | VC_859_0 | 9 | 0.9426 | 1 | 0.9426 | 0 | 0 | 0 | 1 | Ref |
| 12276 | MT897909 | Unassigned | Unassigned | Unassigned | 371_16 | Clustered | 42 | VC_371_16 | 42 | 0.1341 | 1 | 0.1341 | 0 | 0 | 0 | 0.7282 | Ref |
| 12277 | MT897910 | Unassigned | Unassigned | Unassigned | 372_0 | Clustered | 24 | VC_372_0 | 24 | 0.5539 | 1 | 0.5539 | 0 | 0 | 0 | 1 | Ref |
| 12278 | MT899443 | Unassigned | Unassigned | Unassigned | 730_0 | Clustered | 9 | VC_730_0 | 9 | 1 | 1 | 1 | 0 | 0 | 0 | 1 | Ref |
| 12279 | MT899444 | Unassigned | Unassigned | Unassigned | 730_0 | Clustered | 9 | VC_730_0 | 9 | 1 | 1 | 1 | 0 | 0 | 0 | 1 | Ref |
| 12280 | MT899504 | Unassigned | Unassigned | Unassigned | 8_0 | Clustered | 58 | VC_8_0 | 58 | 0.8306 | 1 | 0.8306 | 0 | 0 | 0 | 0.9946 | Ref |
| 12281 | MT900487 | Unassigned | Unassigned | Unassigned | 333_0 | Clustered | 25 | VC_333_0 | 25 | 0.4681 | 1 | 0.4681 | 0 | 0 | 0 | 1 | Ref |
| 12282 | MT900488 | Unassigned | Unassigned | Unassigned | 412_0 | Clustered | 39 | VC_412_0 | 39 | 0.845 | 1 | 0.845 | 0 | 0 | 0 | 1 | Ref |
| 12283 | MT902335 | Unassigned | Unassigned | Unassigned | 422_0 | Clustered | 5 | VC_422_0 | 5 | 0.2592 | 0.99999997 | 0.2592 | 0 | 0 | 0 | 1 | Ref |
| 12284 | MT909815 | Unassigned | Unassigned | Unassigned | 92_0 | Clustered | 22 | VC_92_0 | 22 | 0.5639 | 1 | 0.5639 | 0 | 0 | 0 | 1 | Ref |
| 12285 | MT920315 | Unassigned | Unassigned | Unassigned | 490_6 | Clustered/Singleton | 1 | VC_490_6 | 1 | 0 | 1 | 0 | 0 | 0 | 0 | 1 | Ref |
| 12286 | MT926124 | Unassigned | Unassigned | Unassigned | 96_0 | Clustered | 30 | VC_96_0 | 30 | 0.718 | 1 | 0.718 | 0 | 0 | 0 | 1 | Ref |
| 12287 | MT932211 | Unassigned | Unassigned | Unassigned | 848_0 | Clustered | 28 | VC_848_0 | 28 | 0.7927 | 1 | 0.7927 | 0 | 0 | 0 | 1 | Ref |
| 12288 | MT932212 | Unassigned | Unassigned | Unassigned | 28_1 | Clustered | 53 | VC_28_1 | 53 | 0.7164 | 1 | 0.7164 | 0 | 0 | 0 | 0.9325 | Ref |
| 12289 | MT932213 | Unassigned | Unassigned | Unassigned | 283_93 | Clustered/Singleton | 1 | VC_283_93 | 1 | 0 | 1 | 0 | 0 | 0 | 0 | 1 | Ref |
| 12290 | MT932329 | Unassigned | Unassigned | Unassigned | 843_0 | Clustered | 5 | VC_843_0 | 5 | 0.2843 | 0.99999994 | 0.2843 | 0 | 0 | 0 | 1 | Ref |
| 12291 | MT933737 | Unassigned | Unassigned | Unassigned | 79_0 | Clustered | 64 | VC_79_0 | 64 | 0.4185 | 1 | 0.4185 | 0 | 0 | 0 | 0.7649 | Ref |
| 12292 | MT936332 | Unassigned | Unassigned | Unassigned | 859_0 | Clustered | 9 | VC_859_0 | 9 | 0.9426 | 1 | 0.9426 | 0 | 0 | 0 | 1 | Ref |
| 12293 | MT939241 | Unassigned | Unassigned | Unassigned | 691_0 | Clustered | 2 | VC_691_0 | 2 | 0.1052 | 0.95343038 | 0.1003 | 0 | 0 | 0 | 1 | Ref |
| 12294 | MT939252 | Unassigned | Unassigned | Unassigned | 111_0 | Clustered | 91 | VC_111_0 | 91 | 0.9587 | 1 | 0.9587 | 0 | 0 | 0 | 1 | Ref |
| 12295 | MT939253 | Unassigned | Unassigned | Unassigned | 582_3 | Clustered | 12 | VC_582_3 | 12 | 0.2357 | 1 | 0.2357 | 0 | 0 | 0 | 0.8636 | Ref |
| 12296 | MT939486 | Unassigned | Unassigned | Unassigned | 164_7 | Clustered | 7 | VC_164_7 | 7 | 0.0956 | 0.99998 | 0.0956 | 0 | 0 | 0 | 1 | Ref |
| 12297 | MT939487 | Unassigned | Unassigned | Unassigned | 164_7 | Clustered | 7 | VC_164_7 | 7 | 0.0956 | 0.99998 | 0.0956 | 0 | 0 | 0 | 1 | Ref |
| 12298 | MT939488 | Unassigned | Unassigned | Unassigned | 164_7 | Clustered | 7 | VC_164_7 | 7 | 0.0956 | 0.99998 | 0.0956 | 0 | 0 | 0 | 1 | Ref |
| 12299 | MT941682 | Unassigned | Unassigned | Unassigned | 1027_0 | Clustered | 20 | VC_1027_0 | 20 | 0.7114 | 1 | 0.7114 | 0 | 0 | 0 | 1 | Ref |
| 12300 | MT944117 | Unassigned | Unassigned | Unassigned | 847_1 | Clustered | 26 | VC_847_1 | 26 | 0.6323 | 1 | 0.6323 | 0 | 0 | 0 | 0.9815 | Ref |
| 12301 | MT947439 | Unassigned | Unassigned | Unassigned | 303_0 | Clustered | 155 | VC_303_0 | 155 | 0.9979 | 1 | 0.9979 | 0 | 0 | 0 | 1 | Ref |
| 12302 | MT949699 | Unassigned | Unassigned | Unassigned | 492_2 | Clustered | 8 | VC_492_2 | 8 | 0.1221 | 0.98786574 | 0.1206 | 0 | 0 | 0 | 1 | Ref |
| 12303 | MT951623 | Unassigned | Unassigned | Unassigned | 835_0 | Clustered | 67 | VC_835_0 | 67 | 0.9624 | 1 | 0.9624 | 0 | 0 | 0 | 1 | Ref |
| 12304 | MT952005 | Unassigned | Unassigned | Unassigned | 597_0 | Clustered | 6 | VC_597_0 | 6 | 0.1801 | 1 | 0.1801 | 0 | 0 | 0 | 1 | Ref |
| 12305 | MT952844 | Unassigned | Unassigned | Unassigned | 525_0 | Clustered | 25 | VC_525_0 | 25 | 0.4597 | 1 | 0.4597 | 0 | 0 | 0 | 1 | Ref |
| 12306 | MT952845 | Unassigned | Unassigned | Unassigned | 1206_0 | Clustered | 8 | VC_1206_0 | 8 | 0.1913 | 1 | 0.1913 | 0 | 0 | 0 | 1 | Ref |
| 12307 | MT952846 | Unassigned | Unassigned | Unassigned | 1240_0 | Clustered | 82 | VC_1240_0 | 82 | 0.7945 | 1 | 0.7945 | 0 | 0 | 0 | 1 | Ref |
| 12308 | MT952847 | Unassigned | Unassigned | Unassigned | 325_38 | Clustered | 6 | VC_325_38 | 6 | 0.0267 | 1 | 0.0267 | 0 | 0 | 0 | 0.8 | Ref |
| 12309 | MT952848 | Unassigned | Unassigned | Unassigned | 1440_0 | Clustered | 4 | VC_1440_0 | 4 | 1 | 1 | 1 | 0 | 0 | 0 | 1 | Ref |
| 12310 | MT952849 | Unassigned | Unassigned | Unassigned | 371_12 | Clustered | 4 | VC_371_12 | 4 | 0.0126 | 0.99998489 | 0.0126 | 0 | 0 | 0 | 0.8333 | Ref |

| | | | | | | | | | | | | | | | | | |
|---|---|---|---|---|---|---|---|---|---|---|---|---|---|---|---|---|---|
| 12311 | MT952850 | Unassigned | Unassigned | Unassigned | 1344_0 | Clustered | 78 | VC_1344_0 | 78 | 0.9732 | 1 | 0.9732 | 0 | 0 | 0 | 1 | Ref |
| 12312 | MT952851 | Unassigned | Unassigned | Unassigned | 509_0 | Clustered | 37 | VC_509_0 | 37 | 0.9689 | 1 | 0.9689 | 0 | 0 | 0 | 1 | Ref |
| 12313 | MT952852 | Unassigned | Unassigned | Unassigned | 1072_4 | Clustered | 4 | VC_1072_4 | 4 | 0.0736 | 0.99956559 | 0.0736 | 0 | 0 | 0 | 1 | Ref |
| 12314 | MT952853 | Unassigned | Unassigned | Unassigned | 1166_0 | Clustered | 20 | VC_1166_0 | 20 | 0.99 | 1 | 0.99 | 0 | 0 | 0 | 1 | Ref |
| 12315 | MT952854 | Unassigned | Unassigned | Unassigned | 551_6 | Clustered | 22 | VC_551_6 | 22 | 0.1214 | 1 | 0.1214 | 0 | 0 | 0 | 0.8095 | Ref |
| 12316 | MT956952 | Unassigned | Unassigned | Unassigned | 272_0 | Clustered | 11 | VC_272_0 | 11 | 0.4915 | 1 | 0.4915 | 0 | 0 | 0 | 0.8727 | Ref |
| 12317 | MT966872 | Unassigned | Unassigned | Unassigned | 823_0 | Clustered | 124 | VC_823_0 | 124 | 0.9492 | 1 | 0.9492 | 0 | 0 | 0 | 1 | Ref |
| 12318 | MT966873 | Unassigned | Unassigned | Unassigned | 823_0 | Clustered | 124 | VC_823_0 | 124 | 0.9492 | 1 | 0.9492 | 0 | 0 | 0 | 1 | Ref |
| 12319 | MT968995 | Unassigned | Unassigned | Unassigned | 283_173 | Clustered/Singleton | 1 | VC_283_173 | 1 | 0 | 1 | 0 | 0 | 0 | 0 | 1 | Ref |
| 12320 | MT974436 | Unassigned | Unassigned | Unassigned | 133_7 | Clustered/Singleton | 1 | VC_133_7 | 1 | 0 | 1 | 0 | 0 | 0 | 0 | 1 | Ref |
| 12321 | MT975991 | Unassigned | Unassigned | Unassigned | 1441_0 | Clustered | 2 | VC_1441_0 | 2 | 1 | 1 | 1 | 0 | 0 | 0 | 1 | Ref |
| 12322 | MT980836 | Unassigned | Unassigned | Unassigned | 337_0 | Clustered | 6 | VC_337_0 | 6 | 0.6058 | 1 | 0.6058 | 0 | 0 | 0 | 1 | Ref |
| 12323 | MT980837 | Unassigned | Unassigned | Unassigned | 337_0 | Clustered | 6 | VC_337_0 | 6 | 0.6058 | 1 | 0.6058 | 0 | 0 | 0 | 1 | Ref |
| 12324 | MT980838 | Unassigned | Unassigned | Unassigned | 337_0 | Clustered | 6 | VC_337_0 | 6 | 0.6058 | 1 | 0.6058 | 0 | 0 | 0 | 1 | Ref |
| 12325 | MT980839 | Unassigned | Unassigned | Unassigned | 337_0 | Clustered | 6 | VC_337_0 | 6 | 0.6058 | 1 | 0.6058 | 0 | 0 | 0 | 1 | Ref |
| 12326 | MT980840 | Unassigned | Unassigned | Unassigned | 337_0 | Clustered | 6 | VC_337_0 | 6 | 0.6058 | 1 | 0.6058 | 0 | 0 | 0 | 1 | Ref |
| 12327 | MT980841 | Unassigned | Unassigned | Unassigned | 337_0 | Clustered | 6 | VC_337_0 | 6 | 0.6058 | 1 | 0.6058 | 0 | 0 | 0 | 1 | Ref |
| 12328 | MT984581 | Unassigned | Unassigned | Unassigned | 283_91 | Clustered | 11 | VC_283_91 | 11 | 0.0285 | 1 | 0.0285 | 0 | 0 | 0 | 1 | Ref |
| 12329 | MT986025 | Unassigned | Unassigned | Unassigned | 767_0 | Clustered | 34 | VC_767_0 | 34 | 0.9173 | 1 | 0.9173 | 0 | 0 | 0 | 1 | Ref |
| 12330 | MT986026 | Unassigned | Unassigned | Unassigned | 1240_0 | Clustered | 82 | VC_1240_0 | 82 | 0.7945 | 1 | 0.7945 | 0 | 0 | 0 | 1 | Ref |
| 12331 | MT986027 | Unassigned | Unassigned | Unassigned | 1240_0 | Clustered | 82 | VC_1240_0 | 82 | 0.7945 | 1 | 0.7945 | 0 | 0 | 0 | 1 | Ref |
| 12332 | MT986028 | Unassigned | Unassigned | Unassigned | 857_0 | Clustered | 11 | VC_857_0 | 11 | 0.9712 | 1 | 0.9712 | 0 | 0 | 0 | 1 | Ref |
| 12333 | MT986029 | Unassigned | Unassigned | Unassigned | 868_0 | Clustered | 25 | VC_868_0 | 25 | 0.5791 | 1 | 0.5791 | 0 | 0 | 0 | 1 | Ref |
| 12334 | MT992243 | Unassigned | Unassigned | Unassigned | 608_0 | Clustered | 5 | VC_608_0 | 5 | 0.4331 | 0.99999996 | 0.4331 | 0 | 0 | 0 | 1 | Ref |
| 12335 | MT995926 | Unassigned | Unassigned | Unassigned | 344_0 | Clustered | 21 | VC_344_0 | 21 | 0.5436 | 1 | 0.5436 | 0 | 0 | 0 | 1 | Ref |
| 12336 | MT995927 | Unassigned | Unassigned | Unassigned | 344_0 | Clustered | 21 | VC_344_0 | 21 | 0.5436 | 1 | 0.5436 | 0 | 0 | 0 | 1 | Ref |
| 12337 | MT995928 | Unassigned | Unassigned | Unassigned | 344_0 | Clustered | 21 | VC_344_0 | 21 | 0.5436 | 1 | 0.5436 | 0 | 0 | 0 | 1 | Ref |
| 12338 | MW001214 | Unassigned | Unassigned | Unassigned | 87_0 | Clustered | 8 | VC_87_0 | 8 | 0.5545 | 1 | 0.5545 | 0 | 0 | 0 | 1 | Ref |
| 12339 | MW001769 | Unassigned | Unassigned | Unassigned | 389_0 | Clustered | 3 | VC_389_0 | 3 | 0.0515 | 0.99861557 | 0.0514 | 0 | 0 | 0 | 1 | Ref |
| 12340 | MW004544 | Unassigned | Unassigned | Unassigned | 92_0 | Clustered | 22 | VC_92_0 | 22 | 0.5639 | 1 | 0.5639 | 0 | 0 | 0 | 1 | Ref |
| 12341 | MW004545 | Unassigned | Unassigned | Unassigned | 92_0 | Clustered | 22 | VC_92_0 | 22 | 0.5639 | 1 | 0.5639 | 0 | 0 | 0 | 1 | Ref |
| 12342 | MW006474 | Unassigned | Unassigned | Unassigned | 329_1 | Clustered | 17 | VC_329_1 | 17 | 0.0867 | 1 | 0.0867 | 0 | 0 | 0 | 0.8235 | Ref |
| 12343 | MW006475 | Unassigned | Unassigned | Unassigned | 329_1 | Clustered | 17 | VC_329_1 | 17 | 0.0867 | 1 | 0.0867 | 0 | 0 | 0 | 0.8235 | Ref |
| 12344 | MW006476 | Unassigned | Unassigned | Unassigned | 133_11 | Clustered | 2 | VC_133_11 | 2 | 0.006 | 0.93904954 | 0.0056 | 0 | 0 | 0 | 1 | Ref |
| 12345 | MW006477 | Unassigned | Unassigned | Unassigned | 582_5 | Clustered | 4 | VC_582_5 | 4 | 0.0528 | 0.99935625 | 0.0527 | 0 | 0 | 0 | 0.8333 | Ref |
| 12346 | MW006478 | Unassigned | Unassigned | Unassigned | 191_0 | Clustered | 2 | VC_191_0 | 2 | 0.0067 | 0.95760971 | 0.0064 | 0 | 0 | 0 | 1 | Ref |
| 12347 | MW006479 | Unassigned | Unassigned | Unassigned | 190_40 | Clustered | 2 | VC_190_40 | 2 | 0.0083 | 0.95556395 | 0.0079 | 0 | 0 | 0 | 1 | Ref |
| 12348 | MW006480 | Unassigned | Unassigned | Unassigned | 133_11 | Clustered | 2 | VC_133_11 | 2 | 0.006 | 0.93904954 | 0.0056 | 0 | 0 | 0 | 1 | Ref |
| 12349 | MW006481 | Unassigned | Unassigned | Unassigned | 191_0 | Clustered | 2 | VC_191_0 | 2 | 0.0067 | 0.95760971 | 0.0064 | 0 | 0 | 0 | 1 | Ref |
| 12350 | MW006482 | Unassigned | Unassigned | Unassigned | 329_1 | Clustered | 17 | VC_329_1 | 17 | 0.0867 | 1 | 0.0867 | 0 | 0 | 0 | 0.8235 | Ref |
| 12351 | MW012634 | Unassigned | Unassigned | Unassigned | 818_0 | Clustered | 20 | VC_818_0 | 20 | 0.9601 | 1 | 0.9601 | 0 | 0 | 0 | 1 | Ref |
| 12352 | MW013502 | Unassigned | Unassigned | Unassigned | 28_1 | Clustered | 53 | VC_28_1 | 53 | 0.7164 | 1 | 0.7164 | 0 | 0 | 0 | 0.9325 | Ref |
| 12353 | MW013503 | Unassigned | Unassigned | Unassigned | 28_1 | Clustered | 53 | VC_28_1 | 53 | 0.7164 | 1 | 0.7164 | 0 | 0 | 0 | 0.9325 | Ref |
| 12354 | MW015080 | Unassigned | Unassigned | Unassigned | 340_0 | Clustered | 31 | VC_340_0 | 31 | 0.9549 | 1 | 0.9549 | 0 | 0 | 0 | 1 | Ref |
| 12355 | MW015081 | Unassigned | Unassigned | Unassigned | 432_12 | Clustered/Singleton | 1 | VC_432_12 | 1 | 0 | 1 | 0 | 0 | 0 | 0 | 1 | Ref |
| 12356 | MW021748 | Unassigned | Unassigned | Unassigned | 584_0 | Clustered | 28 | VC_584_0 | 28 | 0.7704 | 1 | 0.7704 | 0 | 0 | 0 | 1 | Ref |
| 12357 | MW021749 | Unassigned | Unassigned | Unassigned | 669_1 | Clustered | 10 | VC_669_1 | 10 | 0.1268 | 1 | 0.1268 | 0 | 0 | 0 | 0.9778 | Ref |
| 12358 | MW021750 | Unassigned | Unassigned | Unassigned | 584_0 | Clustered | 28 | VC_584_0 | 28 | 0.7704 | 1 | 0.7704 | 0 | 0 | 0 | 1 | Ref |
| 12359 | MW021751 | Unassigned | Unassigned | Unassigned | 669_0 | Clustered | 21 | VC_669_0 | 21 | 0.3623 | 1 | 0.3623 | 0 | 0 | 0 | 1 | Ref |
| 12360 | MW021752 | Unassigned | Unassigned | Unassigned | 491_0 | Clustered | 36 | VC_491_0 | 36 | 0.5336 | 1 | 0.5336 | 0 | 0 | 0 | 0.9714 | Ref |
| 12361 | MW021753 | Unassigned | Unassigned | Unassigned | 133_22 | Clustered | 13 | VC_133_22 | 13 | 0.0804 | 1 | 0.0804 | 0 | 0 | 0 | 0.8205 | Ref |
| 12362 | MW021754 | Unassigned | Unassigned | Unassigned | 133_44 | Clustered/Singleton | 1 | VC_133_44 | 1 | 0 | 1 | 0 | 0 | 0 | 0 | 1 | Ref |
| 12363 | MW021755 | Unassigned | Unassigned | Unassigned | 1346_0 | Clustered | 10 | VC_1346_0 | 10 | 0.7784 | 1 | 0.7784 | 0 | 0 | 0 | 1 | Ref |
| 12364 | MW021756 | Unassigned | Unassigned | Unassigned | 669_0 | Clustered | 21 | VC_669_0 | 21 | 0.3623 | 1 | 0.3623 | 0 | 0 | 0 | 1 | Ref |
| 12365 | MW021757 | Unassigned | Unassigned | Unassigned | 133_22 | Clustered | 13 | VC_133_22 | 13 | 0.0804 | 1 | 0.0804 | 0 | 0 | 0 | 0.8205 | Ref |
| 12366 | MW021758 | Unassigned | Unassigned | Unassigned | 488_1 | Clustered | 8 | VC_488_1 | 8 | 0.2927 | 1 | 0.2927 | 0 | 0 | 0 | 1 | Ref |
| 12367 | MW021760 | Unassigned | Unassigned | Unassigned | 133_46 | Clustered/Singleton | 1 | VC_133_46 | 1 | 0 | 1 | 0 | 0 | 0 | 0 | 1 | Ref |
| 12368 | MW021762 | Unassigned | Unassigned | Unassigned | 937_0 | Clustered | 40 | VC_937_0 | 40 | 0.6809 | 1 | 0.6809 | 0 | 0 | 0 | 0.9359 | Ref |
| 12369 | MW021763 | Unassigned | Unassigned | Unassigned | 1346_0 | Clustered | 10 | VC_1346_0 | 10 | 0.7784 | 1 | 0.7784 | 0 | 0 | 0 | 1 | Ref |
| 12370 | MW021764 | Unassigned | Unassigned | Unassigned | 937_0 | Clustered | 40 | VC_937_0 | 40 | 0.6809 | 1 | 0.6809 | 0 | 0 | 0 | 0.9359 | Ref |
| 12371 | MW021765 | Unassigned | Unassigned | Unassigned | 937_0 | Clustered | 40 | VC_937_0 | 40 | 0.6809 | 1 | 0.6809 | 0 | 0 | 0 | 0.9359 | Ref |
| 12372 | MW021766 | Unassigned | Unassigned | Unassigned | 1346_0 | Clustered | 10 | VC_1346_0 | 10 | 0.7784 | 1 | 0.7784 | 0 | 0 | 0 | 1 | Ref |
| 12373 | MW023914 | Unassigned | Unassigned | Unassigned | 481_0 | Clustered | 8 | VC_481_0 | 8 | 0.1348 | 1 | 0.1348 | 0 | 0 | 0 | 1 | Ref |
| 12374 | MW032452 | Unassigned | Unassigned | Unassigned | 585_0 | Clustered | 4 | VC_585_0 | 4 | 0.0533 | 0.99998755 | 0.0533 | 0 | 0 | 0 | 1 | Ref |
| 12375 | MW032477 | Unassigned | Unassigned | Unassigned | 202_0 | Clustered | 102 | VC_202_0 | 102 | 0.4049 | 1 | 0.4049 | 0 | 0 | 0 | 0.7876 | Ref |
| 12376 | MW041632 | Unassigned | Unassigned | Unassigned | 202_0 | Clustered | 102 | VC_202_0 | 102 | 0.4049 | 1 | 0.4049 | 0 | 0 | 0 | 0.7876 | Ref |
| 12377 | MW041633 | Unassigned | Unassigned | Unassigned | 202_0 | Clustered | 102 | VC_202_0 | 102 | 0.4049 | 1 | 0.4049 | 0 | 0 | 0 | 0.7876 | Ref |
| 12378 | MW041634 | Unassigned | Unassigned | Unassigned | 202_0 | Clustered | 102 | VC_202_0 | 102 | 0.4049 | 1 | 0.4049 | 0 | 0 | 0 | 0.7876 | Ref |
| 12379 | MW041635 | Unassigned | Unassigned | Unassigned | 202_0 | Clustered | 102 | VC_202_0 | 102 | 0.4049 | 1 | 0.4049 | 0 | 0 | 0 | 0.7876 | Ref |
| 12380 | MW041636 | Unassigned | Unassigned | Unassigned | 202_0 | Clustered | 102 | VC_202_0 | 102 | 0.4049 | 1 | 0.4049 | 0 | 0 | 0 | 0.7876 | Ref |
| 12381 | MW041637 | Unassigned | Unassigned | Unassigned | 202_0 | Clustered | 102 | VC_202_0 | 102 | 0.4049 | 1 | 0.4049 | 0 | 0 | 0 | 0.7876 | Ref |
| 12382 | MW041638 | Unassigned | Unassigned | Unassigned | 202_0 | Clustered | 102 | VC_202_0 | 102 | 0.4049 | 1 | 0.4049 | 0 | 0 | 0 | 0.7876 | Ref |
| 12383 | MW041639 | Unassigned | Unassigned | Unassigned | 202_0 | Clustered | 102 | VC_202_0 | 102 | 0.4049 | 1 | 0.4049 | 0 | 0 | 0 | 0.7876 | Ref |
| 12384 | MW041640 | Unassigned | Unassigned | Unassigned | 202_0 | Clustered | 102 | VC_202_0 | 102 | 0.4049 | 1 | 0.4049 | 0 | 0 | 0 | 0.7876 | Ref |
| 12385 | MW042787 | Unassigned | Unassigned | Unassigned | 823_0 | Clustered | 124 | VC_823_0 | 124 | 0.9492 | 1 | 0.9492 | 0 | 0 | 0 | 1 | Ref |
| 12386 | MW042788 | Unassigned | Unassigned | Unassigned | 823_0 | Clustered | 124 | VC_823_0 | 124 | 0.9492 | 1 | 0.9492 | 0 | 0 | 0 | 1 | Ref |
| 12387 | MW042789 | Unassigned | Unassigned | Unassigned | 823_0 | Clustered | 124 | VC_823_0 | 124 | 0.9492 | 1 | 0.9492 | 0 | 0 | 0 | 1 | Ref |
| 12388 | MW042790 | Unassigned | Unassigned | Unassigned | 823_0 | Clustered | 124 | VC_823_0 | 124 | 0.9492 | 1 | 0.9492 | 0 | 0 | 0 | 1 | Ref |
| 12389 | MW042791 | Unassigned | Unassigned | Unassigned | 823_0 | Clustered | 124 | VC_823_0 | 124 | 0.9492 | 1 | 0.9492 | 0 | 0 | 0 | 1 | Ref |
| 12390 | MW042792 | Unassigned | Unassigned | Unassigned | 823_0 | Clustered | 124 | VC_823_0 | 124 | 0.9492 | 1 | 0.9492 | 0 | 0 | 0 | 1 | Ref |
| 12391 | MW042793 | Unassigned | Unassigned | Unassigned | 823_0 | Clustered | 124 | VC_823_0 | 124 | 0.9492 | 1 | 0.9492 | 0 | 0 | 0 | 1 | Ref |
| 12392 | MW042794 | Unassigned | Unassigned | Unassigned | 823_0 | Clustered | 124 | VC_823_0 | 124 | 0.9492 | 1 | 0.9492 | 0 | 0 | 0 | 1 | Ref |
| 12393 | MW042795 | Unassigned | Unassigned | Unassigned | 823_0 | Clustered | 124 | VC_823_0 | 124 | 0.9492 | 1 | 0.9492 | 0 | 0 | 0 | 1 | Ref |
| 12394 | MW042796 | Unassigned | Unassigned | Unassigned | 823_0 | Clustered | 124 | VC_823_0 | 124 | 0.9492 | 1 | 0.9492 | 0 | 0 | 0 | 1 | Ref |
| 12395 | MW042797 | Unassigned | Unassigned | Unassigned | 823_0 | Clustered | 124 | VC_823_0 | 124 | 0.9492 | 1 | 0.9492 | 0 | 0 | 0 | 1 | Ref |
| 12396 | MW042798 | Unassigned | Unassigned | Unassigned | 823_0 | Clustered | 124 | VC_823_0 | 124 | 0.9492 | 1 | 0.9492 | 0 | 0 | 0 | 1 | Ref |
| 12397 | MW042799 | Unassigned | Unassigned | Unassigned | 823_0 | Clustered | 124 | VC_823_0 | 124 | 0.9492 | 1 | 0.9492 | 0 | 0 | 0 | 1 | Ref |
| 12398 | MW042800 | Unassigned | Unassigned | Unassigned | 823_0 | Clustered | 124 | VC_823_0 | 124 | 0.9492 | 1 | 0.9492 | 0 | 0 | 0 | 1 | Ref |
| 12399 | MW042801 | Unassigned | Unassigned | Unassigned | 823_0 | Clustered | 124 | VC_823_0 | 124 | 0.9492 | 1 | 0.9492 | 0 | 0 | 0 | 1 | Ref |
| 12400 | MW042802 | Unassigned | Unassigned | Unassigned | 937_0 | Clustered | 40 | VC_937_0 | 40 | 0.6809 | 1 | 0.6809 | 0 | 0 | 0 | 0.9359 | Ref |
| 12401 | MW042803 | Unassigned | Unassigned | Unassigned | 823_0 | Clustered | 124 | VC_823_0 | 124 | 0.9492 | 1 | 0.9492 | 0 | 0 | 0 | 1 | Ref |
| 12402 | MW042804 | Unassigned | Unassigned | Unassigned | 823_0 | Clustered | 124 | VC_823_0 | 124 | 0.9492 | 1 | 0.9492 | 0 | 0 | 0 | 1 | Ref |
| 12403 | MW042805 | Unassigned | Unassigned | Unassigned | 823_0 | Clustered | 124 | VC_823_0 | 124 | 0.9492 | 1 | 0.9492 | 0 | 0 | 0 | 1 | Ref |
| 12404 | MW042806 | Unassigned | Unassigned | Unassigned | 823_0 | Clustered | 124 | VC_823_0 | 124 | 0.9492 | 1 | 0.9492 | 0 | 0 | 0 | 1 | Ref |
| 12405 | MW042807 | Unassigned | Unassigned | Unassigned | 823_0 | Clustered | 124 | VC_823_0 | 124 | 0.9492 | 1 | 0.9492 | 0 | 0 | 0 | 1 | Ref |
| 12406 | MW042808 | Unassigned | Unassigned | Unassigned | 823_0 | Clustered | 124 | VC_823_0 | 124 | 0.9492 | 1 | 0.9492 | 0 | 0 | 0 | 1 | Ref |
| 12407 | MW042809 | Unassigned | Unassigned | Unassigned | 823_0 | Clustered | 124 | VC_823_0 | 124 | 0.9492 | 1 | 0.9492 | 0 | 0 | 0 | 1 | Ref |
| 12408 | MW042810 | Unassigned | Unassigned | Unassigned | 823_0 | Clustered | 124 | VC_823_0 | 124 | 0.9492 | 1 | 0.9492 | 0 | 0 | 0 | 1 | Ref |
| 12409 | MW042811 | Unassigned | Unassigned | Unassigned | 823_0 | Clustered | 124 | VC_823_0 | 124 | 0.9492 | 1 | 0.9492 | 0 | 0 | 0 | 1 | Ref |
| 12410 | MW042812 | Unassigned | Unassigned | Unassigned | 823_0 | Clustered | 124 | VC_823_0 | 124 | 0.9492 | 1 | 0.9492 | 0 | 0 | 0 | 1 | Ref |
| 12411 | MW043865 | Unassigned | Unassigned | Unassigned | 988_0 | Clustered | 2 | VC_988_0 | 2 | 0.1586 | 0.95268934 | 0.1511 | 0 | 0 | 0 | 1 | Ref |
| 12412 | MW055901 | Unassigned | Unassigned | Unassigned | 1177_0 | Clustered | 14 | VC_1177_0 | 14 | 0.7847 | 1 | 0.7847 | 0 | 0 | 0 | 0.989 | Ref |
| 12413 | MW055902 | Unassigned | Unassigned | Unassigned | 519_0 | Clustered | 39 | VC_519_0 | 39 | 0.8156 | 1 | 0.8156 | 0 | 0 | 0 | 1 | Ref |
| 12414 | MW055903 | Unassigned | Unassigned | Unassigned | 1343_0 | Clustered | 21 | VC_1343_0 | 21 | 0.9843 | 1 | 0.9843 | 0 | 0 | 0 | 1 | Ref |
| 12415 | MW055904 | Unassigned | Unassigned | Unassigned | 221_0 | Clustered | 22 | VC_221_0 | 22 | 0.7447 | 1 | 0.7447 | 0 | 0 | 0 | 1 | Ref |
| 12416 | MW055905 | Unassigned | Unassigned | Unassigned | 872_0 | Clustered | 31 | VC_872_0 | 31 | 0.8151 | 1 | 0.8151 | 0 | 0 | 0 | 0.8194 | Ref |
| 12417 | MW055906 | Unassigned | Unassigned | Unassigned | 372_2 | Clustered | 5 | VC_372_2 | 5 | 0.072 | 0.99999997 | 0.072 | 0 | 0 | 0 | 0.9 | Ref |
| 12418 | MW055907 | Unassigned | Unassigned | Unassigned | 1344_0 | Clustered | 78 | VC_1344_0 | 78 | 0.9732 | 1 | 0.9732 | 0 | 0 | 0 | 1 | Ref |
| 12419 | MW055908 | Unassigned | Unassigned | Unassigned | 1175_0 | Clustered | 22 | VC_1175_0 | 22 | 0.4562 | 1 | 0.4562 | 0 | 0 | 0 | 1 | Ref |
| 12420 | MW055909 | Unassigned | Unassigned | Unassigned | 1344_0 | Clustered | 78 | VC_1344_0 | 78 | 0.9732 | 1 | 0.9732 | 0 | 0 | 0 | 1 | Ref |
| 12421 | MW055910 | Unassigned | Unassigned | Unassigned | 1207_0 | Clustered | 29 | VC_1207_0 | 29 | 0.8225 | 1 | 0.8225 | 0 | 0 | 0 | 1 | Ref |
| 12422 | MW055911 | Unassigned | Unassigned | Unassigned | 1240_0 | Clustered | 82 | VC_1240_0 | 82 | 0.7945 | 1 | 0.7945 | 0 | 0 | 0 | 1 | Ref |
| 12423 | MW055912 | Unassigned | Unassigned | Unassigned | 1164_0 | Clustered | 25 | VC_1164_0 | 25 | 0.7312 | 1 | 0.7312 | 0 | 0 | 0 | 1 | Ref |
| 12424 | MW055914 | Unassigned | Unassigned | Unassigned | 1360_0 | Clustered | 31 | VC_1360_0 | 31 | 0.8837 | 1 | 0.8837 | 0 | 0 | 0 | 1 | Ref |
| 12425 | MW055915 | Unassigned | Unassigned | Unassigned | 520_61 | Clustered/Singleton | 1 | VC_520_61 | 1 | 0 | 1 | 0 | 0 | 0 | 0 | 1 | Ref |
| 12426 | MW056501 | Unassigned | Unassigned | Unassigned | 699_0 | Clustered | 71 | VC_699_0 | 71 | 0.9352 | 1 | 0.9352 | 0 | 0 | 0 | 1 | Ref |
| 12427 | MW056502 | Unassigned | Unassigned | Unassigned | 699_0 | Clustered | 71 | VC_699_0 | 71 | 0.9352 | 1 | 0.9352 | 0 | 0 | 0 | 1 | Ref |
| 12428 | MW056503 | Unassigned | Unassigned | Unassigned | 699_0 | Clustered | 71 | VC_699_0 | 71 | 0.9352 | 1 | 0.9352 | 0 | 0 | 0 | 1 | Ref |
| 12429 | MW057854 | Unassigned | Unassigned | Unassigned | 396_0 | Clustered | 2 | VC_396_0 | 2 | 0.0714 | 0.95759359 | 0.0684 | 0 | 0 | 0 | 1 | Ref |
| 12430 | MW057855 | Unassigned | Unassigned | Unassigned | 242_1 | Clustered | 20 | VC_242_1 | 20 | 0.684 | 1 | 0.684 | 0 | 0 | 0 | 0.8368 | Ref |
| 12431 | MW057857 | Unassigned | Unassigned | Unassigned | 193_0 | Clustered | 2 | VC_193_0 | 2 | 0.0321 | 0.95762656 | 0.0308 | 0 | 0 | 0 | 1 | Ref |
| 12432 | MW057858 | Unassigned | Unassigned | Unassigned | 288_4 | Clustered/Singleton | 1 | VC_288_4 | 1 | 0 | 1 | 0 | 0 | 0 | 0 | 1 | Ref |
| 12433 | MW057859 | Unassigned | Unassigned | Unassigned | 51_0 | Clustered | 10 | VC_51_0 | 10 | 0.1741 | 1 | 0.1741 | 0 | 0 | 0 | 1 | Ref |
| 12434 | MW057860 | Unassigned | Unassigned | Unassigned | 592_0 | Clustered | 50 | VC_592_0 | 50 | 0.7314 | 1 | 0.7314 | 0 | 0 | 0 | 1 | Ref |
| 12435 | MW057918 | Unassigned | Unassigned | Unassigned | 456_0 | Clustered | 62 | VC_456_0 | 62 | 0.9026 | 1 | 0.9026 | 0 | 0 | 0 | 1 | Ref |
| 12436 | MW057919 | Unassigned | Unassigned | Unassigned | 344_0 | Clustered | 21 | VC_344_0 | 21 | 0.5436 | 1 | 0.5436 | 0 | 0 | 0 | 1 | Ref |
| 12437 | MW057920 | Unassigned | Unassigned | Unassigned | 898_0 | Clustered | 10 | VC_898_0 | 10 | 0.7284 | 1 | 0.7284 | 0 | 0 | 0 | 1 | Ref |
| 12438 | MW067000 | Unassigned | Unassigned | Unassigned | 673_0 | Clustered | 15 | VC_673_0 | 15 | 0.8234 | 1 | 0.8234 | 0 | 0 | 0 | 0.7619 | Ref |
| 12439 | MW067001 | Unassigned | Unassigned | Unassigned | 673_0 | Clustered | 15 | VC_673_0 | 15 | 0.8234 | 1 | 0.8234 | 0 | 0 | 0 | 0.7619 | Ref |
| 12440 | MW067002 | Unassigned | Unassigned | Unassigned | 673_0 | Clustered | 15 | VC_673_0 | 15 | 0.8234 | 1 | 0.8234 | 0 | 0 | 0 | 0.7619 | Ref |
| 12441 | MW067003 | Unassigned | Unassigned | Unassigned | 673_0 | Clustered | 15 | VC_673_0 | 15 | 0.8234 | 1 | 0.8234 | 0 | 0 | 0 | 0.7619 | Ref |
| 12442 | MW072790 | Unassigned | Unassigned | Unassigned | 3_0 | Clustered | 17 | VC_3_0 | 17 | 0.5639 | 1 | 0.5639 | 0 | 0 | 0 | 1 | Ref |
| 12443 | MW073017 | Unassigned | Unassigned | Unassigned | 478_1 | Clustered/Singleton | 1 | VC_478_1 | 1 | 0 | 1 | 0 | 0 | 0 | 0 | 1 | Ref |
| 12444 | MW073100 | Unassigned | Unassigned | Unassigned | 853_0 | Clustered | 2 | VC_853_0 | 2 | 0.27 | 0.72985431 | 0.1971 | 0 | 0 | 0 | 1 | Ref |
| 12445 | MW074885 | Unassigned | Unassigned | Unassigned | 392_0 | Clustered | 126 | VC_392_0 | 126 | 0.9621 | 1 | 0.9621 | 0 | 0 | 0 | 1 | Ref |
| 12446 | MW084976 | Unassigned | Unassigned | Unassigned | 475_7 | Clustered/Singleton | 1 | VC_475_7 | 1 | 0 | 1 | 0 | 0 | 0 | 0 | 1 | Ref |
| 12447 | MW091529 | Unassigned | Unassigned | Unassigned | 466_0 | Clustered | 3 | VC_466_0 | 3 | 0.3264 | 0.99747305 | 0.3255 | 0 | 0 | 0 | 1 | Ref |
| 12448 | MW114771 | Unassigned | Unassigned | Unassigned | 614_0 | Clustered | 5 | VC_614_0 | 5 | 0.2395 | 0.99999997 | 0.2395 | 0 | 0 | 0 | 1 | Ref |
| 12449 | MW117144 | Unassigned | Unassigned | Unassigned | 423_0 | Clustered | 45 | VC_423_0 | 45 | 0.9484 | 1 | 0.9484 | 0 | 0 | 0 | 1 | Ref |
| 12450 | MW117966 | Unassigned | Unassigned | Unassigned | 433_9 | Clustered/Singleton | 1 | VC_433_9 | 1 | 0 | 1 | 0 | 0 | 0 | 0 | 1 | Ref |
| 12451 | MW119258 | Unassigned | Unassigned | Unassigned | 460_0 | Clustered | 19 | VC_460_0 | 19 | 0.3744 | 1 | 0.3744 | 0 | 0 | 0 | 1 | Ref |
| 12452 | MW119570 | Unassigned | Unassigned | Unassigned | 761_0 | Clustered | 67 | VC_761_0 | 67 | 0.8947 | 1 | 0.8947 | 0 | 0 | 0 | 1 | Ref |
| 12453 | MW132713 | Unassigned | Unassigned | Unassigned | 406_0 | Clustered | 11 | VC_406_0 | 11 | 0.9449 | 1 | 0.9449 | 0 | 0 | 0 | 1 | Ref |
| 12454 | MW132714 | Unassigned | Unassigned | Unassigned | 546_8 | Clustered/Singleton | 1 | VC_546_8 | 1 | 0 | 1 | 0 | 0 | 0 | 0 | 1 | Ref |
| 12455 | MW145139 | Unassigned | Unassigned | Unassigned | 591_0 | Clustered | 14 | VC_591_0 | 14 | 0.5184 | 1 | 0.5184 | 0 | 0 | 0 | 1 | Ref |
| 12456 | MW145539 | Unassigned | Unassigned | Unassigned | 872_0 | Clustered | 31 | VC_872_0 | 31 | 0.8151 | 1 | 0.8151 | 0 | 0 | 0 | 0.8194 | Ref |
| 12457 | MW147366 | Unassigned | Unassigned | Unassigned | 7_0 | Clustered | 28 | VC_7_0 | 28 | 0.3916 | 1 | 0.3916 | 0 | 0 | 0 | 0.9947 | Ref |
| 12458 | MW147367 | Unassigned | Unassigned | Unassigned | 433_7 | Clustered/Singleton | 1 | VC_433_7 | 1 | 0 | 1 | 0 | 0 | 0 | 0 | 1 | Ref |
| 12459 | MW147598 | Unassigned | Unassigned | Unassigned | 792_0 | Clustered | 35 | VC_792_0 | 35 | 0.9828 | 1 | 0.9828 | 0 | 0 | 0 | 1 | Ref |
| 12460 | MW147599 | Unassigned | Unassigned | Unassigned | 792_0 | Clustered | 35 | VC_792_0 | 35 | 0.9828 | 1 | 0.9828 | 0 | 0 | 0 | 1 | Ref |
| 12461 | MW147600 | Unassigned | Unassigned | Unassigned | 792_0 | Clustered | 35 | VC_792_0 | 35 | 0.9828 | 1 | 0.9828 | 0 | 0 | 0 | 1 | Ref |
| 12462 | MW147601 | Unassigned | Unassigned | Unassigned | 792_0 | Clustered | 35 | VC_792_0 | 35 | 0.9828 | 1 | 0.9828 | 0 | 0 | 0 | 1 | Ref |
| 12463 | MW147602 | Unassigned | Unassigned | Unassigned | 792_0 | Clustered | 35 | VC_792_0 | 35 | 0.9828 | 1 | 0.9828 | 0 | 0 | 0 | 1 | Ref |
| 12464 | MW147603 | Unassigned | Unassigned | Unassigned | 386_0 | Clustered | 49 | VC_386_0 | 49 | 0.7941 | 1 | 0.7941 | 0 | 0 | 0 | 1 | Ref |
| 12465 | MW149272 | Unassigned | Unassigned | Unassigned | 592_0 | Clustered | 50 | VC_592_0 | 50 | 0.7314 | 1 | 0.7314 | 0 | 0 | 0 | 1 | Ref |
| 12466 | MW149273 | Unassigned | Unassigned | Unassigned | 190_23 | Clustered | 10 | VC_190_23 | 10 | 0.0704 | 1 | 0.0704 | 0 | 0 | 0 | 0.8444 | Ref |
| 12467 | MW149274 | Unassigned | Unassigned | Unassigned | 290_5 | Clustered | 14 | VC_290_5 | 14 | 0.1422 | 1 | 0.1422 | 0 | 0 | 0 | 0.9231 | Ref |
| 12468 | MW149275 | Unassigned | Unassigned | Unassigned | 190_23 | Clustered | 10 | VC_190_23 | 10 | 0.0704 | 1 | 0.0704 | 0 | 0 | 0 | 0.8444 | Ref |
| 12469 | MW161461 | Unassigned | Unassigned | Unassigned | 765_0 | Clustered | 132 | VC_765_0 | 132 | 1 | 1 | 1 | 0 | 0 | 0 | 1 | Ref |
| 12470 | MW161462 | Unassigned | Unassigned | Unassigned | 765_0 | Clustered | 132 | VC_765_0 | 132 | 1 | 1 | 1 | 0 | 0 | 0 | 1 | Ref |
| 12471 | MW161463 | Unassigned | Unassigned | Unassigned | 765_0 | Clustered | 132 | VC_765_0 | 132 | 1 | 1 | 1 | 0 | 0 | 0 | 1 | Ref |
| 12472 | MW161464 | Unassigned | Unassigned | Unassigned | 765_0 | Clustered | 132 | VC_765_0 | 132 | 1 | 1 | 1 | 0 | 0 | 0 | 1 | Ref |

| | | | | | | | | | | | | | | | | | |
|---|---|---|---|---|---|---|---|---|---|---|---|---|---|---|---|---|---|
| 12473 | MW161465 | Unassigned | Unassigned | Unassigned | 765_0 | Clustered | 132 | VC_765_0 | 132 | 1 | 1 | 1 | 0 | 0 | 0 | 1 | Ref |
| 12474 | MW161466 | Unassigned | Unassigned | Unassigned | 765_0 | Clustered | 132 | VC_765_0 | 132 | 1 | 1 | 1 | 0 | 0 | 0 | 1 | Ref |
| 12475 | MW161467 | Unassigned | Unassigned | Unassigned | 765_0 | Clustered | 132 | VC_765_0 | 132 | 1 | 1 | 1 | 0 | 0 | 0 | 1 | Ref |
| 12476 | MW161468 | Unassigned | Unassigned | Unassigned | 765_0 | Clustered | 132 | VC_765_0 | 132 | 1 | 1 | 1 | 0 | 0 | 0 | 1 | Ref |
| 12477 | MW175414 | Unassigned | Unassigned | Unassigned | 779_0 | Clustered | 37 | VC_779_0 | 37 | 0.5682 | 1 | 0.5682 | 0 | 0 | 0 | 1 | Ref |
| 12478 | MW175491 | Unassigned | Unassigned | Unassigned | 78_3 | Clustered | 11 | VC_78_3 | 11 | 0.2572 | 1 | 0.2572 | 0 | 0 | 0 | 1 | Ref |
| 12479 | MW175890 | Unassigned | Unassigned | Unassigned | 847_1 | Clustered | 26 | VC_847_1 | 26 | 0.6323 | 1 | 0.6323 | 0 | 0 | 0 | 0.9815 | Ref |
| 12480 | MW176032 | Unassigned | Unassigned | Unassigned | 608_0 | Clustered | 5 | VC_608_0 | 5 | 0.4331 | 0.99999996 | 0.4331 | 0 | 0 | 0 | 1 | Ref |
| 12481 | MW176033 | Unassigned | Unassigned | Unassigned | 670_0 | Clustered | 2 | VC_670_0 | 2 | 0.6875 | 0.72985431 | 0.5018 | 0 | 0 | 0 | 1 | Ref |
| 12482 | MW176034 | Unassigned | Unassigned | Unassigned | 608_0 | Clustered | 5 | VC_608_0 | 5 | 0.4331 | 0.99999996 | 0.4331 | 0 | 0 | 0 | 1 | Ref |
| 12483 | MW192778 | Unassigned | Unassigned | Unassigned | 7_0 | Clustered | 28 | VC_7_0 | 28 | 0.3916 | 1 | 0.3916 | 0 | 0 | 0 | 0.9947 | Ref |
| 12484 | MW201959 | Unassigned | Unassigned | Unassigned | 637_0 | Clustered | 115 | VC_637_0 | 115 | 0.731 | 1 | 0.731 | 0 | 0 | 0 | 0.8764 | Ref |
| 12485 | MW202412 | Unassigned | Unassigned | Unassigned | 995_0 | Clustered | 69 | VC_995_0 | 69 | 0.8547 | 1 | 0.8547 | 0 | 0 | 0 | 1 | Ref |
| 12486 | MW202414 | Unassigned | Unassigned | Unassigned | 995_0 | Clustered | 69 | VC_995_0 | 69 | 0.8547 | 1 | 0.8547 | 0 | 0 | 0 | 1 | Ref |
| 12487 | MW202445 | Unassigned | Unassigned | Unassigned | 1077_0 | Clustered | 48 | VC_1077_0 | 48 | 0.8525 | 1 | 0.8525 | 0 | 0 | 0 | 1 | Ref |
| 12488 | MW202455 | Unassigned | Unassigned | Unassigned | 427_0 | Clustered | 3 | VC_427_0 | 3 | 0.0627 | 0.96830121 | 0.0608 | 0 | 0 | 0 | 1 | Ref |
| 12489 | MW202462 | Unassigned | Unassigned | Unassigned | 1004_0 | Clustered | 2 | VC_1004_0 | 2 | 0.4638 | 0.94225536 | 0.437 | 0 | 0 | 0 | 1 | Ref |
| 12490 | MW202464 | Unassigned | Unassigned | Unassigned | 1082_0 | Clustered | 3 | VC_1082_0 | 3 | 0.6925 | 0.61954664 | 0.429 | 0 | 0 | 0 | 1 | Ref |
| 12491 | MW202466 | Unassigned | Unassigned | Unassigned | 995_0 | Clustered | 69 | VC_995_0 | 69 | 0.8547 | 1 | 0.8547 | 0 | 0 | 0 | 1 | Ref |
| 12492 | MW202471 | Unassigned | Unassigned | Unassigned | 995_0 | Clustered | 69 | VC_995_0 | 69 | 0.8547 | 1 | 0.8547 | 0 | 0 | 0 | 1 | Ref |
| 12493 | MW202473 | Unassigned | Unassigned | Unassigned | 527_0 | Clustered | 3 | VC_527_0 | 3 | 0.2387 | 0.99839883 | 0.2383 | 0 | 0 | 0 | 1 | Ref |
| 12494 | MW202475 | Unassigned | Unassigned | Unassigned | 163_0 | Clustered | 2 | VC_163_0 | 2 | 0.2865 | 0.94106069 | 0.2696 | 0 | 0 | 0 | 1 | Ref |
| 12495 | MW202478 | Unassigned | Unassigned | Unassigned | 527_0 | Clustered | 3 | VC_527_0 | 3 | 0.2387 | 0.99839883 | 0.2383 | 0 | 0 | 0 | 1 | Ref |
| 12496 | MW202486 | Unassigned | Unassigned | Unassigned | 1444_0 | Clustered | 3 | VC_1444_0 | 3 | 1 | 0 | 0 | 0 | 0 | 0 | 1 | Ref |
| 12497 | MW202489 | Unassigned | Unassigned | Unassigned | 341_0 | Clustered | 3 | VC_341_0 | 3 | 0.2522 | 0.9396546 | 0.237 | 0 | 0 | 0 | 1 | Ref |
| 12498 | MW202496 | Unassigned | Unassigned | Unassigned | 609_0 | Clustered | 4 | VC_609_0 | 4 | 0.489 | 0.99992419 | 0.489 | 0 | 0 | 0 | 1 | Ref |
| 12499 | MW202520 | Unassigned | Unassigned | Unassigned | 808_0 | Clustered | 2 | VC_808_0 | 2 | 0.3852 | 0.94765994 | 0.3651 | 0 | 0 | 0 | 1 | Ref |
| 12500 | MW202524 | Unassigned | Unassigned | Unassigned | 1446_0 | Clustered | 3 | VC_1446_0 | 3 | 1 | 0 | 0 | 0 | 0 | 0 | 1 | Ref |
| 12501 | MW202535 | Unassigned | Unassigned | Unassigned | 327_0 | Clustered | 12 | VC_327_0 | 12 | 0.8925 | 1 | 0.8925 | 0 | 0 | 0 | 1 | Ref |
| 12502 | MW202544 | Unassigned | Unassigned | Unassigned | 1004_0 | Clustered | 2 | VC_1004_0 | 2 | 0.4638 | 0.94225536 | 0.437 | 0 | 0 | 0 | 1 | Ref |
| 12503 | MW202545 | Unassigned | Unassigned | Unassigned | 1407_0 | Clustered | 2 | VC_1407_0 | 2 | 0.8176 | 0.81445332 | 0.6659 | 0 | 0 | 0 | 1 | Ref |
| 12504 | MW202549 | Unassigned | Unassigned | Unassigned | 995_0 | Clustered | 69 | VC_995_0 | 69 | 0.8547 | 1 | 0.8547 | 0 | 0 | 0 | 1 | Ref |
| 12505 | MW202557 | Unassigned | Unassigned | Unassigned | 806_0 | Clustered | 3 | VC_806_0 | 3 | 0.532 | 0.99824026 | 0.5311 | 0 | 0 | 0 | 1 | Ref |
| 12506 | MW202566 | Unassigned | Unassigned | Unassigned | 1223_0 | Clustered | 2 | VC_1223_0 | 2 | 1 | 1 | 1 | 0 | 0 | 0 | 1 | Ref |
| 12507 | MW202572 | Unassigned | Unassigned | Unassigned | 1445_0 | Clustered | 4 | VC_1445_0 | 4 | 0.9883 | 0.99615702 | 0.9845 | 0 | 0 | 0 | 1 | Ref |
| 12508 | MW202581 | Unassigned | Unassigned | Unassigned | 197_0 | Clustered | 5 | VC_197_0 | 5 | 0.1324 | 0.84698401 | 0.1122 | 0 | 0 | 0 | 1 | Ref |
| 12509 | MW202587 | Unassigned | Unassigned | Unassigned | 995_0 | Clustered | 69 | VC_995_0 | 69 | 0.8547 | 1 | 0.8547 | 0 | 0 | 0 | 1 | Ref |
| 12510 | MW202591 | Unassigned | Unassigned | Unassigned | 1444_0 | Clustered | 3 | VC_1444_0 | 3 | 1 | 0 | 0 | 0 | 0 | 0 | 1 | Ref |
| 12511 | MW202596 | Unassigned | Unassigned | Unassigned | 1222_0 | Clustered | 3 | VC_1222_0 | 3 | 1 | 0 | 0 | 0 | 0 | 0 | 1 | Ref |
| 12512 | MW202601 | Unassigned | Unassigned | Unassigned | 1447_0 | Clustered | 2 | VC_1447_0 | 2 | 1 | 1 | 1 | 0 | 0 | 0 | 1 | Ref |
| 12513 | MW202604 | Unassigned | Unassigned | Unassigned | 1447_0 | Clustered | 2 | VC_1447_0 | 2 | 1 | 1 | 1 | 0 | 0 | 0 | 1 | Ref |
| 12514 | MW202608 | Unassigned | Unassigned | Unassigned | 1448_0 | Clustered | 5 | VC_1448_0 | 5 | 1 | 1 | 1 | 0 | 0 | 0 | 1 | Ref |
| 12515 | MW202609 | Unassigned | Unassigned | Unassigned | 1077_0 | Clustered | 48 | VC_1077_0 | 48 | 0.8525 | 1 | 0.8525 | 0 | 0 | 0 | 1 | Ref |
| 12516 | MW202613 | Unassigned | Unassigned | Unassigned | 1442_0 | Clustered | 2 | VC_1442_0 | 2 | 0.9685 | 0.86822376 | 0.8408 | 0 | 0 | 0 | 1 | Ref |
| 12517 | MW202618 | Unassigned | Unassigned | Unassigned | 1089_0 | Clustered | 2 | VC_1089_0 | 2 | 0.546 | 0.5 | 0.273 | 0 | 0 | 0 | 1 | Ref |
| 12518 | MW202622 | Unassigned | Unassigned | Unassigned | 1449_0 | Clustered | 2 | VC_1449_0 | 2 | 1 | 1 | 1 | 0 | 0 | 0 | 1 | Ref |
| 12519 | MW202631 | Unassigned | Unassigned | Unassigned | 1445_0 | Clustered | 4 | VC_1445_0 | 4 | 0.9883 | 0.99615702 | 0.9845 | 0 | 0 | 0 | 1 | Ref |
| 12520 | MW202637 | Unassigned | Unassigned | Unassigned | 1443_0 | Clustered | 2 | VC_1443_0 | 2 | 0.8611 | 0.76024994 | 0.6547 | 0 | 0 | 0 | 1 | Ref |
| 12521 | MW202642 | Unassigned | Unassigned | Unassigned | 1450_0 | Clustered | 3 | VC_1450_0 | 3 | 1 | 1 | 1 | 0 | 0 | 0 | 1 | Ref |
| 12522 | MW202645 | Unassigned | Unassigned | Unassigned | 1445_0 | Clustered | 4 | VC_1445_0 | 4 | 0.9883 | 0.99615702 | 0.9845 | 0 | 0 | 0 | 1 | Ref |
| 12523 | MW202655 | Unassigned | Unassigned | Unassigned | 806_0 | Clustered | 3 | VC_806_0 | 3 | 0.532 | 0.99824026 | 0.5311 | 0 | 0 | 0 | 1 | Ref |
| 12524 | MW202657 | Unassigned | Unassigned | Unassigned | 1451_0 | Clustered | 2 | VC_1451_0 | 2 | 1 | 1 | 1 | 0 | 0 | 0 | 1 | Ref |
| 12525 | MW202658 | Unassigned | Unassigned | Unassigned | 1449_0 | Clustered | 2 | VC_1449_0 | 2 | 1 | 1 | 1 | 0 | 0 | 0 | 1 | Ref |
| 12526 | MW202665 | Unassigned | Unassigned | Unassigned | 117_0 | Clustered | 9 | VC_117_0 | 9 | 0.9977 | 0.98988637 | 0.9876 | 0 | 0 | 0 | 1 | Ref |
| 12527 | MW202671 | Unassigned | Unassigned | Unassigned | 1082_0 | Clustered | 3 | VC_1082_0 | 3 | 0.6925 | 0.61954664 | 0.429 | 0 | 0 | 0 | 1 | Ref |
| 12528 | MW202674 | Unassigned | Unassigned | Unassigned | 808_0 | Clustered | 2 | VC_808_0 | 2 | 0.3852 | 0.94765994 | 0.3651 | 0 | 0 | 0 | 1 | Ref |
| 12529 | MW202680 | Unassigned | Unassigned | Unassigned | 1368_0 | Clustered | 2 | VC_1368_0 | 2 | 1 | 1 | 1 | 0 | 0 | 0 | 1 | Ref |
| 12530 | MW202703 | Unassigned | Unassigned | Unassigned | 1243_0 | Clustered | 52 | VC_1243_0 | 52 | 0.9126 | 0.92708311 | 0.8461 | 0 | 0 | 0 | 1 | Ref |
| 12531 | MW202711 | Unassigned | Unassigned | Unassigned | 609_0 | Clustered | 4 | VC_609_0 | 4 | 0.489 | 0.99992419 | 0.489 | 0 | 0 | 0 | 1 | Ref |
| 12532 | MW202718 | Unassigned | Unassigned | Unassigned | 1446_0 | Clustered | 3 | VC_1446_0 | 3 | 1 | 0 | 0 | 0 | 0 | 0 | 1 | Ref |
| 12533 | MW202725 | Unassigned | Unassigned | Unassigned | 1448_0 | Clustered | 5 | VC_1448_0 | 5 | 1 | 1 | 1 | 0 | 0 | 0 | 1 | Ref |
| 12534 | MW202732 | Unassigned | Unassigned | Unassigned | 198_0 | Clustered | 28 | VC_198_0 | 28 | 0.2429 | 0.99971175 | 0.2429 | 0 | 0 | 0 | 1 | Ref |
| 12535 | MW202738 | Unassigned | Unassigned | Unassigned | 527_0 | Clustered | 3 | VC_527_0 | 3 | 0.2387 | 0.99839883 | 0.2383 | 0 | 0 | 0 | 1 | Ref |
| 12536 | MW202742 | Unassigned | Unassigned | Unassigned | 1445_0 | Clustered | 4 | VC_1445_0 | 4 | 0.9883 | 0.99615702 | 0.9845 | 0 | 0 | 0 | 1 | Ref |
| 12537 | MW202743 | Unassigned | Unassigned | Unassigned | 1446_0 | Clustered | 3 | VC_1446_0 | 3 | 1 | 0 | 0 | 0 | 0 | 0 | 1 | Ref |
| 12538 | MW202744 | Unassigned | Unassigned | Unassigned | 117_0 | Clustered | 9 | VC_117_0 | 9 | 0.9977 | 0.98988637 | 0.9876 | 0 | 0 | 0 | 1 | Ref |
| 12539 | MW202748 | Unassigned | Unassigned | Unassigned | 1442_0 | Clustered | 2 | VC_1442_0 | 2 | 0.9685 | 0.86822376 | 0.8408 | 0 | 0 | 0 | 1 | Ref |
| 12540 | MW202753 | Unassigned | Unassigned | Unassigned | 1452_0 | Clustered | 2 | VC_1452_0 | 2 | 1 | 1 | 1 | 0 | 0 | 0 | 1 | Ref |
| 12541 | MW202760 | Unassigned | Unassigned | Unassigned | 1077_0 | Clustered | 48 | VC_1077_0 | 48 | 0.8525 | 1 | 0.8525 | 0 | 0 | 0 | 1 | Ref |
| 12542 | MW202766 | Unassigned | Unassigned | Unassigned | 1444_0 | Clustered | 3 | VC_1444_0 | 3 | 1 | 0 | 0 | 0 | 0 | 0 | 1 | Ref |
| 12543 | MW202777 | Unassigned | Unassigned | Unassigned | 466_0 | Clustered | 3 | VC_466_0 | 3 | 0.3264 | 0.99747305 | 0.3255 | 0 | 0 | 0 | 1 | Ref |
| 12544 | MW202785 | Unassigned | Unassigned | Unassigned | 760_0 | Clustered | 3 | VC_760_0 | 3 | 0.7281 | 0.9859071 | 0.7179 | 0 | 0 | 0 | 1 | Ref |
| 12545 | MW202789 | Unassigned | Unassigned | Unassigned | 1443_0 | Clustered | 2 | VC_1443_0 | 2 | 0.8611 | 0.76024994 | 0.6547 | 0 | 0 | 0 | 1 | Ref |
| 12546 | MW202796 | Unassigned | Unassigned | Unassigned | 760_0 | Clustered | 3 | VC_760_0 | 3 | 0.7281 | 0.9859071 | 0.7179 | 0 | 0 | 0 | 1 | Ref |
| 12547 | MW202812 | Unassigned | Unassigned | Unassigned | 1448_0 | Clustered | 5 | VC_1448_0 | 5 | 1 | 1 | 1 | 0 | 0 | 0 | 1 | Ref |
| 12548 | MW202819 | Unassigned | Unassigned | Unassigned | 1077_0 | Clustered | 48 | VC_1077_0 | 48 | 0.8525 | 1 | 0.8525 | 0 | 0 | 0 | 1 | Ref |
| 12549 | MW202824 | Unassigned | Unassigned | Unassigned | 1243_0 | Clustered | 52 | VC_1243_0 | 52 | 0.9126 | 0.92708311 | 0.8461 | 0 | 0 | 0 | 1 | Ref |
| 12550 | MW202825 | Unassigned | Unassigned | Unassigned | 1452_0 | Clustered | 2 | VC_1452_0 | 2 | 1 | 1 | 1 | 0 | 0 | 0 | 1 | Ref |
| 12551 | MW202826 | Unassigned | Unassigned | Unassigned | 1448_0 | Clustered | 5 | VC_1448_0 | 5 | 1 | 1 | 1 | 0 | 0 | 0 | 1 | Ref |
| 12552 | MW202842 | Unassigned | Unassigned | Unassigned | 1448_0 | Clustered | 5 | VC_1448_0 | 5 | 1 | 1 | 1 | 0 | 0 | 0 | 1 | Ref |
| 12553 | MW202843 | Unassigned | Unassigned | Unassigned | 1450_0 | Clustered | 3 | VC_1450_0 | 3 | 1 | 1 | 1 | 0 | 0 | 0 | 1 | Ref |
| 12554 | MW202847 | Unassigned | Unassigned | Unassigned | 995_0 | Clustered | 69 | VC_995_0 | 69 | 0.8547 | 1 | 0.8547 | 0 | 0 | 0 | 1 | Ref |
| 12555 | MW202852 | Unassigned | Unassigned | Unassigned | 1450_0 | Clustered | 3 | VC_1450_0 | 3 | 1 | 1 | 1 | 0 | 0 | 0 | 1 | Ref |
| 12556 | MW202853 | Unassigned | Unassigned | Unassigned | 995_0 | Clustered | 69 | VC_995_0 | 69 | 0.8547 | 1 | 0.8547 | 0 | 0 | 0 | 1 | Ref |
| 12557 | MW202864 | Unassigned | Unassigned | Unassigned | 1451_0 | Clustered | 2 | VC_1451_0 | 2 | 1 | 1 | 1 | 0 | 0 | 0 | 1 | Ref |
| 12558 | MW202867 | Unassigned | Unassigned | Unassigned | 163_0 | Clustered | 2 | VC_163_0 | 2 | 0.2865 | 0.94106069 | 0.2696 | 0 | 0 | 0 | 1 | Ref |
| 12559 | MW202879 | Unassigned | Unassigned | Unassigned | 609_0 | Clustered | 4 | VC_609_0 | 4 | 0.489 | 0.99992419 | 0.489 | 0 | 0 | 0 | 1 | Ref |
| 12560 | MW202883 | Unassigned | Unassigned | Unassigned | 995_0 | Clustered | 69 | VC_995_0 | 69 | 0.8547 | 1 | 0.8547 | 0 | 0 | 0 | 1 | Ref |
| 12561 | MW202889 | Unassigned | Unassigned | Unassigned | 198_0 | Clustered | 28 | VC_198_0 | 28 | 0.2429 | 0.99971175 | 0.2429 | 0 | 0 | 0 | 1 | Ref |
| 12562 | MW202895 | Unassigned | Unassigned | Unassigned | 326_0 | Clustered/Singleton | 1 | VC_326_0 | 1 | 0 | 1 | 0 | 0 | 0 | 0 | 1 | Ref |
| 12563 | MW205203 | Unassigned | Unassigned | Unassigned | 476_0 | Clustered | 19 | VC_476_0 | 19 | 0.7814 | 1 | 0.7814 | 0 | 0 | 0 | 1 | Ref |
| 12564 | MW206381 | Unassigned | Unassigned | Unassigned | 191_25 | Clustered/Singleton | 1 | VC_191_25 | 1 | 0 | 1 | 0 | 0 | 0 | 0 | 1 | Ref |
| 12565 | MW217515 | Unassigned | Unassigned | Unassigned | 530_0 | Clustered | 9 | VC_530_0 | 9 | 0.8027 | 1 | 0.8027 | 0 | 0 | 0 | 1 | Ref |
| 12566 | MW218148 | Unassigned | Unassigned | Unassigned | 449_0 | Clustered | 7 | VC_449_0 | 7 | 0.6015 | 1 | 0.6015 | 0 | 0 | 0 | 1 | Ref |
| 12567 | MW233709 | Unassigned | Unassigned | Unassigned | 290_0 | Clustered | 10 | VC_290_0 | 10 | 0.0981 | 1 | 0.0981 | 0 | 0 | 0 | 1 | Ref |
| 12568 | MW238767 | Unassigned | Unassigned | Unassigned | 12_0 | Clustered | 21 | VC_12_0 | 21 | 0.2274 | 1 | 0.2274 | 0 | 0 | 0 | 0.9571 | Ref |
| 12569 | MW239157 | Unassigned | Unassigned | Unassigned | 491_0 | Clustered | 36 | VC_491_0 | 36 | 0.5336 | 1 | 0.5336 | 0 | 0 | 0 | 0.9714 | Ref |
| 12570 | MW247144 | Unassigned | Unassigned | Unassigned | 99_0 | Clustered | 32 | VC_99_0 | 32 | 0.8828 | 1 | 0.8828 | 0 | 0 | 0 | 1 | Ref |
| 12571 | MW247145 | Unassigned | Unassigned | Unassigned | 99_0 | Clustered | 32 | VC_99_0 | 32 | 0.8828 | 1 | 0.8828 | 0 | 0 | 0 | 1 | Ref |
| 12572 | MW247147 | Unassigned | Unassigned | Unassigned | 99_0 | Clustered | 32 | VC_99_0 | 32 | 0.8828 | 1 | 0.8828 | 0 | 0 | 0 | 1 | Ref |
| 12573 | MW248466 | Unassigned | Unassigned | Unassigned | 449_0 | Clustered | 7 | VC_449_0 | 7 | 0.6015 | 1 | 0.6015 | 0 | 0 | 0 | 1 | Ref |
| 12574 | MW250275 | Unassigned | Unassigned | Unassigned | 156_0 | Clustered | 12 | VC_156_0 | 12 | 0.9062 | 1 | 0.9062 | 0 | 0 | 0 | 1 | Ref |
| 12575 | MW250276 | Unassigned | Unassigned | Unassigned | 878_0 | Clustered | 32 | VC_878_0 | 32 | 0.9965 | 1 | 0.9965 | 0 | 0 | 0 | 1 | Ref |
| 12576 | MW250641 | Unassigned | Unassigned | Unassigned | 837_0 | Clustered | 29 | VC_837_0 | 29 | 0.9955 | 1 | 0.9955 | 0 | 0 | 0 | 1 | Ref |
| 12577 | MW250785 | Unassigned | Unassigned | Unassigned | 669_0 | Clustered | 21 | VC_669_0 | 21 | 0.3623 | 1 | 0.3623 | 0 | 0 | 0 | 1 | Ref |
| 12578 | MW250786 | Unassigned | Unassigned | Unassigned | 849_9 | Clustered/Singleton | 1 | VC_849_9 | 1 | 0 | 1 | 0 | 0 | 0 | 0 | 1 | Ref |
| 12579 | MW250787 | Unassigned | Unassigned | Unassigned | 290_0 | Clustered | 10 | VC_290_0 | 10 | 0.0981 | 1 | 0.0981 | 0 | 0 | 0 | 1 | Ref |
| 12580 | MW258709 | Unassigned | Unassigned | Unassigned | 451_0 | Clustered | 80 | VC_451_0 | 80 | 0.9991 | 1 | 0.9991 | 0 | 0 | 0 | 1 | Ref |
| 12581 | MW258710 | Unassigned | Unassigned | Unassigned | 705_0 | Clustered | 11 | VC_705_0 | 11 | 0.8861 | 1 | 0.8861 | 0 | 0 | 0 | 1 | Ref |
| 12582 | MW258711 | Unassigned | Unassigned | Unassigned | 705_0 | Clustered | 11 | VC_705_0 | 11 | 0.8861 | 1 | 0.8861 | 0 | 0 | 0 | 1 | Ref |
| 12583 | MW269554 | Unassigned | Unassigned | Unassigned | 598_0 | Clustered | 4 | VC_598_0 | 4 | 0.0597 | 0.99355358 | 0.0593 | 0 | 0 | 0 | 1 | Ref |
| 12584 | MW269952 | Unassigned | Unassigned | Unassigned | 290_1 | Clustered | 8 | VC_290_1 | 8 | 0.0757 | 1 | 0.0757 | 0 | 0 | 0 | 0.8571 | Ref |
| 12585 | MW272540 | Unassigned | Unassigned | Unassigned | 443_0 | Clustered | 2 | VC_443_0 | 2 | 0.029 | 0.87409603 | 0.0253 | 0 | 0 | 0 | 1 | Ref |
| 12586 | MW273920 | Unassigned | Unassigned | Unassigned | 486_0 | Clustered | 18 | VC_486_0 | 18 | 0.887 | 1 | 0.887 | 0 | 0 | 0 | 1 | Ref |
| 12587 | MW273922 | Unassigned | Unassigned | Unassigned | 486_0 | Clustered | 18 | VC_486_0 | 18 | 0.887 | 1 | 0.887 | 0 | 0 | 0 | 1 | Ref |
| 12588 | MW273923 | Unassigned | Unassigned | Unassigned | 486_0 | Clustered | 18 | VC_486_0 | 18 | 0.887 | 1 | 0.887 | 0 | 0 | 0 | 1 | Ref |
| 12589 | MW273924 | Unassigned | Unassigned | Unassigned | 486_0 | Clustered | 18 | VC_486_0 | 18 | 0.887 | 1 | 0.887 | 0 | 0 | 0 | 1 | Ref |
| 12590 | MW273925 | Unassigned | Unassigned | Unassigned | 486_0 | Clustered | 18 | VC_486_0 | 18 | 0.887 | 1 | 0.887 | 0 | 0 | 0 | 1 | Ref |
| 12591 | MW273926 | Unassigned | Unassigned | Unassigned | 486_0 | Clustered | 18 | VC_486_0 | 18 | 0.887 | 1 | 0.887 | 0 | 0 | 0 | 1 | Ref |
| 12592 | MW281503 | Unassigned | Unassigned | Unassigned | 883_1 | Clustered | 12 | VC_883_1 | 12 | 0.2441 | 1 | 0.2441 | 0 | 0 | 0 | 1 | Ref |
| 12593 | MW286156 | Unassigned | Unassigned | Unassigned | 455_0 | Clustered | 18 | VC_455_0 | 18 | 0.4598 | 1 | 0.4598 | 0 | 0 | 0 | 1 | Ref |
| 12594 | MW286157 | Unassigned | Unassigned | Unassigned | 283_7 | Clustered/Singleton | 1 | VC_283_7 | 1 | 0 | 1 | 0 | 0 | 0 | 0 | 1 | Ref |
| 12595 | MW286254 | Unassigned | Unassigned | Unassigned | 96_0 | Clustered | 30 | VC_96_0 | 30 | 0.718 | 1 | 0.718 | 0 | 0 | 0 | 1 | Ref |
| 12596 | MW291012 | Unassigned | Unassigned | Unassigned | 558_65 | Clustered | 2 | VC_558_65 | 2 | 0.0032 | 0.61174182 | 0.0019 | 0 | 0 | 0 | 1 | Ref |
| 12597 | MW291013 | Unassigned | Unassigned | Unassigned | 1175_0 | Clustered | 22 | VC_1175_0 | 22 | 0.4562 | 1 | 0.4562 | 0 | 0 | 0 | 1 | Ref |
| 12598 | MW291014 | Unassigned | Unassigned | Unassigned | 1071_0 | Clustered | 20 | VC_1071_0 | 20 | 0.3268 | 1 | 0.3268 | 0 | 0 | 0 | 0.9842 | Ref |
| 12599 | MW291015 | Unassigned | Unassigned | Unassigned | 954_0 | Clustered | 13 | VC_954_0 | 13 | 0.863 | 1 | 0.863 | 0 | 0 | 0 | 1 | Ref |
| 12600 | MW291016 | Unassigned | Unassigned | Unassigned | 1176_0 | Clustered | 41 | VC_1176_0 | 41 | 0.9558 | 1 | 0.9558 | 0 | 0 | 0 | 0.9976 | Ref |
| 12601 | MW291018 | Unassigned | Unassigned | Unassigned | 1360_0 | Clustered | 31 | VC_1360_0 | 31 | 0.8837 | 1 | 0.8837 | 0 | 0 | 0 | 1 | Ref |
| 12602 | MW291019 | Unassigned | Unassigned | Unassigned | 517_1 | Clustered | 108 | VC_517_1 | 108 | 0.8483 | 1 | 0.8483 | 0 | 0 | 0 | 0.9955 | Ref |
| 12603 | MW291020 | Unassigned | Unassigned | Unassigned | 1344_0 | Clustered | 78 | VC_1344_0 | 78 | 0.9732 | 1 | 0.9732 | 0 | 0 | 0 | 1 | Ref |
| 12604 | MW291021 | Unassigned | Unassigned | Unassigned | 1158_0 | Clustered | 3 | VC_1158_0 | 3 | 0.3179 | 0.99335558 | 0.3158 | 0 | 0 | 0 | 1 | Ref |
| 12605 | MW291022 | Unassigned | Unassigned | Unassigned | 799_1 | Clustered | 27 | VC_799_1 | 27 | 0.8611 | 1 | 0.8611 | 0 | 0 | 0 | 1 | Ref |
| 12606 | MW291023 | Unassigned | Unassigned | Unassigned | 551_0 | Clustered | 16 | VC_551_0 | 16 | 0.0915 | 1 | 0.0915 | 0 | 0 | 0 | 0.6667 | Ref |
| 12607 | MW291024 | Unassigned | Unassigned | Unassigned | 889_0 | Clustered | 16 | VC_889_0 | 16 | 0.2969 | 1 | 0.2969 | 0 | 0 | 0 | 1 | Ref |
| 12608 | MW291025 | Unassigned | Unassigned | Unassigned | 1343_0 | Clustered | 21 | VC_1343_0 | 21 | 0.9843 | 1 | 0.9843 | 0 | 0 | 0 | 1 | Ref |
| 12609 | MW291026 | Unassigned | Unassigned | Unassigned | 1240_0 | Clustered | 82 | VC_1240_0 | 82 | 0.7945 | 1 | 0.7945 | 0 | 0 | 0 | 1 | Ref |
| 12610 | MW291027 | Unassigned | Unassigned | Unassigned | 956_0 | Clustered | 39 | VC_956_0 | 39 | 1 | 1 | 1 | 0 | 0 | 0 | 1 | Ref |
| 12611 | MW291028 | Unassigned | Unassigned | Unassigned | 761_0 | Clustered | 67 | VC_761_0 | 67 | 0.8947 | 1 | 0.8947 | 0 | 0 | 0 | 1 | Ref |
| 12612 | MW296032 | Unassigned | Unassigned | Unassigned | 812_0 | Clustered | 3 | VC_812_0 | 3 | 0.1174 | 0.99842379 | 0.1172 | 0 | 0 | 0 | 1 | Ref |
| 12613 | MW296865 | Unassigned | Unassigned | Unassigned | 329_18 | Clustered | 7 | VC_329_18 | 7 | 0.0328 | 1 | 0.0328 | 0 | 0 | 0 | 0.8095 | Ref |
| 12614 | MW296877 | Unassigned | Unassigned | Unassigned | 385_0 | Clustered | 51 | VC_385_0 | 51 | 0.8754 | 1 | 0.8754 | 0 | 0 | 0 | 1 | Ref |
| 12615 | MW302330 | Unassigned | Unassigned | Unassigned | 654_0 | Clustered | 4 | VC_654_0 | 4 | 0.2897 | 0.99998263 | 0.2897 | 0 | 0 | 0 | 1 | Ref |
| 12616 | MW311372 | Unassigned | Unassigned | Unassigned | 329_11 | Clustered | 15 | VC_329_11 | 15 | 0.0791 | 1 | 0.0791 | 0 | 0 | 0 | 0.9333 | Ref |
| 12617 | MW314138 | Unassigned | Unassigned | Unassigned | 830_0 | Clustered | 6 | VC_830_0 | 6 | 0.6536 | 1 | 0.6536 | 0 | 0 | 0 | 1 | Ref |
| 12618 | MW314845 | Unassigned | Unassigned | Unassigned | 556_0 | Clustered | 21 | VC_556_0 | 21 | 0.9912 | 1 | 0.9912 | 0 | 0 | 0 | 1 | Ref |
| 12619 | MW314846 | Unassigned | Unassigned | Unassigned | 546_1 | Clustered | 2 | VC_546_1 | 2 | 0.0136 | 0.68405151 | 0.0093 | 0 | 0 | 0 | 1 | Ref |
| 12620 | MW314847 | Unassigned | Unassigned | Unassigned | 1095_0 | Clustered | 16 | VC_1095_0 | 16 | 0.458 | 1 | 0.458 | 0 | 0 | 0 | 1 | Ref |
| 12621 | MW314848 | Unassigned | Unassigned | Unassigned | 1160_0 | Clustered | 73 | VC_1160_0 | 73 | 0.9835 | 1 | 0.9835 | 0 | 0 | 0 | 1 | Ref |
| 12622 | MW314851 | Unassigned | Unassigned | Unassigned | 1092_0 | Clustered | 12 | VC_1092_0 | 12 | 0.8894 | 1 | 0.8894 | 0 | 0 | 0 | 1 | Ref |
| 12623 | MW314852 | Unassigned | Unassigned | Unassigned | 555_0 | Clustered | 13 | VC_555_0 | 13 | 0.7516 | 1 | 0.7516 | 0 | 0 | 0 | 1 | Ref |
| 12624 | MW314853 | Unassigned | Unassigned | Unassigned | 1092_0 | Clustered | 12 | VC_1092_0 | 12 | 0.8894 | 1 | 0.8894 | 0 | 0 | 0 | 1 | Ref |
| 12625 | MW314854 | Unassigned | Unassigned | Unassigned | 516_0 | Clustered | 3 | VC_516_0 | 3 | 0.4664 | 0.99856798 | 0.4658 | 0 | 0 | 0 | 1 | Ref |
| 12626 | MW314855 | Unassigned | Unassigned | Unassigned | 550_0 | Clustered | 12 | VC_550_0 | 12 | 0.802 | 1 | 0.802 | 0 | 0 | 0 | 1 | Ref |
| 12627 | MW314856 | Unassigned | Unassigned | Unassigned | 550_0 | Clustered | 12 | VC_550_0 | 12 | 0.802 | 1 | 0.802 | 0 | 0 | 0 | 1 | Ref |
| 12628 | MW314857 | Unassigned | Unassigned | Unassigned | 1092_0 | Clustered | 12 | VC_1092_0 | 12 | 0.8894 | 1 | 0.8894 | 0 | 0 | 0 | 1 | Ref |
| 12629 | MW314858 | Unassigned | Unassigned | Unassigned | 550_0 | Clustered | 12 | VC_550_0 | 12 | 0.802 | 1 | 0.802 | 0 | 0 | 0 | 1 | Ref |
| 12630 | MW314859 | Unassigned | Unassigned | Unassigned | 240_0 | Clustered | 4 | VC_240_0 | 4 | 0.7811 | 0.99998265 | 0.7811 | 0 | 0 | 0 | 1 | Ref |
| 12631 | MW316731 | Unassigned | Unassigned | Unassigned | 702_0 | Clustered | 5 | VC_702_0 | 5 | 0.7794 | 0.99999988 | 0.7794 | 0 | 0 | 0 | 1 | Ref |
| 12632 | MW316732 | Unassigned | Unassigned | Unassigned | 702_0 | Clustered | 5 | VC_702_0 | 5 | 0.7794 | 0.99999988 | 0.7794 | 0 | 0 | 0 | 1 | Ref |
| 12633 | MW316733 | Unassigned | Unassigned | Unassigned | 702_0 | Clustered | 5 | VC_702_0 | 5 | 0.7794 | 0.99999988 | 0.7794 | 0 | 0 | 0 | 1 | Ref |
| 12634 | MW321605 | Unassigned | Unassigned | Unassigned | 329_11 | Clustered | 15 | VC_329_11 | 15 | 0.0791 | 1 | 0.0791 | 0 | 0 | 0 | 0.9333 | Ref |

| | | | | | | | | | | | | | | | | | |
|---|---|---|---|---|---|---|---|---|---|---|---|---|---|---|---|---|---|
| 12635 | MW321655 | Unassigned | Unassigned | Unassigned | 1344_0 | Clustered | 78 | VC_1344_0 | 78 | 0.9732 | 1 | 0.9732 | 0 | 0 | 0 | 1 | Ref |
| 12636 | MW329986 | Unassigned | Unassigned | Unassigned | 472_0 | Clustered | 47 | VC_472_0 | 47 | 0.9025 | 1 | 0.9025 | 0 | 0 | 0 | 1 | Ref |
| 12637 | MW331437 | Unassigned | Unassigned | Unassigned | 638_0 | Clustered | 45 | VC_638_0 | 45 | 0.7741 | 1 | 0.7741 | 0 | 0 | 0 | 1 | Ref |
| 12638 | MW331544 | Unassigned | Unassigned | Unassigned | 837_0 | Clustered | 29 | VC_837_0 | 29 | 0.9955 | 1 | 0.9955 | 0 | 0 | 0 | 1 | Ref |
| 12639 | MW341595 | Unassigned | Unassigned | Unassigned | 283_144 | Clustered/Singleton | 1 | VC_283_144 | 1 | 0 | 1 | 0 | 0 | 0 | 0 | 1 | Ref |
| 12640 | MW343794 | Unassigned | Unassigned | Unassigned | 812_2 | Clustered | 2 | VC_812_2 | 2 | 0.0668 | 0.95518138 | 0.0639 | 0 | 0 | 0 | 1 | Ref |
| 12641 | MW344054 | Unassigned | Unassigned | Unassigned | 971_1 | Clustered | 24 | VC_971_1 | 24 | 0.7393 | 1 | 0.7393 | 0 | 0 | 0 | 0.8913 | Ref |
| 12642 | MW344055 | Unassigned | Unassigned | Unassigned | 971_1 | Clustered | 24 | VC_971_1 | 24 | 0.7393 | 1 | 0.7393 | 0 | 0 | 0 | 0.8913 | Ref |
| 12643 | MW344056 | Unassigned | Unassigned | Unassigned | 971_1 | Clustered | 24 | VC_971_1 | 24 | 0.7393 | 1 | 0.7393 | 0 | 0 | 0 | 0.8913 | Ref |
| 12644 | MW344764 | Unassigned | Unassigned | Unassigned | 834_0 | Clustered | 7 | VC_834_0 | 7 | 0.9845 | 0.94276893 | 0.9282 | 0 | 0 | 0 | 1 | Ref |
| 12645 | MW344765 | Unassigned | Unassigned | Unassigned | 351_0 | Clustered | 3 | VC_351_0 | 3 | 0.8899 | 0.98155579 | 0.8735 | 0 | 0 | 0 | 1 | Ref |
| 12646 | MW344766 | Unassigned | Unassigned | Unassigned | 834_0 | Clustered | 7 | VC_834_0 | 7 | 0.9845 | 0.94276893 | 0.9282 | 0 | 0 | 0 | 1 | Ref |
| 12647 | MW344767 | Unassigned | Unassigned | Unassigned | 961_0 | Clustered | 3 | VC_961_0 | 3 | 0.675 | 0.99054823 | 0.6686 | 0 | 0 | 0 | 1 | Ref |
| 12648 | MW345241 | Unassigned | Unassigned | Unassigned | 699_0 | Clustered | 71 | VC_699_0 | 71 | 0.9352 | 1 | 0.9352 | 0 | 0 | 0 | 1 | Ref |
| 12649 | MW345254 | Unassigned | Unassigned | Unassigned | 451_0 | Clustered | 80 | VC_451_0 | 80 | 0.9991 | 1 | 0.9991 | 0 | 0 | 0 | 1 | Ref |
| 12650 | MW348917 | Unassigned | Unassigned | Unassigned | 818_0 | Clustered | 20 | VC_818_0 | 20 | 0.9601 | 1 | 0.9601 | 0 | 0 | 0 | 1 | Ref |
| 12651 | MW349128 | Unassigned | Unassigned | Unassigned | 449_0 | Clustered | 7 | VC_449_0 | 7 | 0.6015 | 1 | 0.6015 | 0 | 0 | 0 | 1 | Ref |
| 12652 | MW349129 | Unassigned | Unassigned | Unassigned | 449_0 | Clustered | 7 | VC_449_0 | 7 | 0.6015 | 1 | 0.6015 | 0 | 0 | 0 | 1 | Ref |
| 12653 | MW349133 | Unassigned | Unassigned | Unassigned | 700_0 | Clustered | 10 | VC_700_0 | 10 | 0.8076 | 1 | 0.8076 | 0 | 0 | 0 | 1 | Ref |
| 12654 | MW349138 | Unassigned | Unassigned | Unassigned | 792_0 | Clustered | 35 | VC_792_0 | 35 | 0.9828 | 1 | 0.9828 | 0 | 0 | 0 | 1 | Ref |
| 12655 | MW353166 | Unassigned | Unassigned | Unassigned | 1453_0 | Clustered | 7 | VC_1453_0 | 7 | 0.912 | 0.99951341 | 0.9115 | 0 | 0 | 0 | 1 | Ref |
| 12656 | MW353167 | Unassigned | Unassigned | Unassigned | 1453_0 | Clustered | 7 | VC_1453_0 | 7 | 0.912 | 0.99951341 | 0.9115 | 0 | 0 | 0 | 1 | Ref |
| 12657 | MW353168 | Unassigned | Unassigned | Unassigned | 1453_0 | Clustered | 7 | VC_1453_0 | 7 | 0.912 | 0.99951341 | 0.9115 | 0 | 0 | 0 | 1 | Ref |
| 12658 | MW353169 | Unassigned | Unassigned | Unassigned | 1453_0 | Clustered | 7 | VC_1453_0 | 7 | 0.912 | 0.99951341 | 0.9115 | 0 | 0 | 0 | 1 | Ref |
| 12659 | MW353170 | Unassigned | Unassigned | Unassigned | 1453_0 | Clustered | 7 | VC_1453_0 | 7 | 0.912 | 0.99951341 | 0.9115 | 0 | 0 | 0 | 1 | Ref |
| 12660 | MW353171 | Unassigned | Unassigned | Unassigned | 1453_0 | Clustered | 7 | VC_1453_0 | 7 | 0.912 | 0.99951341 | 0.9115 | 0 | 0 | 0 | 1 | Ref |
| 12661 | MW353172 | Unassigned | Unassigned | Unassigned | 1453_0 | Clustered | 7 | VC_1453_0 | 7 | 0.912 | 0.99951341 | 0.9115 | 0 | 0 | 0 | 1 | Ref |
| 12662 | MW353175 | Unassigned | Unassigned | Unassigned | 505_0 | Clustered | 3 | VC_505_0 | 3 | 0.8802 | 0.96819872 | 0.8523 | 0 | 0 | 0 | 1 | Ref |
| 12663 | MW353176 | Unassigned | Unassigned | Unassigned | 505_0 | Clustered | 3 | VC_505_0 | 3 | 0.8802 | 0.96819872 | 0.8523 | 0 | 0 | 0 | 1 | Ref |
| 12664 | MW353177 | Unassigned | Unassigned | Unassigned | 505_0 | Clustered | 3 | VC_505_0 | 3 | 0.8802 | 0.96819872 | 0.8523 | 0 | 0 | 0 | 1 | Ref |
| 12665 | MW353181 | Unassigned | Unassigned | Unassigned | 684_0 | Clustered | 5 | VC_684_0 | 5 | 0.3915 | 0.99999995 | 0.3915 | 0 | 0 | 0 | 1 | Ref |
| 12666 | MW354667 | Unassigned | Unassigned | Unassigned | 85_3 | Clustered | 19 | VC_85_3 | 19 | 0.3671 | 1 | 0.3671 | 0 | 0 | 0 | 0.807 | Ref |
| 12667 | MW354668 | Unassigned | Unassigned | Unassigned | 817_0 | Clustered | 14 | VC_817_0 | 14 | 0.6342 | 1 | 0.6342 | 0 | 0 | 0 | 1 | Ref |
| 12668 | MW354669 | Unassigned | Unassigned | Unassigned | 1358_0 | Clustered | 5 | VC_1358_0 | 5 | 1 | 1 | 1 | 0 | 0 | 0 | 1 | Ref |
| 12669 | MW354670 | Unassigned | Unassigned | Unassigned | 817_0 | Clustered | 14 | VC_817_0 | 14 | 0.6342 | 1 | 0.6342 | 0 | 0 | 0 | 1 | Ref |
| 12670 | MW354676 | Unassigned | Unassigned | Unassigned | 1358_0 | Clustered | 5 | VC_1358_0 | 5 | 1 | 1 | 1 | 0 | 0 | 0 | 1 | Ref |
| 12671 | MW354677 | Unassigned | Unassigned | Unassigned | 1358_0 | Clustered | 5 | VC_1358_0 | 5 | 1 | 1 | 1 | 0 | 0 | 0 | 1 | Ref |
| 12672 | MW354678 | Unassigned | Unassigned | Unassigned | 1358_0 | Clustered | 5 | VC_1358_0 | 5 | 1 | 1 | 1 | 0 | 0 | 0 | 1 | Ref |
| 12673 | MW355449 | Unassigned | Unassigned | Unassigned | 592_0 | Clustered | 50 | VC_592_0 | 50 | 0.7314 | 1 | 0.7314 | 0 | 0 | 0 | 1 | Ref |
| 12674 | MW355450 | Unassigned | Unassigned | Unassigned | 133_31 | Clustered | 6 | VC_133_31 | 6 | 0.0263 | 0.99999694 | 0.0263 | 0 | 0 | 0 | 1 | Ref |
| 12675 | MW355451 | Unassigned | Unassigned | Unassigned | 637_1 | Clustered | 27 | VC_637_1 | 27 | 0.2578 | 1 | 0.2578 | 0 | 0 | 0 | 0.9858 | Ref |
| 12676 | MW355452 | Unassigned | Unassigned | Unassigned | 637_1 | Clustered | 27 | VC_637_1 | 27 | 0.2578 | 1 | 0.2578 | 0 | 0 | 0 | 0.9858 | Ref |
| 12677 | MW355453 | Unassigned | Unassigned | Unassigned | 637_1 | Clustered | 27 | VC_637_1 | 27 | 0.2578 | 1 | 0.2578 | 0 | 0 | 0 | 0.9858 | Ref |
| 12678 | MW355454 | Unassigned | Unassigned | Unassigned | 637_1 | Clustered | 27 | VC_637_1 | 27 | 0.2578 | 1 | 0.2578 | 0 | 0 | 0 | 0.9858 | Ref |
| 12679 | MW355455 | Unassigned | Unassigned | Unassigned | 637_1 | Clustered | 27 | VC_637_1 | 27 | 0.2578 | 1 | 0.2578 | 0 | 0 | 0 | 0.9858 | Ref |
| 12680 | MW355456 | Unassigned | Unassigned | Unassigned | 637_1 | Clustered | 27 | VC_637_1 | 27 | 0.2578 | 1 | 0.2578 | 0 | 0 | 0 | 0.9858 | Ref |
| 12681 | MW355457 | Unassigned | Unassigned | Unassigned | 637_1 | Clustered | 27 | VC_637_1 | 27 | 0.2578 | 1 | 0.2578 | 0 | 0 | 0 | 0.9858 | Ref |
| 12682 | MW355458 | Unassigned | Unassigned | Unassigned | 637_1 | Clustered | 27 | VC_637_1 | 27 | 0.2578 | 1 | 0.2578 | 0 | 0 | 0 | 0.9858 | Ref |
| 12683 | MW355459 | Unassigned | Unassigned | Unassigned | 637_1 | Clustered | 27 | VC_637_1 | 27 | 0.2578 | 1 | 0.2578 | 0 | 0 | 0 | 0.9858 | Ref |
| 12684 | MW355460 | Unassigned | Unassigned | Unassigned | 592_0 | Clustered | 50 | VC_592_0 | 50 | 0.7314 | 1 | 0.7314 | 0 | 0 | 0 | 1 | Ref |
| 12685 | MW355461 | Unassigned | Unassigned | Unassigned | 637_1 | Clustered | 27 | VC_637_1 | 27 | 0.2578 | 1 | 0.2578 | 0 | 0 | 0 | 0.9858 | Ref |
| 12686 | MW355462 | Unassigned | Unassigned | Unassigned | 637_1 | Clustered | 27 | VC_637_1 | 27 | 0.2578 | 1 | 0.2578 | 0 | 0 | 0 | 0.9858 | Ref |
| 12687 | MW355463 | Unassigned | Unassigned | Unassigned | 637_0 | Clustered | 115 | VC_637_0 | 115 | 0.731 | 1 | 0.731 | 0 | 0 | 0 | 0.8764 | Ref |
| 12688 | MW355464 | Unassigned | Unassigned | Unassigned | 637_0 | Clustered | 115 | VC_637_0 | 115 | 0.731 | 1 | 0.731 | 0 | 0 | 0 | 0.8764 | Ref |
| 12689 | MW355465 | Unassigned | Unassigned | Unassigned | 637_0 | Clustered | 115 | VC_637_0 | 115 | 0.731 | 1 | 0.731 | 0 | 0 | 0 | 0.8764 | Ref |
| 12690 | MW355466 | Unassigned | Unassigned | Unassigned | 637_1 | Clustered | 27 | VC_637_1 | 27 | 0.2578 | 1 | 0.2578 | 0 | 0 | 0 | 0.9858 | Ref |
| 12691 | MW355467 | Unassigned | Unassigned | Unassigned | 592_0 | Clustered | 50 | VC_592_0 | 50 | 0.7314 | 1 | 0.7314 | 0 | 0 | 0 | 1 | Ref |
| 12692 | MW355468 | Unassigned | Unassigned | Unassigned | 637_1 | Clustered | 27 | VC_637_1 | 27 | 0.2578 | 1 | 0.2578 | 0 | 0 | 0 | 0.9858 | Ref |
| 12693 | MW355469 | Unassigned | Unassigned | Unassigned | 637_1 | Clustered | 27 | VC_637_1 | 27 | 0.2578 | 1 | 0.2578 | 0 | 0 | 0 | 0.9858 | Ref |
| 12694 | MW355470 | Unassigned | Unassigned | Unassigned | 592_0 | Clustered | 50 | VC_592_0 | 50 | 0.7314 | 1 | 0.7314 | 0 | 0 | 0 | 1 | Ref |
| 12695 | MW355471 | Unassigned | Unassigned | Unassigned | 637_1 | Clustered | 27 | VC_637_1 | 27 | 0.2578 | 1 | 0.2578 | 0 | 0 | 0 | 0.9858 | Ref |
| 12696 | MW355472 | Unassigned | Unassigned | Unassigned | 637_1 | Clustered | 27 | VC_637_1 | 27 | 0.2578 | 1 | 0.2578 | 0 | 0 | 0 | 0.9858 | Ref |
| 12697 | MW355473 | Unassigned | Unassigned | Unassigned | 637_1 | Clustered | 27 | VC_637_1 | 27 | 0.2578 | 1 | 0.2578 | 0 | 0 | 0 | 0.9858 | Ref |
| 12698 | MW355474 | Unassigned | Unassigned | Unassigned | 592_0 | Clustered | 50 | VC_592_0 | 50 | 0.7314 | 1 | 0.7314 | 0 | 0 | 0 | 1 | Ref |
| 12699 | MW355475 | Unassigned | Unassigned | Unassigned | 133_31 | Clustered | 6 | VC_133_31 | 6 | 0.0263 | 0.99999694 | 0.0263 | 0 | 0 | 0 | 1 | Ref |
| 12700 | MW355476 | Unassigned | Unassigned | Unassigned | 592_0 | Clustered | 50 | VC_592_0 | 50 | 0.7314 | 1 | 0.7314 | 0 | 0 | 0 | 1 | Ref |
| 12701 | MW355477 | Unassigned | Unassigned | Unassigned | 133_31 | Clustered | 6 | VC_133_31 | 6 | 0.0263 | 0.99999694 | 0.0263 | 0 | 0 | 0 | 1 | Ref |
| 12702 | MW355478 | Unassigned | Unassigned | Unassigned | 133_28 | Clustered | 3 | VC_133_28 | 3 | 0.0105 | 0.97661637 | 0.0103 | 0 | 0 | 0 | 0.6667 | Ref |
| 12703 | MW355479 | Unassigned | Unassigned | Unassigned | 592_0 | Clustered | 50 | VC_592_0 | 50 | 0.7314 | 1 | 0.7314 | 0 | 0 | 0 | 1 | Ref |
| 12704 | MW355480 | Unassigned | Unassigned | Unassigned | 28_1 | Clustered | 53 | VC_28_1 | 53 | 0.7164 | 1 | 0.7164 | 0 | 0 | 0 | 0.9325 | Ref |
| 12705 | MW357609 | Unassigned | Unassigned | Unassigned | 191_19 | Clustered | 12 | VC_191_19 | 12 | 0.0655 | 1 | 0.0655 | 0 | 0 | 0 | 0.8636 | Ref |
| 12706 | MW358927 | Unassigned | Unassigned | Unassigned | 288_0 | Clustered | 3 | VC_288_0 | 3 | 0.042 | 0.99856109 | 0.042 | 0 | 0 | 0 | 1 | Ref |
| 12707 | MW358928 | Unassigned | Unassigned | Unassigned | 396_0 | Clustered | 2 | VC_396_0 | 2 | 0.0714 | 0.95759359 | 0.0684 | 0 | 0 | 0 | 1 | Ref |
| 12708 | MW358930 | Unassigned | Unassigned | Unassigned | 112_0 | Clustered | 8 | VC_112_0 | 8 | 0.2537 | 1 | 0.2537 | 0 | 0 | 0 | 1 | Ref |
| 12709 | MW358931 | Unassigned | Unassigned | Unassigned | 390_0 | Clustered | 4 | VC_390_0 | 4 | 0.0468 | 0.9999882 | 0.0468 | 0 | 0 | 0 | 1 | Ref |
| 12710 | MW362867 | Unassigned | Unassigned | Unassigned | 329_11 | Clustered | 15 | VC_329_11 | 15 | 0.0791 | 1 | 0.0791 | 0 | 0 | 0 | 0.9333 | Ref |
| 12711 | MW363798 | Unassigned | Unassigned | Unassigned | 153_0 | Clustered | 25 | VC_153_0 | 25 | 0.7729 | 1 | 0.7729 | 0 | 0 | 0 | 1 | Ref |
| 12712 | MW363799 | Unassigned | Unassigned | Unassigned | 96_0 | Clustered | 30 | VC_96_0 | 30 | 0.718 | 1 | 0.718 | 0 | 0 | 0 | 1 | Ref |
| 12713 | MW365946 | Unassigned | Unassigned | Unassigned | 1171_0 | Clustered | 21 | VC_1171_0 | 21 | 0.7125 | 1 | 0.7125 | 0 | 0 | 0 | 1 | Ref |
| 12714 | MW365947 | Unassigned | Unassigned | Unassigned | 1360_0 | Clustered | 31 | VC_1360_0 | 31 | 0.8837 | 1 | 0.8837 | 0 | 0 | 0 | 1 | Ref |
| 12715 | MW365948 | Unassigned | Unassigned | Unassigned | 1360_0 | Clustered | 31 | VC_1360_0 | 31 | 0.8837 | 1 | 0.8837 | 0 | 0 | 0 | 1 | Ref |
| 12716 | MW365949 | Unassigned | Unassigned | Unassigned | 1171_0 | Clustered | 21 | VC_1171_0 | 21 | 0.7125 | 1 | 0.7125 | 0 | 0 | 0 | 1 | Ref |
| 12717 | MW365950 | Unassigned | Unassigned | Unassigned | 1171_0 | Clustered | 21 | VC_1171_0 | 21 | 0.7125 | 1 | 0.7125 | 0 | 0 | 0 | 1 | Ref |
| 12718 | MW365951 | Unassigned | Unassigned | Unassigned | 982_0 | Clustered | 62 | VC_982_0 | 62 | 0.8646 | 1 | 0.8646 | 0 | 0 | 0 | 0.743 | Ref |
| 12719 | MW365952 | Unassigned | Unassigned | Unassigned | 1072_2 | Clustered | 5 | VC_1072_2 | 5 | 0.1003 | 0.99999315 | 0.1003 | 0 | 0 | 0 | 1 | Ref |
| 12720 | MW365953 | Unassigned | Unassigned | Unassigned | 982_0 | Clustered | 62 | VC_982_0 | 62 | 0.8646 | 1 | 0.8646 | 0 | 0 | 0 | 0.743 | Ref |
| 12721 | MW366783 | Unassigned | Unassigned | Unassigned | 699_0 | Clustered | 71 | VC_699_0 | 71 | 0.9352 | 1 | 0.9352 | 0 | 0 | 0 | 1 | Ref |
| 12722 | MW366784 | Unassigned | Unassigned | Unassigned | 699_0 | Clustered | 71 | VC_699_0 | 71 | 0.9352 | 1 | 0.9352 | 0 | 0 | 0 | 1 | Ref |
| 12723 | MW366843 | Unassigned | Unassigned | Unassigned | 67_0 | Clustered | 2 | VC_67_0 | 2 | 0.014 | 0.69357611 | 0.0097 | 0 | 0 | 0 | 1 | Ref |
| 12724 | MW367417 | Unassigned | Unassigned | Unassigned | 849_8 | Clustered/Singleton | 1 | VC_849_8 | 1 | 0 | 1 | 0 | 0 | 0 | 0 | 1 | Ref |
| 12725 | MW367418 | Unassigned | Unassigned | Unassigned | 386_0 | Clustered | 49 | VC_386_0 | 49 | 0.7941 | 1 | 0.7941 | 0 | 0 | 0 | 1 | Ref |
| 12726 | MW367898 | Unassigned | Unassigned | Unassigned | 288_3 | Clustered | 3 | VC_288_3 | 3 | 0.0452 | 0.99844025 | 0.0452 | 0 | 0 | 0 | 1 | Ref |
| 12727 | MW380983 | Unassigned | Unassigned | Unassigned | 793_0 | Clustered | 12 | VC_793_0 | 12 | 0.8531 | 1 | 0.8531 | 0 | 0 | 0 | 1 | Ref |
| 12728 | MW380984 | Unassigned | Unassigned | Unassigned | 793_0 | Clustered | 12 | VC_793_0 | 12 | 0.8531 | 1 | 0.8531 | 0 | 0 | 0 | 1 | Ref |
| 12729 | MW380985 | Unassigned | Unassigned | Unassigned | 793_0 | Clustered | 12 | VC_793_0 | 12 | 0.8531 | 1 | 0.8531 | 0 | 0 | 0 | 1 | Ref |
| 12730 | MW384823 | Unassigned | Unassigned | Unassigned | 959_1 | Clustered | 8 | VC_959_1 | 8 | 0.0878 | 0.98435099 | 0.0864 | 0 | 0 | 0 | 1 | Ref |
| 12731 | MW385300 | Unassigned | Unassigned | Unassigned | 288_0 | Clustered | 3 | VC_288_0 | 3 | 0.042 | 0.99856109 | 0.042 | 0 | 0 | 0 | 1 | Ref |
| 12732 | MW387018 | Unassigned | Unassigned | Unassigned | 626_0 | Clustered | 23 | VC_626_0 | 23 | 0.66 | 1 | 0.66 | 0 | 0 | 0 | 0.8379 | Ref |
| 12733 | MW388001 | Unassigned | Unassigned | Unassigned | 492_2 | Clustered | 8 | VC_492_2 | 8 | 0.1221 | 0.98786574 | 0.1206 | 0 | 0 | 0 | 1 | Ref |
| 12734 | MW388002 | Unassigned | Unassigned | Unassigned | 492_2 | Clustered | 8 | VC_492_2 | 8 | 0.1221 | 0.98786574 | 0.1206 | 0 | 0 | 0 | 1 | Ref |
| 12735 | MW388003 | Unassigned | Unassigned | Unassigned | 492_2 | Clustered | 8 | VC_492_2 | 8 | 0.1221 | 0.98786574 | 0.1206 | 0 | 0 | 0 | 1 | Ref |
| 12736 | MW388004 | Unassigned | Unassigned | Unassigned | 492_2 | Clustered | 8 | VC_492_2 | 8 | 0.1221 | 0.98786574 | 0.1206 | 0 | 0 | 0 | 1 | Ref |
| 12737 | MW388005 | Unassigned | Unassigned | Unassigned | 133_15 | Clustered | 2 | VC_133_15 | 2 | 0.0051 | 0.84786437 | 0.0044 | 0 | 0 | 0 | 1 | Ref |
| 12738 | MW390886 | Unassigned | Unassigned | Unassigned | 179_0 | Clustered | 53 | VC_179_0 | 53 | 0.9137 | 1 | 0.9137 | 0 | 0 | 0 | 1 | Ref |
| 12739 | MW392801 | Unassigned | Unassigned | Unassigned | 1006_0 | Clustered | 8 | VC_1006_0 | 8 | 0.7407 | 1 | 0.7407 | 0 | 0 | 0 | 1 | Ref |
| 12740 | MW392802 | Unassigned | Unassigned | Unassigned | 1006_0 | Clustered | 8 | VC_1006_0 | 8 | 0.7407 | 1 | 0.7407 | 0 | 0 | 0 | 1 | Ref |
| 12741 | MW392803 | Unassigned | Unassigned | Unassigned | 884_1 | Clustered | 8 | VC_884_1 | 8 | 0.1102 | 0.9989323 | 0.1101 | 0 | 0 | 0 | 0.75 | Ref |
| 12742 | MW393850 | Unassigned | Unassigned | Unassigned | 671_0 | Clustered | 5 | VC_671_0 | 5 | 0.5289 | 0.99999996 | 0.5289 | 0 | 0 | 0 | 1 | Ref |
| 12743 | MW394389 | Unassigned | Unassigned | Unassigned | 452_0 | Clustered | 66 | VC_452_0 | 66 | 0.9248 | 1 | 0.9248 | 0 | 0 | 0 | 1 | Ref |
| 12744 | MW394391 | Unassigned | Unassigned | Unassigned | 488_0 | Clustered | 8 | VC_488_0 | 8 | 0.2938 | 1 | 0.2938 | 0 | 0 | 0 | 1 | Ref |
| 12745 | MW398869 | Unassigned | Unassigned | Unassigned | 971_1 | Clustered | 24 | VC_971_1 | 24 | 0.7393 | 1 | 0.7393 | 0 | 0 | 0 | 0.8913 | Ref |
| 12746 | MW406974 | Unassigned | Unassigned | Unassigned | 79_1 | Clustered | 2 | VC_79_1 | 2 | 0.006 | 0.95748743 | 0.0057 | 0 | 0 | 0 | 1 | Ref |
| 12747 | MW406975 | Unassigned | Unassigned | Unassigned | 423_0 | Clustered | 45 | VC_423_0 | 45 | 0.9484 | 1 | 0.9484 | 0 | 0 | 0 | 1 | Ref |
| 12748 | MW406976 | Unassigned | Unassigned | Unassigned | 423_0 | Clustered | 45 | VC_423_0 | 45 | 0.9484 | 1 | 0.9484 | 0 | 0 | 0 | 1 | Ref |
| 12749 | MW411578 | Unassigned | Unassigned | Unassigned | 191_9 | Clustered | 5 | VC_191_9 | 5 | 0.0228 | 0.99999662 | 0.0228 | 0 | 0 | 0 | 0.7 | Ref |
| 12750 | MW411580 | Unassigned | Unassigned | Unassigned | 904_2 | Clustered | 7 | VC_904_2 | 7 | 0.2857 | 1 | 0.2857 | 0 | 0 | 0 | 1 | Ref |
| 12751 | MW413353 | Unassigned | Unassigned | Unassigned | 428_0 | Clustered | 33 | VC_428_0 | 33 | 0.9915 | 1 | 0.9915 | 0 | 0 | 0 | 1 | Ref |
| 12752 | MW416012 | Unassigned | Unassigned | Unassigned | 451_0 | Clustered | 80 | VC_451_0 | 80 | 0.9991 | 1 | 0.9991 | 0 | 0 | 0 | 1 | Ref |
| 12753 | MW417503 | Unassigned | Unassigned | Unassigned | 937_0 | Clustered | 40 | VC_937_0 | 40 | 0.6809 | 1 | 0.6809 | 0 | 0 | 0 | 0.9359 | Ref |
| 12754 | MW419084 | Unassigned | Unassigned | Unassigned | 85_1 | Clustered | 4 | VC_85_1 | 4 | 0.0507 | 0.99996692 | 0.0507 | 0 | 0 | 0 | 1 | Ref |
| 12755 | MW419085 | Unassigned | Unassigned | Unassigned | 85_1 | Clustered | 4 | VC_85_1 | 4 | 0.0507 | 0.99996692 | 0.0507 | 0 | 0 | 0 | 1 | Ref |
| 12756 | MW419086 | Unassigned | Unassigned | Unassigned | 85_1 | Clustered | 4 | VC_85_1 | 4 | 0.0507 | 0.99996692 | 0.0507 | 0 | 0 | 0 | 1 | Ref |
| 12757 | MW419087 | Unassigned | Unassigned | Unassigned | 85_1 | Clustered | 4 | VC_85_1 | 4 | 0.0507 | 0.99996692 | 0.0507 | 0 | 0 | 0 | 1 | Ref |
| 12758 | MW419910 | Unassigned | Unassigned | Unassigned | 836_0 | Clustered | 9 | VC_836_0 | 9 | 0.7733 | 1 | 0.7733 | 0 | 0 | 0 | 1 | Ref |
| 12759 | MW421576 | Unassigned | Unassigned | Unassigned | 1023_0 | Clustered | 8 | VC_1023_0 | 8 | 0.8574 | 1 | 0.8574 | 0 | 0 | 0 | 1 | Ref |
| 12760 | MW421577 | Unassigned | Unassigned | Unassigned | 1157_0 | Clustered | 24 | VC_1157_0 | 24 | 1 | 1 | 1 | 0 | 0 | 0 | 1 | Ref |
| 12761 | MW421578 | Unassigned | Unassigned | Unassigned | 1157_0 | Clustered | 24 | VC_1157_0 | 24 | 1 | 1 | 1 | 0 | 0 | 0 | 1 | Ref |
| 12762 | MW421579 | Unassigned | Unassigned | Unassigned | 1157_0 | Clustered | 24 | VC_1157_0 | 24 | 1 | 1 | 1 | 0 | 0 | 0 | 1 | Ref |
| 12763 | MW421580 | Unassigned | Unassigned | Unassigned | 1157_0 | Clustered | 24 | VC_1157_0 | 24 | 1 | 1 | 1 | 0 | 0 | 0 | 1 | Ref |
| 12764 | MW421581 | Unassigned | Unassigned | Unassigned | 1021_0 | Clustered | 10 | VC_1021_0 | 10 | 0.8876 | 1 | 0.8876 | 0 | 0 | 0 | 1 | Ref |
| 12765 | MW421582 | Unassigned | Unassigned | Unassigned | 1021_0 | Clustered | 10 | VC_1021_0 | 10 | 0.8876 | 1 | 0.8876 | 0 | 0 | 0 | 1 | Ref |
| 12766 | MW421583 | Unassigned | Unassigned | Unassigned | 1021_0 | Clustered | 10 | VC_1021_0 | 10 | 0.8876 | 1 | 0.8876 | 0 | 0 | 0 | 1 | Ref |
| 12767 | MW421584 | Unassigned | Unassigned | Unassigned | 1021_0 | Clustered | 10 | VC_1021_0 | 10 | 0.8876 | 1 | 0.8876 | 0 | 0 | 0 | 1 | Ref |
| 12768 | MW423797 | Unassigned | Unassigned | Unassigned | 329_16 | Clustered | 4 | VC_329_16 | 4 | 0.0158 | 0.9999807 | 0.0158 | 0 | 0 | 0 | 1 | Ref |
| 12769 | MW423798 | Unassigned | Unassigned | Unassigned | 191_33 | Clustered/Singleton | 1 | VC_191_33 | 1 | 0 | 1 | 0 | 0 | 0 | 0 | 1 | Ref |
| 12770 | MW427215 | Unassigned | Unassigned | Unassigned | 461_0 | Clustered | 96 | VC_461_0 | 96 | 0.9307 | 1 | 0.9307 | 0 | 0 | 0 | 0.9575 | Ref |
| 12771 | MW430345 | Unassigned | Unassigned | Unassigned | 151_0 | Clustered | 6 | VC_151_0 | 6 | 0.0735 | 1 | 0.0735 | 0 | 0 | 0 | 1 | Ref |
| 12772 | MW435853 | Unassigned | Unassigned | Unassigned | 1072_3 | Clustered | 6 | VC_1072_3 | 6 | 0.1304 | 0.99999994 | 0.1304 | 0 | 0 | 0 | 0.8667 | Ref |
| 12773 | MW435854 | Unassigned | Unassigned | Unassigned | 238_0 | Clustered | 17 | VC_238_0 | 17 | 0.6109 | 1 | 0.6109 | 0 | 0 | 0 | 1 | Ref |
| 12774 | MW435855 | Unassigned | Unassigned | Unassigned | 887_0 | Clustered | 18 | VC_887_0 | 18 | 0.3663 | 1 | 0.3663 | 0 | 0 | 0 | 1 | Ref |
| 12775 | MW435856 | Unassigned | Unassigned | Unassigned | 982_0 | Clustered | 62 | VC_982_0 | 62 | 0.8646 | 1 | 0.8646 | 0 | 0 | 0 | 0.743 | Ref |
| 12776 | MW435857 | Unassigned | Unassigned | Unassigned | 372_0 | Clustered | 24 | VC_372_0 | 24 | 0.5539 | 1 | 0.5539 | 0 | 0 | 0 | 1 | Ref |
| 12777 | MW435858 | Unassigned | Unassigned | Unassigned | 325_32 | Clustered/Singleton | 1 | VC_325_32 | 1 | 0 | 1 | 0 | 0 | 0 | 0 | 1 | Ref |
| 12778 | MW435859 | Unassigned | Unassigned | Unassigned | 1240_0 | Clustered | 82 | VC_1240_0 | 82 | 0.7945 | 1 | 0.7945 | 0 | 0 | 0 | 1 | Ref |
| 12779 | MW442825 | Unassigned | Unassigned | Unassigned | 1157_0 | Clustered | 24 | VC_1157_0 | 24 | 1 | 1 | 1 | 0 | 0 | 0 | 1 | Ref |
| 12780 | MW442826 | Unassigned | Unassigned | Unassigned | 1157_0 | Clustered | 24 | VC_1157_0 | 24 | 1 | 1 | 1 | 0 | 0 | 0 | 1 | Ref |
| 12781 | MW442827 | Unassigned | Unassigned | Unassigned | 1157_0 | Clustered | 24 | VC_1157_0 | 24 | 1 | 1 | 1 | 0 | 0 | 0 | 1 | Ref |
| 12782 | MW442828 | Unassigned | Unassigned | Unassigned | 1157_0 | Clustered | 24 | VC_1157_0 | 24 | 1 | 1 | 1 | 0 | 0 | 0 | 1 | Ref |
| 12783 | MW442829 | Unassigned | Unassigned | Unassigned | 1157_0 | Clustered | 24 | VC_1157_0 | 24 | 1 | 1 | 1 | 0 | 0 | 0 | 1 | Ref |
| 12784 | MW442830 | Unassigned | Unassigned | Unassigned | 1157_0 | Clustered | 24 | VC_1157_0 | 24 | 1 | 1 | 1 | 0 | 0 | 0 | 1 | Ref |
| 12785 | MW448170 | Unassigned | Unassigned | Unassigned | 668_0 | Clustered | 25 | VC_668_0 | 25 | 0.5539 | 1 | 0.5539 | 0 | 0 | 0 | 0.96 | Ref |
| 12786 | MW448539 | Unassigned | Unassigned | Unassigned | 1157_0 | Clustered | 24 | VC_1157_0 | 24 | 1 | 1 | 1 | 0 | 0 | 0 | 1 | Ref |
| 12787 | MW448540 | Unassigned | Unassigned | Unassigned | 1157_0 | Clustered | 24 | VC_1157_0 | 24 | 1 | 1 | 1 | 0 | 0 | 0 | 1 | Ref |
| 12788 | MW448541 | Unassigned | Unassigned | Unassigned | 1157_0 | Clustered | 24 | VC_1157_0 | 24 | 1 | 1 | 1 | 0 | 0 | 0 | 1 | Ref |
| 12789 | MW450817 | Unassigned | Unassigned | Unassigned | 310_0 | Clustered | 33 | VC_310_0 | 33 | 0.8771 | 1 | 0.8771 | 0 | 0 | 0 | 1 | Ref |
| 12790 | MW451246 | Unassigned | Unassigned | Unassigned | 882_0 | Clustered | 20 | VC_882_0 | 20 | 0.9917 | 1 | 0.9917 | 0 | 0 | 0 | 1 | Ref |
| 12791 | MW451247 | Unassigned | Unassigned | Unassigned | 1027_0 | Clustered | 20 | VC_1027_0 | 20 | 0.7114 | 1 | 0.7114 | 0 | 0 | 0 | 1 | Ref |
| 12792 | MW451248 | Unassigned | Unassigned | Unassigned | 530_0 | Clustered | 9 | VC_530_0 | 9 | 0.8027 | 1 | 0.8027 | 0 | 0 | 0 | 1 | Ref |
| 12793 | MW451250 | Unassigned | Unassigned | Unassigned | 845_0 | Clustered | 3 | VC_845_0 | 3 | 0.3348 | 0.99831497 | 0.3343 | 0 | 0 | 0 | 1 | Ref |
| 12794 | MW451251 | Unassigned | Unassigned | Unassigned | 882_0 | Clustered | 20 | VC_882_0 | 20 | 0.9917 | 1 | 0.9917 | 0 | 0 | 0 | 1 | Ref |
| 12795 | MW452562 | Unassigned | Unassigned | Unassigned | 769_0 | Clustered | 2 | VC_769_0 | 2 | 0.0483 | 0.95514297 | 0.0462 | 0 | 0 | 0 | 1 | Ref |
| 12796 | MW452941 | Unassigned | Unassigned | Unassigned | 913_0 | Clustered | 2 | VC_913_0 | 2 | 0.1042 | 0.82306775 | 0.0858 | 0 | 0 | 0 | 1 | Ref |

| | | | | | | | | | | | | | | | | | |
|---|---|---|---|---|---|---|---|---|---|---|---|---|---|---|---|---|---|
| 12797 | MW459163 | Unassigned | Unassigned | Unassigned | 699_0 | Clustered | 71 | VC_699_0 | 71 | 0.9352 | 1 | 0.9352 | 0 | 0 | 0 | 1 | Ref |
| 12798 | MW460245 | Unassigned | Unassigned | Unassigned | 66_2 | Clustered/Singleton | 1 | VC_66_2 | 1 | 0 | 1 | 0 | 0 | 0 | 0 | 1 | Ref |
| 12799 | MW460246 | Unassigned | Unassigned | Unassigned | 536_0 | Clustered | 3 | VC_536_0 | 3 | 0.0878 | 0.9985359 | 0.0877 | 0 | 0 | 0 | 1 | Ref |
| 12800 | MW460248 | Unassigned | Unassigned | Unassigned | 574_0 | Clustered | 7 | VC_574_0 | 7 | 0.2734 | 1 | 0.2734 | 0 | 0 | 0 | 1 | Ref |
| 12801 | MW460250 | Unassigned | Unassigned | Unassigned | 149_8 | Clustered | 66 | VC_149_8 | 66 | 0.4565 | 1 | 0.4565 | 0 | 0 | 0 | 0.8182 | Ref |
| 12802 | MW462191 | Unassigned | Unassigned | Unassigned | 466_0 | Clustered | 3 | VC_466_0 | 3 | 0.3264 | 0.99747305 | 0.3255 | 0 | 0 | 0 | 1 | Ref |
| 12803 | MW462221 | Unassigned | Unassigned | Unassigned | 1454_0 | Clustered | 2 | VC_1454_0 | 2 | 0.6937 | 0.85557782 | 0.5935 | 0 | 0 | 0 | 0 | Ref |
| 12804 | MW476489 | Unassigned | Unassigned | Unassigned | 307_0 | Clustered | 2 | VC_307_0 | 2 | 0.3835 | 0.95251406 | 0.3653 | 0 | 0 | 0 | 1 | Ref |
| 12805 | MW477480 | Unassigned | Unassigned | Unassigned | 817_0 | Clustered | 14 | VC_817_0 | 14 | 0.6342 | 1 | 0.6342 | 0 | 0 | 0 | 1 | Ref |
| 12806 | MW477799 | Unassigned | Unassigned | Unassigned | 461_0 | Clustered | 96 | VC_461_0 | 96 | 0.9307 | 1 | 0.9307 | 0 | 0 | 0 | 0.9575 | Ref |
| 12807 | MW478289 | Unassigned | Unassigned | Unassigned | 417_0 | Clustered | 5 | VC_417_0 | 5 | 0.6629 | 0.9977434 | 0.6614 | 0 | 0 | 0 | 0.8 | Ref |
| 12808 | MW478292 | Unassigned | Unassigned | Unassigned | 542_0 | Clustered | 2 | VC_542_0 | 2 | 0.3753 | 0.94092208 | 0.3531 | 0 | 0 | 0 | 1 | Ref |
| 12809 | MW481326 | Unassigned | Unassigned | Unassigned | 329_7 | Clustered | 18 | VC_329_7 | 18 | 0.0938 | 1 | 0.0938 | 0 | 0 | 0 | 0.7778 | Ref |
| 12810 | MW495853 | Unassigned | Unassigned | Unassigned | 412_0 | Clustered | 39 | VC_412_0 | 39 | 0.845 | 1 | 0.845 | 0 | 0 | 0 | 1 | Ref |
| 12811 | MW505928 | Unassigned | Unassigned | Unassigned | 765_0 | Clustered | 132 | VC_765_0 | 132 | 1 | 1 | 1 | 0 | 0 | 0 | 1 | Ref |
| 12812 | MW507123 | Unassigned | Unassigned | Unassigned | 325_23 | Clustered | 17 | VC_325_23 | 17 | 0.0716 | 1 | 0.0716 | 0 | 0 | 0 | 0.6397 | Ref |
| 12813 | MW507124 | Unassigned | Unassigned | Unassigned | 1360_0 | Clustered | 31 | VC_1360_0 | 31 | 0.8837 | 1 | 0.8837 | 0 | 0 | 0 | 1 | Ref |
| 12814 | MW507125 | Unassigned | Unassigned | Unassigned | 1178_0 | Clustered | 24 | VC_1178_0 | 24 | 0.9524 | 1 | 0.9524 | 0 | 0 | 0 | 1 | Ref |
| 12815 | MW507126 | Unassigned | Unassigned | Unassigned | 1168_0 | Clustered | 2 | VC_1168_0 | 2 | 0.099 | 0.93550199 | 0.0927 | 0 | 0 | 0 | 1 | Ref |
| 12816 | MW507127 | Unassigned | Unassigned | Unassigned | 1344_0 | Clustered | 78 | VC_1344_0 | 78 | 0.9732 | 1 | 0.9732 | 0 | 0 | 0 | 1 | Ref |
| 12817 | MW507128 | Unassigned | Unassigned | Unassigned | 514_0 | Clustered | 75 | VC_514_0 | 75 | 0.9402 | 1 | 0.9402 | 0 | 0 | 0 | 1 | Ref |
| 12818 | MW507129 | Unassigned | Unassigned | Unassigned | 767_0 | Clustered | 34 | VC_767_0 | 34 | 0.9173 | 1 | 0.9173 | 0 | 0 | 0 | 1 | Ref |
| 12819 | MW507130 | Unassigned | Unassigned | Unassigned | 1175_0 | Clustered | 22 | VC_1175_0 | 22 | 0.4562 | 1 | 0.4562 | 0 | 0 | 0 | 1 | Ref |
| 12820 | MW507131 | Unassigned | Unassigned | Unassigned | 1166_0 | Clustered | 20 | VC_1166_0 | 20 | 0.99 | 1 | 0.99 | 0 | 0 | 0 | 1 | Ref |
| 12821 | MW507132 | Unassigned | Unassigned | Unassigned | 868_0 | Clustered | 25 | VC_868_0 | 25 | 0.5791 | 1 | 0.5791 | 0 | 0 | 0 | 1 | Ref |
| 12822 | MW507133 | Unassigned | Unassigned | Unassigned | 868_0 | Clustered | 25 | VC_868_0 | 25 | 0.5791 | 1 | 0.5791 | 0 | 0 | 0 | 1 | Ref |
| 12823 | MW507134 | Unassigned | Unassigned | Unassigned | 1071_0 | Clustered | 20 | VC_1071_0 | 20 | 0.3268 | 1 | 0.3268 | 0 | 0 | 0 | 0.9842 | Ref |
| 12824 | MW507135 | Unassigned | Unassigned | Unassigned | 919_0 | Clustered | 39 | VC_919_0 | 39 | 0.9069 | 1 | 0.9069 | 0 | 0 | 0 | 1 | Ref |
| 12825 | MW507136 | Unassigned | Unassigned | Unassigned | 1071_3 | Clustered | 16 | VC_1071_3 | 16 | 0.2354 | 1 | 0.2354 | 0 | 0 | 0 | 0.9667 | Ref |
| 12826 | MW508890 | Unassigned | Unassigned | Unassigned | 428_0 | Clustered | 33 | VC_428_0 | 33 | 0.9915 | 1 | 0.9915 | 0 | 0 | 0 | 1 | Ref |
| 12827 | MW512264 | Unassigned | Unassigned | Unassigned | 286_0 | Clustered | 2 | VC_286_0 | 2 | 0.0073 | 0.77462993 | 0.0056 | 0 | 0 | 0 | 1 | Ref |
| 12828 | MW512571 | Unassigned | Unassigned | Unassigned | 647_0 | Clustered | 11 | VC_647_0 | 11 | 0.598 | 1 | 0.598 | 0 | 0 | 0 | 1 | Ref |
| 12829 | MW512572 | Unassigned | Unassigned | Unassigned | 647_0 | Clustered | 11 | VC_647_0 | 11 | 0.598 | 1 | 0.598 | 0 | 0 | 0 | 1 | Ref |
| 12830 | MW512830 | Unassigned | Unassigned | Unassigned | 79_6 | Clustered | 11 | VC_79_6 | 11 | 0.0528 | 1 | 0.0528 | 0 | 0 | 0 | 0.7455 | Ref |
| 12831 | MW512831 | Unassigned | Unassigned | Unassigned | 472_0 | Clustered | 47 | VC_472_0 | 47 | 0.9025 | 1 | 0.9025 | 0 | 0 | 0 | 1 | Ref |
| 12832 | MW512832 | Unassigned | Unassigned | Unassigned | 749_0 | Clustered | 40 | VC_749_0 | 40 | 0.7517 | 1 | 0.7517 | 0 | 0 | 0 | 1 | Ref |
| 12833 | MW512833 | Unassigned | Unassigned | Unassigned | 79_0 | Clustered | 64 | VC_79_0 | 64 | 0.4185 | 1 | 0.4185 | 0 | 0 | 0 | 0.7649 | Ref |
| 12834 | MW512834 | Unassigned | Unassigned | Unassigned | 79_6 | Clustered | 11 | VC_79_6 | 11 | 0.0528 | 1 | 0.0528 | 0 | 0 | 0 | 0.7455 | Ref |
| 12835 | MW514246 | Unassigned | Unassigned | Unassigned | 305_0 | Clustered | 12 | VC_305_0 | 12 | 0.5745 | 1 | 0.5745 | 0 | 0 | 0 | 0.8485 | Ref |
| 12836 | MW514247 | Unassigned | Unassigned | Unassigned | 305_0 | Clustered | 12 | VC_305_0 | 12 | 0.5745 | 1 | 0.5745 | 0 | 0 | 0 | 0.8485 | Ref |
| 12837 | MW526259 | Unassigned | Unassigned | Unassigned | 76_0 | Clustered | 37 | VC_76_0 | 37 | 0.9403 | 1 | 0.9403 | 0 | 0 | 0 | 1 | Ref |
| 12838 | MW526260 | Unassigned | Unassigned | Unassigned | 472_0 | Clustered | 47 | VC_472_0 | 47 | 0.9025 | 1 | 0.9025 | 0 | 0 | 0 | 1 | Ref |
| 12839 | MW528836 | Unassigned | Unassigned | Unassigned | 149_8 | Clustered | 66 | VC_149_8 | 66 | 0.4565 | 1 | 0.4565 | 0 | 0 | 0 | 0.8182 | Ref |
| 12840 | MW534374 | Unassigned | Unassigned | Unassigned | 866_0 | Clustered | 4 | VC_866_0 | 4 | 0.0795 | 0.99998595 | 0.0795 | 0 | 0 | 0 | 1 | Ref |
| 12841 | MW534375 | Unassigned | Unassigned | Unassigned | 557_3 | Clustered/Singleton | 1 | VC_557_3 | 1 | 0 | 1 | 0 | 0 | 0 | 0 | 1 | Ref |
| 12842 | MW534376 | Unassigned | Unassigned | Unassigned | 954_0 | Clustered | 13 | VC_954_0 | 13 | 0.863 | 1 | 0.863 | 0 | 0 | 0 | 1 | Ref |
| 12843 | MW534377 | Unassigned | Unassigned | Unassigned | 371_1 | Clustered | 11 | VC_371_1 | 11 | 0.0379 | 1 | 0.0379 | 0 | 0 | 0 | 0.8545 | Ref |
| 12844 | MW534378 | Unassigned | Unassigned | Unassigned | 685_0 | Clustered | 16 | VC_685_0 | 16 | 0.2972 | 1 | 0.2972 | 0 | 0 | 0 | 0.9167 | Ref |
| 12845 | MW534379 | Unassigned | Unassigned | Unassigned | 371_16 | Clustered | 42 | VC_371_16 | 42 | 0.1341 | 1 | 0.1341 | 0 | 0 | 0 | 0.7282 | Ref |
| 12846 | MW534380 | Unassigned | Unassigned | Unassigned | 371_16 | Clustered | 42 | VC_371_16 | 42 | 0.1341 | 1 | 0.1341 | 0 | 0 | 0 | 0.7282 | Ref |
| 12847 | MW534381 | Unassigned | Unassigned | Unassigned | 371_11 | Clustered | 31 | VC_371_11 | 31 | 0.0927 | 1 | 0.0927 | 0 | 0 | 0 | 0.7011 | Ref |
| 12848 | MW544066 | Unassigned | Unassigned | Unassigned | 780_0 | Clustered | 8 | VC_780_0 | 8 | 0.5207 | 1 | 0.5207 | 0 | 0 | 0 | 1 | Ref |
| 12849 | MW546064 | Unassigned | Unassigned | Unassigned | 149_8 | Clustered | 66 | VC_149_8 | 66 | 0.4565 | 1 | 0.4565 | 0 | 0 | 0 | 0.8182 | Ref |
| 12850 | MW546065 | Unassigned | Unassigned | Unassigned | 149_8 | Clustered | 66 | VC_149_8 | 66 | 0.4565 | 1 | 0.4565 | 0 | 0 | 0 | 0.8182 | Ref |
| 12851 | MW546066 | Unassigned | Unassigned | Unassigned | 149_8 | Clustered | 66 | VC_149_8 | 66 | 0.4565 | 1 | 0.4565 | 0 | 0 | 0 | 0.8182 | Ref |
| 12852 | MW546067 | Unassigned | Unassigned | Unassigned | 149_8 | Clustered | 66 | VC_149_8 | 66 | 0.4565 | 1 | 0.4565 | 0 | 0 | 0 | 0.8182 | Ref |
| 12853 | MW546068 | Unassigned | Unassigned | Unassigned | 149_8 | Clustered | 66 | VC_149_8 | 66 | 0.4565 | 1 | 0.4565 | 0 | 0 | 0 | 0.8182 | Ref |
| 12854 | MW546069 | Unassigned | Unassigned | Unassigned | 149_8 | Clustered | 66 | VC_149_8 | 66 | 0.4565 | 1 | 0.4565 | 0 | 0 | 0 | 0.8182 | Ref |
| 12855 | MW546070 | Unassigned | Unassigned | Unassigned | 149_8 | Clustered | 66 | VC_149_8 | 66 | 0.4565 | 1 | 0.4565 | 0 | 0 | 0 | 0.8182 | Ref |
| 12856 | MW546071 | Unassigned | Unassigned | Unassigned | 153_0 | Clustered | 25 | VC_153_0 | 25 | 0.7729 | 1 | 0.7729 | 0 | 0 | 0 | 1 | Ref |
| 12857 | MW546072 | Unassigned | Unassigned | Unassigned | 149_8 | Clustered | 66 | VC_149_8 | 66 | 0.4565 | 1 | 0.4565 | 0 | 0 | 0 | 0.8182 | Ref |
| 12858 | MW546073 | Unassigned | Unassigned | Unassigned | 149_15 | Clustered | 16 | VC_149_15 | 16 | 0.0788 | 1 | 0.0788 | 0 | 0 | 0 | 0.8083 | Ref |
| 12859 | MW546074 | Unassigned | Unassigned | Unassigned | 149_8 | Clustered | 66 | VC_149_8 | 66 | 0.4565 | 1 | 0.4565 | 0 | 0 | 0 | 0.8182 | Ref |
| 12860 | MW546075 | Unassigned | Unassigned | Unassigned | 153_0 | Clustered | 25 | VC_153_0 | 25 | 0.7729 | 1 | 0.7729 | 0 | 0 | 0 | 1 | Ref |
| 12861 | MW546076 | Unassigned | Unassigned | Unassigned | 153_0 | Clustered | 25 | VC_153_0 | 25 | 0.7729 | 1 | 0.7729 | 0 | 0 | 0 | 1 | Ref |
| 12862 | MW546077 | Unassigned | Unassigned | Unassigned | 153_0 | Clustered | 25 | VC_153_0 | 25 | 0.7729 | 1 | 0.7729 | 0 | 0 | 0 | 1 | Ref |
| 12863 | MW554488 | Unassigned | Unassigned | Unassigned | 9_0 | Clustered | 38 | VC_9_0 | 38 | 0.4895 | 1 | 0.4895 | 0 | 0 | 0 | 1 | Ref |
| 12864 | MW557846 | Unassigned | Unassigned | Unassigned | 80_0 | Clustered | 41 | VC_80_0 | 41 | 0.5837 | 1 | 0.5837 | 0 | 0 | 0 | 0.9805 | Ref |
| 12865 | MW557853 | Unassigned | Unassigned | Unassigned | 306_0 | Clustered | 2 | VC_306_0 | 2 | 1 | 1 | 1 | 0 | 0 | 0 | 1 | Ref |
| 12866 | MW567727 | Unassigned | Unassigned | Unassigned | 191_22 | Clustered/Singleton | 1 | VC_191_22 | 1 | 0 | 1 | 0 | 0 | 0 | 0 | 1 | Ref |
| 12867 | MW570842 | Unassigned | Unassigned | Unassigned | 958_0 | Clustered | 3 | VC_958_0 | 3 | 0.1441 | 0.99848014 | 0.1438 | 0 | 0 | 0 | 1 | Ref |
| 12868 | MW574966 | Unassigned | Unassigned | Unassigned | 470_0 | Clustered | 3 | VC_470_0 | 3 | 0.5552 | 0.99823354 | 0.5543 | 0 | 0 | 0 | 1 | Ref |
| 12869 | MW578835 | Unassigned | Unassigned | Unassigned | 888_0 | Clustered | 28 | VC_888_0 | 28 | 0.5497 | 1 | 0.5497 | 0 | 0 | 0 | 1 | Ref |
| 12870 | MW578836 | Unassigned | Unassigned | Unassigned | 763_2 | Clustered | 7 | VC_763_2 | 7 | 0.2612 | 1 | 0.2612 | 0 | 0 | 0 | 1 | Ref |
| 12871 | MW578837 | Unassigned | Unassigned | Unassigned | 1352_0 | Clustered | 31 | VC_1352_0 | 31 | 0.849 | 1 | 0.849 | 0 | 0 | 0 | 1 | Ref |
| 12872 | MW582531 | Unassigned | Unassigned | Unassigned | 769_0 | Clustered | 2 | VC_769_0 | 2 | 0.0483 | 0.95514297 | 0.0462 | 0 | 0 | 0 | 1 | Ref |
| 12873 | MW582634 | Unassigned | Unassigned | Unassigned | 409_0 | Clustered | 3 | VC_409_0 | 3 | 0.1496 | 0.98294101 | 0.147 | 0 | 0 | 0 | 1 | Ref |
| 12874 | MW584148 | Unassigned | Unassigned | Unassigned | 555_0 | Clustered | 13 | VC_555_0 | 13 | 0.7516 | 1 | 0.7516 | 0 | 0 | 0 | 1 | Ref |
| 12875 | MW584149 | Unassigned | Unassigned | Unassigned | 1092_0 | Clustered | 12 | VC_1092_0 | 12 | 0.8894 | 1 | 0.8894 | 0 | 0 | 0 | 1 | Ref |
| 12876 | MW584150 | Unassigned | Unassigned | Unassigned | 555_0 | Clustered | 13 | VC_555_0 | 13 | 0.7516 | 1 | 0.7516 | 0 | 0 | 0 | 1 | Ref |
| 12877 | MW584151 | Unassigned | Unassigned | Unassigned | 1092_0 | Clustered | 12 | VC_1092_0 | 12 | 0.8894 | 1 | 0.8894 | 0 | 0 | 0 | 1 | Ref |
| 12878 | MW584152 | Unassigned | Unassigned | Unassigned | 550_0 | Clustered | 12 | VC_550_0 | 12 | 0.802 | 1 | 0.802 | 0 | 0 | 0 | 1 | Ref |
| 12879 | MW584154 | Unassigned | Unassigned | Unassigned | 555_0 | Clustered | 13 | VC_555_0 | 13 | 0.7516 | 1 | 0.7516 | 0 | 0 | 0 | 1 | Ref |
| 12880 | MW584155 | Unassigned | Unassigned | Unassigned | 564_0 | Clustered | 3 | VC_564_0 | 3 | 0.3421 | 0.99849913 | 0.3415 | 0 | 0 | 0 | 1 | Ref |
| 12881 | MW584156 | Unassigned | Unassigned | Unassigned | 555_0 | Clustered | 13 | VC_555_0 | 13 | 0.7516 | 1 | 0.7516 | 0 | 0 | 0 | 1 | Ref |
| 12882 | MW584157 | Unassigned | Unassigned | Unassigned | 1047_0 | Clustered | 5 | VC_1047_0 | 5 | 0.6609 | 0.99999993 | 0.6609 | 0 | 0 | 0 | 1 | Ref |
| 12883 | MW584158 | Unassigned | Unassigned | Unassigned | 550_0 | Clustered | 12 | VC_550_0 | 12 | 0.802 | 1 | 0.802 | 0 | 0 | 0 | 1 | Ref |
| 12884 | MW584159 | Unassigned | Unassigned | Unassigned | 516_0 | Clustered | 3 | VC_516_0 | 3 | 0.4664 | 0.99856798 | 0.4658 | 0 | 0 | 0 | 1 | Ref |
| 12885 | MW584160 | Unassigned | Unassigned | Unassigned | 763_0 | Clustered | 2 | VC_763_0 | 2 | 0.061 | 0.95095213 | 0.058 | 0 | 0 | 0 | 1 | Ref |
| 12886 | MW584161 | Unassigned | Unassigned | Unassigned | 550_0 | Clustered | 12 | VC_550_0 | 12 | 0.802 | 1 | 0.802 | 0 | 0 | 0 | 1 | Ref |
| 12887 | MW584162 | Unassigned | Unassigned | Unassigned | 555_0 | Clustered | 13 | VC_555_0 | 13 | 0.7516 | 1 | 0.7516 | 0 | 0 | 0 | 1 | Ref |
| 12888 | MW584163 | Unassigned | Unassigned | Unassigned | 555_0 | Clustered | 13 | VC_555_0 | 13 | 0.7516 | 1 | 0.7516 | 0 | 0 | 0 | 1 | Ref |
| 12889 | MW584164 | Unassigned | Unassigned | Unassigned | 550_0 | Clustered | 12 | VC_550_0 | 12 | 0.802 | 1 | 0.802 | 0 | 0 | 0 | 1 | Ref |
| 12890 | MW584165 | Unassigned | Unassigned | Unassigned | 240_0 | Clustered | 4 | VC_240_0 | 4 | 0.7811 | 0.99998265 | 0.7811 | 0 | 0 | 0 | 1 | Ref |
| 12891 | MW584168 | Unassigned | Unassigned | Unassigned | 684_0 | Clustered | 5 | VC_684_0 | 5 | 0.3915 | 0.99999995 | 0.3915 | 0 | 0 | 0 | 1 | Ref |
| 12892 | MW584169 | Unassigned | Unassigned | Unassigned | 240_0 | Clustered | 4 | VC_240_0 | 4 | 0.7811 | 0.99998265 | 0.7811 | 0 | 0 | 0 | 1 | Ref |
| 12893 | MW584170 | Unassigned | Unassigned | Unassigned | 958_0 | Clustered | 3 | VC_958_0 | 3 | 0.1441 | 0.99848014 | 0.1438 | 0 | 0 | 0 | 1 | Ref |
| 12894 | MW584171 | Unassigned | Unassigned | Unassigned | 550_0 | Clustered | 12 | VC_550_0 | 12 | 0.802 | 1 | 0.802 | 0 | 0 | 0 | 1 | Ref |
| 12895 | MW584172 | Unassigned | Unassigned | Unassigned | 684_0 | Clustered | 5 | VC_684_0 | 5 | 0.3915 | 0.99999995 | 0.3915 | 0 | 0 | 0 | 1 | Ref |
| 12896 | MW584173 | Unassigned | Unassigned | Unassigned | 1047_0 | Clustered | 5 | VC_1047_0 | 5 | 0.6609 | 0.99999993 | 0.6609 | 0 | 0 | 0 | 1 | Ref |
| 12897 | MW584175 | Unassigned | Unassigned | Unassigned | 564_0 | Clustered | 3 | VC_564_0 | 3 | 0.3421 | 0.99849913 | 0.3415 | 0 | 0 | 0 | 1 | Ref |
| 12898 | MW584176 | Unassigned | Unassigned | Unassigned | 524_0 | Clustered | 2 | VC_524_0 | 2 | 0.113 | 0.95477567 | 0.1079 | 0 | 0 | 0 | 1 | Ref |
| 12899 | MW584178 | Unassigned | Unassigned | Unassigned | 684_0 | Clustered | 5 | VC_684_0 | 5 | 0.3915 | 0.99999995 | 0.3915 | 0 | 0 | 0 | 1 | Ref |
| 12900 | MW584179 | Unassigned | Unassigned | Unassigned | 550_0 | Clustered | 12 | VC_550_0 | 12 | 0.802 | 1 | 0.802 | 0 | 0 | 0 | 1 | Ref |
| 12901 | MW584180 | Unassigned | Unassigned | Unassigned | 550_0 | Clustered | 12 | VC_550_0 | 12 | 0.802 | 1 | 0.802 | 0 | 0 | 0 | 1 | Ref |
| 12902 | MW584181 | Unassigned | Unassigned | Unassigned | 555_0 | Clustered | 13 | VC_555_0 | 13 | 0.7516 | 1 | 0.7516 | 0 | 0 | 0 | 1 | Ref |
| 12903 | MW584182 | Unassigned | Unassigned | Unassigned | 555_0 | Clustered | 13 | VC_555_0 | 13 | 0.7516 | 1 | 0.7516 | 0 | 0 | 0 | 1 | Ref |
| 12904 | MW584184 | Unassigned | Unassigned | Unassigned | 1092_0 | Clustered | 12 | VC_1092_0 | 12 | 0.8894 | 1 | 0.8894 | 0 | 0 | 0 | 1 | Ref |
| 12905 | MW584188 | Unassigned | Unassigned | Unassigned | 524_1 | Clustered/Singleton | 1 | VC_524_1 | 1 | 0 | 1 | 0 | 0 | 0 | 0 | 1 | Ref |
| 12906 | MW584189 | Unassigned | Unassigned | Unassigned | 1047_0 | Clustered | 5 | VC_1047_0 | 5 | 0.6609 | 0.99999993 | 0.6609 | 0 | 0 | 0 | 1 | Ref |
| 12907 | MW584190 | Unassigned | Unassigned | Unassigned | 684_0 | Clustered | 5 | VC_684_0 | 5 | 0.3915 | 0.99999995 | 0.3915 | 0 | 0 | 0 | 1 | Ref |
| 12908 | MW584191 | Unassigned | Unassigned | Unassigned | 958_0 | Clustered | 3 | VC_958_0 | 3 | 0.1441 | 0.99848014 | 0.1438 | 0 | 0 | 0 | 1 | Ref |
| 12909 | MW584192 | Unassigned | Unassigned | Unassigned | 555_0 | Clustered | 13 | VC_555_0 | 13 | 0.7516 | 1 | 0.7516 | 0 | 0 | 0 | 1 | Ref |
| 12910 | MW584193 | Unassigned | Unassigned | Unassigned | 1092_0 | Clustered | 12 | VC_1092_0 | 12 | 0.8894 | 1 | 0.8894 | 0 | 0 | 0 | 1 | Ref |
| 12911 | MW584194 | Unassigned | Unassigned | Unassigned | 1092_0 | Clustered | 12 | VC_1092_0 | 12 | 0.8894 | 1 | 0.8894 | 0 | 0 | 0 | 1 | Ref |
| 12912 | MW584196 | Unassigned | Unassigned | Unassigned | 550_0 | Clustered | 12 | VC_550_0 | 12 | 0.802 | 1 | 0.802 | 0 | 0 | 0 | 1 | Ref |
| 12913 | MW584197 | Unassigned | Unassigned | Unassigned | 516_0 | Clustered | 3 | VC_516_0 | 3 | 0.4664 | 0.99856798 | 0.4658 | 0 | 0 | 0 | 1 | Ref |
| 12914 | MW584198 | Unassigned | Unassigned | Unassigned | 1092_0 | Clustered | 12 | VC_1092_0 | 12 | 0.8894 | 1 | 0.8894 | 0 | 0 | 0 | 1 | Ref |
| 12915 | MW584199 | Unassigned | Unassigned | Unassigned | 524_0 | Clustered | 2 | VC_524_0 | 2 | 0.113 | 0.95477567 | 0.1079 | 0 | 0 | 0 | 1 | Ref |
| 12916 | MW584200 | Unassigned | Unassigned | Unassigned | 1092_0 | Clustered | 12 | VC_1092_0 | 12 | 0.8894 | 1 | 0.8894 | 0 | 0 | 0 | 1 | Ref |
| 12917 | MW584201 | Unassigned | Unassigned | Unassigned | 564_0 | Clustered | 3 | VC_564_0 | 3 | 0.3421 | 0.99849913 | 0.3415 | 0 | 0 | 0 | 1 | Ref |
| 12918 | MW584202 | Unassigned | Unassigned | Unassigned | 763_0 | Clustered | 2 | VC_763_0 | 2 | 0.061 | 0.95095213 | 0.058 | 0 | 0 | 0 | 1 | Ref |
| 12919 | MW584203 | Unassigned | Unassigned | Unassigned | 555_0 | Clustered | 13 | VC_555_0 | 13 | 0.7516 | 1 | 0.7516 | 0 | 0 | 0 | 1 | Ref |
| 12920 | MW584204 | Unassigned | Unassigned | Unassigned | 550_0 | Clustered | 12 | VC_550_0 | 12 | 0.802 | 1 | 0.802 | 0 | 0 | 0 | 1 | Ref |
| 12921 | MW584205 | Unassigned | Unassigned | Unassigned | 1092_0 | Clustered | 12 | VC_1092_0 | 12 | 0.8894 | 1 | 0.8894 | 0 | 0 | 0 | 1 | Ref |
| 12922 | MW584206 | Unassigned | Unassigned | Unassigned | 1047_0 | Clustered | 5 | VC_1047_0 | 5 | 0.6609 | 0.99999993 | 0.6609 | 0 | 0 | 0 | 1 | Ref |
| 12923 | MW584207 | Unassigned | Unassigned | Unassigned | 240_0 | Clustered | 4 | VC_240_0 | 4 | 0.7811 | 0.99998265 | 0.7811 | 0 | 0 | 0 | 1 | Ref |
| 12924 | MW584208 | Unassigned | Unassigned | Unassigned | 555_0 | Clustered | 13 | VC_555_0 | 13 | 0.7516 | 1 | 0.7516 | 0 | 0 | 0 | 1 | Ref |
| 12925 | MW584209 | Unassigned | Unassigned | Unassigned | 1047_0 | Clustered | 5 | VC_1047_0 | 5 | 0.6609 | 0.99999993 | 0.6609 | 0 | 0 | 0 | 1 | Ref |
| 12926 | MW584210 | Unassigned | Unassigned | Unassigned | 1092_0 | Clustered | 12 | VC_1092_0 | 12 | 0.8894 | 1 | 0.8894 | 0 | 0 | 0 | 1 | Ref |
| 12927 | MW584212 | Unassigned | Unassigned | Unassigned | 555_0 | Clustered | 13 | VC_555_0 | 13 | 0.7516 | 1 | 0.7516 | 0 | 0 | 0 | 1 | Ref |
| 12928 | MW584228 | Unassigned | Unassigned | Unassigned | 884_1 | Clustered | 8 | VC_884_1 | 8 | 0.1102 | 0.9989323 | 0.1101 | 0 | 0 | 0 | 0.75 | Ref |
| 12929 | MW584871 | Unassigned | Unassigned | Unassigned | 600_2 | Clustered | 10 | VC_600_2 | 10 | 0.3041 | 1 | 0.3041 | 0 | 0 | 0 | 1 | Ref |
| 12930 | MW590223 | Unassigned | Unassigned | Unassigned | 1168_0 | Clustered | 2 | VC_1168_0 | 2 | 0.099 | 0.93550199 | 0.0927 | 0 | 0 | 0 | 1 | Ref |
| 12931 | MW590329 | Unassigned | Unassigned | Unassigned | 111_0 | Clustered | 91 | VC_111_0 | 91 | 0.9587 | 1 | 0.9587 | 0 | 0 | 0 | 1 | Ref |
| 12932 | MW590698 | Unassigned | Unassigned | Unassigned | 608_0 | Clustered | 5 | VC_608_0 | 5 | 0.4331 | 0.99999996 | 0.4331 | 0 | 0 | 0 | 1 | Ref |
| 12933 | MW595220 | Unassigned | Unassigned | Unassigned | 79_5 | Clustered | 2 | VC_79_5 | 2 | 0.0046 | 0.92781966 | 0.0043 | 0 | 0 | 0 | 1 | Ref |
| 12934 | MW595221 | Unassigned | Unassigned | Unassigned | 79_0 | Clustered | 64 | VC_79_0 | 64 | 0.4185 | 1 | 0.4185 | 0 | 0 | 0 | 0.7649 | Ref |
| 12935 | MW596414 | Unassigned | Unassigned | Unassigned | 884_1 | Clustered | 8 | VC_884_1 | 8 | 0.1102 | 0.9989323 | 0.1101 | 0 | 0 | 0 | 0.75 | Ref |
| 12936 | MW598258 | Unassigned | Unassigned | Unassigned | 584_0 | Clustered | 28 | VC_584_0 | 28 | 0.7704 | 1 | 0.7704 | 0 | 0 | 0 | 1 | Ref |
| 12937 | MW598459 | Unassigned | Unassigned | Unassigned | 290_0 | Clustered | 10 | VC_290_0 | 10 | 0.0981 | 1 | 0.0981 | 0 | 0 | 0 | 1 | Ref |
| 12938 | MW600722 | Unassigned | Unassigned | Unassigned | 385_0 | Clustered | 51 | VC_385_0 | 51 | 0.8754 | 1 | 0.8754 | 0 | 0 | 0 | 1 | Ref |
| 12939 | MW601216 | Unassigned | Unassigned | Unassigned | 216_5 | Clustered | 2 | VC_216_5 | 2 | 0.0094 | 0.95747812 | 0.009 | 0 | 0 | 0 | 1 | Ref |
| 12940 | MW601217 | Unassigned | Unassigned | Unassigned | 1352_0 | Clustered | 31 | VC_1352_0 | 31 | 0.849 | 1 | 0.849 | 0 | 0 | 0 | 1 | Ref |
| 12941 | MW601218 | Unassigned | Unassigned | Unassigned | 234_0 | Clustered | 21 | VC_234_0 | 21 | 0.8355 | 1 | 0.8355 | 0 | 0 | 0 | 1 | Ref |
| 12942 | MW601219 | Unassigned | Unassigned | Unassigned | 685_0 | Clustered | 16 | VC_685_0 | 16 | 0.2972 | 1 | 0.2972 | 0 | 0 | 0 | 0.9167 | Ref |
| 12943 | MW601220 | Unassigned | Unassigned | Unassigned | 1183_0 | Clustered | 6 | VC_1183_0 | 6 | 0.995 | 0.99996974 | 0.995 | 0 | 0 | 0 | 1 | Ref |
| 12944 | MW601221 | Unassigned | Unassigned | Unassigned | 1234_0 | Clustered | 44 | VC_1234_0 | 44 | 1 | 1 | 1 | 0 | 0 | 0 | 1 | Ref |
| 12945 | MW601222 | Unassigned | Unassigned | Unassigned | 237_1 | Clustered | 13 | VC_237_1 | 13 | 0.1773 | 1 | 0.1773 | 0 | 0 | 0 | 0.7308 | Ref |
| 12946 | MW601223 | Unassigned | Unassigned | Unassigned | 1075_0 | Clustered | 4 | VC_1075_0 | 4 | 0.1483 | 0.99998427 | 0.1483 | 0 | 0 | 0 | 1 | Ref |
| 12947 | MW601224 | Unassigned | Unassigned | Unassigned | 1234_0 | Clustered | 44 | VC_1234_0 | 44 | 1 | 1 | 1 | 0 | 0 | 0 | 1 | Ref |
| 12948 | MW601225 | Unassigned | Unassigned | Unassigned | 1176_0 | Clustered | 41 | VC_1176_0 | 41 | 0.9558 | 1 | 0.9558 | 0 | 0 | 0 | 0.9976 | Ref |
| 12949 | MW602648 | Unassigned | Unassigned | Unassigned | 428_0 | Clustered | 33 | VC_428_0 | 33 | 0.9915 | 1 | 0.9915 | 0 | 0 | 0 | 1 | Ref |
| 12950 | MW623430 | Unassigned | Unassigned | Unassigned | 967_0 | Clustered | 16 | VC_967_0 | 16 | 0.8971 | 1 | 0.8971 | 0 | 0 | 0 | 1 | Ref |
| 12951 | MW627293 | Unassigned | Unassigned | Unassigned | 7_0 | Clustered | 28 | VC_7_0 | 28 | 0.3916 | 1 | 0.3916 | 0 | 0 | 0 | 0.9947 | Ref |
| 12952 | MW627366 | Unassigned | Unassigned | Unassigned | 155_0 | Clustered | 14 | VC_155_0 | 14 | 0.6095 | 1 | 0.6095 | 0 | 0 | 0 | 1 | Ref |
| 12953 | MW629017 | Unassigned | Unassigned | Unassigned | 669_0 | Clustered | 21 | VC_669_0 | 21 | 0.3623 | 1 | 0.3623 | 0 | 0 | 0 | 1 | Ref |
| 12954 | MW630115 | Unassigned | Unassigned | Unassigned | 779_0 | Clustered | 37 | VC_779_0 | 37 | 0.5682 | 1 | 0.5682 | 0 | 0 | 0 | 1 | Ref |
| 12955 | MW630116 | Unassigned | Unassigned | Unassigned | 286_13 | Clustered | 10 | VC_286_13 | 10 | 0.0698 | 0.99999929 | 0.0698 | 0 | 0 | 0 | 0.9556 | Ref |
| 12956 | MW633168 | Unassigned | Unassigned | Unassigned | 91_0 | Clustered | 22 | VC_91_0 | 22 | 0.5318 | 1 | 0.5318 | 0 | 0 | 0 | 1 | Ref |
| 12957 | MW650841 | Unassigned | Unassigned | Unassigned | 7_0 | Clustered | 28 | VC_7_0 | 28 | 0.3916 | 1 | 0.3916 | 0 | 0 | 0 | 0.9947 | Ref |
| 12958 | MW650886 | Unassigned | Unassigned | Unassigned | 131_1 | Clustered | 6 | VC_131_1 | 6 | 0.057 | 0.99999999 | 0.057 | 0 | 0 | 0 | 1 | Ref |

| | | | | | | | | | | | | | | | | | |
|---|---|---|---|---|---|---|---|---|---|---|---|---|---|---|---|---|---|
| 12959 | MW651859 | Unassigned | Unassigned | Unassigned | 355_0 | Clustered | 2 | VC_355_0 | 2 | 0.0513 | 0.93418388 | 0.0479 | 0 | 0 | 0 | 1 | Ref |
| 12960 | MW654182 | Unassigned | Unassigned | Unassigned | 952_0 | Clustered | 2 | VC_952_0 | 2 | 0.9556 | 0.8861 | 0.8468 | 0 | 0 | 0 | 1 | Ref |
| 12961 | MW655991 | Unassigned | Unassigned | Unassigned | 938_0 | Clustered | 99 | VC_938_0 | 99 | 0.9119 | 1 | 0.9119 | 0 | 0 | 0 | 0.9928 | Ref |
| 12962 | MW660583 | Unassigned | Unassigned | Unassigned | 1221_0 | Clustered/Singleton | 1 | VC_1221_0 | 1 | 0 | 1 | 0 | 0 | 0 | 0 | 1 | Ref |
| 12963 | MW672037 | Unassigned | Unassigned | Unassigned | 938_0 | Clustered | 99 | VC_938_0 | 99 | 0.9119 | 1 | 0.9119 | 0 | 0 | 0 | 0.9928 | Ref |
| 12964 | MW672041 | Unassigned | Unassigned | Unassigned | 694_0 | Clustered/Singleton | 1 | VC_694_0 | 1 | 0 | 1 | 0 | 0 | 0 | 0 | 1 | Ref |
| 12965 | MW677132 | Unassigned | Unassigned | Unassigned | 108_0 | Clustered | 33 | VC_108_0 | 33 | 0.949 | 1 | 0.949 | 0 | 0 | 0 | 1 | Ref |
| 12966 | MW677509 | Unassigned | Unassigned | Unassigned | 885_0 | Clustered | 11 | VC_885_0 | 11 | 0.9774 | 1 | 0.9774 | 0 | 0 | 0 | 1 | Ref |
| 12967 | MW677510 | Unassigned | Unassigned | Unassigned | 885_0 | Clustered | 11 | VC_885_0 | 11 | 0.9774 | 1 | 0.9774 | 0 | 0 | 0 | 1 | Ref |
| 12968 | MW677511 | Unassigned | Unassigned | Unassigned | 1029_0 | Clustered | 4 | VC_1029_0 | 4 | 0.9006 | 0.99995764 | 0.9006 | 0 | 0 | 0 | 1 | Ref |
| 12969 | MW677512 | Unassigned | Unassigned | Unassigned | 885_0 | Clustered | 11 | VC_885_0 | 11 | 0.9774 | 1 | 0.9774 | 0 | 0 | 0 | 1 | Ref |
| 12970 | MW677513 | Unassigned | Unassigned | Unassigned | 923_0 | Clustered | 23 | VC_923_0 | 23 | 0.9455 | 1 | 0.9455 | 0 | 0 | 0 | 1 | Ref |
| 12971 | MW677514 | Unassigned | Unassigned | Unassigned | 749_0 | Clustered | 40 | VC_749_0 | 40 | 0.7517 | 1 | 0.7517 | 0 | 0 | 0 | 1 | Ref |
| 12972 | MW677515 | Unassigned | Unassigned | Unassigned | 885_0 | Clustered | 11 | VC_885_0 | 11 | 0.9774 | 1 | 0.9774 | 0 | 0 | 0 | 1 | Ref |
| 12973 | MW677516 | Unassigned | Unassigned | Unassigned | 923_0 | Clustered | 23 | VC_923_0 | 23 | 0.9455 | 1 | 0.9455 | 0 | 0 | 0 | 1 | Ref |
| 12974 | MW677517 | Unassigned | Unassigned | Unassigned | 885_0 | Clustered | 11 | VC_885_0 | 11 | 0.9774 | 1 | 0.9774 | 0 | 0 | 0 | 1 | Ref |
| 12975 | MW677518 | Unassigned | Unassigned | Unassigned | 923_0 | Clustered | 23 | VC_923_0 | 23 | 0.9455 | 1 | 0.9455 | 0 | 0 | 0 | 1 | Ref |
| 12976 | MW677519 | Unassigned | Unassigned | Unassigned | 923_0 | Clustered | 23 | VC_923_0 | 23 | 0.9455 | 1 | 0.9455 | 0 | 0 | 0 | 1 | Ref |
| 12977 | MW677520 | Unassigned | Unassigned | Unassigned | 885_0 | Clustered | 11 | VC_885_0 | 11 | 0.9774 | 1 | 0.9774 | 0 | 0 | 0 | 1 | Ref |
| 12978 | MW677521 | Unassigned | Unassigned | Unassigned | 923_0 | Clustered | 23 | VC_923_0 | 23 | 0.9455 | 1 | 0.9455 | 0 | 0 | 0 | 1 | Ref |
| 12979 | MW677522 | Unassigned | Unassigned | Unassigned | 923_0 | Clustered | 23 | VC_923_0 | 23 | 0.9455 | 1 | 0.9455 | 0 | 0 | 0 | 1 | Ref |
| 12980 | MW677523 | Unassigned | Unassigned | Unassigned | 923_0 | Clustered | 23 | VC_923_0 | 23 | 0.9455 | 1 | 0.9455 | 0 | 0 | 0 | 1 | Ref |
| 12981 | MW677524 | Unassigned | Unassigned | Unassigned | 923_0 | Clustered | 23 | VC_923_0 | 23 | 0.9455 | 1 | 0.9455 | 0 | 0 | 0 | 1 | Ref |
| 12982 | MW677525 | Unassigned | Unassigned | Unassigned | 471_0 | Clustered | 2 | VC_471_0 | 2 | 0.0804 | 0.95665719 | 0.0769 | 0 | 0 | 0 | 1 | Ref |
| 12983 | MW677526 | Unassigned | Unassigned | Unassigned | 749_0 | Clustered | 40 | VC_749_0 | 40 | 0.7517 | 1 | 0.7517 | 0 | 0 | 0 | 1 | Ref |
| 12984 | MW677527 | Unassigned | Unassigned | Unassigned | 1029_0 | Clustered | 4 | VC_1029_0 | 4 | 0.9006 | 0.99995764 | 0.9006 | 0 | 0 | 0 | 1 | Ref |
| 12985 | MW677528 | Unassigned | Unassigned | Unassigned | 602_0 | Clustered | 2 | VC_602_0 | 2 | 0.229 | 0.95560552 | 0.2188 | 0 | 0 | 0 | 1 | Ref |
| 12986 | MW677529 | Unassigned | Unassigned | Unassigned | 886_1 | Clustered/Singleton | 1 | VC_886_1 | 1 | 0 | 1 | 0 | 0 | 0 | 0 | 1 | Ref |
| 12987 | MW679410 | Unassigned | Unassigned | Unassigned | 290_5 | Clustered | 14 | VC_290_5 | 14 | 0.1422 | 1 | 0.1422 | 0 | 0 | 0 | 0.9231 | Ref |
| 12988 | MW689258 | Unassigned | Unassigned | Unassigned | 585_2 | Clustered/Singleton | 1 | VC_585_2 | 1 | 0 | 1 | 0 | 0 | 0 | 0 | 1 | Ref |
| 12989 | MW712717 | Unassigned | Unassigned | Unassigned | 525_0 | Clustered | 25 | VC_525_0 | 25 | 0.4597 | 1 | 0.4597 | 0 | 0 | 0 | 1 | Ref |
| 12990 | MW712718 | Unassigned | Unassigned | Unassigned | 513_0 | Clustered | 32 | VC_513_0 | 32 | 0.6881 | 1 | 0.6881 | 0 | 0 | 0 | 1 | Ref |
| 12991 | MW712719 | Unassigned | Unassigned | Unassigned | 1036_0 | Clustered | 11 | VC_1036_0 | 11 | 0.656 | 1 | 0.656 | 0 | 0 | 0 | 0.8727 | Ref |
| 12992 | MW712720 | Unassigned | Unassigned | Unassigned | 371_11 | Clustered | 31 | VC_371_11 | 31 | 0.0927 | 1 | 0.0927 | 0 | 0 | 0 | 0.7011 | Ref |
| 12993 | MW712721 | Unassigned | Unassigned | Unassigned | 1234_0 | Clustered | 44 | VC_1234_0 | 44 | 1 | 1 | 1 | 0 | 0 | 0 | 1 | Ref |
| 12994 | MW712722 | Unassigned | Unassigned | Unassigned | 236_0 | Clustered | 72 | VC_236_0 | 72 | 0.8104 | 1 | 0.8104 | 0 | 0 | 0 | 1 | Ref |
| 12995 | MW712723 | Unassigned | Unassigned | Unassigned | 236_0 | Clustered | 72 | VC_236_0 | 72 | 0.8104 | 1 | 0.8104 | 0 | 0 | 0 | 1 | Ref |
| 12996 | MW712724 | Unassigned | Unassigned | Unassigned | 1171_0 | Clustered | 21 | VC_1171_0 | 21 | 0.7125 | 1 | 0.7125 | 0 | 0 | 0 | 1 | Ref |
| 12997 | MW712726 | Unassigned | Unassigned | Unassigned | 371_15 | Clustered | 44 | VC_371_15 | 44 | 0.1391 | 1 | 0.1391 | 0 | 0 | 0 | 0.7474 | Ref |
| 12998 | MW712727 | Unassigned | Unassigned | Unassigned | 1171_0 | Clustered | 21 | VC_1171_0 | 21 | 0.7125 | 1 | 0.7125 | 0 | 0 | 0 | 1 | Ref |
| 12999 | MW712728 | Unassigned | Unassigned | Unassigned | 1171_0 | Clustered | 21 | VC_1171_0 | 21 | 0.7125 | 1 | 0.7125 | 0 | 0 | 0 | 1 | Ref |
| 13000 | MW712729 | Unassigned | Unassigned | Unassigned | 558_27 | Clustered | 2 | VC_558_27 | 2 | 0.0032 | 0.64135461 | 0.002 | 0 | 0 | 0 | 1 | Ref |
| 13001 | MW712730 | Unassigned | Unassigned | Unassigned | 236_0 | Clustered | 72 | VC_236_0 | 72 | 0.8104 | 1 | 0.8104 | 0 | 0 | 0 | 1 | Ref |
| 13002 | MW712731 | Unassigned | Unassigned | Unassigned | 508_0 | Clustered | 32 | VC_508_0 | 32 | 0.906 | 1 | 0.906 | 0 | 0 | 0 | 1 | Ref |
| 13003 | MW712732 | Unassigned | Unassigned | Unassigned | 325_23 | Clustered | 17 | VC_325_23 | 17 | 0.0716 | 1 | 0.0716 | 0 | 0 | 0 | 0.6397 | Ref |
| 13004 | MW712733 | Unassigned | Unassigned | Unassigned | 236_0 | Clustered | 72 | VC_236_0 | 72 | 0.8104 | 1 | 0.8104 | 0 | 0 | 0 | 1 | Ref |
| 13005 | MW712734 | Unassigned | Unassigned | Unassigned | 236_0 | Clustered | 72 | VC_236_0 | 72 | 0.8104 | 1 | 0.8104 | 0 | 0 | 0 | 1 | Ref |
| 13006 | MW712735 | Unassigned | Unassigned | Unassigned | 1170_1 | Clustered | 3 | VC_1170_1 | 3 | 0.308 | 0.99836447 | 0.3075 | 0 | 0 | 0 | 1 | Ref |
| 13007 | MW712736 | Unassigned | Unassigned | Unassigned | 872_0 | Clustered | 31 | VC_872_0 | 31 | 0.8151 | 1 | 0.8151 | 0 | 0 | 0 | 0.8194 | Ref |
| 13008 | MW712737 | Unassigned | Unassigned | Unassigned | 556_0 | Clustered | 21 | VC_556_0 | 21 | 0.9912 | 1 | 0.9912 | 0 | 0 | 0 | 1 | Ref |
| 13009 | MW713383 | Unassigned | Unassigned | Unassigned | 763_1 | Clustered | 9 | VC_763_1 | 9 | 0.3706 | 1 | 0.3706 | 0 | 0 | 0 | 1 | Ref |
| 13010 | MW713578 | Unassigned | Unassigned | Unassigned | 192_0 | Clustered | 15 | VC_192_0 | 15 | 0.5071 | 1 | 0.5071 | 0 | 0 | 0 | 1 | Ref |
| 13011 | MW715619 | Unassigned | Unassigned | Unassigned | 637_0 | Clustered | 115 | VC_637_0 | 115 | 0.731 | 1 | 0.731 | 0 | 0 | 0 | 0.8764 | Ref |
| 13012 | MW722080 | Unassigned | Unassigned | Unassigned | 359_0 | Clustered | 7 | VC_359_0 | 7 | 0.3703 | 1 | 0.3703 | 0 | 0 | 0 | 1 | Ref |
| 13013 | MW722081 | Unassigned | Unassigned | Unassigned | 937_0 | Clustered | 40 | VC_937_0 | 40 | 0.6809 | 1 | 0.6809 | 0 | 0 | 0 | 0.9359 | Ref |
| 13014 | MW722082 | Unassigned | Unassigned | Unassigned | 460_0 | Clustered | 19 | VC_460_0 | 19 | 0.3744 | 1 | 0.3744 | 0 | 0 | 0 | 1 | Ref |
| 13015 | MW722083 | Unassigned | Unassigned | Unassigned | 476_0 | Clustered | 19 | VC_476_0 | 19 | 0.7814 | 1 | 0.7814 | 0 | 0 | 0 | 1 | Ref |
| 13016 | MW722521 | Unassigned | Unassigned | Unassigned | 28_0 | Clustered | 2 | VC_28_0 | 2 | 0.0308 | 0.95719664 | 0.0295 | 0 | 0 | 0 | 1 | Ref |
| 13017 | MW722821 | Unassigned | Unassigned | Unassigned | 638_0 | Clustered | 45 | VC_638_0 | 45 | 0.7741 | 1 | 0.7741 | 0 | 0 | 0 | 1 | Ref |
| 13018 | MW725301 | Unassigned | Unassigned | Unassigned | 540_0 | Clustered | 15 | VC_540_0 | 15 | 0.2866 | 1 | 0.2866 | 0 | 0 | 0 | 1 | Ref |
| 13019 | MW741821 | Unassigned | Unassigned | Unassigned | 813_1 | Clustered | 24 | VC_813_1 | 24 | 0.535 | 1 | 0.535 | 0 | 0 | 0 | 1 | Ref |
| 13020 | MW748991 | Unassigned | Unassigned | Unassigned | 451_0 | Clustered | 80 | VC_451_0 | 80 | 0.9991 | 1 | 0.9991 | 0 | 0 | 0 | 1 | Ref |
| 13021 | MW748992 | Unassigned | Unassigned | Unassigned | 530_0 | Clustered | 9 | VC_530_0 | 9 | 0.8027 | 1 | 0.8027 | 0 | 0 | 0 | 1 | Ref |
| 13022 | MW748993 | Unassigned | Unassigned | Unassigned | 297_0 | Clustered | 5 | VC_297_0 | 5 | 0.1056 | 0.99999993 | 0.1056 | 0 | 0 | 0 | 0.9 | Ref |
| 13023 | MW748998 | Unassigned | Unassigned | Unassigned | 772_17 | Clustered | 2 | VC_772_17 | 2 | 0.006 | 0.57025912 | 0.0034 | 0 | 0 | 0 | 1 | Ref |
| 13024 | MW748999 | Unassigned | Unassigned | Unassigned | 590_0 | Clustered | 27 | VC_590_0 | 27 | 0.9947 | 1 | 0.9947 | 0 | 0 | 0 | 1 | Ref |
| 13025 | MW749000 | Unassigned | Unassigned | Unassigned | 283_153 | Clustered/Singleton | 1 | VC_283_153 | 1 | 0 | 1 | 0 | 0 | 0 | 0 | 1 | Ref |
| 13026 | MW749001 | Unassigned | Unassigned | Unassigned | 283_68 | Clustered/Singleton | 1 | VC_283_68 | 1 | 0 | 1 | 0 | 0 | 0 | 0 | 1 | Ref |
| 13027 | MW749002 | Unassigned | Unassigned | Unassigned | 85_2 | Clustered | 3 | VC_85_2 | 3 | 0.0334 | 0.99730296 | 0.0333 | 0 | 0 | 0 | 1 | Ref |
| 13028 | MW749003 | Unassigned | Unassigned | Unassigned | 448_0 | Clustered | 4 | VC_448_0 | 4 | 0.3059 | 0.99978853 | 0.3058 | 0 | 0 | 0 | 1 | Ref |
| 13029 | MW749004 | Unassigned | Unassigned | Unassigned | 332_0 | Clustered | 3 | VC_332_0 | 3 | 0.0411 | 0.99858754 | 0.041 | 0 | 0 | 0 | 1 | Ref |
| 13030 | MW749005 | Unassigned | Unassigned | Unassigned | 835_0 | Clustered | 67 | VC_835_0 | 67 | 0.9624 | 1 | 0.9624 | 0 | 0 | 0 | 1 | Ref |
| 13031 | MW749007 | Unassigned | Unassigned | Unassigned | 85_2 | Clustered | 3 | VC_85_2 | 3 | 0.0334 | 0.99730296 | 0.0333 | 0 | 0 | 0 | 1 | Ref |
| 13032 | MW749010 | Unassigned | Unassigned | Unassigned | 332_0 | Clustered | 3 | VC_332_0 | 3 | 0.0411 | 0.99858754 | 0.041 | 0 | 0 | 0 | 1 | Ref |
| 13033 | MW760841 | Unassigned | Unassigned | Unassigned | 959_2 | Clustered | 5 | VC_959_2 | 5 | 0.0484 | 0.89346426 | 0.0432 | 0 | 0 | 0 | 1 | Ref |
| 13034 | MW767161 | Unassigned | Unassigned | Unassigned | 669_0 | Clustered | 21 | VC_669_0 | 21 | 0.3623 | 1 | 0.3623 | 0 | 0 | 0 | 1 | Ref |
| 13035 | MW767996 | Unassigned | Unassigned | Unassigned | 386_0 | Clustered | 49 | VC_386_0 | 49 | 0.7941 | 1 | 0.7941 | 0 | 0 | 0 | 1 | Ref |
| 13036 | MW787012 | Unassigned | Unassigned | Unassigned | 883_5 | Clustered/Singleton | 1 | VC_883_5 | 1 | 0 | 1 | 0 | 0 | 0 | 0 | 1 | Ref |
| 13037 | MW790495 | Unassigned | Unassigned | Unassigned | 637_0 | Clustered | 115 | VC_637_0 | 115 | 0.731 | 1 | 0.731 | 0 | 0 | 0 | 0.8764 | Ref |
| 13038 | MW790496 | Unassigned | Unassigned | Unassigned | 28_0 | Clustered | 2 | VC_28_0 | 2 | 0.0308 | 0.95719664 | 0.0295 | 0 | 0 | 0 | 1 | Ref |
| 13039 | MW791312 | Unassigned | Unassigned | Unassigned | 689_0 | Clustered | 2 | VC_689_0 | 2 | 0.4402 | 0.95582436 | 0.4208 | 0 | 0 | 0 | 1 | Ref |
| 13040 | MW794140 | Unassigned | Unassigned | Unassigned | 495_1 | Clustered | 41 | VC_495_1 | 41 | 0.328 | 1 | 0.328 | 0 | 0 | 0 | 0.911 | Ref |
| 13041 | MW794141 | Unassigned | Unassigned | Unassigned | 495_1 | Clustered | 41 | VC_495_1 | 41 | 0.328 | 1 | 0.328 | 0 | 0 | 0 | 0.911 | Ref |
| 13042 | MW794142 | Unassigned | Unassigned | Unassigned | 495_1 | Clustered | 41 | VC_495_1 | 41 | 0.328 | 1 | 0.328 | 0 | 0 | 0 | 0.911 | Ref |
| 13043 | MW794143 | Unassigned | Unassigned | Unassigned | 495_1 | Clustered | 41 | VC_495_1 | 41 | 0.328 | 1 | 0.328 | 0 | 0 | 0 | 0.911 | Ref |
| 13044 | MW794144 | Unassigned | Unassigned | Unassigned | 495_1 | Clustered | 41 | VC_495_1 | 41 | 0.328 | 1 | 0.328 | 0 | 0 | 0 | 0.911 | Ref |
| 13045 | MW794145 | Unassigned | Unassigned | Unassigned | 495_1 | Clustered | 41 | VC_495_1 | 41 | 0.328 | 1 | 0.328 | 0 | 0 | 0 | 0.911 | Ref |
| 13046 | MW794146 | Unassigned | Unassigned | Unassigned | 495_1 | Clustered | 41 | VC_495_1 | 41 | 0.328 | 1 | 0.328 | 0 | 0 | 0 | 0.911 | Ref |
| 13047 | MW794147 | Unassigned | Unassigned | Unassigned | 495_1 | Clustered | 41 | VC_495_1 | 41 | 0.328 | 1 | 0.328 | 0 | 0 | 0 | 0.911 | Ref |
| 13048 | MW794148 | Unassigned | Unassigned | Unassigned | 495_1 | Clustered | 41 | VC_495_1 | 41 | 0.328 | 1 | 0.328 | 0 | 0 | 0 | 0.911 | Ref |
| 13049 | MW794149 | Unassigned | Unassigned | Unassigned | 495_1 | Clustered | 41 | VC_495_1 | 41 | 0.328 | 1 | 0.328 | 0 | 0 | 0 | 0.911 | Ref |
| 13050 | MW794150 | Unassigned | Unassigned | Unassigned | 495_0 | Clustered | 39 | VC_495_0 | 39 | 0.308 | 1 | 0.308 | 0 | 0 | 0 | 1 | Ref |
| 13051 | MW794151 | Unassigned | Unassigned | Unassigned | 495_0 | Clustered | 39 | VC_495_0 | 39 | 0.308 | 1 | 0.308 | 0 | 0 | 0 | 1 | Ref |
| 13052 | MW794152 | Unassigned | Unassigned | Unassigned | 495_0 | Clustered | 39 | VC_495_0 | 39 | 0.308 | 1 | 0.308 | 0 | 0 | 0 | 1 | Ref |
| 13053 | MW794153 | Unassigned | Unassigned | Unassigned | 495_0 | Clustered | 39 | VC_495_0 | 39 | 0.308 | 1 | 0.308 | 0 | 0 | 0 | 1 | Ref |
| 13054 | MW794154 | Unassigned | Unassigned | Unassigned | 495_0 | Clustered | 39 | VC_495_0 | 39 | 0.308 | 1 | 0.308 | 0 | 0 | 0 | 1 | Ref |
| 13055 | MW794155 | Unassigned | Unassigned | Unassigned | 495_0 | Clustered | 39 | VC_495_0 | 39 | 0.308 | 1 | 0.308 | 0 | 0 | 0 | 1 | Ref |
| 13056 | MW794156 | Unassigned | Unassigned | Unassigned | 495_0 | Clustered | 39 | VC_495_0 | 39 | 0.308 | 1 | 0.308 | 0 | 0 | 0 | 1 | Ref |
| 13057 | MW794157 | Unassigned | Unassigned | Unassigned | 495_0 | Clustered | 39 | VC_495_0 | 39 | 0.308 | 1 | 0.308 | 0 | 0 | 0 | 1 | Ref |
| 13058 | MW794158 | Unassigned | Unassigned | Unassigned | 495_0 | Clustered | 39 | VC_495_0 | 39 | 0.308 | 1 | 0.308 | 0 | 0 | 0 | 1 | Ref |
| 13059 | MW794159 | Unassigned | Unassigned | Unassigned | 495_0 | Clustered | 39 | VC_495_0 | 39 | 0.308 | 1 | 0.308 | 0 | 0 | 0 | 1 | Ref |
| 13060 | MW794160 | Unassigned | Unassigned | Unassigned | 495_0 | Clustered | 39 | VC_495_0 | 39 | 0.308 | 1 | 0.308 | 0 | 0 | 0 | 1 | Ref |
| 13061 | MW794161 | Unassigned | Unassigned | Unassigned | 495_0 | Clustered | 39 | VC_495_0 | 39 | 0.308 | 1 | 0.308 | 0 | 0 | 0 | 1 | Ref |
| 13062 | MW794162 | Unassigned | Unassigned | Unassigned | 495_0 | Clustered | 39 | VC_495_0 | 39 | 0.308 | 1 | 0.308 | 0 | 0 | 0 | 1 | Ref |
| 13063 | MW794163 | Unassigned | Unassigned | Unassigned | 495_0 | Clustered | 39 | VC_495_0 | 39 | 0.308 | 1 | 0.308 | 0 | 0 | 0 | 1 | Ref |
| 13064 | MW794164 | Unassigned | Unassigned | Unassigned | 495_0 | Clustered | 39 | VC_495_0 | 39 | 0.308 | 1 | 0.308 | 0 | 0 | 0 | 1 | Ref |
| 13065 | MW794165 | Unassigned | Unassigned | Unassigned | 495_0 | Clustered | 39 | VC_495_0 | 39 | 0.308 | 1 | 0.308 | 0 | 0 | 0 | 1 | Ref |
| 13066 | MW794166 | Unassigned | Unassigned | Unassigned | 495_0 | Clustered | 39 | VC_495_0 | 39 | 0.308 | 1 | 0.308 | 0 | 0 | 0 | 1 | Ref |
| 13067 | MW794167 | Unassigned | Unassigned | Unassigned | 495_0 | Clustered | 39 | VC_495_0 | 39 | 0.308 | 1 | 0.308 | 0 | 0 | 0 | 1 | Ref |
| 13068 | MW794168 | Unassigned | Unassigned | Unassigned | 495_0 | Clustered | 39 | VC_495_0 | 39 | 0.308 | 1 | 0.308 | 0 | 0 | 0 | 1 | Ref |
| 13069 | MW794169 | Unassigned | Unassigned | Unassigned | 495_0 | Clustered | 39 | VC_495_0 | 39 | 0.308 | 1 | 0.308 | 0 | 0 | 0 | 1 | Ref |
| 13070 | MW794170 | Unassigned | Unassigned | Unassigned | 495_0 | Clustered | 39 | VC_495_0 | 39 | 0.308 | 1 | 0.308 | 0 | 0 | 0 | 1 | Ref |
| 13071 | MW794171 | Unassigned | Unassigned | Unassigned | 495_0 | Clustered | 39 | VC_495_0 | 39 | 0.308 | 1 | 0.308 | 0 | 0 | 0 | 1 | Ref |
| 13072 | MW794172 | Unassigned | Unassigned | Unassigned | 495_0 | Clustered | 39 | VC_495_0 | 39 | 0.308 | 1 | 0.308 | 0 | 0 | 0 | 1 | Ref |
| 13073 | MW794173 | Unassigned | Unassigned | Unassigned | 495_0 | Clustered | 39 | VC_495_0 | 39 | 0.308 | 1 | 0.308 | 0 | 0 | 0 | 1 | Ref |
| 13074 | MW794174 | Unassigned | Unassigned | Unassigned | 495_1 | Clustered | 41 | VC_495_1 | 41 | 0.328 | 1 | 0.328 | 0 | 0 | 0 | 0.911 | Ref |
| 13075 | MW794175 | Unassigned | Unassigned | Unassigned | 495_1 | Clustered | 41 | VC_495_1 | 41 | 0.328 | 1 | 0.328 | 0 | 0 | 0 | 0.911 | Ref |
| 13076 | MW794176 | Unassigned | Unassigned | Unassigned | 495_0 | Clustered | 39 | VC_495_0 | 39 | 0.308 | 1 | 0.308 | 0 | 0 | 0 | 1 | Ref |
| 13077 | MW794177 | Unassigned | Unassigned | Unassigned | 495_0 | Clustered | 39 | VC_495_0 | 39 | 0.308 | 1 | 0.308 | 0 | 0 | 0 | 1 | Ref |
| 13078 | MW794178 | Unassigned | Unassigned | Unassigned | 495_1 | Clustered | 41 | VC_495_1 | 41 | 0.328 | 1 | 0.328 | 0 | 0 | 0 | 0.911 | Ref |
| 13079 | MW794179 | Unassigned | Unassigned | Unassigned | 495_0 | Clustered | 39 | VC_495_0 | 39 | 0.308 | 1 | 0.308 | 0 | 0 | 0 | 1 | Ref |
| 13080 | MW794180 | Unassigned | Unassigned | Unassigned | 495_0 | Clustered | 39 | VC_495_0 | 39 | 0.308 | 1 | 0.308 | 0 | 0 | 0 | 1 | Ref |
| 13081 | MW794181 | Unassigned | Unassigned | Unassigned | 495_1 | Clustered | 41 | VC_495_1 | 41 | 0.328 | 1 | 0.328 | 0 | 0 | 0 | 0.911 | Ref |
| 13082 | MW794182 | Unassigned | Unassigned | Unassigned | 495_0 | Clustered | 39 | VC_495_0 | 39 | 0.308 | 1 | 0.308 | 0 | 0 | 0 | 1 | Ref |
| 13083 | MW794183 | Unassigned | Unassigned | Unassigned | 495_0 | Clustered | 39 | VC_495_0 | 39 | 0.308 | 1 | 0.308 | 0 | 0 | 0 | 1 | Ref |
| 13084 | MW794184 | Unassigned | Unassigned | Unassigned | 495_1 | Clustered | 41 | VC_495_1 | 41 | 0.328 | 1 | 0.328 | 0 | 0 | 0 | 0.911 | Ref |
| 13085 | MW794185 | Unassigned | Unassigned | Unassigned | 495_1 | Clustered | 41 | VC_495_1 | 41 | 0.328 | 1 | 0.328 | 0 | 0 | 0 | 0.911 | Ref |
| 13086 | MW794186 | Unassigned | Unassigned | Unassigned | 495_1 | Clustered | 41 | VC_495_1 | 41 | 0.328 | 1 | 0.328 | 0 | 0 | 0 | 0.911 | Ref |
| 13087 | MW794187 | Unassigned | Unassigned | Unassigned | 495_1 | Clustered | 41 | VC_495_1 | 41 | 0.328 | 1 | 0.328 | 0 | 0 | 0 | 0.911 | Ref |
| 13088 | MW794188 | Unassigned | Unassigned | Unassigned | 495_0 | Clustered | 39 | VC_495_0 | 39 | 0.308 | 1 | 0.308 | 0 | 0 | 0 | 1 | Ref |
| 13089 | MW794189 | Unassigned | Unassigned | Unassigned | 495_1 | Clustered | 41 | VC_495_1 | 41 | 0.328 | 1 | 0.328 | 0 | 0 | 0 | 0.911 | Ref |
| 13090 | MW794190 | Unassigned | Unassigned | Unassigned | 495_0 | Clustered | 39 | VC_495_0 | 39 | 0.308 | 1 | 0.308 | 0 | 0 | 0 | 1 | Ref |
| 13091 | MW794191 | Unassigned | Unassigned | Unassigned | 495_1 | Clustered | 41 | VC_495_1 | 41 | 0.328 | 1 | 0.328 | 0 | 0 | 0 | 0.911 | Ref |
| 13092 | MW794192 | Unassigned | Unassigned | Unassigned | 495_1 | Clustered | 41 | VC_495_1 | 41 | 0.328 | 1 | 0.328 | 0 | 0 | 0 | 0.911 | Ref |
| 13093 | MW802488 | Unassigned | Unassigned | Unassigned | 116_0 | Clustered | 13 | VC_116_0 | 13 | 1 | 1 | 1 | 0 | 0 | 0 | 1 | Ref |
| 13094 | MW805361 | Unassigned | Unassigned | Unassigned | 581_0 | Clustered/Singleton | 1 | VC_581_0 | 1 | 0 | 1 | 0 | 0 | 0 | 0 | 1 | Ref |
| 13095 | MW805362 | Unassigned | Unassigned | Unassigned | 1026_0 | Clustered | 2 | VC_1026_0 | 2 | 0.2977 | 0.81445332 | 0.2425 | 0 | 0 | 0 | 1 | Ref |
| 13096 | MW812338 | Unassigned | Unassigned | Unassigned | 959_2 | Clustered | 5 | VC_959_2 | 5 | 0.0484 | 0.89346426 | 0.0432 | 0 | 0 | 0 | 1 | Ref |
| 13097 | MW812339 | Unassigned | Unassigned | Unassigned | 959_5 | Clustered | 19 | VC_959_5 | 19 | 0.2626 | 1 | 0.2626 | 0 | 0 | 0 | 1 | Ref |
| 13098 | MW812340 | Unassigned | Unassigned | Unassigned | 959_1 | Clustered | 8 | VC_959_1 | 8 | 0.0878 | 0.98435099 | 0.0864 | 0 | 0 | 0 | 1 | Ref |
| 13099 | MW812341 | Unassigned | Unassigned | Unassigned | 959_2 | Clustered | 5 | VC_959_2 | 5 | 0.0484 | 0.89346426 | 0.0432 | 0 | 0 | 0 | 1 | Ref |
| 13100 | MW812342 | Unassigned | Unassigned | Unassigned | 959_1 | Clustered | 8 | VC_959_1 | 8 | 0.0878 | 0.98435099 | 0.0864 | 0 | 0 | 0 | 1 | Ref |
| 13101 | MW812343 | Unassigned | Unassigned | Unassigned | 959_1 | Clustered | 8 | VC_959_1 | 8 | 0.0878 | 0.98435099 | 0.0864 | 0 | 0 | 0 | 1 | Ref |
| 13102 | MW812345 | Unassigned | Unassigned | Unassigned | 129_0 | Clustered | 43 | VC_129_0 | 43 | 0.886 | 1 | 0.886 | 0 | 0 | 0 | 1 | Ref |
| 13103 | MW815121 | Unassigned | Unassigned | Unassigned | 897_0 | Clustered | 6 | VC_897_0 | 6 | 0.4156 | 1 | 0.4156 | 0 | 0 | 0 | 1 | Ref |
| 13104 | MW822006 | Unassigned | Unassigned | Unassigned | 835_0 | Clustered | 67 | VC_835_0 | 67 | 0.9624 | 1 | 0.9624 | 0 | 0 | 0 | 1 | Ref |
| 13105 | MW822007 | Unassigned | Unassigned | Unassigned | 283_168 | Clustered/Singleton | 1 | VC_283_168 | 1 | 0 | 1 | 0 | 0 | 0 | 0 | 1 | Ref |
| 13106 | MW822008 | Unassigned | Unassigned | Unassigned | 164_11 | Clustered | 10 | VC_164_11 | 10 | 0.1507 | 1 | 0.1507 | 0 | 0 | 0 | 0.9111 | Ref |
| 13107 | MW822011 | Unassigned | Unassigned | Unassigned | 283_175 | Clustered/Singleton | 1 | VC_283_175 | 1 | 0 | 1 | 0 | 0 | 0 | 0 | 1 | Ref |
| 13108 | MW822143 | Unassigned | Unassigned | Unassigned | 223_0 | Clustered | 38 | VC_223_0 | 38 | 0.9095 | 1 | 0.9095 | 0 | 0 | 0 | 1 | Ref |
| 13109 | MW822144 | Unassigned | Unassigned | Unassigned | 920_0 | Clustered | 2 | VC_920_0 | 2 | 0.1671 | 0.95556296 | 0.1597 | 0 | 0 | 0 | 1 | Ref |
| 13110 | MW822145 | Unassigned | Unassigned | Unassigned | 1072_5 | Clustered | 3 | VC_1072_5 | 3 | 0.0475 | 0.98997321 | 0.047 | 0 | 0 | 0 | 1 | Ref |
| 13111 | MW822146 | Unassigned | Unassigned | Unassigned | 1093_0 | Clustered | 40 | VC_1093_0 | 40 | 0.8754 | 1 | 0.8754 | 0 | 0 | 0 | 1 | Ref |
| 13112 | MW822147 | Unassigned | Unassigned | Unassigned | 1175_0 | Clustered | 22 | VC_1175_0 | 22 | 0.4562 | 1 | 0.4562 | 0 | 0 | 0 | 1 | Ref |
| 13113 | MW822148 | Unassigned | Unassigned | Unassigned | 558_76 | Clustered/Singleton | 1 | VC_558_76 | 1 | 0 | 1 | 0 | 0 | 0 | 0 | 1 | Ref |
| 13114 | MW822149 | Unassigned | Unassigned | Unassigned | 887_0 | Clustered | 18 | VC_887_0 | 18 | 0.3663 | 1 | 0.3663 | 0 | 0 | 0 | 1 | Ref |
| 13115 | MW822535 | Unassigned | Unassigned | Unassigned | 116_0 | Clustered | 13 | VC_116_0 | 13 | 1 | 1 | 1 | 0 | 0 | 0 | 1 | Ref |
| 13116 | MW822536 | Unassigned | Unassigned | Unassigned | 116_0 | Clustered | 13 | VC_116_0 | 13 | 1 | 1 | 1 | 0 | 0 | 0 | 1 | Ref |
| 13117 | MW822537 | Unassigned | Unassigned | Unassigned | 116_0 | Clustered | 13 | VC_116_0 | 13 | 1 | 1 | 1 | 0 | 0 | 0 | 1 | Ref |
| 13118 | MW822538 | Unassigned | Unassigned | Unassigned | 116_0 | Clustered | 13 | VC_116_0 | 13 | 1 | 1 | 1 | 0 | 0 | 0 | 1 | Ref |
| 13119 | MW822539 | Unassigned | Unassigned | Unassigned | 116_0 | Clustered | 13 | VC_116_0 | 13 | 1 | 1 | 1 | 0 | 0 | 0 | 1 | Ref |
| 13120 | MW822601 | Unassigned | Unassigned | Unassigned | 482_0 | Clustered | 4 | VC_482_0 | 4 | 0.2975 | 0.99998416 | 0.2975 | 0 | 0 | 0 | 1 | Ref |

| | | | | | | | | | | | | | | | | | |
|---|---|---|---|---|---|---|---|---|---|---|---|---|---|---|---|---|---|
| 13121 | MW824369 | Unassigned | Unassigned | Unassigned | 646_0 | Clustered | 29 | VC_646_0 | 29 | 0.9131 | 1 | 0.9131 | 0 | 0 | 0 | 1 | Ref |
| 13122 | MW824370 | Unassigned | Unassigned | Unassigned | 135_1 | Clustered | 2 | VC_135_1 | 2 | 0.0299 | 0.95557637 | 0.0286 | 0 | 0 | 0 | 1 | Ref |
| 13123 | MW824371 | Unassigned | Unassigned | Unassigned | 493_12 | Clustered | 2 | VC_493_12 | 2 | 0.0328 | 0.95557924 | 0.0313 | 0 | 0 | 0 | 1 | Ref |
| 13124 | MW824372 | Unassigned | Unassigned | Unassigned | 493_12 | Clustered | 2 | VC_493_12 | 2 | 0.0328 | 0.95557924 | 0.0313 | 0 | 0 | 0 | 1 | Ref |
| 13125 | MW824373 | Unassigned | Unassigned | Unassigned | 646_0 | Clustered | 29 | VC_646_0 | 29 | 0.9131 | 1 | 0.9131 | 0 | 0 | 0 | 1 | Ref |
| 13126 | MW824374 | Unassigned | Unassigned | Unassigned | 646_0 | Clustered | 29 | VC_646_0 | 29 | 0.9131 | 1 | 0.9131 | 0 | 0 | 0 | 1 | Ref |
| 13127 | MW824375 | Unassigned | Unassigned | Unassigned | 135_1 | Clustered | 2 | VC_135_1 | 2 | 0.0299 | 0.95557637 | 0.0286 | 0 | 0 | 0 | 1 | Ref |
| 13128 | MW824376 | Unassigned | Unassigned | Unassigned | 493_13 | Clustered/Singleton | 1 | VC_493_13 | 1 | 0 | 1 | 0 | 0 | 0 | 0 | 1 | Ref |
| 13129 | MW824377 | Unassigned | Unassigned | Unassigned | 531_3 | Clustered/Singleton | 1 | VC_531_3 | 1 | 0 | 1 | 0 | 0 | 0 | 0 | 1 | Ref |
| 13130 | MW824378 | Unassigned | Unassigned | Unassigned | 646_0 | Clustered | 29 | VC_646_0 | 29 | 0.9131 | 1 | 0.9131 | 0 | 0 | 0 | 1 | Ref |
| 13131 | MW824379 | Unassigned | Unassigned | Unassigned | 646_0 | Clustered | 29 | VC_646_0 | 29 | 0.9131 | 1 | 0.9131 | 0 | 0 | 0 | 1 | Ref |
| 13132 | MW824380 | Unassigned | Unassigned | Unassigned | 499_0 | Clustered | 24 | VC_499_0 | 24 | 0.8569 | 1 | 0.8569 | 0 | 0 | 0 | 0.9529 | Ref |
| 13133 | MW824381 | Unassigned | Unassigned | Unassigned | 499_0 | Clustered | 24 | VC_499_0 | 24 | 0.8569 | 1 | 0.8569 | 0 | 0 | 0 | 0.9529 | Ref |
| 13134 | MW824382 | Unassigned | Unassigned | Unassigned | 499_0 | Clustered | 24 | VC_499_0 | 24 | 0.8569 | 1 | 0.8569 | 0 | 0 | 0 | 0.9529 | Ref |
| 13135 | MW824383 | Unassigned | Unassigned | Unassigned | 646_0 | Clustered | 29 | VC_646_0 | 29 | 0.9131 | 1 | 0.9131 | 0 | 0 | 0 | 1 | Ref |
| 13136 | MW824384 | Unassigned | Unassigned | Unassigned | 1229_0 | Clustered | 7 | VC_1229_0 | 7 | 0.9924 | 0.99999987 | 0.9924 | 0 | 0 | 0 | 1 | Ref |
| 13137 | MW824385 | Unassigned | Unassigned | Unassigned | 1229_0 | Clustered | 7 | VC_1229_0 | 7 | 0.9924 | 0.99999987 | 0.9924 | 0 | 0 | 0 | 1 | Ref |
| 13138 | MW824386 | Unassigned | Unassigned | Unassigned | 1229_0 | Clustered | 7 | VC_1229_0 | 7 | 0.9924 | 0.99999987 | 0.9924 | 0 | 0 | 0 | 1 | Ref |
| 13139 | MW824387 | Unassigned | Unassigned | Unassigned | 607_0 | Clustered | 16 | VC_607_0 | 16 | 0.9179 | 1 | 0.9179 | 0 | 0 | 0 | 1 | Ref |
| 13140 | MW824388 | Unassigned | Unassigned | Unassigned | 807_0 | Clustered | 17 | VC_807_0 | 17 | 0.907 | 1 | 0.907 | 0 | 0 | 0 | 1 | Ref |
| 13141 | MW824389 | Unassigned | Unassigned | Unassigned | 807_0 | Clustered | 17 | VC_807_0 | 17 | 0.907 | 1 | 0.907 | 0 | 0 | 0 | 1 | Ref |
| 13142 | MW824391 | Unassigned | Unassigned | Unassigned | 529_0 | Clustered | 2 | VC_529_0 | 2 | 0.5141 | 0.95025688 | 0.4886 | 0 | 0 | 0 | 1 | Ref |
| 13143 | MW824392 | Unassigned | Unassigned | Unassigned | 807_0 | Clustered | 17 | VC_807_0 | 17 | 0.907 | 1 | 0.907 | 0 | 0 | 0 | 1 | Ref |
| 13144 | MW824393 | Unassigned | Unassigned | Unassigned | 807_0 | Clustered | 17 | VC_807_0 | 17 | 0.907 | 1 | 0.907 | 0 | 0 | 0 | 1 | Ref |
| 13145 | MW824394 | Unassigned | Unassigned | Unassigned | 807_0 | Clustered | 17 | VC_807_0 | 17 | 0.907 | 1 | 0.907 | 0 | 0 | 0 | 1 | Ref |
| 13146 | MW824395 | Unassigned | Unassigned | Unassigned | 1229_0 | Clustered | 7 | VC_1229_0 | 7 | 0.9924 | 0.99999987 | 0.9924 | 0 | 0 | 0 | 1 | Ref |
| 13147 | MW824396 | Unassigned | Unassigned | Unassigned | 1229_0 | Clustered | 7 | VC_1229_0 | 7 | 0.9924 | 0.99999987 | 0.9924 | 0 | 0 | 0 | 1 | Ref |
| 13148 | MW824397 | Unassigned | Unassigned | Unassigned | 1229_0 | Clustered | 7 | VC_1229_0 | 7 | 0.9924 | 0.99999987 | 0.9924 | 0 | 0 | 0 | 1 | Ref |
| 13149 | MW824398 | Unassigned | Unassigned | Unassigned | 807_0 | Clustered | 17 | VC_807_0 | 17 | 0.907 | 1 | 0.907 | 0 | 0 | 0 | 1 | Ref |
| 13150 | MW824399 | Unassigned | Unassigned | Unassigned | 807_0 | Clustered | 17 | VC_807_0 | 17 | 0.907 | 1 | 0.907 | 0 | 0 | 0 | 1 | Ref |
| 13151 | MW824400 | Unassigned | Unassigned | Unassigned | 901_0 | Clustered | 5 | VC_901_0 | 5 | 0.9265 | 0.99999497 | 0.9265 | 0 | 0 | 0 | 1 | Ref |
| 13152 | MW824401 | Unassigned | Unassigned | Unassigned | 529_0 | Clustered | 2 | VC_529_0 | 2 | 0.5141 | 0.95025688 | 0.4886 | 0 | 0 | 0 | 1 | Ref |
| 13153 | MW824402 | Unassigned | Unassigned | Unassigned | 807_0 | Clustered | 17 | VC_807_0 | 17 | 0.907 | 1 | 0.907 | 0 | 0 | 0 | 1 | Ref |
| 13154 | MW824403 | Unassigned | Unassigned | Unassigned | 807_0 | Clustered | 17 | VC_807_0 | 17 | 0.907 | 1 | 0.907 | 0 | 0 | 0 | 1 | Ref |
| 13155 | MW824404 | Unassigned | Unassigned | Unassigned | 607_0 | Clustered | 16 | VC_607_0 | 16 | 0.9179 | 1 | 0.9179 | 0 | 0 | 0 | 1 | Ref |
| 13156 | MW824405 | Unassigned | Unassigned | Unassigned | 807_0 | Clustered | 17 | VC_807_0 | 17 | 0.907 | 1 | 0.907 | 0 | 0 | 0 | 1 | Ref |
| 13157 | MW824406 | Unassigned | Unassigned | Unassigned | 296_0 | Clustered | 6 | VC_296_0 | 6 | 0.5315 | 1 | 0.5315 | 0 | 0 | 0 | 1 | Ref |
| 13158 | MW824407 | Unassigned | Unassigned | Unassigned | 758_0 | Clustered | 18 | VC_758_0 | 18 | 0.738 | 1 | 0.738 | 0 | 0 | 0 | 1 | Ref |
| 13159 | MW824408 | Unassigned | Unassigned | Unassigned | 758_0 | Clustered | 18 | VC_758_0 | 18 | 0.738 | 1 | 0.738 | 0 | 0 | 0 | 1 | Ref |
| 13160 | MW824409 | Unassigned | Unassigned | Unassigned | 758_0 | Clustered | 18 | VC_758_0 | 18 | 0.738 | 1 | 0.738 | 0 | 0 | 0 | 1 | Ref |
| 13161 | MW824412 | Unassigned | Unassigned | Unassigned | 642_4 | Clustered | 2 | VC_642_4 | 2 | 0.0519 | 0.95604481 | 0.0496 | 0 | 0 | 0 | 1 | Ref |
| 13162 | MW824413 | Unassigned | Unassigned | Unassigned | 642_4 | Clustered | 2 | VC_642_4 | 2 | 0.0519 | 0.95604481 | 0.0496 | 0 | 0 | 0 | 1 | Ref |
| 13163 | MW824414 | Unassigned | Unassigned | Unassigned | 903_0 | Clustered | 7 | VC_903_0 | 7 | 0.7377 | 1 | 0.7377 | 0 | 0 | 0 | 1 | Ref |
| 13164 | MW824415 | Unassigned | Unassigned | Unassigned | 973_0 | Clustered | 8 | VC_973_0 | 8 | 0.472 | 1 | 0.472 | 0 | 0 | 0 | 1 | Ref |
| 13165 | MW824416 | Unassigned | Unassigned | Unassigned | 973_0 | Clustered | 8 | VC_973_0 | 8 | 0.472 | 1 | 0.472 | 0 | 0 | 0 | 1 | Ref |
| 13166 | MW824417 | Unassigned | Unassigned | Unassigned | 901_0 | Clustered | 5 | VC_901_0 | 5 | 0.9265 | 0.99999497 | 0.9265 | 0 | 0 | 0 | 1 | Ref |
| 13167 | MW824418 | Unassigned | Unassigned | Unassigned | 1050_1 | Clustered | 29 | VC_1050_1 | 29 | 0.4861 | 1 | 0.4861 | 0 | 0 | 0 | 0.9581 | Ref |
| 13168 | MW824419 | Unassigned | Unassigned | Unassigned | 973_0 | Clustered | 8 | VC_973_0 | 8 | 0.472 | 1 | 0.472 | 0 | 0 | 0 | 1 | Ref |
| 13169 | MW824420 | Unassigned | Unassigned | Unassigned | 903_0 | Clustered | 7 | VC_903_0 | 7 | 0.7377 | 1 | 0.7377 | 0 | 0 | 0 | 1 | Ref |
| 13170 | MW824421 | Unassigned | Unassigned | Unassigned | 901_0 | Clustered | 5 | VC_901_0 | 5 | 0.9265 | 0.99999497 | 0.9265 | 0 | 0 | 0 | 1 | Ref |
| 13171 | MW824422 | Unassigned | Unassigned | Unassigned | 973_0 | Clustered | 8 | VC_973_0 | 8 | 0.472 | 1 | 0.472 | 0 | 0 | 0 | 1 | Ref |
| 13172 | MW824423 | Unassigned | Unassigned | Unassigned | 973_0 | Clustered | 8 | VC_973_0 | 8 | 0.472 | 1 | 0.472 | 0 | 0 | 0 | 1 | Ref |
| 13173 | MW824424 | Unassigned | Unassigned | Unassigned | 973_0 | Clustered | 8 | VC_973_0 | 8 | 0.472 | 1 | 0.472 | 0 | 0 | 0 | 1 | Ref |
| 13174 | MW824425 | Unassigned | Unassigned | Unassigned | 973_0 | Clustered | 8 | VC_973_0 | 8 | 0.472 | 1 | 0.472 | 0 | 0 | 0 | 1 | Ref |
| 13175 | MW824426 | Unassigned | Unassigned | Unassigned | 973_0 | Clustered | 8 | VC_973_0 | 8 | 0.472 | 1 | 0.472 | 0 | 0 | 0 | 1 | Ref |
| 13176 | MW824427 | Unassigned | Unassigned | Unassigned | 903_0 | Clustered | 7 | VC_903_0 | 7 | 0.7377 | 1 | 0.7377 | 0 | 0 | 0 | 1 | Ref |
| 13177 | MW824428 | Unassigned | Unassigned | Unassigned | 903_0 | Clustered | 7 | VC_903_0 | 7 | 0.7377 | 1 | 0.7377 | 0 | 0 | 0 | 1 | Ref |
| 13178 | MW824429 | Unassigned | Unassigned | Unassigned | 903_0 | Clustered | 7 | VC_903_0 | 7 | 0.7377 | 1 | 0.7377 | 0 | 0 | 0 | 1 | Ref |
| 13179 | MW824430 | Unassigned | Unassigned | Unassigned | 1050_1 | Clustered | 29 | VC_1050_1 | 29 | 0.4861 | 1 | 0.4861 | 0 | 0 | 0 | 0.9581 | Ref |
| 13180 | MW824431 | Unassigned | Unassigned | Unassigned | 901_0 | Clustered | 5 | VC_901_0 | 5 | 0.9265 | 0.99999497 | 0.9265 | 0 | 0 | 0 | 1 | Ref |
| 13181 | MW824432 | Unassigned | Unassigned | Unassigned | 1050_1 | Clustered | 29 | VC_1050_1 | 29 | 0.4861 | 1 | 0.4861 | 0 | 0 | 0 | 0.9581 | Ref |
| 13182 | MW824433 | Unassigned | Unassigned | Unassigned | 1050_1 | Clustered | 29 | VC_1050_1 | 29 | 0.4861 | 1 | 0.4861 | 0 | 0 | 0 | 0.9581 | Ref |
| 13183 | MW824434 | Unassigned | Unassigned | Unassigned | 1050_1 | Clustered | 29 | VC_1050_1 | 29 | 0.4861 | 1 | 0.4861 | 0 | 0 | 0 | 0.9581 | Ref |
| 13184 | MW835180 | Unassigned | Unassigned | Unassigned | 465_0 | Clustered | 7 | VC_465_0 | 7 | 0.3322 | 1 | 0.3322 | 0 | 0 | 0 | 1 | Ref |
| 13185 | MW845758 | Unassigned | Unassigned | Unassigned | 959_3 | Clustered | 3 | VC_959_3 | 3 | 0.0237 | 0.75911548 | 0.018 | 0 | 0 | 0 | 1 | Ref |
| 13186 | MW845759 | Unassigned | Unassigned | Unassigned | 959_0 | Clustered | 5 | VC_959_0 | 5 | 0.0499 | 0.99999231 | 0.0499 | 0 | 0 | 0 | 1 | Ref |
| 13187 | MW845760 | Unassigned | Unassigned | Unassigned | 489_0 | Clustered | 3 | VC_489_0 | 3 | 0.0677 | 0.98259158 | 0.0666 | 0 | 0 | 0 | 1 | Ref |
| 13188 | MW862109 | Unassigned | Unassigned | Unassigned | 298_1 | Clustered/Singleton | 1 | VC_298_1 | 1 | 0 | 1 | 0 | 0 | 0 | 0 | 1 | Ref |
| 13189 | MW862804 | Unassigned | Unassigned | Unassigned | 835_0 | Clustered | 67 | VC_835_0 | 67 | 0.9624 | 1 | 0.9624 | 0 | 0 | 0 | 1 | Ref |
| 13190 | MW862979 | Unassigned | Unassigned | Unassigned | 236_0 | Clustered | 72 | VC_236_0 | 72 | 0.8104 | 1 | 0.8104 | 0 | 0 | 0 | 1 | Ref |
| 13191 | MW862980 | Unassigned | Unassigned | Unassigned | 371_11 | Clustered | 31 | VC_371_11 | 31 | 0.0927 | 1 | 0.0927 | 0 | 0 | 0 | 0.7011 | Ref |
| 13192 | MW862981 | Unassigned | Unassigned | Unassigned | 1206_0 | Clustered | 8 | VC_1206_0 | 8 | 0.1913 | 1 | 0.1913 | 0 | 0 | 0 | 1 | Ref |
| 13193 | MW862982 | Unassigned | Unassigned | Unassigned | 1073_0 | Clustered | 24 | VC_1073_0 | 24 | 0.8388 | 1 | 0.8388 | 0 | 0 | 0 | 1 | Ref |
| 13194 | MW862983 | Unassigned | Unassigned | Unassigned | 325_29 | Clustered | 3 | VC_325_29 | 3 | 0.0115 | 0.99857442 | 0.0115 | 0 | 0 | 0 | 1 | Ref |
| 13195 | MW862984 | Unassigned | Unassigned | Unassigned | 236_0 | Clustered | 72 | VC_236_0 | 72 | 0.8104 | 1 | 0.8104 | 0 | 0 | 0 | 1 | Ref |
| 13196 | MW862985 | Unassigned | Unassigned | Unassigned | 236_0 | Clustered | 72 | VC_236_0 | 72 | 0.8104 | 1 | 0.8104 | 0 | 0 | 0 | 1 | Ref |
| 13197 | MW862986 | Unassigned | Unassigned | Unassigned | 1184_0 | Clustered | 5 | VC_1184_0 | 5 | 0.6591 | 0.99999982 | 0.6591 | 0 | 0 | 0 | 1 | Ref |
| 13198 | MW862987 | Unassigned | Unassigned | Unassigned | 799_1 | Clustered | 27 | VC_799_1 | 27 | 0.8611 | 1 | 0.8611 | 0 | 0 | 0 | 1 | Ref |
| 13199 | MW862988 | Unassigned | Unassigned | Unassigned | 919_0 | Clustered | 39 | VC_919_0 | 39 | 0.9069 | 1 | 0.9069 | 0 | 0 | 0 | 1 | Ref |
| 13200 | MW862990 | Unassigned | Unassigned | Unassigned | 236_0 | Clustered | 72 | VC_236_0 | 72 | 0.8104 | 1 | 0.8104 | 0 | 0 | 0 | 1 | Ref |
| 13201 | MW862991 | Unassigned | Unassigned | Unassigned | 1360_0 | Clustered | 31 | VC_1360_0 | 31 | 0.8837 | 1 | 0.8837 | 0 | 0 | 0 | 1 | Ref |
| 13202 | MW862992 | Unassigned | Unassigned | Unassigned | 1170_2 | Clustered | 2 | VC_1170_2 | 2 | 0.0769 | 0.95418906 | 0.0734 | 0 | 0 | 0 | 1 | Ref |
| 13203 | MW862993 | Unassigned | Unassigned | Unassigned | 371_11 | Clustered | 31 | VC_371_11 | 31 | 0.0927 | 1 | 0.0927 | 0 | 0 | 0 | 0.7011 | Ref |
| 13204 | MW865291 | Unassigned | Unassigned | Unassigned | 1027_0 | Clustered | 20 | VC_1027_0 | 20 | 0.7114 | 1 | 0.7114 | 0 | 0 | 0 | 1 | Ref |
| 13205 | MW865292 | Unassigned | Unassigned | Unassigned | 175_0 | Clustered | 4 | VC_175_0 | 4 | 0.3847 | 0.99998259 | 0.3846 | 0 | 0 | 0 | 1 | Ref |
| 13206 | MW865297 | Unassigned | Unassigned | Unassigned | 758_0 | Clustered | 18 | VC_758_0 | 18 | 0.738 | 1 | 0.738 | 0 | 0 | 0 | 1 | Ref |
| 13207 | MW865299 | Unassigned | Unassigned | Unassigned | 758_0 | Clustered | 18 | VC_758_0 | 18 | 0.738 | 1 | 0.738 | 0 | 0 | 0 | 1 | Ref |
| 13208 | MW865302 | Unassigned | Unassigned | Unassigned | 758_0 | Clustered | 18 | VC_758_0 | 18 | 0.738 | 1 | 0.738 | 0 | 0 | 0 | 1 | Ref |
| 13209 | MW865307 | Unassigned | Unassigned | Unassigned | 758_0 | Clustered | 18 | VC_758_0 | 18 | 0.738 | 1 | 0.738 | 0 | 0 | 0 | 1 | Ref |
| 13210 | MW865319 | Unassigned | Unassigned | Unassigned | 903_0 | Clustered | 7 | VC_903_0 | 7 | 0.7377 | 1 | 0.7377 | 0 | 0 | 0 | 1 | Ref |
| 13211 | MW876471 | Unassigned | Unassigned | Unassigned | 451_0 | Clustered | 80 | VC_451_0 | 80 | 0.9991 | 1 | 0.9991 | 0 | 0 | 0 | 1 | Ref |
| 13212 | MW879340 | Unassigned | Unassigned | Unassigned | 489_0 | Clustered | 3 | VC_489_0 | 3 | 0.0677 | 0.98259158 | 0.0666 | 0 | 0 | 0 | 1 | Ref |
| 13213 | MW879342 | Unassigned | Unassigned | Unassigned | 164_7 | Clustered | 7 | VC_164_7 | 7 | 0.0956 | 0.99998 | 0.0956 | 0 | 0 | 0 | 1 | Ref |
| 13214 | MW879343 | Unassigned | Unassigned | Unassigned | 164_7 | Clustered | 7 | VC_164_7 | 7 | 0.0956 | 0.99998 | 0.0956 | 0 | 0 | 0 | 1 | Ref |
| 13215 | MW879344 | Unassigned | Unassigned | Unassigned | 164_7 | Clustered | 7 | VC_164_7 | 7 | 0.0956 | 0.99998 | 0.0956 | 0 | 0 | 0 | 1 | Ref |
| 13216 | MW882907 | Unassigned | Unassigned | Unassigned | 286_11 | Clustered | 3 | VC_286_11 | 3 | 0.0146 | 0.93460284 | 0.0137 | 0 | 0 | 0 | 1 | Ref |
| 13217 | MW883059 | Unassigned | Unassigned | Unassigned | 283_172 | Clustered/Singleton | 1 | VC_283_172 | 1 | 0 | 1 | 0 | 0 | 0 | 0 | 1 | Ref |
| 13218 | MW883060 | Unassigned | Unassigned | Unassigned | 392_0 | Clustered | 126 | VC_392_0 | 126 | 0.9621 | 1 | 0.9621 | 0 | 0 | 0 | 1 | Ref |
| 13219 | MW883061 | Unassigned | Unassigned | Unassigned | 772_4 | Clustered | 4 | VC_772_4 | 4 | 0.0181 | 0.73018303 | 0.0132 | 0 | 0 | 0 | 1 | Ref |
| 13220 | MW900438 | Unassigned | Unassigned | Unassigned | 1051_0 | Clustered | 2 | VC_1051_0 | 2 | 1 | 1 | 1 | 0 | 0 | 0 | 1 | Ref |
| 13221 | MW911612 | Unassigned | Unassigned | Unassigned | 495_1 | Clustered | 41 | VC_495_1 | 41 | 0.328 | 1 | 0.328 | 0 | 0 | 0 | 0.911 | Ref |
| 13222 | MW911613 | Unassigned | Unassigned | Unassigned | 495_1 | Clustered | 41 | VC_495_1 | 41 | 0.328 | 1 | 0.328 | 0 | 0 | 0 | 0.911 | Ref |
| 13223 | MW911614 | Unassigned | Unassigned | Unassigned | 495_1 | Clustered | 41 | VC_495_1 | 41 | 0.328 | 1 | 0.328 | 0 | 0 | 0 | 0.911 | Ref |
| 13224 | MW911615 | Unassigned | Unassigned | Unassigned | 495_1 | Clustered | 41 | VC_495_1 | 41 | 0.328 | 1 | 0.328 | 0 | 0 | 0 | 0.911 | Ref |
| 13225 | MW916539 | Unassigned | Unassigned | Unassigned | 830_0 | Clustered | 6 | VC_830_0 | 6 | 0.6536 | 1 | 0.6536 | 0 | 0 | 0 | 1 | Ref |
| 13226 | MW924497 | Unassigned | Unassigned | Unassigned | 8_0 | Clustered | 58 | VC_8_0 | 58 | 0.8306 | 1 | 0.8306 | 0 | 0 | 0 | 0.9946 | Ref |
| 13227 | MW924638 | Unassigned | Unassigned | Unassigned | 1171_0 | Clustered | 21 | VC_1171_0 | 21 | 0.7125 | 1 | 0.7125 | 0 | 0 | 0 | 1 | Ref |
| 13228 | MW924639 | Unassigned | Unassigned | Unassigned | 810_0 | Clustered | 18 | VC_810_0 | 18 | 0.9121 | 1 | 0.9121 | 0 | 0 | 0 | 1 | Ref |
| 13229 | MW924640 | Unassigned | Unassigned | Unassigned | 1234_0 | Clustered | 44 | VC_1234_0 | 44 | 1 | 1 | 1 | 0 | 0 | 0 | 1 | Ref |
| 13230 | MW924642 | Unassigned | Unassigned | Unassigned | 1344_0 | Clustered | 78 | VC_1344_0 | 78 | 0.9732 | 1 | 0.9732 | 0 | 0 | 0 | 1 | Ref |
| 13231 | MW924643 | Unassigned | Unassigned | Unassigned | 216_6 | Clustered | 42 | VC_216_6 | 42 | 0.473 | 1 | 0.473 | 0 | 0 | 0 | 0.8351 | Ref |
| 13232 | MW924644 | Unassigned | Unassigned | Unassigned | 509_0 | Clustered | 37 | VC_509_0 | 37 | 0.9689 | 1 | 0.9689 | 0 | 0 | 0 | 1 | Ref |
| 13233 | MW924645 | Unassigned | Unassigned | Unassigned | 1093_0 | Clustered | 40 | VC_1093_0 | 40 | 0.8754 | 1 | 0.8754 | 0 | 0 | 0 | 1 | Ref |
| 13234 | MW924646 | Unassigned | Unassigned | Unassigned | 215_1 | Clustered | 19 | VC_215_1 | 19 | 0.4188 | 1 | 0.4188 | 0 | 0 | 0 | 1 | Ref |
| 13235 | MW924647 | Unassigned | Unassigned | Unassigned | 1234_0 | Clustered | 44 | VC_1234_0 | 44 | 1 | 1 | 1 | 0 | 0 | 0 | 1 | Ref |
| 13236 | MW924648 | Unassigned | Unassigned | Unassigned | 525_0 | Clustered | 25 | VC_525_0 | 25 | 0.4597 | 1 | 0.4597 | 0 | 0 | 0 | 1 | Ref |
| 13237 | MW924649 | Unassigned | Unassigned | Unassigned | 520_31 | Clustered | 11 | VC_520_31 | 11 | 0.0897 | 1 | 0.0897 | 0 | 0 | 0 | 0.6 | Ref |
| 13238 | MW924650 | Unassigned | Unassigned | Unassigned | 525_0 | Clustered | 25 | VC_525_0 | 25 | 0.4597 | 1 | 0.4597 | 0 | 0 | 0 | 1 | Ref |
| 13239 | MW924889 | Unassigned | Unassigned | Unassigned | 10_0 | Clustered | 25 | VC_10_0 | 25 | 0.3467 | 1 | 0.3467 | 0 | 0 | 0 | 0.9733 | Ref |
| 13240 | MW926912 | Unassigned | Unassigned | Unassigned | 699_0 | Clustered | 71 | VC_699_0 | 71 | 0.9352 | 1 | 0.9352 | 0 | 0 | 0 | 1 | Ref |
| 13241 | MW927511 | Unassigned | Unassigned | Unassigned | 708_0 | Clustered | 47 | VC_708_0 | 47 | 0.8654 | 1 | 0.8654 | 0 | 0 | 0 | 1 | Ref |
| 13242 | MW927513 | Unassigned | Unassigned | Unassigned | 708_0 | Clustered | 47 | VC_708_0 | 47 | 0.8654 | 1 | 0.8654 | 0 | 0 | 0 | 1 | Ref |
| 13243 | MW927514 | Unassigned | Unassigned | Unassigned | 708_0 | Clustered | 47 | VC_708_0 | 47 | 0.8654 | 1 | 0.8654 | 0 | 0 | 0 | 1 | Ref |
| 13244 | MW927515 | Unassigned | Unassigned | Unassigned | 708_0 | Clustered | 47 | VC_708_0 | 47 | 0.8654 | 1 | 0.8654 | 0 | 0 | 0 | 1 | Ref |
| 13245 | MW927516 | Unassigned | Unassigned | Unassigned | 708_0 | Clustered | 47 | VC_708_0 | 47 | 0.8654 | 1 | 0.8654 | 0 | 0 | 0 | 1 | Ref |
| 13246 | MW927517 | Unassigned | Unassigned | Unassigned | 708_0 | Clustered | 47 | VC_708_0 | 47 | 0.8654 | 1 | 0.8654 | 0 | 0 | 0 | 1 | Ref |
| 13247 | MW927519 | Unassigned | Unassigned | Unassigned | 708_0 | Clustered | 47 | VC_708_0 | 47 | 0.8654 | 1 | 0.8654 | 0 | 0 | 0 | 1 | Ref |
| 13248 | MW927520 | Unassigned | Unassigned | Unassigned | 708_0 | Clustered | 47 | VC_708_0 | 47 | 0.8654 | 1 | 0.8654 | 0 | 0 | 0 | 1 | Ref |
| 13249 | MW927521 | Unassigned | Unassigned | Unassigned | 708_0 | Clustered | 47 | VC_708_0 | 47 | 0.8654 | 1 | 0.8654 | 0 | 0 | 0 | 1 | Ref |
| 13250 | MW927522 | Unassigned | Unassigned | Unassigned | 708_0 | Clustered | 47 | VC_708_0 | 47 | 0.8654 | 1 | 0.8654 | 0 | 0 | 0 | 1 | Ref |
| 13251 | MW927523 | Unassigned | Unassigned | Unassigned | 708_0 | Clustered | 47 | VC_708_0 | 47 | 0.8654 | 1 | 0.8654 | 0 | 0 | 0 | 1 | Ref |
| 13252 | MW927524 | Unassigned | Unassigned | Unassigned | 708_0 | Clustered | 47 | VC_708_0 | 47 | 0.8654 | 1 | 0.8654 | 0 | 0 | 0 | 1 | Ref |
| 13253 | MW927525 | Unassigned | Unassigned | Unassigned | 708_0 | Clustered | 47 | VC_708_0 | 47 | 0.8654 | 1 | 0.8654 | 0 | 0 | 0 | 1 | Ref |
| 13254 | MW929175 | Unassigned | Unassigned | Unassigned | 755_0 | Clustered | 9 | VC_755_0 | 9 | 0.4489 | 1 | 0.4489 | 0 | 0 | 0 | 1 | Ref |
| 13255 | MW929182 | Unassigned | Unassigned | Unassigned | 813_1 | Clustered | 24 | VC_813_1 | 24 | 0.535 | 1 | 0.535 | 0 | 0 | 0 | 1 | Ref |
| 13256 | MW934263 | Unassigned | Unassigned | Unassigned | 113_0 | Clustered | 9 | VC_113_0 | 9 | 0.519 | 1 | 0.519 | 0 | 0 | 0 | 1 | Ref |
| 13257 | MW960029 | Unassigned | Unassigned | Unassigned | 1403_0 | Clustered | 28 | VC_1403_0 | 28 | 0.753 | 1 | 0.753 | 0 | 0 | 0 | 1 | Ref |
| 13258 | MW960030 | Unassigned | Unassigned | Unassigned | 724_0 | Clustered | 3 | VC_724_0 | 3 | 0.1586 | 0.99815158 | 0.1583 | 0 | 0 | 0 | 1 | Ref |
| 13259 | MW960031 | Unassigned | Unassigned | Unassigned | 124_0 | Clustered | 2 | VC_124_0 | 2 | 0.3934 | 0.95337092 | 0.3751 | 0 | 0 | 0 | 1 | Ref |
| 13260 | MW960032 | Unassigned | Unassigned | Unassigned | 770_0 | Clustered | 5 | VC_770_0 | 5 | 0.8394 | 0.99999405 | 0.8394 | 0 | 0 | 0 | 1 | Ref |
| 13261 | MW960034 | Unassigned | Unassigned | Unassigned | 987_0 | Clustered | 22 | VC_987_0 | 22 | 0.8955 | 1 | 0.8955 | 0 | 0 | 0 | 1 | Ref |
| 13262 | MW960035 | Unassigned | Unassigned | Unassigned | 1403_0 | Clustered | 28 | VC_1403_0 | 28 | 0.753 | 1 | 0.753 | 0 | 0 | 0 | 1 | Ref |
| 13263 | MW960036 | Unassigned | Unassigned | Unassigned | 783_0 | Clustered | 3 | VC_783_0 | 3 | 0.0639 | 0.99854307 | 0.0638 | 0 | 0 | 0 | 1 | Ref |
| 13264 | MW960038 | Unassigned | Unassigned | Unassigned | 783_0 | Clustered | 3 | VC_783_0 | 3 | 0.0639 | 0.99854307 | 0.0638 | 0 | 0 | 0 | 1 | Ref |
| 13265 | MW978786 | Unassigned | Unassigned | Unassigned | 597_0 | Clustered | 6 | VC_597_0 | 6 | 0.1801 | 1 | 0.1801 | 0 | 0 | 0 | 1 | Ref |
| 13266 | MW980062 | Unassigned | Unassigned | Unassigned | 987_0 | Clustered | 22 | VC_987_0 | 22 | 0.8955 | 1 | 0.8955 | 0 | 0 | 0 | 1 | Ref |
| 13267 | MW980063 | Unassigned | Unassigned | Unassigned | 1416_0 | Clustered | 5 | VC_1416_0 | 5 | 0.9955 | 0.99988353 | 0.9954 | 0 | 0 | 0 | 1 | Ref |
| 13268 | MW980064 | Unassigned | Unassigned | Unassigned | 1416_0 | Clustered | 5 | VC_1416_0 | 5 | 0.9955 | 0.99988353 | 0.9954 | 0 | 0 | 0 | 1 | Ref |
| 13269 | MW980065 | Unassigned | Unassigned | Unassigned | 987_0 | Clustered | 22 | VC_987_0 | 22 | 0.8955 | 1 | 0.8955 | 0 | 0 | 0 | 1 | Ref |
| 13270 | MW980067 | Unassigned | Unassigned | Unassigned | 1403_0 | Clustered | 28 | VC_1403_0 | 28 | 0.753 | 1 | 0.753 | 0 | 0 | 0 | 1 | Ref |
| 13271 | MW980068 | Unassigned | Unassigned | Unassigned | 908_0 | Clustered | 3 | VC_908_0 | 3 | 0.5083 | 0.9972453 | 0.5069 | 0 | 0 | 0 | 1 | Ref |
| 13272 | MW980069 | Unassigned | Unassigned | Unassigned | 970_0 | Clustered | 9 | VC_970_0 | 9 | 0.9685 | 1 | 0.9685 | 0 | 0 | 0 | 1 | Ref |
| 13273 | MW980070 | Unassigned | Unassigned | Unassigned | 481_0 | Clustered | 8 | VC_481_0 | 8 | 0.1348 | 1 | 0.1348 | 0 | 0 | 0 | 1 | Ref |
| 13274 | MW980073 | Unassigned | Unassigned | Unassigned | 946_0 | Clustered | 11 | VC_946_0 | 11 | 0.7352 | 1 | 0.7352 | 0 | 0 | 0 | 1 | Ref |
| 13275 | MW980074 | Unassigned | Unassigned | Unassigned | 987_0 | Clustered | 22 | VC_987_0 | 22 | 0.8955 | 1 | 0.8955 | 0 | 0 | 0 | 1 | Ref |
| 13276 | MZ004649 | Unassigned | Unassigned | Unassigned | 540_0 | Clustered | 15 | VC_540_0 | 15 | 0.2866 | 1 | 0.2866 | 0 | 0 | 0 | 1 | Ref |
| 13277 | MZ005670 | Unassigned | Unassigned | Unassigned | 1163_0 | Clustered | 28 | VC_1163_0 | 28 | 0.4923 | 1 | 0.4923 | 0 | 0 | 0 | 1 | Ref |
| 13278 | MZ005671 | Unassigned | Unassigned | Unassigned | 572_0 | Clustered | 16 | VC_572_0 | 16 | 0.7286 | 1 | 0.7286 | 0 | 0 | 0 | 1 | Ref |
| 13279 | MZ005672 | Unassigned | Unassigned | Unassigned | 810_0 | Clustered | 18 | VC_810_0 | 18 | 0.9121 | 1 | 0.9121 | 0 | 0 | 0 | 1 | Ref |
| 13280 | MZ005673 | Unassigned | Unassigned | Unassigned | 517_1 | Clustered | 108 | VC_517_1 | 108 | 0.8483 | 1 | 0.8483 | 0 | 0 | 0 | 0.9955 | Ref |
| 13281 | MZ005674 | Unassigned | Unassigned | Unassigned | 236_0 | Clustered | 72 | VC_236_0 | 72 | 0.8104 | 1 | 0.8104 | 0 | 0 | 0 | 1 | Ref |
| 13282 | MZ005675 | Unassigned | Unassigned | Unassigned | 799_1 | Clustered | 27 | VC_799_1 | 27 | 0.8611 | 1 | 0.8611 | 0 | 0 | 0 | 1 | Ref |

| | | | | | | | | | | | | | | | | | |
|---|---|---|---|---|---|---|---|---|---|---|---|---|---|---|---|---|---|
| 13283 | MZ005676 | Unassigned | Unassigned | Unassigned | 1344_0 | Clustered | 78 | VC_1344_0 | 78 | 0.9732 | 1 | 0.9732 | 0 | 0 | 0 | 1 | Ref |
| 13284 | MZ005677 | Unassigned | Unassigned | Unassigned | 954_0 | Clustered | 13 | VC_954_0 | 13 | 0.863 | 1 | 0.863 | 0 | 0 | 0 | 1 | Ref |
| 13285 | MZ005678 | Unassigned | Unassigned | Unassigned | 1344_0 | Clustered | 78 | VC_1344_0 | 78 | 0.9732 | 1 | 0.9732 | 0 | 0 | 0 | 1 | Ref |
| 13286 | MZ005679 | Unassigned | Unassigned | Unassigned | 888_1 | Clustered | 3 | VC_888_1 | 3 | 0.0289 | 0.99848448 | 0.0289 | 0 | 0 | 0 | 1 | Ref |
| 13287 | MZ005680 | Unassigned | Unassigned | Unassigned | 1178_0 | Clustered | 24 | VC_1178_0 | 24 | 0.9524 | 1 | 0.9524 | 0 | 0 | 0 | 1 | Ref |
| 13288 | MZ005682 | Unassigned | Unassigned | Unassigned | 1207_0 | Clustered | 29 | VC_1207_0 | 29 | 0.8225 | 1 | 0.8225 | 0 | 0 | 0 | 1 | Ref |
| 13289 | MZ005683 | Unassigned | Unassigned | Unassigned | 799_1 | Clustered | 27 | VC_799_1 | 27 | 0.8611 | 1 | 0.8611 | 0 | 0 | 0 | 1 | Ref |
| 13290 | MZ005684 | Unassigned | Unassigned | Unassigned | 1240_0 | Clustered | 82 | VC_1240_0 | 82 | 0.7945 | 1 | 0.7945 | 0 | 0 | 0 | 1 | Ref |
| 13291 | MZ005685 | Unassigned | Unassigned | Unassigned | 223_0 | Clustered | 38 | VC_223_0 | 38 | 0.9095 | 1 | 0.9095 | 0 | 0 | 0 | 1 | Ref |
| 13292 | MZ020222 | Unassigned | Unassigned | Unassigned | 837_0 | Clustered | 29 | VC_837_0 | 29 | 0.9955 | 1 | 0.9955 | 0 | 0 | 0 | 1 | Ref |
| 13293 | MZ028625 | Unassigned | Unassigned | Unassigned | 224_0 | Clustered | 19 | VC_224_0 | 19 | 0.9416 | 1 | 0.9416 | 0 | 0 | 0 | 1 | Ref |
| 13294 | MZ028626 | Unassigned | Unassigned | Unassigned | 558_20 | Clustered | 17 | VC_558_20 | 17 | 0.054 | 1 | 0.054 | 0 | 0 | 0 | 0.9926 | Ref |
| 13295 | MZ028628 | Unassigned | Unassigned | Unassigned | 1344_0 | Clustered | 78 | VC_1344_0 | 78 | 0.9732 | 1 | 0.9732 | 0 | 0 | 0 | 1 | Ref |
| 13296 | MZ031013 | Unassigned | Unassigned | Unassigned | 452_0 | Clustered | 66 | VC_452_0 | 66 | 0.9248 | 1 | 0.9248 | 0 | 0 | 0 | 1 | Ref |
| 13297 | MZ042490 | Unassigned | Unassigned | Unassigned | 506_0 | Clustered | 8 | VC_506_0 | 8 | 0.7376 | 1 | 0.7376 | 0 | 0 | 0 | 1 | Ref |
| 13298 | MZ043613 | Unassigned | Unassigned | Unassigned | 701_1 | Clustered | 2 | VC_701_1 | 2 | 0.1304 | 0.95114164 | 0.124 | 0 | 0 | 0 | 1 | Ref |
| 13299 | MZ043897 | Unassigned | Unassigned | Unassigned | 30_1 | Clustered | 16 | VC_30_1 | 16 | 0.1973 | 1 | 0.1973 | 0 | 0 | 0 | 0.8333 | Ref |
| 13300 | MZ047271 | Unassigned | Unassigned | Unassigned | 1454_0 | Clustered | 2 | VC_1454_0 | 2 | 0.6937 | 0.85557782 | 0.5935 | 0 | 0 | 0 | 0 | Ref |
| 13301 | MZ065353 | Unassigned | Unassigned | Unassigned | 283_150 | Clustered/Singleton | 1 | VC_283_150 | 1 | 0 | 1 | 0 | 0 | 0 | 0 | 1 | Ref |
| 13302 | MZ078141 | Unassigned | Unassigned | Unassigned | 1174_0 | Clustered | 4 | VC_1174_0 | 4 | 1 | 1 | 1 | 0 | 0 | 0 | 1 | Ref |
| 13303 | MZ079855 | Unassigned | Unassigned | Unassigned | 192_1 | Clustered/Singleton | 1 | VC_192_1 | 1 | 0 | 1 | 0 | 0 | 0 | 0 | 1 | Ref |
| 13304 | MZ089728 | Unassigned | Unassigned | Unassigned | 297_0 | Clustered | 5 | VC_297_0 | 5 | 0.1056 | 0.99999993 | 0.1056 | 0 | 0 | 0 | 0.9 | Ref |
| 13305 | MZ089729 | Unassigned | Unassigned | Unassigned | 80_0 | Clustered | 41 | VC_80_0 | 41 | 0.5837 | 1 | 0.5837 | 0 | 0 | 0 | 0.9805 | Ref |
| 13306 | MZ089731 | Unassigned | Unassigned | Unassigned | 755_0 | Clustered | 9 | VC_755_0 | 9 | 0.4489 | 1 | 0.4489 | 0 | 0 | 0 | 1 | Ref |
| 13307 | MZ089732 | Unassigned | Unassigned | Unassigned | 472_0 | Clustered | 47 | VC_472_0 | 47 | 0.9025 | 1 | 0.9025 | 0 | 0 | 0 | 1 | Ref |
| 13308 | MZ089733 | Unassigned | Unassigned | Unassigned | 472_0 | Clustered | 47 | VC_472_0 | 47 | 0.9025 | 1 | 0.9025 | 0 | 0 | 0 | 1 | Ref |
| 13309 | MZ089734 | Unassigned | Unassigned | Unassigned | 166_0 | Clustered | 26 | VC_166_0 | 26 | 0.7923 | 1 | 0.7923 | 0 | 0 | 0 | 1 | Ref |
| 13310 | MZ089738 | Unassigned | Unassigned | Unassigned | 472_0 | Clustered | 47 | VC_472_0 | 47 | 0.9025 | 1 | 0.9025 | 0 | 0 | 0 | 1 | Ref |
| 13311 | MZ089739 | Unassigned | Unassigned | Unassigned | 472_0 | Clustered | 47 | VC_472_0 | 47 | 0.9025 | 1 | 0.9025 | 0 | 0 | 0 | 1 | Ref |
| 13312 | MZ089740 | Unassigned | Unassigned | Unassigned | 472_0 | Clustered | 47 | VC_472_0 | 47 | 0.9025 | 1 | 0.9025 | 0 | 0 | 0 | 1 | Ref |
| 13313 | MZ089741 | Unassigned | Unassigned | Unassigned | 166_0 | Clustered | 26 | VC_166_0 | 26 | 0.7923 | 1 | 0.7923 | 0 | 0 | 0 | 1 | Ref |
| 13314 | MZ089742 | Unassigned | Unassigned | Unassigned | 472_0 | Clustered | 47 | VC_472_0 | 47 | 0.9025 | 1 | 0.9025 | 0 | 0 | 0 | 1 | Ref |
| 13315 | MZ089747 | Unassigned | Unassigned | Unassigned | 1457_0 | Clustered | 2 | VC_1457_0 | 2 | 1 | 1 | 1 | 0 | 0 | 0 | 1 | Ref |
| 13316 | MZ089748 | Unassigned | Unassigned | Unassigned | 1457_0 | Clustered | 2 | VC_1457_0 | 2 | 1 | 1 | 1 | 0 | 0 | 0 | 1 | Ref |
| 13317 | MZ089752 | Unassigned | Unassigned | Unassigned | 1077_0 | Clustered | 48 | VC_1077_0 | 48 | 0.8525 | 1 | 0.8525 | 0 | 0 | 0 | 1 | Ref |
| 13318 | MZ089755 | Unassigned | Unassigned | Unassigned | 1120_0 | Clustered | 7 | VC_1120_0 | 7 | 0.2688 | 0.9951868 | 0.2675 | 0 | 0 | 0 | 1 | Ref |
| 13319 | MZ089756 | Unassigned | Unassigned | Unassigned | 1119_0 | Clustered | 20 | VC_1119_0 | 20 | 0.6751 | 0.99997892 | 0.6751 | 0 | 0 | 0 | 1 | Ref |
| 13320 | MZ089759 | Unassigned | Unassigned | Unassigned | 1120_0 | Clustered | 7 | VC_1120_0 | 7 | 0.2688 | 0.9951868 | 0.2675 | 0 | 0 | 0 | 1 | Ref |
| 13321 | MZ089761 | Unassigned | Unassigned | Unassigned | 1120_0 | Clustered | 7 | VC_1120_0 | 7 | 0.2688 | 0.9951868 | 0.2675 | 0 | 0 | 0 | 1 | Ref |
| 13322 | MZ089764 | Unassigned | Unassigned | Unassigned | 1120_0 | Clustered | 7 | VC_1120_0 | 7 | 0.2688 | 0.9951868 | 0.2675 | 0 | 0 | 0 | 1 | Ref |
| 13323 | MZ089765 | Unassigned | Unassigned | Unassigned | 1104_0 | Clustered | 154 | VC_1104_0 | 154 | 0.8484 | 1 | 0.8484 | 0 | 0 | 0 | 1 | Ref |
| 13324 | MZ089767 | Unassigned | Unassigned | Unassigned | 1104_0 | Clustered | 154 | VC_1104_0 | 154 | 0.8484 | 1 | 0.8484 | 0 | 0 | 0 | 1 | Ref |
| 13325 | MZ089768 | Unassigned | Unassigned | Unassigned | 1303_0 | Clustered | 6 | VC_1303_0 | 6 | 1 | 1 | 1 | 0 | 0 | 0 | 1 | Ref |
| 13326 | MZ089769 | Unassigned | Unassigned | Unassigned | 1458_0 | Clustered | 2 | VC_1458_0 | 2 | 1 | 1 | 1 | 0 | 0 | 0 | 1 | Ref |
| 13327 | MZ089770 | Unassigned | Unassigned | Unassigned | 1321_0 | Clustered | 14 | VC_1321_0 | 14 | 0.826 | 1 | 0.826 | 0 | 0 | 0 | 1 | Ref |
| 13328 | MZ089771 | Unassigned | Unassigned | Unassigned | 1110_0 | Clustered | 8 | VC_1110_0 | 8 | 0.6915 | 1 | 0.6915 | 0 | 0 | 0 | 1 | Ref |
| 13329 | MZ089772 | Unassigned | Unassigned | Unassigned | 1321_0 | Clustered | 14 | VC_1321_0 | 14 | 0.826 | 1 | 0.826 | 0 | 0 | 0 | 1 | Ref |
| 13330 | MZ089773 | Unassigned | Unassigned | Unassigned | 1119_0 | Clustered | 20 | VC_1119_0 | 20 | 0.6751 | 0.99997892 | 0.6751 | 0 | 0 | 0 | 1 | Ref |
| 13331 | MZ089774 | Unassigned | Unassigned | Unassigned | 1104_0 | Clustered | 154 | VC_1104_0 | 154 | 0.8484 | 1 | 0.8484 | 0 | 0 | 0 | 1 | Ref |
| 13332 | MZ089775 | Unassigned | Unassigned | Unassigned | 1104_0 | Clustered | 154 | VC_1104_0 | 154 | 0.8484 | 1 | 0.8484 | 0 | 0 | 0 | 1 | Ref |
| 13333 | MZ089777 | Unassigned | Unassigned | Unassigned | 1134_0 | Clustered | 5 | VC_1134_0 | 5 | 0.129 | 0.7637089 | 0.0985 | 0 | 0 | 0 | 1 | Ref |
| 13334 | MZ089778 | Unassigned | Unassigned | Unassigned | 1133_0 | Clustered | 4 | VC_1133_0 | 4 | 0.1253 | 0.975067 | 0.1222 | 0 | 0 | 0 | 1 | Ref |
| 13335 | MZ089780 | Unassigned | Unassigned | Unassigned | 1321_0 | Clustered | 14 | VC_1321_0 | 14 | 0.826 | 1 | 0.826 | 0 | 0 | 0 | 1 | Ref |
| 13336 | MZ089783 | Unassigned | Unassigned | Unassigned | 1459_0 | Clustered | 2 | VC_1459_0 | 2 | 1 | 1 | 1 | 0 | 0 | 0 | 1 | Ref |
| 13337 | MZ089785 | Unassigned | Unassigned | Unassigned | 1231_0 | Clustered | 2 | VC_1231_0 | 2 | 0.4148 | 0.89331315 | 0.3705 | 0 | 0 | 0 | 1 | Ref |
| 13338 | MZ089786 | Unassigned | Unassigned | Unassigned | 1243_0 | Clustered | 52 | VC_1243_0 | 52 | 0.9126 | 0.92708311 | 0.8461 | 0 | 0 | 0 | 1 | Ref |
| 13339 | MZ089787 | Unassigned | Unassigned | Unassigned | 1460_0 | Clustered | 3 | VC_1460_0 | 3 | 0.7846 | 0.96173875 | 0.7546 | 0 | 0 | 0 | 1 | Ref |
| 13340 | MZ089788 | Unassigned | Unassigned | Unassigned | 1460_0 | Clustered | 3 | VC_1460_0 | 3 | 0.7846 | 0.96173875 | 0.7546 | 0 | 0 | 0 | 1 | Ref |
| 13341 | MZ089789 | Unassigned | Unassigned | Unassigned | 1197_0 | Clustered | 15 | VC_1197_0 | 15 | 0.8991 | 0.76608394 | 0.6888 | 0 | 0 | 0 | 1 | Ref |
| 13342 | MZ089790 | Unassigned | Unassigned | Unassigned | 1461_0 | Clustered | 3 | VC_1461_0 | 3 | 1 | 1 | 1 | 0 | 0 | 0 | 1 | Ref |
| 13343 | MZ089791 | Unassigned | Unassigned | Unassigned | 1243_0 | Clustered | 52 | VC_1243_0 | 52 | 0.9126 | 0.92708311 | 0.8461 | 0 | 0 | 0 | 1 | Ref |
| 13344 | MZ089792 | Unassigned | Unassigned | Unassigned | 1461_0 | Clustered | 3 | VC_1461_0 | 3 | 1 | 1 | 1 | 0 | 0 | 0 | 1 | Ref |
| 13345 | MZ089794 | Unassigned | Unassigned | Unassigned | 1461_0 | Clustered | 3 | VC_1461_0 | 3 | 1 | 1 | 1 | 0 | 0 | 0 | 1 | Ref |
| 13346 | MZ089795 | Unassigned | Unassigned | Unassigned | 1243_0 | Clustered | 52 | VC_1243_0 | 52 | 0.9126 | 0.92708311 | 0.8461 | 0 | 0 | 0 | 1 | Ref |
| 13347 | MZ089796 | Unassigned | Unassigned | Unassigned | 1314_0 | Clustered | 4 | VC_1314_0 | 4 | 0.6744 | 0.9990299 | 0.6738 | 0 | 0 | 0 | 1 | Ref |
| 13348 | MZ089797 | Unassigned | Unassigned | Unassigned | 1314_0 | Clustered | 4 | VC_1314_0 | 4 | 0.6744 | 0.9990299 | 0.6738 | 0 | 0 | 0 | 1 | Ref |
| 13349 | MZ089798 | Unassigned | Unassigned | Unassigned | 1313_0 | Clustered | 13 | VC_1313_0 | 13 | 0.8481 | 0.87456432 | 0.7417 | 0 | 0 | 0 | 1 | Ref |
| 13350 | MZ089799 | Unassigned | Unassigned | Unassigned | 1313_0 | Clustered | 13 | VC_1313_0 | 13 | 0.8481 | 0.87456432 | 0.7417 | 0 | 0 | 0 | 1 | Ref |
| 13351 | MZ089800 | Unassigned | Unassigned | Unassigned | 1313_0 | Clustered | 13 | VC_1313_0 | 13 | 0.8481 | 0.87456432 | 0.7417 | 0 | 0 | 0 | 1 | Ref |
| 13352 | MZ089801 | Unassigned | Unassigned | Unassigned | 1313_0 | Clustered | 13 | VC_1313_0 | 13 | 0.8481 | 0.87456432 | 0.7417 | 0 | 0 | 0 | 1 | Ref |
| 13353 | MZ089802 | Unassigned | Unassigned | Unassigned | 1276_0 | Clustered | 7 | VC_1276_0 | 7 | 0.8695 | 0.99982476 | 0.8693 | 0 | 0 | 0 | 1 | Ref |
| 13354 | MZ089803 | Unassigned | Unassigned | Unassigned | 1313_0 | Clustered | 13 | VC_1313_0 | 13 | 0.8481 | 0.87456432 | 0.7417 | 0 | 0 | 0 | 1 | Ref |
| 13355 | MZ089804 | Unassigned | Unassigned | Unassigned | 1304_0 | Clustered | 7 | VC_1304_0 | 7 | 0.7931 | 0.9999998 | 0.7931 | 0 | 0 | 0 | 1 | Ref |
| 13356 | MZ089805 | Unassigned | Unassigned | Unassigned | 1313_0 | Clustered | 13 | VC_1313_0 | 13 | 0.8481 | 0.87456432 | 0.7417 | 0 | 0 | 0 | 1 | Ref |
| 13357 | MZ089806 | Unassigned | Unassigned | Unassigned | 1243_0 | Clustered | 52 | VC_1243_0 | 52 | 0.9126 | 0.92708311 | 0.8461 | 0 | 0 | 0 | 1 | Ref |
| 13358 | MZ089807 | Unassigned | Unassigned | Unassigned | 1304_0 | Clustered | 7 | VC_1304_0 | 7 | 0.7931 | 0.9999998 | 0.7931 | 0 | 0 | 0 | 1 | Ref |
| 13359 | MZ089808 | Unassigned | Unassigned | Unassigned | 1276_0 | Clustered | 7 | VC_1276_0 | 7 | 0.8695 | 0.99982476 | 0.8693 | 0 | 0 | 0 | 1 | Ref |
| 13360 | MZ089809 | Unassigned | Unassigned | Unassigned | 1276_0 | Clustered | 7 | VC_1276_0 | 7 | 0.8695 | 0.99982476 | 0.8693 | 0 | 0 | 0 | 1 | Ref |
| 13361 | MZ089811 | Unassigned | Unassigned | Unassigned | 1462_0 | Clustered | 4 | VC_1462_0 | 4 | 1 | 1 | 1 | 0 | 0 | 0 | 1 | Ref |
| 13362 | MZ089978 | Unassigned | Unassigned | Unassigned | 424_0 | Clustered | 7 | VC_424_0 | 7 | 0.9044 | 0.9998143 | 0.9043 | 0 | 0 | 0 | 1 | Ref |
| 13363 | MZ089979 | Unassigned | Unassigned | Unassigned | 424_0 | Clustered | 7 | VC_424_0 | 7 | 0.9044 | 0.9998143 | 0.9043 | 0 | 0 | 0 | 1 | Ref |
| 13364 | MZ090946 | Unassigned | Unassigned | Unassigned | 252_0 | Clustered | 121 | VC_252_0 | 121 | 0.7376 | 1 | 0.7376 | 0 | 0 | 0 | 0.9999 | Ref |
| 13365 | MZ099557 | Unassigned | Unassigned | Unassigned | 699_0 | Clustered | 71 | VC_699_0 | 71 | 0.9352 | 1 | 0.9352 | 0 | 0 | 0 | 1 | Ref |
| 13366 | MZ127825 | Unassigned | Unassigned | Unassigned | 428_0 | Clustered | 33 | VC_428_0 | 33 | 0.9915 | 1 | 0.9915 | 0 | 0 | 0 | 1 | Ref |
| 13367 | MZ147816 | Unassigned | Unassigned | Unassigned | 92_0 | Clustered | 22 | VC_92_0 | 22 | 0.5639 | 1 | 0.5639 | 0 | 0 | 0 | 1 | Ref |
| 13368 | MZ150757 | Unassigned | Unassigned | Unassigned | 283_176 | Clustered/Singleton | 1 | VC_283_176 | 1 | 0 | 1 | 0 | 0 | 0 | 0 | 1 | Ref |
| 13369 | MZ150758 | Unassigned | Unassigned | Unassigned | 329_11 | Clustered | 15 | VC_329_11 | 15 | 0.0791 | 1 | 0.0791 | 0 | 0 | 0 | 0.9333 | Ref |
| 13370 | MZ150782 | Unassigned | Unassigned | Unassigned | 1162_0 | Clustered | 10 | VC_1162_0 | 10 | 0.5715 | 1 | 0.5715 | 0 | 0 | 0 | 1 | Ref |
| 13371 | MZ150783 | Unassigned | Unassigned | Unassigned | 1100_0 | Clustered | 2 | VC_1100_0 | 2 | 0.1285 | 0.94673154 | 0.1217 | 0 | 0 | 0 | 1 | Ref |
| 13372 | MZ150784 | Unassigned | Unassigned | Unassigned | 1037_0 | Clustered | 6 | VC_1037_0 | 6 | 0.6798 | 1 | 0.6798 | 0 | 0 | 0 | 1 | Ref |
| 13373 | MZ150785 | Unassigned | Unassigned | Unassigned | 1176_0 | Clustered | 41 | VC_1176_0 | 41 | 0.9558 | 1 | 0.9558 | 0 | 0 | 0 | 0.9976 | Ref |
| 13374 | MZ150786 | Unassigned | Unassigned | Unassigned | 372_4 | Clustered | 3 | VC_372_4 | 3 | 0.033 | 0.99853257 | 0.0329 | 0 | 0 | 0 | 1 | Ref |
| 13375 | MZ150788 | Unassigned | Unassigned | Unassigned | 1175_0 | Clustered | 22 | VC_1175_0 | 22 | 0.4562 | 1 | 0.4562 | 0 | 0 | 0 | 1 | Ref |
| 13376 | MZ150790 | Unassigned | Unassigned | Unassigned | 1440_0 | Clustered | 4 | VC_1440_0 | 4 | 1 | 1 | 1 | 0 | 0 | 0 | 1 | Ref |
| 13377 | MZ150791 | Unassigned | Unassigned | Unassigned | 1178_0 | Clustered | 24 | VC_1178_0 | 24 | 0.9524 | 1 | 0.9524 | 0 | 0 | 0 | 1 | Ref |
| 13378 | MZ150792 | Unassigned | Unassigned | Unassigned | 1352_0 | Clustered | 31 | VC_1352_0 | 31 | 0.849 | 1 | 0.849 | 0 | 0 | 0 | 1 | Ref |
| 13379 | MZ150793 | Unassigned | Unassigned | Unassigned | 919_0 | Clustered | 39 | VC_919_0 | 39 | 0.9069 | 1 | 0.9069 | 0 | 0 | 0 | 1 | Ref |
| 13380 | MZ152914 | Unassigned | Unassigned | Unassigned | 574_0 | Clustered | 7 | VC_574_0 | 7 | 0.2734 | 1 | 0.2734 | 0 | 0 | 0 | 1 | Ref |
| 13381 | MZ156567 | Unassigned | Unassigned | Unassigned | 637_0 | Clustered | 115 | VC_637_0 | 115 | 0.731 | 1 | 0.731 | 0 | 0 | 0 | 0.8764 | Ref |
| 13382 | MZ156568 | Unassigned | Unassigned | Unassigned | 637_0 | Clustered | 115 | VC_637_0 | 115 | 0.731 | 1 | 0.731 | 0 | 0 | 0 | 0.8764 | Ref |
| 13383 | MZ156569 | Unassigned | Unassigned | Unassigned | 637_0 | Clustered | 115 | VC_637_0 | 115 | 0.731 | 1 | 0.731 | 0 | 0 | 0 | 0.8764 | Ref |
| 13384 | MZ156570 | Unassigned | Unassigned | Unassigned | 133_35 | Clustered/Singleton | 1 | VC_133_35 | 1 | 0 | 1 | 0 | 0 | 0 | 0 | 1 | Ref |
| 13385 | MZ170040 | Unassigned | Unassigned | Unassigned | 283_75 | Clustered/Singleton | 1 | VC_283_75 | 1 | 0 | 1 | 0 | 0 | 0 | 0 | 1 | Ref |
| 13386 | MZ170041 | Unassigned | Unassigned | Unassigned | 283_77 | Clustered | 2 | VC_283_77 | 2 | 0.0025 | 0.95407419 | 0.0024 | 0 | 0 | 0 | 1 | Ref |
| 13387 | MZ171370 | Unassigned | Unassigned | Unassigned | 799_1 | Clustered | 27 | VC_799_1 | 27 | 0.8611 | 1 | 0.8611 | 0 | 0 | 0 | 1 | Ref |
| 13388 | MZ171371 | Unassigned | Unassigned | Unassigned | 1175_0 | Clustered | 22 | VC_1175_0 | 22 | 0.4562 | 1 | 0.4562 | 0 | 0 | 0 | 1 | Ref |
| 13389 | MZ171372 | Unassigned | Unassigned | Unassigned | 570_0 | Clustered | 9 | VC_570_0 | 9 | 0.499 | 1 | 0.499 | 0 | 0 | 0 | 1 | Ref |
| 13390 | MZ171373 | Unassigned | Unassigned | Unassigned | 888_0 | Clustered | 28 | VC_888_0 | 28 | 0.5497 | 1 | 0.5497 | 0 | 0 | 0 | 1 | Ref |
| 13391 | MZ171374 | Unassigned | Unassigned | Unassigned | 525_0 | Clustered | 25 | VC_525_0 | 25 | 0.4597 | 1 | 0.4597 | 0 | 0 | 0 | 1 | Ref |
| 13392 | MZ171375 | Unassigned | Unassigned | Unassigned | 509_0 | Clustered | 37 | VC_509_0 | 37 | 0.9689 | 1 | 0.9689 | 0 | 0 | 0 | 1 | Ref |
| 13393 | MZ171376 | Unassigned | Unassigned | Unassigned | 223_0 | Clustered | 38 | VC_223_0 | 38 | 0.9095 | 1 | 0.9095 | 0 | 0 | 0 | 1 | Ref |
| 13394 | MZ171377 | Unassigned | Unassigned | Unassigned | 761_0 | Clustered | 67 | VC_761_0 | 67 | 0.8947 | 1 | 0.8947 | 0 | 0 | 0 | 1 | Ref |
| 13395 | MZ171378 | Unassigned | Unassigned | Unassigned | 520_48 | Clustered/Singleton | 1 | VC_520_48 | 1 | 0 | 1 | 0 | 0 | 0 | 0 | 1 | Ref |
| 13396 | MZ171380 | Unassigned | Unassigned | Unassigned | 517_1 | Clustered | 108 | VC_517_1 | 108 | 0.8483 | 1 | 0.8483 | 0 | 0 | 0 | 0.9955 | Ref |
| 13397 | MZ171381 | Unassigned | Unassigned | Unassigned | 509_0 | Clustered | 37 | VC_509_0 | 37 | 0.9689 | 1 | 0.9689 | 0 | 0 | 0 | 1 | Ref |
| 13398 | MZ172460 | Unassigned | Unassigned | Unassigned | 938_0 | Clustered | 99 | VC_938_0 | 99 | 0.9119 | 1 | 0.9119 | 0 | 0 | 0 | 0.9928 | Ref |
| 13399 | MZ182246 | Unassigned | Unassigned | Unassigned | 695_0 | Clustered | 48 | VC_695_0 | 48 | 0.8162 | 1 | 0.8162 | 0 | 0 | 0 | 1 | Ref |
| 13400 | MZ182247 | Unassigned | Unassigned | Unassigned | 837_0 | Clustered | 29 | VC_837_0 | 29 | 0.9955 | 1 | 0.9955 | 0 | 0 | 0 | 1 | Ref |
| 13401 | MZ188976 | Unassigned | Unassigned | Unassigned | 247_0 | Clustered | 14 | VC_247_0 | 14 | 0.9119 | 1 | 0.9119 | 0 | 0 | 0 | 0.967 | Ref |
| 13402 | MZ189261 | Unassigned | Unassigned | Unassigned | 286_12 | Clustered | 3 | VC_286_12 | 3 | 0.0146 | 0.92328265 | 0.0135 | 0 | 0 | 0 | 1 | Ref |
| 13403 | MZ189262 | Unassigned | Unassigned | Unassigned | 283_166 | Clustered/Singleton | 1 | VC_283_166 | 1 | 0 | 1 | 0 | 0 | 0 | 0 | 1 | Ref |
| 13404 | MZ208805 | Unassigned | Unassigned | Unassigned | 111_0 | Clustered | 91 | VC_111_0 | 91 | 0.9587 | 1 | 0.9587 | 0 | 0 | 0 | 1 | Ref |
| 13405 | MZ209299 | Unassigned | Unassigned | Unassigned | 520_91 | Clustered/Singleton | 1 | VC_520_91 | 1 | 0 | 1 | 0 | 0 | 0 | 0 | 1 | Ref |
| 13406 | MZ209300 | Unassigned | Unassigned | Unassigned | 802_0 | Clustered | 17 | VC_802_0 | 17 | 0.8025 | 1 | 0.8025 | 0 | 0 | 0 | 1 | Ref |
| 13407 | MZ209301 | Unassigned | Unassigned | Unassigned | 799_1 | Clustered | 27 | VC_799_1 | 27 | 0.8611 | 1 | 0.8611 | 0 | 0 | 0 | 1 | Ref |
| 13408 | MZ209302 | Unassigned | Unassigned | Unassigned | 1180_0 | Clustered | 3 | VC_1180_0 | 3 | 0.5161 | 0.9981786 | 0.5151 | 0 | 0 | 0 | 1 | Ref |
| 13409 | MZ209303 | Unassigned | Unassigned | Unassigned | 1164_0 | Clustered | 25 | VC_1164_0 | 25 | 0.7312 | 1 | 0.7312 | 0 | 0 | 0 | 1 | Ref |
| 13410 | MZ209304 | Unassigned | Unassigned | Unassigned | 1344_0 | Clustered | 78 | VC_1344_0 | 78 | 0.9732 | 1 | 0.9732 | 0 | 0 | 0 | 1 | Ref |
| 13411 | MZ209305 | Unassigned | Unassigned | Unassigned | 216_5 | Clustered | 2 | VC_216_5 | 2 | 0.0094 | 0.95747812 | 0.009 | 0 | 0 | 0 | 1 | Ref |
| 13412 | MZ209306 | Unassigned | Unassigned | Unassigned | 556_0 | Clustered | 21 | VC_556_0 | 21 | 0.9912 | 1 | 0.9912 | 0 | 0 | 0 | 1 | Ref |
| 13413 | MZ209307 | Unassigned | Unassigned | Unassigned | 802_0 | Clustered | 17 | VC_802_0 | 17 | 0.8025 | 1 | 0.8025 | 0 | 0 | 0 | 1 | Ref |
| 13414 | MZ220766 | Unassigned | Unassigned | Unassigned | 392_0 | Clustered | 126 | VC_392_0 | 126 | 0.9621 | 1 | 0.9621 | 0 | 0 | 0 | 1 | Ref |
| 13415 | MZ221764 | Unassigned | Unassigned | Unassigned | 938_0 | Clustered | 99 | VC_938_0 | 99 | 0.9119 | 1 | 0.9119 | 0 | 0 | 0 | 0.9928 | Ref |
| 13416 | MZ223858 | Unassigned | Unassigned | Unassigned | 451_0 | Clustered | 80 | VC_451_0 | 80 | 0.9991 | 1 | 0.9991 | 0 | 0 | 0 | 1 | Ref |
| 13417 | MZ230992 | Unassigned | Unassigned | Unassigned | 1359_0 | Clustered | 11 | VC_1359_0 | 11 | 1 | 0 | 0 | 0 | 0 | 0 | 1 | Ref |
| 13418 | MZ230993 | Unassigned | Unassigned | Unassigned | 1359_0 | Clustered | 11 | VC_1359_0 | 11 | 1 | 0 | 0 | 0 | 0 | 0 | 1 | Ref |
| 13419 | MZ230994 | Unassigned | Unassigned | Unassigned | 1359_0 | Clustered | 11 | VC_1359_0 | 11 | 1 | 0 | 0 | 0 | 0 | 0 | 1 | Ref |
| 13420 | MZ230995 | Unassigned | Unassigned | Unassigned | 1359_0 | Clustered | 11 | VC_1359_0 | 11 | 1 | 0 | 0 | 0 | 0 | 0 | 1 | Ref |
| 13421 | MZ230996 | Unassigned | Unassigned | Unassigned | 1359_0 | Clustered | 11 | VC_1359_0 | 11 | 1 | 0 | 0 | 0 | 0 | 0 | 1 | Ref |
| 13422 | MZ230997 | Unassigned | Unassigned | Unassigned | 1359_0 | Clustered | 11 | VC_1359_0 | 11 | 1 | 0 | 0 | 0 | 0 | 0 | 1 | Ref |
| 13423 | MZ230998 | Unassigned | Unassigned | Unassigned | 1359_0 | Clustered | 11 | VC_1359_0 | 11 | 1 | 0 | 0 | 0 | 0 | 0 | 1 | Ref |
| 13424 | MZ230999 | Unassigned | Unassigned | Unassigned | 1359_0 | Clustered | 11 | VC_1359_0 | 11 | 1 | 0 | 0 | 0 | 0 | 0 | 1 | Ref |
| 13425 | MZ231000 | Unassigned | Unassigned | Unassigned | 1359_0 | Clustered | 11 | VC_1359_0 | 11 | 1 | 0 | 0 | 0 | 0 | 0 | 1 | Ref |
| 13426 | MZ231001 | Unassigned | Unassigned | Unassigned | 1359_0 | Clustered | 11 | VC_1359_0 | 11 | 1 | 0 | 0 | 0 | 0 | 0 | 1 | Ref |
| 13427 | MZ234008 | Unassigned | Unassigned | Unassigned | 835_0 | Clustered | 67 | VC_835_0 | 67 | 0.9624 | 1 | 0.9624 | 0 | 0 | 0 | 1 | Ref |
| 13428 | MZ234009 | Unassigned | Unassigned | Unassigned | 451_0 | Clustered | 80 | VC_451_0 | 80 | 0.9991 | 1 | 0.9991 | 0 | 0 | 0 | 1 | Ref |
| 13429 | MZ234010 | Unassigned | Unassigned | Unassigned | 638_0 | Clustered | 45 | VC_638_0 | 45 | 0.7741 | 1 | 0.7741 | 0 | 0 | 0 | 1 | Ref |
| 13430 | MZ234011 | Unassigned | Unassigned | Unassigned | 451_0 | Clustered | 80 | VC_451_0 | 80 | 0.9991 | 1 | 0.9991 | 0 | 0 | 0 | 1 | Ref |
| 13431 | MZ234012 | Unassigned | Unassigned | Unassigned | 452_0 | Clustered | 66 | VC_452_0 | 66 | 0.9248 | 1 | 0.9248 | 0 | 0 | 0 | 1 | Ref |
| 13432 | MZ234013 | Unassigned | Unassigned | Unassigned | 283_105 | Clustered/Singleton | 1 | VC_283_105 | 1 | 0 | 1 | 0 | 0 | 0 | 0 | 1 | Ref |
| 13433 | MZ234014 | Unassigned | Unassigned | Unassigned | 835_0 | Clustered | 67 | VC_835_0 | 67 | 0.9624 | 1 | 0.9624 | 0 | 0 | 0 | 1 | Ref |
| 13434 | MZ234015 | Unassigned | Unassigned | Unassigned | 813_0 | Clustered | 12 | VC_813_0 | 12 | 0.2347 | 1 | 0.2347 | 0 | 0 | 0 | 1 | Ref |
| 13435 | MZ234016 | Unassigned | Unassigned | Unassigned | 835_0 | Clustered | 67 | VC_835_0 | 67 | 0.9624 | 1 | 0.9624 | 0 | 0 | 0 | 1 | Ref |
| 13436 | MZ234017 | Unassigned | Unassigned | Unassigned | 813_0 | Clustered | 12 | VC_813_0 | 12 | 0.2347 | 1 | 0.2347 | 0 | 0 | 0 | 1 | Ref |
| 13437 | MZ234018 | Unassigned | Unassigned | Unassigned | 283_40 | Clustered/Singleton | 1 | VC_283_40 | 1 | 0 | 1 | 0 | 0 | 0 | 0 | 1 | Ref |
| 13438 | MZ234019 | Unassigned | Unassigned | Unassigned | 452_0 | Clustered | 66 | VC_452_0 | 66 | 0.9248 | 1 | 0.9248 | 0 | 0 | 0 | 1 | Ref |
| 13439 | MZ234020 | Unassigned | Unassigned | Unassigned | 452_0 | Clustered | 66 | VC_452_0 | 66 | 0.9248 | 1 | 0.9248 | 0 | 0 | 0 | 1 | Ref |
| 13440 | MZ234021 | Unassigned | Unassigned | Unassigned | 451_0 | Clustered | 80 | VC_451_0 | 80 | 0.9991 | 1 | 0.9991 | 0 | 0 | 0 | 1 | Ref |
| 13441 | MZ234022 | Unassigned | Unassigned | Unassigned | 451_0 | Clustered | 80 | VC_451_0 | 80 | 0.9991 | 1 | 0.9991 | 0 | 0 | 0 | 1 | Ref |
| 13442 | MZ234023 | Unassigned | Unassigned | Unassigned | 451_0 | Clustered | 80 | VC_451_0 | 80 | 0.9991 | 1 | 0.9991 | 0 | 0 | 0 | 1 | Ref |
| 13443 | MZ234024 | Unassigned | Unassigned | Unassigned | 451_0 | Clustered | 80 | VC_451_0 | 80 | 0.9991 | 1 | 0.9991 | 0 | 0 | 0 | 1 | Ref |
| 13444 | MZ234025 | Unassigned | Unassigned | Unassigned | 286_1 | Clustered | 21 | VC_286_1 | 21 | 0.1696 | 1 | 0.1696 | 0 | 0 | 0 | 0.9048 | Ref |

| | | | | | | | | | | | | | | | | | |
|---|---|---|---|---|---|---|---|---|---|---|---|---|---|---|---|---|---|
| 13445 | MZ234026 | Unassigned | Unassigned | Unassigned | 835_0 | Clustered | 67 | VC_835_0 | 67 | 0.9624 | 1 | 0.9624 | 0 | 0 | 0 | 1 | Ref |
| 13446 | MZ234027 | Unassigned | Unassigned | Unassigned | 283_78 | Clustered | 13 | VC_283_78 | 13 | 0.035 | 1 | 0.035 | 0 | 0 | 0 | 0.7949 | Ref |
| 13447 | MZ234028 | Unassigned | Unassigned | Unassigned | 813_1 | Clustered | 24 | VC_813_1 | 24 | 0.535 | 1 | 0.535 | 0 | 0 | 0 | 1 | Ref |
| 13448 | MZ234029 | Unassigned | Unassigned | Unassigned | 835_0 | Clustered | 67 | VC_835_0 | 67 | 0.9624 | 1 | 0.9624 | 0 | 0 | 0 | 1 | Ref |
| 13449 | MZ234030 | Unassigned | Unassigned | Unassigned | 283_78 | Clustered | 13 | VC_283_78 | 13 | 0.035 | 1 | 0.035 | 0 | 0 | 0 | 0.7949 | Ref |
| 13450 | MZ234031 | Unassigned | Unassigned | Unassigned | 283_117 | Clustered | 3 | VC_283_117 | 3 | 0.0053 | 0.99752037 | 0.0053 | 0 | 0 | 0 | 1 | Ref |
| 13451 | MZ234032 | Unassigned | Unassigned | Unassigned | 452_0 | Clustered | 66 | VC_452_0 | 66 | 0.9248 | 1 | 0.9248 | 0 | 0 | 0 | 1 | Ref |
| 13452 | MZ234033 | Unassigned | Unassigned | Unassigned | 942_0 | Clustered | 36 | VC_942_0 | 36 | 0.7848 | 1 | 0.7848 | 0 | 0 | 0 | 1 | Ref |
| 13453 | MZ234034 | Unassigned | Unassigned | Unassigned | 283_78 | Clustered | 13 | VC_283_78 | 13 | 0.035 | 1 | 0.035 | 0 | 0 | 0 | 0.7949 | Ref |
| 13454 | MZ234035 | Unassigned | Unassigned | Unassigned | 283_117 | Clustered | 3 | VC_283_117 | 3 | 0.0053 | 0.99752037 | 0.0053 | 0 | 0 | 0 | 1 | Ref |
| 13455 | MZ234036 | Unassigned | Unassigned | Unassigned | 283_78 | Clustered | 13 | VC_283_78 | 13 | 0.035 | 1 | 0.035 | 0 | 0 | 0 | 0.7949 | Ref |
| 13456 | MZ234037 | Unassigned | Unassigned | Unassigned | 283_78 | Clustered | 13 | VC_283_78 | 13 | 0.035 | 1 | 0.035 | 0 | 0 | 0 | 0.7949 | Ref |
| 13457 | MZ234038 | Unassigned | Unassigned | Unassigned | 283_117 | Clustered | 3 | VC_283_117 | 3 | 0.0053 | 0.99752037 | 0.0053 | 0 | 0 | 0 | 1 | Ref |
| 13458 | MZ234039 | Unassigned | Unassigned | Unassigned | 283_78 | Clustered | 13 | VC_283_78 | 13 | 0.035 | 1 | 0.035 | 0 | 0 | 0 | 0.7949 | Ref |
| 13459 | MZ234040 | Unassigned | Unassigned | Unassigned | 283_78 | Clustered | 13 | VC_283_78 | 13 | 0.035 | 1 | 0.035 | 0 | 0 | 0 | 0.7949 | Ref |
| 13460 | MZ234041 | Unassigned | Unassigned | Unassigned | 283_78 | Clustered | 13 | VC_283_78 | 13 | 0.035 | 1 | 0.035 | 0 | 0 | 0 | 0.7949 | Ref |
| 13461 | MZ234042 | Unassigned | Unassigned | Unassigned | 283_78 | Clustered | 13 | VC_283_78 | 13 | 0.035 | 1 | 0.035 | 0 | 0 | 0 | 0.7949 | Ref |
| 13462 | MZ234043 | Unassigned | Unassigned | Unassigned | 283_78 | Clustered | 13 | VC_283_78 | 13 | 0.035 | 1 | 0.035 | 0 | 0 | 0 | 0.7949 | Ref |
| 13463 | MZ234044 | Unassigned | Unassigned | Unassigned | 451_0 | Clustered | 80 | VC_451_0 | 80 | 0.9991 | 1 | 0.9991 | 0 | 0 | 0 | 1 | Ref |
| 13464 | MZ234045 | Unassigned | Unassigned | Unassigned | 451_0 | Clustered | 80 | VC_451_0 | 80 | 0.9991 | 1 | 0.9991 | 0 | 0 | 0 | 1 | Ref |
| 13465 | MZ234046 | Unassigned | Unassigned | Unassigned | 451_0 | Clustered | 80 | VC_451_0 | 80 | 0.9991 | 1 | 0.9991 | 0 | 0 | 0 | 1 | Ref |
| 13466 | MZ234047 | Unassigned | Unassigned | Unassigned | 451_0 | Clustered | 80 | VC_451_0 | 80 | 0.9991 | 1 | 0.9991 | 0 | 0 | 0 | 1 | Ref |
| 13467 | MZ234048 | Unassigned | Unassigned | Unassigned | 638_0 | Clustered | 45 | VC_638_0 | 45 | 0.7741 | 1 | 0.7741 | 0 | 0 | 0 | 1 | Ref |
| 13468 | MZ234049 | Unassigned | Unassigned | Unassigned | 452_0 | Clustered | 66 | VC_452_0 | 66 | 0.9248 | 1 | 0.9248 | 0 | 0 | 0 | 1 | Ref |
| 13469 | MZ234050 | Unassigned | Unassigned | Unassigned | 452_0 | Clustered | 66 | VC_452_0 | 66 | 0.9248 | 1 | 0.9248 | 0 | 0 | 0 | 1 | Ref |
| 13470 | MZ234051 | Unassigned | Unassigned | Unassigned | 283_78 | Clustered | 13 | VC_283_78 | 13 | 0.035 | 1 | 0.035 | 0 | 0 | 0 | 0.7949 | Ref |
| 13471 | MZ234052 | Unassigned | Unassigned | Unassigned | 283_78 | Clustered | 13 | VC_283_78 | 13 | 0.035 | 1 | 0.035 | 0 | 0 | 0 | 0.7949 | Ref |
| 13472 | MZ234053 | Unassigned | Unassigned | Unassigned | 283_78 | Clustered | 13 | VC_283_78 | 13 | 0.035 | 1 | 0.035 | 0 | 0 | 0 | 0.7949 | Ref |
| 13473 | MZ272341 | Unassigned | Unassigned | Unassigned | 695_0 | Clustered | 48 | VC_695_0 | 48 | 0.8162 | 1 | 0.8162 | 0 | 0 | 0 | 1 | Ref |
| 13474 | MZ274225 | Unassigned | Unassigned | Unassigned | 637_0 | Clustered | 115 | VC_637_0 | 115 | 0.731 | 1 | 0.731 | 0 | 0 | 0 | 0.8764 | Ref |
| 13475 | MZ274304 | Unassigned | Unassigned | Unassigned | 525_0 | Clustered | 25 | VC_525_0 | 25 | 0.4597 | 1 | 0.4597 | 0 | 0 | 0 | 1 | Ref |
| 13476 | MZ274305 | Unassigned | Unassigned | Unassigned | 551_2 | Clustered | 17 | VC_551_2 | 17 | 0.0973 | 1 | 0.0973 | 0 | 0 | 0 | 0.9779 | Ref |
| 13477 | MZ274306 | Unassigned | Unassigned | Unassigned | 520_46 | Clustered/Singleton | 1 | VC_520_46 | 1 | 0 | 1 | 0 | 0 | 0 | 0 | 1 | Ref |
| 13478 | MZ274307 | Unassigned | Unassigned | Unassigned | 1344_0 | Clustered | 78 | VC_1344_0 | 78 | 0.9732 | 1 | 0.9732 | 0 | 0 | 0 | 1 | Ref |
| 13479 | MZ274308 | Unassigned | Unassigned | Unassigned | 1037_0 | Clustered | 6 | VC_1037_0 | 6 | 0.6798 | 1 | 0.6798 | 0 | 0 | 0 | 1 | Ref |
| 13480 | MZ274309 | Unassigned | Unassigned | Unassigned | 1164_0 | Clustered | 25 | VC_1164_0 | 25 | 0.7312 | 1 | 0.7312 | 0 | 0 | 0 | 1 | Ref |
| 13481 | MZ285878 | Unassigned | Unassigned | Unassigned | 79_7 | Clustered | 33 | VC_79_7 | 33 | 0.216 | 1 | 0.216 | 0 | 0 | 0 | 0.8409 | Ref |
| 13482 | MZ285879 | Unassigned | Unassigned | Unassigned | 630_0 | Clustered | 20 | VC_630_0 | 20 | 0.8513 | 1 | 0.8513 | 0 | 0 | 0 | 1 | Ref |
| 13483 | MZ286294 | Unassigned | Unassigned | Unassigned | 201_0 | Clustered | 3 | VC_201_0 | 3 | 0.0998 | 0.69690761 | 0.0695 | 0 | 0 | 0 | 1 | Ref |
| 13484 | MZ291552 | Unassigned | Unassigned | Unassigned | 283_4 | Clustered | 5 | VC_283_4 | 5 | 0.0112 | 0.99999969 | 0.0112 | 0 | 0 | 0 | 1 | Ref |
| 13485 | MZ292654 | Unassigned | Unassigned | Unassigned | 495_0 | Clustered | 39 | VC_495_0 | 39 | 0.308 | 1 | 0.308 | 0 | 0 | 0 | 1 | Ref |
| 13486 | MZ311861 | Unassigned | Unassigned | Unassigned | 451_0 | Clustered | 80 | VC_451_0 | 80 | 0.9991 | 1 | 0.9991 | 0 | 0 | 0 | 1 | Ref |
| 13487 | MZ311862 | Unassigned | Unassigned | Unassigned | 451_0 | Clustered | 80 | VC_451_0 | 80 | 0.9991 | 1 | 0.9991 | 0 | 0 | 0 | 1 | Ref |
| 13488 | MZ311863 | Unassigned | Unassigned | Unassigned | 329_22 | Clustered | 5 | VC_329_22 | 5 | 0.022 | 0.99999994 | 0.022 | 0 | 0 | 0 | 1 | Ref |
| 13489 | MZ311864 | Unassigned | Unassigned | Unassigned | 582_4 | Clustered | 3 | VC_582_4 | 3 | 0.0348 | 0.98173503 | 0.0341 | 0 | 0 | 0 | 1 | Ref |
| 13490 | MZ311865 | Unassigned | Unassigned | Unassigned | 286_13 | Clustered | 10 | VC_286_13 | 10 | 0.0698 | 0.99999929 | 0.0698 | 0 | 0 | 0 | 0.9556 | Ref |
| 13491 | MZ318361 | Unassigned | Unassigned | Unassigned | 392_0 | Clustered | 126 | VC_392_0 | 126 | 0.9621 | 1 | 0.9621 | 0 | 0 | 0 | 1 | Ref |
| 13492 | MZ318362 | Unassigned | Unassigned | Unassigned | 392_0 | Clustered | 126 | VC_392_0 | 126 | 0.9621 | 1 | 0.9621 | 0 | 0 | 0 | 1 | Ref |
| 13493 | MZ318363 | Unassigned | Unassigned | Unassigned | 392_0 | Clustered | 126 | VC_392_0 | 126 | 0.9621 | 1 | 0.9621 | 0 | 0 | 0 | 1 | Ref |
| 13494 | MZ318367 | Unassigned | Unassigned | Unassigned | 460_0 | Clustered | 19 | VC_460_0 | 19 | 0.3744 | 1 | 0.3744 | 0 | 0 | 0 | 1 | Ref |
| 13495 | MZ322004 | Unassigned | Unassigned | Unassigned | 325_35 | Clustered | 17 | VC_325_35 | 17 | 0.0732 | 1 | 0.0732 | 0 | 0 | 0 | 0.7721 | Ref |
| 13496 | MZ322006 | Unassigned | Unassigned | Unassigned | 1344_0 | Clustered | 78 | VC_1344_0 | 78 | 0.9732 | 1 | 0.9732 | 0 | 0 | 0 | 1 | Ref |
| 13497 | MZ322007 | Unassigned | Unassigned | Unassigned | 223_0 | Clustered | 38 | VC_223_0 | 38 | 0.9095 | 1 | 0.9095 | 0 | 0 | 0 | 1 | Ref |
| 13498 | MZ322008 | Unassigned | Unassigned | Unassigned | 1344_0 | Clustered | 78 | VC_1344_0 | 78 | 0.9732 | 1 | 0.9732 | 0 | 0 | 0 | 1 | Ref |
| 13499 | MZ322009 | Unassigned | Unassigned | Unassigned | 1095_0 | Clustered | 16 | VC_1095_0 | 16 | 0.458 | 1 | 0.458 | 0 | 0 | 0 | 1 | Ref |
| 13500 | MZ322010 | Unassigned | Unassigned | Unassigned | 767_0 | Clustered | 34 | VC_767_0 | 34 | 0.9173 | 1 | 0.9173 | 0 | 0 | 0 | 1 | Ref |
| 13501 | MZ322011 | Unassigned | Unassigned | Unassigned | 1171_0 | Clustered | 21 | VC_1171_0 | 21 | 0.7125 | 1 | 0.7125 | 0 | 0 | 0 | 1 | Ref |
| 13502 | MZ322012 | Unassigned | Unassigned | Unassigned | 1234_0 | Clustered | 44 | VC_1234_0 | 44 | 1 | 1 | 1 | 0 | 0 | 0 | 1 | Ref |
| 13503 | MZ322013 | Unassigned | Unassigned | Unassigned | 919_0 | Clustered | 39 | VC_919_0 | 39 | 0.9069 | 1 | 0.9069 | 0 | 0 | 0 | 1 | Ref |
| 13504 | MZ322014 | Unassigned | Unassigned | Unassigned | 1344_0 | Clustered | 78 | VC_1344_0 | 78 | 0.9732 | 1 | 0.9732 | 0 | 0 | 0 | 1 | Ref |
| 13505 | MZ322015 | Unassigned | Unassigned | Unassigned | 1093_0 | Clustered | 40 | VC_1093_0 | 40 | 0.8754 | 1 | 0.8754 | 0 | 0 | 0 | 1 | Ref |
| 13506 | MZ322016 | Unassigned | Unassigned | Unassigned | 238_0 | Clustered | 17 | VC_238_0 | 17 | 0.6109 | 1 | 0.6109 | 0 | 0 | 0 | 1 | Ref |
| 13507 | MZ322017 | Unassigned | Unassigned | Unassigned | 515_0 | Clustered | 16 | VC_515_0 | 16 | 0.3793 | 1 | 0.3793 | 0 | 0 | 0 | 1 | Ref |
| 13508 | MZ322018 | Unassigned | Unassigned | Unassigned | 1171_0 | Clustered | 21 | VC_1171_0 | 21 | 0.7125 | 1 | 0.7125 | 0 | 0 | 0 | 1 | Ref |
| 13509 | MZ322019 | Unassigned | Unassigned | Unassigned | 216_4 | Clustered | 13 | VC_216_4 | 13 | 0.1657 | 1 | 0.1657 | 0 | 0 | 0 | 1 | Ref |
| 13510 | MZ322020 | Unassigned | Unassigned | Unassigned | 1095_0 | Clustered | 16 | VC_1095_0 | 16 | 0.458 | 1 | 0.458 | 0 | 0 | 0 | 1 | Ref |
| 13511 | MZ322021 | Unassigned | Unassigned | Unassigned | 955_0 | Clustered | 3 | VC_955_0 | 3 | 0.7745 | 0.99134622 | 0.7678 | 0 | 0 | 0 | 1 | Ref |
| 13512 | MZ322022 | Unassigned | Unassigned | Unassigned | 767_0 | Clustered | 34 | VC_767_0 | 34 | 0.9173 | 1 | 0.9173 | 0 | 0 | 0 | 1 | Ref |
| 13513 | MZ322023 | Unassigned | Unassigned | Unassigned | 517_0 | Clustered | 18 | VC_517_0 | 18 | 0.1599 | 1 | 0.1599 | 0 | 0 | 0 | 1 | Ref |
| 13514 | MZ322024 | Unassigned | Unassigned | Unassigned | 551_7 | Clustered | 28 | VC_551_7 | 28 | 0.1661 | 1 | 0.1661 | 0 | 0 | 0 | 0.8519 | Ref |
| 13515 | MZ322025 | Unassigned | Unassigned | Unassigned | 517_1 | Clustered | 108 | VC_517_1 | 108 | 0.8483 | 1 | 0.8483 | 0 | 0 | 0 | 0.9955 | Ref |
| 13516 | MZ322026 | Unassigned | Unassigned | Unassigned | 1352_0 | Clustered | 31 | VC_1352_0 | 31 | 0.849 | 1 | 0.849 | 0 | 0 | 0 | 1 | Ref |
| 13517 | MZ322027 | Unassigned | Unassigned | Unassigned | 1344_0 | Clustered | 78 | VC_1344_0 | 78 | 0.9732 | 1 | 0.9732 | 0 | 0 | 0 | 1 | Ref |
| 13518 | MZ322028 | Unassigned | Unassigned | Unassigned | 685_0 | Clustered | 16 | VC_685_0 | 16 | 0.2972 | 1 | 0.2972 | 0 | 0 | 0 | 0.9167 | Ref |
| 13519 | MZ322895 | Unassigned | Unassigned | Unassigned | 668_0 | Clustered | 25 | VC_668_0 | 25 | 0.5539 | 1 | 0.5539 | 0 | 0 | 0 | 0.96 | Ref |
| 13520 | MZ326168 | Unassigned | Unassigned | Unassigned | 582_5 | Clustered | 4 | VC_582_5 | 4 | 0.0528 | 0.99935625 | 0.0527 | 0 | 0 | 0 | 0.8333 | Ref |
| 13521 | MZ326855 | Unassigned | Unassigned | Unassigned | 172_0 | Clustered | 5 | VC_172_0 | 5 | 0.3579 | 0.99999997 | 0.3579 | 0 | 0 | 0 | 1 | Ref |
| 13522 | MZ326856 | Unassigned | Unassigned | Unassigned | 779_3 | Clustered | 7 | VC_779_3 | 7 | 0.0766 | 1 | 0.0766 | 0 | 0 | 0 | 0.7143 | Ref |
| 13523 | MZ326857 | Unassigned | Unassigned | Unassigned | 779_3 | Clustered | 7 | VC_779_3 | 7 | 0.0766 | 1 | 0.0766 | 0 | 0 | 0 | 0.7143 | Ref |
| 13524 | MZ326859 | Unassigned | Unassigned | Unassigned | 671_0 | Clustered | 5 | VC_671_0 | 5 | 0.5289 | 0.99999996 | 0.5289 | 0 | 0 | 0 | 1 | Ref |
| 13525 | MZ326861 | Unassigned | Unassigned | Unassigned | 779_3 | Clustered | 7 | VC_779_3 | 7 | 0.0766 | 1 | 0.0766 | 0 | 0 | 0 | 0.7143 | Ref |
| 13526 | MZ326862 | Unassigned | Unassigned | Unassigned | 905_0 | Clustered | 3 | VC_905_0 | 3 | 0.3424 | 0.99823775 | 0.3417 | 0 | 0 | 0 | 1 | Ref |
| 13527 | MZ326863 | Unassigned | Unassigned | Unassigned | 70_0 | Clustered | 27 | VC_70_0 | 27 | 0.5981 | 1 | 0.5981 | 0 | 0 | 0 | 0.9886 | Ref |
| 13528 | MZ326864 | Unassigned | Unassigned | Unassigned | 615_0 | Clustered | 2 | VC_615_0 | 2 | 0.2668 | 0.94178001 | 0.2513 | 0 | 0 | 0 | 1 | Ref |
| 13529 | MZ326865 | Unassigned | Unassigned | Unassigned | 695_0 | Clustered | 48 | VC_695_0 | 48 | 0.8162 | 1 | 0.8162 | 0 | 0 | 0 | 1 | Ref |
| 13530 | MZ326866 | Unassigned | Unassigned | Unassigned | 316_0 | Clustered | 3 | VC_316_0 | 3 | 0.774 | 0.99820498 | 0.7726 | 0 | 0 | 0 | 1 | Ref |
| 13531 | MZ326868 | Unassigned | Unassigned | Unassigned | 506_0 | Clustered | 8 | VC_506_0 | 8 | 0.7376 | 1 | 0.7376 | 0 | 0 | 0 | 1 | Ref |
| 13532 | MZ327261 | Unassigned | Unassigned | Unassigned | 329_1 | Clustered | 17 | VC_329_1 | 17 | 0.0867 | 1 | 0.0867 | 0 | 0 | 0 | 0.8235 | Ref |
| 13533 | MZ327262 | Unassigned | Unassigned | Unassigned | 329_1 | Clustered | 17 | VC_329_1 | 17 | 0.0867 | 1 | 0.0867 | 0 | 0 | 0 | 0.8235 | Ref |
| 13534 | MZ333134 | Unassigned | Unassigned | Unassigned | 744_0 | Clustered | 10 | VC_744_0 | 10 | 0.7049 | 1 | 0.7049 | 0 | 0 | 0 | 1 | Ref |
| 13535 | MZ333135 | Unassigned | Unassigned | Unassigned | 110_0 | Clustered | 4 | VC_110_0 | 4 | 0.1143 | 0.99998732 | 0.1143 | 0 | 0 | 0 | 1 | Ref |
| 13536 | MZ333136 | Unassigned | Unassigned | Unassigned | 744_0 | Clustered | 10 | VC_744_0 | 10 | 0.7049 | 1 | 0.7049 | 0 | 0 | 0 | 1 | Ref |
| 13537 | MZ333457 | Unassigned | Unassigned | Unassigned | 108_0 | Clustered | 33 | VC_108_0 | 33 | 0.949 | 1 | 0.949 | 0 | 0 | 0 | 1 | Ref |
| 13538 | MZ333461 | Unassigned | Unassigned | Unassigned | 695_0 | Clustered | 48 | VC_695_0 | 48 | 0.8162 | 1 | 0.8162 | 0 | 0 | 0 | 1 | Ref |
| 13539 | MZ333462 | Unassigned | Unassigned | Unassigned | 92_0 | Clustered | 22 | VC_92_0 | 22 | 0.5639 | 1 | 0.5639 | 0 | 0 | 0 | 1 | Ref |
| 13540 | MZ334492 | Unassigned | Unassigned | Unassigned | 310_0 | Clustered | 33 | VC_310_0 | 33 | 0.8771 | 1 | 0.8771 | 0 | 0 | 0 | 1 | Ref |
| 13541 | MZ334493 | Unassigned | Unassigned | Unassigned | 310_0 | Clustered | 33 | VC_310_0 | 33 | 0.8771 | 1 | 0.8771 | 0 | 0 | 0 | 1 | Ref |
| 13542 | MZ334494 | Unassigned | Unassigned | Unassigned | 310_0 | Clustered | 33 | VC_310_0 | 33 | 0.8771 | 1 | 0.8771 | 0 | 0 | 0 | 1 | Ref |
| 13543 | MZ334495 | Unassigned | Unassigned | Unassigned | 310_0 | Clustered | 33 | VC_310_0 | 33 | 0.8771 | 1 | 0.8771 | 0 | 0 | 0 | 1 | Ref |
| 13544 | MZ334496 | Unassigned | Unassigned | Unassigned | 310_0 | Clustered | 33 | VC_310_0 | 33 | 0.8771 | 1 | 0.8771 | 0 | 0 | 0 | 1 | Ref |
| 13545 | MZ334497 | Unassigned | Unassigned | Unassigned | 310_0 | Clustered | 33 | VC_310_0 | 33 | 0.8771 | 1 | 0.8771 | 0 | 0 | 0 | 1 | Ref |
| 13546 | MZ334498 | Unassigned | Unassigned | Unassigned | 310_0 | Clustered | 33 | VC_310_0 | 33 | 0.8771 | 1 | 0.8771 | 0 | 0 | 0 | 1 | Ref |
| 13547 | MZ334499 | Unassigned | Unassigned | Unassigned | 310_0 | Clustered | 33 | VC_310_0 | 33 | 0.8771 | 1 | 0.8771 | 0 | 0 | 0 | 1 | Ref |
| 13548 | MZ334500 | Unassigned | Unassigned | Unassigned | 310_0 | Clustered | 33 | VC_310_0 | 33 | 0.8771 | 1 | 0.8771 | 0 | 0 | 0 | 1 | Ref |
| 13549 | MZ334501 | Unassigned | Unassigned | Unassigned | 310_0 | Clustered | 33 | VC_310_0 | 33 | 0.8771 | 1 | 0.8771 | 0 | 0 | 0 | 1 | Ref |
| 13550 | MZ334502 | Unassigned | Unassigned | Unassigned | 310_0 | Clustered | 33 | VC_310_0 | 33 | 0.8771 | 1 | 0.8771 | 0 | 0 | 0 | 1 | Ref |
| 13551 | MZ334503 | Unassigned | Unassigned | Unassigned | 310_0 | Clustered | 33 | VC_310_0 | 33 | 0.8771 | 1 | 0.8771 | 0 | 0 | 0 | 1 | Ref |
| 13552 | MZ334504 | Unassigned | Unassigned | Unassigned | 310_0 | Clustered | 33 | VC_310_0 | 33 | 0.8771 | 1 | 0.8771 | 0 | 0 | 0 | 1 | Ref |
| 13553 | MZ334505 | Unassigned | Unassigned | Unassigned | 310_0 | Clustered | 33 | VC_310_0 | 33 | 0.8771 | 1 | 0.8771 | 0 | 0 | 0 | 1 | Ref |
| 13554 | MZ334506 | Unassigned | Unassigned | Unassigned | 310_0 | Clustered | 33 | VC_310_0 | 33 | 0.8771 | 1 | 0.8771 | 0 | 0 | 0 | 1 | Ref |
| 13555 | MZ334507 | Unassigned | Unassigned | Unassigned | 310_0 | Clustered | 33 | VC_310_0 | 33 | 0.8771 | 1 | 0.8771 | 0 | 0 | 0 | 1 | Ref |
| 13556 | MZ334508 | Unassigned | Unassigned | Unassigned | 310_0 | Clustered | 33 | VC_310_0 | 33 | 0.8771 | 1 | 0.8771 | 0 | 0 | 0 | 1 | Ref |
| 13557 | MZ334509 | Unassigned | Unassigned | Unassigned | 310_0 | Clustered | 33 | VC_310_0 | 33 | 0.8771 | 1 | 0.8771 | 0 | 0 | 0 | 1 | Ref |
| 13558 | MZ334510 | Unassigned | Unassigned | Unassigned | 310_0 | Clustered | 33 | VC_310_0 | 33 | 0.8771 | 1 | 0.8771 | 0 | 0 | 0 | 1 | Ref |
| 13559 | MZ334511 | Unassigned | Unassigned | Unassigned | 310_0 | Clustered | 33 | VC_310_0 | 33 | 0.8771 | 1 | 0.8771 | 0 | 0 | 0 | 1 | Ref |
| 13560 | MZ334512 | Unassigned | Unassigned | Unassigned | 310_0 | Clustered | 33 | VC_310_0 | 33 | 0.8771 | 1 | 0.8771 | 0 | 0 | 0 | 1 | Ref |
| 13561 | MZ334513 | Unassigned | Unassigned | Unassigned | 310_0 | Clustered | 33 | VC_310_0 | 33 | 0.8771 | 1 | 0.8771 | 0 | 0 | 0 | 1 | Ref |
| 13562 | MZ334514 | Unassigned | Unassigned | Unassigned | 310_0 | Clustered | 33 | VC_310_0 | 33 | 0.8771 | 1 | 0.8771 | 0 | 0 | 0 | 1 | Ref |
| 13563 | MZ334515 | Unassigned | Unassigned | Unassigned | 310_0 | Clustered | 33 | VC_310_0 | 33 | 0.8771 | 1 | 0.8771 | 0 | 0 | 0 | 1 | Ref |
| 13564 | MZ334516 | Unassigned | Unassigned | Unassigned | 310_0 | Clustered | 33 | VC_310_0 | 33 | 0.8771 | 1 | 0.8771 | 0 | 0 | 0 | 1 | Ref |
| 13565 | MZ334517 | Unassigned | Unassigned | Unassigned | 311_0 | Clustered | 6 | VC_311_0 | 6 | 0.1997 | 1 | 0.1997 | 0 | 0 | 0 | 1 | Ref |
| 13566 | MZ334518 | Unassigned | Unassigned | Unassigned | 311_0 | Clustered | 6 | VC_311_0 | 6 | 0.1997 | 1 | 0.1997 | 0 | 0 | 0 | 1 | Ref |
| 13567 | MZ334519 | Unassigned | Unassigned | Unassigned | 311_0 | Clustered | 6 | VC_311_0 | 6 | 0.1997 | 1 | 0.1997 | 0 | 0 | 0 | 1 | Ref |
| 13568 | MZ334520 | Unassigned | Unassigned | Unassigned | 311_0 | Clustered | 6 | VC_311_0 | 6 | 0.1997 | 1 | 0.1997 | 0 | 0 | 0 | 1 | Ref |
| 13569 | MZ334523 | Unassigned | Unassigned | Unassigned | 313_1 | Clustered | 3 | VC_313_1 | 3 | 0.3511 | 0.99802018 | 0.3504 | 0 | 0 | 0 | 1 | Ref |
| 13570 | MZ334524 | Unassigned | Unassigned | Unassigned | 313_0 | Clustered | 2 | VC_313_0 | 2 | 0.1721 | 0.90744399 | 0.1562 | 0 | 0 | 0 | 1 | Ref |
| 13571 | MZ334525 | Unassigned | Unassigned | Unassigned | 312_0 | Clustered | 2 | VC_312_0 | 2 | 0.0307 | 0.95436427 | 0.0293 | 0 | 0 | 0 | 1 | Ref |
| 13572 | MZ334526 | Unassigned | Unassigned | Unassigned | 459_0 | Clustered | 3 | VC_459_0 | 3 | 0.8542 | 0.99054823 | 0.8461 | 0 | 0 | 0 | 1 | Ref |
| 13573 | MZ334527 | Unassigned | Unassigned | Unassigned | 1002_0 | Clustered | 2 | VC_1002_0 | 2 | 0.1809 | 0.72090768 | 0.1304 | 0 | 0 | 0 | 1 | Ref |
| 13574 | MZ336020 | Unassigned | Unassigned | Unassigned | 793_0 | Clustered | 12 | VC_793_0 | 12 | 0.8531 | 1 | 0.8531 | 0 | 0 | 0 | 1 | Ref |
| 13575 | MZ342906 | Unassigned | Unassigned | Unassigned | 813_0 | Clustered | 12 | VC_813_0 | 12 | 0.2347 | 1 | 0.2347 | 0 | 0 | 0 | 1 | Ref |
| 13576 | MZ345003 | Unassigned | Unassigned | Unassigned | 125_0 | Clustered | 17 | VC_125_0 | 17 | 0.9244 | 1 | 0.9244 | 0 | 0 | 0 | 1 | Ref |
| 13577 | MZ348421 | Unassigned | Unassigned | Unassigned | 841_0 | Clustered | 10 | VC_841_0 | 10 | 0.5997 | 1 | 0.5997 | 0 | 0 | 0 | 1 | Ref |
| 13578 | MZ348422 | Unassigned | Unassigned | Unassigned | 442_2 | Clustered | 4 | VC_442_2 | 4 | 0.0844 | 0.99988129 | 0.0844 | 0 | 0 | 0 | 0.8333 | Ref |
| 13579 | MZ348423 | Unassigned | Unassigned | Unassigned | 490_1 | Clustered | 5 | VC_490_1 | 5 | 0.0447 | 0.99999996 | 0.0447 | 0 | 0 | 0 | 1 | Ref |
| 13580 | MZ348425 | Unassigned | Unassigned | Unassigned | 697_0 | Clustered | 5 | VC_697_0 | 5 | 0.8303 | 0.99999971 | 0.8303 | 0 | 0 | 0 | 1 | Ref |
| 13581 | MZ348426 | Unassigned | Unassigned | Unassigned | 78_3 | Clustered | 11 | VC_78_3 | 11 | 0.2572 | 1 | 0.2572 | 0 | 0 | 0 | 1 | Ref |
| 13582 | MZ355721 | Unassigned | Unassigned | Unassigned | 236_0 | Clustered | 72 | VC_236_0 | 72 | 0.8104 | 1 | 0.8104 | 0 | 0 | 0 | 1 | Ref |
| 13583 | MZ355722 | Unassigned | Unassigned | Unassigned | 325_55 | Clustered | 2 | VC_325_55 | 2 | 0.0061 | 0.95750995 | 0.0058 | 0 | 0 | 0 | 1 | Ref |
| 13584 | MZ355723 | Unassigned | Unassigned | Unassigned | 515_0 | Clustered | 16 | VC_515_0 | 16 | 0.3793 | 1 | 0.3793 | 0 | 0 | 0 | 1 | Ref |
| 13585 | MZ355724 | Unassigned | Unassigned | Unassigned | 856_0 | Clustered | 7 | VC_856_0 | 7 | 0.3463 | 1 | 0.3463 | 0 | 0 | 0 | 1 | Ref |
| 13586 | MZ355725 | Unassigned | Unassigned | Unassigned | 517_1 | Clustered | 108 | VC_517_1 | 108 | 0.8483 | 1 | 0.8483 | 0 | 0 | 0 | 0.9955 | Ref |
| 13587 | MZ355726 | Unassigned | Unassigned | Unassigned | 325_39 | Clustered | 2 | VC_325_39 | 2 | 0.0045 | 0.95750889 | 0.0043 | 0 | 0 | 0 | 1 | Ref |
| 13588 | MZ355728 | Unassigned | Unassigned | Unassigned | 216_6 | Clustered | 42 | VC_216_6 | 42 | 0.473 | 1 | 0.473 | 0 | 0 | 0 | 0.8351 | Ref |
| 13589 | MZ358387 | Unassigned | Unassigned | Unassigned | 779_0 | Clustered | 37 | VC_779_0 | 37 | 0.5682 | 1 | 0.5682 | 0 | 0 | 0 | 1 | Ref |
| 13590 | MZ359091 | Unassigned | Unassigned | Unassigned | 96_0 | Clustered | 30 | VC_96_0 | 30 | 0.718 | 1 | 0.718 | 0 | 0 | 0 | 1 | Ref |
| 13591 | MZ359670 | Unassigned | Unassigned | Unassigned | 385_0 | Clustered | 51 | VC_385_0 | 51 | 0.8754 | 1 | 0.8754 | 0 | 0 | 0 | 1 | Ref |
| 13592 | MZ361825 | Unassigned | Unassigned | Unassigned | 378_0 | Clustered | 9 | VC_378_0 | 9 | 0.7603 | 1 | 0.7603 | 0 | 0 | 0 | 1 | Ref |
| 13593 | MZ361826 | Unassigned | Unassigned | Unassigned | 378_0 | Clustered | 9 | VC_378_0 | 9 | 0.7603 | 1 | 0.7603 | 0 | 0 | 0 | 1 | Ref |
| 13594 | MZ361827 | Unassigned | Unassigned | Unassigned | 378_0 | Clustered | 9 | VC_378_0 | 9 | 0.7603 | 1 | 0.7603 | 0 | 0 | 0 | 1 | Ref |
| 13595 | MZ364222 | Unassigned | Unassigned | Unassigned | 1066_0 | Clustered | 52 | VC_1066_0 | 52 | 0.7682 | 1 | 0.7682 | 0 | 0 | 0 | 1 | Ref |
| 13596 | MZ364223 | Unassigned | Unassigned | Unassigned | 1066_0 | Clustered | 52 | VC_1066_0 | 52 | 0.7682 | 1 | 0.7682 | 0 | 0 | 0 | 1 | Ref |
| 13597 | MZ364224 | Unassigned | Unassigned | Unassigned | 1077_0 | Clustered | 48 | VC_1077_0 | 48 | 0.8525 | 1 | 0.8525 | 0 | 0 | 0 | 1 | Ref |
| 13598 | MZ364225 | Unassigned | Unassigned | Unassigned | 1077_0 | Clustered | 48 | VC_1077_0 | 48 | 0.8525 | 1 | 0.8525 | 0 | 0 | 0 | 1 | Ref |
| 13599 | MZ364226 | Unassigned | Unassigned | Unassigned | 1000_0 | Clustered | 21 | VC_1000_0 | 21 | 0.8437 | 0.99999998 | 0.8437 | 0 | 0 | 0 | 1 | Ref |
| 13600 | MZ364227 | Unassigned | Unassigned | Unassigned | 1077_0 | Clustered | 48 | VC_1077_0 | 48 | 0.8525 | 1 | 0.8525 | 0 | 0 | 0 | 1 | Ref |
| 13601 | MZ364228 | Unassigned | Unassigned | Unassigned | 995_0 | Clustered | 69 | VC_995_0 | 69 | 0.8547 | 1 | 0.8547 | 0 | 0 | 0 | 1 | Ref |
| 13602 | MZ364231 | Unassigned | Unassigned | Unassigned | 1068_0 | Clustered | 5 | VC_1068_0 | 5 | 1 | 1 | 1 | 0 | 0 | 0 | 1 | Ref |
| 13603 | MZ364232 | Unassigned | Unassigned | Unassigned | 1066_0 | Clustered | 52 | VC_1066_0 | 52 | 0.7682 | 1 | 0.7682 | 0 | 0 | 0 | 1 | Ref |
| 13604 | MZ364233 | Unassigned | Unassigned | Unassigned | 995_0 | Clustered | 69 | VC_995_0 | 69 | 0.8547 | 1 | 0.8547 | 0 | 0 | 0 | 1 | Ref |
| 13605 | MZ364234 | Unassigned | Unassigned | Unassigned | 1324_0 | Clustered | 4 | VC_1324_0 | 4 | 0.6008 | 0.54407361 | 0.3269 | 0 | 0 | 0 | 1 | Ref |
| 13606 | MZ364235 | Unassigned | Unassigned | Unassigned | 1463_0 | Clustered | 6 | VC_1463_0 | 6 | 1 | 1 | 1 | 0 | 0 | 0 | 1 | Ref |

| | | | | | | | | | | | | | | | | | |
|---|---|---|---|---|---|---|---|---|---|---|---|---|---|---|---|---|---|
| 13607 | MZ364236 | Unassigned | Unassigned | Unassigned | 1463_0 | Clustered | 6 | VC_1463_0 | 6 | 1 | 1 | 1 | 0 | 0 | 0 | 1 | Ref |
| 13608 | MZ364238 | Unassigned | Unassigned | Unassigned | 1463_0 | Clustered | 6 | VC_1463_0 | 6 | 1 | 1 | 1 | 0 | 0 | 0 | 1 | Ref |
| 13609 | MZ364239 | Unassigned | Unassigned | Unassigned | 1463_0 | Clustered | 6 | VC_1463_0 | 6 | 1 | 1 | 1 | 0 | 0 | 0 | 1 | Ref |
| 13610 | MZ364240 | Unassigned | Unassigned | Unassigned | 995_0 | Clustered | 69 | VC_995_0 | 69 | 0.8547 | 1 | 0.8547 | 0 | 0 | 0 | 1 | Ref |
| 13611 | MZ364241 | Unassigned | Unassigned | Unassigned | 1000_0 | Clustered | 21 | VC_1000_0 | 21 | 0.8437 | 0.99999998 | 0.8437 | 0 | 0 | 0 | 1 | Ref |
| 13612 | MZ364242 | Unassigned | Unassigned | Unassigned | 1062_0 | Clustered | 45 | VC_1062_0 | 45 | 0.9734 | 0.50949847 | 0.4959 | 0 | 0 | 0 | 1 | Ref |
| 13613 | MZ364243 | Unassigned | Unassigned | Unassigned | 995_0 | Clustered | 69 | VC_995_0 | 69 | 0.8547 | 1 | 0.8547 | 0 | 0 | 0 | 1 | Ref |
| 13614 | MZ364244 | Unassigned | Unassigned | Unassigned | 1000_0 | Clustered | 21 | VC_1000_0 | 21 | 0.8437 | 0.99999998 | 0.8437 | 0 | 0 | 0 | 1 | Ref |
| 13615 | MZ364245 | Unassigned | Unassigned | Unassigned | 1000_0 | Clustered | 21 | VC_1000_0 | 21 | 0.8437 | 0.99999998 | 0.8437 | 0 | 0 | 0 | 1 | Ref |
| 13616 | MZ364246 | Unassigned | Unassigned | Unassigned | 1000_0 | Clustered | 21 | VC_1000_0 | 21 | 0.8437 | 0.99999998 | 0.8437 | 0 | 0 | 0 | 1 | Ref |
| 13617 | MZ364247 | Unassigned | Unassigned | Unassigned | 1000_0 | Clustered | 21 | VC_1000_0 | 21 | 0.8437 | 0.99999998 | 0.8437 | 0 | 0 | 0 | 1 | Ref |
| 13618 | MZ364248 | Unassigned | Unassigned | Unassigned | 1066_0 | Clustered | 52 | VC_1066_0 | 52 | 0.7682 | 1 | 0.7682 | 0 | 0 | 0 | 1 | Ref |
| 13619 | MZ364249 | Unassigned | Unassigned | Unassigned | 1000_0 | Clustered | 21 | VC_1000_0 | 21 | 0.8437 | 0.99999998 | 0.8437 | 0 | 0 | 0 | 1 | Ref |
| 13620 | MZ364250 | Unassigned | Unassigned | Unassigned | 1000_0 | Clustered | 21 | VC_1000_0 | 21 | 0.8437 | 0.99999998 | 0.8437 | 0 | 0 | 0 | 1 | Ref |
| 13621 | MZ364251 | Unassigned | Unassigned | Unassigned | 1464_0 | Clustered | 3 | VC_1464_0 | 3 | 1 | 1 | 1 | 0 | 0 | 0 | 1 | Ref |
| 13622 | MZ364252 | Unassigned | Unassigned | Unassigned | 1338_0 | Clustered | 4 | VC_1338_0 | 4 | 1 | 1 | 1 | 0 | 0 | 0 | 1 | Ref |
| 13623 | MZ364254 | Unassigned | Unassigned | Unassigned | 1066_0 | Clustered | 52 | VC_1066_0 | 52 | 0.7682 | 1 | 0.7682 | 0 | 0 | 0 | 1 | Ref |
| 13624 | MZ364255 | Unassigned | Unassigned | Unassigned | 1066_0 | Clustered | 52 | VC_1066_0 | 52 | 0.7682 | 1 | 0.7682 | 0 | 0 | 0 | 1 | Ref |
| 13625 | MZ364256 | Unassigned | Unassigned | Unassigned | 995_0 | Clustered | 69 | VC_995_0 | 69 | 0.8547 | 1 | 0.8547 | 0 | 0 | 0 | 1 | Ref |
| 13626 | MZ364257 | Unassigned | Unassigned | Unassigned | 1463_0 | Clustered | 6 | VC_1463_0 | 6 | 1 | 1 | 1 | 0 | 0 | 0 | 1 | Ref |
| 13627 | MZ364258 | Unassigned | Unassigned | Unassigned | 995_0 | Clustered | 69 | VC_995_0 | 69 | 0.8547 | 1 | 0.8547 | 0 | 0 | 0 | 1 | Ref |
| 13628 | MZ364259 | Unassigned | Unassigned | Unassigned | 995_0 | Clustered | 69 | VC_995_0 | 69 | 0.8547 | 1 | 0.8547 | 0 | 0 | 0 | 1 | Ref |
| 13629 | MZ364260 | Unassigned | Unassigned | Unassigned | 1000_0 | Clustered | 21 | VC_1000_0 | 21 | 0.8437 | 0.99999998 | 0.8437 | 0 | 0 | 0 | 1 | Ref |
| 13630 | MZ364261 | Unassigned | Unassigned | Unassigned | 1066_0 | Clustered | 52 | VC_1066_0 | 52 | 0.7682 | 1 | 0.7682 | 0 | 0 | 0 | 1 | Ref |
| 13631 | MZ364262 | Unassigned | Unassigned | Unassigned | 1066_0 | Clustered | 52 | VC_1066_0 | 52 | 0.7682 | 1 | 0.7682 | 0 | 0 | 0 | 1 | Ref |
| 13632 | MZ364263 | Unassigned | Unassigned | Unassigned | 1000_0 | Clustered | 21 | VC_1000_0 | 21 | 0.8437 | 0.99999998 | 0.8437 | 0 | 0 | 0 | 1 | Ref |
| 13633 | MZ364264 | Unassigned | Unassigned | Unassigned | 1066_0 | Clustered | 52 | VC_1066_0 | 52 | 0.7682 | 1 | 0.7682 | 0 | 0 | 0 | 1 | Ref |
| 13634 | MZ364265 | Unassigned | Unassigned | Unassigned | 1066_0 | Clustered | 52 | VC_1066_0 | 52 | 0.7682 | 1 | 0.7682 | 0 | 0 | 0 | 1 | Ref |
| 13635 | MZ364266 | Unassigned | Unassigned | Unassigned | 1000_0 | Clustered | 21 | VC_1000_0 | 21 | 0.8437 | 0.99999998 | 0.8437 | 0 | 0 | 0 | 1 | Ref |
| 13636 | MZ364267 | Unassigned | Unassigned | Unassigned | 995_0 | Clustered | 69 | VC_995_0 | 69 | 0.8547 | 1 | 0.8547 | 0 | 0 | 0 | 1 | Ref |
| 13637 | MZ364268 | Unassigned | Unassigned | Unassigned | 1066_0 | Clustered | 52 | VC_1066_0 | 52 | 0.7682 | 1 | 0.7682 | 0 | 0 | 0 | 1 | Ref |
| 13638 | MZ364269 | Unassigned | Unassigned | Unassigned | 1066_0 | Clustered | 52 | VC_1066_0 | 52 | 0.7682 | 1 | 0.7682 | 0 | 0 | 0 | 1 | Ref |
| 13639 | MZ364270 | Unassigned | Unassigned | Unassigned | 1000_0 | Clustered | 21 | VC_1000_0 | 21 | 0.8437 | 0.99999998 | 0.8437 | 0 | 0 | 0 | 1 | Ref |
| 13640 | MZ364271 | Unassigned | Unassigned | Unassigned | 1066_0 | Clustered | 52 | VC_1066_0 | 52 | 0.7682 | 1 | 0.7682 | 0 | 0 | 0 | 1 | Ref |
| 13641 | MZ364272 | Unassigned | Unassigned | Unassigned | 1066_0 | Clustered | 52 | VC_1066_0 | 52 | 0.7682 | 1 | 0.7682 | 0 | 0 | 0 | 1 | Ref |
| 13642 | MZ364273 | Unassigned | Unassigned | Unassigned | 1066_0 | Clustered | 52 | VC_1066_0 | 52 | 0.7682 | 1 | 0.7682 | 0 | 0 | 0 | 1 | Ref |
| 13643 | MZ364274 | Unassigned | Unassigned | Unassigned | 995_0 | Clustered | 69 | VC_995_0 | 69 | 0.8547 | 1 | 0.8547 | 0 | 0 | 0 | 1 | Ref |
| 13644 | MZ364275 | Unassigned | Unassigned | Unassigned | 1066_0 | Clustered | 52 | VC_1066_0 | 52 | 0.7682 | 1 | 0.7682 | 0 | 0 | 0 | 1 | Ref |
| 13645 | MZ364276 | Unassigned | Unassigned | Unassigned | 1066_0 | Clustered | 52 | VC_1066_0 | 52 | 0.7682 | 1 | 0.7682 | 0 | 0 | 0 | 1 | Ref |
| 13646 | MZ364277 | Unassigned | Unassigned | Unassigned | 1077_0 | Clustered | 48 | VC_1077_0 | 48 | 0.8525 | 1 | 0.8525 | 0 | 0 | 0 | 1 | Ref |
| 13647 | MZ364278 | Unassigned | Unassigned | Unassigned | 1066_0 | Clustered | 52 | VC_1066_0 | 52 | 0.7682 | 1 | 0.7682 | 0 | 0 | 0 | 1 | Ref |
| 13648 | MZ364279 | Unassigned | Unassigned | Unassigned | 1077_0 | Clustered | 48 | VC_1077_0 | 48 | 0.8525 | 1 | 0.8525 | 0 | 0 | 0 | 1 | Ref |
| 13649 | MZ364280 | Unassigned | Unassigned | Unassigned | 1068_0 | Clustered | 5 | VC_1068_0 | 5 | 1 | 1 | 1 | 0 | 0 | 0 | 1 | Ref |
| 13650 | MZ364282 | Unassigned | Unassigned | Unassigned | 1464_0 | Clustered | 3 | VC_1464_0 | 3 | 1 | 1 | 1 | 0 | 0 | 0 | 1 | Ref |
| 13651 | MZ364283 | Unassigned | Unassigned | Unassigned | 1077_0 | Clustered | 48 | VC_1077_0 | 48 | 0.8525 | 1 | 0.8525 | 0 | 0 | 0 | 1 | Ref |
| 13652 | MZ364284 | Unassigned | Unassigned | Unassigned | 1068_0 | Clustered | 5 | VC_1068_0 | 5 | 1 | 1 | 1 | 0 | 0 | 0 | 1 | Ref |
| 13653 | MZ364285 | Unassigned | Unassigned | Unassigned | 1077_0 | Clustered | 48 | VC_1077_0 | 48 | 0.8525 | 1 | 0.8525 | 0 | 0 | 0 | 1 | Ref |
| 13654 | MZ364286 | Unassigned | Unassigned | Unassigned | 1077_0 | Clustered | 48 | VC_1077_0 | 48 | 0.8525 | 1 | 0.8525 | 0 | 0 | 0 | 1 | Ref |
| 13655 | MZ364289 | Unassigned | Unassigned | Unassigned | 1463_0 | Clustered | 6 | VC_1463_0 | 6 | 1 | 1 | 1 | 0 | 0 | 0 | 1 | Ref |
| 13656 | MZ364291 | Unassigned | Unassigned | Unassigned | 1077_0 | Clustered | 48 | VC_1077_0 | 48 | 0.8525 | 1 | 0.8525 | 0 | 0 | 0 | 1 | Ref |
| 13657 | MZ364293 | Unassigned | Unassigned | Unassigned | 1338_0 | Clustered | 4 | VC_1338_0 | 4 | 1 | 1 | 1 | 0 | 0 | 0 | 1 | Ref |
| 13658 | MZ364294 | Unassigned | Unassigned | Unassigned | 1464_0 | Clustered | 3 | VC_1464_0 | 3 | 1 | 1 | 1 | 0 | 0 | 0 | 1 | Ref |
| 13659 | MZ368140 | Unassigned | Unassigned | Unassigned | 209_0 | Clustered | 4 | VC_209_0 | 4 | 0.1136 | 0.99998779 | 0.1136 | 0 | 0 | 0 | 1 | Ref |
| 13660 | MZ369152 | Unassigned | Unassigned | Unassigned | 948_0 | Clustered | 9 | VC_948_0 | 9 | 0.5443 | 1 | 0.5443 | 0 | 0 | 0 | 1 | Ref |
| 13661 | MZ374361 | Unassigned | Unassigned | Unassigned | 123_0 | Clustered | 23 | VC_123_0 | 23 | 0.817 | 1 | 0.817 | 0 | 0 | 0 | 1 | Ref |
| 13662 | MZ375232 | Unassigned | Unassigned | Unassigned | 461_0 | Clustered | 96 | VC_461_0 | 96 | 0.9307 | 1 | 0.9307 | 0 | 0 | 0 | 0.9575 | Ref |
| 13663 | MZ375233 | Unassigned | Unassigned | Unassigned | 392_0 | Clustered | 126 | VC_392_0 | 126 | 0.9621 | 1 | 0.9621 | 0 | 0 | 0 | 1 | Ref |
| 13664 | MZ375234 | Unassigned | Unassigned | Unassigned | 392_0 | Clustered | 126 | VC_392_0 | 126 | 0.9621 | 1 | 0.9621 | 0 | 0 | 0 | 1 | Ref |
| 13665 | MZ375235 | Unassigned | Unassigned | Unassigned | 392_0 | Clustered | 126 | VC_392_0 | 126 | 0.9621 | 1 | 0.9621 | 0 | 0 | 0 | 1 | Ref |
| 13666 | MZ375236 | Unassigned | Unassigned | Unassigned | 392_0 | Clustered | 126 | VC_392_0 | 126 | 0.9621 | 1 | 0.9621 | 0 | 0 | 0 | 1 | Ref |
| 13667 | MZ375237 | Unassigned | Unassigned | Unassigned | 392_0 | Clustered | 126 | VC_392_0 | 126 | 0.9621 | 1 | 0.9621 | 0 | 0 | 0 | 1 | Ref |
| 13668 | MZ375238 | Unassigned | Unassigned | Unassigned | 392_0 | Clustered | 126 | VC_392_0 | 126 | 0.9621 | 1 | 0.9621 | 0 | 0 | 0 | 1 | Ref |
| 13669 | MZ375239 | Unassigned | Unassigned | Unassigned | 392_0 | Clustered | 126 | VC_392_0 | 126 | 0.9621 | 1 | 0.9621 | 0 | 0 | 0 | 1 | Ref |
| 13670 | MZ375240 | Unassigned | Unassigned | Unassigned | 392_0 | Clustered | 126 | VC_392_0 | 126 | 0.9621 | 1 | 0.9621 | 0 | 0 | 0 | 1 | Ref |
| 13671 | MZ375241 | Unassigned | Unassigned | Unassigned | 392_0 | Clustered | 126 | VC_392_0 | 126 | 0.9621 | 1 | 0.9621 | 0 | 0 | 0 | 1 | Ref |
| 13672 | MZ375242 | Unassigned | Unassigned | Unassigned | 392_0 | Clustered | 126 | VC_392_0 | 126 | 0.9621 | 1 | 0.9621 | 0 | 0 | 0 | 1 | Ref |
| 13673 | MZ375243 | Unassigned | Unassigned | Unassigned | 392_0 | Clustered | 126 | VC_392_0 | 126 | 0.9621 | 1 | 0.9621 | 0 | 0 | 0 | 1 | Ref |
| 13674 | MZ375244 | Unassigned | Unassigned | Unassigned | 392_0 | Clustered | 126 | VC_392_0 | 126 | 0.9621 | 1 | 0.9621 | 0 | 0 | 0 | 1 | Ref |
| 13675 | MZ375245 | Unassigned | Unassigned | Unassigned | 392_0 | Clustered | 126 | VC_392_0 | 126 | 0.9621 | 1 | 0.9621 | 0 | 0 | 0 | 1 | Ref |
| 13676 | MZ375246 | Unassigned | Unassigned | Unassigned | 392_0 | Clustered | 126 | VC_392_0 | 126 | 0.9621 | 1 | 0.9621 | 0 | 0 | 0 | 1 | Ref |
| 13677 | MZ375247 | Unassigned | Unassigned | Unassigned | 392_0 | Clustered | 126 | VC_392_0 | 126 | 0.9621 | 1 | 0.9621 | 0 | 0 | 0 | 1 | Ref |
| 13678 | MZ375248 | Unassigned | Unassigned | Unassigned | 392_0 | Clustered | 126 | VC_392_0 | 126 | 0.9621 | 1 | 0.9621 | 0 | 0 | 0 | 1 | Ref |
| 13679 | MZ375249 | Unassigned | Unassigned | Unassigned | 392_0 | Clustered | 126 | VC_392_0 | 126 | 0.9621 | 1 | 0.9621 | 0 | 0 | 0 | 1 | Ref |
| 13680 | MZ375250 | Unassigned | Unassigned | Unassigned | 392_0 | Clustered | 126 | VC_392_0 | 126 | 0.9621 | 1 | 0.9621 | 0 | 0 | 0 | 1 | Ref |
| 13681 | MZ375251 | Unassigned | Unassigned | Unassigned | 392_0 | Clustered | 126 | VC_392_0 | 126 | 0.9621 | 1 | 0.9621 | 0 | 0 | 0 | 1 | Ref |
| 13682 | MZ375252 | Unassigned | Unassigned | Unassigned | 392_0 | Clustered | 126 | VC_392_0 | 126 | 0.9621 | 1 | 0.9621 | 0 | 0 | 0 | 1 | Ref |
| 13683 | MZ375253 | Unassigned | Unassigned | Unassigned | 392_0 | Clustered | 126 | VC_392_0 | 126 | 0.9621 | 1 | 0.9621 | 0 | 0 | 0 | 1 | Ref |
| 13684 | MZ375254 | Unassigned | Unassigned | Unassigned | 392_0 | Clustered | 126 | VC_392_0 | 126 | 0.9621 | 1 | 0.9621 | 0 | 0 | 0 | 1 | Ref |
| 13685 | MZ375255 | Unassigned | Unassigned | Unassigned | 392_0 | Clustered | 126 | VC_392_0 | 126 | 0.9621 | 1 | 0.9621 | 0 | 0 | 0 | 1 | Ref |
| 13686 | MZ375256 | Unassigned | Unassigned | Unassigned | 392_0 | Clustered | 126 | VC_392_0 | 126 | 0.9621 | 1 | 0.9621 | 0 | 0 | 0 | 1 | Ref |
| 13687 | MZ375257 | Unassigned | Unassigned | Unassigned | 392_0 | Clustered | 126 | VC_392_0 | 126 | 0.9621 | 1 | 0.9621 | 0 | 0 | 0 | 1 | Ref |
| 13688 | MZ375258 | Unassigned | Unassigned | Unassigned | 392_0 | Clustered | 126 | VC_392_0 | 126 | 0.9621 | 1 | 0.9621 | 0 | 0 | 0 | 1 | Ref |
| 13689 | MZ375259 | Unassigned | Unassigned | Unassigned | 392_0 | Clustered | 126 | VC_392_0 | 126 | 0.9621 | 1 | 0.9621 | 0 | 0 | 0 | 1 | Ref |
| 13690 | MZ375260 | Unassigned | Unassigned | Unassigned | 392_0 | Clustered | 126 | VC_392_0 | 126 | 0.9621 | 1 | 0.9621 | 0 | 0 | 0 | 1 | Ref |
| 13691 | MZ375261 | Unassigned | Unassigned | Unassigned | 392_0 | Clustered | 126 | VC_392_0 | 126 | 0.9621 | 1 | 0.9621 | 0 | 0 | 0 | 1 | Ref |
| 13692 | MZ375262 | Unassigned | Unassigned | Unassigned | 392_0 | Clustered | 126 | VC_392_0 | 126 | 0.9621 | 1 | 0.9621 | 0 | 0 | 0 | 1 | Ref |
| 13693 | MZ375263 | Unassigned | Unassigned | Unassigned | 392_0 | Clustered | 126 | VC_392_0 | 126 | 0.9621 | 1 | 0.9621 | 0 | 0 | 0 | 1 | Ref |
| 13694 | MZ375264 | Unassigned | Unassigned | Unassigned | 392_0 | Clustered | 126 | VC_392_0 | 126 | 0.9621 | 1 | 0.9621 | 0 | 0 | 0 | 1 | Ref |
| 13695 | MZ375265 | Unassigned | Unassigned | Unassigned | 392_0 | Clustered | 126 | VC_392_0 | 126 | 0.9621 | 1 | 0.9621 | 0 | 0 | 0 | 1 | Ref |
| 13696 | MZ375266 | Unassigned | Unassigned | Unassigned | 392_0 | Clustered | 126 | VC_392_0 | 126 | 0.9621 | 1 | 0.9621 | 0 | 0 | 0 | 1 | Ref |
| 13697 | MZ375267 | Unassigned | Unassigned | Unassigned | 392_0 | Clustered | 126 | VC_392_0 | 126 | 0.9621 | 1 | 0.9621 | 0 | 0 | 0 | 1 | Ref |
| 13698 | MZ375268 | Unassigned | Unassigned | Unassigned | 392_0 | Clustered | 126 | VC_392_0 | 126 | 0.9621 | 1 | 0.9621 | 0 | 0 | 0 | 1 | Ref |
| 13699 | MZ375269 | Unassigned | Unassigned | Unassigned | 392_0 | Clustered | 126 | VC_392_0 | 126 | 0.9621 | 1 | 0.9621 | 0 | 0 | 0 | 1 | Ref |
| 13700 | MZ375270 | Unassigned | Unassigned | Unassigned | 392_0 | Clustered | 126 | VC_392_0 | 126 | 0.9621 | 1 | 0.9621 | 0 | 0 | 0 | 1 | Ref |
| 13701 | MZ375271 | Unassigned | Unassigned | Unassigned | 392_0 | Clustered | 126 | VC_392_0 | 126 | 0.9621 | 1 | 0.9621 | 0 | 0 | 0 | 1 | Ref |
| 13702 | MZ375272 | Unassigned | Unassigned | Unassigned | 392_0 | Clustered | 126 | VC_392_0 | 126 | 0.9621 | 1 | 0.9621 | 0 | 0 | 0 | 1 | Ref |
| 13703 | MZ375273 | Unassigned | Unassigned | Unassigned | 392_0 | Clustered | 126 | VC_392_0 | 126 | 0.9621 | 1 | 0.9621 | 0 | 0 | 0 | 1 | Ref |
| 13704 | MZ375274 | Unassigned | Unassigned | Unassigned | 392_0 | Clustered | 126 | VC_392_0 | 126 | 0.9621 | 1 | 0.9621 | 0 | 0 | 0 | 1 | Ref |
| 13705 | MZ375275 | Unassigned | Unassigned | Unassigned | 392_0 | Clustered | 126 | VC_392_0 | 126 | 0.9621 | 1 | 0.9621 | 0 | 0 | 0 | 1 | Ref |
| 13706 | MZ375276 | Unassigned | Unassigned | Unassigned | 392_0 | Clustered | 126 | VC_392_0 | 126 | 0.9621 | 1 | 0.9621 | 0 | 0 | 0 | 1 | Ref |
| 13707 | MZ375277 | Unassigned | Unassigned | Unassigned | 392_0 | Clustered | 126 | VC_392_0 | 126 | 0.9621 | 1 | 0.9621 | 0 | 0 | 0 | 1 | Ref |
| 13708 | MZ375278 | Unassigned | Unassigned | Unassigned | 392_0 | Clustered | 126 | VC_392_0 | 126 | 0.9621 | 1 | 0.9621 | 0 | 0 | 0 | 1 | Ref |
| 13709 | MZ375279 | Unassigned | Unassigned | Unassigned | 392_0 | Clustered | 126 | VC_392_0 | 126 | 0.9621 | 1 | 0.9621 | 0 | 0 | 0 | 1 | Ref |
| 13710 | MZ375280 | Unassigned | Unassigned | Unassigned | 392_0 | Clustered | 126 | VC_392_0 | 126 | 0.9621 | 1 | 0.9621 | 0 | 0 | 0 | 1 | Ref |
| 13711 | MZ375281 | Unassigned | Unassigned | Unassigned | 392_0 | Clustered | 126 | VC_392_0 | 126 | 0.9621 | 1 | 0.9621 | 0 | 0 | 0 | 1 | Ref |
| 13712 | MZ375282 | Unassigned | Unassigned | Unassigned | 392_0 | Clustered | 126 | VC_392_0 | 126 | 0.9621 | 1 | 0.9621 | 0 | 0 | 0 | 1 | Ref |
| 13713 | MZ375283 | Unassigned | Unassigned | Unassigned | 392_0 | Clustered | 126 | VC_392_0 | 126 | 0.9621 | 1 | 0.9621 | 0 | 0 | 0 | 1 | Ref |
| 13714 | MZ375284 | Unassigned | Unassigned | Unassigned | 392_0 | Clustered | 126 | VC_392_0 | 126 | 0.9621 | 1 | 0.9621 | 0 | 0 | 0 | 1 | Ref |
| 13715 | MZ375285 | Unassigned | Unassigned | Unassigned | 392_0 | Clustered | 126 | VC_392_0 | 126 | 0.9621 | 1 | 0.9621 | 0 | 0 | 0 | 1 | Ref |
| 13716 | MZ375286 | Unassigned | Unassigned | Unassigned | 392_0 | Clustered | 126 | VC_392_0 | 126 | 0.9621 | 1 | 0.9621 | 0 | 0 | 0 | 1 | Ref |
| 13717 | MZ375287 | Unassigned | Unassigned | Unassigned | 392_0 | Clustered | 126 | VC_392_0 | 126 | 0.9621 | 1 | 0.9621 | 0 | 0 | 0 | 1 | Ref |
| 13718 | MZ375288 | Unassigned | Unassigned | Unassigned | 392_0 | Clustered | 126 | VC_392_0 | 126 | 0.9621 | 1 | 0.9621 | 0 | 0 | 0 | 1 | Ref |
| 13719 | MZ375289 | Unassigned | Unassigned | Unassigned | 392_0 | Clustered | 126 | VC_392_0 | 126 | 0.9621 | 1 | 0.9621 | 0 | 0 | 0 | 1 | Ref |
| 13720 | MZ375290 | Unassigned | Unassigned | Unassigned | 392_0 | Clustered | 126 | VC_392_0 | 126 | 0.9621 | 1 | 0.9621 | 0 | 0 | 0 | 1 | Ref |
| 13721 | MZ375291 | Unassigned | Unassigned | Unassigned | 392_0 | Clustered | 126 | VC_392_0 | 126 | 0.9621 | 1 | 0.9621 | 0 | 0 | 0 | 1 | Ref |
| 13722 | MZ375292 | Unassigned | Unassigned | Unassigned | 392_0 | Clustered | 126 | VC_392_0 | 126 | 0.9621 | 1 | 0.9621 | 0 | 0 | 0 | 1 | Ref |
| 13723 | MZ375293 | Unassigned | Unassigned | Unassigned | 392_0 | Clustered | 126 | VC_392_0 | 126 | 0.9621 | 1 | 0.9621 | 0 | 0 | 0 | 1 | Ref |
| 13724 | MZ375294 | Unassigned | Unassigned | Unassigned | 392_0 | Clustered | 126 | VC_392_0 | 126 | 0.9621 | 1 | 0.9621 | 0 | 0 | 0 | 1 | Ref |
| 13725 | MZ375295 | Unassigned | Unassigned | Unassigned | 461_0 | Clustered | 96 | VC_461_0 | 96 | 0.9307 | 1 | 0.9307 | 0 | 0 | 0 | 0.9575 | Ref |
| 13726 | MZ375296 | Unassigned | Unassigned | Unassigned | 461_0 | Clustered | 96 | VC_461_0 | 96 | 0.9307 | 1 | 0.9307 | 0 | 0 | 0 | 0.9575 | Ref |
| 13727 | MZ375297 | Unassigned | Unassigned | Unassigned | 461_0 | Clustered | 96 | VC_461_0 | 96 | 0.9307 | 1 | 0.9307 | 0 | 0 | 0 | 0.9575 | Ref |
| 13728 | MZ375298 | Unassigned | Unassigned | Unassigned | 461_0 | Clustered | 96 | VC_461_0 | 96 | 0.9307 | 1 | 0.9307 | 0 | 0 | 0 | 0.9575 | Ref |
| 13729 | MZ375299 | Unassigned | Unassigned | Unassigned | 461_0 | Clustered | 96 | VC_461_0 | 96 | 0.9307 | 1 | 0.9307 | 0 | 0 | 0 | 0.9575 | Ref |
| 13730 | MZ375300 | Unassigned | Unassigned | Unassigned | 461_0 | Clustered | 96 | VC_461_0 | 96 | 0.9307 | 1 | 0.9307 | 0 | 0 | 0 | 0.9575 | Ref |
| 13731 | MZ375301 | Unassigned | Unassigned | Unassigned | 461_0 | Clustered | 96 | VC_461_0 | 96 | 0.9307 | 1 | 0.9307 | 0 | 0 | 0 | 0.9575 | Ref |
| 13732 | MZ375302 | Unassigned | Unassigned | Unassigned | 461_0 | Clustered | 96 | VC_461_0 | 96 | 0.9307 | 1 | 0.9307 | 0 | 0 | 0 | 0.9575 | Ref |
| 13733 | MZ375303 | Unassigned | Unassigned | Unassigned | 461_0 | Clustered | 96 | VC_461_0 | 96 | 0.9307 | 1 | 0.9307 | 0 | 0 | 0 | 0.9575 | Ref |
| 13734 | MZ375304 | Unassigned | Unassigned | Unassigned | 461_0 | Clustered | 96 | VC_461_0 | 96 | 0.9307 | 1 | 0.9307 | 0 | 0 | 0 | 0.9575 | Ref |
| 13735 | MZ375305 | Unassigned | Unassigned | Unassigned | 461_0 | Clustered | 96 | VC_461_0 | 96 | 0.9307 | 1 | 0.9307 | 0 | 0 | 0 | 0.9575 | Ref |
| 13736 | MZ375306 | Unassigned | Unassigned | Unassigned | 461_0 | Clustered | 96 | VC_461_0 | 96 | 0.9307 | 1 | 0.9307 | 0 | 0 | 0 | 0.9575 | Ref |
| 13737 | MZ375307 | Unassigned | Unassigned | Unassigned | 461_0 | Clustered | 96 | VC_461_0 | 96 | 0.9307 | 1 | 0.9307 | 0 | 0 | 0 | 0.9575 | Ref |
| 13738 | MZ375308 | Unassigned | Unassigned | Unassigned | 461_0 | Clustered | 96 | VC_461_0 | 96 | 0.9307 | 1 | 0.9307 | 0 | 0 | 0 | 0.9575 | Ref |
| 13739 | MZ375309 | Unassigned | Unassigned | Unassigned | 461_0 | Clustered | 96 | VC_461_0 | 96 | 0.9307 | 1 | 0.9307 | 0 | 0 | 0 | 0.9575 | Ref |
| 13740 | MZ375310 | Unassigned | Unassigned | Unassigned | 461_0 | Clustered | 96 | VC_461_0 | 96 | 0.9307 | 1 | 0.9307 | 0 | 0 | 0 | 0.9575 | Ref |
| 13741 | MZ375311 | Unassigned | Unassigned | Unassigned | 461_0 | Clustered | 96 | VC_461_0 | 96 | 0.9307 | 1 | 0.9307 | 0 | 0 | 0 | 0.9575 | Ref |
| 13742 | MZ375312 | Unassigned | Unassigned | Unassigned | 461_0 | Clustered | 96 | VC_461_0 | 96 | 0.9307 | 1 | 0.9307 | 0 | 0 | 0 | 0.9575 | Ref |
| 13743 | MZ375313 | Unassigned | Unassigned | Unassigned | 461_0 | Clustered | 96 | VC_461_0 | 96 | 0.9307 | 1 | 0.9307 | 0 | 0 | 0 | 0.9575 | Ref |
| 13744 | MZ375314 | Unassigned | Unassigned | Unassigned | 461_0 | Clustered | 96 | VC_461_0 | 96 | 0.9307 | 1 | 0.9307 | 0 | 0 | 0 | 0.9575 | Ref |
| 13745 | MZ375315 | Unassigned | Unassigned | Unassigned | 461_0 | Clustered | 96 | VC_461_0 | 96 | 0.9307 | 1 | 0.9307 | 0 | 0 | 0 | 0.9575 | Ref |
| 13746 | MZ375316 | Unassigned | Unassigned | Unassigned | 461_0 | Clustered | 96 | VC_461_0 | 96 | 0.9307 | 1 | 0.9307 | 0 | 0 | 0 | 0.9575 | Ref |
| 13747 | MZ375317 | Unassigned | Unassigned | Unassigned | 461_0 | Clustered | 96 | VC_461_0 | 96 | 0.9307 | 1 | 0.9307 | 0 | 0 | 0 | 0.9575 | Ref |
| 13748 | MZ375318 | Unassigned | Unassigned | Unassigned | 461_0 | Clustered | 96 | VC_461_0 | 96 | 0.9307 | 1 | 0.9307 | 0 | 0 | 0 | 0.9575 | Ref |
| 13749 | MZ375319 | Unassigned | Unassigned | Unassigned | 461_0 | Clustered | 96 | VC_461_0 | 96 | 0.9307 | 1 | 0.9307 | 0 | 0 | 0 | 0.9575 | Ref |
| 13750 | MZ375320 | Unassigned | Unassigned | Unassigned | 461_0 | Clustered | 96 | VC_461_0 | 96 | 0.9307 | 1 | 0.9307 | 0 | 0 | 0 | 0.9575 | Ref |
| 13751 | MZ375321 | Unassigned | Unassigned | Unassigned | 461_0 | Clustered | 96 | VC_461_0 | 96 | 0.9307 | 1 | 0.9307 | 0 | 0 | 0 | 0.9575 | Ref |
| 13752 | MZ375322 | Unassigned | Unassigned | Unassigned | 461_0 | Clustered | 96 | VC_461_0 | 96 | 0.9307 | 1 | 0.9307 | 0 | 0 | 0 | 0.9575 | Ref |
| 13753 | MZ375323 | Unassigned | Unassigned | Unassigned | 461_0 | Clustered | 96 | VC_461_0 | 96 | 0.9307 | 1 | 0.9307 | 0 | 0 | 0 | 0.9575 | Ref |
| 13754 | MZ375324 | Unassigned | Unassigned | Unassigned | 461_0 | Clustered | 96 | VC_461_0 | 96 | 0.9307 | 1 | 0.9307 | 0 | 0 | 0 | 0.9575 | Ref |
| 13755 | MZ375325 | Unassigned | Unassigned | Unassigned | 461_0 | Clustered | 96 | VC_461_0 | 96 | 0.9307 | 1 | 0.9307 | 0 | 0 | 0 | 0.9575 | Ref |
| 13756 | MZ375326 | Unassigned | Unassigned | Unassigned | 461_0 | Clustered | 96 | VC_461_0 | 96 | 0.9307 | 1 | 0.9307 | 0 | 0 | 0 | 0.9575 | Ref |
| 13757 | MZ375327 | Unassigned | Unassigned | Unassigned | 461_0 | Clustered | 96 | VC_461_0 | 96 | 0.9307 | 1 | 0.9307 | 0 | 0 | 0 | 0.9575 | Ref |
| 13758 | MZ375328 | Unassigned | Unassigned | Unassigned | 461_0 | Clustered | 96 | VC_461_0 | 96 | 0.9307 | 1 | 0.9307 | 0 | 0 | 0 | 0.9575 | Ref |
| 13759 | MZ375329 | Unassigned | Unassigned | Unassigned | 461_0 | Clustered | 96 | VC_461_0 | 96 | 0.9307 | 1 | 0.9307 | 0 | 0 | 0 | 0.9575 | Ref |
| 13760 | MZ375330 | Unassigned | Unassigned | Unassigned | 461_0 | Clustered | 96 | VC_461_0 | 96 | 0.9307 | 1 | 0.9307 | 0 | 0 | 0 | 0.9575 | Ref |
| 13761 | MZ375331 | Unassigned | Unassigned | Unassigned | 461_0 | Clustered | 96 | VC_461_0 | 96 | 0.9307 | 1 | 0.9307 | 0 | 0 | 0 | 0.9575 | Ref |
| 13762 | MZ375332 | Unassigned | Unassigned | Unassigned | 461_0 | Clustered | 96 | VC_461_0 | 96 | 0.9307 | 1 | 0.9307 | 0 | 0 | 0 | 0.9575 | Ref |
| 13763 | MZ375333 | Unassigned | Unassigned | Unassigned | 461_0 | Clustered | 96 | VC_461_0 | 96 | 0.9307 | 1 | 0.9307 | 0 | 0 | 0 | 0.9575 | Ref |
| 13764 | MZ375334 | Unassigned | Unassigned | Unassigned | 461_0 | Clustered | 96 | VC_461_0 | 96 | 0.9307 | 1 | 0.9307 | 0 | 0 | 0 | 0.9575 | Ref |
| 13765 | MZ375335 | Unassigned | Unassigned | Unassigned | 461_0 | Clustered | 96 | VC_461_0 | 96 | 0.9307 | 1 | 0.9307 | 0 | 0 | 0 | 0.9575 | Ref |
| 13766 | MZ375336 | Unassigned | Unassigned | Unassigned | 461_0 | Clustered | 96 | VC_461_0 | 96 | 0.9307 | 1 | 0.9307 | 0 | 0 | 0 | 0.9575 | Ref |
| 13767 | MZ375337 | Unassigned | Unassigned | Unassigned | 461_0 | Clustered | 96 | VC_461_0 | 96 | 0.9307 | 1 | 0.9307 | 0 | 0 | 0 | 0.9575 | Ref |
| 13768 | MZ375338 | Unassigned | Unassigned | Unassigned | 461_0 | Clustered | 96 | VC_461_0 | 96 | 0.9307 | 1 | 0.9307 | 0 | 0 | 0 | 0.9575 | Ref |

| | | | | | | | | | | | | | | | | | |
|---|---|---|---|---|---|---|---|---|---|---|---|---|---|---|---|---|---|
| 13769 | MZ375339 | Unassigned | Unassigned | Unassigned | 461_0 | Clustered | 96 | VC_461_0 | 96 | 0.9307 | 1 | 0.9307 | 0 | 0 | 0 | 0.9575 | Ref |
| 13770 | MZ375340 | Unassigned | Unassigned | Unassigned | 461_0 | Clustered | 96 | VC_461_0 | 96 | 0.9307 | 1 | 0.9307 | 0 | 0 | 0 | 0.9575 | Ref |
| 13771 | MZ375341 | Unassigned | Unassigned | Unassigned | 461_0 | Clustered | 96 | VC_461_0 | 96 | 0.9307 | 1 | 0.9307 | 0 | 0 | 0 | 0.9575 | Ref |
| 13772 | MZ375342 | Unassigned | Unassigned | Unassigned | 461_0 | Clustered | 96 | VC_461_0 | 96 | 0.9307 | 1 | 0.9307 | 0 | 0 | 0 | 0.9575 | Ref |
| 13773 | MZ375343 | Unassigned | Unassigned | Unassigned | 461_0 | Clustered | 96 | VC_461_0 | 96 | 0.9307 | 1 | 0.9307 | 0 | 0 | 0 | 0.9575 | Ref |
| 13774 | MZ375344 | Unassigned | Unassigned | Unassigned | 461_0 | Clustered | 96 | VC_461_0 | 96 | 0.9307 | 1 | 0.9307 | 0 | 0 | 0 | 0.9575 | Ref |
| 13775 | MZ375345 | Unassigned | Unassigned | Unassigned | 461_0 | Clustered | 96 | VC_461_0 | 96 | 0.9307 | 1 | 0.9307 | 0 | 0 | 0 | 0.9575 | Ref |
| 13776 | MZ375346 | Unassigned | Unassigned | Unassigned | 461_0 | Clustered | 96 | VC_461_0 | 96 | 0.9307 | 1 | 0.9307 | 0 | 0 | 0 | 0.9575 | Ref |
| 13777 | MZ375347 | Unassigned | Unassigned | Unassigned | 461_0 | Clustered | 96 | VC_461_0 | 96 | 0.9307 | 1 | 0.9307 | 0 | 0 | 0 | 0.9575 | Ref |
| 13778 | MZ375348 | Unassigned | Unassigned | Unassigned | 461_0 | Clustered | 96 | VC_461_0 | 96 | 0.9307 | 1 | 0.9307 | 0 | 0 | 0 | 0.9575 | Ref |
| 13779 | MZ375349 | Unassigned | Unassigned | Unassigned | 461_0 | Clustered | 96 | VC_461_0 | 96 | 0.9307 | 1 | 0.9307 | 0 | 0 | 0 | 0.9575 | Ref |
| 13780 | MZ375350 | Unassigned | Unassigned | Unassigned | 461_0 | Clustered | 96 | VC_461_0 | 96 | 0.9307 | 1 | 0.9307 | 0 | 0 | 0 | 0.9575 | Ref |
| 13781 | MZ375351 | Unassigned | Unassigned | Unassigned | 461_0 | Clustered | 96 | VC_461_0 | 96 | 0.9307 | 1 | 0.9307 | 0 | 0 | 0 | 0.9575 | Ref |
| 13782 | MZ375352 | Unassigned | Unassigned | Unassigned | 461_0 | Clustered | 96 | VC_461_0 | 96 | 0.9307 | 1 | 0.9307 | 0 | 0 | 0 | 0.9575 | Ref |
| 13783 | MZ375353 | Unassigned | Unassigned | Unassigned | 461_0 | Clustered | 96 | VC_461_0 | 96 | 0.9307 | 1 | 0.9307 | 0 | 0 | 0 | 0.9575 | Ref |
| 13784 | MZ375354 | Unassigned | Unassigned | Unassigned | 461_0 | Clustered | 96 | VC_461_0 | 96 | 0.9307 | 1 | 0.9307 | 0 | 0 | 0 | 0.9575 | Ref |
| 13785 | MZ375355 | Unassigned | Unassigned | Unassigned | 461_0 | Clustered | 96 | VC_461_0 | 96 | 0.9307 | 1 | 0.9307 | 0 | 0 | 0 | 0.9575 | Ref |
| 13786 | MZ375356 | Unassigned | Unassigned | Unassigned | 283_91 | Clustered | 11 | VC_283_91 | 11 | 0.0285 | 1 | 0.0285 | 0 | 0 | 0 | 1 | Ref |
| 13787 | MZ375357 | Unassigned | Unassigned | Unassigned | 133_10 | Clustered | 10 | VC_133_10 | 10 | 0.0527 | 1 | 0.0527 | 0 | 0 | 0 | 1 | Ref |
| 13788 | MZ375358 | Unassigned | Unassigned | Unassigned | 133_10 | Clustered | 10 | VC_133_10 | 10 | 0.0527 | 1 | 0.0527 | 0 | 0 | 0 | 1 | Ref |
| 13789 | MZ375754 | Unassigned | Unassigned | Unassigned | 3_0 | Clustered | 17 | VC_3_0 | 17 | 0.5639 | 1 | 0.5639 | 0 | 0 | 0 | 1 | Ref |
| 13790 | MZ384014 | Unassigned | Unassigned | Unassigned | 818_0 | Clustered | 20 | VC_818_0 | 20 | 0.9601 | 1 | 0.9601 | 0 | 0 | 0 | 1 | Ref |
| 13791 | MZ388551 | Unassigned | Unassigned | Unassigned | 558_58 | Clustered/Singleton | 1 | VC_558_58 | 1 | 0 | 1 | 0 | 0 | 0 | 0 | 1 | Ref |
| 13792 | MZ388552 | Unassigned | Unassigned | Unassigned | 1181_1 | Clustered | 10 | VC_1181_1 | 10 | 0.5828 | 1 | 0.5828 | 0 | 0 | 0 | 1 | Ref |
| 13793 | MZ388553 | Unassigned | Unassigned | Unassigned | 557_1 | Clustered | 11 | VC_557_1 | 11 | 0.4945 | 1 | 0.4945 | 0 | 0 | 0 | 0.7818 | Ref |
| 13794 | MZ388554 | Unassigned | Unassigned | Unassigned | 1360_0 | Clustered | 31 | VC_1360_0 | 31 | 0.8837 | 1 | 0.8837 | 0 | 0 | 0 | 1 | Ref |
| 13795 | MZ388556 | Unassigned | Unassigned | Unassigned | 799_1 | Clustered | 27 | VC_799_1 | 27 | 0.8611 | 1 | 0.8611 | 0 | 0 | 0 | 1 | Ref |
| 13796 | MZ394712 | Unassigned | Unassigned | Unassigned | 779_0 | Clustered | 37 | VC_779_0 | 37 | 0.5682 | 1 | 0.5682 | 0 | 0 | 0 | 1 | Ref |
| 13797 | MZ398021 | Unassigned | Unassigned | Unassigned | 123_0 | Clustered | 23 | VC_123_0 | 23 | 0.817 | 1 | 0.817 | 0 | 0 | 0 | 1 | Ref |
| 13798 | MZ398135 | Unassigned | Unassigned | Unassigned | 109_0 | Clustered | 3 | VC_109_0 | 3 | 0.1706 | 0.99835623 | 0.1703 | 0 | 0 | 0 | 1 | Ref |
| 13799 | MZ398136 | Unassigned | Unassigned | Unassigned | 109_0 | Clustered | 3 | VC_109_0 | 3 | 0.1706 | 0.99835623 | 0.1703 | 0 | 0 | 0 | 1 | Ref |
| 13800 | MZ398240 | Unassigned | Unassigned | Unassigned | 671_0 | Clustered | 5 | VC_671_0 | 5 | 0.5289 | 0.99999996 | 0.5289 | 0 | 0 | 0 | 1 | Ref |
| 13801 | MZ398241 | Unassigned | Unassigned | Unassigned | 671_0 | Clustered | 5 | VC_671_0 | 5 | 0.5289 | 0.99999996 | 0.5289 | 0 | 0 | 0 | 1 | Ref |
| 13802 | MZ398242 | Unassigned | Unassigned | Unassigned | 938_0 | Clustered | 99 | VC_938_0 | 99 | 0.9119 | 1 | 0.9119 | 0 | 0 | 0 | 0.9928 | Ref |
| 13803 | MZ398243 | Unassigned | Unassigned | Unassigned | 813_0 | Clustered | 12 | VC_813_0 | 12 | 0.2347 | 1 | 0.2347 | 0 | 0 | 0 | 1 | Ref |
| 13804 | MZ398244 | Unassigned | Unassigned | Unassigned | 452_0 | Clustered | 66 | VC_452_0 | 66 | 0.9248 | 1 | 0.9248 | 0 | 0 | 0 | 1 | Ref |
| 13805 | MZ398245 | Unassigned | Unassigned | Unassigned | 123_0 | Clustered | 23 | VC_123_0 | 23 | 0.817 | 1 | 0.817 | 0 | 0 | 0 | 1 | Ref |
| 13806 | MZ398246 | Unassigned | Unassigned | Unassigned | 190_24 | Clustered | 5 | VC_190_24 | 5 | 0.0293 | 0.9999996 | 0.0293 | 0 | 0 | 0 | 0.7 | Ref |
| 13807 | MZ398247 | Unassigned | Unassigned | Unassigned | 451_0 | Clustered | 80 | VC_451_0 | 80 | 0.9991 | 1 | 0.9991 | 0 | 0 | 0 | 1 | Ref |
| 13808 | MZ398248 | Unassigned | Unassigned | Unassigned | 506_0 | Clustered | 8 | VC_506_0 | 8 | 0.7376 | 1 | 0.7376 | 0 | 0 | 0 | 1 | Ref |
| 13809 | MZ399596 | Unassigned | Unassigned | Unassigned | 109_0 | Clustered | 3 | VC_109_0 | 3 | 0.1706 | 0.99835623 | 0.1703 | 0 | 0 | 0 | 1 | Ref |
| 13810 | MZ401005 | Unassigned | Unassigned | Unassigned | 898_0 | Clustered | 10 | VC_898_0 | 10 | 0.7284 | 1 | 0.7284 | 0 | 0 | 0 | 1 | Ref |
| 13811 | MZ401006 | Unassigned | Unassigned | Unassigned | 815_0 | Clustered | 8 | VC_815_0 | 8 | 0.9801 | 1 | 0.9801 | 0 | 0 | 0 | 1 | Ref |
| 13812 | MZ401007 | Unassigned | Unassigned | Unassigned | 898_0 | Clustered | 10 | VC_898_0 | 10 | 0.7284 | 1 | 0.7284 | 0 | 0 | 0 | 1 | Ref |
| 13813 | MZ401008 | Unassigned | Unassigned | Unassigned | 898_0 | Clustered | 10 | VC_898_0 | 10 | 0.7284 | 1 | 0.7284 | 0 | 0 | 0 | 1 | Ref |
| 13814 | MZ401009 | Unassigned | Unassigned | Unassigned | 898_0 | Clustered | 10 | VC_898_0 | 10 | 0.7284 | 1 | 0.7284 | 0 | 0 | 0 | 1 | Ref |
| 13815 | MZ402608 | Unassigned | Unassigned | Unassigned | 956_0 | Clustered | 39 | VC_956_0 | 39 | 1 | 1 | 1 | 0 | 0 | 0 | 1 | Ref |
| 13816 | MZ417315 | Unassigned | Unassigned | Unassigned | 149_25 | Clustered | 14 | VC_149_25 | 14 | 0.095 | 1 | 0.095 | 0 | 0 | 0 | 0.8791 | Ref |
| 13817 | MZ417319 | Unassigned | Unassigned | Unassigned | 149_25 | Clustered | 14 | VC_149_25 | 14 | 0.095 | 1 | 0.095 | 0 | 0 | 0 | 0.8791 | Ref |
| 13818 | MZ417320 | Unassigned | Unassigned | Unassigned | 149_25 | Clustered | 14 | VC_149_25 | 14 | 0.095 | 1 | 0.095 | 0 | 0 | 0 | 0.8791 | Ref |
| 13819 | MZ417321 | Unassigned | Unassigned | Unassigned | 149_25 | Clustered | 14 | VC_149_25 | 14 | 0.095 | 1 | 0.095 | 0 | 0 | 0 | 0.8791 | Ref |
| 13820 | MZ417322 | Unassigned | Unassigned | Unassigned | 149_24 | Clustered/Singleton | 1 | VC_149_24 | 1 | 0 | 1 | 0 | 0 | 0 | 0 | 1 | Ref |
| 13821 | MZ417323 | Unassigned | Unassigned | Unassigned | 150_0 | Clustered | 13 | VC_150_0 | 13 | 0.2417 | 1 | 0.2417 | 0 | 0 | 0 | 0.9872 | Ref |
| 13822 | MZ417325 | Unassigned | Unassigned | Unassigned | 150_0 | Clustered | 13 | VC_150_0 | 13 | 0.2417 | 1 | 0.2417 | 0 | 0 | 0 | 0.9872 | Ref |
| 13823 | MZ417326 | Unassigned | Unassigned | Unassigned | 152_1 | Clustered/Singleton | 1 | VC_152_1 | 1 | 0 | 1 | 0 | 0 | 0 | 0 | 1 | Ref |
| 13824 | MZ417327 | Unassigned | Unassigned | Unassigned | 149_25 | Clustered | 14 | VC_149_25 | 14 | 0.095 | 1 | 0.095 | 0 | 0 | 0 | 0.8791 | Ref |
| 13825 | MZ417328 | Unassigned | Unassigned | Unassigned | 149_25 | Clustered | 14 | VC_149_25 | 14 | 0.095 | 1 | 0.095 | 0 | 0 | 0 | 0.8791 | Ref |
| 13826 | MZ417329 | Unassigned | Unassigned | Unassigned | 151_0 | Clustered | 6 | VC_151_0 | 6 | 0.0735 | 1 | 0.0735 | 0 | 0 | 0 | 1 | Ref |
| 13827 | MZ417330 | Unassigned | Unassigned | Unassigned | 151_0 | Clustered | 6 | VC_151_0 | 6 | 0.0735 | 1 | 0.0735 | 0 | 0 | 0 | 1 | Ref |
| 13828 | MZ417331 | Unassigned | Unassigned | Unassigned | 149_26 | Clustered/Singleton | 1 | VC_149_26 | 1 | 0 | 1 | 0 | 0 | 0 | 0 | 1 | Ref |
| 13829 | MZ417332 | Unassigned | Unassigned | Unassigned | 149_25 | Clustered | 14 | VC_149_25 | 14 | 0.095 | 1 | 0.095 | 0 | 0 | 0 | 0.8791 | Ref |
| 13830 | MZ417336 | Unassigned | Unassigned | Unassigned | 667_0 | Clustered | 7 | VC_667_0 | 7 | 0.4101 | 1 | 0.4101 | 0 | 0 | 0 | 1 | Ref |
| 13831 | MZ417337 | Unassigned | Unassigned | Unassigned | 149_25 | Clustered | 14 | VC_149_25 | 14 | 0.095 | 1 | 0.095 | 0 | 0 | 0 | 0.8791 | Ref |
| 13832 | MZ417339 | Unassigned | Unassigned | Unassigned | 667_0 | Clustered | 7 | VC_667_0 | 7 | 0.4101 | 1 | 0.4101 | 0 | 0 | 0 | 1 | Ref |
| 13833 | MZ417340 | Unassigned | Unassigned | Unassigned | 149_25 | Clustered | 14 | VC_149_25 | 14 | 0.095 | 1 | 0.095 | 0 | 0 | 0 | 0.8791 | Ref |
| 13834 | MZ417343 | Unassigned | Unassigned | Unassigned | 667_0 | Clustered | 7 | VC_667_0 | 7 | 0.4101 | 1 | 0.4101 | 0 | 0 | 0 | 1 | Ref |
| 13835 | MZ417344 | Unassigned | Unassigned | Unassigned | 151_0 | Clustered | 6 | VC_151_0 | 6 | 0.0735 | 1 | 0.0735 | 0 | 0 | 0 | 1 | Ref |
| 13836 | MZ417345 | Unassigned | Unassigned | Unassigned | 149_25 | Clustered | 14 | VC_149_25 | 14 | 0.095 | 1 | 0.095 | 0 | 0 | 0 | 0.8791 | Ref |
| 13837 | MZ417346 | Unassigned | Unassigned | Unassigned | 149_25 | Clustered | 14 | VC_149_25 | 14 | 0.095 | 1 | 0.095 | 0 | 0 | 0 | 0.8791 | Ref |
| 13838 | MZ417347 | Unassigned | Unassigned | Unassigned | 149_25 | Clustered | 14 | VC_149_25 | 14 | 0.095 | 1 | 0.095 | 0 | 0 | 0 | 0.8791 | Ref |
| 13839 | MZ417348 | Unassigned | Unassigned | Unassigned | 667_0 | Clustered | 7 | VC_667_0 | 7 | 0.4101 | 1 | 0.4101 | 0 | 0 | 0 | 1 | Ref |
| 13840 | MZ417350 | Unassigned | Unassigned | Unassigned | 149_25 | Clustered | 14 | VC_149_25 | 14 | 0.095 | 1 | 0.095 | 0 | 0 | 0 | 0.8791 | Ref |
| 13841 | MZ417354 | Unassigned | Unassigned | Unassigned | 149_25 | Clustered | 14 | VC_149_25 | 14 | 0.095 | 1 | 0.095 | 0 | 0 | 0 | 0.8791 | Ref |
| 13842 | MZ417521 | Unassigned | Unassigned | Unassigned | 283_174 | Clustered/Singleton | 1 | VC_283_174 | 1 | 0 | 1 | 0 | 0 | 0 | 0 | 1 | Ref |
| 13843 | MZ420150 | Unassigned | Unassigned | Unassigned | 91_0 | Clustered | 22 | VC_91_0 | 22 | 0.5318 | 1 | 0.5318 | 0 | 0 | 0 | 1 | Ref |
| 13844 | MZ420230 | Unassigned | Unassigned | Unassigned | 228_0 | Clustered | 60 | VC_228_0 | 60 | 0.942 | 1 | 0.942 | 0 | 0 | 0 | 1 | Ref |
| 13845 | MZ424863 | Unassigned | Unassigned | Unassigned | 426_1 | Clustered | 3 | VC_426_1 | 3 | 0.1152 | 0.99804615 | 0.1149 | 0 | 0 | 0 | 1 | Ref |
| 13846 | MZ424864 | Unassigned | Unassigned | Unassigned | 111_0 | Clustered | 91 | VC_111_0 | 91 | 0.9587 | 1 | 0.9587 | 0 | 0 | 0 | 1 | Ref |
| 13847 | MZ424865 | Unassigned | Unassigned | Unassigned | 823_0 | Clustered | 124 | VC_823_0 | 124 | 0.9492 | 1 | 0.9492 | 0 | 0 | 0 | 1 | Ref |
| 13848 | MZ427930 | Unassigned | Unassigned | Unassigned | 149_15 | Clustered | 16 | VC_149_15 | 16 | 0.0788 | 1 | 0.0788 | 0 | 0 | 0 | 0.8083 | Ref |
| 13849 | MZ428221 | Unassigned | Unassigned | Unassigned | 937_0 | Clustered | 40 | VC_937_0 | 40 | 0.6809 | 1 | 0.6809 | 0 | 0 | 0 | 0.9359 | Ref |
| 13850 | MZ428222 | Unassigned | Unassigned | Unassigned | 938_0 | Clustered | 99 | VC_938_0 | 99 | 0.9119 | 1 | 0.9119 | 0 | 0 | 0 | 0.9928 | Ref |
| 13851 | MZ428223 | Unassigned | Unassigned | Unassigned | 938_0 | Clustered | 99 | VC_938_0 | 99 | 0.9119 | 1 | 0.9119 | 0 | 0 | 0 | 0.9928 | Ref |
| 13852 | MZ428224 | Unassigned | Unassigned | Unassigned | 937_0 | Clustered | 40 | VC_937_0 | 40 | 0.6809 | 1 | 0.6809 | 0 | 0 | 0 | 0.9359 | Ref |
| 13853 | MZ428225 | Unassigned | Unassigned | Unassigned | 938_0 | Clustered | 99 | VC_938_0 | 99 | 0.9119 | 1 | 0.9119 | 0 | 0 | 0 | 0.9928 | Ref |
| 13854 | MZ428226 | Unassigned | Unassigned | Unassigned | 938_0 | Clustered | 99 | VC_938_0 | 99 | 0.9119 | 1 | 0.9119 | 0 | 0 | 0 | 0.9928 | Ref |
| 13855 | MZ428227 | Unassigned | Unassigned | Unassigned | 938_0 | Clustered | 99 | VC_938_0 | 99 | 0.9119 | 1 | 0.9119 | 0 | 0 | 0 | 0.9928 | Ref |
| 13856 | MZ428228 | Unassigned | Unassigned | Unassigned | 938_0 | Clustered | 99 | VC_938_0 | 99 | 0.9119 | 1 | 0.9119 | 0 | 0 | 0 | 0.9928 | Ref |
| 13857 | MZ428229 | Unassigned | Unassigned | Unassigned | 385_0 | Clustered | 51 | VC_385_0 | 51 | 0.8754 | 1 | 0.8754 | 0 | 0 | 0 | 1 | Ref |
| 13858 | MZ436830 | Unassigned | Unassigned | Unassigned | 283_32 | Clustered/Singleton | 1 | VC_283_32 | 1 | 0 | 1 | 0 | 0 | 0 | 0 | 1 | Ref |
| 13859 | MZ440881 | Unassigned | Unassigned | Unassigned | 460_0 | Clustered | 19 | VC_460_0 | 19 | 0.3744 | 1 | 0.3744 | 0 | 0 | 0 | 1 | Ref |
| 13860 | MZ443769 | Unassigned | Unassigned | Unassigned | 67_2 | Clustered | 2 | VC_67_2 | 2 | 0.014 | 0.69357611 | 0.0097 | 0 | 0 | 0 | 1 | Ref |
| 13861 | MZ443770 | Unassigned | Unassigned | Unassigned | 67_3 | Clustered | 20 | VC_67_3 | 20 | 0.3542 | 1 | 0.3542 | 0 | 0 | 0 | 1 | Ref |
| 13862 | MZ443771 | Unassigned | Unassigned | Unassigned | 67_3 | Clustered | 20 | VC_67_3 | 20 | 0.3542 | 1 | 0.3542 | 0 | 0 | 0 | 1 | Ref |
| 13863 | MZ443772 | Unassigned | Unassigned | Unassigned | 67_3 | Clustered | 20 | VC_67_3 | 20 | 0.3542 | 1 | 0.3542 | 0 | 0 | 0 | 1 | Ref |
| 13864 | MZ443773 | Unassigned | Unassigned | Unassigned | 67_3 | Clustered | 20 | VC_67_3 | 20 | 0.3542 | 1 | 0.3542 | 0 | 0 | 0 | 1 | Ref |
| 13865 | MZ443774 | Unassigned | Unassigned | Unassigned | 67_3 | Clustered | 20 | VC_67_3 | 20 | 0.3542 | 1 | 0.3542 | 0 | 0 | 0 | 1 | Ref |
| 13866 | MZ443776 | Unassigned | Unassigned | Unassigned | 67_3 | Clustered | 20 | VC_67_3 | 20 | 0.3542 | 1 | 0.3542 | 0 | 0 | 0 | 1 | Ref |
| 13867 | MZ443777 | Unassigned | Unassigned | Unassigned | 67_3 | Clustered | 20 | VC_67_3 | 20 | 0.3542 | 1 | 0.3542 | 0 | 0 | 0 | 1 | Ref |
| 13868 | MZ443778 | Unassigned | Unassigned | Unassigned | 67_3 | Clustered | 20 | VC_67_3 | 20 | 0.3542 | 1 | 0.3542 | 0 | 0 | 0 | 1 | Ref |
| 13869 | MZ443779 | Unassigned | Unassigned | Unassigned | 67_3 | Clustered | 20 | VC_67_3 | 20 | 0.3542 | 1 | 0.3542 | 0 | 0 | 0 | 1 | Ref |
| 13870 | MZ443780 | Unassigned | Unassigned | Unassigned | 67_3 | Clustered | 20 | VC_67_3 | 20 | 0.3542 | 1 | 0.3542 | 0 | 0 | 0 | 1 | Ref |
| 13871 | MZ443782 | Unassigned | Unassigned | Unassigned | 67_3 | Clustered | 20 | VC_67_3 | 20 | 0.3542 | 1 | 0.3542 | 0 | 0 | 0 | 1 | Ref |
| 13872 | MZ443783 | Unassigned | Unassigned | Unassigned | 67_3 | Clustered | 20 | VC_67_3 | 20 | 0.3542 | 1 | 0.3542 | 0 | 0 | 0 | 1 | Ref |
| 13873 | MZ443784 | Unassigned | Unassigned | Unassigned | 67_3 | Clustered | 20 | VC_67_3 | 20 | 0.3542 | 1 | 0.3542 | 0 | 0 | 0 | 1 | Ref |
| 13874 | MZ443785 | Unassigned | Unassigned | Unassigned | 67_3 | Clustered | 20 | VC_67_3 | 20 | 0.3542 | 1 | 0.3542 | 0 | 0 | 0 | 1 | Ref |
| 13875 | MZ443786 | Unassigned | Unassigned | Unassigned | 67_3 | Clustered | 20 | VC_67_3 | 20 | 0.3542 | 1 | 0.3542 | 0 | 0 | 0 | 1 | Ref |
| 13876 | MZ443787 | Unassigned | Unassigned | Unassigned | 67_3 | Clustered | 20 | VC_67_3 | 20 | 0.3542 | 1 | 0.3542 | 0 | 0 | 0 | 1 | Ref |
| 13877 | MZ443788 | Unassigned | Unassigned | Unassigned | 67_3 | Clustered | 20 | VC_67_3 | 20 | 0.3542 | 1 | 0.3542 | 0 | 0 | 0 | 1 | Ref |
| 13878 | MZ443789 | Unassigned | Unassigned | Unassigned | 67_3 | Clustered | 20 | VC_67_3 | 20 | 0.3542 | 1 | 0.3542 | 0 | 0 | 0 | 1 | Ref |
| 13879 | MZ444140 | Unassigned | Unassigned | Unassigned | 344_0 | Clustered | 21 | VC_344_0 | 21 | 0.5436 | 1 | 0.5436 | 0 | 0 | 0 | 1 | Ref |
| 13880 | MZ447863 | Unassigned | Unassigned | Unassigned | 1465_0 | Clustered | 2 | VC_1465_0 | 2 | 1 | 1 | 1 | 0 | 0 | 0 | 1 | Ref |
| 13881 | MZ456034 | Unassigned | Unassigned | Unassigned | 223_0 | Clustered | 38 | VC_223_0 | 38 | 0.9095 | 1 | 0.9095 | 0 | 0 | 0 | 1 | Ref |
| 13882 | MZ456035 | Unassigned | Unassigned | Unassigned | 856_0 | Clustered | 7 | VC_856_0 | 7 | 0.3463 | 1 | 0.3463 | 0 | 0 | 0 | 1 | Ref |
| 13883 | MZ456036 | Unassigned | Unassigned | Unassigned | 508_0 | Clustered | 32 | VC_508_0 | 32 | 0.906 | 1 | 0.906 | 0 | 0 | 0 | 1 | Ref |
| 13884 | MZ456037 | Unassigned | Unassigned | Unassigned | 1348_0 | Clustered | 5 | VC_1348_0 | 5 | 0.5462 | 0.99999985 | 0.5462 | 0 | 0 | 0 | 1 | Ref |
| 13885 | MZ462995 | Unassigned | Unassigned | Unassigned | 987_0 | Clustered | 22 | VC_987_0 | 22 | 0.8955 | 1 | 0.8955 | 0 | 0 | 0 | 1 | Ref |
| 13886 | MZ474488 | Unassigned | Unassigned | Unassigned | 1466_0 | Clustered | 2 | VC_1466_0 | 2 | 1 | 1 | 1 | 0 | 0 | 0 | 1 | Ref |
| 13887 | MZ474489 | Unassigned | Unassigned | Unassigned | 1466_0 | Clustered | 2 | VC_1466_0 | 2 | 1 | 1 | 1 | 0 | 0 | 0 | 1 | Ref |
| 13888 | MZ475891 | Unassigned | Unassigned | Unassigned | 67_1 | Clustered | 8 | VC_67_1 | 8 | 0.1067 | 0.99893246 | 0.1065 | 0 | 0 | 0 | 1 | Ref |
| 13889 | MZ475892 | Unassigned | Unassigned | Unassigned | 67_1 | Clustered | 8 | VC_67_1 | 8 | 0.1067 | 0.99893246 | 0.1065 | 0 | 0 | 0 | 1 | Ref |
| 13890 | MZ475893 | Unassigned | Unassigned | Unassigned | 67_1 | Clustered | 8 | VC_67_1 | 8 | 0.1067 | 0.99893246 | 0.1065 | 0 | 0 | 0 | 1 | Ref |
| 13891 | MZ475894 | Unassigned | Unassigned | Unassigned | 67_1 | Clustered | 8 | VC_67_1 | 8 | 0.1067 | 0.99893246 | 0.1065 | 0 | 0 | 0 | 1 | Ref |
| 13892 | MZ475895 | Unassigned | Unassigned | Unassigned | 67_1 | Clustered | 8 | VC_67_1 | 8 | 0.1067 | 0.99893246 | 0.1065 | 0 | 0 | 0 | 1 | Ref |
| 13893 | MZ475896 | Unassigned | Unassigned | Unassigned | 67_1 | Clustered | 8 | VC_67_1 | 8 | 0.1067 | 0.99893246 | 0.1065 | 0 | 0 | 0 | 1 | Ref |
| 13894 | MZ475897 | Unassigned | Unassigned | Unassigned | 67_1 | Clustered | 8 | VC_67_1 | 8 | 0.1067 | 0.99893246 | 0.1065 | 0 | 0 | 0 | 1 | Ref |
| 13895 | MZ475898 | Unassigned | Unassigned | Unassigned | 67_1 | Clustered | 8 | VC_67_1 | 8 | 0.1067 | 0.99893246 | 0.1065 | 0 | 0 | 0 | 1 | Ref |
| 13896 | MZ477002 | Unassigned | Unassigned | Unassigned | 701_0 | Clustered | 2 | VC_701_0 | 2 | 0.1487 | 0.95249915 | 0.1417 | 0 | 0 | 0 | 1 | Ref |
| 13897 | MZ489634 | Unassigned | Unassigned | Unassigned | 461_0 | Clustered | 96 | VC_461_0 | 96 | 0.9307 | 1 | 0.9307 | 0 | 0 | 0 | 0.9575 | Ref |
| 13898 | MZ496293 | Unassigned | Unassigned | Unassigned | 231_0 | Clustered | 3 | VC_231_0 | 3 | 0.236 | 0.99824503 | 0.2356 | 0 | 0 | 0 | 1 | Ref |
| 13899 | MZ501046 | Unassigned | Unassigned | Unassigned | 283_41 | Clustered/Singleton | 1 | VC_283_41 | 1 | 0 | 1 | 0 | 0 | 0 | 0 | 1 | Ref |
| 13900 | MZ501047 | Unassigned | Unassigned | Unassigned | 286_0 | Clustered | 2 | VC_286_0 | 2 | 0.0073 | 0.77462993 | 0.0056 | 0 | 0 | 0 | 1 | Ref |
| 13901 | MZ501048 | Unassigned | Unassigned | Unassigned | 772_11 | Clustered | 6 | VC_772_11 | 6 | 0.0305 | 0.84253803 | 0.0257 | 0 | 0 | 0 | 0.8 | Ref |
| 13902 | MZ501049 | Unassigned | Unassigned | Unassigned | 779_0 | Clustered | 37 | VC_779_0 | 37 | 0.5682 | 1 | 0.5682 | 0 | 0 | 0 | 1 | Ref |
| 13903 | MZ501050 | Unassigned | Unassigned | Unassigned | 283_42 | Clustered/Singleton | 1 | VC_283_42 | 1 | 0 | 1 | 0 | 0 | 0 | 0 | 1 | Ref |
| 13904 | MZ501051 | Unassigned | Unassigned | Unassigned | 456_0 | Clustered | 62 | VC_456_0 | 62 | 0.9026 | 1 | 0.9026 | 0 | 0 | 0 | 1 | Ref |
| 13905 | MZ501052 | Unassigned | Unassigned | Unassigned | 283_9 | Clustered/Singleton | 1 | VC_283_9 | 1 | 0 | 1 | 0 | 0 | 0 | 0 | 1 | Ref |
| 13906 | MZ501053 | Unassigned | Unassigned | Unassigned | 576_0 | Clustered | 4 | VC_576_0 | 4 | 0.0606 | 0.99998766 | 0.0606 | 0 | 0 | 0 | 1 | Ref |
| 13907 | MZ501054 | Unassigned | Unassigned | Unassigned | 772_19 | Clustered | 16 | VC_772_19 | 16 | 0.0975 | 0.9970466 | 0.0972 | 0 | 0 | 0 | 0.8083 | Ref |
| 13908 | MZ501055 | Unassigned | Unassigned | Unassigned | 386_0 | Clustered | 49 | VC_386_0 | 49 | 0.7941 | 1 | 0.7941 | 0 | 0 | 0 | 1 | Ref |
| 13909 | MZ501056 | Unassigned | Unassigned | Unassigned | 286_10 | Clustered | 9 | VC_286_10 | 9 | 0.0615 | 0.99999878 | 0.0615 | 0 | 0 | 0 | 0.6667 | Ref |
| 13910 | MZ501057 | Unassigned | Unassigned | Unassigned | 722_0 | Clustered | 13 | VC_722_0 | 13 | 0.5104 | 1 | 0.5104 | 0 | 0 | 0 | 1 | Ref |
| 13911 | MZ501058 | Unassigned | Unassigned | Unassigned | 190_32 | Clustered/Singleton | 1 | VC_190_32 | 1 | 0 | 1 | 0 | 0 | 0 | 0 | 1 | Ref |
| 13912 | MZ501059 | Unassigned | Unassigned | Unassigned | 584_0 | Clustered | 28 | VC_584_0 | 28 | 0.7704 | 1 | 0.7704 | 0 | 0 | 0 | 1 | Ref |
| 13913 | MZ501060 | Unassigned | Unassigned | Unassigned | 772_2 | Clustered/Singleton | 1 | VC_772_2 | 1 | 0 | 1 | 0 | 0 | 0 | 0 | 1 | Ref |
| 13914 | MZ501061 | Unassigned | Unassigned | Unassigned | 772_18 | Clustered/Singleton | 1 | VC_772_18 | 1 | 0 | 1 | 0 | 0 | 0 | 0 | 1 | Ref |
| 13915 | MZ501062 | Unassigned | Unassigned | Unassigned | 772_19 | Clustered | 16 | VC_772_19 | 16 | 0.0975 | 0.9970466 | 0.0972 | 0 | 0 | 0 | 0.8083 | Ref |
| 13916 | MZ501063 | Unassigned | Unassigned | Unassigned | 848_0 | Clustered | 28 | VC_848_0 | 28 | 0.7927 | 1 | 0.7927 | 0 | 0 | 0 | 1 | Ref |
| 13917 | MZ501064 | Unassigned | Unassigned | Unassigned | 386_0 | Clustered | 49 | VC_386_0 | 49 | 0.7941 | 1 | 0.7941 | 0 | 0 | 0 | 1 | Ref |
| 13918 | MZ501065 | Unassigned | Unassigned | Unassigned | 283_36 | Clustered/Singleton | 1 | VC_283_36 | 1 | 0 | 1 | 0 | 0 | 0 | 0 | 1 | Ref |
| 13919 | MZ501066 | Unassigned | Unassigned | Unassigned | 283_46 | Clustered/Singleton | 1 | VC_283_46 | 1 | 0 | 1 | 0 | 0 | 0 | 0 | 1 | Ref |
| 13920 | MZ501067 | Unassigned | Unassigned | Unassigned | 283_47 | Clustered/Singleton | 1 | VC_283_47 | 1 | 0 | 1 | 0 | 0 | 0 | 0 | 1 | Ref |
| 13921 | MZ501068 | Unassigned | Unassigned | Unassigned | 722_0 | Clustered | 13 | VC_722_0 | 13 | 0.5104 | 1 | 0.5104 | 0 | 0 | 0 | 1 | Ref |
| 13922 | MZ501069 | Unassigned | Unassigned | Unassigned | 942_0 | Clustered | 36 | VC_942_0 | 36 | 0.7848 | 1 | 0.7848 | 0 | 0 | 0 | 1 | Ref |
| 13923 | MZ501070 | Unassigned | Unassigned | Unassigned | 835_0 | Clustered | 67 | VC_835_0 | 67 | 0.9624 | 1 | 0.9624 | 0 | 0 | 0 | 1 | Ref |
| 13924 | MZ501071 | Unassigned | Unassigned | Unassigned | 795_0 | Clustered | 19 | VC_795_0 | 19 | 0.6848 | 1 | 0.6848 | 0 | 0 | 0 | 1 | Ref |
| 13925 | MZ501072 | Unassigned | Unassigned | Unassigned | 191_18 | Clustered | 2 | VC_191_18 | 2 | 0.0057 | 0.91903589 | 0.0053 | 0 | 0 | 0 | 0 | Ref |
| 13926 | MZ501073 | Unassigned | Unassigned | Unassigned | 772_17 | Clustered | 2 | VC_772_17 | 2 | 0.006 | 0.57025912 | 0.0034 | 0 | 0 | 0 | 1 | Ref |
| 13927 | MZ501074 | Unassigned | Unassigned | Unassigned | 190_6 | Clustered/Singleton | 1 | VC_190_6 | 1 | 0 | 1 | 0 | 0 | 0 | 0 | 1 | Ref |
| 13928 | MZ501075 | Unassigned | Unassigned | Unassigned | 190_19 | Clustered/Singleton | 1 | VC_190_19 | 1 | 0 | 1 | 0 | 0 | 0 | 0 | 1 | Ref |
| 13929 | MZ501076 | Unassigned | Unassigned | Unassigned | 191_21 | Clustered/Singleton | 1 | VC_191_21 | 1 | 0 | 1 | 0 | 0 | 0 | 0 | 1 | Ref |
| 13930 | MZ501077 | Unassigned | Unassigned | Unassigned | 585_0 | Clustered | 4 | VC_585_0 | 4 | 0.0533 | 0.99998755 | 0.0533 | 0 | 0 | 0 | 1 | Ref |

| | | | | | | | | | | | | | | | | | |
|---|---|---|---|---|---|---|---|---|---|---|---|---|---|---|---|---|---|
| 13931 | MZ501078 | Unassigned | Unassigned | Unassigned | 392_0 | Clustered | 126 | VC_392_0 | 126 | 0.9621 | 1 | 0.9621 | 0 | 0 | 0 | 1 | Ref |
| 13932 | MZ501079 | Unassigned | Unassigned | Unassigned | 941_0 | Clustered | 40 | VC_941_0 | 40 | 0.6903 | 1 | 0.6903 | 0 | 0 | 0 | 1 | Ref |
| 13933 | MZ501080 | Unassigned | Unassigned | Unassigned | 456_0 | Clustered | 62 | VC_456_0 | 62 | 0.9026 | 1 | 0.9026 | 0 | 0 | 0 | 1 | Ref |
| 13934 | MZ501081 | Unassigned | Unassigned | Unassigned | 392_0 | Clustered | 126 | VC_392_0 | 126 | 0.9621 | 1 | 0.9621 | 0 | 0 | 0 | 1 | Ref |
| 13935 | MZ501082 | Unassigned | Unassigned | Unassigned | 772_10 | Clustered | 8 | VC_772_10 | 8 | 0.0433 | 0.9088183 | 0.0394 | 0 | 0 | 0 | 0.9286 | Ref |
| 13936 | MZ501083 | Unassigned | Unassigned | Unassigned | 772_10 | Clustered | 8 | VC_772_10 | 8 | 0.0433 | 0.9088183 | 0.0394 | 0 | 0 | 0 | 0.9286 | Ref |
| 13937 | MZ501084 | Unassigned | Unassigned | Unassigned | 848_0 | Clustered | 28 | VC_848_0 | 28 | 0.7927 | 1 | 0.7927 | 0 | 0 | 0 | 1 | Ref |
| 13938 | MZ501085 | Unassigned | Unassigned | Unassigned | 585_0 | Clustered | 4 | VC_585_0 | 4 | 0.0533 | 0.99998755 | 0.0533 | 0 | 0 | 0 | 1 | Ref |
| 13939 | MZ501086 | Unassigned | Unassigned | Unassigned | 329_3 | Clustered | 24 | VC_329_3 | 24 | 0.1206 | 1 | 0.1206 | 0 | 0 | 0 | 0.7283 | Ref |
| 13940 | MZ501087 | Unassigned | Unassigned | Unassigned | 456_0 | Clustered | 62 | VC_456_0 | 62 | 0.9026 | 1 | 0.9026 | 0 | 0 | 0 | 1 | Ref |
| 13941 | MZ501088 | Unassigned | Unassigned | Unassigned | 835_0 | Clustered | 67 | VC_835_0 | 67 | 0.9624 | 1 | 0.9624 | 0 | 0 | 0 | 1 | Ref |
| 13942 | MZ501089 | Unassigned | Unassigned | Unassigned | 283_116 | Clustered/Singleton | 1 | VC_283_116 | 1 | 0 | 1 | 0 | 0 | 0 | 0 | 1 | Ref |
| 13943 | MZ501090 | Unassigned | Unassigned | Unassigned | 941_0 | Clustered | 40 | VC_941_0 | 40 | 0.6903 | 1 | 0.6903 | 0 | 0 | 0 | 1 | Ref |
| 13944 | MZ501091 | Unassigned | Unassigned | Unassigned | 795_0 | Clustered | 19 | VC_795_0 | 19 | 0.6848 | 1 | 0.6848 | 0 | 0 | 0 | 1 | Ref |
| 13945 | MZ501092 | Unassigned | Unassigned | Unassigned | 575_0 | Clustered | 35 | VC_575_0 | 35 | 0.8815 | 1 | 0.8815 | 0 | 0 | 0 | 1 | Ref |
| 13946 | MZ501093 | Unassigned | Unassigned | Unassigned | 772_3 | Clustered/Singleton | 1 | VC_772_3 | 1 | 0 | 1 | 0 | 0 | 0 | 0 | 1 | Ref |
| 13947 | MZ501094 | Unassigned | Unassigned | Unassigned | 772_11 | Clustered | 6 | VC_772_11 | 6 | 0.0305 | 0.84253803 | 0.0257 | 0 | 0 | 0 | 0.8 | Ref |
| 13948 | MZ501095 | Unassigned | Unassigned | Unassigned | 835_0 | Clustered | 67 | VC_835_0 | 67 | 0.9624 | 1 | 0.9624 | 0 | 0 | 0 | 1 | Ref |
| 13949 | MZ501096 | Unassigned | Unassigned | Unassigned | 283_147 | Clustered/Singleton | 1 | VC_283_147 | 1 | 0 | 1 | 0 | 0 | 0 | 0 | 1 | Ref |
| 13950 | MZ501097 | Unassigned | Unassigned | Unassigned | 835_0 | Clustered | 67 | VC_835_0 | 67 | 0.9624 | 1 | 0.9624 | 0 | 0 | 0 | 1 | Ref |
| 13951 | MZ501098 | Unassigned | Unassigned | Unassigned | 283_70 | Clustered/Singleton | 1 | VC_283_70 | 1 | 0 | 1 | 0 | 0 | 0 | 0 | 1 | Ref |
| 13952 | MZ501099 | Unassigned | Unassigned | Unassigned | 585_1 | Clustered | 3 | VC_585_1 | 3 | 0.0313 | 0.99857668 | 0.0312 | 0 | 0 | 0 | 1 | Ref |
| 13953 | MZ501100 | Unassigned | Unassigned | Unassigned | 848_0 | Clustered | 28 | VC_848_0 | 28 | 0.7927 | 1 | 0.7927 | 0 | 0 | 0 | 1 | Ref |
| 13954 | MZ501101 | Unassigned | Unassigned | Unassigned | 942_0 | Clustered | 36 | VC_942_0 | 36 | 0.7848 | 1 | 0.7848 | 0 | 0 | 0 | 1 | Ref |
| 13955 | MZ501102 | Unassigned | Unassigned | Unassigned | 772_10 | Clustered | 8 | VC_772_10 | 8 | 0.0433 | 0.9088183 | 0.0394 | 0 | 0 | 0 | 0.9286 | Ref |
| 13956 | MZ501103 | Unassigned | Unassigned | Unassigned | 190_33 | Clustered/Singleton | 1 | VC_190_33 | 1 | 0 | 1 | 0 | 0 | 0 | 0 | 1 | Ref |
| 13957 | MZ501104 | Unassigned | Unassigned | Unassigned | 795_0 | Clustered | 19 | VC_795_0 | 19 | 0.6848 | 1 | 0.6848 | 0 | 0 | 0 | 1 | Ref |
| 13958 | MZ501105 | Unassigned | Unassigned | Unassigned | 191_17 | Clustered | 4 | VC_191_17 | 4 | 0.0166 | 0.99930063 | 0.0166 | 0 | 0 | 0 | 0.8333 | Ref |
| 13959 | MZ501106 | Unassigned | Unassigned | Unassigned | 283_50 | Clustered/Singleton | 1 | VC_283_50 | 1 | 0 | 1 | 0 | 0 | 0 | 0 | 1 | Ref |
| 13960 | MZ501107 | Unassigned | Unassigned | Unassigned | 835_0 | Clustered | 67 | VC_835_0 | 67 | 0.9624 | 1 | 0.9624 | 0 | 0 | 0 | 1 | Ref |
| 13961 | MZ501108 | Unassigned | Unassigned | Unassigned | 191_28 | Clustered/Singleton | 1 | VC_191_28 | 1 | 0 | 1 | 0 | 0 | 0 | 0 | 1 | Ref |
| 13962 | MZ501109 | Unassigned | Unassigned | Unassigned | 191_2 | Clustered/Singleton | 1 | VC_191_2 | 1 | 0 | 1 | 0 | 0 | 0 | 0 | 1 | Ref |
| 13963 | MZ501110 | Unassigned | Unassigned | Unassigned | 795_0 | Clustered | 19 | VC_795_0 | 19 | 0.6848 | 1 | 0.6848 | 0 | 0 | 0 | 1 | Ref |
| 13964 | MZ501111 | Unassigned | Unassigned | Unassigned | 772_6 | Clustered | 3 | VC_772_6 | 3 | 0.012 | 0.63979507 | 0.0077 | 0 | 0 | 0 | 1 | Ref |
| 13965 | MZ501112 | Unassigned | Unassigned | Unassigned | 795_0 | Clustered | 19 | VC_795_0 | 19 | 0.6848 | 1 | 0.6848 | 0 | 0 | 0 | 1 | Ref |
| 13966 | MZ501113 | Unassigned | Unassigned | Unassigned | 283_118 | Clustered | 3 | VC_283_118 | 3 | 0.0054 | 0.99110781 | 0.0053 | 0 | 0 | 0 | 1 | Ref |
| 13967 | MZ501260 | Unassigned | Unassigned | Unassigned | 85_3 | Clustered | 19 | VC_85_3 | 19 | 0.3671 | 1 | 0.3671 | 0 | 0 | 0 | 0.807 | Ref |
| 13968 | MZ501261 | Unassigned | Unassigned | Unassigned | 85_3 | Clustered | 19 | VC_85_3 | 19 | 0.3671 | 1 | 0.3671 | 0 | 0 | 0 | 0.807 | Ref |
| 13969 | MZ501262 | Unassigned | Unassigned | Unassigned | 85_3 | Clustered | 19 | VC_85_3 | 19 | 0.3671 | 1 | 0.3671 | 0 | 0 | 0 | 0.807 | Ref |
| 13970 | MZ501263 | Unassigned | Unassigned | Unassigned | 85_3 | Clustered | 19 | VC_85_3 | 19 | 0.3671 | 1 | 0.3671 | 0 | 0 | 0 | 0.807 | Ref |
| 13971 | MZ501265 | Unassigned | Unassigned | Unassigned | 85_3 | Clustered | 19 | VC_85_3 | 19 | 0.3671 | 1 | 0.3671 | 0 | 0 | 0 | 0.807 | Ref |
| 13972 | MZ501266 | Unassigned | Unassigned | Unassigned | 749_0 | Clustered | 40 | VC_749_0 | 40 | 0.7517 | 1 | 0.7517 | 0 | 0 | 0 | 1 | Ref |
| 13973 | MZ501267 | Unassigned | Unassigned | Unassigned | 442_2 | Clustered | 4 | VC_442_2 | 4 | 0.0844 | 0.99988129 | 0.0844 | 0 | 0 | 0 | 0.8333 | Ref |
| 13974 | MZ501268 | Unassigned | Unassigned | Unassigned | 489_1 | Clustered | 11 | VC_489_1 | 11 | 0.4621 | 1 | 0.4621 | 0 | 0 | 0 | 1 | Ref |
| 13975 | MZ501269 | Unassigned | Unassigned | Unassigned | 540_0 | Clustered | 15 | VC_540_0 | 15 | 0.2866 | 1 | 0.2866 | 0 | 0 | 0 | 1 | Ref |
| 13976 | MZ501270 | Unassigned | Unassigned | Unassigned | 45_0 | Clustered | 12 | VC_45_0 | 12 | 0.35 | 1 | 0.35 | 0 | 0 | 0 | 1 | Ref |
| 13977 | MZ501272 | Unassigned | Unassigned | Unassigned | 129_0 | Clustered | 43 | VC_129_0 | 43 | 0.886 | 1 | 0.886 | 0 | 0 | 0 | 1 | Ref |
| 13978 | MZ502379 | Unassigned | Unassigned | Unassigned | 283_1 | Clustered/Singleton | 1 | VC_283_1 | 1 | 0 | 1 | 0 | 0 | 0 | 0 | 1 | Ref |
| 13979 | MZ502380 | Unassigned | Unassigned | Unassigned | 283_164 | Clustered | 2 | VC_283_164 | 2 | 0.003 | 0.93475788 | 0.0028 | 0 | 0 | 0 | 1 | Ref |
| 13980 | MZ503613 | Unassigned | Unassigned | Unassigned | 417_0 | Clustered | 5 | VC_417_0 | 5 | 0.6629 | 0.9977434 | 0.6614 | 0 | 0 | 0 | 0.8 | Ref |
| 13981 | MZ504876 | Unassigned | Unassigned | Unassigned | 590_0 | Clustered | 27 | VC_590_0 | 27 | 0.9947 | 1 | 0.9947 | 0 | 0 | 0 | 1 | Ref |
| 13982 | MZ504877 | Unassigned | Unassigned | Unassigned | 590_0 | Clustered | 27 | VC_590_0 | 27 | 0.9947 | 1 | 0.9947 | 0 | 0 | 0 | 1 | Ref |
| 13983 | MZ504878 | Unassigned | Unassigned | Unassigned | 590_0 | Clustered | 27 | VC_590_0 | 27 | 0.9947 | 1 | 0.9947 | 0 | 0 | 0 | 1 | Ref |
| 13984 | MZ504995 | Unassigned | Unassigned | Unassigned | 123_0 | Clustered | 23 | VC_123_0 | 23 | 0.817 | 1 | 0.817 | 0 | 0 | 0 | 1 | Ref |
| 13985 | MZ514874 | Unassigned | Unassigned | Unassigned | 696_0 | Clustered | 2 | VC_696_0 | 2 | 0.1624 | 0.94573679 | 0.1536 | 0 | 0 | 0 | 1 | Ref |
| 13986 | MZ516827 | Unassigned | Unassigned | Unassigned | 837_0 | Clustered | 29 | VC_837_0 | 29 | 0.9955 | 1 | 0.9955 | 0 | 0 | 0 | 1 | Ref |
| 13987 | MZ520832 | Unassigned | Unassigned | Unassigned | 849_1 | Clustered/Singleton | 1 | VC_849_1 | 1 | 0 | 1 | 0 | 0 | 0 | 0 | 1 | Ref |
| 13988 | MZ520833 | Unassigned | Unassigned | Unassigned | 228_0 | Clustered | 60 | VC_228_0 | 60 | 0.942 | 1 | 0.942 | 0 | 0 | 0 | 1 | Ref |
| 13989 | MZ547449 | Unassigned | Unassigned | Unassigned | 184_0 | Clustered | 2 | VC_184_0 | 2 | 0.7964 | 0.93668477 | 0.746 | 0 | 0 | 0 | 1 | Ref |
| 13990 | MZ553930 | Unassigned | Unassigned | Unassigned | 79_0 | Clustered | 64 | VC_79_0 | 64 | 0.4185 | 1 | 0.4185 | 0 | 0 | 0 | 0.7649 | Ref |
| 13991 | MZ553931 | Unassigned | Unassigned | Unassigned | 423_0 | Clustered | 45 | VC_423_0 | 45 | 0.9484 | 1 | 0.9484 | 0 | 0 | 0 | 1 | Ref |
| 13992 | MZ560701 | Unassigned | Unassigned | Unassigned | 286_13 | Clustered | 10 | VC_286_13 | 10 | 0.0698 | 0.99999929 | 0.0698 | 0 | 0 | 0 | 0.9556 | Ref |
| 13993 | MZ560702 | Unassigned | Unassigned | Unassigned | 491_0 | Clustered | 36 | VC_491_0 | 36 | 0.5336 | 1 | 0.5336 | 0 | 0 | 0 | 0.9714 | Ref |
| 13994 | MZ561532 | Unassigned | Unassigned | Unassigned | 290_5 | Clustered | 14 | VC_290_5 | 14 | 0.1422 | 1 | 0.1422 | 0 | 0 | 0 | 0.9231 | Ref |
| 13995 | MZ568826 | Unassigned | Unassigned | Unassigned | 910_0 | Clustered | 2 | VC_910_0 | 2 | 0.9891 | 0.5 | 0.4946 | 0 | 0 | 0 | 1 | Ref |
| 13996 | MZ568827 | Unassigned | Unassigned | Unassigned | 183_0 | Clustered | 2 | VC_183_0 | 2 | 0.0667 | 0.95489893 | 0.0637 | 0 | 0 | 0 | 1 | Ref |
| 13997 | MZ568828 | Unassigned | Unassigned | Unassigned | 248_0 | Clustered | 11 | VC_248_0 | 11 | 0.7031 | 1 | 0.7031 | 0 | 0 | 0 | 1 | Ref |
| 13998 | MZ570151 | Unassigned | Unassigned | Unassigned | 191_24 | Clustered/Singleton | 1 | VC_191_24 | 1 | 0 | 1 | 0 | 0 | 0 | 0 | 1 | Ref |
| 13999 | MZ570263 | Unassigned | Unassigned | Unassigned | 817_0 | Clustered | 14 | VC_817_0 | 14 | 0.6342 | 1 | 0.6342 | 0 | 0 | 0 | 1 | Ref |
| 14000 | MZ570427 | Unassigned | Unassigned | Unassigned | 1465_0 | Clustered | 2 | VC_1465_0 | 2 | 1 | 1 | 1 | 0 | 0 | 0 | 1 | Ref |
| 14001 | MZ571827 | Unassigned | Unassigned | Unassigned | 111_0 | Clustered | 91 | VC_111_0 | 91 | 0.9587 | 1 | 0.9587 | 0 | 0 | 0 | 1 | Ref |
| 14002 | MZ571828 | Unassigned | Unassigned | Unassigned | 192_0 | Clustered | 15 | VC_192_0 | 15 | 0.5071 | 1 | 0.5071 | 0 | 0 | 0 | 1 | Ref |
| 14003 | MZ571830 | Unassigned | Unassigned | Unassigned | 192_0 | Clustered | 15 | VC_192_0 | 15 | 0.5071 | 1 | 0.5071 | 0 | 0 | 0 | 1 | Ref |
| 14004 | MZ571831 | Unassigned | Unassigned | Unassigned | 938_0 | Clustered | 99 | VC_938_0 | 99 | 0.9119 | 1 | 0.9119 | 0 | 0 | 0 | 0.9928 | Ref |
| 14005 | MZ571832 | Unassigned | Unassigned | Unassigned | 823_0 | Clustered | 124 | VC_823_0 | 124 | 0.9492 | 1 | 0.9492 | 0 | 0 | 0 | 1 | Ref |
| 14006 | MZ571833 | Unassigned | Unassigned | Unassigned | 938_0 | Clustered | 99 | VC_938_0 | 99 | 0.9119 | 1 | 0.9119 | 0 | 0 | 0 | 0.9928 | Ref |
| 14007 | MZ571834 | Unassigned | Unassigned | Unassigned | 938_0 | Clustered | 99 | VC_938_0 | 99 | 0.9119 | 1 | 0.9119 | 0 | 0 | 0 | 0.9928 | Ref |
| 14008 | MZ573923 | Unassigned | Unassigned | Unassigned | 149_15 | Clustered | 16 | VC_149_15 | 16 | 0.0788 | 1 | 0.0788 | 0 | 0 | 0 | 0.8083 | Ref |
| 14009 | MZ573924 | Unassigned | Unassigned | Unassigned | 452_0 | Clustered | 66 | VC_452_0 | 66 | 0.9248 | 1 | 0.9248 | 0 | 0 | 0 | 1 | Ref |
| 14010 | MZ576217 | Unassigned | Unassigned | Unassigned | 638_0 | Clustered | 45 | VC_638_0 | 45 | 0.7741 | 1 | 0.7741 | 0 | 0 | 0 | 1 | Ref |
| 14011 | MZ576218 | Unassigned | Unassigned | Unassigned | 638_0 | Clustered | 45 | VC_638_0 | 45 | 0.7741 | 1 | 0.7741 | 0 | 0 | 0 | 1 | Ref |
| 14012 | MZ577095 | Unassigned | Unassigned | Unassigned | 436_0 | Clustered | 4 | VC_436_0 | 4 | 0.3318 | 0.99998719 | 0.3318 | 0 | 0 | 0 | 1 | Ref |
| 14013 | MZ577096 | Unassigned | Unassigned | Unassigned | 436_0 | Clustered | 4 | VC_436_0 | 4 | 0.3318 | 0.99998719 | 0.3318 | 0 | 0 | 0 | 1 | Ref |
| 14014 | MZ577097 | Unassigned | Unassigned | Unassigned | 436_0 | Clustered | 4 | VC_436_0 | 4 | 0.3318 | 0.99998719 | 0.3318 | 0 | 0 | 0 | 1 | Ref |
| 14015 | MZ577098 | Unassigned | Unassigned | Unassigned | 436_0 | Clustered | 4 | VC_436_0 | 4 | 0.3318 | 0.99998719 | 0.3318 | 0 | 0 | 0 | 1 | Ref |
| 14016 | MZ592920 | Unassigned | Unassigned | Unassigned | 878_0 | Clustered | 32 | VC_878_0 | 32 | 0.9965 | 1 | 0.9965 | 0 | 0 | 0 | 1 | Ref |
| 14017 | MZ592921 | Unassigned | Unassigned | Unassigned | 837_0 | Clustered | 29 | VC_837_0 | 29 | 0.9955 | 1 | 0.9955 | 0 | 0 | 0 | 1 | Ref |
| 14018 | MZ592922 | Unassigned | Unassigned | Unassigned | 882_0 | Clustered | 20 | VC_882_0 | 20 | 0.9917 | 1 | 0.9917 | 0 | 0 | 0 | 1 | Ref |
| 14019 | MZ593174 | Unassigned | Unassigned | Unassigned | 699_0 | Clustered | 71 | VC_699_0 | 71 | 0.9352 | 1 | 0.9352 | 0 | 0 | 0 | 1 | Ref |
| 14020 | MZ593728 | Unassigned | Unassigned | Unassigned | 698_0 | Clustered | 26 | VC_698_0 | 26 | 0.9366 | 1 | 0.9366 | 0 | 0 | 0 | 1 | Ref |
| 14021 | MZ598487 | Unassigned | Unassigned | Unassigned | 129_0 | Clustered | 43 | VC_129_0 | 43 | 0.886 | 1 | 0.886 | 0 | 0 | 0 | 1 | Ref |
| 14022 | MZ598515 | Unassigned | Unassigned | Unassigned | 937_0 | Clustered | 40 | VC_937_0 | 40 | 0.6809 | 1 | 0.6809 | 0 | 0 | 0 | 0.9359 | Ref |
| 14023 | MZ602146 | Unassigned | Unassigned | Unassigned | 111_0 | Clustered | 91 | VC_111_0 | 91 | 0.9587 | 1 | 0.9587 | 0 | 0 | 0 | 1 | Ref |
| 14024 | MZ602147 | Unassigned | Unassigned | Unassigned | 582_3 | Clustered | 12 | VC_582_3 | 12 | 0.2357 | 1 | 0.2357 | 0 | 0 | 0 | 0.8636 | Ref |
| 14025 | MZ602148 | Unassigned | Unassigned | Unassigned | 491_0 | Clustered | 36 | VC_491_0 | 36 | 0.5336 | 1 | 0.5336 | 0 | 0 | 0 | 0.9714 | Ref |
| 14026 | MZ605292 | Unassigned | Unassigned | Unassigned | 1030_0 | Clustered | 3 | VC_1030_0 | 3 | 0.7895 | 0.99542416 | 0.7859 | 0 | 0 | 0 | 1 | Ref |
| 14027 | MZ605293 | Unassigned | Unassigned | Unassigned | 78_0 | Clustered | 2 | VC_78_0 | 2 | 0.0306 | 0.95617387 | 0.0293 | 0 | 0 | 0 | 1 | Ref |
| 14028 | MZ611865 | Unassigned | Unassigned | Unassigned | 316_0 | Clustered | 3 | VC_316_0 | 3 | 0.774 | 0.99820498 | 0.7726 | 0 | 0 | 0 | 1 | Ref |
| 14029 | MZ612130 | Unassigned | Unassigned | Unassigned | 668_0 | Clustered | 25 | VC_668_0 | 25 | 0.5539 | 1 | 0.5539 | 0 | 0 | 0 | 0.96 | Ref |
| 14030 | MZ618622 | Unassigned | Unassigned | Unassigned | 698_0 | Clustered | 26 | VC_698_0 | 26 | 0.9366 | 1 | 0.9366 | 0 | 0 | 0 | 1 | Ref |
| 14031 | MZ622157 | Unassigned | Unassigned | Unassigned | 551_6 | Clustered | 22 | VC_551_6 | 22 | 0.1214 | 1 | 0.1214 | 0 | 0 | 0 | 0.8095 | Ref |
| 14032 | MZ622158 | Unassigned | Unassigned | Unassigned | 551_7 | Clustered | 28 | VC_551_7 | 28 | 0.1661 | 1 | 0.1661 | 0 | 0 | 0 | 0.8519 | Ref |
| 14033 | MZ622159 | Unassigned | Unassigned | Unassigned | 515_0 | Clustered | 16 | VC_515_0 | 16 | 0.3793 | 1 | 0.3793 | 0 | 0 | 0 | 1 | Ref |
| 14034 | MZ622160 | Unassigned | Unassigned | Unassigned | 1224_0 | Clustered | 19 | VC_1224_0 | 19 | 1 | 1 | 1 | 0 | 0 | 0 | 1 | Ref |
| 14035 | MZ622161 | Unassigned | Unassigned | Unassigned | 685_0 | Clustered | 16 | VC_685_0 | 16 | 0.2972 | 1 | 0.2972 | 0 | 0 | 0 | 0.9167 | Ref |
| 14036 | MZ622162 | Unassigned | Unassigned | Unassigned | 521_0 | Clustered | 37 | VC_521_0 | 37 | 0.912 | 1 | 0.912 | 0 | 0 | 0 | 1 | Ref |
| 14037 | MZ622163 | Unassigned | Unassigned | Unassigned | 551_7 | Clustered | 28 | VC_551_7 | 28 | 0.1661 | 1 | 0.1661 | 0 | 0 | 0 | 0.8519 | Ref |
| 14038 | MZ622164 | Unassigned | Unassigned | Unassigned | 1176_0 | Clustered | 41 | VC_1176_0 | 41 | 0.9558 | 1 | 0.9558 | 0 | 0 | 0 | 0.9976 | Ref |
| 14039 | MZ622165 | Unassigned | Unassigned | Unassigned | 685_0 | Clustered | 16 | VC_685_0 | 16 | 0.2972 | 1 | 0.2972 | 0 | 0 | 0 | 0.9167 | Ref |
| 14040 | MZ622166 | Unassigned | Unassigned | Unassigned | 856_1 | Clustered | 13 | VC_856_1 | 13 | 0.5848 | 1 | 0.5848 | 0 | 0 | 0 | 1 | Ref |
| 14041 | MZ622167 | Unassigned | Unassigned | Unassigned | 919_0 | Clustered | 39 | VC_919_0 | 39 | 0.9069 | 1 | 0.9069 | 0 | 0 | 0 | 1 | Ref |
| 14042 | MZ622168 | Unassigned | Unassigned | Unassigned | 799_1 | Clustered | 27 | VC_799_1 | 27 | 0.8611 | 1 | 0.8611 | 0 | 0 | 0 | 1 | Ref |
| 14043 | MZ622169 | Unassigned | Unassigned | Unassigned | 546_4 | Clustered | 16 | VC_546_4 | 16 | 0.2517 | 0.9999998 | 0.2517 | 0 | 0 | 0 | 0.9417 | Ref |
| 14044 | MZ622170 | Unassigned | Unassigned | Unassigned | 1348_0 | Clustered | 5 | VC_1348_0 | 5 | 0.5462 | 0.99999985 | 0.5462 | 0 | 0 | 0 | 1 | Ref |
| 14045 | MZ622171 | Unassigned | Unassigned | Unassigned | 216_6 | Clustered | 42 | VC_216_6 | 42 | 0.473 | 1 | 0.473 | 0 | 0 | 0 | 0.8351 | Ref |
| 14046 | MZ622172 | Unassigned | Unassigned | Unassigned | 371_19 | Clustered/Singleton | 1 | VC_371_19 | 1 | 0 | 1 | 0 | 0 | 0 | 0 | 1 | Ref |
| 14047 | MZ622173 | Unassigned | Unassigned | Unassigned | 371_16 | Clustered | 42 | VC_371_16 | 42 | 0.1341 | 1 | 0.1341 | 0 | 0 | 0 | 0.7282 | Ref |
| 14048 | MZ622176 | Unassigned | Unassigned | Unassigned | 224_0 | Clustered | 19 | VC_224_0 | 19 | 0.9416 | 1 | 0.9416 | 0 | 0 | 0 | 1 | Ref |
| 14049 | MZ622177 | Unassigned | Unassigned | Unassigned | 1178_0 | Clustered | 24 | VC_1178_0 | 24 | 0.9524 | 1 | 0.9524 | 0 | 0 | 0 | 1 | Ref |
| 14050 | MZ622178 | Unassigned | Unassigned | Unassigned | 767_0 | Clustered | 34 | VC_767_0 | 34 | 0.9173 | 1 | 0.9173 | 0 | 0 | 0 | 1 | Ref |
| 14051 | MZ622179 | Unassigned | Unassigned | Unassigned | 1352_0 | Clustered | 31 | VC_1352_0 | 31 | 0.849 | 1 | 0.849 | 0 | 0 | 0 | 1 | Ref |
| 14052 | MZ622180 | Unassigned | Unassigned | Unassigned | 1344_0 | Clustered | 78 | VC_1344_0 | 78 | 0.9732 | 1 | 0.9732 | 0 | 0 | 0 | 1 | Ref |
| 14053 | MZ622181 | Unassigned | Unassigned | Unassigned | 810_0 | Clustered | 18 | VC_810_0 | 18 | 0.9121 | 1 | 0.9121 | 0 | 0 | 0 | 1 | Ref |
| 14054 | MZ622182 | Unassigned | Unassigned | Unassigned | 216_6 | Clustered | 42 | VC_216_6 | 42 | 0.473 | 1 | 0.473 | 0 | 0 | 0 | 0.8351 | Ref |
| 14055 | MZ622183 | Unassigned | Unassigned | Unassigned | 889_0 | Clustered | 16 | VC_889_0 | 16 | 0.2969 | 1 | 0.2969 | 0 | 0 | 0 | 1 | Ref |
| 14056 | MZ634338 | Unassigned | Unassigned | Unassigned | 111_0 | Clustered | 91 | VC_111_0 | 91 | 0.9587 | 1 | 0.9587 | 0 | 0 | 0 | 1 | Ref |
| 14057 | MZ634339 | Unassigned | Unassigned | Unassigned | 111_0 | Clustered | 91 | VC_111_0 | 91 | 0.9587 | 1 | 0.9587 | 0 | 0 | 0 | 1 | Ref |
| 14058 | MZ634340 | Unassigned | Unassigned | Unassigned | 111_0 | Clustered | 91 | VC_111_0 | 91 | 0.9587 | 1 | 0.9587 | 0 | 0 | 0 | 1 | Ref |
| 14059 | MZ634341 | Unassigned | Unassigned | Unassigned | 132_1 | Clustered | 17 | VC_132_1 | 17 | 0.1738 | 1 | 0.1738 | 0 | 0 | 0 | 0.9853 | Ref |
| 14060 | MZ634342 | Unassigned | Unassigned | Unassigned | 111_0 | Clustered | 91 | VC_111_0 | 91 | 0.9587 | 1 | 0.9587 | 0 | 0 | 0 | 1 | Ref |
| 14061 | MZ634343 | Unassigned | Unassigned | Unassigned | 111_0 | Clustered | 91 | VC_111_0 | 91 | 0.9587 | 1 | 0.9587 | 0 | 0 | 0 | 1 | Ref |
| 14062 | MZ634344 | Unassigned | Unassigned | Unassigned | 192_0 | Clustered | 15 | VC_192_0 | 15 | 0.5071 | 1 | 0.5071 | 0 | 0 | 0 | 1 | Ref |
| 14063 | MZ634345 | Unassigned | Unassigned | Unassigned | 938_0 | Clustered | 99 | VC_938_0 | 99 | 0.9119 | 1 | 0.9119 | 0 | 0 | 0 | 0.9928 | Ref |
| 14064 | MZ634346 | Unassigned | Unassigned | Unassigned | 938_0 | Clustered | 99 | VC_938_0 | 99 | 0.9119 | 1 | 0.9119 | 0 | 0 | 0 | 0.9928 | Ref |
| 14065 | MZ634348 | Unassigned | Unassigned | Unassigned | 937_0 | Clustered | 40 | VC_937_0 | 40 | 0.6809 | 1 | 0.6809 | 0 | 0 | 0 | 0.9359 | Ref |
| 14066 | MZ634350 | Unassigned | Unassigned | Unassigned | 937_0 | Clustered | 40 | VC_937_0 | 40 | 0.6809 | 1 | 0.6809 | 0 | 0 | 0 | 0.9359 | Ref |
| 14067 | MZ634351 | Unassigned | Unassigned | Unassigned | 937_0 | Clustered | 40 | VC_937_0 | 40 | 0.6809 | 1 | 0.6809 | 0 | 0 | 0 | 0.9359 | Ref |
| 14068 | MZ643272 | Unassigned | Unassigned | Unassigned | 153_0 | Clustered | 25 | VC_153_0 | 25 | 0.7729 | 1 | 0.7729 | 0 | 0 | 0 | 1 | Ref |
| 14069 | MZ648031 | Unassigned | Unassigned | Unassigned | 1344_0 | Clustered | 78 | VC_1344_0 | 78 | 0.9732 | 1 | 0.9732 | 0 | 0 | 0 | 1 | Ref |
| 14070 | MZ648032 | Unassigned | Unassigned | Unassigned | 982_0 | Clustered | 62 | VC_982_0 | 62 | 0.8646 | 1 | 0.8646 | 0 | 0 | 0 | 0.743 | Ref |
| 14071 | MZ648033 | Unassigned | Unassigned | Unassigned | 859_0 | Clustered | 9 | VC_859_0 | 9 | 0.9426 | 1 | 0.9426 | 0 | 0 | 0 | 1 | Ref |
| 14072 | MZ648034 | Unassigned | Unassigned | Unassigned | 325_25 | Clustered/Singleton | 1 | VC_325_25 | 1 | 0 | 1 | 0 | 0 | 0 | 0 | 1 | Ref |
| 14073 | MZ648035 | Unassigned | Unassigned | Unassigned | 1352_0 | Clustered | 31 | VC_1352_0 | 31 | 0.849 | 1 | 0.849 | 0 | 0 | 0 | 1 | Ref |
| 14074 | MZ648036 | Unassigned | Unassigned | Unassigned | 859_0 | Clustered | 9 | VC_859_0 | 9 | 0.9426 | 1 | 0.9426 | 0 | 0 | 0 | 1 | Ref |
| 14075 | MZ648037 | Unassigned | Unassigned | Unassigned | 236_0 | Clustered | 72 | VC_236_0 | 72 | 0.8104 | 1 | 0.8104 | 0 | 0 | 0 | 1 | Ref |
| 14076 | MZ648038 | Unassigned | Unassigned | Unassigned | 371_11 | Clustered | 31 | VC_371_11 | 31 | 0.0927 | 1 | 0.0927 | 0 | 0 | 0 | 0.7011 | Ref |
| 14077 | MZ648039 | Unassigned | Unassigned | Unassigned | 982_0 | Clustered | 62 | VC_982_0 | 62 | 0.8646 | 1 | 0.8646 | 0 | 0 | 0 | 0.743 | Ref |
| 14078 | MZ648040 | Unassigned | Unassigned | Unassigned | 982_0 | Clustered | 62 | VC_982_0 | 62 | 0.8646 | 1 | 0.8646 | 0 | 0 | 0 | 0.743 | Ref |
| 14079 | MZ648041 | Unassigned | Unassigned | Unassigned | 371_8 | Clustered/Singleton | 1 | VC_371_8 | 1 | 0 | 1 | 0 | 0 | 0 | 0 | 1 | Ref |
| 14080 | MZ648214 | Unassigned | Unassigned | Unassigned | 813_1 | Clustered | 24 | VC_813_1 | 24 | 0.535 | 1 | 0.535 | 0 | 0 | 0 | 1 | Ref |
| 14081 | MZ648215 | Unassigned | Unassigned | Unassigned | 451_0 | Clustered | 80 | VC_451_0 | 80 | 0.9991 | 1 | 0.9991 | 0 | 0 | 0 | 1 | Ref |
| 14082 | MZ666938 | Unassigned | Unassigned | Unassigned | 481_0 | Clustered | 8 | VC_481_0 | 8 | 0.1348 | 1 | 0.1348 | 0 | 0 | 0 | 1 | Ref |
| 14083 | MZ666965 | Unassigned | Unassigned | Unassigned | 75_0 | Clustered | 2 | VC_75_0 | 2 | 0.0563 | 0.95619279 | 0.0538 | 0 | 0 | 0 | 1 | Ref |
| 14084 | MZ666966 | Unassigned | Unassigned | Unassigned | 75_0 | Clustered | 2 | VC_75_0 | 2 | 0.0563 | 0.95619279 | 0.0538 | 0 | 0 | 0 | 1 | Ref |
| 14085 | MZ675741 | Unassigned | Unassigned | Unassigned | 696_1 | Clustered | 8 | VC_696_1 | 8 | 0.5452 | 1 | 0.5452 | 0 | 0 | 0 | 0.9286 | Ref |
| 14086 | MZ681505 | Unassigned | Unassigned | Unassigned | 551_6 | Clustered | 22 | VC_551_6 | 22 | 0.1214 | 1 | 0.1214 | 0 | 0 | 0 | 0.8095 | Ref |
| 14087 | MZ681506 | Unassigned | Unassigned | Unassigned | 1207_0 | Clustered | 29 | VC_1207_0 | 29 | 0.8225 | 1 | 0.8225 | 0 | 0 | 0 | 1 | Ref |
| 14088 | MZ681507 | Unassigned | Unassigned | Unassigned | 410_0 | Clustered | 7 | VC_410_0 | 7 | 0.7905 | 1 | 0.7905 | 0 | 0 | 0 | 1 | Ref |
| 14089 | MZ681508 | Unassigned | Unassigned | Unassigned | 224_0 | Clustered | 19 | VC_224_0 | 19 | 0.9416 | 1 | 0.9416 | 0 | 0 | 0 | 1 | Ref |
| 14090 | MZ681509 | Unassigned | Unassigned | Unassigned | 1175_0 | Clustered | 22 | VC_1175_0 | 22 | 0.4562 | 1 | 0.4562 | 0 | 0 | 0 | 1 | Ref |
| 14091 | MZ681510 | Unassigned | Unassigned | Unassigned | 1093_0 | Clustered | 40 | VC_1093_0 | 40 | 0.8754 | 1 | 0.8754 | 0 | 0 | 0 | 1 | Ref |
| 14092 | MZ681511 | Unassigned | Unassigned | Unassigned | 558_20 | Clustered | 17 | VC_558_20 | 17 | 0.054 | 1 | 0.054 | 0 | 0 | 0 | 0.9926 | Ref |

| | | | | | | | | | | | | | | | | | |
|---|---|---|---|---|---|---|---|---|---|---|---|---|---|---|---|---|---|
| 14093 | MZ681512 | Unassigned | Unassigned | Unassigned | 520_59 | Clustered | 5 | VC_520_59 | 5 | 0.0459 | 0.99999997 | 0.0459 | 0 | 0 | 0 | 1 | Ref |
| 14094 | MZ681513 | Unassigned | Unassigned | Unassigned | 1343_0 | Clustered | 21 | VC_1343_0 | 21 | 0.9843 | 1 | 0.9843 | 0 | 0 | 0 | 1 | Ref |
| 14095 | MZ681516 | Unassigned | Unassigned | Unassigned | 223_0 | Clustered | 38 | VC_223_0 | 38 | 0.9095 | 1 | 0.9095 | 0 | 0 | 0 | 1 | Ref |
| 14096 | MZ681518 | Unassigned | Unassigned | Unassigned | 514_0 | Clustered | 75 | VC_514_0 | 75 | 0.9402 | 1 | 0.9402 | 0 | 0 | 0 | 1 | Ref |
| 14097 | MZ681519 | Unassigned | Unassigned | Unassigned | 520_31 | Clustered | 11 | VC_520_31 | 11 | 0.0897 | 1 | 0.0897 | 0 | 0 | 0 | 0.6 | Ref |
| 14098 | MZ681520 | Unassigned | Unassigned | Unassigned | 371_9 | Clustered | 4 | VC_371_9 | 4 | 0.0109 | 0.9999766 | 0.0109 | 0 | 0 | 0 | 0.6667 | Ref |
| 14099 | MZ681930 | Unassigned | Unassigned | Unassigned | 283_155 | Clustered | 2 | VC_283_155 | 2 | 0.0027 | 0.9528577 | 0.0026 | 0 | 0 | 0 | 1 | Ref |
| 14100 | MZ687409 | Unassigned | Unassigned | Unassigned | 423_0 | Clustered | 45 | VC_423_0 | 45 | 0.9484 | 1 | 0.9484 | 0 | 0 | 0 | 1 | Ref |
| 14101 | MZ701794 | Unassigned | Unassigned | Unassigned | 113_0 | Clustered | 9 | VC_113_0 | 9 | 0.519 | 1 | 0.519 | 0 | 0 | 0 | 1 | Ref |
| 14102 | MZ707156 | Unassigned | Unassigned | Unassigned | 668_0 | Clustered | 25 | VC_668_0 | 25 | 0.5539 | 1 | 0.5539 | 0 | 0 | 0 | 0.96 | Ref |
| 14103 | MZ707157 | Unassigned | Unassigned | Unassigned | 668_0 | Clustered | 25 | VC_668_0 | 25 | 0.5539 | 1 | 0.5539 | 0 | 0 | 0 | 0.96 | Ref |
| 14104 | MZ712044 | Unassigned | Unassigned | Unassigned | 698_0 | Clustered | 26 | VC_698_0 | 26 | 0.9366 | 1 | 0.9366 | 0 | 0 | 0 | 1 | Ref |
| 14105 | MZ726791 | Unassigned | Unassigned | Unassigned | 329_6 | Clustered | 3 | VC_329_6 | 3 | 0.0107 | 0.99833469 | 0.0107 | 0 | 0 | 0 | 1 | Ref |
| 14106 | MZ726792 | Unassigned | Unassigned | Unassigned | 329_6 | Clustered | 3 | VC_329_6 | 3 | 0.0107 | 0.99833469 | 0.0107 | 0 | 0 | 0 | 1 | Ref |
| 14107 | MZ726793 | Unassigned | Unassigned | Unassigned | 847_1 | Clustered | 26 | VC_847_1 | 26 | 0.6323 | 1 | 0.6323 | 0 | 0 | 0 | 0.9815 | Ref |
| 14108 | MZ726794 | Unassigned | Unassigned | Unassigned | 451_0 | Clustered | 80 | VC_451_0 | 80 | 0.9991 | 1 | 0.9991 | 0 | 0 | 0 | 1 | Ref |
| 14109 | MZ726795 | Unassigned | Unassigned | Unassigned | 283_56 | Clustered/Singleton | 1 | VC_283_56 | 1 | 0 | 1 | 0 | 0 | 0 | 0 | 1 | Ref |
| 14110 | MZ726796 | Unassigned | Unassigned | Unassigned | 461_0 | Clustered | 96 | VC_461_0 | 96 | 0.9307 | 1 | 0.9307 | 0 | 0 | 0 | 0.9575 | Ref |
| 14111 | MZ726797 | Unassigned | Unassigned | Unassigned | 1049_0 | Clustered | 8 | VC_1049_0 | 8 | 0.6413 | 1 | 0.6413 | 0 | 0 | 0 | 1 | Ref |
| 14112 | MZ727333 | Unassigned | Unassigned | Unassigned | 755_0 | Clustered | 9 | VC_755_0 | 9 | 0.4489 | 1 | 0.4489 | 0 | 0 | 0 | 1 | Ref |
| 14113 | MZ747508 | Unassigned | Unassigned | Unassigned | 761_0 | Clustered | 67 | VC_761_0 | 67 | 0.8947 | 1 | 0.8947 | 0 | 0 | 0 | 1 | Ref |
| 14114 | MZ747509 | Unassigned | Unassigned | Unassigned | 517_1 | Clustered | 108 | VC_517_1 | 108 | 0.8483 | 1 | 0.8483 | 0 | 0 | 0 | 0.9955 | Ref |
| 14115 | MZ747510 | Unassigned | Unassigned | Unassigned | 557_1 | Clustered | 11 | VC_557_1 | 11 | 0.4945 | 1 | 0.4945 | 0 | 0 | 0 | 0.7818 | Ref |
| 14116 | MZ747511 | Unassigned | Unassigned | Unassigned | 556_0 | Clustered | 21 | VC_556_0 | 21 | 0.9912 | 1 | 0.9912 | 0 | 0 | 0 | 1 | Ref |
| 14117 | MZ747512 | Unassigned | Unassigned | Unassigned | 520_86 | Clustered/Singleton | 1 | VC_520_86 | 1 | 0 | 1 | 0 | 0 | 0 | 0 | 1 | Ref |
| 14118 | MZ747513 | Unassigned | Unassigned | Unassigned | 1234_0 | Clustered | 44 | VC_1234_0 | 44 | 1 | 1 | 1 | 0 | 0 | 0 | 1 | Ref |
| 14119 | MZ747514 | Unassigned | Unassigned | Unassigned | 514_0 | Clustered | 75 | VC_514_0 | 75 | 0.9402 | 1 | 0.9402 | 0 | 0 | 0 | 1 | Ref |
| 14120 | MZ747515 | Unassigned | Unassigned | Unassigned | 1162_0 | Clustered | 10 | VC_1162_0 | 10 | 0.5715 | 1 | 0.5715 | 0 | 0 | 0 | 1 | Ref |
| 14121 | MZ747516 | Unassigned | Unassigned | Unassigned | 1207_0 | Clustered | 29 | VC_1207_0 | 29 | 0.8225 | 1 | 0.8225 | 0 | 0 | 0 | 1 | Ref |
| 14122 | MZ747517 | Unassigned | Unassigned | Unassigned | 221_0 | Clustered | 22 | VC_221_0 | 22 | 0.7447 | 1 | 0.7447 | 0 | 0 | 0 | 1 | Ref |
| 14123 | MZ747518 | Unassigned | Unassigned | Unassigned | 452_0 | Clustered | 66 | VC_452_0 | 66 | 0.9248 | 1 | 0.9248 | 0 | 0 | 0 | 1 | Ref |
| 14124 | MZ747519 | Unassigned | Unassigned | Unassigned | 520_65 | Clustered/Singleton | 1 | VC_520_65 | 1 | 0 | 1 | 0 | 0 | 0 | 0 | 1 | Ref |
| 14125 | MZ747520 | Unassigned | Unassigned | Unassigned | 802_0 | Clustered | 17 | VC_802_0 | 17 | 0.8025 | 1 | 0.8025 | 0 | 0 | 0 | 1 | Ref |
| 14126 | MZ747521 | Unassigned | Unassigned | Unassigned | 221_0 | Clustered | 22 | VC_221_0 | 22 | 0.7447 | 1 | 0.7447 | 0 | 0 | 0 | 1 | Ref |
| 14127 | MZ747522 | Unassigned | Unassigned | Unassigned | 1075_0 | Clustered | 4 | VC_1075_0 | 4 | 0.1483 | 0.99998427 | 0.1483 | 0 | 0 | 0 | 1 | Ref |
| 14128 | MZ751040 | Unassigned | Unassigned | Unassigned | 87_0 | Clustered | 8 | VC_87_0 | 8 | 0.5545 | 1 | 0.5545 | 0 | 0 | 0 | 1 | Ref |
| 14129 | MZ753803 | Unassigned | Unassigned | Unassigned | 283_106 | Clustered/Singleton | 1 | VC_283_106 | 1 | 0 | 1 | 0 | 0 | 0 | 0 | 1 | Ref |
| 14130 | MZ773648 | Unassigned | Unassigned | Unassigned | 833_0 | Clustered | 16 | VC_833_0 | 16 | 0.9351 | 1 | 0.9351 | 0 | 0 | 0 | 1 | Ref |
| 14131 | MZ773939 | Unassigned | Unassigned | Unassigned | 297_1 | Clustered | 20 | VC_297_1 | 20 | 0.5653 | 1 | 0.5653 | 0 | 0 | 0 | 0.7684 | Ref |
| 14132 | MZ779062 | Unassigned | Unassigned | Unassigned | 723_0 | Clustered | 9 | VC_723_0 | 9 | 0.7138 | 1 | 0.7138 | 0 | 0 | 0 | 1 | Ref |
| 14133 | MZ779063 | Unassigned | Unassigned | Unassigned | 449_0 | Clustered | 7 | VC_449_0 | 7 | 0.6015 | 1 | 0.6015 | 0 | 0 | 0 | 1 | Ref |
| 14134 | MZ803112 | Unassigned | Unassigned | Unassigned | 340_0 | Clustered | 31 | VC_340_0 | 31 | 0.9549 | 1 | 0.9549 | 0 | 0 | 0 | 1 | Ref |
| 14135 | MZ820083 | Unassigned | Unassigned | Unassigned | 1234_0 | Clustered | 44 | VC_1234_0 | 44 | 1 | 1 | 1 | 0 | 0 | 0 | 1 | Ref |
| 14136 | MZ820085 | Unassigned | Unassigned | Unassigned | 559_1 | Clustered/Singleton | 1 | VC_559_1 | 1 | 0 | 1 | 0 | 0 | 0 | 0 | 1 | Ref |
| 14137 | MZ820087 | Unassigned | Unassigned | Unassigned | 1207_0 | Clustered | 29 | VC_1207_0 | 29 | 0.8225 | 1 | 0.8225 | 0 | 0 | 0 | 1 | Ref |
| 14138 | MZ820088 | Unassigned | Unassigned | Unassigned | 982_0 | Clustered | 62 | VC_982_0 | 62 | 0.8646 | 1 | 0.8646 | 0 | 0 | 0 | 0.743 | Ref |
| 14139 | MZ820089 | Unassigned | Unassigned | Unassigned | 1096_0 | Clustered | 5 | VC_1096_0 | 5 | 0.8566 | 0.99869977 | 0.8555 | 0 | 0 | 0 | 1 | Ref |
| 14140 | MZ820090 | Unassigned | Unassigned | Unassigned | 216_6 | Clustered | 42 | VC_216_6 | 42 | 0.473 | 1 | 0.473 | 0 | 0 | 0 | 0.8351 | Ref |
| 14141 | MZ820091 | Unassigned | Unassigned | Unassigned | 982_0 | Clustered | 62 | VC_982_0 | 62 | 0.8646 | 1 | 0.8646 | 0 | 0 | 0 | 0.743 | Ref |
| 14142 | MZ820092 | Unassigned | Unassigned | Unassigned | 982_0 | Clustered | 62 | VC_982_0 | 62 | 0.8646 | 1 | 0.8646 | 0 | 0 | 0 | 0.743 | Ref |
| 14143 | MZ820093 | Unassigned | Unassigned | Unassigned | 1072_3 | Clustered | 6 | VC_1072_3 | 6 | 0.1304 | 0.99999994 | 0.1304 | 0 | 0 | 0 | 0.8667 | Ref |
| 14144 | MZ820094 | Unassigned | Unassigned | Unassigned | 1072_3 | Clustered | 6 | VC_1072_3 | 6 | 0.1304 | 0.99999994 | 0.1304 | 0 | 0 | 0 | 0.8667 | Ref |
| 14145 | MZ820095 | Unassigned | Unassigned | Unassigned | 1072_2 | Clustered | 5 | VC_1072_2 | 5 | 0.1003 | 0.99999315 | 0.1003 | 0 | 0 | 0 | 1 | Ref |
| 14146 | MZ821031 | Unassigned | Unassigned | Unassigned | 1127_0 | Clustered | 19 | VC_1127_0 | 19 | 0.7611 | 1 | 0.7611 | 0 | 0 | 0 | 1 | Ref |
| 14147 | MZ826344 | Unassigned | Unassigned | Unassigned | 129_0 | Clustered | 43 | VC_129_0 | 43 | 0.886 | 1 | 0.886 | 0 | 0 | 0 | 1 | Ref |
| 14148 | MZ826345 | Unassigned | Unassigned | Unassigned | 129_0 | Clustered | 43 | VC_129_0 | 43 | 0.886 | 1 | 0.886 | 0 | 0 | 0 | 1 | Ref |
| 14149 | MZ826346 | Unassigned | Unassigned | Unassigned | 129_0 | Clustered | 43 | VC_129_0 | 43 | 0.886 | 1 | 0.886 | 0 | 0 | 0 | 1 | Ref |
| 14150 | MZ826347 | Unassigned | Unassigned | Unassigned | 129_0 | Clustered | 43 | VC_129_0 | 43 | 0.886 | 1 | 0.886 | 0 | 0 | 0 | 1 | Ref |
| 14151 | MZ826348 | Unassigned | Unassigned | Unassigned | 129_0 | Clustered | 43 | VC_129_0 | 43 | 0.886 | 1 | 0.886 | 0 | 0 | 0 | 1 | Ref |
| 14152 | MZ826349 | Unassigned | Unassigned | Unassigned | 129_0 | Clustered | 43 | VC_129_0 | 43 | 0.886 | 1 | 0.886 | 0 | 0 | 0 | 1 | Ref |
| 14153 | MZ826350 | Unassigned | Unassigned | Unassigned | 78_5 | Clustered | 6 | VC_78_5 | 6 | 0.1087 | 0.99978082 | 0.1087 | 0 | 0 | 0 | 1 | Ref |
| 14154 | MZ826351 | Unassigned | Unassigned | Unassigned | 129_0 | Clustered | 43 | VC_129_0 | 43 | 0.886 | 1 | 0.886 | 0 | 0 | 0 | 1 | Ref |
| 14155 | MZ826352 | Unassigned | Unassigned | Unassigned | 129_0 | Clustered | 43 | VC_129_0 | 43 | 0.886 | 1 | 0.886 | 0 | 0 | 0 | 1 | Ref |
| 14156 | MZ826353 | Unassigned | Unassigned | Unassigned | 78_5 | Clustered | 6 | VC_78_5 | 6 | 0.1087 | 0.99978082 | 0.1087 | 0 | 0 | 0 | 1 | Ref |
| 14157 | MZ826354 | Unassigned | Unassigned | Unassigned | 129_0 | Clustered | 43 | VC_129_0 | 43 | 0.886 | 1 | 0.886 | 0 | 0 | 0 | 1 | Ref |
| 14158 | MZ832314 | Unassigned | Unassigned | Unassigned | 638_0 | Clustered | 45 | VC_638_0 | 45 | 0.7741 | 1 | 0.7741 | 0 | 0 | 0 | 1 | Ref |
| 14159 | MZ833439 | Unassigned | Unassigned | Unassigned | 392_0 | Clustered | 126 | VC_392_0 | 126 | 0.9621 | 1 | 0.9621 | 0 | 0 | 0 | 1 | Ref |
| 14160 | MZ836210 | Unassigned | Unassigned | Unassigned | 460_0 | Clustered | 19 | VC_460_0 | 19 | 0.3744 | 1 | 0.3744 | 0 | 0 | 0 | 1 | Ref |
| 14161 | MZ856343 | Unassigned | Unassigned | Unassigned | 558_42 | Clustered/Singleton | 1 | VC_558_42 | 1 | 0 | 1 | 0 | 0 | 0 | 0 | 1 | Ref |
| 14162 | MZ868638 | Unassigned | Unassigned | Unassigned | 286_13 | Clustered | 10 | VC_286_13 | 10 | 0.0698 | 0.99999929 | 0.0698 | 0 | 0 | 0 | 0.9556 | Ref |
| 14163 | MZ868713 | Unassigned | Unassigned | Unassigned | 78_3 | Clustered | 11 | VC_78_3 | 11 | 0.2572 | 1 | 0.2572 | 0 | 0 | 0 | 1 | Ref |
| 14164 | MZ868714 | Unassigned | Unassigned | Unassigned | 155_0 | Clustered | 14 | VC_155_0 | 14 | 0.6095 | 1 | 0.6095 | 0 | 0 | 0 | 1 | Ref |
| 14165 | MZ868715 | Unassigned | Unassigned | Unassigned | 155_0 | Clustered | 14 | VC_155_0 | 14 | 0.6095 | 1 | 0.6095 | 0 | 0 | 0 | 1 | Ref |
| 14166 | MZ868716 | Unassigned | Unassigned | Unassigned | 155_0 | Clustered | 14 | VC_155_0 | 14 | 0.6095 | 1 | 0.6095 | 0 | 0 | 0 | 1 | Ref |
| 14167 | MZ868717 | Unassigned | Unassigned | Unassigned | 155_0 | Clustered | 14 | VC_155_0 | 14 | 0.6095 | 1 | 0.6095 | 0 | 0 | 0 | 1 | Ref |
| 14168 | MZ868718 | Unassigned | Unassigned | Unassigned | 697_0 | Clustered | 5 | VC_697_0 | 5 | 0.8303 | 0.99999971 | 0.8303 | 0 | 0 | 0 | 1 | Ref |
| 14169 | MZ868724 | Unassigned | Unassigned | Unassigned | 699_0 | Clustered | 71 | VC_699_0 | 71 | 0.9352 | 1 | 0.9352 | 0 | 0 | 0 | 1 | Ref |
| 14170 | MZ868725 | Unassigned | Unassigned | Unassigned | 699_0 | Clustered | 71 | VC_699_0 | 71 | 0.9352 | 1 | 0.9352 | 0 | 0 | 0 | 1 | Ref |
| 14171 | MZ868726 | Unassigned | Unassigned | Unassigned | 699_0 | Clustered | 71 | VC_699_0 | 71 | 0.9352 | 1 | 0.9352 | 0 | 0 | 0 | 1 | Ref |
| 14172 | MZ890186 | Unassigned | Unassigned | Unassigned | 491_0 | Clustered | 36 | VC_491_0 | 36 | 0.5336 | 1 | 0.5336 | 0 | 0 | 0 | 0.9714 | Ref |
| 14173 | MZ892987 | Unassigned | Unassigned | Unassigned | 305_0 | Clustered | 12 | VC_305_0 | 12 | 0.5745 | 1 | 0.5745 | 0 | 0 | 0 | 0.8485 | Ref |
| 14174 | MZ892988 | Unassigned | Unassigned | Unassigned | 305_0 | Clustered | 12 | VC_305_0 | 12 | 0.5745 | 1 | 0.5745 | 0 | 0 | 0 | 0.8485 | Ref |
| 14175 | MZ892989 | Unassigned | Unassigned | Unassigned | 305_0 | Clustered | 12 | VC_305_0 | 12 | 0.5745 | 1 | 0.5745 | 0 | 0 | 0 | 0.8485 | Ref |
| 14176 | MZ892990 | Unassigned | Unassigned | Unassigned | 305_0 | Clustered | 12 | VC_305_0 | 12 | 0.5745 | 1 | 0.5745 | 0 | 0 | 0 | 0.8485 | Ref |
| 14177 | MZ892991 | Unassigned | Unassigned | Unassigned | 305_0 | Clustered | 12 | VC_305_0 | 12 | 0.5745 | 1 | 0.5745 | 0 | 0 | 0 | 0.8485 | Ref |
| 14178 | MZ892992 | Unassigned | Unassigned | Unassigned | 305_0 | Clustered | 12 | VC_305_0 | 12 | 0.5745 | 1 | 0.5745 | 0 | 0 | 0 | 0.8485 | Ref |
| 14179 | MZ923531 | Unassigned | Unassigned | Unassigned | 451_0 | Clustered | 80 | VC_451_0 | 80 | 0.9991 | 1 | 0.9991 | 0 | 0 | 0 | 1 | Ref |
| 14180 | MZ926748 | Unassigned | Unassigned | Unassigned | 464_0 | Clustered | 8 | VC_464_0 | 8 | 0.2629 | 1 | 0.2629 | 0 | 0 | 0 | 1 | Ref |
| 14181 | MZ927745 | Unassigned | Unassigned | Unassigned | 749_0 | Clustered | 40 | VC_749_0 | 40 | 0.7517 | 1 | 0.7517 | 0 | 0 | 0 | 1 | Ref |
| 14182 | MZ927746 | Unassigned | Unassigned | Unassigned | 749_0 | Clustered | 40 | VC_749_0 | 40 | 0.7517 | 1 | 0.7517 | 0 | 0 | 0 | 1 | Ref |
| 14183 | MZ936314 | Unassigned | Unassigned | Unassigned | 699_0 | Clustered | 71 | VC_699_0 | 71 | 0.9352 | 1 | 0.9352 | 0 | 0 | 0 | 1 | Ref |
| 14184 | MZ936315 | Unassigned | Unassigned | Unassigned | 699_0 | Clustered | 71 | VC_699_0 | 71 | 0.9352 | 1 | 0.9352 | 0 | 0 | 0 | 1 | Ref |
| 14185 | MZ936316 | Unassigned | Unassigned | Unassigned | 699_0 | Clustered | 71 | VC_699_0 | 71 | 0.9352 | 1 | 0.9352 | 0 | 0 | 0 | 1 | Ref |
| 14186 | MZ955866 | Unassigned | Unassigned | Unassigned | 637_0 | Clustered | 115 | VC_637_0 | 115 | 0.731 | 1 | 0.731 | 0 | 0 | 0 | 0.8764 | Ref |
| 14187 | MZ955867 | Unassigned | Unassigned | Unassigned | 838_0 | Clustered | 29 | VC_838_0 | 29 | 0.8611 | 1 | 0.8611 | 0 | 0 | 0 | 1 | Ref |
| 14188 | MZ955869 | Unassigned | Unassigned | Unassigned | 690_0 | Clustered | 50 | VC_690_0 | 50 | 0.9805 | 1 | 0.9805 | 0 | 0 | 0 | 1 | Ref |
| 14189 | MZ955870 | Unassigned | Unassigned | Unassigned | 1159_0 | Clustered | 3 | VC_1159_0 | 3 | 0.3304 | 0.98548359 | 0.3256 | 0 | 0 | 0 | 1 | Ref |
| 14190 | MZ958744 | Unassigned | Unassigned | Unassigned | 520_8 | Clustered/Singleton | 1 | VC_520_8 | 1 | 0 | 1 | 0 | 0 | 0 | 0 | 1 | Ref |
| 14191 | MZ958745 | Unassigned | Unassigned | Unassigned | 558_22 | Clustered | 4 | VC_558_22 | 4 | 0.01 | 0.99312819 | 0.0099 | 0 | 0 | 0 | 0.8333 | Ref |
| 14192 | MZ958746 | Unassigned | Unassigned | Unassigned | 887_0 | Clustered | 18 | VC_887_0 | 18 | 0.3663 | 1 | 0.3663 | 0 | 0 | 0 | 1 | Ref |
| 14193 | MZ958747 | Unassigned | Unassigned | Unassigned | 238_0 | Clustered | 17 | VC_238_0 | 17 | 0.6109 | 1 | 0.6109 | 0 | 0 | 0 | 1 | Ref |
| 14194 | MZ958748 | Unassigned | Unassigned | Unassigned | 546_4 | Clustered | 16 | VC_546_4 | 16 | 0.2517 | 0.9999998 | 0.2517 | 0 | 0 | 0 | 0.9417 | Ref |
| 14195 | MZ958749 | Unassigned | Unassigned | Unassigned | 1240_0 | Clustered | 82 | VC_1240_0 | 82 | 0.7945 | 1 | 0.7945 | 0 | 0 | 0 | 1 | Ref |
| 14196 | MZ958750 | Unassigned | Unassigned | Unassigned | 1071_3 | Clustered | 16 | VC_1071_3 | 16 | 0.2354 | 1 | 0.2354 | 0 | 0 | 0 | 0.9667 | Ref |
| 14197 | MZ967493 | Unassigned | Unassigned | Unassigned | 699_0 | Clustered | 71 | VC_699_0 | 71 | 0.9352 | 1 | 0.9352 | 0 | 0 | 0 | 1 | Ref |
| 14198 | MZ969646 | Unassigned | Unassigned | Unassigned | 89_5 | Clustered/Singleton | 1 | VC_89_5 | 1 | 0 | 1 | 0 | 0 | 0 | 0 | 1 | Ref |
| 14199 | MZ995503 | Unassigned | Unassigned | Unassigned | 422_0 | Clustered | 5 | VC_422_0 | 5 | 0.2592 | 0.99999997 | 0.2592 | 0 | 0 | 0 | 1 | Ref |
| 14200 | MZ995504 | Unassigned | Unassigned | Unassigned | 916_0 | Clustered | 4 | VC_916_0 | 4 | 0.8472 | 0.99985574 | 0.8471 | 0 | 0 | 0 | 1 | Ref |
| 14201 | MZ995505 | Unassigned | Unassigned | Unassigned | 815_0 | Clustered | 8 | VC_815_0 | 8 | 0.9801 | 1 | 0.9801 | 0 | 0 | 0 | 1 | Ref |
| 14202 | MZ995506 | Unassigned | Unassigned | Unassigned | 422_0 | Clustered | 5 | VC_422_0 | 5 | 0.2592 | 0.99999997 | 0.2592 | 0 | 0 | 0 | 1 | Ref |
| 14203 | MZ997838 | Unassigned | Unassigned | Unassigned | 638_0 | Clustered | 45 | VC_638_0 | 45 | 0.7741 | 1 | 0.7741 | 0 | 0 | 0 | 1 | Ref |
| 14204 | MZ997839 | Unassigned | Unassigned | Unassigned | 452_0 | Clustered | 66 | VC_452_0 | 66 | 0.9248 | 1 | 0.9248 | 0 | 0 | 0 | 1 | Ref |
| 14205 | MZ997840 | Unassigned | Unassigned | Unassigned | 452_0 | Clustered | 66 | VC_452_0 | 66 | 0.9248 | 1 | 0.9248 | 0 | 0 | 0 | 1 | Ref |
| 14206 | MZ997841 | Unassigned | Unassigned | Unassigned | 452_0 | Clustered | 66 | VC_452_0 | 66 | 0.9248 | 1 | 0.9248 | 0 | 0 | 0 | 1 | Ref |
| 14207 | OK018184 | Unassigned | Unassigned | Unassigned | 283_165 | Clustered/Singleton | 1 | VC_283_165 | 1 | 0 | 1 | 0 | 0 | 0 | 0 | 1 | Ref |
| 14208 | OK019720 | Unassigned | Unassigned | Unassigned | 586_0 | Clustered | 3 | VC_586_0 | 3 | 0.0495 | 0.99859933 | 0.0495 | 0 | 0 | 0 | 1 | Ref |
| 14209 | OK040170 | Unassigned | Unassigned | Unassigned | 488_1 | Clustered | 8 | VC_488_1 | 8 | 0.2927 | 1 | 0.2927 | 0 | 0 | 0 | 1 | Ref |
| 14210 | OK040171 | Unassigned | Unassigned | Unassigned | 78_2 | Clustered/Singleton | 1 | VC_78_2 | 1 | 0 | 1 | 0 | 0 | 0 | 0 | 1 | Ref |
| 14211 | OK040777 | Unassigned | Unassigned | Unassigned | 1207_0 | Clustered | 29 | VC_1207_0 | 29 | 0.8225 | 1 | 0.8225 | 0 | 0 | 0 | 1 | Ref |
| 14212 | OK040778 | Unassigned | Unassigned | Unassigned | 236_0 | Clustered | 72 | VC_236_0 | 72 | 0.8104 | 1 | 0.8104 | 0 | 0 | 0 | 1 | Ref |
| 14213 | OK040779 | Unassigned | Unassigned | Unassigned | 1206_0 | Clustered | 8 | VC_1206_0 | 8 | 0.1913 | 1 | 0.1913 | 0 | 0 | 0 | 1 | Ref |
| 14214 | OK040781 | Unassigned | Unassigned | Unassigned | 1344_0 | Clustered | 78 | VC_1344_0 | 78 | 0.9732 | 1 | 0.9732 | 0 | 0 | 0 | 1 | Ref |
| 14215 | OK040782 | Unassigned | Unassigned | Unassigned | 236_0 | Clustered | 72 | VC_236_0 | 72 | 0.8104 | 1 | 0.8104 | 0 | 0 | 0 | 1 | Ref |
| 14216 | OK040783 | Unassigned | Unassigned | Unassigned | 1344_0 | Clustered | 78 | VC_1344_0 | 78 | 0.9732 | 1 | 0.9732 | 0 | 0 | 0 | 1 | Ref |
| 14217 | OK040784 | Unassigned | Unassigned | Unassigned | 236_0 | Clustered | 72 | VC_236_0 | 72 | 0.8104 | 1 | 0.8104 | 0 | 0 | 0 | 1 | Ref |
| 14218 | OK040785 | Unassigned | Unassigned | Unassigned | 919_0 | Clustered | 39 | VC_919_0 | 39 | 0.9069 | 1 | 0.9069 | 0 | 0 | 0 | 1 | Ref |
| 14219 | OK040786 | Unassigned | Unassigned | Unassigned | 890_1 | Clustered | 4 | VC_890_1 | 4 | 0.4613 | 0.99998458 | 0.4613 | 0 | 0 | 0 | 1 | Ref |
| 14220 | OK040788 | Unassigned | Unassigned | Unassigned | 1343_0 | Clustered | 21 | VC_1343_0 | 21 | 0.9843 | 1 | 0.9843 | 0 | 0 | 0 | 1 | Ref |
| 14221 | OK040789 | Unassigned | Unassigned | Unassigned | 558_50 | Clustered | 15 | VC_558_50 | 15 | 0.0468 | 1 | 0.0468 | 0 | 0 | 0 | 0.9238 | Ref |
| 14222 | OK040790 | Unassigned | Unassigned | Unassigned | 1100_2 | Clustered/Singleton | 1 | VC_1100_2 | 1 | 0 | 1 | 0 | 0 | 0 | 0 | 1 | Ref |
| 14223 | OK040791 | Unassigned | Unassigned | Unassigned | 325_30 | Clustered | 4 | VC_325_30 | 4 | 0.0144 | 0.99998616 | 0.0144 | 0 | 0 | 0 | 0.8333 | Ref |
| 14224 | OK040792 | Unassigned | Unassigned | Unassigned | 1349_0 | Clustered | 7 | VC_1349_0 | 7 | 0.2495 | 1 | 0.2495 | 0 | 0 | 0 | 1 | Ref |
| 14225 | OK040793 | Unassigned | Unassigned | Unassigned | 1100_1 | Clustered | 2 | VC_1100_1 | 2 | 0.1223 | 0.94673154 | 0.1158 | 0 | 0 | 0 | 1 | Ref |
| 14226 | OK040794 | Unassigned | Unassigned | Unassigned | 1099_1 | Clustered/Singleton | 1 | VC_1099_1 | 1 | 0 | 1 | 0 | 0 | 0 | 0 | 1 | Ref |
| 14227 | OK040806 | Unassigned | Unassigned | Unassigned | 329_17 | Clustered | 4 | VC_329_17 | 4 | 0.0163 | 0.99996247 | 0.0163 | 0 | 0 | 0 | 1 | Ref |
| 14228 | OK040807 | Unassigned | Unassigned | Unassigned | 329_17 | Clustered | 4 | VC_329_17 | 4 | 0.0163 | 0.99996247 | 0.0163 | 0 | 0 | 0 | 1 | Ref |
| 14229 | OK040907 | Unassigned | Unassigned | Unassigned | 283_3 | Clustered | 3 | VC_283_3 | 3 | 0.0055 | 0.9968017 | 0.0055 | 0 | 0 | 0 | 1 | Ref |
| 14230 | OK041468 | Unassigned | Unassigned | Unassigned | 603_0 | Clustered | 9 | VC_603_0 | 9 | 0.9925 | 0.99999994 | 0.9925 | 0 | 0 | 0 | 1 | Ref |
| 14231 | OK041469 | Unassigned | Unassigned | Unassigned | 594_0 | Clustered | 2 | VC_594_0 | 2 | 0.0737 | 0.95685859 | 0.0705 | 0 | 0 | 0 | 1 | Ref |
| 14232 | OK042080 | Unassigned | Unassigned | Unassigned | 392_0 | Clustered | 126 | VC_392_0 | 126 | 0.9621 | 1 | 0.9621 | 0 | 0 | 0 | 1 | Ref |
| 14233 | OK073976 | Unassigned | Unassigned | Unassigned | 792_0 | Clustered | 35 | VC_792_0 | 35 | 0.9828 | 1 | 0.9828 | 0 | 0 | 0 | 1 | Ref |
| 14234 | OK076929 | Unassigned | Unassigned | Unassigned | 283_128 | Clustered/Singleton | 1 | VC_283_128 | 1 | 0 | 1 | 0 | 0 | 0 | 0 | 1 | Ref |
| 14235 | OK085710 | Unassigned | Unassigned | Unassigned | 341_0 | Clustered | 3 | VC_341_0 | 3 | 0.2522 | 0.9396546 | 0.237 | 0 | 0 | 0 | 1 | Ref |
| 14236 | OK094519 | Unassigned | Unassigned | Unassigned | 130_0 | Clustered | 19 | VC_130_0 | 19 | 0.6068 | 1 | 0.6068 | 0 | 0 | 0 | 1 | Ref |
| 14237 | OK094665 | Unassigned | Unassigned | Unassigned | 749_0 | Clustered | 40 | VC_749_0 | 40 | 0.7517 | 1 | 0.7517 | 0 | 0 | 0 | 1 | Ref |
| 14238 | OK094707 | Unassigned | Unassigned | Unassigned | 174_0 | Clustered | 3 | VC_174_0 | 3 | 0.9624 | 0.99333557 | 0.9559 | 0 | 0 | 0 | 1 | Ref |
| 14239 | OK095358 | Unassigned | Unassigned | Unassigned | 353_0 | Clustered | 6 | VC_353_0 | 6 | 0.545 | 1 | 0.545 | 0 | 0 | 0 | 1 | Ref |
| 14240 | OK108607 | Unassigned | Unassigned | Unassigned | 190_23 | Clustered | 10 | VC_190_23 | 10 | 0.0704 | 1 | 0.0704 | 0 | 0 | 0 | 0.8444 | Ref |
| 14241 | OK108608 | Unassigned | Unassigned | Unassigned | 637_0 | Clustered | 115 | VC_637_0 | 115 | 0.731 | 1 | 0.731 | 0 | 0 | 0 | 0.8764 | Ref |
| 14242 | OK108609 | Unassigned | Unassigned | Unassigned | 637_0 | Clustered | 115 | VC_637_0 | 115 | 0.731 | 1 | 0.731 | 0 | 0 | 0 | 0.8764 | Ref |
| 14243 | OK129343 | Unassigned | Unassigned | Unassigned | 792_0 | Clustered | 35 | VC_792_0 | 35 | 0.9828 | 1 | 0.9828 | 0 | 0 | 0 | 1 | Ref |
| 14244 | OK136180 | Unassigned | Unassigned | Unassigned | 582_1 | Clustered | 3 | VC_582_1 | 3 | 0.0621 | 0.99815566 | 0.062 | 0 | 0 | 0 | 1 | Ref |
| 14245 | OK136181 | Unassigned | Unassigned | Unassigned | 850_0 | Clustered | 3 | VC_850_0 | 3 | 0.0897 | 0.99847793 | 0.0896 | 0 | 0 | 0 | 1 | Ref |
| 14246 | OK138555 | Unassigned | Unassigned | Unassigned | 938_0 | Clustered | 99 | VC_938_0 | 99 | 0.9119 | 1 | 0.9119 | 0 | 0 | 0 | 0.9928 | Ref |
| 14247 | OK138556 | Unassigned | Unassigned | Unassigned | 823_0 | Clustered | 124 | VC_823_0 | 124 | 0.9492 | 1 | 0.9492 | 0 | 0 | 0 | 1 | Ref |
| 14248 | OK143207 | Unassigned | Unassigned | Unassigned | 190_24 | Clustered | 5 | VC_190_24 | 5 | 0.0293 | 0.9999996 | 0.0293 | 0 | 0 | 0 | 0.7 | Ref |
| 14249 | OK143508 | Unassigned | Unassigned | Unassigned | 637_0 | Clustered | 115 | VC_637_0 | 115 | 0.731 | 1 | 0.731 | 0 | 0 | 0 | 0.8764 | Ref |
| 14250 | OK148439 | Unassigned | Unassigned | Unassigned | 792_0 | Clustered | 35 | VC_792_0 | 35 | 0.9828 | 1 | 0.9828 | 0 | 0 | 0 | 1 | Ref |
| 14251 | OK148440 | Unassigned | Unassigned | Unassigned | 813_0 | Clustered | 12 | VC_813_0 | 12 | 0.2347 | 1 | 0.2347 | 0 | 0 | 0 | 1 | Ref |
| 14252 | OK148441 | Unassigned | Unassigned | Unassigned | 283_108 | Clustered/Singleton | 1 | VC_283_108 | 1 | 0 | 1 | 0 | 0 | 0 | 0 | 1 | Ref |
| 14253 | OK149215 | Unassigned | Unassigned | Unassigned | 385_0 | Clustered | 51 | VC_385_0 | 51 | 0.8754 | 1 | 0.8754 | 0 | 0 | 0 | 1 | Ref |
| 14254 | OK169616 | Unassigned | Unassigned | Unassigned | 637_1 | Clustered | 27 | VC_637_1 | 27 | 0.2578 | 1 | 0.2578 | 0 | 0 | 0 | 0.9858 | Ref |

| | | | | | | | | | | | | | | | | | |
|---|---|---|---|---|---|---|---|---|---|---|---|---|---|---|---|---|---|
| 14255 | OK172329 | Unassigned | Unassigned | Unassigned | 760_0 | Clustered | 3 | VC_760_0 | 3 | 0.7281 | 0.9859071 | 0.7179 | 0 | 0 | 0 | 1 | Ref |
| 14256 | OK173051 | Unassigned | Unassigned | Unassigned | 414_0 | Clustered | 37 | VC_414_0 | 37 | 0.585 | 1 | 0.585 | 0 | 0 | 0 | 0.9474 | Ref |
| 14257 | OK181858 | Unassigned | Unassigned | Unassigned | 582_1 | Clustered | 3 | VC_582_1 | 3 | 0.0621 | 0.99815566 | 0.062 | 0 | 0 | 0 | 1 | Ref |
| 14258 | OK181860 | Unassigned | Unassigned | Unassigned | 850_0 | Clustered | 3 | VC_850_0 | 3 | 0.0897 | 0.99847793 | 0.0896 | 0 | 0 | 0 | 1 | Ref |
| 14259 | OK181861 | Unassigned | Unassigned | Unassigned | 582_1 | Clustered | 3 | VC_582_1 | 3 | 0.0621 | 0.99815566 | 0.062 | 0 | 0 | 0 | 1 | Ref |
| 14260 | OK181862 | Unassigned | Unassigned | Unassigned | 850_0 | Clustered | 3 | VC_850_0 | 3 | 0.0897 | 0.99847793 | 0.0896 | 0 | 0 | 0 | 1 | Ref |
| 14261 | OK210074 | Unassigned | Unassigned | Unassigned | 451_0 | Clustered | 80 | VC_451_0 | 80 | 0.9991 | 1 | 0.9991 | 0 | 0 | 0 | 1 | Ref |
| 14262 | OK210075 | Unassigned | Unassigned | Unassigned | 669_4 | Clustered/Singleton | 1 | VC_669_4 | 1 | 0 | 1 | 0 | 0 | 0 | 0 | 1 | Ref |
| 14263 | OK210077 | Unassigned | Unassigned | Unassigned | 848_0 | Clustered | 28 | VC_848_0 | 28 | 0.7927 | 1 | 0.7927 | 0 | 0 | 0 | 1 | Ref |
| 14264 | OK216879 | Unassigned | Unassigned | Unassigned | 510_0 | Clustered | 12 | VC_510_0 | 12 | 0.3963 | 1 | 0.3963 | 0 | 0 | 0 | 0.9697 | Ref |
| 14265 | OK247671 | Unassigned | Unassigned | Unassigned | 542_0 | Clustered | 2 | VC_542_0 | 2 | 0.3753 | 0.94092208 | 0.3531 | 0 | 0 | 0 | 1 | Ref |
| 14266 | OK254197 | Unassigned | Unassigned | Unassigned | 190_23 | Clustered | 10 | VC_190_23 | 10 | 0.0704 | 1 | 0.0704 | 0 | 0 | 0 | 0.8444 | Ref |
| 14267 | OK254198 | Unassigned | Unassigned | Unassigned | 290_1 | Clustered | 8 | VC_290_1 | 8 | 0.0757 | 1 | 0.0757 | 0 | 0 | 0 | 0.8571 | Ref |
| 14268 | OK258138 | Unassigned | Unassigned | Unassigned | 74_0 | Clustered | 15 | VC_74_0 | 15 | 0.5204 | 1 | 0.5204 | 0 | 0 | 0 | 1 | Ref |
| 14269 | OK258139 | Unassigned | Unassigned | Unassigned | 78_5 | Clustered | 6 | VC_78_5 | 6 | 0.1087 | 0.99978082 | 0.1087 | 0 | 0 | 0 | 1 | Ref |
| 14270 | OK258140 | Unassigned | Unassigned | Unassigned | 113_0 | Clustered | 9 | VC_113_0 | 9 | 0.519 | 1 | 0.519 | 0 | 0 | 0 | 1 | Ref |
| 14271 | OK268422 | Unassigned | Unassigned | Unassigned | 1040_0 | Clustered | 23 | VC_1040_0 | 23 | 0.9605 | 1 | 0.9605 | 0 | 0 | 0 | 1 | Ref |
| 14272 | OK268423 | Unassigned | Unassigned | Unassigned | 1040_0 | Clustered | 23 | VC_1040_0 | 23 | 0.9605 | 1 | 0.9605 | 0 | 0 | 0 | 1 | Ref |
| 14273 | OK268424 | Unassigned | Unassigned | Unassigned | 1040_0 | Clustered | 23 | VC_1040_0 | 23 | 0.9605 | 1 | 0.9605 | 0 | 0 | 0 | 1 | Ref |
| 14274 | OK268425 | Unassigned | Unassigned | Unassigned | 1040_0 | Clustered | 23 | VC_1040_0 | 23 | 0.9605 | 1 | 0.9605 | 0 | 0 | 0 | 1 | Ref |
| 14275 | OK268426 | Unassigned | Unassigned | Unassigned | 1040_0 | Clustered | 23 | VC_1040_0 | 23 | 0.9605 | 1 | 0.9605 | 0 | 0 | 0 | 1 | Ref |
| 14276 | OK268427 | Unassigned | Unassigned | Unassigned | 1040_0 | Clustered | 23 | VC_1040_0 | 23 | 0.9605 | 1 | 0.9605 | 0 | 0 | 0 | 1 | Ref |
| 14277 | OK272470 | Unassigned | Unassigned | Unassigned | 848_0 | Clustered | 28 | VC_848_0 | 28 | 0.7927 | 1 | 0.7927 | 0 | 0 | 0 | 1 | Ref |
| 14278 | OK272471 | Unassigned | Unassigned | Unassigned | 942_0 | Clustered | 36 | VC_942_0 | 36 | 0.7848 | 1 | 0.7848 | 0 | 0 | 0 | 1 | Ref |
| 14279 | OK272472 | Unassigned | Unassigned | Unassigned | 329_12 | Clustered | 2 | VC_329_12 | 2 | 0.0041 | 0.72605466 | 0.003 | 0 | 0 | 0 | 1 | Ref |
| 14280 | OK272473 | Unassigned | Unassigned | Unassigned | 283_59 | Clustered/Singleton | 1 | VC_283_59 | 1 | 0 | 1 | 0 | 0 | 0 | 0 | 1 | Ref |
| 14281 | OK272474 | Unassigned | Unassigned | Unassigned | 835_0 | Clustered | 67 | VC_835_0 | 67 | 0.9624 | 1 | 0.9624 | 0 | 0 | 0 | 1 | Ref |
| 14282 | OK272476 | Unassigned | Unassigned | Unassigned | 283_60 | Clustered/Singleton | 1 | VC_283_60 | 1 | 0 | 1 | 0 | 0 | 0 | 0 | 1 | Ref |
| 14283 | OK272477 | Unassigned | Unassigned | Unassigned | 190_35 | Clustered/Singleton | 1 | VC_190_35 | 1 | 0 | 1 | 0 | 0 | 0 | 0 | 1 | Ref |
| 14284 | OK272478 | Unassigned | Unassigned | Unassigned | 942_0 | Clustered | 36 | VC_942_0 | 36 | 0.7848 | 1 | 0.7848 | 0 | 0 | 0 | 1 | Ref |
| 14285 | OK272479 | Unassigned | Unassigned | Unassigned | 941_0 | Clustered | 40 | VC_941_0 | 40 | 0.6903 | 1 | 0.6903 | 0 | 0 | 0 | 1 | Ref |
| 14286 | OK272480 | Unassigned | Unassigned | Unassigned | 190_16 | Clustered/Singleton | 1 | VC_190_16 | 1 | 0 | 1 | 0 | 0 | 0 | 0 | 1 | Ref |
| 14287 | OK272481 | Unassigned | Unassigned | Unassigned | 942_0 | Clustered | 36 | VC_942_0 | 36 | 0.7848 | 1 | 0.7848 | 0 | 0 | 0 | 1 | Ref |
| 14288 | OK272482 | Unassigned | Unassigned | Unassigned | 772_19 | Clustered | 16 | VC_772_19 | 16 | 0.0975 | 0.9970466 | 0.0972 | 0 | 0 | 0 | 0.8083 | Ref |
| 14289 | OK272483 | Unassigned | Unassigned | Unassigned | 942_0 | Clustered | 36 | VC_942_0 | 36 | 0.7848 | 1 | 0.7848 | 0 | 0 | 0 | 1 | Ref |
| 14290 | OK272484 | Unassigned | Unassigned | Unassigned | 283_62 | Clustered/Singleton | 1 | VC_283_62 | 1 | 0 | 1 | 0 | 0 | 0 | 0 | 1 | Ref |
| 14291 | OK272485 | Unassigned | Unassigned | Unassigned | 942_0 | Clustered | 36 | VC_942_0 | 36 | 0.7848 | 1 | 0.7848 | 0 | 0 | 0 | 1 | Ref |
| 14292 | OK272486 | Unassigned | Unassigned | Unassigned | 942_0 | Clustered | 36 | VC_942_0 | 36 | 0.7848 | 1 | 0.7848 | 0 | 0 | 0 | 1 | Ref |
| 14293 | OK272487 | Unassigned | Unassigned | Unassigned | 941_0 | Clustered | 40 | VC_941_0 | 40 | 0.6903 | 1 | 0.6903 | 0 | 0 | 0 | 1 | Ref |
| 14294 | OK272488 | Unassigned | Unassigned | Unassigned | 835_0 | Clustered | 67 | VC_835_0 | 67 | 0.9624 | 1 | 0.9624 | 0 | 0 | 0 | 1 | Ref |
| 14295 | OK272489 | Unassigned | Unassigned | Unassigned | 942_0 | Clustered | 36 | VC_942_0 | 36 | 0.7848 | 1 | 0.7848 | 0 | 0 | 0 | 1 | Ref |
| 14296 | OK272490 | Unassigned | Unassigned | Unassigned | 329_3 | Clustered | 24 | VC_329_3 | 24 | 0.1206 | 1 | 0.1206 | 0 | 0 | 0 | 0.7283 | Ref |
| 14297 | OK274152 | Unassigned | Unassigned | Unassigned | 283_122 | Clustered/Singleton | 1 | VC_283_122 | 1 | 0 | 1 | 0 | 0 | 0 | 0 | 1 | Ref |
| 14298 | OK274245 | Unassigned | Unassigned | Unassigned | 83_0 | Clustered | 6 | VC_83_0 | 6 | 0.6436 | 1 | 0.6436 | 0 | 0 | 0 | 1 | Ref |
| 14299 | OK275491 | Unassigned | Unassigned | Unassigned | 78_5 | Clustered | 6 | VC_78_5 | 6 | 0.1087 | 0.99978082 | 0.1087 | 0 | 0 | 0 | 1 | Ref |
| 14300 | OK275492 | Unassigned | Unassigned | Unassigned | 169_0 | Clustered | 20 | VC_169_0 | 20 | 0.7529 | 1 | 0.7529 | 0 | 0 | 0 | 1 | Ref |
| 14301 | OK275493 | Unassigned | Unassigned | Unassigned | 169_0 | Clustered | 20 | VC_169_0 | 20 | 0.7529 | 1 | 0.7529 | 0 | 0 | 0 | 1 | Ref |
| 14302 | OK275494 | Unassigned | Unassigned | Unassigned | 364_0 | Clustered | 16 | VC_364_0 | 16 | 0.6491 | 1 | 0.6491 | 0 | 0 | 0 | 1 | Ref |
| 14303 | OK275495 | Unassigned | Unassigned | Unassigned | 74_0 | Clustered | 15 | VC_74_0 | 15 | 0.5204 | 1 | 0.5204 | 0 | 0 | 0 | 1 | Ref |
| 14304 | OK275722 | Unassigned | Unassigned | Unassigned | 283_109 | Clustered | 2 | VC_283_109 | 2 | 0.0027 | 0.91535723 | 0.0024 | 0 | 0 | 0 | 1 | Ref |
| 14305 | OK283618 | Unassigned | Unassigned | Unassigned | 332_0 | Clustered | 3 | VC_332_0 | 3 | 0.0411 | 0.99858754 | 0.041 | 0 | 0 | 0 | 1 | Ref |
| 14306 | OK283619 | Unassigned | Unassigned | Unassigned | 133_34 | Clustered | 3 | VC_133_34 | 3 | 0.0102 | 0.97369044 | 0.01 | 0 | 0 | 0 | 0.6667 | Ref |
| 14307 | OK288022 | Unassigned | Unassigned | Unassigned | 191_19 | Clustered | 12 | VC_191_19 | 12 | 0.0655 | 1 | 0.0655 | 0 | 0 | 0 | 0.8636 | Ref |
| 14308 | OK310502 | Unassigned | Unassigned | Unassigned | 1071_3 | Clustered | 16 | VC_1071_3 | 16 | 0.2354 | 1 | 0.2354 | 0 | 0 | 0 | 0.9667 | Ref |
| 14309 | OK310503 | Unassigned | Unassigned | Unassigned | 1164_0 | Clustered | 25 | VC_1164_0 | 25 | 0.7312 | 1 | 0.7312 | 0 | 0 | 0 | 1 | Ref |
| 14310 | OK310504 | Unassigned | Unassigned | Unassigned | 1344_0 | Clustered | 78 | VC_1344_0 | 78 | 0.9732 | 1 | 0.9732 | 0 | 0 | 0 | 1 | Ref |
| 14311 | OK310505 | Unassigned | Unassigned | Unassigned | 236_0 | Clustered | 72 | VC_236_0 | 72 | 0.8104 | 1 | 0.8104 | 0 | 0 | 0 | 1 | Ref |
| 14312 | OK310506 | Unassigned | Unassigned | Unassigned | 1240_0 | Clustered | 82 | VC_1240_0 | 82 | 0.7945 | 1 | 0.7945 | 0 | 0 | 0 | 1 | Ref |
| 14313 | OK310512 | Unassigned | Unassigned | Unassigned | 835_0 | Clustered | 67 | VC_835_0 | 67 | 0.9624 | 1 | 0.9624 | 0 | 0 | 0 | 1 | Ref |
| 14314 | OK310513 | Unassigned | Unassigned | Unassigned | 835_0 | Clustered | 67 | VC_835_0 | 67 | 0.9624 | 1 | 0.9624 | 0 | 0 | 0 | 1 | Ref |
| 14315 | OK310514 | Unassigned | Unassigned | Unassigned | 835_0 | Clustered | 67 | VC_835_0 | 67 | 0.9624 | 1 | 0.9624 | 0 | 0 | 0 | 1 | Ref |
| 14316 | OK318957 | Unassigned | Unassigned | Unassigned | 1343_0 | Clustered | 21 | VC_1343_0 | 21 | 0.9843 | 1 | 0.9843 | 0 | 0 | 0 | 1 | Ref |
| 14317 | OK318958 | Unassigned | Unassigned | Unassigned | 1234_0 | Clustered | 44 | VC_1234_0 | 44 | 1 | 1 | 1 | 0 | 0 | 0 | 1 | Ref |
| 14318 | OK318959 | Unassigned | Unassigned | Unassigned | 514_0 | Clustered | 75 | VC_514_0 | 75 | 0.9402 | 1 | 0.9402 | 0 | 0 | 0 | 1 | Ref |
| 14319 | OK318960 | Unassigned | Unassigned | Unassigned | 509_0 | Clustered | 37 | VC_509_0 | 37 | 0.9689 | 1 | 0.9689 | 0 | 0 | 0 | 1 | Ref |
| 14320 | OK318961 | Unassigned | Unassigned | Unassigned | 1163_0 | Clustered | 28 | VC_1163_0 | 28 | 0.4923 | 1 | 0.4923 | 0 | 0 | 0 | 1 | Ref |
| 14321 | OK318991 | Unassigned | Unassigned | Unassigned | 83_0 | Clustered | 6 | VC_83_0 | 6 | 0.6436 | 1 | 0.6436 | 0 | 0 | 0 | 1 | Ref |
| 14322 | OK319016 | Unassigned | Unassigned | Unassigned | 70_0 | Clustered | 27 | VC_70_0 | 27 | 0.5981 | 1 | 0.5981 | 0 | 0 | 0 | 0.9886 | Ref |
| 14323 | OK322699 | Unassigned | Unassigned | Unassigned | 133_22 | Clustered | 13 | VC_133_22 | 13 | 0.0804 | 1 | 0.0804 | 0 | 0 | 0 | 0.8205 | Ref |
| 14324 | OK330455 | Unassigned | Unassigned | Unassigned | 749_0 | Clustered | 40 | VC_749_0 | 40 | 0.7517 | 1 | 0.7517 | 0 | 0 | 0 | 1 | Ref |
| 14325 | OK330456 | Unassigned | Unassigned | Unassigned | 749_0 | Clustered | 40 | VC_749_0 | 40 | 0.7517 | 1 | 0.7517 | 0 | 0 | 0 | 1 | Ref |
| 14326 | OK335775 | Unassigned | Unassigned | Unassigned | 290_5 | Clustered | 14 | VC_290_5 | 14 | 0.1422 | 1 | 0.1422 | 0 | 0 | 0 | 0.9231 | Ref |
| 14327 | OK335783 | Unassigned | Unassigned | Unassigned | 70_0 | Clustered | 27 | VC_70_0 | 27 | 0.5981 | 1 | 0.5981 | 0 | 0 | 0 | 0.9886 | Ref |
| 14328 | OK338687 | Unassigned | Unassigned | Unassigned | 363_0 | Clustered | 2 | VC_363_0 | 2 | 0.8022 | 0.72985431 | 0.5855 | 0 | 0 | 0 | 1 | Ref |
| 14329 | OK340817 | Unassigned | Unassigned | Unassigned | 1040_0 | Clustered | 23 | VC_1040_0 | 23 | 0.9605 | 1 | 0.9605 | 0 | 0 | 0 | 1 | Ref |
| 14330 | OK340818 | Unassigned | Unassigned | Unassigned | 1040_0 | Clustered | 23 | VC_1040_0 | 23 | 0.9605 | 1 | 0.9605 | 0 | 0 | 0 | 1 | Ref |
| 14331 | OK340819 | Unassigned | Unassigned | Unassigned | 1040_0 | Clustered | 23 | VC_1040_0 | 23 | 0.9605 | 1 | 0.9605 | 0 | 0 | 0 | 1 | Ref |
| 14332 | OK340820 | Unassigned | Unassigned | Unassigned | 1157_0 | Clustered | 24 | VC_1157_0 | 24 | 1 | 1 | 1 | 0 | 0 | 0 | 1 | Ref |
| 14333 | OK340821 | Unassigned | Unassigned | Unassigned | 1157_0 | Clustered | 24 | VC_1157_0 | 24 | 1 | 1 | 1 | 0 | 0 | 0 | 1 | Ref |
| 14334 | OK340822 | Unassigned | Unassigned | Unassigned | 1157_0 | Clustered | 24 | VC_1157_0 | 24 | 1 | 1 | 1 | 0 | 0 | 0 | 1 | Ref |
| 14335 | OK349506 | Unassigned | Unassigned | Unassigned | 837_0 | Clustered | 29 | VC_837_0 | 29 | 0.9955 | 1 | 0.9955 | 0 | 0 | 0 | 1 | Ref |
| 14336 | OK349510 | Unassigned | Unassigned | Unassigned | 85_4 | Clustered | 3 | VC_85_4 | 3 | 0.0294 | 0.98125255 | 0.0288 | 0 | 0 | 0 | 1 | Ref |
| 14337 | OK412919 | Unassigned | Unassigned | Unassigned | 981_1 | Clustered | 26 | VC_981_1 | 26 | 0.7009 | 1 | 0.7009 | 0 | 0 | 0 | 1 | Ref |
| 14338 | OK428535 | Unassigned | Unassigned | Unassigned | 959_5 | Clustered | 19 | VC_959_5 | 19 | 0.2626 | 1 | 0.2626 | 0 | 0 | 0 | 1 | Ref |
| 14339 | OK428602 | Unassigned | Unassigned | Unassigned | 134_0 | Clustered | 3 | VC_134_0 | 3 | 0.0956 | 0.99858494 | 0.0955 | 0 | 0 | 0 | 1 | Ref |
| 14340 | OK483199 | Unassigned | Unassigned | Unassigned | 904_2 | Clustered | 7 | VC_904_2 | 7 | 0.2857 | 1 | 0.2857 | 0 | 0 | 0 | 1 | Ref |
| 14341 | OK483201 | Unassigned | Unassigned | Unassigned | 882_0 | Clustered | 20 | VC_882_0 | 20 | 0.9917 | 1 | 0.9917 | 0 | 0 | 0 | 1 | Ref |
| 14342 | OK483342 | Unassigned | Unassigned | Unassigned | 328_0 | Clustered | 11 | VC_328_0 | 11 | 0.4707 | 1 | 0.4707 | 0 | 0 | 0 | 1 | Ref |
| 14343 | OK490394 | Unassigned | Unassigned | Unassigned | 229_0 | Clustered | 32 | VC_229_0 | 32 | 0.9119 | 1 | 0.9119 | 0 | 0 | 0 | 1 | Ref |
| 14344 | OK490395 | Unassigned | Unassigned | Unassigned | 6_0 | Clustered | 44 | VC_6_0 | 44 | 0.908 | 1 | 0.908 | 0 | 0 | 0 | 1 | Ref |
| 14345 | OK490396 | Unassigned | Unassigned | Unassigned | 229_0 | Clustered | 32 | VC_229_0 | 32 | 0.9119 | 1 | 0.9119 | 0 | 0 | 0 | 1 | Ref |
| 14346 | OK490397 | Unassigned | Unassigned | Unassigned | 6_0 | Clustered | 44 | VC_6_0 | 44 | 0.908 | 1 | 0.908 | 0 | 0 | 0 | 1 | Ref |
| 14347 | OK490398 | Unassigned | Unassigned | Unassigned | 229_0 | Clustered | 32 | VC_229_0 | 32 | 0.9119 | 1 | 0.9119 | 0 | 0 | 0 | 1 | Ref |
| 14348 | OK490399 | Unassigned | Unassigned | Unassigned | 6_0 | Clustered | 44 | VC_6_0 | 44 | 0.908 | 1 | 0.908 | 0 | 0 | 0 | 1 | Ref |
| 14349 | OK490400 | Unassigned | Unassigned | Unassigned | 229_0 | Clustered | 32 | VC_229_0 | 32 | 0.9119 | 1 | 0.9119 | 0 | 0 | 0 | 1 | Ref |
| 14350 | OK490401 | Unassigned | Unassigned | Unassigned | 6_0 | Clustered | 44 | VC_6_0 | 44 | 0.908 | 1 | 0.908 | 0 | 0 | 0 | 1 | Ref |
| 14351 | OK490402 | Unassigned | Unassigned | Unassigned | 229_0 | Clustered | 32 | VC_229_0 | 32 | 0.9119 | 1 | 0.9119 | 0 | 0 | 0 | 1 | Ref |
| 14352 | OK490403 | Unassigned | Unassigned | Unassigned | 6_0 | Clustered | 44 | VC_6_0 | 44 | 0.908 | 1 | 0.908 | 0 | 0 | 0 | 1 | Ref |
| 14353 | OK490404 | Unassigned | Unassigned | Unassigned | 229_0 | Clustered | 32 | VC_229_0 | 32 | 0.9119 | 1 | 0.9119 | 0 | 0 | 0 | 1 | Ref |
| 14354 | OK490405 | Unassigned | Unassigned | Unassigned | 6_0 | Clustered | 44 | VC_6_0 | 44 | 0.908 | 1 | 0.908 | 0 | 0 | 0 | 1 | Ref |
| 14355 | OK490406 | Unassigned | Unassigned | Unassigned | 229_0 | Clustered | 32 | VC_229_0 | 32 | 0.9119 | 1 | 0.9119 | 0 | 0 | 0 | 1 | Ref |
| 14356 | OK490407 | Unassigned | Unassigned | Unassigned | 6_0 | Clustered | 44 | VC_6_0 | 44 | 0.908 | 1 | 0.908 | 0 | 0 | 0 | 1 | Ref |
| 14357 | OK490408 | Unassigned | Unassigned | Unassigned | 228_0 | Clustered | 60 | VC_228_0 | 60 | 0.942 | 1 | 0.942 | 0 | 0 | 0 | 1 | Ref |
| 14358 | OK490410 | Unassigned | Unassigned | Unassigned | 31_0 | Clustered | 9 | VC_31_0 | 9 | 0.3435 | 1 | 0.3435 | 0 | 0 | 0 | 0.6944 | Ref |
| 14359 | OK490411 | Unassigned | Unassigned | Unassigned | 6_0 | Clustered | 44 | VC_6_0 | 44 | 0.908 | 1 | 0.908 | 0 | 0 | 0 | 1 | Ref |
| 14360 | OK490412 | Unassigned | Unassigned | Unassigned | 229_0 | Clustered | 32 | VC_229_0 | 32 | 0.9119 | 1 | 0.9119 | 0 | 0 | 0 | 1 | Ref |
| 14361 | OK490413 | Unassigned | Unassigned | Unassigned | 6_0 | Clustered | 44 | VC_6_0 | 44 | 0.908 | 1 | 0.908 | 0 | 0 | 0 | 1 | Ref |
| 14362 | OK490414 | Unassigned | Unassigned | Unassigned | 229_0 | Clustered | 32 | VC_229_0 | 32 | 0.9119 | 1 | 0.9119 | 0 | 0 | 0 | 1 | Ref |
| 14363 | OK490415 | Unassigned | Unassigned | Unassigned | 31_0 | Clustered | 9 | VC_31_0 | 9 | 0.3435 | 1 | 0.3435 | 0 | 0 | 0 | 0.6944 | Ref |
| 14364 | OK490416 | Unassigned | Unassigned | Unassigned | 31_0 | Clustered | 9 | VC_31_0 | 9 | 0.3435 | 1 | 0.3435 | 0 | 0 | 0 | 0.6944 | Ref |
| 14365 | OK490417 | Unassigned | Unassigned | Unassigned | 476_0 | Clustered | 19 | VC_476_0 | 19 | 0.7814 | 1 | 0.7814 | 0 | 0 | 0 | 1 | Ref |
| 14366 | OK490418 | Unassigned | Unassigned | Unassigned | 6_0 | Clustered | 44 | VC_6_0 | 44 | 0.908 | 1 | 0.908 | 0 | 0 | 0 | 1 | Ref |
| 14367 | OK490422 | Unassigned | Unassigned | Unassigned | 229_0 | Clustered | 32 | VC_229_0 | 32 | 0.9119 | 1 | 0.9119 | 0 | 0 | 0 | 1 | Ref |
| 14368 | OK490423 | Unassigned | Unassigned | Unassigned | 6_0 | Clustered | 44 | VC_6_0 | 44 | 0.908 | 1 | 0.908 | 0 | 0 | 0 | 1 | Ref |
| 14369 | OK490424 | Unassigned | Unassigned | Unassigned | 229_0 | Clustered | 32 | VC_229_0 | 32 | 0.9119 | 1 | 0.9119 | 0 | 0 | 0 | 1 | Ref |
| 14370 | OK490425 | Unassigned | Unassigned | Unassigned | 6_0 | Clustered | 44 | VC_6_0 | 44 | 0.908 | 1 | 0.908 | 0 | 0 | 0 | 1 | Ref |
| 14371 | OK490426 | Unassigned | Unassigned | Unassigned | 229_0 | Clustered | 32 | VC_229_0 | 32 | 0.9119 | 1 | 0.9119 | 0 | 0 | 0 | 1 | Ref |
| 14372 | OK490427 | Unassigned | Unassigned | Unassigned | 6_0 | Clustered | 44 | VC_6_0 | 44 | 0.908 | 1 | 0.908 | 0 | 0 | 0 | 1 | Ref |
| 14373 | OK490428 | Unassigned | Unassigned | Unassigned | 229_0 | Clustered | 32 | VC_229_0 | 32 | 0.9119 | 1 | 0.9119 | 0 | 0 | 0 | 1 | Ref |
| 14374 | OK490429 | Unassigned | Unassigned | Unassigned | 6_0 | Clustered | 44 | VC_6_0 | 44 | 0.908 | 1 | 0.908 | 0 | 0 | 0 | 1 | Ref |
| 14375 | OK490430 | Unassigned | Unassigned | Unassigned | 244_0 | Clustered | 12 | VC_244_0 | 12 | 0.4039 | 1 | 0.4039 | 0 | 0 | 0 | 1 | Ref |
| 14376 | OK490431 | Unassigned | Unassigned | Unassigned | 6_0 | Clustered | 44 | VC_6_0 | 44 | 0.908 | 1 | 0.908 | 0 | 0 | 0 | 1 | Ref |
| 14377 | OK490432 | Unassigned | Unassigned | Unassigned | 229_0 | Clustered | 32 | VC_229_0 | 32 | 0.9119 | 1 | 0.9119 | 0 | 0 | 0 | 1 | Ref |
| 14378 | OK490433 | Unassigned | Unassigned | Unassigned | 244_0 | Clustered | 12 | VC_244_0 | 12 | 0.4039 | 1 | 0.4039 | 0 | 0 | 0 | 1 | Ref |
| 14379 | OK490434 | Unassigned | Unassigned | Unassigned | 6_0 | Clustered | 44 | VC_6_0 | 44 | 0.908 | 1 | 0.908 | 0 | 0 | 0 | 1 | Ref |
| 14380 | OK490435 | Unassigned | Unassigned | Unassigned | 229_0 | Clustered | 32 | VC_229_0 | 32 | 0.9119 | 1 | 0.9119 | 0 | 0 | 0 | 1 | Ref |
| 14381 | OK490436 | Unassigned | Unassigned | Unassigned | 6_0 | Clustered | 44 | VC_6_0 | 44 | 0.908 | 1 | 0.908 | 0 | 0 | 0 | 1 | Ref |
| 14382 | OK490437 | Unassigned | Unassigned | Unassigned | 229_0 | Clustered | 32 | VC_229_0 | 32 | 0.9119 | 1 | 0.9119 | 0 | 0 | 0 | 1 | Ref |
| 14383 | OK490438 | Unassigned | Unassigned | Unassigned | 244_0 | Clustered | 12 | VC_244_0 | 12 | 0.4039 | 1 | 0.4039 | 0 | 0 | 0 | 1 | Ref |
| 14384 | OK490439 | Unassigned | Unassigned | Unassigned | 6_0 | Clustered | 44 | VC_6_0 | 44 | 0.908 | 1 | 0.908 | 0 | 0 | 0 | 1 | Ref |
| 14385 | OK490440 | Unassigned | Unassigned | Unassigned | 229_0 | Clustered | 32 | VC_229_0 | 32 | 0.9119 | 1 | 0.9119 | 0 | 0 | 0 | 1 | Ref |
| 14386 | OK490441 | Unassigned | Unassigned | Unassigned | 244_0 | Clustered | 12 | VC_244_0 | 12 | 0.4039 | 1 | 0.4039 | 0 | 0 | 0 | 1 | Ref |
| 14387 | OK490442 | Unassigned | Unassigned | Unassigned | 6_0 | Clustered | 44 | VC_6_0 | 44 | 0.908 | 1 | 0.908 | 0 | 0 | 0 | 1 | Ref |
| 14388 | OK490443 | Unassigned | Unassigned | Unassigned | 229_0 | Clustered | 32 | VC_229_0 | 32 | 0.9119 | 1 | 0.9119 | 0 | 0 | 0 | 1 | Ref |
| 14389 | OK490444 | Unassigned | Unassigned | Unassigned | 244_0 | Clustered | 12 | VC_244_0 | 12 | 0.4039 | 1 | 0.4039 | 0 | 0 | 0 | 1 | Ref |
| 14390 | OK490445 | Unassigned | Unassigned | Unassigned | 6_0 | Clustered | 44 | VC_6_0 | 44 | 0.908 | 1 | 0.908 | 0 | 0 | 0 | 1 | Ref |
| 14391 | OK490446 | Unassigned | Unassigned | Unassigned | 229_0 | Clustered | 32 | VC_229_0 | 32 | 0.9119 | 1 | 0.9119 | 0 | 0 | 0 | 1 | Ref |
| 14392 | OK490447 | Unassigned | Unassigned | Unassigned | 244_0 | Clustered | 12 | VC_244_0 | 12 | 0.4039 | 1 | 0.4039 | 0 | 0 | 0 | 1 | Ref |
| 14393 | OK490448 | Unassigned | Unassigned | Unassigned | 6_0 | Clustered | 44 | VC_6_0 | 44 | 0.908 | 1 | 0.908 | 0 | 0 | 0 | 1 | Ref |
| 14394 | OK490449 | Unassigned | Unassigned | Unassigned | 229_0 | Clustered | 32 | VC_229_0 | 32 | 0.9119 | 1 | 0.9119 | 0 | 0 | 0 | 1 | Ref |
| 14395 | OK490450 | Unassigned | Unassigned | Unassigned | 244_0 | Clustered | 12 | VC_244_0 | 12 | 0.4039 | 1 | 0.4039 | 0 | 0 | 0 | 1 | Ref |
| 14396 | OK490451 | Unassigned | Unassigned | Unassigned | 244_0 | Clustered | 12 | VC_244_0 | 12 | 0.4039 | 1 | 0.4039 | 0 | 0 | 0 | 1 | Ref |
| 14397 | OK490452 | Unassigned | Unassigned | Unassigned | 6_0 | Clustered | 44 | VC_6_0 | 44 | 0.908 | 1 | 0.908 | 0 | 0 | 0 | 1 | Ref |
| 14398 | OK490453 | Unassigned | Unassigned | Unassigned | 229_0 | Clustered | 32 | VC_229_0 | 32 | 0.9119 | 1 | 0.9119 | 0 | 0 | 0 | 1 | Ref |
| 14399 | OK490454 | Unassigned | Unassigned | Unassigned | 244_0 | Clustered | 12 | VC_244_0 | 12 | 0.4039 | 1 | 0.4039 | 0 | 0 | 0 | 1 | Ref |
| 14400 | OK490455 | Unassigned | Unassigned | Unassigned | 6_0 | Clustered | 44 | VC_6_0 | 44 | 0.908 | 1 | 0.908 | 0 | 0 | 0 | 1 | Ref |
| 14401 | OK490456 | Unassigned | Unassigned | Unassigned | 229_0 | Clustered | 32 | VC_229_0 | 32 | 0.9119 | 1 | 0.9119 | 0 | 0 | 0 | 1 | Ref |
| 14402 | OK490457 | Unassigned | Unassigned | Unassigned | 6_0 | Clustered | 44 | VC_6_0 | 44 | 0.908 | 1 | 0.908 | 0 | 0 | 0 | 1 | Ref |
| 14403 | OK490458 | Unassigned | Unassigned | Unassigned | 244_0 | Clustered | 12 | VC_244_0 | 12 | 0.4039 | 1 | 0.4039 | 0 | 0 | 0 | 1 | Ref |
| 14404 | OK490459 | Unassigned | Unassigned | Unassigned | 229_0 | Clustered | 32 | VC_229_0 | 32 | 0.9119 | 1 | 0.9119 | 0 | 0 | 0 | 1 | Ref |
| 14405 | OK490460 | Unassigned | Unassigned | Unassigned | 60_0 | Clustered | 5 | VC_60_0 | 5 | 0.4237 | 0.99999997 | 0.4237 | 0 | 0 | 0 | 1 | Ref |
| 14406 | OK490461 | Unassigned | Unassigned | Unassigned | 228_0 | Clustered | 60 | VC_228_0 | 60 | 0.942 | 1 | 0.942 | 0 | 0 | 0 | 1 | Ref |
| 14407 | OK490463 | Unassigned | Unassigned | Unassigned | 60_0 | Clustered | 5 | VC_60_0 | 5 | 0.4237 | 0.99999997 | 0.4237 | 0 | 0 | 0 | 1 | Ref |
| 14408 | OK490464 | Unassigned | Unassigned | Unassigned | 228_0 | Clustered | 60 | VC_228_0 | 60 | 0.942 | 1 | 0.942 | 0 | 0 | 0 | 1 | Ref |
| 14409 | OK490466 | Unassigned | Unassigned | Unassigned | 60_0 | Clustered | 5 | VC_60_0 | 5 | 0.4237 | 0.99999997 | 0.4237 | 0 | 0 | 0 | 1 | Ref |
| 14410 | OK490467 | Unassigned | Unassigned | Unassigned | 228_0 | Clustered | 60 | VC_228_0 | 60 | 0.942 | 1 | 0.942 | 0 | 0 | 0 | 1 | Ref |
| 14411 | OK490469 | Unassigned | Unassigned | Unassigned | 60_0 | Clustered | 5 | VC_60_0 | 5 | 0.4237 | 0.99999997 | 0.4237 | 0 | 0 | 0 | 1 | Ref |
| 14412 | OK490470 | Unassigned | Unassigned | Unassigned | 228_0 | Clustered | 60 | VC_228_0 | 60 | 0.942 | 1 | 0.942 | 0 | 0 | 0 | 1 | Ref |
| 14413 | OK490472 | Unassigned | Unassigned | Unassigned | 60_0 | Clustered | 5 | VC_60_0 | 5 | 0.4237 | 0.99999997 | 0.4237 | 0 | 0 | 0 | 1 | Ref |
| 14414 | OK490473 | Unassigned | Unassigned | Unassigned | 228_0 | Clustered | 60 | VC_228_0 | 60 | 0.942 | 1 | 0.942 | 0 | 0 | 0 | 1 | Ref |
| 14415 | OK499971 | Unassigned | Unassigned | Unassigned | 31_1 | Clustered/Singleton | 1 | VC_31_1 | 1 | 0 | 1 | 0 | 0 | 0 | 0 | 1 | Ref |
| 14416 | OK499972 | Unassigned | Unassigned | Unassigned | 475_1 | Clustered | 3 | VC_475_1 | 3 | 0.0582 | 0.98866009 | 0.0575 | 0 | 0 | 0 | 1 | Ref |

| | | | | | | | | | | | | | | | | | |
|---|---|---|---|---|---|---|---|---|---|---|---|---|---|---|---|---|---|
| 14417 | OK499973 | Unassigned | Unassigned | Unassigned | 584_0 | Clustered | 28 | VC_584_0 | 28 | 0.7704 | 1 | 0.7704 | 0 | 0 | 0 | 1 | Ref |
| 14418 | OK499974 | Unassigned | Unassigned | Unassigned | 591_0 | Clustered | 14 | VC_591_0 | 14 | 0.5184 | 1 | 0.5184 | 0 | 0 | 0 | 1 | Ref |
| 14419 | OK499975 | Unassigned | Unassigned | Unassigned | 591_0 | Clustered | 14 | VC_591_0 | 14 | 0.5184 | 1 | 0.5184 | 0 | 0 | 0 | 1 | Ref |
| 14420 | OK499976 | Unassigned | Unassigned | Unassigned | 883_1 | Clustered | 12 | VC_883_1 | 12 | 0.2441 | 1 | 0.2441 | 0 | 0 | 0 | 1 | Ref |
| 14421 | OK499977 | Unassigned | Unassigned | Unassigned | 187_0 | Clustered | 4 | VC_187_0 | 4 | 0.0661 | 0.99994098 | 0.0661 | 0 | 0 | 0 | 1 | Ref |
| 14422 | OK499978 | Unassigned | Unassigned | Unassigned | 123_0 | Clustered | 23 | VC_123_0 | 23 | 0.817 | 1 | 0.817 | 0 | 0 | 0 | 1 | Ref |
| 14423 | OK499979 | Unassigned | Unassigned | Unassigned | 704_0 | Clustered | 16 | VC_704_0 | 16 | 0.7274 | 1 | 0.7274 | 0 | 0 | 0 | 1 | Ref |
| 14424 | OK499980 | Unassigned | Unassigned | Unassigned | 491_0 | Clustered | 36 | VC_491_0 | 36 | 0.5336 | 1 | 0.5336 | 0 | 0 | 0 | 0.9714 | Ref |
| 14425 | OK499981 | Unassigned | Unassigned | Unassigned | 491_0 | Clustered | 36 | VC_491_0 | 36 | 0.5336 | 1 | 0.5336 | 0 | 0 | 0 | 0.9714 | Ref |
| 14426 | OK499982 | Unassigned | Unassigned | Unassigned | 390_0 | Clustered | 4 | VC_390_0 | 4 | 0.0468 | 0.9999882 | 0.0468 | 0 | 0 | 0 | 1 | Ref |
| 14427 | OK499983 | Unassigned | Unassigned | Unassigned | 187_0 | Clustered | 4 | VC_187_0 | 4 | 0.0661 | 0.99994098 | 0.0661 | 0 | 0 | 0 | 1 | Ref |
| 14428 | OK499984 | Unassigned | Unassigned | Unassigned | 584_0 | Clustered | 28 | VC_584_0 | 28 | 0.7704 | 1 | 0.7704 | 0 | 0 | 0 | 1 | Ref |
| 14429 | OK499985 | Unassigned | Unassigned | Unassigned | 835_0 | Clustered | 67 | VC_835_0 | 67 | 0.9624 | 1 | 0.9624 | 0 | 0 | 0 | 1 | Ref |
| 14430 | OK499986 | Unassigned | Unassigned | Unassigned | 591_0 | Clustered | 14 | VC_591_0 | 14 | 0.5184 | 1 | 0.5184 | 0 | 0 | 0 | 1 | Ref |
| 14431 | OK499987 | Unassigned | Unassigned | Unassigned | 475_1 | Clustered | 3 | VC_475_1 | 3 | 0.0582 | 0.98866009 | 0.0575 | 0 | 0 | 0 | 1 | Ref |
| 14432 | OK499988 | Unassigned | Unassigned | Unassigned | 835_0 | Clustered | 67 | VC_835_0 | 67 | 0.9624 | 1 | 0.9624 | 0 | 0 | 0 | 1 | Ref |
| 14433 | OK499989 | Unassigned | Unassigned | Unassigned | 288_0 | Clustered | 3 | VC_288_0 | 3 | 0.042 | 0.99856109 | 0.042 | 0 | 0 | 0 | 1 | Ref |
| 14434 | OK499990 | Unassigned | Unassigned | Unassigned | 883_1 | Clustered | 12 | VC_883_1 | 12 | 0.2441 | 1 | 0.2441 | 0 | 0 | 0 | 1 | Ref |
| 14435 | OK499991 | Unassigned | Unassigned | Unassigned | 475_8 | Clustered | 2 | VC_475_8 | 2 | 0.0291 | 0.90184699 | 0.0262 | 0 | 0 | 0 | 1 | Ref |
| 14436 | OK499992 | Unassigned | Unassigned | Unassigned | 475_10 | Clustered | 2 | VC_475_10 | 2 | 0.0285 | 0.87721192 | 0.025 | 0 | 0 | 0 | 1 | Ref |
| 14437 | OK499993 | Unassigned | Unassigned | Unassigned | 835_0 | Clustered | 67 | VC_835_0 | 67 | 0.9624 | 1 | 0.9624 | 0 | 0 | 0 | 1 | Ref |
| 14438 | OK499994 | Unassigned | Unassigned | Unassigned | 475_1 | Clustered | 3 | VC_475_1 | 3 | 0.0582 | 0.98866009 | 0.0575 | 0 | 0 | 0 | 1 | Ref |
| 14439 | OK499995 | Unassigned | Unassigned | Unassigned | 966_0 | Clustered | 3 | VC_966_0 | 3 | 0.3549 | 0.99840333 | 0.3543 | 0 | 0 | 0 | 1 | Ref |
| 14440 | OK499996 | Unassigned | Unassigned | Unassigned | 491_0 | Clustered | 36 | VC_491_0 | 36 | 0.5336 | 1 | 0.5336 | 0 | 0 | 0 | 0.9714 | Ref |
| 14441 | OK499997 | Unassigned | Unassigned | Unassigned | 491_0 | Clustered | 36 | VC_491_0 | 36 | 0.5336 | 1 | 0.5336 | 0 | 0 | 0 | 0.9714 | Ref |
| 14442 | OK499998 | Unassigned | Unassigned | Unassigned | 591_0 | Clustered | 14 | VC_591_0 | 14 | 0.5184 | 1 | 0.5184 | 0 | 0 | 0 | 1 | Ref |
| 14443 | OK499999 | Unassigned | Unassigned | Unassigned | 704_0 | Clustered | 16 | VC_704_0 | 16 | 0.7274 | 1 | 0.7274 | 0 | 0 | 0 | 1 | Ref |
| 14444 | OK500000 | Unassigned | Unassigned | Unassigned | 591_0 | Clustered | 14 | VC_591_0 | 14 | 0.5184 | 1 | 0.5184 | 0 | 0 | 0 | 1 | Ref |
| 14445 | OK500001 | Unassigned | Unassigned | Unassigned | 884_0 | Clustered | 28 | VC_884_0 | 28 | 0.6211 | 1 | 0.6211 | 0 | 0 | 0 | 0.9841 | Ref |
| 14446 | OK523998 | Unassigned | Unassigned | Unassigned | 129_0 | Clustered | 43 | VC_129_0 | 43 | 0.886 | 1 | 0.886 | 0 | 0 | 0 | 1 | Ref |
| 14447 | OK523999 | Unassigned | Unassigned | Unassigned | 129_0 | Clustered | 43 | VC_129_0 | 43 | 0.886 | 1 | 0.886 | 0 | 0 | 0 | 1 | Ref |
| 14448 | OK539620 | Unassigned | Unassigned | Unassigned | 638_0 | Clustered | 45 | VC_638_0 | 45 | 0.7741 | 1 | 0.7741 | 0 | 0 | 0 | 1 | Ref |
| 14449 | OK539824 | Unassigned | Unassigned | Unassigned | 423_0 | Clustered | 45 | VC_423_0 | 45 | 0.9484 | 1 | 0.9484 | 0 | 0 | 0 | 1 | Ref |
| 14450 | OK539825 | Unassigned | Unassigned | Unassigned | 749_0 | Clustered | 40 | VC_749_0 | 40 | 0.7517 | 1 | 0.7517 | 0 | 0 | 0 | 1 | Ref |
| 14451 | OK539826 | Unassigned | Unassigned | Unassigned | 76_0 | Clustered | 37 | VC_76_0 | 37 | 0.9403 | 1 | 0.9403 | 0 | 0 | 0 | 1 | Ref |
| 14452 | OK539913 | Unassigned | Unassigned | Unassigned | 330_0 | Clustered | 27 | VC_330_0 | 27 | 0.5088 | 1 | 0.5088 | 0 | 0 | 0 | 0.8661 | Ref |
| 14453 | OK546191 | Unassigned | Unassigned | Unassigned | 699_0 | Clustered | 71 | VC_699_0 | 71 | 0.9352 | 1 | 0.9352 | 0 | 0 | 0 | 1 | Ref |
| 14454 | OK562429 | Unassigned | Unassigned | Unassigned | 111_0 | Clustered | 91 | VC_111_0 | 91 | 0.9587 | 1 | 0.9587 | 0 | 0 | 0 | 1 | Ref |
| 14455 | OK570184 | Unassigned | Unassigned | Unassigned | 453_0 | Clustered | 6 | VC_453_0 | 6 | 0.114 | 1 | 0.114 | 0 | 0 | 0 | 1 | Ref |
| 14456 | OK570186 | Unassigned | Unassigned | Unassigned | 846_0 | Clustered | 2 | VC_846_0 | 2 | 0.5455 | 0.92357512 | 0.5038 | 0 | 0 | 0 | 1 | Ref |
| 14457 | OK570375 | Unassigned | Unassigned | Unassigned | 283_102 | Clustered/Singleton | 1 | VC_283_102 | 1 | 0 | 1 | 0 | 0 | 0 | 0 | 1 | Ref |
| 14458 | OK583892 | Unassigned | Unassigned | Unassigned | 111_0 | Clustered | 91 | VC_111_0 | 91 | 0.9587 | 1 | 0.9587 | 0 | 0 | 0 | 1 | Ref |
| 14459 | OK583893 | Unassigned | Unassigned | Unassigned | 111_0 | Clustered | 91 | VC_111_0 | 91 | 0.9587 | 1 | 0.9587 | 0 | 0 | 0 | 1 | Ref |
| 14460 | OK585158 | Unassigned | Unassigned | Unassigned | 837_0 | Clustered | 29 | VC_837_0 | 29 | 0.9955 | 1 | 0.9955 | 0 | 0 | 0 | 1 | Ref |
| 14461 | OK585159 | Unassigned | Unassigned | Unassigned | 837_0 | Clustered | 29 | VC_837_0 | 29 | 0.9955 | 1 | 0.9955 | 0 | 0 | 0 | 1 | Ref |
| 14462 | OK585160 | Unassigned | Unassigned | Unassigned | 837_0 | Clustered | 29 | VC_837_0 | 29 | 0.9955 | 1 | 0.9955 | 0 | 0 | 0 | 1 | Ref |
| 14463 | OK625527 | Unassigned | Unassigned | Unassigned | 452_0 | Clustered | 66 | VC_452_0 | 66 | 0.9248 | 1 | 0.9248 | 0 | 0 | 0 | 1 | Ref |
| 14464 | OK625546 | Unassigned | Unassigned | Unassigned | 882_0 | Clustered | 20 | VC_882_0 | 20 | 0.9917 | 1 | 0.9917 | 0 | 0 | 0 | 1 | Ref |
| 14465 | OK631811 | Unassigned | Unassigned | Unassigned | 491_0 | Clustered | 36 | VC_491_0 | 36 | 0.5336 | 1 | 0.5336 | 0 | 0 | 0 | 0.9714 | Ref |
| 14466 | OK631812 | Unassigned | Unassigned | Unassigned | 668_0 | Clustered | 25 | VC_668_0 | 25 | 0.5539 | 1 | 0.5539 | 0 | 0 | 0 | 0.96 | Ref |
| 14467 | OK631813 | Unassigned | Unassigned | Unassigned | 668_0 | Clustered | 25 | VC_668_0 | 25 | 0.5539 | 1 | 0.5539 | 0 | 0 | 0 | 0.96 | Ref |
| 14468 | OK632025 | Unassigned | Unassigned | Unassigned | 831_0 | Clustered | 5 | VC_831_0 | 5 | 0.4369 | 0.99999952 | 0.4369 | 0 | 0 | 0 | 1 | Ref |
| 14469 | OK638201 | Unassigned | Unassigned | Unassigned | 1028_0 | Clustered | 8 | VC_1028_0 | 8 | 0.9323 | 1 | 0.9323 | 0 | 0 | 0 | 1 | Ref |
| 14470 | OK638202 | Unassigned | Unassigned | Unassigned | 1028_0 | Clustered | 8 | VC_1028_0 | 8 | 0.9323 | 1 | 0.9323 | 0 | 0 | 0 | 1 | Ref |
| 14471 | OK638203 | Unassigned | Unassigned | Unassigned | 1028_0 | Clustered | 8 | VC_1028_0 | 8 | 0.9323 | 1 | 0.9323 | 0 | 0 | 0 | 1 | Ref |
| 14472 | OK649958 | Unassigned | Unassigned | Unassigned | 310_0 | Clustered | 33 | VC_310_0 | 33 | 0.8771 | 1 | 0.8771 | 0 | 0 | 0 | 1 | Ref |
| 14473 | OK655936 | Unassigned | Unassigned | Unassigned | 938_0 | Clustered | 99 | VC_938_0 | 99 | 0.9119 | 1 | 0.9119 | 0 | 0 | 0 | 0.9928 | Ref |
| 14474 | OK663600 | Unassigned | Unassigned | Unassigned | 386_0 | Clustered | 49 | VC_386_0 | 49 | 0.7941 | 1 | 0.7941 | 0 | 0 | 0 | 1 | Ref |
| 14475 | OK665488 | Unassigned | Unassigned | Unassigned | 838_0 | Clustered | 29 | VC_838_0 | 29 | 0.8611 | 1 | 0.8611 | 0 | 0 | 0 | 1 | Ref |
| 14476 | OK665835 | Unassigned | Unassigned | Unassigned | 190_28 | Clustered | 5 | VC_190_28 | 5 | 0.0304 | 0.99999994 | 0.0304 | 0 | 0 | 0 | 1 | Ref |
| 14477 | OK665842 | Unassigned | Unassigned | Unassigned | 242_1 | Clustered | 20 | VC_242_1 | 20 | 0.684 | 1 | 0.684 | 0 | 0 | 0 | 0.8368 | Ref |
| 14478 | OK905446 | Unassigned | Unassigned | Unassigned | 531_1 | Clustered | 6 | VC_531_1 | 6 | 0.1481 | 0.99978665 | 0.148 | 0 | 0 | 0 | 1 | Ref |
| 14479 | OK913679 | Unassigned | Unassigned | Unassigned | 116_0 | Clustered | 13 | VC_116_0 | 13 | 1 | 1 | 1 | 0 | 0 | 0 | 1 | Ref |
| 14480 | OK913680 | Unassigned | Unassigned | Unassigned | 116_0 | Clustered | 13 | VC_116_0 | 13 | 1 | 1 | 1 | 0 | 0 | 0 | 1 | Ref |
| 14481 | OK999978 | Unassigned | Unassigned | Unassigned | 514_0 | Clustered | 75 | VC_514_0 | 75 | 0.9402 | 1 | 0.9402 | 0 | 0 | 0 | 1 | Ref |
| 14482 | OK999979 | Unassigned | Unassigned | Unassigned | 1234_0 | Clustered | 44 | VC_1234_0 | 44 | 1 | 1 | 1 | 0 | 0 | 0 | 1 | Ref |
| 14483 | OK999980 | Unassigned | Unassigned | Unassigned | 1073_0 | Clustered | 24 | VC_1073_0 | 24 | 0.8388 | 1 | 0.8388 | 0 | 0 | 0 | 1 | Ref |
| 14484 | OK999981 | Unassigned | Unassigned | Unassigned | 1234_0 | Clustered | 44 | VC_1234_0 | 44 | 1 | 1 | 1 | 0 | 0 | 0 | 1 | Ref |
| 14485 | OL310488 | Unassigned | Unassigned | Unassigned | 283_104 | Clustered/Singleton | 1 | VC_283_104 | 1 | 0 | 1 | 0 | 0 | 0 | 0 | 1 | Ref |
| 14486 | OL314400 | Unassigned | Unassigned | Unassigned | 882_0 | Clustered | 20 | VC_882_0 | 20 | 0.9917 | 1 | 0.9917 | 0 | 0 | 0 | 1 | Ref |
| 14487 | OL343755 | Unassigned | Unassigned | Unassigned | 1028_0 | Clustered | 8 | VC_1028_0 | 8 | 0.9323 | 1 | 0.9323 | 0 | 0 | 0 | 1 | Ref |
| 14488 | OL362041 | Unassigned | Unassigned | Unassigned | 283_123 | Clustered/Singleton | 1 | VC_283_123 | 1 | 0 | 1 | 0 | 0 | 0 | 0 | 1 | Ref |
| 14489 | OL362268 | Unassigned | Unassigned | Unassigned | 79_0 | Clustered | 64 | VC_79_0 | 64 | 0.4185 | 1 | 0.4185 | 0 | 0 | 0 | 0.7649 | Ref |
| 14490 | OL362269 | Unassigned | Unassigned | Unassigned | 79_0 | Clustered | 64 | VC_79_0 | 64 | 0.4185 | 1 | 0.4185 | 0 | 0 | 0 | 0.7649 | Ref |
| 14491 | OL362270 | Unassigned | Unassigned | Unassigned | 878_0 | Clustered | 32 | VC_878_0 | 32 | 0.9965 | 1 | 0.9965 | 0 | 0 | 0 | 1 | Ref |
| 14492 | OL362272 | Unassigned | Unassigned | Unassigned | 452_0 | Clustered | 66 | VC_452_0 | 66 | 0.9248 | 1 | 0.9248 | 0 | 0 | 0 | 1 | Ref |
| 14493 | OL362273 | Unassigned | Unassigned | Unassigned | 452_0 | Clustered | 66 | VC_452_0 | 66 | 0.9248 | 1 | 0.9248 | 0 | 0 | 0 | 1 | Ref |
| 14494 | OL362274 | Unassigned | Unassigned | Unassigned | 461_0 | Clustered | 96 | VC_461_0 | 96 | 0.9307 | 1 | 0.9307 | 0 | 0 | 0 | 0.9575 | Ref |
| 14495 | OL362275 | Unassigned | Unassigned | Unassigned | 451_0 | Clustered | 80 | VC_451_0 | 80 | 0.9991 | 1 | 0.9991 | 0 | 0 | 0 | 1 | Ref |
| 14496 | OL362276 | Unassigned | Unassigned | Unassigned | 451_0 | Clustered | 80 | VC_451_0 | 80 | 0.9991 | 1 | 0.9991 | 0 | 0 | 0 | 1 | Ref |
| 14497 | OL362277 | Unassigned | Unassigned | Unassigned | 460_0 | Clustered | 19 | VC_460_0 | 19 | 0.3744 | 1 | 0.3744 | 0 | 0 | 0 | 1 | Ref |
| 14498 | OL362278 | Unassigned | Unassigned | Unassigned | 111_0 | Clustered | 91 | VC_111_0 | 91 | 0.9587 | 1 | 0.9587 | 0 | 0 | 0 | 1 | Ref |
| 14499 | OL362279 | Unassigned | Unassigned | Unassigned | 111_0 | Clustered | 91 | VC_111_0 | 91 | 0.9587 | 1 | 0.9587 | 0 | 0 | 0 | 1 | Ref |
| 14500 | OL362280 | Unassigned | Unassigned | Unassigned | 149_8 | Clustered | 66 | VC_149_8 | 66 | 0.4565 | 1 | 0.4565 | 0 | 0 | 0 | 0.8182 | Ref |
| 14501 | OL362281 | Unassigned | Unassigned | Unassigned | 385_0 | Clustered | 51 | VC_385_0 | 51 | 0.8754 | 1 | 0.8754 | 0 | 0 | 0 | 1 | Ref |
| 14502 | OL362282 | Unassigned | Unassigned | Unassigned | 385_0 | Clustered | 51 | VC_385_0 | 51 | 0.8754 | 1 | 0.8754 | 0 | 0 | 0 | 1 | Ref |
| 14503 | OL362283 | Unassigned | Unassigned | Unassigned | 385_0 | Clustered | 51 | VC_385_0 | 51 | 0.8754 | 1 | 0.8754 | 0 | 0 | 0 | 1 | Ref |
| 14504 | OL396571 | Unassigned | Unassigned | Unassigned | 37_1 | Clustered | 3 | VC_37_1 | 3 | 0.0833 | 0.9484635 | 0.079 | 0 | 0 | 0 | 1 | Ref |
| 14505 | OL409034 | Unassigned | Unassigned | Unassigned | 823_0 | Clustered | 124 | VC_823_0 | 124 | 0.9492 | 1 | 0.9492 | 0 | 0 | 0 | 1 | Ref |
| 14506 | OL416096 | Unassigned | Unassigned | Unassigned | 112_0 | Clustered | 8 | VC_112_0 | 8 | 0.2537 | 1 | 0.2537 | 0 | 0 | 0 | 1 | Ref |
| 14507 | OL436139 | Unassigned | Unassigned | Unassigned | 637_0 | Clustered | 115 | VC_637_0 | 115 | 0.731 | 1 | 0.731 | 0 | 0 | 0 | 0.8764 | Ref |
| 14508 | OL441337 | Unassigned | Unassigned | Unassigned | 472_0 | Clustered | 47 | VC_472_0 | 47 | 0.9025 | 1 | 0.9025 | 0 | 0 | 0 | 1 | Ref |
| 14509 | OL451225 | Unassigned | Unassigned | Unassigned | 290_1 | Clustered | 8 | VC_290_1 | 8 | 0.0757 | 1 | 0.0757 | 0 | 0 | 0 | 0.8571 | Ref |
| 14510 | OL451946 | Unassigned | Unassigned | Unassigned | 191_15 | Clustered | 2 | VC_191_15 | 2 | 0.0057 | 0.93631009 | 0.0053 | 0 | 0 | 0 | 1 | Ref |
| 14511 | OL455887 | Unassigned | Unassigned | Unassigned | 572_0 | Clustered | 16 | VC_572_0 | 16 | 0.7286 | 1 | 0.7286 | 0 | 0 | 0 | 1 | Ref |
| 14512 | OL455888 | Unassigned | Unassigned | Unassigned | 919_0 | Clustered | 39 | VC_919_0 | 39 | 0.9069 | 1 | 0.9069 | 0 | 0 | 0 | 1 | Ref |
| 14513 | OL455889 | Unassigned | Unassigned | Unassigned | 940_0 | Clustered | 34 | VC_940_0 | 34 | 0.7959 | 1 | 0.7959 | 0 | 0 | 0 | 1 | Ref |
| 14514 | OL455890 | Unassigned | Unassigned | Unassigned | 572_0 | Clustered | 16 | VC_572_0 | 16 | 0.7286 | 1 | 0.7286 | 0 | 0 | 0 | 1 | Ref |
| 14515 | OL455891 | Unassigned | Unassigned | Unassigned | 982_0 | Clustered | 62 | VC_982_0 | 62 | 0.8646 | 1 | 0.8646 | 0 | 0 | 0 | 0.743 | Ref |
| 14516 | OL455892 | Unassigned | Unassigned | Unassigned | 1440_0 | Clustered | 4 | VC_1440_0 | 4 | 1 | 1 | 1 | 0 | 0 | 0 | 1 | Ref |
| 14517 | OL455893 | Unassigned | Unassigned | Unassigned | 372_2 | Clustered | 5 | VC_372_2 | 5 | 0.072 | 0.99999997 | 0.072 | 0 | 0 | 0 | 0.9 | Ref |
| 14518 | OL455894 | Unassigned | Unassigned | Unassigned | 982_0 | Clustered | 62 | VC_982_0 | 62 | 0.8646 | 1 | 0.8646 | 0 | 0 | 0 | 0.743 | Ref |
| 14519 | OL455895 | Unassigned | Unassigned | Unassigned | 572_0 | Clustered | 16 | VC_572_0 | 16 | 0.7286 | 1 | 0.7286 | 0 | 0 | 0 | 1 | Ref |
| 14520 | OL455896 | Unassigned | Unassigned | Unassigned | 1207_0 | Clustered | 29 | VC_1207_0 | 29 | 0.8225 | 1 | 0.8225 | 0 | 0 | 0 | 1 | Ref |
| 14521 | OL455897 | Unassigned | Unassigned | Unassigned | 1224_0 | Clustered | 19 | VC_1224_0 | 19 | 1 | 1 | 1 | 0 | 0 | 0 | 1 | Ref |
| 14522 | OL455898 | Unassigned | Unassigned | Unassigned | 325_62 | Clustered | 3 | VC_325_62 | 3 | 0.0188 | 0.99859194 | 0.0188 | 0 | 0 | 0 | 1 | Ref |
| 14523 | OL455899 | Unassigned | Unassigned | Unassigned | 888_0 | Clustered | 28 | VC_888_0 | 28 | 0.5497 | 1 | 0.5497 | 0 | 0 | 0 | 1 | Ref |
| 14524 | OL473897 | Unassigned | Unassigned | Unassigned | 1028_0 | Clustered | 8 | VC_1028_0 | 8 | 0.9323 | 1 | 0.9323 | 0 | 0 | 0 | 1 | Ref |
| 14525 | OL474125 | Unassigned | Unassigned | Unassigned | 111_0 | Clustered | 91 | VC_111_0 | 91 | 0.9587 | 1 | 0.9587 | 0 | 0 | 0 | 1 | Ref |
| 14526 | OL474141 | Unassigned | Unassigned | Unassigned | 329_11 | Clustered | 15 | VC_329_11 | 15 | 0.0791 | 1 | 0.0791 | 0 | 0 | 0 | 0.9333 | Ref |
| 14527 | OL502173 | Unassigned | Unassigned | Unassigned | 191_15 | Clustered | 2 | VC_191_15 | 2 | 0.0057 | 0.93631009 | 0.0053 | 0 | 0 | 0 | 1 | Ref |
| 14528 | OL505078 | Unassigned | Unassigned | Unassigned | 847_1 | Clustered | 26 | VC_847_1 | 26 | 0.6323 | 1 | 0.6323 | 0 | 0 | 0 | 0.9815 | Ref |
| 14529 | OL505084 | Unassigned | Unassigned | Unassigned | 695_0 | Clustered | 48 | VC_695_0 | 48 | 0.8162 | 1 | 0.8162 | 0 | 0 | 0 | 1 | Ref |
| 14530 | OL505085 | Unassigned | Unassigned | Unassigned | 108_0 | Clustered | 33 | VC_108_0 | 33 | 0.949 | 1 | 0.949 | 0 | 0 | 0 | 1 | Ref |
| 14531 | OL519842 | Unassigned | Unassigned | Unassigned | 839_0 | Clustered | 5 | VC_839_0 | 5 | 0.165 | 0.99999995 | 0.165 | 0 | 0 | 0 | 1 | Ref |
| 14532 | OL519843 | Unassigned | Unassigned | Unassigned | 637_0 | Clustered | 115 | VC_637_0 | 115 | 0.731 | 1 | 0.731 | 0 | 0 | 0 | 0.8764 | Ref |
| 14533 | OL519844 | Unassigned | Unassigned | Unassigned | 838_0 | Clustered | 29 | VC_838_0 | 29 | 0.8611 | 1 | 0.8611 | 0 | 0 | 0 | 1 | Ref |
| 14534 | OL539438 | Unassigned | Unassigned | Unassigned | 1346_0 | Clustered | 10 | VC_1346_0 | 10 | 0.7784 | 1 | 0.7784 | 0 | 0 | 0 | 1 | Ref |
| 14535 | OL539439 | Unassigned | Unassigned | Unassigned | 1346_0 | Clustered | 10 | VC_1346_0 | 10 | 0.7784 | 1 | 0.7784 | 0 | 0 | 0 | 1 | Ref |
| 14536 | OL539440 | Unassigned | Unassigned | Unassigned | 836_0 | Clustered | 9 | VC_836_0 | 9 | 0.7733 | 1 | 0.7733 | 0 | 0 | 0 | 1 | Ref |
| 14537 | OL539442 | Unassigned | Unassigned | Unassigned | 1346_0 | Clustered | 10 | VC_1346_0 | 10 | 0.7784 | 1 | 0.7784 | 0 | 0 | 0 | 1 | Ref |
| 14538 | OL539443 | Unassigned | Unassigned | Unassigned | 584_0 | Clustered | 28 | VC_584_0 | 28 | 0.7704 | 1 | 0.7704 | 0 | 0 | 0 | 1 | Ref |
| 14539 | OL539444 | Unassigned | Unassigned | Unassigned | 584_0 | Clustered | 28 | VC_584_0 | 28 | 0.7704 | 1 | 0.7704 | 0 | 0 | 0 | 1 | Ref |
| 14540 | OL539445 | Unassigned | Unassigned | Unassigned | 942_0 | Clustered | 36 | VC_942_0 | 36 | 0.7848 | 1 | 0.7848 | 0 | 0 | 0 | 1 | Ref |
| 14541 | OL539446 | Unassigned | Unassigned | Unassigned | 942_0 | Clustered | 36 | VC_942_0 | 36 | 0.7848 | 1 | 0.7848 | 0 | 0 | 0 | 1 | Ref |
| 14542 | OL539447 | Unassigned | Unassigned | Unassigned | 455_0 | Clustered | 18 | VC_455_0 | 18 | 0.4598 | 1 | 0.4598 | 0 | 0 | 0 | 1 | Ref |
| 14543 | OL539448 | Unassigned | Unassigned | Unassigned | 938_0 | Clustered | 99 | VC_938_0 | 99 | 0.9119 | 1 | 0.9119 | 0 | 0 | 0 | 0.9928 | Ref |
| 14544 | OL539449 | Unassigned | Unassigned | Unassigned | 695_0 | Clustered | 48 | VC_695_0 | 48 | 0.8162 | 1 | 0.8162 | 0 | 0 | 0 | 1 | Ref |
| 14545 | OL539450 | Unassigned | Unassigned | Unassigned | 937_0 | Clustered | 40 | VC_937_0 | 40 | 0.6809 | 1 | 0.6809 | 0 | 0 | 0 | 0.9359 | Ref |
| 14546 | OL539451 | Unassigned | Unassigned | Unassigned | 455_0 | Clustered | 18 | VC_455_0 | 18 | 0.4598 | 1 | 0.4598 | 0 | 0 | 0 | 1 | Ref |
| 14547 | OL539452 | Unassigned | Unassigned | Unassigned | 966_0 | Clustered | 3 | VC_966_0 | 3 | 0.3549 | 0.99840333 | 0.3543 | 0 | 0 | 0 | 1 | Ref |
| 14548 | OL539453 | Unassigned | Unassigned | Unassigned | 455_0 | Clustered | 18 | VC_455_0 | 18 | 0.4598 | 1 | 0.4598 | 0 | 0 | 0 | 1 | Ref |
| 14549 | OL539454 | Unassigned | Unassigned | Unassigned | 795_0 | Clustered | 19 | VC_795_0 | 19 | 0.6848 | 1 | 0.6848 | 0 | 0 | 0 | 1 | Ref |
| 14550 | OL539455 | Unassigned | Unassigned | Unassigned | 1346_0 | Clustered | 10 | VC_1346_0 | 10 | 0.7784 | 1 | 0.7784 | 0 | 0 | 0 | 1 | Ref |
| 14551 | OL539456 | Unassigned | Unassigned | Unassigned | 1346_0 | Clustered | 10 | VC_1346_0 | 10 | 0.7784 | 1 | 0.7784 | 0 | 0 | 0 | 1 | Ref |
| 14552 | OL539458 | Unassigned | Unassigned | Unassigned | 491_0 | Clustered | 36 | VC_491_0 | 36 | 0.5336 | 1 | 0.5336 | 0 | 0 | 0 | 0.9714 | Ref |
| 14553 | OL539459 | Unassigned | Unassigned | Unassigned | 941_0 | Clustered | 40 | VC_941_0 | 40 | 0.6903 | 1 | 0.6903 | 0 | 0 | 0 | 1 | Ref |
| 14554 | OL539460 | Unassigned | Unassigned | Unassigned | 938_0 | Clustered | 99 | VC_938_0 | 99 | 0.9119 | 1 | 0.9119 | 0 | 0 | 0 | 0.9928 | Ref |
| 14555 | OL539461 | Unassigned | Unassigned | Unassigned | 584_0 | Clustered | 28 | VC_584_0 | 28 | 0.7704 | 1 | 0.7704 | 0 | 0 | 0 | 1 | Ref |
| 14556 | OL539462 | Unassigned | Unassigned | Unassigned | 584_0 | Clustered | 28 | VC_584_0 | 28 | 0.7704 | 1 | 0.7704 | 0 | 0 | 0 | 1 | Ref |
| 14557 | OL539464 | Unassigned | Unassigned | Unassigned | 1346_0 | Clustered | 10 | VC_1346_0 | 10 | 0.7784 | 1 | 0.7784 | 0 | 0 | 0 | 1 | Ref |
| 14558 | OL539465 | Unassigned | Unassigned | Unassigned | 584_0 | Clustered | 28 | VC_584_0 | 28 | 0.7704 | 1 | 0.7704 | 0 | 0 | 0 | 1 | Ref |
| 14559 | OL539466 | Unassigned | Unassigned | Unassigned | 584_0 | Clustered | 28 | VC_584_0 | 28 | 0.7704 | 1 | 0.7704 | 0 | 0 | 0 | 1 | Ref |
| 14560 | OL539467 | Unassigned | Unassigned | Unassigned | 455_0 | Clustered | 18 | VC_455_0 | 18 | 0.4598 | 1 | 0.4598 | 0 | 0 | 0 | 1 | Ref |
| 14561 | OL539468 | Unassigned | Unassigned | Unassigned | 488_1 | Clustered | 8 | VC_488_1 | 8 | 0.2927 | 1 | 0.2927 | 0 | 0 | 0 | 1 | Ref |
| 14562 | OL539470 | Unassigned | Unassigned | Unassigned | 669_0 | Clustered | 21 | VC_669_0 | 21 | 0.3623 | 1 | 0.3623 | 0 | 0 | 0 | 1 | Ref |
| 14563 | OL539471 | Unassigned | Unassigned | Unassigned | 669_0 | Clustered | 21 | VC_669_0 | 21 | 0.3623 | 1 | 0.3623 | 0 | 0 | 0 | 1 | Ref |
| 14564 | OL539472 | Unassigned | Unassigned | Unassigned | 937_0 | Clustered | 40 | VC_937_0 | 40 | 0.6809 | 1 | 0.6809 | 0 | 0 | 0 | 0.9359 | Ref |
| 14565 | OL539727 | Unassigned | Unassigned | Unassigned | 638_0 | Clustered | 45 | VC_638_0 | 45 | 0.7741 | 1 | 0.7741 | 0 | 0 | 0 | 1 | Ref |
| 14566 | OL539729 | Unassigned | Unassigned | Unassigned | 451_0 | Clustered | 80 | VC_451_0 | 80 | 0.9991 | 1 | 0.9991 | 0 | 0 | 0 | 1 | Ref |
| 14567 | OL547738 | Unassigned | Unassigned | Unassigned | 898_0 | Clustered | 10 | VC_898_0 | 10 | 0.7284 | 1 | 0.7284 | 0 | 0 | 0 | 1 | Ref |
| 14568 | OL549188 | Unassigned | Unassigned | Unassigned | 1207_0 | Clustered | 29 | VC_1207_0 | 29 | 0.8225 | 1 | 0.8225 | 0 | 0 | 0 | 1 | Ref |
| 14569 | OL549189 | Unassigned | Unassigned | Unassigned | 1207_0 | Clustered | 29 | VC_1207_0 | 29 | 0.8225 | 1 | 0.8225 | 0 | 0 | 0 | 1 | Ref |
| 14570 | OL549190 | Unassigned | Unassigned | Unassigned | 325_18 | Clustered/Singleton | 1 | VC_325_18 | 1 | 0 | 1 | 0 | 0 | 0 | 0 | 1 | Ref |
| 14571 | OL549191 | Unassigned | Unassigned | Unassigned | 1207_0 | Clustered | 29 | VC_1207_0 | 29 | 0.8225 | 1 | 0.8225 | 0 | 0 | 0 | 1 | Ref |
| 14572 | OL549192 | Unassigned | Unassigned | Unassigned | 1207_0 | Clustered | 29 | VC_1207_0 | 29 | 0.8225 | 1 | 0.8225 | 0 | 0 | 0 | 1 | Ref |
| 14573 | OL551674 | Unassigned | Unassigned | Unassigned | 669_0 | Clustered | 21 | VC_669_0 | 21 | 0.3623 | 1 | 0.3623 | 0 | 0 | 0 | 1 | Ref |
| 14574 | OL580764 | Unassigned | Unassigned | Unassigned | 89_1 | Clustered | 2 | VC_89_1 | 2 | 0.0447 | 0.9464689 | 0.0423 | 0 | 0 | 0 | 1 | Ref |
| 14575 | OL581611 | Unassigned | Unassigned | Unassigned | 129_0 | Clustered | 43 | VC_129_0 | 43 | 0.886 | 1 | 0.886 | 0 | 0 | 0 | 1 | Ref |
| 14576 | OL604152 | Unassigned | Unassigned | Unassigned | 1049_0 | Clustered | 8 | VC_1049_0 | 8 | 0.6413 | 1 | 0.6413 | 0 | 0 | 0 | 1 | Ref |
| 14577 | OL606627 | Unassigned | Unassigned | Unassigned | 592_0 | Clustered | 50 | VC_592_0 | 50 | 0.7314 | 1 | 0.7314 | 0 | 0 | 0 | 1 | Ref |
| 14578 | OL614104 | Unassigned | Unassigned | Unassigned | 127_1 | Clustered/Singleton | 1 | VC_127_1 | 1 | 0 | 1 | 0 | 0 | 0 | 0 | 1 | Ref |

| | | | | | | | | | | | | | | | | | |
|---|---|---|---|---|---|---|---|---|---|---|---|---|---|---|---|---|---|
| 14579 | OL615010 | Unassigned | Unassigned | Unassigned | 835_0 | Clustered | 67 | VC_835_0 | 67 | 0.9624 | 1 | 0.9624 | 0 | 0 | 0 | 1 | Ref |
| 14580 | OL615011 | Unassigned | Unassigned | Unassigned | 131_2 | Clustered | 7 | VC_131_2 | 7 | 0.0685 | 1 | 0.0685 | 0 | 0 | 0 | 0.9048 | Ref |
| 14581 | OL615012 | Unassigned | Unassigned | Unassigned | 283_26 | Clustered/Singleton | 1 | VC_283_26 | 1 | 0 | 1 | 0 | 0 | 0 | 0 | 1 | Ref |
| 14582 | OL615013 | Unassigned | Unassigned | Unassigned | 131_2 | Clustered | 7 | VC_131_2 | 7 | 0.0685 | 1 | 0.0685 | 0 | 0 | 0 | 0.9048 | Ref |
| 14583 | OL617020 | Unassigned | Unassigned | Unassigned | 1402_0 | Clustered | 14 | VC_1402_0 | 14 | 0.1673 | 0.51390354 | 0.086 | 0 | 0 | 0 | 1 | Ref |
| 14584 | OL617021 | Unassigned | Unassigned | Unassigned | 1403_0 | Clustered | 28 | VC_1403_0 | 28 | 0.753 | 1 | 0.753 | 0 | 0 | 0 | 1 | Ref |
| 14585 | OL617022 | Unassigned | Unassigned | Unassigned | 1403_0 | Clustered | 28 | VC_1403_0 | 28 | 0.753 | 1 | 0.753 | 0 | 0 | 0 | 1 | Ref |
| 14586 | OL617023 | Unassigned | Unassigned | Unassigned | 1233_0 | Clustered | 16 | VC_1233_0 | 16 | 0.8971 | 1 | 0.8971 | 0 | 0 | 0 | 1 | Ref |
| 14587 | OL617024 | Unassigned | Unassigned | Unassigned | 1402_0 | Clustered | 14 | VC_1402_0 | 14 | 0.1673 | 0.51390354 | 0.086 | 0 | 0 | 0 | 1 | Ref |
| 14588 | OL617025 | Unassigned | Unassigned | Unassigned | 1403_0 | Clustered | 28 | VC_1403_0 | 28 | 0.753 | 1 | 0.753 | 0 | 0 | 0 | 1 | Ref |
| 14589 | OL617026 | Unassigned | Unassigned | Unassigned | 1402_0 | Clustered | 14 | VC_1402_0 | 14 | 0.1673 | 0.51390354 | 0.086 | 0 | 0 | 0 | 1 | Ref |
| 14590 | OL617027 | Unassigned | Unassigned | Unassigned | 1403_0 | Clustered | 28 | VC_1403_0 | 28 | 0.753 | 1 | 0.753 | 0 | 0 | 0 | 1 | Ref |
| 14591 | OL617028 | Unassigned | Unassigned | Unassigned | 1243_0 | Clustered | 52 | VC_1243_0 | 52 | 0.9126 | 0.92708311 | 0.8461 | 0 | 0 | 0 | 1 | Ref |
| 14592 | OL617029 | Unassigned | Unassigned | Unassigned | 995_0 | Clustered | 69 | VC_995_0 | 69 | 0.8547 | 1 | 0.8547 | 0 | 0 | 0 | 1 | Ref |
| 14593 | OL617030 | Unassigned | Unassigned | Unassigned | 1077_0 | Clustered | 48 | VC_1077_0 | 48 | 0.8525 | 1 | 0.8525 | 0 | 0 | 0 | 1 | Ref |
| 14594 | OL617031 | Unassigned | Unassigned | Unassigned | 1077_0 | Clustered | 48 | VC_1077_0 | 48 | 0.8525 | 1 | 0.8525 | 0 | 0 | 0 | 1 | Ref |
| 14595 | OL617032 | Unassigned | Unassigned | Unassigned | 1062_0 | Clustered | 45 | VC_1062_0 | 45 | 0.9734 | 0.50949847 | 0.4959 | 0 | 0 | 0 | 1 | Ref |
| 14596 | OL617033 | Unassigned | Unassigned | Unassigned | 1062_0 | Clustered | 45 | VC_1062_0 | 45 | 0.9734 | 0.50949847 | 0.4959 | 0 | 0 | 0 | 1 | Ref |
| 14597 | OL617035 | Unassigned | Unassigned | Unassigned | 198_0 | Clustered | 28 | VC_198_0 | 28 | 0.2429 | 0.99971175 | 0.2429 | 0 | 0 | 0 | 1 | Ref |
| 14598 | OL617036 | Unassigned | Unassigned | Unassigned | 1285_0 | Clustered | 25 | VC_1285_0 | 25 | 1 | 1 | 1 | 0 | 0 | 0 | 1 | Ref |
| 14599 | OL617037 | Unassigned | Unassigned | Unassigned | 1222_0 | Clustered | 3 | VC_1222_0 | 3 | 1 | 0 | 0 | 0 | 0 | 0 | 1 | Ref |
| 14600 | OL617041 | Unassigned | Unassigned | Unassigned | 460_0 | Clustered | 19 | VC_460_0 | 19 | 0.3744 | 1 | 0.3744 | 0 | 0 | 0 | 1 | Ref |
| 14601 | OL617042 | Unassigned | Unassigned | Unassigned | 35_1 | Clustered/Singleton | 1 | VC_35_1 | 1 | 0 | 1 | 0 | 0 | 0 | 0 | 1 | Ref |
| 14602 | OL622073 | Unassigned | Unassigned | Unassigned | 1028_0 | Clustered | 8 | VC_1028_0 | 8 | 0.9323 | 1 | 0.9323 | 0 | 0 | 0 | 1 | Ref |
| 14603 | OL634959 | Unassigned | Unassigned | Unassigned | 154_0 | Clustered | 4 | VC_154_0 | 4 | 0.1766 | 0.99995106 | 0.1766 | 0 | 0 | 0 | 1 | Ref |
| 14604 | OL652658 | Unassigned | Unassigned | Unassigned | 637_0 | Clustered | 115 | VC_637_0 | 115 | 0.731 | 1 | 0.731 | 0 | 0 | 0 | 0.8764 | Ref |
| 14605 | OL656105 | Unassigned | Unassigned | Unassigned | 1201_0 | Clustered | 2 | VC_1201_0 | 2 | 0.8919 | 0.81445332 | 0.7264 | 0 | 0 | 0 | 1 | Ref |
| 14606 | OL656110 | Unassigned | Unassigned | Unassigned | 131_1 | Clustered | 6 | VC_131_1 | 6 | 0.057 | 0.99999999 | 0.057 | 0 | 0 | 0 | 1 | Ref |
| 14607 | OL657226 | Unassigned | Unassigned | Unassigned | 43_0 | Clustered | 38 | VC_43_0 | 38 | 0.5885 | 1 | 0.5885 | 0 | 0 | 0 | 0.9659 | Ref |
| 14608 | OL657227 | Unassigned | Unassigned | Unassigned | 43_0 | Clustered | 38 | VC_43_0 | 38 | 0.5885 | 1 | 0.5885 | 0 | 0 | 0 | 0.9659 | Ref |
| 14609 | OL657228 | Unassigned | Unassigned | Unassigned | 43_0 | Clustered | 38 | VC_43_0 | 38 | 0.5885 | 1 | 0.5885 | 0 | 0 | 0 | 0.9659 | Ref |
| 14610 | OL658619 | Unassigned | Unassigned | Unassigned | 498_0 | Clustered | 8 | VC_498_0 | 8 | 0.3987 | 1 | 0.3987 | 0 | 0 | 0 | 1 | Ref |
| 14611 | OL674249 | Unassigned | Unassigned | Unassigned | 938_0 | Clustered | 99 | VC_938_0 | 99 | 0.9119 | 1 | 0.9119 | 0 | 0 | 0 | 0.9928 | Ref |
| 14612 | OL674541 | Unassigned | Unassigned | Unassigned | 169_0 | Clustered | 20 | VC_169_0 | 20 | 0.7529 | 1 | 0.7529 | 0 | 0 | 0 | 1 | Ref |
| 14613 | OL676770 | Unassigned | Unassigned | Unassigned | 758_0 | Clustered | 18 | VC_758_0 | 18 | 0.738 | 1 | 0.738 | 0 | 0 | 0 | 1 | Ref |
| 14614 | OL689226 | Unassigned | Unassigned | Unassigned | 1467_0 | Clustered | 3 | VC_1467_0 | 3 | 1 | 1 | 1 | 0 | 0 | 0 | 1 | Ref |
| 14615 | OL689227 | Unassigned | Unassigned | Unassigned | 1467_0 | Clustered | 3 | VC_1467_0 | 3 | 1 | 1 | 1 | 0 | 0 | 0 | 1 | Ref |
| 14616 | OL689230 | Unassigned | Unassigned | Unassigned | 1467_0 | Clustered | 3 | VC_1467_0 | 3 | 1 | 1 | 1 | 0 | 0 | 0 | 1 | Ref |
| 14617 | OL690335 | Unassigned | Unassigned | Unassigned | 8_0 | Clustered | 58 | VC_8_0 | 58 | 0.8306 | 1 | 0.8306 | 0 | 0 | 0 | 0.9946 | Ref |
| 14618 | OL691920 | Unassigned | Unassigned | Unassigned | 385_0 | Clustered | 51 | VC_385_0 | 51 | 0.8754 | 1 | 0.8754 | 0 | 0 | 0 | 1 | Ref |
| 14619 | OL699877 | Unassigned | Unassigned | Unassigned | 592_0 | Clustered | 50 | VC_592_0 | 50 | 0.7314 | 1 | 0.7314 | 0 | 0 | 0 | 1 | Ref |
| 14620 | OL702938 | Unassigned | Unassigned | Unassigned | 938_0 | Clustered | 99 | VC_938_0 | 99 | 0.9119 | 1 | 0.9119 | 0 | 0 | 0 | 0.9928 | Ref |
| 14621 | OL702939 | Unassigned | Unassigned | Unassigned | 506_0 | Clustered | 8 | VC_506_0 | 8 | 0.7376 | 1 | 0.7376 | 0 | 0 | 0 | 1 | Ref |
| 14622 | OL739525 | Unassigned | Unassigned | Unassigned | 847_1 | Clustered | 26 | VC_847_1 | 26 | 0.6323 | 1 | 0.6323 | 0 | 0 | 0 | 0.9815 | Ref |
| 14623 | OL741057 | Unassigned | Unassigned | Unassigned | 149_4 | Clustered/Singleton | 1 | VC_149_4 | 1 | 0 | 1 | 0 | 0 | 0 | 0 | 1 | Ref |
| 14624 | OL741059 | Unassigned | Unassigned | Unassigned | 792_0 | Clustered | 35 | VC_792_0 | 35 | 0.9828 | 1 | 0.9828 | 0 | 0 | 0 | 1 | Ref |
| 14625 | OL741431 | Unassigned | Unassigned | Unassigned | 591_0 | Clustered | 14 | VC_591_0 | 14 | 0.5184 | 1 | 0.5184 | 0 | 0 | 0 | 1 | Ref |
| 14626 | OL741432 | Unassigned | Unassigned | Unassigned | 283_118 | Clustered | 3 | VC_283_118 | 3 | 0.0054 | 0.99110781 | 0.0053 | 0 | 0 | 0 | 1 | Ref |
| 14627 | OL742559 | Unassigned | Unassigned | Unassigned | 551_6 | Clustered | 22 | VC_551_6 | 22 | 0.1214 | 1 | 0.1214 | 0 | 0 | 0 | 0.8095 | Ref |
| 14628 | OL742560 | Unassigned | Unassigned | Unassigned | 1075_0 | Clustered | 4 | VC_1075_0 | 4 | 0.1483 | 0.99998427 | 0.1483 | 0 | 0 | 0 | 1 | Ref |
| 14629 | OL742561 | Unassigned | Unassigned | Unassigned | 521_0 | Clustered | 37 | VC_521_0 | 37 | 0.912 | 1 | 0.912 | 0 | 0 | 0 | 1 | Ref |
| 14630 | OL742562 | Unassigned | Unassigned | Unassigned | 1234_0 | Clustered | 44 | VC_1234_0 | 44 | 1 | 1 | 1 | 0 | 0 | 0 | 1 | Ref |
| 14631 | OL742563 | Unassigned | Unassigned | Unassigned | 982_0 | Clustered | 62 | VC_982_0 | 62 | 0.8646 | 1 | 0.8646 | 0 | 0 | 0 | 0.743 | Ref |
| 14632 | OL743187 | Unassigned | Unassigned | Unassigned | 698_0 | Clustered | 26 | VC_698_0 | 26 | 0.9366 | 1 | 0.9366 | 0 | 0 | 0 | 1 | Ref |
| 14633 | OL744111 | Unassigned | Unassigned | Unassigned | 817_0 | Clustered | 14 | VC_817_0 | 14 | 0.6342 | 1 | 0.6342 | 0 | 0 | 0 | 1 | Ref |
| 14634 | OL744112 | Unassigned | Unassigned | Unassigned | 818_0 | Clustered | 20 | VC_818_0 | 20 | 0.9601 | 1 | 0.9601 | 0 | 0 | 0 | 1 | Ref |
| 14635 | OL744209 | Unassigned | Unassigned | Unassigned | 385_0 | Clustered | 51 | VC_385_0 | 51 | 0.8754 | 1 | 0.8754 | 0 | 0 | 0 | 1 | Ref |
| 14636 | OL744210 | Unassigned | Unassigned | Unassigned | 938_0 | Clustered | 99 | VC_938_0 | 99 | 0.9119 | 1 | 0.9119 | 0 | 0 | 0 | 0.9928 | Ref |
| 14637 | OL744211 | Unassigned | Unassigned | Unassigned | 849_6 | Clustered | 7 | VC_849_6 | 7 | 0.164 | 0.99999999 | 0.164 | 0 | 0 | 0 | 0.9524 | Ref |
| 14638 | OL744212 | Unassigned | Unassigned | Unassigned | 385_0 | Clustered | 51 | VC_385_0 | 51 | 0.8754 | 1 | 0.8754 | 0 | 0 | 0 | 1 | Ref |
| 14639 | OL744213 | Unassigned | Unassigned | Unassigned | 938_0 | Clustered | 99 | VC_938_0 | 99 | 0.9119 | 1 | 0.9119 | 0 | 0 | 0 | 0.9928 | Ref |
| 14640 | OL744214 | Unassigned | Unassigned | Unassigned | 849_6 | Clustered | 7 | VC_849_6 | 7 | 0.164 | 0.99999999 | 0.164 | 0 | 0 | 0 | 0.9524 | Ref |
| 14641 | OL744215 | Unassigned | Unassigned | Unassigned | 452_0 | Clustered | 66 | VC_452_0 | 66 | 0.9248 | 1 | 0.9248 | 0 | 0 | 0 | 1 | Ref |
| 14642 | OL744216 | Unassigned | Unassigned | Unassigned | 849_6 | Clustered | 7 | VC_849_6 | 7 | 0.164 | 0.99999999 | 0.164 | 0 | 0 | 0 | 0.9524 | Ref |
| 14643 | OL744217 | Unassigned | Unassigned | Unassigned | 385_0 | Clustered | 51 | VC_385_0 | 51 | 0.8754 | 1 | 0.8754 | 0 | 0 | 0 | 1 | Ref |
| 14644 | OL744218 | Unassigned | Unassigned | Unassigned | 452_0 | Clustered | 66 | VC_452_0 | 66 | 0.9248 | 1 | 0.9248 | 0 | 0 | 0 | 1 | Ref |
| 14645 | OL744219 | Unassigned | Unassigned | Unassigned | 849_6 | Clustered | 7 | VC_849_6 | 7 | 0.164 | 0.99999999 | 0.164 | 0 | 0 | 0 | 0.9524 | Ref |
| 14646 | OL744220 | Unassigned | Unassigned | Unassigned | 849_6 | Clustered | 7 | VC_849_6 | 7 | 0.164 | 0.99999999 | 0.164 | 0 | 0 | 0 | 0.9524 | Ref |
| 14647 | OL754588 | Unassigned | Unassigned | Unassigned | 79_0 | Clustered | 64 | VC_79_0 | 64 | 0.4185 | 1 | 0.4185 | 0 | 0 | 0 | 0.7649 | Ref |
| 14648 | OL754589 | Unassigned | Unassigned | Unassigned | 423_0 | Clustered | 45 | VC_423_0 | 45 | 0.9484 | 1 | 0.9484 | 0 | 0 | 0 | 1 | Ref |
| 14649 | OL762410 | Unassigned | Unassigned | Unassigned | 807_0 | Clustered | 17 | VC_807_0 | 17 | 0.907 | 1 | 0.907 | 0 | 0 | 0 | 1 | Ref |
| 14650 | OL763419 | Unassigned | Unassigned | Unassigned | 166_0 | Clustered | 26 | VC_166_0 | 26 | 0.7923 | 1 | 0.7923 | 0 | 0 | 0 | 1 | Ref |
| 14651 | OL763420 | Unassigned | Unassigned | Unassigned | 492_1 | Clustered | 20 | VC_492_1 | 20 | 0.4176 | 1 | 0.4176 | 0 | 0 | 0 | 1 | Ref |
| 14652 | OL770073 | Unassigned | Unassigned | Unassigned | 283_177 | Clustered/Singleton | 1 | VC_283_177 | 1 | 0 | 1 | 0 | 0 | 0 | 0 | 1 | Ref |
| 14653 | OL770074 | Unassigned | Unassigned | Unassigned | 286_12 | Clustered | 3 | VC_286_12 | 3 | 0.0146 | 0.92328265 | 0.0135 | 0 | 0 | 0 | 1 | Ref |
| 14654 | OL770107 | Unassigned | Unassigned | Unassigned | 290_6 | Clustered | 12 | VC_290_6 | 12 | 0.1149 | 1 | 0.1149 | 0 | 0 | 0 | 0.8182 | Ref |
| 14655 | OL770108 | Unassigned | Unassigned | Unassigned | 575_0 | Clustered | 35 | VC_575_0 | 35 | 0.8815 | 1 | 0.8815 | 0 | 0 | 0 | 1 | Ref |
| 14656 | OL770258 | Unassigned | Unassigned | Unassigned | 492_1 | Clustered | 20 | VC_492_1 | 20 | 0.4176 | 1 | 0.4176 | 0 | 0 | 0 | 1 | Ref |
| 14657 | OL770259 | Unassigned | Unassigned | Unassigned | 492_1 | Clustered | 20 | VC_492_1 | 20 | 0.4176 | 1 | 0.4176 | 0 | 0 | 0 | 1 | Ref |
| 14658 | OL770260 | Unassigned | Unassigned | Unassigned | 492_2 | Clustered | 8 | VC_492_2 | 8 | 0.1221 | 0.98786574 | 0.1206 | 0 | 0 | 0 | 1 | Ref |
| 14659 | OL770261 | Unassigned | Unassigned | Unassigned | 492_1 | Clustered | 20 | VC_492_1 | 20 | 0.4176 | 1 | 0.4176 | 0 | 0 | 0 | 1 | Ref |
| 14660 | OL770262 | Unassigned | Unassigned | Unassigned | 492_1 | Clustered | 20 | VC_492_1 | 20 | 0.4176 | 1 | 0.4176 | 0 | 0 | 0 | 1 | Ref |
| 14661 | OL770263 | Unassigned | Unassigned | Unassigned | 699_0 | Clustered | 71 | VC_699_0 | 71 | 0.9352 | 1 | 0.9352 | 0 | 0 | 0 | 1 | Ref |
| 14662 | OL770276 | Unassigned | Unassigned | Unassigned | 637_0 | Clustered | 115 | VC_637_0 | 115 | 0.731 | 1 | 0.731 | 0 | 0 | 0 | 0.8764 | Ref |
| 14663 | OL770278 | Unassigned | Unassigned | Unassigned | 835_0 | Clustered | 67 | VC_835_0 | 67 | 0.9624 | 1 | 0.9624 | 0 | 0 | 0 | 1 | Ref |
| 14664 | OL770310 | Unassigned | Unassigned | Unassigned | 191_17 | Clustered | 4 | VC_191_17 | 4 | 0.0166 | 0.99930063 | 0.0166 | 0 | 0 | 0 | 0.8333 | Ref |
| 14665 | OL770365 | Unassigned | Unassigned | Unassigned | 793_0 | Clustered | 12 | VC_793_0 | 12 | 0.8531 | 1 | 0.8531 | 0 | 0 | 0 | 1 | Ref |
| 14666 | OL773676 | Unassigned | Unassigned | Unassigned | 451_0 | Clustered | 80 | VC_451_0 | 80 | 0.9991 | 1 | 0.9991 | 0 | 0 | 0 | 1 | Ref |
| 14667 | OL773677 | Unassigned | Unassigned | Unassigned | 451_0 | Clustered | 80 | VC_451_0 | 80 | 0.9991 | 1 | 0.9991 | 0 | 0 | 0 | 1 | Ref |
| 14668 | OL774866 | Unassigned | Unassigned | Unassigned | 1187_0 | Clustered | 3 | VC_1187_0 | 3 | 0.924 | 0.92554266 | 0.8552 | 0 | 0 | 0 | 1 | Ref |
| 14669 | OL774867 | Unassigned | Unassigned | Unassigned | 137_0 | Clustered | 3 | VC_137_0 | 3 | 0.1129 | 0.998547 | 0.1128 | 0 | 0 | 0 | 1 | Ref |
| 14670 | OL774868 | Unassigned | Unassigned | Unassigned | 333_0 | Clustered | 25 | VC_333_0 | 25 | 0.4681 | 1 | 0.4681 | 0 | 0 | 0 | 1 | Ref |
| 14671 | OL774869 | Unassigned | Unassigned | Unassigned | 317_2 | Clustered | 20 | VC_317_2 | 20 | 0.1976 | 1 | 0.1976 | 0 | 0 | 0 | 0.6842 | Ref |
| 14672 | OL774870 | Unassigned | Unassigned | Unassigned | 293_3 | Clustered | 19 | VC_293_3 | 19 | 0.2627 | 1 | 0.2627 | 0 | 0 | 0 | 0.8187 | Ref |
| 14673 | OL774872 | Unassigned | Unassigned | Unassigned | 323_1 | Clustered | 9 | VC_323_1 | 9 | 0.1798 | 1 | 0.1798 | 0 | 0 | 0 | 0.75 | Ref |
| 14674 | OL774873 | Unassigned | Unassigned | Unassigned | 412_0 | Clustered | 39 | VC_412_0 | 39 | 0.845 | 1 | 0.845 | 0 | 0 | 0 | 1 | Ref |
| 14675 | OL774876 | Unassigned | Unassigned | Unassigned | 1187_0 | Clustered | 3 | VC_1187_0 | 3 | 0.924 | 0.92554266 | 0.8552 | 0 | 0 | 0 | 1 | Ref |
| 14676 | OL774877 | Unassigned | Unassigned | Unassigned | 317_2 | Clustered | 20 | VC_317_2 | 20 | 0.1976 | 1 | 0.1976 | 0 | 0 | 0 | 0.6842 | Ref |
| 14677 | OL780484 | Unassigned | Unassigned | Unassigned | 290_5 | Clustered | 14 | VC_290_5 | 14 | 0.1422 | 1 | 0.1422 | 0 | 0 | 0 | 0.9231 | Ref |
| 14678 | OL791266 | Unassigned | Unassigned | Unassigned | 242_1 | Clustered | 20 | VC_242_1 | 20 | 0.684 | 1 | 0.684 | 0 | 0 | 0 | 0.8368 | Ref |
| 14679 | OL799256 | Unassigned | Unassigned | Unassigned | 695_0 | Clustered | 48 | VC_695_0 | 48 | 0.8162 | 1 | 0.8162 | 0 | 0 | 0 | 1 | Ref |
| 14680 | OL799257 | Unassigned | Unassigned | Unassigned | 695_0 | Clustered | 48 | VC_695_0 | 48 | 0.8162 | 1 | 0.8162 | 0 | 0 | 0 | 1 | Ref |
| 14681 | OL799258 | Unassigned | Unassigned | Unassigned | 92_0 | Clustered | 22 | VC_92_0 | 22 | 0.5639 | 1 | 0.5639 | 0 | 0 | 0 | 1 | Ref |
| 14682 | OL799259 | Unassigned | Unassigned | Unassigned | 968_0 | Clustered | 4 | VC_968_0 | 4 | 0.2959 | 0.99997642 | 0.2959 | 0 | 0 | 0 | 1 | Ref |
| 14683 | OL799260 | Unassigned | Unassigned | Unassigned | 92_0 | Clustered | 22 | VC_92_0 | 22 | 0.5639 | 1 | 0.5639 | 0 | 0 | 0 | 1 | Ref |
| 14684 | OL799327 | Unassigned | Unassigned | Unassigned | 812_3 | Clustered/Singleton | 1 | VC_812_3 | 1 | 0 | 1 | 0 | 0 | 0 | 0 | 1 | Ref |
| 14685 | OL799328 | Unassigned | Unassigned | Unassigned | 823_0 | Clustered | 124 | VC_823_0 | 124 | 0.9492 | 1 | 0.9492 | 0 | 0 | 0 | 1 | Ref |
| 14686 | OL800603 | Unassigned | Unassigned | Unassigned | 386_0 | Clustered | 49 | VC_386_0 | 49 | 0.7941 | 1 | 0.7941 | 0 | 0 | 0 | 1 | Ref |
| 14687 | OL800605 | Unassigned | Unassigned | Unassigned | 385_0 | Clustered | 51 | VC_385_0 | 51 | 0.8754 | 1 | 0.8754 | 0 | 0 | 0 | 1 | Ref |
| 14688 | OL800606 | Unassigned | Unassigned | Unassigned | 329_11 | Clustered | 15 | VC_329_11 | 15 | 0.0791 | 1 | 0.0791 | 0 | 0 | 0 | 0.9333 | Ref |
| 14689 | OL800706 | Unassigned | Unassigned | Unassigned | 575_0 | Clustered | 35 | VC_575_0 | 35 | 0.8815 | 1 | 0.8815 | 0 | 0 | 0 | 1 | Ref |
| 14690 | OL802210 | Unassigned | Unassigned | Unassigned | 423_0 | Clustered | 45 | VC_423_0 | 45 | 0.9484 | 1 | 0.9484 | 0 | 0 | 0 | 1 | Ref |
| 14691 | OL802211 | Unassigned | Unassigned | Unassigned | 749_0 | Clustered | 40 | VC_749_0 | 40 | 0.7517 | 1 | 0.7517 | 0 | 0 | 0 | 1 | Ref |
| 14692 | OL825705 | Unassigned | Unassigned | Unassigned | 329_11 | Clustered | 15 | VC_329_11 | 15 | 0.0791 | 1 | 0.0791 | 0 | 0 | 0 | 0.9333 | Ref |
| 14693 | OL829977 | Unassigned | Unassigned | Unassigned | 919_0 | Clustered | 39 | VC_919_0 | 39 | 0.9069 | 1 | 0.9069 | 0 | 0 | 0 | 1 | Ref |
| 14694 | OL829978 | Unassigned | Unassigned | Unassigned | 1071_1 | Clustered/Singleton | 1 | VC_1071_1 | 1 | 0 | 1 | 0 | 0 | 0 | 0 | 1 | Ref |
| 14695 | OL840290 | Unassigned | Unassigned | Unassigned | 8_0 | Clustered | 58 | VC_8_0 | 58 | 0.8306 | 1 | 0.8306 | 0 | 0 | 0 | 0.9946 | Ref |
| 14696 | OL840900 | Unassigned | Unassigned | Unassigned | 600_2 | Clustered | 10 | VC_600_2 | 10 | 0.3041 | 1 | 0.3041 | 0 | 0 | 0 | 1 | Ref |
| 14697 | OL840901 | Unassigned | Unassigned | Unassigned | 597_0 | Clustered | 6 | VC_597_0 | 6 | 0.1801 | 1 | 0.1801 | 0 | 0 | 0 | 1 | Ref |
| 14698 | OL856099 | Unassigned | Unassigned | Unassigned | 618_0 | Clustered | 13 | VC_618_0 | 13 | 0.6252 | 1 | 0.6252 | 0 | 0 | 0 | 1 | Ref |
| 14699 | OL856139 | Unassigned | Unassigned | Unassigned | 386_0 | Clustered | 49 | VC_386_0 | 49 | 0.7941 | 1 | 0.7941 | 0 | 0 | 0 | 1 | Ref |
| 14700 | OL865411 | Unassigned | Unassigned | Unassigned | 937_0 | Clustered | 40 | VC_937_0 | 40 | 0.6809 | 1 | 0.6809 | 0 | 0 | 0 | 0.9359 | Ref |
| 14701 | OL870316 | Unassigned | Unassigned | Unassigned | 283_118 | Clustered | 3 | VC_283_118 | 3 | 0.0054 | 0.99110781 | 0.0053 | 0 | 0 | 0 | 1 | Ref |
| 14702 | OL870317 | Unassigned | Unassigned | Unassigned | 283_112 | Clustered/Singleton | 1 | VC_283_112 | 1 | 0 | 1 | 0 | 0 | 0 | 0 | 1 | Ref |
| 14703 | OL870318 | Unassigned | Unassigned | Unassigned | 491_0 | Clustered | 36 | VC_491_0 | 36 | 0.5336 | 1 | 0.5336 | 0 | 0 | 0 | 0.9714 | Ref |
| 14704 | OL870319 | Unassigned | Unassigned | Unassigned | 491_0 | Clustered | 36 | VC_491_0 | 36 | 0.5336 | 1 | 0.5336 | 0 | 0 | 0 | 0.9714 | Ref |
| 14705 | OL870611 | Unassigned | Unassigned | Unassigned | 92_0 | Clustered | 22 | VC_92_0 | 22 | 0.5639 | 1 | 0.5639 | 0 | 0 | 0 | 1 | Ref |
| 14706 | OL870612 | Unassigned | Unassigned | Unassigned | 108_0 | Clustered | 33 | VC_108_0 | 33 | 0.949 | 1 | 0.949 | 0 | 0 | 0 | 1 | Ref |
| 14707 | OL955261 | Unassigned | Unassigned | Unassigned | 978_0 | Clustered | 2 | VC_978_0 | 2 | 0.0595 | 0.95411371 | 0.0568 | 0 | 0 | 0 | 1 | Ref |
| 14708 | OL956808 | Unassigned | Unassigned | Unassigned | 283_65 | Clustered/Singleton | 1 | VC_283_65 | 1 | 0 | 1 | 0 | 0 | 0 | 0 | 1 | Ref |
| 14709 | OL959999 | Unassigned | Unassigned | Unassigned | 117_0 | Clustered | 9 | VC_117_0 | 9 | 0.9977 | 0.98988637 | 0.9876 | 0 | 0 | 0 | 1 | Ref |
| 14710 | OL960029 | Unassigned | Unassigned | Unassigned | 469_0 | Clustered | 2 | VC_469_0 | 2 | 0.1492 | 0.95658144 | 0.1427 | 0 | 0 | 0 | 1 | Ref |
| 14711 | OL960030 | Unassigned | Unassigned | Unassigned | 700_0 | Clustered | 10 | VC_700_0 | 10 | 0.8076 | 1 | 0.8076 | 0 | 0 | 0 | 1 | Ref |
| 14712 | OL960567 | Unassigned | Unassigned | Unassigned | 149_8 | Clustered | 66 | VC_149_8 | 66 | 0.4565 | 1 | 0.4565 | 0 | 0 | 0 | 0.8182 | Ref |
| 14713 | OL960568 | Unassigned | Unassigned | Unassigned | 149_8 | Clustered | 66 | VC_149_8 | 66 | 0.4565 | 1 | 0.4565 | 0 | 0 | 0 | 0.8182 | Ref |
| 14714 | OL960569 | Unassigned | Unassigned | Unassigned | 149_8 | Clustered | 66 | VC_149_8 | 66 | 0.4565 | 1 | 0.4565 | 0 | 0 | 0 | 0.8182 | Ref |
| 14715 | OL960570 | Unassigned | Unassigned | Unassigned | 149_8 | Clustered | 66 | VC_149_8 | 66 | 0.4565 | 1 | 0.4565 | 0 | 0 | 0 | 0.8182 | Ref |
| 14716 | OL960573 | Unassigned | Unassigned | Unassigned | 795_0 | Clustered | 19 | VC_795_0 | 19 | 0.6848 | 1 | 0.6848 | 0 | 0 | 0 | 1 | Ref |
| 14717 | OL960574 | Unassigned | Unassigned | Unassigned | 792_0 | Clustered | 35 | VC_792_0 | 35 | 0.9828 | 1 | 0.9828 | 0 | 0 | 0 | 1 | Ref |
| 14718 | OL960575 | Unassigned | Unassigned | Unassigned | 795_0 | Clustered | 19 | VC_795_0 | 19 | 0.6848 | 1 | 0.6848 | 0 | 0 | 0 | 1 | Ref |
| 14719 | OL960576 | Unassigned | Unassigned | Unassigned | 286_1 | Clustered | 21 | VC_286_1 | 21 | 0.1696 | 1 | 0.1696 | 0 | 0 | 0 | 0.9048 | Ref |
| 14720 | OL960577 | Unassigned | Unassigned | Unassigned | 286_1 | Clustered | 21 | VC_286_1 | 21 | 0.1696 | 1 | 0.1696 | 0 | 0 | 0 | 0.9048 | Ref |
| 14721 | OL960578 | Unassigned | Unassigned | Unassigned | 575_0 | Clustered | 35 | VC_575_0 | 35 | 0.8815 | 1 | 0.8815 | 0 | 0 | 0 | 1 | Ref |
| 14722 | OL960579 | Unassigned | Unassigned | Unassigned | 575_0 | Clustered | 35 | VC_575_0 | 35 | 0.8815 | 1 | 0.8815 | 0 | 0 | 0 | 1 | Ref |
| 14723 | OL960580 | Unassigned | Unassigned | Unassigned | 283_152 | Clustered/Singleton | 1 | VC_283_152 | 1 | 0 | 1 | 0 | 0 | 0 | 0 | 1 | Ref |
| 14724 | OL960581 | Unassigned | Unassigned | Unassigned | 283_157 | Clustered/Singleton | 1 | VC_283_157 | 1 | 0 | 1 | 0 | 0 | 0 | 0 | 1 | Ref |
| 14725 | OL960582 | Unassigned | Unassigned | Unassigned | 575_0 | Clustered | 35 | VC_575_0 | 35 | 0.8815 | 1 | 0.8815 | 0 | 0 | 0 | 1 | Ref |
| 14726 | OL963731 | Unassigned | Unassigned | Unassigned | 978_1 | Clustered/Singleton | 1 | VC_978_1 | 1 | 0 | 1 | 0 | 0 | 0 | 0 | 1 | Ref |
| 14727 | OL964058 | Unassigned | Unassigned | Unassigned | 187_3 | Clustered | 9 | VC_187_3 | 9 | 0.1971 | 1 | 0.1971 | 0 | 0 | 0 | 0.8611 | Ref |
| 14728 | OL964734 | Unassigned | Unassigned | Unassigned | 461_0 | Clustered | 96 | VC_461_0 | 96 | 0.9307 | 1 | 0.9307 | 0 | 0 | 0 | 0.9575 | Ref |
| 14729 | OL964735 | Unassigned | Unassigned | Unassigned | 283_91 | Clustered | 11 | VC_283_91 | 11 | 0.0285 | 1 | 0.0285 | 0 | 0 | 0 | 1 | Ref |
| 14730 | OL964736 | Unassigned | Unassigned | Unassigned | 133_10 | Clustered | 10 | VC_133_10 | 10 | 0.0527 | 1 | 0.0527 | 0 | 0 | 0 | 1 | Ref |
| 14731 | OL964737 | Unassigned | Unassigned | Unassigned | 133_10 | Clustered | 10 | VC_133_10 | 10 | 0.0527 | 1 | 0.0527 | 0 | 0 | 0 | 1 | Ref |
| 14732 | OL964738 | Unassigned | Unassigned | Unassigned | 133_10 | Clustered | 10 | VC_133_10 | 10 | 0.0527 | 1 | 0.0527 | 0 | 0 | 0 | 1 | Ref |
| 14733 | OL964739 | Unassigned | Unassigned | Unassigned | 392_0 | Clustered | 126 | VC_392_0 | 126 | 0.9621 | 1 | 0.9621 | 0 | 0 | 0 | 1 | Ref |
| 14734 | OL964740 | Unassigned | Unassigned | Unassigned | 392_0 | Clustered | 126 | VC_392_0 | 126 | 0.9621 | 1 | 0.9621 | 0 | 0 | 0 | 1 | Ref |
| 14735 | OL964741 | Unassigned | Unassigned | Unassigned | 392_0 | Clustered | 126 | VC_392_0 | 126 | 0.9621 | 1 | 0.9621 | 0 | 0 | 0 | 1 | Ref |
| 14736 | OL964742 | Unassigned | Unassigned | Unassigned | 461_0 | Clustered | 96 | VC_461_0 | 96 | 0.9307 | 1 | 0.9307 | 0 | 0 | 0 | 0.9575 | Ref |
| 14737 | OL964743 | Unassigned | Unassigned | Unassigned | 461_0 | Clustered | 96 | VC_461_0 | 96 | 0.9307 | 1 | 0.9307 | 0 | 0 | 0 | 0.9575 | Ref |
| 14738 | OL964744 | Unassigned | Unassigned | Unassigned | 461_0 | Clustered | 96 | VC_461_0 | 96 | 0.9307 | 1 | 0.9307 | 0 | 0 | 0 | 0.9575 | Ref |
| 14739 | OL964745 | Unassigned | Unassigned | Unassigned | 283_91 | Clustered | 11 | VC_283_91 | 11 | 0.0285 | 1 | 0.0285 | 0 | 0 | 0 | 1 | Ref |
| 14740 | OL964746 | Unassigned | Unassigned | Unassigned | 133_10 | Clustered | 10 | VC_133_10 | 10 | 0.0527 | 1 | 0.0527 | 0 | 0 | 0 | 1 | Ref |

| | | | | | | | | | | | | | | | | | |
|---|---|---|---|---|---|---|---|---|---|---|---|---|---|---|---|---|---|
| 14741 | OL964747 | Unassigned | Unassigned | Unassigned | 133_10 | Clustered | 10 | VC_133_10 | 10 | 0.0527 | 1 | 0.0527 | 0 | 0 | 0 | 1 | Ref |
| 14742 | OL964748 | Unassigned | Unassigned | Unassigned | 392_0 | Clustered | 126 | VC_392_0 | 126 | 0.9621 | 1 | 0.9621 | 0 | 0 | 0 | 1 | Ref |
| 14743 | OL964749 | Unassigned | Unassigned | Unassigned | 392_0 | Clustered | 126 | VC_392_0 | 126 | 0.9621 | 1 | 0.9621 | 0 | 0 | 0 | 1 | Ref |
| 14744 | OL964750 | Unassigned | Unassigned | Unassigned | 392_0 | Clustered | 126 | VC_392_0 | 126 | 0.9621 | 1 | 0.9621 | 0 | 0 | 0 | 1 | Ref |
| 14745 | OL964751 | Unassigned | Unassigned | Unassigned | 461_0 | Clustered | 96 | VC_461_0 | 96 | 0.9307 | 1 | 0.9307 | 0 | 0 | 0 | 0.9575 | Ref |
| 14746 | OL964752 | Unassigned | Unassigned | Unassigned | 461_0 | Clustered | 96 | VC_461_0 | 96 | 0.9307 | 1 | 0.9307 | 0 | 0 | 0 | 0.9575 | Ref |
| 14747 | OL964753 | Unassigned | Unassigned | Unassigned | 133_10 | Clustered | 10 | VC_133_10 | 10 | 0.0527 | 1 | 0.0527 | 0 | 0 | 0 | 1 | Ref |
| 14748 | OL964754 | Unassigned | Unassigned | Unassigned | 597_0 | Clustered | 6 | VC_597_0 | 6 | 0.1801 | 1 | 0.1801 | 0 | 0 | 0 | 1 | Ref |
| 14749 | OL964755 | Unassigned | Unassigned | Unassigned | 230_0 | Clustered | 3 | VC_230_0 | 3 | 0.0739 | 0.9934951 | 0.0734 | 0 | 0 | 0 | 0.6667 | Ref |
| 14750 | OL964756 | Unassigned | Unassigned | Unassigned | 442_3 | Clustered | 3 | VC_442_3 | 3 | 0.0529 | 0.99675181 | 0.0527 | 0 | 0 | 0 | 1 | Ref |
| 14751 | OL964948 | Unassigned | Unassigned | Unassigned | 699_0 | Clustered | 71 | VC_699_0 | 71 | 0.9352 | 1 | 0.9352 | 0 | 0 | 0 | 1 | Ref |
| 14752 | OL976437 | Unassigned | Unassigned | Unassigned | 111_0 | Clustered | 91 | VC_111_0 | 91 | 0.9587 | 1 | 0.9587 | 0 | 0 | 0 | 1 | Ref |
| 14753 | OL979478 | Unassigned | Unassigned | Unassigned | 111_0 | Clustered | 91 | VC_111_0 | 91 | 0.9587 | 1 | 0.9587 | 0 | 0 | 0 | 1 | Ref |
| 14754 | OL979479 | Unassigned | Unassigned | Unassigned | 111_0 | Clustered | 91 | VC_111_0 | 91 | 0.9587 | 1 | 0.9587 | 0 | 0 | 0 | 1 | Ref |
| 14755 | OL989991 | Unassigned | Unassigned | Unassigned | 490_1 | Clustered | 5 | VC_490_1 | 5 | 0.0447 | 0.99999996 | 0.0447 | 0 | 0 | 0 | 1 | Ref |
| 14756 | OM022251 | Unassigned | Unassigned | Unassigned | 30_1 | Clustered | 16 | VC_30_1 | 16 | 0.1973 | 1 | 0.1973 | 0 | 0 | 0 | 0.8333 | Ref |
| 14757 | OM025231 | Unassigned | Unassigned | Unassigned | 592_0 | Clustered | 50 | VC_592_0 | 50 | 0.7314 | 1 | 0.7314 | 0 | 0 | 0 | 1 | Ref |
| 14758 | OM030343 | Unassigned | Unassigned | Unassigned | 149_8 | Clustered | 66 | VC_149_8 | 66 | 0.4565 | 1 | 0.4565 | 0 | 0 | 0 | 0.8182 | Ref |
| 14759 | OM032871 | Unassigned | Unassigned | Unassigned | 938_0 | Clustered | 99 | VC_938_0 | 99 | 0.9119 | 1 | 0.9119 | 0 | 0 | 0 | 0.9928 | Ref |
| 14760 | OM033133 | Unassigned | Unassigned | Unassigned | 639_0 | Clustered | 2 | VC_639_0 | 2 | 0.2791 | 0.95526917 | 0.2666 | 0 | 0 | 0 | 1 | Ref |
| 14761 | OM033134 | Unassigned | Unassigned | Unassigned | 865_3 | Clustered | 2 | VC_865_3 | 2 | 0.0748 | 0.93469061 | 0.0699 | 0 | 0 | 0 | 1 | Ref |
| 14762 | OM033135 | Unassigned | Unassigned | Unassigned | 865_3 | Clustered | 2 | VC_865_3 | 2 | 0.0748 | 0.93469061 | 0.0699 | 0 | 0 | 0 | 1 | Ref |
| 14763 | OM033136 | Unassigned | Unassigned | Unassigned | 865_4 | Clustered/Singleton | 1 | VC_865_4 | 1 | 0 | 1 | 0 | 0 | 0 | 0 | 1 | Ref |
| 14764 | OM046621 | Unassigned | Unassigned | Unassigned | 37_0 | Clustered | 5 | VC_37_0 | 5 | 0.5393 | 0.99999987 | 0.5393 | 0 | 0 | 0 | 1 | Ref |
| 14765 | OM046622 | Unassigned | Unassigned | Unassigned | 37_0 | Clustered | 5 | VC_37_0 | 5 | 0.5393 | 0.99999987 | 0.5393 | 0 | 0 | 0 | 1 | Ref |
| 14766 | OM046623 | Unassigned | Unassigned | Unassigned | 37_0 | Clustered | 5 | VC_37_0 | 5 | 0.5393 | 0.99999987 | 0.5393 | 0 | 0 | 0 | 1 | Ref |
| 14767 | OM046625 | Unassigned | Unassigned | Unassigned | 37_0 | Clustered | 5 | VC_37_0 | 5 | 0.5393 | 0.99999987 | 0.5393 | 0 | 0 | 0 | 1 | Ref |
| 14768 | OM046626 | Unassigned | Unassigned | Unassigned | 228_0 | Clustered | 60 | VC_228_0 | 60 | 0.942 | 1 | 0.942 | 0 | 0 | 0 | 1 | Ref |
| 14769 | OM046627 | Unassigned | Unassigned | Unassigned | 228_0 | Clustered | 60 | VC_228_0 | 60 | 0.942 | 1 | 0.942 | 0 | 0 | 0 | 1 | Ref |
| 14770 | OM046628 | Unassigned | Unassigned | Unassigned | 228_0 | Clustered | 60 | VC_228_0 | 60 | 0.942 | 1 | 0.942 | 0 | 0 | 0 | 1 | Ref |
| 14771 | OM046629 | Unassigned | Unassigned | Unassigned | 37_0 | Clustered | 5 | VC_37_0 | 5 | 0.5393 | 0.99999987 | 0.5393 | 0 | 0 | 0 | 1 | Ref |
| 14772 | OM055665 | Unassigned | Unassigned | Unassigned | 363_0 | Clustered | 2 | VC_363_0 | 2 | 0.8022 | 0.72985431 | 0.5855 | 0 | 0 | 0 | 1 | Ref |
| 14773 | OM065837 | Unassigned | Unassigned | Unassigned | 937_0 | Clustered | 40 | VC_937_0 | 40 | 0.6809 | 1 | 0.6809 | 0 | 0 | 0 | 0.9359 | Ref |
| 14774 | OM065838 | Unassigned | Unassigned | Unassigned | 937_0 | Clustered | 40 | VC_937_0 | 40 | 0.6809 | 1 | 0.6809 | 0 | 0 | 0 | 0.9359 | Ref |
| 14775 | OM065839 | Unassigned | Unassigned | Unassigned | 937_0 | Clustered | 40 | VC_937_0 | 40 | 0.6809 | 1 | 0.6809 | 0 | 0 | 0 | 0.9359 | Ref |
| 14776 | OM065840 | Unassigned | Unassigned | Unassigned | 938_0 | Clustered | 99 | VC_938_0 | 99 | 0.9119 | 1 | 0.9119 | 0 | 0 | 0 | 0.9928 | Ref |
| 14777 | OM065841 | Unassigned | Unassigned | Unassigned | 937_0 | Clustered | 40 | VC_937_0 | 40 | 0.6809 | 1 | 0.6809 | 0 | 0 | 0 | 0.9359 | Ref |
| 14778 | OM065842 | Unassigned | Unassigned | Unassigned | 938_0 | Clustered | 99 | VC_938_0 | 99 | 0.9119 | 1 | 0.9119 | 0 | 0 | 0 | 0.9928 | Ref |
| 14779 | OM065843 | Unassigned | Unassigned | Unassigned | 938_0 | Clustered | 99 | VC_938_0 | 99 | 0.9119 | 1 | 0.9119 | 0 | 0 | 0 | 0.9928 | Ref |
| 14780 | OM095401 | Unassigned | Unassigned | Unassigned | 5_0 | Clustered | 30 | VC_5_0 | 30 | 0.9647 | 1 | 0.9647 | 0 | 0 | 0 | 1 | Ref |
| 14781 | OM103621 | Unassigned | Unassigned | Unassigned | 668_0 | Clustered | 25 | VC_668_0 | 25 | 0.5539 | 1 | 0.5539 | 0 | 0 | 0 | 0.96 | Ref |
| 14782 | OM105884 | Unassigned | Unassigned | Unassigned | 325_62 | Clustered | 3 | VC_325_62 | 3 | 0.0188 | 0.99859194 | 0.0188 | 0 | 0 | 0 | 1 | Ref |
| 14783 | OM105885 | Unassigned | Unassigned | Unassigned | 520_54 | Clustered/Singleton | 1 | VC_520_54 | 1 | 0 | 1 | 0 | 0 | 0 | 0 | 1 | Ref |
| 14784 | OM105886 | Unassigned | Unassigned | Unassigned | 510_0 | Clustered | 12 | VC_510_0 | 12 | 0.3963 | 1 | 0.3963 | 0 | 0 | 0 | 0.9697 | Ref |
| 14785 | OM131411 | Unassigned | Unassigned | Unassigned | 130_0 | Clustered | 19 | VC_130_0 | 19 | 0.6068 | 1 | 0.6068 | 0 | 0 | 0 | 1 | Ref |
| 14786 | OM135583 | Unassigned | Unassigned | Unassigned | 283_167 | Clustered/Singleton | 1 | VC_283_167 | 1 | 0 | 1 | 0 | 0 | 0 | 0 | 1 | Ref |
| 14787 | OM135608 | Unassigned | Unassigned | Unassigned | 1346_0 | Clustered | 10 | VC_1346_0 | 10 | 0.7784 | 1 | 0.7784 | 0 | 0 | 0 | 1 | Ref |
| 14788 | OM135992 | Unassigned | Unassigned | Unassigned | 94_0 | Clustered | 2 | VC_94_0 | 2 | 0.1304 | 0.95723134 | 0.1248 | 0 | 0 | 0 | 1 | Ref |
| 14789 | OM140653 | Unassigned | Unassigned | Unassigned | 228_0 | Clustered | 60 | VC_228_0 | 60 | 0.942 | 1 | 0.942 | 0 | 0 | 0 | 1 | Ref |
| 14790 | OM140839 | Unassigned | Unassigned | Unassigned | 725_0 | Clustered | 4 | VC_725_0 | 4 | 0.4268 | 0.99997355 | 0.4268 | 0 | 0 | 0 | 1 | Ref |
| 14791 | OM141125 | Unassigned | Unassigned | Unassigned | 176_2 | Clustered/Singleton | 1 | VC_176_2 | 1 | 0 | 1 | 0 | 0 | 0 | 0 | 1 | Ref |
| 14792 | OM194188 | Unassigned | Unassigned | Unassigned | 823_0 | Clustered | 124 | VC_823_0 | 124 | 0.9492 | 1 | 0.9492 | 0 | 0 | 0 | 1 | Ref |
| 14793 | OM194189 | Unassigned | Unassigned | Unassigned | 823_0 | Clustered | 124 | VC_823_0 | 124 | 0.9492 | 1 | 0.9492 | 0 | 0 | 0 | 1 | Ref |
| 14794 | OM194190 | Unassigned | Unassigned | Unassigned | 823_0 | Clustered | 124 | VC_823_0 | 124 | 0.9492 | 1 | 0.9492 | 0 | 0 | 0 | 1 | Ref |
| 14795 | OM203156 | Unassigned | Unassigned | Unassigned | 869_0 | Clustered | 9 | VC_869_0 | 9 | 0.2624 | 1 | 0.2624 | 0 | 0 | 0 | 1 | Ref |
| 14796 | OM203157 | Unassigned | Unassigned | Unassigned | 1171_0 | Clustered | 21 | VC_1171_0 | 21 | 0.7125 | 1 | 0.7125 | 0 | 0 | 0 | 1 | Ref |
| 14797 | OM203158 | Unassigned | Unassigned | Unassigned | 222_0 | Clustered | 5 | VC_222_0 | 5 | 0.0565 | 0.99999997 | 0.0565 | 0 | 0 | 0 | 1 | Ref |
| 14798 | OM203159 | Unassigned | Unassigned | Unassigned | 216_4 | Clustered | 13 | VC_216_4 | 13 | 0.1657 | 1 | 0.1657 | 0 | 0 | 0 | 1 | Ref |
| 14799 | OM203160 | Unassigned | Unassigned | Unassigned | 1175_0 | Clustered | 22 | VC_1175_0 | 22 | 0.4562 | 1 | 0.4562 | 0 | 0 | 0 | 1 | Ref |
| 14800 | OM203161 | Unassigned | Unassigned | Unassigned | 220_0 | Clustered | 4 | VC_220_0 | 4 | 0.0615 | 0.99998802 | 0.0615 | 0 | 0 | 0 | 1 | Ref |
| 14801 | OM203162 | Unassigned | Unassigned | Unassigned | 325_33 | Clustered | 4 | VC_325_33 | 4 | 0.0149 | 0.99998565 | 0.0149 | 0 | 0 | 0 | 1 | Ref |
| 14802 | OM210019 | Unassigned | Unassigned | Unassigned | 637_0 | Clustered | 115 | VC_637_0 | 115 | 0.731 | 1 | 0.731 | 0 | 0 | 0 | 0.8764 | Ref |
| 14803 | OM223115 | Unassigned | Unassigned | Unassigned | 725_0 | Clustered | 4 | VC_725_0 | 4 | 0.4268 | 0.99997355 | 0.4268 | 0 | 0 | 0 | 1 | Ref |
| 14804 | OM234791 | Unassigned | Unassigned | Unassigned | 749_0 | Clustered | 40 | VC_749_0 | 40 | 0.7517 | 1 | 0.7517 | 0 | 0 | 0 | 1 | Ref |
| 14805 | OM234792 | Unassigned | Unassigned | Unassigned | 838_0 | Clustered | 29 | VC_838_0 | 29 | 0.8611 | 1 | 0.8611 | 0 | 0 | 0 | 1 | Ref |
| 14806 | OM234793 | Unassigned | Unassigned | Unassigned | 112_0 | Clustered | 8 | VC_112_0 | 8 | 0.2537 | 1 | 0.2537 | 0 | 0 | 0 | 1 | Ref |
| 14807 | OM236513 | Unassigned | Unassigned | Unassigned | 87_0 | Clustered | 8 | VC_87_0 | 8 | 0.5545 | 1 | 0.5545 | 0 | 0 | 0 | 1 | Ref |
| 14808 | OM236514 | Unassigned | Unassigned | Unassigned | 18_0 | Clustered | 4 | VC_18_0 | 4 | 0.9098 | 0.99982347 | 0.9096 | 0 | 0 | 0 | 1 | Ref |
| 14809 | OM236515 | Unassigned | Unassigned | Unassigned | 89_6 | Clustered/Singleton | 1 | VC_89_6 | 1 | 0 | 1 | 0 | 0 | 0 | 0 | 1 | Ref |
| 14810 | OM236516 | Unassigned | Unassigned | Unassigned | 475_0 | Clustered | 3 | VC_475_0 | 3 | 0.0778 | 0.99798375 | 0.0776 | 0 | 0 | 0 | 1 | Ref |
| 14811 | OM249648 | Unassigned | Unassigned | Unassigned | 845_0 | Clustered | 3 | VC_845_0 | 3 | 0.3348 | 0.99831497 | 0.3343 | 0 | 0 | 0 | 1 | Ref |
| 14812 | OM249958 | Unassigned | Unassigned | Unassigned | 817_0 | Clustered | 14 | VC_817_0 | 14 | 0.6342 | 1 | 0.6342 | 0 | 0 | 0 | 1 | Ref |
| 14813 | OM256450 | Unassigned | Unassigned | Unassigned | 129_0 | Clustered | 43 | VC_129_0 | 43 | 0.886 | 1 | 0.886 | 0 | 0 | 0 | 1 | Ref |
| 14814 | OM256482 | Unassigned | Unassigned | Unassigned | 451_0 | Clustered | 80 | VC_451_0 | 80 | 0.9991 | 1 | 0.9991 | 0 | 0 | 0 | 1 | Ref |
| 14815 | OM258170 | Unassigned | Unassigned | Unassigned | 456_0 | Clustered | 62 | VC_456_0 | 62 | 0.9026 | 1 | 0.9026 | 0 | 0 | 0 | 1 | Ref |
| 14816 | OM262540 | Unassigned | Unassigned | Unassigned | 835_0 | Clustered | 67 | VC_835_0 | 67 | 0.9624 | 1 | 0.9624 | 0 | 0 | 0 | 1 | Ref |
| 14817 | OM262541 | Unassigned | Unassigned | Unassigned | 835_0 | Clustered | 67 | VC_835_0 | 67 | 0.9624 | 1 | 0.9624 | 0 | 0 | 0 | 1 | Ref |
| 14818 | OM272052 | Unassigned | Unassigned | Unassigned | 638_0 | Clustered | 45 | VC_638_0 | 45 | 0.7741 | 1 | 0.7741 | 0 | 0 | 0 | 1 | Ref |
| 14819 | OM282085 | Unassigned | Unassigned | Unassigned | 129_0 | Clustered | 43 | VC_129_0 | 43 | 0.886 | 1 | 0.886 | 0 | 0 | 0 | 1 | Ref |
| 14820 | OM283127 | Unassigned | Unassigned | Unassigned | 162_0 | Clustered | 18 | VC_162_0 | 18 | 0.7403 | 1 | 0.7403 | 0 | 0 | 0 | 0.8627 | Ref |
| 14821 | OM284015 | Unassigned | Unassigned | Unassigned | 242_1 | Clustered | 20 | VC_242_1 | 20 | 0.684 | 1 | 0.684 | 0 | 0 | 0 | 0.8368 | Ref |
| 14822 | OM287554 | Unassigned | Unassigned | Unassigned | 500_0 | Clustered | 3 | VC_500_0 | 3 | 0.8278 | 0.9969176 | 0.8253 | 0 | 0 | 0 | 1 | Ref |
| 14823 | OM291373 | Unassigned | Unassigned | Unassigned | 637_0 | Clustered | 115 | VC_637_0 | 115 | 0.731 | 1 | 0.731 | 0 | 0 | 0 | 0.8764 | Ref |
| 14824 | OM293948 | Unassigned | Unassigned | Unassigned | 95_0 | Clustered | 3 | VC_95_0 | 3 | 0.2092 | 0.99852577 | 0.2089 | 0 | 0 | 0 | 1 | Ref |
| 14825 | OM304382 | Unassigned | Unassigned | Unassigned | 3_0 | Clustered | 17 | VC_3_0 | 17 | 0.5639 | 1 | 0.5639 | 0 | 0 | 0 | 1 | Ref |
| 14826 | OM304856 | Unassigned | Unassigned | Unassigned | 8_0 | Clustered | 58 | VC_8_0 | 58 | 0.8306 | 1 | 0.8306 | 0 | 0 | 0 | 0.9946 | Ref |
| 14827 | OM313460 | Unassigned | Unassigned | Unassigned | 428_0 | Clustered | 33 | VC_428_0 | 33 | 0.9915 | 1 | 0.9915 | 0 | 0 | 0 | 1 | Ref |
| 14828 | OM313461 | Unassigned | Unassigned | Unassigned | 428_0 | Clustered | 33 | VC_428_0 | 33 | 0.9915 | 1 | 0.9915 | 0 | 0 | 0 | 1 | Ref |
| 14829 | OM317910 | Unassigned | Unassigned | Unassigned | 585_3 | Clustered/Singleton | 1 | VC_585_3 | 1 | 0 | 1 | 0 | 0 | 0 | 0 | 1 | Ref |
| 14830 | OM319461 | Unassigned | Unassigned | Unassigned | 346_1 | Clustered | 2 | VC_346_1 | 2 | 0.1238 | 0.95353448 | 0.1181 | 0 | 0 | 0 | 1 | Ref |
| 14831 | OM328157 | Unassigned | Unassigned | Unassigned | 937_0 | Clustered | 40 | VC_937_0 | 40 | 0.6809 | 1 | 0.6809 | 0 | 0 | 0 | 0.9359 | Ref |
| 14832 | OM337876 | Unassigned | Unassigned | Unassigned | 491_0 | Clustered | 36 | VC_491_0 | 36 | 0.5336 | 1 | 0.5336 | 0 | 0 | 0 | 0.9714 | Ref |
| 14833 | OM337877 | Unassigned | Unassigned | Unassigned | 491_0 | Clustered | 36 | VC_491_0 | 36 | 0.5336 | 1 | 0.5336 | 0 | 0 | 0 | 0.9714 | Ref |
| 14834 | OM339528 | Unassigned | Unassigned | Unassigned | 283_92 | Clustered | 4 | VC_283_92 | 4 | 0.0079 | 0.9999582 | 0.0079 | 0 | 0 | 0 | 1 | Ref |
| 14835 | OM339548 | Unassigned | Unassigned | Unassigned | 637_0 | Clustered | 115 | VC_637_0 | 115 | 0.731 | 1 | 0.731 | 0 | 0 | 0 | 0.8764 | Ref |
| 14836 | OM339718 | Unassigned | Unassigned | Unassigned | 443_2 | Clustered | 10 | VC_443_2 | 10 | 0.3344 | 1 | 0.3344 | 0 | 0 | 0 | 0.9333 | Ref |
| 14837 | OM350011 | Unassigned | Unassigned | Unassigned | 883_1 | Clustered | 12 | VC_883_1 | 12 | 0.2441 | 1 | 0.2441 | 0 | 0 | 0 | 1 | Ref |
| 14838 | OM350012 | Unassigned | Unassigned | Unassigned | 884_0 | Clustered | 28 | VC_884_0 | 28 | 0.6211 | 1 | 0.6211 | 0 | 0 | 0 | 0.9841 | Ref |
| 14839 | OM350013 | Unassigned | Unassigned | Unassigned | 884_0 | Clustered | 28 | VC_884_0 | 28 | 0.6211 | 1 | 0.6211 | 0 | 0 | 0 | 0.9841 | Ref |
| 14840 | OM355495 | Unassigned | Unassigned | Unassigned | 451_0 | Clustered | 80 | VC_451_0 | 80 | 0.9991 | 1 | 0.9991 | 0 | 0 | 0 | 1 | Ref |
| 14841 | OM362522 | Unassigned | Unassigned | Unassigned | 837_0 | Clustered | 29 | VC_837_0 | 29 | 0.9955 | 1 | 0.9955 | 0 | 0 | 0 | 1 | Ref |
| 14842 | OM362897 | Unassigned | Unassigned | Unassigned | 847_1 | Clustered | 26 | VC_847_1 | 26 | 0.6323 | 1 | 0.6323 | 0 | 0 | 0 | 0.9815 | Ref |
| 14843 | OM367911 | Unassigned | Unassigned | Unassigned | 133_20 | Clustered/Singleton | 1 | VC_133_20 | 1 | 0 | 1 | 0 | 0 | 0 | 0 | 1 | Ref |
| 14844 | OM373202 | Unassigned | Unassigned | Unassigned | 682_0 | Clustered | 2 | VC_682_0 | 2 | 1 | 1 | 1 | 0 | 0 | 0 | 1 | Ref |
| 14845 | OM373550 | Unassigned | Unassigned | Unassigned | 15_1 | Clustered | 9 | VC_15_1 | 9 | 0.182 | 1 | 0.182 | 0 | 0 | 0 | 0.8889 | Ref |
| 14846 | OM373551 | Unassigned | Unassigned | Unassigned | 11_0 | Clustered | 19 | VC_11_0 | 19 | 0.7562 | 1 | 0.7562 | 0 | 0 | 0 | 1 | Ref |
| 14847 | OM373552 | Unassigned | Unassigned | Unassigned | 11_0 | Clustered | 19 | VC_11_0 | 19 | 0.7562 | 1 | 0.7562 | 0 | 0 | 0 | 1 | Ref |
| 14848 | OM373553 | Unassigned | Unassigned | Unassigned | 11_0 | Clustered | 19 | VC_11_0 | 19 | 0.7562 | 1 | 0.7562 | 0 | 0 | 0 | 1 | Ref |
| 14849 | OM373554 | Unassigned | Unassigned | Unassigned | 15_1 | Clustered | 9 | VC_15_1 | 9 | 0.182 | 1 | 0.182 | 0 | 0 | 0 | 0.8889 | Ref |
| 14850 | OM373555 | Unassigned | Unassigned | Unassigned | 15_1 | Clustered | 9 | VC_15_1 | 9 | 0.182 | 1 | 0.182 | 0 | 0 | 0 | 0.8889 | Ref |
| 14851 | OM373556 | Unassigned | Unassigned | Unassigned | 15_1 | Clustered | 9 | VC_15_1 | 9 | 0.182 | 1 | 0.182 | 0 | 0 | 0 | 0.8889 | Ref |
| 14852 | OM373557 | Unassigned | Unassigned | Unassigned | 149_8 | Clustered | 66 | VC_149_8 | 66 | 0.4565 | 1 | 0.4565 | 0 | 0 | 0 | 0.8182 | Ref |
| 14853 | OM373558 | Unassigned | Unassigned | Unassigned | 15_0 | Clustered | 3 | VC_15_0 | 3 | 0.0481 | 0.9985346 | 0.048 | 0 | 0 | 0 | 1 | Ref |
| 14854 | OM373561 | Unassigned | Unassigned | Unassigned | 15_1 | Clustered | 9 | VC_15_1 | 9 | 0.182 | 1 | 0.182 | 0 | 0 | 0 | 0.8889 | Ref |
| 14855 | OM386652 | Unassigned | Unassigned | Unassigned | 722_0 | Clustered | 13 | VC_722_0 | 13 | 0.5104 | 1 | 0.5104 | 0 | 0 | 0 | 1 | Ref |
| 14856 | OM386653 | Unassigned | Unassigned | Unassigned | 848_0 | Clustered | 28 | VC_848_0 | 28 | 0.7927 | 1 | 0.7927 | 0 | 0 | 0 | 1 | Ref |
| 14857 | OM386654 | Unassigned | Unassigned | Unassigned | 848_0 | Clustered | 28 | VC_848_0 | 28 | 0.7927 | 1 | 0.7927 | 0 | 0 | 0 | 1 | Ref |
| 14858 | OM386655 | Unassigned | Unassigned | Unassigned | 290_4 | Clustered | 2 | VC_290_4 | 2 | 0.0092 | 0.90346068 | 0.0083 | 0 | 0 | 0 | 1 | Ref |
| 14859 | OM386656 | Unassigned | Unassigned | Unassigned | 847_1 | Clustered | 26 | VC_847_1 | 26 | 0.6323 | 1 | 0.6323 | 0 | 0 | 0 | 0.9815 | Ref |
| 14860 | OM386657 | Unassigned | Unassigned | Unassigned | 190_5 | Clustered/Singleton | 1 | VC_190_5 | 1 | 0 | 1 | 0 | 0 | 0 | 0 | 1 | Ref |
| 14861 | OM386658 | Unassigned | Unassigned | Unassigned | 848_0 | Clustered | 28 | VC_848_0 | 28 | 0.7927 | 1 | 0.7927 | 0 | 0 | 0 | 1 | Ref |
| 14862 | OM386659 | Unassigned | Unassigned | Unassigned | 847_1 | Clustered | 26 | VC_847_1 | 26 | 0.6323 | 1 | 0.6323 | 0 | 0 | 0 | 0.9815 | Ref |
| 14863 | OM386660 | Unassigned | Unassigned | Unassigned | 190_8 | Clustered/Singleton | 1 | VC_190_8 | 1 | 0 | 1 | 0 | 0 | 0 | 0 | 1 | Ref |
| 14864 | OM386661 | Unassigned | Unassigned | Unassigned | 329_22 | Clustered | 5 | VC_329_22 | 5 | 0.022 | 0.99999994 | 0.022 | 0 | 0 | 0 | 1 | Ref |
| 14865 | OM386662 | Unassigned | Unassigned | Unassigned | 286_9 | Clustered/Singleton | 1 | VC_286_9 | 1 | 0 | 1 | 0 | 0 | 0 | 0 | 1 | Ref |
| 14866 | OM386663 | Unassigned | Unassigned | Unassigned | 329_22 | Clustered | 5 | VC_329_22 | 5 | 0.022 | 0.99999994 | 0.022 | 0 | 0 | 0 | 1 | Ref |
| 14867 | OM386664 | Unassigned | Unassigned | Unassigned | 329_22 | Clustered | 5 | VC_329_22 | 5 | 0.022 | 0.99999994 | 0.022 | 0 | 0 | 0 | 1 | Ref |
| 14868 | OM386665 | Unassigned | Unassigned | Unassigned | 329_22 | Clustered | 5 | VC_329_22 | 5 | 0.022 | 0.99999994 | 0.022 | 0 | 0 | 0 | 1 | Ref |
| 14869 | OM386666 | Unassigned | Unassigned | Unassigned | 283_126 | Clustered/Singleton | 1 | VC_283_126 | 1 | 0 | 1 | 0 | 0 | 0 | 0 | 1 | Ref |
| 14870 | OM386667 | Unassigned | Unassigned | Unassigned | 847_1 | Clustered | 26 | VC_847_1 | 26 | 0.6323 | 1 | 0.6323 | 0 | 0 | 0 | 0.9815 | Ref |
| 14871 | OM401712 | Unassigned | Unassigned | Unassigned | 348_0 | Clustered | 22 | VC_348_0 | 22 | 0.9602 | 1 | 0.9602 | 0 | 0 | 0 | 1 | Ref |
| 14872 | OM401713 | Unassigned | Unassigned | Unassigned | 348_0 | Clustered | 22 | VC_348_0 | 22 | 0.9602 | 1 | 0.9602 | 0 | 0 | 0 | 1 | Ref |
| 14873 | OM401714 | Unassigned | Unassigned | Unassigned | 348_0 | Clustered | 22 | VC_348_0 | 22 | 0.9602 | 1 | 0.9602 | 0 | 0 | 0 | 1 | Ref |
| 14874 | OM401812 | Unassigned | Unassigned | Unassigned | 848_0 | Clustered | 28 | VC_848_0 | 28 | 0.7927 | 1 | 0.7927 | 0 | 0 | 0 | 1 | Ref |
| 14875 | OM416763 | Unassigned | Unassigned | Unassigned | 479_0 | Clustered | 17 | VC_479_0 | 17 | 0.9103 | 1 | 0.9103 | 0 | 0 | 0 | 1 | Ref |
| 14876 | OM416764 | Unassigned | Unassigned | Unassigned | 479_0 | Clustered | 17 | VC_479_0 | 17 | 0.9103 | 1 | 0.9103 | 0 | 0 | 0 | 1 | Ref |
| 14877 | OM416765 | Unassigned | Unassigned | Unassigned | 479_0 | Clustered | 17 | VC_479_0 | 17 | 0.9103 | 1 | 0.9103 | 0 | 0 | 0 | 1 | Ref |
| 14878 | OM416766 | Unassigned | Unassigned | Unassigned | 479_0 | Clustered | 17 | VC_479_0 | 17 | 0.9103 | 1 | 0.9103 | 0 | 0 | 0 | 1 | Ref |
| 14879 | OM416767 | Unassigned | Unassigned | Unassigned | 479_0 | Clustered | 17 | VC_479_0 | 17 | 0.9103 | 1 | 0.9103 | 0 | 0 | 0 | 1 | Ref |
| 14880 | OM416768 | Unassigned | Unassigned | Unassigned | 479_0 | Clustered | 17 | VC_479_0 | 17 | 0.9103 | 1 | 0.9103 | 0 | 0 | 0 | 1 | Ref |
| 14881 | OM416769 | Unassigned | Unassigned | Unassigned | 479_0 | Clustered | 17 | VC_479_0 | 17 | 0.9103 | 1 | 0.9103 | 0 | 0 | 0 | 1 | Ref |
| 14882 | OM416770 | Unassigned | Unassigned | Unassigned | 479_0 | Clustered | 17 | VC_479_0 | 17 | 0.9103 | 1 | 0.9103 | 0 | 0 | 0 | 1 | Ref |
| 14883 | OM416771 | Unassigned | Unassigned | Unassigned | 479_0 | Clustered | 17 | VC_479_0 | 17 | 0.9103 | 1 | 0.9103 | 0 | 0 | 0 | 1 | Ref |
| 14884 | OM416772 | Unassigned | Unassigned | Unassigned | 479_0 | Clustered | 17 | VC_479_0 | 17 | 0.9103 | 1 | 0.9103 | 0 | 0 | 0 | 1 | Ref |
| 14885 | OM416773 | Unassigned | Unassigned | Unassigned | 479_0 | Clustered | 17 | VC_479_0 | 17 | 0.9103 | 1 | 0.9103 | 0 | 0 | 0 | 1 | Ref |
| 14886 | OM416774 | Unassigned | Unassigned | Unassigned | 479_0 | Clustered | 17 | VC_479_0 | 17 | 0.9103 | 1 | 0.9103 | 0 | 0 | 0 | 1 | Ref |
| 14887 | OM416775 | Unassigned | Unassigned | Unassigned | 479_0 | Clustered | 17 | VC_479_0 | 17 | 0.9103 | 1 | 0.9103 | 0 | 0 | 0 | 1 | Ref |
| 14888 | OM416776 | Unassigned | Unassigned | Unassigned | 479_0 | Clustered | 17 | VC_479_0 | 17 | 0.9103 | 1 | 0.9103 | 0 | 0 | 0 | 1 | Ref |
| 14889 | OM416777 | Unassigned | Unassigned | Unassigned | 479_0 | Clustered | 17 | VC_479_0 | 17 | 0.9103 | 1 | 0.9103 | 0 | 0 | 0 | 1 | Ref |
| 14890 | OM416778 | Unassigned | Unassigned | Unassigned | 479_0 | Clustered | 17 | VC_479_0 | 17 | 0.9103 | 1 | 0.9103 | 0 | 0 | 0 | 1 | Ref |
| 14891 | OM416779 | Unassigned | Unassigned | Unassigned | 479_0 | Clustered | 17 | VC_479_0 | 17 | 0.9103 | 1 | 0.9103 | 0 | 0 | 0 | 1 | Ref |
| 14892 | OM418625 | Unassigned | Unassigned | Unassigned | 43_0 | Clustered | 38 | VC_43_0 | 38 | 0.5885 | 1 | 0.5885 | 0 | 0 | 0 | 0.9659 | Ref |
| 14893 | OM418626 | Unassigned | Unassigned | Unassigned | 43_0 | Clustered | 38 | VC_43_0 | 38 | 0.5885 | 1 | 0.5885 | 0 | 0 | 0 | 0.9659 | Ref |
| 14894 | OM418627 | Unassigned | Unassigned | Unassigned | 43_0 | Clustered | 38 | VC_43_0 | 38 | 0.5885 | 1 | 0.5885 | 0 | 0 | 0 | 0.9659 | Ref |
| 14895 | OM421595 | Unassigned | Unassigned | Unassigned | 77_0 | Clustered | 20 | VC_77_0 | 20 | 0.9657 | 1 | 0.9657 | 0 | 0 | 0 | 1 | Ref |
| 14896 | OM439662 | Unassigned | Unassigned | Unassigned | 9_0 | Clustered | 38 | VC_9_0 | 38 | 0.4895 | 1 | 0.4895 | 0 | 0 | 0 | 1 | Ref |
| 14897 | OM439663 | Unassigned | Unassigned | Unassigned | 12_0 | Clustered | 21 | VC_12_0 | 21 | 0.2274 | 1 | 0.2274 | 0 | 0 | 0 | 0.9571 | Ref |
| 14898 | OM439664 | Unassigned | Unassigned | Unassigned | 7_0 | Clustered | 28 | VC_7_0 | 28 | 0.3916 | 1 | 0.3916 | 0 | 0 | 0 | 0.9947 | Ref |
| 14899 | OM439665 | Unassigned | Unassigned | Unassigned | 8_0 | Clustered | 58 | VC_8_0 | 58 | 0.8306 | 1 | 0.8306 | 0 | 0 | 0 | 0.9946 | Ref |
| 14900 | OM439666 | Unassigned | Unassigned | Unassigned | 9_0 | Clustered | 38 | VC_9_0 | 38 | 0.4895 | 1 | 0.4895 | 0 | 0 | 0 | 1 | Ref |
| 14901 | OM439667 | Unassigned | Unassigned | Unassigned | 8_0 | Clustered | 58 | VC_8_0 | 58 | 0.8306 | 1 | 0.8306 | 0 | 0 | 0 | 0.9946 | Ref |
| 14902 | OM439668 | Unassigned | Unassigned | Unassigned | 9_0 | Clustered | 38 | VC_9_0 | 38 | 0.4895 | 1 | 0.4895 | 0 | 0 | 0 | 1 | Ref |

| | | | | | | | | | | | | | | | | | |
|---|---|---|---|---|---|---|---|---|---|---|---|---|---|---|---|---|---|
| 14903 | OM439669 | Unassigned | Unassigned | Unassigned | 12_0 | Clustered | 21 | VC_12_0 | 21 | 0.2274 | 1 | 0.2274 | 0 | 0 | 0 | 0.9571 | Ref |
| 14904 | OM439670 | Unassigned | Unassigned | Unassigned | 9_0 | Clustered | 38 | VC_9_0 | 38 | 0.4895 | 1 | 0.4895 | 0 | 0 | 0 | 1 | Ref |
| 14905 | OM439671 | Unassigned | Unassigned | Unassigned | 9_0 | Clustered | 38 | VC_9_0 | 38 | 0.4895 | 1 | 0.4895 | 0 | 0 | 0 | 1 | Ref |
| 14906 | OM439672 | Unassigned | Unassigned | Unassigned | 12_0 | Clustered | 21 | VC_12_0 | 21 | 0.2274 | 1 | 0.2274 | 0 | 0 | 0 | 0.9571 | Ref |
| 14907 | OM439673 | Unassigned | Unassigned | Unassigned | 12_0 | Clustered | 21 | VC_12_0 | 21 | 0.2274 | 1 | 0.2274 | 0 | 0 | 0 | 0.9571 | Ref |
| 14908 | OM439674 | Unassigned | Unassigned | Unassigned | 12_0 | Clustered | 21 | VC_12_0 | 21 | 0.2274 | 1 | 0.2274 | 0 | 0 | 0 | 0.9571 | Ref |
| 14909 | OM439675 | Unassigned | Unassigned | Unassigned | 9_0 | Clustered | 38 | VC_9_0 | 38 | 0.4895 | 1 | 0.4895 | 0 | 0 | 0 | 1 | Ref |
| 14910 | OM456492 | Unassigned | Unassigned | Unassigned | 959_5 | Clustered | 19 | VC_959_5 | 19 | 0.2626 | 1 | 0.2626 | 0 | 0 | 0 | 1 | Ref |
| 14911 | OM457001 | Unassigned | Unassigned | Unassigned | 284_0 | Clustered | 3 | VC_284_0 | 3 | 0.0495 | 0.99860966 | 0.0494 | 0 | 0 | 0 | 1 | Ref |
| 14912 | OM457002 | Unassigned | Unassigned | Unassigned | 451_0 | Clustered | 80 | VC_451_0 | 80 | 0.9991 | 1 | 0.9991 | 0 | 0 | 0 | 1 | Ref |
| 14913 | OM471789 | Unassigned | Unassigned | Unassigned | 129_0 | Clustered | 43 | VC_129_0 | 43 | 0.886 | 1 | 0.886 | 0 | 0 | 0 | 1 | Ref |
| 14914 | OM471864 | Unassigned | Unassigned | Unassigned | 698_0 | Clustered | 26 | VC_698_0 | 26 | 0.9366 | 1 | 0.9366 | 0 | 0 | 0 | 1 | Ref |
| 14915 | OM475426 | Unassigned | Unassigned | Unassigned | 228_0 | Clustered | 60 | VC_228_0 | 60 | 0.942 | 1 | 0.942 | 0 | 0 | 0 | 1 | Ref |
| 14916 | OM475427 | Unassigned | Unassigned | Unassigned | 43_0 | Clustered | 38 | VC_43_0 | 38 | 0.5885 | 1 | 0.5885 | 0 | 0 | 0 | 0.9659 | Ref |
| 14917 | OM475428 | Unassigned | Unassigned | Unassigned | 43_0 | Clustered | 38 | VC_43_0 | 38 | 0.5885 | 1 | 0.5885 | 0 | 0 | 0 | 0.9659 | Ref |
| 14918 | OM475429 | Unassigned | Unassigned | Unassigned | 228_0 | Clustered | 60 | VC_228_0 | 60 | 0.942 | 1 | 0.942 | 0 | 0 | 0 | 1 | Ref |
| 14919 | OM475430 | Unassigned | Unassigned | Unassigned | 43_0 | Clustered | 38 | VC_43_0 | 38 | 0.5885 | 1 | 0.5885 | 0 | 0 | 0 | 0.9659 | Ref |
| 14920 | OM475431 | Unassigned | Unassigned | Unassigned | 228_0 | Clustered | 60 | VC_228_0 | 60 | 0.942 | 1 | 0.942 | 0 | 0 | 0 | 1 | Ref |
| 14921 | OM475432 | Unassigned | Unassigned | Unassigned | 43_0 | Clustered | 38 | VC_43_0 | 38 | 0.5885 | 1 | 0.5885 | 0 | 0 | 0 | 0.9659 | Ref |
| 14922 | OM475433 | Unassigned | Unassigned | Unassigned | 228_0 | Clustered | 60 | VC_228_0 | 60 | 0.942 | 1 | 0.942 | 0 | 0 | 0 | 1 | Ref |
| 14923 | OM475434 | Unassigned | Unassigned | Unassigned | 228_0 | Clustered | 60 | VC_228_0 | 60 | 0.942 | 1 | 0.942 | 0 | 0 | 0 | 1 | Ref |
| 14924 | OM475435 | Unassigned | Unassigned | Unassigned | 43_0 | Clustered | 38 | VC_43_0 | 38 | 0.5885 | 1 | 0.5885 | 0 | 0 | 0 | 0.9659 | Ref |
| 14925 | OM486945 | Unassigned | Unassigned | Unassigned | 453_0 | Clustered | 6 | VC_453_0 | 6 | 0.114 | 1 | 0.114 | 0 | 0 | 0 | 1 | Ref |
| 14926 | OM513679 | Unassigned | Unassigned | Unassigned | 110_0 | Clustered | 4 | VC_110_0 | 4 | 0.1143 | 0.99998732 | 0.1143 | 0 | 0 | 0 | 1 | Ref |
| 14927 | OM513680 | Unassigned | Unassigned | Unassigned | 110_0 | Clustered | 4 | VC_110_0 | 4 | 0.1143 | 0.99998732 | 0.1143 | 0 | 0 | 0 | 1 | Ref |
| 14928 | OM515228 | Unassigned | Unassigned | Unassigned | 99_0 | Clustered | 32 | VC_99_0 | 32 | 0.8828 | 1 | 0.8828 | 0 | 0 | 0 | 1 | Ref |
| 14929 | OM519319 | Unassigned | Unassigned | Unassigned | 613_0 | Clustered | 2 | VC_613_0 | 2 | 0.0357 | 0.94194318 | 0.0336 | 0 | 0 | 0 | 1 | Ref |
| 14930 | OM522317 | Unassigned | Unassigned | Unassigned | 328_0 | Clustered | 11 | VC_328_0 | 11 | 0.4707 | 1 | 0.4707 | 0 | 0 | 0 | 1 | Ref |
| 14931 | OM522999 | Unassigned | Unassigned | Unassigned | 1054_0 | Clustered | 12 | VC_1054_0 | 12 | 0.2863 | 0.99746644 | 0.2855 | 0 | 0 | 0 | 1 | Ref |
| 14932 | OM523001 | Unassigned | Unassigned | Unassigned | 1054_0 | Clustered | 12 | VC_1054_0 | 12 | 0.2863 | 0.99746644 | 0.2855 | 0 | 0 | 0 | 1 | Ref |
| 14933 | OM523002 | Unassigned | Unassigned | Unassigned | 995_0 | Clustered | 69 | VC_995_0 | 69 | 0.8547 | 1 | 0.8547 | 0 | 0 | 0 | 1 | Ref |
| 14934 | OM541307 | Unassigned | Unassigned | Unassigned | 179_0 | Clustered | 53 | VC_179_0 | 53 | 0.9137 | 1 | 0.9137 | 0 | 0 | 0 | 1 | Ref |
| 14935 | OM621814 | Unassigned | Unassigned | Unassigned | 962_0 | Clustered | 2 | VC_962_0 | 2 | 0.3572 | 0.92263554 | 0.3295 | 0 | 0 | 0 | 1 | Ref |
| 14936 | OM621816 | Unassigned | Unassigned | Unassigned | 242_1 | Clustered | 20 | VC_242_1 | 20 | 0.684 | 1 | 0.684 | 0 | 0 | 0 | 0.8368 | Ref |
| 14937 | OM630219 | Unassigned | Unassigned | Unassigned | 491_0 | Clustered | 36 | VC_491_0 | 36 | 0.5336 | 1 | 0.5336 | 0 | 0 | 0 | 0.9714 | Ref |
| 14938 | OM630585 | Unassigned | Unassigned | Unassigned | 329_3 | Clustered | 24 | VC_329_3 | 24 | 0.1206 | 1 | 0.1206 | 0 | 0 | 0 | 0.7283 | Ref |
| 14939 | OM630586 | Unassigned | Unassigned | Unassigned | 392_0 | Clustered | 126 | VC_392_0 | 126 | 0.9621 | 1 | 0.9621 | 0 | 0 | 0 | 1 | Ref |
| 14940 | OM630588 | Unassigned | Unassigned | Unassigned | 603_0 | Clustered | 9 | VC_603_0 | 9 | 0.9925 | 0.99999994 | 0.9925 | 0 | 0 | 0 | 1 | Ref |
| 14941 | OM630590 | Unassigned | Unassigned | Unassigned | 129_0 | Clustered | 43 | VC_129_0 | 43 | 0.886 | 1 | 0.886 | 0 | 0 | 0 | 1 | Ref |
| 14942 | OM632664 | Unassigned | Unassigned | Unassigned | 779_0 | Clustered | 37 | VC_779_0 | 37 | 0.5682 | 1 | 0.5682 | 0 | 0 | 0 | 1 | Ref |
| 14943 | OM632665 | Unassigned | Unassigned | Unassigned | 779_0 | Clustered | 37 | VC_779_0 | 37 | 0.5682 | 1 | 0.5682 | 0 | 0 | 0 | 1 | Ref |
| 14944 | OM632666 | Unassigned | Unassigned | Unassigned | 779_0 | Clustered | 37 | VC_779_0 | 37 | 0.5682 | 1 | 0.5682 | 0 | 0 | 0 | 1 | Ref |
| 14945 | OM632667 | Unassigned | Unassigned | Unassigned | 638_0 | Clustered | 45 | VC_638_0 | 45 | 0.7741 | 1 | 0.7741 | 0 | 0 | 0 | 1 | Ref |
| 14946 | OM632668 | Unassigned | Unassigned | Unassigned | 638_0 | Clustered | 45 | VC_638_0 | 45 | 0.7741 | 1 | 0.7741 | 0 | 0 | 0 | 1 | Ref |
| 14947 | OM632669 | Unassigned | Unassigned | Unassigned | 779_0 | Clustered | 37 | VC_779_0 | 37 | 0.5682 | 1 | 0.5682 | 0 | 0 | 0 | 1 | Ref |
| 14948 | OM632670 | Unassigned | Unassigned | Unassigned | 779_0 | Clustered | 37 | VC_779_0 | 37 | 0.5682 | 1 | 0.5682 | 0 | 0 | 0 | 1 | Ref |
| 14949 | OM632671 | Unassigned | Unassigned | Unassigned | 286_7 | Clustered/Singleton | 1 | VC_286_7 | 1 | 0 | 1 | 0 | 0 | 0 | 0 | 1 | Ref |
| 14950 | OM632672 | Unassigned | Unassigned | Unassigned | 779_0 | Clustered | 37 | VC_779_0 | 37 | 0.5682 | 1 | 0.5682 | 0 | 0 | 0 | 1 | Ref |
| 14951 | OM634653 | Unassigned | Unassigned | Unassigned | 475_0 | Clustered | 3 | VC_475_0 | 3 | 0.0778 | 0.99798375 | 0.0776 | 0 | 0 | 0 | 1 | Ref |
| 14952 | OM634661 | Unassigned | Unassigned | Unassigned | 90_1 | Clustered/Singleton | 1 | VC_90_1 | 1 | 0 | 1 | 0 | 0 | 0 | 0 | 1 | Ref |
| 14953 | OM638103 | Unassigned | Unassigned | Unassigned | 580_0 | Clustered | 2 | VC_580_0 | 2 | 0.0632 | 0.95763305 | 0.0605 | 0 | 0 | 0 | 1 | Ref |
| 14954 | OM638104 | Unassigned | Unassigned | Unassigned | 899_0 | Clustered | 4 | VC_899_0 | 4 | 0.2655 | 0.99997104 | 0.2655 | 0 | 0 | 0 | 1 | Ref |
| 14955 | OM638608 | Unassigned | Unassigned | Unassigned | 3_0 | Clustered | 17 | VC_3_0 | 17 | 0.5639 | 1 | 0.5639 | 0 | 0 | 0 | 1 | Ref |
| 14956 | OM638609 | Unassigned | Unassigned | Unassigned | 3_0 | Clustered | 17 | VC_3_0 | 17 | 0.5639 | 1 | 0.5639 | 0 | 0 | 0 | 1 | Ref |
| 14957 | OM638610 | Unassigned | Unassigned | Unassigned | 3_0 | Clustered | 17 | VC_3_0 | 17 | 0.5639 | 1 | 0.5639 | 0 | 0 | 0 | 1 | Ref |
| 14958 | OM638611 | Unassigned | Unassigned | Unassigned | 3_0 | Clustered | 17 | VC_3_0 | 17 | 0.5639 | 1 | 0.5639 | 0 | 0 | 0 | 1 | Ref |
| 14959 | OM654379 | Unassigned | Unassigned | Unassigned | 214_1 | Clustered/Singleton | 1 | VC_214_1 | 1 | 0 | 1 | 0 | 0 | 0 | 0 | 1 | Ref |
| 14960 | OM654404 | Unassigned | Unassigned | Unassigned | 878_0 | Clustered | 32 | VC_878_0 | 32 | 0.9965 | 1 | 0.9965 | 0 | 0 | 0 | 1 | Ref |
| 14961 | OM681507 | Unassigned | Unassigned | Unassigned | 133_14 | Clustered | 3 | VC_133_14 | 3 | 0.0102 | 0.96327256 | 0.0098 | 0 | 0 | 0 | 1 | Ref |
| 14962 | OM687511 | Unassigned | Unassigned | Unassigned | 779_3 | Clustered | 7 | VC_779_3 | 7 | 0.0766 | 1 | 0.0766 | 0 | 0 | 0 | 0.7143 | Ref |
| 14963 | OM687892 | Unassigned | Unassigned | Unassigned | 836_0 | Clustered | 9 | VC_836_0 | 9 | 0.7733 | 1 | 0.7733 | 0 | 0 | 0 | 1 | Ref |
| 14964 | OM687893 | Unassigned | Unassigned | Unassigned | 938_0 | Clustered | 99 | VC_938_0 | 99 | 0.9119 | 1 | 0.9119 | 0 | 0 | 0 | 0.9928 | Ref |
| 14965 | OM688977 | Unassigned | Unassigned | Unassigned | 133_32 | Clustered/Singleton | 1 | VC_133_32 | 1 | 0 | 1 | 0 | 0 | 0 | 0 | 1 | Ref |
| 14966 | OM732336 | Unassigned | Unassigned | Unassigned | 500_0 | Clustered | 3 | VC_500_0 | 3 | 0.8278 | 0.9969176 | 0.8253 | 0 | 0 | 0 | 1 | Ref |
| 14967 | OM732337 | Unassigned | Unassigned | Unassigned | 500_0 | Clustered | 3 | VC_500_0 | 3 | 0.8278 | 0.9969176 | 0.8253 | 0 | 0 | 0 | 1 | Ref |
| 14968 | OM735690 | Unassigned | Unassigned | Unassigned | 937_0 | Clustered | 40 | VC_937_0 | 40 | 0.6809 | 1 | 0.6809 | 0 | 0 | 0 | 0.9359 | Ref |
| 14969 | OM743306 | Unassigned | Unassigned | Unassigned | 883_6 | Clustered/Singleton | 1 | VC_883_6 | 1 | 0 | 1 | 0 | 0 | 0 | 0 | 1 | Ref |
| 14970 | OM782324 | Unassigned | Unassigned | Unassigned | 1402_0 | Clustered | 14 | VC_1402_0 | 14 | 0.1673 | 0.51390354 | 0.086 | 0 | 0 | 0 | 1 | Ref |
| 14971 | OM782452 | Unassigned | Unassigned | Unassigned | 839_0 | Clustered | 5 | VC_839_0 | 5 | 0.165 | 0.99999995 | 0.165 | 0 | 0 | 0 | 1 | Ref |
| 14972 | OM793623 | Unassigned | Unassigned | Unassigned | 574_0 | Clustered | 7 | VC_574_0 | 7 | 0.2734 | 1 | 0.2734 | 0 | 0 | 0 | 1 | Ref |
| 14973 | OM799543 | Unassigned | Unassigned | Unassigned | 837_0 | Clustered | 29 | VC_837_0 | 29 | 0.9955 | 1 | 0.9955 | 0 | 0 | 0 | 1 | Ref |
| 14974 | OM809160 | Unassigned | Unassigned | Unassigned | 166_0 | Clustered | 26 | VC_166_0 | 26 | 0.7923 | 1 | 0.7923 | 0 | 0 | 0 | 1 | Ref |
| 14975 | OM810255 | Unassigned | Unassigned | Unassigned | 638_0 | Clustered | 45 | VC_638_0 | 45 | 0.7741 | 1 | 0.7741 | 0 | 0 | 0 | 1 | Ref |
| 14976 | OM810291 | Unassigned | Unassigned | Unassigned | 793_0 | Clustered | 12 | VC_793_0 | 12 | 0.8531 | 1 | 0.8531 | 0 | 0 | 0 | 1 | Ref |
| 14977 | OM810292 | Unassigned | Unassigned | Unassigned | 442_4 | Clustered | 2 | VC_442_4 | 2 | 0.0254 | 0.91859498 | 0.0233 | 0 | 0 | 0 | 1 | Ref |
| 14978 | OM818327 | Unassigned | Unassigned | Unassigned | 1180_0 | Clustered | 3 | VC_1180_0 | 3 | 0.5161 | 0.9981786 | 0.5151 | 0 | 0 | 0 | 1 | Ref |
| 14979 | OM818328 | Unassigned | Unassigned | Unassigned | 224_0 | Clustered | 19 | VC_224_0 | 19 | 0.9416 | 1 | 0.9416 | 0 | 0 | 0 | 1 | Ref |
| 14980 | OM818329 | Unassigned | Unassigned | Unassigned | 1234_0 | Clustered | 44 | VC_1234_0 | 44 | 1 | 1 | 1 | 0 | 0 | 0 | 1 | Ref |
| 14981 | OM818330 | Unassigned | Unassigned | Unassigned | 236_0 | Clustered | 72 | VC_236_0 | 72 | 0.8104 | 1 | 0.8104 | 0 | 0 | 0 | 1 | Ref |
| 14982 | OM818331 | Unassigned | Unassigned | Unassigned | 1170_2 | Clustered | 2 | VC_1170_2 | 2 | 0.0769 | 0.95418906 | 0.0734 | 0 | 0 | 0 | 1 | Ref |
| 14983 | OM837731 | Unassigned | Unassigned | Unassigned | 909_0 | Clustered | 5 | VC_909_0 | 5 | 0.7406 | 0.9999861 | 0.7406 | 0 | 0 | 0 | 1 | Ref |
| 14984 | OM850391 | Unassigned | Unassigned | Unassigned | 149_1 | Clustered | 3 | VC_149_1 | 3 | 0.0128 | 0.99303953 | 0.0127 | 0 | 0 | 0 | 1 | Ref |
| 14985 | OM850392 | Unassigned | Unassigned | Unassigned | 149_1 | Clustered | 3 | VC_149_1 | 3 | 0.0128 | 0.99303953 | 0.0127 | 0 | 0 | 0 | 1 | Ref |
| 14986 | OM858838 | Unassigned | Unassigned | Unassigned | 780_0 | Clustered | 8 | VC_780_0 | 8 | 0.5207 | 1 | 0.5207 | 0 | 0 | 0 | 1 | Ref |
| 14987 | OM864357 | Unassigned | Unassigned | Unassigned | 111_0 | Clustered | 91 | VC_111_0 | 91 | 0.9587 | 1 | 0.9587 | 0 | 0 | 0 | 1 | Ref |
| 14988 | OM867676 | Unassigned | Unassigned | Unassigned | 451_0 | Clustered | 80 | VC_451_0 | 80 | 0.9991 | 1 | 0.9991 | 0 | 0 | 0 | 1 | Ref |
| 14989 | OM869494 | Unassigned | Unassigned | Unassigned | 1275_0 | Clustered | 4 | VC_1275_0 | 4 | 1 | 1 | 1 | 0 | 0 | 0 | 1 | Ref |
| 14990 | OM869497 | Unassigned | Unassigned | Unassigned | 1069_0 | Clustered | 4 | VC_1069_0 | 4 | 1 | 1 | 1 | 0 | 0 | 0 | 1 | Ref |
| 14991 | OM869498 | Unassigned | Unassigned | Unassigned | 1308_0 | Clustered | 12 | VC_1308_0 | 12 | 0.8184 | 0.9810731 | 0.8029 | 0 | 0 | 0 | 1 | Ref |
| 14992 | OM869500 | Unassigned | Unassigned | Unassigned | 1282_0 | Clustered | 3 | VC_1282_0 | 3 | 1 | 1 | 1 | 0 | 0 | 0 | 1 | Ref |
| 14993 | OM869502 | Unassigned | Unassigned | Unassigned | 1308_0 | Clustered | 12 | VC_1308_0 | 12 | 0.8184 | 0.9810731 | 0.8029 | 0 | 0 | 0 | 1 | Ref |
| 14994 | OM869503 | Unassigned | Unassigned | Unassigned | 1119_0 | Clustered | 20 | VC_1119_0 | 20 | 0.6751 | 0.99997892 | 0.6751 | 0 | 0 | 0 | 1 | Ref |
| 14995 | OM869505 | Unassigned | Unassigned | Unassigned | 1308_0 | Clustered | 12 | VC_1308_0 | 12 | 0.8184 | 0.9810731 | 0.8029 | 0 | 0 | 0 | 1 | Ref |
| 14996 | OM869510 | Unassigned | Unassigned | Unassigned | 1468_0 | Clustered | 2 | VC_1468_0 | 2 | 1 | 1 | 1 | 0 | 0 | 0 | 1 | Ref |
| 14997 | OM869512 | Unassigned | Unassigned | Unassigned | 1468_0 | Clustered | 2 | VC_1468_0 | 2 | 1 | 1 | 1 | 0 | 0 | 0 | 1 | Ref |
| 14998 | OM869513 | Unassigned | Unassigned | Unassigned | 1145_0 | Clustered | 5 | VC_1145_0 | 5 | 0.8807 | 0.99921062 | 0.8801 | 0 | 0 | 0 | 1 | Ref |
| 14999 | OM869514 | Unassigned | Unassigned | Unassigned | 1321_0 | Clustered | 14 | VC_1321_0 | 14 | 0.826 | 1 | 0.826 | 0 | 0 | 0 | 1 | Ref |
| 15000 | OM869516 | Unassigned | Unassigned | Unassigned | 1145_0 | Clustered | 5 | VC_1145_0 | 5 | 0.8807 | 0.99921062 | 0.8801 | 0 | 0 | 0 | 1 | Ref |
| 15001 | OM869517 | Unassigned | Unassigned | Unassigned | 1133_0 | Clustered | 4 | VC_1133_0 | 4 | 0.1253 | 0.975067 | 0.1222 | 0 | 0 | 0 | 1 | Ref |
| 15002 | OM869519 | Unassigned | Unassigned | Unassigned | 1145_0 | Clustered | 5 | VC_1145_0 | 5 | 0.8807 | 0.99921062 | 0.8801 | 0 | 0 | 0 | 1 | Ref |
| 15003 | OM869521 | Unassigned | Unassigned | Unassigned | 1280_0 | Clustered | 2 | VC_1280_0 | 2 | 0.0695 | 0.94944661 | 0.066 | 0 | 0 | 0 | 1 | Ref |
| 15004 | OM869524 | Unassigned | Unassigned | Unassigned | 1104_0 | Clustered | 154 | VC_1104_0 | 154 | 0.8484 | 1 | 0.8484 | 0 | 0 | 0 | 1 | Ref |
| 15005 | OM869525 | Unassigned | Unassigned | Unassigned | 1104_0 | Clustered | 154 | VC_1104_0 | 154 | 0.8484 | 1 | 0.8484 | 0 | 0 | 0 | 1 | Ref |
| 15006 | OM869526 | Unassigned | Unassigned | Unassigned | 1104_0 | Clustered | 154 | VC_1104_0 | 154 | 0.8484 | 1 | 0.8484 | 0 | 0 | 0 | 1 | Ref |
| 15007 | OM869527 | Unassigned | Unassigned | Unassigned | 1104_0 | Clustered | 154 | VC_1104_0 | 154 | 0.8484 | 1 | 0.8484 | 0 | 0 | 0 | 1 | Ref |
| 15008 | OM869528 | Unassigned | Unassigned | Unassigned | 1104_0 | Clustered | 154 | VC_1104_0 | 154 | 0.8484 | 1 | 0.8484 | 0 | 0 | 0 | 1 | Ref |
| 15009 | OM869529 | Unassigned | Unassigned | Unassigned | 1321_0 | Clustered | 14 | VC_1321_0 | 14 | 0.826 | 1 | 0.826 | 0 | 0 | 0 | 1 | Ref |
| 15010 | OM869531 | Unassigned | Unassigned | Unassigned | 1104_0 | Clustered | 154 | VC_1104_0 | 154 | 0.8484 | 1 | 0.8484 | 0 | 0 | 0 | 1 | Ref |
| 15011 | OM869532 | Unassigned | Unassigned | Unassigned | 1058_0 | Clustered | 2 | VC_1058_0 | 2 | 0.5063 | 0.72985431 | 0.3695 | 0 | 0 | 0 | 1 | Ref |
| 15012 | OM869533 | Unassigned | Unassigned | Unassigned | 1104_0 | Clustered | 154 | VC_1104_0 | 154 | 0.8484 | 1 | 0.8484 | 0 | 0 | 0 | 1 | Ref |
| 15013 | OM869534 | Unassigned | Unassigned | Unassigned | 1104_0 | Clustered | 154 | VC_1104_0 | 154 | 0.8484 | 1 | 0.8484 | 0 | 0 | 0 | 1 | Ref |
| 15014 | OM869535 | Unassigned | Unassigned | Unassigned | 1104_0 | Clustered | 154 | VC_1104_0 | 154 | 0.8484 | 1 | 0.8484 | 0 | 0 | 0 | 1 | Ref |
| 15015 | OM869537 | Unassigned | Unassigned | Unassigned | 1104_0 | Clustered | 154 | VC_1104_0 | 154 | 0.8484 | 1 | 0.8484 | 0 | 0 | 0 | 1 | Ref |
| 15016 | OM869538 | Unassigned | Unassigned | Unassigned | 1104_0 | Clustered | 154 | VC_1104_0 | 154 | 0.8484 | 1 | 0.8484 | 0 | 0 | 0 | 1 | Ref |
| 15017 | OM869540 | Unassigned | Unassigned | Unassigned | 1321_0 | Clustered | 14 | VC_1321_0 | 14 | 0.826 | 1 | 0.826 | 0 | 0 | 0 | 1 | Ref |
| 15018 | OM869542 | Unassigned | Unassigned | Unassigned | 1321_0 | Clustered | 14 | VC_1321_0 | 14 | 0.826 | 1 | 0.826 | 0 | 0 | 0 | 1 | Ref |
| 15019 | OM869543 | Unassigned | Unassigned | Unassigned | 1104_0 | Clustered | 154 | VC_1104_0 | 154 | 0.8484 | 1 | 0.8484 | 0 | 0 | 0 | 1 | Ref |
| 15020 | OM869544 | Unassigned | Unassigned | Unassigned | 1321_0 | Clustered | 14 | VC_1321_0 | 14 | 0.826 | 1 | 0.826 | 0 | 0 | 0 | 1 | Ref |
| 15021 | OM869545 | Unassigned | Unassigned | Unassigned | 1134_0 | Clustered | 5 | VC_1134_0 | 5 | 0.129 | 0.7637089 | 0.0985 | 0 | 0 | 0 | 1 | Ref |
| 15022 | OM869546 | Unassigned | Unassigned | Unassigned | 1104_0 | Clustered | 154 | VC_1104_0 | 154 | 0.8484 | 1 | 0.8484 | 0 | 0 | 0 | 1 | Ref |
| 15023 | OM869547 | Unassigned | Unassigned | Unassigned | 1104_0 | Clustered | 154 | VC_1104_0 | 154 | 0.8484 | 1 | 0.8484 | 0 | 0 | 0 | 1 | Ref |
| 15024 | OM869548 | Unassigned | Unassigned | Unassigned | 1321_0 | Clustered | 14 | VC_1321_0 | 14 | 0.826 | 1 | 0.826 | 0 | 0 | 0 | 1 | Ref |
| 15025 | OM869549 | Unassigned | Unassigned | Unassigned | 1243_0 | Clustered | 52 | VC_1243_0 | 52 | 0.9126 | 0.92708311 | 0.8461 | 0 | 0 | 0 | 1 | Ref |
| 15026 | OM869550 | Unassigned | Unassigned | Unassigned | 1104_0 | Clustered | 154 | VC_1104_0 | 154 | 0.8484 | 1 | 0.8484 | 0 | 0 | 0 | 1 | Ref |
| 15027 | OM869552 | Unassigned | Unassigned | Unassigned | 1230_0 | Clustered | 3 | VC_1230_0 | 3 | 0.4802 | 0.82711071 | 0.3971 | 0 | 0 | 0 | 1 | Ref |
| 15028 | OM869553 | Unassigned | Unassigned | Unassigned | 1321_0 | Clustered | 14 | VC_1321_0 | 14 | 0.826 | 1 | 0.826 | 0 | 0 | 0 | 1 | Ref |
| 15029 | OM869555 | Unassigned | Unassigned | Unassigned | 1231_0 | Clustered | 2 | VC_1231_0 | 2 | 0.4148 | 0.89331315 | 0.3705 | 0 | 0 | 0 | 1 | Ref |
| 15030 | OM869557 | Unassigned | Unassigned | Unassigned | 1313_0 | Clustered | 13 | VC_1313_0 | 13 | 0.8481 | 0.87456432 | 0.7417 | 0 | 0 | 0 | 1 | Ref |
| 15031 | OM869558 | Unassigned | Unassigned | Unassigned | 1125_0 | Clustered | 20 | VC_1125_0 | 20 | 0.5599 | 1 | 0.5599 | 0 | 0 | 0 | 1 | Ref |
| 15032 | OM869559 | Unassigned | Unassigned | Unassigned | 1243_0 | Clustered | 52 | VC_1243_0 | 52 | 0.9126 | 0.92708311 | 0.8461 | 0 | 0 | 0 | 1 | Ref |
| 15033 | OM869560 | Unassigned | Unassigned | Unassigned | 1243_0 | Clustered | 52 | VC_1243_0 | 52 | 0.9126 | 0.92708311 | 0.8461 | 0 | 0 | 0 | 1 | Ref |
| 15034 | OM869561 | Unassigned | Unassigned | Unassigned | 1211_0 | Clustered | 25 | VC_1211_0 | 25 | 1 | 1 | 1 | 0 | 0 | 0 | 1 | Ref |
| 15035 | OM869562 | Unassigned | Unassigned | Unassigned | 1243_0 | Clustered | 52 | VC_1243_0 | 52 | 0.9126 | 0.92708311 | 0.8461 | 0 | 0 | 0 | 1 | Ref |
| 15036 | OM869563 | Unassigned | Unassigned | Unassigned | 1299_0 | Clustered | 2 | VC_1299_0 | 2 | 1 | 1 | 1 | 0 | 0 | 0 | 1 | Ref |
| 15037 | OM869564 | Unassigned | Unassigned | Unassigned | 1197_0 | Clustered | 15 | VC_1197_0 | 15 | 0.8991 | 0.76608394 | 0.6888 | 0 | 0 | 0 | 1 | Ref |
| 15038 | OM869565 | Unassigned | Unassigned | Unassigned | 1197_0 | Clustered | 15 | VC_1197_0 | 15 | 0.8991 | 0.76608394 | 0.6888 | 0 | 0 | 0 | 1 | Ref |
| 15039 | OM869566 | Unassigned | Unassigned | Unassigned | 1314_0 | Clustered | 4 | VC_1314_0 | 4 | 0.6744 | 0.9990299 | 0.6738 | 0 | 0 | 0 | 1 | Ref |
| 15040 | OM869568 | Unassigned | Unassigned | Unassigned | 1314_0 | Clustered | 4 | VC_1314_0 | 4 | 0.6744 | 0.9990299 | 0.6738 | 0 | 0 | 0 | 1 | Ref |
| 15041 | OM869569 | Unassigned | Unassigned | Unassigned | 1243_0 | Clustered | 52 | VC_1243_0 | 52 | 0.9126 | 0.92708311 | 0.8461 | 0 | 0 | 0 | 1 | Ref |
| 15042 | OM869570 | Unassigned | Unassigned | Unassigned | 1197_0 | Clustered | 15 | VC_1197_0 | 15 | 0.8991 | 0.76608394 | 0.6888 | 0 | 0 | 0 | 1 | Ref |
| 15043 | OM869571 | Unassigned | Unassigned | Unassigned | 1294_0 | Clustered | 6 | VC_1294_0 | 6 | 0.6626 | 0.99999272 | 0.6626 | 0 | 0 | 0 | 1 | Ref |
| 15044 | OM869572 | Unassigned | Unassigned | Unassigned | 1469_0 | Clustered | 2 | VC_1469_0 | 2 | 1 | 1 | 1 | 0 | 0 | 0 | 1 | Ref |
| 15045 | OM869573 | Unassigned | Unassigned | Unassigned | 1469_0 | Clustered | 2 | VC_1469_0 | 2 | 1 | 1 | 1 | 0 | 0 | 0 | 1 | Ref |
| 15046 | OM869574 | Unassigned | Unassigned | Unassigned | 1304_0 | Clustered | 7 | VC_1304_0 | 7 | 0.7931 | 0.9999998 | 0.7931 | 0 | 0 | 0 | 1 | Ref |
| 15047 | OM869576 | Unassigned | Unassigned | Unassigned | 1304_0 | Clustered | 7 | VC_1304_0 | 7 | 0.7931 | 0.9999998 | 0.7931 | 0 | 0 | 0 | 1 | Ref |
| 15048 | OM869577 | Unassigned | Unassigned | Unassigned | 1304_0 | Clustered | 7 | VC_1304_0 | 7 | 0.7931 | 0.9999998 | 0.7931 | 0 | 0 | 0 | 1 | Ref |
| 15049 | OM869578 | Unassigned | Unassigned | Unassigned | 1371_0 | Clustered | 5 | VC_1371_0 | 5 | 0.7004 | 0.52188382 | 0.3656 | 0 | 0 | 0 | 1 | Ref |
| 15050 | OM869583 | Unassigned | Unassigned | Unassigned | 1470_0 | Clustered | 3 | VC_1470_0 | 3 | 1 | 1 | 1 | 0 | 0 | 0 | 1 | Ref |
| 15051 | OM869587 | Unassigned | Unassigned | Unassigned | 1308_0 | Clustered | 12 | VC_1308_0 | 12 | 0.8184 | 0.9810731 | 0.8029 | 0 | 0 | 0 | 1 | Ref |
| 15052 | OM869588 | Unassigned | Unassigned | Unassigned | 1458_0 | Clustered | 2 | VC_1458_0 | 2 | 1 | 1 | 1 | 0 | 0 | 0 | 1 | Ref |
| 15053 | OM869589 | Unassigned | Unassigned | Unassigned | 1321_0 | Clustered | 14 | VC_1321_0 | 14 | 0.826 | 1 | 0.826 | 0 | 0 | 0 | 1 | Ref |
| 15054 | OM869590 | Unassigned | Unassigned | Unassigned | 1321_0 | Clustered | 14 | VC_1321_0 | 14 | 0.826 | 1 | 0.826 | 0 | 0 | 0 | 1 | Ref |
| 15055 | OM869592 | Unassigned | Unassigned | Unassigned | 1304_0 | Clustered | 7 | VC_1304_0 | 7 | 0.7931 | 0.9999998 | 0.7931 | 0 | 0 | 0 | 1 | Ref |
| 15056 | OM869593 | Unassigned | Unassigned | Unassigned | 1462_0 | Clustered | 4 | VC_1462_0 | 4 | 1 | 1 | 1 | 0 | 0 | 0 | 1 | Ref |
| 15057 | OM869594 | Unassigned | Unassigned | Unassigned | 1471_0 | Clustered | 2 | VC_1471_0 | 2 | 1 | 1 | 1 | 0 | 0 | 0 | 1 | Ref |
| 15058 | OM869604 | Unassigned | Unassigned | Unassigned | 1308_0 | Clustered | 12 | VC_1308_0 | 12 | 0.8184 | 0.9810731 | 0.8029 | 0 | 0 | 0 | 1 | Ref |
| 15059 | OM869606 | Unassigned | Unassigned | Unassigned | 1104_0 | Clustered | 154 | VC_1104_0 | 154 | 0.8484 | 1 | 0.8484 | 0 | 0 | 0 | 1 | Ref |
| 15060 | OM869610 | Unassigned | Unassigned | Unassigned | 1211_0 | Clustered | 25 | VC_1211_0 | 25 | 1 | 1 | 1 | 0 | 0 | 0 | 1 | Ref |
| 15061 | OM869612 | Unassigned | Unassigned | Unassigned | 1277_0 | Clustered | 14 | VC_1277_0 | 14 | 0.8218 | 1 | 0.8218 | 0 | 0 | 0 | 1 | Ref |
| 15062 | OM869623 | Unassigned | Unassigned | Unassigned | 1285_0 | Clustered | 25 | VC_1285_0 | 25 | 1 | 1 | 1 | 0 | 0 | 0 | 1 | Ref |
| 15063 | OM869624 | Unassigned | Unassigned | Unassigned | 1054_0 | Clustered | 12 | VC_1054_0 | 12 | 0.2863 | 0.99746644 | 0.2855 | 0 | 0 | 0 | 1 | Ref |
| 15064 | OM869626 | Unassigned | Unassigned | Unassigned | 1309_0 | Clustered | 2 | VC_1309_0 | 2 | 0.309 | 0.72985431 | 0.2255 | 0 | 0 | 0 | 1 | Ref |

| | | | | | | | | | | | | | | | | | |
|---|---|---|---|---|---|---|---|---|---|---|---|---|---|---|---|---|---|
| 15065 | OM869627 | Unassigned | Unassigned | Unassigned | 1120_0 | Clustered | 7 | VC_1120_0 | 7 | 0.2688 | 0.9951868 | 0.2675 | 0 | 0 | 0 | 1 | Ref |
| 15066 | OM869629 | Unassigned | Unassigned | Unassigned | 1120_0 | Clustered | 7 | VC_1120_0 | 7 | 0.2688 | 0.9951868 | 0.2675 | 0 | 0 | 0 | 1 | Ref |
| 15067 | OM869632 | Unassigned | Unassigned | Unassigned | 1394_0 | Clustered | 2 | VC_1394_0 | 2 | 1 | 1 | 1 | 0 | 0 | 0 | 1 | Ref |
| 15068 | OM869633 | Unassigned | Unassigned | Unassigned | 1308_0 | Clustered | 12 | VC_1308_0 | 12 | 0.8184 | 0.9810731 | 0.8029 | 0 | 0 | 0 | 1 | Ref |
| 15069 | OM869634 | Unassigned | Unassigned | Unassigned | 1145_0 | Clustered | 5 | VC_1145_0 | 5 | 0.8807 | 0.99921062 | 0.8801 | 0 | 0 | 0 | 1 | Ref |
| 15070 | OM869635 | Unassigned | Unassigned | Unassigned | 1104_0 | Clustered | 154 | VC_1104_0 | 154 | 0.8484 | 1 | 0.8484 | 0 | 0 | 0 | 1 | Ref |
| 15071 | OM869637 | Unassigned | Unassigned | Unassigned | 1145_0 | Clustered | 5 | VC_1145_0 | 5 | 0.8807 | 0.99921062 | 0.8801 | 0 | 0 | 0 | 1 | Ref |
| 15072 | OM869638 | Unassigned | Unassigned | Unassigned | 1321_0 | Clustered | 14 | VC_1321_0 | 14 | 0.826 | 1 | 0.826 | 0 | 0 | 0 | 1 | Ref |
| 15073 | OM869639 | Unassigned | Unassigned | Unassigned | 1332_0 | Clustered | 2 | VC_1332_0 | 2 | 0.4405 | 0.9197097 | 0.4052 | 0 | 0 | 0 | 1 | Ref |
| 15074 | OM869640 | Unassigned | Unassigned | Unassigned | 1133_0 | Clustered | 4 | VC_1133_0 | 4 | 0.1253 | 0.975067 | 0.1222 | 0 | 0 | 0 | 1 | Ref |
| 15075 | OM869641 | Unassigned | Unassigned | Unassigned | 1104_0 | Clustered | 154 | VC_1104_0 | 154 | 0.8484 | 1 | 0.8484 | 0 | 0 | 0 | 1 | Ref |
| 15076 | OM869642 | Unassigned | Unassigned | Unassigned | 1104_0 | Clustered | 154 | VC_1104_0 | 154 | 0.8484 | 1 | 0.8484 | 0 | 0 | 0 | 1 | Ref |
| 15077 | OM869643 | Unassigned | Unassigned | Unassigned | 1104_0 | Clustered | 154 | VC_1104_0 | 154 | 0.8484 | 1 | 0.8484 | 0 | 0 | 0 | 1 | Ref |
| 15078 | OM869644 | Unassigned | Unassigned | Unassigned | 1104_0 | Clustered | 154 | VC_1104_0 | 154 | 0.8484 | 1 | 0.8484 | 0 | 0 | 0 | 1 | Ref |
| 15079 | OM869645 | Unassigned | Unassigned | Unassigned | 1104_0 | Clustered | 154 | VC_1104_0 | 154 | 0.8484 | 1 | 0.8484 | 0 | 0 | 0 | 1 | Ref |
| 15080 | OM869648 | Unassigned | Unassigned | Unassigned | 1313_0 | Clustered | 13 | VC_1313_0 | 13 | 0.8481 | 0.87456432 | 0.7417 | 0 | 0 | 0 | 1 | Ref |
| 15081 | OM869650 | Unassigned | Unassigned | Unassigned | 1243_0 | Clustered | 52 | VC_1243_0 | 52 | 0.9126 | 0.92708311 | 0.8461 | 0 | 0 | 0 | 1 | Ref |
| 15082 | OM869652 | Unassigned | Unassigned | Unassigned | 1313_0 | Clustered | 13 | VC_1313_0 | 13 | 0.8481 | 0.87456432 | 0.7417 | 0 | 0 | 0 | 1 | Ref |
| 15083 | OM869653 | Unassigned | Unassigned | Unassigned | 1304_0 | Clustered | 7 | VC_1304_0 | 7 | 0.7931 | 0.9999998 | 0.7931 | 0 | 0 | 0 | 1 | Ref |
| 15084 | OM869654 | Unassigned | Unassigned | Unassigned | 1313_0 | Clustered | 13 | VC_1313_0 | 13 | 0.8481 | 0.87456432 | 0.7417 | 0 | 0 | 0 | 1 | Ref |
| 15085 | OM869655 | Unassigned | Unassigned | Unassigned | 1313_0 | Clustered | 13 | VC_1313_0 | 13 | 0.8481 | 0.87456432 | 0.7417 | 0 | 0 | 0 | 1 | Ref |
| 15086 | OM869663 | Unassigned | Unassigned | Unassigned | 1125_0 | Clustered | 20 | VC_1125_0 | 20 | 0.5599 | 1 | 0.5599 | 0 | 0 | 0 | 1 | Ref |
| 15087 | OM869664 | Unassigned | Unassigned | Unassigned | 1104_0 | Clustered | 154 | VC_1104_0 | 154 | 0.8484 | 1 | 0.8484 | 0 | 0 | 0 | 1 | Ref |
| 15088 | OM869666 | Unassigned | Unassigned | Unassigned | 1313_0 | Clustered | 13 | VC_1313_0 | 13 | 0.8481 | 0.87456432 | 0.7417 | 0 | 0 | 0 | 1 | Ref |
| 15089 | OM869668 | Unassigned | Unassigned | Unassigned | 1277_0 | Clustered | 14 | VC_1277_0 | 14 | 0.8218 | 1 | 0.8218 | 0 | 0 | 0 | 1 | Ref |
| 15090 | OM869669 | Unassigned | Unassigned | Unassigned | 1462_0 | Clustered | 4 | VC_1462_0 | 4 | 1 | 1 | 1 | 0 | 0 | 0 | 1 | Ref |
| 15091 | OM869672 | Unassigned | Unassigned | Unassigned | 1470_0 | Clustered | 3 | VC_1470_0 | 3 | 1 | 1 | 1 | 0 | 0 | 0 | 1 | Ref |
| 15092 | OM869677 | Unassigned | Unassigned | Unassigned | 1154_0 | Clustered | 5 | VC_1154_0 | 5 | 1 | 1 | 1 | 0 | 0 | 0 | 1 | Ref |
| 15093 | OM869679 | Unassigned | Unassigned | Unassigned | 1332_0 | Clustered | 2 | VC_1332_0 | 2 | 0.4405 | 0.9197097 | 0.4052 | 0 | 0 | 0 | 1 | Ref |
| 15094 | OM869684 | Unassigned | Unassigned | Unassigned | 1313_0 | Clustered | 13 | VC_1313_0 | 13 | 0.8481 | 0.87456432 | 0.7417 | 0 | 0 | 0 | 1 | Ref |
| 15095 | OM869688 | Unassigned | Unassigned | Unassigned | 1470_0 | Clustered | 3 | VC_1470_0 | 3 | 1 | 1 | 1 | 0 | 0 | 0 | 1 | Ref |
| 15096 | OM869691 | Unassigned | Unassigned | Unassigned | 1308_0 | Clustered | 12 | VC_1308_0 | 12 | 0.8184 | 0.9810731 | 0.8029 | 0 | 0 | 0 | 1 | Ref |
| 15097 | OM869696 | Unassigned | Unassigned | Unassigned | 1303_0 | Clustered | 6 | VC_1303_0 | 6 | 1 | 1 | 1 | 0 | 0 | 0 | 1 | Ref |
| 15098 | OM869697 | Unassigned | Unassigned | Unassigned | 1104_0 | Clustered | 154 | VC_1104_0 | 154 | 0.8484 | 1 | 0.8484 | 0 | 0 | 0 | 1 | Ref |
| 15099 | OM869698 | Unassigned | Unassigned | Unassigned | 1104_0 | Clustered | 154 | VC_1104_0 | 154 | 0.8484 | 1 | 0.8484 | 0 | 0 | 0 | 1 | Ref |
| 15100 | OM869700 | Unassigned | Unassigned | Unassigned | 1104_0 | Clustered | 154 | VC_1104_0 | 154 | 0.8484 | 1 | 0.8484 | 0 | 0 | 0 | 1 | Ref |
| 15101 | OM869701 | Unassigned | Unassigned | Unassigned | 1104_0 | Clustered | 154 | VC_1104_0 | 154 | 0.8484 | 1 | 0.8484 | 0 | 0 | 0 | 1 | Ref |
| 15102 | OM869702 | Unassigned | Unassigned | Unassigned | 1058_0 | Clustered | 2 | VC_1058_0 | 2 | 0.5063 | 0.72985431 | 0.3695 | 0 | 0 | 0 | 1 | Ref |
| 15103 | OM869703 | Unassigned | Unassigned | Unassigned | 1104_0 | Clustered | 154 | VC_1104_0 | 154 | 0.8484 | 1 | 0.8484 | 0 | 0 | 0 | 1 | Ref |
| 15104 | OM869704 | Unassigned | Unassigned | Unassigned | 1104_0 | Clustered | 154 | VC_1104_0 | 154 | 0.8484 | 1 | 0.8484 | 0 | 0 | 0 | 1 | Ref |
| 15105 | OM869706 | Unassigned | Unassigned | Unassigned | 1459_0 | Clustered | 2 | VC_1459_0 | 2 | 1 | 1 | 1 | 0 | 0 | 0 | 1 | Ref |
| 15106 | OM869707 | Unassigned | Unassigned | Unassigned | 1321_0 | Clustered | 14 | VC_1321_0 | 14 | 0.826 | 1 | 0.826 | 0 | 0 | 0 | 1 | Ref |
| 15107 | OM869708 | Unassigned | Unassigned | Unassigned | 1460_0 | Clustered | 3 | VC_1460_0 | 3 | 0.7846 | 0.96173875 | 0.7546 | 0 | 0 | 0 | 1 | Ref |
| 15108 | OM869709 | Unassigned | Unassigned | Unassigned | 1197_0 | Clustered | 15 | VC_1197_0 | 15 | 0.8991 | 0.76608394 | 0.6888 | 0 | 0 | 0 | 1 | Ref |
| 15109 | OM869711 | Unassigned | Unassigned | Unassigned | 1371_0 | Clustered | 5 | VC_1371_0 | 5 | 0.7004 | 0.52188382 | 0.3656 | 0 | 0 | 0 | 1 | Ref |
| 15110 | OM869712 | Unassigned | Unassigned | Unassigned | 1462_0 | Clustered | 4 | VC_1462_0 | 4 | 1 | 1 | 1 | 0 | 0 | 0 | 1 | Ref |
| 15111 | OM869713 | Unassigned | Unassigned | Unassigned | 1471_0 | Clustered | 2 | VC_1471_0 | 2 | 1 | 1 | 1 | 0 | 0 | 0 | 1 | Ref |
| 15112 | OM892879 | Unassigned | Unassigned | Unassigned | 353_0 | Clustered | 6 | VC_353_0 | 6 | 0.545 | 1 | 0.545 | 0 | 0 | 0 | 1 | Ref |
| 15113 | OM897575 | Unassigned | Unassigned | Unassigned | 778_0 | Clustered | 4 | VC_778_0 | 4 | 0.8115 | 0.99985435 | 0.8114 | 0 | 0 | 0 | 1 | Ref |
| 15114 | OM908436 | Unassigned | Unassigned | Unassigned | 451_0 | Clustered | 80 | VC_451_0 | 80 | 0.9991 | 1 | 0.9991 | 0 | 0 | 0 | 1 | Ref |
| 15115 | OM908766 | Unassigned | Unassigned | Unassigned | 780_0 | Clustered | 8 | VC_780_0 | 8 | 0.5207 | 1 | 0.5207 | 0 | 0 | 0 | 1 | Ref |
| 15116 | OM913584 | Unassigned | Unassigned | Unassigned | 236_0 | Clustered | 72 | VC_236_0 | 72 | 0.8104 | 1 | 0.8104 | 0 | 0 | 0 | 1 | Ref |
| 15117 | OM913585 | Unassigned | Unassigned | Unassigned | 223_0 | Clustered | 38 | VC_223_0 | 38 | 0.9095 | 1 | 0.9095 | 0 | 0 | 0 | 1 | Ref |
| 15118 | OM913586 | Unassigned | Unassigned | Unassigned | 1044_0 | Clustered | 3 | VC_1044_0 | 3 | 0.0843 | 0.99852363 | 0.0842 | 0 | 0 | 0 | 1 | Ref |
| 15119 | OM913587 | Unassigned | Unassigned | Unassigned | 1360_0 | Clustered | 31 | VC_1360_0 | 31 | 0.8837 | 1 | 0.8837 | 0 | 0 | 0 | 1 | Ref |
| 15120 | OM913597 | Unassigned | Unassigned | Unassigned | 159_0 | Clustered | 2 | VC_159_0 | 2 | 0.1337 | 0.95628623 | 0.1279 | 0 | 0 | 0 | 1 | Ref |
| 15121 | OM913598 | Unassigned | Unassigned | Unassigned | 159_0 | Clustered | 2 | VC_159_0 | 2 | 0.1337 | 0.95628623 | 0.1279 | 0 | 0 | 0 | 1 | Ref |
| 15122 | OM913894 | Unassigned | Unassigned | Unassigned | 113_0 | Clustered | 9 | VC_113_0 | 9 | 0.519 | 1 | 0.519 | 0 | 0 | 0 | 1 | Ref |
| 15123 | OM925528 | Unassigned | Unassigned | Unassigned | 699_0 | Clustered | 71 | VC_699_0 | 71 | 0.9352 | 1 | 0.9352 | 0 | 0 | 0 | 1 | Ref |
| 15124 | OM937123 | Unassigned | Unassigned | Unassigned | 451_0 | Clustered | 80 | VC_451_0 | 80 | 0.9991 | 1 | 0.9991 | 0 | 0 | 0 | 1 | Ref |
| 15125 | OM937766 | Unassigned | Unassigned | Unassigned | 838_0 | Clustered | 29 | VC_838_0 | 29 | 0.8611 | 1 | 0.8611 | 0 | 0 | 0 | 1 | Ref |
| 15126 | OM938991 | Unassigned | Unassigned | Unassigned | 938_0 | Clustered | 99 | VC_938_0 | 99 | 0.9119 | 1 | 0.9119 | 0 | 0 | 0 | 0.9928 | Ref |
| 15127 | OM938992 | Unassigned | Unassigned | Unassigned | 938_0 | Clustered | 99 | VC_938_0 | 99 | 0.9119 | 1 | 0.9119 | 0 | 0 | 0 | 0.9928 | Ref |
| 15128 | OM953433 | Unassigned | Unassigned | Unassigned | 330_0 | Clustered | 27 | VC_330_0 | 27 | 0.5088 | 1 | 0.5088 | 0 | 0 | 0 | 0.8661 | Ref |
| 15129 | OM953790 | Unassigned | Unassigned | Unassigned | 79_0 | Clustered | 64 | VC_79_0 | 64 | 0.4185 | 1 | 0.4185 | 0 | 0 | 0 | 0.7649 | Ref |
| 15130 | OM960734 | Unassigned | Unassigned | Unassigned | 286_1 | Clustered | 21 | VC_286_1 | 21 | 0.1696 | 1 | 0.1696 | 0 | 0 | 0 | 0.9048 | Ref |
| 15131 | OM962992 | Unassigned | Unassigned | Unassigned | 591_0 | Clustered | 14 | VC_591_0 | 14 | 0.5184 | 1 | 0.5184 | 0 | 0 | 0 | 1 | Ref |
| 15132 | OM964875 | Unassigned | Unassigned | Unassigned | 938_0 | Clustered | 99 | VC_938_0 | 99 | 0.9119 | 1 | 0.9119 | 0 | 0 | 0 | 0.9928 | Ref |
| 15133 | OM966901 | Unassigned | Unassigned | Unassigned | 92_0 | Clustered | 22 | VC_92_0 | 22 | 0.5639 | 1 | 0.5639 | 0 | 0 | 0 | 1 | Ref |
| 15134 | OM971648 | Unassigned | Unassigned | Unassigned | 668_0 | Clustered | 25 | VC_668_0 | 25 | 0.5539 | 1 | 0.5539 | 0 | 0 | 0 | 0.96 | Ref |
| 15135 | OM971649 | Unassigned | Unassigned | Unassigned | 668_0 | Clustered | 25 | VC_668_0 | 25 | 0.5539 | 1 | 0.5539 | 0 | 0 | 0 | 0.96 | Ref |
| 15136 | OM982619 | Unassigned | Unassigned | Unassigned | 965_0 | Clustered | 2 | VC_965_0 | 2 | 0.2408 | 0.95689931 | 0.2304 | 0 | 0 | 0 | 1 | Ref |
| 15137 | OM982620 | Unassigned | Unassigned | Unassigned | 78_5 | Clustered | 6 | VC_78_5 | 6 | 0.1087 | 0.99978082 | 0.1087 | 0 | 0 | 0 | 1 | Ref |
| 15138 | OM982621 | Unassigned | Unassigned | Unassigned | 581_1 | Clustered/Singleton | 1 | VC_581_1 | 1 | 0 | 1 | 0 | 0 | 0 | 0 | 1 | Ref |
| 15139 | OM982646 | Unassigned | Unassigned | Unassigned | 779_0 | Clustered | 37 | VC_779_0 | 37 | 0.5682 | 1 | 0.5682 | 0 | 0 | 0 | 1 | Ref |
| 15140 | OM982647 | Unassigned | Unassigned | Unassigned | 286_10 | Clustered | 9 | VC_286_10 | 9 | 0.0615 | 0.99999878 | 0.0615 | 0 | 0 | 0 | 0.6667 | Ref |
| 15141 | OM982668 | Unassigned | Unassigned | Unassigned | 85_3 | Clustered | 19 | VC_85_3 | 19 | 0.3671 | 1 | 0.3671 | 0 | 0 | 0 | 0.807 | Ref |
| 15142 | OM982669 | Unassigned | Unassigned | Unassigned | 85_3 | Clustered | 19 | VC_85_3 | 19 | 0.3671 | 1 | 0.3671 | 0 | 0 | 0 | 0.807 | Ref |
| 15143 | OM982670 | Unassigned | Unassigned | Unassigned | 85_3 | Clustered | 19 | VC_85_3 | 19 | 0.3671 | 1 | 0.3671 | 0 | 0 | 0 | 0.807 | Ref |
| 15144 | OM982671 | Unassigned | Unassigned | Unassigned | 85_3 | Clustered | 19 | VC_85_3 | 19 | 0.3671 | 1 | 0.3671 | 0 | 0 | 0 | 0.807 | Ref |
| 15145 | OM982672 | Unassigned | Unassigned | Unassigned | 85_3 | Clustered | 19 | VC_85_3 | 19 | 0.3671 | 1 | 0.3671 | 0 | 0 | 0 | 0.807 | Ref |
| 15146 | OM982673 | Unassigned | Unassigned | Unassigned | 85_3 | Clustered | 19 | VC_85_3 | 19 | 0.3671 | 1 | 0.3671 | 0 | 0 | 0 | 0.807 | Ref |
| 15147 | OM982674 | Unassigned | Unassigned | Unassigned | 85_3 | Clustered | 19 | VC_85_3 | 19 | 0.3671 | 1 | 0.3671 | 0 | 0 | 0 | 0.807 | Ref |
| 15148 | ON000910 | Unassigned | Unassigned | Unassigned | 531_0 | Clustered | 9 | VC_531_0 | 9 | 0.26 | 1 | 0.26 | 0 | 0 | 0 | 1 | Ref |
| 15149 | ON001686 | Unassigned | Unassigned | Unassigned | 638_0 | Clustered | 45 | VC_638_0 | 45 | 0.7741 | 1 | 0.7741 | 0 | 0 | 0 | 1 | Ref |
| 15150 | ON005621 | Unassigned | Unassigned | Unassigned | 169_0 | Clustered | 20 | VC_169_0 | 20 | 0.7529 | 1 | 0.7529 | 0 | 0 | 0 | 1 | Ref |
| 15151 | ON014753 | Unassigned | Unassigned | Unassigned | 1472_0 | Clustered | 2 | VC_1472_0 | 2 | 1 | 1 | 1 | 0 | 0 | 0 | 1 | Ref |
| 15152 | ON014754 | Unassigned | Unassigned | Unassigned | 1433_0 | Clustered | 2 | VC_1433_0 | 2 | 1 | 1 | 1 | 0 | 0 | 0 | 1 | Ref |
| 15153 | ON014755 | Unassigned | Unassigned | Unassigned | 468_0 | Clustered | 4 | VC_468_0 | 4 | 0.143 | 0.99998478 | 0.143 | 0 | 0 | 0 | 1 | Ref |
| 15154 | ON014757 | Unassigned | Unassigned | Unassigned | 1472_0 | Clustered | 2 | VC_1472_0 | 2 | 1 | 1 | 1 | 0 | 0 | 0 | 1 | Ref |
| 15155 | ON015836 | Unassigned | Unassigned | Unassigned | 904_1 | Clustered | 4 | VC_904_1 | 4 | 0.1486 | 0.99997723 | 0.1486 | 0 | 0 | 0 | 1 | Ref |
| 15156 | ON018986 | Unassigned | Unassigned | Unassigned | 247_0 | Clustered | 14 | VC_247_0 | 14 | 0.9119 | 1 | 0.9119 | 0 | 0 | 0 | 0.967 | Ref |
| 15157 | ON036882 | Unassigned | Unassigned | Unassigned | 698_0 | Clustered | 26 | VC_698_0 | 26 | 0.9366 | 1 | 0.9366 | 0 | 0 | 0 | 1 | Ref |
| 15158 | ON036883 | Unassigned | Unassigned | Unassigned | 698_0 | Clustered | 26 | VC_698_0 | 26 | 0.9366 | 1 | 0.9366 | 0 | 0 | 0 | 1 | Ref |
| 15159 | ON042478 | Unassigned | Unassigned | Unassigned | 600_0 | Clustered | 3 | VC_600_0 | 3 | 0.0541 | 0.97110069 | 0.0526 | 0 | 0 | 0 | 1 | Ref |
| 15160 | ON042750 | Unassigned | Unassigned | Unassigned | 817_0 | Clustered | 14 | VC_817_0 | 14 | 0.6342 | 1 | 0.6342 | 0 | 0 | 0 | 1 | Ref |
| 15161 | ON045001 | Unassigned | Unassigned | Unassigned | 586_0 | Clustered | 3 | VC_586_0 | 3 | 0.0495 | 0.99859933 | 0.0495 | 0 | 0 | 0 | 1 | Ref |
| 15162 | ON080940 | Unassigned | Unassigned | Unassigned | 837_0 | Clustered | 29 | VC_837_0 | 29 | 0.9955 | 1 | 0.9955 | 0 | 0 | 0 | 1 | Ref |
| 15163 | ON080941 | Unassigned | Unassigned | Unassigned | 837_0 | Clustered | 29 | VC_837_0 | 29 | 0.9955 | 1 | 0.9955 | 0 | 0 | 0 | 1 | Ref |
| 15164 | ON081052 | Unassigned | Unassigned | Unassigned | 456_0 | Clustered | 62 | VC_456_0 | 62 | 0.9026 | 1 | 0.9026 | 0 | 0 | 0 | 1 | Ref |
| 15165 | ON081328 | Unassigned | Unassigned | Unassigned | 1360_0 | Clustered | 31 | VC_1360_0 | 31 | 0.8837 | 1 | 0.8837 | 0 | 0 | 0 | 1 | Ref |
| 15166 | ON081329 | Unassigned | Unassigned | Unassigned | 236_0 | Clustered | 72 | VC_236_0 | 72 | 0.8104 | 1 | 0.8104 | 0 | 0 | 0 | 1 | Ref |
| 15167 | ON081330 | Unassigned | Unassigned | Unassigned | 1352_0 | Clustered | 31 | VC_1352_0 | 31 | 0.849 | 1 | 0.849 | 0 | 0 | 0 | 1 | Ref |
| 15168 | ON081331 | Unassigned | Unassigned | Unassigned | 236_0 | Clustered | 72 | VC_236_0 | 72 | 0.8104 | 1 | 0.8104 | 0 | 0 | 0 | 1 | Ref |
| 15169 | ON081332 | Unassigned | Unassigned | Unassigned | 1093_0 | Clustered | 40 | VC_1093_0 | 40 | 0.8754 | 1 | 0.8754 | 0 | 0 | 0 | 1 | Ref |
| 15170 | ON081333 | Unassigned | Unassigned | Unassigned | 510_0 | Clustered | 12 | VC_510_0 | 12 | 0.3963 | 1 | 0.3963 | 0 | 0 | 0 | 0.9697 | Ref |
| 15171 | ON081334 | Unassigned | Unassigned | Unassigned | 1344_0 | Clustered | 78 | VC_1344_0 | 78 | 0.9732 | 1 | 0.9732 | 0 | 0 | 0 | 1 | Ref |
| 15172 | ON081335 | Unassigned | Unassigned | Unassigned | 558_20 | Clustered | 17 | VC_558_20 | 17 | 0.054 | 1 | 0.054 | 0 | 0 | 0 | 0.9926 | Ref |
| 15173 | ON086804 | Unassigned | Unassigned | Unassigned | 123_0 | Clustered | 23 | VC_123_0 | 23 | 0.817 | 1 | 0.817 | 0 | 0 | 0 | 1 | Ref |
| 15174 | ON086985 | Unassigned | Unassigned | Unassigned | 108_0 | Clustered | 33 | VC_108_0 | 33 | 0.949 | 1 | 0.949 | 0 | 0 | 0 | 1 | Ref |
| 15175 | ON107264 | Unassigned | Unassigned | Unassigned | 89_4 | Clustered/Singleton | 1 | VC_89_4 | 1 | 0 | 1 | 0 | 0 | 0 | 0 | 1 | Ref |
| 15176 | ON108640 | Unassigned | Unassigned | Unassigned | 221_0 | Clustered | 22 | VC_221_0 | 22 | 0.7447 | 1 | 0.7447 | 0 | 0 | 0 | 1 | Ref |
| 15177 | ON108642 | Unassigned | Unassigned | Unassigned | 1344_0 | Clustered | 78 | VC_1344_0 | 78 | 0.9732 | 1 | 0.9732 | 0 | 0 | 0 | 1 | Ref |
| 15178 | ON108643 | Unassigned | Unassigned | Unassigned | 573_0 | Clustered | 2 | VC_573_0 | 2 | 0.11 | 0.95639318 | 0.1052 | 0 | 0 | 0 | 1 | Ref |
| 15179 | ON108644 | Unassigned | Unassigned | Unassigned | 510_0 | Clustered | 12 | VC_510_0 | 12 | 0.3963 | 1 | 0.3963 | 0 | 0 | 0 | 0.9697 | Ref |
| 15180 | ON108645 | Unassigned | Unassigned | Unassigned | 574_0 | Clustered | 7 | VC_574_0 | 7 | 0.2734 | 1 | 0.2734 | 0 | 0 | 0 | 1 | Ref |
| 15181 | ON108646 | Unassigned | Unassigned | Unassigned | 872_0 | Clustered | 31 | VC_872_0 | 31 | 0.8151 | 1 | 0.8151 | 0 | 0 | 0 | 0.8194 | Ref |
| 15182 | ON108647 | Unassigned | Unassigned | Unassigned | 1224_0 | Clustered | 19 | VC_1224_0 | 19 | 1 | 1 | 1 | 0 | 0 | 0 | 1 | Ref |
| 15183 | ON108648 | Unassigned | Unassigned | Unassigned | 1344_0 | Clustered | 78 | VC_1344_0 | 78 | 0.9732 | 1 | 0.9732 | 0 | 0 | 0 | 1 | Ref |
| 15184 | ON108649 | Unassigned | Unassigned | Unassigned | 569_0 | Clustered | 3 | VC_569_0 | 3 | 0.2094 | 0.99849867 | 0.2091 | 0 | 0 | 0 | 1 | Ref |
| 15185 | ON108650 | Unassigned | Unassigned | Unassigned | 1071_3 | Clustered | 16 | VC_1071_3 | 16 | 0.2354 | 1 | 0.2354 | 0 | 0 | 0 | 0.9667 | Ref |
| 15186 | ON108651 | Unassigned | Unassigned | Unassigned | 685_0 | Clustered | 16 | VC_685_0 | 16 | 0.2972 | 1 | 0.2972 | 0 | 0 | 0 | 0.9167 | Ref |
| 15187 | ON109291 | Unassigned | Unassigned | Unassigned | 130_0 | Clustered | 19 | VC_130_0 | 19 | 0.6068 | 1 | 0.6068 | 0 | 0 | 0 | 1 | Ref |
| 15188 | ON113167 | Unassigned | Unassigned | Unassigned | 91_0 | Clustered | 22 | VC_91_0 | 22 | 0.5318 | 1 | 0.5318 | 0 | 0 | 0 | 1 | Ref |
| 15189 | ON113168 | Unassigned | Unassigned | Unassigned | 91_0 | Clustered | 22 | VC_91_0 | 22 | 0.5318 | 1 | 0.5318 | 0 | 0 | 0 | 1 | Ref |
| 15190 | ON113169 | Unassigned | Unassigned | Unassigned | 91_0 | Clustered | 22 | VC_91_0 | 22 | 0.5318 | 1 | 0.5318 | 0 | 0 | 0 | 1 | Ref |
| 15191 | ON113170 | Unassigned | Unassigned | Unassigned | 92_0 | Clustered | 22 | VC_92_0 | 22 | 0.5639 | 1 | 0.5639 | 0 | 0 | 0 | 1 | Ref |
| 15192 | ON113171 | Unassigned | Unassigned | Unassigned | 108_0 | Clustered | 33 | VC_108_0 | 33 | 0.949 | 1 | 0.949 | 0 | 0 | 0 | 1 | Ref |
| 15193 | ON113172 | Unassigned | Unassigned | Unassigned | 108_0 | Clustered | 33 | VC_108_0 | 33 | 0.949 | 1 | 0.949 | 0 | 0 | 0 | 1 | Ref |
| 15194 | ON113173 | Unassigned | Unassigned | Unassigned | 108_0 | Clustered | 33 | VC_108_0 | 33 | 0.949 | 1 | 0.949 | 0 | 0 | 0 | 1 | Ref |
| 15195 | ON113174 | Unassigned | Unassigned | Unassigned | 108_0 | Clustered | 33 | VC_108_0 | 33 | 0.949 | 1 | 0.949 | 0 | 0 | 0 | 1 | Ref |
| 15196 | ON113175 | Unassigned | Unassigned | Unassigned | 108_0 | Clustered | 33 | VC_108_0 | 33 | 0.949 | 1 | 0.949 | 0 | 0 | 0 | 1 | Ref |
| 15197 | ON113176 | Unassigned | Unassigned | Unassigned | 695_0 | Clustered | 48 | VC_695_0 | 48 | 0.8162 | 1 | 0.8162 | 0 | 0 | 0 | 1 | Ref |
| 15198 | ON113177 | Unassigned | Unassigned | Unassigned | 108_0 | Clustered | 33 | VC_108_0 | 33 | 0.949 | 1 | 0.949 | 0 | 0 | 0 | 1 | Ref |
| 15199 | ON113178 | Unassigned | Unassigned | Unassigned | 108_0 | Clustered | 33 | VC_108_0 | 33 | 0.949 | 1 | 0.949 | 0 | 0 | 0 | 1 | Ref |
| 15200 | ON113179 | Unassigned | Unassigned | Unassigned | 108_0 | Clustered | 33 | VC_108_0 | 33 | 0.949 | 1 | 0.949 | 0 | 0 | 0 | 1 | Ref |
| 15201 | ON113180 | Unassigned | Unassigned | Unassigned | 108_0 | Clustered | 33 | VC_108_0 | 33 | 0.949 | 1 | 0.949 | 0 | 0 | 0 | 1 | Ref |
| 15202 | ON113181 | Unassigned | Unassigned | Unassigned | 968_0 | Clustered | 4 | VC_968_0 | 4 | 0.2959 | 0.99997642 | 0.2959 | 0 | 0 | 0 | 1 | Ref |
| 15203 | ON113334 | Unassigned | Unassigned | Unassigned | 695_0 | Clustered | 48 | VC_695_0 | 48 | 0.8162 | 1 | 0.8162 | 0 | 0 | 0 | 1 | Ref |
| 15204 | ON113344 | Unassigned | Unassigned | Unassigned | 149_1 | Clustered | 3 | VC_149_1 | 3 | 0.0128 | 0.99303953 | 0.0127 | 0 | 0 | 0 | 1 | Ref |
| 15205 | ON117106 | Unassigned | Unassigned | Unassigned | 634_1 | Clustered | 6 | VC_634_1 | 6 | 0.4228 | 1 | 0.4228 | 0 | 0 | 0 | 1 | Ref |
| 15206 | ON125307 | Unassigned | Unassigned | Unassigned | 92_0 | Clustered | 22 | VC_92_0 | 22 | 0.5639 | 1 | 0.5639 | 0 | 0 | 0 | 1 | Ref |
| 15207 | ON132389 | Unassigned | Unassigned | Unassigned | 111_0 | Clustered | 91 | VC_111_0 | 91 | 0.9587 | 1 | 0.9587 | 0 | 0 | 0 | 1 | Ref |
| 15208 | ON133270 | Unassigned | Unassigned | Unassigned | 475_0 | Clustered | 3 | VC_475_0 | 3 | 0.0778 | 0.99798375 | 0.0776 | 0 | 0 | 0 | 1 | Ref |
| 15209 | ON135435 | Unassigned | Unassigned | Unassigned | 3_0 | Clustered | 17 | VC_3_0 | 17 | 0.5639 | 1 | 0.5639 | 0 | 0 | 0 | 1 | Ref |
| 15210 | ON146439 | Unassigned | Unassigned | Unassigned | 228_0 | Clustered | 60 | VC_228_0 | 60 | 0.942 | 1 | 0.942 | 0 | 0 | 0 | 1 | Ref |
| 15211 | ON146440 | Unassigned | Unassigned | Unassigned | 228_0 | Clustered | 60 | VC_228_0 | 60 | 0.942 | 1 | 0.942 | 0 | 0 | 0 | 1 | Ref |
| 15212 | ON146449 | Unassigned | Unassigned | Unassigned | 111_0 | Clustered | 91 | VC_111_0 | 91 | 0.9587 | 1 | 0.9587 | 0 | 0 | 0 | 1 | Ref |
| 15213 | ON148527 | Unassigned | Unassigned | Unassigned | 452_0 | Clustered | 66 | VC_452_0 | 66 | 0.9248 | 1 | 0.9248 | 0 | 0 | 0 | 1 | Ref |
| 15214 | ON148528 | Unassigned | Unassigned | Unassigned | 823_0 | Clustered | 124 | VC_823_0 | 124 | 0.9492 | 1 | 0.9492 | 0 | 0 | 0 | 1 | Ref |
| 15215 | ON148529 | Unassigned | Unassigned | Unassigned | 823_0 | Clustered | 124 | VC_823_0 | 124 | 0.9492 | 1 | 0.9492 | 0 | 0 | 0 | 1 | Ref |
| 15216 | ON156559 | Unassigned | Unassigned | Unassigned | 472_0 | Clustered | 47 | VC_472_0 | 47 | 0.9025 | 1 | 0.9025 | 0 | 0 | 0 | 1 | Ref |
| 15217 | ON157416 | Unassigned | Unassigned | Unassigned | 669_0 | Clustered | 21 | VC_669_0 | 21 | 0.3623 | 1 | 0.3623 | 0 | 0 | 0 | 1 | Ref |
| 15218 | ON165415 | Unassigned | Unassigned | Unassigned | 111_0 | Clustered | 91 | VC_111_0 | 91 | 0.9587 | 1 | 0.9587 | 0 | 0 | 0 | 1 | Ref |
| 15219 | ON165687 | Unassigned | Unassigned | Unassigned | 166_0 | Clustered | 26 | VC_166_0 | 26 | 0.7923 | 1 | 0.7923 | 0 | 0 | 0 | 1 | Ref |
| 15220 | ON167532 | Unassigned | Unassigned | Unassigned | 637_0 | Clustered | 115 | VC_637_0 | 115 | 0.731 | 1 | 0.731 | 0 | 0 | 0 | 0.8764 | Ref |
| 15221 | ON169961 | Unassigned | Unassigned | Unassigned | 133_50 | Clustered/Singleton | 1 | VC_133_50 | 1 | 0 | 1 | 0 | 0 | 0 | 0 | 1 | Ref |
| 15222 | ON169962 | Unassigned | Unassigned | Unassigned | 133_51 | Clustered/Singleton | 1 | VC_133_51 | 1 | 0 | 1 | 0 | 0 | 0 | 0 | 1 | Ref |
| 15223 | ON169963 | Unassigned | Unassigned | Unassigned | 133_49 | Clustered/Singleton | 1 | VC_133_49 | 1 | 0 | 1 | 0 | 0 | 0 | 0 | 1 | Ref |
| 15224 | ON169972 | Unassigned | Unassigned | Unassigned | 76_0 | Clustered | 37 | VC_76_0 | 37 | 0.9403 | 1 | 0.9403 | 0 | 0 | 0 | 1 | Ref |
| 15225 | ON170017 | Unassigned | Unassigned | Unassigned | 25_0 | Clustered | 12 | VC_25_0 | 12 | 0.7945 | 1 | 0.7945 | 0 | 0 | 0 | 1 | Ref |
| 15226 | ON170019 | Unassigned | Unassigned | Unassigned | 826_0 | Clustered | 91 | VC_826_0 | 91 | 0.9935 | 1 | 0.9935 | 0 | 0 | 0 | 1 | Ref |

| | | | | | | | | | | | | | | | | | |
|---|---|---|---|---|---|---|---|---|---|---|---|---|---|---|---|---|---|
| 15227 | ON170020 | Unassigned | Unassigned | Unassigned | 52_0 | Clustered | 87 | VC_52_0 | 87 | 0.9858 | 1 | 0.9858 | 0 | 0 | 0 | 1 | Ref |
| 15228 | ON170022 | Unassigned | Unassigned | Unassigned | 826_0 | Clustered | 91 | VC_826_0 | 91 | 0.9935 | 1 | 0.9935 | 0 | 0 | 0 | 1 | Ref |
| 15229 | ON170023 | Unassigned | Unassigned | Unassigned | 52_0 | Clustered | 87 | VC_52_0 | 87 | 0.9858 | 1 | 0.9858 | 0 | 0 | 0 | 1 | Ref |
| 15230 | ON170024 | Unassigned | Unassigned | Unassigned | 357_0 | Clustered | 53 | VC_357_0 | 53 | 0.8979 | 1 | 0.8979 | 0 | 0 | 0 | 1 | Ref |
| 15231 | ON170025 | Unassigned | Unassigned | Unassigned | 53_0 | Clustered | 5 | VC_53_0 | 5 | 0.4977 | 0.99999808 | 0.4977 | 0 | 0 | 0 | 1 | Ref |
| 15232 | ON170026 | Unassigned | Unassigned | Unassigned | 1_0 | Clustered | 54 | VC_1_0 | 54 | 0.8527 | 1 | 0.8527 | 0 | 0 | 0 | 1 | Ref |
| 15233 | ON170027 | Unassigned | Unassigned | Unassigned | 357_0 | Clustered | 53 | VC_357_0 | 53 | 0.8979 | 1 | 0.8979 | 0 | 0 | 0 | 1 | Ref |
| 15234 | ON170028 | Unassigned | Unassigned | Unassigned | 826_0 | Clustered | 91 | VC_826_0 | 91 | 0.9935 | 1 | 0.9935 | 0 | 0 | 0 | 1 | Ref |
| 15235 | ON170029 | Unassigned | Unassigned | Unassigned | 52_0 | Clustered | 87 | VC_52_0 | 87 | 0.9858 | 1 | 0.9858 | 0 | 0 | 0 | 1 | Ref |
| 15236 | ON170030 | Unassigned | Unassigned | Unassigned | 52_0 | Clustered | 87 | VC_52_0 | 87 | 0.9858 | 1 | 0.9858 | 0 | 0 | 0 | 1 | Ref |
| 15237 | ON170031 | Unassigned | Unassigned | Unassigned | 357_0 | Clustered | 53 | VC_357_0 | 53 | 0.8979 | 1 | 0.8979 | 0 | 0 | 0 | 1 | Ref |
| 15238 | ON170032 | Unassigned | Unassigned | Unassigned | 1_0 | Clustered | 54 | VC_1_0 | 54 | 0.8527 | 1 | 0.8527 | 0 | 0 | 0 | 1 | Ref |
| 15239 | ON170033 | Unassigned | Unassigned | Unassigned | 826_0 | Clustered | 91 | VC_826_0 | 91 | 0.9935 | 1 | 0.9935 | 0 | 0 | 0 | 1 | Ref |
| 15240 | ON170034 | Unassigned | Unassigned | Unassigned | 52_0 | Clustered | 87 | VC_52_0 | 87 | 0.9858 | 1 | 0.9858 | 0 | 0 | 0 | 1 | Ref |
| 15241 | ON170036 | Unassigned | Unassigned | Unassigned | 826_0 | Clustered | 91 | VC_826_0 | 91 | 0.9935 | 1 | 0.9935 | 0 | 0 | 0 | 1 | Ref |
| 15242 | ON170037 | Unassigned | Unassigned | Unassigned | 52_0 | Clustered | 87 | VC_52_0 | 87 | 0.9858 | 1 | 0.9858 | 0 | 0 | 0 | 1 | Ref |
| 15243 | ON170038 | Unassigned | Unassigned | Unassigned | 826_0 | Clustered | 91 | VC_826_0 | 91 | 0.9935 | 1 | 0.9935 | 0 | 0 | 0 | 1 | Ref |
| 15244 | ON170040 | Unassigned | Unassigned | Unassigned | 826_0 | Clustered | 91 | VC_826_0 | 91 | 0.9935 | 1 | 0.9935 | 0 | 0 | 0 | 1 | Ref |
| 15245 | ON170041 | Unassigned | Unassigned | Unassigned | 52_0 | Clustered | 87 | VC_52_0 | 87 | 0.9858 | 1 | 0.9858 | 0 | 0 | 0 | 1 | Ref |
| 15246 | ON170042 | Unassigned | Unassigned | Unassigned | 25_0 | Clustered | 12 | VC_25_0 | 12 | 0.7945 | 1 | 0.7945 | 0 | 0 | 0 | 1 | Ref |
| 15247 | ON170049 | Unassigned | Unassigned | Unassigned | 826_0 | Clustered | 91 | VC_826_0 | 91 | 0.9935 | 1 | 0.9935 | 0 | 0 | 0 | 1 | Ref |
| 15248 | ON170050 | Unassigned | Unassigned | Unassigned | 52_0 | Clustered | 87 | VC_52_0 | 87 | 0.9858 | 1 | 0.9858 | 0 | 0 | 0 | 1 | Ref |
| 15249 | ON170051 | Unassigned | Unassigned | Unassigned | 25_0 | Clustered | 12 | VC_25_0 | 12 | 0.7945 | 1 | 0.7945 | 0 | 0 | 0 | 1 | Ref |
| 15250 | ON170052 | Unassigned | Unassigned | Unassigned | 357_0 | Clustered | 53 | VC_357_0 | 53 | 0.8979 | 1 | 0.8979 | 0 | 0 | 0 | 1 | Ref |
| 15251 | ON170053 | Unassigned | Unassigned | Unassigned | 1_0 | Clustered | 54 | VC_1_0 | 54 | 0.8527 | 1 | 0.8527 | 0 | 0 | 0 | 1 | Ref |
| 15252 | ON170054 | Unassigned | Unassigned | Unassigned | 826_0 | Clustered | 91 | VC_826_0 | 91 | 0.9935 | 1 | 0.9935 | 0 | 0 | 0 | 1 | Ref |
| 15253 | ON170055 | Unassigned | Unassigned | Unassigned | 52_0 | Clustered | 87 | VC_52_0 | 87 | 0.9858 | 1 | 0.9858 | 0 | 0 | 0 | 1 | Ref |
| 15254 | ON170056 | Unassigned | Unassigned | Unassigned | 25_0 | Clustered | 12 | VC_25_0 | 12 | 0.7945 | 1 | 0.7945 | 0 | 0 | 0 | 1 | Ref |
| 15255 | ON170057 | Unassigned | Unassigned | Unassigned | 357_0 | Clustered | 53 | VC_357_0 | 53 | 0.8979 | 1 | 0.8979 | 0 | 0 | 0 | 1 | Ref |
| 15256 | ON170058 | Unassigned | Unassigned | Unassigned | 357_0 | Clustered | 53 | VC_357_0 | 53 | 0.8979 | 1 | 0.8979 | 0 | 0 | 0 | 1 | Ref |
| 15257 | ON170059 | Unassigned | Unassigned | Unassigned | 1_0 | Clustered | 54 | VC_1_0 | 54 | 0.8527 | 1 | 0.8527 | 0 | 0 | 0 | 1 | Ref |
| 15258 | ON170060 | Unassigned | Unassigned | Unassigned | 25_0 | Clustered | 12 | VC_25_0 | 12 | 0.7945 | 1 | 0.7945 | 0 | 0 | 0 | 1 | Ref |
| 15259 | ON170061 | Unassigned | Unassigned | Unassigned | 1_0 | Clustered | 54 | VC_1_0 | 54 | 0.8527 | 1 | 0.8527 | 0 | 0 | 0 | 1 | Ref |
| 15260 | ON170062 | Unassigned | Unassigned | Unassigned | 249_0 | Clustered | 5 | VC_249_0 | 5 | 0.5515 | 0.9999999 | 0.5515 | 0 | 0 | 0 | 1 | Ref |
| 15261 | ON170066 | Unassigned | Unassigned | Unassigned | 25_0 | Clustered | 12 | VC_25_0 | 12 | 0.7945 | 1 | 0.7945 | 0 | 0 | 0 | 1 | Ref |
| 15262 | ON170067 | Unassigned | Unassigned | Unassigned | 1_0 | Clustered | 54 | VC_1_0 | 54 | 0.8527 | 1 | 0.8527 | 0 | 0 | 0 | 1 | Ref |
| 15263 | ON170068 | Unassigned | Unassigned | Unassigned | 357_0 | Clustered | 53 | VC_357_0 | 53 | 0.8979 | 1 | 0.8979 | 0 | 0 | 0 | 1 | Ref |
| 15264 | ON170069 | Unassigned | Unassigned | Unassigned | 53_0 | Clustered | 5 | VC_53_0 | 5 | 0.4977 | 0.99999808 | 0.4977 | 0 | 0 | 0 | 1 | Ref |
| 15265 | ON170070 | Unassigned | Unassigned | Unassigned | 1_0 | Clustered | 54 | VC_1_0 | 54 | 0.8527 | 1 | 0.8527 | 0 | 0 | 0 | 1 | Ref |
| 15266 | ON170071 | Unassigned | Unassigned | Unassigned | 357_0 | Clustered | 53 | VC_357_0 | 53 | 0.8979 | 1 | 0.8979 | 0 | 0 | 0 | 1 | Ref |
| 15267 | ON170072 | Unassigned | Unassigned | Unassigned | 25_0 | Clustered | 12 | VC_25_0 | 12 | 0.7945 | 1 | 0.7945 | 0 | 0 | 0 | 1 | Ref |
| 15268 | ON170073 | Unassigned | Unassigned | Unassigned | 1_0 | Clustered | 54 | VC_1_0 | 54 | 0.8527 | 1 | 0.8527 | 0 | 0 | 0 | 1 | Ref |
| 15269 | ON170074 | Unassigned | Unassigned | Unassigned | 826_0 | Clustered | 91 | VC_826_0 | 91 | 0.9935 | 1 | 0.9935 | 0 | 0 | 0 | 1 | Ref |
| 15270 | ON170075 | Unassigned | Unassigned | Unassigned | 357_0 | Clustered | 53 | VC_357_0 | 53 | 0.8979 | 1 | 0.8979 | 0 | 0 | 0 | 1 | Ref |
| 15271 | ON170076 | Unassigned | Unassigned | Unassigned | 1_0 | Clustered | 54 | VC_1_0 | 54 | 0.8527 | 1 | 0.8527 | 0 | 0 | 0 | 1 | Ref |
| 15272 | ON170077 | Unassigned | Unassigned | Unassigned | 357_0 | Clustered | 53 | VC_357_0 | 53 | 0.8979 | 1 | 0.8979 | 0 | 0 | 0 | 1 | Ref |
| 15273 | ON170078 | Unassigned | Unassigned | Unassigned | 1_0 | Clustered | 54 | VC_1_0 | 54 | 0.8527 | 1 | 0.8527 | 0 | 0 | 0 | 1 | Ref |
| 15274 | ON170079 | Unassigned | Unassigned | Unassigned | 357_0 | Clustered | 53 | VC_357_0 | 53 | 0.8979 | 1 | 0.8979 | 0 | 0 | 0 | 1 | Ref |
| 15275 | ON170080 | Unassigned | Unassigned | Unassigned | 1_0 | Clustered | 54 | VC_1_0 | 54 | 0.8527 | 1 | 0.8527 | 0 | 0 | 0 | 1 | Ref |
| 15276 | ON170081 | Unassigned | Unassigned | Unassigned | 1_0 | Clustered | 54 | VC_1_0 | 54 | 0.8527 | 1 | 0.8527 | 0 | 0 | 0 | 1 | Ref |
| 15277 | ON170082 | Unassigned | Unassigned | Unassigned | 1_0 | Clustered | 54 | VC_1_0 | 54 | 0.8527 | 1 | 0.8527 | 0 | 0 | 0 | 1 | Ref |
| 15278 | ON170084 | Unassigned | Unassigned | Unassigned | 1_0 | Clustered | 54 | VC_1_0 | 54 | 0.8527 | 1 | 0.8527 | 0 | 0 | 0 | 1 | Ref |
| 15279 | ON170086 | Unassigned | Unassigned | Unassigned | 826_0 | Clustered | 91 | VC_826_0 | 91 | 0.9935 | 1 | 0.9935 | 0 | 0 | 0 | 1 | Ref |
| 15280 | ON170087 | Unassigned | Unassigned | Unassigned | 52_0 | Clustered | 87 | VC_52_0 | 87 | 0.9858 | 1 | 0.9858 | 0 | 0 | 0 | 1 | Ref |
| 15281 | ON170088 | Unassigned | Unassigned | Unassigned | 826_0 | Clustered | 91 | VC_826_0 | 91 | 0.9935 | 1 | 0.9935 | 0 | 0 | 0 | 1 | Ref |
| 15282 | ON170089 | Unassigned | Unassigned | Unassigned | 52_0 | Clustered | 87 | VC_52_0 | 87 | 0.9858 | 1 | 0.9858 | 0 | 0 | 0 | 1 | Ref |
| 15283 | ON170090 | Unassigned | Unassigned | Unassigned | 826_0 | Clustered | 91 | VC_826_0 | 91 | 0.9935 | 1 | 0.9935 | 0 | 0 | 0 | 1 | Ref |
| 15284 | ON170091 | Unassigned | Unassigned | Unassigned | 52_0 | Clustered | 87 | VC_52_0 | 87 | 0.9858 | 1 | 0.9858 | 0 | 0 | 0 | 1 | Ref |
| 15285 | ON170092 | Unassigned | Unassigned | Unassigned | 826_0 | Clustered | 91 | VC_826_0 | 91 | 0.9935 | 1 | 0.9935 | 0 | 0 | 0 | 1 | Ref |
| 15286 | ON170093 | Unassigned | Unassigned | Unassigned | 52_0 | Clustered | 87 | VC_52_0 | 87 | 0.9858 | 1 | 0.9858 | 0 | 0 | 0 | 1 | Ref |
| 15287 | ON170094 | Unassigned | Unassigned | Unassigned | 826_0 | Clustered | 91 | VC_826_0 | 91 | 0.9935 | 1 | 0.9935 | 0 | 0 | 0 | 1 | Ref |
| 15288 | ON170095 | Unassigned | Unassigned | Unassigned | 52_0 | Clustered | 87 | VC_52_0 | 87 | 0.9858 | 1 | 0.9858 | 0 | 0 | 0 | 1 | Ref |
| 15289 | ON170096 | Unassigned | Unassigned | Unassigned | 826_0 | Clustered | 91 | VC_826_0 | 91 | 0.9935 | 1 | 0.9935 | 0 | 0 | 0 | 1 | Ref |
| 15290 | ON170097 | Unassigned | Unassigned | Unassigned | 52_0 | Clustered | 87 | VC_52_0 | 87 | 0.9858 | 1 | 0.9858 | 0 | 0 | 0 | 1 | Ref |
| 15291 | ON170098 | Unassigned | Unassigned | Unassigned | 826_0 | Clustered | 91 | VC_826_0 | 91 | 0.9935 | 1 | 0.9935 | 0 | 0 | 0 | 1 | Ref |
| 15292 | ON170100 | Unassigned | Unassigned | Unassigned | 826_0 | Clustered | 91 | VC_826_0 | 91 | 0.9935 | 1 | 0.9935 | 0 | 0 | 0 | 1 | Ref |
| 15293 | ON170101 | Unassigned | Unassigned | Unassigned | 52_0 | Clustered | 87 | VC_52_0 | 87 | 0.9858 | 1 | 0.9858 | 0 | 0 | 0 | 1 | Ref |
| 15294 | ON170102 | Unassigned | Unassigned | Unassigned | 826_0 | Clustered | 91 | VC_826_0 | 91 | 0.9935 | 1 | 0.9935 | 0 | 0 | 0 | 1 | Ref |
| 15295 | ON170103 | Unassigned | Unassigned | Unassigned | 52_0 | Clustered | 87 | VC_52_0 | 87 | 0.9858 | 1 | 0.9858 | 0 | 0 | 0 | 1 | Ref |
| 15296 | ON170104 | Unassigned | Unassigned | Unassigned | 826_0 | Clustered | 91 | VC_826_0 | 91 | 0.9935 | 1 | 0.9935 | 0 | 0 | 0 | 1 | Ref |
| 15297 | ON170105 | Unassigned | Unassigned | Unassigned | 52_0 | Clustered | 87 | VC_52_0 | 87 | 0.9858 | 1 | 0.9858 | 0 | 0 | 0 | 1 | Ref |
| 15298 | ON170106 | Unassigned | Unassigned | Unassigned | 826_0 | Clustered | 91 | VC_826_0 | 91 | 0.9935 | 1 | 0.9935 | 0 | 0 | 0 | 1 | Ref |
| 15299 | ON170107 | Unassigned | Unassigned | Unassigned | 52_0 | Clustered | 87 | VC_52_0 | 87 | 0.9858 | 1 | 0.9858 | 0 | 0 | 0 | 1 | Ref |
| 15300 | ON170108 | Unassigned | Unassigned | Unassigned | 826_0 | Clustered | 91 | VC_826_0 | 91 | 0.9935 | 1 | 0.9935 | 0 | 0 | 0 | 1 | Ref |
| 15301 | ON170109 | Unassigned | Unassigned | Unassigned | 52_0 | Clustered | 87 | VC_52_0 | 87 | 0.9858 | 1 | 0.9858 | 0 | 0 | 0 | 1 | Ref |
| 15302 | ON170110 | Unassigned | Unassigned | Unassigned | 826_0 | Clustered | 91 | VC_826_0 | 91 | 0.9935 | 1 | 0.9935 | 0 | 0 | 0 | 1 | Ref |
| 15303 | ON170111 | Unassigned | Unassigned | Unassigned | 52_0 | Clustered | 87 | VC_52_0 | 87 | 0.9858 | 1 | 0.9858 | 0 | 0 | 0 | 1 | Ref |
| 15304 | ON170112 | Unassigned | Unassigned | Unassigned | 826_0 | Clustered | 91 | VC_826_0 | 91 | 0.9935 | 1 | 0.9935 | 0 | 0 | 0 | 1 | Ref |
| 15305 | ON170113 | Unassigned | Unassigned | Unassigned | 52_0 | Clustered | 87 | VC_52_0 | 87 | 0.9858 | 1 | 0.9858 | 0 | 0 | 0 | 1 | Ref |
| 15306 | ON170114 | Unassigned | Unassigned | Unassigned | 826_0 | Clustered | 91 | VC_826_0 | 91 | 0.9935 | 1 | 0.9935 | 0 | 0 | 0 | 1 | Ref |
| 15307 | ON170115 | Unassigned | Unassigned | Unassigned | 52_0 | Clustered | 87 | VC_52_0 | 87 | 0.9858 | 1 | 0.9858 | 0 | 0 | 0 | 1 | Ref |
| 15308 | ON170116 | Unassigned | Unassigned | Unassigned | 826_0 | Clustered | 91 | VC_826_0 | 91 | 0.9935 | 1 | 0.9935 | 0 | 0 | 0 | 1 | Ref |
| 15309 | ON170117 | Unassigned | Unassigned | Unassigned | 52_0 | Clustered | 87 | VC_52_0 | 87 | 0.9858 | 1 | 0.9858 | 0 | 0 | 0 | 1 | Ref |
| 15310 | ON170118 | Unassigned | Unassigned | Unassigned | 826_0 | Clustered | 91 | VC_826_0 | 91 | 0.9935 | 1 | 0.9935 | 0 | 0 | 0 | 1 | Ref |
| 15311 | ON170119 | Unassigned | Unassigned | Unassigned | 52_0 | Clustered | 87 | VC_52_0 | 87 | 0.9858 | 1 | 0.9858 | 0 | 0 | 0 | 1 | Ref |
| 15312 | ON170120 | Unassigned | Unassigned | Unassigned | 826_0 | Clustered | 91 | VC_826_0 | 91 | 0.9935 | 1 | 0.9935 | 0 | 0 | 0 | 1 | Ref |
| 15313 | ON170121 | Unassigned | Unassigned | Unassigned | 52_0 | Clustered | 87 | VC_52_0 | 87 | 0.9858 | 1 | 0.9858 | 0 | 0 | 0 | 1 | Ref |
| 15314 | ON170122 | Unassigned | Unassigned | Unassigned | 826_0 | Clustered | 91 | VC_826_0 | 91 | 0.9935 | 1 | 0.9935 | 0 | 0 | 0 | 1 | Ref |
| 15315 | ON170124 | Unassigned | Unassigned | Unassigned | 826_0 | Clustered | 91 | VC_826_0 | 91 | 0.9935 | 1 | 0.9935 | 0 | 0 | 0 | 1 | Ref |
| 15316 | ON170125 | Unassigned | Unassigned | Unassigned | 52_0 | Clustered | 87 | VC_52_0 | 87 | 0.9858 | 1 | 0.9858 | 0 | 0 | 0 | 1 | Ref |
| 15317 | ON170126 | Unassigned | Unassigned | Unassigned | 826_0 | Clustered | 91 | VC_826_0 | 91 | 0.9935 | 1 | 0.9935 | 0 | 0 | 0 | 1 | Ref |
| 15318 | ON170127 | Unassigned | Unassigned | Unassigned | 52_0 | Clustered | 87 | VC_52_0 | 87 | 0.9858 | 1 | 0.9858 | 0 | 0 | 0 | 1 | Ref |
| 15319 | ON170128 | Unassigned | Unassigned | Unassigned | 1_0 | Clustered | 54 | VC_1_0 | 54 | 0.8527 | 1 | 0.8527 | 0 | 0 | 0 | 1 | Ref |
| 15320 | ON170129 | Unassigned | Unassigned | Unassigned | 357_0 | Clustered | 53 | VC_357_0 | 53 | 0.8979 | 1 | 0.8979 | 0 | 0 | 0 | 1 | Ref |
| 15321 | ON170130 | Unassigned | Unassigned | Unassigned | 1_0 | Clustered | 54 | VC_1_0 | 54 | 0.8527 | 1 | 0.8527 | 0 | 0 | 0 | 1 | Ref |
| 15322 | ON170131 | Unassigned | Unassigned | Unassigned | 1_0 | Clustered | 54 | VC_1_0 | 54 | 0.8527 | 1 | 0.8527 | 0 | 0 | 0 | 1 | Ref |
| 15323 | ON170132 | Unassigned | Unassigned | Unassigned | 52_0 | Clustered | 87 | VC_52_0 | 87 | 0.9858 | 1 | 0.9858 | 0 | 0 | 0 | 1 | Ref |
| 15324 | ON170133 | Unassigned | Unassigned | Unassigned | 357_0 | Clustered | 53 | VC_357_0 | 53 | 0.8979 | 1 | 0.8979 | 0 | 0 | 0 | 1 | Ref |
| 15325 | ON170134 | Unassigned | Unassigned | Unassigned | 357_0 | Clustered | 53 | VC_357_0 | 53 | 0.8979 | 1 | 0.8979 | 0 | 0 | 0 | 1 | Ref |
| 15326 | ON170135 | Unassigned | Unassigned | Unassigned | 1_0 | Clustered | 54 | VC_1_0 | 54 | 0.8527 | 1 | 0.8527 | 0 | 0 | 0 | 1 | Ref |
| 15327 | ON170136 | Unassigned | Unassigned | Unassigned | 357_0 | Clustered | 53 | VC_357_0 | 53 | 0.8979 | 1 | 0.8979 | 0 | 0 | 0 | 1 | Ref |
| 15328 | ON170139 | Unassigned | Unassigned | Unassigned | 53_0 | Clustered | 5 | VC_53_0 | 5 | 0.4977 | 0.99999808 | 0.4977 | 0 | 0 | 0 | 1 | Ref |
| 15329 | ON170142 | Unassigned | Unassigned | Unassigned | 826_0 | Clustered | 91 | VC_826_0 | 91 | 0.9935 | 1 | 0.9935 | 0 | 0 | 0 | 1 | Ref |
| 15330 | ON170143 | Unassigned | Unassigned | Unassigned | 52_0 | Clustered | 87 | VC_52_0 | 87 | 0.9858 | 1 | 0.9858 | 0 | 0 | 0 | 1 | Ref |
| 15331 | ON170144 | Unassigned | Unassigned | Unassigned | 826_0 | Clustered | 91 | VC_826_0 | 91 | 0.9935 | 1 | 0.9935 | 0 | 0 | 0 | 1 | Ref |
| 15332 | ON170145 | Unassigned | Unassigned | Unassigned | 52_0 | Clustered | 87 | VC_52_0 | 87 | 0.9858 | 1 | 0.9858 | 0 | 0 | 0 | 1 | Ref |
| 15333 | ON170146 | Unassigned | Unassigned | Unassigned | 826_0 | Clustered | 91 | VC_826_0 | 91 | 0.9935 | 1 | 0.9935 | 0 | 0 | 0 | 1 | Ref |
| 15334 | ON170147 | Unassigned | Unassigned | Unassigned | 52_0 | Clustered | 87 | VC_52_0 | 87 | 0.9858 | 1 | 0.9858 | 0 | 0 | 0 | 1 | Ref |
| 15335 | ON170148 | Unassigned | Unassigned | Unassigned | 180_0 | Clustered | 4 | VC_180_0 | 4 | 0.2056 | 0.9999862 | 0.2056 | 0 | 0 | 0 | 1 | Ref |
| 15336 | ON170149 | Unassigned | Unassigned | Unassigned | 358_0 | Clustered | 3 | VC_358_0 | 3 | 0.5146 | 0.97657291 | 0.5026 | 0 | 0 | 0 | 1 | Ref |
| 15337 | ON170150 | Unassigned | Unassigned | Unassigned | 357_0 | Clustered | 53 | VC_357_0 | 53 | 0.8979 | 1 | 0.8979 | 0 | 0 | 0 | 1 | Ref |
| 15338 | ON170151 | Unassigned | Unassigned | Unassigned | 1_0 | Clustered | 54 | VC_1_0 | 54 | 0.8527 | 1 | 0.8527 | 0 | 0 | 0 | 1 | Ref |
| 15339 | ON170152 | Unassigned | Unassigned | Unassigned | 826_0 | Clustered | 91 | VC_826_0 | 91 | 0.9935 | 1 | 0.9935 | 0 | 0 | 0 | 1 | Ref |
| 15340 | ON170153 | Unassigned | Unassigned | Unassigned | 52_0 | Clustered | 87 | VC_52_0 | 87 | 0.9858 | 1 | 0.9858 | 0 | 0 | 0 | 1 | Ref |
| 15341 | ON170154 | Unassigned | Unassigned | Unassigned | 826_0 | Clustered | 91 | VC_826_0 | 91 | 0.9935 | 1 | 0.9935 | 0 | 0 | 0 | 1 | Ref |
| 15342 | ON170155 | Unassigned | Unassigned | Unassigned | 52_0 | Clustered | 87 | VC_52_0 | 87 | 0.9858 | 1 | 0.9858 | 0 | 0 | 0 | 1 | Ref |
| 15343 | ON170156 | Unassigned | Unassigned | Unassigned | 826_0 | Clustered | 91 | VC_826_0 | 91 | 0.9935 | 1 | 0.9935 | 0 | 0 | 0 | 1 | Ref |
| 15344 | ON170157 | Unassigned | Unassigned | Unassigned | 52_0 | Clustered | 87 | VC_52_0 | 87 | 0.9858 | 1 | 0.9858 | 0 | 0 | 0 | 1 | Ref |
| 15345 | ON170158 | Unassigned | Unassigned | Unassigned | 826_0 | Clustered | 91 | VC_826_0 | 91 | 0.9935 | 1 | 0.9935 | 0 | 0 | 0 | 1 | Ref |
| 15346 | ON170159 | Unassigned | Unassigned | Unassigned | 52_0 | Clustered | 87 | VC_52_0 | 87 | 0.9858 | 1 | 0.9858 | 0 | 0 | 0 | 1 | Ref |
| 15347 | ON170160 | Unassigned | Unassigned | Unassigned | 826_0 | Clustered | 91 | VC_826_0 | 91 | 0.9935 | 1 | 0.9935 | 0 | 0 | 0 | 1 | Ref |
| 15348 | ON170161 | Unassigned | Unassigned | Unassigned | 52_0 | Clustered | 87 | VC_52_0 | 87 | 0.9858 | 1 | 0.9858 | 0 | 0 | 0 | 1 | Ref |
| 15349 | ON170162 | Unassigned | Unassigned | Unassigned | 826_0 | Clustered | 91 | VC_826_0 | 91 | 0.9935 | 1 | 0.9935 | 0 | 0 | 0 | 1 | Ref |
| 15350 | ON170163 | Unassigned | Unassigned | Unassigned | 52_0 | Clustered | 87 | VC_52_0 | 87 | 0.9858 | 1 | 0.9858 | 0 | 0 | 0 | 1 | Ref |
| 15351 | ON170164 | Unassigned | Unassigned | Unassigned | 826_0 | Clustered | 91 | VC_826_0 | 91 | 0.9935 | 1 | 0.9935 | 0 | 0 | 0 | 1 | Ref |
| 15352 | ON170165 | Unassigned | Unassigned | Unassigned | 52_0 | Clustered | 87 | VC_52_0 | 87 | 0.9858 | 1 | 0.9858 | 0 | 0 | 0 | 1 | Ref |
| 15353 | ON170166 | Unassigned | Unassigned | Unassigned | 25_0 | Clustered | 12 | VC_25_0 | 12 | 0.7945 | 1 | 0.7945 | 0 | 0 | 0 | 1 | Ref |
| 15354 | ON170167 | Unassigned | Unassigned | Unassigned | 826_0 | Clustered | 91 | VC_826_0 | 91 | 0.9935 | 1 | 0.9935 | 0 | 0 | 0 | 1 | Ref |
| 15355 | ON170168 | Unassigned | Unassigned | Unassigned | 52_0 | Clustered | 87 | VC_52_0 | 87 | 0.9858 | 1 | 0.9858 | 0 | 0 | 0 | 1 | Ref |
| 15356 | ON170169 | Unassigned | Unassigned | Unassigned | 25_0 | Clustered | 12 | VC_25_0 | 12 | 0.7945 | 1 | 0.7945 | 0 | 0 | 0 | 1 | Ref |
| 15357 | ON170170 | Unassigned | Unassigned | Unassigned | 180_0 | Clustered | 4 | VC_180_0 | 4 | 0.2056 | 0.9999862 | 0.2056 | 0 | 0 | 0 | 1 | Ref |
| 15358 | ON170171 | Unassigned | Unassigned | Unassigned | 358_0 | Clustered | 3 | VC_358_0 | 3 | 0.5146 | 0.97657291 | 0.5026 | 0 | 0 | 0 | 1 | Ref |
| 15359 | ON170172 | Unassigned | Unassigned | Unassigned | 357_0 | Clustered | 53 | VC_357_0 | 53 | 0.8979 | 1 | 0.8979 | 0 | 0 | 0 | 1 | Ref |
| 15360 | ON170173 | Unassigned | Unassigned | Unassigned | 1_0 | Clustered | 54 | VC_1_0 | 54 | 0.8527 | 1 | 0.8527 | 0 | 0 | 0 | 1 | Ref |
| 15361 | ON170174 | Unassigned | Unassigned | Unassigned | 70_0 | Clustered | 27 | VC_70_0 | 27 | 0.5981 | 1 | 0.5981 | 0 | 0 | 0 | 0.9886 | Ref |
| 15362 | ON170175 | Unassigned | Unassigned | Unassigned | 357_0 | Clustered | 53 | VC_357_0 | 53 | 0.8979 | 1 | 0.8979 | 0 | 0 | 0 | 1 | Ref |
| 15363 | ON170176 | Unassigned | Unassigned | Unassigned | 826_0 | Clustered | 91 | VC_826_0 | 91 | 0.9935 | 1 | 0.9935 | 0 | 0 | 0 | 1 | Ref |
| 15364 | ON170178 | Unassigned | Unassigned | Unassigned | 70_0 | Clustered | 27 | VC_70_0 | 27 | 0.5981 | 1 | 0.5981 | 0 | 0 | 0 | 0.9886 | Ref |
| 15365 | ON170179 | Unassigned | Unassigned | Unassigned | 357_0 | Clustered | 53 | VC_357_0 | 53 | 0.8979 | 1 | 0.8979 | 0 | 0 | 0 | 1 | Ref |
| 15366 | ON170180 | Unassigned | Unassigned | Unassigned | 826_0 | Clustered | 91 | VC_826_0 | 91 | 0.9935 | 1 | 0.9935 | 0 | 0 | 0 | 1 | Ref |
| 15367 | ON170181 | Unassigned | Unassigned | Unassigned | 357_0 | Clustered | 53 | VC_357_0 | 53 | 0.8979 | 1 | 0.8979 | 0 | 0 | 0 | 1 | Ref |
| 15368 | ON170182 | Unassigned | Unassigned | Unassigned | 357_0 | Clustered | 53 | VC_357_0 | 53 | 0.8979 | 1 | 0.8979 | 0 | 0 | 0 | 1 | Ref |
| 15369 | ON170184 | Unassigned | Unassigned | Unassigned | 70_0 | Clustered | 27 | VC_70_0 | 27 | 0.5981 | 1 | 0.5981 | 0 | 0 | 0 | 0.9886 | Ref |
| 15370 | ON170185 | Unassigned | Unassigned | Unassigned | 357_0 | Clustered | 53 | VC_357_0 | 53 | 0.8979 | 1 | 0.8979 | 0 | 0 | 0 | 1 | Ref |
| 15371 | ON170186 | Unassigned | Unassigned | Unassigned | 826_0 | Clustered | 91 | VC_826_0 | 91 | 0.9935 | 1 | 0.9935 | 0 | 0 | 0 | 1 | Ref |
| 15372 | ON170187 | Unassigned | Unassigned | Unassigned | 357_0 | Clustered | 53 | VC_357_0 | 53 | 0.8979 | 1 | 0.8979 | 0 | 0 | 0 | 1 | Ref |
| 15373 | ON170188 | Unassigned | Unassigned | Unassigned | 250_0 | Clustered | 4 | VC_250_0 | 4 | 0.2395 | 0.99998241 | 0.2395 | 0 | 0 | 0 | 1 | Ref |
| 15374 | ON170189 | Unassigned | Unassigned | Unassigned | 826_0 | Clustered | 91 | VC_826_0 | 91 | 0.9935 | 1 | 0.9935 | 0 | 0 | 0 | 1 | Ref |
| 15375 | ON170190 | Unassigned | Unassigned | Unassigned | 70_0 | Clustered | 27 | VC_70_0 | 27 | 0.5981 | 1 | 0.5981 | 0 | 0 | 0 | 0.9886 | Ref |
| 15376 | ON170192 | Unassigned | Unassigned | Unassigned | 826_0 | Clustered | 91 | VC_826_0 | 91 | 0.9935 | 1 | 0.9935 | 0 | 0 | 0 | 1 | Ref |
| 15377 | ON170193 | Unassigned | Unassigned | Unassigned | 52_0 | Clustered | 87 | VC_52_0 | 87 | 0.9858 | 1 | 0.9858 | 0 | 0 | 0 | 1 | Ref |
| 15378 | ON170194 | Unassigned | Unassigned | Unassigned | 826_0 | Clustered | 91 | VC_826_0 | 91 | 0.9935 | 1 | 0.9935 | 0 | 0 | 0 | 1 | Ref |
| 15379 | ON170195 | Unassigned | Unassigned | Unassigned | 52_0 | Clustered | 87 | VC_52_0 | 87 | 0.9858 | 1 | 0.9858 | 0 | 0 | 0 | 1 | Ref |
| 15380 | ON170196 | Unassigned | Unassigned | Unassigned | 826_0 | Clustered | 91 | VC_826_0 | 91 | 0.9935 | 1 | 0.9935 | 0 | 0 | 0 | 1 | Ref |
| 15381 | ON170197 | Unassigned | Unassigned | Unassigned | 52_0 | Clustered | 87 | VC_52_0 | 87 | 0.9858 | 1 | 0.9858 | 0 | 0 | 0 | 1 | Ref |
| 15382 | ON170198 | Unassigned | Unassigned | Unassigned | 826_0 | Clustered | 91 | VC_826_0 | 91 | 0.9935 | 1 | 0.9935 | 0 | 0 | 0 | 1 | Ref |
| 15383 | ON170199 | Unassigned | Unassigned | Unassigned | 52_0 | Clustered | 87 | VC_52_0 | 87 | 0.9858 | 1 | 0.9858 | 0 | 0 | 0 | 1 | Ref |
| 15384 | ON170200 | Unassigned | Unassigned | Unassigned | 826_0 | Clustered | 91 | VC_826_0 | 91 | 0.9935 | 1 | 0.9935 | 0 | 0 | 0 | 1 | Ref |
| 15385 | ON170201 | Unassigned | Unassigned | Unassigned | 52_0 | Clustered | 87 | VC_52_0 | 87 | 0.9858 | 1 | 0.9858 | 0 | 0 | 0 | 1 | Ref |
| 15386 | ON170202 | Unassigned | Unassigned | Unassigned | 826_0 | Clustered | 91 | VC_826_0 | 91 | 0.9935 | 1 | 0.9935 | 0 | 0 | 0 | 1 | Ref |
| 15387 | ON170203 | Unassigned | Unassigned | Unassigned | 52_0 | Clustered | 87 | VC_52_0 | 87 | 0.9858 | 1 | 0.9858 | 0 | 0 | 0 | 1 | Ref |
| 15388 | ON170204 | Unassigned | Unassigned | Unassigned | 826_0 | Clustered | 91 | VC_826_0 | 91 | 0.9935 | 1 | 0.9935 | 0 | 0 | 0 | 1 | Ref |

| | | | | | | | | | | | | | | | | | |
|---|---|---|---|---|---|---|---|---|---|---|---|---|---|---|---|---|---|
| 15389 | ON170205 | Unassigned | Unassigned | Unassigned | 52_0 | Clustered | 87 | VC_52_0 | 87 | 0.9858 | 1 | 0.9858 | 0 | 0 | 0 | 1 | Ref |
| 15390 | ON170206 | Unassigned | Unassigned | Unassigned | 826_0 | Clustered | 91 | VC_826_0 | 91 | 0.9935 | 1 | 0.9935 | 0 | 0 | 0 | 1 | Ref |
| 15391 | ON170207 | Unassigned | Unassigned | Unassigned | 52_0 | Clustered | 87 | VC_52_0 | 87 | 0.9858 | 1 | 0.9858 | 0 | 0 | 0 | 1 | Ref |
| 15392 | ON170208 | Unassigned | Unassigned | Unassigned | 357_0 | Clustered | 53 | VC_357_0 | 53 | 0.8979 | 1 | 0.8979 | 0 | 0 | 0 | 1 | Ref |
| 15393 | ON170209 | Unassigned | Unassigned | Unassigned | 249_0 | Clustered | 5 | VC_249_0 | 5 | 0.5515 | 0.9999999 | 0.5515 | 0 | 0 | 0 | 1 | Ref |
| 15394 | ON170210 | Unassigned | Unassigned | Unassigned | 1_0 | Clustered | 54 | VC_1_0 | 54 | 0.8527 | 1 | 0.8527 | 0 | 0 | 0 | 1 | Ref |
| 15395 | ON170211 | Unassigned | Unassigned | Unassigned | 826_0 | Clustered | 91 | VC_826_0 | 91 | 0.9935 | 1 | 0.9935 | 0 | 0 | 0 | 1 | Ref |
| 15396 | ON170212 | Unassigned | Unassigned | Unassigned | 52_0 | Clustered | 87 | VC_52_0 | 87 | 0.9858 | 1 | 0.9858 | 0 | 0 | 0 | 1 | Ref |
| 15397 | ON170213 | Unassigned | Unassigned | Unassigned | 1_0 | Clustered | 54 | VC_1_0 | 54 | 0.8527 | 1 | 0.8527 | 0 | 0 | 0 | 1 | Ref |
| 15398 | ON170214 | Unassigned | Unassigned | Unassigned | 357_0 | Clustered | 53 | VC_357_0 | 53 | 0.8979 | 1 | 0.8979 | 0 | 0 | 0 | 1 | Ref |
| 15399 | ON170215 | Unassigned | Unassigned | Unassigned | 826_0 | Clustered | 91 | VC_826_0 | 91 | 0.9935 | 1 | 0.9935 | 0 | 0 | 0 | 1 | Ref |
| 15400 | ON170216 | Unassigned | Unassigned | Unassigned | 52_0 | Clustered | 87 | VC_52_0 | 87 | 0.9858 | 1 | 0.9858 | 0 | 0 | 0 | 1 | Ref |
| 15401 | ON170217 | Unassigned | Unassigned | Unassigned | 826_0 | Clustered | 91 | VC_826_0 | 91 | 0.9935 | 1 | 0.9935 | 0 | 0 | 0 | 1 | Ref |
| 15402 | ON170218 | Unassigned | Unassigned | Unassigned | 52_0 | Clustered | 87 | VC_52_0 | 87 | 0.9858 | 1 | 0.9858 | 0 | 0 | 0 | 1 | Ref |
| 15403 | ON170219 | Unassigned | Unassigned | Unassigned | 826_0 | Clustered | 91 | VC_826_0 | 91 | 0.9935 | 1 | 0.9935 | 0 | 0 | 0 | 1 | Ref |
| 15404 | ON170220 | Unassigned | Unassigned | Unassigned | 52_0 | Clustered | 87 | VC_52_0 | 87 | 0.9858 | 1 | 0.9858 | 0 | 0 | 0 | 1 | Ref |
| 15405 | ON170221 | Unassigned | Unassigned | Unassigned | 826_0 | Clustered | 91 | VC_826_0 | 91 | 0.9935 | 1 | 0.9935 | 0 | 0 | 0 | 1 | Ref |
| 15406 | ON170222 | Unassigned | Unassigned | Unassigned | 826_0 | Clustered | 91 | VC_826_0 | 91 | 0.9935 | 1 | 0.9935 | 0 | 0 | 0 | 1 | Ref |
| 15407 | ON170223 | Unassigned | Unassigned | Unassigned | 52_0 | Clustered | 87 | VC_52_0 | 87 | 0.9858 | 1 | 0.9858 | 0 | 0 | 0 | 1 | Ref |
| 15408 | ON170225 | Unassigned | Unassigned | Unassigned | 826_0 | Clustered | 91 | VC_826_0 | 91 | 0.9935 | 1 | 0.9935 | 0 | 0 | 0 | 1 | Ref |
| 15409 | ON170226 | Unassigned | Unassigned | Unassigned | 52_0 | Clustered | 87 | VC_52_0 | 87 | 0.9858 | 1 | 0.9858 | 0 | 0 | 0 | 1 | Ref |
| 15410 | ON170227 | Unassigned | Unassigned | Unassigned | 1_0 | Clustered | 54 | VC_1_0 | 54 | 0.8527 | 1 | 0.8527 | 0 | 0 | 0 | 1 | Ref |
| 15411 | ON170228 | Unassigned | Unassigned | Unassigned | 357_0 | Clustered | 53 | VC_357_0 | 53 | 0.8979 | 1 | 0.8979 | 0 | 0 | 0 | 1 | Ref |
| 15412 | ON170229 | Unassigned | Unassigned | Unassigned | 357_0 | Clustered | 53 | VC_357_0 | 53 | 0.8979 | 1 | 0.8979 | 0 | 0 | 0 | 1 | Ref |
| 15413 | ON170230 | Unassigned | Unassigned | Unassigned | 1_0 | Clustered | 54 | VC_1_0 | 54 | 0.8527 | 1 | 0.8527 | 0 | 0 | 0 | 1 | Ref |
| 15414 | ON170231 | Unassigned | Unassigned | Unassigned | 249_0 | Clustered | 5 | VC_249_0 | 5 | 0.5515 | 0.9999999 | 0.5515 | 0 | 0 | 0 | 1 | Ref |
| 15415 | ON170232 | Unassigned | Unassigned | Unassigned | 357_0 | Clustered | 53 | VC_357_0 | 53 | 0.8979 | 1 | 0.8979 | 0 | 0 | 0 | 1 | Ref |
| 15416 | ON170233 | Unassigned | Unassigned | Unassigned | 1_0 | Clustered | 54 | VC_1_0 | 54 | 0.8527 | 1 | 0.8527 | 0 | 0 | 0 | 1 | Ref |
| 15417 | ON170234 | Unassigned | Unassigned | Unassigned | 357_0 | Clustered | 53 | VC_357_0 | 53 | 0.8979 | 1 | 0.8979 | 0 | 0 | 0 | 1 | Ref |
| 15418 | ON170235 | Unassigned | Unassigned | Unassigned | 1_0 | Clustered | 54 | VC_1_0 | 54 | 0.8527 | 1 | 0.8527 | 0 | 0 | 0 | 1 | Ref |
| 15419 | ON170236 | Unassigned | Unassigned | Unassigned | 357_0 | Clustered | 53 | VC_357_0 | 53 | 0.8979 | 1 | 0.8979 | 0 | 0 | 0 | 1 | Ref |
| 15420 | ON170237 | Unassigned | Unassigned | Unassigned | 1_0 | Clustered | 54 | VC_1_0 | 54 | 0.8527 | 1 | 0.8527 | 0 | 0 | 0 | 1 | Ref |
| 15421 | ON170238 | Unassigned | Unassigned | Unassigned | 357_0 | Clustered | 53 | VC_357_0 | 53 | 0.8979 | 1 | 0.8979 | 0 | 0 | 0 | 1 | Ref |
| 15422 | ON170239 | Unassigned | Unassigned | Unassigned | 1_0 | Clustered | 54 | VC_1_0 | 54 | 0.8527 | 1 | 0.8527 | 0 | 0 | 0 | 1 | Ref |
| 15423 | ON170240 | Unassigned | Unassigned | Unassigned | 357_0 | Clustered | 53 | VC_357_0 | 53 | 0.8979 | 1 | 0.8979 | 0 | 0 | 0 | 1 | Ref |
| 15424 | ON170241 | Unassigned | Unassigned | Unassigned | 1_0 | Clustered | 54 | VC_1_0 | 54 | 0.8527 | 1 | 0.8527 | 0 | 0 | 0 | 1 | Ref |
| 15425 | ON170242 | Unassigned | Unassigned | Unassigned | 357_0 | Clustered | 53 | VC_357_0 | 53 | 0.8979 | 1 | 0.8979 | 0 | 0 | 0 | 1 | Ref |
| 15426 | ON170243 | Unassigned | Unassigned | Unassigned | 1_0 | Clustered | 54 | VC_1_0 | 54 | 0.8527 | 1 | 0.8527 | 0 | 0 | 0 | 1 | Ref |
| 15427 | ON170244 | Unassigned | Unassigned | Unassigned | 826_0 | Clustered | 91 | VC_826_0 | 91 | 0.9935 | 1 | 0.9935 | 0 | 0 | 0 | 1 | Ref |
| 15428 | ON170245 | Unassigned | Unassigned | Unassigned | 52_0 | Clustered | 87 | VC_52_0 | 87 | 0.9858 | 1 | 0.9858 | 0 | 0 | 0 | 1 | Ref |
| 15429 | ON170246 | Unassigned | Unassigned | Unassigned | 826_0 | Clustered | 91 | VC_826_0 | 91 | 0.9935 | 1 | 0.9935 | 0 | 0 | 0 | 1 | Ref |
| 15430 | ON170247 | Unassigned | Unassigned | Unassigned | 52_0 | Clustered | 87 | VC_52_0 | 87 | 0.9858 | 1 | 0.9858 | 0 | 0 | 0 | 1 | Ref |
| 15431 | ON170248 | Unassigned | Unassigned | Unassigned | 826_0 | Clustered | 91 | VC_826_0 | 91 | 0.9935 | 1 | 0.9935 | 0 | 0 | 0 | 1 | Ref |
| 15432 | ON170249 | Unassigned | Unassigned | Unassigned | 52_0 | Clustered | 87 | VC_52_0 | 87 | 0.9858 | 1 | 0.9858 | 0 | 0 | 0 | 1 | Ref |
| 15433 | ON170250 | Unassigned | Unassigned | Unassigned | 826_0 | Clustered | 91 | VC_826_0 | 91 | 0.9935 | 1 | 0.9935 | 0 | 0 | 0 | 1 | Ref |
| 15434 | ON170251 | Unassigned | Unassigned | Unassigned | 52_0 | Clustered | 87 | VC_52_0 | 87 | 0.9858 | 1 | 0.9858 | 0 | 0 | 0 | 1 | Ref |
| 15435 | ON170252 | Unassigned | Unassigned | Unassigned | 826_0 | Clustered | 91 | VC_826_0 | 91 | 0.9935 | 1 | 0.9935 | 0 | 0 | 0 | 1 | Ref |
| 15436 | ON170253 | Unassigned | Unassigned | Unassigned | 52_0 | Clustered | 87 | VC_52_0 | 87 | 0.9858 | 1 | 0.9858 | 0 | 0 | 0 | 1 | Ref |
| 15437 | ON170254 | Unassigned | Unassigned | Unassigned | 357_0 | Clustered | 53 | VC_357_0 | 53 | 0.8979 | 1 | 0.8979 | 0 | 0 | 0 | 1 | Ref |
| 15438 | ON170257 | Unassigned | Unassigned | Unassigned | 357_0 | Clustered | 53 | VC_357_0 | 53 | 0.8979 | 1 | 0.8979 | 0 | 0 | 0 | 1 | Ref |
| 15439 | ON170258 | Unassigned | Unassigned | Unassigned | 1_0 | Clustered | 54 | VC_1_0 | 54 | 0.8527 | 1 | 0.8527 | 0 | 0 | 0 | 1 | Ref |
| 15440 | ON170261 | Unassigned | Unassigned | Unassigned | 1_0 | Clustered | 54 | VC_1_0 | 54 | 0.8527 | 1 | 0.8527 | 0 | 0 | 0 | 1 | Ref |
| 15441 | ON170262 | Unassigned | Unassigned | Unassigned | 357_0 | Clustered | 53 | VC_357_0 | 53 | 0.8979 | 1 | 0.8979 | 0 | 0 | 0 | 1 | Ref |
| 15442 | ON170263 | Unassigned | Unassigned | Unassigned | 1_0 | Clustered | 54 | VC_1_0 | 54 | 0.8527 | 1 | 0.8527 | 0 | 0 | 0 | 1 | Ref |
| 15443 | ON170264 | Unassigned | Unassigned | Unassigned | 357_0 | Clustered | 53 | VC_357_0 | 53 | 0.8979 | 1 | 0.8979 | 0 | 0 | 0 | 1 | Ref |
| 15444 | ON170265 | Unassigned | Unassigned | Unassigned | 357_0 | Clustered | 53 | VC_357_0 | 53 | 0.8979 | 1 | 0.8979 | 0 | 0 | 0 | 1 | Ref |
| 15445 | ON170266 | Unassigned | Unassigned | Unassigned | 1_0 | Clustered | 54 | VC_1_0 | 54 | 0.8527 | 1 | 0.8527 | 0 | 0 | 0 | 1 | Ref |
| 15446 | ON170267 | Unassigned | Unassigned | Unassigned | 357_0 | Clustered | 53 | VC_357_0 | 53 | 0.8979 | 1 | 0.8979 | 0 | 0 | 0 | 1 | Ref |
| 15447 | ON170268 | Unassigned | Unassigned | Unassigned | 1_0 | Clustered | 54 | VC_1_0 | 54 | 0.8527 | 1 | 0.8527 | 0 | 0 | 0 | 1 | Ref |
| 15448 | ON170270 | Unassigned | Unassigned | Unassigned | 1_0 | Clustered | 54 | VC_1_0 | 54 | 0.8527 | 1 | 0.8527 | 0 | 0 | 0 | 1 | Ref |
| 15449 | ON170271 | Unassigned | Unassigned | Unassigned | 826_0 | Clustered | 91 | VC_826_0 | 91 | 0.9935 | 1 | 0.9935 | 0 | 0 | 0 | 1 | Ref |
| 15450 | ON170272 | Unassigned | Unassigned | Unassigned | 1_0 | Clustered | 54 | VC_1_0 | 54 | 0.8527 | 1 | 0.8527 | 0 | 0 | 0 | 1 | Ref |
| 15451 | ON170273 | Unassigned | Unassigned | Unassigned | 52_0 | Clustered | 87 | VC_52_0 | 87 | 0.9858 | 1 | 0.9858 | 0 | 0 | 0 | 1 | Ref |
| 15452 | ON170274 | Unassigned | Unassigned | Unassigned | 25_0 | Clustered | 12 | VC_25_0 | 12 | 0.7945 | 1 | 0.7945 | 0 | 0 | 0 | 1 | Ref |
| 15453 | ON170275 | Unassigned | Unassigned | Unassigned | 357_0 | Clustered | 53 | VC_357_0 | 53 | 0.8979 | 1 | 0.8979 | 0 | 0 | 0 | 1 | Ref |
| 15454 | ON170276 | Unassigned | Unassigned | Unassigned | 1_0 | Clustered | 54 | VC_1_0 | 54 | 0.8527 | 1 | 0.8527 | 0 | 0 | 0 | 1 | Ref |
| 15455 | ON170277 | Unassigned | Unassigned | Unassigned | 52_0 | Clustered | 87 | VC_52_0 | 87 | 0.9858 | 1 | 0.9858 | 0 | 0 | 0 | 1 | Ref |
| 15456 | ON170278 | Unassigned | Unassigned | Unassigned | 25_0 | Clustered | 12 | VC_25_0 | 12 | 0.7945 | 1 | 0.7945 | 0 | 0 | 0 | 1 | Ref |
| 15457 | ON170279 | Unassigned | Unassigned | Unassigned | 357_0 | Clustered | 53 | VC_357_0 | 53 | 0.8979 | 1 | 0.8979 | 0 | 0 | 0 | 1 | Ref |
| 15458 | ON170280 | Unassigned | Unassigned | Unassigned | 826_0 | Clustered | 91 | VC_826_0 | 91 | 0.9935 | 1 | 0.9935 | 0 | 0 | 0 | 1 | Ref |
| 15459 | ON170281 | Unassigned | Unassigned | Unassigned | 52_0 | Clustered | 87 | VC_52_0 | 87 | 0.9858 | 1 | 0.9858 | 0 | 0 | 0 | 1 | Ref |
| 15460 | ON170282 | Unassigned | Unassigned | Unassigned | 826_0 | Clustered | 91 | VC_826_0 | 91 | 0.9935 | 1 | 0.9935 | 0 | 0 | 0 | 1 | Ref |
| 15461 | ON170283 | Unassigned | Unassigned | Unassigned | 52_0 | Clustered | 87 | VC_52_0 | 87 | 0.9858 | 1 | 0.9858 | 0 | 0 | 0 | 1 | Ref |
| 15462 | ON170284 | Unassigned | Unassigned | Unassigned | 826_0 | Clustered | 91 | VC_826_0 | 91 | 0.9935 | 1 | 0.9935 | 0 | 0 | 0 | 1 | Ref |
| 15463 | ON170285 | Unassigned | Unassigned | Unassigned | 52_0 | Clustered | 87 | VC_52_0 | 87 | 0.9858 | 1 | 0.9858 | 0 | 0 | 0 | 1 | Ref |
| 15464 | ON170286 | Unassigned | Unassigned | Unassigned | 357_0 | Clustered | 53 | VC_357_0 | 53 | 0.8979 | 1 | 0.8979 | 0 | 0 | 0 | 1 | Ref |
| 15465 | ON170287 | Unassigned | Unassigned | Unassigned | 357_0 | Clustered | 53 | VC_357_0 | 53 | 0.8979 | 1 | 0.8979 | 0 | 0 | 0 | 1 | Ref |
| 15466 | ON170289 | Unassigned | Unassigned | Unassigned | 826_0 | Clustered | 91 | VC_826_0 | 91 | 0.9935 | 1 | 0.9935 | 0 | 0 | 0 | 1 | Ref |
| 15467 | ON170290 | Unassigned | Unassigned | Unassigned | 52_0 | Clustered | 87 | VC_52_0 | 87 | 0.9858 | 1 | 0.9858 | 0 | 0 | 0 | 1 | Ref |
| 15468 | ON170291 | Unassigned | Unassigned | Unassigned | 826_0 | Clustered | 91 | VC_826_0 | 91 | 0.9935 | 1 | 0.9935 | 0 | 0 | 0 | 1 | Ref |
| 15469 | ON170292 | Unassigned | Unassigned | Unassigned | 52_0 | Clustered | 87 | VC_52_0 | 87 | 0.9858 | 1 | 0.9858 | 0 | 0 | 0 | 1 | Ref |
| 15470 | ON170293 | Unassigned | Unassigned | Unassigned | 826_0 | Clustered | 91 | VC_826_0 | 91 | 0.9935 | 1 | 0.9935 | 0 | 0 | 0 | 1 | Ref |
| 15471 | ON170294 | Unassigned | Unassigned | Unassigned | 52_0 | Clustered | 87 | VC_52_0 | 87 | 0.9858 | 1 | 0.9858 | 0 | 0 | 0 | 1 | Ref |
| 15472 | ON170295 | Unassigned | Unassigned | Unassigned | 826_0 | Clustered | 91 | VC_826_0 | 91 | 0.9935 | 1 | 0.9935 | 0 | 0 | 0 | 1 | Ref |
| 15473 | ON170296 | Unassigned | Unassigned | Unassigned | 52_0 | Clustered | 87 | VC_52_0 | 87 | 0.9858 | 1 | 0.9858 | 0 | 0 | 0 | 1 | Ref |
| 15474 | ON170297 | Unassigned | Unassigned | Unassigned | 826_0 | Clustered | 91 | VC_826_0 | 91 | 0.9935 | 1 | 0.9935 | 0 | 0 | 0 | 1 | Ref |
| 15475 | ON170298 | Unassigned | Unassigned | Unassigned | 826_0 | Clustered | 91 | VC_826_0 | 91 | 0.9935 | 1 | 0.9935 | 0 | 0 | 0 | 1 | Ref |
| 15476 | ON170299 | Unassigned | Unassigned | Unassigned | 52_0 | Clustered | 87 | VC_52_0 | 87 | 0.9858 | 1 | 0.9858 | 0 | 0 | 0 | 1 | Ref |
| 15477 | ON170300 | Unassigned | Unassigned | Unassigned | 826_0 | Clustered | 91 | VC_826_0 | 91 | 0.9935 | 1 | 0.9935 | 0 | 0 | 0 | 1 | Ref |
| 15478 | ON170301 | Unassigned | Unassigned | Unassigned | 52_0 | Clustered | 87 | VC_52_0 | 87 | 0.9858 | 1 | 0.9858 | 0 | 0 | 0 | 1 | Ref |
| 15479 | ON170302 | Unassigned | Unassigned | Unassigned | 249_0 | Clustered | 5 | VC_249_0 | 5 | 0.5515 | 0.9999999 | 0.5515 | 0 | 0 | 0 | 1 | Ref |
| 15480 | ON170303 | Unassigned | Unassigned | Unassigned | 357_0 | Clustered | 53 | VC_357_0 | 53 | 0.8979 | 1 | 0.8979 | 0 | 0 | 0 | 1 | Ref |
| 15481 | ON170304 | Unassigned | Unassigned | Unassigned | 1_0 | Clustered | 54 | VC_1_0 | 54 | 0.8527 | 1 | 0.8527 | 0 | 0 | 0 | 1 | Ref |
| 15482 | ON170305 | Unassigned | Unassigned | Unassigned | 1_0 | Clustered | 54 | VC_1_0 | 54 | 0.8527 | 1 | 0.8527 | 0 | 0 | 0 | 1 | Ref |
| 15483 | ON170306 | Unassigned | Unassigned | Unassigned | 826_0 | Clustered | 91 | VC_826_0 | 91 | 0.9935 | 1 | 0.9935 | 0 | 0 | 0 | 1 | Ref |
| 15484 | ON170307 | Unassigned | Unassigned | Unassigned | 826_0 | Clustered | 91 | VC_826_0 | 91 | 0.9935 | 1 | 0.9935 | 0 | 0 | 0 | 1 | Ref |
| 15485 | ON170308 | Unassigned | Unassigned | Unassigned | 52_0 | Clustered | 87 | VC_52_0 | 87 | 0.9858 | 1 | 0.9858 | 0 | 0 | 0 | 1 | Ref |
| 15486 | ON170311 | Unassigned | Unassigned | Unassigned | 826_0 | Clustered | 91 | VC_826_0 | 91 | 0.9935 | 1 | 0.9935 | 0 | 0 | 0 | 1 | Ref |
| 15487 | ON170312 | Unassigned | Unassigned | Unassigned | 52_0 | Clustered | 87 | VC_52_0 | 87 | 0.9858 | 1 | 0.9858 | 0 | 0 | 0 | 1 | Ref |
| 15488 | ON170313 | Unassigned | Unassigned | Unassigned | 826_0 | Clustered | 91 | VC_826_0 | 91 | 0.9935 | 1 | 0.9935 | 0 | 0 | 0 | 1 | Ref |
| 15489 | ON170314 | Unassigned | Unassigned | Unassigned | 52_0 | Clustered | 87 | VC_52_0 | 87 | 0.9858 | 1 | 0.9858 | 0 | 0 | 0 | 1 | Ref |
| 15490 | ON170315 | Unassigned | Unassigned | Unassigned | 826_0 | Clustered | 91 | VC_826_0 | 91 | 0.9935 | 1 | 0.9935 | 0 | 0 | 0 | 1 | Ref |
| 15491 | ON170316 | Unassigned | Unassigned | Unassigned | 52_0 | Clustered | 87 | VC_52_0 | 87 | 0.9858 | 1 | 0.9858 | 0 | 0 | 0 | 1 | Ref |
| 15492 | ON170317 | Unassigned | Unassigned | Unassigned | 357_0 | Clustered | 53 | VC_357_0 | 53 | 0.8979 | 1 | 0.8979 | 0 | 0 | 0 | 1 | Ref |
| 15493 | ON170318 | Unassigned | Unassigned | Unassigned | 70_0 | Clustered | 27 | VC_70_0 | 27 | 0.5981 | 1 | 0.5981 | 0 | 0 | 0 | 0.9886 | Ref |
| 15494 | ON170320 | Unassigned | Unassigned | Unassigned | 53_0 | Clustered | 5 | VC_53_0 | 5 | 0.4977 | 0.99999808 | 0.4977 | 0 | 0 | 0 | 1 | Ref |
| 15495 | ON170321 | Unassigned | Unassigned | Unassigned | 1_0 | Clustered | 54 | VC_1_0 | 54 | 0.8527 | 1 | 0.8527 | 0 | 0 | 0 | 1 | Ref |
| 15496 | ON170322 | Unassigned | Unassigned | Unassigned | 358_0 | Clustered | 3 | VC_358_0 | 3 | 0.5146 | 0.97657291 | 0.5026 | 0 | 0 | 0 | 1 | Ref |
| 15497 | ON170323 | Unassigned | Unassigned | Unassigned | 357_0 | Clustered | 53 | VC_357_0 | 53 | 0.8979 | 1 | 0.8979 | 0 | 0 | 0 | 1 | Ref |
| 15498 | ON170324 | Unassigned | Unassigned | Unassigned | 357_0 | Clustered | 53 | VC_357_0 | 53 | 0.8979 | 1 | 0.8979 | 0 | 0 | 0 | 1 | Ref |
| 15499 | ON170325 | Unassigned | Unassigned | Unassigned | 1_0 | Clustered | 54 | VC_1_0 | 54 | 0.8527 | 1 | 0.8527 | 0 | 0 | 0 | 1 | Ref |
| 15500 | ON170326 | Unassigned | Unassigned | Unassigned | 357_0 | Clustered | 53 | VC_357_0 | 53 | 0.8979 | 1 | 0.8979 | 0 | 0 | 0 | 1 | Ref |
| 15501 | ON170327 | Unassigned | Unassigned | Unassigned | 357_0 | Clustered | 53 | VC_357_0 | 53 | 0.8979 | 1 | 0.8979 | 0 | 0 | 0 | 1 | Ref |
| 15502 | ON170328 | Unassigned | Unassigned | Unassigned | 249_0 | Clustered | 5 | VC_249_0 | 5 | 0.5515 | 0.9999999 | 0.5515 | 0 | 0 | 0 | 1 | Ref |
| 15503 | ON170329 | Unassigned | Unassigned | Unassigned | 1_0 | Clustered | 54 | VC_1_0 | 54 | 0.8527 | 1 | 0.8527 | 0 | 0 | 0 | 1 | Ref |
| 15504 | ON170330 | Unassigned | Unassigned | Unassigned | 826_0 | Clustered | 91 | VC_826_0 | 91 | 0.9935 | 1 | 0.9935 | 0 | 0 | 0 | 1 | Ref |
| 15505 | ON170331 | Unassigned | Unassigned | Unassigned | 52_0 | Clustered | 87 | VC_52_0 | 87 | 0.9858 | 1 | 0.9858 | 0 | 0 | 0 | 1 | Ref |
| 15506 | ON170332 | Unassigned | Unassigned | Unassigned | 826_0 | Clustered | 91 | VC_826_0 | 91 | 0.9935 | 1 | 0.9935 | 0 | 0 | 0 | 1 | Ref |
| 15507 | ON170333 | Unassigned | Unassigned | Unassigned | 52_0 | Clustered | 87 | VC_52_0 | 87 | 0.9858 | 1 | 0.9858 | 0 | 0 | 0 | 1 | Ref |
| 15508 | ON170334 | Unassigned | Unassigned | Unassigned | 826_0 | Clustered | 91 | VC_826_0 | 91 | 0.9935 | 1 | 0.9935 | 0 | 0 | 0 | 1 | Ref |
| 15509 | ON170335 | Unassigned | Unassigned | Unassigned | 52_0 | Clustered | 87 | VC_52_0 | 87 | 0.9858 | 1 | 0.9858 | 0 | 0 | 0 | 1 | Ref |
| 15510 | ON170336 | Unassigned | Unassigned | Unassigned | 826_0 | Clustered | 91 | VC_826_0 | 91 | 0.9935 | 1 | 0.9935 | 0 | 0 | 0 | 1 | Ref |
| 15511 | ON170337 | Unassigned | Unassigned | Unassigned | 52_0 | Clustered | 87 | VC_52_0 | 87 | 0.9858 | 1 | 0.9858 | 0 | 0 | 0 | 1 | Ref |
| 15512 | ON170338 | Unassigned | Unassigned | Unassigned | 826_0 | Clustered | 91 | VC_826_0 | 91 | 0.9935 | 1 | 0.9935 | 0 | 0 | 0 | 1 | Ref |
| 15513 | ON170339 | Unassigned | Unassigned | Unassigned | 52_0 | Clustered | 87 | VC_52_0 | 87 | 0.9858 | 1 | 0.9858 | 0 | 0 | 0 | 1 | Ref |
| 15514 | ON170340 | Unassigned | Unassigned | Unassigned | 826_0 | Clustered | 91 | VC_826_0 | 91 | 0.9935 | 1 | 0.9935 | 0 | 0 | 0 | 1 | Ref |
| 15515 | ON170341 | Unassigned | Unassigned | Unassigned | 52_0 | Clustered | 87 | VC_52_0 | 87 | 0.9858 | 1 | 0.9858 | 0 | 0 | 0 | 1 | Ref |
| 15516 | ON170343 | Unassigned | Unassigned | Unassigned | 357_0 | Clustered | 53 | VC_357_0 | 53 | 0.8979 | 1 | 0.8979 | 0 | 0 | 0 | 1 | Ref |
| 15517 | ON170344 | Unassigned | Unassigned | Unassigned | 1_0 | Clustered | 54 | VC_1_0 | 54 | 0.8527 | 1 | 0.8527 | 0 | 0 | 0 | 1 | Ref |
| 15518 | ON170347 | Unassigned | Unassigned | Unassigned | 1_0 | Clustered | 54 | VC_1_0 | 54 | 0.8527 | 1 | 0.8527 | 0 | 0 | 0 | 1 | Ref |
| 15519 | ON170348 | Unassigned | Unassigned | Unassigned | 1_0 | Clustered | 54 | VC_1_0 | 54 | 0.8527 | 1 | 0.8527 | 0 | 0 | 0 | 1 | Ref |
| 15520 | ON170351 | Unassigned | Unassigned | Unassigned | 1_0 | Clustered | 54 | VC_1_0 | 54 | 0.8527 | 1 | 0.8527 | 0 | 0 | 0 | 1 | Ref |
| 15521 | ON170352 | Unassigned | Unassigned | Unassigned | 826_0 | Clustered | 91 | VC_826_0 | 91 | 0.9935 | 1 | 0.9935 | 0 | 0 | 0 | 1 | Ref |
| 15522 | ON170353 | Unassigned | Unassigned | Unassigned | 52_0 | Clustered | 87 | VC_52_0 | 87 | 0.9858 | 1 | 0.9858 | 0 | 0 | 0 | 1 | Ref |
| 15523 | ON170354 | Unassigned | Unassigned | Unassigned | 180_0 | Clustered | 4 | VC_180_0 | 4 | 0.2056 | 0.9999862 | 0.2056 | 0 | 0 | 0 | 1 | Ref |
| 15524 | ON170355 | Unassigned | Unassigned | Unassigned | 357_0 | Clustered | 53 | VC_357_0 | 53 | 0.8979 | 1 | 0.8979 | 0 | 0 | 0 | 1 | Ref |
| 15525 | ON170356 | Unassigned | Unassigned | Unassigned | 826_0 | Clustered | 91 | VC_826_0 | 91 | 0.9935 | 1 | 0.9935 | 0 | 0 | 0 | 1 | Ref |
| 15526 | ON170357 | Unassigned | Unassigned | Unassigned | 25_0 | Clustered | 12 | VC_25_0 | 12 | 0.7945 | 1 | 0.7945 | 0 | 0 | 0 | 1 | Ref |
| 15527 | ON170358 | Unassigned | Unassigned | Unassigned | 70_0 | Clustered | 27 | VC_70_0 | 27 | 0.5981 | 1 | 0.5981 | 0 | 0 | 0 | 0.9886 | Ref |
| 15528 | ON170359 | Unassigned | Unassigned | Unassigned | 357_0 | Clustered | 53 | VC_357_0 | 53 | 0.8979 | 1 | 0.8979 | 0 | 0 | 0 | 1 | Ref |
| 15529 | ON170360 | Unassigned | Unassigned | Unassigned | 826_0 | Clustered | 91 | VC_826_0 | 91 | 0.9935 | 1 | 0.9935 | 0 | 0 | 0 | 1 | Ref |
| 15530 | ON184124 | Unassigned | Unassigned | Unassigned | 835_0 | Clustered | 67 | VC_835_0 | 67 | 0.9624 | 1 | 0.9624 | 0 | 0 | 0 | 1 | Ref |
| 15531 | ON184125 | Unassigned | Unassigned | Unassigned | 131_1 | Clustered | 6 | VC_131_1 | 6 | 0.057 | 0.99999999 | 0.057 | 0 | 0 | 0 | 1 | Ref |
| 15532 | ON184126 | Unassigned | Unassigned | Unassigned | 131_1 | Clustered | 6 | VC_131_1 | 6 | 0.057 | 0.99999999 | 0.057 | 0 | 0 | 0 | 1 | Ref |
| 15533 | ON185577 | Unassigned | Unassigned | Unassigned | 575_0 | Clustered | 35 | VC_575_0 | 35 | 0.8815 | 1 | 0.8815 | 0 | 0 | 0 | 1 | Ref |
| 15534 | ON185580 | Unassigned | Unassigned | Unassigned | 452_0 | Clustered | 66 | VC_452_0 | 66 | 0.9248 | 1 | 0.9248 | 0 | 0 | 0 | 1 | Ref |
| 15535 | ON185581 | Unassigned | Unassigned | Unassigned | 190_28 | Clustered | 5 | VC_190_28 | 5 | 0.0304 | 0.99999994 | 0.0304 | 0 | 0 | 0 | 1 | Ref |
| 15536 | ON185582 | Unassigned | Unassigned | Unassigned | 190_28 | Clustered | 5 | VC_190_28 | 5 | 0.0304 | 0.99999994 | 0.0304 | 0 | 0 | 0 | 1 | Ref |
| 15537 | ON185583 | Unassigned | Unassigned | Unassigned | 190_28 | Clustered | 5 | VC_190_28 | 5 | 0.0304 | 0.99999994 | 0.0304 | 0 | 0 | 0 | 1 | Ref |
| 15538 | ON185584 | Unassigned | Unassigned | Unassigned | 190_28 | Clustered | 5 | VC_190_28 | 5 | 0.0304 | 0.99999994 | 0.0304 | 0 | 0 | 0 | 1 | Ref |
| 15539 | ON185585 | Unassigned | Unassigned | Unassigned | 190_24 | Clustered | 5 | VC_190_24 | 5 | 0.0293 | 0.9999996 | 0.0293 | 0 | 0 | 0 | 0.7 | Ref |
| 15540 | ON185586 | Unassigned | Unassigned | Unassigned | 942_0 | Clustered | 36 | VC_942_0 | 36 | 0.7848 | 1 | 0.7848 | 0 | 0 | 0 | 1 | Ref |
| 15541 | ON185587 | Unassigned | Unassigned | Unassigned | 942_0 | Clustered | 36 | VC_942_0 | 36 | 0.7848 | 1 | 0.7848 | 0 | 0 | 0 | 1 | Ref |
| 15542 | ON185588 | Unassigned | Unassigned | Unassigned | 942_0 | Clustered | 36 | VC_942_0 | 36 | 0.7848 | 1 | 0.7848 | 0 | 0 | 0 | 1 | Ref |
| 15543 | ON185589 | Unassigned | Unassigned | Unassigned | 941_0 | Clustered | 40 | VC_941_0 | 40 | 0.6903 | 1 | 0.6903 | 0 | 0 | 0 | 1 | Ref |
| 15544 | ON189042 | Unassigned | Unassigned | Unassigned | 749_0 | Clustered | 40 | VC_749_0 | 40 | 0.7517 | 1 | 0.7517 | 0 | 0 | 0 | 1 | Ref |
| 15545 | ON189043 | Unassigned | Unassigned | Unassigned | 749_0 | Clustered | 40 | VC_749_0 | 40 | 0.7517 | 1 | 0.7517 | 0 | 0 | 0 | 1 | Ref |
| 15546 | ON189044 | Unassigned | Unassigned | Unassigned | 749_0 | Clustered | 40 | VC_749_0 | 40 | 0.7517 | 1 | 0.7517 | 0 | 0 | 0 | 1 | Ref |
| 15547 | ON189045 | Unassigned | Unassigned | Unassigned | 169_0 | Clustered | 20 | VC_169_0 | 20 | 0.7529 | 1 | 0.7529 | 0 | 0 | 0 | 1 | Ref |
| 15548 | ON189046 | Unassigned | Unassigned | Unassigned | 129_0 | Clustered | 43 | VC_129_0 | 43 | 0.886 | 1 | 0.886 | 0 | 0 | 0 | 1 | Ref |
| 15549 | ON189047 | Unassigned | Unassigned | Unassigned | 169_0 | Clustered | 20 | VC_169_0 | 20 | 0.7529 | 1 | 0.7529 | 0 | 0 | 0 | 1 | Ref |
| 15550 | ON189048 | Unassigned | Unassigned | Unassigned | 172_0 | Clustered | 5 | VC_172_0 | 5 | 0.3579 | 0.99999997 | 0.3579 | 0 | 0 | 0 | 1 | Ref |

| | | | | | | | | | | | | | | | | | |
|---|---|---|---|---|---|---|---|---|---|---|---|---|---|---|---|---|---|
| 15551 | ON190024 | Unassigned | Unassigned | Unassigned | 531_1 | Clustered | 6 | VC_531_1 | 6 | 0.1481 | 0.99978665 | 0.148 | 0 | 0 | 0 | 1 | Ref |
| 15552 | ON190025 | Unassigned | Unassigned | Unassigned | 531_0 | Clustered | 9 | VC_531_0 | 9 | 0.26 | 1 | 0.26 | 0 | 0 | 0 | 1 | Ref |
| 15553 | ON202820 | Unassigned | Unassigned | Unassigned | 132_1 | Clustered | 17 | VC_132_1 | 17 | 0.1738 | 1 | 0.1738 | 0 | 0 | 0 | 0.9853 | Ref |
| 15554 | ON208829 | Unassigned | Unassigned | Unassigned | 325_57 | Clustered/Singleton | 1 | VC_325_57 | 1 | 0 | 1 | 0 | 0 | 0 | 0 | 1 | Ref |
| 15555 | ON208830 | Unassigned | Unassigned | Unassigned | 1072_3 | Clustered | 6 | VC_1072_3 | 6 | 0.1304 | 0.99999994 | 0.1304 | 0 | 0 | 0 | 0.8667 | Ref |
| 15556 | ON208832 | Unassigned | Unassigned | Unassigned | 982_0 | Clustered | 62 | VC_982_0 | 62 | 0.8646 | 1 | 0.8646 | 0 | 0 | 0 | 0.743 | Ref |
| 15557 | ON208833 | Unassigned | Unassigned | Unassigned | 1207_0 | Clustered | 29 | VC_1207_0 | 29 | 0.8225 | 1 | 0.8225 | 0 | 0 | 0 | 1 | Ref |
| 15558 | ON210136 | Unassigned | Unassigned | Unassigned | 835_0 | Clustered | 67 | VC_835_0 | 67 | 0.9624 | 1 | 0.9624 | 0 | 0 | 0 | 1 | Ref |
| 15559 | ON210137 | Unassigned | Unassigned | Unassigned | 618_0 | Clustered | 13 | VC_618_0 | 13 | 0.6252 | 1 | 0.6252 | 0 | 0 | 0 | 1 | Ref |
| 15560 | ON210138 | Unassigned | Unassigned | Unassigned | 329_20 | Clustered | 4 | VC_329_20 | 4 | 0.0174 | 0.99998777 | 0.0174 | 0 | 0 | 0 | 1 | Ref |
| 15561 | ON210139 | Unassigned | Unassigned | Unassigned | 283_98 | Clustered/Singleton | 1 | VC_283_98 | 1 | 0 | 1 | 0 | 0 | 0 | 0 | 1 | Ref |
| 15562 | ON210142 | Unassigned | Unassigned | Unassigned | 699_0 | Clustered | 71 | VC_699_0 | 71 | 0.9352 | 1 | 0.9352 | 0 | 0 | 0 | 1 | Ref |
| 15563 | ON210144 | Unassigned | Unassigned | Unassigned | 452_0 | Clustered | 66 | VC_452_0 | 66 | 0.9248 | 1 | 0.9248 | 0 | 0 | 0 | 1 | Ref |
| 15564 | ON210145 | Unassigned | Unassigned | Unassigned | 452_0 | Clustered | 66 | VC_452_0 | 66 | 0.9248 | 1 | 0.9248 | 0 | 0 | 0 | 1 | Ref |
| 15565 | ON210834 | Unassigned | Unassigned | Unassigned | 214_0 | Clustered | 2 | VC_214_0 | 2 | 0.2179 | 0.95358803 | 0.2078 | 0 | 0 | 0 | 1 | Ref |
| 15566 | ON210835 | Unassigned | Unassigned | Unassigned | 214_0 | Clustered | 2 | VC_214_0 | 2 | 0.2179 | 0.95358803 | 0.2078 | 0 | 0 | 0 | 1 | Ref |
| 15567 | ON212266 | Unassigned | Unassigned | Unassigned | 131_1 | Clustered | 6 | VC_131_1 | 6 | 0.057 | 0.99999999 | 0.057 | 0 | 0 | 0 | 1 | Ref |
| 15568 | ON229909 | Unassigned | Unassigned | Unassigned | 283_160 | Clustered/Singleton | 1 | VC_283_160 | 1 | 0 | 1 | 0 | 0 | 0 | 0 | 1 | Ref |
| 15569 | ON237674 | Unassigned | Unassigned | Unassigned | 698_0 | Clustered | 26 | VC_698_0 | 26 | 0.9366 | 1 | 0.9366 | 0 | 0 | 0 | 1 | Ref |
| 15570 | ON239128 | Unassigned | Unassigned | Unassigned | 329_11 | Clustered | 15 | VC_329_11 | 15 | 0.0791 | 1 | 0.0791 | 0 | 0 | 0 | 0.9333 | Ref |
| 15571 | ON239131 | Unassigned | Unassigned | Unassigned | 228_0 | Clustered | 60 | VC_228_0 | 60 | 0.942 | 1 | 0.942 | 0 | 0 | 0 | 1 | Ref |
| 15572 | ON239132 | Unassigned | Unassigned | Unassigned | 461_0 | Clustered | 96 | VC_461_0 | 96 | 0.9307 | 1 | 0.9307 | 0 | 0 | 0 | 0.9575 | Ref |
| 15573 | ON260812 | Unassigned | Unassigned | Unassigned | 1344_0 | Clustered | 78 | VC_1344_0 | 78 | 0.9732 | 1 | 0.9732 | 0 | 0 | 0 | 1 | Ref |
| 15574 | ON260813 | Unassigned | Unassigned | Unassigned | 510_0 | Clustered | 12 | VC_510_0 | 12 | 0.3963 | 1 | 0.3963 | 0 | 0 | 0 | 0.9697 | Ref |
| 15575 | ON260814 | Unassigned | Unassigned | Unassigned | 1344_0 | Clustered | 78 | VC_1344_0 | 78 | 0.9732 | 1 | 0.9732 | 0 | 0 | 0 | 1 | Ref |
| 15576 | ON260815 | Unassigned | Unassigned | Unassigned | 1352_0 | Clustered | 31 | VC_1352_0 | 31 | 0.849 | 1 | 0.849 | 0 | 0 | 0 | 1 | Ref |
| 15577 | ON260816 | Unassigned | Unassigned | Unassigned | 236_0 | Clustered | 72 | VC_236_0 | 72 | 0.8104 | 1 | 0.8104 | 0 | 0 | 0 | 1 | Ref |
| 15578 | ON260817 | Unassigned | Unassigned | Unassigned | 1361_0 | Clustered | 6 | VC_1361_0 | 6 | 0.2371 | 1 | 0.2371 | 0 | 0 | 0 | 1 | Ref |
| 15579 | ON260818 | Unassigned | Unassigned | Unassigned | 686_0 | Clustered | 2 | VC_686_0 | 2 | 0.2147 | 0.9539194 | 0.2048 | 0 | 0 | 0 | 1 | Ref |
| 15580 | ON260819 | Unassigned | Unassigned | Unassigned | 1071_0 | Clustered | 20 | VC_1071_0 | 20 | 0.3268 | 1 | 0.3268 | 0 | 0 | 0 | 0.9842 | Ref |
| 15581 | ON260820 | Unassigned | Unassigned | Unassigned | 1352_0 | Clustered | 31 | VC_1352_0 | 31 | 0.849 | 1 | 0.849 | 0 | 0 | 0 | 1 | Ref |
| 15582 | ON260821 | Unassigned | Unassigned | Unassigned | 1162_0 | Clustered | 10 | VC_1162_0 | 10 | 0.5715 | 1 | 0.5715 | 0 | 0 | 0 | 1 | Ref |
| 15583 | ON260822 | Unassigned | Unassigned | Unassigned | 1166_0 | Clustered | 20 | VC_1166_0 | 20 | 0.99 | 1 | 0.99 | 0 | 0 | 0 | 1 | Ref |
| 15584 | ON260823 | Unassigned | Unassigned | Unassigned | 1240_0 | Clustered | 82 | VC_1240_0 | 82 | 0.7945 | 1 | 0.7945 | 0 | 0 | 0 | 1 | Ref |
| 15585 | ON260824 | Unassigned | Unassigned | Unassigned | 558_20 | Clustered | 17 | VC_558_20 | 17 | 0.054 | 1 | 0.054 | 0 | 0 | 0 | 0.9926 | Ref |
| 15586 | ON260825 | Unassigned | Unassigned | Unassigned | 1348_0 | Clustered | 5 | VC_1348_0 | 5 | 0.5462 | 0.99999985 | 0.5462 | 0 | 0 | 0 | 1 | Ref |
| 15587 | ON260826 | Unassigned | Unassigned | Unassigned | 406_0 | Clustered | 11 | VC_406_0 | 11 | 0.9449 | 1 | 0.9449 | 0 | 0 | 0 | 1 | Ref |
| 15588 | ON260827 | Unassigned | Unassigned | Unassigned | 856_0 | Clustered | 7 | VC_856_0 | 7 | 0.3463 | 1 | 0.3463 | 0 | 0 | 0 | 1 | Ref |
| 15589 | ON260828 | Unassigned | Unassigned | Unassigned | 1071_0 | Clustered | 20 | VC_1071_0 | 20 | 0.3268 | 1 | 0.3268 | 0 | 0 | 0 | 0.9842 | Ref |
| 15590 | ON260829 | Unassigned | Unassigned | Unassigned | 556_0 | Clustered | 21 | VC_556_0 | 21 | 0.9912 | 1 | 0.9912 | 0 | 0 | 0 | 1 | Ref |
| 15591 | ON286972 | Unassigned | Unassigned | Unassigned | 164_3 | Clustered/Singleton | 1 | VC_164_3 | 1 | 0 | 1 | 0 | 0 | 0 | 0 | 1 | Ref |
| 15592 | ON286973 | Unassigned | Unassigned | Unassigned | 772_8 | Clustered | 2 | VC_772_8 | 2 | 0.006 | 0.58293682 | 0.0035 | 0 | 0 | 0 | 1 | Ref |
| 15593 | ON286974 | Unassigned | Unassigned | Unassigned | 590_0 | Clustered | 27 | VC_590_0 | 27 | 0.9947 | 1 | 0.9947 | 0 | 0 | 0 | 1 | Ref |
| 15594 | ON286975 | Unassigned | Unassigned | Unassigned | 772_1 | Clustered | 2 | VC_772_1 | 2 | 0.006 | 0.57682361 | 0.0034 | 0 | 0 | 0 | 1 | Ref |
| 15595 | ON286976 | Unassigned | Unassigned | Unassigned | 91_0 | Clustered | 22 | VC_91_0 | 22 | 0.5318 | 1 | 0.5318 | 0 | 0 | 0 | 1 | Ref |
| 15596 | ON287369 | Unassigned | Unassigned | Unassigned | 849_0 | Clustered | 3 | VC_849_0 | 3 | 0.069 | 0.9984794 | 0.0689 | 0 | 0 | 0 | 1 | Ref |
| 15597 | ON287370 | Unassigned | Unassigned | Unassigned | 442_1 | Clustered | 3 | VC_442_1 | 3 | 0.0538 | 0.99419552 | 0.0535 | 0 | 0 | 0 | 1 | Ref |
| 15598 | ON287372 | Unassigned | Unassigned | Unassigned | 488_1 | Clustered | 8 | VC_488_1 | 8 | 0.2927 | 1 | 0.2927 | 0 | 0 | 0 | 1 | Ref |
| 15599 | ON287373 | Unassigned | Unassigned | Unassigned | 196_0 | Clustered | 2 | VC_196_0 | 2 | 0.0218 | 0.95762696 | 0.0209 | 0 | 0 | 0 | 1 | Ref |
| 15600 | ON287374 | Unassigned | Unassigned | Unassigned | 849_0 | Clustered | 3 | VC_849_0 | 3 | 0.069 | 0.9984794 | 0.0689 | 0 | 0 | 0 | 1 | Ref |
| 15601 | ON287375 | Unassigned | Unassigned | Unassigned | 442_1 | Clustered | 3 | VC_442_1 | 3 | 0.0538 | 0.99419552 | 0.0535 | 0 | 0 | 0 | 1 | Ref |
| 15602 | ON287376 | Unassigned | Unassigned | Unassigned | 488_1 | Clustered | 8 | VC_488_1 | 8 | 0.2927 | 1 | 0.2927 | 0 | 0 | 0 | 1 | Ref |
| 15603 | ON287377 | Unassigned | Unassigned | Unassigned | 878_0 | Clustered | 32 | VC_878_0 | 32 | 0.9965 | 1 | 0.9965 | 0 | 0 | 0 | 1 | Ref |
| 15604 | ON287378 | Unassigned | Unassigned | Unassigned | 393_0 | Clustered | 7 | VC_393_0 | 7 | 0.1775 | 1 | 0.1775 | 0 | 0 | 0 | 1 | Ref |
| 15605 | ON323462 | Unassigned | Unassigned | Unassigned | 132_1 | Clustered | 17 | VC_132_1 | 17 | 0.1738 | 1 | 0.1738 | 0 | 0 | 0 | 0.9853 | Ref |
| 15606 | ON323491 | Unassigned | Unassigned | Unassigned | 1090_0 | Clustered | 4 | VC_1090_0 | 4 | 1 | 1 | 1 | 0 | 0 | 0 | 1 | Ref |
| 15607 | ON324181 | Unassigned | Unassigned | Unassigned | 79_0 | Clustered | 64 | VC_79_0 | 64 | 0.4185 | 1 | 0.4185 | 0 | 0 | 0 | 0.7649 | Ref |
| 15608 | ON331942 | Unassigned | Unassigned | Unassigned | 169_0 | Clustered | 20 | VC_169_0 | 20 | 0.7529 | 1 | 0.7529 | 0 | 0 | 0 | 1 | Ref |
| 15609 | ON337194 | Unassigned | Unassigned | Unassigned | 1240_0 | Clustered | 82 | VC_1240_0 | 82 | 0.7945 | 1 | 0.7945 | 0 | 0 | 0 | 1 | Ref |
| 15610 | ON337195 | Unassigned | Unassigned | Unassigned | 1167_0 | Clustered | 6 | VC_1167_0 | 6 | 0.8642 | 0.99999946 | 0.8642 | 0 | 0 | 0 | 1 | Ref |
| 15611 | ON337196 | Unassigned | Unassigned | Unassigned | 515_0 | Clustered | 16 | VC_515_0 | 16 | 0.3793 | 1 | 0.3793 | 0 | 0 | 0 | 1 | Ref |
| 15612 | ON352054 | Unassigned | Unassigned | Unassigned | 967_0 | Clustered | 16 | VC_967_0 | 16 | 0.8971 | 1 | 0.8971 | 0 | 0 | 0 | 1 | Ref |
| 15613 | ON361134 | Unassigned | Unassigned | Unassigned | 80_0 | Clustered | 41 | VC_80_0 | 41 | 0.5837 | 1 | 0.5837 | 0 | 0 | 0 | 0.9805 | Ref |
| 15614 | ON361135 | Unassigned | Unassigned | Unassigned | 76_0 | Clustered | 37 | VC_76_0 | 37 | 0.9403 | 1 | 0.9403 | 0 | 0 | 0 | 1 | Ref |
| 15615 | ON375838 | Unassigned | Unassigned | Unassigned | 79_0 | Clustered | 64 | VC_79_0 | 64 | 0.4185 | 1 | 0.4185 | 0 | 0 | 0 | 0.7649 | Ref |
| 15616 | ON375839 | Unassigned | Unassigned | Unassigned | 79_0 | Clustered | 64 | VC_79_0 | 64 | 0.4185 | 1 | 0.4185 | 0 | 0 | 0 | 0.7649 | Ref |
| 15617 | ON375840 | Unassigned | Unassigned | Unassigned | 79_0 | Clustered | 64 | VC_79_0 | 64 | 0.4185 | 1 | 0.4185 | 0 | 0 | 0 | 0.7649 | Ref |
| 15618 | ON375847 | Unassigned | Unassigned | Unassigned | 488_1 | Clustered | 8 | VC_488_1 | 8 | 0.2927 | 1 | 0.2927 | 0 | 0 | 0 | 1 | Ref |
| 15619 | ON376263 | Unassigned | Unassigned | Unassigned | 472_0 | Clustered | 47 | VC_472_0 | 47 | 0.9025 | 1 | 0.9025 | 0 | 0 | 0 | 1 | Ref |
| 15620 | ON381767 | Unassigned | Unassigned | Unassigned | 744_0 | Clustered | 10 | VC_744_0 | 10 | 0.7049 | 1 | 0.7049 | 0 | 0 | 0 | 1 | Ref |
| 15621 | ON381768 | Unassigned | Unassigned | Unassigned | 637_0 | Clustered | 115 | VC_637_0 | 115 | 0.731 | 1 | 0.731 | 0 | 0 | 0 | 0.8764 | Ref |
| 15622 | ON391668 | Unassigned | Unassigned | Unassigned | 638_0 | Clustered | 45 | VC_638_0 | 45 | 0.7741 | 1 | 0.7741 | 0 | 0 | 0 | 1 | Ref |
| 15623 | ON391949 | Unassigned | Unassigned | Unassigned | 696_1 | Clustered | 8 | VC_696_1 | 8 | 0.5452 | 1 | 0.5452 | 0 | 0 | 0 | 0.9286 | Ref |
| 15624 | ON392159 | Unassigned | Unassigned | Unassigned | 1095_0 | Clustered | 16 | VC_1095_0 | 16 | 0.458 | 1 | 0.458 | 0 | 0 | 0 | 1 | Ref |
| 15625 | ON392160 | Unassigned | Unassigned | Unassigned | 761_0 | Clustered | 67 | VC_761_0 | 67 | 0.8947 | 1 | 0.8947 | 0 | 0 | 0 | 1 | Ref |
| 15626 | ON392161 | Unassigned | Unassigned | Unassigned | 1344_0 | Clustered | 78 | VC_1344_0 | 78 | 0.9732 | 1 | 0.9732 | 0 | 0 | 0 | 1 | Ref |
| 15627 | ON392162 | Unassigned | Unassigned | Unassigned | 761_0 | Clustered | 67 | VC_761_0 | 67 | 0.8947 | 1 | 0.8947 | 0 | 0 | 0 | 1 | Ref |
| 15628 | ON392166 | Unassigned | Unassigned | Unassigned | 1037_0 | Clustered | 6 | VC_1037_0 | 6 | 0.6798 | 1 | 0.6798 | 0 | 0 | 0 | 1 | Ref |
| 15629 | ON399185 | Unassigned | Unassigned | Unassigned | 132_1 | Clustered | 17 | VC_132_1 | 17 | 0.1738 | 1 | 0.1738 | 0 | 0 | 0 | 0.9853 | Ref |
| 15630 | ON409568 | Unassigned | Unassigned | Unassigned | 637_0 | Clustered | 115 | VC_637_0 | 115 | 0.731 | 1 | 0.731 | 0 | 0 | 0 | 0.8764 | Ref |
| 15631 | ON409569 | Unassigned | Unassigned | Unassigned | 637_0 | Clustered | 115 | VC_637_0 | 115 | 0.731 | 1 | 0.731 | 0 | 0 | 0 | 0.8764 | Ref |
| 15632 | ON409570 | Unassigned | Unassigned | Unassigned | 637_0 | Clustered | 115 | VC_637_0 | 115 | 0.731 | 1 | 0.731 | 0 | 0 | 0 | 0.8764 | Ref |
| 15633 | ON416862 | Unassigned | Unassigned | Unassigned | 40_0 | Clustered | 5 | VC_40_0 | 5 | 0.0844 | 0.99999995 | 0.0844 | 0 | 0 | 0 | 1 | Ref |
| 15634 | ON454038 | Unassigned | Unassigned | Unassigned | 637_0 | Clustered | 115 | VC_637_0 | 115 | 0.731 | 1 | 0.731 | 0 | 0 | 0 | 0.8764 | Ref |
| 15635 | ON456151 | Unassigned | Unassigned | Unassigned | 378_0 | Clustered | 9 | VC_378_0 | 9 | 0.7603 | 1 | 0.7603 | 0 | 0 | 0 | 1 | Ref |
| 15636 | ON456330 | Unassigned | Unassigned | Unassigned | 371_13 | Clustered | 33 | VC_371_13 | 33 | 0.179 | 1 | 0.179 | 0 | 0 | 0 | 0.8769 | Ref |
| 15637 | ON456331 | Unassigned | Unassigned | Unassigned | 868_0 | Clustered | 25 | VC_868_0 | 25 | 0.5791 | 1 | 0.5791 | 0 | 0 | 0 | 1 | Ref |
| 15638 | ON456332 | Unassigned | Unassigned | Unassigned | 371_12 | Clustered | 4 | VC_371_12 | 4 | 0.0126 | 0.99998489 | 0.0126 | 0 | 0 | 0 | 0.8333 | Ref |
| 15639 | ON456333 | Unassigned | Unassigned | Unassigned | 767_0 | Clustered | 34 | VC_767_0 | 34 | 0.9173 | 1 | 0.9173 | 0 | 0 | 0 | 1 | Ref |
| 15640 | ON456334 | Unassigned | Unassigned | Unassigned | 557_1 | Clustered | 11 | VC_557_1 | 11 | 0.4945 | 1 | 0.4945 | 0 | 0 | 0 | 0.7818 | Ref |
| 15641 | ON456335 | Unassigned | Unassigned | Unassigned | 1164_0 | Clustered | 25 | VC_1164_0 | 25 | 0.7312 | 1 | 0.7312 | 0 | 0 | 0 | 1 | Ref |
| 15642 | ON456336 | Unassigned | Unassigned | Unassigned | 1093_0 | Clustered | 40 | VC_1093_0 | 40 | 0.8754 | 1 | 0.8754 | 0 | 0 | 0 | 1 | Ref |
| 15643 | ON456337 | Unassigned | Unassigned | Unassigned | 1095_0 | Clustered | 16 | VC_1095_0 | 16 | 0.458 | 1 | 0.458 | 0 | 0 | 0 | 1 | Ref |
| 15644 | ON456338 | Unassigned | Unassigned | Unassigned | 1353_0 | Clustered | 10 | VC_1353_0 | 10 | 1 | 1 | 1 | 0 | 0 | 0 | 1 | Ref |
| 15645 | ON456339 | Unassigned | Unassigned | Unassigned | 236_0 | Clustered | 72 | VC_236_0 | 72 | 0.8104 | 1 | 0.8104 | 0 | 0 | 0 | 1 | Ref |
| 15646 | ON456340 | Unassigned | Unassigned | Unassigned | 1164_0 | Clustered | 25 | VC_1164_0 | 25 | 0.7312 | 1 | 0.7312 | 0 | 0 | 0 | 1 | Ref |
| 15647 | ON456342 | Unassigned | Unassigned | Unassigned | 371_0 | Clustered | 2 | VC_371_0 | 2 | 0.0032 | 0.94731972 | 0.003 | 0 | 0 | 0 | 1 | Ref |
| 15648 | ON456343 | Unassigned | Unassigned | Unassigned | 1072_2 | Clustered | 5 | VC_1072_2 | 5 | 0.1003 | 0.99999315 | 0.1003 | 0 | 0 | 0 | 1 | Ref |
| 15649 | ON456344 | Unassigned | Unassigned | Unassigned | 558_70 | Clustered/Singleton | 1 | VC_558_70 | 1 | 0 | 1 | 0 | 0 | 0 | 0 | 1 | Ref |
| 15650 | ON456345 | Unassigned | Unassigned | Unassigned | 551_7 | Clustered | 28 | VC_551_7 | 28 | 0.1661 | 1 | 0.1661 | 0 | 0 | 0 | 0.8519 | Ref |
| 15651 | ON456346 | Unassigned | Unassigned | Unassigned | 1360_0 | Clustered | 31 | VC_1360_0 | 31 | 0.8837 | 1 | 0.8837 | 0 | 0 | 0 | 1 | Ref |
| 15652 | ON456348 | Unassigned | Unassigned | Unassigned | 1353_0 | Clustered | 10 | VC_1353_0 | 10 | 1 | 1 | 1 | 0 | 0 | 0 | 1 | Ref |
| 15653 | ON456349 | Unassigned | Unassigned | Unassigned | 1095_0 | Clustered | 16 | VC_1095_0 | 16 | 0.458 | 1 | 0.458 | 0 | 0 | 0 | 1 | Ref |
| 15654 | ON456350 | Unassigned | Unassigned | Unassigned | 1344_0 | Clustered | 78 | VC_1344_0 | 78 | 0.9732 | 1 | 0.9732 | 0 | 0 | 0 | 1 | Ref |
| 15655 | ON456351 | Unassigned | Unassigned | Unassigned | 237_2 | Clustered | 11 | VC_237_2 | 11 | 0.1409 | 1 | 0.1409 | 0 | 0 | 0 | 0.9818 | Ref |
| 15656 | ON456352 | Unassigned | Unassigned | Unassigned | 410_0 | Clustered | 7 | VC_410_0 | 7 | 0.7905 | 1 | 0.7905 | 0 | 0 | 0 | 1 | Ref |
| 15657 | ON456353 | Unassigned | Unassigned | Unassigned | 236_0 | Clustered | 72 | VC_236_0 | 72 | 0.8104 | 1 | 0.8104 | 0 | 0 | 0 | 1 | Ref |
| 15658 | ON456354 | Unassigned | Unassigned | Unassigned | 1240_0 | Clustered | 82 | VC_1240_0 | 82 | 0.7945 | 1 | 0.7945 | 0 | 0 | 0 | 1 | Ref |
| 15659 | ON456356 | Unassigned | Unassigned | Unassigned | 520_43 | Clustered | 3 | VC_520_43 | 3 | 0.0193 | 0.99837043 | 0.0193 | 0 | 0 | 0 | 1 | Ref |
| 15660 | ON457554 | Unassigned | Unassigned | Unassigned | 837_0 | Clustered | 29 | VC_837_0 | 29 | 0.9955 | 1 | 0.9955 | 0 | 0 | 0 | 1 | Ref |
| 15661 | ON457555 | Unassigned | Unassigned | Unassigned | 837_0 | Clustered | 29 | VC_837_0 | 29 | 0.9955 | 1 | 0.9955 | 0 | 0 | 0 | 1 | Ref |
| 15662 | ON457556 | Unassigned | Unassigned | Unassigned | 837_0 | Clustered | 29 | VC_837_0 | 29 | 0.9955 | 1 | 0.9955 | 0 | 0 | 0 | 1 | Ref |
| 15663 | ON457557 | Unassigned | Unassigned | Unassigned | 837_0 | Clustered | 29 | VC_837_0 | 29 | 0.9955 | 1 | 0.9955 | 0 | 0 | 0 | 1 | Ref |
| 15664 | ON457558 | Unassigned | Unassigned | Unassigned | 614_0 | Clustered | 5 | VC_614_0 | 5 | 0.2395 | 0.99999997 | 0.2395 | 0 | 0 | 0 | 1 | Ref |
| 15665 | ON457559 | Unassigned | Unassigned | Unassigned | 614_0 | Clustered | 5 | VC_614_0 | 5 | 0.2395 | 0.99999997 | 0.2395 | 0 | 0 | 0 | 1 | Ref |
| 15666 | ON461912 | Unassigned | Unassigned | Unassigned | 452_0 | Clustered | 66 | VC_452_0 | 66 | 0.9248 | 1 | 0.9248 | 0 | 0 | 0 | 1 | Ref |
| 15667 | ON464735 | Unassigned | Unassigned | Unassigned | 82_0 | Clustered | 24 | VC_82_0 | 24 | 0.7351 | 1 | 0.7351 | 0 | 0 | 0 | 1 | Ref |
| 15668 | ON464736 | Unassigned | Unassigned | Unassigned | 82_0 | Clustered | 24 | VC_82_0 | 24 | 0.7351 | 1 | 0.7351 | 0 | 0 | 0 | 1 | Ref |
| 15669 | ON464759 | Unassigned | Unassigned | Unassigned | 1474_0 | Clustered | 3 | VC_1474_0 | 3 | 1 | 0 | 0 | 0 | 0 | 0 | 1 | Ref |
| 15670 | ON464760 | Unassigned | Unassigned | Unassigned | 713_0 | Clustered | 3 | VC_713_0 | 3 | 0.5082 | 0.99827116 | 0.5074 | 0 | 0 | 0 | 1 | Ref |
| 15671 | ON464761 | Unassigned | Unassigned | Unassigned | 713_0 | Clustered | 3 | VC_713_0 | 3 | 0.5082 | 0.99827116 | 0.5074 | 0 | 0 | 0 | 1 | Ref |
| 15672 | ON464762 | Unassigned | Unassigned | Unassigned | 713_0 | Clustered | 3 | VC_713_0 | 3 | 0.5082 | 0.99827116 | 0.5074 | 0 | 0 | 0 | 1 | Ref |
| 15673 | ON464763 | Unassigned | Unassigned | Unassigned | 1474_0 | Clustered | 3 | VC_1474_0 | 3 | 1 | 0 | 0 | 0 | 0 | 0 | 1 | Ref |
| 15674 | ON464764 | Unassigned | Unassigned | Unassigned | 1474_0 | Clustered | 3 | VC_1474_0 | 3 | 1 | 0 | 0 | 0 | 0 | 0 | 1 | Ref |
| 15675 | ON470578 | Unassigned | Unassigned | Unassigned | 51_0 | Clustered | 10 | VC_51_0 | 10 | 0.1741 | 1 | 0.1741 | 0 | 0 | 0 | 1 | Ref |
| 15676 | ON470579 | Unassigned | Unassigned | Unassigned | 242_1 | Clustered | 20 | VC_242_1 | 20 | 0.684 | 1 | 0.684 | 0 | 0 | 0 | 0.8368 | Ref |
| 15677 | ON470580 | Unassigned | Unassigned | Unassigned | 245_0 | Clustered | 10 | VC_245_0 | 10 | 0.1805 | 1 | 0.1805 | 0 | 0 | 0 | 0.9778 | Ref |
| 15678 | ON470582 | Unassigned | Unassigned | Unassigned | 242_1 | Clustered | 20 | VC_242_1 | 20 | 0.684 | 1 | 0.684 | 0 | 0 | 0 | 0.8368 | Ref |
| 15679 | ON470583 | Unassigned | Unassigned | Unassigned | 245_0 | Clustered | 10 | VC_245_0 | 10 | 0.1805 | 1 | 0.1805 | 0 | 0 | 0 | 0.9778 | Ref |
| 15680 | ON470584 | Unassigned | Unassigned | Unassigned | 45_0 | Clustered | 12 | VC_45_0 | 12 | 0.35 | 1 | 0.35 | 0 | 0 | 0 | 1 | Ref |
| 15681 | ON470586 | Unassigned | Unassigned | Unassigned | 245_0 | Clustered | 10 | VC_245_0 | 10 | 0.1805 | 1 | 0.1805 | 0 | 0 | 0 | 0.9778 | Ref |
| 15682 | ON470588 | Unassigned | Unassigned | Unassigned | 242_1 | Clustered | 20 | VC_242_1 | 20 | 0.684 | 1 | 0.684 | 0 | 0 | 0 | 0.8368 | Ref |
| 15683 | ON470589 | Unassigned | Unassigned | Unassigned | 30_1 | Clustered | 16 | VC_30_1 | 16 | 0.1973 | 1 | 0.1973 | 0 | 0 | 0 | 0.8333 | Ref |
| 15684 | ON470591 | Unassigned | Unassigned | Unassigned | 710_0 | Clustered | 10 | VC_710_0 | 10 | 0.2446 | 1 | 0.2446 | 0 | 0 | 0 | 0.8222 | Ref |
| 15685 | ON470593 | Unassigned | Unassigned | Unassigned | 242_1 | Clustered | 20 | VC_242_1 | 20 | 0.684 | 1 | 0.684 | 0 | 0 | 0 | 0.8368 | Ref |
| 15686 | ON470594 | Unassigned | Unassigned | Unassigned | 30_1 | Clustered | 16 | VC_30_1 | 16 | 0.1973 | 1 | 0.1973 | 0 | 0 | 0 | 0.8333 | Ref |
| 15687 | ON470596 | Unassigned | Unassigned | Unassigned | 57_0 | Clustered | 4 | VC_57_0 | 4 | 0.4183 | 0.99998673 | 0.4183 | 0 | 0 | 0 | 1 | Ref |
| 15688 | ON470597 | Unassigned | Unassigned | Unassigned | 41_0 | Clustered | 16 | VC_41_0 | 16 | 0.2025 | 1 | 0.2025 | 0 | 0 | 0 | 0.9667 | Ref |
| 15689 | ON470598 | Unassigned | Unassigned | Unassigned | 51_0 | Clustered | 10 | VC_51_0 | 10 | 0.1741 | 1 | 0.1741 | 0 | 0 | 0 | 1 | Ref |
| 15690 | ON470600 | Unassigned | Unassigned | Unassigned | 245_0 | Clustered | 10 | VC_245_0 | 10 | 0.1805 | 1 | 0.1805 | 0 | 0 | 0 | 0.9778 | Ref |
| 15691 | ON470601 | Unassigned | Unassigned | Unassigned | 228_0 | Clustered | 60 | VC_228_0 | 60 | 0.942 | 1 | 0.942 | 0 | 0 | 0 | 1 | Ref |
| 15692 | ON470603 | Unassigned | Unassigned | Unassigned | 45_0 | Clustered | 12 | VC_45_0 | 12 | 0.35 | 1 | 0.35 | 0 | 0 | 0 | 1 | Ref |
| 15693 | ON470604 | Unassigned | Unassigned | Unassigned | 57_0 | Clustered | 4 | VC_57_0 | 4 | 0.4183 | 0.99998673 | 0.4183 | 0 | 0 | 0 | 1 | Ref |
| 15694 | ON470605 | Unassigned | Unassigned | Unassigned | 30_1 | Clustered | 16 | VC_30_1 | 16 | 0.1973 | 1 | 0.1973 | 0 | 0 | 0 | 0.8333 | Ref |
| 15695 | ON470606 | Unassigned | Unassigned | Unassigned | 228_0 | Clustered | 60 | VC_228_0 | 60 | 0.942 | 1 | 0.942 | 0 | 0 | 0 | 1 | Ref |
| 15696 | ON470607 | Unassigned | Unassigned | Unassigned | 245_0 | Clustered | 10 | VC_245_0 | 10 | 0.1805 | 1 | 0.1805 | 0 | 0 | 0 | 0.9778 | Ref |
| 15697 | ON470612 | Unassigned | Unassigned | Unassigned | 245_0 | Clustered | 10 | VC_245_0 | 10 | 0.1805 | 1 | 0.1805 | 0 | 0 | 0 | 0.9778 | Ref |
| 15698 | ON470613 | Unassigned | Unassigned | Unassigned | 228_0 | Clustered | 60 | VC_228_0 | 60 | 0.942 | 1 | 0.942 | 0 | 0 | 0 | 1 | Ref |
| 15699 | ON470615 | Unassigned | Unassigned | Unassigned | 247_0 | Clustered | 14 | VC_247_0 | 14 | 0.9119 | 1 | 0.9119 | 0 | 0 | 0 | 0.967 | Ref |
| 15700 | ON470617 | Unassigned | Unassigned | Unassigned | 247_0 | Clustered | 14 | VC_247_0 | 14 | 0.9119 | 1 | 0.9119 | 0 | 0 | 0 | 0.967 | Ref |
| 15701 | ON470618 | Unassigned | Unassigned | Unassigned | 51_0 | Clustered | 10 | VC_51_0 | 10 | 0.1741 | 1 | 0.1741 | 0 | 0 | 0 | 1 | Ref |
| 15702 | ON470620 | Unassigned | Unassigned | Unassigned | 6_0 | Clustered | 44 | VC_6_0 | 44 | 0.908 | 1 | 0.908 | 0 | 0 | 0 | 1 | Ref |
| 15703 | ON470621 | Unassigned | Unassigned | Unassigned | 245_0 | Clustered | 10 | VC_245_0 | 10 | 0.1805 | 1 | 0.1805 | 0 | 0 | 0 | 0.9778 | Ref |
| 15704 | ON470624 | Unassigned | Unassigned | Unassigned | 57_0 | Clustered | 4 | VC_57_0 | 4 | 0.4183 | 0.99998673 | 0.4183 | 0 | 0 | 0 | 1 | Ref |
| 15705 | ON470625 | Unassigned | Unassigned | Unassigned | 41_0 | Clustered | 16 | VC_41_0 | 16 | 0.2025 | 1 | 0.2025 | 0 | 0 | 0 | 0.9667 | Ref |
| 15706 | ON470627 | Unassigned | Unassigned | Unassigned | 57_0 | Clustered | 4 | VC_57_0 | 4 | 0.4183 | 0.99998673 | 0.4183 | 0 | 0 | 0 | 1 | Ref |
| 15707 | ON480525 | Unassigned | Unassigned | Unassigned | 698_0 | Clustered | 26 | VC_698_0 | 26 | 0.9366 | 1 | 0.9366 | 0 | 0 | 0 | 1 | Ref |
| 15708 | ON489264 | Unassigned | Unassigned | Unassigned | 938_0 | Clustered | 99 | VC_938_0 | 99 | 0.9119 | 1 | 0.9119 | 0 | 0 | 0 | 0.9928 | Ref |
| 15709 | ON506924 | Unassigned | Unassigned | Unassigned | 283_96 | Clustered/Singleton | 1 | VC_283_96 | 1 | 0 | 1 | 0 | 0 | 0 | 0 | 1 | Ref |
| 15710 | ON506926 | Unassigned | Unassigned | Unassigned | 691_0 | Clustered | 2 | VC_691_0 | 2 | 0.1052 | 0.95343038 | 0.1003 | 0 | 0 | 0 | 1 | Ref |
| 15711 | ON506927 | Unassigned | Unassigned | Unassigned | 92_0 | Clustered | 22 | VC_92_0 | 22 | 0.5639 | 1 | 0.5639 | 0 | 0 | 0 | 1 | Ref |
| 15712 | ON506928 | Unassigned | Unassigned | Unassigned | 92_0 | Clustered | 22 | VC_92_0 | 22 | 0.5639 | 1 | 0.5639 | 0 | 0 | 0 | 1 | Ref |

| | | | | | | | | | | | | | | | | | |
|---|---|---|---|---|---|---|---|---|---|---|---|---|---|---|---|---|---|
| 15713 | ON513429 | Unassigned | Unassigned | Unassigned | 699_0 | Clustered | 71 | VC_699_0 | 71 | 0.9352 | 1 | 0.9352 | 0 | 0 | 0 | 1 | Ref |
| 15714 | ON515458 | Unassigned | Unassigned | Unassigned | 668_0 | Clustered | 25 | VC_668_0 | 25 | 0.5539 | 1 | 0.5539 | 0 | 0 | 0 | 0.96 | Ref |
| 15715 | ON526838 | Unassigned | Unassigned | Unassigned | 637_0 | Clustered | 115 | VC_637_0 | 115 | 0.731 | 1 | 0.731 | 0 | 0 | 0 | 0.8764 | Ref |
| 15716 | ON526839 | Unassigned | Unassigned | Unassigned | 452_0 | Clustered | 66 | VC_452_0 | 66 | 0.9248 | 1 | 0.9248 | 0 | 0 | 0 | 1 | Ref |
| 15717 | ON526840 | Unassigned | Unassigned | Unassigned | 191_32 | Clustered | 12 | VC_191_32 | 12 | 0.0683 | 1 | 0.0683 | 0 | 0 | 0 | 0.7576 | Ref |
| 15718 | ON526969 | Unassigned | Unassigned | Unassigned | 1031_0 | Clustered | 4 | VC_1031_0 | 4 | 0.7884 | 0.99997621 | 0.7884 | 0 | 0 | 0 | 1 | Ref |
| 15719 | ON526970 | Unassigned | Unassigned | Unassigned | 981_0 | Clustered | 3 | VC_981_0 | 3 | 0.0531 | 0.99853685 | 0.053 | 0 | 0 | 0 | 1 | Ref |
| 15720 | ON526971 | Unassigned | Unassigned | Unassigned | 799_1 | Clustered | 27 | VC_799_1 | 27 | 0.8611 | 1 | 0.8611 | 0 | 0 | 0 | 1 | Ref |
| 15721 | ON526972 | Unassigned | Unassigned | Unassigned | 1163_0 | Clustered | 28 | VC_1163_0 | 28 | 0.4923 | 1 | 0.4923 | 0 | 0 | 0 | 1 | Ref |
| 15722 | ON526973 | Unassigned | Unassigned | Unassigned | 866_0 | Clustered | 4 | VC_866_0 | 4 | 0.0795 | 0.99998595 | 0.0795 | 0 | 0 | 0 | 1 | Ref |
| 15723 | ON526974 | Unassigned | Unassigned | Unassigned | 981_0 | Clustered | 3 | VC_981_0 | 3 | 0.0531 | 0.99853685 | 0.053 | 0 | 0 | 0 | 1 | Ref |
| 15724 | ON526975 | Unassigned | Unassigned | Unassigned | 867_0 | Clustered | 6 | VC_867_0 | 6 | 0.3202 | 1 | 0.3202 | 0 | 0 | 0 | 1 | Ref |
| 15725 | ON526976 | Unassigned | Unassigned | Unassigned | 520_30 | Clustered | 2 | VC_520_30 | 2 | 0.0073 | 0.9544492 | 0.0069 | 0 | 0 | 0 | 1 | Ref |
| 15726 | ON526977 | Unassigned | Unassigned | Unassigned | 1343_0 | Clustered | 21 | VC_1343_0 | 21 | 0.9843 | 1 | 0.9843 | 0 | 0 | 0 | 1 | Ref |
| 15727 | ON526978 | Unassigned | Unassigned | Unassigned | 234_0 | Clustered | 21 | VC_234_0 | 21 | 0.8355 | 1 | 0.8355 | 0 | 0 | 0 | 1 | Ref |
| 15728 | ON526979 | Unassigned | Unassigned | Unassigned | 975_0 | Clustered | 11 | VC_975_0 | 11 | 0.9687 | 1 | 0.9687 | 0 | 0 | 0 | 1 | Ref |
| 15729 | ON526980 | Unassigned | Unassigned | Unassigned | 1361_0 | Clustered | 6 | VC_1361_0 | 6 | 0.2371 | 1 | 0.2371 | 0 | 0 | 0 | 1 | Ref |
| 15730 | ON526981 | Unassigned | Unassigned | Unassigned | 557_1 | Clustered | 11 | VC_557_1 | 11 | 0.4945 | 1 | 0.4945 | 0 | 0 | 0 | 0.7818 | Ref |
| 15731 | ON526982 | Unassigned | Unassigned | Unassigned | 558_14 | Clustered | 8 | VC_558_14 | 8 | 0.0228 | 0.99945246 | 0.0228 | 0 | 0 | 0 | 0.8214 | Ref |
| 15732 | ON526984 | Unassigned | Unassigned | Unassigned | 236_0 | Clustered | 72 | VC_236_0 | 72 | 0.8104 | 1 | 0.8104 | 0 | 0 | 0 | 1 | Ref |
| 15733 | ON526985 | Unassigned | Unassigned | Unassigned | 982_0 | Clustered | 62 | VC_982_0 | 62 | 0.8646 | 1 | 0.8646 | 0 | 0 | 0 | 0.743 | Ref |
| 15734 | ON528715 | Unassigned | Unassigned | Unassigned | 392_0 | Clustered | 126 | VC_392_0 | 126 | 0.9621 | 1 | 0.9621 | 0 | 0 | 0 | 1 | Ref |
| 15735 | ON528716 | Unassigned | Unassigned | Unassigned | 392_0 | Clustered | 126 | VC_392_0 | 126 | 0.9621 | 1 | 0.9621 | 0 | 0 | 0 | 1 | Ref |
| 15736 | ON528717 | Unassigned | Unassigned | Unassigned | 392_0 | Clustered | 126 | VC_392_0 | 126 | 0.9621 | 1 | 0.9621 | 0 | 0 | 0 | 1 | Ref |
| 15737 | ON528718 | Unassigned | Unassigned | Unassigned | 941_0 | Clustered | 40 | VC_941_0 | 40 | 0.6903 | 1 | 0.6903 | 0 | 0 | 0 | 1 | Ref |
| 15738 | ON528719 | Unassigned | Unassigned | Unassigned | 392_0 | Clustered | 126 | VC_392_0 | 126 | 0.9621 | 1 | 0.9621 | 0 | 0 | 0 | 1 | Ref |
| 15739 | ON528720 | Unassigned | Unassigned | Unassigned | 385_0 | Clustered | 51 | VC_385_0 | 51 | 0.8754 | 1 | 0.8754 | 0 | 0 | 0 | 1 | Ref |
| 15740 | ON528721 | Unassigned | Unassigned | Unassigned | 385_0 | Clustered | 51 | VC_385_0 | 51 | 0.8754 | 1 | 0.8754 | 0 | 0 | 0 | 1 | Ref |
| 15741 | ON528722 | Unassigned | Unassigned | Unassigned | 385_0 | Clustered | 51 | VC_385_0 | 51 | 0.8754 | 1 | 0.8754 | 0 | 0 | 0 | 1 | Ref |
| 15742 | ON528723 | Unassigned | Unassigned | Unassigned | 385_0 | Clustered | 51 | VC_385_0 | 51 | 0.8754 | 1 | 0.8754 | 0 | 0 | 0 | 1 | Ref |
| 15743 | ON528724 | Unassigned | Unassigned | Unassigned | 385_0 | Clustered | 51 | VC_385_0 | 51 | 0.8754 | 1 | 0.8754 | 0 | 0 | 0 | 1 | Ref |
| 15744 | ON528725 | Unassigned | Unassigned | Unassigned | 286_15 | Clustered | 2 | VC_286_15 | 2 | 0.0073 | 0.75626308 | 0.0055 | 0 | 0 | 0 | 1 | Ref |
| 15745 | ON528726 | Unassigned | Unassigned | Unassigned | 283_142 | Clustered | 2 | VC_283_142 | 2 | 0.0029 | 0.94283912 | 0.0027 | 0 | 0 | 0 | 1 | Ref |
| 15746 | ON528727 | Unassigned | Unassigned | Unassigned | 283_29 | Clustered/Singleton | 1 | VC_283_29 | 1 | 0 | 1 | 0 | 0 | 0 | 0 | 1 | Ref |
| 15747 | ON528728 | Unassigned | Unassigned | Unassigned | 283_30 | Clustered/Singleton | 1 | VC_283_30 | 1 | 0 | 1 | 0 | 0 | 0 | 0 | 1 | Ref |
| 15748 | ON528729 | Unassigned | Unassigned | Unassigned | 330_0 | Clustered | 27 | VC_330_0 | 27 | 0.5088 | 1 | 0.5088 | 0 | 0 | 0 | 0.8661 | Ref |
| 15749 | ON528730 | Unassigned | Unassigned | Unassigned | 330_0 | Clustered | 27 | VC_330_0 | 27 | 0.5088 | 1 | 0.5088 | 0 | 0 | 0 | 0.8661 | Ref |
| 15750 | ON528731 | Unassigned | Unassigned | Unassigned | 330_0 | Clustered | 27 | VC_330_0 | 27 | 0.5088 | 1 | 0.5088 | 0 | 0 | 0 | 0.8661 | Ref |
| 15751 | ON528732 | Unassigned | Unassigned | Unassigned | 330_0 | Clustered | 27 | VC_330_0 | 27 | 0.5088 | 1 | 0.5088 | 0 | 0 | 0 | 0.8661 | Ref |
| 15752 | ON528733 | Unassigned | Unassigned | Unassigned | 451_0 | Clustered | 80 | VC_451_0 | 80 | 0.9991 | 1 | 0.9991 | 0 | 0 | 0 | 1 | Ref |
| 15753 | ON528734 | Unassigned | Unassigned | Unassigned | 283_86 | Clustered/Singleton | 1 | VC_283_86 | 1 | 0 | 1 | 0 | 0 | 0 | 0 | 1 | Ref |
| 15754 | ON528735 | Unassigned | Unassigned | Unassigned | 283_103 | Clustered/Singleton | 1 | VC_283_103 | 1 | 0 | 1 | 0 | 0 | 0 | 0 | 1 | Ref |
| 15755 | ON528736 | Unassigned | Unassigned | Unassigned | 283_149 | Clustered/Singleton | 1 | VC_283_149 | 1 | 0 | 1 | 0 | 0 | 0 | 0 | 1 | Ref |
| 15756 | ON528737 | Unassigned | Unassigned | Unassigned | 286_10 | Clustered | 9 | VC_286_10 | 9 | 0.0615 | 0.99999878 | 0.0615 | 0 | 0 | 0 | 0.6667 | Ref |
| 15757 | ON528738 | Unassigned | Unassigned | Unassigned | 283_87 | Clustered/Singleton | 1 | VC_283_87 | 1 | 0 | 1 | 0 | 0 | 0 | 0 | 1 | Ref |
| 15758 | ON528739 | Unassigned | Unassigned | Unassigned | 283_107 | Clustered/Singleton | 1 | VC_283_107 | 1 | 0 | 1 | 0 | 0 | 0 | 0 | 1 | Ref |
| 15759 | ON528740 | Unassigned | Unassigned | Unassigned | 283_132 | Clustered/Singleton | 1 | VC_283_132 | 1 | 0 | 1 | 0 | 0 | 0 | 0 | 1 | Ref |
| 15760 | ON528741 | Unassigned | Unassigned | Unassigned | 283_11 | Clustered/Singleton | 1 | VC_283_11 | 1 | 0 | 1 | 0 | 0 | 0 | 0 | 1 | Ref |
| 15761 | ON528742 | Unassigned | Unassigned | Unassigned | 283_186 | Clustered/Singleton | 1 | VC_283_186 | 1 | 0 | 1 | 0 | 0 | 0 | 0 | 1 | Ref |
| 15762 | ON528743 | Unassigned | Unassigned | Unassigned | 283_55 | Clustered/Singleton | 1 | VC_283_55 | 1 | 0 | 1 | 0 | 0 | 0 | 0 | 1 | Ref |
| 15763 | ON528744 | Unassigned | Unassigned | Unassigned | 283_142 | Clustered | 2 | VC_283_142 | 2 | 0.0029 | 0.94283912 | 0.0027 | 0 | 0 | 0 | 1 | Ref |
| 15764 | ON528745 | Unassigned | Unassigned | Unassigned | 283_13 | Clustered/Singleton | 1 | VC_283_13 | 1 | 0 | 1 | 0 | 0 | 0 | 0 | 1 | Ref |
| 15765 | ON528746 | Unassigned | Unassigned | Unassigned | 283_146 | Clustered/Singleton | 1 | VC_283_146 | 1 | 0 | 1 | 0 | 0 | 0 | 0 | 1 | Ref |
| 15766 | ON528933 | Unassigned | Unassigned | Unassigned | 1207_0 | Clustered | 29 | VC_1207_0 | 29 | 0.8225 | 1 | 0.8225 | 0 | 0 | 0 | 1 | Ref |
| 15767 | ON528935 | Unassigned | Unassigned | Unassigned | 18_0 | Clustered | 4 | VC_18_0 | 4 | 0.9098 | 0.99982347 | 0.9096 | 0 | 0 | 0 | 1 | Ref |
| 15768 | ON529291 | Unassigned | Unassigned | Unassigned | 79_0 | Clustered | 64 | VC_79_0 | 64 | 0.4185 | 1 | 0.4185 | 0 | 0 | 0 | 0.7649 | Ref |
| 15769 | ON529850 | Unassigned | Unassigned | Unassigned | 976_0 | Clustered | 2 | VC_976_0 | 2 | 0.0306 | 0.92749365 | 0.0284 | 0 | 0 | 0 | 1 | Ref |
| 15770 | ON529851 | Unassigned | Unassigned | Unassigned | 976_1 | Clustered | 5 | VC_976_1 | 5 | 0.1328 | 0.99999971 | 0.1328 | 0 | 0 | 0 | 0.9 | Ref |
| 15771 | ON529852 | Unassigned | Unassigned | Unassigned | 976_1 | Clustered | 5 | VC_976_1 | 5 | 0.1328 | 0.99999971 | 0.1328 | 0 | 0 | 0 | 0.9 | Ref |
| 15772 | ON529853 | Unassigned | Unassigned | Unassigned | 976_1 | Clustered | 5 | VC_976_1 | 5 | 0.1328 | 0.99999971 | 0.1328 | 0 | 0 | 0 | 0.9 | Ref |
| 15773 | ON529854 | Unassigned | Unassigned | Unassigned | 169_0 | Clustered | 20 | VC_169_0 | 20 | 0.7529 | 1 | 0.7529 | 0 | 0 | 0 | 1 | Ref |
| 15774 | ON529855 | Unassigned | Unassigned | Unassigned | 976_1 | Clustered | 5 | VC_976_1 | 5 | 0.1328 | 0.99999971 | 0.1328 | 0 | 0 | 0 | 0.9 | Ref |
| 15775 | ON529856 | Unassigned | Unassigned | Unassigned | 467_0 | Clustered | 5 | VC_467_0 | 5 | 0.6874 | 0.99999996 | 0.6874 | 0 | 0 | 0 | 1 | Ref |
| 15776 | ON529857 | Unassigned | Unassigned | Unassigned | 976_0 | Clustered | 2 | VC_976_0 | 2 | 0.0306 | 0.92749365 | 0.0284 | 0 | 0 | 0 | 1 | Ref |
| 15777 | ON529858 | Unassigned | Unassigned | Unassigned | 976_5 | Clustered/Singleton | 1 | VC_976_5 | 1 | 0 | 1 | 0 | 0 | 0 | 0 | 1 | Ref |
| 15778 | ON529859 | Unassigned | Unassigned | Unassigned | 467_0 | Clustered | 5 | VC_467_0 | 5 | 0.6874 | 0.99999996 | 0.6874 | 0 | 0 | 0 | 1 | Ref |
| 15779 | ON529860 | Unassigned | Unassigned | Unassigned | 976_1 | Clustered | 5 | VC_976_1 | 5 | 0.1328 | 0.99999971 | 0.1328 | 0 | 0 | 0 | 0.9 | Ref |
| 15780 | ON529862 | Unassigned | Unassigned | Unassigned | 467_0 | Clustered | 5 | VC_467_0 | 5 | 0.6874 | 0.99999996 | 0.6874 | 0 | 0 | 0 | 1 | Ref |
| 15781 | ON529863 | Unassigned | Unassigned | Unassigned | 1434_0 | Clustered | 2 | VC_1434_0 | 2 | 0.8742 | 0.85557782 | 0.7479 | 0 | 0 | 0 | 1 | Ref |
| 15782 | ON529864 | Unassigned | Unassigned | Unassigned | 467_0 | Clustered | 5 | VC_467_0 | 5 | 0.6874 | 0.99999996 | 0.6874 | 0 | 0 | 0 | 1 | Ref |
| 15783 | ON529865 | Unassigned | Unassigned | Unassigned | 1475_0 | Clustered | 2 | VC_1475_0 | 2 | 1 | 1 | 1 | 0 | 0 | 0 | 1 | Ref |
| 15784 | ON529866 | Unassigned | Unassigned | Unassigned | 1475_0 | Clustered | 2 | VC_1475_0 | 2 | 1 | 1 | 1 | 0 | 0 | 0 | 1 | Ref |
| 15785 | ON529867 | Unassigned | Unassigned | Unassigned | 1434_0 | Clustered | 2 | VC_1434_0 | 2 | 0.8742 | 0.85557782 | 0.7479 | 0 | 0 | 0 | 1 | Ref |
| 15786 | ON529868 | Unassigned | Unassigned | Unassigned | 70_0 | Clustered | 27 | VC_70_0 | 27 | 0.5981 | 1 | 0.5981 | 0 | 0 | 0 | 0.9886 | Ref |
| 15787 | ON540300 | Unassigned | Unassigned | Unassigned | 329_18 | Clustered | 7 | VC_329_18 | 7 | 0.0328 | 1 | 0.0328 | 0 | 0 | 0 | 0.8095 | Ref |
| 15788 | ON540301 | Unassigned | Unassigned | Unassigned | 637_0 | Clustered | 115 | VC_637_0 | 115 | 0.731 | 1 | 0.731 | 0 | 0 | 0 | 0.8764 | Ref |
| 15789 | ON548417 | Unassigned | Unassigned | Unassigned | 475_5 | Clustered | 2 | VC_475_5 | 2 | 0.0294 | 0.93987345 | 0.0276 | 0 | 0 | 0 | 1 | Ref |
| 15790 | ON548418 | Unassigned | Unassigned | Unassigned | 475_10 | Clustered | 2 | VC_475_10 | 2 | 0.0285 | 0.87721192 | 0.025 | 0 | 0 | 0 | 1 | Ref |
| 15791 | ON548419 | Unassigned | Unassigned | Unassigned | 475_5 | Clustered | 2 | VC_475_5 | 2 | 0.0294 | 0.93987345 | 0.0276 | 0 | 0 | 0 | 1 | Ref |
| 15792 | ON548420 | Unassigned | Unassigned | Unassigned | 475_2 | Clustered | 3 | VC_475_2 | 3 | 0.0585 | 0.98883022 | 0.0578 | 0 | 0 | 0 | 1 | Ref |
| 15793 | ON548421 | Unassigned | Unassigned | Unassigned | 475_9 | Clustered/Singleton | 1 | VC_475_9 | 1 | 0 | 1 | 0 | 0 | 0 | 0 | 1 | Ref |
| 15794 | ON548432 | Unassigned | Unassigned | Unassigned | 638_0 | Clustered | 45 | VC_638_0 | 45 | 0.7741 | 1 | 0.7741 | 0 | 0 | 0 | 1 | Ref |
| 15795 | ON548433 | Unassigned | Unassigned | Unassigned | 451_0 | Clustered | 80 | VC_451_0 | 80 | 0.9991 | 1 | 0.9991 | 0 | 0 | 0 | 1 | Ref |
| 15796 | ON550260 | Unassigned | Unassigned | Unassigned | 491_0 | Clustered | 36 | VC_491_0 | 36 | 0.5336 | 1 | 0.5336 | 0 | 0 | 0 | 0.9714 | Ref |
| 15797 | ON551388 | Unassigned | Unassigned | Unassigned | 558_75 | Clustered/Singleton | 1 | VC_558_75 | 1 | 0 | 1 | 0 | 0 | 0 | 0 | 1 | Ref |
| 15798 | ON556632 | Unassigned | Unassigned | Unassigned | 638_0 | Clustered | 45 | VC_638_0 | 45 | 0.7741 | 1 | 0.7741 | 0 | 0 | 0 | 1 | Ref |
| 15799 | ON568193 | Unassigned | Unassigned | Unassigned | 392_0 | Clustered | 126 | VC_392_0 | 126 | 0.9621 | 1 | 0.9621 | 0 | 0 | 0 | 1 | Ref |
| 15800 | ON568341 | Unassigned | Unassigned | Unassigned | 164_11 | Clustered | 10 | VC_164_11 | 10 | 0.1507 | 1 | 0.1507 | 0 | 0 | 0 | 0.9111 | Ref |
| 15801 | ON571554 | Unassigned | Unassigned | Unassigned | 823_0 | Clustered | 124 | VC_823_0 | 124 | 0.9492 | 1 | 0.9492 | 0 | 0 | 0 | 1 | Ref |
| 15802 | ON571632 | Unassigned | Unassigned | Unassigned | 12_2 | Clustered | 19 | VC_12_2 | 19 | 0.2018 | 1 | 0.2018 | 0 | 0 | 0 | 1 | Ref |
| 15803 | ON602723 | Unassigned | Unassigned | Unassigned | 111_0 | Clustered | 91 | VC_111_0 | 91 | 0.9587 | 1 | 0.9587 | 0 | 0 | 0 | 1 | Ref |
| 15804 | ON602724 | Unassigned | Unassigned | Unassigned | 111_0 | Clustered | 91 | VC_111_0 | 91 | 0.9587 | 1 | 0.9587 | 0 | 0 | 0 | 1 | Ref |
| 15805 | ON602725 | Unassigned | Unassigned | Unassigned | 111_0 | Clustered | 91 | VC_111_0 | 91 | 0.9587 | 1 | 0.9587 | 0 | 0 | 0 | 1 | Ref |
| 15806 | ON602726 | Unassigned | Unassigned | Unassigned | 111_0 | Clustered | 91 | VC_111_0 | 91 | 0.9587 | 1 | 0.9587 | 0 | 0 | 0 | 1 | Ref |
| 15807 | ON602727 | Unassigned | Unassigned | Unassigned | 111_0 | Clustered | 91 | VC_111_0 | 91 | 0.9587 | 1 | 0.9587 | 0 | 0 | 0 | 1 | Ref |
| 15808 | ON602728 | Unassigned | Unassigned | Unassigned | 111_0 | Clustered | 91 | VC_111_0 | 91 | 0.9587 | 1 | 0.9587 | 0 | 0 | 0 | 1 | Ref |
| 15809 | ON602729 | Unassigned | Unassigned | Unassigned | 582_3 | Clustered | 12 | VC_582_3 | 12 | 0.2357 | 1 | 0.2357 | 0 | 0 | 0 | 0.8636 | Ref |
| 15810 | ON602730 | Unassigned | Unassigned | Unassigned | 460_0 | Clustered | 19 | VC_460_0 | 19 | 0.3744 | 1 | 0.3744 | 0 | 0 | 0 | 1 | Ref |
| 15811 | ON602732 | Unassigned | Unassigned | Unassigned | 111_0 | Clustered | 91 | VC_111_0 | 91 | 0.9587 | 1 | 0.9587 | 0 | 0 | 0 | 1 | Ref |
| 15812 | ON602733 | Unassigned | Unassigned | Unassigned | 388_0 | Clustered | 4 | VC_388_0 | 4 | 0.0345 | 0.99998812 | 0.0345 | 0 | 0 | 0 | 1 | Ref |
| 15813 | ON602734 | Unassigned | Unassigned | Unassigned | 836_0 | Clustered | 9 | VC_836_0 | 9 | 0.7733 | 1 | 0.7733 | 0 | 0 | 0 | 1 | Ref |
| 15814 | ON602735 | Unassigned | Unassigned | Unassigned | 938_0 | Clustered | 99 | VC_938_0 | 99 | 0.9119 | 1 | 0.9119 | 0 | 0 | 0 | 0.9928 | Ref |
| 15815 | ON602736 | Unassigned | Unassigned | Unassigned | 582_3 | Clustered | 12 | VC_582_3 | 12 | 0.2357 | 1 | 0.2357 | 0 | 0 | 0 | 0.8636 | Ref |
| 15816 | ON602737 | Unassigned | Unassigned | Unassigned | 460_0 | Clustered | 19 | VC_460_0 | 19 | 0.3744 | 1 | 0.3744 | 0 | 0 | 0 | 1 | Ref |
| 15817 | ON602738 | Unassigned | Unassigned | Unassigned | 460_0 | Clustered | 19 | VC_460_0 | 19 | 0.3744 | 1 | 0.3744 | 0 | 0 | 0 | 1 | Ref |
| 15818 | ON602739 | Unassigned | Unassigned | Unassigned | 111_0 | Clustered | 91 | VC_111_0 | 91 | 0.9587 | 1 | 0.9587 | 0 | 0 | 0 | 1 | Ref |
| 15819 | ON602740 | Unassigned | Unassigned | Unassigned | 111_0 | Clustered | 91 | VC_111_0 | 91 | 0.9587 | 1 | 0.9587 | 0 | 0 | 0 | 1 | Ref |
| 15820 | ON602741 | Unassigned | Unassigned | Unassigned | 938_0 | Clustered | 99 | VC_938_0 | 99 | 0.9119 | 1 | 0.9119 | 0 | 0 | 0 | 0.9928 | Ref |
| 15821 | ON602742 | Unassigned | Unassigned | Unassigned | 823_0 | Clustered | 124 | VC_823_0 | 124 | 0.9492 | 1 | 0.9492 | 0 | 0 | 0 | 1 | Ref |
| 15822 | ON602743 | Unassigned | Unassigned | Unassigned | 111_0 | Clustered | 91 | VC_111_0 | 91 | 0.9587 | 1 | 0.9587 | 0 | 0 | 0 | 1 | Ref |
| 15823 | ON602744 | Unassigned | Unassigned | Unassigned | 460_0 | Clustered | 19 | VC_460_0 | 19 | 0.3744 | 1 | 0.3744 | 0 | 0 | 0 | 1 | Ref |
| 15824 | ON602746 | Unassigned | Unassigned | Unassigned | 1049_0 | Clustered | 8 | VC_1049_0 | 8 | 0.6413 | 1 | 0.6413 | 0 | 0 | 0 | 1 | Ref |
| 15825 | ON602747 | Unassigned | Unassigned | Unassigned | 836_0 | Clustered | 9 | VC_836_0 | 9 | 0.7733 | 1 | 0.7733 | 0 | 0 | 0 | 1 | Ref |
| 15826 | ON602748 | Unassigned | Unassigned | Unassigned | 111_0 | Clustered | 91 | VC_111_0 | 91 | 0.9587 | 1 | 0.9587 | 0 | 0 | 0 | 1 | Ref |
| 15827 | ON602749 | Unassigned | Unassigned | Unassigned | 779_0 | Clustered | 37 | VC_779_0 | 37 | 0.5682 | 1 | 0.5682 | 0 | 0 | 0 | 1 | Ref |
| 15828 | ON602750 | Unassigned | Unassigned | Unassigned | 938_0 | Clustered | 99 | VC_938_0 | 99 | 0.9119 | 1 | 0.9119 | 0 | 0 | 0 | 0.9928 | Ref |
| 15829 | ON602751 | Unassigned | Unassigned | Unassigned | 779_0 | Clustered | 37 | VC_779_0 | 37 | 0.5682 | 1 | 0.5682 | 0 | 0 | 0 | 1 | Ref |
| 15830 | ON602752 | Unassigned | Unassigned | Unassigned | 111_0 | Clustered | 91 | VC_111_0 | 91 | 0.9587 | 1 | 0.9587 | 0 | 0 | 0 | 1 | Ref |
| 15831 | ON602753 | Unassigned | Unassigned | Unassigned | 823_0 | Clustered | 124 | VC_823_0 | 124 | 0.9492 | 1 | 0.9492 | 0 | 0 | 0 | 1 | Ref |
| 15832 | ON602754 | Unassigned | Unassigned | Unassigned | 111_0 | Clustered | 91 | VC_111_0 | 91 | 0.9587 | 1 | 0.9587 | 0 | 0 | 0 | 1 | Ref |
| 15833 | ON602755 | Unassigned | Unassigned | Unassigned | 823_0 | Clustered | 124 | VC_823_0 | 124 | 0.9492 | 1 | 0.9492 | 0 | 0 | 0 | 1 | Ref |
| 15834 | ON602756 | Unassigned | Unassigned | Unassigned | 111_0 | Clustered | 91 | VC_111_0 | 91 | 0.9587 | 1 | 0.9587 | 0 | 0 | 0 | 1 | Ref |
| 15835 | ON602757 | Unassigned | Unassigned | Unassigned | 111_0 | Clustered | 91 | VC_111_0 | 91 | 0.9587 | 1 | 0.9587 | 0 | 0 | 0 | 1 | Ref |
| 15836 | ON602758 | Unassigned | Unassigned | Unassigned | 123_0 | Clustered | 23 | VC_123_0 | 23 | 0.817 | 1 | 0.817 | 0 | 0 | 0 | 1 | Ref |
| 15837 | ON602759 | Unassigned | Unassigned | Unassigned | 460_0 | Clustered | 19 | VC_460_0 | 19 | 0.3744 | 1 | 0.3744 | 0 | 0 | 0 | 1 | Ref |
| 15838 | ON602760 | Unassigned | Unassigned | Unassigned | 460_0 | Clustered | 19 | VC_460_0 | 19 | 0.3744 | 1 | 0.3744 | 0 | 0 | 0 | 1 | Ref |
| 15839 | ON602761 | Unassigned | Unassigned | Unassigned | 111_0 | Clustered | 91 | VC_111_0 | 91 | 0.9587 | 1 | 0.9587 | 0 | 0 | 0 | 1 | Ref |
| 15840 | ON602762 | Unassigned | Unassigned | Unassigned | 111_0 | Clustered | 91 | VC_111_0 | 91 | 0.9587 | 1 | 0.9587 | 0 | 0 | 0 | 1 | Ref |
| 15841 | ON602763 | Unassigned | Unassigned | Unassigned | 452_0 | Clustered | 66 | VC_452_0 | 66 | 0.9248 | 1 | 0.9248 | 0 | 0 | 0 | 1 | Ref |
| 15842 | ON602764 | Unassigned | Unassigned | Unassigned | 452_0 | Clustered | 66 | VC_452_0 | 66 | 0.9248 | 1 | 0.9248 | 0 | 0 | 0 | 1 | Ref |
| 15843 | ON602765 | Unassigned | Unassigned | Unassigned | 452_0 | Clustered | 66 | VC_452_0 | 66 | 0.9248 | 1 | 0.9248 | 0 | 0 | 0 | 1 | Ref |
| 15844 | ON602766 | Unassigned | Unassigned | Unassigned | 836_0 | Clustered | 9 | VC_836_0 | 9 | 0.7733 | 1 | 0.7733 | 0 | 0 | 0 | 1 | Ref |
| 15845 | ON602767 | Unassigned | Unassigned | Unassigned | 111_0 | Clustered | 91 | VC_111_0 | 91 | 0.9587 | 1 | 0.9587 | 0 | 0 | 0 | 1 | Ref |
| 15846 | ON630910 | Unassigned | Unassigned | Unassigned | 490_1 | Clustered | 5 | VC_490_1 | 5 | 0.0447 | 0.99999996 | 0.0447 | 0 | 0 | 0 | 1 | Ref |
| 15847 | ON631220 | Unassigned | Unassigned | Unassigned | 344_0 | Clustered | 21 | VC_344_0 | 21 | 0.5436 | 1 | 0.5436 | 0 | 0 | 0 | 1 | Ref |
| 15848 | ON637170 | Unassigned | Unassigned | Unassigned | 192_0 | Clustered | 15 | VC_192_0 | 15 | 0.5071 | 1 | 0.5071 | 0 | 0 | 0 | 1 | Ref |
| 15849 | ON637248 | Unassigned | Unassigned | Unassigned | 97_0 | Clustered | 5 | VC_97_0 | 5 | 0.0696 | 0.98113835 | 0.0683 | 0 | 0 | 0 | 1 | Ref |
| 15850 | ON637250 | Unassigned | Unassigned | Unassigned | 451_0 | Clustered | 80 | VC_451_0 | 80 | 0.9991 | 1 | 0.9991 | 0 | 0 | 0 | 1 | Ref |
| 15851 | ON637251 | Unassigned | Unassigned | Unassigned | 461_0 | Clustered | 96 | VC_461_0 | 96 | 0.9307 | 1 | 0.9307 | 0 | 0 | 0 | 0.9575 | Ref |
| 15852 | ON637759 | Unassigned | Unassigned | Unassigned | 761_0 | Clustered | 67 | VC_761_0 | 67 | 0.8947 | 1 | 0.8947 | 0 | 0 | 0 | 1 | Ref |
| 15853 | ON637760 | Unassigned | Unassigned | Unassigned | 519_0 | Clustered | 39 | VC_519_0 | 39 | 0.8156 | 1 | 0.8156 | 0 | 0 | 0 | 1 | Ref |
| 15854 | ON637761 | Unassigned | Unassigned | Unassigned | 1093_0 | Clustered | 40 | VC_1093_0 | 40 | 0.8754 | 1 | 0.8754 | 0 | 0 | 0 | 1 | Ref |
| 15855 | ON637762 | Unassigned | Unassigned | Unassigned | 954_0 | Clustered | 13 | VC_954_0 | 13 | 0.863 | 1 | 0.863 | 0 | 0 | 0 | 1 | Ref |
| 15856 | ON637763 | Unassigned | Unassigned | Unassigned | 954_0 | Clustered | 13 | VC_954_0 | 13 | 0.863 | 1 | 0.863 | 0 | 0 | 0 | 1 | Ref |
| 15857 | ON637764 | Unassigned | Unassigned | Unassigned | 558_19 | Clustered/Singleton | 1 | VC_558_19 | 1 | 0 | 1 | 0 | 0 | 0 | 0 | 1 | Ref |
| 15858 | ON637765 | Unassigned | Unassigned | Unassigned | 761_0 | Clustered | 67 | VC_761_0 | 67 | 0.8947 | 1 | 0.8947 | 0 | 0 | 0 | 1 | Ref |
| 15859 | ON637766 | Unassigned | Unassigned | Unassigned | 761_0 | Clustered | 67 | VC_761_0 | 67 | 0.8947 | 1 | 0.8947 | 0 | 0 | 0 | 1 | Ref |
| 15860 | ON637767 | Unassigned | Unassigned | Unassigned | 1343_0 | Clustered | 21 | VC_1343_0 | 21 | 0.9843 | 1 | 0.9843 | 0 | 0 | 0 | 1 | Ref |
| 15861 | ON637768 | Unassigned | Unassigned | Unassigned | 558_14 | Clustered | 8 | VC_558_14 | 8 | 0.0228 | 0.99945246 | 0.0228 | 0 | 0 | 0 | 0.8214 | Ref |
| 15862 | ON637769 | Unassigned | Unassigned | Unassigned | 371_11 | Clustered | 31 | VC_371_11 | 31 | 0.0927 | 1 | 0.0927 | 0 | 0 | 0 | 0.7011 | Ref |
| 15863 | ON637770 | Unassigned | Unassigned | Unassigned | 1100_1 | Clustered | 2 | VC_1100_1 | 2 | 0.1223 | 0.94673154 | 0.1158 | 0 | 0 | 0 | 1 | Ref |
| 15864 | ON637771 | Unassigned | Unassigned | Unassigned | 517_1 | Clustered | 108 | VC_517_1 | 108 | 0.8483 | 1 | 0.8483 | 0 | 0 | 0 | 0.9955 | Ref |
| 15865 | ON637772 | Unassigned | Unassigned | Unassigned | 939_0 | Clustered | 15 | VC_939_0 | 15 | 1 | 0 | 0 | 0 | 0 | 0 | 1 | Ref |
| 15866 | ON642070 | Unassigned | Unassigned | Unassigned | 1_0 | Clustered | 54 | VC_1_0 | 54 | 0.8527 | 1 | 0.8527 | 0 | 0 | 0 | 1 | Ref |
| 15867 | ON645337 | Unassigned | Unassigned | Unassigned | 761_0 | Clustered | 67 | VC_761_0 | 67 | 0.8947 | 1 | 0.8947 | 0 | 0 | 0 | 1 | Ref |
| 15868 | ON645338 | Unassigned | Unassigned | Unassigned | 1164_0 | Clustered | 25 | VC_1164_0 | 25 | 0.7312 | 1 | 0.7312 | 0 | 0 | 0 | 1 | Ref |
| 15869 | ON645339 | Unassigned | Unassigned | Unassigned | 1072_5 | Clustered | 3 | VC_1072_5 | 3 | 0.0475 | 0.98997321 | 0.047 | 0 | 0 | 0 | 1 | Ref |
| 15870 | ON645341 | Unassigned | Unassigned | Unassigned | 1344_0 | Clustered | 78 | VC_1344_0 | 78 | 0.9732 | 1 | 0.9732 | 0 | 0 | 0 | 1 | Ref |
| 15871 | ON645342 | Unassigned | Unassigned | Unassigned | 234_0 | Clustered | 21 | VC_234_0 | 21 | 0.8355 | 1 | 0.8355 | 0 | 0 | 0 | 1 | Ref |
| 15872 | ON645343 | Unassigned | Unassigned | Unassigned | 1175_0 | Clustered | 22 | VC_1175_0 | 22 | 0.4562 | 1 | 0.4562 | 0 | 0 | 0 | 1 | Ref |
| 15873 | ON645344 | Unassigned | Unassigned | Unassigned | 522_0 | Clustered | 3 | VC_522_0 | 3 | 0.0836 | 0.9985266 | 0.0835 | 0 | 0 | 0 | 1 | Ref |
| 15874 | ON645345 | Unassigned | Unassigned | Unassigned | 1344_0 | Clustered | 78 | VC_1344_0 | 78 | 0.9732 | 1 | 0.9732 | 0 | 0 | 0 | 1 | Ref |

| | | | | | | | | | | | | | | | | | |
|---|---|---|---|---|---|---|---|---|---|---|---|---|---|---|---|---|---|
| 15875 | ON645346 | Unassigned | Unassigned | Unassigned | 1034_0 | Clustered | 2 | VC_1034_0 | 2 | 0.071 | 0.95326364 | 0.0677 | 0 | 0 | 0 | 1 | Ref |
| 15876 | ON645936 | Unassigned | Unassigned | Unassigned | 283_178 | Clustered/Singleton | 1 | VC_283_178 | 1 | 0 | 1 | 0 | 0 | 0 | 0 | 1 | Ref |
| 15877 | ON653033 | Unassigned | Unassigned | Unassigned | 77_0 | Clustered | 20 | VC_77_0 | 20 | 0.9657 | 1 | 0.9657 | 0 | 0 | 0 | 1 | Ref |
| 15878 | ON677304 | Unassigned | Unassigned | Unassigned | 325_34 | Clustered/Singleton | 1 | VC_325_34 | 1 | 0 | 1 | 0 | 0 | 0 | 0 | 1 | Ref |
| 15879 | ON677538 | Unassigned | Unassigned | Unassigned | 778_0 | Clustered | 4 | VC_778_0 | 4 | 0.8115 | 0.99985435 | 0.8114 | 0 | 0 | 0 | 1 | Ref |
| 15880 | ON687735 | Unassigned | Unassigned | Unassigned | 236_0 | Clustered | 72 | VC_236_0 | 72 | 0.8104 | 1 | 0.8104 | 0 | 0 | 0 | 1 | Ref |
| 15881 | ON687736 | Unassigned | Unassigned | Unassigned | 236_0 | Clustered | 72 | VC_236_0 | 72 | 0.8104 | 1 | 0.8104 | 0 | 0 | 0 | 1 | Ref |
| 15882 | ON699007 | Unassigned | Unassigned | Unassigned | 143_0 | Clustered | 2 | VC_143_0 | 2 | 0.1189 | 0.94995671 | 0.1129 | 0 | 0 | 0 | 1 | Ref |
| 15883 | ON706965 | Unassigned | Unassigned | Unassigned | 328_0 | Clustered | 11 | VC_328_0 | 11 | 0.4707 | 1 | 0.4707 | 0 | 0 | 0 | 1 | Ref |
| 15884 | ON711395 | Unassigned | Unassigned | Unassigned | 942_0 | Clustered | 36 | VC_942_0 | 36 | 0.7848 | 1 | 0.7848 | 0 | 0 | 0 | 1 | Ref |
| 15885 | ON711490 | Unassigned | Unassigned | Unassigned | 316_0 | Clustered | 3 | VC_316_0 | 3 | 0.774 | 0.99820498 | 0.7726 | 0 | 0 | 0 | 1 | Ref |
| 15886 | ON712643 | Unassigned | Unassigned | Unassigned | 849_7 | Clustered/Singleton | 1 | VC_849_7 | 1 | 0 | 1 | 0 | 0 | 0 | 0 | 1 | Ref |
| 15887 | ON714422 | Unassigned | Unassigned | Unassigned | 642_2 | Clustered | 6 | VC_642_2 | 6 | 0.1679 | 1 | 0.1679 | 0 | 0 | 0 | 1 | Ref |
| 15888 | ON715521 | Unassigned | Unassigned | Unassigned | 387_0 | Clustered | 3 | VC_387_0 | 3 | 0.0424 | 0.99861589 | 0.0423 | 0 | 0 | 0 | 1 | Ref |
| 15889 | ON720975 | Unassigned | Unassigned | Unassigned | 283_162 | Clustered/Singleton | 1 | VC_283_162 | 1 | 0 | 1 | 0 | 0 | 0 | 0 | 1 | Ref |
| 15890 | ON720976 | Unassigned | Unassigned | Unassigned | 540_0 | Clustered | 15 | VC_540_0 | 15 | 0.2866 | 1 | 0.2866 | 0 | 0 | 0 | 1 | Ref |
| 15891 | ON720977 | Unassigned | Unassigned | Unassigned | 637_0 | Clustered | 115 | VC_637_0 | 115 | 0.731 | 1 | 0.731 | 0 | 0 | 0 | 0.8764 | Ref |
| 15892 | ON721384 | Unassigned | Unassigned | Unassigned | 1009_0 | Clustered | 3 | VC_1009_0 | 3 | 0.127 | 0.99814461 | 0.1267 | 0 | 0 | 0 | 1 | Ref |
| 15893 | ON721385 | Unassigned | Unassigned | Unassigned | 1009_0 | Clustered | 3 | VC_1009_0 | 3 | 0.127 | 0.99814461 | 0.1267 | 0 | 0 | 0 | 1 | Ref |
| 15894 | ON724008 | Unassigned | Unassigned | Unassigned | 1176_0 | Clustered | 41 | VC_1176_0 | 41 | 0.9558 | 1 | 0.9558 | 0 | 0 | 0 | 0.9976 | Ref |
| 15895 | ON724009 | Unassigned | Unassigned | Unassigned | 1184_0 | Clustered | 5 | VC_1184_0 | 5 | 0.6591 | 0.99999982 | 0.6591 | 0 | 0 | 0 | 1 | Ref |
| 15896 | ON724010 | Unassigned | Unassigned | Unassigned | 744_0 | Clustered | 10 | VC_744_0 | 10 | 0.7049 | 1 | 0.7049 | 0 | 0 | 0 | 1 | Ref |
| 15897 | ON724011 | Unassigned | Unassigned | Unassigned | 1176_0 | Clustered | 41 | VC_1176_0 | 41 | 0.9558 | 1 | 0.9558 | 0 | 0 | 0 | 0.9976 | Ref |
| 15898 | ON724012 | Unassigned | Unassigned | Unassigned | 1176_0 | Clustered | 41 | VC_1176_0 | 41 | 0.9558 | 1 | 0.9558 | 0 | 0 | 0 | 0.9976 | Ref |
| 15899 | ON724013 | Unassigned | Unassigned | Unassigned | 236_0 | Clustered | 72 | VC_236_0 | 72 | 0.8104 | 1 | 0.8104 | 0 | 0 | 0 | 1 | Ref |
| 15900 | ON724014 | Unassigned | Unassigned | Unassigned | 1093_0 | Clustered | 40 | VC_1093_0 | 40 | 0.8754 | 1 | 0.8754 | 0 | 0 | 0 | 1 | Ref |
| 15901 | ON724015 | Unassigned | Unassigned | Unassigned | 866_1 | Clustered | 20 | VC_866_1 | 20 | 0.4814 | 1 | 0.4814 | 0 | 0 | 0 | 1 | Ref |
| 15902 | ON724016 | Unassigned | Unassigned | Unassigned | 225_0 | Clustered | 32 | VC_225_0 | 32 | 0.848 | 1 | 0.848 | 0 | 0 | 0 | 0.996 | Ref |
| 15903 | ON724017 | Unassigned | Unassigned | Unassigned | 1036_0 | Clustered | 11 | VC_1036_0 | 11 | 0.656 | 1 | 0.656 | 0 | 0 | 0 | 0.8727 | Ref |
| 15904 | ON724018 | Unassigned | Unassigned | Unassigned | 1234_0 | Clustered | 44 | VC_1234_0 | 44 | 1 | 1 | 1 | 0 | 0 | 0 | 1 | Ref |
| 15905 | ON754979 | Unassigned | Unassigned | Unassigned | 386_0 | Clustered | 49 | VC_386_0 | 49 | 0.7941 | 1 | 0.7941 | 0 | 0 | 0 | 1 | Ref |
| 15906 | ON755181 | Unassigned | Unassigned | Unassigned | 889_0 | Clustered | 16 | VC_889_0 | 16 | 0.2969 | 1 | 0.2969 | 0 | 0 | 0 | 1 | Ref |
| 15907 | ON755182 | Unassigned | Unassigned | Unassigned | 1344_0 | Clustered | 78 | VC_1344_0 | 78 | 0.9732 | 1 | 0.9732 | 0 | 0 | 0 | 1 | Ref |
| 15908 | ON755183 | Unassigned | Unassigned | Unassigned | 1343_0 | Clustered | 21 | VC_1343_0 | 21 | 0.9843 | 1 | 0.9843 | 0 | 0 | 0 | 1 | Ref |
| 15909 | ON755184 | Unassigned | Unassigned | Unassigned | 1096_0 | Clustered | 5 | VC_1096_0 | 5 | 0.8566 | 0.99869977 | 0.8555 | 0 | 0 | 0 | 1 | Ref |
| 15910 | ON755185 | Unassigned | Unassigned | Unassigned | 572_0 | Clustered | 16 | VC_572_0 | 16 | 0.7286 | 1 | 0.7286 | 0 | 0 | 0 | 1 | Ref |
| 15911 | ON755186 | Unassigned | Unassigned | Unassigned | 1095_0 | Clustered | 16 | VC_1095_0 | 16 | 0.458 | 1 | 0.458 | 0 | 0 | 0 | 1 | Ref |
| 15912 | ON755187 | Unassigned | Unassigned | Unassigned | 572_0 | Clustered | 16 | VC_572_0 | 16 | 0.7286 | 1 | 0.7286 | 0 | 0 | 0 | 1 | Ref |
| 15913 | ON755188 | Unassigned | Unassigned | Unassigned | 1344_0 | Clustered | 78 | VC_1344_0 | 78 | 0.9732 | 1 | 0.9732 | 0 | 0 | 0 | 1 | Ref |
| 15914 | ON755189 | Unassigned | Unassigned | Unassigned | 558_16 | Clustered/Singleton | 1 | VC_558_16 | 1 | 0 | 1 | 0 | 0 | 0 | 0 | 1 | Ref |
| 15915 | ON758385 | Unassigned | Unassigned | Unassigned | 506_1 | Clustered/Singleton | 1 | VC_506_1 | 1 | 0 | 1 | 0 | 0 | 0 | 0 | 1 | Ref |
| 15916 | ON778007 | Unassigned | Unassigned | Unassigned | 297_0 | Clustered | 5 | VC_297_0 | 5 | 0.1056 | 0.99999993 | 0.1056 | 0 | 0 | 0 | 0.9 | Ref |
| 15917 | ON778463 | Unassigned | Unassigned | Unassigned | 792_0 | Clustered | 35 | VC_792_0 | 35 | 0.9828 | 1 | 0.9828 | 0 | 0 | 0 | 1 | Ref |
| 15918 | ON782130 | Unassigned | Unassigned | Unassigned | 228_0 | Clustered | 60 | VC_228_0 | 60 | 0.942 | 1 | 0.942 | 0 | 0 | 0 | 1 | Ref |
| 15919 | ON782582 | Unassigned | Unassigned | Unassigned | 283_6 | Clustered/Singleton | 1 | VC_283_6 | 1 | 0 | 1 | 0 | 0 | 0 | 0 | 1 | Ref |
| 15920 | ON807605 | Unassigned | Unassigned | Unassigned | 123_0 | Clustered | 23 | VC_123_0 | 23 | 0.817 | 1 | 0.817 | 0 | 0 | 0 | 1 | Ref |
| 15921 | ON809559 | Unassigned | Unassigned | Unassigned | 111_0 | Clustered | 91 | VC_111_0 | 91 | 0.9587 | 1 | 0.9587 | 0 | 0 | 0 | 1 | Ref |
| 15922 | ON809560 | Unassigned | Unassigned | Unassigned | 123_0 | Clustered | 23 | VC_123_0 | 23 | 0.817 | 1 | 0.817 | 0 | 0 | 0 | 1 | Ref |
| 15923 | ON814134 | Unassigned | Unassigned | Unassigned | 153_0 | Clustered | 25 | VC_153_0 | 25 | 0.7729 | 1 | 0.7729 | 0 | 0 | 0 | 1 | Ref |
| 15924 | ON814135 | Unassigned | Unassigned | Unassigned | 153_0 | Clustered | 25 | VC_153_0 | 25 | 0.7729 | 1 | 0.7729 | 0 | 0 | 0 | 1 | Ref |
| 15925 | ON814136 | Unassigned | Unassigned | Unassigned | 153_0 | Clustered | 25 | VC_153_0 | 25 | 0.7729 | 1 | 0.7729 | 0 | 0 | 0 | 1 | Ref |
| 15926 | ON815901 | Unassigned | Unassigned | Unassigned | 838_0 | Clustered | 29 | VC_838_0 | 29 | 0.8611 | 1 | 0.8611 | 0 | 0 | 0 | 1 | Ref |
| 15927 | ON815902 | Unassigned | Unassigned | Unassigned | 838_0 | Clustered | 29 | VC_838_0 | 29 | 0.8611 | 1 | 0.8611 | 0 | 0 | 0 | 1 | Ref |
| 15928 | ON815903 | Unassigned | Unassigned | Unassigned | 838_0 | Clustered | 29 | VC_838_0 | 29 | 0.8611 | 1 | 0.8611 | 0 | 0 | 0 | 1 | Ref |
| 15929 | ON843697 | Unassigned | Unassigned | Unassigned | 584_0 | Clustered | 28 | VC_584_0 | 28 | 0.7704 | 1 | 0.7704 | 0 | 0 | 0 | 1 | Ref |
| 15930 | ON853583 | Unassigned | Unassigned | Unassigned | 125_0 | Clustered | 17 | VC_125_0 | 17 | 0.9244 | 1 | 0.9244 | 0 | 0 | 0 | 1 | Ref |
| 15931 | ON856257 | Unassigned | Unassigned | Unassigned | 8_0 | Clustered | 58 | VC_8_0 | 58 | 0.8306 | 1 | 0.8306 | 0 | 0 | 0 | 0.9946 | Ref |
| 15932 | ON856258 | Unassigned | Unassigned | Unassigned | 8_0 | Clustered | 58 | VC_8_0 | 58 | 0.8306 | 1 | 0.8306 | 0 | 0 | 0 | 0.9946 | Ref |
| 15933 | ON857925 | Unassigned | Unassigned | Unassigned | 79_7 | Clustered | 33 | VC_79_7 | 33 | 0.216 | 1 | 0.216 | 0 | 0 | 0 | 0.8409 | Ref |
| 15934 | ON857926 | Unassigned | Unassigned | Unassigned | 423_0 | Clustered | 45 | VC_423_0 | 45 | 0.9484 | 1 | 0.9484 | 0 | 0 | 0 | 1 | Ref |
| 15935 | ON857927 | Unassigned | Unassigned | Unassigned | 472_0 | Clustered | 47 | VC_472_0 | 47 | 0.9025 | 1 | 0.9025 | 0 | 0 | 0 | 1 | Ref |
| 15936 | ON857928 | Unassigned | Unassigned | Unassigned | 423_0 | Clustered | 45 | VC_423_0 | 45 | 0.9484 | 1 | 0.9484 | 0 | 0 | 0 | 1 | Ref |
| 15937 | ON857929 | Unassigned | Unassigned | Unassigned | 79_7 | Clustered | 33 | VC_79_7 | 33 | 0.216 | 1 | 0.216 | 0 | 0 | 0 | 0.8409 | Ref |
| 15938 | ON857930 | Unassigned | Unassigned | Unassigned | 79_0 | Clustered | 64 | VC_79_0 | 64 | 0.4185 | 1 | 0.4185 | 0 | 0 | 0 | 0.7649 | Ref |
| 15939 | ON857931 | Unassigned | Unassigned | Unassigned | 295_0 | Clustered | 2 | VC_295_0 | 2 | 0.0336 | 0.94464231 | 0.0317 | 0 | 0 | 0 | 1 | Ref |
| 15940 | ON857932 | Unassigned | Unassigned | Unassigned | 755_0 | Clustered | 9 | VC_755_0 | 9 | 0.4489 | 1 | 0.4489 | 0 | 0 | 0 | 1 | Ref |
| 15941 | ON857933 | Unassigned | Unassigned | Unassigned | 423_0 | Clustered | 45 | VC_423_0 | 45 | 0.9484 | 1 | 0.9484 | 0 | 0 | 0 | 1 | Ref |
| 15942 | ON857934 | Unassigned | Unassigned | Unassigned | 79_0 | Clustered | 64 | VC_79_0 | 64 | 0.4185 | 1 | 0.4185 | 0 | 0 | 0 | 0.7649 | Ref |
| 15943 | ON857935 | Unassigned | Unassigned | Unassigned | 423_0 | Clustered | 45 | VC_423_0 | 45 | 0.9484 | 1 | 0.9484 | 0 | 0 | 0 | 1 | Ref |
| 15944 | ON857936 | Unassigned | Unassigned | Unassigned | 472_0 | Clustered | 47 | VC_472_0 | 47 | 0.9025 | 1 | 0.9025 | 0 | 0 | 0 | 1 | Ref |
| 15945 | ON857937 | Unassigned | Unassigned | Unassigned | 79_7 | Clustered | 33 | VC_79_7 | 33 | 0.216 | 1 | 0.216 | 0 | 0 | 0 | 0.8409 | Ref |
| 15946 | ON857938 | Unassigned | Unassigned | Unassigned | 79_7 | Clustered | 33 | VC_79_7 | 33 | 0.216 | 1 | 0.216 | 0 | 0 | 0 | 0.8409 | Ref |
| 15947 | ON857939 | Unassigned | Unassigned | Unassigned | 1028_0 | Clustered | 8 | VC_1028_0 | 8 | 0.9323 | 1 | 0.9323 | 0 | 0 | 0 | 1 | Ref |
| 15948 | ON857940 | Unassigned | Unassigned | Unassigned | 176_1 | Clustered | 2 | VC_176_1 | 2 | 0.0363 | 0.95657568 | 0.0347 | 0 | 0 | 0 | 1 | Ref |
| 15949 | ON857941 | Unassigned | Unassigned | Unassigned | 612_0 | Clustered | 9 | VC_612_0 | 9 | 0.9545 | 1 | 0.9545 | 0 | 0 | 0 | 1 | Ref |
| 15950 | ON857942 | Unassigned | Unassigned | Unassigned | 79_0 | Clustered | 64 | VC_79_0 | 64 | 0.4185 | 1 | 0.4185 | 0 | 0 | 0 | 0.7649 | Ref |
| 15951 | ON857943 | Unassigned | Unassigned | Unassigned | 838_0 | Clustered | 29 | VC_838_0 | 29 | 0.8611 | 1 | 0.8611 | 0 | 0 | 0 | 1 | Ref |
| 15952 | ON862890 | Unassigned | Unassigned | Unassigned | 131_1 | Clustered | 6 | VC_131_1 | 6 | 0.057 | 0.99999999 | 0.057 | 0 | 0 | 0 | 1 | Ref |
| 15953 | ON864052 | Unassigned | Unassigned | Unassigned | 837_0 | Clustered | 29 | VC_837_0 | 29 | 0.9955 | 1 | 0.9955 | 0 | 0 | 0 | 1 | Ref |
| 15954 | ON866946 | Unassigned | Unassigned | Unassigned | 153_0 | Clustered | 25 | VC_153_0 | 25 | 0.7729 | 1 | 0.7729 | 0 | 0 | 0 | 1 | Ref |
| 15955 | ON868915 | Unassigned | Unassigned | Unassigned | 329_19 | Clustered/Singleton | 1 | VC_329_19 | 1 | 0 | 1 | 0 | 0 | 0 | 0 | 1 | Ref |
| 15956 | ON872379 | Unassigned | Unassigned | Unassigned | 837_0 | Clustered | 29 | VC_837_0 | 29 | 0.9955 | 1 | 0.9955 | 0 | 0 | 0 | 1 | Ref |
| 15957 | ON880501 | Unassigned | Unassigned | Unassigned | 823_0 | Clustered | 124 | VC_823_0 | 124 | 0.9492 | 1 | 0.9492 | 0 | 0 | 0 | 1 | Ref |
| 15958 | ON881243 | Unassigned | Unassigned | Unassigned | 475_3 | Clustered/Singleton | 1 | VC_475_3 | 1 | 0 | 1 | 0 | 0 | 0 | 0 | 1 | Ref |
| 15959 | ON881905 | Unassigned | Unassigned | Unassigned | 938_0 | Clustered | 99 | VC_938_0 | 99 | 0.9119 | 1 | 0.9119 | 0 | 0 | 0 | 0.9928 | Ref |
| 15960 | ON911538 | Unassigned | Unassigned | Unassigned | 749_0 | Clustered | 40 | VC_749_0 | 40 | 0.7517 | 1 | 0.7517 | 0 | 0 | 0 | 1 | Ref |
| 15961 | ON911539 | Unassigned | Unassigned | Unassigned | 74_0 | Clustered | 15 | VC_74_0 | 15 | 0.5204 | 1 | 0.5204 | 0 | 0 | 0 | 1 | Ref |
| 15962 | ON911540 | Unassigned | Unassigned | Unassigned | 465_0 | Clustered | 7 | VC_465_0 | 7 | 0.3322 | 1 | 0.3322 | 0 | 0 | 0 | 1 | Ref |
| 15963 | ON911714 | Unassigned | Unassigned | Unassigned | 9_0 | Clustered | 38 | VC_9_0 | 38 | 0.4895 | 1 | 0.4895 | 0 | 0 | 0 | 1 | Ref |
| 15964 | ON911715 | Unassigned | Unassigned | Unassigned | 9_0 | Clustered | 38 | VC_9_0 | 38 | 0.4895 | 1 | 0.4895 | 0 | 0 | 0 | 1 | Ref |
| 15965 | ON911716 | Unassigned | Unassigned | Unassigned | 9_0 | Clustered | 38 | VC_9_0 | 38 | 0.4895 | 1 | 0.4895 | 0 | 0 | 0 | 1 | Ref |
| 15966 | ON911717 | Unassigned | Unassigned | Unassigned | 9_0 | Clustered | 38 | VC_9_0 | 38 | 0.4895 | 1 | 0.4895 | 0 | 0 | 0 | 1 | Ref |
| 15967 | ON911718 | Unassigned | Unassigned | Unassigned | 9_0 | Clustered | 38 | VC_9_0 | 38 | 0.4895 | 1 | 0.4895 | 0 | 0 | 0 | 1 | Ref |
| 15968 | ON922917 | Unassigned | Unassigned | Unassigned | 916_0 | Clustered | 4 | VC_916_0 | 4 | 0.8472 | 0.99985574 | 0.8471 | 0 | 0 | 0 | 1 | Ref |
| 15969 | ON922918 | Unassigned | Unassigned | Unassigned | 722_0 | Clustered | 13 | VC_722_0 | 13 | 0.5104 | 1 | 0.5104 | 0 | 0 | 0 | 1 | Ref |
| 15970 | ON922919 | Unassigned | Unassigned | Unassigned | 779_0 | Clustered | 37 | VC_779_0 | 37 | 0.5682 | 1 | 0.5682 | 0 | 0 | 0 | 1 | Ref |
| 15971 | ON922920 | Unassigned | Unassigned | Unassigned | 386_0 | Clustered | 49 | VC_386_0 | 49 | 0.7941 | 1 | 0.7941 | 0 | 0 | 0 | 1 | Ref |
| 15972 | ON922990 | Unassigned | Unassigned | Unassigned | 493_9 | Clustered/Singleton | 1 | VC_493_9 | 1 | 0 | 1 | 0 | 0 | 0 | 0 | 1 | Ref |
| 15973 | ON932078 | Unassigned | Unassigned | Unassigned | 74_0 | Clustered | 15 | VC_74_0 | 15 | 0.5204 | 1 | 0.5204 | 0 | 0 | 0 | 1 | Ref |
| 15974 | ON932079 | Unassigned | Unassigned | Unassigned | 169_0 | Clustered | 20 | VC_169_0 | 20 | 0.7529 | 1 | 0.7529 | 0 | 0 | 0 | 1 | Ref |
| 15975 | ON932080 | Unassigned | Unassigned | Unassigned | 169_0 | Clustered | 20 | VC_169_0 | 20 | 0.7529 | 1 | 0.7529 | 0 | 0 | 0 | 1 | Ref |
| 15976 | ON932081 | Unassigned | Unassigned | Unassigned | 74_0 | Clustered | 15 | VC_74_0 | 15 | 0.5204 | 1 | 0.5204 | 0 | 0 | 0 | 1 | Ref |
| 15977 | ON932082 | Unassigned | Unassigned | Unassigned | 749_0 | Clustered | 40 | VC_749_0 | 40 | 0.7517 | 1 | 0.7517 | 0 | 0 | 0 | 1 | Ref |
| 15978 | ON932083 | Unassigned | Unassigned | Unassigned | 74_0 | Clustered | 15 | VC_74_0 | 15 | 0.5204 | 1 | 0.5204 | 0 | 0 | 0 | 1 | Ref |
| 15979 | ON932084 | Unassigned | Unassigned | Unassigned | 169_0 | Clustered | 20 | VC_169_0 | 20 | 0.7529 | 1 | 0.7529 | 0 | 0 | 0 | 1 | Ref |
| 15980 | ON932085 | Unassigned | Unassigned | Unassigned | 749_0 | Clustered | 40 | VC_749_0 | 40 | 0.7517 | 1 | 0.7517 | 0 | 0 | 0 | 1 | Ref |
| 15981 | ON933637 | Unassigned | Unassigned | Unassigned | 916_0 | Clustered | 4 | VC_916_0 | 4 | 0.8472 | 0.99985574 | 0.8471 | 0 | 0 | 0 | 1 | Ref |
| 15982 | ON950090 | Unassigned | Unassigned | Unassigned | 385_0 | Clustered | 51 | VC_385_0 | 51 | 0.8754 | 1 | 0.8754 | 0 | 0 | 0 | 1 | Ref |
| 15983 | ON960072 | Unassigned | Unassigned | Unassigned | 74_0 | Clustered | 15 | VC_74_0 | 15 | 0.5204 | 1 | 0.5204 | 0 | 0 | 0 | 1 | Ref |
| 15984 | ON970561 | Unassigned | Unassigned | Unassigned | 1035_0 | Clustered | 6 | VC_1035_0 | 6 | 0.4854 | 1 | 0.4854 | 0 | 0 | 0 | 1 | Ref |
| 15985 | ON970562 | Unassigned | Unassigned | Unassigned | 520_43 | Clustered | 3 | VC_520_43 | 3 | 0.0193 | 0.99837043 | 0.0193 | 0 | 0 | 0 | 1 | Ref |
| 15986 | ON970563 | Unassigned | Unassigned | Unassigned | 1352_0 | Clustered | 31 | VC_1352_0 | 31 | 0.849 | 1 | 0.849 | 0 | 0 | 0 | 1 | Ref |
| 15987 | ON970564 | Unassigned | Unassigned | Unassigned | 1044_0 | Clustered | 3 | VC_1044_0 | 3 | 0.0843 | 0.99852363 | 0.0842 | 0 | 0 | 0 | 1 | Ref |
| 15988 | ON970565 | Unassigned | Unassigned | Unassigned | 517_1 | Clustered | 108 | VC_517_1 | 108 | 0.8483 | 1 | 0.8483 | 0 | 0 | 0 | 0.9955 | Ref |
| 15989 | ON970566 | Unassigned | Unassigned | Unassigned | 520_62 | Clustered/Singleton | 1 | VC_520_62 | 1 | 0 | 1 | 0 | 0 | 0 | 0 | 1 | Ref |
| 15990 | ON970567 | Unassigned | Unassigned | Unassigned | 1176_0 | Clustered | 41 | VC_1176_0 | 41 | 0.9558 | 1 | 0.9558 | 0 | 0 | 0 | 0.9976 | Ref |
| 15991 | ON970568 | Unassigned | Unassigned | Unassigned | 1206_0 | Clustered | 8 | VC_1206_0 | 8 | 0.1913 | 1 | 0.1913 | 0 | 0 | 0 | 1 | Ref |
| 15992 | ON970569 | Unassigned | Unassigned | Unassigned | 1224_0 | Clustered | 19 | VC_1224_0 | 19 | 1 | 1 | 1 | 0 | 0 | 0 | 1 | Ref |
| 15993 | ON970571 | Unassigned | Unassigned | Unassigned | 1163_0 | Clustered | 28 | VC_1163_0 | 28 | 0.4923 | 1 | 0.4923 | 0 | 0 | 0 | 1 | Ref |
| 15994 | ON970572 | Unassigned | Unassigned | Unassigned | 799_1 | Clustered | 27 | VC_799_1 | 27 | 0.8611 | 1 | 0.8611 | 0 | 0 | 0 | 1 | Ref |
| 15995 | ON970573 | Unassigned | Unassigned | Unassigned | 1344_0 | Clustered | 78 | VC_1344_0 | 78 | 0.9732 | 1 | 0.9732 | 0 | 0 | 0 | 1 | Ref |
| 15996 | ON970574 | Unassigned | Unassigned | Unassigned | 519_0 | Clustered | 39 | VC_519_0 | 39 | 0.8156 | 1 | 0.8156 | 0 | 0 | 0 | 1 | Ref |
| 15997 | ON970575 | Unassigned | Unassigned | Unassigned | 372_0 | Clustered | 24 | VC_372_0 | 24 | 0.5539 | 1 | 0.5539 | 0 | 0 | 0 | 1 | Ref |
| 15998 | ON970576 | Unassigned | Unassigned | Unassigned | 919_0 | Clustered | 39 | VC_919_0 | 39 | 0.9069 | 1 | 0.9069 | 0 | 0 | 0 | 1 | Ref |
| 15999 | ON970577 | Unassigned | Unassigned | Unassigned | 371_9 | Clustered | 4 | VC_371_9 | 4 | 0.0109 | 0.9999766 | 0.0109 | 0 | 0 | 0 | 0.6667 | Ref |
| 16000 | ON970578 | Unassigned | Unassigned | Unassigned | 955_0 | Clustered | 3 | VC_955_0 | 3 | 0.7745 | 0.99134622 | 0.7678 | 0 | 0 | 0 | 1 | Ref |
| 16001 | ON970579 | Unassigned | Unassigned | Unassigned | 1071_0 | Clustered | 20 | VC_1071_0 | 20 | 0.3268 | 1 | 0.3268 | 0 | 0 | 0 | 0.9842 | Ref |
| 16002 | ON970580 | Unassigned | Unassigned | Unassigned | 950_0 | Clustered | 38 | VC_950_0 | 38 | 0.9443 | 1 | 0.9443 | 0 | 0 | 0 | 1 | Ref |
| 16003 | ON970581 | Unassigned | Unassigned | Unassigned | 1240_0 | Clustered | 82 | VC_1240_0 | 82 | 0.7945 | 1 | 0.7945 | 0 | 0 | 0 | 1 | Ref |
| 16004 | ON970582 | Unassigned | Unassigned | Unassigned | 866_1 | Clustered | 20 | VC_866_1 | 20 | 0.4814 | 1 | 0.4814 | 0 | 0 | 0 | 1 | Ref |
| 16005 | ON970583 | Unassigned | Unassigned | Unassigned | 1162_0 | Clustered | 10 | VC_1162_0 | 10 | 0.5715 | 1 | 0.5715 | 0 | 0 | 0 | 1 | Ref |
| 16006 | ON970584 | Unassigned | Unassigned | Unassigned | 1360_0 | Clustered | 31 | VC_1360_0 | 31 | 0.8837 | 1 | 0.8837 | 0 | 0 | 0 | 1 | Ref |
| 16007 | ON970585 | Unassigned | Unassigned | Unassigned | 520_84 | Clustered/Singleton | 1 | VC_520_84 | 1 | 0 | 1 | 0 | 0 | 0 | 0 | 1 | Ref |
| 16008 | ON970586 | Unassigned | Unassigned | Unassigned | 325_0 | Clustered | 2 | VC_325_0 | 2 | 0.0041 | 0.95574439 | 0.0039 | 0 | 0 | 0 | 1 | Ref |
| 16009 | ON970587 | Unassigned | Unassigned | Unassigned | 508_0 | Clustered | 32 | VC_508_0 | 32 | 0.906 | 1 | 0.906 | 0 | 0 | 0 | 1 | Ref |
| 16010 | ON970588 | Unassigned | Unassigned | Unassigned | 866_1 | Clustered | 20 | VC_866_1 | 20 | 0.4814 | 1 | 0.4814 | 0 | 0 | 0 | 1 | Ref |
| 16011 | ON970589 | Unassigned | Unassigned | Unassigned | 1344_0 | Clustered | 78 | VC_1344_0 | 78 | 0.9732 | 1 | 0.9732 | 0 | 0 | 0 | 1 | Ref |
| 16012 | ON970590 | Unassigned | Unassigned | Unassigned | 1071_0 | Clustered | 20 | VC_1071_0 | 20 | 0.3268 | 1 | 0.3268 | 0 | 0 | 0 | 0.9842 | Ref |
| 16013 | ON970591 | Unassigned | Unassigned | Unassigned | 1071_2 | Clustered | 5 | VC_1071_2 | 5 | 0.0534 | 0.99997362 | 0.0534 | 0 | 0 | 0 | 0.9 | Ref |
| 16014 | ON970592 | Unassigned | Unassigned | Unassigned | 1350_0 | Clustered | 2 | VC_1350_0 | 2 | 0.1631 | 0.95304852 | 0.1555 | 0 | 0 | 0 | 1 | Ref |
| 16015 | ON970593 | Unassigned | Unassigned | Unassigned | 508_0 | Clustered | 32 | VC_508_0 | 32 | 0.906 | 1 | 0.906 | 0 | 0 | 0 | 1 | Ref |
| 16016 | ON970594 | Unassigned | Unassigned | Unassigned | 371_7 | Clustered/Singleton | 1 | VC_371_7 | 1 | 0 | 1 | 0 | 0 | 0 | 0 | 1 | Ref |
| 16017 | ON970595 | Unassigned | Unassigned | Unassigned | 520_58 | Clustered | 3 | VC_520_58 | 3 | 0.021 | 0.9985993 | 0.021 | 0 | 0 | 0 | 1 | Ref |
| 16018 | ON970596 | Unassigned | Unassigned | Unassigned | 866_1 | Clustered | 20 | VC_866_1 | 20 | 0.4814 | 1 | 0.4814 | 0 | 0 | 0 | 1 | Ref |
| 16019 | ON970597 | Unassigned | Unassigned | Unassigned | 551_6 | Clustered | 22 | VC_551_6 | 22 | 0.1214 | 1 | 0.1214 | 0 | 0 | 0 | 0.8095 | Ref |
| 16020 | ON970598 | Unassigned | Unassigned | Unassigned | 1036_0 | Clustered | 11 | VC_1036_0 | 11 | 0.656 | 1 | 0.656 | 0 | 0 | 0 | 0.8727 | Ref |
| 16021 | ON970599 | Unassigned | Unassigned | Unassigned | 919_0 | Clustered | 39 | VC_919_0 | 39 | 0.9069 | 1 | 0.9069 | 0 | 0 | 0 | 1 | Ref |
| 16022 | ON970600 | Unassigned | Unassigned | Unassigned | 551_6 | Clustered | 22 | VC_551_6 | 22 | 0.1214 | 1 | 0.1214 | 0 | 0 | 0 | 0.8095 | Ref |
| 16023 | ON970601 | Unassigned | Unassigned | Unassigned | 1073_0 | Clustered | 24 | VC_1073_0 | 24 | 0.8388 | 1 | 0.8388 | 0 | 0 | 0 | 1 | Ref |
| 16024 | ON970602 | Unassigned | Unassigned | Unassigned | 1178_0 | Clustered | 24 | VC_1178_0 | 24 | 0.9524 | 1 | 0.9524 | 0 | 0 | 0 | 1 | Ref |
| 16025 | ON970603 | Unassigned | Unassigned | Unassigned | 856_0 | Clustered | 7 | VC_856_0 | 7 | 0.3463 | 1 | 0.3463 | 0 | 0 | 0 | 1 | Ref |
| 16026 | ON970604 | Unassigned | Unassigned | Unassigned | 1166_0 | Clustered | 20 | VC_1166_0 | 20 | 0.99 | 1 | 0.99 | 0 | 0 | 0 | 1 | Ref |
| 16027 | ON970605 | Unassigned | Unassigned | Unassigned | 1240_0 | Clustered | 82 | VC_1240_0 | 82 | 0.7945 | 1 | 0.7945 | 0 | 0 | 0 | 1 | Ref |
| 16028 | ON970606 | Unassigned | Unassigned | Unassigned | 1240_0 | Clustered | 82 | VC_1240_0 | 82 | 0.7945 | 1 | 0.7945 | 0 | 0 | 0 | 1 | Ref |
| 16029 | ON970607 | Unassigned | Unassigned | Unassigned | 866_1 | Clustered | 20 | VC_866_1 | 20 | 0.4814 | 1 | 0.4814 | 0 | 0 | 0 | 1 | Ref |
| 16030 | ON970608 | Unassigned | Unassigned | Unassigned | 1044_0 | Clustered | 3 | VC_1044_0 | 3 | 0.0843 | 0.99852363 | 0.0842 | 0 | 0 | 0 | 1 | Ref |
| 16031 | ON970609 | Unassigned | Unassigned | Unassigned | 919_0 | Clustered | 39 | VC_919_0 | 39 | 0.9069 | 1 | 0.9069 | 0 | 0 | 0 | 1 | Ref |
| 16032 | ON970610 | Unassigned | Unassigned | Unassigned | 520_67 | Clustered/Singleton | 1 | VC_520_67 | 1 | 0 | 1 | 0 | 0 | 0 | 0 | 1 | Ref |
| 16033 | ON970611 | Unassigned | Unassigned | Unassigned | 572_0 | Clustered | 16 | VC_572_0 | 16 | 0.7286 | 1 | 0.7286 | 0 | 0 | 0 | 1 | Ref |
| 16034 | ON970612 | Unassigned | Unassigned | Unassigned | 1352_0 | Clustered | 31 | VC_1352_0 | 31 | 0.849 | 1 | 0.849 | 0 | 0 | 0 | 1 | Ref |
| 16035 | ON970613 | Unassigned | Unassigned | Unassigned | 1344_0 | Clustered | 78 | VC_1344_0 | 78 | 0.9732 | 1 | 0.9732 | 0 | 0 | 0 | 1 | Ref |
| 16036 | ON970614 | Unassigned | Unassigned | Unassigned | 517_0 | Clustered | 18 | VC_517_0 | 18 | 0.1599 | 1 | 0.1599 | 0 | 0 | 0 | 1 | Ref |

| | | | | | | | | | | | | | | | | | |
|---|---|---|---|---|---|---|---|---|---|---|---|---|---|---|---|---|---|
| 16037 | ON970616 | Unassigned | Unassigned | Unassigned | 1207_0 | Clustered | 29 | VC_1207_0 | 29 | 0.8225 | 1 | 0.8225 | 0 | 0 | 0 | 1 | Ref |
| 16038 | ON970618 | Unassigned | Unassigned | Unassigned | 761_0 | Clustered | 67 | VC_761_0 | 67 | 0.8947 | 1 | 0.8947 | 0 | 0 | 0 | 1 | Ref |
| 16039 | ON970619 | Unassigned | Unassigned | Unassigned | 1178_0 | Clustered | 24 | VC_1178_0 | 24 | 0.9524 | 1 | 0.9524 | 0 | 0 | 0 | 1 | Ref |
| 16040 | ON970620 | Unassigned | Unassigned | Unassigned | 570_0 | Clustered | 9 | VC_570_0 | 9 | 0.499 | 1 | 0.499 | 0 | 0 | 0 | 1 | Ref |
| 16041 | ON970621 | Unassigned | Unassigned | Unassigned | 919_0 | Clustered | 39 | VC_919_0 | 39 | 0.9069 | 1 | 0.9069 | 0 | 0 | 0 | 1 | Ref |
| 16042 | ON970622 | Unassigned | Unassigned | Unassigned | 799_1 | Clustered | 27 | VC_799_1 | 27 | 0.8611 | 1 | 0.8611 | 0 | 0 | 0 | 1 | Ref |
| 16043 | ON970623 | Unassigned | Unassigned | Unassigned | 513_0 | Clustered | 32 | VC_513_0 | 32 | 0.6881 | 1 | 0.6881 | 0 | 0 | 0 | 1 | Ref |
| 16044 | ON970624 | Unassigned | Unassigned | Unassigned | 325_58 | Clustered/Singleton | 1 | VC_325_58 | 1 | 0 | 1 | 0 | 0 | 0 | 0 | 1 | Ref |
| 16045 | ON970625 | Unassigned | Unassigned | Unassigned | 940_0 | Clustered | 34 | VC_940_0 | 34 | 0.7959 | 1 | 0.7959 | 0 | 0 | 0 | 1 | Ref |
| 16046 | ON970626 | Unassigned | Unassigned | Unassigned | 940_0 | Clustered | 34 | VC_940_0 | 34 | 0.7959 | 1 | 0.7959 | 0 | 0 | 0 | 1 | Ref |
| 16047 | ON970627 | Unassigned | Unassigned | Unassigned | 325_23 | Clustered | 17 | VC_325_23 | 17 | 0.0716 | 1 | 0.0716 | 0 | 0 | 0 | 0.6397 | Ref |
| 16048 | ON989481 | Unassigned | Unassigned | Unassigned | 8_0 | Clustered | 58 | VC_8_0 | 58 | 0.8306 | 1 | 0.8306 | 0 | 0 | 0 | 0.9946 | Ref |
| 16049 | ON995367 | Unassigned | Unassigned | Unassigned | 389_0 | Clustered | 3 | VC_389_0 | 3 | 0.0515 | 0.99861557 | 0.0514 | 0 | 0 | 0 | 1 | Ref |
| 16050 | ON996339 | Unassigned | Unassigned | Unassigned | 637_0 | Clustered | 115 | VC_637_0 | 115 | 0.731 | 1 | 0.731 | 0 | 0 | 0 | 0.8764 | Ref |
| 16051 | ON996340 | Unassigned | Unassigned | Unassigned | 364_0 | Clustered | 16 | VC_364_0 | 16 | 0.6491 | 1 | 0.6491 | 0 | 0 | 0 | 1 | Ref |
| 16052 | OP009265 | Unassigned | Unassigned | Unassigned | 923_0 | Clustered | 23 | VC_923_0 | 23 | 0.9455 | 1 | 0.9455 | 0 | 0 | 0 | 1 | Ref |
| 16053 | OP009266 | Unassigned | Unassigned | Unassigned | 923_0 | Clustered | 23 | VC_923_0 | 23 | 0.9455 | 1 | 0.9455 | 0 | 0 | 0 | 1 | Ref |
| 16054 | OP009267 | Unassigned | Unassigned | Unassigned | 923_0 | Clustered | 23 | VC_923_0 | 23 | 0.9455 | 1 | 0.9455 | 0 | 0 | 0 | 1 | Ref |
| 16055 | OP009268 | Unassigned | Unassigned | Unassigned | 923_0 | Clustered | 23 | VC_923_0 | 23 | 0.9455 | 1 | 0.9455 | 0 | 0 | 0 | 1 | Ref |
| 16056 | OP009269 | Unassigned | Unassigned | Unassigned | 923_0 | Clustered | 23 | VC_923_0 | 23 | 0.9455 | 1 | 0.9455 | 0 | 0 | 0 | 1 | Ref |
| 16057 | OP009270 | Unassigned | Unassigned | Unassigned | 923_0 | Clustered | 23 | VC_923_0 | 23 | 0.9455 | 1 | 0.9455 | 0 | 0 | 0 | 1 | Ref |
| 16058 | OP009271 | Unassigned | Unassigned | Unassigned | 923_0 | Clustered | 23 | VC_923_0 | 23 | 0.9455 | 1 | 0.9455 | 0 | 0 | 0 | 1 | Ref |
| 16059 | OP009272 | Unassigned | Unassigned | Unassigned | 885_0 | Clustered | 11 | VC_885_0 | 11 | 0.9774 | 1 | 0.9774 | 0 | 0 | 0 | 1 | Ref |
| 16060 | OP009273 | Unassigned | Unassigned | Unassigned | 923_0 | Clustered | 23 | VC_923_0 | 23 | 0.9455 | 1 | 0.9455 | 0 | 0 | 0 | 1 | Ref |
| 16061 | OP009274 | Unassigned | Unassigned | Unassigned | 1029_0 | Clustered | 4 | VC_1029_0 | 4 | 0.9006 | 0.99995764 | 0.9006 | 0 | 0 | 0 | 1 | Ref |
| 16062 | OP009275 | Unassigned | Unassigned | Unassigned | 885_0 | Clustered | 11 | VC_885_0 | 11 | 0.9774 | 1 | 0.9774 | 0 | 0 | 0 | 1 | Ref |
| 16063 | OP009276 | Unassigned | Unassigned | Unassigned | 923_0 | Clustered | 23 | VC_923_0 | 23 | 0.9455 | 1 | 0.9455 | 0 | 0 | 0 | 1 | Ref |
| 16064 | OP009277 | Unassigned | Unassigned | Unassigned | 923_0 | Clustered | 23 | VC_923_0 | 23 | 0.9455 | 1 | 0.9455 | 0 | 0 | 0 | 1 | Ref |
| 16065 | OP009278 | Unassigned | Unassigned | Unassigned | 885_0 | Clustered | 11 | VC_885_0 | 11 | 0.9774 | 1 | 0.9774 | 0 | 0 | 0 | 1 | Ref |
| 16066 | OP009279 | Unassigned | Unassigned | Unassigned | 923_0 | Clustered | 23 | VC_923_0 | 23 | 0.9455 | 1 | 0.9455 | 0 | 0 | 0 | 1 | Ref |
| 16067 | OP009280 | Unassigned | Unassigned | Unassigned | 885_0 | Clustered | 11 | VC_885_0 | 11 | 0.9774 | 1 | 0.9774 | 0 | 0 | 0 | 1 | Ref |
| 16068 | OP009281 | Unassigned | Unassigned | Unassigned | 1029_0 | Clustered | 4 | VC_1029_0 | 4 | 0.9006 | 0.99995764 | 0.9006 | 0 | 0 | 0 | 1 | Ref |
| 16069 | OP009282 | Unassigned | Unassigned | Unassigned | 749_0 | Clustered | 40 | VC_749_0 | 40 | 0.7517 | 1 | 0.7517 | 0 | 0 | 0 | 1 | Ref |
| 16070 | OP009283 | Unassigned | Unassigned | Unassigned | 923_0 | Clustered | 23 | VC_923_0 | 23 | 0.9455 | 1 | 0.9455 | 0 | 0 | 0 | 1 | Ref |
| 16071 | OP009284 | Unassigned | Unassigned | Unassigned | 923_0 | Clustered | 23 | VC_923_0 | 23 | 0.9455 | 1 | 0.9455 | 0 | 0 | 0 | 1 | Ref |
| 16072 | OP009285 | Unassigned | Unassigned | Unassigned | 471_0 | Clustered | 2 | VC_471_0 | 2 | 0.0804 | 0.95665719 | 0.0769 | 0 | 0 | 0 | 1 | Ref |
| 16073 | OP009286 | Unassigned | Unassigned | Unassigned | 885_0 | Clustered | 11 | VC_885_0 | 11 | 0.9774 | 1 | 0.9774 | 0 | 0 | 0 | 1 | Ref |
| 16074 | OP009287 | Unassigned | Unassigned | Unassigned | 923_0 | Clustered | 23 | VC_923_0 | 23 | 0.9455 | 1 | 0.9455 | 0 | 0 | 0 | 1 | Ref |
| 16075 | OP009288 | Unassigned | Unassigned | Unassigned | 923_0 | Clustered | 23 | VC_923_0 | 23 | 0.9455 | 1 | 0.9455 | 0 | 0 | 0 | 1 | Ref |
| 16076 | OP013026 | Unassigned | Unassigned | Unassigned | 823_0 | Clustered | 124 | VC_823_0 | 124 | 0.9492 | 1 | 0.9492 | 0 | 0 | 0 | 1 | Ref |
| 16077 | OP018999 | Unassigned | Unassigned | Unassigned | 452_0 | Clustered | 66 | VC_452_0 | 66 | 0.9248 | 1 | 0.9248 | 0 | 0 | 0 | 1 | Ref |
| 16078 | OP019135 | Unassigned | Unassigned | Unassigned | 699_0 | Clustered | 71 | VC_699_0 | 71 | 0.9352 | 1 | 0.9352 | 0 | 0 | 0 | 1 | Ref |
| 16079 | OP021677 | Unassigned | Unassigned | Unassigned | 325_62 | Clustered | 3 | VC_325_62 | 3 | 0.0188 | 0.99859194 | 0.0188 | 0 | 0 | 0 | 1 | Ref |
| 16080 | OP021678 | Unassigned | Unassigned | Unassigned | 1224_0 | Clustered | 19 | VC_1224_0 | 19 | 1 | 1 | 1 | 0 | 0 | 0 | 1 | Ref |
| 16081 | OP021679 | Unassigned | Unassigned | Unassigned | 224_0 | Clustered | 19 | VC_224_0 | 19 | 0.9416 | 1 | 0.9416 | 0 | 0 | 0 | 1 | Ref |
| 16082 | OP021680 | Unassigned | Unassigned | Unassigned | 1071_0 | Clustered | 20 | VC_1071_0 | 20 | 0.3268 | 1 | 0.3268 | 0 | 0 | 0 | 0.9842 | Ref |
| 16083 | OP021681 | Unassigned | Unassigned | Unassigned | 558_24 | Clustered/Singleton | 1 | VC_558_24 | 1 | 0 | 1 | 0 | 0 | 0 | 0 | 1 | Ref |
| 16084 | OP021682 | Unassigned | Unassigned | Unassigned | 558_67 | Clustered/Singleton | 1 | VC_558_67 | 1 | 0 | 1 | 0 | 0 | 0 | 0 | 1 | Ref |
| 16085 | OP021683 | Unassigned | Unassigned | Unassigned | 1240_0 | Clustered | 82 | VC_1240_0 | 82 | 0.7945 | 1 | 0.7945 | 0 | 0 | 0 | 1 | Ref |
| 16086 | OP021684 | Unassigned | Unassigned | Unassigned | 558_50 | Clustered | 15 | VC_558_50 | 15 | 0.0468 | 1 | 0.0468 | 0 | 0 | 0 | 0.9238 | Ref |
| 16087 | OP022426 | Unassigned | Unassigned | Unassigned | 897_0 | Clustered | 6 | VC_897_0 | 6 | 0.4156 | 1 | 0.4156 | 0 | 0 | 0 | 1 | Ref |
| 16088 | OP028995 | Unassigned | Unassigned | Unassigned | 935_0 | Clustered | 2 | VC_935_0 | 2 | 0.7079 | 0.72985431 | 0.5167 | 0 | 0 | 0 | 1 | Ref |
| 16089 | OP038318 | Unassigned | Unassigned | Unassigned | 8_0 | Clustered | 58 | VC_8_0 | 58 | 0.8306 | 1 | 0.8306 | 0 | 0 | 0 | 0.9946 | Ref |
| 16090 | OP038319 | Unassigned | Unassigned | Unassigned | 8_0 | Clustered | 58 | VC_8_0 | 58 | 0.8306 | 1 | 0.8306 | 0 | 0 | 0 | 0.9946 | Ref |
| 16091 | OP038320 | Unassigned | Unassigned | Unassigned | 10_0 | Clustered | 25 | VC_10_0 | 25 | 0.3467 | 1 | 0.3467 | 0 | 0 | 0 | 0.9733 | Ref |
| 16092 | OP038547 | Unassigned | Unassigned | Unassigned | 452_0 | Clustered | 66 | VC_452_0 | 66 | 0.9248 | 1 | 0.9248 | 0 | 0 | 0 | 1 | Ref |
| 16093 | OP045496 | Unassigned | Unassigned | Unassigned | 823_0 | Clustered | 124 | VC_823_0 | 124 | 0.9492 | 1 | 0.9492 | 0 | 0 | 0 | 1 | Ref |
| 16094 | OP045497 | Unassigned | Unassigned | Unassigned | 668_0 | Clustered | 25 | VC_668_0 | 25 | 0.5539 | 1 | 0.5539 | 0 | 0 | 0 | 0.96 | Ref |
| 16095 | OP045498 | Unassigned | Unassigned | Unassigned | 191_19 | Clustered | 12 | VC_191_19 | 12 | 0.0655 | 1 | 0.0655 | 0 | 0 | 0 | 0.8636 | Ref |
| 16096 | OP046317 | Unassigned | Unassigned | Unassigned | 835_0 | Clustered | 67 | VC_835_0 | 67 | 0.9624 | 1 | 0.9624 | 0 | 0 | 0 | 1 | Ref |
| 16097 | OP053360 | Unassigned | Unassigned | Unassigned | 664_0 | Clustered | 4 | VC_664_0 | 4 | 0.263 | 0.99998747 | 0.263 | 0 | 0 | 0 | 1 | Ref |
| 16098 | OP053361 | Unassigned | Unassigned | Unassigned | 664_0 | Clustered | 4 | VC_664_0 | 4 | 0.263 | 0.99998747 | 0.263 | 0 | 0 | 0 | 1 | Ref |
| 16099 | OP053362 | Unassigned | Unassigned | Unassigned | 664_0 | Clustered | 4 | VC_664_0 | 4 | 0.263 | 0.99998747 | 0.263 | 0 | 0 | 0 | 1 | Ref |
| 16100 | OP056089 | Unassigned | Unassigned | Unassigned | 793_0 | Clustered | 12 | VC_793_0 | 12 | 0.8531 | 1 | 0.8531 | 0 | 0 | 0 | 1 | Ref |
| 16101 | OP056300 | Unassigned | Unassigned | Unassigned | 778_0 | Clustered | 4 | VC_778_0 | 4 | 0.8115 | 0.99985435 | 0.8114 | 0 | 0 | 0 | 1 | Ref |
| 16102 | OP067662 | Unassigned | Unassigned | Unassigned | 364_0 | Clustered | 16 | VC_364_0 | 16 | 0.6491 | 1 | 0.6491 | 0 | 0 | 0 | 1 | Ref |
| 16103 | OP068331 | Unassigned | Unassigned | Unassigned | 1171_0 | Clustered | 21 | VC_1171_0 | 21 | 0.7125 | 1 | 0.7125 | 0 | 0 | 0 | 1 | Ref |
| 16104 | OP068332 | Unassigned | Unassigned | Unassigned | 1171_0 | Clustered | 21 | VC_1171_0 | 21 | 0.7125 | 1 | 0.7125 | 0 | 0 | 0 | 1 | Ref |
| 16105 | OP068333 | Unassigned | Unassigned | Unassigned | 856_1 | Clustered | 13 | VC_856_1 | 13 | 0.5848 | 1 | 0.5848 | 0 | 0 | 0 | 1 | Ref |
| 16106 | OP068334 | Unassigned | Unassigned | Unassigned | 1440_0 | Clustered | 4 | VC_1440_0 | 4 | 1 | 1 | 1 | 0 | 0 | 0 | 1 | Ref |
| 16107 | OP068335 | Unassigned | Unassigned | Unassigned | 1361_0 | Clustered | 6 | VC_1361_0 | 6 | 0.2371 | 1 | 0.2371 | 0 | 0 | 0 | 1 | Ref |
| 16108 | OP068336 | Unassigned | Unassigned | Unassigned | 1171_0 | Clustered | 21 | VC_1171_0 | 21 | 0.7125 | 1 | 0.7125 | 0 | 0 | 0 | 1 | Ref |
| 16109 | OP068337 | Unassigned | Unassigned | Unassigned | 1171_0 | Clustered | 21 | VC_1171_0 | 21 | 0.7125 | 1 | 0.7125 | 0 | 0 | 0 | 1 | Ref |
| 16110 | OP068338 | Unassigned | Unassigned | Unassigned | 1352_0 | Clustered | 31 | VC_1352_0 | 31 | 0.849 | 1 | 0.849 | 0 | 0 | 0 | 1 | Ref |
| 16111 | OP068339 | Unassigned | Unassigned | Unassigned | 1163_0 | Clustered | 28 | VC_1163_0 | 28 | 0.4923 | 1 | 0.4923 | 0 | 0 | 0 | 1 | Ref |
| 16112 | OP068340 | Unassigned | Unassigned | Unassigned | 520_22 | Clustered/Singleton | 1 | VC_520_22 | 1 | 0 | 1 | 0 | 0 | 0 | 0 | 1 | Ref |
| 16113 | OP068341 | Unassigned | Unassigned | Unassigned | 558_49 | Clustered | 7 | VC_558_49 | 7 | 0.0194 | 0.99510369 | 0.0193 | 0 | 0 | 0 | 0.8571 | Ref |
| 16114 | OP068342 | Unassigned | Unassigned | Unassigned | 1095_0 | Clustered | 16 | VC_1095_0 | 16 | 0.458 | 1 | 0.458 | 0 | 0 | 0 | 1 | Ref |
| 16115 | OP068343 | Unassigned | Unassigned | Unassigned | 1100_3 | Clustered/Singleton | 1 | VC_1100_3 | 1 | 0 | 1 | 0 | 0 | 0 | 0 | 1 | Ref |
| 16116 | OP068344 | Unassigned | Unassigned | Unassigned | 1171_0 | Clustered | 21 | VC_1171_0 | 21 | 0.7125 | 1 | 0.7125 | 0 | 0 | 0 | 1 | Ref |
| 16117 | OP079918 | Unassigned | Unassigned | Unassigned | 452_0 | Clustered | 66 | VC_452_0 | 66 | 0.9248 | 1 | 0.9248 | 0 | 0 | 0 | 1 | Ref |
| 16118 | OP087522 | Unassigned | Unassigned | Unassigned | 456_0 | Clustered | 62 | VC_456_0 | 62 | 0.9026 | 1 | 0.9026 | 0 | 0 | 0 | 1 | Ref |
| 16119 | OP094641 | Unassigned | Unassigned | Unassigned | 575_0 | Clustered | 35 | VC_575_0 | 35 | 0.8815 | 1 | 0.8815 | 0 | 0 | 0 | 1 | Ref |
| 16120 | OP103917 | Unassigned | Unassigned | Unassigned | 823_0 | Clustered | 124 | VC_823_0 | 124 | 0.9492 | 1 | 0.9492 | 0 | 0 | 0 | 1 | Ref |
| 16121 | OP103918 | Unassigned | Unassigned | Unassigned | 823_0 | Clustered | 124 | VC_823_0 | 124 | 0.9492 | 1 | 0.9492 | 0 | 0 | 0 | 1 | Ref |
| 16122 | OP103919 | Unassigned | Unassigned | Unassigned | 823_0 | Clustered | 124 | VC_823_0 | 124 | 0.9492 | 1 | 0.9492 | 0 | 0 | 0 | 1 | Ref |
| 16123 | OP103920 | Unassigned | Unassigned | Unassigned | 823_0 | Clustered | 124 | VC_823_0 | 124 | 0.9492 | 1 | 0.9492 | 0 | 0 | 0 | 1 | Ref |
| 16124 | OP103921 | Unassigned | Unassigned | Unassigned | 823_0 | Clustered | 124 | VC_823_0 | 124 | 0.9492 | 1 | 0.9492 | 0 | 0 | 0 | 1 | Ref |
| 16125 | OP103922 | Unassigned | Unassigned | Unassigned | 823_0 | Clustered | 124 | VC_823_0 | 124 | 0.9492 | 1 | 0.9492 | 0 | 0 | 0 | 1 | Ref |
| 16126 | OP103923 | Unassigned | Unassigned | Unassigned | 823_0 | Clustered | 124 | VC_823_0 | 124 | 0.9492 | 1 | 0.9492 | 0 | 0 | 0 | 1 | Ref |
| 16127 | OP114730 | Unassigned | Unassigned | Unassigned | 123_0 | Clustered | 23 | VC_123_0 | 23 | 0.817 | 1 | 0.817 | 0 | 0 | 0 | 1 | Ref |
| 16128 | OP114731 | Unassigned | Unassigned | Unassigned | 111_0 | Clustered | 91 | VC_111_0 | 91 | 0.9587 | 1 | 0.9587 | 0 | 0 | 0 | 1 | Ref |
| 16129 | OP114732 | Unassigned | Unassigned | Unassigned | 242_1 | Clustered | 20 | VC_242_1 | 20 | 0.684 | 1 | 0.684 | 0 | 0 | 0 | 0.8368 | Ref |
| 16130 | OP117450 | Unassigned | Unassigned | Unassigned | 837_0 | Clustered | 29 | VC_837_0 | 29 | 0.9955 | 1 | 0.9955 | 0 | 0 | 0 | 1 | Ref |
| 16131 | OP121182 | Unassigned | Unassigned | Unassigned | 231_0 | Clustered | 3 | VC_231_0 | 3 | 0.236 | 0.99824503 | 0.2356 | 0 | 0 | 0 | 1 | Ref |
| 16132 | OP125534 | Unassigned | Unassigned | Unassigned | 354_0 | Clustered | 4 | VC_354_0 | 4 | 0.1749 | 0.99998684 | 0.1749 | 0 | 0 | 0 | 0.8333 | Ref |
| 16133 | OP131293 | Unassigned | Unassigned | Unassigned | 486_0 | Clustered | 18 | VC_486_0 | 18 | 0.887 | 1 | 0.887 | 0 | 0 | 0 | 1 | Ref |
| 16134 | OP131294 | Unassigned | Unassigned | Unassigned | 486_0 | Clustered | 18 | VC_486_0 | 18 | 0.887 | 1 | 0.887 | 0 | 0 | 0 | 1 | Ref |
| 16135 | OP131295 | Unassigned | Unassigned | Unassigned | 486_0 | Clustered | 18 | VC_486_0 | 18 | 0.887 | 1 | 0.887 | 0 | 0 | 0 | 1 | Ref |
| 16136 | OP131296 | Unassigned | Unassigned | Unassigned | 486_0 | Clustered | 18 | VC_486_0 | 18 | 0.887 | 1 | 0.887 | 0 | 0 | 0 | 1 | Ref |
| 16137 | OP131297 | Unassigned | Unassigned | Unassigned | 486_0 | Clustered | 18 | VC_486_0 | 18 | 0.887 | 1 | 0.887 | 0 | 0 | 0 | 1 | Ref |
| 16138 | OP131299 | Unassigned | Unassigned | Unassigned | 486_0 | Clustered | 18 | VC_486_0 | 18 | 0.887 | 1 | 0.887 | 0 | 0 | 0 | 1 | Ref |
| 16139 | OP131300 | Unassigned | Unassigned | Unassigned | 486_0 | Clustered | 18 | VC_486_0 | 18 | 0.887 | 1 | 0.887 | 0 | 0 | 0 | 1 | Ref |
| 16140 | OP131301 | Unassigned | Unassigned | Unassigned | 486_0 | Clustered | 18 | VC_486_0 | 18 | 0.887 | 1 | 0.887 | 0 | 0 | 0 | 1 | Ref |
| 16141 | OP131302 | Unassigned | Unassigned | Unassigned | 486_0 | Clustered | 18 | VC_486_0 | 18 | 0.887 | 1 | 0.887 | 0 | 0 | 0 | 1 | Ref |
| 16142 | OP132241 | Unassigned | Unassigned | Unassigned | 428_0 | Clustered | 33 | VC_428_0 | 33 | 0.9915 | 1 | 0.9915 | 0 | 0 | 0 | 1 | Ref |
| 16143 | OP136131 | Unassigned | Unassigned | Unassigned | 8_0 | Clustered | 58 | VC_8_0 | 58 | 0.8306 | 1 | 0.8306 | 0 | 0 | 0 | 0.9946 | Ref |
| 16144 | OP136132 | Unassigned | Unassigned | Unassigned | 8_0 | Clustered | 58 | VC_8_0 | 58 | 0.8306 | 1 | 0.8306 | 0 | 0 | 0 | 0.9946 | Ref |
| 16145 | OP136133 | Unassigned | Unassigned | Unassigned | 7_0 | Clustered | 28 | VC_7_0 | 28 | 0.3916 | 1 | 0.3916 | 0 | 0 | 0 | 0.9947 | Ref |
| 16146 | OP136134 | Unassigned | Unassigned | Unassigned | 8_0 | Clustered | 58 | VC_8_0 | 58 | 0.8306 | 1 | 0.8306 | 0 | 0 | 0 | 0.9946 | Ref |
| 16147 | OP136135 | Unassigned | Unassigned | Unassigned | 10_0 | Clustered | 25 | VC_10_0 | 25 | 0.3467 | 1 | 0.3467 | 0 | 0 | 0 | 0.9733 | Ref |
| 16148 | OP136151 | Unassigned | Unassigned | Unassigned | 1228_0 | Clustered | 2 | VC_1228_0 | 2 | 0.9902 | 0.72985431 | 0.7227 | 0 | 0 | 0 | 1 | Ref |
| 16149 | OP168678 | Unassigned | Unassigned | Unassigned | 11_0 | Clustered | 19 | VC_11_0 | 19 | 0.7562 | 1 | 0.7562 | 0 | 0 | 0 | 1 | Ref |
| 16150 | OP168898 | Unassigned | Unassigned | Unassigned | 283_135 | Clustered/Singleton | 1 | VC_283_135 | 1 | 0 | 1 | 0 | 0 | 0 | 0 | 1 | Ref |
| 16151 | OP171942 | Unassigned | Unassigned | Unassigned | 699_0 | Clustered | 71 | VC_699_0 | 71 | 0.9352 | 1 | 0.9352 | 0 | 0 | 0 | 1 | Ref |
| 16152 | OP171943 | Unassigned | Unassigned | Unassigned | 192_0 | Clustered | 15 | VC_192_0 | 15 | 0.5071 | 1 | 0.5071 | 0 | 0 | 0 | 1 | Ref |
| 16153 | OP172867 | Unassigned | Unassigned | Unassigned | 519_0 | Clustered | 39 | VC_519_0 | 39 | 0.8156 | 1 | 0.8156 | 0 | 0 | 0 | 1 | Ref |
| 16154 | OP172868 | Unassigned | Unassigned | Unassigned | 572_0 | Clustered | 16 | VC_572_0 | 16 | 0.7286 | 1 | 0.7286 | 0 | 0 | 0 | 1 | Ref |
| 16155 | OP172870 | Unassigned | Unassigned | Unassigned | 572_0 | Clustered | 16 | VC_572_0 | 16 | 0.7286 | 1 | 0.7286 | 0 | 0 | 0 | 1 | Ref |
| 16156 | OP172872 | Unassigned | Unassigned | Unassigned | 761_0 | Clustered | 67 | VC_761_0 | 67 | 0.8947 | 1 | 0.8947 | 0 | 0 | 0 | 1 | Ref |
| 16157 | OP172873 | Unassigned | Unassigned | Unassigned | 511_0 | Clustered | 3 | VC_511_0 | 3 | 0.1104 | 0.99858683 | 0.1103 | 0 | 0 | 0 | 1 | Ref |
| 16158 | OP172874 | Unassigned | Unassigned | Unassigned | 511_0 | Clustered | 3 | VC_511_0 | 3 | 0.1104 | 0.99858683 | 0.1103 | 0 | 0 | 0 | 1 | Ref |
| 16159 | OP172875 | Unassigned | Unassigned | Unassigned | 1234_0 | Clustered | 44 | VC_1234_0 | 44 | 1 | 1 | 1 | 0 | 0 | 0 | 1 | Ref |
| 16160 | OP172876 | Unassigned | Unassigned | Unassigned | 572_0 | Clustered | 16 | VC_572_0 | 16 | 0.7286 | 1 | 0.7286 | 0 | 0 | 0 | 1 | Ref |
| 16161 | OP172877 | Unassigned | Unassigned | Unassigned | 1093_0 | Clustered | 40 | VC_1093_0 | 40 | 0.8754 | 1 | 0.8754 | 0 | 0 | 0 | 1 | Ref |
| 16162 | OP172878 | Unassigned | Unassigned | Unassigned | 1200_0 | Clustered | 4 | VC_1200_0 | 4 | 0.8691 | 0.9999992 | 0.8691 | 0 | 0 | 0 | 1 | Ref |
| 16163 | OP177727 | Unassigned | Unassigned | Unassigned | 848_0 | Clustered | 28 | VC_848_0 | 28 | 0.7927 | 1 | 0.7927 | 0 | 0 | 0 | 1 | Ref |
| 16164 | OP186293 | Unassigned | Unassigned | Unassigned | 977_2 | Clustered/Singleton | 1 | VC_977_2 | 1 | 0 | 1 | 0 | 0 | 0 | 0 | 1 | Ref |
| 16165 | OP197928 | Unassigned | Unassigned | Unassigned | 283_22 | Clustered/Singleton | 1 | VC_283_22 | 1 | 0 | 1 | 0 | 0 | 0 | 0 | 1 | Ref |
| 16166 | OP209174 | Unassigned | Unassigned | Unassigned | 1116_0 | Clustered | 14 | VC_1116_0 | 14 | 0.7927 | 1 | 0.7927 | 0 | 0 | 0 | 1 | Ref |
| 16167 | OP209176 | Unassigned | Unassigned | Unassigned | 1294_0 | Clustered | 6 | VC_1294_0 | 6 | 0.6626 | 0.99999272 | 0.6626 | 0 | 0 | 0 | 1 | Ref |
| 16168 | OP235318 | Unassigned | Unassigned | Unassigned | 149_15 | Clustered | 16 | VC_149_15 | 16 | 0.0788 | 1 | 0.0788 | 0 | 0 | 0 | 0.8083 | Ref |
| 16169 | OP251154 | Unassigned | Unassigned | Unassigned | 456_0 | Clustered | 62 | VC_456_0 | 62 | 0.9026 | 1 | 0.9026 | 0 | 0 | 0 | 1 | Ref |
| 16170 | OP254195 | Unassigned | Unassigned | Unassigned | 695_0 | Clustered | 48 | VC_695_0 | 48 | 0.8162 | 1 | 0.8162 | 0 | 0 | 0 | 1 | Ref |
| 16171 | OP256047 | Unassigned | Unassigned | Unassigned | 823_0 | Clustered | 124 | VC_823_0 | 124 | 0.9492 | 1 | 0.9492 | 0 | 0 | 0 | 1 | Ref |
| 16172 | OP256049 | Unassigned | Unassigned | Unassigned | 899_0 | Clustered | 4 | VC_899_0 | 4 | 0.2655 | 0.99997104 | 0.2655 | 0 | 0 | 0 | 1 | Ref |
| 16173 | OP263967 | Unassigned | Unassigned | Unassigned | 149_9 | Clustered | 3 | VC_149_9 | 3 | 0.0084 | 0.97854556 | 0.0082 | 0 | 0 | 0 | 1 | Ref |
| 16174 | OP263968 | Unassigned | Unassigned | Unassigned | 149_19 | Clustered/Singleton | 1 | VC_149_19 | 1 | 0 | 1 | 0 | 0 | 0 | 0 | 1 | Ref |
| 16175 | OP263969 | Unassigned | Unassigned | Unassigned | 153_0 | Clustered | 25 | VC_153_0 | 25 | 0.7729 | 1 | 0.7729 | 0 | 0 | 0 | 1 | Ref |
| 16176 | OP265889 | Unassigned | Unassigned | Unassigned | 28_1 | Clustered | 53 | VC_28_1 | 53 | 0.7164 | 1 | 0.7164 | 0 | 0 | 0 | 0.9325 | Ref |
| 16177 | OP267563 | Unassigned | Unassigned | Unassigned | 938_0 | Clustered | 99 | VC_938_0 | 99 | 0.9119 | 1 | 0.9119 | 0 | 0 | 0 | 0.9928 | Ref |
| 16178 | OP279344 | Unassigned | Unassigned | Unassigned | 247_0 | Clustered | 14 | VC_247_0 | 14 | 0.9119 | 1 | 0.9119 | 0 | 0 | 0 | 0.967 | Ref |
| 16179 | OP291336 | Unassigned | Unassigned | Unassigned | 699_0 | Clustered | 71 | VC_699_0 | 71 | 0.9352 | 1 | 0.9352 | 0 | 0 | 0 | 1 | Ref |
| 16180 | OP292222 | Unassigned | Unassigned | Unassigned | 290_6 | Clustered | 12 | VC_290_6 | 12 | 0.1149 | 1 | 0.1149 | 0 | 0 | 0 | 0.8182 | Ref |
| 16181 | OP292288 | Unassigned | Unassigned | Unassigned | 423_0 | Clustered | 45 | VC_423_0 | 45 | 0.9484 | 1 | 0.9484 | 0 | 0 | 0 | 1 | Ref |
| 16182 | OP292654 | Unassigned | Unassigned | Unassigned | 164_6 | Clustered/Singleton | 1 | VC_164_6 | 1 | 0 | 1 | 0 | 0 | 0 | 0 | 1 | Ref |
| 16183 | OP292673 | Unassigned | Unassigned | Unassigned | 584_0 | Clustered | 28 | VC_584_0 | 28 | 0.7704 | 1 | 0.7704 | 0 | 0 | 0 | 1 | Ref |
| 16184 | OP292674 | Unassigned | Unassigned | Unassigned | 584_0 | Clustered | 28 | VC_584_0 | 28 | 0.7704 | 1 | 0.7704 | 0 | 0 | 0 | 1 | Ref |
| 16185 | OP293079 | Unassigned | Unassigned | Unassigned | 353_0 | Clustered | 6 | VC_353_0 | 6 | 0.545 | 1 | 0.545 | 0 | 0 | 0 | 1 | Ref |
| 16186 | OP293233 | Unassigned | Unassigned | Unassigned | 813_0 | Clustered | 12 | VC_813_0 | 12 | 0.2347 | 1 | 0.2347 | 0 | 0 | 0 | 1 | Ref |
| 16187 | OP294982 | Unassigned | Unassigned | Unassigned | 761_0 | Clustered | 67 | VC_761_0 | 67 | 0.8947 | 1 | 0.8947 | 0 | 0 | 0 | 1 | Ref |
| 16188 | OP296941 | Unassigned | Unassigned | Unassigned | 582_3 | Clustered | 12 | VC_582_3 | 12 | 0.2357 | 1 | 0.2357 | 0 | 0 | 0 | 0.8636 | Ref |
| 16189 | OP297178 | Unassigned | Unassigned | Unassigned | 149_6 | Clustered/Singleton | 1 | VC_149_6 | 1 | 0 | 1 | 0 | 0 | 0 | 0 | 1 | Ref |
| 16190 | OP297528 | Unassigned | Unassigned | Unassigned | 520_78 | Clustered | 2 | VC_520_78 | 2 | 0.008 | 0.95761153 | 0.0076 | 0 | 0 | 0 | 1 | Ref |
| 16191 | OP297529 | Unassigned | Unassigned | Unassigned | 517_1 | Clustered | 108 | VC_517_1 | 108 | 0.8483 | 1 | 0.8483 | 0 | 0 | 0 | 0.9955 | Ref |
| 16192 | OP297531 | Unassigned | Unassigned | Unassigned | 919_0 | Clustered | 39 | VC_919_0 | 39 | 0.9069 | 1 | 0.9069 | 0 | 0 | 0 | 1 | Ref |
| 16193 | OP297532 | Unassigned | Unassigned | Unassigned | 221_0 | Clustered | 22 | VC_221_0 | 22 | 0.7447 | 1 | 0.7447 | 0 | 0 | 0 | 1 | Ref |
| 16194 | OP297533 | Unassigned | Unassigned | Unassigned | 810_0 | Clustered | 18 | VC_810_0 | 18 | 0.9121 | 1 | 0.9121 | 0 | 0 | 0 | 1 | Ref |
| 16195 | OP297534 | Unassigned | Unassigned | Unassigned | 558_39 | Clustered | 3 | VC_558_39 | 3 | 0.0063 | 0.73849574 | 0.0047 | 0 | 0 | 0 | 0.6667 | Ref |
| 16196 | OP297535 | Unassigned | Unassigned | Unassigned | 955_0 | Clustered | 3 | VC_955_0 | 3 | 0.7745 | 0.99134622 | 0.7678 | 0 | 0 | 0 | 1 | Ref |
| 16197 | OP297536 | Unassigned | Unassigned | Unassigned | 520_31 | Clustered | 11 | VC_520_31 | 11 | 0.0897 | 1 | 0.0897 | 0 | 0 | 0 | 0.6 | Ref |
| 16198 | OP297537 | Unassigned | Unassigned | Unassigned | 216_6 | Clustered | 42 | VC_216_6 | 42 | 0.473 | 1 | 0.473 | 0 | 0 | 0 | 0.8351 | Ref |

| | | | | | | | | | | | | | | | | | |
|---|---|---|---|---|---|---|---|---|---|---|---|---|---|---|---|---|---|
| 16199 | OP297538 | Unassigned | Unassigned | Unassigned | 1360_0 | Clustered | 31 | VC_1360_0 | 31 | 0.8837 | 1 | 0.8837 | 0 | 0 | 0 | 1 | Ref |
| 16200 | OP297539 | Unassigned | Unassigned | Unassigned | 1098_0 | Clustered | 4 | VC_1098_0 | 4 | 0.5465 | 0.99996462 | 0.5465 | 0 | 0 | 0 | 1 | Ref |
| 16201 | OP297540 | Unassigned | Unassigned | Unassigned | 521_0 | Clustered | 37 | VC_521_0 | 37 | 0.912 | 1 | 0.912 | 0 | 0 | 0 | 1 | Ref |
| 16202 | OP297541 | Unassigned | Unassigned | Unassigned | 372_4 | Clustered | 3 | VC_372_4 | 3 | 0.033 | 0.99853257 | 0.0329 | 0 | 0 | 0 | 1 | Ref |
| 16203 | OP297542 | Unassigned | Unassigned | Unassigned | 520_55 | Clustered/Singleton | 1 | VC_520_55 | 1 | 0 | 1 | 0 | 0 | 0 | 0 | 1 | Ref |
| 16204 | OP297543 | Unassigned | Unassigned | Unassigned | 1352_0 | Clustered | 31 | VC_1352_0 | 31 | 0.849 | 1 | 0.849 | 0 | 0 | 0 | 1 | Ref |
| 16205 | OP297544 | Unassigned | Unassigned | Unassigned | 1175_0 | Clustered | 22 | VC_1175_0 | 22 | 0.4562 | 1 | 0.4562 | 0 | 0 | 0 | 1 | Ref |
| 16206 | OP297546 | Unassigned | Unassigned | Unassigned | 525_0 | Clustered | 25 | VC_525_0 | 25 | 0.4597 | 1 | 0.4597 | 0 | 0 | 0 | 1 | Ref |
| 16207 | OP297547 | Unassigned | Unassigned | Unassigned | 866_1 | Clustered | 20 | VC_866_1 | 20 | 0.4814 | 1 | 0.4814 | 0 | 0 | 0 | 1 | Ref |
| 16208 | OP297548 | Unassigned | Unassigned | Unassigned | 522_1 | Clustered | 5 | VC_522_1 | 5 | 0.1017 | 0.99999997 | 0.1017 | 0 | 0 | 0 | 0.8 | Ref |
| 16209 | OP297549 | Unassigned | Unassigned | Unassigned | 221_0 | Clustered | 22 | VC_221_0 | 22 | 0.7447 | 1 | 0.7447 | 0 | 0 | 0 | 1 | Ref |
| 16210 | OP297550 | Unassigned | Unassigned | Unassigned | 1166_0 | Clustered | 20 | VC_1166_0 | 20 | 0.99 | 1 | 0.99 | 0 | 0 | 0 | 1 | Ref |
| 16211 | OP297551 | Unassigned | Unassigned | Unassigned | 763_1 | Clustered | 9 | VC_763_1 | 9 | 0.3706 | 1 | 0.3706 | 0 | 0 | 0 | 1 | Ref |
| 16212 | OP297552 | Unassigned | Unassigned | Unassigned | 1166_0 | Clustered | 20 | VC_1166_0 | 20 | 0.99 | 1 | 0.99 | 0 | 0 | 0 | 1 | Ref |
| 16213 | OP297553 | Unassigned | Unassigned | Unassigned | 215_1 | Clustered | 19 | VC_215_1 | 19 | 0.4188 | 1 | 0.4188 | 0 | 0 | 0 | 1 | Ref |
| 16214 | OP297622 | Unassigned | Unassigned | Unassigned | 0_0 | Clustered | 3 | VC_0_0 | 3 | 1 | 1 | 1 | 0 | 0 | 0 | 1 | Ref |
| 16215 | OP310970 | Unassigned | Unassigned | Unassigned | 79_7 | Clustered | 33 | VC_79_7 | 33 | 0.216 | 1 | 0.216 | 0 | 0 | 0 | 0.8409 | Ref |
| 16216 | OP310971 | Unassigned | Unassigned | Unassigned | 79_7 | Clustered | 33 | VC_79_7 | 33 | 0.216 | 1 | 0.216 | 0 | 0 | 0 | 0.8409 | Ref |
| 16217 | OP310972 | Unassigned | Unassigned | Unassigned | 79_7 | Clustered | 33 | VC_79_7 | 33 | 0.216 | 1 | 0.216 | 0 | 0 | 0 | 0.8409 | Ref |
| 16218 | OP310973 | Unassigned | Unassigned | Unassigned | 79_7 | Clustered | 33 | VC_79_7 | 33 | 0.216 | 1 | 0.216 | 0 | 0 | 0 | 0.8409 | Ref |
| 16219 | OP310974 | Unassigned | Unassigned | Unassigned | 79_6 | Clustered | 11 | VC_79_6 | 11 | 0.0528 | 1 | 0.0528 | 0 | 0 | 0 | 0.7455 | Ref |
| 16220 | OP310975 | Unassigned | Unassigned | Unassigned | 79_0 | Clustered | 64 | VC_79_0 | 64 | 0.4185 | 1 | 0.4185 | 0 | 0 | 0 | 0.7649 | Ref |
| 16221 | OP310976 | Unassigned | Unassigned | Unassigned | 79_7 | Clustered | 33 | VC_79_7 | 33 | 0.216 | 1 | 0.216 | 0 | 0 | 0 | 0.8409 | Ref |
| 16222 | OP310977 | Unassigned | Unassigned | Unassigned | 79_0 | Clustered | 64 | VC_79_0 | 64 | 0.4185 | 1 | 0.4185 | 0 | 0 | 0 | 0.7649 | Ref |
| 16223 | OP310978 | Unassigned | Unassigned | Unassigned | 79_7 | Clustered | 33 | VC_79_7 | 33 | 0.216 | 1 | 0.216 | 0 | 0 | 0 | 0.8409 | Ref |
| 16224 | OP310979 | Unassigned | Unassigned | Unassigned | 79_3 | Clustered/Singleton | 1 | VC_79_3 | 1 | 0 | 1 | 0 | 0 | 0 | 0 | 1 | Ref |
| 16225 | OP310980 | Unassigned | Unassigned | Unassigned | 79_7 | Clustered | 33 | VC_79_7 | 33 | 0.216 | 1 | 0.216 | 0 | 0 | 0 | 0.8409 | Ref |
| 16226 | OP310981 | Unassigned | Unassigned | Unassigned | 79_7 | Clustered | 33 | VC_79_7 | 33 | 0.216 | 1 | 0.216 | 0 | 0 | 0 | 0.8409 | Ref |
| 16227 | OP310982 | Unassigned | Unassigned | Unassigned | 79_7 | Clustered | 33 | VC_79_7 | 33 | 0.216 | 1 | 0.216 | 0 | 0 | 0 | 0.8409 | Ref |
| 16228 | OP310983 | Unassigned | Unassigned | Unassigned | 344_0 | Clustered | 21 | VC_344_0 | 21 | 0.5436 | 1 | 0.5436 | 0 | 0 | 0 | 1 | Ref |
| 16229 | OP310984 | Unassigned | Unassigned | Unassigned | 344_0 | Clustered | 21 | VC_344_0 | 21 | 0.5436 | 1 | 0.5436 | 0 | 0 | 0 | 1 | Ref |
| 16230 | OP312987 | Unassigned | Unassigned | Unassigned | 779_0 | Clustered | 37 | VC_779_0 | 37 | 0.5682 | 1 | 0.5682 | 0 | 0 | 0 | 1 | Ref |
| 16231 | OP313111 | Unassigned | Unassigned | Unassigned | 83_0 | Clustered | 6 | VC_83_0 | 6 | 0.6436 | 1 | 0.6436 | 0 | 0 | 0 | 1 | Ref |
| 16232 | OP313112 | Unassigned | Unassigned | Unassigned | 784_0 | Clustered | 6 | VC_784_0 | 6 | 0.5517 | 1 | 0.5517 | 0 | 0 | 0 | 1 | Ref |
| 16233 | OP314870 | Unassigned | Unassigned | Unassigned | 79_0 | Clustered | 64 | VC_79_0 | 64 | 0.4185 | 1 | 0.4185 | 0 | 0 | 0 | 0.7649 | Ref |
| 16234 | OP314871 | Unassigned | Unassigned | Unassigned | 79_0 | Clustered | 64 | VC_79_0 | 64 | 0.4185 | 1 | 0.4185 | 0 | 0 | 0 | 0.7649 | Ref |
| 16235 | OP314872 | Unassigned | Unassigned | Unassigned | 79_7 | Clustered | 33 | VC_79_7 | 33 | 0.216 | 1 | 0.216 | 0 | 0 | 0 | 0.8409 | Ref |
| 16236 | OP323059 | Unassigned | Unassigned | Unassigned | 149_15 | Clustered | 16 | VC_149_15 | 16 | 0.0788 | 1 | 0.0788 | 0 | 0 | 0 | 0.8083 | Ref |
| 16237 | OP328420 | Unassigned | Unassigned | Unassigned | 8_0 | Clustered | 58 | VC_8_0 | 58 | 0.8306 | 1 | 0.8306 | 0 | 0 | 0 | 0.9946 | Ref |
| 16238 | OP328422 | Unassigned | Unassigned | Unassigned | 8_0 | Clustered | 58 | VC_8_0 | 58 | 0.8306 | 1 | 0.8306 | 0 | 0 | 0 | 0.9946 | Ref |
| 16239 | OP328423 | Unassigned | Unassigned | Unassigned | 10_0 | Clustered | 25 | VC_10_0 | 25 | 0.3467 | 1 | 0.3467 | 0 | 0 | 0 | 0.9733 | Ref |
| 16240 | OP331213 | Unassigned | Unassigned | Unassigned | 491_0 | Clustered | 36 | VC_491_0 | 36 | 0.5336 | 1 | 0.5336 | 0 | 0 | 0 | 0.9714 | Ref |
| 16241 | OP331230 | Unassigned | Unassigned | Unassigned | 392_0 | Clustered | 126 | VC_392_0 | 126 | 0.9621 | 1 | 0.9621 | 0 | 0 | 0 | 1 | Ref |
| 16242 | OP341624 | Unassigned | Unassigned | Unassigned | 107_0 | Clustered | 5 | VC_107_0 | 5 | 0.4515 | 0.99999995 | 0.4515 | 0 | 0 | 0 | 1 | Ref |
| 16243 | OP341625 | Unassigned | Unassigned | Unassigned | 101_0 | Clustered | 3 | VC_101_0 | 3 | 0.3621 | 0.99764884 | 0.3612 | 0 | 0 | 0 | 1 | Ref |
| 16244 | OP341626 | Unassigned | Unassigned | Unassigned | 107_0 | Clustered | 5 | VC_107_0 | 5 | 0.4515 | 0.99999995 | 0.4515 | 0 | 0 | 0 | 1 | Ref |
| 16245 | OP341627 | Unassigned | Unassigned | Unassigned | 107_0 | Clustered | 5 | VC_107_0 | 5 | 0.4515 | 0.99999995 | 0.4515 | 0 | 0 | 0 | 1 | Ref |
| 16246 | OP341628 | Unassigned | Unassigned | Unassigned | 107_0 | Clustered | 5 | VC_107_0 | 5 | 0.4515 | 0.99999995 | 0.4515 | 0 | 0 | 0 | 1 | Ref |
| 16247 | OP341630 | Unassigned | Unassigned | Unassigned | 107_0 | Clustered | 5 | VC_107_0 | 5 | 0.4515 | 0.99999995 | 0.4515 | 0 | 0 | 0 | 1 | Ref |
| 16248 | OP342787 | Unassigned | Unassigned | Unassigned | 472_0 | Clustered | 47 | VC_472_0 | 47 | 0.9025 | 1 | 0.9025 | 0 | 0 | 0 | 1 | Ref |
| 16249 | OP352608 | Unassigned | Unassigned | Unassigned | 618_0 | Clustered | 13 | VC_618_0 | 13 | 0.6252 | 1 | 0.6252 | 0 | 0 | 0 | 1 | Ref |
| 16250 | OP361299 | Unassigned | Unassigned | Unassigned | 169_0 | Clustered | 20 | VC_169_0 | 20 | 0.7529 | 1 | 0.7529 | 0 | 0 | 0 | 1 | Ref |
| 16251 | OP373729 | Unassigned | Unassigned | Unassigned | 460_0 | Clustered | 19 | VC_460_0 | 19 | 0.3744 | 1 | 0.3744 | 0 | 0 | 0 | 1 | Ref |
| 16252 | OP374154 | Unassigned | Unassigned | Unassigned | 8_0 | Clustered | 58 | VC_8_0 | 58 | 0.8306 | 1 | 0.8306 | 0 | 0 | 0 | 0.9946 | Ref |
| 16253 | OP374155 | Unassigned | Unassigned | Unassigned | 8_0 | Clustered | 58 | VC_8_0 | 58 | 0.8306 | 1 | 0.8306 | 0 | 0 | 0 | 0.9946 | Ref |
| 16254 | OP374156 | Unassigned | Unassigned | Unassigned | 8_0 | Clustered | 58 | VC_8_0 | 58 | 0.8306 | 1 | 0.8306 | 0 | 0 | 0 | 0.9946 | Ref |
| 16255 | OP375635 | Unassigned | Unassigned | Unassigned | 642_2 | Clustered | 6 | VC_642_2 | 6 | 0.1679 | 1 | 0.1679 | 0 | 0 | 0 | 1 | Ref |
| 16256 | OP377721 | Unassigned | Unassigned | Unassigned | 28_1 | Clustered | 53 | VC_28_1 | 53 | 0.7164 | 1 | 0.7164 | 0 | 0 | 0 | 0.9325 | Ref |
| 16257 | OP380269 | Unassigned | Unassigned | Unassigned | 169_0 | Clustered | 20 | VC_169_0 | 20 | 0.7529 | 1 | 0.7529 | 0 | 0 | 0 | 1 | Ref |
| 16258 | OP381444 | Unassigned | Unassigned | Unassigned | 1022_0 | Clustered | 4 | VC_1022_0 | 4 | 0.8047 | 0.99996089 | 0.8046 | 0 | 0 | 0 | 1 | Ref |
| 16259 | OP394141 | Unassigned | Unassigned | Unassigned | 190_23 | Clustered | 10 | VC_190_23 | 10 | 0.0704 | 1 | 0.0704 | 0 | 0 | 0 | 0.8444 | Ref |
| 16260 | OP394178 | Unassigned | Unassigned | Unassigned | 38_0 | Clustered | 3 | VC_38_0 | 3 | 1 | 0 | 0 | 0 | 0 | 0 | 1 | Ref |
| 16261 | OP413828 | Unassigned | Unassigned | Unassigned | 452_0 | Clustered | 66 | VC_452_0 | 66 | 0.9248 | 1 | 0.9248 | 0 | 0 | 0 | 1 | Ref |
| 16262 | OP413829 | Unassigned | Unassigned | Unassigned | 111_0 | Clustered | 91 | VC_111_0 | 91 | 0.9587 | 1 | 0.9587 | 0 | 0 | 0 | 1 | Ref |
| 16263 | OP413830 | Unassigned | Unassigned | Unassigned | 452_0 | Clustered | 66 | VC_452_0 | 66 | 0.9248 | 1 | 0.9248 | 0 | 0 | 0 | 1 | Ref |
| 16264 | OP413832 | Unassigned | Unassigned | Unassigned | 938_0 | Clustered | 99 | VC_938_0 | 99 | 0.9119 | 1 | 0.9119 | 0 | 0 | 0 | 0.9928 | Ref |
| 16265 | OP422637 | Unassigned | Unassigned | Unassigned | 749_0 | Clustered | 40 | VC_749_0 | 40 | 0.7517 | 1 | 0.7517 | 0 | 0 | 0 | 1 | Ref |
| 16266 | OP433453 | Unassigned | Unassigned | Unassigned | 1441_0 | Clustered | 2 | VC_1441_0 | 2 | 1 | 1 | 1 | 0 | 0 | 0 | 1 | Ref |
| 16267 | OP434439 | Unassigned | Unassigned | Unassigned | 1234_0 | Clustered | 44 | VC_1234_0 | 44 | 1 | 1 | 1 | 0 | 0 | 0 | 1 | Ref |
| 16268 | OP434440 | Unassigned | Unassigned | Unassigned | 525_0 | Clustered | 25 | VC_525_0 | 25 | 0.4597 | 1 | 0.4597 | 0 | 0 | 0 | 1 | Ref |
| 16269 | OP434441 | Unassigned | Unassigned | Unassigned | 509_0 | Clustered | 37 | VC_509_0 | 37 | 0.9689 | 1 | 0.9689 | 0 | 0 | 0 | 1 | Ref |
| 16270 | OP434442 | Unassigned | Unassigned | Unassigned | 1036_0 | Clustered | 11 | VC_1036_0 | 11 | 0.656 | 1 | 0.656 | 0 | 0 | 0 | 0.8727 | Ref |
| 16271 | OP434443 | Unassigned | Unassigned | Unassigned | 1234_0 | Clustered | 44 | VC_1234_0 | 44 | 1 | 1 | 1 | 0 | 0 | 0 | 1 | Ref |
| 16272 | OP434444 | Unassigned | Unassigned | Unassigned | 1160_0 | Clustered | 73 | VC_1160_0 | 73 | 0.9835 | 1 | 0.9835 | 0 | 0 | 0 | 1 | Ref |
| 16273 | OP434445 | Unassigned | Unassigned | Unassigned | 1234_0 | Clustered | 44 | VC_1234_0 | 44 | 1 | 1 | 1 | 0 | 0 | 0 | 1 | Ref |
| 16274 | OP434447 | Unassigned | Unassigned | Unassigned | 223_0 | Clustered | 38 | VC_223_0 | 38 | 0.9095 | 1 | 0.9095 | 0 | 0 | 0 | 1 | Ref |
| 16275 | OP434448 | Unassigned | Unassigned | Unassigned | 869_0 | Clustered | 9 | VC_869_0 | 9 | 0.2624 | 1 | 0.2624 | 0 | 0 | 0 | 1 | Ref |
| 16276 | OP434450 | Unassigned | Unassigned | Unassigned | 866_1 | Clustered | 20 | VC_866_1 | 20 | 0.4814 | 1 | 0.4814 | 0 | 0 | 0 | 1 | Ref |
| 16277 | OP434451 | Unassigned | Unassigned | Unassigned | 802_0 | Clustered | 17 | VC_802_0 | 17 | 0.8025 | 1 | 0.8025 | 0 | 0 | 0 | 1 | Ref |
| 16278 | OP434452 | Unassigned | Unassigned | Unassigned | 1170_0 | Clustered | 3 | VC_1170_0 | 3 | 0.3275 | 0.99834213 | 0.327 | 0 | 0 | 0 | 1 | Ref |
| 16279 | OP434453 | Unassigned | Unassigned | Unassigned | 1369_0 | Clustered | 3 | VC_1369_0 | 3 | 0.9809 | 0.9595722 | 0.9412 | 0 | 0 | 0 | 1 | Ref |
| 16280 | OP434454 | Unassigned | Unassigned | Unassigned | 519_0 | Clustered | 39 | VC_519_0 | 39 | 0.8156 | 1 | 0.8156 | 0 | 0 | 0 | 1 | Ref |
| 16281 | OP434455 | Unassigned | Unassigned | Unassigned | 1207_0 | Clustered | 29 | VC_1207_0 | 29 | 0.8225 | 1 | 0.8225 | 0 | 0 | 0 | 1 | Ref |
| 16282 | OP434456 | Unassigned | Unassigned | Unassigned | 1093_0 | Clustered | 40 | VC_1093_0 | 40 | 0.8754 | 1 | 0.8754 | 0 | 0 | 0 | 1 | Ref |
| 16283 | OP434457 | Unassigned | Unassigned | Unassigned | 522_1 | Clustered | 5 | VC_522_1 | 5 | 0.1017 | 0.99999997 | 0.1017 | 0 | 0 | 0 | 0.8 | Ref |
| 16284 | OP434458 | Unassigned | Unassigned | Unassigned | 557_0 | Clustered | 3 | VC_557_0 | 3 | 0.0814 | 0.99853279 | 0.0812 | 0 | 0 | 0 | 0.6667 | Ref |
| 16285 | OP434459 | Unassigned | Unassigned | Unassigned | 371_11 | Clustered | 31 | VC_371_11 | 31 | 0.0927 | 1 | 0.0927 | 0 | 0 | 0 | 0.7011 | Ref |
| 16286 | OP434460 | Unassigned | Unassigned | Unassigned | 685_0 | Clustered | 16 | VC_685_0 | 16 | 0.2972 | 1 | 0.2972 | 0 | 0 | 0 | 0.9167 | Ref |
| 16287 | OP434461 | Unassigned | Unassigned | Unassigned | 510_0 | Clustered | 12 | VC_510_0 | 12 | 0.3963 | 1 | 0.3963 | 0 | 0 | 0 | 0.9697 | Ref |
| 16288 | OP434462 | Unassigned | Unassigned | Unassigned | 1164_0 | Clustered | 25 | VC_1164_0 | 25 | 0.7312 | 1 | 0.7312 | 0 | 0 | 0 | 1 | Ref |
| 16289 | OP434463 | Unassigned | Unassigned | Unassigned | 1036_0 | Clustered | 11 | VC_1036_0 | 11 | 0.656 | 1 | 0.656 | 0 | 0 | 0 | 0.8727 | Ref |
| 16290 | OP453108 | Unassigned | Unassigned | Unassigned | 592_0 | Clustered | 50 | VC_592_0 | 50 | 0.7314 | 1 | 0.7314 | 0 | 0 | 0 | 1 | Ref |
| 16291 | OP455115 | Unassigned | Unassigned | Unassigned | 575_0 | Clustered | 35 | VC_575_0 | 35 | 0.8815 | 1 | 0.8815 | 0 | 0 | 0 | 1 | Ref |
| 16292 | OP480062 | Unassigned | Unassigned | Unassigned | 582_0 | Clustered | 4 | VC_582_0 | 4 | 0.0589 | 0.99996986 | 0.0589 | 0 | 0 | 0 | 1 | Ref |
| 16293 | OP480063 | Unassigned | Unassigned | Unassigned | 582_0 | Clustered | 4 | VC_582_0 | 4 | 0.0589 | 0.99996986 | 0.0589 | 0 | 0 | 0 | 1 | Ref |
| 16294 | OP481875 | Unassigned | Unassigned | Unassigned | 460_0 | Clustered | 19 | VC_460_0 | 19 | 0.3744 | 1 | 0.3744 | 0 | 0 | 0 | 1 | Ref |
| 16295 | OP484843 | Unassigned | Unassigned | Unassigned | 5_0 | Clustered | 30 | VC_5_0 | 30 | 0.9647 | 1 | 0.9647 | 0 | 0 | 0 | 1 | Ref |
| 16296 | OP484850 | Unassigned | Unassigned | Unassigned | 637_0 | Clustered | 115 | VC_637_0 | 115 | 0.731 | 1 | 0.731 | 0 | 0 | 0 | 0.8764 | Ref |
| 16297 | OP484857 | Unassigned | Unassigned | Unassigned | 770_0 | Clustered | 5 | VC_770_0 | 5 | 0.8394 | 0.99999405 | 0.8394 | 0 | 0 | 0 | 1 | Ref |
| 16298 | OP484858 | Unassigned | Unassigned | Unassigned | 770_0 | Clustered | 5 | VC_770_0 | 5 | 0.8394 | 0.99999405 | 0.8394 | 0 | 0 | 0 | 1 | Ref |
| 16299 | OP493557 | Unassigned | Unassigned | Unassigned | 7_0 | Clustered | 28 | VC_7_0 | 28 | 0.3916 | 1 | 0.3916 | 0 | 0 | 0 | 0.9947 | Ref |
| 16300 | OP493558 | Unassigned | Unassigned | Unassigned | 10_0 | Clustered | 25 | VC_10_0 | 25 | 0.3467 | 1 | 0.3467 | 0 | 0 | 0 | 0.9733 | Ref |
| 16301 | OP494211 | Unassigned | Unassigned | Unassigned | 637_0 | Clustered | 115 | VC_637_0 | 115 | 0.731 | 1 | 0.731 | 0 | 0 | 0 | 0.8764 | Ref |
| 16302 | OP494865 | Unassigned | Unassigned | Unassigned | 620_0 | Clustered | 27 | VC_620_0 | 27 | 0.879 | 1 | 0.879 | 0 | 0 | 0 | 1 | Ref |
| 16303 | OP503543 | Unassigned | Unassigned | Unassigned | 620_0 | Clustered | 27 | VC_620_0 | 27 | 0.879 | 1 | 0.879 | 0 | 0 | 0 | 1 | Ref |
| 16304 | OP503968 | Unassigned | Unassigned | Unassigned | 620_0 | Clustered | 27 | VC_620_0 | 27 | 0.879 | 1 | 0.879 | 0 | 0 | 0 | 1 | Ref |
| 16305 | OP503969 | Unassigned | Unassigned | Unassigned | 620_0 | Clustered | 27 | VC_620_0 | 27 | 0.879 | 1 | 0.879 | 0 | 0 | 0 | 1 | Ref |
| 16306 | OP503970 | Unassigned | Unassigned | Unassigned | 620_0 | Clustered | 27 | VC_620_0 | 27 | 0.879 | 1 | 0.879 | 0 | 0 | 0 | 1 | Ref |
| 16307 | OP503971 | Unassigned | Unassigned | Unassigned | 620_0 | Clustered | 27 | VC_620_0 | 27 | 0.879 | 1 | 0.879 | 0 | 0 | 0 | 1 | Ref |
| 16308 | OP503972 | Unassigned | Unassigned | Unassigned | 620_0 | Clustered | 27 | VC_620_0 | 27 | 0.879 | 1 | 0.879 | 0 | 0 | 0 | 1 | Ref |
| 16309 | OP503973 | Unassigned | Unassigned | Unassigned | 620_0 | Clustered | 27 | VC_620_0 | 27 | 0.879 | 1 | 0.879 | 0 | 0 | 0 | 1 | Ref |
| 16310 | OP503974 | Unassigned | Unassigned | Unassigned | 620_0 | Clustered | 27 | VC_620_0 | 27 | 0.879 | 1 | 0.879 | 0 | 0 | 0 | 1 | Ref |
| 16311 | OP503975 | Unassigned | Unassigned | Unassigned | 620_0 | Clustered | 27 | VC_620_0 | 27 | 0.879 | 1 | 0.879 | 0 | 0 | 0 | 1 | Ref |
| 16312 | OP503976 | Unassigned | Unassigned | Unassigned | 620_0 | Clustered | 27 | VC_620_0 | 27 | 0.879 | 1 | 0.879 | 0 | 0 | 0 | 1 | Ref |
| 16313 | OP503977 | Unassigned | Unassigned | Unassigned | 620_0 | Clustered | 27 | VC_620_0 | 27 | 0.879 | 1 | 0.879 | 0 | 0 | 0 | 1 | Ref |
| 16314 | OP503978 | Unassigned | Unassigned | Unassigned | 620_0 | Clustered | 27 | VC_620_0 | 27 | 0.879 | 1 | 0.879 | 0 | 0 | 0 | 1 | Ref |
| 16315 | OP503979 | Unassigned | Unassigned | Unassigned | 620_0 | Clustered | 27 | VC_620_0 | 27 | 0.879 | 1 | 0.879 | 0 | 0 | 0 | 1 | Ref |
| 16316 | OP503980 | Unassigned | Unassigned | Unassigned | 620_0 | Clustered | 27 | VC_620_0 | 27 | 0.879 | 1 | 0.879 | 0 | 0 | 0 | 1 | Ref |
| 16317 | OP503981 | Unassigned | Unassigned | Unassigned | 620_0 | Clustered | 27 | VC_620_0 | 27 | 0.879 | 1 | 0.879 | 0 | 0 | 0 | 1 | Ref |
| 16318 | OP503982 | Unassigned | Unassigned | Unassigned | 620_0 | Clustered | 27 | VC_620_0 | 27 | 0.879 | 1 | 0.879 | 0 | 0 | 0 | 1 | Ref |
| 16319 | OP503983 | Unassigned | Unassigned | Unassigned | 620_0 | Clustered | 27 | VC_620_0 | 27 | 0.879 | 1 | 0.879 | 0 | 0 | 0 | 1 | Ref |
| 16320 | OP503984 | Unassigned | Unassigned | Unassigned | 620_0 | Clustered | 27 | VC_620_0 | 27 | 0.879 | 1 | 0.879 | 0 | 0 | 0 | 1 | Ref |
| 16321 | OP503985 | Unassigned | Unassigned | Unassigned | 620_0 | Clustered | 27 | VC_620_0 | 27 | 0.879 | 1 | 0.879 | 0 | 0 | 0 | 1 | Ref |
| 16322 | OP503986 | Unassigned | Unassigned | Unassigned | 620_0 | Clustered | 27 | VC_620_0 | 27 | 0.879 | 1 | 0.879 | 0 | 0 | 0 | 1 | Ref |
| 16323 | OP503987 | Unassigned | Unassigned | Unassigned | 620_0 | Clustered | 27 | VC_620_0 | 27 | 0.879 | 1 | 0.879 | 0 | 0 | 0 | 1 | Ref |
| 16324 | OP503988 | Unassigned | Unassigned | Unassigned | 620_0 | Clustered | 27 | VC_620_0 | 27 | 0.879 | 1 | 0.879 | 0 | 0 | 0 | 1 | Ref |
| 16325 | OP503992 | Unassigned | Unassigned | Unassigned | 708_0 | Clustered | 47 | VC_708_0 | 47 | 0.8654 | 1 | 0.8654 | 0 | 0 | 0 | 1 | Ref |
| 16326 | OP503993 | Unassigned | Unassigned | Unassigned | 708_0 | Clustered | 47 | VC_708_0 | 47 | 0.8654 | 1 | 0.8654 | 0 | 0 | 0 | 1 | Ref |
| 16327 | OP503994 | Unassigned | Unassigned | Unassigned | 708_0 | Clustered | 47 | VC_708_0 | 47 | 0.8654 | 1 | 0.8654 | 0 | 0 | 0 | 1 | Ref |
| 16328 | OP503995 | Unassigned | Unassigned | Unassigned | 708_0 | Clustered | 47 | VC_708_0 | 47 | 0.8654 | 1 | 0.8654 | 0 | 0 | 0 | 1 | Ref |
| 16329 | OP508218 | Unassigned | Unassigned | Unassigned | 699_0 | Clustered | 71 | VC_699_0 | 71 | 0.9352 | 1 | 0.9352 | 0 | 0 | 0 | 1 | Ref |
| 16330 | OP535471 | Unassigned | Unassigned | Unassigned | 592_0 | Clustered | 50 | VC_592_0 | 50 | 0.7314 | 1 | 0.7314 | 0 | 0 | 0 | 1 | Ref |
| 16331 | OP535472 | Unassigned | Unassigned | Unassigned | 592_0 | Clustered | 50 | VC_592_0 | 50 | 0.7314 | 1 | 0.7314 | 0 | 0 | 0 | 1 | Ref |
| 16332 | OP547477 | Unassigned | Unassigned | Unassigned | 882_0 | Clustered | 20 | VC_882_0 | 20 | 0.9917 | 1 | 0.9917 | 0 | 0 | 0 | 1 | Ref |
| 16333 | OP547478 | Unassigned | Unassigned | Unassigned | 904_2 | Clustered | 7 | VC_904_2 | 7 | 0.2857 | 1 | 0.2857 | 0 | 0 | 0 | 1 | Ref |
| 16334 | OP549864 | Unassigned | Unassigned | Unassigned | 1336_0 | Clustered | 3 | VC_1336_0 | 3 | 0.4948 | 0.99528984 | 0.4925 | 0 | 0 | 0 | 1 | Ref |
| 16335 | OP549865 | Unassigned | Unassigned | Unassigned | 1336_0 | Clustered | 3 | VC_1336_0 | 3 | 0.4948 | 0.99528984 | 0.4925 | 0 | 0 | 0 | 1 | Ref |
| 16336 | OP549866 | Unassigned | Unassigned | Unassigned | 1336_0 | Clustered | 3 | VC_1336_0 | 3 | 0.4948 | 0.99528984 | 0.4925 | 0 | 0 | 0 | 1 | Ref |
| 16337 | OP549867 | Unassigned | Unassigned | Unassigned | 1276_0 | Clustered | 7 | VC_1276_0 | 7 | 0.8695 | 0.99982476 | 0.8693 | 0 | 0 | 0 | 1 | Ref |
| 16338 | OP549868 | Unassigned | Unassigned | Unassigned | 1243_0 | Clustered | 52 | VC_1243_0 | 52 | 0.9126 | 0.92708311 | 0.8461 | 0 | 0 | 0 | 1 | Ref |
| 16339 | OP549869 | Unassigned | Unassigned | Unassigned | 1104_0 | Clustered | 154 | VC_1104_0 | 154 | 0.8484 | 1 | 0.8484 | 0 | 0 | 0 | 1 | Ref |
| 16340 | OP549870 | Unassigned | Unassigned | Unassigned | 1151_0 | Clustered | 9 | VC_1151_0 | 9 | 0.8548 | 1 | 0.8548 | 0 | 0 | 0 | 1 | Ref |
| 16341 | OP549871 | Unassigned | Unassigned | Unassigned | 1243_0 | Clustered | 52 | VC_1243_0 | 52 | 0.9126 | 0.92708311 | 0.8461 | 0 | 0 | 0 | 1 | Ref |
| 16342 | OP549872 | Unassigned | Unassigned | Unassigned | 1104_0 | Clustered | 154 | VC_1104_0 | 154 | 0.8484 | 1 | 0.8484 | 0 | 0 | 0 | 1 | Ref |
| 16343 | OP549873 | Unassigned | Unassigned | Unassigned | 1151_0 | Clustered | 9 | VC_1151_0 | 9 | 0.8548 | 1 | 0.8548 | 0 | 0 | 0 | 1 | Ref |
| 16344 | OP549874 | Unassigned | Unassigned | Unassigned | 1243_0 | Clustered | 52 | VC_1243_0 | 52 | 0.9126 | 0.92708311 | 0.8461 | 0 | 0 | 0 | 1 | Ref |
| 16345 | OP549875 | Unassigned | Unassigned | Unassigned | 1104_0 | Clustered | 154 | VC_1104_0 | 154 | 0.8484 | 1 | 0.8484 | 0 | 0 | 0 | 1 | Ref |
| 16346 | OP549876 | Unassigned | Unassigned | Unassigned | 1104_0 | Clustered | 154 | VC_1104_0 | 154 | 0.8484 | 1 | 0.8484 | 0 | 0 | 0 | 1 | Ref |
| 16347 | OP549877 | Unassigned | Unassigned | Unassigned | 1151_0 | Clustered | 9 | VC_1151_0 | 9 | 0.8548 | 1 | 0.8548 | 0 | 0 | 0 | 1 | Ref |
| 16348 | OP549878 | Unassigned | Unassigned | Unassigned | 1476_0 | Clustered | 2 | VC_1476_0 | 2 | 1 | 1 | 1 | 0 | 0 | 0 | 1 | Ref |
| 16349 | OP549879 | Unassigned | Unassigned | Unassigned | 1476_0 | Clustered | 2 | VC_1476_0 | 2 | 1 | 1 | 1 | 0 | 0 | 0 | 1 | Ref |
| 16350 | OP549880 | Unassigned | Unassigned | Unassigned | 1477_0 | Clustered | 3 | VC_1477_0 | 3 | 1 | 1 | 1 | 0 | 0 | 0 | 1 | Ref |
| 16351 | OP555980 | Unassigned | Unassigned | Unassigned | 283_4 | Clustered | 5 | VC_283_4 | 5 | 0.0112 | 0.99999969 | 0.0112 | 0 | 0 | 0 | 1 | Ref |
| 16352 | OP555981 | Unassigned | Unassigned | Unassigned | 283_4 | Clustered | 5 | VC_283_4 | 5 | 0.0112 | 0.99999969 | 0.0112 | 0 | 0 | 0 | 1 | Ref |
| 16353 | OP555982 | Unassigned | Unassigned | Unassigned | 283_4 | Clustered | 5 | VC_283_4 | 5 | 0.0112 | 0.99999969 | 0.0112 | 0 | 0 | 0 | 1 | Ref |
| 16354 | OP556577 | Unassigned | Unassigned | Unassigned | 423_0 | Clustered | 45 | VC_423_0 | 45 | 0.9484 | 1 | 0.9484 | 0 | 0 | 0 | 1 | Ref |
| 16355 | OP559177 | Unassigned | Unassigned | Unassigned | 695_0 | Clustered | 48 | VC_695_0 | 48 | 0.8162 | 1 | 0.8162 | 0 | 0 | 0 | 1 | Ref |
| 16356 | OP559178 | Unassigned | Unassigned | Unassigned | 91_1 | Clustered/Singleton | 1 | VC_91_1 | 1 | 0 | 1 | 0 | 0 | 0 | 0 | 1 | Ref |
| 16357 | OP562383 | Unassigned | Unassigned | Unassigned | 847_1 | Clustered | 26 | VC_847_1 | 26 | 0.6323 | 1 | 0.6323 | 0 | 0 | 0 | 0.9815 | Ref |
| 16358 | OP572230 | Unassigned | Unassigned | Unassigned | 283_4 | Clustered | 5 | VC_283_4 | 5 | 0.0112 | 0.99999969 | 0.0112 | 0 | 0 | 0 | 1 | Ref |
| 16359 | OP580516 | Unassigned | Unassigned | Unassigned | 1034_0 | Clustered | 2 | VC_1034_0 | 2 | 0.071 | 0.95326364 | 0.0677 | 0 | 0 | 0 | 1 | Ref |
| 16360 | OP580517 | Unassigned | Unassigned | Unassigned | 1343_0 | Clustered | 21 | VC_1343_0 | 21 | 0.9843 | 1 | 0.9843 | 0 | 0 | 0 | 1 | Ref |

| | | | | | | | | | | | | | | | | | |
|---|---|---|---|---|---|---|---|---|---|---|---|---|---|---|---|---|---|
| 16361 | OP580518 | Unassigned | Unassigned | Unassigned | 1344_0 | Clustered | 78 | VC_1344_0 | 78 | 0.9732 | 1 | 0.9732 | 0 | 0 | 0 | 1 | Ref |
| 16362 | OP584903 | Unassigned | Unassigned | Unassigned | 8_0 | Clustered | 58 | VC_8_0 | 58 | 0.8306 | 1 | 0.8306 | 0 | 0 | 0 | 0.9946 | Ref |
| 16363 | OP584904 | Unassigned | Unassigned | Unassigned | 8_0 | Clustered | 58 | VC_8_0 | 58 | 0.8306 | 1 | 0.8306 | 0 | 0 | 0 | 0.9946 | Ref |
| 16364 | OP585103 | Unassigned | Unassigned | Unassigned | 492_1 | Clustered | 20 | VC_492_1 | 20 | 0.4176 | 1 | 0.4176 | 0 | 0 | 0 | 1 | Ref |
| 16365 | OP585104 | Unassigned | Unassigned | Unassigned | 698_0 | Clustered | 26 | VC_698_0 | 26 | 0.9366 | 1 | 0.9366 | 0 | 0 | 0 | 1 | Ref |
| 16366 | OP585105 | Unassigned | Unassigned | Unassigned | 700_0 | Clustered | 10 | VC_700_0 | 10 | 0.8076 | 1 | 0.8076 | 0 | 0 | 0 | 1 | Ref |
| 16367 | OP595601 | Unassigned | Unassigned | Unassigned | 882_0 | Clustered | 20 | VC_882_0 | 20 | 0.9917 | 1 | 0.9917 | 0 | 0 | 0 | 1 | Ref |
| 16368 | OP610151 | Unassigned | Unassigned | Unassigned | 133_22 | Clustered | 13 | VC_133_22 | 13 | 0.0804 | 1 | 0.0804 | 0 | 0 | 0 | 0.8205 | Ref |
| 16369 | OP611406 | Unassigned | Unassigned | Unassigned | 937_0 | Clustered | 40 | VC_937_0 | 40 | 0.6809 | 1 | 0.6809 | 0 | 0 | 0 | 0.9359 | Ref |
| 16370 | OP617560 | Unassigned | Unassigned | Unassigned | 83_0 | Clustered | 6 | VC_83_0 | 6 | 0.6436 | 1 | 0.6436 | 0 | 0 | 0 | 1 | Ref |
| 16371 | OP617742 | Unassigned | Unassigned | Unassigned | 938_0 | Clustered | 99 | VC_938_0 | 99 | 0.9119 | 1 | 0.9119 | 0 | 0 | 0 | 0.9928 | Ref |
| 16372 | OP617743 | Unassigned | Unassigned | Unassigned | 848_1 | Clustered/Singleton | 1 | VC_848_1 | 1 | 0 | 1 | 0 | 0 | 0 | 0 | 1 | Ref |
| 16373 | OP617744 | Unassigned | Unassigned | Unassigned | 1049_0 | Clustered | 8 | VC_1049_0 | 8 | 0.6413 | 1 | 0.6413 | 0 | 0 | 0 | 1 | Ref |
| 16374 | OP617745 | Unassigned | Unassigned | Unassigned | 164_10 | Clustered | 5 | VC_164_10 | 5 | 0.0615 | 0.96483582 | 0.0593 | 0 | 0 | 0 | 1 | Ref |
| 16375 | OP617746 | Unassigned | Unassigned | Unassigned | 668_1 | Clustered/Singleton | 1 | VC_668_1 | 1 | 0 | 1 | 0 | 0 | 0 | 0 | 1 | Ref |
| 16376 | OP617747 | Unassigned | Unassigned | Unassigned | 385_0 | Clustered | 51 | VC_385_0 | 51 | 0.8754 | 1 | 0.8754 | 0 | 0 | 0 | 1 | Ref |
| 16377 | OP617748 | Unassigned | Unassigned | Unassigned | 491_0 | Clustered | 36 | VC_491_0 | 36 | 0.5336 | 1 | 0.5336 | 0 | 0 | 0 | 0.9714 | Ref |
| 16378 | OP650937 | Unassigned | Unassigned | Unassigned | 300_0 | Clustered | 3 | VC_300_0 | 3 | 0.1218 | 0.99841701 | 0.1216 | 0 | 0 | 0 | 0.6667 | Ref |
| 16379 | OP684128 | Unassigned | Unassigned | Unassigned | 668_0 | Clustered | 25 | VC_668_0 | 25 | 0.5539 | 1 | 0.5539 | 0 | 0 | 0 | 0.96 | Ref |
| 16380 | OU342754 | Unassigned | Unassigned | Unassigned | 823_0 | Clustered | 124 | VC_823_0 | 124 | 0.9492 | 1 | 0.9492 | 0 | 0 | 0 | 1 | Ref |
| 16381 | OU342755 | Unassigned | Unassigned | Unassigned | 938_0 | Clustered | 99 | VC_938_0 | 99 | 0.9119 | 1 | 0.9119 | 0 | 0 | 0 | 0.9928 | Ref |
| 16382 | OU343167 | Unassigned | Unassigned | Unassigned | 344_1 | Clustered | 3 | VC_344_1 | 3 | 0.0359 | 0.93348877 | 0.0335 | 0 | 0 | 0 | 1 | Ref |
| 16383 | OU509533 | Unassigned | Unassigned | Unassigned | 111_0 | Clustered | 91 | VC_111_0 | 91 | 0.9587 | 1 | 0.9587 | 0 | 0 | 0 | 1 | Ref |
| 16384 | OU509534 | Unassigned | Unassigned | Unassigned | 111_0 | Clustered | 91 | VC_111_0 | 91 | 0.9587 | 1 | 0.9587 | 0 | 0 | 0 | 1 | Ref |
| 16385 | OU509535 | Unassigned | Unassigned | Unassigned | 164_10 | Clustered | 5 | VC_164_10 | 5 | 0.0615 | 0.96483582 | 0.0593 | 0 | 0 | 0 | 1 | Ref |
| 16386 | OU509536 | Unassigned | Unassigned | Unassigned | 491_0 | Clustered | 36 | VC_491_0 | 36 | 0.5336 | 1 | 0.5336 | 0 | 0 | 0 | 0.9714 | Ref |
| 16387 | OU734268 | Unassigned | Unassigned | Unassigned | 942_0 | Clustered | 36 | VC_942_0 | 36 | 0.7848 | 1 | 0.7848 | 0 | 0 | 0 | 1 | Ref |
| 16388 | OU784549 | Unassigned | Unassigned | Unassigned | 228_0 | Clustered | 60 | VC_228_0 | 60 | 0.942 | 1 | 0.942 | 0 | 0 | 0 | 1 | Ref |
| 16389 | OU784746 | Unassigned | Unassigned | Unassigned | 228_0 | Clustered | 60 | VC_228_0 | 60 | 0.942 | 1 | 0.942 | 0 | 0 | 0 | 1 | Ref |
| 16390 | OU784750 | Unassigned | Unassigned | Unassigned | 228_0 | Clustered | 60 | VC_228_0 | 60 | 0.942 | 1 | 0.942 | 0 | 0 | 0 | 1 | Ref |
| 16391 | OV032860 | Unassigned | Unassigned | Unassigned | 904_2 | Clustered | 7 | VC_904_2 | 7 | 0.2857 | 1 | 0.2857 | 0 | 0 | 0 | 1 | Ref |
| 16392 | OV032902 | Unassigned | Unassigned | Unassigned | 346_2 | Clustered/Singleton | 1 | VC_346_2 | 1 | 0 | 1 | 0 | 0 | 0 | 0 | 1 | Ref |
| 16393 | OV049961 | Unassigned | Unassigned | Unassigned | 779_0 | Clustered | 37 | VC_779_0 | 37 | 0.5682 | 1 | 0.5682 | 0 | 0 | 0 | 1 | Ref |
| 16394 | OV101294 | Unassigned | Unassigned | Unassigned | 772_19 | Clustered | 16 | VC_772_19 | 16 | 0.0975 | 0.9970466 | 0.0972 | 0 | 0 | 0 | 0.8083 | Ref |
| 16395 | OV121131 | Unassigned | Unassigned | Unassigned | 162_0 | Clustered | 18 | VC_162_0 | 18 | 0.7403 | 1 | 0.7403 | 0 | 0 | 0 | 0.8627 | Ref |
| 16396 | OV192280 | Unassigned | Unassigned | Unassigned | 228_0 | Clustered | 60 | VC_228_0 | 60 | 0.942 | 1 | 0.942 | 0 | 0 | 0 | 1 | Ref |
| 16397 | OV696608 | Unassigned | Unassigned | Unassigned | 30_1 | Clustered | 16 | VC_30_1 | 16 | 0.1973 | 1 | 0.1973 | 0 | 0 | 0 | 0.8333 | Ref |
| 16398 | OV696610 | Unassigned | Unassigned | Unassigned | 51_0 | Clustered | 10 | VC_51_0 | 10 | 0.1741 | 1 | 0.1741 | 0 | 0 | 0 | 1 | Ref |
| 16399 | OV696611 | Unassigned | Unassigned | Unassigned | 6_0 | Clustered | 44 | VC_6_0 | 44 | 0.908 | 1 | 0.908 | 0 | 0 | 0 | 1 | Ref |
| 16400 | OV696612 | Unassigned | Unassigned | Unassigned | 43_0 | Clustered | 38 | VC_43_0 | 38 | 0.5885 | 1 | 0.5885 | 0 | 0 | 0 | 0.9659 | Ref |
| 16401 | OV696614 | Unassigned | Unassigned | Unassigned | 723_0 | Clustered | 9 | VC_723_0 | 9 | 0.7138 | 1 | 0.7138 | 0 | 0 | 0 | 1 | Ref |
| 16402 | OV696617 | Unassigned | Unassigned | Unassigned | 744_0 | Clustered | 10 | VC_744_0 | 10 | 0.7049 | 1 | 0.7049 | 0 | 0 | 0 | 1 | Ref |
| 16403 | OV696619 | Unassigned | Unassigned | Unassigned | 1159_0 | Clustered | 3 | VC_1159_0 | 3 | 0.3304 | 0.98548359 | 0.3256 | 0 | 0 | 0 | 1 | Ref |
| 16404 | OV696621 | Unassigned | Unassigned | Unassigned | 861_0 | Clustered | 9 | VC_861_0 | 9 | 0.5869 | 1 | 0.5869 | 0 | 0 | 0 | 1 | Ref |
| 16405 | OV754623 | Unassigned | Unassigned | Unassigned | 286_13 | Clustered | 10 | VC_286_13 | 10 | 0.0698 | 0.99999929 | 0.0698 | 0 | 0 | 0 | 0.9556 | Ref |
| 16406 | OV876900 | Unassigned | Unassigned | Unassigned | 283_140 | Clustered | 2 | VC_283_140 | 2 | 0.0028 | 0.93811915 | 0.0026 | 0 | 0 | 0 | 1 | Ref |
| 16407 | OV876907 | Unassigned | Unassigned | Unassigned | 290_6 | Clustered | 12 | VC_290_6 | 12 | 0.1149 | 1 | 0.1149 | 0 | 0 | 0 | 0.8182 | Ref |
| 16408 | OV877085 | Unassigned | Unassigned | Unassigned | 283_140 | Clustered | 2 | VC_283_140 | 2 | 0.0028 | 0.93811915 | 0.0026 | 0 | 0 | 0 | 1 | Ref |
| 16409 | OV877339 | Unassigned | Unassigned | Unassigned | 937_0 | Clustered | 40 | VC_937_0 | 40 | 0.6809 | 1 | 0.6809 | 0 | 0 | 0 | 0.9359 | Ref |
| 16410 | OV877387 | Unassigned | Unassigned | Unassigned | 937_0 | Clustered | 40 | VC_937_0 | 40 | 0.6809 | 1 | 0.6809 | 0 | 0 | 0 | 0.9359 | Ref |
| 16411 | OV877511 | Unassigned | Unassigned | Unassigned | 286_13 | Clustered | 10 | VC_286_13 | 10 | 0.0698 | 0.99999929 | 0.0698 | 0 | 0 | 0 | 0.9556 | Ref |
| 16412 | OV877765 | Unassigned | Unassigned | Unassigned | 290_6 | Clustered | 12 | VC_290_6 | 12 | 0.1149 | 1 | 0.1149 | 0 | 0 | 0 | 0.8182 | Ref |
| 16413 | OV877766 | Unassigned | Unassigned | Unassigned | 937_0 | Clustered | 40 | VC_937_0 | 40 | 0.6809 | 1 | 0.6809 | 0 | 0 | 0 | 0.9359 | Ref |
| 16414 | OW251746 | Unassigned | Unassigned | Unassigned | 938_0 | Clustered | 99 | VC_938_0 | 99 | 0.9119 | 1 | 0.9119 | 0 | 0 | 0 | 0.9928 | Ref |
| 16415 | OW991345 | Unassigned | Unassigned | Unassigned | 779_0 | Clustered | 37 | VC_779_0 | 37 | 0.5682 | 1 | 0.5682 | 0 | 0 | 0 | 1 | Ref |
| 16416 | OX001802 | Unassigned | Unassigned | Unassigned | 590_0 | Clustered | 27 | VC_590_0 | 27 | 0.9947 | 1 | 0.9947 | 0 | 0 | 0 | 1 | Ref |
| 16417 | OX016465 | Unassigned | Unassigned | Unassigned | 329_3 | Clustered | 24 | VC_329_3 | 24 | 0.1206 | 1 | 0.1206 | 0 | 0 | 0 | 0.7283 | Ref |
| 16418 | OX090893 | Unassigned | Unassigned | Unassigned | 848_0 | Clustered | 28 | VC_848_0 | 28 | 0.7927 | 1 | 0.7927 | 0 | 0 | 0 | 1 | Ref |
| 16419 | OX241425 | Unassigned | Unassigned | Unassigned | 115_0 | Clustered | 2 | VC_115_0 | 2 | 0.4117 | 0.90744399 | 0.3736 | 0 | 0 | 0 | 1 | Ref |
| 16420 | OX241426 | Unassigned | Unassigned | Unassigned | 501_0 | Clustered | 16 | VC_501_0 | 16 | 0.9985 | 1 | 0.9985 | 0 | 0 | 0 | 1 | Ref |
| 16421 | OX241427 | Unassigned | Unassigned | Unassigned | 1050_1 | Clustered | 29 | VC_1050_1 | 29 | 0.4861 | 1 | 0.4861 | 0 | 0 | 0 | 0.9581 | Ref |
| 16422 | OX241428 | Unassigned | Unassigned | Unassigned | 501_0 | Clustered | 16 | VC_501_0 | 16 | 0.9985 | 1 | 0.9985 | 0 | 0 | 0 | 1 | Ref |
| 16423 | OX241429 | Unassigned | Unassigned | Unassigned | 646_0 | Clustered | 29 | VC_646_0 | 29 | 0.9131 | 1 | 0.9131 | 0 | 0 | 0 | 1 | Ref |
| 16424 | OX241430 | Unassigned | Unassigned | Unassigned | 501_0 | Clustered | 16 | VC_501_0 | 16 | 0.9985 | 1 | 0.9985 | 0 | 0 | 0 | 1 | Ref |
| 16425 | OX241431 | Unassigned | Unassigned | Unassigned | 501_0 | Clustered | 16 | VC_501_0 | 16 | 0.9985 | 1 | 0.9985 | 0 | 0 | 0 | 1 | Ref |
| 16426 | OX241432 | Unassigned | Unassigned | Unassigned | 501_0 | Clustered | 16 | VC_501_0 | 16 | 0.9985 | 1 | 0.9985 | 0 | 0 | 0 | 1 | Ref |
| 16427 | OX241434 | Unassigned | Unassigned | Unassigned | 903_0 | Clustered | 7 | VC_903_0 | 7 | 0.7377 | 1 | 0.7377 | 0 | 0 | 0 | 1 | Ref |
| 16428 | OX241435 | Unassigned | Unassigned | Unassigned | 758_0 | Clustered | 18 | VC_758_0 | 18 | 0.738 | 1 | 0.738 | 0 | 0 | 0 | 1 | Ref |
| 16429 | OX241436 | Unassigned | Unassigned | Unassigned | 758_0 | Clustered | 18 | VC_758_0 | 18 | 0.738 | 1 | 0.738 | 0 | 0 | 0 | 1 | Ref |
| 16430 | OX241437 | Unassigned | Unassigned | Unassigned | 493_3 | Clustered/Singleton | 1 | VC_493_3 | 1 | 0 | 1 | 0 | 0 | 0 | 0 | 1 | Ref |
| 16431 | OX241438 | Unassigned | Unassigned | Unassigned | 646_0 | Clustered | 29 | VC_646_0 | 29 | 0.9131 | 1 | 0.9131 | 0 | 0 | 0 | 1 | Ref |
| 16432 | OX241441 | Unassigned | Unassigned | Unassigned | 501_0 | Clustered | 16 | VC_501_0 | 16 | 0.9985 | 1 | 0.9985 | 0 | 0 | 0 | 1 | Ref |
| 16433 | OX241445 | Unassigned | Unassigned | Unassigned | 758_0 | Clustered | 18 | VC_758_0 | 18 | 0.738 | 1 | 0.738 | 0 | 0 | 0 | 1 | Ref |
| 16434 | OX241446 | Unassigned | Unassigned | Unassigned | 758_0 | Clustered | 18 | VC_758_0 | 18 | 0.738 | 1 | 0.738 | 0 | 0 | 0 | 1 | Ref |
| 16435 | OX241447 | Unassigned | Unassigned | Unassigned | 758_0 | Clustered | 18 | VC_758_0 | 18 | 0.738 | 1 | 0.738 | 0 | 0 | 0 | 1 | Ref |
| 16436 | OX241448 | Unassigned | Unassigned | Unassigned | 501_0 | Clustered | 16 | VC_501_0 | 16 | 0.9985 | 1 | 0.9985 | 0 | 0 | 0 | 1 | Ref |
| 16437 | OX241449 | Unassigned | Unassigned | Unassigned | 501_0 | Clustered | 16 | VC_501_0 | 16 | 0.9985 | 1 | 0.9985 | 0 | 0 | 0 | 1 | Ref |
| 16438 | OX241455 | Unassigned | Unassigned | Unassigned | 493_4 | Clustered/Singleton | 1 | VC_493_4 | 1 | 0 | 1 | 0 | 0 | 0 | 0 | 1 | Ref |
| 16439 | OX241494 | Unassigned | Unassigned | Unassigned | 501_0 | Clustered | 16 | VC_501_0 | 16 | 0.9985 | 1 | 0.9985 | 0 | 0 | 0 | 1 | Ref |
| 16440 | OX241498 | Unassigned | Unassigned | Unassigned | 646_0 | Clustered | 29 | VC_646_0 | 29 | 0.9131 | 1 | 0.9131 | 0 | 0 | 0 | 1 | Ref |
| 16441 | OX241501 | Unassigned | Unassigned | Unassigned | 758_0 | Clustered | 18 | VC_758_0 | 18 | 0.738 | 1 | 0.738 | 0 | 0 | 0 | 1 | Ref |
| 16442 | OX241504 | Unassigned | Unassigned | Unassigned | 758_0 | Clustered | 18 | VC_758_0 | 18 | 0.738 | 1 | 0.738 | 0 | 0 | 0 | 1 | Ref |
| 16443 | OX241556 | Unassigned | Unassigned | Unassigned | 1050_1 | Clustered | 29 | VC_1050_1 | 29 | 0.4861 | 1 | 0.4861 | 0 | 0 | 0 | 0.9581 | Ref |
| 16444 | OX241557 | Unassigned | Unassigned | Unassigned | 646_0 | Clustered | 29 | VC_646_0 | 29 | 0.9131 | 1 | 0.9131 | 0 | 0 | 0 | 1 | Ref |
| 16445 | OX241558 | Unassigned | Unassigned | Unassigned | 646_0 | Clustered | 29 | VC_646_0 | 29 | 0.9131 | 1 | 0.9131 | 0 | 0 | 0 | 1 | Ref |
| 16446 | OX241559 | Unassigned | Unassigned | Unassigned | 758_0 | Clustered | 18 | VC_758_0 | 18 | 0.738 | 1 | 0.738 | 0 | 0 | 0 | 1 | Ref |
| 16447 | OX241560 | Unassigned | Unassigned | Unassigned | 758_0 | Clustered | 18 | VC_758_0 | 18 | 0.738 | 1 | 0.738 | 0 | 0 | 0 | 1 | Ref |
| 16448 | OX241561 | Unassigned | Unassigned | Unassigned | 1041_0 | Clustered | 9 | VC_1041_0 | 9 | 0.6541 | 1 | 0.6541 | 0 | 0 | 0 | 1 | Ref |
| 16449 | OX241562 | Unassigned | Unassigned | Unassigned | 478_0 | Clustered | 2 | VC_478_0 | 2 | 0.3505 | 0.94766489 | 0.3321 | 0 | 0 | 0 | 1 | Ref |
| 16450 | OX241563 | Unassigned | Unassigned | Unassigned | 1050_1 | Clustered | 29 | VC_1050_1 | 29 | 0.4861 | 1 | 0.4861 | 0 | 0 | 0 | 0.9581 | Ref |
| 16451 | OX241566 | Unassigned | Unassigned | Unassigned | 926_0 | Clustered | 3 | VC_926_0 | 3 | 0.9445 | 0.99430386 | 0.9391 | 0 | 0 | 0 | 1 | Ref |
| 16452 | OX241567 | Unassigned | Unassigned | Unassigned | 501_0 | Clustered | 16 | VC_501_0 | 16 | 0.9985 | 1 | 0.9985 | 0 | 0 | 0 | 1 | Ref |
| 16453 | OX241568 | Unassigned | Unassigned | Unassigned | 501_0 | Clustered | 16 | VC_501_0 | 16 | 0.9985 | 1 | 0.9985 | 0 | 0 | 0 | 1 | Ref |
| 16454 | OX241569 | Unassigned | Unassigned | Unassigned | 1041_0 | Clustered | 9 | VC_1041_0 | 9 | 0.6541 | 1 | 0.6541 | 0 | 0 | 0 | 1 | Ref |
| 16455 | OX241570 | Unassigned | Unassigned | Unassigned | 926_0 | Clustered | 3 | VC_926_0 | 3 | 0.9445 | 0.99430386 | 0.9391 | 0 | 0 | 0 | 1 | Ref |
| 16456 | OX241572 | Unassigned | Unassigned | Unassigned | 175_0 | Clustered | 4 | VC_175_0 | 4 | 0.3847 | 0.99998259 | 0.3846 | 0 | 0 | 0 | 1 | Ref |
| 16457 | OX241573 | Unassigned | Unassigned | Unassigned | 642_5 | Clustered | 4 | VC_642_5 | 4 | 0.0902 | 0.99996664 | 0.0902 | 0 | 0 | 0 | 1 | Ref |
| 16458 | OX241574 | Unassigned | Unassigned | Unassigned | 642_1 | Clustered/Singleton | 1 | VC_642_1 | 1 | 0 | 1 | 0 | 0 | 0 | 0 | 1 | Ref |
| 16459 | OX241575 | Unassigned | Unassigned | Unassigned | 646_0 | Clustered | 29 | VC_646_0 | 29 | 0.9131 | 1 | 0.9131 | 0 | 0 | 0 | 1 | Ref |
| 16460 | OX241591 | Unassigned | Unassigned | Unassigned | 646_0 | Clustered | 29 | VC_646_0 | 29 | 0.9131 | 1 | 0.9131 | 0 | 0 | 0 | 1 | Ref |
| 16461 | OX335359 | Unassigned | Unassigned | Unassigned | 823_0 | Clustered | 124 | VC_823_0 | 124 | 0.9492 | 1 | 0.9492 | 0 | 0 | 0 | 1 | Ref |
| 16462 | OX335360 | Unassigned | Unassigned | Unassigned | 823_0 | Clustered | 124 | VC_823_0 | 124 | 0.9492 | 1 | 0.9492 | 0 | 0 | 0 | 1 | Ref |
| 16463 | OX335361 | Unassigned | Unassigned | Unassigned | 937_0 | Clustered | 40 | VC_937_0 | 40 | 0.6809 | 1 | 0.6809 | 0 | 0 | 0 | 0.9359 | Ref |
| 16464 | OX335369 | Unassigned | Unassigned | Unassigned | 938_0 | Clustered | 99 | VC_938_0 | 99 | 0.9119 | 1 | 0.9119 | 0 | 0 | 0 | 0.9928 | Ref |
| 16465 | OX335370 | Unassigned | Unassigned | Unassigned | 938_0 | Clustered | 99 | VC_938_0 | 99 | 0.9119 | 1 | 0.9119 | 0 | 0 | 0 | 0.9928 | Ref |
| 16466 | OX335371 | Unassigned | Unassigned | Unassigned | 938_0 | Clustered | 99 | VC_938_0 | 99 | 0.9119 | 1 | 0.9119 | 0 | 0 | 0 | 0.9928 | Ref |
| 16467 | OX335375 | Unassigned | Unassigned | Unassigned | 823_0 | Clustered | 124 | VC_823_0 | 124 | 0.9492 | 1 | 0.9492 | 0 | 0 | 0 | 1 | Ref |
| 16468 | OX335376 | Unassigned | Unassigned | Unassigned | 823_0 | Clustered | 124 | VC_823_0 | 124 | 0.9492 | 1 | 0.9492 | 0 | 0 | 0 | 1 | Ref |
| 16469 | OX335377 | Unassigned | Unassigned | Unassigned | 823_0 | Clustered | 124 | VC_823_0 | 124 | 0.9492 | 1 | 0.9492 | 0 | 0 | 0 | 1 | Ref |
| 16470 | OX335378 | Unassigned | Unassigned | Unassigned | 388_0 | Clustered | 4 | VC_388_0 | 4 | 0.0345 | 0.99998812 | 0.0345 | 0 | 0 | 0 | 1 | Ref |
| 16471 | OX335379 | Unassigned | Unassigned | Unassigned | 111_0 | Clustered | 91 | VC_111_0 | 91 | 0.9587 | 1 | 0.9587 | 0 | 0 | 0 | 1 | Ref |
| 16472 | OX335380 | Unassigned | Unassigned | Unassigned | 938_0 | Clustered | 99 | VC_938_0 | 99 | 0.9119 | 1 | 0.9119 | 0 | 0 | 0 | 0.9928 | Ref |
| 16473 | OX335381 | Unassigned | Unassigned | Unassigned | 938_0 | Clustered | 99 | VC_938_0 | 99 | 0.9119 | 1 | 0.9119 | 0 | 0 | 0 | 0.9928 | Ref |
| 16474 | OX335382 | Unassigned | Unassigned | Unassigned | 452_0 | Clustered | 66 | VC_452_0 | 66 | 0.9248 | 1 | 0.9248 | 0 | 0 | 0 | 1 | Ref |
| 16475 | OX335383 | Unassigned | Unassigned | Unassigned | 938_0 | Clustered | 99 | VC_938_0 | 99 | 0.9119 | 1 | 0.9119 | 0 | 0 | 0 | 0.9928 | Ref |
| 16476 | OX335384 | Unassigned | Unassigned | Unassigned | 938_0 | Clustered | 99 | VC_938_0 | 99 | 0.9119 | 1 | 0.9119 | 0 | 0 | 0 | 0.9928 | Ref |
| 16477 | OX335385 | Unassigned | Unassigned | Unassigned | 938_0 | Clustered | 99 | VC_938_0 | 99 | 0.9119 | 1 | 0.9119 | 0 | 0 | 0 | 0.9928 | Ref |
| 16478 | OX335386 | Unassigned | Unassigned | Unassigned | 938_0 | Clustered | 99 | VC_938_0 | 99 | 0.9119 | 1 | 0.9119 | 0 | 0 | 0 | 0.9928 | Ref |
| 16479 | OX335387 | Unassigned | Unassigned | Unassigned | 938_0 | Clustered | 99 | VC_938_0 | 99 | 0.9119 | 1 | 0.9119 | 0 | 0 | 0 | 0.9928 | Ref |
| 16480 | OX335388 | Unassigned | Unassigned | Unassigned | 938_0 | Clustered | 99 | VC_938_0 | 99 | 0.9119 | 1 | 0.9119 | 0 | 0 | 0 | 0.9928 | Ref |
| 16481 | OX335389 | Unassigned | Unassigned | Unassigned | 823_0 | Clustered | 124 | VC_823_0 | 124 | 0.9492 | 1 | 0.9492 | 0 | 0 | 0 | 1 | Ref |
| 16482 | OX335390 | Unassigned | Unassigned | Unassigned | 823_0 | Clustered | 124 | VC_823_0 | 124 | 0.9492 | 1 | 0.9492 | 0 | 0 | 0 | 1 | Ref |
| 16483 | OX335391 | Unassigned | Unassigned | Unassigned | 938_0 | Clustered | 99 | VC_938_0 | 99 | 0.9119 | 1 | 0.9119 | 0 | 0 | 0 | 0.9928 | Ref |
| 16484 | OX335392 | Unassigned | Unassigned | Unassigned | 823_0 | Clustered | 124 | VC_823_0 | 124 | 0.9492 | 1 | 0.9492 | 0 | 0 | 0 | 1 | Ref |
| 16485 | OX335393 | Unassigned | Unassigned | Unassigned | 938_0 | Clustered | 99 | VC_938_0 | 99 | 0.9119 | 1 | 0.9119 | 0 | 0 | 0 | 0.9928 | Ref |
| 16486 | OX335394 | Unassigned | Unassigned | Unassigned | 823_0 | Clustered | 124 | VC_823_0 | 124 | 0.9492 | 1 | 0.9492 | 0 | 0 | 0 | 1 | Ref |
| 16487 | OX335395 | Unassigned | Unassigned | Unassigned | 937_0 | Clustered | 40 | VC_937_0 | 40 | 0.6809 | 1 | 0.6809 | 0 | 0 | 0 | 0.9359 | Ref |
| 16488 | OX335396 | Unassigned | Unassigned | Unassigned | 668_0 | Clustered | 25 | VC_668_0 | 25 | 0.5539 | 1 | 0.5539 | 0 | 0 | 0 | 0.96 | Ref |
| 16489 | OX335398 | Unassigned | Unassigned | Unassigned | 823_0 | Clustered | 124 | VC_823_0 | 124 | 0.9492 | 1 | 0.9492 | 0 | 0 | 0 | 1 | Ref |
| 16490 | OX335400 | Unassigned | Unassigned | Unassigned | 938_0 | Clustered | 99 | VC_938_0 | 99 | 0.9119 | 1 | 0.9119 | 0 | 0 | 0 | 0.9928 | Ref |
| 16491 | OX335401 | Unassigned | Unassigned | Unassigned | 823_0 | Clustered | 124 | VC_823_0 | 124 | 0.9492 | 1 | 0.9492 | 0 | 0 | 0 | 1 | Ref |
| 16492 | OX335402 | Unassigned | Unassigned | Unassigned | 132_1 | Clustered | 17 | VC_132_1 | 17 | 0.1738 | 1 | 0.1738 | 0 | 0 | 0 | 0.9853 | Ref |
| 16493 | OX335403 | Unassigned | Unassigned | Unassigned | 132_1 | Clustered | 17 | VC_132_1 | 17 | 0.1738 | 1 | 0.1738 | 0 | 0 | 0 | 0.9853 | Ref |
| 16494 | OX335404 | Unassigned | Unassigned | Unassigned | 938_0 | Clustered | 99 | VC_938_0 | 99 | 0.9119 | 1 | 0.9119 | 0 | 0 | 0 | 0.9928 | Ref |
| 16495 | OX335405 | Unassigned | Unassigned | Unassigned | 132_1 | Clustered | 17 | VC_132_1 | 17 | 0.1738 | 1 | 0.1738 | 0 | 0 | 0 | 0.9853 | Ref |
| 16496 | OX335407 | Unassigned | Unassigned | Unassigned | 668_0 | Clustered | 25 | VC_668_0 | 25 | 0.5539 | 1 | 0.5539 | 0 | 0 | 0 | 0.96 | Ref |
| 16497 | OX335408 | Unassigned | Unassigned | Unassigned | 823_0 | Clustered | 124 | VC_823_0 | 124 | 0.9492 | 1 | 0.9492 | 0 | 0 | 0 | 1 | Ref |
| 16498 | OX335409 | Unassigned | Unassigned | Unassigned | 823_0 | Clustered | 124 | VC_823_0 | 124 | 0.9492 | 1 | 0.9492 | 0 | 0 | 0 | 1 | Ref |
| 16499 | OX335410 | Unassigned | Unassigned | Unassigned | 937_0 | Clustered | 40 | VC_937_0 | 40 | 0.6809 | 1 | 0.6809 | 0 | 0 | 0 | 0.9359 | Ref |
| 16500 | OX335411 | Unassigned | Unassigned | Unassigned | 938_0 | Clustered | 99 | VC_938_0 | 99 | 0.9119 | 1 | 0.9119 | 0 | 0 | 0 | 0.9928 | Ref |
| 16501 | OX335412 | Unassigned | Unassigned | Unassigned | 938_0 | Clustered | 99 | VC_938_0 | 99 | 0.9119 | 1 | 0.9119 | 0 | 0 | 0 | 0.9928 | Ref |
| 16502 | OX335413 | Unassigned | Unassigned | Unassigned | 823_0 | Clustered | 124 | VC_823_0 | 124 | 0.9492 | 1 | 0.9492 | 0 | 0 | 0 | 1 | Ref |
| 16503 | OX335414 | Unassigned | Unassigned | Unassigned | 823_0 | Clustered | 124 | VC_823_0 | 124 | 0.9492 | 1 | 0.9492 | 0 | 0 | 0 | 1 | Ref |
| 16504 | OX335416 | Unassigned | Unassigned | Unassigned | 132_1 | Clustered | 17 | VC_132_1 | 17 | 0.1738 | 1 | 0.1738 | 0 | 0 | 0 | 0.9853 | Ref |
| 16505 | OX335417 | Unassigned | Unassigned | Unassigned | 938_0 | Clustered | 99 | VC_938_0 | 99 | 0.9119 | 1 | 0.9119 | 0 | 0 | 0 | 0.9928 | Ref |
| 16506 | OX335418 | Unassigned | Unassigned | Unassigned | 823_0 | Clustered | 124 | VC_823_0 | 124 | 0.9492 | 1 | 0.9492 | 0 | 0 | 0 | 1 | Ref |
| 16507 | OX335419 | Unassigned | Unassigned | Unassigned | 385_0 | Clustered | 51 | VC_385_0 | 51 | 0.8754 | 1 | 0.8754 | 0 | 0 | 0 | 1 | Ref |
| 16508 | OX335420 | Unassigned | Unassigned | Unassigned | 385_0 | Clustered | 51 | VC_385_0 | 51 | 0.8754 | 1 | 0.8754 | 0 | 0 | 0 | 1 | Ref |
| 16509 | OX335421 | Unassigned | Unassigned | Unassigned | 938_0 | Clustered | 99 | VC_938_0 | 99 | 0.9119 | 1 | 0.9119 | 0 | 0 | 0 | 0.9928 | Ref |
| 16510 | OX335422 | Unassigned | Unassigned | Unassigned | 938_0 | Clustered | 99 | VC_938_0 | 99 | 0.9119 | 1 | 0.9119 | 0 | 0 | 0 | 0.9928 | Ref |
| 16511 | OX335423 | Unassigned | Unassigned | Unassigned | 938_0 | Clustered | 99 | VC_938_0 | 99 | 0.9119 | 1 | 0.9119 | 0 | 0 | 0 | 0.9928 | Ref |
| 16512 | OX335426 | Unassigned | Unassigned | Unassigned | 823_0 | Clustered | 124 | VC_823_0 | 124 | 0.9492 | 1 | 0.9492 | 0 | 0 | 0 | 1 | Ref |
| 16513 | OX335427 | Unassigned | Unassigned | Unassigned | 823_0 | Clustered | 124 | VC_823_0 | 124 | 0.9492 | 1 | 0.9492 | 0 | 0 | 0 | 1 | Ref |
| 16514 | OX335428 | Unassigned | Unassigned | Unassigned | 111_0 | Clustered | 91 | VC_111_0 | 91 | 0.9587 | 1 | 0.9587 | 0 | 0 | 0 | 1 | Ref |
| 16515 | OX335429 | Unassigned | Unassigned | Unassigned | 938_0 | Clustered | 99 | VC_938_0 | 99 | 0.9119 | 1 | 0.9119 | 0 | 0 | 0 | 0.9928 | Ref |
| 16516 | OX335430 | Unassigned | Unassigned | Unassigned | 938_0 | Clustered | 99 | VC_938_0 | 99 | 0.9119 | 1 | 0.9119 | 0 | 0 | 0 | 0.9928 | Ref |
| 16517 | OX335431 | Unassigned | Unassigned | Unassigned | 385_0 | Clustered | 51 | VC_385_0 | 51 | 0.8754 | 1 | 0.8754 | 0 | 0 | 0 | 1 | Ref |
| 16518 | OX335432 | Unassigned | Unassigned | Unassigned | 938_0 | Clustered | 99 | VC_938_0 | 99 | 0.9119 | 1 | 0.9119 | 0 | 0 | 0 | 0.9928 | Ref |
| 16519 | OX335433 | Unassigned | Unassigned | Unassigned | 823_0 | Clustered | 124 | VC_823_0 | 124 | 0.9492 | 1 | 0.9492 | 0 | 0 | 0 | 1 | Ref |
| 16520 | OX335434 | Unassigned | Unassigned | Unassigned | 385_0 | Clustered | 51 | VC_385_0 | 51 | 0.8754 | 1 | 0.8754 | 0 | 0 | 0 | 1 | Ref |
| 16521 | OX335435 | Unassigned | Unassigned | Unassigned | 938_0 | Clustered | 99 | VC_938_0 | 99 | 0.9119 | 1 | 0.9119 | 0 | 0 | 0 | 0.9928 | Ref |
| 16522 | OX335437 | Unassigned | Unassigned | Unassigned | 385_0 | Clustered | 51 | VC_385_0 | 51 | 0.8754 | 1 | 0.8754 | 0 | 0 | 0 | 1 | Ref |

| | | | | | | | | | | | | | | | | | |
|---|---|---|---|---|---|---|---|---|---|---|---|---|---|---|---|---|---|
| 16523 | OX335438 | Unassigned | Unassigned | Unassigned | 823_0 | Clustered | 124 | VC_823_0 | 124 | 0.9492 | 1 | 0.9492 | 0 | 0 | 0 | 1 | Ref |
| 16524 | OX335439 | Unassigned | Unassigned | Unassigned | 938_0 | Clustered | 99 | VC_938_0 | 99 | 0.9119 | 1 | 0.9119 | 0 | 0 | 0 | 0.9928 | Ref |
| 16525 | OX335440 | Unassigned | Unassigned | Unassigned | 823_0 | Clustered | 124 | VC_823_0 | 124 | 0.9492 | 1 | 0.9492 | 0 | 0 | 0 | 1 | Ref |
| 16526 | OX335441 | Unassigned | Unassigned | Unassigned | 388_0 | Clustered | 4 | VC_388_0 | 4 | 0.0345 | 0.99998812 | 0.0345 | 0 | 0 | 0 | 1 | Ref |
| 16527 | OX335443 | Unassigned | Unassigned | Unassigned | 823_0 | Clustered | 124 | VC_823_0 | 124 | 0.9492 | 1 | 0.9492 | 0 | 0 | 0 | 1 | Ref |
| 16528 | PF3COMN | Unassigned | Unassigned | Unassigned | 1478_0 | Clustered | 2 | VC_1478_0 | 2 | 1 | 1 | 1 | 0 | 0 | 0 | 1 | Ref |
| 16529 | PF3COMNY | Unassigned | Unassigned | Unassigned | 1478_0 | Clustered | 2 | VC_1478_0 | 2 | 1 | 1 | 1 | 0 | 0 | 0 | 1 | Ref |
| 16530 | PFDCG | Unassigned | Unassigned | Unassigned | 61_0 | Clustered | 12 | VC_61_0 | 12 | 0.7633 | 1 | 0.7633 | 0 | 0 | 0 | 1 | Ref |
| 16531 | PX1CG | Unassigned | Unassigned | Unassigned | 303_0 | Clustered | 155 | VC_303_0 | 155 | 0.9979 | 1 | 0.9979 | 0 | 0 | 0 | 1 | Ref |
| 16532 | PZACG | Unassigned | Unassigned | Unassigned | 817_0 | Clustered | 14 | VC_817_0 | 14 | 0.6342 | 1 | 0.6342 | 0 | 0 | 0 | 1 | Ref |
| 16533 | S1-wms-combined_107073 | Unassigned | Unassigned | Unassigned | 610_0 | Clustered | 4 | VC_610_0 | 4 | 0.8779 | 0.9998533 | 0.8778 | 0 | 0 | 0 | 1 | ME |
| 16534 | S1-wms-combined_118167 | Unassigned | Unassigned | Unassigned | 825_0 | Clustered | 7 | VC_825_0 | 7 | 0.5674 | 1 | 0.5674 | 0 | 0 | 0 | 0.8571 | ME |
| 16535 | S1-wms-combined_120533 | Unassigned | Unassigned | Unassigned | 1481_0 | Clustered | 9 | VC_1481_0 | 9 | 0.9416 | 0.77917309 | 0.7337 | 0 | 0 | 0 | 1 | ME |
| 16536 | S1-wms-combined_132934 | Unassigned | Unassigned | Unassigned | 707_0 | Clustered | 6 | VC_707_0 | 6 | 0.2616 | 1 | 0.2616 | 0 | 0 | 0 | 0.8667 | ME |
| 16537 | S1-wms-combined_151708 | Unassigned | Unassigned | Unassigned | 1484_0 | Clustered | 10 | VC_1484_0 | 10 | 0.6918 | 0.99999998 | 0.6918 | 0 | 0 | 0 | 0.8889 | ME |
| 16538 | S1-wms-combined_156637 | Unassigned | Unassigned | Unassigned | 1487_0 | Clustered | 2 | VC_1487_0 | 2 | 1 | 1 | 1 | 0 | 0 | 0 | 1 | ME |
| 16539 | S1-wms-combined_159544 | Unassigned | Unassigned | Unassigned | 5_0 | Clustered | 30 | VC_5_0 | 30 | 0.9647 | 1 | 0.9647 | 0 | 0 | 0 | 1 | ME |
| 16540 | S1-wms-combined_164485 | Unassigned | Unassigned | Unassigned | 1484_0 | Clustered | 10 | VC_1484_0 | 10 | 0.6918 | 0.99999998 | 0.6918 | 0 | 0 | 0 | 0.8889 | ME |
| 16541 | S1-wms-combined_169879 | Unassigned | Unassigned | Unassigned | 825_0 | Clustered | 7 | VC_825_0 | 7 | 0.5674 | 1 | 0.5674 | 0 | 0 | 0 | 0.8571 | ME |
| 16542 | S1-wms-combined_185336 | Unassigned | Unassigned | Unassigned | 1491_0 | Clustered | 2 | VC_1491_0 | 2 | 1 | 1 | 1 | 0 | 0 | 0 | 1 | ME |
| 16543 | S1-wms-combined_19031 | Unassigned | Unassigned | Unassigned | 181_0 | Clustered | 5 | VC_181_0 | 5 | 0.1168 | 0.99999995 | 0.1168 | 0 | 0 | 0 | 1 | ME |
| 16544 | S1-wms-combined_197401 | Unassigned | Unassigned | Unassigned | 1408_0 | Clustered | 2 | VC_1408_0 | 2 | 0.1695 | 0.59870633 | 0.1015 | 0 | 0 | 0 | 1 | ME |
| 16545 | S1-wms-combined_220022 | Unassigned | Unassigned | Unassigned | 1408_1 | Clustered/Singleton | 1 | VC_1408_1 | 1 | 0 | 1 | 0 | 0 | 0 | 0 | 1 | ME |
| 16546 | S1-wms-combined_224578 | Unassigned | Unassigned | Unassigned | 418_0 | Clustered | 6 | VC_418_0 | 6 | 0.4575 | 0.9999995 | 0.4575 | 0 | 0 | 0 | 0.8 | ME |
| 16547 | S1-wms-combined_23248 | Unassigned | Unassigned | Unassigned | 621_0 | Clustered | 7 | VC_621_0 | 7 | 0.5783 | 1 | 0.5783 | 0 | 0 | 0 | 1 | ME |
| 16548 | S1-wms-combined_246923 | Unassigned | Unassigned | Unassigned | 679_0 | Clustered | 4 | VC_679_0 | 4 | 0.2754 | 0.9999774 | 0.2754 | 0 | 0 | 0 | 1 | ME |
| 16549 | S1-wms-combined_248622 | Unassigned | Unassigned | Unassigned | 1494_0 | Clustered | 2 | VC_1494_0 | 2 | 0.9538 | 0.5 | 0.4769 | 0 | 0 | 0 | 1 | ME |
| 16550 | S1-wms-combined_263562 | Unassigned | Unassigned | Unassigned | 1437_0 | Clustered | 3 | VC_1437_0 | 3 | 0.742 | 0.99188498 | 0.7359 | 0 | 0 | 0 | 1 | ME |
| 16551 | S1-wms-combined_302166 | Unassigned | Unassigned | Unassigned | 715_0 | Clustered | 3 | VC_715_0 | 3 | 0.2991 | 0.99813417 | 0.2985 | 0 | 0 | 0 | 1 | ME |
| 16552 | S1-wms-combined_304377 | Unassigned | Unassigned | Unassigned | 804_0 | Clustered | 2 | VC_804_0 | 2 | 0.0559 | 0.95690755 | 0.0535 | 0 | 0 | 0 | 1 | ME |
| 16553 | S1-wms-combined_307323 | Unassigned | Unassigned | Unassigned | 798_0 | Clustered | 2 | VC_798_0 | 2 | 0.1818 | 0.94346013 | 0.1715 | 0 | 0 | 0 | 1 | ME |
| 16554 | S1-wms-combined_313851 | Unassigned | Unassigned | Unassigned | 1496_0 | Clustered | 5 | VC_1496_0 | 5 | 0.9228 | 0.98875286 | 0.9124 | 0 | 0 | 0 | 1 | ME |
| 16555 | S1-wms-combined_324053 | Unassigned | Unassigned | Unassigned | 427_0 | Clustered | 3 | VC_427_0 | 3 | 0.0627 | 0.96830121 | 0.0608 | 0 | 0 | 0 | 1 | ME |
| 16556 | S1-wms-combined_331544 | Unassigned | Unassigned | Unassigned | 1497_0 | Clustered | 3 | VC_1497_0 | 3 | 1 | 1 | 1 | 0 | 0 | 0 | 1 | ME |
| 16557 | S1-wms-combined_331896 | Unassigned | Unassigned | Unassigned | 1481_0 | Clustered | 9 | VC_1481_0 | 9 | 0.9416 | 0.77917309 | 0.7337 | 0 | 0 | 0 | 1 | ME |
| 16558 | S1-wms-combined_332236 | Unassigned | Unassigned | Unassigned | 626_0 | Clustered | 23 | VC_626_0 | 23 | 0.66 | 1 | 0.66 | 0 | 0 | 0 | 0.8379 | ME |
| 16559 | S1-wms-combined_340558 | Unassigned | Unassigned | Unassigned | 679_0 | Clustered | 4 | VC_679_0 | 4 | 0.2754 | 0.9999774 | 0.2754 | 0 | 0 | 0 | 1 | ME |
| 16560 | S1-wms-combined_353757 | Unassigned | Unassigned | Unassigned | 650_0 | Clustered | 10 | VC_650_0 | 10 | 0.4538 | 1 | 0.4538 | 0 | 0 | 0 | 1 | ME |
| 16561 | S1-wms-combined_354899 | Unassigned | Unassigned | Unassigned | 1498_0 | Clustered | 2 | VC_1498_0 | 2 | 0.9131 | 0.5 | 0.4566 | 0 | 0 | 0 | 1 | ME |
| 16562 | S1-wms-combined_357373 | Unassigned | Unassigned | Unassigned | 1243_0 | Clustered | 52 | VC_1243_0 | 52 | 0.9126 | 0.92708311 | 0.8461 | 0 | 0 | 0 | 1 | ME |
| 16563 | S1-wms-combined_360285 | Unassigned | Unassigned | Unassigned | 716_0 | Clustered | 2 | VC_716_0 | 2 | 0.1025 | 0.95462977 | 0.0978 | 0 | 0 | 0 | 1 | ME |
| 16564 | S1-wms-combined_394574 | Unassigned | Unassigned | Unassigned | 831_0 | Clustered | 5 | VC_831_0 | 5 | 0.4369 | 0.99999952 | 0.4369 | 0 | 0 | 0 | 1 | ME |
| 16565 | S1-wms-combined_394628 | Unassigned | Unassigned | Unassigned | 1500_0 | Clustered | 2 | VC_1500_0 | 2 | 0.9749 | 0.5 | 0.4875 | 0 | 0 | 0 | 1 | ME |
| 16566 | S1-wms-combined_40115 | Unassigned | Unassigned | Unassigned | 1491_0 | Clustered | 2 | VC_1491_0 | 2 | 1 | 1 | 1 | 0 | 0 | 0 | 1 | ME |
| 16567 | S1-wms-combined_41369 | Unassigned | Unassigned | Unassigned | 1209_0 | Clustered | 3 | VC_1209_0 | 3 | 0.1553 | 0.99152019 | 0.154 | 0 | 0 | 0 | 1 | ME |
| 16568 | S1-wms-combined_52643 | Unassigned | Unassigned | Unassigned | 679_0 | Clustered | 4 | VC_679_0 | 4 | 0.2754 | 0.9999774 | 0.2754 | 0 | 0 | 0 | 1 | ME |
| 16569 | S1-wms-combined_68998 | Unassigned | Unassigned | Unassigned | 711_0 | Clustered | 3 | VC_711_0 | 3 | 0.2312 | 0.99842495 | 0.2309 | 0 | 0 | 0 | 1 | ME |
| 16570 | S1-wms-combined_72167 | Unassigned | Unassigned | Unassigned | 420_0 | Clustered | 6 | VC_420_0 | 6 | 0.2282 | 0.97462081 | 0.2224 | 0 | 0 | 0 | 1 | ME |
| 16571 | S1-wms-combined_85225 | Unassigned | Unassigned | Unassigned | 798_1 | Clustered/Singleton | 1 | VC_798_1 | 1 | 0 | 1 | 0 | 0 | 0 | 0 | 1 | ME |
| 16572 | S1-wms_100010 | Unassigned | Unassigned | Unassigned | 1492_0 | Clustered | 7 | VC_1492_0 | 7 | 0.7322 | 1 | 0.7322 | 0 | 0 | 0 | 0.8571 | ME |
| 16573 | S1-wms_102342 | Unassigned | Unassigned | Unassigned | 719_0 | Clustered/Singleton | 1 | VC_719_0 | 1 | 0 | 1 | 0 | 0 | 0 | 0 | 1 | ME |
| 16574 | S1-wms_130214 | Unassigned | Unassigned | Unassigned | 1209_0 | Clustered | 3 | VC_1209_0 | 3 | 0.1553 | 0.99152019 | 0.154 | 0 | 0 | 0 | 1 | ME |
| 16575 | S1-wms_170115 | Unassigned | Unassigned | Unassigned | 627_0 | Clustered | 3 | VC_627_0 | 3 | 0.2127 | 0.99411368 | 0.2115 | 0 | 0 | 0 | 1 | ME |
| 16576 | S1-wms_185046 | Unassigned | Unassigned | Unassigned | 1239_0 | Clustered | 6 | VC_1239_0 | 6 | 0.4699 | 0.99999972 | 0.4699 | 0 | 0 | 0 | 1 | ME |
| 16577 | S1-wms_212940 | Unassigned | Unassigned | Unassigned | 804_2 | Clustered | 6 | VC_804_2 | 6 | 0.1161 | 1 | 0.1161 | 0 | 0 | 0 | 0.8667 | ME |
| 16578 | S1-wms_225995 | Unassigned | Unassigned | Unassigned | 1492_0 | Clustered | 7 | VC_1492_0 | 7 | 0.7322 | 1 | 0.7322 | 0 | 0 | 0 | 0.8571 | ME |
| 16579 | S1-wms_244836 | Unassigned | Unassigned | Unassigned | 1473_0 | Clustered | 3 | VC_1473_0 | 3 | 0.7237 | 0.99613166 | 0.7209 | 0 | 0 | 0 | 0.6667 | ME |
| 16580 | S1-wms_296850 | Unassigned | Unassigned | Unassigned | 710_0 | Clustered | 10 | VC_710_0 | 10 | 0.2446 | 1 | 0.2446 | 0 | 0 | 0 | 0.8222 | ME |
| 16581 | S1-wms_313991 | Unassigned | Unassigned | Unassigned | 1496_0 | Clustered | 5 | VC_1496_0 | 5 | 0.9228 | 0.98875286 | 0.9124 | 0 | 0 | 0 | 1 | ME |
| 16582 | S1-wms_4277 | Unassigned | Unassigned | Unassigned | 1235_0 | Clustered | 12 | VC_1235_0 | 12 | 0.7997 | 1 | 0.7997 | 0 | 0 | 0 | 1 | ME |
| 16583 | S11-wms-combined_119215 | Unassigned | Unassigned | Unassigned | 650_0 | Clustered | 10 | VC_650_0 | 10 | 0.4538 | 1 | 0.4538 | 0 | 0 | 0 | 1 | ME |
| 16584 | S11-wms-combined_125176 | Unassigned | Unassigned | Unassigned | 1456_0 | Clustered | 4 | VC_1456_0 | 4 | 0.6121 | 0.99987744 | 0.612 | 0 | 0 | 0 | 1 | ME |
| 16585 | S11-wms-combined_140009 | Unassigned | Unassigned | Unassigned | 621_0 | Clustered | 7 | VC_621_0 | 7 | 0.5783 | 1 | 0.5783 | 0 | 0 | 0 | 1 | ME |
| 16586 | S11-wms-combined_154845 | Unassigned | Unassigned | Unassigned | 5_0 | Clustered | 30 | VC_5_0 | 30 | 0.9647 | 1 | 0.9647 | 0 | 0 | 0 | 1 | ME |
| 16587 | S11-wms-combined_17029 | Unassigned | Unassigned | Unassigned | 1502_0 | Clustered | 2 | VC_1502_0 | 2 | 0.3993 | 0.91813282 | 0.3666 | 0 | 0 | 0 | 1 | ME |
| 16588 | S11-wms-combined_172889 | Unassigned | Unassigned | Unassigned | 679_0 | Clustered | 4 | VC_679_0 | 4 | 0.2754 | 0.9999774 | 0.2754 | 0 | 0 | 0 | 1 | ME |
| 16589 | S11-wms-combined_193148 | Unassigned | Unassigned | Unassigned | 616_0 | Clustered | 2 | VC_616_0 | 2 | 0.7322 | 0.81445332 | 0.5964 | 0 | 0 | 0 | 1 | ME |
| 16590 | S11-wms-combined_194205 | Unassigned | Unassigned | Unassigned | 626_0 | Clustered | 23 | VC_626_0 | 23 | 0.66 | 1 | 0.66 | 0 | 0 | 0 | 0.8379 | ME |
| 16591 | S11-wms-combined_195425 | Unassigned | Unassigned | Unassigned | 677_0 | Clustered | 9 | VC_677_0 | 9 | 0.3787 | 1 | 0.3787 | 0 | 0 | 0 | 0.8333 | ME |
| 16592 | S11-wms-combined_198512 | Unassigned | Unassigned | Unassigned | 715_0 | Clustered | 3 | VC_715_0 | 3 | 0.2991 | 0.99813417 | 0.2985 | 0 | 0 | 0 | 1 | ME |
| 16593 | S11-wms-combined_200084 | Unassigned | Unassigned | Unassigned | 825_0 | Clustered | 7 | VC_825_0 | 7 | 0.5674 | 1 | 0.5674 | 0 | 0 | 0 | 0.8571 | ME |
| 16594 | S11-wms-combined_204522 | Unassigned | Unassigned | Unassigned | 616_0 | Clustered | 2 | VC_616_0 | 2 | 0.7322 | 0.81445332 | 0.5964 | 0 | 0 | 0 | 1 | ME |
| 16595 | S11-wms-combined_217790 | Unassigned | Unassigned | Unassigned | 1503_0 | Clustered | 3 | VC_1503_0 | 3 | 0.9111 | 0.99054823 | 0.9025 | 0 | 0 | 0 | 1 | ME |
| 16596 | S11-wms-combined_221310 | Unassigned | Unassigned | Unassigned | 1504_0 | Clustered | 2 | VC_1504_0 | 2 | 0.9646 | 0.5 | 0.4823 | 0 | 0 | 0 | 1 | ME |
| 16597 | S11-wms-combined_264050 | Unassigned | Unassigned | Unassigned | 707_3 | Clustered/Singleton | 1 | VC_707_3 | 1 | 0 | 1 | 0 | 0 | 0 | 0 | 1 | ME |
| 16598 | S11-wms-combined_267993 | Unassigned | Unassigned | Unassigned | 825_1 | Clustered/Singleton | 1 | VC_825_1 | 1 | 0 | 1 | 0 | 0 | 0 | 0 | 1 | ME |
| 16599 | S11-wms-combined_268967 | Unassigned | Unassigned | Unassigned | 181_0 | Clustered | 5 | VC_181_0 | 5 | 0.1168 | 0.99999995 | 0.1168 | 0 | 0 | 0 | 1 | ME |
| 16600 | S11-wms-combined_283220 | Unassigned | Unassigned | Unassigned | 1235_0 | Clustered | 12 | VC_1235_0 | 12 | 0.7997 | 1 | 0.7997 | 0 | 0 | 0 | 1 | ME |
| 16601 | S11-wms-combined_299954 | Unassigned | Unassigned | Unassigned | 650_0 | Clustered | 10 | VC_650_0 | 10 | 0.4538 | 1 | 0.4538 | 0 | 0 | 0 | 1 | ME |
| 16602 | S11-wms-combined_304239 | Unassigned | Unassigned | Unassigned | 707_0 | Clustered | 6 | VC_707_0 | 6 | 0.2616 | 1 | 0.2616 | 0 | 0 | 0 | 0.8667 | ME |
| 16603 | S11-wms-combined_3507 | Unassigned | Unassigned | Unassigned | 1209_2 | Clustered/Singleton | 1 | VC_1209_2 | 1 | 0 | 1 | 0 | 0 | 0 | 0 | 1 | ME |
| 16604 | S11-wms-combined_41367 | Unassigned | Unassigned | Unassigned | 1505_0 | Clustered | 4 | VC_1505_0 | 4 | 0.9495 | 0.99879268 | 0.9484 | 0 | 0 | 0 | 1 | ME |
| 16605 | S11-wms-combined_46816 | Unassigned | Unassigned | Unassigned | 893_0 | Clustered | 2 | VC_893_0 | 2 | 0.1729 | 0.93846104 | 0.1623 | 0 | 0 | 0 | 1 | ME |
| 16606 | S11-wms-combined_48246 | Unassigned | Unassigned | Unassigned | 917_0 | Clustered | 2 | VC_917_0 | 2 | 0.3004 | 0.94854123 | 0.2849 | 0 | 0 | 0 | 1 | ME |
| 16607 | S11-wms-combined_49632 | Unassigned | Unassigned | Unassigned | 650_0 | Clustered | 10 | VC_650_0 | 10 | 0.4538 | 1 | 0.4538 | 0 | 0 | 0 | 1 | ME |
| 16608 | S11-wms-combined_58538 | Unassigned | Unassigned | Unassigned | 271_0 | Clustered | 5 | VC_271_0 | 5 | 0.3916 | 0.99999838 | 0.3916 | 0 | 0 | 0 | 0.8 | ME |
| 16609 | S11-wms-combined_73204 | Unassigned | Unassigned | Unassigned | 1397_0 | Clustered | 2 | VC_1397_0 | 2 | 0.6905 | 0.93463371 | 0.6454 | 0 | 0 | 0 | 1 | ME |
| 16610 | S11-wms-combined_74983 | Unassigned | Unassigned | Unassigned | 710_0 | Clustered | 10 | VC_710_0 | 10 | 0.2446 | 1 | 0.2446 | 0 | 0 | 0 | 0.8222 | ME |
| 16611 | S11-wms-combined_83571 | Unassigned | Unassigned | Unassigned | 676_1 | Clustered | 4 | VC_676_1 | 4 | 0.3759 | 0.9999693 | 0.3759 | 0 | 0 | 0 | 0.6667 | ME |
| 16612 | S11-wms-combined_84326 | Unassigned | Unassigned | Unassigned | 716_0 | Clustered | 2 | VC_716_0 | 2 | 0.1025 | 0.95462977 | 0.0978 | 0 | 0 | 0 | 1 | ME |
| 16613 | S11-wms-combined_92114 | Unassigned | Unassigned | Unassigned | 1014_0 | Clustered | 3 | VC_1014_0 | 3 | 0.2026 | 0.85591218 | 0.1734 | 0 | 0 | 0 | 0.6667 | ME |
| 16614 | S11-wms_14390 | Unassigned | Unassigned | Unassigned | 271_0 | Clustered | 5 | VC_271_0 | 5 | 0.3916 | 0.99999838 | 0.3916 | 0 | 0 | 0 | 0.8 | ME |
| 16615 | S11-wms_303398 | Unassigned | Unassigned | Unassigned | 1239_0 | Clustered | 6 | VC_1239_0 | 6 | 0.4699 | 0.99999972 | 0.4699 | 0 | 0 | 0 | 1 | ME |
| 16616 | S11-wms_3116 | Unassigned | Unassigned | Unassigned | 1235_0 | Clustered | 12 | VC_1235_0 | 12 | 0.7997 | 1 | 0.7997 | 0 | 0 | 0 | 1 | ME |
| 16617 | S11-wms_320555 | Unassigned | Unassigned | Unassigned | 918_0 | Clustered | 2 | VC_918_0 | 2 | 0.1414 | 0.92876662 | 0.1314 | 0 | 0 | 0 | 1 | ME |
| 16618 | S11-wms_330479 | Unassigned | Unassigned | Unassigned | 1010_0 | Clustered | 2 | VC_1010_0 | 2 | 0.2036 | 0.77337265 | 0.1574 | 0 | 0 | 0 | 1 | ME |
| 16619 | S11-wms_55610 | Unassigned | Unassigned | Unassigned | 606_0 | Clustered | 4 | VC_606_0 | 4 | 0.4507 | 0.99995146 | 0.4507 | 0 | 0 | 0 | 1 | ME |
| 16620 | S12-wms-combined_107942 | Unassigned | Unassigned | Unassigned | 804_2 | Clustered | 6 | VC_804_2 | 6 | 0.1161 | 1 | 0.1161 | 0 | 0 | 0 | 0.8667 | SHHC |
| 16621 | S12-wms-combined_111524 | Unassigned | Unassigned | Unassigned | 1503_0 | Clustered | 3 | VC_1503_0 | 3 | 0.9111 | 0.99054823 | 0.9025 | 0 | 0 | 0 | 1 | SHHC |
| 16622 | S12-wms-combined_120133 | Unassigned | Unassigned | Unassigned | 338_0 | Clustered | 5 | VC_338_0 | 5 | 0.3309 | 0.99999987 | 0.3309 | 0 | 0 | 0 | 1 | SHHC |
| 16623 | S12-wms-combined_129554 | Unassigned | Unassigned | Unassigned | 707_1 | Clustered | 9 | VC_707_1 | 9 | 0.2702 | 1 | 0.2702 | 0 | 0 | 0 | 0.8889 | SHHC |
| 16624 | S12-wms-combined_129799 | Unassigned | Unassigned | Unassigned | 707_2 | Clustered/Singleton | 1 | VC_707_2 | 1 | 0 | 1 | 0 | 0 | 0 | 0 | 1 | SHHC |
| 16625 | S12-wms-combined_147628 | Unassigned | Unassigned | Unassigned | 1238_0 | Clustered | 2 | VC_1238_0 | 2 | 0.1055 | 0.91454824 | 0.0965 | 0 | 0 | 0 | 1 | SHHC |
| 16626 | S12-wms-combined_156234 | Unassigned | Unassigned | Unassigned | 710_0 | Clustered | 10 | VC_710_0 | 10 | 0.2446 | 1 | 0.2446 | 0 | 0 | 0 | 0.8222 | SHHC |
| 16627 | S12-wms-combined_157167 | Unassigned | Unassigned | Unassigned | 1489_0 | Clustered/Singleton | 1 | VC_1489_0 | 1 | 0 | 1 | 0 | 0 | 0 | 0 | 1 | SHHC |
| 16628 | S12-wms-combined_168259 | Unassigned | Unassigned | Unassigned | 736_0 | Clustered | 3 | VC_736_0 | 3 | 0.5056 | 0.95195384 | 0.4813 | 0 | 0 | 0 | 1 | SHHC |
| 16629 | S12-wms-combined_17617 | Unassigned | Unassigned | Unassigned | 707_0 | Clustered | 6 | VC_707_0 | 6 | 0.2616 | 1 | 0.2616 | 0 | 0 | 0 | 0.8667 | SHHC |
| 16630 | S12-wms-combined_18287 | Unassigned | Unassigned | Unassigned | 707_0 | Clustered | 6 | VC_707_0 | 6 | 0.2616 | 1 | 0.2616 | 0 | 0 | 0 | 0.8667 | SHHC |
| 16631 | S12-wms-combined_191570 | Unassigned | Unassigned | Unassigned | 732_0 | Clustered | 2 | VC_732_0 | 2 | 0.0524 | 0.90742814 | 0.0476 | 0 | 0 | 0 | 1 | SHHC |
| 16632 | S12-wms-combined_201125 | Unassigned | Unassigned | Unassigned | 918_2 | Clustered/Singleton | 1 | VC_918_2 | 1 | 0 | 1 | 0 | 0 | 0 | 0 | 1 | SHHC |
| 16633 | S12-wms-combined_210942 | Unassigned | Unassigned | Unassigned | 1487_0 | Clustered | 2 | VC_1487_0 | 2 | 1 | 1 | 1 | 0 | 0 | 0 | 1 | SHHC |
| 16634 | S12-wms-combined_23517 | Unassigned | Unassigned | Unassigned | 1209_1 | Clustered/Singleton | 1 | VC_1209_1 | 1 | 0 | 1 | 0 | 0 | 0 | 0 | 1 | SHHC |
| 16635 | S12-wms-combined_2450 | Unassigned | Unassigned | Unassigned | 676_0 | Clustered | 2 | VC_676_0 | 2 | 0.1026 | 0.95156715 | 0.0977 | 0 | 0 | 0 | 1 | SHHC |
| 16636 | S12-wms-combined_245755 | Unassigned | Unassigned | Unassigned | 876_0 | Clustered | 3 | VC_876_0 | 3 | 0.1628 | 0.99746631 | 0.1624 | 0 | 0 | 0 | 0.6667 | SHHC |
| 16637 | S12-wms-combined_250551 | Unassigned | Unassigned | Unassigned | 677_0 | Clustered | 9 | VC_677_0 | 9 | 0.3787 | 1 | 0.3787 | 0 | 0 | 0 | 0.8333 | SHHC |
| 16638 | S12-wms-combined_256299 | Unassigned | Unassigned | Unassigned | 1505_0 | Clustered | 4 | VC_1505_0 | 4 | 0.9495 | 0.99879268 | 0.9484 | 0 | 0 | 0 | 1 | SHHC |
| 16639 | S12-wms-combined_259783 | Unassigned | Unassigned | Unassigned | 717_0 | Clustered | 7 | VC_717_0 | 7 | 0.5813 | 1 | 0.5813 | 0 | 0 | 0 | 0.9048 | SHHC |
| 16640 | S12-wms-combined_260036 | Unassigned | Unassigned | Unassigned | 1439_0 | Clustered | 3 | VC_1439_0 | 3 | 0.4461 | 0.98736442 | 0.4405 | 0 | 0 | 0 | 1 | SHHC |
| 16641 | S12-wms-combined_263377 | Unassigned | Unassigned | Unassigned | 212_0 | Clustered | 2 | VC_212_0 | 2 | 0.3005 | 0.9466884 | 0.2845 | 0 | 0 | 0 | 1 | SHHC |
| 16642 | S12-wms-combined_264425 | Unassigned | Unassigned | Unassigned | 1492_0 | Clustered | 7 | VC_1492_0 | 7 | 0.7322 | 1 | 0.7322 | 0 | 0 | 0 | 0.8571 | SHHC |
| 16643 | S12-wms-combined_264845 | Unassigned | Unassigned | Unassigned | 1507_0 | Clustered | 2 | VC_1507_0 | 2 | 0.543 | 0.72985431 | 0.3963 | 0 | 0 | 0 | 1 | SHHC |
| 16644 | S12-wms-combined_282594 | Unassigned | Unassigned | Unassigned | 736_0 | Clustered | 3 | VC_736_0 | 3 | 0.5056 | 0.95195384 | 0.4813 | 0 | 0 | 0 | 1 | SHHC |
| 16645 | S12-wms-combined_299293 | Unassigned | Unassigned | Unassigned | 1492_0 | Clustered | 7 | VC_1492_0 | 7 | 0.7322 | 1 | 0.7322 | 0 | 0 | 0 | 0.8571 | SHHC |
| 16646 | S12-wms-combined_299872 | Unassigned | Unassigned | Unassigned | 1235_0 | Clustered | 12 | VC_1235_0 | 12 | 0.7997 | 1 | 0.7997 | 0 | 0 | 0 | 1 | SHHC |
| 16647 | S12-wms-combined_320727 | Unassigned | Unassigned | Unassigned | 1397_0 | Clustered | 2 | VC_1397_0 | 2 | 0.6905 | 0.93463371 | 0.6454 | 0 | 0 | 0 | 1 | SHHC |
| 16648 | S12-wms-combined_321174 | Unassigned | Unassigned | Unassigned | 915_0 | Clustered | 3 | VC_915_0 | 3 | 0.2219 | 0.99819349 | 0.2215 | 0 | 0 | 0 | 1 | SHHC |
| 16649 | S12-wms-combined_323943 | Unassigned | Unassigned | Unassigned | 711_1 | Clustered/Singleton | 1 | VC_711_1 | 1 | 0 | 1 | 0 | 0 | 0 | 0 | 1 | SHHC |
| 16650 | S12-wms-combined_329242 | Unassigned | Unassigned | Unassigned | 1484_0 | Clustered | 10 | VC_1484_0 | 10 | 0.6918 | 0.99999998 | 0.6918 | 0 | 0 | 0 | 0.8889 | SHHC |
| 16651 | S12-wms-combined_334806 | Unassigned | Unassigned | Unassigned | 1239_0 | Clustered | 6 | VC_1239_0 | 6 | 0.4699 | 0.99999972 | 0.4699 | 0 | 0 | 0 | 1 | SHHC |
| 16652 | S12-wms-combined_345948 | Unassigned | Unassigned | Unassigned | 621_0 | Clustered | 7 | VC_621_0 | 7 | 0.5783 | 1 | 0.5783 | 0 | 0 | 0 | 1 | SHHC |
| 16653 | S12-wms-combined_351847 | Unassigned | Unassigned | Unassigned | 1436_0 | Clustered | 4 | VC_1436_0 | 4 | 0.571 | 0.96369953 | 0.5502 | 0 | 0 | 0 | 1 | SHHC |
| 16654 | S12-wms-combined_358010 | Unassigned | Unassigned | Unassigned | 715_0 | Clustered | 3 | VC_715_0 | 3 | 0.2991 | 0.99813417 | 0.2985 | 0 | 0 | 0 | 1 | SHHC |
| 16655 | S12-wms-combined_358797 | Unassigned | Unassigned | Unassigned | 678_0 | Clustered/Singleton | 1 | VC_678_0 | 1 | 0 | 1 | 0 | 0 | 0 | 0 | 1 | SHHC |
| 16656 | S12-wms-combined_368462 | Unassigned | Unassigned | Unassigned | 626_0 | Clustered | 23 | VC_626_0 | 23 | 0.66 | 1 | 0.66 | 0 | 0 | 0 | 0.8379 | SHHC |
| 16657 | S12-wms-combined_375343 | Unassigned | Unassigned | Unassigned | 1502_0 | Clustered | 2 | VC_1502_0 | 2 | 0.3993 | 0.91813282 | 0.3666 | 0 | 0 | 0 | 1 | SHHC |
| 16658 | S12-wms-combined_376326 | Unassigned | Unassigned | Unassigned | 1076_0 | Clustered | 2 | VC_1076_0 | 2 | 0.3461 | 0.6381632 | 0.2209 | 0 | 0 | 0 | 1 | SHHC |
| 16659 | S12-wms-combined_393703 | Unassigned | Unassigned | Unassigned | 1481_0 | Clustered | 9 | VC_1481_0 | 9 | 0.9416 | 0.77917309 | 0.7337 | 0 | 0 | 0 | 1 | SHHC |
| 16660 | S12-wms-combined_401379 | Unassigned | Unassigned | Unassigned | 1500_0 | Clustered | 2 | VC_1500_0 | 2 | 0.9749 | 0.5 | 0.4875 | 0 | 0 | 0 | 1 | SHHC |
| 16661 | S12-wms-combined_401383 | Unassigned | Unassigned | Unassigned | 1510_0 | Clustered | 2 | VC_1510_0 | 2 | 1 | 1 | 1 | 0 | 0 | 0 | 1 | SHHC |
| 16662 | S12-wms-combined_41160 | Unassigned | Unassigned | Unassigned | 1238_0 | Clustered | 2 | VC_1238_0 | 2 | 0.1055 | 0.91454824 | 0.0965 | 0 | 0 | 0 | 1 | SHHC |
| 16663 | S12-wms-combined_51076 | Unassigned | Unassigned | Unassigned | 1479_0 | Clustered | 2 | VC_1479_0 | 2 | 0.4887 | 0.93846104 | 0.4586 | 0 | 0 | 0 | 1 | SHHC |
| 16664 | S12-wms-combined_71235 | Unassigned | Unassigned | Unassigned | 621_0 | Clustered | 7 | VC_621_0 | 7 | 0.5783 | 1 | 0.5783 | 0 | 0 | 0 | 1 | SHHC |
| 16665 | S12-wms-combined_7174 | Unassigned | Unassigned | Unassigned | 626_0 | Clustered | 23 | VC_626_0 | 23 | 0.66 | 1 | 0.66 | 0 | 0 | 0 | 0.8379 | SHHC |
| 16666 | S12-wms-combined_91231 | Unassigned | Unassigned | Unassigned | 1439_0 | Clustered | 3 | VC_1439_0 | 3 | 0.4461 | 0.98736442 | 0.4405 | 0 | 0 | 0 | 1 | SHHC |
| 16667 | S12-wms-combined_98046 | Unassigned | Unassigned | Unassigned | 626_2 | Clustered/Singleton | 1 | VC_626_2 | 1 | 0 | 1 | 0 | 0 | 0 | 0 | 1 | SHHC |
| 16668 | S12-wms-combined_99704 | Unassigned | Unassigned | Unassigned | 626_0 | Clustered | 23 | VC_626_0 | 23 | 0.66 | 1 | 0.66 | 0 | 0 | 0 | 0.8379 | SHHC |
| 16669 | S12-wms_12658 | Unassigned | Unassigned | Unassigned | 918_0 | Clustered | 2 | VC_918_0 | 2 | 0.1414 | 0.92876662 | 0.1314 | 0 | 0 | 0 | 1 | SHHC |
| 16670 | S12-wms_15700 | Unassigned | Unassigned | Unassigned | 1511_0 | Clustered | 2 | VC_1511_0 | 2 | 1 | 1 | 1 | 0 | 0 | 0 | 1 | SHHC |
| 16671 | S12-wms_179388 | Unassigned | Unassigned | Unassigned | 1492_0 | Clustered | 7 | VC_1492_0 | 7 | 0.7322 | 1 | 0.7322 | 0 | 0 | 0 | 0.8571 | SHHC |
| 16672 | S12-wms_193565 | Unassigned | Unassigned | Unassigned | 1235_0 | Clustered | 12 | VC_1235_0 | 12 | 0.7997 | 1 | 0.7997 | 0 | 0 | 0 | 1 | SHHC |
| 16673 | S12-wms_373775 | Unassigned | Unassigned | Unassigned | 1507_0 | Clustered | 2 | VC_1507_0 | 2 | 0.543 | 0.72985431 | 0.3963 | 0 | 0 | 0 | 1 | SHHC |
| 16674 | S12-wms_383851 | Unassigned | Unassigned | Unassigned | 1436_0 | Clustered | 4 | VC_1436_0 | 4 | 0.571 | 0.96369953 | 0.5502 | 0 | 0 | 0 | 1 | SHHC |
| 16675 | S12-wms_386692 | Unassigned | Unassigned | Unassigned | 1456_0 | Clustered | 4 | VC_1456_0 | 4 | 0.6121 | 0.99987744 | 0.612 | 0 | 0 | 0 | 1 | SHHC |
| 16676 | S12-wms_401366 | Unassigned | Unassigned | Unassigned | 1492_0 | Clustered | 7 | VC_1492_0 | 7 | 0.7322 | 1 | 0.7322 | 0 | 0 | 0 | 0.8571 | SHHC |
| 16677 | S12-wms_401372 | Unassigned | Unassigned | Unassigned | 1420_0 | Clustered | 5 | VC_1420_0 | 5 | 1 | 1 | 1 | 0 | 0 | 0 | 1 | SHHC |
| 16678 | S12-wms_44817 | Unassigned | Unassigned | Unassigned | 1492_0 | Clustered | 7 | VC_1492_0 | 7 | 0.7322 | 1 | 0.7322 | 0 | 0 | 0 | 0.8571 | SHHC |
| 16679 | S13CG | Unassigned | Unassigned | Unassigned | 303_0 | Clustered | 155 | VC_303_0 | 155 | 0.9979 | 1 | 0.9979 | 0 | 0 | 0 | 1 | Ref |
| 16680 | S17-wms-combined_126600 | Unassigned | Unassigned | Unassigned | 1481_0 | Clustered | 9 | VC_1481_0 | 9 | 0.9416 | 0.77917309 | 0.7337 | 0 | 0 | 0 | 1 | ME |
| 16681 | S17-wms-combined_126774 | Unassigned | Unassigned | Unassigned | 1484_0 | Clustered | 10 | VC_1484_0 | 10 | 0.6918 | 0.99999998 | 0.6918 | 0 | 0 | 0 | 0.8889 | ME |
| 16682 | S17-wms-combined_127688 | Unassigned | Unassigned | Unassigned | 707_1 | Clustered | 9 | VC_707_1 | 9 | 0.2702 | 1 | 0.2702 | 0 | 0 | 0 | 0.8889 | ME |
| 16683 | S17-wms-combined_130215 | Unassigned | Unassigned | Unassigned | 338_0 | Clustered | 5 | VC_338_0 | 5 | 0.3309 | 0.99999987 | 0.3309 | 0 | 0 | 0 | 1 | ME |
| 16684 | S17-wms-combined_136622 | Unassigned | Unassigned | Unassigned | 650_0 | Clustered | 10 | VC_650_0 | 10 | 0.4538 | 1 | 0.4538 | 0 | 0 | 0 | 1 | ME |

| | | | | | | | | | | | | | | | | | |
|---|---|---|---|---|---|---|---|---|---|---|---|---|---|---|---|---|---|
| 16685 | S17-wms-combined_146850 | Unassigned | Unassigned | Unassigned | 1484_0 | Clustered | 10 | VC_1484_0 | 10 | 0.6918 | 0.99999998 | 0.6918 | 0 | 0 | 0 | 0.8889 | ME |
| 16686 | S17-wms-combined_157468 | Unassigned | Unassigned | Unassigned | 1484_0 | Clustered | 10 | VC_1484_0 | 10 | 0.6918 | 0.99999998 | 0.6918 | 0 | 0 | 0 | 0.8889 | ME |
| 16687 | S17-wms-combined_158354 | Unassigned | Unassigned | Unassigned | 1505_0 | Clustered | 4 | VC_1505_0 | 4 | 0.9495 | 0.99879268 | 0.9484 | 0 | 0 | 0 | 1 | ME |
| 16688 | S17-wms-combined_162786 | Unassigned | Unassigned | Unassigned | 1408_0 | Clustered | 2 | VC_1408_0 | 2 | 0.1695 | 0.59870633 | 0.1015 | 0 | 0 | 0 | 1 | ME |
| 16689 | S17-wms-combined_16309 | Unassigned | Unassigned | Unassigned | 1435_0 | Clustered | 9 | VC_1435_0 | 9 | 0.6931 | 0.99999995 | 0.6931 | 0 | 0 | 0 | 1 | ME |
| 16690 | S17-wms-combined_175924 | Unassigned | Unassigned | Unassigned | 1235_0 | Clustered | 12 | VC_1235_0 | 12 | 0.7997 | 1 | 0.7997 | 0 | 0 | 0 | 1 | ME |
| 16691 | S17-wms-combined_179771 | Unassigned | Unassigned | Unassigned | 1509_0 | Clustered | 2 | VC_1509_0 | 2 | 0.5962 | 0.91237788 | 0.5439 | 0 | 0 | 0 | 1 | ME |
| 16692 | S17-wms-combined_186314 | Unassigned | Unassigned | Unassigned | 798_2 | Clustered/Singleton | 1 | VC_798_2 | 1 | 0 | 1 | 0 | 0 | 0 | 0 | 1 | ME |
| 16693 | S17-wms-combined_193958 | Unassigned | Unassigned | Unassigned | 1235_0 | Clustered | 12 | VC_1235_0 | 12 | 0.7997 | 1 | 0.7997 | 0 | 0 | 0 | 1 | ME |
| 16694 | S17-wms-combined_199440 | Unassigned | Unassigned | Unassigned | 707_0 | Clustered | 6 | VC_707_0 | 6 | 0.2616 | 1 | 0.2616 | 0 | 0 | 0 | 0.8667 | ME |
| 16695 | S17-wms-combined_199746 | Unassigned | Unassigned | Unassigned | 1014_0 | Clustered | 3 | VC_1014_0 | 3 | 0.2026 | 0.85591218 | 0.1734 | 0 | 0 | 0 | 0.6667 | ME |
| 16696 | S17-wms-combined_203701 | Unassigned | Unassigned | Unassigned | 1508_0 | Clustered | 3 | VC_1508_0 | 3 | 0.8977 | 0.9595722 | 0.8614 | 0 | 0 | 0 | 0.6667 | ME |
| 16697 | S17-wms-combined_2234 | Unassigned | Unassigned | Unassigned | 711_2 | Clustered/Singleton | 1 | VC_711_2 | 1 | 0 | 1 | 0 | 0 | 0 | 0 | 1 | ME |
| 16698 | S17-wms-combined_223748 | Unassigned | Unassigned | Unassigned | 1512_0 | Clustered | 2 | VC_1512_0 | 2 | 1 | 1 | 1 | 0 | 0 | 0 | 1 | ME |
| 16699 | S17-wms-combined_225356 | Unassigned | Unassigned | Unassigned | 1513_0 | Clustered | 2 | VC_1513_0 | 2 | 0.9263 | 0.5 | 0.4632 | 0 | 0 | 0 | 1 | ME |
| 16700 | S17-wms-combined_233761 | Unassigned | Unassigned | Unassigned | 1495_0 | Clustered | 3 | VC_1495_0 | 3 | 0.7112 | 0.99363432 | 0.7067 | 0 | 0 | 0 | 1 | ME |
| 16701 | S17-wms-combined_247494 | Unassigned | Unassigned | Unassigned | 650_0 | Clustered | 10 | VC_650_0 | 10 | 0.4538 | 1 | 0.4538 | 0 | 0 | 0 | 1 | ME |
| 16702 | S17-wms-combined_248321 | Unassigned | Unassigned | Unassigned | 717_1 | Clustered/Singleton | 1 | VC_717_1 | 1 | 0 | 1 | 0 | 0 | 0 | 0 | 1 | ME |
| 16703 | S17-wms-combined_259811 | Unassigned | Unassigned | Unassigned | 804_1 | Clustered | 2 | VC_804_1 | 2 | 0.0413 | 0.95352218 | 0.0394 | 0 | 0 | 0 | 1 | ME |
| 16704 | S17-wms-combined_260343 | Unassigned | Unassigned | Unassigned | 710_0 | Clustered | 10 | VC_710_0 | 10 | 0.2446 | 1 | 0.2446 | 0 | 0 | 0 | 0.8222 | ME |
| 16705 | S17-wms-combined_30308 | Unassigned | Unassigned | Unassigned | 1482_0 | Clustered | 2 | VC_1482_0 | 2 | 0.3552 | 0.85557782 | 0.3039 | 0 | 0 | 0 | 1 | ME |
| 16706 | S17-wms-combined_47860 | Unassigned | Unassigned | Unassigned | 804_1 | Clustered | 2 | VC_804_1 | 2 | 0.0413 | 0.95352218 | 0.0394 | 0 | 0 | 0 | 1 | ME |
| 16707 | S17-wms-combined_70983 | Unassigned | Unassigned | Unassigned | 1494_0 | Clustered | 2 | VC_1494_0 | 2 | 0.9538 | 0.5 | 0.4769 | 0 | 0 | 0 | 1 | ME |
| 16708 | S17-wms-combined_79709 | Unassigned | Unassigned | Unassigned | 650_0 | Clustered | 10 | VC_650_0 | 10 | 0.4538 | 1 | 0.4538 | 0 | 0 | 0 | 1 | ME |
| 16709 | S17-wms-combined_90378 | Unassigned | Unassigned | Unassigned | 621_0 | Clustered | 7 | VC_621_0 | 7 | 0.5783 | 1 | 0.5783 | 0 | 0 | 0 | 1 | ME |
| 16710 | S17-wms-combined_95360 | Unassigned | Unassigned | Unassigned | 732_1 | Clustered/Singleton | 1 | VC_732_1 | 1 | 0 | 1 | 0 | 0 | 0 | 0 | 1 | ME |
| 16711 | S17-wms-combined_9864 | Unassigned | Unassigned | Unassigned | 626_0 | Clustered | 23 | VC_626_0 | 23 | 0.66 | 1 | 0.66 | 0 | 0 | 0 | 0.8379 | ME |
| 16712 | S17-wms_124308 | Unassigned | Unassigned | Unassigned | 1435_0 | Clustered | 9 | VC_1435_0 | 9 | 0.6931 | 0.99999995 | 0.6931 | 0 | 0 | 0 | 1 | ME |
| 16713 | S17-wms_254872 | Unassigned | Unassigned | Unassigned | 1276_0 | Clustered | 7 | VC_1276_0 | 7 | 0.8695 | 0.99982476 | 0.8693 | 0 | 0 | 0 | 1 | ME |
| 16714 | S17-wms_266511 | Unassigned | Unassigned | Unassigned | 540_0 | Clustered | 15 | VC_540_0 | 15 | 0.2866 | 1 | 0.2866 | 0 | 0 | 0 | 1 | ME |
| 16715 | S2-wms-combined_101826 | Unassigned | Unassigned | Unassigned | 734_0 | Clustered | 6 | VC_734_0 | 6 | 0.1914 | 0.99996888 | 0.1914 | 0 | 0 | 0 | 1 | SHHC |
| 16716 | S2-wms-combined_113619 | Unassigned | Unassigned | Unassigned | 832_0 | Clustered | 2 | VC_832_0 | 2 | 0.2344 | 0.94526139 | 0.2215 | 0 | 0 | 0 | 1 | SHHC |
| 16717 | S2-wms-combined_116221 | Unassigned | Unassigned | Unassigned | 876_1 | Clustered/Singleton | 1 | VC_876_1 | 1 | 0 | 1 | 0 | 0 | 0 | 0 | 1 | SHHC |
| 16718 | S2-wms-combined_126134 | Unassigned | Unassigned | Unassigned | 650_0 | Clustered | 10 | VC_650_0 | 10 | 0.4538 | 1 | 0.4538 | 0 | 0 | 0 | 1 | SHHC |
| 16719 | S2-wms-combined_13246 | Unassigned | Unassigned | Unassigned | 710_4 | Clustered/Singleton | 1 | VC_710_4 | 1 | 0 | 1 | 0 | 0 | 0 | 0 | 1 | SHHC |
| 16720 | S2-wms-combined_132966 | Unassigned | Unassigned | Unassigned | 1422_0 | Clustered | 2 | VC_1422_0 | 2 | 1 | 1 | 1 | 0 | 0 | 0 | 1 | SHHC |
| 16721 | S2-wms-combined_135139 | Unassigned | Unassigned | Unassigned | 738_0 | Clustered | 2 | VC_738_0 | 2 | 0.6709 | 0.85557782 | 0.574 | 0 | 0 | 0 | 1 | SHHC |
| 16722 | S2-wms-combined_135154 | Unassigned | Unassigned | Unassigned | 947_0 | Clustered | 3 | VC_947_0 | 3 | 0.0374 | 0.84591675 | 0.0316 | 0 | 0 | 0 | 1 | SHHC |
| 16723 | S2-wms-combined_137119 | Unassigned | Unassigned | Unassigned | 710_1 | Clustered/Singleton | 1 | VC_710_1 | 1 | 0 | 1 | 0 | 0 | 0 | 0 | 1 | SHHC |
| 16724 | S2-wms-combined_140996 | Unassigned | Unassigned | Unassigned | 711_3 | Clustered/Singleton | 1 | VC_711_3 | 1 | 0 | 1 | 0 | 0 | 0 | 0 | 1 | SHHC |
| 16725 | S2-wms-combined_157934 | Unassigned | Unassigned | Unassigned | 1235_1 | Clustered/Singleton | 1 | VC_1235_1 | 1 | 0 | 1 | 0 | 0 | 0 | 0 | 1 | SHHC |
| 16726 | S2-wms-combined_165833 | Unassigned | Unassigned | Unassigned | 1235_0 | Clustered | 12 | VC_1235_0 | 12 | 0.7997 | 1 | 0.7997 | 0 | 0 | 0 | 1 | SHHC |
| 16727 | S2-wms-combined_166665 | Unassigned | Unassigned | Unassigned | 324_0 | Clustered | 2 | VC_324_0 | 2 | 0.2055 | 0.95514459 | 0.1963 | 0 | 0 | 0 | 1 | SHHC |
| 16728 | S2-wms-combined_174241 | Unassigned | Unassigned | Unassigned | 626_0 | Clustered | 23 | VC_626_0 | 23 | 0.66 | 1 | 0.66 | 0 | 0 | 0 | 0.8379 | SHHC |
| 16729 | S2-wms-combined_176376 | Unassigned | Unassigned | Unassigned | 825_0 | Clustered | 7 | VC_825_0 | 7 | 0.5674 | 1 | 0.5674 | 0 | 0 | 0 | 0.8571 | SHHC |
| 16730 | S2-wms-combined_180640 | Unassigned | Unassigned | Unassigned | 707_1 | Clustered | 9 | VC_707_1 | 9 | 0.2702 | 1 | 0.2702 | 0 | 0 | 0 | 0.8889 | SHHC |
| 16731 | S2-wms-combined_181095 | Unassigned | Unassigned | Unassigned | 1481_0 | Clustered | 9 | VC_1481_0 | 9 | 0.9416 | 0.77917309 | 0.7337 | 0 | 0 | 0 | 1 | SHHC |
| 16732 | S2-wms-combined_196783 | Unassigned | Unassigned | Unassigned | 629_0 | Clustered | 2 | VC_629_0 | 2 | 0.0041 | 0.93549515 | 0.0038 | 0 | 0 | 0 | 1 | SHHC |
| 16733 | S2-wms-combined_197577 | Unassigned | Unassigned | Unassigned | 1420_0 | Clustered | 5 | VC_1420_0 | 5 | 1 | 1 | 1 | 0 | 0 | 0 | 1 | SHHC |
| 16734 | S2-wms-combined_207832 | Unassigned | Unassigned | Unassigned | 717_0 | Clustered | 7 | VC_717_0 | 7 | 0.5813 | 1 | 0.5813 | 0 | 0 | 0 | 0.9048 | SHHC |
| 16735 | S2-wms-combined_210822 | Unassigned | Unassigned | Unassigned | 1495_0 | Clustered | 3 | VC_1495_0 | 3 | 0.7112 | 0.99363432 | 0.7067 | 0 | 0 | 0 | 1 | SHHC |
| 16736 | S2-wms-combined_218002 | Unassigned | Unassigned | Unassigned | 148_1 | Clustered | 3 | VC_148_1 | 3 | 0.4008 | 0.99752212 | 0.3998 | 0 | 0 | 0 | 1 | SHHC |
| 16737 | S2-wms-combined_227800 | Unassigned | Unassigned | Unassigned | 420_0 | Clustered | 6 | VC_420_0 | 6 | 0.2282 | 0.97462081 | 0.2224 | 0 | 0 | 0 | 1 | SHHC |
| 16738 | S2-wms-combined_229529 | Unassigned | Unassigned | Unassigned | 1473_0 | Clustered | 3 | VC_1473_0 | 3 | 0.7237 | 0.99613166 | 0.7209 | 0 | 0 | 0 | 0.6667 | SHHC |
| 16739 | S2-wms-combined_233107 | Unassigned | Unassigned | Unassigned | 1485_0 | Clustered/Singleton | 1 | VC_1485_0 | 1 | 0 | 1 | 0 | 0 | 0 | 0 | 1 | SHHC |
| 16740 | S2-wms-combined_238929 | Unassigned | Unassigned | Unassigned | 732_0 | Clustered | 2 | VC_732_0 | 2 | 0.0524 | 0.90742814 | 0.0476 | 0 | 0 | 0 | 1 | SHHC |
| 16741 | S2-wms-combined_244564 | Unassigned | Unassigned | Unassigned | 626_0 | Clustered | 23 | VC_626_0 | 23 | 0.66 | 1 | 0.66 | 0 | 0 | 0 | 0.8379 | SHHC |
| 16742 | S2-wms-combined_24549 | Unassigned | Unassigned | Unassigned | 651_0 | Clustered/Singleton | 1 | VC_651_0 | 1 | 0 | 1 | 0 | 0 | 0 | 0 | 1 | SHHC |
| 16743 | S2-wms-combined_246480 | Unassigned | Unassigned | Unassigned | 418_0 | Clustered | 6 | VC_418_0 | 6 | 0.4575 | 0.9999995 | 0.4575 | 0 | 0 | 0 | 0.8 | SHHC |
| 16744 | S2-wms-combined_248318 | Unassigned | Unassigned | Unassigned | 1498_0 | Clustered | 2 | VC_1498_0 | 2 | 0.9131 | 0.5 | 0.4566 | 0 | 0 | 0 | 1 | SHHC |
| 16745 | S2-wms-combined_248329 | Unassigned | Unassigned | Unassigned | 734_0 | Clustered | 6 | VC_734_0 | 6 | 0.1914 | 0.99996888 | 0.1914 | 0 | 0 | 0 | 1 | SHHC |
| 16746 | S2-wms-combined_31084 | Unassigned | Unassigned | Unassigned | 212_1 | Clustered | 2 | VC_212_1 | 2 | 0.164 | 0.95199974 | 0.1561 | 0 | 0 | 0 | 1 | SHHC |
| 16747 | S2-wms-combined_4685 | Unassigned | Unassigned | Unassigned | 1484_0 | Clustered | 10 | VC_1484_0 | 10 | 0.6918 | 0.99999998 | 0.6918 | 0 | 0 | 0 | 0.8889 | SHHC |
| 16748 | S2-wms-combined_55495 | Unassigned | Unassigned | Unassigned | 338_0 | Clustered | 5 | VC_338_0 | 5 | 0.3309 | 0.99999987 | 0.3309 | 0 | 0 | 0 | 1 | SHHC |
| 16749 | S2-wms-combined_58121 | Unassigned | Unassigned | Unassigned | 1436_0 | Clustered | 4 | VC_1436_0 | 4 | 0.571 | 0.96369953 | 0.5502 | 0 | 0 | 0 | 1 | SHHC |
| 16750 | S2-wms-combined_62377 | Unassigned | Unassigned | Unassigned | 804_2 | Clustered | 6 | VC_804_2 | 6 | 0.1161 | 1 | 0.1161 | 0 | 0 | 0 | 0.8667 | SHHC |
| 16751 | S2-wms-combined_62519 | Unassigned | Unassigned | Unassigned | 148_0 | Clustered | 2 | VC_148_0 | 2 | 0.2043 | 0.95366539 | 0.1949 | 0 | 0 | 0 | 1 | SHHC |
| 16752 | S2-wms-combined_6605 | Unassigned | Unassigned | Unassigned | 1484_0 | Clustered | 10 | VC_1484_0 | 10 | 0.6918 | 0.99999998 | 0.6918 | 0 | 0 | 0 | 0.8889 | SHHC |
| 16753 | S2-wms-combined_85381 | Unassigned | Unassigned | Unassigned | 707_1 | Clustered | 9 | VC_707_1 | 9 | 0.2702 | 1 | 0.2702 | 0 | 0 | 0 | 0.8889 | SHHC |
| 16754 | S2-wms-combined_89742 | Unassigned | Unassigned | Unassigned | 610_0 | Clustered | 4 | VC_610_0 | 4 | 0.8779 | 0.9998533 | 0.8778 | 0 | 0 | 0 | 1 | SHHC |
| 16755 | S2-wms-combined_93845 | Unassigned | Unassigned | Unassigned | 271_0 | Clustered | 5 | VC_271_0 | 5 | 0.3916 | 0.99999838 | 0.3916 | 0 | 0 | 0 | 0.8 | SHHC |
| 16756 | S2-wms-combined_98888 | Unassigned | Unassigned | Unassigned | 804_3 | Clustered/Singleton | 1 | VC_804_3 | 1 | 0 | 1 | 0 | 0 | 0 | 0 | 1 | SHHC |
| 16757 | S2-wms_107329 | Unassigned | Unassigned | Unassigned | 606_0 | Clustered | 4 | VC_606_0 | 4 | 0.4507 | 0.99995146 | 0.4507 | 0 | 0 | 0 | 1 | SHHC |
| 16758 | S2-wms_110761 | Unassigned | Unassigned | Unassigned | 1496_0 | Clustered | 5 | VC_1496_0 | 5 | 0.9228 | 0.98875286 | 0.9124 | 0 | 0 | 0 | 1 | SHHC |
| 16759 | S2-wms_128831 | Unassigned | Unassigned | Unassigned | 1420_0 | Clustered | 5 | VC_1420_0 | 5 | 1 | 1 | 1 | 0 | 0 | 0 | 1 | SHHC |
| 16760 | S2-wms_128962 | Unassigned | Unassigned | Unassigned | 502_0 | Clustered | 2 | VC_502_0 | 2 | 0.0737 | 0.95171362 | 0.0702 | 0 | 0 | 0 | 1 | SHHC |
| 16761 | S2-wms_130687 | Unassigned | Unassigned | Unassigned | 417_0 | Clustered | 5 | VC_417_0 | 5 | 0.6629 | 0.9977434 | 0.6614 | 0 | 0 | 0 | 0.8 | SHHC |
| 16762 | S2-wms_140933 | Unassigned | Unassigned | Unassigned | 418_0 | Clustered | 6 | VC_418_0 | 6 | 0.4575 | 0.9999995 | 0.4575 | 0 | 0 | 0 | 0.8 | SHHC |
| 16763 | S2-wms_156439 | Unassigned | Unassigned | Unassigned | 1435_0 | Clustered | 9 | VC_1435_0 | 9 | 0.6931 | 0.99999995 | 0.6931 | 0 | 0 | 0 | 1 | SHHC |
| 16764 | S2-wms_23562 | Unassigned | Unassigned | Unassigned | 1496_0 | Clustered | 5 | VC_1496_0 | 5 | 0.9228 | 0.98875286 | 0.9124 | 0 | 0 | 0 | 1 | SHHC |
| 16765 | S2-wms_248315 | Unassigned | Unassigned | Unassigned | 804_2 | Clustered | 6 | VC_804_2 | 6 | 0.1161 | 1 | 0.1161 | 0 | 0 | 0 | 0.8667 | SHHC |
| 16766 | S2-wms_53881 | Unassigned | Unassigned | Unassigned | 821_0 | Clustered | 4 | VC_821_0 | 4 | 0.0504 | 0.65965187 | 0.0333 | 0 | 0 | 0 | 1 | SHHC |
| 16767 | S2-wms_79304 | Unassigned | Unassigned | Unassigned | 744_0 | Clustered | 10 | VC_744_0 | 10 | 0.7049 | 1 | 0.7049 | 0 | 0 | 0 | 1 | SHHC |
| 16768 | S2-wms_79944 | Unassigned | Unassigned | Unassigned | 825_0 | Clustered | 7 | VC_825_0 | 7 | 0.5674 | 1 | 0.5674 | 0 | 0 | 0 | 0.8571 | SHHC |
| 16769 | S3-wms-combined_73262 | Unassigned | Unassigned | Unassigned | 677_0 | Clustered | 9 | VC_677_0 | 9 | 0.3787 | 1 | 0.3787 | 0 | 0 | 0 | 0.8333 | ME |
| 16770 | S3-wms_22744 | Unassigned | Unassigned | Unassigned | 717_0 | Clustered | 7 | VC_717_0 | 7 | 0.5813 | 1 | 0.5813 | 0 | 0 | 0 | 0.9048 | ME |
| 16771 | S3-wms_23958 | Unassigned | Unassigned | Unassigned | 915_0 | Clustered | 3 | VC_915_0 | 3 | 0.2219 | 0.99819349 | 0.2215 | 0 | 0 | 0 | 1 | ME |
| 16772 | S4-wms-combined_113721 | Unassigned | Unassigned | Unassigned | 893_0 | Clustered | 2 | VC_893_0 | 2 | 0.1729 | 0.93846104 | 0.1623 | 0 | 0 | 0 | 1 | SHHC |
| 16773 | S4-wms-combined_143879 | Unassigned | Unassigned | Unassigned | 1482_0 | Clustered | 2 | VC_1482_0 | 2 | 0.3552 | 0.85557782 | 0.3039 | 0 | 0 | 0 | 1 | SHHC |
| 16774 | S4-wms-combined_14411 | Unassigned | Unassigned | Unassigned | 831_0 | Clustered | 5 | VC_831_0 | 5 | 0.4369 | 0.99999952 | 0.4369 | 0 | 0 | 0 | 1 | SHHC |
| 16775 | S4-wms-combined_17284 | Unassigned | Unassigned | Unassigned | 852_0 | Clustered | 2 | VC_852_0 | 2 | 0.4064 | 0.94189878 | 0.3828 | 0 | 0 | 0 | 1 | SHHC |
| 16776 | S4-wms-combined_41490 | Unassigned | Unassigned | Unassigned | 610_0 | Clustered | 4 | VC_610_0 | 4 | 0.8779 | 0.9998533 | 0.8778 | 0 | 0 | 0 | 1 | SHHC |
| 16777 | S4-wms-combined_42647 | Unassigned | Unassigned | Unassigned | 621_0 | Clustered | 7 | VC_621_0 | 7 | 0.5783 | 1 | 0.5783 | 0 | 0 | 0 | 1 | SHHC |
| 16778 | S4-wms-combined_47565 | Unassigned | Unassigned | Unassigned | 1495_0 | Clustered | 3 | VC_1495_0 | 3 | 0.7112 | 0.99363432 | 0.7067 | 0 | 0 | 0 | 1 | SHHC |
| 16779 | S4-wms-combined_51079 | Unassigned | Unassigned | Unassigned | 1497_0 | Clustered | 3 | VC_1497_0 | 3 | 1 | 1 | 1 | 0 | 0 | 0 | 1 | SHHC |
| 16780 | S4-wms-combined_57123 | Unassigned | Unassigned | Unassigned | 1508_0 | Clustered | 3 | VC_1508_0 | 3 | 0.8977 | 0.9595722 | 0.8614 | 0 | 0 | 0 | 0.6667 | SHHC |
| 16781 | S4-wms-combined_64632 | Unassigned | Unassigned | Unassigned | 418_0 | Clustered | 6 | VC_418_0 | 6 | 0.4575 | 0.9999995 | 0.4575 | 0 | 0 | 0 | 0.8 | SHHC |
| 16782 | S4-wms_107182 | Unassigned | Unassigned | Unassigned | 1513_0 | Clustered | 2 | VC_1513_0 | 2 | 0.9263 | 0.5 | 0.4632 | 0 | 0 | 0 | 1 | SHHC |
| 16783 | S4-wms_118690 | Unassigned | Unassigned | Unassigned | 535_0 | Clustered | 2 | VC_535_0 | 2 | 0.1332 | 0.94063346 | 0.1253 | 0 | 0 | 0 | 1 | SHHC |
| 16784 | S4-wms_141607 | Unassigned | Unassigned | Unassigned | 1008_0 | Clustered | 4 | VC_1008_0 | 4 | 0.6602 | 0.99993542 | 0.6601 | 0 | 0 | 0 | 1 | SHHC |
| 16785 | S4-wms_161443 | Unassigned | Unassigned | Unassigned | 1477_0 | Clustered | 3 | VC_1477_0 | 3 | 1 | 1 | 1 | 0 | 0 | 0 | 1 | SHHC |
| 16786 | S4-wms_45128 | Unassigned | Unassigned | Unassigned | 1439_0 | Clustered | 3 | VC_1439_0 | 3 | 0.4461 | 0.98736442 | 0.4405 | 0 | 0 | 0 | 1 | SHHC |
| 16787 | S4-wms_62635 | Unassigned | Unassigned | Unassigned | 418_0 | Clustered | 6 | VC_418_0 | 6 | 0.4575 | 0.9999995 | 0.4575 | 0 | 0 | 0 | 0.8 | SHHC |
| 16788 | S4-wms_73990 | Unassigned | Unassigned | Unassigned | 502_1 | Clustered | 6 | VC_502_1 | 6 | 0.234 | 1 | 0.234 | 0 | 0 | 0 | 1 | SHHC |
| 16789 | S5-wms-combined_108794 | Unassigned | Unassigned | Unassigned | 626_3 | Clustered/Singleton | 1 | VC_626_3 | 1 | 0 | 1 | 0 | 0 | 0 | 0 | 1 | ME |
| 16790 | S5-wms-combined_117213 | Unassigned | Unassigned | Unassigned | 854_0 | Clustered | 2 | VC_854_0 | 2 | 0.4056 | 0.9446828 | 0.3831 | 0 | 0 | 0 | 1 | ME |
| 16791 | S5-wms-combined_134381 | Unassigned | Unassigned | Unassigned | 717_0 | Clustered | 7 | VC_717_0 | 7 | 0.5813 | 1 | 0.5813 | 0 | 0 | 0 | 0.9048 | ME |
| 16792 | S5-wms-combined_134743 | Unassigned | Unassigned | Unassigned | 825_2 | Clustered/Singleton | 1 | VC_825_2 | 1 | 0 | 1 | 0 | 0 | 0 | 0 | 1 | ME |
| 16793 | S5-wms-combined_145067 | Unassigned | Unassigned | Unassigned | 650_0 | Clustered | 10 | VC_650_0 | 10 | 0.4538 | 1 | 0.4538 | 0 | 0 | 0 | 1 | ME |
| 16794 | S5-wms-combined_178432 | Unassigned | Unassigned | Unassigned | 1506_0 | Clustered | 2 | VC_1506_0 | 2 | 0.2109 | 0.80863346 | 0.1706 | 0 | 0 | 0 | 1 | ME |
| 16795 | S5-wms-combined_18356 | Unassigned | Unassigned | Unassigned | 5_0 | Clustered | 30 | VC_5_0 | 30 | 0.9647 | 1 | 0.9647 | 0 | 0 | 0 | 1 | ME |
| 16796 | S5-wms-combined_206050 | Unassigned | Unassigned | Unassigned | 626_0 | Clustered | 23 | VC_626_0 | 23 | 0.66 | 1 | 0.66 | 0 | 0 | 0 | 0.8379 | ME |
| 16797 | S5-wms-combined_207291 | Unassigned | Unassigned | Unassigned | 1437_0 | Clustered | 3 | VC_1437_0 | 3 | 0.742 | 0.99188498 | 0.7359 | 0 | 0 | 0 | 1 | ME |
| 16798 | S5-wms-combined_26971 | Unassigned | Unassigned | Unassigned | 876_0 | Clustered | 3 | VC_876_0 | 3 | 0.1628 | 0.99746631 | 0.1624 | 0 | 0 | 0 | 0.6667 | ME |
| 16799 | S5-wms-combined_29272 | Unassigned | Unassigned | Unassigned | 626_0 | Clustered | 23 | VC_626_0 | 23 | 0.66 | 1 | 0.66 | 0 | 0 | 0 | 0.8379 | ME |
| 16800 | S5-wms-combined_29447 | Unassigned | Unassigned | Unassigned | 676_0 | Clustered | 2 | VC_676_0 | 2 | 0.1026 | 0.95156715 | 0.0977 | 0 | 0 | 0 | 1 | ME |
| 16801 | S5-wms-combined_39793 | Unassigned | Unassigned | Unassigned | 1514_0 | Clustered | 2 | VC_1514_0 | 2 | 1 | 1 | 1 | 0 | 0 | 0 | 1 | ME |
| 16802 | S5-wms-combined_52741 | Unassigned | Unassigned | Unassigned | 736_0 | Clustered | 3 | VC_736_0 | 3 | 0.5056 | 0.95195384 | 0.4813 | 0 | 0 | 0 | 1 | ME |
| 16803 | S5-wms-combined_59064 | Unassigned | Unassigned | Unassigned | 677_0 | Clustered | 9 | VC_677_0 | 9 | 0.3787 | 1 | 0.3787 | 0 | 0 | 0 | 0.8333 | ME |
| 16804 | S5-wms-combined_66017 | Unassigned | Unassigned | Unassigned | 1499_0 | Clustered | 2 | VC_1499_0 | 2 | 0.3148 | 0.5 | 0.1574 | 0 | 0 | 0 | 1 | ME |
| 16805 | S5-wms-combined_66662 | Unassigned | Unassigned | Unassigned | 1219_1 | Clustered/Singleton | 1 | VC_1219_1 | 1 | 0 | 1 | 0 | 0 | 0 | 0 | 1 | ME |
| 16806 | S5-wms-combined_72410 | Unassigned | Unassigned | Unassigned | 804_4 | Clustered/Singleton | 1 | VC_804_4 | 1 | 0 | 1 | 0 | 0 | 0 | 0 | 1 | ME |
| 16807 | S5-wms-combined_7292 | Unassigned | Unassigned | Unassigned | 734_0 | Clustered | 6 | VC_734_0 | 6 | 0.1914 | 0.99996888 | 0.1914 | 0 | 0 | 0 | 1 | ME |
| 16808 | S5-wms-combined_73301 | Unassigned | Unassigned | Unassigned | 627_0 | Clustered | 3 | VC_627_0 | 3 | 0.2127 | 0.99411368 | 0.2115 | 0 | 0 | 0 | 1 | ME |
| 16809 | S5-wms-combined_78434 | Unassigned | Unassigned | Unassigned | 1479_0 | Clustered | 2 | VC_1479_0 | 2 | 0.4887 | 0.93846104 | 0.4586 | 0 | 0 | 0 | 1 | ME |
| 16810 | S5-wms-combined_83370 | Unassigned | Unassigned | Unassigned | 680_0 | Clustered | 2 | VC_680_0 | 2 | 0.1163 | 0.94939599 | 0.1104 | 0 | 0 | 0 | 1 | ME |
| 16811 | S5-wms-combined_83412 | Unassigned | Unassigned | Unassigned | 19_0 | Clustered | 2 | VC_19_0 | 2 | 0.0418 | 0.92044806 | 0.0385 | 0 | 0 | 0 | 1 | ME |
| 16812 | S5-wms-combined_90315 | Unassigned | Unassigned | Unassigned | 917_1 | Clustered/Singleton | 1 | VC_917_1 | 1 | 0 | 1 | 0 | 0 | 0 | 0 | 1 | ME |
| 16813 | S5-wms-combined_98202 | Unassigned | Unassigned | Unassigned | 1503_0 | Clustered | 3 | VC_1503_0 | 3 | 0.9111 | 0.99054823 | 0.9025 | 0 | 0 | 0 | 1 | ME |
| 16814 | S5-wms_210460 | Unassigned | Unassigned | Unassigned | 1499_0 | Clustered | 2 | VC_1499_0 | 2 | 0.3148 | 0.5 | 0.1574 | 0 | 0 | 0 | 1 | ME |
| 16815 | S5-wms_49310 | Unassigned | Unassigned | Unassigned | 417_0 | Clustered | 5 | VC_417_0 | 5 | 0.6629 | 0.9977434 | 0.6614 | 0 | 0 | 0 | 0.8 | ME |
| 16816 | S5-wms_52566 | Unassigned | Unassigned | Unassigned | 1481_0 | Clustered | 9 | VC_1481_0 | 9 | 0.9416 | 0.77917309 | 0.7337 | 0 | 0 | 0 | 1 | ME |
| 16817 | S5-wms_67169 | Unassigned | Unassigned | Unassigned | 914_0 | Clustered/Singleton | 1 | VC_914_0 | 1 | 0 | 1 | 0 | 0 | 0 | 0 | 1 | ME |
| 16818 | S5-wms_85650 | Unassigned | Unassigned | Unassigned | 1505_0 | Clustered | 4 | VC_1505_0 | 4 | 0.9495 | 0.99879268 | 0.9484 | 0 | 0 | 0 | 1 | ME |
| 16819 | S5-wms_94965 | Unassigned | Unassigned | Unassigned | 1198_0 | Clustered | 3 | VC_1198_0 | 3 | 0.2794 | 0.95148141 | 0.2658 | 0 | 0 | 0 | 1 | ME |
| 16820 | S6-wms-combined_113986 | Unassigned | Unassigned | Unassigned | 627_0 | Clustered | 3 | VC_627_0 | 3 | 0.2127 | 0.99411368 | 0.2115 | 0 | 0 | 0 | 1 | SHHC |
| 16821 | S6-wms-combined_117756 | Unassigned | Unassigned | Unassigned | 707_1 | Clustered | 9 | VC_707_1 | 9 | 0.2702 | 1 | 0.2702 | 0 | 0 | 0 | 0.8889 | SHHC |
| 16822 | S6-wms-combined_118427 | Unassigned | Unassigned | Unassigned | 710_3 | Clustered/Singleton | 1 | VC_710_3 | 1 | 0 | 1 | 0 | 0 | 0 | 0 | 1 | SHHC |
| 16823 | S6-wms-combined_118484 | Unassigned | Unassigned | Unassigned | 338_0 | Clustered | 5 | VC_338_0 | 5 | 0.3309 | 0.99999987 | 0.3309 | 0 | 0 | 0 | 1 | SHHC |
| 16824 | S6-wms-combined_119017 | Unassigned | Unassigned | Unassigned | 5_0 | Clustered | 30 | VC_5_0 | 30 | 0.9647 | 1 | 0.9647 | 0 | 0 | 0 | 1 | SHHC |
| 16825 | S6-wms-combined_119138 | Unassigned | Unassigned | Unassigned | 148_1 | Clustered | 3 | VC_148_1 | 3 | 0.4008 | 0.99752212 | 0.3998 | 0 | 0 | 0 | 1 | SHHC |
| 16826 | S6-wms-combined_126236 | Unassigned | Unassigned | Unassigned | 710_0 | Clustered | 10 | VC_710_0 | 10 | 0.2446 | 1 | 0.2446 | 0 | 0 | 0 | 0.8222 | SHHC |
| 16827 | S6-wms-combined_135223 | Unassigned | Unassigned | Unassigned | 1508_0 | Clustered | 3 | VC_1508_0 | 3 | 0.8977 | 0.9595722 | 0.8614 | 0 | 0 | 0 | 0.6667 | SHHC |
| 16828 | S6-wms-combined_140731 | Unassigned | Unassigned | Unassigned | 825_0 | Clustered | 7 | VC_825_0 | 7 | 0.5674 | 1 | 0.5674 | 0 | 0 | 0 | 0.8571 | SHHC |
| 16829 | S6-wms-combined_32530 | Unassigned | Unassigned | Unassigned | 148_1 | Clustered | 3 | VC_148_1 | 3 | 0.4008 | 0.99752212 | 0.3998 | 0 | 0 | 0 | 1 | SHHC |
| 16830 | S6-wms-combined_41246 | Unassigned | Unassigned | Unassigned | 1484_0 | Clustered | 10 | VC_1484_0 | 10 | 0.6918 | 0.99999998 | 0.6918 | 0 | 0 | 0 | 0.8889 | SHHC |
| 16831 | S6-wms-combined_49909 | Unassigned | Unassigned | Unassigned | 324_0 | Clustered | 2 | VC_324_0 | 2 | 0.2055 | 0.95514459 | 0.1963 | 0 | 0 | 0 | 1 | SHHC |
| 16832 | S6-wms-combined_50721 | Unassigned | Unassigned | Unassigned | 676_1 | Clustered | 4 | VC_676_1 | 4 | 0.3759 | 0.9999693 | 0.3759 | 0 | 0 | 0 | 0.6667 | SHHC |
| 16833 | S6-wms-combined_72228 | Unassigned | Unassigned | Unassigned | 1506_0 | Clustered | 2 | VC_1506_0 | 2 | 0.2109 | 0.80863346 | 0.1706 | 0 | 0 | 0 | 1 | SHHC |
| 16834 | S6-wms-combined_82597 | Unassigned | Unassigned | Unassigned | 1435_1 | Clustered/Singleton | 1 | VC_1435_1 | 1 | 0 | 1 | 0 | 0 | 0 | 0 | 1 | SHHC |
| 16835 | S6-wms-combined_8508 | Unassigned | Unassigned | Unassigned | 707_0 | Clustered | 6 | VC_707_0 | 6 | 0.2616 | 1 | 0.2616 | 0 | 0 | 0 | 0.8667 | SHHC |
| 16836 | S6-wms-combined_91039 | Unassigned | Unassigned | Unassigned | 1456_0 | Clustered | 4 | VC_1456_0 | 4 | 0.6121 | 0.99987744 | 0.612 | 0 | 0 | 0 | 1 | SHHC |
| 16837 | S6-wms_132588 | Unassigned | Unassigned | Unassigned | 212_0 | Clustered | 2 | VC_212_0 | 2 | 0.3005 | 0.9466884 | 0.2845 | 0 | 0 | 0 | 1 | SHHC |
| 16838 | S6-wms_137591 | Unassigned | Unassigned | Unassigned | 1496_0 | Clustered | 5 | VC_1496_0 | 5 | 0.9228 | 0.98875286 | 0.9124 | 0 | 0 | 0 | 1 | SHHC |
| 16839 | S6-wms_138342 | Unassigned | Unassigned | Unassigned | 502_0 | Clustered | 2 | VC_502_0 | 2 | 0.0737 | 0.95171362 | 0.0702 | 0 | 0 | 0 | 1 | SHHC |
| 16840 | S6-wms_54578 | Unassigned | Unassigned | Unassigned | 831_0 | Clustered | 5 | VC_831_0 | 5 | 0.4369 | 0.99999952 | 0.4369 | 0 | 0 | 0 | 1 | SHHC |
| 16841 | S7-wms-combined_104872 | Unassigned | Unassigned | Unassigned | 1437_0 | Clustered | 3 | VC_1437_0 | 3 | 0.742 | 0.99188498 | 0.7359 | 0 | 0 | 0 | 1 | ME |
| 16842 | S7-wms-combined_107679_0 | Unassigned | Unassigned | Unassigned | 676_1 | Clustered | 4 | VC_676_1 | 4 | 0.3759 | 0.9999693 | 0.3759 | 0 | 0 | 0 | 0.6667 | ME |
| 16843 | S7-wms-combined_111004 | Unassigned | Unassigned | Unassigned | 711_0 | Clustered | 3 | VC_711_0 | 3 | 0.2312 | 0.99842495 | 0.2309 | 0 | 0 | 0 | 1 | ME |
| 16844 | S7-wms-combined_134458 | Unassigned | Unassigned | Unassigned | 710_0 | Clustered | 10 | VC_710_0 | 10 | 0.2446 | 1 | 0.2446 | 0 | 0 | 0 | 0.8222 | ME |
| 16845 | S7-wms-combined_136642 | Unassigned | Unassigned | Unassigned | 1497_0 | Clustered | 3 | VC_1497_0 | 3 | 1 | 1 | 1 | 0 | 0 | 0 | 1 | ME |
| 16846 | S7-wms-combined_139683 | Unassigned | Unassigned | Unassigned | 876_0 | Clustered | 3 | VC_876_0 | 3 | 0.1628 | 0.99746631 | 0.1624 | 0 | 0 | 0 | 0.6667 | ME |

| | | | | | | | | | | | | | | | | | |
|---|---|---|---|---|---|---|---|---|---|---|---|---|---|---|---|---|---|
| 16847 | S7-wms-combined_14267 | Unassigned | Unassigned | Unassigned | 1456_0 | Clustered | 4 | VC_1456_0 | 4 | 0.6121 | 0.99987744 | 0.612 | 0 | 0 | 0 | 1 | ME |
| 16848 | S7-wms-combined_143218 | Unassigned | Unassigned | Unassigned | 1514_0 | Clustered | 2 | VC_1514_0 | 2 | 1 | 1 | 1 | 0 | 0 | 0 | 1 | ME |
| 16849 | S7-wms-combined_147158 | Unassigned | Unassigned | Unassigned | 798_0 | Clustered | 2 | VC_798_0 | 2 | 0.1818 | 0.94346013 | 0.1715 | 0 | 0 | 0 | 1 | ME |
| 16850 | S7-wms-combined_157260 | Unassigned | Unassigned | Unassigned | 710_2 | Clustered/Singleton | 1 | VC_710_2 | 1 | 0 | 1 | 0 | 0 | 0 | 0 | 1 | ME |
| 16851 | S7-wms-combined_163980 | Unassigned | Unassigned | Unassigned | 1014_0 | Clustered | 3 | VC_1014_0 | 3 | 0.2026 | 0.85591218 | 0.1734 | 0 | 0 | 0 | 0.6667 | ME |
| 16852 | S7-wms-combined_169375 | Unassigned | Unassigned | Unassigned | 1483_0 | Clustered/Singleton | 1 | VC_1483_0 | 1 | 0 | 1 | 0 | 0 | 0 | 0 | 1 | ME |
| 16853 | S7-wms-combined_185016 | Unassigned | Unassigned | Unassigned | 1435_0 | Clustered | 9 | VC_1435_0 | 9 | 0.6931 | 0.99999995 | 0.6931 | 0 | 0 | 0 | 1 | ME |
| 16854 | S7-wms-combined_189250 | Unassigned | Unassigned | Unassigned | 1198_0 | Clustered | 3 | VC_1198_0 | 3 | 0.2794 | 0.95148141 | 0.2658 | 0 | 0 | 0 | 1 | ME |
| 16855 | S7-wms-combined_20921 | Unassigned | Unassigned | Unassigned | 719_1 | Clustered/Singleton | 1 | VC_719_1 | 1 | 0 | 1 | 0 | 0 | 0 | 0 | 1 | ME |
| 16856 | S7-wms-combined_225260 | Unassigned | Unassigned | Unassigned | 822_0 | Clustered | 2 | VC_822_0 | 2 | 0.038 | 0.89489263 | 0.034 | 0 | 0 | 0 | 1 | ME |
| 16857 | S7-wms-combined_235165 | Unassigned | Unassigned | Unassigned | 1109_0 | Clustered | 2 | VC_1109_0 | 2 | 0.6449 | 0.85557782 | 0.5517 | 0 | 0 | 0 | 1 | ME |
| 16858 | S7-wms-combined_238231 | Unassigned | Unassigned | Unassigned | 271_2 | Clustered/Singleton | 1 | VC_271_2 | 1 | 0 | 1 | 0 | 0 | 0 | 0 | 1 | ME |
| 16859 | S7-wms-combined_239784 | Unassigned | Unassigned | Unassigned | 1436_0 | Clustered | 4 | VC_1436_0 | 4 | 0.571 | 0.96369953 | 0.5502 | 0 | 0 | 0 | 1 | ME |
| 16860 | S7-wms-combined_26217 | Unassigned | Unassigned | Unassigned | 918_1 | Clustered/Singleton | 1 | VC_918_1 | 1 | 0 | 1 | 0 | 0 | 0 | 0 | 1 | ME |
| 16861 | S7-wms-combined_28138 | Unassigned | Unassigned | Unassigned | 420_0 | Clustered | 6 | VC_420_0 | 6 | 0.2282 | 0.97462081 | 0.2224 | 0 | 0 | 0 | 1 | ME |
| 16862 | S7-wms-combined_31068 | Unassigned | Unassigned | Unassigned | 650_0 | Clustered | 10 | VC_650_0 | 10 | 0.4538 | 1 | 0.4538 | 0 | 0 | 0 | 1 | ME |
| 16863 | S7-wms-combined_3697 | Unassigned | Unassigned | Unassigned | 832_1 | Clustered/Singleton | 1 | VC_832_1 | 1 | 0 | 1 | 0 | 0 | 0 | 0 | 1 | ME |
| 16864 | S7-wms-combined_40779 | Unassigned | Unassigned | Unassigned | 1239_0 | Clustered | 6 | VC_1239_0 | 6 | 0.4699 | 0.99999972 | 0.4699 | 0 | 0 | 0 | 1 | ME |
| 16865 | S7-wms-combined_43950 | Unassigned | Unassigned | Unassigned | 677_0 | Clustered | 9 | VC_677_0 | 9 | 0.3787 | 1 | 0.3787 | 0 | 0 | 0 | 0.8333 | ME |
| 16866 | S7-wms-combined_5625 | Unassigned | Unassigned | Unassigned | 677_0 | Clustered | 9 | VC_677_0 | 9 | 0.3787 | 1 | 0.3787 | 0 | 0 | 0 | 0.8333 | ME |
| 16867 | S7-wms-combined_62119 | Unassigned | Unassigned | Unassigned | 1221_1 | Clustered/Singleton | 1 | VC_1221_1 | 1 | 0 | 1 | 0 | 0 | 0 | 0 | 1 | ME |
| 16868 | S7-wms-combined_7205 | Unassigned | Unassigned | Unassigned | 212_1 | Clustered | 2 | VC_212_1 | 2 | 0.164 | 0.95199974 | 0.1561 | 0 | 0 | 0 | 1 | ME |
| 16869 | S7-wms-combined_72070 | Unassigned | Unassigned | Unassigned | 1509_0 | Clustered | 2 | VC_1509_0 | 2 | 0.5962 | 0.91237788 | 0.5439 | 0 | 0 | 0 | 1 | ME |
| 16870 | S7-wms-combined_84688 | Unassigned | Unassigned | Unassigned | 1484_0 | Clustered | 10 | VC_1484_0 | 10 | 0.6918 | 0.99999998 | 0.6918 | 0 | 0 | 0 | 0.8889 | ME |
| 16871 | S7-wms-combined_90393 | Unassigned | Unassigned | Unassigned | 1435_0 | Clustered | 9 | VC_1435_0 | 9 | 0.6931 | 0.99999995 | 0.6931 | 0 | 0 | 0 | 1 | ME |
| 16872 | S7-wms-combined_90728 | Unassigned | Unassigned | Unassigned | 1435_0 | Clustered | 9 | VC_1435_0 | 9 | 0.6931 | 0.99999995 | 0.6931 | 0 | 0 | 0 | 1 | ME |
| 16873 | S7-wms_113137 | Unassigned | Unassigned | Unassigned | 1235_0 | Clustered | 12 | VC_1235_0 | 12 | 0.7997 | 1 | 0.7997 | 0 | 0 | 0 | 1 | ME |
| 16874 | S7-wms_118607 | Unassigned | Unassigned | Unassigned | 717_0 | Clustered | 7 | VC_717_0 | 7 | 0.5813 | 1 | 0.5813 | 0 | 0 | 0 | 0.9048 | ME |
| 16875 | S7-wms_123954 | Unassigned | Unassigned | Unassigned | 1435_0 | Clustered | 9 | VC_1435_0 | 9 | 0.6931 | 0.99999995 | 0.6931 | 0 | 0 | 0 | 1 | ME |
| 16876 | S7-wms_127801 | Unassigned | Unassigned | Unassigned | 420_0 | Clustered | 6 | VC_420_0 | 6 | 0.2282 | 0.97462081 | 0.2224 | 0 | 0 | 0 | 1 | ME |
| 16877 | S7-wms_157264 | Unassigned | Unassigned | Unassigned | 1477_0 | Clustered | 3 | VC_1477_0 | 3 | 1 | 1 | 1 | 0 | 0 | 0 | 1 | ME |
| 16878 | S7-wms_159432 | Unassigned | Unassigned | Unassigned | 707_1 | Clustered | 9 | VC_707_1 | 9 | 0.2702 | 1 | 0.2702 | 0 | 0 | 0 | 0.8889 | ME |
| 16879 | S7-wms_200936 | Unassigned | Unassigned | Unassigned | 1235_0 | Clustered | 12 | VC_1235_0 | 12 | 0.7997 | 1 | 0.7997 | 0 | 0 | 0 | 1 | ME |
| 16880 | S7-wms_239833 | Unassigned | Unassigned | Unassigned | 821_0 | Clustered | 4 | VC_821_0 | 4 | 0.0504 | 0.65965187 | 0.0333 | 0 | 0 | 0 | 1 | ME |
| 16881 | S7-wms_244425 | Unassigned | Unassigned | Unassigned | 675_0 | Clustered | 2 | VC_675_0 | 2 | 0.081 | 0.95018582 | 0.077 | 0 | 0 | 0 | 1 | ME |
| 16882 | S7-wms_244866 | Unassigned | Unassigned | Unassigned | 1009_0 | Clustered | 3 | VC_1009_0 | 3 | 0.127 | 0.99814461 | 0.1267 | 0 | 0 | 0 | 1 | ME |
| 16883 | S7-wms_53338 | Unassigned | Unassigned | Unassigned | 821_0 | Clustered | 4 | VC_821_0 | 4 | 0.0504 | 0.65965187 | 0.0333 | 0 | 0 | 0 | 1 | ME |
| 16884 | S7-wms_80255 | Unassigned | Unassigned | Unassigned | 629_0 | Clustered | 2 | VC_629_0 | 2 | 0.0041 | 0.93549515 | 0.0038 | 0 | 0 | 0 | 1 | ME |
| 16885 | S8-wms-combined_108917 | Unassigned | Unassigned | Unassigned | 626_0 | Clustered | 23 | VC_626_0 | 23 | 0.66 | 1 | 0.66 | 0 | 0 | 0 | 0.8379 | SHHC |
| 16886 | S8-wms-combined_134098 | Unassigned | Unassigned | Unassigned | 1239_0 | Clustered | 6 | VC_1239_0 | 6 | 0.4699 | 0.99999972 | 0.4699 | 0 | 0 | 0 | 1 | SHHC |
| 16887 | S8-wms-combined_134284 | Unassigned | Unassigned | Unassigned | 1236_0 | Clustered | 3 | VC_1236_0 | 3 | 0.1415 | 0.99774774 | 0.1412 | 0 | 0 | 0 | 1 | SHHC |
| 16888 | S8-wms-combined_140059 | Unassigned | Unassigned | Unassigned | 420_0 | Clustered | 6 | VC_420_0 | 6 | 0.2282 | 0.97462081 | 0.2224 | 0 | 0 | 0 | 1 | SHHC |
| 16889 | S8-wms-combined_146562 | Unassigned | Unassigned | Unassigned | 271_0 | Clustered | 5 | VC_271_0 | 5 | 0.3916 | 0.99999838 | 0.3916 | 0 | 0 | 0 | 0.8 | SHHC |
| 16890 | S8-wms-combined_161181 | Unassigned | Unassigned | Unassigned | 148_0 | Clustered | 2 | VC_148_0 | 2 | 0.2043 | 0.95366539 | 0.1949 | 0 | 0 | 0 | 1 | SHHC |
| 16891 | S8-wms-combined_169835 | Unassigned | Unassigned | Unassigned | 915_0 | Clustered | 3 | VC_915_0 | 3 | 0.2219 | 0.99819349 | 0.2215 | 0 | 0 | 0 | 1 | SHHC |
| 16892 | S8-wms-combined_186054 | Unassigned | Unassigned | Unassigned | 626_0 | Clustered | 23 | VC_626_0 | 23 | 0.66 | 1 | 0.66 | 0 | 0 | 0 | 0.8379 | SHHC |
| 16893 | S8-wms-combined_200168 | Unassigned | Unassigned | Unassigned | 707_1 | Clustered | 9 | VC_707_1 | 9 | 0.2702 | 1 | 0.2702 | 0 | 0 | 0 | 0.8889 | SHHC |
| 16894 | S8-wms-combined_211119 | Unassigned | Unassigned | Unassigned | 1236_0 | Clustered | 3 | VC_1236_0 | 3 | 0.1415 | 0.99774774 | 0.1412 | 0 | 0 | 0 | 1 | SHHC |
| 16895 | S8-wms-combined_215102 | Unassigned | Unassigned | Unassigned | 1504_0 | Clustered | 2 | VC_1504_0 | 2 | 0.9646 | 0.5 | 0.4823 | 0 | 0 | 0 | 1 | SHHC |
| 16896 | S8-wms-combined_217866 | Unassigned | Unassigned | Unassigned | 707_1 | Clustered | 9 | VC_707_1 | 9 | 0.2702 | 1 | 0.2702 | 0 | 0 | 0 | 0.8889 | SHHC |
| 16897 | S8-wms-combined_225114 | Unassigned | Unassigned | Unassigned | 271_1 | Clustered/Singleton | 1 | VC_271_1 | 1 | 0 | 1 | 0 | 0 | 0 | 0 | 1 | SHHC |
| 16898 | S8-wms-combined_226076 | Unassigned | Unassigned | Unassigned | 1511_0 | Clustered | 2 | VC_1511_0 | 2 | 1 | 1 | 1 | 0 | 0 | 0 | 1 | SHHC |
| 16899 | S8-wms-combined_234473 | Unassigned | Unassigned | Unassigned | 1481_0 | Clustered | 9 | VC_1481_0 | 9 | 0.9416 | 0.77917309 | 0.7337 | 0 | 0 | 0 | 1 | SHHC |
| 16900 | S8-wms-combined_246185 | Unassigned | Unassigned | Unassigned | 738_0 | Clustered | 2 | VC_738_0 | 2 | 0.6709 | 0.85557782 | 0.574 | 0 | 0 | 0 | 1 | SHHC |
| 16901 | S8-wms-combined_25607 | Unassigned | Unassigned | Unassigned | 1239_0 | Clustered | 6 | VC_1239_0 | 6 | 0.4699 | 0.99999972 | 0.4699 | 0 | 0 | 0 | 1 | SHHC |
| 16902 | S8-wms-combined_26087 | Unassigned | Unassigned | Unassigned | 610_0 | Clustered | 4 | VC_610_0 | 4 | 0.8779 | 0.9998533 | 0.8778 | 0 | 0 | 0 | 1 | SHHC |
| 16903 | S8-wms-combined_264711 | Unassigned | Unassigned | Unassigned | 1481_0 | Clustered | 9 | VC_1481_0 | 9 | 0.9416 | 0.77917309 | 0.7337 | 0 | 0 | 0 | 1 | SHHC |
| 16904 | S8-wms-combined_270362 | Unassigned | Unassigned | Unassigned | 917_0 | Clustered | 2 | VC_917_0 | 2 | 0.3004 | 0.94854123 | 0.2849 | 0 | 0 | 0 | 1 | SHHC |
| 16905 | S8-wms-combined_276204 | Unassigned | Unassigned | Unassigned | 1501_0 | Clustered | 2 | VC_1501_0 | 2 | 0.1152 | 0.71936208 | 0.0829 | 0 | 0 | 0 | 1 | SHHC |
| 16906 | S8-wms-combined_276211 | Unassigned | Unassigned | Unassigned | 1209_0 | Clustered | 3 | VC_1209_0 | 3 | 0.1553 | 0.99152019 | 0.154 | 0 | 0 | 0 | 1 | SHHC |
| 16907 | S8-wms-combined_40666 | Unassigned | Unassigned | Unassigned | 1483_1 | Clustered/Singleton | 1 | VC_1483_1 | 1 | 0 | 1 | 0 | 0 | 0 | 0 | 1 | SHHC |
| 16908 | S8-wms-combined_49369 | Unassigned | Unassigned | Unassigned | 1236_0 | Clustered | 3 | VC_1236_0 | 3 | 0.1415 | 0.99774774 | 0.1412 | 0 | 0 | 0 | 1 | SHHC |
| 16909 | S8-wms-combined_68780 | Unassigned | Unassigned | Unassigned | 271_0 | Clustered | 5 | VC_271_0 | 5 | 0.3916 | 0.99999838 | 0.3916 | 0 | 0 | 0 | 0.8 | SHHC |
| 16910 | S8-wms-combined_72911 | Unassigned | Unassigned | Unassigned | 1076_0 | Clustered | 2 | VC_1076_0 | 2 | 0.3461 | 0.6381632 | 0.2209 | 0 | 0 | 0 | 1 | SHHC |
| 16911 | S8-wms-combined_97848 | Unassigned | Unassigned | Unassigned | 677_0 | Clustered | 9 | VC_677_0 | 9 | 0.3787 | 1 | 0.3787 | 0 | 0 | 0 | 0.8333 | SHHC |
| 16912 | S8-wms_111056 | Unassigned | Unassigned | Unassigned | 674_0 | Clustered/Singleton | 1 | VC_674_0 | 1 | 0 | 1 | 0 | 0 | 0 | 0 | 1 | SHHC |
| 16913 | S8-wms_127191 | Unassigned | Unassigned | Unassigned | 804_2 | Clustered | 6 | VC_804_2 | 6 | 0.1161 | 1 | 0.1161 | 0 | 0 | 0 | 0.8667 | SHHC |
| 16914 | S8-wms_152831 | Unassigned | Unassigned | Unassigned | 710_0 | Clustered | 10 | VC_710_0 | 10 | 0.2446 | 1 | 0.2446 | 0 | 0 | 0 | 0.8222 | SHHC |
| 16915 | S8-wms_244525 | Unassigned | Unassigned | Unassigned | 821_0 | Clustered | 4 | VC_821_0 | 4 | 0.0504 | 0.65965187 | 0.0333 | 0 | 0 | 0 | 1 | SHHC |
| 16916 | S8-wms_30498 | Unassigned | Unassigned | Unassigned | 1235_0 | Clustered | 12 | VC_1235_0 | 12 | 0.7997 | 1 | 0.7997 | 0 | 0 | 0 | 1 | SHHC |
| 16917 | S8-wms_48935 | Unassigned | Unassigned | Unassigned | 710_0 | Clustered | 10 | VC_710_0 | 10 | 0.2446 | 1 | 0.2446 | 0 | 0 | 0 | 0.8222 | SHHC |
| 16918 | S8-wms_57121 | Unassigned | Unassigned | Unassigned | 1501_0 | Clustered | 2 | VC_1501_0 | 2 | 0.1152 | 0.71936208 | 0.0829 | 0 | 0 | 0 | 1 | SHHC |
| 16919 | S8-wms_82880 | Unassigned | Unassigned | Unassigned | 606_0 | Clustered | 4 | VC_606_0 | 4 | 0.4507 | 0.99995146 | 0.4507 | 0 | 0 | 0 | 1 | SHHC |
| 16920 | S9-wms-combined_105462 | Unassigned | Unassigned | Unassigned | 825_0 | Clustered | 7 | VC_825_0 | 7 | 0.5674 | 1 | 0.5674 | 0 | 0 | 0 | 0.8571 | ME |
| 16921 | S9-wms-combined_20485 | Unassigned | Unassigned | Unassigned | 255_1 | Clustered/Singleton | 1 | VC_255_1 | 1 | 0 | 1 | 0 | 0 | 0 | 0 | 1 | ME |
| 16922 | S9-wms-combined_25007 | Unassigned | Unassigned | Unassigned | 626_0 | Clustered | 23 | VC_626_0 | 23 | 0.66 | 1 | 0.66 | 0 | 0 | 0 | 0.8379 | ME |
| 16923 | S9-wms-combined_26410 | Unassigned | Unassigned | Unassigned | 804_2 | Clustered | 6 | VC_804_2 | 6 | 0.1161 | 1 | 0.1161 | 0 | 0 | 0 | 0.8667 | ME |
| 16924 | S9-wms-combined_28805 | Unassigned | Unassigned | Unassigned | 676_1 | Clustered | 4 | VC_676_1 | 4 | 0.3759 | 0.9999693 | 0.3759 | 0 | 0 | 0 | 0.6667 | ME |
| 16925 | S9-wms-combined_29611 | Unassigned | Unassigned | Unassigned | 606_1 | Clustered/Singleton | 1 | VC_606_1 | 1 | 0 | 1 | 0 | 0 | 0 | 0 | 1 | ME |
| 16926 | S9-wms-combined_31456 | Unassigned | Unassigned | Unassigned | 677_0 | Clustered | 9 | VC_677_0 | 9 | 0.3787 | 1 | 0.3787 | 0 | 0 | 0 | 0.8333 | ME |
| 16927 | S9-wms-combined_33229 | Unassigned | Unassigned | Unassigned | 1435_0 | Clustered | 9 | VC_1435_0 | 9 | 0.6931 | 0.99999995 | 0.6931 | 0 | 0 | 0 | 1 | ME |
| 16928 | S9-wms-combined_33230 | Unassigned | Unassigned | Unassigned | 1435_0 | Clustered | 9 | VC_1435_0 | 9 | 0.6931 | 0.99999995 | 0.6931 | 0 | 0 | 0 | 1 | ME |
| 16929 | S9-wms-combined_36459 | Unassigned | Unassigned | Unassigned | 338_0 | Clustered | 5 | VC_338_0 | 5 | 0.3309 | 0.99999987 | 0.3309 | 0 | 0 | 0 | 1 | ME |
| 16930 | S9-wms-combined_37587 | Unassigned | Unassigned | Unassigned | 1473_0 | Clustered | 3 | VC_1473_0 | 3 | 0.7237 | 0.99613166 | 0.7209 | 0 | 0 | 0 | 0.6667 | ME |
| 16931 | S9-wms-combined_3878 | Unassigned | Unassigned | Unassigned | 1510_0 | Clustered | 2 | VC_1510_0 | 2 | 1 | 1 | 1 | 0 | 0 | 0 | 1 | ME |
| 16932 | S9-wms-combined_44402 | Unassigned | Unassigned | Unassigned | 680_0 | Clustered | 2 | VC_680_0 | 2 | 0.1163 | 0.94939599 | 0.1104 | 0 | 0 | 0 | 1 | ME |
| 16933 | S9-wms-combined_53991 | Unassigned | Unassigned | Unassigned | 947_0 | Clustered | 3 | VC_947_0 | 3 | 0.0374 | 0.84591675 | 0.0316 | 0 | 0 | 0 | 1 | ME |
| 16934 | S9-wms-combined_5526 | Unassigned | Unassigned | Unassigned | 832_0 | Clustered | 2 | VC_832_0 | 2 | 0.2344 | 0.94526139 | 0.2215 | 0 | 0 | 0 | 1 | ME |
| 16935 | S9-wms-combined_56773 | Unassigned | Unassigned | Unassigned | 1481_0 | Clustered | 9 | VC_1481_0 | 9 | 0.9416 | 0.77917309 | 0.7337 | 0 | 0 | 0 | 1 | ME |
| 16936 | S9-wms-combined_64277 | Unassigned | Unassigned | Unassigned | 250_0 | Clustered | 4 | VC_250_0 | 4 | 0.2395 | 0.99998241 | 0.2395 | 0 | 0 | 0 | 1 | ME |
| 16937 | S9-wms-combined_71647 | Unassigned | Unassigned | Unassigned | 420_0 | Clustered | 6 | VC_420_0 | 6 | 0.2282 | 0.97462081 | 0.2224 | 0 | 0 | 0 | 1 | ME |
| 16938 | S9-wms-combined_76274 | Unassigned | Unassigned | Unassigned | 854_0 | Clustered | 2 | VC_854_0 | 2 | 0.4056 | 0.9446828 | 0.3831 | 0 | 0 | 0 | 1 | ME |
| 16939 | S9-wms-combined_89249 | Unassigned | Unassigned | Unassigned | 947_0 | Clustered | 3 | VC_947_0 | 3 | 0.0374 | 0.84591675 | 0.0316 | 0 | 0 | 0 | 1 | ME |
| 16940 | S9-wms-combined_9931 | Unassigned | Unassigned | Unassigned | 626_1 | Clustered/Singleton | 1 | VC_626_1 | 1 | 0 | 1 | 0 | 0 | 0 | 0 | 1 | ME |
| 16941 | S9-wms_104169 | Unassigned | Unassigned | Unassigned | 1423_0 | Clustered | 8 | VC_1423_0 | 8 | 0.896 | 0.99983932 | 0.8959 | 0 | 0 | 0 | 1 | ME |
| 16942 | S9-wms_128232 | Unassigned | Unassigned | Unassigned | 1243_0 | Clustered | 52 | VC_1243_0 | 52 | 0.9126 | 0.92708311 | 0.8461 | 0 | 0 | 0 | 1 | ME |
| 16943 | S9-wms_19025 | Unassigned | Unassigned | Unassigned | 535_0 | Clustered | 2 | VC_535_0 | 2 | 0.1332 | 0.94063346 | 0.1253 | 0 | 0 | 0 | 1 | ME |
| 16944 | S9-wms_64472 | Unassigned | Unassigned | Unassigned | 1512_0 | Clustered | 2 | VC_1512_0 | 2 | 1 | 1 | 1 | 0 | 0 | 0 | 1 | ME |
| 16945 | SFU82619 | Unassigned | Unassigned | Unassigned | 45_0 | Clustered | 12 | VC_45_0 | 12 | 0.35 | 1 | 0.35 | 0 | 0 | 0 | 1 | Ref |
| 16946 | SVU28974 | Unassigned | Unassigned | Unassigned | 794_0 | Clustered | 3 | VC_794_0 | 3 | 1 | 1 | 1 | 0 | 0 | 0 | 1 | Ref |
| 16947 | U88974 | Unassigned | Unassigned | Unassigned | 254_0 | Clustered | 46 | VC_254_0 | 46 | 0.5159 | 1 | 0.5159 | 0 | 0 | 0 | 1 | Ref |
| 16948 | V01146 | Unassigned | Unassigned | Unassigned | 392_0 | Clustered | 126 | VC_392_0 | 126 | 0.9621 | 1 | 0.9621 | 0 | 0 | 0 | 1 | Ref |
| 16949 | X07234 | Unassigned | Unassigned | Unassigned | 599_0 | Clustered | 50 | VC_599_0 | 50 | 0.9989 | 1 | 0.9989 | 0 | 0 | 0 | 1 | Ref |
| 16950 | X51344 | Unassigned | Unassigned | Unassigned | 794_0 | Clustered | 3 | VC_794_0 | 3 | 1 | 1 | 1 | 0 | 0 | 0 | 1 | Ref |
| 16951 | X60322 | Unassigned | Unassigned | Unassigned | 743_0 | Clustered | 15 | VC_743_0 | 15 | 1 | 1 | 1 | 0 | 0 | 0 | 1 | Ref |
| 16952 | X60323 | Unassigned | Unassigned | Unassigned | 743_0 | Clustered | 15 | VC_743_0 | 15 | 1 | 1 | 1 | 0 | 0 | 0 | 1 | Ref |
| 16953 | X97918 | Unassigned | Unassigned | Unassigned | 104_0 | Clustered | 6 | VC_104_0 | 6 | 0.8291 | 1 | 0.8291 | 0 | 0 | 0 | 1 | Ref |
| 16954 | X98106 | Unassigned | Unassigned | Unassigned | 309_0 | Clustered | 3 | VC_309_0 | 3 | 0.1265 | 0.99498896 | 0.1258 | 0 | 0 | 0 | 1 | Ref |
| 16955 | X99260 | Unassigned | Unassigned | Unassigned | 817_0 | Clustered | 14 | VC_817_0 | 14 | 0.6342 | 1 | 0.6342 | 0 | 0 | 0 | 1 | Ref |
| 16956 | Y13918 | Unassigned | Unassigned | Unassigned | 2_0 | Clustered | 4 | VC_2_0 | 4 | 0.1311 | 0.99998692 | 0.1311 | 0 | 0 | 0 | 1 | Ref |
| 16957 | Z18946 | Unassigned | Unassigned | Unassigned | 215_2 | Clustered | 5 | VC_215_2 | 5 | 0.0572 | 0.99999997 | 0.0572 | 0 | 0 | 0 | 1 | Ref |
| 16958 | Z47794 | Unassigned | Unassigned | Unassigned | 1048_0 | Clustered | 3 | VC_1048_0 | 3 | 1 | 1 | 1 | 0 | 0 | 0 | 1 | Ref |
| 15 | AB255436 | Unassigned | Unassigned | Unassigned | | Outlier | | | | | | | | | | | Ref |
| 29 | AB451219 | Unassigned | Unassigned | Unassigned | | Overlap (VC_71/VC_72) | | | | | | | | | | | Ref |
| 34 | AB605730 | Unassigned | Unassigned | Unassigned | | Overlap (VC_87/VC_88) | | | | | | | | | | | Ref |
| 39 | AB711120 | Unassigned | Unassigned | Unassigned | | Outlier | | | | | | | | | | | Ref |
| 48 | AB767244 | Unassigned | Unassigned | Unassigned | | Overlap (VC_121/VC_124) | | | | | | | | | | | Ref |
| 52 | AB823818 | Unassigned | Unassigned | Unassigned | | Overlap (VC_138/VC_146) | | | | | | | | | | | Ref |
| 68 | AB967974 | Unassigned | Unassigned | Unassigned | | Outlier | | | | | | | | | | | Ref |
| 82 | AF064539 | Unassigned | Unassigned | Unassigned | | Outlier | | | | | | | | | | | Ref |
| 84 | AF068845 | Unassigned | Unassigned | Unassigned | | Overlap (VC_236/VC_238) | | | | | | | | | | | Ref |
| 85 | AF069308 | Unassigned | Unassigned | Unassigned | | Overlap (VC_31/VC_34) | | | | | | | | | | | Ref |
| 86 | AF069529 | Unassigned | Unassigned | Unassigned | | Overlap (VC_31/VC_34) | | | | | | | | | | | Ref |
| 87 | AF074945 | Unassigned | Unassigned | Unassigned | | Outlier | | | | | | | | | | | Ref |
| 90 | AF109874 | Unassigned | Unassigned | Unassigned | | Overlap (VC_257/VC_262) | | | | | | | | | | | Ref |
| 96 | AF143772 | Unassigned | Unassigned | Unassigned | | Singleton | | | | | | | | | | | Ref |
| 113 | AF191796 | Unassigned | Unassigned | Unassigned | | Singleton | | | | | | | | | | | Ref |
| 122 | AF242738 | Unassigned | Unassigned | Unassigned | | Overlap (VC_263/VC_318) | | | | | | | | | | | Ref |
| 123 | AF246223 | Unassigned | Unassigned | Unassigned | | Singleton | | | | | | | | | | | Ref |
| 142 | AF304433 | Unassigned | Unassigned | Unassigned | | Overlap (VC_257/VC_262) | | | | | | | | | | | Ref |
| 147 | AF323670 | Unassigned | Unassigned | Unassigned | | Overlap (VC_261/VC_265) | | | | | | | | | | | Ref |
| 149 | AF323672 | Unassigned | Unassigned | Unassigned | | Outlier | | | | | | | | | | | Ref |
| 150 | AF323673 | Unassigned | Unassigned | Unassigned | | Outlier | | | | | | | | | | | Ref |
| 154 | AF349457 | Unassigned | Unassigned | Unassigned | | Overlap (VC_257/VC_262) | | | | | | | | | | | Ref |
| 160 | AF428260 | Unassigned | Unassigned | Unassigned | | Singleton | | | | | | | | | | | Ref |
| 164 | AF448724 | Unassigned | Unassigned | Unassigned | | Outlier | | | | | | | | | | | Ref |
| 174 | AF543311 | Unassigned | Unassigned | Unassigned | | Overlap (VC_365/VC_366) | | | | | | | | | | | Ref |
| 187 | AJ304858 | Unassigned | Unassigned | Unassigned | | Outlier | | | | | | | | | | | Ref |
| 199 | AJ564013 | Unassigned | Unassigned | Unassigned | | Outlier | | | | | | | | | | | Ref |
| 200 | AJ567472 | Unassigned | Unassigned | Unassigned | | Outlier | | | | | | | | | | | Ref |
| 206 | AJ697969 | Unassigned | Unassigned | Unassigned | | Overlap (VC_442/VC_443) | | | | | | | | | | | Ref |
| 209 | AJ842011 | Unassigned | Unassigned | Unassigned | | Outlier | | | | | | | | | | | Ref |
| 214 | AJ972879 | Unassigned | Unassigned | Unassigned | | Outlier | | | | | | | | | | | Ref |
| 225 | AM265639 | Unassigned | Unassigned | Unassigned | | Outlier | | | | | | | | | | | Ref |
| 233 | AP000400 | Unassigned | Unassigned | Unassigned | | Outlier | | | | | | | | | | | Ref |
| 247 | AP009624 | Unassigned | Unassigned | Unassigned | | Overlap (VC_177/VC_296) | | | | | | | | | | | Ref |
| 258 | AP012533 | Unassigned | Unassigned | Unassigned | | Outlier | | | | | | | | | | | Ref |
| 261 | AP012536 | Unassigned | Unassigned | Unassigned | | Overlap (VC_42/VC_57) | | | | | | | | | | | Ref |
| 262 | AP012537 | Unassigned | Unassigned | Unassigned | | Overlap (VC_42/VC_57) | | | | | | | | | | | Ref |
| 267 | AP013057 | Unassigned | Unassigned | Unassigned | | Overlap (VC_121/VC_124) | | | | | | | | | | | Ref |
| 270 | AP013540 | Unassigned | Unassigned | Unassigned | | Overlap (VC_481/VC_482) | | | | | | | | | | | Ref |
| 273 | AP013543 | Unassigned | Unassigned | Unassigned | | Overlap (VC_481/VC_482) | | | | | | | | | | | Ref |
| 279 | AP013549 | Unassigned | Unassigned | Unassigned | | Outlier | | | | | | | | | | | Ref |
| 280 | AP014629 | Unassigned | Unassigned | Unassigned | | Overlap (VC_367/VC_369) | | | | | | | | | | | Ref |
| 281 | AP014684 | Unassigned | Unassigned | Unassigned | | Outlier | | | | | | | | | | | Ref |
| 282 | AP014693 | Unassigned | Unassigned | Unassigned | | Overlap (VC_344/VC_346) | | | | | | | | | | | Ref |
| 288 | AP014927 | Unassigned | Unassigned | Unassigned | | Overlap (VC_344/VC_346) | | | | | | | | | | | Ref |
| 289 | AP017903 | Unassigned | Unassigned | Unassigned | | Outlier | | | | | | | | | | | Ref |
| 292 | AP017972 | Unassigned | Unassigned | Unassigned | | Overlap (VC_442/VC_443) | | | | | | | | | | | Ref |
| 307 | AP018486 | Unassigned | Unassigned | Unassigned | | Outlier | | | | | | | | | | | Ref |
| 315 | AP019415 | Unassigned | Unassigned | Unassigned | | Outlier | | | | | | | | | | | Ref |
| 334 | AY082070 | Unassigned | Unassigned | Unassigned | | Overlap (VC_20/VC_542) | | | | | | | | | | | Ref |

| | | | | | | |
|---|---|---|---|---|---|---|
| 335 | AY095314 | Unassigned | Unassigned | Unassigned | Outlier | Ref |
| 342 | AY129336 | Unassigned | Unassigned | Unassigned | Overlap (VC_524/VC_546) | Ref |
| 365 | AY266303 | Unassigned | Unassigned | Unassigned | Overlap (VC_444/VC_580) | Ref |
| 378 | AY357582 | Unassigned | Unassigned | Unassigned | Outlier | Ref |
| 379 | AY368235 | Unassigned | Unassigned | Unassigned | Overlap (VC_365/VC_366) | Ref |
| 380 | AY369265 | Unassigned | Unassigned | Unassigned | Overlap (VC_365/VC_366) | Ref |
| 384 | AY374414 | Unassigned | Unassigned | Unassigned | Outlier | Ref |
| 392 | AY459533 | Unassigned | Unassigned | Unassigned | Outlier | Ref |
| 393 | AY459535 | Unassigned | Unassigned | Unassigned | Outlier | Ref |
| 405 | AY576273 | Unassigned | Unassigned | Unassigned | Outlier | Ref |
| 406 | AY576796 | Unassigned | Unassigned | Unassigned | Singleton | Ref |
| 413 | AY616033 | Unassigned | Unassigned | Unassigned | Outlier | Ref |
| 414 | AY616446 | Unassigned | Unassigned | Unassigned | Outlier | Ref |
| 416 | AY657002 | Unassigned | Unassigned | Unassigned | Overlap (VC_633/VC_634) | Ref |
| 424 | AY736146 | Unassigned | Unassigned | Unassigned | Outlier | Ref |
| 427 | AY766464 | Unassigned | Unassigned | Unassigned | Outlier | Ref |
| 437 | AY894696 | Unassigned | Unassigned | Unassigned | Outlier | Ref |
| 442 | AY954948 | Unassigned | Unassigned | Unassigned | Outlier | Ref |
| 444 | AY954950 | Unassigned | Unassigned | Unassigned | Outlier | Ref |
| 452 | AY954958 | Unassigned | Unassigned | Unassigned | Outlier | Ref |
| 453 | AY954959 | Unassigned | Unassigned | Unassigned | Outlier | Ref |
| 464 | AY954970 | Unassigned | Unassigned | Unassigned | Outlier | Ref |
| 467 | AY986977 | Unassigned | Unassigned | Unassigned | Outlier | Ref |
| 469 | B2U02303 | Unassigned | Unassigned | Unassigned | Outlier | Ref |
| 470 | B4PORFSX | Unassigned | Unassigned | Unassigned | Outlier | Ref |
| 481 | BK059157 | Unassigned | Unassigned | Unassigned | Singleton | Ref |
| 482 | BK059162 | Unassigned | Unassigned | Unassigned | Singleton | Ref |
| 489 | BL2CG | Unassigned | Unassigned | Unassigned | Singleton | Ref |
| 490 | BL5GENM | Unassigned | Unassigned | Unassigned | Singleton | Ref |
| 491 | BX571876 | Unassigned | Unassigned | Unassigned | Outlier | Ref |
| 498 | CAJDKB010000002 | Unassigned | Unassigned | Unassigned | Overlap (VC_695/VC_696) | Ref |
| 504 | CAKLQB020000002 | Unassigned | Unassigned | Unassigned | Outlier | Ref |
| 518 | CP002889 | Unassigned | Unassigned | Unassigned | Outlier | Ref |
| 519 | CP003186 | Unassigned | Unassigned | Unassigned | Outlier | Ref |
| 523 | CP011103 | Unassigned | Unassigned | Unassigned | Outlier | Ref |
| 527 | CP014526 | Unassigned | Unassigned | Unassigned | Outlier | Ref |
| 528 | CP017837 | Unassigned | Unassigned | Unassigned | Outlier | Ref |
| 529 | CP017838 | Unassigned | Unassigned | Unassigned | Singleton | Ref |
| 532 | CP018841 | Unassigned | Unassigned | Unassigned | Outlier | Ref |
| 534 | CP019719 | Unassigned | Unassigned | Unassigned | Outlier | Ref |
| 537 | CP025712 | Unassigned | Unassigned | Unassigned | Outlier | Ref |
| 538 | CP025713 | Unassigned | Unassigned | Unassigned | Outlier | Ref |
| 540 | CP025887 | Unassigned | Unassigned | Unassigned | Outlier | Ref |
| 543 | CP025902 | Unassigned | Unassigned | Unassigned | Outlier | Ref |
| 544 | CP025907 | Unassigned | Unassigned | Unassigned | Outlier | Ref |
| 548 | CP038625 | Unassigned | Unassigned | Unassigned | Outlier | Ref |
| 550 | CP050061 | Unassigned | Unassigned | Unassigned | Overlap (VC_257/VC_262) | Ref |
| 552 | CP051275 | Unassigned | Unassigned | Unassigned | Outlier | Ref |
| 558 | CP052843 | Unassigned | Unassigned | Unassigned | Outlier | Ref |
| 562 | CP058906 | Unassigned | Unassigned | Unassigned | Outlier | Ref |
| 576 | CP062450 | Unassigned | Unassigned | Unassigned | Outlier | Ref |
| 583 | CP063417 | Unassigned | Unassigned | Unassigned | Outlier | Ref |
| 584 | CP063964 | Unassigned | Unassigned | Unassigned | Outlier | Ref |
| 585 | CP063968 | Unassigned | Unassigned | Unassigned | Outlier | Ref |
| 592 | CP084202 | Unassigned | Unassigned | Unassigned | Overlap (VC_737/VC_738) | Ref |
| 596 | CP103976 | Unassigned | Unassigned | Unassigned | Outlier | Ref |
| 597 | CU468217 | Unassigned | Unassigned | Unassigned | Singleton | Ref |
| 609 | DQ079869 | Unassigned | Unassigned | Unassigned | Overlap (VC_362/VC_363) | Ref |
| 617 | DQ079877 | Unassigned | Unassigned | Unassigned | Overlap (VC_361/VC_363) | Ref |
| 638 | DQ079898 | Unassigned | Unassigned | Unassigned | Overlap (VC_361/VC_363) | Ref |
| 645 | DQ079905 | Unassigned | Unassigned | Unassigned | Overlap (VC_361/VC_363) | Ref |
| 647 | DQ079907 | Unassigned | Unassigned | Unassigned | Overlap (VC_361/VC_363) | Ref |
| 658 | DQ149023 | Unassigned | Unassigned | Unassigned | Overlap (VC_435/VC_656) | Ref |
| 666 | DQ203855 | Unassigned | Unassigned | Unassigned | Outlier | Ref |
| 676 | DQ372923 | Unassigned | Unassigned | Unassigned | Outlier | Ref |
| 678 | DQ394806 | Unassigned | Unassigned | Unassigned | Overlap (VC_257/VC_262) | Ref |
| 699 | DQ453159 | Unassigned | Unassigned | Unassigned | Outlier | Ref |
| 702 | DQ499600 | Unassigned | Unassigned | Unassigned | Outlier | Ref |
| 710 | DQ535032 | Unassigned | Unassigned | Unassigned | Singleton | Ref |
| 715 | DQ831957 | Unassigned | Unassigned | Unassigned | Overlap (VC_13/VC_70) | Ref |
| 717 | DQ834250 | Unassigned | Unassigned | Unassigned | Overlap (VC_13/VC_70) | Ref |
| 719 | DQ838728 | Unassigned | Unassigned | Unassigned | Overlap (VC_257/VC_262) | Ref |
| 730 | EF071488 | Unassigned | Unassigned | Unassigned | Singleton | Ref |
| 733 | EF116926 | Unassigned | Unassigned | Unassigned | Overlap (VC_319/VC_626) | Ref |
| 739 | EF207558 | Unassigned | Unassigned | Unassigned | Overlap (VC_324/VC_539) | Ref |
| 767 | EF455602 | Unassigned | Unassigned | Unassigned | Overlap (VC_310/VC_620) | Ref |
| 774 | EF523948 | Unassigned | Unassigned | Unassigned | Outlier | Ref |
| 778 | EF579802 | Unassigned | Unassigned | Unassigned | Outlier | Ref |
| 780 | EF602154 | Unassigned | Unassigned | Unassigned | Overlap (VC_365/VC_366) | Ref |
| 797 | EU307292 | Unassigned | Unassigned | Unassigned | Outlier | Ref |
| 804 | EU409559 | Unassigned | Unassigned | Unassigned | Outlier | Ref |
| 806 | EU438902 | Unassigned | Unassigned | Unassigned | Singleton | Ref |
| 818 | EU719189 | Unassigned | Unassigned | Unassigned | Overlap (VC_140/VC_649) | Ref |
| 837 | EU822883 | Unassigned | Unassigned | Unassigned | Outlier | Ref |
| 844 | EU849489 | Unassigned | Unassigned | Unassigned | Outlier | Ref |
| 850 | EU871039 | Unassigned | Unassigned | Unassigned | Outlier | Ref |
| 852 | EU876853 | Unassigned | Unassigned | Unassigned | Outlier | Ref |
| 855 | EU982300 | Unassigned | Unassigned | Unassigned | Outlier | Ref |
| 865 | FJ174692 | Unassigned | Unassigned | Unassigned | Overlap (VC_521/VC_548) | Ref |
| 897 | FJ792813 | Unassigned | Unassigned | Unassigned | Outlier | Ref |
| 920 | FJ982340 | Unassigned | Unassigned | Unassigned | Outlier | Ref |
| 921 | FM163528 | Unassigned | Unassigned | Unassigned | Outlier | Ref |
| 935 | FN436268 | Unassigned | Unassigned | Unassigned | Singleton | Ref |
| 936 | FN582354 | Unassigned | Unassigned | Unassigned | Outlier | Ref |
| 942 | FPLJ01000048 | Unassigned | Unassigned | Unassigned | Singleton | Ref |
| 946 | FQ482085 | Unassigned | Unassigned | Unassigned | Outlier | Ref |
| 959 | FR725450 | Unassigned | Unassigned | Unassigned | Singleton | Ref |
| 988 | GQ334450 | Unassigned | Unassigned | Unassigned | Overlap (VC_436/VC_859) | Ref |
| 989 | GQ357915 | Unassigned | Unassigned | Unassigned | Outlier | Ref |
| 1009 | GQ478086 | Unassigned | Unassigned | Unassigned | Overlap (VC_606/VC_775) | Ref |
| 1010 | GQ478087 | Unassigned | Unassigned | Unassigned | Overlap (VC_606/VC_775) | Ref |
| 1011 | GQ478088 | Unassigned | Unassigned | Unassigned | Overlap (VC_431/VC_742) | Ref |
| 1014 | GQ900512 | Unassigned | Unassigned | Unassigned | Outlier | Ref |
| 1021 | GU071090 | Unassigned | Unassigned | Unassigned | Overlap (VC_436/VC_859) | Ref |
| 1027 | GU071096 | Unassigned | Unassigned | Unassigned | Overlap (VC_656/VC_661) | Ref |
| 1032 | GU071101 | Unassigned | Unassigned | Unassigned | Outlier | Ref |
| 1039 | GU071108 | Unassigned | Unassigned | Unassigned | Overlap (VC_656/VC_662/VC_664) | Ref |
| 1040 | GU075905 | Unassigned | Unassigned | Unassigned | Outlier | Ref |
| 1049 | GU233956 | Unassigned | Unassigned | Unassigned | Outlier | Ref |
| 1056 | GU323318 | Unassigned | Unassigned | Unassigned | Overlap (VC_286/VC_290/VC_491) | Ref |
| 1061 | GU396103 | Unassigned | Unassigned | Unassigned | Overlap (VC_444/VC_580) | Ref |
| 1065 | GU568037 | Unassigned | Unassigned | Unassigned | Outlier | Ref |
| 1067 | GU580940 | Unassigned | Unassigned | Unassigned | Outlier | Ref |
| 1070 | GU580943 | Unassigned | Unassigned | Unassigned | Outlier | Ref |
| 1073 | GU722197 | Unassigned | Unassigned | Unassigned | Singleton | Ref |
| 1075 | GU815339 | Unassigned | Unassigned | Unassigned | Outlier | Ref |
| 1080 | GU936714 | Unassigned | Unassigned | Unassigned | Outlier | Ref |
| 1083 | GU943031 | Unassigned | Unassigned | Unassigned | Outlier | Ref |
| 1085 | GU943068 | Unassigned | Unassigned | Unassigned | Outlier | Ref |
| 1086 | GU949551 | Unassigned | Unassigned | Unassigned | Outlier | Ref |
| 1087 | GU967410 | Unassigned | Unassigned | Unassigned | Outlier | Ref |
| 1089 | HE580238 | Unassigned | Unassigned | Unassigned | Singleton | Ref |
| 1093 | HE611333 | Unassigned | Unassigned | Unassigned | Outlier | Ref |
| 1094 | HE614281 | Unassigned | Unassigned | Unassigned | Overlap (VC_654/VC_747) | Ref |
| 1095 | HE614282 | Unassigned | Unassigned | Unassigned | Overlap (VC_654/VC_747) | Ref |
| 1097 | HE681887 | Unassigned | Unassigned | Unassigned | Singleton | Ref |
| 1139 | HG798806 | Unassigned | Unassigned | Unassigned | Overlap (VC_177/VC_296) | Ref |
| 1167 | HM114277 | Unassigned | Unassigned | Unassigned | Overlap (VC_560/VC_562/VC_563) | Ref |
| 1171 | HM144385 | Unassigned | Unassigned | Unassigned | Outlier | Ref |
| 1172 | HM144386 | Unassigned | Unassigned | Unassigned | Outlier | Ref |
| 1175 | HM151342 | Unassigned | Unassigned | Unassigned | Overlap (VC_169/VC_472) | Ref |
| 1181 | HM159959 | Unassigned | Unassigned | Unassigned | Outlier | Ref |
| 1182 | HM173637 | Unassigned | Unassigned | Unassigned | Overlap (VC_31/VC_34) | Ref |
| 1186 | HM246720 | Unassigned | Unassigned | Unassigned | Outlier | Ref |
| 1187 | HM246722 | Unassigned | Unassigned | Unassigned | Outlier | Ref |
| 1188 | HM246723 | Unassigned | Unassigned | Unassigned | Outlier | Ref |
| 1189 | HM246724 | Unassigned | Unassigned | Unassigned | Outlier | Ref |
| 1192 | HM452126 | Unassigned | Unassigned | Unassigned | Overlap (VC_444/VC_580) | Ref |
| 1196 | HM486077 | Unassigned | Unassigned | Unassigned | Outlier | Ref |
| 1201 | HM596271 | Unassigned | Unassigned | Unassigned | Outlier | Ref |
| 1285 | HQ225832 | Unassigned | Unassigned | Unassigned | Singleton | Ref |
| 1288 | HQ264138 | Unassigned | Unassigned | Unassigned | Singleton | Ref |
| 1291 | HQ316580 | Unassigned | Unassigned | Unassigned | Outlier | Ref |
| 1301 | HQ317383 | Unassigned | Unassigned | Unassigned | Overlap (VC_656/VC_663) | Ref |
| 1303 | HQ317385 | Unassigned | Unassigned | Unassigned | Overlap (VC_435/VC_656) | Ref |
| 1312 | HQ331142 | Unassigned | Unassigned | Unassigned | Overlap (VC_286/VC_290/VC_491) | Ref |
| 1316 | HQ332139 | Unassigned | Unassigned | Unassigned | Outlier | Ref |
| 1329 | HQ615693 | Unassigned | Unassigned | Unassigned | Outlier | Ref |
| 1339 | HQ634153 | Unassigned | Unassigned | Unassigned | Outlier | Ref |
| 1342 | HQ634174 | Unassigned | Unassigned | Unassigned | Outlier | Ref |
| 1346 | HQ634178 | Unassigned | Unassigned | Unassigned | Overlap (VC_656/VC_662/VC_664) | Ref |
| 1351 | HQ634191 | Unassigned | Unassigned | Unassigned | Overlap (VC_435/VC_656) | Ref |
| 1353 | HQ634193 | Unassigned | Unassigned | Unassigned | Outlier | Ref |
| 1354 | HQ634194 | Unassigned | Unassigned | Unassigned | Overlap (VC_788/VC_931) | Ref |
| 1356 | HQ634196 | Unassigned | Unassigned | Unassigned | Overlap (VC_788/VC_931) | Ref |
| 1372 | HQ641380 | Unassigned | Unassigned | Unassigned | Outlier | Ref |
| 1382 | HQ728266 | Unassigned | Unassigned | Unassigned | Outlier | Ref |
| 1390 | JF270478 | Unassigned | Unassigned | Unassigned | Outlier | Ref |
| 1391 | JF314845 | Unassigned | Unassigned | Unassigned | Outlier | Ref |
| 1392 | JF317274 | Unassigned | Unassigned | Unassigned | Outlier | Ref |
| 1393 | JF344709 | Unassigned | Unassigned | Unassigned | Outlier | Ref |
| 1394 | JF412294 | Unassigned | Unassigned | Unassigned | Outlier | Ref |

| | | | | | | |
|---|---|---|---|---|---|---|
| 1395 | JF412295 | Unassigned | Unassigned | Unassigned | Outlier | Ref |
| 1397 | JF412297 | Unassigned | Unassigned | Unassigned | Outlier | Ref |
| 1398 | JF412298 | Unassigned | Unassigned | Unassigned | Singleton | Ref |
| 1399 | JF412299 | Unassigned | Unassigned | Unassigned | Outlier | Ref |
| 1400 | JF412300 | Unassigned | Unassigned | Unassigned | Singleton | Ref |
| 1401 | JF412301 | Unassigned | Unassigned | Unassigned | Singleton | Ref |
| 1402 | JF412302 | Unassigned | Unassigned | Unassigned | Outlier | Ref |
| 1429 | JF704115 | Unassigned | Unassigned | Unassigned | Overlap (VC_524/VC_546) | Ref |
| 1454 | JF912400 | Unassigned | Unassigned | Unassigned | Overlap (VC_584/VC_586) | Ref |
| 1489 | JF974288 | Unassigned | Unassigned | Unassigned | Overlap (VC_435/VC_656) | Ref |
| 1490 | JF974289 | Unassigned | Unassigned | Unassigned | Overlap (VC_435/VC_656) | Ref |
| 1491 | JF974292 | Unassigned | Unassigned | Unassigned | Overlap (VC_656/VC_661) | Ref |
| 1492 | JF974293 | Unassigned | Unassigned | Unassigned | Overlap (VC_435/VC_656) | Ref |
| 1495 | JF974296 | Unassigned | Unassigned | Unassigned | Overlap (VC_788/VC_931) | Ref |
| 1501 | JF974315 | Unassigned | Unassigned | Unassigned | Outlier | Ref |
| 1521 | JN116825 | Unassigned | Unassigned | Unassigned | Singleton | Ref |
| 1532 | JN191664 | Unassigned | Unassigned | Unassigned | Overlap (VC_705/VC_747) | Ref |
| 1533 | JN192400 | Unassigned | Unassigned | Unassigned | Overlap (VC_13/VC_70) | Ref |
| 1534 | JN192401 | Unassigned | Unassigned | Unassigned | Overlap (VC_13/VC_70) | Ref |
| 1547 | JN371768 | Unassigned | Unassigned | Unassigned | Overlap (VC_656/VC_664) | Ref |
| 1567 | JN564907 | Unassigned | Unassigned | Unassigned | Outlier | Ref |
| 1578 | JN638751 | Unassigned | Unassigned | Unassigned | Outlier | Ref |
| 1626 | JN700519 | Unassigned | Unassigned | Unassigned | Outlier | Ref |
| 1627 | JN700520 | Unassigned | Unassigned | Unassigned | Outlier | Ref |
| 1628 | JN700521 | Unassigned | Unassigned | Unassigned | Overlap (VC_14/VC_15) | Ref |
| 1638 | JN831654 | Unassigned | Unassigned | Unassigned | Overlap (VC_227/VC_941) | Ref |
| 1643 | JN880423 | Unassigned | Unassigned | Unassigned | Singleton | Ref |
| 1645 | JN882265 | Unassigned | Unassigned | Unassigned | Outlier | Ref |
| 1660 | JN991020 | Unassigned | Unassigned | Unassigned | Overlap (VC_71/VC_74) | Ref |
| 1663 | JQ011316 | Unassigned | Unassigned | Unassigned | Outlier | Ref |
| 1669 | JQ066768 | Unassigned | Unassigned | Unassigned | Outlier | Ref |
| 1676 | JQ086369 | Unassigned | Unassigned | Unassigned | Overlap (VC_31/VC_34) | Ref |
| 1677 | JQ086370 | Unassigned | Unassigned | Unassigned | Overlap (VC_31/VC_34) | Ref |
| 1678 | JQ086371 | Unassigned | Unassigned | Unassigned | Outlier | Ref |
| 1679 | JQ086372 | Unassigned | Unassigned | Unassigned | Overlap (VC_31/VC_34) | Ref |
| 1680 | JQ086373 | Unassigned | Unassigned | Unassigned | Overlap (VC_31/VC_34) | Ref |
| 1681 | JQ086374 | Unassigned | Unassigned | Unassigned | Overlap (VC_31/VC_34) | Ref |
| 1684 | JQ086377 | Unassigned | Unassigned | Unassigned | Overlap (VC_31/VC_34) | Ref |
| 1686 | JQ177062 | Unassigned | Unassigned | Unassigned | Outlier | Ref |
| 1689 | JQ177065 | Unassigned | Unassigned | Unassigned | Outlier | Ref |
| 1690 | JQ182726 | Unassigned | Unassigned | Unassigned | Overlap (VC_31/VC_34) | Ref |
| 1691 | JQ182727 | Unassigned | Unassigned | Unassigned | Overlap (VC_31/VC_34) | Ref |
| 1694 | JQ182730 | Unassigned | Unassigned | Unassigned | Outlier | Ref |
| 1695 | JQ182731 | Unassigned | Unassigned | Unassigned | Overlap (VC_31/VC_34) | Ref |
| 1696 | JQ182732 | Unassigned | Unassigned | Unassigned | Overlap (VC_31/VC_34) | Ref |
| 1702 | JQ246028 | Unassigned | Unassigned | Unassigned | Outlier | Ref |
| 1703 | JQ267518 | Unassigned | Unassigned | Unassigned | Outlier | Ref |
| 1705 | JQ288021 | Unassigned | Unassigned | Unassigned | Outlier | Ref |
| 1711 | JQ340088 | Unassigned | Unassigned | Unassigned | Outlier | Ref |
| 1714 | JQ341413 | Unassigned | Unassigned | Unassigned | Singleton | Ref |
| 1716 | JQ362498 | Unassigned | Unassigned | Unassigned | Outlier | Ref |
| 1718 | JQ446452 | Unassigned | Unassigned | Unassigned | Outlier | Ref |
| 1724 | JQ660954 | Unassigned | Unassigned | Unassigned | Overlap (VC_20/VC_542) | Ref |
| 1735 | JQ740787 | Unassigned | Unassigned | Unassigned | Overlap (VC_203/VC_204) | Ref |
| 1736 | JQ740788 | Unassigned | Unassigned | Unassigned | Overlap (VC_203/VC_204) | Ref |
| 1737 | JQ740789 | Unassigned | Unassigned | Unassigned | Overlap (VC_203/VC_205) | Ref |
| 1738 | JQ740790 | Unassigned | Unassigned | Unassigned | Overlap (VC_203/VC_205) | Ref |
| 1739 | JQ740791 | Unassigned | Unassigned | Unassigned | Overlap (VC_203/VC_204) | Ref |
| 1740 | JQ740792 | Unassigned | Unassigned | Unassigned | Overlap (VC_203/VC_205) | Ref |
| 1741 | JQ740793 | Unassigned | Unassigned | Unassigned | Overlap (VC_203/VC_204) | Ref |
| 1742 | JQ740794 | Unassigned | Unassigned | Unassigned | Overlap (VC_203/VC_204) | Ref |
| 1743 | JQ740795 | Unassigned | Unassigned | Unassigned | Overlap (VC_203/VC_205) | Ref |
| 1744 | JQ740796 | Unassigned | Unassigned | Unassigned | Overlap (VC_203/VC_205) | Ref |
| 1745 | JQ740797 | Unassigned | Unassigned | Unassigned | Overlap (VC_203/VC_205) | Ref |
| 1746 | JQ740798 | Unassigned | Unassigned | Unassigned | Overlap (VC_203/VC_204) | Ref |
| 1747 | JQ740799 | Unassigned | Unassigned | Unassigned | Overlap (VC_203/VC_204) | Ref |
| 1748 | JQ740800 | Unassigned | Unassigned | Unassigned | Overlap (VC_203/VC_204) | Ref |
| 1749 | JQ740801 | Unassigned | Unassigned | Unassigned | Overlap (VC_203/VC_204) | Ref |
| 1750 | JQ740802 | Unassigned | Unassigned | Unassigned | Overlap (VC_203/VC_204) | Ref |
| 1751 | JQ740803 | Unassigned | Unassigned | Unassigned | Overlap (VC_203/VC_204) | Ref |
| 1752 | JQ740804 | Unassigned | Unassigned | Unassigned | Overlap (VC_203/VC_205) | Ref |
| 1753 | JQ740805 | Unassigned | Unassigned | Unassigned | Overlap (VC_203/VC_205) | Ref |
| 1754 | JQ740806 | Unassigned | Unassigned | Unassigned | Overlap (VC_203/VC_204) | Ref |
| 1755 | JQ740807 | Unassigned | Unassigned | Unassigned | Overlap (VC_203/VC_205) | Ref |
| 1756 | JQ740808 | Unassigned | Unassigned | Unassigned | Overlap (VC_203/VC_205) | Ref |
| 1757 | JQ740809 | Unassigned | Unassigned | Unassigned | Overlap (VC_203/VC_204) | Ref |
| 1758 | JQ740810 | Unassigned | Unassigned | Unassigned | Overlap (VC_203/VC_204) | Ref |
| 1759 | JQ740811 | Unassigned | Unassigned | Unassigned | Overlap (VC_203/VC_204) | Ref |
| 1760 | JQ740812 | Unassigned | Unassigned | Unassigned | Overlap (VC_203/VC_205) | Ref |
| 1761 | JQ740813 | Unassigned | Unassigned | Unassigned | Overlap (VC_203/VC_205) | Ref |
| 1762 | JQ740814 | Unassigned | Unassigned | Unassigned | Overlap (VC_203/VC_204) | Ref |
| 1769 | JQ768459 | Unassigned | Unassigned | Unassigned | Outlier | Ref |
| 1775 | JQ801337 | Unassigned | Unassigned | Unassigned | Outlier | Ref |
| 1777 | JQ806764 | Unassigned | Unassigned | Unassigned | Outlier | Ref |
| 1779 | JQ809663 | Unassigned | Unassigned | Unassigned | Outlier | Ref |
| 1781 | JQ809702 | Unassigned | Unassigned | Unassigned | Overlap (VC_524/VC_546) | Ref |
| 1786 | JQ867099 | Unassigned | Unassigned | Unassigned | Outlier | Ref |
| 1787 | JQ867100 | Unassigned | Unassigned | Unassigned | Overlap (VC_897/VC_926) | Ref |
| 1800 | JX006077 | Unassigned | Unassigned | Unassigned | Outlier | Ref |
| 1802 | JX015524 | Unassigned | Unassigned | Unassigned | Overlap (VC_227/VC_941) | Ref |
| 1832 | JX145341 | Unassigned | Unassigned | Unassigned | Overlap (VC_140/VC_649) | Ref |
| 1837 | JX181814 | Unassigned | Unassigned | Unassigned | Outlier | Ref |
| 1838 | JX181815 | Unassigned | Unassigned | Unassigned | Outlier | Ref |
| 1839 | JX181816 | Unassigned | Unassigned | Unassigned | Outlier | Ref |
| 1840 | JX181817 | Unassigned | Unassigned | Unassigned | Outlier | Ref |
| 1842 | JX181823 | Unassigned | Unassigned | Unassigned | Outlier | Ref |
| 1844 | JX181825 | Unassigned | Unassigned | Unassigned | Overlap (VC_286/VC_290/VC_491) | Ref |
| 1854 | JX195166 | Unassigned | Unassigned | Unassigned | Outlier | Ref |
| 1883 | JX409894 | Unassigned | Unassigned | Unassigned | Overlap (VC_294/VC_320) | Ref |
| 1884 | JX409895 | Unassigned | Unassigned | Unassigned | Overlap (VC_294/VC_320) | Ref |
| 1886 | JX411620 | Unassigned | Unassigned | Unassigned | Overlap (VC_521/VC_548) | Ref |
| 1912 | JX509734 | Unassigned | Unassigned | Unassigned | Outlier | Ref |
| 1916 | JX560521 | Unassigned | Unassigned | Unassigned | Outlier | Ref |
| 1942 | JX676771 | Unassigned | Unassigned | Unassigned | Outlier | Ref |
| 1950 | JX871397 | Unassigned | Unassigned | Unassigned | Outlier | Ref |
| 1963 | JX944686 | Unassigned | Unassigned | Unassigned | Outlier | Ref |
| 1980 | KC117378 | Unassigned | Unassigned | Unassigned | Outlier | Ref |
| 1984 | KC131024 | Unassigned | Unassigned | Unassigned | Overlap (VC_998/VC_1000/VC_1002) | Ref |
| 1987 | KC131130 | Unassigned | Unassigned | Unassigned | Overlap (VC_442/VC_443) | Ref |
| 1992 | KC139516 | Unassigned | Unassigned | Unassigned | Outlier | Ref |
| 2022 | KC292025 | Unassigned | Unassigned | Unassigned | Singleton | Ref |
| 2043 | KC348598 | Unassigned | Unassigned | Unassigned | Overlap (VC_318/VC_319) | Ref |
| 2044 | KC348599 | Unassigned | Unassigned | Unassigned | Overlap (VC_294/VC_383) | Ref |
| 2049 | KC348604 | Unassigned | Unassigned | Unassigned | Outlier | Ref |
| 2051 | KC413987 | Unassigned | Unassigned | Unassigned | Outlier | Ref |
| 2056 | KC460990 | Unassigned | Unassigned | Unassigned | Outlier | Ref |
| 2057 | KC462197 | Unassigned | Unassigned | Unassigned | Outlier | Ref |
| 2059 | KC465899 | Unassigned | Unassigned | Unassigned | Outlier | Ref |
| 2060 | KC465900 | Unassigned | Unassigned | Unassigned | Overlap (VC_481/VC_482) | Ref |
| 2082 | KC595516 | Unassigned | Unassigned | Unassigned | Outlier | Ref |
| 2101 | KC688701 | Unassigned | Unassigned | Unassigned | Outlier | Ref |
| 2122 | KC787101 | Unassigned | Unassigned | Unassigned | Overlap (VC_236/VC_238) | Ref |
| 2142 | KC821611 | Unassigned | Unassigned | Unassigned | Singleton | Ref |
| 2155 | KC821624 | Unassigned | Unassigned | Unassigned | Outlier | Ref |
| 2157 | KC821626 | Unassigned | Unassigned | Unassigned | Singleton | Ref |
| 2166 | KC832325 | Unassigned | Unassigned | Unassigned | Outlier | Ref |
| 2167 | KC847113 | Unassigned | Unassigned | Unassigned | Outlier | Ref |
| 2195 | KF017003 | Unassigned | Unassigned | Unassigned | Overlap (VC_524/VC_546) | Ref |
| 2196 | KF017004 | Unassigned | Unassigned | Unassigned | Overlap (VC_521/VC_549) | Ref |
| 2197 | KF017005 | Unassigned | Unassigned | Unassigned | Overlap (VC_521/VC_548) | Ref |
| 2214 | KF030445 | Unassigned | Unassigned | Unassigned | Outlier | Ref |
| 2215 | KF044309 | Unassigned | Unassigned | Unassigned | Overlap (VC_362/VC_363) | Ref |
| 2216 | KF044310 | Unassigned | Unassigned | Unassigned | Overlap (VC_362/VC_363) | Ref |
| 2223 | KF114878 | Unassigned | Unassigned | Unassigned | Singleton | Ref |
| 2226 | KF147891 | Unassigned | Unassigned | Unassigned | Outlier | Ref |
| 2227 | KF147927 | Unassigned | Unassigned | Unassigned | Overlap (VC_267/VC_606) | Ref |
| 2231 | KF156339 | Unassigned | Unassigned | Unassigned | Overlap (VC_656/VC_664) | Ref |
| 2232 | KF156340 | Unassigned | Unassigned | Unassigned | Overlap (VC_656/VC_666) | Ref |
| 2233 | KF183314 | Unassigned | Unassigned | Unassigned | Overlap (VC_267/VC_606) | Ref |
| 2234 | KF183315 | Unassigned | Unassigned | Unassigned | Overlap (VC_267/VC_606) | Ref |
| 2235 | KF188409 | Unassigned | Unassigned | Unassigned | Overlap (VC_310/VC_620) | Ref |
| 2238 | KF188458 | Unassigned | Unassigned | Unassigned | Outlier | Ref |
| 2246 | KF279414 | Unassigned | Unassigned | Unassigned | Overlap (VC_227/VC_941) | Ref |
| 2252 | KF296718 | Unassigned | Unassigned | Unassigned | Overlap (VC_705/VC_747) | Ref |
| 2253 | KF301602 | Unassigned | Unassigned | Unassigned | Outlier | Ref |
| 2264 | KF356198 | Unassigned | Unassigned | Unassigned | Outlier | Ref |
| 2280 | KF547926 | Unassigned | Unassigned | Unassigned | Singleton | Ref |
| 2297 | KF589919 | Unassigned | Unassigned | Unassigned | Overlap (VC_14/VC_15) | Ref |
| 2306 | KF626666 | Unassigned | Unassigned | Unassigned | Overlap (VC_75/VC_115) | Ref |
| 2318 | KF669656 | Unassigned | Unassigned | Unassigned | Outlier | Ref |
| 2320 | KF669658 | Unassigned | Unassigned | Unassigned | Outlier | Ref |
| 2323 | KF669661 | Unassigned | Unassigned | Unassigned | Overlap (VC_1006/VC_1007) | Ref |
| 2325 | KF669663 | Unassigned | Unassigned | Unassigned | Overlap (VC_1006/VC_1007) | Ref |
| 2346 | KF772233 | Unassigned | Unassigned | Unassigned | Outlier | Ref |
| 2347 | KF772234 | Unassigned | Unassigned | Unassigned | Outlier | Ref |
| 2353 | KF811200 | Unassigned | Unassigned | Unassigned | Overlap (VC_750/VC_751) | Ref |
| 2375 | KJ000058 | Unassigned | Unassigned | Unassigned | Overlap (VC_286/VC_290/VC_491) | Ref |
| 2378 | KJ010489 | Unassigned | Unassigned | Unassigned | Overlap (VC_695/VC_696) | Ref |
| 2381 | KJ018209 | Unassigned | Unassigned | Unassigned | Outlier | Ref |
| 2382 | KJ018210 | Unassigned | Unassigned | Unassigned | Outlier | Ref |
| 2383 | KJ018211 | Unassigned | Unassigned | Unassigned | Outlier | Ref |
| 2384 | KJ018212 | Unassigned | Unassigned | Unassigned | Outlier | Ref |

| | | | | | | |
|---|---|---|---|---|---|---|
| 2386 | KJ018214 | Unassigned | Unassigned | Unassigned | Outlier | Ref |
| 2387 | KJ019026 | Unassigned | Unassigned | Unassigned | Overlap (VC_656/VC_666) | Ref |
| 2388 | KJ019027 | Unassigned | Unassigned | Unassigned | Overlap (VC_656/VC_664) | Ref |
| 2391 | KJ019030 | Unassigned | Unassigned | Unassigned | Overlap (VC_656/VC_666) | Ref |
| 2394 | KJ019033 | Unassigned | Unassigned | Unassigned | Overlap (VC_656/VC_666) | Ref |
| 2399 | KJ019038 | Unassigned | Unassigned | Unassigned | Overlap (VC_656/VC_666) | Ref |
| 2400 | KJ019039 | Unassigned | Unassigned | Unassigned | Overlap (VC_656/VC_666) | Ref |
| 2401 | KJ019040 | Unassigned | Unassigned | Unassigned | Overlap (VC_656/VC_664) | Ref |
| 2402 | KJ019041 | Unassigned | Unassigned | Unassigned | Overlap (VC_656/VC_664) | Ref |
| 2403 | KJ019042 | Unassigned | Unassigned | Unassigned | Overlap (VC_656/VC_664) | Ref |
| 2404 | KJ019043 | Unassigned | Unassigned | Unassigned | Overlap (VC_656/VC_664) | Ref |
| 2405 | KJ019044 | Unassigned | Unassigned | Unassigned | Overlap (VC_656/VC_664) | Ref |
| 2410 | KJ019049 | Unassigned | Unassigned | Unassigned | Overlap (VC_656/VC_664) | Ref |
| 2412 | KJ019051 | Unassigned | Unassigned | Unassigned | Overlap (VC_656/VC_664) | Ref |
| 2416 | KJ019055 | Unassigned | Unassigned | Unassigned | Overlap (VC_656/VC_666) | Ref |
| 2421 | KJ019060 | Unassigned | Unassigned | Unassigned | Overlap (VC_656/VC_664) | Ref |
| 2422 | KJ019061 | Unassigned | Unassigned | Unassigned | Overlap (VC_656/VC_664) | Ref |
| 2424 | KJ019063 | Unassigned | Unassigned | Unassigned | Overlap (VC_656/VC_664) | Ref |
| 2425 | KJ019064 | Unassigned | Unassigned | Unassigned | Overlap (VC_656/VC_664) | Ref |
| 2426 | KJ019065 | Unassigned | Unassigned | Unassigned | Overlap (VC_656/VC_666) | Ref |
| 2428 | KJ019067 | Unassigned | Unassigned | Unassigned | Overlap (VC_656/VC_666) | Ref |
| 2429 | KJ019068 | Unassigned | Unassigned | Unassigned | Overlap (VC_656/VC_666) | Ref |
| 2432 | KJ019071 | Unassigned | Unassigned | Unassigned | Overlap (VC_656/VC_664) | Ref |
| 2437 | KJ019076 | Unassigned | Unassigned | Unassigned | Overlap (VC_656/VC_666) | Ref |
| 2442 | KJ019081 | Unassigned | Unassigned | Unassigned | Overlap (VC_656/VC_666) | Ref |
| 2445 | KJ019084 | Unassigned | Unassigned | Unassigned | Overlap (VC_656/VC_666) | Ref |
| 2448 | KJ019087 | Unassigned | Unassigned | Unassigned | Overlap (VC_656/VC_666) | Ref |
| 2449 | KJ019088 | Unassigned | Unassigned | Unassigned | Overlap (VC_656/VC_666) | Ref |
| 2465 | KJ019104 | Unassigned | Unassigned | Unassigned | Overlap (VC_656/VC_664) | Ref |
| 2469 | KJ019108 | Unassigned | Unassigned | Unassigned | Overlap (VC_656/VC_664) | Ref |
| 2470 | KJ019109 | Unassigned | Unassigned | Unassigned | Overlap (VC_656/VC_664) | Ref |
| 2471 | KJ019110 | Unassigned | Unassigned | Unassigned | Overlap (VC_656/VC_664) | Ref |
| 2475 | KJ019114 | Unassigned | Unassigned | Unassigned | Overlap (VC_656/VC_666) | Ref |
| 2477 | KJ019116 | Unassigned | Unassigned | Unassigned | Overlap (VC_656/VC_666) | Ref |
| 2483 | KJ019122 | Unassigned | Unassigned | Unassigned | Overlap (VC_656/VC_666) | Ref |
| 2489 | KJ019128 | Unassigned | Unassigned | Unassigned | Overlap (VC_656/VC_664) | Ref |
| 2493 | KJ019132 | Unassigned | Unassigned | Unassigned | Overlap (VC_656/VC_664) | Ref |
| 2494 | KJ019133 | Unassigned | Unassigned | Unassigned | Overlap (VC_656/VC_664) | Ref |
| 2495 | KJ019134 | Unassigned | Unassigned | Unassigned | Overlap (VC_656/VC_664) | Ref |
| 2496 | KJ019135 | Unassigned | Unassigned | Unassigned | Overlap (VC_656/VC_666) | Ref |
| 2498 | KJ019137 | Unassigned | Unassigned | Unassigned | Overlap (VC_656/VC_666) | Ref |
| 2499 | KJ019138 | Unassigned | Unassigned | Unassigned | Overlap (VC_656/VC_666) | Ref |
| 2514 | KJ019153 | Unassigned | Unassigned | Unassigned | Overlap (VC_656/VC_666) | Ref |
| 2515 | KJ019154 | Unassigned | Unassigned | Unassigned | Overlap (VC_656/VC_664) | Ref |
| 2518 | KJ019157 | Unassigned | Unassigned | Unassigned | Overlap (VC_656/VC_666) | Ref |
| 2519 | KJ019158 | Unassigned | Unassigned | Unassigned | Overlap (VC_656/VC_666) | Ref |
| 2522 | KJ019161 | Unassigned | Unassigned | Unassigned | Overlap (VC_656/VC_664) | Ref |
| 2524 | KJ019163 | Unassigned | Unassigned | Unassigned | Overlap (VC_656/VC_666) | Ref |
| 2528 | KJ024807 | Unassigned | Unassigned | Unassigned | Overlap (VC_654/VC_747) | Ref |
| 2542 | KJ101592 | Unassigned | Unassigned | Unassigned | Overlap (VC_286/VC_290/VC_491) | Ref |
| 2579 | KJ192196 | Unassigned | Unassigned | Unassigned | Outlier | Ref |
| 2580 | KJ192399 | Unassigned | Unassigned | Unassigned | Outlier | Ref |
| 2594 | KJ395778 | Unassigned | Unassigned | Unassigned | Outlier | Ref |
| 2611 | KJ473423 | Unassigned | Unassigned | Unassigned | Outlier | Ref |
| 2629 | KJ510412 | Unassigned | Unassigned | Unassigned | Overlap (VC_236/VC_238) | Ref |
| 2634 | KJ534580 | Unassigned | Unassigned | Unassigned | Singleton | Ref |
| 2640 | KJ545483 | Unassigned | Unassigned | Unassigned | Overlap (VC_129/VC_711) | Ref |
| 2644 | KJ567041 | Unassigned | Unassigned | Unassigned | Overlap (VC_524/VC_546) | Ref |
| 2651 | KJ572845 | Unassigned | Unassigned | Unassigned | Overlap (VC_129/VC_711) | Ref |
| 2695 | KJ608188 | Unassigned | Unassigned | Unassigned | Outlier | Ref |
| 2710 | KJ743987 | Unassigned | Unassigned | Unassigned | Outlier | Ref |
| 2713 | KJ749827 | Unassigned | Unassigned | Unassigned | Overlap (VC_455/VC_1028) | Ref |
| 2721 | KJ847052 | Unassigned | Unassigned | Unassigned | Overlap (VC_897/VC_926) | Ref |
| 2725 | KJ920400 | Unassigned | Unassigned | Unassigned | Overlap (VC_654/VC_747) | Ref |
| 2727 | KJ944830 | Unassigned | Unassigned | Unassigned | Outlier | Ref |
| 2732 | KM000061 | Unassigned | Unassigned | Unassigned | Outlier | Ref |
| 2736 | KM066034 | Unassigned | Unassigned | Unassigned | Overlap (VC_521/VC_548) | Ref |
| 2738 | KM083128 | Unassigned | Unassigned | Unassigned | Outlier | Ref |
| 2740 | KM091443 | Unassigned | Unassigned | Unassigned | Overlap (VC_203/VC_206) | Ref |
| 2741 | KM091444 | Unassigned | Unassigned | Unassigned | Overlap (VC_203/VC_206) | Ref |
| 2749 | KM101124 | Unassigned | Unassigned | Unassigned | Outlier | Ref |
| 2756 | KM209271 | Unassigned | Unassigned | Unassigned | Outlier | Ref |
| 2757 | KM209294 | Unassigned | Unassigned | Unassigned | Outlier | Ref |
| 2759 | KM215148 | Unassigned | Unassigned | Unassigned | Overlap (VC_521/VC_549) | Ref |
| 2770 | KM236240 | Unassigned | Unassigned | Unassigned | Overlap (VC_286/VC_290/VC_491) | Ref |
| 2801 | KM378617 | Unassigned | Unassigned | Unassigned | Overlap (VC_615/VC_754) | Ref |
| 2818 | KM514685 | Unassigned | Unassigned | Unassigned | Outlier | Ref |
| 2825 | KM589501 | Unassigned | Unassigned | Unassigned | Overlap (VC_995/VC_996) | Ref |
| 2830 | KM589506 | Unassigned | Unassigned | Unassigned | Outlier | Ref |
| 2875 | KM879221 | Unassigned | Unassigned | Unassigned | Outlier | Ref |
| 2878 | KM882824 | Unassigned | Unassigned | Unassigned | Overlap (VC_334/VC_416/VC_430) | Ref |
| 2886 | KM983327 | Unassigned | Unassigned | Unassigned | Overlap (VC_138/VC_146) | Ref |
| 2888 | KM983329 | Unassigned | Unassigned | Unassigned | Overlap (VC_138/VC_146) | Ref |
| 2891 | KM983332 | Unassigned | Unassigned | Unassigned | Outlier | Ref |
| 2903 | KP027015 | Unassigned | Unassigned | Unassigned | Outlier | Ref |
| 2915 | KP027206 | Unassigned | Unassigned | Unassigned | Overlap (VC_236/VC_238) | Ref |
| 2921 | KP037007 | Unassigned | Unassigned | Unassigned | Outlier | Ref |
| 2922 | KP054477 | Unassigned | Unassigned | Unassigned | Outlier | Ref |
| 2926 | KP063902 | Unassigned | Unassigned | Unassigned | Overlap (VC_649/VC_825) | Ref |
| 2927 | KP063903 | Unassigned | Unassigned | Unassigned | Overlap (VC_649/VC_825) | Ref |
| 2930 | KP087937 | Unassigned | Unassigned | Unassigned | Singleton | Ref |
| 2933 | KP087940 | Unassigned | Unassigned | Unassigned | Singleton | Ref |
| 2934 | KP087941 | Unassigned | Unassigned | Unassigned | Overlap (VC_199/VC_398) | Ref |
| 2937 | KP087944 | Unassigned | Unassigned | Unassigned | Outlier | Ref |
| 2939 | KP087946 | Unassigned | Unassigned | Unassigned | Overlap (VC_199/VC_399) | Ref |
| 2940 | KP087947 | Unassigned | Unassigned | Unassigned | Singleton | Ref |
| 2942 | KP087949 | Unassigned | Unassigned | Unassigned | Overlap (VC_199/VC_399) | Ref |
| 2943 | KP087950 | Unassigned | Unassigned | Unassigned | Singleton | Ref |
| 2945 | KP087952 | Unassigned | Unassigned | Unassigned | Outlier | Ref |
| 2949 | KP087956 | Unassigned | Unassigned | Unassigned | Outlier | Ref |
| 2965 | KP162168 | Unassigned | Unassigned | Unassigned | Outlier | Ref |
| 2973 | KP211958 | Unassigned | Unassigned | Unassigned | Outlier | Ref |
| 2982 | KP282676 | Unassigned | Unassigned | Unassigned | Outlier | Ref |
| 2988 | KP296792 | Unassigned | Unassigned | Unassigned | Outlier | Ref |
| 3015 | KP682381 | Unassigned | Unassigned | Unassigned | Outlier | Ref |
| 3029 | KP696447 | Unassigned | Unassigned | Unassigned | Overlap (VC_1006/VC_1007) | Ref |
| 3030 | KP696448 | Unassigned | Unassigned | Unassigned | Overlap (VC_1006/VC_1007) | Ref |
| 3031 | KP703175 | Unassigned | Unassigned | Unassigned | Singleton | Ref |
| 3037 | KP735928 | Unassigned | Unassigned | Unassigned | Overlap (VC_14/VC_15) | Ref |
| 3038 | KP774835 | Unassigned | Unassigned | Unassigned | Outlier | Ref |
| 3046 | KP792622 | Unassigned | Unassigned | Unassigned | Singleton | Ref |
| 3048 | KP793101 | Unassigned | Unassigned | Unassigned | Overlap (VC_203/VC_207) | Ref |
| 3049 | KP793102 | Unassigned | Unassigned | Unassigned | Overlap (VC_203/VC_207) | Ref |
| 3050 | KP793103 | Unassigned | Unassigned | Unassigned | Overlap (VC_203/VC_207) | Ref |
| 3053 | KP793106 | Unassigned | Unassigned | Unassigned | Overlap (VC_203/VC_207) | Ref |
| 3054 | KP793107 | Unassigned | Unassigned | Unassigned | Overlap (VC_203/VC_208) | Ref |
| 3057 | KP793110 | Unassigned | Unassigned | Unassigned | Overlap (VC_203/VC_208) | Ref |
| 3059 | KP793112 | Unassigned | Unassigned | Unassigned | Overlap (VC_203/VC_208) | Ref |
| 3061 | KP793114 | Unassigned | Unassigned | Unassigned | Overlap (VC_203/VC_206) | Ref |
| 3066 | KP793119 | Unassigned | Unassigned | Unassigned | Overlap (VC_203/VC_209) | Ref |
| 3067 | KP793120 | Unassigned | Unassigned | Unassigned | Overlap (VC_203/VC_206) | Ref |
| 3068 | KP793121 | Unassigned | Unassigned | Unassigned | Overlap (VC_203/VC_206) | Ref |
| 3069 | KP793122 | Unassigned | Unassigned | Unassigned | Overlap (VC_203/VC_206) | Ref |
| 3072 | KP793125 | Unassigned | Unassigned | Unassigned | Overlap (VC_203/VC_206) | Ref |
| 3073 | KP793126 | Unassigned | Unassigned | Unassigned | Overlap (VC_203/VC_206) | Ref |
| 3074 | KP793127 | Unassigned | Unassigned | Unassigned | Overlap (VC_203/VC_206) | Ref |
| 3075 | KP793128 | Unassigned | Unassigned | Unassigned | Overlap (VC_203/VC_206) | Ref |
| 3077 | KP793130 | Unassigned | Unassigned | Unassigned | Overlap (VC_203/VC_206) | Ref |
| 3078 | KP793131 | Unassigned | Unassigned | Unassigned | Overlap (VC_203/VC_206) | Ref |
| 3079 | KP793132 | Unassigned | Unassigned | Unassigned | Overlap (VC_203/VC_206) | Ref |
| 3080 | KP793133 | Unassigned | Unassigned | Unassigned | Overlap (VC_203/VC_207) | Ref |
| 3082 | KP793135 | Unassigned | Unassigned | Unassigned | Overlap (VC_203/VC_206) | Ref |
| 3116 | KR011061 | Unassigned | Unassigned | Unassigned | Outlier | Ref |
| 3129 | KR053194 | Unassigned | Unassigned | Unassigned | Outlier | Ref |
| 3131 | KR053196 | Unassigned | Unassigned | Unassigned | Outlier | Ref |
| 3134 | KR053199 | Unassigned | Unassigned | Unassigned | Outlier | Ref |
| 3135 | KR053200 | Unassigned | Unassigned | Unassigned | Overlap (VC_1095/VC_1096) | Ref |
| 3150 | KR072689 | Unassigned | Unassigned | Unassigned | Outlier | Ref |
| 3167 | KR091942 | Unassigned | Unassigned | Unassigned | Outlier | Ref |
| 3176 | KR093632 | Unassigned | Unassigned | Unassigned | Outlier | Ref |
| 3182 | KR093638 | Unassigned | Unassigned | Unassigned | Outlier | Ref |
| 3201 | KR131710 | Unassigned | Unassigned | Unassigned | Outlier | Ref |
| 3208 | KR153873 | Unassigned | Unassigned | Unassigned | Outlier | Ref |
| 3216 | KR296684 | Unassigned | Unassigned | Unassigned | Outlier | Ref |
| 3217 | KR296685 | Unassigned | Unassigned | Unassigned | Outlier | Ref |
| 3222 | KR296690 | Unassigned | Unassigned | Unassigned | Outlier | Ref |
| 3225 | KR296693 | Unassigned | Unassigned | Unassigned | Outlier | Ref |
| 3226 | KR296694 | Unassigned | Unassigned | Unassigned | Outlier | Ref |
| 3227 | KR296695 | Unassigned | Unassigned | Unassigned | Outlier | Ref |
| 3253 | KR704914 | Unassigned | Unassigned | Unassigned | Outlier | Ref |
| 3275 | KR905066 | Unassigned | Unassigned | Unassigned | Overlap (VC_273/VC_384) | Ref |
| 3276 | KR905067 | Unassigned | Unassigned | Unassigned | Overlap (VC_273/VC_384) | Ref |
| 3278 | KR905069 | Unassigned | Unassigned | Unassigned | Overlap (VC_273/VC_384) | Ref |
| 3279 | KR905070 | Unassigned | Unassigned | Unassigned | Overlap (VC_273/VC_384) | Ref |
| 3298 | KT001915 | Unassigned | Unassigned | Unassigned | Overlap (VC_286/VC_290/VC_491) | Ref |
| 3307 | KT021004 | Unassigned | Unassigned | Unassigned | Outlier | Ref |
| 3310 | KT070868 | Unassigned | Unassigned | Unassigned | Outlier | Ref |
| 3334 | KT184694 | Unassigned | Unassigned | Unassigned | Overlap (VC_227/VC_941) | Ref |
| 3339 | KT206225 | Unassigned | Unassigned | Unassigned | Outlier | Ref |
| 3341 | KT221033 | Unassigned | Unassigned | Unassigned | Overlap (VC_376/VC_588) | Ref |
| 3379 | KT264770 | Unassigned | Unassigned | Unassigned | Overlap (VC_1106/VC_1107) | Ref |
| 3380 | KT264771 | Unassigned | Unassigned | Unassigned | Outlier | Ref |

| | | | | | | |
|---|---|---|---|---|---|---|
| 3381 | KT264772 | Unassigned | Unassigned | Unassigned | Outlier | Ref |
| 3382 | KT264773 | Unassigned | Unassigned | Unassigned | Outlier | Ref |
| 3383 | KT264774 | Unassigned | Unassigned | Unassigned | Outlier | Ref |
| 3386 | KT264777 | Unassigned | Unassigned | Unassigned | Overlap (VC_1116/VC_1117) | Ref |
| 3387 | KT264778 | Unassigned | Unassigned | Unassigned | Overlap (VC_1116/VC_1117) | Ref |
| 3389 | KT264780 | Unassigned | Unassigned | Unassigned | Singleton | Ref |
| 3395 | KT264786 | Unassigned | Unassigned | Unassigned | Singleton | Ref |
| 3396 | KT264787 | Unassigned | Unassigned | Unassigned | Singleton | Ref |
| 3413 | KT264804 | Unassigned | Unassigned | Unassigned | Singleton | Ref |
| 3425 | KT264816 | Unassigned | Unassigned | Unassigned | Overlap (VC_1057/VC_1143) | Ref |
| 3426 | KT264817 | Unassigned | Unassigned | Unassigned | Overlap (VC_1057/VC_1143) | Ref |
| 3441 | KT264832 | Unassigned | Unassigned | Unassigned | Singleton | Ref |
| 3444 | KT264835 | Unassigned | Unassigned | Unassigned | Outlier | Ref |
| 3445 | KT266805 | Unassigned | Unassigned | Unassigned | Overlap (VC_169/VC_472) | Ref |
| 3448 | KT281789 | Unassigned | Unassigned | Unassigned | Overlap (VC_238/VC_240) | Ref |
| 3460 | KT319602 | Unassigned | Unassigned | Unassigned | Outlier | Ref |
| 3461 | KT319607 | Unassigned | Unassigned | Unassigned | Outlier | Ref |
| 3462 | KT319620 | Unassigned | Unassigned | Unassigned | Outlier | Ref |
| 3463 | KT319626 | Unassigned | Unassigned | Unassigned | Outlier | Ref |
| 3486 | KT337365 | Unassigned | Unassigned | Unassigned | Outlier | Ref |
| 3487 | KT337370 | Unassigned | Unassigned | Unassigned | Overlap (VC_334/VC_416) | Ref |
| 3488 | KT339177 | Unassigned | Unassigned | Unassigned | Outlier | Ref |
| 3503 | KT361651 | Unassigned | Unassigned | Unassigned | Overlap (VC_710/VC_713) | Ref |
| 3505 | KT361653 | Unassigned | Unassigned | Unassigned | Overlap (VC_710/VC_713) | Ref |
| 3548 | KT429161 | Unassigned | Unassigned | Unassigned | Overlap (VC_14/VC_15) | Ref |
| 3554 | KT588072 | Unassigned | Unassigned | Unassigned | Overlap (VC_695/VC_696) | Ref |
| 3561 | KT591490 | Unassigned | Unassigned | Unassigned | Overlap (VC_236/VC_238) | Ref |
| 3568 | KT630645 | Unassigned | Unassigned | Unassigned | Outlier | Ref |
| 3569 | KT630646 | Unassigned | Unassigned | Unassigned | Outlier | Ref |
| 3571 | KT630648 | Unassigned | Unassigned | Unassigned | Outlier | Ref |
| 3573 | KT716399 | Unassigned | Unassigned | Unassigned | Outlier | Ref |
| 3578 | KT724718 | Unassigned | Unassigned | Unassigned | Outlier | Ref |
| 3585 | KT780304 | Unassigned | Unassigned | Unassigned | Outlier | Ref |
| 3587 | KT803877 | Unassigned | Unassigned | Unassigned | Overlap (VC_368/VC_369) | Ref |
| 3594 | KT820175 | Unassigned | Unassigned | Unassigned | Outlier | Ref |
| 3596 | KT852574 | Unassigned | Unassigned | Unassigned | Outlier | Ref |
| 3611 | KT898133 | Unassigned | Unassigned | Unassigned | Outlier | Ref |
| 3612 | KT898134 | Unassigned | Unassigned | Unassigned | Outlier | Ref |
| 3630 | KT967075 | Unassigned | Unassigned | Unassigned | Overlap (VC_705/VC_747) | Ref |
| 3633 | KT970645 | Unassigned | Unassigned | Unassigned | Overlap (VC_654/VC_747) | Ref |
| 3634 | KT970646 | Unassigned | Unassigned | Unassigned | Overlap (VC_654/VC_747) | Ref |
| 3641 | KU052038 | Unassigned | Unassigned | Unassigned | Outlier | Ref |
| 3652 | KU160494 | Unassigned | Unassigned | Unassigned | Outlier | Ref |
| 3653 | KU160495 | Unassigned | Unassigned | Unassigned | Outlier | Ref |
| 3654 | KU160496 | Unassigned | Unassigned | Unassigned | Outlier | Ref |
| 3708 | KU238067 | Unassigned | Unassigned | Unassigned | Outlier | Ref |
| 3713 | KU252585 | Unassigned | Unassigned | Unassigned | Overlap (VC_526/VC_554) | Ref |
| 3721 | KU310944 | Unassigned | Unassigned | Unassigned | Outlier | Ref |
| 3737 | KU594605 | Unassigned | Unassigned | Unassigned | Overlap (VC_435/VC_656) | Ref |
| 3743 | KU598975 | Unassigned | Unassigned | Unassigned | Overlap (VC_13/VC_70) | Ref |
| 3759 | KU645528 | Unassigned | Unassigned | Unassigned | Outlier | Ref |
| 3764 | KU665491 | Unassigned | Unassigned | Unassigned | Outlier | Ref |
| 3770 | KU682439 | Unassigned | Unassigned | Unassigned | Outlier | Ref |
| 3782 | KU686203 | Unassigned | Unassigned | Unassigned | Overlap (VC_656/VC_662/VC_664) | Ref |
| 3789 | KU686210 | Unassigned | Unassigned | Unassigned | Outlier | Ref |
| 3801 | KU708005 | Unassigned | Unassigned | Unassigned | Singleton | Ref |
| 3824 | KU848186 | Unassigned | Unassigned | Unassigned | Overlap (VC_273/VC_384) | Ref |
| 3829 | KU867307 | Unassigned | Unassigned | Unassigned | Overlap (VC_286/VC_290/VC_491) | Ref |
| 3833 | KU873925 | Unassigned | Unassigned | Unassigned | Outlier | Ref |
| 3850 | KU892558 | Unassigned | Unassigned | Unassigned | Outlier | Ref |
| 3862 | KU935715 | Unassigned | Unassigned | Unassigned | Outlier | Ref |
| 3877 | KU963248 | Unassigned | Unassigned | Unassigned | Outlier | Ref |
| 3893 | KU977419 | Unassigned | Unassigned | Unassigned | Outlier | Ref |
| 3894 | KU977420 | Unassigned | Unassigned | Unassigned | Outlier | Ref |
| 3897 | KU984979 | Unassigned | Unassigned | Unassigned | Overlap (VC_1186/VC_1187) | Ref |
| 3898 | KU984980 | Unassigned | Unassigned | Unassigned | Overlap (VC_1186/VC_1187) | Ref |
| 3908 | KU998233 | Unassigned | Unassigned | Unassigned | Outlier | Ref |
| 3940 | KX066068 | Unassigned | Unassigned | Unassigned | Overlap (VC_71/VC_74) | Ref |
| 3942 | KX077889 | Unassigned | Unassigned | Unassigned | Overlap (VC_319/VC_790) | Ref |
| 3943 | KX077890 | Unassigned | Unassigned | Unassigned | Outlier | Ref |
| 3944 | KX077891 | Unassigned | Unassigned | Unassigned | Outlier | Ref |
| 3946 | KX077893 | Unassigned | Unassigned | Unassigned | Outlier | Ref |
| 3948 | KX077895 | Unassigned | Unassigned | Unassigned | Outlier | Ref |
| 3959 | KX119178 | Unassigned | Unassigned | Unassigned | Outlier | Ref |
| 3960 | KX119181 | Unassigned | Unassigned | Unassigned | Outlier | Ref |
| 3962 | KX119183 | Unassigned | Unassigned | Unassigned | Outlier | Ref |
| 3983 | KX130668 | Unassigned | Unassigned | Unassigned | Overlap (VC_458/VC_586) | Ref |
| 3995 | KX156762 | Unassigned | Unassigned | Unassigned | Overlap (VC_14/VC_15) | Ref |
| 3998 | KX160205 | Unassigned | Unassigned | Unassigned | Overlap (VC_257/VC_262) | Ref |
| 4000 | KX160207 | Unassigned | Unassigned | Unassigned | Overlap (VC_257/VC_262) | Ref |
| 4001 | KX160208 | Unassigned | Unassigned | Unassigned | Overlap (VC_257/VC_262) | Ref |
| 4005 | KX160212 | Unassigned | Unassigned | Unassigned | Overlap (VC_257/VC_262) | Ref |
| 4006 | KX160213 | Unassigned | Unassigned | Unassigned | Overlap (VC_257/VC_262) | Ref |
| 4007 | KX160214 | Unassigned | Unassigned | Unassigned | Overlap (VC_257/VC_262) | Ref |
| 4008 | KX160215 | Unassigned | Unassigned | Unassigned | Overlap (VC_257/VC_262) | Ref |
| 4009 | KX160216 | Unassigned | Unassigned | Unassigned | Overlap (VC_257/VC_262/VC_271) | Ref |
| 4010 | KX160217 | Unassigned | Unassigned | Unassigned | Overlap (VC_257/VC_262/VC_271) | Ref |
| 4011 | KX160218 | Unassigned | Unassigned | Unassigned | Overlap (VC_257/VC_262/VC_271) | Ref |
| 4022 | KX190832 | Unassigned | Unassigned | Unassigned | Outlier | Ref |
| 4027 | KX198612 | Unassigned | Unassigned | Unassigned | Outlier | Ref |
| 4037 | KX228399 | Unassigned | Unassigned | Unassigned | Overlap (VC_140/VC_649) | Ref |
| 4038 | KX228400 | Unassigned | Unassigned | Unassigned | Overlap (VC_140/VC_649) | Ref |
| 4391 | KX268652 | Unassigned | Unassigned | Unassigned | Outlier | Ref |
| 4398 | KX346236 | Unassigned | Unassigned | Unassigned | Overlap (VC_203/VC_209) | Ref |
| 4399 | KX346237 | Unassigned | Unassigned | Unassigned | Overlap (VC_203/VC_209) | Ref |
| 4400 | KX346238 | Unassigned | Unassigned | Unassigned | Overlap (VC_203/VC_209) | Ref |
| 4401 | KX346239 | Unassigned | Unassigned | Unassigned | Overlap (VC_203/VC_209) | Ref |
| 4402 | KX346240 | Unassigned | Unassigned | Unassigned | Overlap (VC_203/VC_209) | Ref |
| 4403 | KX346241 | Unassigned | Unassigned | Unassigned | Overlap (VC_203/VC_209) | Ref |
| 4404 | KX346242 | Unassigned | Unassigned | Unassigned | Overlap (VC_203/VC_206) | Ref |
| 4405 | KX346243 | Unassigned | Unassigned | Unassigned | Overlap (VC_203/VC_209) | Ref |
| 4406 | KX346244 | Unassigned | Unassigned | Unassigned | Overlap (VC_203/VC_209) | Ref |
| 4407 | KX346245 | Unassigned | Unassigned | Unassigned | Overlap (VC_203/VC_209) | Ref |
| 4409 | KX346247 | Unassigned | Unassigned | Unassigned | Overlap (VC_203/VC_209) | Ref |
| 4410 | KX346248 | Unassigned | Unassigned | Unassigned | Overlap (VC_203/VC_209) | Ref |
| 4411 | KX346249 | Unassigned | Unassigned | Unassigned | Overlap (VC_203/VC_206) | Ref |
| 4412 | KX346250 | Unassigned | Unassigned | Unassigned | Overlap (VC_203/VC_206) | Ref |
| 4472 | KX349285 | Unassigned | Unassigned | Unassigned | Overlap (VC_435/VC_656) | Ref |
| 4473 | KX349286 | Unassigned | Unassigned | Unassigned | Overlap (VC_435/VC_656) | Ref |
| 4474 | KX349287 | Unassigned | Unassigned | Unassigned | Overlap (VC_435/VC_656) | Ref |
| 4475 | KX349288 | Unassigned | Unassigned | Unassigned | Overlap (VC_435/VC_656) | Ref |
| 4476 | KX349289 | Unassigned | Unassigned | Unassigned | Overlap (VC_435/VC_656) | Ref |
| 4477 | KX349290 | Unassigned | Unassigned | Unassigned | Overlap (VC_435/VC_656) | Ref |
| 4485 | KX349298 | Unassigned | Unassigned | Unassigned | Overlap (VC_656/VC_661) | Ref |
| 4486 | KX349299 | Unassigned | Unassigned | Unassigned | Overlap (VC_656/VC_661) | Ref |
| 4487 | KX349300 | Unassigned | Unassigned | Unassigned | Overlap (VC_656/VC_661) | Ref |
| 4488 | KX349301 | Unassigned | Unassigned | Unassigned | Overlap (VC_656/VC_661) | Ref |
| 4489 | KX349302 | Unassigned | Unassigned | Unassigned | Overlap (VC_656/VC_661) | Ref |
| 4490 | KX349303 | Unassigned | Unassigned | Unassigned | Overlap (VC_656/VC_661) | Ref |
| 4491 | KX349304 | Unassigned | Unassigned | Unassigned | Overlap (VC_656/VC_661) | Ref |
| 4492 | KX349305 | Unassigned | Unassigned | Unassigned | Overlap (VC_656/VC_661) | Ref |
| 4493 | KX349306 | Unassigned | Unassigned | Unassigned | Overlap (VC_656/VC_661) | Ref |
| 4494 | KX349307 | Unassigned | Unassigned | Unassigned | Overlap (VC_656/VC_662/VC_664) | Ref |
| 4495 | KX349308 | Unassigned | Unassigned | Unassigned | Overlap (VC_656/VC_662/VC_664) | Ref |
| 4496 | KX349309 | Unassigned | Unassigned | Unassigned | Overlap (VC_656/VC_662/VC_664) | Ref |
| 4497 | KX349310 | Unassigned | Unassigned | Unassigned | Overlap (VC_656/VC_662/VC_664) | Ref |
| 4498 | KX349311 | Unassigned | Unassigned | Unassigned | Overlap (VC_656/VC_662/VC_664) | Ref |
| 4499 | KX349312 | Unassigned | Unassigned | Unassigned | Overlap (VC_656/VC_662/VC_664) | Ref |
| 4500 | KX349313 | Unassigned | Unassigned | Unassigned | Overlap (VC_656/VC_662/VC_664) | Ref |
| 4501 | KX349314 | Unassigned | Unassigned | Unassigned | Overlap (VC_656/VC_662/VC_664) | Ref |
| 4502 | KX349315 | Unassigned | Unassigned | Unassigned | Overlap (VC_656/VC_662/VC_664) | Ref |
| 4503 | KX349316 | Unassigned | Unassigned | Unassigned | Overlap (VC_656/VC_662/VC_664) | Ref |
| 4504 | KX349317 | Unassigned | Unassigned | Unassigned | Overlap (VC_656/VC_662/VC_664) | Ref |
| 4505 | KX349318 | Unassigned | Unassigned | Unassigned | Overlap (VC_656/VC_662/VC_664) | Ref |
| 4506 | KX349319 | Unassigned | Unassigned | Unassigned | Overlap (VC_656/VC_662/VC_664) | Ref |
| 4507 | KX349320 | Unassigned | Unassigned | Unassigned | Overlap (VC_656/VC_662/VC_664) | Ref |
| 4508 | KX349321 | Unassigned | Unassigned | Unassigned | Overlap (VC_656/VC_662/VC_664) | Ref |
| 4509 | KX349322 | Unassigned | Unassigned | Unassigned | Overlap (VC_656/VC_662/VC_664) | Ref |
| 4510 | KX349323 | Unassigned | Unassigned | Unassigned | Overlap (VC_656/VC_662/VC_664) | Ref |
| 4511 | KX349324 | Unassigned | Unassigned | Unassigned | Overlap (VC_656/VC_662/VC_664) | Ref |
| 4512 | KX349325 | Unassigned | Unassigned | Unassigned | Overlap (VC_656/VC_662/VC_664) | Ref |
| 4513 | KX349326 | Unassigned | Unassigned | Unassigned | Overlap (VC_656/VC_662/VC_664) | Ref |
| 4514 | KX349327 | Unassigned | Unassigned | Unassigned | Overlap (VC_656/VC_662/VC_664) | Ref |
| 4536 | KX379664 | Unassigned | Unassigned | Unassigned | Overlap (VC_203/VC_209) | Ref |
| 4537 | KX379665 | Unassigned | Unassigned | Unassigned | Overlap (VC_203/VC_208) | Ref |
| 4538 | KX379666 | Unassigned | Unassigned | Unassigned | Overlap (VC_203/VC_209) | Ref |
| 4541 | KX379669 | Unassigned | Unassigned | Unassigned | Overlap (VC_203/VC_209) | Ref |
| 4544 | KX379672 | Unassigned | Unassigned | Unassigned | Overlap (VC_203/VC_206) | Ref |
| 4545 | KX379673 | Unassigned | Unassigned | Unassigned | Overlap (VC_203/VC_206) | Ref |
| 4546 | KX397280 | Unassigned | Unassigned | Unassigned | Outlier | Ref |
| 4578 | KX456207 | Unassigned | Unassigned | Unassigned | Overlap (VC_263/VC_318) | Ref |
| 4583 | KX456212 | Unassigned | Unassigned | Unassigned | Overlap (VC_263/VC_318) | Ref |
| 4587 | KX458241 | Unassigned | Unassigned | Unassigned | Overlap (VC_71/VC_74) | Ref |
| 4609 | KX513876 | Unassigned | Unassigned | Unassigned | Outlier | Ref |
| 4639 | KX557277 | Unassigned | Unassigned | Unassigned | Outlier | Ref |
| 4650 | KX557288 | Unassigned | Unassigned | Unassigned | Outlier | Ref |
| 4697 | KX620748 | Unassigned | Unassigned | Unassigned | Overlap (VC_798/VC_1187) | Ref |
| 4698 | KX620749 | Unassigned | Unassigned | Unassigned | Overlap (VC_798/VC_1187) | Ref |
| 4701 | KX620752 | Unassigned | Unassigned | Unassigned | Overlap (VC_798/VC_1187) | Ref |
| 4718 | KX648391 | Unassigned | Unassigned | Unassigned | Outlier | Ref |
| 4731 | KX669658 | Unassigned | Unassigned | Unassigned | Outlier | Ref |
| 4746 | KX688047 | Unassigned | Unassigned | Unassigned | Overlap (VC_236/VC_238) | Ref |

| | | | | | | |
|---|---|---|---|---|---|---|
| 4778 | KX808132 | Unassigned | Unassigned | Unassigned | Overlap (VC_238/VC_240) | Ref |
| 4779 | KX815270 | Unassigned | Unassigned | Unassigned | Singleton | Ref |
| 4794 | KX833905 | Unassigned | Unassigned | Unassigned | Outlier | Ref |
| 4797 | KX853513 | Unassigned | Unassigned | Unassigned | Singleton | Ref |
| 4798 | KX853514 | Unassigned | Unassigned | Unassigned | Singleton | Ref |
| 4808 | KX879752 | Unassigned | Unassigned | Unassigned | Outlier | Ref |
| 4813 | KX905163 | Unassigned | Unassigned | Unassigned | Outlier | Ref |
| 4814 | KX911187 | Unassigned | Unassigned | Unassigned | Outlier | Ref |
| 4822 | KX965989 | Unassigned | Unassigned | Unassigned | Outlier | Ref |
| 4823 | KX982260 | Unassigned | Unassigned | Unassigned | Overlap (VC_897/VC_926) | Ref |
| 4843 | KY012363 | Unassigned | Unassigned | Unassigned | Overlap (VC_521/VC_548) | Ref |
| 4847 | KY038360 | Unassigned | Unassigned | Unassigned | Outlier | Ref |
| 4849 | KY053532 | Unassigned | Unassigned | Unassigned | Outlier | Ref |
| 4863 | KY065457 | Unassigned | Unassigned | Unassigned | Outlier | Ref |
| 4870 | KY065464 | Unassigned | Unassigned | Unassigned | Outlier | Ref |
| 4873 | KY065467 | Unassigned | Unassigned | Unassigned | Outlier | Ref |
| 4887 | KY065481 | Unassigned | Unassigned | Unassigned | Overlap (VC_334/VC_416) | Ref |
| 4888 | KY065482 | Unassigned | Unassigned | Unassigned | Overlap (VC_334/VC_416) | Ref |
| 4889 | KY065483 | Unassigned | Unassigned | Unassigned | Overlap (VC_334/VC_416) | Ref |
| 4890 | KY065484 | Unassigned | Unassigned | Unassigned | Overlap (VC_334/VC_416) | Ref |
| 4895 | KY065489 | Unassigned | Unassigned | Unassigned | Overlap (VC_334/VC_416) | Ref |
| 4934 | KY124276 | Unassigned | Unassigned | Unassigned | Overlap (VC_392/VC_395) | Ref |
| 4952 | KY245949 | Unassigned | Unassigned | Unassigned | Outlier | Ref |
| 4953 | KY245950 | Unassigned | Unassigned | Unassigned | Outlier | Ref |
| 4955 | KY250034 | Unassigned | Unassigned | Unassigned | Overlap (VC_455/VC_1028) | Ref |
| 4956 | KY250035 | Unassigned | Unassigned | Unassigned | Overlap (VC_392/VC_395) | Ref |
| 4966 | KY271400 | Unassigned | Unassigned | Unassigned | Outlier | Ref |
| 5000 | KY363359 | Unassigned | Unassigned | Unassigned | Overlap (VC_334/VC_416/VC_430/VC_777) | Ref |
| 5020 | KY417925 | Unassigned | Unassigned | Unassigned | Outlier | Ref |
| 5021 | KY420199 | Unassigned | Unassigned | Unassigned | Outlier | Ref |
| 5022 | KY421186 | Unassigned | Unassigned | Unassigned | Outlier | Ref |
| 5025 | KY442063 | Unassigned | Unassigned | Unassigned | Overlap (VC_97/VC_99) | Ref |
| 5034 | KY471386 | Unassigned | Unassigned | Unassigned | Overlap (VC_97/VC_99) | Ref |
| 5037 | KY499642 | Unassigned | Unassigned | Unassigned | Outlier | Ref |
| 5078 | KY606587 | Unassigned | Unassigned | Unassigned | Outlier | Ref |
| 5084 | KY619305 | Unassigned | Unassigned | Unassigned | Overlap (VC_458/VC_586) | Ref |
| 5106 | KY653116 | Unassigned | Unassigned | Unassigned | Outlier | Ref |
| 5107 | KY653117 | Unassigned | Unassigned | Unassigned | Outlier | Ref |
| 5108 | KY653118 | Unassigned | Unassigned | Unassigned | Outlier | Ref |
| 5110 | KY653120 | Unassigned | Unassigned | Unassigned | Overlap (VC_13/VC_70) | Ref |
| 5112 | KY653124 | Unassigned | Unassigned | Unassigned | Overlap (VC_14/VC_15) | Ref |
| 5114 | KY653126 | Unassigned | Unassigned | Unassigned | Overlap (VC_14/VC_15) | Ref |
| 5115 | KY653127 | Unassigned | Unassigned | Unassigned | Outlier | Ref |
| 5116 | KY653129 | Unassigned | Unassigned | Unassigned | Outlier | Ref |
| 5138 | KY695239 | Unassigned | Unassigned | Unassigned | Outlier | Ref |
| 5139 | KY695241 | Unassigned | Unassigned | Unassigned | Outlier | Ref |
| 5140 | KY697807 | Unassigned | Unassigned | Unassigned | Singleton | Ref |
| 5141 | KY702574 | Unassigned | Unassigned | Unassigned | Overlap (VC_521/VC_549) | Ref |
| 5186 | KY709687 | Unassigned | Unassigned | Unassigned | Outlier | Ref |
| 5187 | KY742649 | Unassigned | Unassigned | Unassigned | Overlap (VC_932/VC_1050) | Ref |
| 5210 | KY798216 | Unassigned | Unassigned | Unassigned | Outlier | Ref |
| 5212 | KY821088 | Unassigned | Unassigned | Unassigned | Outlier | Ref |
| 5244 | KY888885 | Unassigned | Unassigned | Unassigned | Outlier | Ref |
| 5246 | KY914485 | Unassigned | Unassigned | Unassigned | Outlier | Ref |
| 5247 | KY921761 | Unassigned | Unassigned | Unassigned | Outlier | Ref |
| 5253 | KY939598 | Unassigned | Unassigned | Unassigned | Outlier | Ref |
| 5254 | KY940711 | Unassigned | Unassigned | Unassigned | Outlier | Ref |
| 5257 | KY945241 | Unassigned | Unassigned | Unassigned | Overlap (VC_435/VC_656) | Ref |
| 5274 | KY971609 | Unassigned | Unassigned | Unassigned | Overlap (VC_596/VC_599) | Ref |
| 5277 | KY979108 | Unassigned | Unassigned | Unassigned | Overlap (VC_31/VC_34) | Ref |
| 5278 | KY979132 | Unassigned | Unassigned | Unassigned | Outlier | Ref |
| 5292 | KY981271 | Unassigned | Unassigned | Unassigned | Outlier | Ref |
| 5312 | LC121084 | Unassigned | Unassigned | Unassigned | Outlier | Ref |
| 5314 | LC208019 | Unassigned | Unassigned | Unassigned | Singleton | Ref |
| 5316 | LC216347 | Unassigned | Unassigned | Unassigned | Overlap (VC_392/VC_395) | Ref |
| 5319 | LC371242 | Unassigned | Unassigned | Unassigned | Overlap (VC_286/VC_491) | Ref |
| 5331 | LC483177 | Unassigned | Unassigned | Unassigned | Outlier | Ref |
| 5333 | LC487996 | Unassigned | Unassigned | Unassigned | Outlier | Ref |
| 5334 | LC487997 | Unassigned | Unassigned | Unassigned | Outlier | Ref |
| 5339 | LC513943 | Unassigned | Unassigned | Unassigned | Overlap (VC_695/VC_696) | Ref |
| 5354 | LC554890 | Unassigned | Unassigned | Unassigned | Outlier | Ref |
| 5355 | LC557520 | Unassigned | Unassigned | Unassigned | Outlier | Ref |
| 5356 | LC567818 | Unassigned | Unassigned | Unassigned | Overlap (VC_41/VC_42) | Ref |
| 5357 | LC567819 | Unassigned | Unassigned | Unassigned | Overlap (VC_41/VC_42) | Ref |
| 5358 | LC567820 | Unassigned | Unassigned | Unassigned | Overlap (VC_41/VC_42) | Ref |
| 5359 | LC567821 | Unassigned | Unassigned | Unassigned | Overlap (VC_41/VC_42) | Ref |
| 5360 | LC567822 | Unassigned | Unassigned | Unassigned | Overlap (VC_41/VC_42/VC_426) | Ref |
| 5361 | LC567823 | Unassigned | Unassigned | Unassigned | Overlap (VC_41/VC_42) | Ref |
| 5362 | LC567824 | Unassigned | Unassigned | Unassigned | Overlap (VC_41/VC_42) | Ref |
| 5363 | LC567825 | Unassigned | Unassigned | Unassigned | Overlap (VC_41/VC_42) | Ref |
| 5364 | LC567826 | Unassigned | Unassigned | Unassigned | Overlap (VC_41/VC_42/VC_426) | Ref |
| 5365 | LC567827 | Unassigned | Unassigned | Unassigned | Overlap (VC_41/VC_42) | Ref |
| 5366 | LC567828 | Unassigned | Unassigned | Unassigned | Overlap (VC_41/VC_42/VC_426) | Ref |
| 5367 | LC567829 | Unassigned | Unassigned | Unassigned | Overlap (VC_41/VC_42) | Ref |
| 5369 | LC567831 | Unassigned | Unassigned | Unassigned | Overlap (VC_41/VC_42) | Ref |
| 5370 | LC567832 | Unassigned | Unassigned | Unassigned | Overlap (VC_41/VC_42/VC_426) | Ref |
| 5371 | LC567833 | Unassigned | Unassigned | Unassigned | Overlap (VC_41/VC_42) | Ref |
| 5372 | LC567834 | Unassigned | Unassigned | Unassigned | Overlap (VC_41/VC_42) | Ref |
| 5373 | LC567835 | Unassigned | Unassigned | Unassigned | Overlap (VC_41/VC_42) | Ref |
| 5374 | LC567836 | Unassigned | Unassigned | Unassigned | Overlap (VC_41/VC_42/VC_426) | Ref |
| 5375 | LC567837 | Unassigned | Unassigned | Unassigned | Overlap (VC_41/VC_42) | Ref |
| 5377 | LC567839 | Unassigned | Unassigned | Unassigned | Overlap (VC_41/VC_42/VC_426) | Ref |
| 5378 | LC567840 | Unassigned | Unassigned | Unassigned | Overlap (VC_41/VC_42) | Ref |
| 5379 | LC567841 | Unassigned | Unassigned | Unassigned | Overlap (VC_41/VC_42) | Ref |
| 5384 | LC592711 | Unassigned | Unassigned | Unassigned | Overlap (VC_344/VC_346) | Ref |
| 5443 | LC644971 | Unassigned | Unassigned | Unassigned | Outlier | Ref |
| 5445 | LC644973 | Unassigned | Unassigned | Unassigned | Outlier | Ref |
| 5480 | LC645439 | Unassigned | Unassigned | Unassigned | Outlier | Ref |
| 5481 | LC645440 | Unassigned | Unassigned | Unassigned | Outlier | Ref |
| 5482 | LC645441 | Unassigned | Unassigned | Unassigned | Outlier | Ref |
| 5497 | LC667450 | Unassigned | Unassigned | Unassigned | Overlap (VC_344/VC_346) | Ref |
| 5498 | LC667451 | Unassigned | Unassigned | Unassigned | Outlier | Ref |
| 5546 | LC727876 | Unassigned | Unassigned | Unassigned | Overlap (VC_42/VC_57) | Ref |
| 5547 | LC727877 | Unassigned | Unassigned | Unassigned | Overlap (VC_42/VC_57) | Ref |
| 5548 | LC727878 | Unassigned | Unassigned | Unassigned | Overlap (VC_42/VC_57) | Ref |
| 5549 | LC727879 | Unassigned | Unassigned | Unassigned | Overlap (VC_42/VC_57) | Ref |
| 5550 | LC727880 | Unassigned | Unassigned | Unassigned | Overlap (VC_42/VC_57) | Ref |
| 5551 | LC727881 | Unassigned | Unassigned | Unassigned | Overlap (VC_42/VC_57) | Ref |
| 5552 | LC727882 | Unassigned | Unassigned | Unassigned | Overlap (VC_42/VC_57) | Ref |
| 5553 | LC727883 | Unassigned | Unassigned | Unassigned | Overlap (VC_42/VC_57) | Ref |
| 5554 | LC727884 | Unassigned | Unassigned | Unassigned | Overlap (VC_42/VC_57) | Ref |
| 5555 | LC727885 | Unassigned | Unassigned | Unassigned | Overlap (VC_42/VC_57) | Ref |
| 5556 | LC727886 | Unassigned | Unassigned | Unassigned | Overlap (VC_42/VC_57) | Ref |
| 5557 | LC727887 | Unassigned | Unassigned | Unassigned | Overlap (VC_42/VC_57) | Ref |
| 5558 | LC727888 | Unassigned | Unassigned | Unassigned | Overlap (VC_42/VC_57) | Ref |
| 5559 | LC727889 | Unassigned | Unassigned | Unassigned | Overlap (VC_42/VC_57) | Ref |
| 5560 | LC727890 | Unassigned | Unassigned | Unassigned | Overlap (VC_42/VC_57) | Ref |
| 5574 | LC727904 | Unassigned | Unassigned | Unassigned | Outlier | Ref |
| 5624 | LN681539 | Unassigned | Unassigned | Unassigned | Overlap (VC_140/VC_649) | Ref |
| 5645 | LN885237 | Unassigned | Unassigned | Unassigned | Outlier | Ref |
| 5649 | LN889991 | Unassigned | Unassigned | Unassigned | Overlap (VC_32/VC_33/VC_901) | Ref |
| 5650 | LN889992 | Unassigned | Unassigned | Unassigned | Overlap (VC_32/VC_33/VC_901) | Ref |
| 5651 | LN889993 | Unassigned | Unassigned | Unassigned | Outlier | Ref |
| 5653 | LN890663 | Unassigned | Unassigned | Unassigned | Overlap (VC_750/VC_751) | Ref |
| 5706 | LR595862 | Unassigned | Unassigned | Unassigned | Outlier | Ref |
| 5734 | LR597645 | Unassigned | Unassigned | Unassigned | Outlier | Ref |
| 5764 | LR745208 | Unassigned | Unassigned | Unassigned | Singleton | Ref |
| 5773 | LR756500 | Unassigned | Unassigned | Unassigned | Outlier | Ref |
| 5778 | LR756508 | Unassigned | Unassigned | Unassigned | Outlier | Ref |
| 5779 | LR756511 | Unassigned | Unassigned | Unassigned | Outlier | Ref |
| 5801 | LR881103 | Unassigned | Unassigned | Unassigned | Outlier | Ref |
| 5805 | LR990833 | Unassigned | Unassigned | Unassigned | Overlap (VC_695/VC_696) | Ref |
| 5806 | LR990834 | Unassigned | Unassigned | Unassigned | Outlier | Ref |
| 5808 | LR991625 | Unassigned | Unassigned | Unassigned | Overlap (VC_695/VC_696) | Ref |
| 5819 | LT614807 | Unassigned | Unassigned | Unassigned | Overlap (VC_286/VC_290/VC_491) | Ref |
| 5829 | LT960607 | Unassigned | Unassigned | Unassigned | Outlier | Ref |
| 5832 | LT960610 | Unassigned | Unassigned | Unassigned | Overlap (VC_87/VC_88) | Ref |
| 5851 | LT985570 | Unassigned | Unassigned | Unassigned | Overlap (VC_257/VC_262) | Ref |
| 5854 | LT985573 | Unassigned | Unassigned | Unassigned | Outlier | Ref |
| 5858 | LT985577 | Unassigned | Unassigned | Unassigned | Outlier | Ref |
| 5860 | LT985579 | Unassigned | Unassigned | Unassigned | Outlier | Ref |
| 5861 | LT985580 | Unassigned | Unassigned | Unassigned | Singleton | Ref |
| 5863 | LT985582 | Unassigned | Unassigned | Unassigned | Overlap (VC_271/VC_292) | Ref |
| 5866 | LT985585 | Unassigned | Unassigned | Unassigned | Overlap (VC_271/VC_292) | Ref |
| 5869 | LT985588 | Unassigned | Unassigned | Unassigned | Outlier | Ref |
| 5871 | LT985590 | Unassigned | Unassigned | Unassigned | Singleton | Ref |
| 5872 | LT985591 | Unassigned | Unassigned | Unassigned | Outlier | Ref |
| 5873 | LT985592 | Unassigned | Unassigned | Unassigned | Outlier | Ref |
| 5877 | LT992259 | Unassigned | Unassigned | Unassigned | Outlier | Ref |
| 5879 | LT993225 | Unassigned | Unassigned | Unassigned | Outlier | Ref |
| 5883 | LT996072 | Unassigned | Unassigned | Unassigned | Overlap (VC_271/VC_292) | Ref |
| 5884 | LT996073 | Unassigned | Unassigned | Unassigned | Outlier | Ref |
| 5885 | LT996074 | Unassigned | Unassigned | Unassigned | Outlier | Ref |
| 5886 | LT996075 | Unassigned | Unassigned | Unassigned | Overlap (VC_257/VC_262) | Ref |
| 5887 | LT996076 | Unassigned | Unassigned | Unassigned | Overlap (VC_261/VC_265) | Ref |
| 5888 | LT996077 | Unassigned | Unassigned | Unassigned | Outlier | Ref |
| 5889 | LT996078 | Unassigned | Unassigned | Unassigned | Overlap (VC_257/VC_262) | Ref |
| 5890 | LT996079 | Unassigned | Unassigned | Unassigned | Outlier | Ref |
| 5906 | MF001367 | Unassigned | Unassigned | Unassigned | Outlier | Ref |
| 5913 | MF036690 | Unassigned | Unassigned | Unassigned | Overlap (VC_286/VC_491) | Ref |
| 5914 | MF036691 | Unassigned | Unassigned | Unassigned | Overlap (VC_286/VC_491) | Ref |
| 5915 | MF036692 | Unassigned | Unassigned | Unassigned | Overlap (VC_286/VC_491) | Ref |

| | | | | | | |
|---|---|---|---|---|---|---|
| 5920 | MF042361 | Unassigned | Unassigned | Unassigned | Outlier | Ref |
| 5942 | MF140408 | Unassigned | Unassigned | Unassigned | Overlap (VC_236/VC_238) | Ref |
| 5945 | MF140411 | Unassigned | Unassigned | Unassigned | Overlap (VC_236/VC_238) | Ref |
| 5970 | MF141539 | Unassigned | Unassigned | Unassigned | Outlier | Ref |
| 6013 | MF186604 | Unassigned | Unassigned | Unassigned | Singleton | Ref |
| 6025 | MF190168 | Unassigned | Unassigned | Unassigned | Overlap (VC_521/VC_548) | Ref |
| 6079 | MF360958 | Unassigned | Unassigned | Unassigned | Outlier | Ref |
| 6080 | MF361639 | Unassigned | Unassigned | Unassigned | Singleton | Ref |
| 6098 | MF403008 | Unassigned | Unassigned | Unassigned | Outlier | Ref |
| 6099 | MF403009 | Unassigned | Unassigned | Unassigned | Outlier | Ref |
| 6102 | MF415410 | Unassigned | Unassigned | Unassigned | Overlap (VC_593/VC_597) | Ref |
| 6103 | MF415411 | Unassigned | Unassigned | Unassigned | Overlap (VC_593/VC_597) | Ref |
| 6105 | MF415413 | Unassigned | Unassigned | Unassigned | Overlap (VC_593/VC_597) | Ref |
| 6106 | MF417929 | Unassigned | Unassigned | Unassigned | Outlier | Ref |
| 6160 | MF459646 | Unassigned | Unassigned | Unassigned | Overlap (VC_442/VC_443) | Ref |
| 6161 | MF459647 | Unassigned | Unassigned | Unassigned | Overlap (VC_442/VC_443) | Ref |
| 6166 | MF472893 | Unassigned | Unassigned | Unassigned | Outlier | Ref |
| 6168 | MF472895 | Unassigned | Unassigned | Unassigned | Outlier | Ref |
| 6170 | MF476924 | Unassigned | Unassigned | Unassigned | Overlap (VC_593/VC_597) | Ref |
| 6172 | MF477236 | Unassigned | Unassigned | Unassigned | Outlier | Ref |
| 6181 | MF497422 | Unassigned | Unassigned | Unassigned | Overlap (VC_342/VC_1215) | Ref |
| 6235 | MF630922 | Unassigned | Unassigned | Unassigned | Outlier | Ref |
| 6243 | MF668271 | Unassigned | Unassigned | Unassigned | Overlap (VC_521/VC_548) | Ref |
| 6253 | MF668281 | Unassigned | Unassigned | Unassigned | Overlap (VC_521/VC_548) | Ref |
| 6258 | MF668286 | Unassigned | Unassigned | Unassigned | Outlier | Ref |
| 6284 | MF774687 | Unassigned | Unassigned | Unassigned | Outlier | Ref |
| 6286 | MF774689 | Unassigned | Unassigned | Unassigned | Outlier | Ref |
| 6287 | MF775669 | Unassigned | Unassigned | Unassigned | Overlap (VC_203/VC_211) | Ref |
| 6288 | MF775670 | Unassigned | Unassigned | Unassigned | Overlap (VC_203/VC_211) | Ref |
| 6289 | MF775671 | Unassigned | Unassigned | Unassigned | Overlap (VC_203/VC_211) | Ref |
| 6290 | MF775672 | Unassigned | Unassigned | Unassigned | Overlap (VC_203/VC_211) | Ref |
| 6291 | MF775673 | Unassigned | Unassigned | Unassigned | Overlap (VC_203/VC_211) | Ref |
| 6292 | MF775674 | Unassigned | Unassigned | Unassigned | Overlap (VC_203/VC_211) | Ref |
| 6293 | MF775675 | Unassigned | Unassigned | Unassigned | Overlap (VC_203/VC_211) | Ref |
| 6294 | MF775676 | Unassigned | Unassigned | Unassigned | Overlap (VC_203/VC_211) | Ref |
| 6295 | MF775677 | Unassigned | Unassigned | Unassigned | Overlap (VC_203/VC_211) | Ref |
| 6296 | MF775678 | Unassigned | Unassigned | Unassigned | Overlap (VC_203/VC_211) | Ref |
| 6297 | MF775679 | Unassigned | Unassigned | Unassigned | Overlap (VC_203/VC_211) | Ref |
| 6298 | MF775680 | Unassigned | Unassigned | Unassigned | Overlap (VC_203/VC_211) | Ref |
| 6299 | MF775681 | Unassigned | Unassigned | Unassigned | Overlap (VC_203/VC_211) | Ref |
| 6300 | MF775682 | Unassigned | Unassigned | Unassigned | Overlap (VC_203/VC_211) | Ref |
| 6301 | MF775683 | Unassigned | Unassigned | Unassigned | Overlap (VC_203/VC_211) | Ref |
| 6302 | MF775684 | Unassigned | Unassigned | Unassigned | Overlap (VC_203/VC_211) | Ref |
| 6303 | MF775685 | Unassigned | Unassigned | Unassigned | Overlap (VC_203/VC_211) | Ref |
| 6304 | MF775686 | Unassigned | Unassigned | Unassigned | Overlap (VC_203/VC_211) | Ref |
| 6305 | MF775687 | Unassigned | Unassigned | Unassigned | Overlap (VC_203/VC_211) | Ref |
| 6306 | MF775688 | Unassigned | Unassigned | Unassigned | Overlap (VC_203/VC_211) | Ref |
| 6307 | MF775689 | Unassigned | Unassigned | Unassigned | Overlap (VC_203/VC_211) | Ref |
| 6308 | MF775690 | Unassigned | Unassigned | Unassigned | Overlap (VC_203/VC_211) | Ref |
| 6309 | MF775691 | Unassigned | Unassigned | Unassigned | Overlap (VC_203/VC_211) | Ref |
| 6310 | MF775692 | Unassigned | Unassigned | Unassigned | Overlap (VC_203/VC_211) | Ref |
| 6311 | MF775693 | Unassigned | Unassigned | Unassigned | Overlap (VC_203/VC_211) | Ref |
| 6312 | MF775694 | Unassigned | Unassigned | Unassigned | Overlap (VC_203/VC_211) | Ref |
| 6313 | MF775695 | Unassigned | Unassigned | Unassigned | Overlap (VC_203/VC_211) | Ref |
| 6314 | MF775696 | Unassigned | Unassigned | Unassigned | Overlap (VC_203/VC_211) | Ref |
| 6315 | MF775697 | Unassigned | Unassigned | Unassigned | Overlap (VC_203/VC_211) | Ref |
| 6316 | MF775698 | Unassigned | Unassigned | Unassigned | Overlap (VC_203/VC_211) | Ref |
| 6317 | MF775699 | Unassigned | Unassigned | Unassigned | Overlap (VC_203/VC_211) | Ref |
| 6318 | MF775700 | Unassigned | Unassigned | Unassigned | Overlap (VC_203/VC_211) | Ref |
| 6319 | MF775701 | Unassigned | Unassigned | Unassigned | Overlap (VC_203/VC_211) | Ref |
| 6320 | MF775702 | Unassigned | Unassigned | Unassigned | Overlap (VC_203/VC_211) | Ref |
| 6329 | MF893271 | Unassigned | Unassigned | Unassigned | Outlier | Ref |
| 6331 | MF893341 | Unassigned | Unassigned | Unassigned | Outlier | Ref |
| 6340 | MF919498 | Unassigned | Unassigned | Unassigned | Outlier | Ref |
| 6344 | MF919502 | Unassigned | Unassigned | Unassigned | Overlap (VC_524/VC_546) | Ref |
| 6360 | MF919518 | Unassigned | Unassigned | Unassigned | Overlap (VC_238/VC_240) | Ref |
| 6385 | MF939898 | Unassigned | Unassigned | Unassigned | Outlier | Ref |
| 6386 | MF957259 | Unassigned | Unassigned | Unassigned | Overlap (VC_286/VC_290/VC_491) | Ref |
| 6387 | MF959998 | Unassigned | Unassigned | Unassigned | Outlier | Ref |
| 6392 | MF975637 | Unassigned | Unassigned | Unassigned | Overlap (VC_872/VC_873) | Ref |
| 6395 | MF975721 | Unassigned | Unassigned | Unassigned | Outlier | Ref |
| 6396 | MF979559 | Unassigned | Unassigned | Unassigned | Overlap (VC_481/VC_482/VC_484) | Ref |
| 6401 | MF979564 | Unassigned | Unassigned | Unassigned | Outlier | Ref |
| 6402 | MF988720 | Unassigned | Unassigned | Unassigned | Outlier | Ref |
| 6407 | MG009575 | Unassigned | Unassigned | Unassigned | Outlier | Ref |
| 6409 | MG018926 | Unassigned | Unassigned | Unassigned | Outlier | Ref |
| 6429 | MG030347 | Unassigned | Unassigned | Unassigned | Overlap (VC_194/VC_195) | Ref |
| 6434 | MG065637 | Unassigned | Unassigned | Unassigned | Outlier | Ref |
| 6435 | MG065638 | Unassigned | Unassigned | Unassigned | Outlier | Ref |
| 6448 | MG065652 | Unassigned | Unassigned | Unassigned | Outlier | Ref |
| 6463 | MG065668 | Unassigned | Unassigned | Unassigned | Outlier | Ref |
| 6471 | MG065676 | Unassigned | Unassigned | Unassigned | Outlier | Ref |
| 6477 | MG065684 | Unassigned | Unassigned | Unassigned | Outlier | Ref |
| 6478 | MG065685 | Unassigned | Unassigned | Unassigned | Outlier | Ref |
| 6506 | MG159787 | Unassigned | Unassigned | Unassigned | Outlier | Ref |
| 6509 | MG197996 | Unassigned | Unassigned | Unassigned | Outlier | Ref |
| 6525 | MG214783 | Unassigned | Unassigned | Unassigned | Outlier | Ref |
| 6527 | MG250483 | Unassigned | Unassigned | Unassigned | Overlap (VC_444/VC_580) | Ref |
| 6528 | MG250484 | Unassigned | Unassigned | Unassigned | Overlap (VC_286/VC_290/VC_491) | Ref |
| 6531 | MG251388 | Unassigned | Unassigned | Unassigned | Outlier | Ref |
| 6554 | MG366114 | Unassigned | Unassigned | Unassigned | Outlier | Ref |
| 6564 | MG450654 | Unassigned | Unassigned | Unassigned | Outlier | Ref |
| 6586 | MG575420 | Unassigned | Unassigned | Unassigned | Overlap (VC_932/VC_1050) | Ref |
| 6589 | MG584725 | Unassigned | Unassigned | Unassigned | Outlier | Ref |
| 6617 | MG592413 | Unassigned | Unassigned | Unassigned | Outlier | Ref |
| 6634 | MG592433 | Unassigned | Unassigned | Unassigned | Outlier | Ref |
| 6650 | MG592452 | Unassigned | Unassigned | Unassigned | Overlap (VC_644/VC_946) | Ref |
| 6684 | MG592491 | Unassigned | Unassigned | Unassigned | Overlap (VC_615/VC_754) | Ref |
| 6691 | MG592498 | Unassigned | Unassigned | Unassigned | Outlier | Ref |
| 6706 | MG592515 | Unassigned | Unassigned | Unassigned | Outlier | Ref |
| 6728 | MG592537 | Unassigned | Unassigned | Unassigned | Outlier | Ref |
| 6739 | MG592548 | Unassigned | Unassigned | Unassigned | Outlier | Ref |
| 6765 | MG592574 | Unassigned | Unassigned | Unassigned | Outlier | Ref |
| 6766 | MG592575 | Unassigned | Unassigned | Unassigned | Overlap (VC_45/VC_47) | Ref |
| 6768 | MG592577 | Unassigned | Unassigned | Unassigned | Overlap (VC_615/VC_754) | Ref |
| 6771 | MG592580 | Unassigned | Unassigned | Unassigned | Overlap (VC_644/VC_946) | Ref |
| 6806 | MG592615 | Unassigned | Unassigned | Unassigned | Outlier | Ref |
| 6832 | MG592643 | Unassigned | Unassigned | Unassigned | Outlier | Ref |
| 6855 | MG592667 | Unassigned | Unassigned | Unassigned | Overlap (VC_615/VC_754) | Ref |
| 6862 | MG593801 | Unassigned | Unassigned | Unassigned | Outlier | Ref |
| 6870 | MG603697 | Unassigned | Unassigned | Unassigned | Outlier | Ref |
| 6874 | MG641181 | Unassigned | Unassigned | Unassigned | Singleton | Ref |
| 6875 | MG641182 | Unassigned | Unassigned | Unassigned | Outlier | Ref |
| 6880 | MG641187 | Unassigned | Unassigned | Unassigned | Overlap (VC_1078/VC_1079) | Ref |
| 6882 | MG641885 | Unassigned | Unassigned | Unassigned | Overlap (VC_463/VC_609) | Ref |
| 6890 | MG652450 | Unassigned | Unassigned | Unassigned | Overlap (VC_71/VC_72) | Ref |
| 6909 | MG696115 | Unassigned | Unassigned | Unassigned | Outlier | Ref |
| 6915 | MG710484 | Unassigned | Unassigned | Unassigned | Overlap (VC_705/VC_747) | Ref |
| 6925 | MG711516 | Unassigned | Unassigned | Unassigned | Overlap (VC_481/VC_482/VC_484) | Ref |
| 6926 | MG720308 | Unassigned | Unassigned | Unassigned | Overlap (VC_442/VC_443) | Ref |
| 6931 | MG727697 | Unassigned | Unassigned | Unassigned | Outlier | Ref |
| 6937 | MG732930 | Unassigned | Unassigned | Unassigned | Outlier | Ref |
| 6940 | MG746602 | Unassigned | Unassigned | Unassigned | Overlap (VC_491/VC_492) | Ref |
| 6945 | MG752970 | Unassigned | Unassigned | Unassigned | Outlier | Ref |
| 6973 | MG770213 | Unassigned | Unassigned | Unassigned | Overlap (VC_521/VC_548) | Ref |
| 6983 | MG775259 | Unassigned | Unassigned | Unassigned | Outlier | Ref |
| 6985 | MG775261 | Unassigned | Unassigned | Unassigned | Overlap (VC_71/VC_74) | Ref |
| 6986 | MG779474 | Unassigned | Unassigned | Unassigned | Overlap (VC_203/VC_206) | Ref |
| 7019 | MG835450 | Unassigned | Unassigned | Unassigned | Singleton | Ref |
| 7021 | MG835858 | Unassigned | Unassigned | Unassigned | Overlap (VC_593/VC_597) | Ref |
| 7027 | MG839019 | Unassigned | Unassigned | Unassigned | Overlap (VC_1241/VC_1242) | Ref |
| 7032 | MG839024 | Unassigned | Unassigned | Unassigned | Overlap (VC_1241/VC_1242) | Ref |
| 7033 | MG839025 | Unassigned | Unassigned | Unassigned | Overlap (VC_1241/VC_1242) | Ref |
| 7038 | MG839030 | Unassigned | Unassigned | Unassigned | Overlap (VC_1241/VC_1242) | Ref |
| 7039 | MG845393 | Unassigned | Unassigned | Unassigned | Overlap (VC_526/VC_554) | Ref |
| 7040 | MG845683 | Unassigned | Unassigned | Unassigned | Outlier | Ref |
| 7062 | MG883711 | Unassigned | Unassigned | Unassigned | Overlap (VC_1106/VC_1107) | Ref |
| 7071 | MG883721 | Unassigned | Unassigned | Unassigned | Singleton | Ref |
| 7131 | MG945132 | Unassigned | Unassigned | Unassigned | Overlap (VC_1054/VC_1055) | Ref |
| 7132 | MG945133 | Unassigned | Unassigned | Unassigned | Overlap (VC_1255/VC_1256) | Ref |
| 7133 | MG945134 | Unassigned | Unassigned | Unassigned | Overlap (VC_1106/VC_1107) | Ref |
| 7136 | MG945137 | Unassigned | Unassigned | Unassigned | Singleton | Ref |
| 7144 | MG945155 | Unassigned | Unassigned | Unassigned | Outlier | Ref |
| 7145 | MG945156 | Unassigned | Unassigned | Unassigned | Overlap (VC_1056/VC_1057/VC_1061) | Ref |
| 7159 | MG945180 | Unassigned | Unassigned | Unassigned | Singleton | Ref |
| 7161 | MG945182 | Unassigned | Unassigned | Unassigned | Singleton | Ref |
| 7163 | MG945184 | Unassigned | Unassigned | Unassigned | Singleton | Ref |
| 7166 | MG945187 | Unassigned | Unassigned | Unassigned | Overlap (VC_1265/VC_1266) | Ref |
| 7170 | MG945191 | Unassigned | Unassigned | Unassigned | Outlier | Ref |
| 7185 | MG945216 | Unassigned | Unassigned | Unassigned | Singleton | Ref |
| 7198 | MG945239 | Unassigned | Unassigned | Unassigned | Outlier | Ref |
| 7199 | MG945240 | Unassigned | Unassigned | Unassigned | Outlier | Ref |
| 7202 | MG945253 | Unassigned | Unassigned | Unassigned | Outlier | Ref |
| 7208 | MG945259 | Unassigned | Unassigned | Unassigned | Singleton | Ref |
| 7211 | MG945262 | Unassigned | Unassigned | Unassigned | Singleton | Ref |
| 7213 | MG945264 | Unassigned | Unassigned | Unassigned | Outlier | Ref |
| 7214 | MG945265 | Unassigned | Unassigned | Unassigned | Overlap (VC_1054/VC_1056/VC_1057/VC_1061) | Ref |
| 7215 | MG945266 | Unassigned | Unassigned | Unassigned | Singleton | Ref |
| 7218 | MG945277 | Unassigned | Unassigned | Unassigned | Singleton | Ref |
| 7222 | MG945281 | Unassigned | Unassigned | Unassigned | Outlier | Ref |
| 7224 | MG945283 | Unassigned | Unassigned | Unassigned | Overlap (VC_1130/VC_1132/VC_1249) | Ref |
| 7228 | MG945306 | Unassigned | Unassigned | Unassigned | Outlier | Ref |

| | | | | | | |
|---|---|---|---|---|---|---|
| 7229 | MG945307 | Unassigned | Unassigned | Unassigned | Outlier | Ref |
| 7230 | MG945308 | Unassigned | Unassigned | Unassigned | Singleton | Ref |
| 7232 | MG945310 | Unassigned | Unassigned | Unassigned | Overlap (VC_1126/VC_1131) | Ref |
| 7233 | MG945311 | Unassigned | Unassigned | Unassigned | Overlap (VC_1126/VC_1131) | Ref |
| 7234 | MG945312 | Unassigned | Unassigned | Unassigned | Overlap (VC_1126/VC_1131) | Ref |
| 7235 | MG945313 | Unassigned | Unassigned | Unassigned | Overlap (VC_199/VC_398) | Ref |
| 7237 | MG945315 | Unassigned | Unassigned | Unassigned | Outlier | Ref |
| 7239 | MG945317 | Unassigned | Unassigned | Unassigned | Outlier | Ref |
| 7243 | MG945321 | Unassigned | Unassigned | Unassigned | Overlap (VC_1265/VC_1266) | Ref |
| 7244 | MG945322 | Unassigned | Unassigned | Unassigned | Overlap (VC_199/VC_398) | Ref |
| 7248 | MG945326 | Unassigned | Unassigned | Unassigned | Singleton | Ref |
| 7262 | MG945340 | Unassigned | Unassigned | Unassigned | Outlier | Ref |
| 7264 | MG945342 | Unassigned | Unassigned | Unassigned | Outlier | Ref |
| 7275 | MG945353 | Unassigned | Unassigned | Unassigned | Overlap (VC_1265/VC_1266) | Ref |
| 7278 | MG945356 | Unassigned | Unassigned | Unassigned | Outlier | Ref |
| 7280 | MG945358 | Unassigned | Unassigned | Unassigned | Singleton | Ref |
| 7281 | MG945359 | Unassigned | Unassigned | Unassigned | Overlap (VC_1106/VC_1108) | Ref |
| 7306 | MG945399 | Unassigned | Unassigned | Unassigned | Overlap (VC_1106/VC_1107) | Ref |
| 7307 | MG945400 | Unassigned | Unassigned | Unassigned | Singleton | Ref |
| 7313 | MG945406 | Unassigned | Unassigned | Unassigned | Outlier | Ref |
| 7329 | MG945422 | Unassigned | Unassigned | Unassigned | Singleton | Ref |
| 7330 | MG945423 | Unassigned | Unassigned | Unassigned | Outlier | Ref |
| 7335 | MG945428 | Unassigned | Unassigned | Unassigned | Outlier | Ref |
| 7338 | MG945431 | Unassigned | Unassigned | Unassigned | Overlap (VC_1106/VC_1108) | Ref |
| 7343 | MG945436 | Unassigned | Unassigned | Unassigned | Singleton | Ref |
| 7347 | MG945440 | Unassigned | Unassigned | Unassigned | Outlier | Ref |
| 7349 | MG945442 | Unassigned | Unassigned | Unassigned | Outlier | Ref |
| 7350 | MG945443 | Unassigned | Unassigned | Unassigned | Singleton | Ref |
| 7353 | MG945446 | Unassigned | Unassigned | Unassigned | Outlier | Ref |
| 7357 | MG945450 | Unassigned | Unassigned | Unassigned | Overlap (VC_1056/VC_1057/VC_1061) | Ref |
| 7358 | MG945451 | Unassigned | Unassigned | Unassigned | Singleton | Ref |
| 7361 | MG945454 | Unassigned | Unassigned | Unassigned | Outlier | Ref |
| 7373 | MG945466 | Unassigned | Unassigned | Unassigned | Outlier | Ref |
| 7378 | MG945471 | Unassigned | Unassigned | Unassigned | Singleton | Ref |
| 7381 | MG945474 | Unassigned | Unassigned | Unassigned | Overlap (VC_199/VC_399/VC_400) | Ref |
| 7386 | MG945479 | Unassigned | Unassigned | Unassigned | Overlap (VC_1055/VC_1056/VC_1057) | Ref |
| 7391 | MG945484 | Unassigned | Unassigned | Unassigned | Outlier | Ref |
| 7398 | MG945491 | Unassigned | Unassigned | Unassigned | Overlap (VC_1055/VC_1056/VC_1057) | Ref |
| 7400 | MG945493 | Unassigned | Unassigned | Unassigned | Outlier | Ref |
| 7404 | MG945497 | Unassigned | Unassigned | Unassigned | Outlier | Ref |
| 7410 | MG945503 | Unassigned | Unassigned | Unassigned | Outlier | Ref |
| 7417 | MG945510 | Unassigned | Unassigned | Unassigned | Singleton | Ref |
| 7420 | MG945513 | Unassigned | Unassigned | Unassigned | Outlier | Ref |
| 7423 | MG945516 | Unassigned | Unassigned | Unassigned | Singleton | Ref |
| 7428 | MG945521 | Unassigned | Unassigned | Unassigned | Singleton | Ref |
| 7429 | MG945522 | Unassigned | Unassigned | Unassigned | Overlap (VC_1255/VC_1256) | Ref |
| 7434 | MG945527 | Unassigned | Unassigned | Unassigned | Singleton | Ref |
| 7435 | MG945528 | Unassigned | Unassigned | Unassigned | Overlap (VC_1055/VC_1057) | Ref |
| 7437 | MG945530 | Unassigned | Unassigned | Unassigned | Outlier | Ref |
| 7438 | MG945531 | Unassigned | Unassigned | Unassigned | Outlier | Ref |
| 7439 | MG945532 | Unassigned | Unassigned | Unassigned | Singleton | Ref |
| 7442 | MG945535 | Unassigned | Unassigned | Unassigned | Outlier | Ref |
| 7443 | MG945536 | Unassigned | Unassigned | Unassigned | Outlier | Ref |
| 7444 | MG945537 | Unassigned | Unassigned | Unassigned | Singleton | Ref |
| 7450 | MG945543 | Unassigned | Unassigned | Unassigned | Overlap (VC_1130/VC_1132/VC_1249) | Ref |
| 7452 | MG945545 | Unassigned | Unassigned | Unassigned | Overlap (VC_1130/VC_1135) | Ref |
| 7454 | MG945547 | Unassigned | Unassigned | Unassigned | Singleton | Ref |
| 7459 | MG945552 | Unassigned | Unassigned | Unassigned | Overlap (VC_1114/VC_1136/VC_1318) | Ref |
| 7460 | MG945553 | Unassigned | Unassigned | Unassigned | Outlier | Ref |
| 7462 | MG945555 | Unassigned | Unassigned | Unassigned | Singleton | Ref |
| 7465 | MG945558 | Unassigned | Unassigned | Unassigned | Overlap (VC_1192/VC_1196) | Ref |
| 7484 | MG945577 | Unassigned | Unassigned | Unassigned | Overlap (VC_1120/VC_1121) | Ref |
| 7488 | MG945581 | Unassigned | Unassigned | Unassigned | Overlap (VC_1192/VC_1196) | Ref |
| 7490 | MG945583 | Unassigned | Unassigned | Unassigned | Overlap (VC_1055/VC_1056/VC_1057) | Ref |
| 7495 | MG945588 | Unassigned | Unassigned | Unassigned | Outlier | Ref |
| 7497 | MG945590 | Unassigned | Unassigned | Unassigned | Overlap (VC_1318/VC_1319) | Ref |
| 7499 | MG945592 | Unassigned | Unassigned | Unassigned | Outlier | Ref |
| 7500 | MG945593 | Unassigned | Unassigned | Unassigned | Outlier | Ref |
| 7502 | MG945595 | Unassigned | Unassigned | Unassigned | Singleton | Ref |
| 7505 | MG945598 | Unassigned | Unassigned | Unassigned | Outlier | Ref |
| 7506 | MG945599 | Unassigned | Unassigned | Unassigned | Overlap (VC_1056/VC_1057/VC_1061) | Ref |
| 7507 | MG945600 | Unassigned | Unassigned | Unassigned | Outlier | Ref |
| 7508 | MG945601 | Unassigned | Unassigned | Unassigned | Overlap (VC_1056/VC_1057/VC_1061/VC_1142) | Ref |
| 7510 | MG945603 | Unassigned | Unassigned | Unassigned | Overlap (VC_1056/VC_1057) | Ref |
| 7511 | MG945604 | Unassigned | Unassigned | Unassigned | Singleton | Ref |
| 7513 | MG945606 | Unassigned | Unassigned | Unassigned | Outlier | Ref |
| 7514 | MG945607 | Unassigned | Unassigned | Unassigned | Overlap (VC_1130/VC_1135) | Ref |
| 7515 | MG945608 | Unassigned | Unassigned | Unassigned | Singleton | Ref |
| 7517 | MG945610 | Unassigned | Unassigned | Unassigned | Singleton | Ref |
| 7531 | MG945624 | Unassigned | Unassigned | Unassigned | Outlier | Ref |
| 7533 | MG945626 | Unassigned | Unassigned | Unassigned | Outlier | Ref |
| 7534 | MG945627 | Unassigned | Unassigned | Unassigned | Outlier | Ref |
| 7537 | MG945630 | Unassigned | Unassigned | Unassigned | Singleton | Ref |
| 7543 | MG945636 | Unassigned | Unassigned | Unassigned | Singleton | Ref |
| 7545 | MG945638 | Unassigned | Unassigned | Unassigned | Singleton | Ref |
| 7559 | MG945666 | Unassigned | Unassigned | Unassigned | Outlier | Ref |
| 7565 | MG945672 | Unassigned | Unassigned | Unassigned | Overlap (VC_1126/VC_1131) | Ref |
| 7568 | MG945675 | Unassigned | Unassigned | Unassigned | Outlier | Ref |
| 7572 | MG945679 | Unassigned | Unassigned | Unassigned | Overlap (VC_1054/VC_1055) | Ref |
| 7577 | MG945684 | Unassigned | Unassigned | Unassigned | Outlier | Ref |
| 7581 | MG945688 | Unassigned | Unassigned | Unassigned | Outlier | Ref |
| 7588 | MG945695 | Unassigned | Unassigned | Unassigned | Singleton | Ref |
| 7593 | MG945700 | Unassigned | Unassigned | Unassigned | Overlap (VC_1106/VC_1108) | Ref |
| 7594 | MG945701 | Unassigned | Unassigned | Unassigned | Overlap (VC_1054/VC_1055) | Ref |
| 7600 | MG945707 | Unassigned | Unassigned | Unassigned | Overlap (VC_1106/VC_1107) | Ref |
| 7602 | MG945709 | Unassigned | Unassigned | Unassigned | Singleton | Ref |
| 7604 | MG945711 | Unassigned | Unassigned | Unassigned | Overlap (VC_1055/VC_1057) | Ref |
| 7607 | MG945714 | Unassigned | Unassigned | Unassigned | Singleton | Ref |
| 7608 | MG945715 | Unassigned | Unassigned | Unassigned | Outlier | Ref |
| 7613 | MG945720 | Unassigned | Unassigned | Unassigned | Overlap (VC_1114/VC_1136/VC_1318/VC_1319) | Ref |
| 7615 | MG945722 | Unassigned | Unassigned | Unassigned | Outlier | Ref |
| 7625 | MG945732 | Unassigned | Unassigned | Unassigned | Overlap (VC_1255/VC_1256) | Ref |
| 7630 | MG945737 | Unassigned | Unassigned | Unassigned | Outlier | Ref |
| 7642 | MG945749 | Unassigned | Unassigned | Unassigned | Outlier | Ref |
| 7655 | MG945762 | Unassigned | Unassigned | Unassigned | Singleton | Ref |
| 7656 | MG945763 | Unassigned | Unassigned | Unassigned | Singleton | Ref |
| 7665 | MG945772 | Unassigned | Unassigned | Unassigned | Singleton | Ref |
| 7666 | MG945773 | Unassigned | Unassigned | Unassigned | Singleton | Ref |
| 7670 | MG945777 | Unassigned | Unassigned | Unassigned | Overlap (VC_1056/VC_1057/VC_1061/VC_1142) | Ref |
| 7672 | MG945779 | Unassigned | Unassigned | Unassigned | Singleton | Ref |
| 7726 | MG945833 | Unassigned | Unassigned | Unassigned | Singleton | Ref |
| 7727 | MG945834 | Unassigned | Unassigned | Unassigned | Overlap (VC_406/VC_1250) | Ref |
| 7734 | MG945841 | Unassigned | Unassigned | Unassigned | Singleton | Ref |
| 7737 | MG945844 | Unassigned | Unassigned | Unassigned | Singleton | Ref |
| 7740 | MG945847 | Unassigned | Unassigned | Unassigned | Outlier | Ref |
| 7744 | MG945851 | Unassigned | Unassigned | Unassigned | Singleton | Ref |
| 7747 | MG945854 | Unassigned | Unassigned | Unassigned | Overlap (VC_1078/VC_1079) | Ref |
| 7749 | MG945856 | Unassigned | Unassigned | Unassigned | Outlier | Ref |
| 7753 | MG945860 | Unassigned | Unassigned | Unassigned | Outlier | Ref |
| 7754 | MG945861 | Unassigned | Unassigned | Unassigned | Overlap (VC_1078/VC_1082) | Ref |
| 7756 | MG945863 | Unassigned | Unassigned | Unassigned | Overlap (VC_1078/VC_1079) | Ref |
| 7758 | MG945865 | Unassigned | Unassigned | Unassigned | Singleton | Ref |
| 7761 | MG945868 | Unassigned | Unassigned | Unassigned | Singleton | Ref |
| 7765 | MG948468 | Unassigned | Unassigned | Unassigned | Outlier | Ref |
| 7773 | MG962366 | Unassigned | Unassigned | Unassigned | Overlap (VC_560/VC_561/VC_562/VC_563) | Ref |
| 7793 | MG969407 | Unassigned | Unassigned | Unassigned | Outlier | Ref |
| 7799 | MG969413 | Unassigned | Unassigned | Unassigned | Outlier | Ref |
| 7800 | MG969414 | Unassigned | Unassigned | Unassigned | Outlier | Ref |
| 7802 | MG969427 | Unassigned | Unassigned | Unassigned | Outlier | Ref |
| 7804 | MG973030 | Unassigned | Unassigned | Unassigned | Outlier | Ref |
| 7809 | MG986485 | Unassigned | Unassigned | Unassigned | Outlier | Ref |
| 7810 | MG999954 | Unassigned | Unassigned | Unassigned | Overlap (VC_286/VC_290/VC_491) | Ref |
| 7819 | MH001449 | Unassigned | Unassigned | Unassigned | Outlier | Ref |
| 7851 | MH020242 | Unassigned | Unassigned | Unassigned | Overlap (VC_521/VC_550) | Ref |
| 7852 | MH020243 | Unassigned | Unassigned | Unassigned | Overlap (VC_521/VC_550) | Ref |
| 7854 | MH020245 | Unassigned | Unassigned | Unassigned | Overlap (VC_238/VC_240) | Ref |
| 7863 | MH028956 | Unassigned | Unassigned | Unassigned | Outlier | Ref |
| 7867 | MH045555 | Unassigned | Unassigned | Unassigned | Overlap (VC_1241/VC_1242) | Ref |
| 7868 | MH045556 | Unassigned | Unassigned | Unassigned | Outlier | Ref |
| 7871 | MH045559 | Unassigned | Unassigned | Unassigned | Overlap (VC_1241/VC_1242) | Ref |
| 7882 | MH047631 | Unassigned | Unassigned | Unassigned | Outlier | Ref |
| 7887 | MH051248 | Unassigned | Unassigned | Unassigned | Overlap (VC_521/VC_549) | Ref |
| 7916 | MH059633 | Unassigned | Unassigned | Unassigned | Overlap (VC_538/VC_619) | Ref |
| 7917 | MH059634 | Unassigned | Unassigned | Unassigned | Outlier | Ref |
| 7920 | MH059637 | Unassigned | Unassigned | Unassigned | Outlier | Ref |
| 7932 | MH077585 | Unassigned | Unassigned | Unassigned | Outlier | Ref |
| 7933 | MH078572 | Unassigned | Unassigned | Unassigned | Outlier | Ref |
| 7936 | MH105080 | Unassigned | Unassigned | Unassigned | Singleton | Ref |
| 7940 | MH107246 | Unassigned | Unassigned | Unassigned | Overlap (VC_435/VC_656) | Ref |
| 7971 | MH155867 | Unassigned | Unassigned | Unassigned | Overlap (VC_526/VC_554) | Ref |
| 7973 | MH155869 | Unassigned | Unassigned | Unassigned | Overlap (VC_872/VC_873) | Ref |
| 7976 | MH155872 | Unassigned | Unassigned | Unassigned | Overlap (VC_872/VC_873) | Ref |
| 7982 | MH155878 | Unassigned | Unassigned | Unassigned | Overlap (VC_872/VC_873) | Ref |
| 8016 | MH191395 | Unassigned | Unassigned | Unassigned | Overlap (VC_538/VC_619) | Ref |
| 8038 | MH230879 | Unassigned | Unassigned | Unassigned | Outlier | Ref |
| 8039 | MH238466 | Unassigned | Unassigned | Unassigned | Outlier | Ref |
| 8040 | MH238467 | Unassigned | Unassigned | Unassigned | Outlier | Ref |
| 8097 | MH319722 | Unassigned | Unassigned | Unassigned | Singleton | Ref |
| 8098 | MH319723 | Unassigned | Unassigned | Unassigned | Outlier | Ref |
| 8099 | MH319724 | Unassigned | Unassigned | Unassigned | Outlier | Ref |
| 8100 | MH319725 | Unassigned | Unassigned | Unassigned | Singleton | Ref |
| 8101 | MH319732 | Unassigned | Unassigned | Unassigned | Outlier | Ref |

| | | | | | | |
|---|---|---|---|---|---|---|
| 8103 | MH319734 | Unassigned | Unassigned | Unassigned | Overlap (VC_440/VC_659) | Ref |
| 8105 | MH319738 | Unassigned | Unassigned | Unassigned | Outlier | Ref |
| 8106 | MH319741 | Unassigned | Unassigned | Unassigned | Outlier | Ref |
| 8108 | MH319743 | Unassigned | Unassigned | Unassigned | Outlier | Ref |
| 8109 | MH319745 | Unassigned | Unassigned | Unassigned | Outlier | Ref |
| 8113 | MH319751 | Unassigned | Unassigned | Unassigned | Overlap (VC_440/VC_659) | Ref |
| 8114 | MH319752 | Unassigned | Unassigned | Unassigned | Outlier | Ref |
| 8117 | MH319755 | Unassigned | Unassigned | Unassigned | Overlap (VC_440/VC_659) | Ref |
| 8120 | MH319760 | Unassigned | Unassigned | Unassigned | Singleton | Ref |
| 8121 | MH319764 | Unassigned | Unassigned | Unassigned | Outlier | Ref |
| 8122 | MH319765 | Unassigned | Unassigned | Unassigned | Outlier | Ref |
| 8127 | MH327485 | Unassigned | Unassigned | Unassigned | Outlier | Ref |
| 8128 | MH327486 | Unassigned | Unassigned | Unassigned | Outlier | Ref |
| 8130 | MH327490 | Unassigned | Unassigned | Unassigned | Outlier | Ref |
| 8131 | MH327491 | Unassigned | Unassigned | Unassigned | Outlier | Ref |
| 8151 | MH362766 | Unassigned | Unassigned | Unassigned | Overlap (VC_750/VC_751) | Ref |
| 8160 | MH370364 | Unassigned | Unassigned | Unassigned | Outlier | Ref |
| 8178 | MH370382 | Unassigned | Unassigned | Unassigned | Outlier | Ref |
| 8179 | MH370383 | Unassigned | Unassigned | Unassigned | Outlier | Ref |
| 8219 | MH382836 | Unassigned | Unassigned | Unassigned | Outlier | Ref |
| 8226 | MH399770 | Unassigned | Unassigned | Unassigned | Overlap (VC_238/VC_240) | Ref |
| 8260 | MH431930 | Unassigned | Unassigned | Unassigned | Outlier | Ref |
| 8270 | MH443101 | Unassigned | Unassigned | Unassigned | Overlap (VC_32/VC_33) | Ref |
| 8275 | MH445500 | Unassigned | Unassigned | Unassigned | Outlier | Ref |
| 8287 | MH450123 | Unassigned | Unassigned | Unassigned | Overlap (VC_1034/VC_1035/VC_1039) | Ref |
| 8303 | MH454081 | Unassigned | Unassigned | Unassigned | Outlier | Ref |
| 8307 | MH458951 | Unassigned | Unassigned | Unassigned | Outlier | Ref |
| 8332 | MH479922 | Unassigned | Unassigned | Unassigned | Overlap (VC_1241/VC_1242) | Ref |
| 8343 | MH509441 | Unassigned | Unassigned | Unassigned | Outlier | Ref |
| 8344 | MH509442 | Unassigned | Unassigned | Unassigned | Outlier | Ref |
| 8367 | MH517022 | Unassigned | Unassigned | Unassigned | Outlier | Ref |
| 8368 | MH518298 | Unassigned | Unassigned | Unassigned | Outlier | Ref |
| 8387 | MH536826 | Unassigned | Unassigned | Unassigned | Outlier | Ref |
| 8392 | MH538296 | Unassigned | Unassigned | Unassigned | Outlier | Ref |
| 8398 | MH553517 | Unassigned | Unassigned | Unassigned | Outlier | Ref |
| 8403 | MH572270 | Unassigned | Unassigned | Unassigned | Overlap (VC_199/VC_398) | Ref |
| 8404 | MH572271 | Unassigned | Unassigned | Unassigned | Overlap (VC_406/VC_1250) | Ref |
| 8405 | MH572272 | Unassigned | Unassigned | Unassigned | Overlap (VC_406/VC_1250) | Ref |
| 8406 | MH572273 | Unassigned | Unassigned | Unassigned | Overlap (VC_406/VC_1250) | Ref |
| 8411 | MH572278 | Unassigned | Unassigned | Unassigned | Outlier | Ref |
| 8414 | MH572281 | Unassigned | Unassigned | Unassigned | Overlap (VC_199/VC_399/VC_400) | Ref |
| 8422 | MH572289 | Unassigned | Unassigned | Unassigned | Overlap (VC_406/VC_999) | Ref |
| 8430 | MH572297 | Unassigned | Unassigned | Unassigned | Overlap (VC_400/VC_994/VC_1084) | Ref |
| 8431 | MH572298 | Unassigned | Unassigned | Unassigned | Outlier | Ref |
| 8437 | MH572304 | Unassigned | Unassigned | Unassigned | Outlier | Ref |
| 8455 | MH572322 | Unassigned | Unassigned | Unassigned | Overlap (VC_199/VC_399) | Ref |
| 8456 | MH572323 | Unassigned | Unassigned | Unassigned | Overlap (VC_199/VC_399) | Ref |
| 8457 | MH572324 | Unassigned | Unassigned | Unassigned | Overlap (VC_199/VC_399) | Ref |
| 8460 | MH572327 | Unassigned | Unassigned | Unassigned | Overlap (VC_199/VC_399) | Ref |
| 8464 | MH572331 | Unassigned | Unassigned | Unassigned | Outlier | Ref |
| 8465 | MH572332 | Unassigned | Unassigned | Unassigned | Overlap (VC_199/VC_399) | Ref |
| 8466 | MH572333 | Unassigned | Unassigned | Unassigned | Overlap (VC_398/VC_403) | Ref |
| 8468 | MH572335 | Unassigned | Unassigned | Unassigned | Overlap (VC_199/VC_399) | Ref |
| 8470 | MH572337 | Unassigned | Unassigned | Unassigned | Overlap (VC_199/VC_399) | Ref |
| 8471 | MH572338 | Unassigned | Unassigned | Unassigned | Overlap (VC_400/VC_994/VC_1084) | Ref |
| 8472 | MH572339 | Unassigned | Unassigned | Unassigned | Outlier | Ref |
| 8477 | MH572344 | Unassigned | Unassigned | Unassigned | Outlier | Ref |
| 8479 | MH572346 | Unassigned | Unassigned | Unassigned | Outlier | Ref |
| 8483 | MH572350 | Unassigned | Unassigned | Unassigned | Overlap (VC_199/VC_399) | Ref |
| 8485 | MH572352 | Unassigned | Unassigned | Unassigned | Singleton | Ref |
| 8489 | MH572356 | Unassigned | Unassigned | Unassigned | Outlier | Ref |
| 8492 | MH572359 | Unassigned | Unassigned | Unassigned | Outlier | Ref |
| 8496 | MH572363 | Unassigned | Unassigned | Unassigned | Outlier | Ref |
| 8498 | MH572365 | Unassigned | Unassigned | Unassigned | Singleton | Ref |
| 8503 | MH572370 | Unassigned | Unassigned | Unassigned | Overlap (VC_1066/VC_1335) | Ref |
| 8505 | MH572372 | Unassigned | Unassigned | Unassigned | Singleton | Ref |
| 8514 | MH572381 | Unassigned | Unassigned | Unassigned | Outlier | Ref |
| 8531 | MH572398 | Unassigned | Unassigned | Unassigned | Outlier | Ref |
| 8546 | MH572413 | Unassigned | Unassigned | Unassigned | Overlap (VC_1079/VC_1081) | Ref |
| 8548 | MH572415 | Unassigned | Unassigned | Unassigned | Overlap (VC_1324/VC_1335) | Ref |
| 8549 | MH572416 | Unassigned | Unassigned | Unassigned | Overlap (VC_406/VC_1250) | Ref |
| 8550 | MH572417 | Unassigned | Unassigned | Unassigned | Outlier | Ref |
| 8553 | MH572420 | Unassigned | Unassigned | Unassigned | Outlier | Ref |
| 8554 | MH572421 | Unassigned | Unassigned | Unassigned | Overlap (VC_199/VC_399) | Ref |
| 8564 | MH572432 | Unassigned | Unassigned | Unassigned | Overlap (VC_400/VC_994/VC_999/VC_1084) | Ref |
| 8565 | MH572433 | Unassigned | Unassigned | Unassigned | Overlap (VC_202/VC_401) | Ref |
| 8572 | MH572440 | Unassigned | Unassigned | Unassigned | Overlap (VC_199/VC_399) | Ref |
| 8574 | MH572442 | Unassigned | Unassigned | Unassigned | Overlap (VC_399/VC_400/VC_1084) | Ref |
| 8579 | MH572447 | Unassigned | Unassigned | Unassigned | Outlier | Ref |
| 8581 | MH572449 | Unassigned | Unassigned | Unassigned | Overlap (VC_400/VC_994/VC_998/VC_1002/VC_1084) | Ref |
| 8582 | MH572450 | Unassigned | Unassigned | Unassigned | Overlap (VC_399/VC_400/VC_1084) | Ref |
| 8588 | MH572456 | Unassigned | Unassigned | Unassigned | Singleton | Ref |
| 8596 | MH572464 | Unassigned | Unassigned | Unassigned | Overlap (VC_1066/VC_1335) | Ref |
| 8606 | MH572474 | Unassigned | Unassigned | Unassigned | Overlap (VC_199/VC_399) | Ref |
| 8608 | MH572476 | Unassigned | Unassigned | Unassigned | Overlap (VC_402/VC_406) | Ref |
| 8609 | MH572477 | Unassigned | Unassigned | Unassigned | Overlap (VC_402/VC_406) | Ref |
| 8610 | MH572478 | Unassigned | Unassigned | Unassigned | Overlap (VC_402/VC_406) | Ref |
| 8613 | MH572481 | Unassigned | Unassigned | Unassigned | Outlier | Ref |
| 8620 | MH572488 | Unassigned | Unassigned | Unassigned | Outlier | Ref |
| 8633 | MH572501 | Unassigned | Unassigned | Unassigned | Overlap (VC_400/VC_994/VC_999/VC_1084) | Ref |
| 8634 | MH572502 | Unassigned | Unassigned | Unassigned | Outlier | Ref |
| 8655 | MH572523 | Unassigned | Unassigned | Unassigned | Overlap (VC_1324/VC_1335) | Ref |
| 8695 | MH588546 | Unassigned | Unassigned | Unassigned | Overlap (VC_977/VC_980) | Ref |
| 8696 | MH588547 | Unassigned | Unassigned | Unassigned | Overlap (VC_977/VC_980) | Ref |
| 8700 | MH590590 | Unassigned | Unassigned | Unassigned | Overlap (VC_1241/VC_1242) | Ref |
| 8720 | MH598799 | Unassigned | Unassigned | Unassigned | Overlap (VC_481/VC_482) | Ref |
| 8722 | MH598801 | Unassigned | Unassigned | Unassigned | Overlap (VC_481/VC_482) | Ref |
| 8727 | MH598806 | Unassigned | Unassigned | Unassigned | Overlap (VC_481/VC_482) | Ref |
| 8733 | MH626557 | Unassigned | Unassigned | Unassigned | Singleton | Ref |
| 8740 | MH632120 | Unassigned | Unassigned | Unassigned | Outlier | Ref |
| 8745 | MH636380 | Unassigned | Unassigned | Unassigned | Singleton | Ref |
| 8761 | MH651173 | Unassigned | Unassigned | Unassigned | Overlap (VC_227/VC_941) | Ref |
| 8775 | MH651187 | Unassigned | Unassigned | Unassigned | Outlier | Ref |
| 8784 | MH669005 | Unassigned | Unassigned | Unassigned | Overlap (VC_521/VC_548) | Ref |
| 8788 | MH669009 | Unassigned | Unassigned | Unassigned | Overlap (VC_521/VC_548) | Ref |
| 8789 | MH669010 | Unassigned | Unassigned | Unassigned | Overlap (VC_521/VC_548) | Ref |
| 8798 | MH673671 | Unassigned | Unassigned | Unassigned | Singleton | Ref |
| 8799 | MH673672 | Unassigned | Unassigned | Unassigned | Outlier | Ref |
| 8800 | MH673673 | Unassigned | Unassigned | Unassigned | Outlier | Ref |
| 8801 | MH673674 | Unassigned | Unassigned | Unassigned | Outlier | Ref |
| 8822 | MH697589 | Unassigned | Unassigned | Unassigned | Overlap (VC_1164/VC_1243) | Ref |
| 8827 | MH700630 | Unassigned | Unassigned | Unassigned | Outlier | Ref |
| 8828 | MH706966 | Unassigned | Unassigned | Unassigned | Outlier | Ref |
| 8838 | MH719082 | Unassigned | Unassigned | Unassigned | Outlier | Ref |
| 8849 | MH727545 | Unassigned | Unassigned | Unassigned | Overlap (VC_236/VC_238) | Ref |
| 8854 | MH727550 | Unassigned | Unassigned | Unassigned | Outlier | Ref |
| 8860 | MH727556 | Unassigned | Unassigned | Unassigned | Overlap (VC_1172/VC_1173) | Ref |
| 8870 | MH729379 | Unassigned | Unassigned | Unassigned | Outlier | Ref |
| 8888 | MH745368 | Unassigned | Unassigned | Unassigned | Outlier | Ref |
| 8895 | MH766655 | Unassigned | Unassigned | Unassigned | Outlier | Ref |
| 8933 | MH791401 | Unassigned | Unassigned | Unassigned | Overlap (VC_444/VC_580) | Ref |
| 8939 | MH791407 | Unassigned | Unassigned | Unassigned | Overlap (VC_444/VC_580) | Ref |
| 8967 | MH809532 | Unassigned | Unassigned | Unassigned | Overlap (VC_368/VC_369) | Ref |
| 8968 | MH809533 | Unassigned | Unassigned | Unassigned | Overlap (VC_368/VC_369) | Ref |
| 8974 | MH817999 | Unassigned | Unassigned | Unassigned | Overlap (VC_593/VC_597) | Ref |
| 8987 | MH825708 | Unassigned | Unassigned | Unassigned | Overlap (VC_227/VC_941) | Ref |
| 8991 | MH825712 | Unassigned | Unassigned | Unassigned | Outlier | Ref |
| 8993 | MH830339 | Unassigned | Unassigned | Unassigned | Overlap (VC_194/VC_195) | Ref |
| 9001 | MH834601 | Unassigned | Unassigned | Unassigned | Overlap (VC_236/VC_238) | Ref |
| 9036 | MH844558 | Unassigned | Unassigned | Unassigned | Outlier | Ref |
| 9037 | MH845412 | Unassigned | Unassigned | Unassigned | Overlap (VC_584/VC_586) | Ref |
| 9038 | MH853355 | Unassigned | Unassigned | Unassigned | Overlap (VC_318/VC_320) | Ref |
| 9039 | MH853356 | Unassigned | Unassigned | Unassigned | Overlap (VC_294/VC_320) | Ref |
| 9040 | MH853357 | Unassigned | Unassigned | Unassigned | Overlap (VC_334/VC_416/VC_430/VC_777) | Ref |
| 9041 | MH853358 | Unassigned | Unassigned | Unassigned | Overlap (VC_318/VC_320) | Ref |
| 9047 | MH884509 | Unassigned | Unassigned | Unassigned | Outlier | Ref |
| 9048 | MH884510 | Unassigned | Unassigned | Unassigned | Outlier | Ref |
| 9053 | MH884648 | Unassigned | Unassigned | Unassigned | Singleton | Ref |
| 9091 | MH898687 | Unassigned | Unassigned | Unassigned | Overlap (VC_367/VC_369) | Ref |
| 9102 | MH920638 | Unassigned | Unassigned | Unassigned | Outlier | Ref |
| 9108 | MH925093 | Unassigned | Unassigned | Unassigned | Overlap (VC_615/VC_965) | Ref |
| 9179 | MH972261 | Unassigned | Unassigned | Unassigned | Overlap (VC_97/VC_99) | Ref |
| 9180 | MH972262 | Unassigned | Unassigned | Unassigned | Overlap (VC_97/VC_99) | Ref |
| 9181 | MH972263 | Unassigned | Unassigned | Unassigned | Overlap (VC_97/VC_99) | Ref |
| 9214 | MH992162 | Unassigned | Unassigned | Unassigned | Overlap (VC_1065/VC_1367) | Ref |
| 9218 | MH992166 | Unassigned | Unassigned | Unassigned | Outlier | Ref |
| 9220 | MH992168 | Unassigned | Unassigned | Unassigned | Singleton | Ref |
| 9224 | MH992172 | Unassigned | Unassigned | Unassigned | Overlap (VC_1065/VC_1367) | Ref |
| 9228 | MH992176 | Unassigned | Unassigned | Unassigned | Overlap (VC_1065/VC_1367) | Ref |
| 9231 | MH992179 | Unassigned | Unassigned | Unassigned | Overlap (VC_1065/VC_1367) | Ref |
| 9235 | MH992183 | Unassigned | Unassigned | Unassigned | Overlap (VC_327/VC_401) | Ref |
| 9238 | MH992186 | Unassigned | Unassigned | Unassigned | Singleton | Ref |
| 9239 | MH992187 | Unassigned | Unassigned | Unassigned | Overlap (VC_1065/VC_1367) | Ref |
| 9240 | MH992188 | Unassigned | Unassigned | Unassigned | Overlap (VC_199/VC_399/VC_400) | Ref |
| 9242 | MH992190 | Unassigned | Unassigned | Unassigned | Overlap (VC_199/VC_401) | Ref |
| 9250 | MH992198 | Unassigned | Unassigned | Unassigned | Overlap (VC_1065/VC_1367) | Ref |
| 9253 | MH992201 | Unassigned | Unassigned | Unassigned | Outlier | Ref |
| 9255 | MH992203 | Unassigned | Unassigned | Unassigned | Overlap (VC_201/VC_398/VC_401) | Ref |
| 9256 | MH992204 | Unassigned | Unassigned | Unassigned | Outlier | Ref |
| 9261 | MH992209 | Unassigned | Unassigned | Unassigned | Overlap (VC_199/VC_401) | Ref |
| 9264 | MH992212 | Unassigned | Unassigned | Unassigned | Overlap (VC_406/VC_1250) | Ref |
| 9273 | MH992221 | Unassigned | Unassigned | Unassigned | Overlap (VC_1066/VC_1335) | Ref |

| | | | | | | |
|---|---|---|---|---|---|---|
| 9275 | MH992223 | Unassigned | Unassigned | Unassigned | Outlier | Ref |
| 9286 | MK016493 | Unassigned | Unassigned | Unassigned | Outlier | Ref |
| 9299 | MK016663 | Unassigned | Unassigned | Unassigned | Outlier | Ref |
| 9307 | MK044828 | Unassigned | Unassigned | Unassigned | Overlap (VC_256/VC_414) | Ref |
| 9308 | MK044829 | Unassigned | Unassigned | Unassigned | Outlier | Ref |
| 9348 | MK061406 | Unassigned | Unassigned | Unassigned | Overlap (VC_526/VC_554) | Ref |
| 9358 | MK061416 | Unassigned | Unassigned | Unassigned | Outlier | Ref |
| 9364 | MK069556 | Unassigned | Unassigned | Unassigned | Outlier | Ref |
| 9370 | MK075005 | Unassigned | Unassigned | Unassigned | Outlier | Ref |
| 9397 | MK095605 | Unassigned | Unassigned | Unassigned | Outlier | Ref |
| 9398 | MK095606 | Unassigned | Unassigned | Unassigned | Outlier | Ref |
| 9446 | MK170447 | Unassigned | Unassigned | Unassigned | Singleton | Ref |
| 9451 | MK203850 | Unassigned | Unassigned | Unassigned | Outlier | Ref |
| 9464 | MK241539 | Unassigned | Unassigned | Unassigned | Outlier | Ref |
| 9469 | MK249130 | Unassigned | Unassigned | Unassigned | Overlap (VC_1067/VC_1068) | Ref |
| 9473 | MK249134 | Unassigned | Unassigned | Unassigned | Overlap (VC_995/VC_1064) | Ref |
| 9475 | MK249136 | Unassigned | Unassigned | Unassigned | Overlap (VC_1067/VC_1068) | Ref |
| 9476 | MK249137 | Unassigned | Unassigned | Unassigned | Overlap (VC_1067/VC_1068) | Ref |
| 9479 | MK249140 | Unassigned | Unassigned | Unassigned | Overlap (VC_1067/VC_1068) | Ref |
| 9481 | MK249142 | Unassigned | Unassigned | Unassigned | Overlap (VC_1067/VC_1068) | Ref |
| 9484 | MK249145 | Unassigned | Unassigned | Unassigned | Overlap (VC_1067/VC_1068) | Ref |
| 9485 | MK249146 | Unassigned | Unassigned | Unassigned | Overlap (VC_1056/VC_1057/VC_1061) | Ref |
| 9491 | MK249152 | Unassigned | Unassigned | Unassigned | Overlap (VC_1067/VC_1068) | Ref |
| 9496 | MK249157 | Unassigned | Unassigned | Unassigned | Outlier | Ref |
| 9500 | MK249161 | Unassigned | Unassigned | Unassigned | Overlap (VC_1067/VC_1068) | Ref |
| 9503 | MK249164 | Unassigned | Unassigned | Unassigned | Overlap (VC_1067/VC_1068) | Ref |
| 9508 | MK249169 | Unassigned | Unassigned | Unassigned | Overlap (VC_1067/VC_1068) | Ref |
| 9509 | MK249170 | Unassigned | Unassigned | Unassigned | Overlap (VC_1067/VC_1068) | Ref |
| 9510 | MK249171 | Unassigned | Unassigned | Unassigned | Outlier | Ref |
| 9511 | MK249172 | Unassigned | Unassigned | Unassigned | Overlap (VC_998/VC_1000/VC_1002) | Ref |
| 9512 | MK249173 | Unassigned | Unassigned | Unassigned | Overlap (VC_995/VC_996) | Ref |
| 9514 | MK249175 | Unassigned | Unassigned | Unassigned | Overlap (VC_1067/VC_1068) | Ref |
| 9516 | MK249177 | Unassigned | Unassigned | Unassigned | Overlap (VC_201/VC_398/VC_401) | Ref |
| 9521 | MK249182 | Unassigned | Unassigned | Unassigned | Overlap (VC_998/VC_1002) | Ref |
| 9522 | MK249183 | Unassigned | Unassigned | Unassigned | Outlier | Ref |
| 9523 | MK249184 | Unassigned | Unassigned | Unassigned | Overlap (VC_1067/VC_1068) | Ref |
| 9525 | MK249186 | Unassigned | Unassigned | Unassigned | Overlap (VC_998/VC_1002) | Ref |
| 9526 | MK249187 | Unassigned | Unassigned | Unassigned | Overlap (VC_998/VC_1002) | Ref |
| 9532 | MK249193 | Unassigned | Unassigned | Unassigned | Singleton | Ref |
| 9539 | MK249200 | Unassigned | Unassigned | Unassigned | Overlap (VC_398/VC_403) | Ref |
| 9545 | MK249206 | Unassigned | Unassigned | Unassigned | Overlap (VC_998/VC_1002) | Ref |
| 9548 | MK249209 | Unassigned | Unassigned | Unassigned | Outlier | Ref |
| 9549 | MK249210 | Unassigned | Unassigned | Unassigned | Overlap (VC_995/VC_1064) | Ref |
| 9559 | MK249220 | Unassigned | Unassigned | Unassigned | Outlier | Ref |
| 9563 | MK249224 | Unassigned | Unassigned | Unassigned | Overlap (VC_201/VC_398) | Ref |
| 9564 | MK249225 | Unassigned | Unassigned | Unassigned | Outlier | Ref |
| 9565 | MK249226 | Unassigned | Unassigned | Unassigned | Overlap (VC_400/VC_994/VC_1084) | Ref |
| 9566 | MK249227 | Unassigned | Unassigned | Unassigned | Overlap (VC_201/VC_398) | Ref |
| 9569 | MK249230 | Unassigned | Unassigned | Unassigned | Overlap (VC_201/VC_398) | Ref |
| 9573 | MK249234 | Unassigned | Unassigned | Unassigned | Overlap (VC_1066/VC_1335) | Ref |
| 9575 | MK249871 | Unassigned | Unassigned | Unassigned | Outlier | Ref |
| 9612 | MK279848 | Unassigned | Unassigned | Unassigned | Overlap (VC_526/VC_554) | Ref |
| 9663 | MK279899 | Unassigned | Unassigned | Unassigned | Overlap (VC_1035/VC_1039) | Ref |
| 9691 | MK290739 | Unassigned | Unassigned | Unassigned | Outlier | Ref |
| 9694 | MK291442 | Unassigned | Unassigned | Unassigned | Outlier | Ref |
| 9696 | MK291444 | Unassigned | Unassigned | Unassigned | Outlier | Ref |
| 9697 | MK291445 | Unassigned | Unassigned | Unassigned | Outlier | Ref |
| 9700 | MK295205 | Unassigned | Unassigned | Unassigned | Outlier | Ref |
| 9702 | MK301438 | Unassigned | Unassigned | Unassigned | Overlap (VC_203/VC_206) | Ref |
| 9703 | MK301439 | Unassigned | Unassigned | Unassigned | Overlap (VC_203/VC_207) | Ref |
| 9704 | MK301440 | Unassigned | Unassigned | Unassigned | Overlap (VC_203/VC_206) | Ref |
| 9705 | MK301441 | Unassigned | Unassigned | Unassigned | Overlap (VC_203/VC_206) | Ref |
| 9709 | MK301608 | Unassigned | Unassigned | Unassigned | Outlier | Ref |
| 9712 | MK305886 | Unassigned | Unassigned | Unassigned | Overlap (VC_521/VC_548) | Ref |
| 9738 | MK310226 | Unassigned | Unassigned | Unassigned | Outlier | Ref |
| 9753 | MK327938 | Unassigned | Unassigned | Unassigned | Outlier | Ref |
| 9771 | MK358448 | Unassigned | Unassigned | Unassigned | Overlap (VC_442/VC_443) | Ref |
| 9843 | MK368614 | Unassigned | Unassigned | Unassigned | Overlap (VC_442/VC_443) | Ref |
| 9904 | MK411820 | Unassigned | Unassigned | Unassigned | Overlap (VC_750/VC_751) | Ref |
| 9914 | MK416016 | Unassigned | Unassigned | Unassigned | Outlier | Ref |
| 9919 | MK416021 | Unassigned | Unassigned | Unassigned | Outlier | Ref |
| 9927 | MK422450 | Unassigned | Unassigned | Unassigned | Outlier | Ref |
| 9929 | MK422452 | Unassigned | Unassigned | Unassigned | Outlier | Ref |
| 9939 | MK433264 | Unassigned | Unassigned | Unassigned | Overlap (VC_1095/VC_1096) | Ref |
| 9961 | MK433583 | Unassigned | Unassigned | Unassigned | Outlier | Ref |
| 9965 | MK448228 | Unassigned | Unassigned | Unassigned | Outlier | Ref |
| 9968 | MK448232 | Unassigned | Unassigned | Unassigned | Overlap (VC_59/VC_245) | Ref |
| 9972 | MK448236 | Unassigned | Unassigned | Unassigned | Outlier | Ref |
| 9974 | MK448665 | Unassigned | Unassigned | Unassigned | Outlier | Ref |
| 9975 | MK448666 | Unassigned | Unassigned | Unassigned | Overlap (VC_633/VC_634) | Ref |
| 9977 | MK448668 | Unassigned | Unassigned | Unassigned | Overlap (VC_633/VC_634) | Ref |
| 9978 | MK448669 | Unassigned | Unassigned | Unassigned | Overlap (VC_256/VC_414) | Ref |
| 9979 | MK448670 | Unassigned | Unassigned | Unassigned | Overlap (VC_633/VC_634) | Ref |
| 9981 | MK448672 | Unassigned | Unassigned | Unassigned | Outlier | Ref |
| 9986 | MK448677 | Unassigned | Unassigned | Unassigned | Overlap (VC_324/VC_539) | Ref |
| 9988 | MK448679 | Unassigned | Unassigned | Unassigned | Overlap (VC_324/VC_539) | Ref |
| 9991 | MK448682 | Unassigned | Unassigned | Unassigned | Overlap (VC_324/VC_539) | Ref |
| 9996 | MK448687 | Unassigned | Unassigned | Unassigned | Overlap (VC_633/VC_634) | Ref |
| 10000 | MK448691 | Unassigned | Unassigned | Unassigned | Overlap (VC_432/VC_774) | Ref |
| 10004 | MK448695 | Unassigned | Unassigned | Unassigned | Overlap (VC_633/VC_634) | Ref |
| 10010 | MK448701 | Unassigned | Unassigned | Unassigned | Overlap (VC_318/VC_321) | Ref |
| 10012 | MK448703 | Unassigned | Unassigned | Unassigned | Overlap (VC_324/VC_335/VC_636) | Ref |
| 10014 | MK448705 | Unassigned | Unassigned | Unassigned | Overlap (VC_322/VC_431) | Ref |
| 10021 | MK448712 | Unassigned | Unassigned | Unassigned | Overlap (VC_254/VC_258) | Ref |
| 10022 | MK448713 | Unassigned | Unassigned | Unassigned | Overlap (VC_633/VC_634) | Ref |
| 10024 | MK448715 | Unassigned | Unassigned | Unassigned | Overlap (VC_256/VC_414) | Ref |
| 10026 | MK448717 | Unassigned | Unassigned | Unassigned | Overlap (VC_633/VC_634) | Ref |
| 10027 | MK448718 | Unassigned | Unassigned | Unassigned | Outlier | Ref |
| 10028 | MK448719 | Unassigned | Unassigned | Unassigned | Overlap (VC_633/VC_634) | Ref |
| 10031 | MK448722 | Unassigned | Unassigned | Unassigned | Overlap (VC_633/VC_634) | Ref |
| 10032 | MK448723 | Unassigned | Unassigned | Unassigned | Overlap (VC_637/VC_986) | Ref |
| 10034 | MK448725 | Unassigned | Unassigned | Unassigned | Outlier | Ref |
| 10035 | MK448726 | Unassigned | Unassigned | Unassigned | Overlap (VC_633/VC_634) | Ref |
| 10036 | MK448727 | Unassigned | Unassigned | Unassigned | Outlier | Ref |
| 10037 | MK448728 | Unassigned | Unassigned | Unassigned | Overlap (VC_633/VC_634) | Ref |
| 10038 | MK448729 | Unassigned | Unassigned | Unassigned | Overlap (VC_432/VC_774) | Ref |
| 10040 | MK448731 | Unassigned | Unassigned | Unassigned | Overlap (VC_334/VC_416/VC_430/VC_777) | Ref |
| 10043 | MK448734 | Unassigned | Unassigned | Unassigned | Overlap (VC_334/VC_416/VC_430) | Ref |
| 10045 | MK448736 | Unassigned | Unassigned | Unassigned | Overlap (VC_318/VC_320) | Ref |
| 10046 | MK448737 | Unassigned | Unassigned | Unassigned | Outlier | Ref |
| 10047 | MK448738 | Unassigned | Unassigned | Unassigned | Outlier | Ref |
| 10051 | MK448742 | Unassigned | Unassigned | Unassigned | Overlap (VC_318/VC_320) | Ref |
| 10052 | MK448743 | Unassigned | Unassigned | Unassigned | Overlap (VC_324/VC_337) | Ref |
| 10055 | MK448746 | Unassigned | Unassigned | Unassigned | Overlap (VC_637/VC_986) | Ref |
| 10056 | MK448747 | Unassigned | Unassigned | Unassigned | Overlap (VC_254/VC_259) | Ref |
| 10057 | MK448748 | Unassigned | Unassigned | Unassigned | Overlap (VC_254/VC_259) | Ref |
| 10058 | MK448749 | Unassigned | Unassigned | Unassigned | Overlap (VC_294/VC_320) | Ref |
| 10060 | MK448751 | Unassigned | Unassigned | Unassigned | Overlap (VC_254/VC_259) | Ref |
| 10062 | MK448753 | Unassigned | Unassigned | Unassigned | Overlap (VC_637/VC_986) | Ref |
| 10063 | MK448754 | Unassigned | Unassigned | Unassigned | Overlap (VC_416/VC_430/VC_987) | Ref |
| 10064 | MK448755 | Unassigned | Unassigned | Unassigned | Overlap (VC_318/VC_320/VC_323) | Ref |
| 10065 | MK448756 | Unassigned | Unassigned | Unassigned | Overlap (VC_318/VC_320/VC_323) | Ref |
| 10069 | MK448760 | Unassigned | Unassigned | Unassigned | Overlap (VC_294/VC_385) | Ref |
| 10070 | MK448761 | Unassigned | Unassigned | Unassigned | Overlap (VC_294/VC_385) | Ref |
| 10073 | MK448764 | Unassigned | Unassigned | Unassigned | Overlap (VC_416/VC_430/VC_777) | Ref |
| 10077 | MK448768 | Unassigned | Unassigned | Unassigned | Overlap (VC_432/VC_774) | Ref |
| 10079 | MK448770 | Unassigned | Unassigned | Unassigned | Overlap (VC_324/VC_539) | Ref |
| 10085 | MK448776 | Unassigned | Unassigned | Unassigned | Overlap (VC_633/VC_634) | Ref |
| 10089 | MK448780 | Unassigned | Unassigned | Unassigned | Overlap (VC_633/VC_634) | Ref |
| 10090 | MK448781 | Unassigned | Unassigned | Unassigned | Overlap (VC_318/VC_320) | Ref |
| 10094 | MK448785 | Unassigned | Unassigned | Unassigned | Overlap (VC_324/VC_539) | Ref |
| 10096 | MK448787 | Unassigned | Unassigned | Unassigned | Overlap (VC_633/VC_634) | Ref |
| 10101 | MK448792 | Unassigned | Unassigned | Unassigned | Overlap (VC_324/VC_539) | Ref |
| 10105 | MK448796 | Unassigned | Unassigned | Unassigned | Outlier | Ref |
| 10108 | MK448799 | Unassigned | Unassigned | Unassigned | Overlap (VC_256/VC_276) | Ref |
| 10109 | MK448800 | Unassigned | Unassigned | Unassigned | Overlap (VC_256/VC_276) | Ref |
| 10110 | MK448801 | Unassigned | Unassigned | Unassigned | Overlap (VC_294/VC_383) | Ref |
| 10111 | MK448802 | Unassigned | Unassigned | Unassigned | Overlap (VC_633/VC_634) | Ref |
| 10112 | MK448803 | Unassigned | Unassigned | Unassigned | Overlap (VC_318/VC_319) | Ref |
| 10115 | MK448806 | Unassigned | Unassigned | Unassigned | Overlap (VC_324/VC_335/VC_624) | Ref |
| 10118 | MK448809 | Unassigned | Unassigned | Unassigned | Overlap (VC_633/VC_634) | Ref |
| 10119 | MK448810 | Unassigned | Unassigned | Unassigned | Overlap (VC_319/VC_626) | Ref |
| 10121 | MK448812 | Unassigned | Unassigned | Unassigned | Overlap (VC_324/VC_335/VC_624) | Ref |
| 10122 | MK448813 | Unassigned | Unassigned | Unassigned | Overlap (VC_319/VC_790) | Ref |
| 10125 | MK448816 | Unassigned | Unassigned | Unassigned | Overlap (VC_318/VC_319) | Ref |
| 10126 | MK448817 | Unassigned | Unassigned | Unassigned | Outlier | Ref |
| 10129 | MK448820 | Unassigned | Unassigned | Unassigned | Outlier | Ref |
| 10132 | MK448823 | Unassigned | Unassigned | Unassigned | Overlap (VC_259/VC_625/VC_626) | Ref |
| 10134 | MK448825 | Unassigned | Unassigned | Unassigned | Overlap (VC_633/VC_634) | Ref |
| 10135 | MK448826 | Unassigned | Unassigned | Unassigned | Overlap (VC_414/VC_791) | Ref |
| 10136 | MK448827 | Unassigned | Unassigned | Unassigned | Overlap (VC_633/VC_634) | Ref |
| 10138 | MK448829 | Unassigned | Unassigned | Unassigned | Overlap (VC_318/VC_320) | Ref |
| 10139 | MK448830 | Unassigned | Unassigned | Unassigned | Outlier | Ref |
| 10144 | MK448835 | Unassigned | Unassigned | Unassigned | Overlap (VC_633/VC_634) | Ref |
| 10145 | MK448836 | Unassigned | Unassigned | Unassigned | Overlap (VC_633/VC_634) | Ref |
| 10146 | MK448837 | Unassigned | Unassigned | Unassigned | Overlap (VC_318/VC_320) | Ref |
| 10147 | MK448838 | Unassigned | Unassigned | Unassigned | Outlier | Ref |
| 10149 | MK448840 | Unassigned | Unassigned | Unassigned | Overlap (VC_633/VC_634) | Ref |
| 10151 | MK448842 | Unassigned | Unassigned | Unassigned | Outlier | Ref |
| 10153 | MK448844 | Unassigned | Unassigned | Unassigned | Outlier | Ref |
| 10155 | MK448846 | Unassigned | Unassigned | Unassigned | Overlap (VC_633/VC_634) | Ref |
| 10156 | MK448847 | Unassigned | Unassigned | Unassigned | Overlap (VC_633/VC_634) | Ref |

| | | | | | | |
|---|---|---|---|---|---|---|
| 10157 | MK448848 | Unassigned | Unassigned | Unassigned | Overlap (VC_324/VC_539) | Ref |
| 10158 | MK448849 | Unassigned | Unassigned | Unassigned | Outlier | Ref |
| 10160 | MK448851 | Unassigned | Unassigned | Unassigned | Overlap (VC_320/VC_324) | Ref |
| 10165 | MK448856 | Unassigned | Unassigned | Unassigned | Outlier | Ref |
| 10166 | MK448857 | Unassigned | Unassigned | Unassigned | Overlap (VC_324/VC_539) | Ref |
| 10169 | MK448860 | Unassigned | Unassigned | Unassigned | Overlap (VC_324/VC_539) | Ref |
| 10172 | MK448863 | Unassigned | Unassigned | Unassigned | Overlap (VC_334/VC_416/VC_430/VC_777) | Ref |
| 10174 | MK448865 | Unassigned | Unassigned | Unassigned | Overlap (VC_318/VC_321) | Ref |
| 10178 | MK448869 | Unassigned | Unassigned | Unassigned | Overlap (VC_318/VC_321) | Ref |
| 10179 | MK448870 | Unassigned | Unassigned | Unassigned | Overlap (VC_318/VC_321) | Ref |
| 10181 | MK448872 | Unassigned | Unassigned | Unassigned | Overlap (VC_633/VC_634) | Ref |
| 10182 | MK448873 | Unassigned | Unassigned | Unassigned | Overlap (VC_322/VC_431) | Ref |
| 10185 | MK448876 | Unassigned | Unassigned | Unassigned | Overlap (VC_324/VC_335/VC_636) | Ref |
| 10187 | MK448878 | Unassigned | Unassigned | Unassigned | Outlier | Ref |
| 10188 | MK448879 | Unassigned | Unassigned | Unassigned | Outlier | Ref |
| 10189 | MK448880 | Unassigned | Unassigned | Unassigned | Overlap (VC_254/VC_258) | Ref |
| 10192 | MK448883 | Unassigned | Unassigned | Unassigned | Overlap (VC_633/VC_634) | Ref |
| 10193 | MK448884 | Unassigned | Unassigned | Unassigned | Outlier | Ref |
| 10194 | MK448885 | Unassigned | Unassigned | Unassigned | Outlier | Ref |
| 10197 | MK448888 | Unassigned | Unassigned | Unassigned | Overlap (VC_334/VC_416/VC_430/VC_777) | Ref |
| 10198 | MK448889 | Unassigned | Unassigned | Unassigned | Outlier | Ref |
| 10200 | MK448891 | Unassigned | Unassigned | Unassigned | Outlier | Ref |
| 10201 | MK448892 | Unassigned | Unassigned | Unassigned | Overlap (VC_256/VC_414) | Ref |
| 10203 | MK448894 | Unassigned | Unassigned | Unassigned | Outlier | Ref |
| 10212 | MK448903 | Unassigned | Unassigned | Unassigned | Overlap (VC_324/VC_337) | Ref |
| 10215 | MK448906 | Unassigned | Unassigned | Unassigned | Overlap (VC_633/VC_634) | Ref |
| 10216 | MK448907 | Unassigned | Unassigned | Unassigned | Overlap (VC_633/VC_634) | Ref |
| 10217 | MK448908 | Unassigned | Unassigned | Unassigned | Outlier | Ref |
| 10220 | MK448911 | Unassigned | Unassigned | Unassigned | Overlap (VC_318/VC_320) | Ref |
| 10222 | MK448913 | Unassigned | Unassigned | Unassigned | Overlap (VC_633/VC_634) | Ref |
| 10225 | MK448916 | Unassigned | Unassigned | Unassigned | Overlap (VC_318/VC_320) | Ref |
| 10228 | MK448919 | Unassigned | Unassigned | Unassigned | Overlap (VC_324/VC_337) | Ref |
| 10230 | MK448921 | Unassigned | Unassigned | Unassigned | Overlap (VC_324/VC_337) | Ref |
| 10231 | MK448922 | Unassigned | Unassigned | Unassigned | Overlap (VC_633/VC_634) | Ref |
| 10233 | MK448924 | Unassigned | Unassigned | Unassigned | Overlap (VC_637/VC_986) | Ref |
| 10235 | MK448926 | Unassigned | Unassigned | Unassigned | Overlap (VC_254/VC_259) | Ref |
| 10237 | MK448928 | Unassigned | Unassigned | Unassigned | Overlap (VC_633/VC_634) | Ref |
| 10238 | MK448929 | Unassigned | Unassigned | Unassigned | Overlap (VC_416/VC_430/VC_987) | Ref |
| 10241 | MK448932 | Unassigned | Unassigned | Unassigned | Overlap (VC_256/VC_414) | Ref |
| 10244 | MK448935 | Unassigned | Unassigned | Unassigned | Overlap (VC_633/VC_634) | Ref |
| 10245 | MK448936 | Unassigned | Unassigned | Unassigned | Overlap (VC_633/VC_634) | Ref |
| 10246 | MK448937 | Unassigned | Unassigned | Unassigned | Outlier | Ref |
| 10247 | MK448938 | Unassigned | Unassigned | Unassigned | Overlap (VC_318/VC_320) | Ref |
| 10249 | MK448940 | Unassigned | Unassigned | Unassigned | Overlap (VC_256/VC_414) | Ref |
| 10257 | MK448948 | Unassigned | Unassigned | Unassigned | Overlap (VC_432/VC_774) | Ref |
| 10265 | MK448956 | Unassigned | Unassigned | Unassigned | Overlap (VC_320/VC_324) | Ref |
| 10280 | MK448971 | Unassigned | Unassigned | Unassigned | Overlap (VC_320/VC_324) | Ref |
| 10287 | MK448978 | Unassigned | Unassigned | Unassigned | Overlap (VC_324/VC_539) | Ref |
| 10288 | MK448979 | Unassigned | Unassigned | Unassigned | Overlap (VC_256/VC_276) | Ref |
| 10289 | MK448980 | Unassigned | Unassigned | Unassigned | Outlier | Ref |
| 10290 | MK448981 | Unassigned | Unassigned | Unassigned | Overlap (VC_318/VC_319) | Ref |
| 10293 | MK448984 | Unassigned | Unassigned | Unassigned | Overlap (VC_318/VC_319) | Ref |
| 10294 | MK448985 | Unassigned | Unassigned | Unassigned | Overlap (VC_318/VC_319) | Ref |
| 10295 | MK448986 | Unassigned | Unassigned | Unassigned | Outlier | Ref |
| 10296 | MK448987 | Unassigned | Unassigned | Unassigned | Outlier | Ref |
| 10299 | MK448990 | Unassigned | Unassigned | Unassigned | Overlap (VC_324/VC_335/VC_624) | Ref |
| 10301 | MK448992 | Unassigned | Unassigned | Unassigned | Overlap (VC_334/VC_416/VC_430/VC_777) | Ref |
| 10302 | MK448993 | Unassigned | Unassigned | Unassigned | Outlier | Ref |
| 10303 | MK448994 | Unassigned | Unassigned | Unassigned | Overlap (VC_633/VC_634) | Ref |
| 10305 | MK448996 | Unassigned | Unassigned | Unassigned | Overlap (VC_259/VC_625/VC_626) | Ref |
| 10307 | MK448998 | Unassigned | Unassigned | Unassigned | Outlier | Ref |
| 10308 | MK448999 | Unassigned | Unassigned | Unassigned | Overlap (VC_633/VC_634) | Ref |
| 10310 | MK449001 | Unassigned | Unassigned | Unassigned | Overlap (VC_414/VC_791) | Ref |
| 10314 | MK449005 | Unassigned | Unassigned | Unassigned | Outlier | Ref |
| 10316 | MK449007 | Unassigned | Unassigned | Unassigned | Overlap (VC_633/VC_634) | Ref |
| 10320 | MK449011 | Unassigned | Unassigned | Unassigned | Overlap (VC_633/VC_634) | Ref |
| 10355 | MK493321 | Unassigned | Unassigned | Unassigned | Overlap (VC_435/VC_656) | Ref |
| 10356 | MK493322 | Unassigned | Unassigned | Unassigned | Overlap (VC_435/VC_656) | Ref |
| 10357 | MK493323 | Unassigned | Unassigned | Unassigned | Overlap (VC_435/VC_656) | Ref |
| 10358 | MK493324 | Unassigned | Unassigned | Unassigned | Overlap (VC_435/VC_656) | Ref |
| 10359 | MK493325 | Unassigned | Unassigned | Unassigned | Overlap (VC_435/VC_656) | Ref |
| 10393 | MK494120 | Unassigned | Unassigned | Unassigned | Overlap (VC_521/VC_550) | Ref |
| 10411 | MK496684 | Unassigned | Unassigned | Unassigned | Singleton | Ref |
| 10414 | MK496687 | Unassigned | Unassigned | Unassigned | Singleton | Ref |
| 10418 | MK496691 | Unassigned | Unassigned | Unassigned | Outlier | Ref |
| 10422 | MK496695 | Unassigned | Unassigned | Unassigned | Singleton | Ref |
| 10430 | MK496703 | Unassigned | Unassigned | Unassigned | Outlier | Ref |
| 10432 | MK496705 | Unassigned | Unassigned | Unassigned | Singleton | Ref |
| 10433 | MK496706 | Unassigned | Unassigned | Unassigned | Outlier | Ref |
| 10434 | MK496707 | Unassigned | Unassigned | Unassigned | Overlap (VC_1372/VC_1373) | Ref |
| 10435 | MK496708 | Unassigned | Unassigned | Unassigned | Singleton | Ref |
| 10436 | MK496709 | Unassigned | Unassigned | Unassigned | Singleton | Ref |
| 10440 | MK496713 | Unassigned | Unassigned | Unassigned | Singleton | Ref |
| 10442 | MK496715 | Unassigned | Unassigned | Unassigned | Overlap (VC_1270/VC_1271) | Ref |
| 10445 | MK496718 | Unassigned | Unassigned | Unassigned | Singleton | Ref |
| 10449 | MK496722 | Unassigned | Unassigned | Unassigned | Outlier | Ref |
| 10451 | MK496724 | Unassigned | Unassigned | Unassigned | Outlier | Ref |
| 10453 | MK496726 | Unassigned | Unassigned | Unassigned | Overlap (VC_1106/VC_1107) | Ref |
| 10454 | MK496727 | Unassigned | Unassigned | Unassigned | Singleton | Ref |
| 10455 | MK496728 | Unassigned | Unassigned | Unassigned | Outlier | Ref |
| 10457 | MK496730 | Unassigned | Unassigned | Unassigned | Singleton | Ref |
| 10458 | MK496731 | Unassigned | Unassigned | Unassigned | Outlier | Ref |
| 10460 | MK496733 | Unassigned | Unassigned | Unassigned | Overlap (VC_1054/VC_1058) | Ref |
| 10461 | MK496734 | Unassigned | Unassigned | Unassigned | Outlier | Ref |
| 10462 | MK496735 | Unassigned | Unassigned | Unassigned | Outlier | Ref |
| 10463 | MK496736 | Unassigned | Unassigned | Unassigned | Singleton | Ref |
| 10464 | MK496737 | Unassigned | Unassigned | Unassigned | Outlier | Ref |
| 10466 | MK496739 | Unassigned | Unassigned | Unassigned | Overlap (VC_1054/VC_1058) | Ref |
| 10467 | MK496740 | Unassigned | Unassigned | Unassigned | Outlier | Ref |
| 10468 | MK496741 | Unassigned | Unassigned | Unassigned | Overlap (VC_1130/VC_1135) | Ref |
| 10469 | MK496742 | Unassigned | Unassigned | Unassigned | Overlap (VC_1054/VC_1055) | Ref |
| 10474 | MK496747 | Unassigned | Unassigned | Unassigned | Outlier | Ref |
| 10476 | MK496749 | Unassigned | Unassigned | Unassigned | Singleton | Ref |
| 10478 | MK496751 | Unassigned | Unassigned | Unassigned | Outlier | Ref |
| 10479 | MK496752 | Unassigned | Unassigned | Unassigned | Overlap (VC_1067/VC_1068) | Ref |
| 10482 | MK496755 | Unassigned | Unassigned | Unassigned | Singleton | Ref |
| 10486 | MK496759 | Unassigned | Unassigned | Unassigned | Singleton | Ref |
| 10489 | MK496762 | Unassigned | Unassigned | Unassigned | Singleton | Ref |
| 10490 | MK496763 | Unassigned | Unassigned | Unassigned | Outlier | Ref |
| 10496 | MK496769 | Unassigned | Unassigned | Unassigned | Singleton | Ref |
| 10500 | MK496773 | Unassigned | Unassigned | Unassigned | Singleton | Ref |
| 10504 | MK496777 | Unassigned | Unassigned | Unassigned | Singleton | Ref |
| 10505 | MK496778 | Unassigned | Unassigned | Unassigned | Overlap (VC_1130/VC_1135) | Ref |
| 10510 | MK496783 | Unassigned | Unassigned | Unassigned | Singleton | Ref |
| 10515 | MK496788 | Unassigned | Unassigned | Unassigned | Overlap (VC_1130/VC_1132/VC_1249) | Ref |
| 10516 | MK496789 | Unassigned | Unassigned | Unassigned | Overlap (VC_1270/VC_1271) | Ref |
| 10518 | MK496791 | Unassigned | Unassigned | Unassigned | Singleton | Ref |
| 10520 | MK496793 | Unassigned | Unassigned | Unassigned | Outlier | Ref |
| 10521 | MK496794 | Unassigned | Unassigned | Unassigned | Singleton | Ref |
| 10522 | MK496795 | Unassigned | Unassigned | Unassigned | Outlier | Ref |
| 10524 | MK496797 | Unassigned | Unassigned | Unassigned | Overlap (VC_1130/VC_1249) | Ref |
| 10525 | MK496798 | Unassigned | Unassigned | Unassigned | Outlier | Ref |
| 10526 | MK496799 | Unassigned | Unassigned | Unassigned | Outlier | Ref |
| 10527 | MK496800 | Unassigned | Unassigned | Unassigned | Singleton | Ref |
| 10528 | MK496801 | Unassigned | Unassigned | Unassigned | Outlier | Ref |
| 10529 | MK496802 | Unassigned | Unassigned | Unassigned | Outlier | Ref |
| 10530 | MK496803 | Unassigned | Unassigned | Unassigned | Overlap (VC_1133/VC_1135) | Ref |
| 10533 | MK496806 | Unassigned | Unassigned | Unassigned | Overlap (VC_1114/VC_1136) | Ref |
| 10534 | MK496807 | Unassigned | Unassigned | Unassigned | Outlier | Ref |
| 10536 | MK496809 | Unassigned | Unassigned | Unassigned | Outlier | Ref |
| 10537 | MK496810 | Unassigned | Unassigned | Unassigned | Outlier | Ref |
| 10541 | MK496814 | Unassigned | Unassigned | Unassigned | Outlier | Ref |
| 10544 | MK496817 | Unassigned | Unassigned | Unassigned | Singleton | Ref |
| 10546 | MK496819 | Unassigned | Unassigned | Unassigned | Singleton | Ref |
| 10548 | MK496821 | Unassigned | Unassigned | Unassigned | Outlier | Ref |
| 10549 | MK496822 | Unassigned | Unassigned | Unassigned | Overlap (VC_996/VC_1065) | Ref |
| 10550 | MK496823 | Unassigned | Unassigned | Unassigned | Overlap (VC_996/VC_1065) | Ref |
| 10553 | MK496826 | Unassigned | Unassigned | Unassigned | Outlier | Ref |
| 10559 | MK504445 | Unassigned | Unassigned | Unassigned | Outlier | Ref |
| 10672 | MK524492 | Unassigned | Unassigned | Unassigned | Overlap (VC_227/VC_941) | Ref |
| 10712 | MK533143 | Unassigned | Unassigned | Unassigned | Outlier | Ref |
| 10715 | MK552105 | Unassigned | Unassigned | Unassigned | Outlier | Ref |
| 10719 | MK554696 | Unassigned | Unassigned | Unassigned | Outlier | Ref |
| 10720 | MK557849 | Unassigned | Unassigned | Unassigned | Singleton | Ref |
| 10725 | MK560763 | Unassigned | Unassigned | Unassigned | Outlier | Ref |
| 10735 | MK570056 | Unassigned | Unassigned | Unassigned | Singleton | Ref |
| 10736 | MK570057 | Unassigned | Unassigned | Unassigned | Outlier | Ref |
| 10741 | MK575466 | Unassigned | Unassigned | Unassigned | Overlap (VC_129/VC_711) | Ref |
| 10761 | MK613347 | Unassigned | Unassigned | Unassigned | Outlier | Ref |
| 10762 | MK613348 | Unassigned | Unassigned | Unassigned | Outlier | Ref |
| 10763 | MK613349 | Unassigned | Unassigned | Unassigned | Outlier | Ref |
| 10768 | MK618717 | Unassigned | Unassigned | Unassigned | Overlap (VC_82/VC_619) | Ref |
| 10775 | MK620899 | Unassigned | Unassigned | Unassigned | Overlap (VC_1098/VC_1099) | Ref |
| 10782 | MK629525 | Unassigned | Unassigned | Unassigned | Outlier | Ref |
| 10783 | MK629526 | Unassigned | Unassigned | Unassigned | Outlier | Ref |
| 10786 | MK629529 | Unassigned | Unassigned | Unassigned | Outlier | Ref |
| 10794 | MK660713 | Unassigned | Unassigned | Unassigned | Outlier | Ref |
| 10800 | MK672802 | Unassigned | Unassigned | Unassigned | Overlap (VC_127/VC_789) | Ref |
| 10803 | MK672805 | Unassigned | Unassigned | Unassigned | Overlap (VC_127/VC_789) | Ref |
| 10805 | MK675901 | Unassigned | Unassigned | Unassigned | Outlier | Ref |
| 10810 | MK686070 | Unassigned | Unassigned | Unassigned | Overlap (VC_872/VC_873) | Ref |
| 10812 | MK686072 | Unassigned | Unassigned | Unassigned | Overlap (VC_872/VC_873) | Ref |

| | | | | | | |
|---|---|---|---|---|---|---|
| 10950 | MK765554 | Unassigned | Unassigned | Unassigned | Overlap (VC_1284/VC_1285) | Ref |
| 10962 | MK765566 | Unassigned | Unassigned | Unassigned | Overlap (VC_1284/VC_1285) | Ref |
| 10966 | MK765570 | Unassigned | Unassigned | Unassigned | Singleton | Ref |
| 10980 | MK765584 | Unassigned | Unassigned | Unassigned | Singleton | Ref |
| 10981 | MK765585 | Unassigned | Unassigned | Unassigned | Outlier | Ref |
| 10982 | MK765586 | Unassigned | Unassigned | Unassigned | Overlap (VC_1284/VC_1285) | Ref |
| 10991 | MK765595 | Unassigned | Unassigned | Unassigned | Overlap (VC_1403/VC_1405) | Ref |
| 10993 | MK765597 | Unassigned | Unassigned | Unassigned | Overlap (VC_1284/VC_1285) | Ref |
| 11004 | MK765608 | Unassigned | Unassigned | Unassigned | Singleton | Ref |
| 11010 | MK765614 | Unassigned | Unassigned | Unassigned | Overlap (VC_1284/VC_1285) | Ref |
| 11012 | MK765616 | Unassigned | Unassigned | Unassigned | Overlap (VC_1284/VC_1285) | Ref |
| 11014 | MK765618 | Unassigned | Unassigned | Unassigned | Overlap (VC_1284/VC_1285) | Ref |
| 11015 | MK765619 | Unassigned | Unassigned | Unassigned | Outlier | Ref |
| 11020 | MK765624 | Unassigned | Unassigned | Unassigned | Overlap (VC_1403/VC_1405) | Ref |
| 11023 | MK765627 | Unassigned | Unassigned | Unassigned | Overlap (VC_1403/VC_1405) | Ref |
| 11025 | MK765629 | Unassigned | Unassigned | Unassigned | Overlap (VC_1403/VC_1405) | Ref |
| 11028 | MK765632 | Unassigned | Unassigned | Unassigned | Outlier | Ref |
| 11031 | MK765635 | Unassigned | Unassigned | Unassigned | Outlier | Ref |
| 11032 | MK765636 | Unassigned | Unassigned | Unassigned | Overlap (VC_1284/VC_1285) | Ref |
| 11033 | MK765637 | Unassigned | Unassigned | Unassigned | Overlap (VC_1284/VC_1285) | Ref |
| 11043 | MK765647 | Unassigned | Unassigned | Unassigned | Overlap (VC_1284/VC_1285) | Ref |
| 11048 | MK765652 | Unassigned | Unassigned | Unassigned | Overlap (VC_1284/VC_1285) | Ref |
| 11061 | MK765665 | Unassigned | Unassigned | Unassigned | Overlap (VC_1284/VC_1285) | Ref |
| 11066 | MK770119 | Unassigned | Unassigned | Unassigned | Outlier | Ref |
| 11078 | MK791318 | Unassigned | Unassigned | Unassigned | Outlier | Ref |
| 11103 | MK801729 | Unassigned | Unassigned | Unassigned | Overlap (VC_526/VC_554) | Ref |
| 11119 | MK813942 | Unassigned | Unassigned | Unassigned | Outlier | Ref |
| 11120 | MK813943 | Unassigned | Unassigned | Unassigned | Overlap (VC_602/VC_813) | Ref |
| 11154 | MK838112 | Unassigned | Unassigned | Unassigned | Overlap (VC_602/VC_813) | Ref |
| 11156 | MK838114 | Unassigned | Unassigned | Unassigned | Overlap (VC_602/VC_813) | Ref |
| 11157 | MK838115 | Unassigned | Unassigned | Unassigned | Overlap (VC_602/VC_813) | Ref |
| 11159 | MK843319 | Unassigned | Unassigned | Unassigned | Overlap (VC_654/VC_747) | Ref |
| 11169 | MK875792 | Unassigned | Unassigned | Unassigned | Overlap (VC_521/VC_548) | Ref |
| 11171 | MK875794 | Unassigned | Unassigned | Unassigned | Overlap (VC_1164/VC_1243) | Ref |
| 11185 | MK878902 | Unassigned | Unassigned | Unassigned | Overlap (VC_526/VC_554) | Ref |
| 11190 | MK882925 | Unassigned | Unassigned | Unassigned | Outlier | Ref |
| 11200 | MK894437 | Unassigned | Unassigned | Unassigned | Overlap (VC_1241/VC_1242) | Ref |
| 11207 | MK903280 | Unassigned | Unassigned | Unassigned | Overlap (VC_75/VC_115) | Ref |
| 11211 | MK905543 | Unassigned | Unassigned | Unassigned | Overlap (VC_442/VC_443) | Ref |
| 11265 | MK911750 | Unassigned | Unassigned | Unassigned | Overlap (VC_629/VC_637/VC_986) | Ref |
| 11301 | MK937592 | Unassigned | Unassigned | Unassigned | Overlap (VC_560/VC_562) | Ref |
| 11314 | MK937605 | Unassigned | Unassigned | Unassigned | Overlap (VC_227/VC_941) | Ref |
| 11318 | MK937609 | Unassigned | Unassigned | Unassigned | Overlap (VC_526/VC_554) | Ref |
| 11324 | MK942064 | Unassigned | Unassigned | Unassigned | Overlap (VC_279/VC_283) | Ref |
| 11359 | MK967380 | Unassigned | Unassigned | Unassigned | Outlier | Ref |
| 11385 | MK972688 | Unassigned | Unassigned | Unassigned | Overlap (VC_29/VC_37) | Ref |
| 11386 | MK972689 | Unassigned | Unassigned | Unassigned | Overlap (VC_29/VC_37) | Ref |
| 11392 | MK972695 | Unassigned | Unassigned | Unassigned | Overlap (VC_29/VC_37) | Ref |
| 11393 | MK972696 | Unassigned | Unassigned | Unassigned | Overlap (VC_29/VC_37) | Ref |
| 11394 | MK972697 | Unassigned | Unassigned | Unassigned | Overlap (VC_29/VC_37) | Ref |
| 11418 | MK977695 | Unassigned | Unassigned | Unassigned | Overlap (VC_560/VC_563) | Ref |
| 11419 | MK977696 | Unassigned | Unassigned | Unassigned | Overlap (VC_560/VC_563) | Ref |
| 11426 | MK977703 | Unassigned | Unassigned | Unassigned | Overlap (VC_1034/VC_1035/VC_1039) | Ref |
| 11427 | MK977704 | Unassigned | Unassigned | Unassigned | Overlap (VC_1095/VC_1096) | Ref |
| 11430 | MK977707 | Unassigned | Unassigned | Unassigned | Outlier | Ref |
| 11440 | MK984681 | Unassigned | Unassigned | Unassigned | Overlap (VC_482/VC_484) | Ref |
| 11442 | MK994522 | Unassigned | Unassigned | Unassigned | Singleton | Ref |
| 11458 | MN013081 | Unassigned | Unassigned | Unassigned | Overlap (VC_491/VC_492) | Ref |
| 11461 | MN013084 | Unassigned | Unassigned | Unassigned | Overlap (VC_286/VC_290/VC_491) | Ref |
| 11467 | MN013090 | Unassigned | Unassigned | Unassigned | Overlap (VC_82/VC_619) | Ref |
| 11468 | MN013189 | Unassigned | Unassigned | Unassigned | Singleton | Ref |
| 11504 | MN044033 | Unassigned | Unassigned | Unassigned | Overlap (VC_491/VC_492) | Ref |
| 11518 | MN062189 | Unassigned | Unassigned | Unassigned | Overlap (VC_82/VC_619) | Ref |
| 11521 | MN062703 | Unassigned | Unassigned | Unassigned | Overlap (VC_1241/VC_1242) | Ref |
| 11523 | MN062705 | Unassigned | Unassigned | Unassigned | Overlap (VC_983/VC_984) | Ref |
| 11538 | MN062720 | Unassigned | Unassigned | Unassigned | Outlier | Ref |
| 11547 | MN082625 | Unassigned | Unassigned | Unassigned | Overlap (VC_818/VC_820) | Ref |
| 11556 | MN096267 | Unassigned | Unassigned | Unassigned | Overlap (VC_177/VC_296) | Ref |
| 11557 | MN096355 | Unassigned | Unassigned | Unassigned | Outlier | Ref |
| 11564 | MN096362 | Unassigned | Unassigned | Unassigned | Outlier | Ref |
| 11571 | MN096369 | Unassigned | Unassigned | Unassigned | Overlap (VC_1098/VC_1099) | Ref |
| 11606 | MN102376 | Unassigned | Unassigned | Unassigned | Overlap (VC_615/VC_965) | Ref |
| 11607 | MN103401 | Unassigned | Unassigned | Unassigned | Outlier | Ref |
| 11608 | MN103533 | Unassigned | Unassigned | Unassigned | Outlier | Ref |
| 11611 | MN106244 | Unassigned | Unassigned | Unassigned | Overlap (VC_593/VC_597) | Ref |
| 11612 | MN106245 | Unassigned | Unassigned | Unassigned | Overlap (VC_491/VC_492) | Ref |
| 11646 | MN175679 | Unassigned | Unassigned | Unassigned | Overlap (VC_615/VC_965) | Ref |
| 11649 | MN176220 | Unassigned | Unassigned | Unassigned | Overlap (VC_101/VC_106) | Ref |
| 11650 | MN176221 | Unassigned | Unassigned | Unassigned | Overlap (VC_101/VC_106) | Ref |
| 11659 | MN176230 | Unassigned | Unassigned | Unassigned | Overlap (VC_101/VC_106) | Ref |
| 11660 | MN176231 | Unassigned | Unassigned | Unassigned | Overlap (VC_101/VC_106) | Ref |
| 11661 | MN176232 | Unassigned | Unassigned | Unassigned | Overlap (VC_101/VC_106) | Ref |
| 11670 | MN180253 | Unassigned | Unassigned | Unassigned | Outlier | Ref |
| 11680 | MN184886 | Unassigned | Unassigned | Unassigned | Outlier | Ref |
| 11698 | MN204502 | Unassigned | Unassigned | Unassigned | Overlap (VC_521/VC_548) | Ref |
| 11708 | MN224566 | Unassigned | Unassigned | Unassigned | Outlier | Ref |
| 11720 | MN234170 | Unassigned | Unassigned | Unassigned | Outlier | Ref |
| 11722 | MN234172 | Unassigned | Unassigned | Unassigned | Outlier | Ref |
| 11723 | MN234173 | Unassigned | Unassigned | Unassigned | Overlap (VC_1165/VC_1166) | Ref |
| 11725 | MN234175 | Unassigned | Unassigned | Unassigned | Overlap (VC_1165/VC_1166) | Ref |
| 11728 | MN234178 | Unassigned | Unassigned | Unassigned | Outlier | Ref |
| 11730 | MN234180 | Unassigned | Unassigned | Unassigned | Overlap (VC_1165/VC_1166) | Ref |
| 11734 | MN234184 | Unassigned | Unassigned | Unassigned | Outlier | Ref |
| 11736 | MN234186 | Unassigned | Unassigned | Unassigned | Overlap (VC_1165/VC_1166) | Ref |
| 11738 | MN234188 | Unassigned | Unassigned | Unassigned | Outlier | Ref |
| 11739 | MN234189 | Unassigned | Unassigned | Unassigned | Overlap (VC_983/VC_984) | Ref |
| 11741 | MN234191 | Unassigned | Unassigned | Unassigned | Outlier | Ref |
| 11752 | MN234202 | Unassigned | Unassigned | Unassigned | Overlap (VC_1165/VC_1166) | Ref |
| 11760 | MN234210 | Unassigned | Unassigned | Unassigned | Overlap (VC_1165/VC_1166) | Ref |
| 11768 | MN234218 | Unassigned | Unassigned | Unassigned | Overlap (VC_526/VC_554) | Ref |
| 11779 | MN234229 | Unassigned | Unassigned | Unassigned | Overlap (VC_1165/VC_1166) | Ref |
| 11790 | MN266305 | Unassigned | Unassigned | Unassigned | Outlier | Ref |
| 11815 | MN270889 | Unassigned | Unassigned | Unassigned | Overlap (VC_481/VC_482/VC_484) | Ref |
| 11816 | MN270890 | Unassigned | Unassigned | Unassigned | Overlap (VC_481/VC_482/VC_484) | Ref |
| 11833 | MN284903 | Unassigned | Unassigned | Unassigned | Outlier | Ref |
| 11842 | MN308080 | Unassigned | Unassigned | Unassigned | Overlap (VC_392/VC_395) | Ref |
| 11862 | MN327636 | Unassigned | Unassigned | Unassigned | Overlap (VC_392/VC_395) | Ref |
| 11871 | MN335165 | Unassigned | Unassigned | Unassigned | Singleton | Ref |
| 11917 | MN379461 | Unassigned | Unassigned | Unassigned | Outlier | Ref |
| 11920 | MN379631 | Unassigned | Unassigned | Unassigned | Outlier | Ref |
| 11923 | MN379634 | Unassigned | Unassigned | Unassigned | Overlap (VC_1056/VC_1057/VC_1061/VC_1143) | Ref |
| 11924 | MN379635 | Unassigned | Unassigned | Unassigned | Overlap (VC_1120/VC_1121) | Ref |
| 11931 | MN379642 | Unassigned | Unassigned | Unassigned | Overlap (VC_1055/VC_1057) | Ref |
| 11933 | MN379644 | Unassigned | Unassigned | Unassigned | Overlap (VC_1079/VC_1081) | Ref |
| 11938 | MN379649 | Unassigned | Unassigned | Unassigned | Overlap (VC_1079/VC_1081) | Ref |
| 11951 | MN395284 | Unassigned | Unassigned | Unassigned | Overlap (VC_491/VC_492) | Ref |
| 11955 | MN399336 | Unassigned | Unassigned | Unassigned | Overlap (VC_702/VC_704) | Ref |
| 11960 | MN428047 | Unassigned | Unassigned | Unassigned | Overlap (VC_521/VC_548) | Ref |
| 11961 | MN428048 | Unassigned | Unassigned | Unassigned | Overlap (VC_526/VC_554) | Ref |
| 11968 | MN428055 | Unassigned | Unassigned | Unassigned | Overlap (VC_526/VC_554) | Ref |
| 11970 | MN428057 | Unassigned | Unassigned | Unassigned | Overlap (VC_526/VC_554) | Ref |
| 11982 | MN434092 | Unassigned | Unassigned | Unassigned | Overlap (VC_491/VC_492) | Ref |
| 12005 | MN478374 | Unassigned | Unassigned | Unassigned | Overlap (VC_596/VC_599) | Ref |
| 12007 | MN478376 | Unassigned | Unassigned | Unassigned | Overlap (VC_455/VC_1028) | Ref |
| 12020 | MN497414 | Unassigned | Unassigned | Unassigned | Outlier | Ref |
| 12021 | MN497415 | Unassigned | Unassigned | Unassigned | Overlap (VC_702/VC_704) | Ref |
| 12025 | MN505213 | Unassigned | Unassigned | Unassigned | Outlier | Ref |
| 12026 | MN508356 | Unassigned | Unassigned | Unassigned | Outlier | Ref |
| 12044 | MN518139 | Unassigned | Unassigned | Unassigned | Outlier | Ref |
| 12047 | MN524844 | Unassigned | Unassigned | Unassigned | Overlap (VC_818/VC_821) | Ref |
| 12048 | MN524845 | Unassigned | Unassigned | Unassigned | Overlap (VC_818/VC_821) | Ref |
| 12050 | MN530981 | Unassigned | Unassigned | Unassigned | Outlier | Ref |
| 12051 | MN534315 | Unassigned | Unassigned | Unassigned | Overlap (VC_257/VC_262) | Ref |
| 12054 | MN534318 | Unassigned | Unassigned | Unassigned | Overlap (VC_257/VC_262) | Ref |
| 12055 | MN534319 | Unassigned | Unassigned | Unassigned | Overlap (VC_257/VC_262) | Ref |
| 12058 | MN536026 | Unassigned | Unassigned | Unassigned | Outlier | Ref |
| 12060 | MN539721 | Unassigned | Unassigned | Unassigned | Outlier | Ref |
| 12064 | MN545971 | Unassigned | Unassigned | Unassigned | Outlier | Ref |
| 12082 | MN562221 | Unassigned | Unassigned | Unassigned | Overlap (VC_442/VC_443) | Ref |
| 12090 | MN584918 | Unassigned | Unassigned | Unassigned | Outlier | Ref |
| 12092 | MN585195 | Unassigned | Unassigned | Unassigned | Outlier | Ref |
| 12150 | MN586020 | Unassigned | Unassigned | Unassigned | Outlier | Ref |
| 12155 | MN586025 | Unassigned | Unassigned | Unassigned | Overlap (VC_238/VC_240) | Ref |
| 12163 | MN586033 | Unassigned | Unassigned | Unassigned | Outlier | Ref |
| 12170 | MN586040 | Unassigned | Unassigned | Unassigned | Overlap (VC_560/VC_561/VC_562/VC_563) | Ref |
| 12200 | MN602881 | Unassigned | Unassigned | Unassigned | Outlier | Ref |
| 12207 | MN605466 | Unassigned | Unassigned | Unassigned | Outlier | Ref |
| 12215 | MN617835 | Unassigned | Unassigned | Unassigned | Overlap (VC_286/VC_290/VC_491) | Ref |
| 12230 | MN685189 | Unassigned | Unassigned | Unassigned | Overlap (VC_481/VC_482/VC_484) | Ref |
| 12231 | MN685190 | Unassigned | Unassigned | Unassigned | Overlap (VC_481/VC_482/VC_484) | Ref |
| 12232 | MN685191 | Unassigned | Unassigned | Unassigned | Overlap (VC_481/VC_482/VC_484) | Ref |
| 12268 | MN692199 | Unassigned | Unassigned | Unassigned | Outlier | Ref |
| 12269 | MN692200 | Unassigned | Unassigned | Unassigned | Outlier | Ref |
| 12279 | MN698244 | Unassigned | Unassigned | Unassigned | Outlier | Ref |
| 12301 | MN718199 | Unassigned | Unassigned | Unassigned | Outlier | Ref |
| 12318 | MN734436 | Unassigned | Unassigned | Unassigned | Outlier | Ref |
| 12326 | MN735433 | Unassigned | Unassigned | Unassigned | Overlap (VC_521/VC_548) | Ref |
| 12331 | MN746332 | Unassigned | Unassigned | Unassigned | Overlap (VC_279/VC_283) | Ref |
| 12332 | MN781108 | Unassigned | Unassigned | Unassigned | Overlap (VC_491/VC_492) | Ref |
| 12343 | MN794238 | Unassigned | Unassigned | Unassigned | Overlap (VC_905/VC_907) | Ref |
| 12359 | MN812211 | Unassigned | Unassigned | Unassigned | Outlier | Ref |
| 12387 | MN812239 | Unassigned | Unassigned | Unassigned | Outlier | Ref |

| | | | | | | |
|---|---|---|---|---|---|---|
| 12394 | MN812689 | Unassigned | Unassigned | Unassigned | Outlier | Ref |
| 12413 | MN813681 | Unassigned | Unassigned | Unassigned | Overlap (VC_227/VC_941) | Ref |
| 12417 | MN813685 | Unassigned | Unassigned | Unassigned | Outlier | Ref |
| 12418 | MN813686 | Unassigned | Unassigned | Unassigned | Outlier | Ref |
| 12433 | MN820898 | Unassigned | Unassigned | Unassigned | Outlier | Ref |
| 12434 | MN830252 | Unassigned | Unassigned | Unassigned | Overlap (VC_274/VC_382) | Ref |
| 12435 | MN830253 | Unassigned | Unassigned | Unassigned | Overlap (VC_274/VC_382) | Ref |
| 12439 | MN830257 | Unassigned | Unassigned | Unassigned | Outlier | Ref |
| 12441 | MN830259 | Unassigned | Unassigned | Unassigned | Overlap (VC_273/VC_384) | Ref |
| 12444 | MN840487 | Unassigned | Unassigned | Unassigned | Overlap (VC_932/VC_1050) | Ref |
| 12481 | MN850599 | Unassigned | Unassigned | Unassigned | Outlier | Ref |
| 12534 | MN850656 | Unassigned | Unassigned | Unassigned | Outlier | Ref |
| 12536 | MN857473 | Unassigned | Unassigned | Unassigned | Outlier | Ref |
| 12537 | MN857617 | Unassigned | Unassigned | Unassigned | Overlap (VC_818/VC_820) | Ref |
| 12540 | MN862334 | Unassigned | Unassigned | Unassigned | Overlap (VC_406/VC_1250) | Ref |
| 12542 | MN862336 | Unassigned | Unassigned | Unassigned | Overlap (VC_199/VC_401) | Ref |
| 12543 | MN862337 | Unassigned | Unassigned | Unassigned | Overlap (VC_998/VC_1002) | Ref |
| 12548 | MN862342 | Unassigned | Unassigned | Unassigned | Overlap (VC_199/VC_401) | Ref |
| 12550 | MN862344 | Unassigned | Unassigned | Unassigned | Outlier | Ref |
| 12553 | MN862347 | Unassigned | Unassigned | Unassigned | Singleton | Ref |
| 12556 | MN871440 | Unassigned | Unassigned | Unassigned | Overlap (VC_444/VC_580) | Ref |
| 12593 | MN871477 | Unassigned | Unassigned | Unassigned | Outlier | Ref |
| 12632 | MN877442 | Unassigned | Unassigned | Unassigned | Outlier | Ref |
| 12635 | MN882550 | Unassigned | Unassigned | Unassigned | Overlap (VC_877/VC_1413) | Ref |
| 12638 | MN882553 | Unassigned | Unassigned | Unassigned | Overlap (VC_817/VC_1210) | Ref |
| 12639 | MN882554 | Unassigned | Unassigned | Unassigned | Overlap (VC_877/VC_1413) | Ref |
| 12651 | MN892485 | Unassigned | Unassigned | Unassigned | Outlier | Ref |
| 12668 | MN908684 | Unassigned | Unassigned | Unassigned | Overlap (VC_1035/VC_1039) | Ref |
| 12670 | MN908686 | Unassigned | Unassigned | Unassigned | Overlap (VC_1164/VC_1243) | Ref |
| 12671 | MN908687 | Unassigned | Unassigned | Unassigned | Overlap (VC_560/VC_563) | Ref |
| 12672 | MN908688 | Unassigned | Unassigned | Unassigned | Outlier | Ref |
| 12678 | MN908694 | Unassigned | Unassigned | Unassigned | Overlap (VC_1172/VC_1173) | Ref |
| 12679 | MN917146 | Unassigned | Unassigned | Unassigned | Overlap (VC_674/VC_675) | Ref |
| 12680 | MN922296 | Unassigned | Unassigned | Unassigned | Overlap (VC_355/VC_618) | Ref |
| 12685 | MN935203 | Unassigned | Unassigned | Unassigned | Overlap (VC_194/VC_195) | Ref |
| 12687 | MN938931 | Unassigned | Unassigned | Unassigned | Overlap (VC_629/VC_637/VC_986) | Ref |
| 12693 | MN945900 | Unassigned | Unassigned | Unassigned | Overlap (VC_521/VC_548) | Ref |
| 12702 | MN956515 | Unassigned | Unassigned | Unassigned | Overlap (VC_355/VC_618) | Ref |
| 12714 | MN988464 | Unassigned | Unassigned | Unassigned | Overlap (VC_1403/VC_1406) | Ref |
| 12726 | MN988476 | Unassigned | Unassigned | Unassigned | Overlap (VC_1403/VC_1406) | Ref |
| 12727 | MN988477 | Unassigned | Unassigned | Unassigned | Overlap (VC_1403/VC_1406) | Ref |
| 12733 | MN988483 | Unassigned | Unassigned | Unassigned | Overlap (VC_784/VC_786) | Ref |
| 12737 | MN988487 | Unassigned | Unassigned | Unassigned | Outlier | Ref |
| 12747 | MN988497 | Unassigned | Unassigned | Unassigned | Overlap (VC_784/VC_786) | Ref |
| 12767 | MN988517 | Unassigned | Unassigned | Unassigned | Overlap (VC_784/VC_786) | Ref |
| 12769 | MN988519 | Unassigned | Unassigned | Unassigned | Outlier | Ref |
| 12789 | MN988539 | Unassigned | Unassigned | Unassigned | Outlier | Ref |
| 12792 | MN988542 | Unassigned | Unassigned | Unassigned | Outlier | Ref |
| 12805 | MN988555 | Unassigned | Unassigned | Unassigned | Overlap (VC_784/VC_786) | Ref |
| 12822 | MN995824 | Unassigned | Unassigned | Unassigned | Overlap (VC_431/VC_742) | Ref |
| 12826 | MT002873 | Unassigned | Unassigned | Unassigned | Outlier | Ref |
| 12834 | MT006236 | Unassigned | Unassigned | Unassigned | Outlier | Ref |
| 12844 | MT024859 | Unassigned | Unassigned | Unassigned | Outlier | Ref |
| 12847 | MT024862 | Unassigned | Unassigned | Unassigned | Overlap (VC_1164/VC_1243) | Ref |
| 12859 | MT028491 | Unassigned | Unassigned | Unassigned | Outlier | Ref |
| 12860 | MT028492 | Unassigned | Unassigned | Unassigned | Outlier | Ref |
| 12974 | MT094430 | Unassigned | Unassigned | Unassigned | Overlap (VC_71/VC_74) | Ref |
| 12976 | MT104122 | Unassigned | Unassigned | Unassigned | Outlier | Ref |
| 12979 | MT104467 | Unassigned | Unassigned | Unassigned | Overlap (VC_455/VC_1028) | Ref |
| 12986 | MT104474 | Unassigned | Unassigned | Unassigned | Outlier | Ref |
| 13005 | MT114163 | Unassigned | Unassigned | Unassigned | Overlap (VC_560/VC_562) | Ref |
| 13018 | MT118296 | Unassigned | Unassigned | Unassigned | Outlier | Ref |
| 13053 | MT121968 | Unassigned | Unassigned | Unassigned | Outlier | Ref |
| 13073 | MT135260 | Unassigned | Unassigned | Unassigned | Outlier | Ref |
| 13081 | MT135268 | Unassigned | Unassigned | Unassigned | Outlier | Ref |
| 13083 | MT135270 | Unassigned | Unassigned | Unassigned | Outlier | Ref |
| 13092 | MT135279 | Unassigned | Unassigned | Unassigned | Outlier | Ref |
| 13093 | MT135280 | Unassigned | Unassigned | Unassigned | Outlier | Ref |
| 13107 | MT135295 | Unassigned | Unassigned | Unassigned | Singleton | Ref |
| 13112 | MT135300 | Unassigned | Unassigned | Unassigned | Outlier | Ref |
| 13124 | MT135312 | Unassigned | Unassigned | Unassigned | Outlier | Ref |
| 13125 | MT135313 | Unassigned | Unassigned | Unassigned | Singleton | Ref |
| 13128 | MT135327 | Unassigned | Unassigned | Unassigned | Outlier | Ref |
| 13142 | MT135341 | Unassigned | Unassigned | Unassigned | Singleton | Ref |
| 13157 | MT135356 | Unassigned | Unassigned | Unassigned | Outlier | Ref |
| 13161 | MT135360 | Unassigned | Unassigned | Unassigned | Singleton | Ref |
| 13163 | MT135362 | Unassigned | Unassigned | Unassigned | Overlap (VC_1253/VC_1317) | Ref |
| 13165 | MT135364 | Unassigned | Unassigned | Unassigned | Singleton | Ref |
| 13187 | MT135386 | Unassigned | Unassigned | Unassigned | Singleton | Ref |
| 13191 | MT135390 | Unassigned | Unassigned | Unassigned | Outlier | Ref |
| 13199 | MT135398 | Unassigned | Unassigned | Unassigned | Overlap (VC_1253/VC_1317) | Ref |
| 13218 | MT135417 | Unassigned | Unassigned | Unassigned | Outlier | Ref |
| 13229 | MT135428 | Unassigned | Unassigned | Unassigned | Singleton | Ref |
| 13243 | MT135442 | Unassigned | Unassigned | Unassigned | Singleton | Ref |
| 13249 | MT135448 | Unassigned | Unassigned | Unassigned | Overlap (VC_1287/VC_1290) | Ref |
| 13250 | MT135449 | Unassigned | Unassigned | Unassigned | Singleton | Ref |
| 13251 | MT135450 | Unassigned | Unassigned | Unassigned | Outlier | Ref |
| 13253 | MT135452 | Unassigned | Unassigned | Unassigned | Outlier | Ref |
| 13265 | MT135464 | Unassigned | Unassigned | Unassigned | Singleton | Ref |
| 13268 | MT151386 | Unassigned | Unassigned | Unassigned | Outlier | Ref |
| 13277 | MT161385 | Unassigned | Unassigned | Unassigned | Outlier | Ref |
| 13278 | MT161386 | Unassigned | Unassigned | Unassigned | Overlap (VC_538/VC_619) | Ref |
| 13279 | MT161387 | Unassigned | Unassigned | Unassigned | Overlap (VC_538/VC_619) | Ref |
| 13280 | MT161459 | Unassigned | Unassigned | Unassigned | Overlap (VC_444/VC_580) | Ref |
| 13283 | MT162468 | Unassigned | Unassigned | Unassigned | Overlap (VC_435/VC_656) | Ref |
| 13287 | MT176427 | Unassigned | Unassigned | Unassigned | Overlap (VC_458/VC_586) | Ref |
| 13293 | MT185428 | Unassigned | Unassigned | Unassigned | Overlap (VC_1056/VC_1057/VC_1061/VC_1151) | Ref |
| 13300 | MT188704 | Unassigned | Unassigned | Unassigned | Outlier | Ref |
| 13301 | MT192033 | Unassigned | Unassigned | Unassigned | Outlier | Ref |
| 13306 | MT210152 | Unassigned | Unassigned | Unassigned | Outlier | Ref |
| 13309 | MT225100 | Unassigned | Unassigned | Unassigned | Overlap (VC_42/VC_57) | Ref |
| 13310 | MT225101 | Unassigned | Unassigned | Unassigned | Outlier | Ref |
| 13316 | MT233525 | Unassigned | Unassigned | Unassigned | Outlier | Ref |
| 13330 | MT259468 | Unassigned | Unassigned | Unassigned | Outlier | Ref |
| 13335 | MT303952 | Unassigned | Unassigned | Unassigned | Overlap (VC_633/VC_634) | Ref |
| 13343 | MT310857 | Unassigned | Unassigned | Unassigned | Overlap (VC_227/VC_941) | Ref |
| 13348 | MT310862 | Unassigned | Unassigned | Unassigned | Overlap (VC_983/VC_984) | Ref |
| 13380 | MT310894 | Unassigned | Unassigned | Unassigned | Overlap (VC_1172/VC_1173) | Ref |
| 13386 | MT311967 | Unassigned | Unassigned | Unassigned | Overlap (VC_633/VC_634) | Ref |
| 13387 | MT311968 | Unassigned | Unassigned | Unassigned | Overlap (VC_633/VC_634) | Ref |
| 13396 | MT330372 | Unassigned | Unassigned | Unassigned | Outlier | Ref |
| 13404 | MT345684 | Unassigned | Unassigned | Unassigned | Outlier | Ref |
| 13405 | MT349887 | Unassigned | Unassigned | Unassigned | Outlier | Ref |
| 13413 | MT360682 | Unassigned | Unassigned | Unassigned | Outlier | Ref |
| 13416 | MT362618 | Unassigned | Unassigned | Unassigned | Outlier | Ref |
| 13452 | MT375522 | Unassigned | Unassigned | Unassigned | Outlier | Ref |
| 13453 | MT375523 | Unassigned | Unassigned | Unassigned | Overlap (VC_481/VC_482) | Ref |
| 13454 | MT375524 | Unassigned | Unassigned | Unassigned | Overlap (VC_481/VC_482) | Ref |
| 13467 | MT410774 | Unassigned | Unassigned | Unassigned | Outlier | Ref |
| 13471 | MT416612 | Unassigned | Unassigned | Unassigned | Overlap (VC_87/VC_88) | Ref |
| 13494 | MT446395 | Unassigned | Unassigned | Unassigned | Overlap (VC_458/VC_586) | Ref |
| 13500 | MT446401 | Unassigned | Unassigned | Unassigned | Overlap (VC_458/VC_586) | Ref |
| 13503 | MT446404 | Unassigned | Unassigned | Unassigned | Overlap (VC_458/VC_586) | Ref |
| 13523 | MT448615 | Unassigned | Unassigned | Unassigned | Outlier | Ref |
| 13527 | MT451980 | Unassigned | Unassigned | Unassigned | Overlap (VC_728/VC_730) | Ref |
| 13528 | MT451981 | Unassigned | Unassigned | Unassigned | Overlap (VC_728/VC_730) | Ref |
| 13529 | MT451982 | Unassigned | Unassigned | Unassigned | Overlap (VC_728/VC_730) | Ref |
| 13530 | MT451983 | Unassigned | Unassigned | Unassigned | Overlap (VC_1164/VC_1243) | Ref |
| 13531 | MT451984 | Unassigned | Unassigned | Unassigned | Overlap (VC_1164/VC_1243) | Ref |
| 13684 | MT498043 | Unassigned | Unassigned | Unassigned | Outlier | Ref |
| 13699 | MT498058 | Unassigned | Unassigned | Unassigned | Outlier | Ref |
| 13703 | MT498062 | Unassigned | Unassigned | Unassigned | Overlap (VC_1241/VC_1242) | Ref |
| 13704 | MT498063 | Unassigned | Unassigned | Unassigned | Overlap (VC_1164/VC_1243) | Ref |
| 13712 | MT500539 | Unassigned | Unassigned | Unassigned | Outlier | Ref |
| 13726 | MT521991 | Unassigned | Unassigned | Unassigned | Overlap (VC_728/VC_730) | Ref |
| 13727 | MT521992 | Unassigned | Unassigned | Unassigned | Outlier | Ref |
| 13728 | MT521993 | Unassigned | Unassigned | Unassigned | Overlap (VC_560/VC_561/VC_562/VC_563) | Ref |
| 13735 | MT522000 | Unassigned | Unassigned | Unassigned | Overlap (VC_521/VC_524) | Ref |
| 13757 | MT553342 | Unassigned | Unassigned | Unassigned | Overlap (VC_1172/VC_1173) | Ref |
| 13762 | MT553347 | Unassigned | Unassigned | Unassigned | Overlap (VC_521/VC_550) | Ref |
| 13774 | MT584811 | Unassigned | Unassigned | Unassigned | Outlier | Ref |
| 13821 | MT596497 | Unassigned | Unassigned | Unassigned | Overlap (VC_13/VC_70) | Ref |
| 13822 | MT596498 | Unassigned | Unassigned | Unassigned | Overlap (VC_13/VC_70) | Ref |
| 13824 | MT596500 | Unassigned | Unassigned | Unassigned | Overlap (VC_97/VC_99) | Ref |
| 13831 | MT597419 | Unassigned | Unassigned | Unassigned | Outlier | Ref |
| 13870 | MT647606 | Unassigned | Unassigned | Unassigned | Outlier | Ref |
| 13901 | MT663720 | Unassigned | Unassigned | Unassigned | Outlier | Ref |
| 13902 | MT664721 | Unassigned | Unassigned | Unassigned | Outlier | Ref |
| 13919 | MT679222 | Unassigned | Unassigned | Unassigned | Overlap (VC_194/VC_195) | Ref |
| 13920 | MT679223 | Unassigned | Unassigned | Unassigned | Overlap (VC_194/VC_195) | Ref |
| 13960 | MT700412 | Unassigned | Unassigned | Unassigned | Outlier | Ref |
| 13965 | MT701590 | Unassigned | Unassigned | Unassigned | Overlap (VC_442/VC_443) | Ref |
| 13982 | MT711370 | Unassigned | Unassigned | Unassigned | Singleton | Ref |
| 13995 | MT711977 | Unassigned | Unassigned | Unassigned | Singleton | Ref |
| 14014 | MT724048 | Unassigned | Unassigned | Unassigned | Outlier | Ref |
| 14039 | MT732457 | Unassigned | Unassigned | Unassigned | Outlier | Ref |
| 14055 | MT732473 | Unassigned | Unassigned | Unassigned | Outlier | Ref |
| 14056 | MT732474 | Unassigned | Unassigned | Unassigned | Outlier | Ref |
| 14057 | MT732475 | Unassigned | Unassigned | Unassigned | Outlier | Ref |
| 14059 | MT735630 | Unassigned | Unassigned | Unassigned | Overlap (VC_905/VC_907) | Ref |
| 14061 | MT740245 | Unassigned | Unassigned | Unassigned | Overlap (VC_194/VC_195) | Ref |
| 14063 | MT740307 | Unassigned | Unassigned | Unassigned | Outlier | Ref |

| | | | | | | |
|---|---|---|---|---|---|---|
| 14065 | MT740315 | Unassigned | Unassigned | Unassigned | Outlier | Ref |
| 14066 | MT740725 | Unassigned | Unassigned | Unassigned | Overlap (VC_365/VC_371) | Ref |
| 14070 | MT740729 | Unassigned | Unassigned | Unassigned | Overlap (VC_365/VC_371) | Ref |
| 14076 | MT740735 | Unassigned | Unassigned | Unassigned | Overlap (VC_365/VC_371) | Ref |
| 14085 | MT740744 | Unassigned | Unassigned | Unassigned | Overlap (VC_365/VC_371) | Ref |
| 14087 | MT740746 | Unassigned | Unassigned | Unassigned | Overlap (VC_365/VC_371) | Ref |
| 14088 | MT740747 | Unassigned | Unassigned | Unassigned | Overlap (VC_365/VC_371) | Ref |
| 14089 | MT740748 | Unassigned | Unassigned | Unassigned | Overlap (VC_83/VC_85) | Ref |
| 14096 | MT747435 | Unassigned | Unassigned | Unassigned | Overlap (VC_431/VC_742) | Ref |
| 14104 | MT764845 | Unassigned | Unassigned | Unassigned | Outlier | Ref |
| 14113 | MT771343 | Unassigned | Unassigned | Unassigned | Outlier | Ref |
| 14116 | MT771346 | Unassigned | Unassigned | Unassigned | Overlap (VC_526/VC_554) | Ref |
| 14123 | MT774377 | Unassigned | Unassigned | Unassigned | Overlap (VC_674/VC_675) | Ref |
| 14148 | MT774402 | Unassigned | Unassigned | Unassigned | Overlap (VC_503/VC_504/VC_505) | Ref |
| 14152 | MT774406 | Unassigned | Unassigned | Unassigned | Overlap (VC_503/VC_504/VC_505) | Ref |
| 14154 | MT774408 | Unassigned | Unassigned | Unassigned | Overlap (VC_503/VC_505) | Ref |
| 14155 | MT774409 | Unassigned | Unassigned | Unassigned | Overlap (VC_503/VC_505) | Ref |
| 14156 | MT774410 | Unassigned | Unassigned | Unassigned | Overlap (VC_503/VC_505) | Ref |
| 14157 | MT774411 | Unassigned | Unassigned | Unassigned | Overlap (VC_503/VC_505) | Ref |
| 14173 | MT783706 | Unassigned | Unassigned | Unassigned | Outlier | Ref |
| 14180 | MT806187 | Unassigned | Unassigned | Unassigned | Outlier | Ref |
| 14215 | MT843276 | Unassigned | Unassigned | Unassigned | Outlier | Ref |
| 14216 | MT855965 | Unassigned | Unassigned | Unassigned | Singleton | Ref |
| 14239 | MT872956 | Unassigned | Unassigned | Unassigned | Outlier | Ref |
| 14240 | MT880870 | Unassigned | Unassigned | Unassigned | Outlier | Ref |
| 14241 | MT880871 | Unassigned | Unassigned | Unassigned | Outlier | Ref |
| 14267 | MT889377 | Unassigned | Unassigned | Unassigned | Overlap (VC_1164/VC_1243) | Ref |
| 14270 | MT889380 | Unassigned | Unassigned | Unassigned | Overlap (VC_521/VC_549) | Ref |
| 14277 | MT889387 | Unassigned | Unassigned | Unassigned | Outlier | Ref |
| 14306 | MT901797 | Unassigned | Unassigned | Unassigned | Overlap (VC_62/VC_63) | Ref |
| 14307 | MT901798 | Unassigned | Unassigned | Unassigned | Overlap (VC_62/VC_63) | Ref |
| 14308 | MT901799 | Unassigned | Unassigned | Unassigned | Overlap (VC_62/VC_63) | Ref |
| 14309 | MT901800 | Unassigned | Unassigned | Unassigned | Overlap (VC_62/VC_63) | Ref |
| 14320 | MT939240 | Unassigned | Unassigned | Unassigned | Outlier | Ref |
| 14322 | MT939242 | Unassigned | Unassigned | Unassigned | Overlap (VC_695/VC_696) | Ref |
| 14328 | MT939492 | Unassigned | Unassigned | Unassigned | Outlier | Ref |
| 14333 | MT951568 | Unassigned | Unassigned | Unassigned | Overlap (VC_75/VC_115) | Ref |
| 14336 | MT952843 | Unassigned | Unassigned | Unassigned | Overlap (VC_1095/VC_1096) | Ref |
| 14383 | MW009675 | Unassigned | Unassigned | Unassigned | Overlap (VC_355/VC_618) | Ref |
| 14400 | MW021759 | Unassigned | Unassigned | Unassigned | Overlap (VC_82/VC_619) | Ref |
| 14402 | MW021761 | Unassigned | Unassigned | Unassigned | Overlap (VC_1347/VC_1348) | Ref |
| 14408 | MW021767 | Unassigned | Unassigned | Unassigned | Overlap (VC_1347/VC_1348) | Ref |
| 14460 | MW055913 | Unassigned | Unassigned | Unassigned | Outlier | Ref |
| 14468 | MW057856 | Unassigned | Unassigned | Unassigned | Outlier | Ref |
| 14473 | MW057861 | Unassigned | Unassigned | Unassigned | Outlier | Ref |
| 14474 | MW057862 | Unassigned | Unassigned | Unassigned | Outlier | Ref |
| 14486 | MW082583 | Unassigned | Unassigned | Unassigned | Outlier | Ref |
| 14487 | MW082584 | Unassigned | Unassigned | Unassigned | Outlier | Ref |
| 14489 | MW091528 | Unassigned | Unassigned | Unassigned | Outlier | Ref |
| 14493 | MW117965 | Unassigned | Unassigned | Unassigned | Outlier | Ref |
| 14499 | MW145136 | Unassigned | Unassigned | Unassigned | Overlap (VC_71/VC_74) | Ref |
| 14500 | MW145137 | Unassigned | Unassigned | Unassigned | Outlier | Ref |
| 14501 | MW145138 | Unassigned | Unassigned | Unassigned | Overlap (VC_463/VC_609) | Ref |
| 14516 | MW151244 | Unassigned | Unassigned | Unassigned | Overlap (VC_750/VC_751) | Ref |
| 14535 | MW202423 | Unassigned | Unassigned | Unassigned | Singleton | Ref |
| 14536 | MW202426 | Unassigned | Unassigned | Unassigned | Singleton | Ref |
| 14537 | MW202444 | Unassigned | Unassigned | Unassigned | Singleton | Ref |
| 14540 | MW202458 | Unassigned | Unassigned | Unassigned | Singleton | Ref |
| 14547 | MW202476 | Unassigned | Unassigned | Unassigned | Singleton | Ref |
| 14548 | MW202477 | Unassigned | Unassigned | Unassigned | Outlier | Ref |
| 14550 | MW202481 | Unassigned | Unassigned | Unassigned | Outlier | Ref |
| 14551 | MW202482 | Unassigned | Unassigned | Unassigned | Singleton | Ref |
| 14552 | MW202483 | Unassigned | Unassigned | Unassigned | Outlier | Ref |
| 14553 | MW202484 | Unassigned | Unassigned | Unassigned | Singleton | Ref |
| 14554 | MW202485 | Unassigned | Unassigned | Unassigned | Outlier | Ref |
| 14557 | MW202491 | Unassigned | Unassigned | Unassigned | Outlier | Ref |
| 14559 | MW202497 | Unassigned | Unassigned | Unassigned | Singleton | Ref |
| 14560 | MW202510 | Unassigned | Unassigned | Unassigned | Outlier | Ref |
| 14561 | MW202512 | Unassigned | Unassigned | Unassigned | Outlier | Ref |
| 14568 | MW202554 | Unassigned | Unassigned | Unassigned | Singleton | Ref |
| 14570 | MW202558 | Unassigned | Unassigned | Unassigned | Outlier | Ref |
| 14571 | MW202561 | Unassigned | Unassigned | Unassigned | Outlier | Ref |
| 14573 | MW202567 | Unassigned | Unassigned | Unassigned | Overlap (VC_199/VC_399/VC_400) | Ref |
| 14574 | MW202568 | Unassigned | Unassigned | Unassigned | Singleton | Ref |
| 14576 | MW202573 | Unassigned | Unassigned | Unassigned | Outlier | Ref |
| 14577 | MW202575 | Unassigned | Unassigned | Unassigned | Overlap (VC_199/VC_401) | Ref |
| 14578 | MW202577 | Unassigned | Unassigned | Unassigned | Singleton | Ref |
| 14581 | MW202590 | Unassigned | Unassigned | Unassigned | Overlap (VC_996/VC_1065) | Ref |
| 14584 | MW202597 | Unassigned | Unassigned | Unassigned | Overlap (VC_995/VC_996) | Ref |
| 14585 | MW202600 | Unassigned | Unassigned | Unassigned | Overlap (VC_996/VC_1065) | Ref |
| 14591 | MW202615 | Unassigned | Unassigned | Unassigned | Outlier | Ref |
| 14593 | MW202621 | Unassigned | Unassigned | Unassigned | Outlier | Ref |
| 14595 | MW202627 | Unassigned | Unassigned | Unassigned | Singleton | Ref |
| 14598 | MW202640 | Unassigned | Unassigned | Unassigned | Overlap (VC_279/VC_529) | Ref |
| 14601 | MW202647 | Unassigned | Unassigned | Unassigned | Singleton | Ref |
| 14608 | MW202675 | Unassigned | Unassigned | Unassigned | Outlier | Ref |
| 14609 | MW202679 | Unassigned | Unassigned | Unassigned | Outlier | Ref |
| 14611 | MW202691 | Unassigned | Unassigned | Unassigned | Outlier | Ref |
| 14612 | MW202692 | Unassigned | Unassigned | Unassigned | Singleton | Ref |
| 14613 | MW202693 | Unassigned | Unassigned | Unassigned | Outlier | Ref |
| 14614 | MW202694 | Unassigned | Unassigned | Unassigned | Singleton | Ref |
| 14615 | MW202697 | Unassigned | Unassigned | Unassigned | Outlier | Ref |
| 14616 | MW202698 | Unassigned | Unassigned | Unassigned | Overlap (VC_199/VC_399) | Ref |
| 14617 | MW202701 | Unassigned | Unassigned | Unassigned | Outlier | Ref |
| 14619 | MW202710 | Unassigned | Unassigned | Unassigned | Outlier | Ref |
| 14621 | MW202713 | Unassigned | Unassigned | Unassigned | Outlier | Ref |
| 14623 | MW202722 | Unassigned | Unassigned | Unassigned | Overlap (VC_279/VC_529) | Ref |
| 14626 | MW202733 | Unassigned | Unassigned | Unassigned | Outlier | Ref |
| 14635 | MW202768 | Unassigned | Unassigned | Unassigned | Overlap (VC_199/VC_401) | Ref |
| 14637 | MW202779 | Unassigned | Unassigned | Unassigned | Overlap (VC_199/VC_399/VC_400) | Ref |
| 14638 | MW202783 | Unassigned | Unassigned | Unassigned | Singleton | Ref |
| 14641 | MW202795 | Unassigned | Unassigned | Unassigned | Outlier | Ref |
| 14643 | MW202801 | Unassigned | Unassigned | Unassigned | Singleton | Ref |
| 14644 | MW202803 | Unassigned | Unassigned | Unassigned | Singleton | Ref |
| 14646 | MW202816 | Unassigned | Unassigned | Unassigned | Outlier | Ref |
| 14651 | MW202828 | Unassigned | Unassigned | Unassigned | Outlier | Ref |
| 14659 | MW202869 | Unassigned | Unassigned | Unassigned | Outlier | Ref |
| 14661 | MW202882 | Unassigned | Unassigned | Unassigned | Singleton | Ref |
| 14664 | MW202893 | Unassigned | Unassigned | Unassigned | Overlap (VC_199/VC_399) | Ref |
| 14666 | MW202897 | Unassigned | Unassigned | Unassigned | Outlier | Ref |
| 14667 | MW202898 | Unassigned | Unassigned | Unassigned | Outlier | Ref |
| 14672 | MW221967 | Unassigned | Unassigned | Unassigned | Outlier | Ref |
| 14675 | MW239124 | Unassigned | Unassigned | Unassigned | Overlap (VC_286/VC_290/VC_491) | Ref |
| 14677 | MW247143 | Unassigned | Unassigned | Unassigned | Outlier | Ref |
| 14680 | MW247146 | Unassigned | Unassigned | Unassigned | Outlier | Ref |
| 14692 | MW258712 | Unassigned | Unassigned | Unassigned | Outlier | Ref |
| 14697 | MW273921 | Unassigned | Unassigned | Unassigned | Outlier | Ref |
| 14707 | MW286266 | Unassigned | Unassigned | Unassigned | Overlap (VC_155/VC_159) | Ref |
| 14708 | MW286267 | Unassigned | Unassigned | Unassigned | Overlap (VC_155/VC_159) | Ref |
| 14709 | MW286268 | Unassigned | Unassigned | Unassigned | Overlap (VC_155/VC_159) | Ref |
| 14715 | MW291017 | Unassigned | Unassigned | Unassigned | Outlier | Ref |
| 14727 | MW291508 | Unassigned | Unassigned | Unassigned | Outlier | Ref |
| 14738 | MW314849 | Unassigned | Unassigned | Unassigned | Overlap (VC_1095/VC_1096) | Ref |
| 14739 | MW314850 | Unassigned | Unassigned | Unassigned | Outlier | Ref |
| 14752 | MW317014 | Unassigned | Unassigned | Unassigned | Outlier | Ref |
| 14755 | MW325771 | Unassigned | Unassigned | Unassigned | Overlap (VC_286/VC_290/VC_491) | Ref |
| 14759 | MW334946 | Unassigned | Unassigned | Unassigned | Outlier | Ref |
| 14769 | MW344869 | Unassigned | Unassigned | Unassigned | Overlap (VC_286/VC_290/VC_491) | Ref |
| 14828 | MW358880 | Unassigned | Unassigned | Unassigned | Outlier | Ref |
| 14831 | MW358929 | Unassigned | Unassigned | Unassigned | Outlier | Ref |
| 14837 | MW364971 | Unassigned | Unassigned | Unassigned | Overlap (VC_13/VC_70) | Ref |
| 14838 | MW364972 | Unassigned | Unassigned | Unassigned | Overlap (VC_13/VC_70) | Ref |
| 14839 | MW364973 | Unassigned | Unassigned | Unassigned | Overlap (VC_13/VC_70) | Ref |
| 14840 | MW364974 | Unassigned | Unassigned | Unassigned | Overlap (VC_13/VC_70) | Ref |
| 14841 | MW364975 | Unassigned | Unassigned | Unassigned | Overlap (VC_13/VC_70) | Ref |
| 14872 | MW394388 | Unassigned | Unassigned | Unassigned | Overlap (VC_841/VC_842) | Ref |
| 14874 | MW394390 | Unassigned | Unassigned | Unassigned | Outlier | Ref |
| 14876 | MW394467 | Unassigned | Unassigned | Unassigned | Overlap (VC_101/VC_106) | Ref |
| 14890 | MW419775 | Unassigned | Unassigned | Unassigned | Overlap (VC_818/VC_821) | Ref |
| 14901 | MW423737 | Unassigned | Unassigned | Unassigned | Outlier | Ref |
| 14902 | MW423738 | Unassigned | Unassigned | Unassigned | Outlier | Ref |
| 14903 | MW423739 | Unassigned | Unassigned | Unassigned | Outlier | Ref |
| 14929 | MW451249 | Unassigned | Unassigned | Unassigned | Overlap (VC_615/VC_754) | Ref |
| 14937 | MW460247 | Unassigned | Unassigned | Unassigned | Outlier | Ref |
| 14939 | MW460249 | Unassigned | Unassigned | Unassigned | Outlier | Ref |
| 14943 | MW464860 | Unassigned | Unassigned | Unassigned | Outlier | Ref |
| 14945 | MW476490 | Unassigned | Unassigned | Unassigned | Outlier | Ref |
| 14949 | MW478290 | Unassigned | Unassigned | Unassigned | Overlap (VC_20/VC_542) | Ref |
| 14950 | MW478291 | Unassigned | Unassigned | Unassigned | Outlier | Ref |
| 14953 | MW495044 | Unassigned | Unassigned | Unassigned | Overlap (VC_841/VC_842) | Ref |
| 14972 | MW512570 | Unassigned | Unassigned | Unassigned | Outlier | Ref |
| 14975 | MW512573 | Unassigned | Unassigned | Unassigned | Overlap (VC_140/VC_649) | Ref |
| 14983 | MW526258 | Unassigned | Unassigned | Unassigned | Outlier | Ref |
| 15013 | MW560978 | Unassigned | Unassigned | Unassigned | Outlier | Ref |
| 15015 | MW570730 | Unassigned | Unassigned | Unassigned | Outlier | Ref |
| 15022 | MW582532 | Unassigned | Unassigned | Unassigned | Outlier | Ref |
| 15029 | MW584153 | Unassigned | Unassigned | Unassigned | Outlier | Ref |
| 15042 | MW584166 | Unassigned | Unassigned | Unassigned | Overlap (VC_512/VC_513) | Ref |
| 15043 | MW584167 | Unassigned | Unassigned | Unassigned | Outlier | Ref |
| 15050 | MW584174 | Unassigned | Unassigned | Unassigned | Overlap (VC_512/VC_513) | Ref |
| 15053 | MW584177 | Unassigned | Unassigned | Unassigned | Overlap (VC_553/VC_959) | Ref |
| 15059 | MW584183 | Unassigned | Unassigned | Unassigned | Overlap (VC_553/VC_959) | Ref |

| | | | | | | |
|---|---|---|---|---|---|---|
| 15061 | MW584185 | Unassigned | Unassigned | Unassigned | Outlier | Ref |
| 15062 | MW584186 | Unassigned | Unassigned | Unassigned | Overlap (VC_553/VC_959) | Ref |
| 15063 | MW584187 | Unassigned | Unassigned | Unassigned | Outlier | Ref |
| 15071 | MW584195 | Unassigned | Unassigned | Unassigned | Outlier | Ref |
| 15087 | MW584211 | Unassigned | Unassigned | Unassigned | Outlier | Ref |
| 15091 | MW589261 | Unassigned | Unassigned | Unassigned | Overlap (VC_668/VC_694) | Ref |
| 15125 | MW671054 | Unassigned | Unassigned | Unassigned | Overlap (VC_83/VC_85) | Ref |
| 15160 | MW712725 | Unassigned | Unassigned | Unassigned | Overlap (VC_1095/VC_1096) | Ref |
| 15183 | MW735835 | Unassigned | Unassigned | Unassigned | Outlier | Ref |
| 15196 | MW749006 | Unassigned | Unassigned | Unassigned | Outlier | Ref |
| 15198 | MW749008 | Unassigned | Unassigned | Unassigned | Outlier | Ref |
| 15199 | MW749009 | Unassigned | Unassigned | Unassigned | Outlier | Ref |
| 15205 | MW788655 | Unassigned | Unassigned | Unassigned | Singleton | Ref |
| 15206 | MW788656 | Unassigned | Unassigned | Unassigned | Outlier | Ref |
| 15266 | MW805363 | Unassigned | Unassigned | Unassigned | Outlier | Ref |
| 15267 | MW805364 | Unassigned | Unassigned | Unassigned | Outlier | Ref |
| 15314 | MW824390 | Unassigned | Unassigned | Unassigned | Overlap (VC_45/VC_47) | Ref |
| 15334 | MW824410 | Unassigned | Unassigned | Unassigned | Overlap (VC_1042/VC_1043) | Ref |
| 15335 | MW824411 | Unassigned | Unassigned | Unassigned | Overlap (VC_1042/VC_1043) | Ref |
| 15359 | MW825358 | Unassigned | Unassigned | Unassigned | Outlier | Ref |
| 15360 | MW831865 | Unassigned | Unassigned | Unassigned | Overlap (VC_75/VC_115) | Ref |
| 15377 | MW862989 | Unassigned | Unassigned | Unassigned | Overlap (VC_521/VC_548) | Ref |
| 15384 | MW865293 | Unassigned | Unassigned | Unassigned | Outlier | Ref |
| 15390 | MW865327 | Unassigned | Unassigned | Unassigned | Outlier | Ref |
| 15391 | MW865341 | Unassigned | Unassigned | Unassigned | Outlier | Ref |
| 15394 | MW879341 | Unassigned | Unassigned | Unassigned | Outlier | Ref |
| 15412 | MW924641 | Unassigned | Unassigned | Unassigned | Overlap (VC_1172/VC_1173) | Ref |
| 15425 | MW927512 | Unassigned | Unassigned | Unassigned | Overlap (VC_709/VC_713) | Ref |
| 15431 | MW927518 | Unassigned | Unassigned | Unassigned | Overlap (VC_709/VC_713) | Ref |
| 15441 | MW929183 | Unassigned | Unassigned | Unassigned | Outlier | Ref |
| 15447 | MW960033 | Unassigned | Unassigned | Unassigned | Outlier | Ref |
| 15451 | MW960037 | Unassigned | Unassigned | Unassigned | Outlier | Ref |
| 15453 | MW960043 | Unassigned | Unassigned | Unassigned | Overlap (VC_75/VC_115) | Ref |
| 15454 | MW965453 | Unassigned | Unassigned | Unassigned | Overlap (VC_71/VC_72) | Ref |
| 15456 | MW980061 | Unassigned | Unassigned | Unassigned | Outlier | Ref |
| 15461 | MW980066 | Unassigned | Unassigned | Unassigned | Overlap (VC_784/VC_786) | Ref |
| 15466 | MW980071 | Unassigned | Unassigned | Unassigned | Overlap (VC_784/VC_786) | Ref |
| 15467 | MW980072 | Unassigned | Unassigned | Unassigned | Overlap (VC_784/VC_786) | Ref |
| 15482 | MZ005681 | Unassigned | Unassigned | Unassigned | Overlap (VC_1241/VC_1242) | Ref |
| 15490 | MZ028627 | Unassigned | Unassigned | Unassigned | Outlier | Ref |
| 15498 | MZ073369 | Unassigned | Unassigned | Unassigned | Outlier | Ref |
| 15503 | MZ089730 | Unassigned | Unassigned | Unassigned | Outlier | Ref |
| 15508 | MZ089736 | Unassigned | Unassigned | Unassigned | Outlier | Ref |
| 15509 | MZ089737 | Unassigned | Unassigned | Unassigned | Overlap (VC_463/VC_609) | Ref |
| 15517 | MZ089749 | Unassigned | Unassigned | Unassigned | Overlap (VC_1119/VC_1121/VC_1321) | Ref |
| 15518 | MZ089750 | Unassigned | Unassigned | Unassigned | Overlap (VC_1056/VC_1057/VC_1061/VC_1151) | Ref |
| 15519 | MZ089751 | Unassigned | Unassigned | Unassigned | Singleton | Ref |
| 15521 | MZ089753 | Unassigned | Unassigned | Unassigned | Overlap (VC_1119/VC_1121) | Ref |
| 15522 | MZ089754 | Unassigned | Unassigned | Unassigned | Overlap (VC_1119/VC_1121/VC_1321) | Ref |
| 15525 | MZ089757 | Unassigned | Unassigned | Unassigned | Overlap (VC_1120/VC_1121) | Ref |
| 15526 | MZ089758 | Unassigned | Unassigned | Unassigned | Singleton | Ref |
| 15528 | MZ089760 | Unassigned | Unassigned | Unassigned | Singleton | Ref |
| 15530 | MZ089762 | Unassigned | Unassigned | Unassigned | Overlap (VC_1119/VC_1321) | Ref |
| 15531 | MZ089763 | Unassigned | Unassigned | Unassigned | Singleton | Ref |
| 15534 | MZ089766 | Unassigned | Unassigned | Unassigned | Outlier | Ref |
| 15544 | MZ089776 | Unassigned | Unassigned | Unassigned | Outlier | Ref |
| 15547 | MZ089779 | Unassigned | Unassigned | Unassigned | Overlap (VC_1114/VC_1136/VC_1318) | Ref |
| 15549 | MZ089781 | Unassigned | Unassigned | Unassigned | Singleton | Ref |
| 15550 | MZ089782 | Unassigned | Unassigned | Unassigned | Singleton | Ref |
| 15552 | MZ089784 | Unassigned | Unassigned | Unassigned | Outlier | Ref |
| 15561 | MZ089793 | Unassigned | Unassigned | Unassigned | Outlier | Ref |
| 15578 | MZ089810 | Unassigned | Unassigned | Unassigned | Singleton | Ref |
| 15584 | MZ099574 | Unassigned | Unassigned | Unassigned | Singleton | Ref |
| 15586 | MZ127829 | Unassigned | Unassigned | Unassigned | Outlier | Ref |
| 15595 | MZ150787 | Unassigned | Unassigned | Unassigned | Overlap (VC_1164/VC_1243) | Ref |
| 15597 | MZ150789 | Unassigned | Unassigned | Unassigned | Outlier | Ref |
| 15603 | MZ152915 | Unassigned | Unassigned | Unassigned | Overlap (VC_97/VC_99) | Ref |
| 15610 | MZ171369 | Unassigned | Unassigned | Unassigned | Outlier | Ref |
| 15620 | MZ171379 | Unassigned | Unassigned | Unassigned | Overlap (VC_1241/VC_1242) | Ref |
| 15626 | MZ188970 | Unassigned | Unassigned | Unassigned | Outlier | Ref |
| 15640 | MZ220765 | Unassigned | Unassigned | Unassigned | Overlap (VC_121/VC_124) | Ref |
| 15708 | MZ274310 | Unassigned | Unassigned | Unassigned | Outlier | Ref |
| 15714 | MZ308445 | Unassigned | Unassigned | Unassigned | Overlap (VC_257/VC_262) | Ref |
| 15725 | MZ322005 | Unassigned | Unassigned | Unassigned | Overlap (VC_227/VC_941) | Ref |
| 15751 | MZ326854 | Unassigned | Unassigned | Unassigned | Overlap (VC_75/VC_115) | Ref |
| 15755 | MZ326858 | Unassigned | Unassigned | Unassigned | Overlap (VC_75/VC_115) | Ref |
| 15757 | MZ326860 | Unassigned | Unassigned | Unassigned | Outlier | Ref |
| 15764 | MZ326867 | Unassigned | Unassigned | Unassigned | Outlier | Ref |
| 15768 | MZ333131 | Unassigned | Unassigned | Unassigned | Outlier | Ref |
| 15769 | MZ333132 | Unassigned | Unassigned | Unassigned | Outlier | Ref |
| 15770 | MZ333133 | Unassigned | Unassigned | Unassigned | Outlier | Ref |
| 15774 | MZ333456 | Unassigned | Unassigned | Unassigned | Overlap (VC_268/VC_864) | Ref |
| 15776 | MZ333458 | Unassigned | Unassigned | Unassigned | Overlap (VC_268/VC_864) | Ref |
| 15808 | MZ334521 | Unassigned | Unassigned | Unassigned | Outlier | Ref |
| 15809 | MZ334522 | Unassigned | Unassigned | Unassigned | Outlier | Ref |
| 15815 | MZ334528 | Unassigned | Unassigned | Unassigned | Outlier | Ref |
| 15818 | MZ343158 | Unassigned | Unassigned | Unassigned | Overlap (VC_524/VC_546) | Ref |
| 15823 | MZ348424 | Unassigned | Unassigned | Unassigned | Overlap (VC_286/VC_491) | Ref |
| 15832 | MZ355727 | Unassigned | Unassigned | Unassigned | Overlap (VC_1164/VC_1243) | Ref |
| 15834 | MZ357096 | Unassigned | Unassigned | Unassigned | Outlier | Ref |
| 15848 | MZ364229 | Unassigned | Unassigned | Unassigned | Overlap (VC_1151/VC_1433) | Ref |
| 15849 | MZ364230 | Unassigned | Unassigned | Unassigned | Overlap (VC_406/VC_1250) | Ref |
| 15856 | MZ364237 | Unassigned | Unassigned | Unassigned | Outlier | Ref |
| 15872 | MZ364253 | Unassigned | Unassigned | Unassigned | Outlier | Ref |
| 15900 | MZ364281 | Unassigned | Unassigned | Unassigned | Overlap (VC_1067/VC_1068) | Ref |
| 15906 | MZ364287 | Unassigned | Unassigned | Unassigned | Outlier | Ref |
| 15907 | MZ364288 | Unassigned | Unassigned | Unassigned | Overlap (VC_1151/VC_1433) | Ref |
| 15909 | MZ364290 | Unassigned | Unassigned | Unassigned | Singleton | Ref |
| 15911 | MZ364292 | Unassigned | Unassigned | Unassigned | Outlier | Ref |
| 16050 | MZ388555 | Unassigned | Unassigned | Unassigned | Overlap (VC_521/VC_548) | Ref |
| 16073 | MZ417316 | Unassigned | Unassigned | Unassigned | Overlap (VC_152/VC_153) | Ref |
| 16074 | MZ417317 | Unassigned | Unassigned | Unassigned | Overlap (VC_152/VC_153) | Ref |
| 16075 | MZ417318 | Unassigned | Unassigned | Unassigned | Overlap (VC_152/VC_153) | Ref |
| 16081 | MZ417324 | Unassigned | Unassigned | Unassigned | Overlap (VC_152/VC_153) | Ref |
| 16090 | MZ417333 | Unassigned | Unassigned | Unassigned | Overlap (VC_152/VC_153) | Ref |
| 16091 | MZ417334 | Unassigned | Unassigned | Unassigned | Overlap (VC_152/VC_153) | Ref |
| 16092 | MZ417335 | Unassigned | Unassigned | Unassigned | Overlap (VC_152/VC_153) | Ref |
| 16095 | MZ417338 | Unassigned | Unassigned | Unassigned | Overlap (VC_668/VC_694) | Ref |
| 16098 | MZ417341 | Unassigned | Unassigned | Unassigned | Overlap (VC_668/VC_694) | Ref |
| 16099 | MZ417342 | Unassigned | Unassigned | Unassigned | Overlap (VC_668/VC_694) | Ref |
| 16106 | MZ417349 | Unassigned | Unassigned | Unassigned | Overlap (VC_13/VC_70) | Ref |
| 16108 | MZ417351 | Unassigned | Unassigned | Unassigned | Overlap (VC_13/VC_70) | Ref |
| 16109 | MZ417352 | Unassigned | Unassigned | Unassigned | Overlap (VC_13/VC_70) | Ref |
| 16110 | MZ417353 | Unassigned | Unassigned | Unassigned | Overlap (VC_13/VC_70) | Ref |
| 16115 | MZ422438 | Unassigned | Unassigned | Unassigned | Outlier | Ref |
| 16129 | MZ428230 | Unassigned | Unassigned | Unassigned | Outlier | Ref |
| 16130 | MZ436628 | Unassigned | Unassigned | Unassigned | Outlier | Ref |
| 16153 | MZ447858 | Unassigned | Unassigned | Unassigned | Outlier | Ref |
| 16159 | MZ456038 | Unassigned | Unassigned | Unassigned | Overlap (VC_526/VC_554) | Ref |
| 16160 | MZ456039 | Unassigned | Unassigned | Unassigned | Overlap (VC_526/VC_554) | Ref |
| 16161 | MZ456352 | Unassigned | Unassigned | Unassigned | Outlier | Ref |
| 16165 | MZ474490 | Unassigned | Unassigned | Unassigned | Singleton | Ref |
| 16166 | MZ474491 | Unassigned | Unassigned | Unassigned | Outlier | Ref |
| 16176 | MZ488273 | Unassigned | Unassigned | Unassigned | Overlap (VC_14/VC_15) | Ref |
| 16178 | MZ496292 | Unassigned | Unassigned | Unassigned | Outlier | Ref |
| 16180 | MZ496294 | Unassigned | Unassigned | Unassigned | Outlier | Ref |
| 16253 | MZ501264 | Unassigned | Unassigned | Unassigned | Overlap (VC_101/VC_106) | Ref |
| 16260 | MZ501271 | Unassigned | Unassigned | Unassigned | Outlier | Ref |
| 16264 | MZ503612 | Unassigned | Unassigned | Unassigned | Outlier | Ref |
| 16270 | MZ506873 | Unassigned | Unassigned | Unassigned | Outlier | Ref |
| 16272 | MZ516527 | Unassigned | Unassigned | Unassigned | Outlier | Ref |
| 16281 | MZ560703 | Unassigned | Unassigned | Unassigned | Outlier | Ref |
| 16286 | MZ568829 | Unassigned | Unassigned | Unassigned | Outlier | Ref |
| 16292 | MZ571829 | Unassigned | Unassigned | Unassigned | Overlap (VC_491/VC_492) | Ref |
| 16300 | MZ574429 | Unassigned | Unassigned | Unassigned | Overlap (VC_977/VC_980) | Ref |
| 16301 | MZ574430 | Unassigned | Unassigned | Unassigned | Overlap (VC_977/VC_980) | Ref |
| 16302 | MZ574431 | Unassigned | Unassigned | Unassigned | Overlap (VC_977/VC_980) | Ref |
| 16303 | MZ574432 | Unassigned | Unassigned | Unassigned | Overlap (VC_977/VC_980) | Ref |
| 16304 | MZ574433 | Unassigned | Unassigned | Unassigned | Overlap (VC_977/VC_980) | Ref |
| 16324 | MZ612110 | Unassigned | Unassigned | Unassigned | Outlier | Ref |
| 16326 | MZ614453 | Unassigned | Unassigned | Unassigned | Outlier | Ref |
| 16345 | MZ622174 | Unassigned | Unassigned | Unassigned | Overlap (VC_521/VC_549) | Ref |
| 16346 | MZ622175 | Unassigned | Unassigned | Unassigned | Overlap (VC_1164/VC_1243) | Ref |
| 16355 | MZ622713 | Unassigned | Unassigned | Unassigned | Overlap (VC_442/VC_443) | Ref |
| 16365 | MZ634347 | Unassigned | Unassigned | Unassigned | Overlap (VC_491/VC_492) | Ref |
| 16367 | MZ634349 | Unassigned | Unassigned | Unassigned | Overlap (VC_491/VC_492) | Ref |
| 16370 | MZ643249 | Unassigned | Unassigned | Unassigned | Overlap (VC_932/VC_1050) | Ref |
| 16385 | MZ666130 | Unassigned | Unassigned | Unassigned | Overlap (VC_695/VC_696) | Ref |
| 16387 | MZ666939 | Unassigned | Unassigned | Unassigned | Singleton | Ref |
| 16390 | MZ669808 | Unassigned | Unassigned | Unassigned | Outlier | Ref |
| 16401 | MZ681514 | Unassigned | Unassigned | Unassigned | Outlier | Ref |
| 16402 | MZ681515 | Unassigned | Unassigned | Unassigned | Overlap (VC_526/VC_554) | Ref |
| 16404 | MZ681517 | Unassigned | Unassigned | Unassigned | Overlap (VC_526/VC_554) | Ref |
| 16421 | MZ727332 | Unassigned | Unassigned | Unassigned | Singleton | Ref |
| 16423 | MZ727334 | Unassigned | Unassigned | Unassigned | Outlier | Ref |
| 16446 | MZ818790 | Unassigned | Unassigned | Unassigned | Singleton | Ref |
| 16448 | MZ820084 | Unassigned | Unassigned | Unassigned | Overlap (VC_1164/VC_1243) | Ref |
| 16450 | MZ820086 | Unassigned | Unassigned | Unassigned | Overlap (VC_872/VC_873) | Ref |
| 16472 | MZ826699 | Unassigned | Unassigned | Unassigned | Overlap (VC_458/VC_586) | Ref |
| 16476 | MZ851152 | Unassigned | Unassigned | Unassigned | Overlap (VC_83/VC_85) | Ref |
| 16495 | MZ892993 | Unassigned | Unassigned | Unassigned | Outlier | Ref |
| 16496 | MZ923504 | Unassigned | Unassigned | Unassigned | Outlier | Ref |

| | | | | | | |
|---|---|---|---|---|---|---|
| 16516 | MZ969012 | Unassigned | Unassigned | Unassigned | Outlier | Ref |
| 16518 | MZ983385 | Unassigned | Unassigned | Unassigned | Overlap (VC_138/VC_146) | Ref |
| 16527 | OK018136 | Unassigned | Unassigned | Unassigned | Overlap (VC_75/VC_115) | Ref |
| 16535 | OK040780 | Unassigned | Unassigned | Unassigned | Overlap (VC_1035/VC_1039) | Ref |
| 16542 | OK040787 | Unassigned | Unassigned | Unassigned | Overlap (VC_1172/VC_1173) | Ref |
| 16553 | OK041467 | Unassigned | Unassigned | Unassigned | Outlier | Ref |
| 16557 | OK042081 | Unassigned | Unassigned | Unassigned | Outlier | Ref |
| 16562 | OK094664 | Unassigned | Unassigned | Unassigned | Overlap (VC_38/VC_812) | Ref |
| 16579 | OK149171 | Unassigned | Unassigned | Unassigned | Outlier | Ref |
| 16581 | OK149216 | Unassigned | Unassigned | Unassigned | Outlier | Ref |
| 16582 | OK169294 | Unassigned | Unassigned | Unassigned | Outlier | Ref |
| 16583 | OK169295 | Unassigned | Unassigned | Unassigned | Outlier | Ref |
| 16587 | OK181169 | Unassigned | Unassigned | Unassigned | Outlier | Ref |
| 16594 | OK210076 | Unassigned | Unassigned | Unassigned | Overlap (VC_286/VC_290/VC_491) | Ref |
| 16614 | OK272475 | Unassigned | Unassigned | Unassigned | Outlier | Ref |
| 16669 | OK349507 | Unassigned | Unassigned | Unassigned | Overlap (VC_615/VC_965) | Ref |
| 16677 | OK490361 | Unassigned | Unassigned | Unassigned | Outlier | Ref |
| 16693 | OK490409 | Unassigned | Unassigned | Unassigned | Outlier | Ref |
| 16703 | OK490419 | Unassigned | Unassigned | Unassigned | Outlier | Ref |
| 16704 | OK490420 | Unassigned | Unassigned | Unassigned | Outlier | Ref |
| 16705 | OK490421 | Unassigned | Unassigned | Unassigned | Outlier | Ref |
| 16746 | OK490462 | Unassigned | Unassigned | Unassigned | Overlap (VC_59/VC_245) | Ref |
| 16749 | OK490465 | Unassigned | Unassigned | Unassigned | Overlap (VC_59/VC_245) | Ref |
| 16752 | OK490468 | Unassigned | Unassigned | Unassigned | Overlap (VC_59/VC_245) | Ref |
| 16755 | OK490471 | Unassigned | Unassigned | Unassigned | Overlap (VC_59/VC_245) | Ref |
| 16758 | OK490474 | Unassigned | Unassigned | Unassigned | Overlap (VC_59/VC_245) | Ref |
| 16759 | OK490494 | Unassigned | Unassigned | Unassigned | Overlap (VC_75/VC_115) | Ref |
| 16791 | OK500002 | Unassigned | Unassigned | Unassigned | Outlier | Ref |
| 16801 | OK562670 | Unassigned | Unassigned | Unassigned | Outlier | Ref |
| 16803 | OK570185 | Unassigned | Unassigned | Unassigned | Overlap (VC_455/VC_1028) | Ref |
| 16805 | OK570374 | Unassigned | Unassigned | Unassigned | Overlap (VC_286/VC_290/VC_491) | Ref |
| 16807 | OK574338 | Unassigned | Unassigned | Unassigned | Overlap (VC_97/VC_99) | Ref |
| 16813 | OK597184 | Unassigned | Unassigned | Unassigned | Outlier | Ref |
| 16814 | OK624832 | Unassigned | Unassigned | Unassigned | Outlier | Ref |
| 16821 | OK632026 | Unassigned | Unassigned | Unassigned | Outlier | Ref |
| 16830 | OK665841 | Unassigned | Unassigned | Unassigned | Outlier | Ref |
| 16842 | OL348188 | Unassigned | Unassigned | Unassigned | Outlier | Ref |
| 16847 | OL362271 | Unassigned | Unassigned | Unassigned | Overlap (VC_455/VC_1028) | Ref |
| 16862 | OL412002 | Unassigned | Unassigned | Unassigned | Outlier | Ref |
| 16866 | OL449681 | Unassigned | Unassigned | Unassigned | Overlap (VC_458/VC_586) | Ref |
| 16869 | OL455886 | Unassigned | Unassigned | Unassigned | Overlap (VC_728/VC_730) | Ref |
| 16883 | OL455900 | Unassigned | Unassigned | Unassigned | Outlier | Ref |
| 16884 | OL473597 | Unassigned | Unassigned | Unassigned | Outlier | Ref |
| 16892 | OL512804 | Unassigned | Unassigned | Unassigned | Overlap (VC_32/VC_33) | Ref |
| 16893 | OL512807 | Unassigned | Unassigned | Unassigned | Outlier | Ref |
| 16900 | OL539441 | Unassigned | Unassigned | Unassigned | Overlap (VC_1347/VC_1348) | Ref |
| 16916 | OL539457 | Unassigned | Unassigned | Unassigned | Overlap (VC_458/VC_586) | Ref |
| 16927 | OL539469 | Unassigned | Unassigned | Unassigned | Outlier | Ref |
| 16942 | OL581612 | Unassigned | Unassigned | Unassigned | Overlap (VC_71/VC_74) | Ref |
| 16950 | OL617014 | Unassigned | Unassigned | Unassigned | Outlier | Ref |
| 16951 | OL617015 | Unassigned | Unassigned | Unassigned | Singleton | Ref |
| 16952 | OL617016 | Unassigned | Unassigned | Unassigned | Overlap (VC_1403/VC_1406) | Ref |
| 16953 | OL617017 | Unassigned | Unassigned | Unassigned | Overlap (VC_1403/VC_1406) | Ref |
| 16954 | OL617018 | Unassigned | Unassigned | Unassigned | Overlap (VC_1403/VC_1406) | Ref |
| 16955 | OL617019 | Unassigned | Unassigned | Unassigned | Overlap (VC_1403/VC_1406) | Ref |
| 16970 | OL617034 | Unassigned | Unassigned | Unassigned | Singleton | Ref |
| 16974 | OL617038 | Unassigned | Unassigned | Unassigned | Outlier | Ref |
| 16975 | OL617039 | Unassigned | Unassigned | Unassigned | Outlier | Ref |
| 16976 | OL617040 | Unassigned | Unassigned | Unassigned | Overlap (VC_932/VC_1050) | Ref |
| 17041 | OL770309 | Unassigned | Unassigned | Unassigned | Overlap (VC_286/VC_491) | Ref |
| 17046 | OL774570 | Unassigned | Unassigned | Unassigned | Outlier | Ref |
| 17052 | OL774871 | Unassigned | Unassigned | Unassigned | Outlier | Ref |
| 17055 | OL774874 | Unassigned | Unassigned | Unassigned | Outlier | Ref |
| 17056 | OL774875 | Unassigned | Unassigned | Unassigned | Outlier | Ref |
| 17061 | OL791267 | Unassigned | Unassigned | Unassigned | Outlier | Ref |
| 17062 | OL791268 | Unassigned | Unassigned | Unassigned | Overlap (VC_279/VC_529) | Ref |
| 17063 | OL799250 | Unassigned | Unassigned | Unassigned | Outlier | Ref |
| 17072 | OL800604 | Unassigned | Unassigned | Unassigned | Outlier | Ref |
| 17079 | OL828290 | Unassigned | Unassigned | Unassigned | Overlap (VC_286/VC_290/VC_491) | Ref |
| 17080 | OL828291 | Unassigned | Unassigned | Unassigned | Overlap (VC_286/VC_290/VC_491) | Ref |
| 17086 | OL849997 | Unassigned | Unassigned | Unassigned | Overlap (VC_286/VC_290/VC_491) | Ref |
| 17096 | OL944604 | Unassigned | Unassigned | Unassigned | Outlier | Ref |
| 17116 | OL963531 | Unassigned | Unassigned | Unassigned | Singleton | Ref |
| 17117 | OL963532 | Unassigned | Unassigned | Unassigned | Singleton | Ref |
| 17159 | OM046624 | Unassigned | Unassigned | Unassigned | Outlier | Ref |
| 17178 | OM112209 | Unassigned | Unassigned | Unassigned | Outlier | Ref |
| 17179 | OM112210 | Unassigned | Unassigned | Unassigned | Outlier | Ref |
| 17187 | OM158235 | Unassigned | Unassigned | Unassigned | Outlier | Ref |
| 17228 | OM334891 | Unassigned | Unassigned | Unassigned | Outlier | Ref |
| 17242 | OM373548 | Unassigned | Unassigned | Unassigned | Outlier | Ref |
| 17243 | OM373549 | Unassigned | Unassigned | Unassigned | Overlap (VC_668/VC_694) | Ref |
| 17253 | OM373559 | Unassigned | Unassigned | Unassigned | Outlier | Ref |
| 17254 | OM373560 | Unassigned | Unassigned | Unassigned | Overlap (VC_97/VC_99) | Ref |
| 17293 | OM416780 | Unassigned | Unassigned | Unassigned | Outlier | Ref |
| 17297 | OM418631 | Unassigned | Unassigned | Unassigned | Outlier | Ref |
| 17334 | OM522998 | Unassigned | Unassigned | Unassigned | Singleton | Ref |
| 17336 | OM523000 | Unassigned | Unassigned | Unassigned | Overlap (VC_400/VC_994/VC_1084) | Ref |
| 17345 | OM630587 | Unassigned | Unassigned | Unassigned | Overlap (VC_71/VC_74) | Ref |
| 17347 | OM630589 | Unassigned | Unassigned | Unassigned | Overlap (VC_71/VC_74) | Ref |
| 17360 | OM638088 | Unassigned | Unassigned | Unassigned | Outlier | Ref |
| 17367 | OM654374 | Unassigned | Unassigned | Unassigned | Outlier | Ref |
| 17368 | OM654375 | Unassigned | Unassigned | Unassigned | Outlier | Ref |
| 17369 | OM654376 | Unassigned | Unassigned | Unassigned | Outlier | Ref |
| 17370 | OM654377 | Unassigned | Unassigned | Unassigned | Outlier | Ref |
| 17371 | OM654378 | Unassigned | Unassigned | Unassigned | Outlier | Ref |
| 17374 | OM681334 | Unassigned | Unassigned | Unassigned | Outlier | Ref |
| 17380 | OM716000 | Unassigned | Unassigned | Unassigned | Singleton | Ref |
| 17381 | OM716001 | Unassigned | Unassigned | Unassigned | Singleton | Ref |
| 17382 | OM716002 | Unassigned | Unassigned | Unassigned | Singleton | Ref |
| 17383 | OM716005 | Unassigned | Unassigned | Unassigned | Overlap (VC_83/VC_85) | Ref |
| 17386 | OM735461 | Unassigned | Unassigned | Unassigned | Outlier | Ref |
| 17387 | OM735686 | Unassigned | Unassigned | Unassigned | Overlap (VC_75/VC_115) | Ref |
| 17388 | OM735687 | Unassigned | Unassigned | Unassigned | Overlap (VC_431/VC_743) | Ref |
| 17389 | OM735688 | Unassigned | Unassigned | Unassigned | Overlap (VC_75/VC_115) | Ref |
| 17390 | OM735689 | Unassigned | Unassigned | Unassigned | Overlap (VC_431/VC_743) | Ref |
| 17412 | OM869493 | Unassigned | Unassigned | Unassigned | Singleton | Ref |
| 17414 | OM869495 | Unassigned | Unassigned | Unassigned | Singleton | Ref |
| 17415 | OM869496 | Unassigned | Unassigned | Unassigned | Singleton | Ref |
| 17418 | OM869499 | Unassigned | Unassigned | Unassigned | Singleton | Ref |
| 17420 | OM869501 | Unassigned | Unassigned | Unassigned | Outlier | Ref |
| 17423 | OM869504 | Unassigned | Unassigned | Unassigned | Singleton | Ref |
| 17425 | OM869506 | Unassigned | Unassigned | Unassigned | Overlap (VC_1119/VC_1122/VC_1321) | Ref |
| 17426 | OM869507 | Unassigned | Unassigned | Unassigned | Outlier | Ref |
| 17427 | OM869508 | Unassigned | Unassigned | Unassigned | Overlap (VC_1119/VC_1122/VC_1321) | Ref |
| 17428 | OM869509 | Unassigned | Unassigned | Unassigned | Overlap (VC_1054/VC_1055) | Ref |
| 17430 | OM869511 | Unassigned | Unassigned | Unassigned | Singleton | Ref |
| 17434 | OM869515 | Unassigned | Unassigned | Unassigned | Outlier | Ref |
| 17437 | OM869518 | Unassigned | Unassigned | Unassigned | Overlap (VC_1114/VC_1136) | Ref |
| 17439 | OM869520 | Unassigned | Unassigned | Unassigned | Singleton | Ref |
| 17441 | OM869522 | Unassigned | Unassigned | Unassigned | Overlap (VC_1056/VC_1057/VC_1061) | Ref |
| 17442 | OM869523 | Unassigned | Unassigned | Unassigned | Outlier | Ref |
| 17449 | OM869530 | Unassigned | Unassigned | Unassigned | Outlier | Ref |
| 17455 | OM869536 | Unassigned | Unassigned | Unassigned | Outlier | Ref |
| 17458 | OM869539 | Unassigned | Unassigned | Unassigned | Singleton | Ref |
| 17460 | OM869541 | Unassigned | Unassigned | Unassigned | Outlier | Ref |
| 17470 | OM869551 | Unassigned | Unassigned | Unassigned | Outlier | Ref |
| 17473 | OM869554 | Unassigned | Unassigned | Unassigned | Overlap (VC_1130/VC_1135) | Ref |
| 17475 | OM869556 | Unassigned | Unassigned | Unassigned | Outlier | Ref |
| 17486 | OM869567 | Unassigned | Unassigned | Unassigned | Outlier | Ref |
| 17494 | OM869575 | Unassigned | Unassigned | Unassigned | Outlier | Ref |
| 17498 | OM869579 | Unassigned | Unassigned | Unassigned | Singleton | Ref |
| 17499 | OM869580 | Unassigned | Unassigned | Unassigned | Singleton | Ref |
| 17501 | OM869585 | Unassigned | Unassigned | Unassigned | Singleton | Ref |
| 17502 | OM869586 | Unassigned | Unassigned | Unassigned | Singleton | Ref |
| 17507 | OM869591 | Unassigned | Unassigned | Unassigned | Outlier | Ref |
| 17511 | OM869603 | Unassigned | Unassigned | Unassigned | Overlap (VC_1119/VC_1122/VC_1321) | Ref |
| 17513 | OM869605 | Unassigned | Unassigned | Unassigned | Outlier | Ref |
| 17515 | OM869607 | Unassigned | Unassigned | Unassigned | Singleton | Ref |
| 17516 | OM869608 | Unassigned | Unassigned | Unassigned | Singleton | Ref |
| 17517 | OM869609 | Unassigned | Unassigned | Unassigned | Singleton | Ref |
| 17519 | OM869611 | Unassigned | Unassigned | Unassigned | Singleton | Ref |
| 17523 | OM869625 | Unassigned | Unassigned | Unassigned | Singleton | Ref |
| 17526 | OM869628 | Unassigned | Unassigned | Unassigned | Overlap (VC_1119/VC_1121) | Ref |
| 17528 | OM869630 | Unassigned | Unassigned | Unassigned | Singleton | Ref |
| 17529 | OM869631 | Unassigned | Unassigned | Unassigned | Overlap (VC_1119/VC_1122/VC_1321) | Ref |
| 17534 | OM869636 | Unassigned | Unassigned | Unassigned | Singleton | Ref |
| 17544 | OM869646 | Unassigned | Unassigned | Unassigned | Outlier | Ref |
| 17545 | OM869647 | Unassigned | Unassigned | Unassigned | Singleton | Ref |
| 17547 | OM869649 | Unassigned | Unassigned | Unassigned | Outlier | Ref |
| 17549 | OM869651 | Unassigned | Unassigned | Unassigned | Outlier | Ref |
| 17554 | OM869656 | Unassigned | Unassigned | Unassigned | Singleton | Ref |
| 17555 | OM869659 | Unassigned | Unassigned | Unassigned | Outlier | Ref |
| 17556 | OM869660 | Unassigned | Unassigned | Unassigned | Outlier | Ref |
| 17557 | OM869661 | Unassigned | Unassigned | Unassigned | Outlier | Ref |
| 17558 | OM869662 | Unassigned | Unassigned | Unassigned | Outlier | Ref |
| 17561 | OM869665 | Unassigned | Unassigned | Unassigned | Outlier | Ref |
| 17563 | OM869667 | Unassigned | Unassigned | Unassigned | Outlier | Ref |
| 17568 | OM869678 | Unassigned | Unassigned | Unassigned | Singleton | Ref |
| 17570 | OM869680 | Unassigned | Unassigned | Unassigned | Singleton | Ref |
| 17571 | OM869681 | Unassigned | Unassigned | Unassigned | Singleton | Ref |
| 17572 | OM869682 | Unassigned | Unassigned | Unassigned | Outlier | Ref |
| 17573 | OM869683 | Unassigned | Unassigned | Unassigned | Outlier | Ref |

| | | | | | | |
|---|---|---|---|---|---|---|
| 17575 | OM869685 | Unassigned | Unassigned | Unassigned | Outlier | Ref |
| 17577 | OM869690 | Unassigned | Unassigned | Unassigned | Outlier | Ref |
| 17579 | OM869692 | Unassigned | Unassigned | Unassigned | Singleton | Ref |
| 17580 | OM869693 | Unassigned | Unassigned | Unassigned | Outlier | Ref |
| 17581 | OM869694 | Unassigned | Unassigned | Unassigned | Overlap (VC_1119/VC_1121/VC_1321) | Ref |
| 17582 | OM869695 | Unassigned | Unassigned | Unassigned | Singleton | Ref |
| 17586 | OM869699 | Unassigned | Unassigned | Unassigned | Outlier | Ref |
| 17592 | OM869705 | Unassigned | Unassigned | Unassigned | Singleton | Ref |
| 17597 | OM869710 | Unassigned | Unassigned | Unassigned | Outlier | Ref |
| 17601 | OM869714 | Unassigned | Unassigned | Unassigned | Outlier | Ref |
| 17602 | OM869715 | Unassigned | Unassigned | Unassigned | Singleton | Ref |
| 17607 | OM912978 | Unassigned | Unassigned | Unassigned | Overlap (VC_286/VC_290/VC_491) | Ref |
| 17608 | OM912979 | Unassigned | Unassigned | Unassigned | Outlier | Ref |
| 17609 | OM913583 | Unassigned | Unassigned | Unassigned | Overlap (VC_238/VC_240) | Ref |
| 17616 | OM913599 | Unassigned | Unassigned | Unassigned | Overlap (VC_455/VC_1028) | Ref |
| 17627 | OM962999 | Unassigned | Unassigned | Unassigned | Overlap (VC_71/VC_74) | Ref |
| 17646 | ON004963 | Unassigned | Unassigned | Unassigned | Overlap (VC_841/VC_842) | Ref |
| 17651 | ON014756 | Unassigned | Unassigned | Unassigned | Outlier | Ref |
| 17655 | ON032529 | Unassigned | Unassigned | Unassigned | Overlap (VC_36/VC_60) | Ref |
| 17658 | ON041214 | Unassigned | Unassigned | Unassigned | Outlier | Ref |
| 17662 | ON062054 | Unassigned | Unassigned | Unassigned | Overlap (VC_458/VC_586) | Ref |
| 17663 | ON067806 | Unassigned | Unassigned | Unassigned | Singleton | Ref |
| 17664 | ON075462 | Unassigned | Unassigned | Unassigned | Overlap (VC_905/VC_907) | Ref |
| 17665 | ON075799 | Unassigned | Unassigned | Unassigned | Overlap (VC_583/VC_851) | Ref |
| 17677 | ON081336 | Unassigned | Unassigned | Unassigned | Overlap (VC_872/VC_873) | Ref |
| 17680 | ON087563 | Unassigned | Unassigned | Unassigned | Outlier | Ref |
| 17683 | ON108641 | Unassigned | Unassigned | Unassigned | Overlap (VC_1172/VC_1173) | Ref |
| 17712 | ON113345 | Unassigned | Unassigned | Unassigned | Overlap (VC_14/VC_15) | Ref |
| 17718 | ON146294 | Unassigned | Unassigned | Unassigned | Outlier | Ref |
| 17756 | ON170044 | Unassigned | Unassigned | Unassigned | Outlier | Ref |
| 17757 | ON170045 | Unassigned | Unassigned | Unassigned | Outlier | Ref |
| 17758 | ON170047 | Unassigned | Unassigned | Unassigned | Outlier | Ref |
| 17759 | ON170048 | Unassigned | Unassigned | Unassigned | Outlier | Ref |
| 17774 | ON170063 | Unassigned | Unassigned | Unassigned | Outlier | Ref |
| 17775 | ON170065 | Unassigned | Unassigned | Unassigned | Outlier | Ref |
| 17794 | ON170085 | Unassigned | Unassigned | Unassigned | Outlier | Ref |
| 17844 | ON170138 | Unassigned | Unassigned | Unassigned | Outlier | Ref |
| 17846 | ON170140 | Unassigned | Unassigned | Unassigned | Outlier | Ref |
| 17882 | ON170177 | Unassigned | Unassigned | Unassigned | Outlier | Ref |
| 17888 | ON170183 | Unassigned | Unassigned | Unassigned | Outlier | Ref |
| 17896 | ON170191 | Unassigned | Unassigned | Unassigned | Overlap (VC_182/VC_187) | Ref |
| 17959 | ON170255 | Unassigned | Unassigned | Unassigned | Outlier | Ref |
| 17960 | ON170256 | Unassigned | Unassigned | Unassigned | Outlier | Ref |
| 17963 | ON170259 | Unassigned | Unassigned | Unassigned | Outlier | Ref |
| 18010 | ON170310 | Unassigned | Unassigned | Unassigned | Outlier | Ref |
| 18041 | ON170342 | Unassigned | Unassigned | Unassigned | Overlap (VC_182/VC_187) | Ref |
| 18044 | ON170345 | Unassigned | Unassigned | Unassigned | Outlier | Ref |
| 18080 | ON191531 | Unassigned | Unassigned | Unassigned | Outlier | Ref |
| 18081 | ON191532 | Unassigned | Unassigned | Unassigned | Outlier | Ref |
| 18085 | ON208831 | Unassigned | Unassigned | Unassigned | Overlap (VC_1164/VC_1243) | Ref |
| 18097 | ON212265 | Unassigned | Unassigned | Unassigned | Overlap (VC_286/VC_290/VC_491) | Ref |
| 18099 | ON212267 | Unassigned | Unassigned | Unassigned | Overlap (VC_286/VC_290/VC_491) | Ref |
| 18105 | ON245412 | Unassigned | Unassigned | Unassigned | Outlier | Ref |
| 18124 | ON260830 | Unassigned | Unassigned | Unassigned | Overlap (VC_526/VC_554) | Ref |
| 18130 | ON287368 | Unassigned | Unassigned | Unassigned | Outlier | Ref |
| 18133 | ON287371 | Unassigned | Unassigned | Unassigned | Outlier | Ref |
| 18144 | ON325435 | Unassigned | Unassigned | Unassigned | Overlap (VC_14/VC_107) | Ref |
| 18165 | ON392163 | Unassigned | Unassigned | Unassigned | Overlap (VC_1241/VC_1242) | Ref |
| 18166 | ON392164 | Unassigned | Unassigned | Unassigned | Overlap (VC_728/VC_730) | Ref |
| 18167 | ON392165 | Unassigned | Unassigned | Unassigned | Overlap (VC_728/VC_730) | Ref |
| 18169 | ON392167 | Unassigned | Unassigned | Unassigned | Outlier | Ref |
| 18188 | ON456341 | Unassigned | Unassigned | Unassigned | Overlap (VC_728/VC_730) | Ref |
| 18194 | ON456347 | Unassigned | Unassigned | Unassigned | Outlier | Ref |
| 18202 | ON456355 | Unassigned | Unassigned | Unassigned | Overlap (VC_526/VC_554) | Ref |
| 18222 | ON470581 | Unassigned | Unassigned | Unassigned | Overlap (VC_36/VC_60) | Ref |
| 18226 | ON470585 | Unassigned | Unassigned | Unassigned | Overlap (VC_36/VC_60) | Ref |
| 18228 | ON470587 | Unassigned | Unassigned | Unassigned | Overlap (VC_36/VC_60) | Ref |
| 18234 | ON470595 | Unassigned | Unassigned | Unassigned | Overlap (VC_36/VC_60) | Ref |
| 18238 | ON470599 | Unassigned | Unassigned | Unassigned | Outlier | Ref |
| 18241 | ON470602 | Unassigned | Unassigned | Unassigned | Overlap (VC_36/VC_60) | Ref |
| 18247 | ON470608 | Unassigned | Unassigned | Unassigned | Overlap (VC_244/VC_245) | Ref |
| 18248 | ON470609 | Unassigned | Unassigned | Unassigned | Overlap (VC_36/VC_60) | Ref |
| 18249 | ON470611 | Unassigned | Unassigned | Unassigned | Outlier | Ref |
| 18252 | ON470614 | Unassigned | Unassigned | Unassigned | Overlap (VC_36/VC_60) | Ref |
| 18254 | ON470616 | Unassigned | Unassigned | Unassigned | Overlap (VC_36/VC_60) | Ref |
| 18259 | ON470622 | Unassigned | Unassigned | Unassigned | Overlap (VC_244/VC_245) | Ref |
| 18288 | ON526983 | Unassigned | Unassigned | Unassigned | Overlap (VC_983/VC_984) | Ref |
| 18337 | ON529861 | Unassigned | Unassigned | Unassigned | Outlier | Ref |
| 18355 | ON550478 | Unassigned | Unassigned | Unassigned | Overlap (VC_14/VC_107) | Ref |
| 18362 | ON585039 | Unassigned | Unassigned | Unassigned | Overlap (VC_286/VC_290/VC_491) | Ref |
| 18371 | ON602731 | Unassigned | Unassigned | Unassigned | Overlap (VC_463/VC_609) | Ref |
| 18385 | ON602745 | Unassigned | Unassigned | Unassigned | Outlier | Ref |
| 18408 | ON604651 | Unassigned | Unassigned | Unassigned | Overlap (VC_83/VC_85) | Ref |
| 18409 | ON615601 | Unassigned | Unassigned | Unassigned | Outlier | Ref |
| 18410 | ON623732 | Unassigned | Unassigned | Unassigned | Overlap (VC_593/VC_597) | Ref |
| 18411 | ON624112 | Unassigned | Unassigned | Unassigned | Overlap (VC_455/VC_1028) | Ref |
| 18436 | ON645340 | Unassigned | Unassigned | Unassigned | Overlap (VC_1172/VC_1173) | Ref |
| 18473 | ON755180 | Unassigned | Unassigned | Unassigned | Outlier | Ref |
| 18484 | ON778006 | Unassigned | Unassigned | Unassigned | Overlap (VC_177/VC_296) | Ref |
| 18500 | ON854455 | Unassigned | Unassigned | Unassigned | Outlier | Ref |
| 18526 | ON872163 | Unassigned | Unassigned | Unassigned | Outlier | Ref |
| 18531 | ON886116 | Unassigned | Unassigned | Unassigned | Outlier | Ref |
| 18540 | ON920540 | Unassigned | Unassigned | Unassigned | Outlier | Ref |
| 18555 | ON950054 | Unassigned | Unassigned | Unassigned | Outlier | Ref |
| 18567 | ON970570 | Unassigned | Unassigned | Unassigned | Overlap (VC_1164/VC_1243) | Ref |
| 18612 | ON970615 | Unassigned | Unassigned | Unassigned | Outlier | Ref |
| 18614 | ON970617 | Unassigned | Unassigned | Unassigned | Overlap (VC_872/VC_873) | Ref |
| 18625 | ON981384 | Unassigned | Unassigned | Unassigned | Outlier | Ref |
| 18654 | OP012469 | Unassigned | Unassigned | Unassigned | Outlier | Ref |
| 18656 | OP018674 | Unassigned | Unassigned | Unassigned | Overlap (VC_695/VC_696) | Ref |
| 18682 | OP066531 | Unassigned | Unassigned | Unassigned | Outlier | Ref |
| 18684 | OP068330 | Unassigned | Unassigned | Unassigned | Overlap (VC_1164/VC_1243) | Ref |
| 18700 | OP079919 | Unassigned | Unassigned | Unassigned | Overlap (VC_442/VC_443) | Ref |
| 18701 | OP087422 | Unassigned | Unassigned | Unassigned | Overlap (VC_71/VC_72) | Ref |
| 18702 | OP087423 | Unassigned | Unassigned | Unassigned | Overlap (VC_71/VC_72) | Ref |
| 18716 | OP120783 | Unassigned | Unassigned | Unassigned | Overlap (VC_584/VC_586) | Ref |
| 18718 | OP123707 | Unassigned | Unassigned | Unassigned | Overlap (VC_442/VC_443) | Ref |
| 18720 | OP125547 | Unassigned | Unassigned | Unassigned | Outlier | Ref |
| 18737 | OP142323 | Unassigned | Unassigned | Unassigned | Overlap (VC_14/VC_107) | Ref |
| 18744 | OP172869 | Unassigned | Unassigned | Unassigned | Overlap (VC_1165/VC_1166) | Ref |
| 18746 | OP172871 | Unassigned | Unassigned | Unassigned | Overlap (VC_1172/VC_1173) | Ref |
| 18758 | OP209175 | Unassigned | Unassigned | Unassigned | Outlier | Ref |
| 18760 | OP209177 | Unassigned | Unassigned | Unassigned | Singleton | Ref |
| 18761 | OP209178 | Unassigned | Unassigned | Unassigned | Outlier | Ref |
| 18782 | OP296522 | Unassigned | Unassigned | Unassigned | Outlier | Ref |
| 18785 | OP297179 | Unassigned | Unassigned | Unassigned | Overlap (VC_97/VC_99) | Ref |
| 18788 | OP297530 | Unassigned | Unassigned | Unassigned | Overlap (VC_238/VC_240) | Ref |
| 18803 | OP297545 | Unassigned | Unassigned | Unassigned | Outlier | Ref |
| 18835 | OP323071 | Unassigned | Unassigned | Unassigned | Overlap (VC_257/VC_262) | Ref |
| 18836 | OP323072 | Unassigned | Unassigned | Unassigned | Overlap (VC_257/VC_262) | Ref |
| 18837 | OP323073 | Unassigned | Unassigned | Unassigned | Overlap (VC_257/VC_262) | Ref |
| 18838 | OP323074 | Unassigned | Unassigned | Unassigned | Overlap (VC_257/VC_262) | Ref |
| 18839 | OP323075 | Unassigned | Unassigned | Unassigned | Overlap (VC_257/VC_262) | Ref |
| 18840 | OP323076 | Unassigned | Unassigned | Unassigned | Overlap (VC_257/VC_262) | Ref |
| 18841 | OP323077 | Unassigned | Unassigned | Unassigned | Overlap (VC_257/VC_262) | Ref |
| 18843 | OP328421 | Unassigned | Unassigned | Unassigned | Outlier | Ref |
| 18848 | OP341623 | Unassigned | Unassigned | Unassigned | Outlier | Ref |
| 18854 | OP341629 | Unassigned | Unassigned | Unassigned | Outlier | Ref |
| 18866 | OP380492 | Unassigned | Unassigned | Unassigned | Overlap (VC_87/VC_88) | Ref |
| 18867 | OP380511 | Unassigned | Unassigned | Unassigned | Outlier | Ref |
| 18871 | OP413827 | Unassigned | Unassigned | Unassigned | Overlap (VC_841/VC_842) | Ref |
| 18875 | OP413831 | Unassigned | Unassigned | Unassigned | Overlap (VC_841/VC_842) | Ref |
| 18879 | OP433492 | Unassigned | Unassigned | Unassigned | Overlap (VC_818/VC_820) | Ref |
| 18887 | OP434446 | Unassigned | Unassigned | Unassigned | Outlier | Ref |
| 18890 | OP434449 | Unassigned | Unassigned | Unassigned | Outlier | Ref |
| 18905 | OP434518 | Unassigned | Unassigned | Unassigned | Outlier | Ref |
| 18910 | OP481051 | Unassigned | Unassigned | Unassigned | Outlier | Ref |
| 18916 | OP490597 | Unassigned | Unassigned | Unassigned | Overlap (VC_82/VC_619) | Ref |
| 18943 | OP503989 | Unassigned | Unassigned | Unassigned | Overlap (VC_710/VC_713) | Ref |
| 18944 | OP503990 | Unassigned | Unassigned | Unassigned | Overlap (VC_710/VC_713) | Ref |
| 18945 | OP503991 | Unassigned | Unassigned | Unassigned | Overlap (VC_710/VC_713) | Ref |
| 18951 | OP535465 | Unassigned | Unassigned | Unassigned | Outlier | Ref |
| 18979 | OP559179 | Unassigned | Unassigned | Unassigned | Outlier | Ref |
| 18993 | OP617331 | Unassigned | Unassigned | Unassigned | Overlap (VC_286/VC_491) | Ref |
| 18995 | OP617741 | Unassigned | Unassigned | Unassigned | Outlier | Ref |
| 19003 | OP650935 | Unassigned | Unassigned | Unassigned | Outlier | Ref |
| 19004 | OP650936 | Unassigned | Unassigned | Unassigned | Overlap (VC_38/VC_812) | Ref |
| 19007 | OP714164 | Unassigned | Unassigned | Unassigned | Outlier | Ref |
| 19011 | OU343168 | Unassigned | Unassigned | Unassigned | Overlap (VC_442/VC_443) | Ref |
| 19016 | OU509537 | Unassigned | Unassigned | Unassigned | Overlap (VC_593/VC_597) | Ref |
| 19034 | OV696620 | Unassigned | Unassigned | Unassigned | Overlap (VC_702/VC_704) | Ref |
| 19036 | OV696622 | Unassigned | Unassigned | Unassigned | Outlier | Ref |
| 19048 | OW991346 | Unassigned | Unassigned | Unassigned | Overlap (VC_83/VC_85) | Ref |
| 19049 | OX001577 | Unassigned | Unassigned | Unassigned | Overlap (VC_83/VC_85) | Ref |
| 19052 | OX090892 | Unassigned | Unassigned | Unassigned | Overlap (VC_83/VC_85) | Ref |
| 19054 | OX241423 | Unassigned | Unassigned | Unassigned | Overlap (VC_494/VC_495) | Ref |
| 19069 | OX241442 | Unassigned | Unassigned | Unassigned | Overlap (VC_494/VC_495) | Ref |
| 19070 | OX241443 | Unassigned | Unassigned | Unassigned | Outlier | Ref |
| 19071 | OX241444 | Unassigned | Unassigned | Unassigned | Overlap (VC_494/VC_495) | Ref |
| 19078 | OX241464 | Unassigned | Unassigned | Unassigned | Overlap (VC_494/VC_495) | Ref |
| 19083 | OX241555 | Unassigned | Unassigned | Unassigned | Overlap (VC_1042/VC_1043) | Ref |
| 19101 | OX241576 | Unassigned | Unassigned | Unassigned | Overlap (VC_494/VC_495) | Ref |

| | | | | | | |
|---|---|---|---|---|---|---|
| 19138 | OX335406 | Unassigned | Unassigned | Unassigned | Overlap (VC_841/VC_842) | Ref |
| 19174 | PMULEFTEN | Unassigned | Unassigned | Unassigned | Outlier | Ref |
| 19177 | S1-wms-combined_100430 | Unassigned | Unassigned | Unassigned | Overlap (VC_711/VC_715) | ME |
| 19178 | S1-wms-combined_102801 | Unassigned | Unassigned | Unassigned | Outlier | ME |
| 19180 | S1-wms-combined_107766 | Unassigned | Unassigned | Unassigned | Overlap (VC_652/VC_1481) | ME |
| 19181 | S1-wms-combined_116938 | Unassigned | Unassigned | Unassigned | Outlier | ME |
| 19184 | S1-wms-combined_131270 | Unassigned | Unassigned | Unassigned | Outlier | ME |
| 19186 | S1-wms-combined_136097 | Unassigned | Unassigned | Unassigned | Outlier | ME |
| 19187 | S1-wms-combined_143459 | Unassigned | Unassigned | Unassigned | Outlier | ME |
| 19189 | S1-wms-combined_153670 | Unassigned | Unassigned | Unassigned | Overlap (VC_1245/VC_1487) | ME |
| 19193 | S1-wms-combined_164894 | Unassigned | Unassigned | Unassigned | Outlier | ME |
| 19194 | S1-wms-combined_166392 | Unassigned | Unassigned | Unassigned | Overlap (VC_893/VC_894) | ME |
| 19195 | S1-wms-combined_169788 | Unassigned | Unassigned | Unassigned | Singleton | ME |
| 19197 | S1-wms-combined_17651 | Unassigned | Unassigned | Unassigned | Overlap (VC_1490/VC_1491) | ME |
| 19199 | S1-wms-combined_186878 | Unassigned | Unassigned | Unassigned | Outlier | ME |
| 19202 | S1-wms-combined_212892 | Unassigned | Unassigned | Unassigned | Outlier | ME |
| 19205 | S1-wms-combined_227003 | Unassigned | Unassigned | Unassigned | Outlier | ME |
| 19206 | S1-wms-combined_227917 | Unassigned | Unassigned | Unassigned | Outlier | ME |
| 19207 | S1-wms-combined_231234 | Unassigned | Unassigned | Unassigned | Singleton | ME |
| 19208 | S1-wms-combined_231411 | Unassigned | Unassigned | Unassigned | Singleton | ME |
| 19212 | S1-wms-combined_252439 | Unassigned | Unassigned | Unassigned | Outlier | ME |
| 19214 | S1-wms-combined_279492 | Unassigned | Unassigned | Unassigned | Outlier | ME |
| 19215 | S1-wms-combined_285012 | Unassigned | Unassigned | Unassigned | Overlap (VC_711/VC_715) | ME |
| 19219 | S1-wms-combined_308848 | Unassigned | Unassigned | Unassigned | Outlier | ME |
| 19222 | S1-wms-combined_32863 | Unassigned | Unassigned | Unassigned | Singleton | ME |
| 19223 | S1-wms-combined_330329 | Unassigned | Unassigned | Unassigned | Overlap (VC_678/VC_679) | ME |
| 19228 | S1-wms-combined_351889 | Unassigned | Unassigned | Unassigned | Overlap (VC_817/VC_1210) | ME |
| 19233 | S1-wms-combined_373308 | Unassigned | Unassigned | Unassigned | Overlap (VC_1483/VC_1490) | ME |
| 19234 | S1-wms-combined_374219 | Unassigned | Unassigned | Unassigned | Overlap (VC_651/VC_736) | ME |
| 19240 | S1-wms-combined_52708 | Unassigned | Unassigned | Unassigned | Overlap (VC_1221/VC_1237) | ME |
| 19241 | S1-wms-combined_57370 | Unassigned | Unassigned | Unassigned | Outlier | ME |
| 19245 | S1-wms-combined_92523 | Unassigned | Unassigned | Unassigned | Outlier | ME |
| 19246 | S1-wms-combined_94988 | Unassigned | Unassigned | Unassigned | Outlier | ME |
| 19249 | S1-wms_106115 | Unassigned | Unassigned | Unassigned | Outlier | ME |
| 19251 | S1-wms_153615 | Unassigned | Unassigned | Unassigned | Overlap (VC_1245/VC_1487) | ME |
| 19254 | S1-wms_208875 | Unassigned | Unassigned | Unassigned | Outlier | ME |
| 19256 | S1-wms_223620 | Unassigned | Unassigned | Unassigned | Overlap (VC_893/VC_894) | ME |
| 19258 | S1-wms_236105 | Unassigned | Unassigned | Unassigned | Outlier | ME |
| 19260 | S1-wms_265285 | Unassigned | Unassigned | Unassigned | Overlap (VC_138/VC_146/VC_147) | ME |
| 19261 | S1-wms_29288 | Unassigned | Unassigned | Unassigned | Outlier | ME |
| 19264 | S1-wms_32878 | Unassigned | Unassigned | Unassigned | Singleton | ME |
| 19265 | S1-wms_388292 | Unassigned | Unassigned | Unassigned | Singleton | ME |
| 19266 | S1-wms_394571 | Unassigned | Unassigned | Unassigned | Outlier | ME |
| 19268 | S11-wms-combined_101337 | Unassigned | Unassigned | Unassigned | Outlier | ME |
| 19269 | S11-wms-combined_112147 | Unassigned | Unassigned | Unassigned | Outlier | ME |
| 19270 | S11-wms-combined_112733 | Unassigned | Unassigned | Unassigned | Outlier | ME |
| 19271 | S11-wms-combined_116589 | Unassigned | Unassigned | Unassigned | Outlier | ME |
| 19272 | S11-wms-combined_117756 | Unassigned | Unassigned | Unassigned | Singleton | ME |
| 19274 | S11-wms-combined_120257 | Unassigned | Unassigned | Unassigned | Overlap (VC_711/VC_715) | ME |
| 19277 | S11-wms-combined_14636 | Unassigned | Unassigned | Unassigned | Outlier | ME |
| 19279 | S11-wms-combined_168048 | Unassigned | Unassigned | Unassigned | Outlier | ME |
| 19282 | S11-wms-combined_179605 | Unassigned | Unassigned | Unassigned | Overlap (VC_893/VC_894/VC_1489/VC_1494) | ME |
| 19289 | S11-wms-combined_215723 | Unassigned | Unassigned | Unassigned | Outlier | ME |
| 19292 | S11-wms-combined_226165 | Unassigned | Unassigned | Unassigned | Overlap (VC_678/VC_679) | ME |
| 19293 | S11-wms-combined_236214 | Unassigned | Unassigned | Unassigned | Outlier | ME |
| 19295 | S11-wms-combined_265453 | Unassigned | Unassigned | Unassigned | Singleton | ME |
| 19298 | S11-wms-combined_282227 | Unassigned | Unassigned | Unassigned | Outlier | ME |
| 19302 | S11-wms-combined_320710 | Unassigned | Unassigned | Unassigned | Overlap (VC_342/VC_1215) | ME |
| 19303 | S11-wms-combined_321938 | Unassigned | Unassigned | Unassigned | Overlap (VC_678/VC_679) | ME |
| 19304 | S11-wms-combined_329493 | Unassigned | Unassigned | Unassigned | Outlier | ME |
| 19311 | S11-wms-combined_63818 | Unassigned | Unassigned | Unassigned | Outlier | ME |
| 19312 | S11-wms-combined_64392 | Unassigned | Unassigned | Unassigned | Overlap (VC_678/VC_679) | ME |
| 19322 | S11-wms_32691 | Unassigned | Unassigned | Unassigned | Singleton | ME |
| 19325 | S11-wms_6203 | Unassigned | Unassigned | Unassigned | Singleton | ME |
| 19326 | S11-wms_87293 | Unassigned | Unassigned | Unassigned | Outlier | ME |
| 19329 | S12-wms-combined_117447 | Unassigned | Unassigned | Unassigned | Singleton | SHHC |
| 19330 | S12-wms-combined_117628 | Unassigned | Unassigned | Unassigned | Overlap (VC_20/VC_542) | SHHC |
| 19334 | S12-wms-combined_132907 | Unassigned | Unassigned | Unassigned | Overlap (VC_138/VC_146) | SHHC |
| 19335 | S12-wms-combined_133608 | Unassigned | Unassigned | Unassigned | Overlap (VC_1490/VC_1491) | SHHC |
| 19337 | S12-wms-combined_148741 | Unassigned | Unassigned | Unassigned | Overlap (VC_1103/VC_1240) | SHHC |
| 19338 | S12-wms-combined_151843 | Unassigned | Unassigned | Unassigned | Outlier | SHHC |
| 19341 | S12-wms-combined_164656 | Unassigned | Unassigned | Unassigned | Outlier | SHHC |
| 19345 | S12-wms-combined_184338 | Unassigned | Unassigned | Unassigned | Overlap (VC_652/VC_894) | SHHC |
| 19346 | S12-wms-combined_189323 | Unassigned | Unassigned | Unassigned | Overlap (VC_678/VC_679) | SHHC |
| 19347 | S12-wms-combined_190114 | Unassigned | Unassigned | Unassigned | Outlier | SHHC |
| 19351 | S12-wms-combined_21504 | Unassigned | Unassigned | Unassigned | Outlier | SHHC |
| 19352 | S12-wms-combined_223043 | Unassigned | Unassigned | Unassigned | Outlier | SHHC |
| 19357 | S12-wms-combined_25459 | Unassigned | Unassigned | Unassigned | Overlap (VC_652/VC_1481) | SHHC |
| 19362 | S12-wms-combined_264077 | Unassigned | Unassigned | Unassigned | Outlier | SHHC |
| 19365 | S12-wms-combined_270839 | Unassigned | Unassigned | Unassigned | Overlap (VC_1103/VC_1240) | SHHC |
| 19367 | S12-wms-combined_293354 | Unassigned | Unassigned | Unassigned | Outlier | SHHC |
| 19368 | S12-wms-combined_297545 | Unassigned | Unassigned | Unassigned | Overlap (VC_918/VC_1456) | SHHC |
| 19371 | S12-wms-combined_302876 | Unassigned | Unassigned | Unassigned | Outlier | SHHC |
| 19372 | S12-wms-combined_313953 | Unassigned | Unassigned | Unassigned | Outlier | SHHC |
| 19373 | S12-wms-combined_31761 | Unassigned | Unassigned | Unassigned | Overlap (VC_877/VC_1413) | SHHC |
| 19374 | S12-wms-combined_320262 | Unassigned | Unassigned | Unassigned | Overlap (VC_652/VC_1481) | SHHC |
| 19378 | S12-wms-combined_325418 | Unassigned | Unassigned | Unassigned | Outlier | SHHC |
| 19379 | S12-wms-combined_328554 | Unassigned | Unassigned | Unassigned | Outlier | SHHC |
| 19386 | S12-wms-combined_359190 | Unassigned | Unassigned | Unassigned | Outlier | SHHC |
| 19387 | S12-wms-combined_368190 | Unassigned | Unassigned | Unassigned | Overlap (VC_918/VC_1456) | SHHC |
| 19389 | S12-wms-combined_368562 | Unassigned | Unassigned | Unassigned | Outlier | SHHC |
| 19390 | S12-wms-combined_371541 | Unassigned | Unassigned | Unassigned | Outlier | SHHC |
| 19393 | S12-wms-combined_385270 | Unassigned | Unassigned | Unassigned | Overlap (VC_630/VC_642) | SHHC |
| 19394 | S12-wms-combined_390985 | Unassigned | Unassigned | Unassigned | Singleton | SHHC |
| 19399 | S12-wms-combined_48648 | Unassigned | Unassigned | Unassigned | Outlier | SHHC |
| 19401 | S12-wms-combined_52101 | Unassigned | Unassigned | Unassigned | Overlap (VC_138/VC_146) | SHHC |
| 19402 | S12-wms-combined_63056 | Unassigned | Unassigned | Unassigned | Singleton | SHHC |
| 19403 | S12-wms-combined_63649 | Unassigned | Unassigned | Unassigned | Outlier | SHHC |
| 19406 | S12-wms-combined_75733 | Unassigned | Unassigned | Unassigned | Overlap (VC_1244/VC_1245) | SHHC |
| 19411 | S12-wms_135291 | Unassigned | Unassigned | Unassigned | Outlier | SHHC |
| 19413 | S12-wms_161239 | Unassigned | Unassigned | Unassigned | Overlap (VC_138/VC_146/VC_147) | SHHC |
| 19415 | S12-wms_17950 | Unassigned | Unassigned | Unassigned | Outlier | SHHC |
| 19416 | S12-wms_184276 | Unassigned | Unassigned | Unassigned | Overlap (VC_735/VC_915) | SHHC |
| 19418 | S12-wms_202803 | Unassigned | Unassigned | Unassigned | Outlier | SHHC |
| 19419 | S12-wms_223235 | Unassigned | Unassigned | Unassigned | Singleton | SHHC |
| 19420 | S12-wms_235125 | Unassigned | Unassigned | Unassigned | Outlier | SHHC |
| 19421 | S12-wms_336780 | Unassigned | Unassigned | Unassigned | Singleton | SHHC |
| 19422 | S12-wms_352846 | Unassigned | Unassigned | Unassigned | Singleton | SHHC |
| 19430 | S17-wms-combined_106262 | Unassigned | Unassigned | Unassigned | Overlap (VC_711/VC_715) | ME |
| 19431 | S17-wms-combined_106940 | Unassigned | Unassigned | Unassigned | Outlier | ME |
| 19437 | S17-wms-combined_140517 | Unassigned | Unassigned | Unassigned | Outlier | ME |
| 19438 | S17-wms-combined_146050 | Unassigned | Unassigned | Unassigned | Outlier | ME |
| 19444 | S17-wms-combined_171305 | Unassigned | Unassigned | Unassigned | Outlier | ME |
| 19446 | S17-wms-combined_179363 | Unassigned | Unassigned | Unassigned | Overlap (VC_630/VC_642) | ME |
| 19448 | S17-wms-combined_185239 | Unassigned | Unassigned | Unassigned | Outlier | ME |
| 19460 | S17-wms-combined_253499 | Unassigned | Unassigned | Unassigned | Overlap (VC_833/VC_1015) | ME |
| 19461 | S17-wms-combined_258087 | Unassigned | Unassigned | Unassigned | Overlap (VC_138/VC_146) | ME |
| 19465 | S17-wms-combined_39687 | Unassigned | Unassigned | Unassigned | Overlap (VC_733/VC_893/VC_1494) | ME |
| 19467 | S17-wms-combined_56006 | Unassigned | Unassigned | Unassigned | Overlap (VC_607/VC_1238) | ME |
| 19468 | S17-wms-combined_56061 | Unassigned | Unassigned | Unassigned | Overlap (VC_1221/VC_1237) | ME |
| 19469 | S17-wms-combined_66123 | Unassigned | Unassigned | Unassigned | Overlap (VC_138/VC_146/VC_148) | ME |
| 19471 | S17-wms-combined_72837 | Unassigned | Unassigned | Unassigned | Outlier | ME |
| 19472 | S17-wms-combined_75892 | Unassigned | Unassigned | Unassigned | Overlap (VC_138/VC_146/VC_148) | ME |
| 19477 | S17-wms_106671 | Unassigned | Unassigned | Unassigned | Overlap (VC_138/VC_146) | ME |
| 19478 | S17-wms_116248 | Unassigned | Unassigned | Unassigned | Overlap (VC_138/VC_146/VC_147) | ME |
| 19480 | S17-wms_235536 | Unassigned | Unassigned | Unassigned | Outlier | ME |
| 19483 | S17-wms_29082 | Unassigned | Unassigned | Unassigned | Overlap (VC_138/VC_146/VC_147) | ME |
| 19485 | S2-wms-combined_110215 | Unassigned | Unassigned | Unassigned | Outlier | SHHC |
| 19487 | S2-wms-combined_115352 | Unassigned | Unassigned | Unassigned | Singleton | SHHC |
| 19489 | S2-wms-combined_122457 | Unassigned | Unassigned | Unassigned | Overlap (VC_138/VC_146/VC_148) | SHHC |
| 19491 | S2-wms-combined_126440 | Unassigned | Unassigned | Unassigned | Outlier | SHHC |
| 19492 | S2-wms-combined_127080 | Unassigned | Unassigned | Unassigned | Outlier | SHHC |
| 19495 | S2-wms-combined_135014 | Unassigned | Unassigned | Unassigned | Overlap (VC_138/VC_146) | SHHC |
| 19500 | S2-wms-combined_143737 | Unassigned | Unassigned | Unassigned | Outlier | SHHC |
| 19501 | S2-wms-combined_147065 | Unassigned | Unassigned | Unassigned | Overlap (VC_678/VC_679) | SHHC |
| 19503 | S2-wms-combined_158821 | Unassigned | Unassigned | Unassigned | Overlap (VC_422/VC_1413) | SHHC |
| 19504 | S2-wms-combined_159670 | Unassigned | Unassigned | Unassigned | Outlier | SHHC |
| 19507 | S2-wms-combined_170845 | Unassigned | Unassigned | Unassigned | Outlier | SHHC |
| 19512 | S2-wms-combined_181391 | Unassigned | Unassigned | Unassigned | Outlier | SHHC |
| 19513 | S2-wms-combined_181849 | Unassigned | Unassigned | Unassigned | Overlap (VC_1221/VC_1237) | SHHC |
| 19514 | S2-wms-combined_185494 | Unassigned | Unassigned | Unassigned | Outlier | SHHC |
| 19515 | S2-wms-combined_190624 | Unassigned | Unassigned | Unassigned | Outlier | SHHC |
| 19516 | S2-wms-combined_191613 | Unassigned | Unassigned | Unassigned | Outlier | SHHC |
| 19519 | S2-wms-combined_199384 | Unassigned | Unassigned | Unassigned | Outlier | SHHC |
| 19520 | S2-wms-combined_200390 | Unassigned | Unassigned | Unassigned | Outlier | SHHC |
| 19521 | S2-wms-combined_201205 | Unassigned | Unassigned | Unassigned | Overlap (VC_737/VC_738) | SHHC |
| 19522 | S2-wms-combined_204225 | Unassigned | Unassigned | Unassigned | Singleton | SHHC |
| 19523 | S2-wms-combined_207000 | Unassigned | Unassigned | Unassigned | Overlap (VC_138/VC_146) | SHHC |
| 19525 | S2-wms-combined_208559 | Unassigned | Unassigned | Unassigned | Overlap (VC_652/VC_1481) | SHHC |
| 19528 | S2-wms-combined_222535 | Unassigned | Unassigned | Unassigned | Singleton | SHHC |
| 19529 | S2-wms-combined_225908 | Unassigned | Unassigned | Unassigned | Outlier | SHHC |
| 19532 | S2-wms-combined_230282 | Unassigned | Unassigned | Unassigned | Singleton | SHHC |
| 19540 | S2-wms-combined_25727 | Unassigned | Unassigned | Unassigned | Outlier | SHHC |
| 19541 | S2-wms-combined_30750 | Unassigned | Unassigned | Unassigned | Singleton | SHHC |
| 19544 | S2-wms-combined_48661 | Unassigned | Unassigned | Unassigned | Singleton | SHHC |
| 19545 | S2-wms-combined_51446 | Unassigned | Unassigned | Unassigned | Outlier | SHHC |
| 19546 | S2-wms-combined_52006 | Unassigned | Unassigned | Unassigned | Singleton | SHHC |
| 19547 | S2-wms-combined_52568 | Unassigned | Unassigned | Unassigned | Outlier | SHHC |
| 19549 | S2-wms-combined_57293 | Unassigned | Unassigned | Unassigned | Overlap (VC_1221/VC_1237) | SHHC |
| 19550 | S2-wms-combined_5791 | Unassigned | Unassigned | Unassigned | Outlier | SHHC |
| 19552 | S2-wms-combined_58563 | Unassigned | Unassigned | Unassigned | Overlap (VC_138/VC_146) | SHHC |
| 19555 | S2-wms-combined_62839 | Unassigned | Unassigned | Unassigned | Outlier | SHHC |

| | | | | | | |
|---|---|---|---|---|---|---|
| 19556 | S2-wms-combined_64692 | Unassigned | Unassigned | Unassigned | Overlap (VC_817/VC_1210) | SHHC |
| 19558 | S2-wms-combined_72725 | Unassigned | Unassigned | Unassigned | Outlier | SHHC |
| 19559 | S2-wms-combined_73327 | Unassigned | Unassigned | Unassigned | Singleton | SHHC |
| 19560 | S2-wms-combined_74387 | Unassigned | Unassigned | Unassigned | Outlier | SHHC |
| 19561 | S2-wms-combined_75144 | Unassigned | Unassigned | Unassigned | Outlier | SHHC |
| 19562 | S2-wms-combined_7756 | Unassigned | Unassigned | Unassigned | Outlier | SHHC |
| 19563 | S2-wms-combined_78670 | Unassigned | Unassigned | Unassigned | Outlier | SHHC |
| 19564 | S2-wms-combined_84608 | Unassigned | Unassigned | Unassigned | Outlier | SHHC |
| 19568 | S2-wms-combined_95987 | Unassigned | Unassigned | Unassigned | Singleton | SHHC |
| 19570 | S2-wms-combined_99446 | Unassigned | Unassigned | Unassigned | Overlap (VC_630/VC_642) | SHHC |
| 19573 | S2-wms_124416 | Unassigned | Unassigned | Unassigned | Overlap (VC_607/VC_1238) | SHHC |
| 19579 | S2-wms_161986 | Unassigned | Unassigned | Unassigned | Overlap (VC_20/VC_542) | SHHC |
| 19580 | S2-wms_192667 | Unassigned | Unassigned | Unassigned | Overlap (VC_20/VC_542) | SHHC |
| 19582 | S2-wms_245232 | Unassigned | Unassigned | Unassigned | Overlap (VC_138/VC_146) | SHHC |
| 19583 | S2-wms_246296 | Unassigned | Unassigned | Unassigned | Outlier | SHHC |
| 19585 | S2-wms_248324 | Unassigned | Unassigned | Unassigned | Outlier | SHHC |
| 19586 | S2-wms_248327 | Unassigned | Unassigned | Unassigned | Outlier | SHHC |
| 19588 | S2-wms_6424 | Unassigned | Unassigned | Unassigned | Outlier | SHHC |
| 19591 | S3-wms-combined_135428 | Unassigned | Unassigned | Unassigned | Outlier | ME |
| 19592 | S3-wms-combined_45305 | Unassigned | Unassigned | Unassigned | Outlier | ME |
| 19596 | S3-wms_94494 | Unassigned | Unassigned | Unassigned | Outlier | ME |
| 19597 | S4-wms-combined_10758 | Unassigned | Unassigned | Unassigned | Outlier | SHHC |
| 19598 | S4-wms-combined_112093 | Unassigned | Unassigned | Unassigned | Overlap (VC_422/VC_1413) | SHHC |
| 19600 | S4-wms-combined_118531 | Unassigned | Unassigned | Unassigned | Outlier | SHHC |
| 19601 | S4-wms-combined_121978 | Unassigned | Unassigned | Unassigned | Outlier | SHHC |
| 19602 | S4-wms-combined_1246 | Unassigned | Unassigned | Unassigned | Outlier | SHHC |
| 19606 | S4-wms-combined_22012 | Unassigned | Unassigned | Unassigned | Outlier | SHHC |
| 19607 | S4-wms-combined_24659 | Unassigned | Unassigned | Unassigned | Singleton | SHHC |
| 19608 | S4-wms-combined_25201 | Unassigned | Unassigned | Unassigned | Outlier | SHHC |
| 19614 | S4-wms-combined_57963 | Unassigned | Unassigned | Unassigned | Overlap (VC_138/VC_146/VC_147) | SHHC |
| 19616 | S4-wms-combined_83492 | Unassigned | Unassigned | Unassigned | Outlier | SHHC |
| 19618 | S4-wms_114120 | Unassigned | Unassigned | Unassigned | Outlier | SHHC |
| 19623 | S4-wms_50475 | Unassigned | Unassigned | Unassigned | Outlier | SHHC |
| 19626 | S5-wms-combined_106597 | Unassigned | Unassigned | Unassigned | Singleton | ME |
| 19628 | S5-wms-combined_117095 | Unassigned | Unassigned | Unassigned | Outlier | ME |
| 19633 | S5-wms-combined_147993 | Unassigned | Unassigned | Unassigned | Overlap (VC_630/VC_642) | ME |
| 19634 | S5-wms-combined_156329 | Unassigned | Unassigned | Unassigned | Outlier | ME |
| 19635 | S5-wms-combined_161191 | Unassigned | Unassigned | Unassigned | Outlier | ME |
| 19636 | S5-wms-combined_163165 | Unassigned | Unassigned | Unassigned | Overlap (VC_1485/VC_1486) | ME |
| 19637 | S5-wms-combined_175145 | Unassigned | Unassigned | Unassigned | Outlier | ME |
| 19639 | S5-wms-combined_179241 | Unassigned | Unassigned | Unassigned | Outlier | ME |
| 19640 | S5-wms-combined_183113 | Unassigned | Unassigned | Unassigned | Overlap (VC_652/VC_1481) | ME |
| 19642 | S5-wms-combined_197186 | Unassigned | Unassigned | Unassigned | Singleton | ME |
| 19643 | S5-wms-combined_199135 | Unassigned | Unassigned | Unassigned | Overlap (VC_733/VC_1494) | ME |
| 19644 | S5-wms-combined_204782 | Unassigned | Unassigned | Unassigned | Outlier | ME |
| 19645 | S5-wms-combined_205046 | Unassigned | Unassigned | Unassigned | Singleton | ME |
| 19648 | S5-wms-combined_210959 | Unassigned | Unassigned | Unassigned | Overlap (VC_678/VC_679) | ME |
| 19649 | S5-wms-combined_26061 | Unassigned | Unassigned | Unassigned | Overlap (VC_877/VC_1413) | ME |
| 19653 | S5-wms-combined_3201 | Unassigned | Unassigned | Unassigned | Outlier | ME |
| 19659 | S5-wms-combined_67650 | Unassigned | Unassigned | Unassigned | Overlap (VC_678/VC_679) | ME |
| 19667 | S5-wms-combined_94482 | Unassigned | Unassigned | Unassigned | Outlier | ME |
| 19668 | S5-wms-combined_94786 | Unassigned | Unassigned | Unassigned | Overlap (VC_630/VC_642) | ME |
| 19670 | S5-wms_10783 | Unassigned | Unassigned | Unassigned | Singleton | ME |
| 19671 | S5-wms_123499 | Unassigned | Unassigned | Unassigned | Singleton | ME |
| 19672 | S5-wms_164032 | Unassigned | Unassigned | Unassigned | Outlier | ME |
| 19674 | S5-wms_212596 | Unassigned | Unassigned | Unassigned | Singleton | ME |
| 19677 | S5-wms_62679 | Unassigned | Unassigned | Unassigned | Overlap (VC_735/VC_915) | ME |
| 19681 | S6-wms-combined_110950 | Unassigned | Unassigned | Unassigned | Overlap (VC_1221/VC_1237) | SHHC |
| 19689 | S6-wms-combined_128972 | Unassigned | Unassigned | Unassigned | Overlap (VC_651/VC_736) | SHHC |
| 19691 | S6-wms-combined_138710 | Unassigned | Unassigned | Unassigned | Overlap (VC_1221/VC_1237) | SHHC |
| 19697 | S6-wms-combined_56464 | Unassigned | Unassigned | Unassigned | Outlier | SHHC |
| 19698 | S6-wms-combined_65225 | Unassigned | Unassigned | Unassigned | Singleton | SHHC |
| 19700 | S6-wms-combined_745 | Unassigned | Unassigned | Unassigned | Singleton | SHHC |
| 19703 | S6-wms-combined_88414 | Unassigned | Unassigned | Unassigned | Overlap (VC_877/VC_1413) | SHHC |
| 19708 | S6-wms_140743 | Unassigned | Unassigned | Unassigned | Outlier | SHHC |
| 19711 | S7-wms-combined_106253 | Unassigned | Unassigned | Unassigned | Outlier | ME |
| 19713 | S7-wms-combined_107679_1 | Unassigned | Unassigned | Unassigned | Outlier | ME |
| 19715 | S7-wms-combined_121062 | Unassigned | Unassigned | Unassigned | Singleton | ME |
| 19716 | S7-wms-combined_123085 | Unassigned | Unassigned | Unassigned | Singleton | ME |
| 19717 | S7-wms-combined_134452 | Unassigned | Unassigned | Unassigned | Singleton | ME |
| 19719 | S7-wms-combined_135154 | Unassigned | Unassigned | Unassigned | Outlier | ME |
| 19721 | S7-wms-combined_137858 | Unassigned | Unassigned | Unassigned | Overlap (VC_1221/VC_1237) | ME |
| 19725 | S7-wms-combined_14529 | Unassigned | Unassigned | Unassigned | Outlier | ME |
| 19727 | S7-wms-combined_156351 | Unassigned | Unassigned | Unassigned | Outlier | ME |
| 19729 | S7-wms-combined_157434 | Unassigned | Unassigned | Unassigned | Singleton | ME |
| 19731 | S7-wms-combined_167867 | Unassigned | Unassigned | Unassigned | Singleton | ME |
| 19733 | S7-wms-combined_174935 | Unassigned | Unassigned | Unassigned | Outlier | ME |
| 19736 | S7-wms-combined_190281 | Unassigned | Unassigned | Unassigned | Outlier | ME |
| 19738 | S7-wms-combined_2122 | Unassigned | Unassigned | Unassigned | Outlier | ME |
| 19739 | S7-wms-combined_21693 | Unassigned | Unassigned | Unassigned | Outlier | ME |
| 19740 | S7-wms-combined_217226 | Unassigned | Unassigned | Unassigned | Outlier | ME |
| 19741 | S7-wms-combined_218379 | Unassigned | Unassigned | Unassigned | Outlier | ME |
| 19742 | S7-wms-combined_224523 | Unassigned | Unassigned | Unassigned | Outlier | ME |
| 19750 | S7-wms-combined_3669 | Unassigned | Unassigned | Unassigned | Outlier | ME |
| 19756 | S7-wms-combined_69024 | Unassigned | Unassigned | Unassigned | Outlier | ME |
| 19757 | S7-wms-combined_71592 | Unassigned | Unassigned | Unassigned | Outlier | ME |
| 19760 | S7-wms-combined_74829 | Unassigned | Unassigned | Unassigned | Outlier | ME |
| 19768 | S7-wms_151112 | Unassigned | Unassigned | Unassigned | Outlier | ME |
| 19776 | S7-wms_76437 | Unassigned | Unassigned | Unassigned | Outlier | ME |
| 19779 | S8-wms-combined_117281 | Unassigned | Unassigned | Unassigned | Outlier | SHHC |
| 19785 | S8-wms-combined_164289 | Unassigned | Unassigned | Unassigned | Outlier | SHHC |
| 19787 | S8-wms-combined_180957 | Unassigned | Unassigned | Unassigned | Outlier | SHHC |
| 19788 | S8-wms-combined_185611 | Unassigned | Unassigned | Unassigned | Outlier | SHHC |
| 19794 | S8-wms-combined_219796 | Unassigned | Unassigned | Unassigned | Overlap (VC_138/VC_146) | SHHC |
| 19798 | S8-wms-combined_23583 | Unassigned | Unassigned | Unassigned | Outlier | SHHC |
| 19799 | S8-wms-combined_239791 | Unassigned | Unassigned | Unassigned | Outlier | SHHC |
| 19801 | S8-wms-combined_246389 | Unassigned | Unassigned | Unassigned | Overlap (VC_817/VC_1210) | SHHC |
| 19809 | S8-wms-combined_48046 | Unassigned | Unassigned | Unassigned | Outlier | SHHC |
| 19813 | S8-wms-combined_74275 | Unassigned | Unassigned | Unassigned | Outlier | SHHC |
| 19814 | S8-wms-combined_81597 | Unassigned | Unassigned | Unassigned | Outlier | SHHC |
| 19817 | S8-wms_113693 | Unassigned | Unassigned | Unassigned | Outlier | SHHC |
| 19820 | S8-wms_190781 | Unassigned | Unassigned | Unassigned | Outlier | SHHC |
| 19821 | S8-wms_20528 | Unassigned | Unassigned | Unassigned | Overlap (VC_138/VC_146) | SHHC |
| 19823 | S8-wms_270 | Unassigned | Unassigned | Unassigned | Overlap (VC_1244/VC_1245) | SHHC |
| 19827 | S8-wms_58515 | Unassigned | Unassigned | Unassigned | Outlier | SHHC |
| 19830 | S9-wms-combined_105822 | Unassigned | Unassigned | Unassigned | Overlap (VC_138/VC_146) | ME |
| 19831 | S9-wms-combined_109322 | Unassigned | Unassigned | Unassigned | Outlier | ME |
| 19832 | S9-wms-combined_10990 | Unassigned | Unassigned | Unassigned | Overlap (VC_1489/VC_1494) | ME |
| 19833 | S9-wms-combined_122742 | Unassigned | Unassigned | Unassigned | Outlier | ME |
| 19834 | S9-wms-combined_124857 | Unassigned | Unassigned | Unassigned | Outlier | ME |
| 19835 | S9-wms-combined_2042 | Unassigned | Unassigned | Unassigned | Outlier | ME |
| 19839 | S9-wms-combined_27311 | Unassigned | Unassigned | Unassigned | Singleton | ME |
| 19845 | S9-wms-combined_3461 | Unassigned | Unassigned | Unassigned | Outlier | ME |
| 19857 | S9-wms-combined_93232 | Unassigned | Unassigned | Unassigned | Outlier | ME |
| 19858 | S9-wms-combined_97755 | Unassigned | Unassigned | Unassigned | Overlap (VC_138/VC_146) | ME |
| 19861 | S9-wms_10480 | Unassigned | Unassigned | Unassigned | Singleton | ME |
| 19862 | S9-wms_114971 | Unassigned | Unassigned | Unassigned | Outlier | ME |
| 19867 | SPVDNA | Unassigned | Unassigned | Unassigned | Singleton | Ref |
| 19873 | X51522 | Unassigned | Unassigned | Unassigned | Singleton | Ref |
| 19874 | X58839 | Unassigned | Unassigned | Unassigned | Singleton | Ref |
| 19877 | X80191 | Unassigned | Unassigned | Unassigned | Outlier | Ref |
| 19878 | X96987 | Unassigned | Unassigned | Unassigned | Overlap (VC_818/VC_820) | Ref |

| VC_reference_genomes_only | Members |
|---|---|
| VC_0_0 | AB002632,KC357596,OP297622 |
| VC_1_0 | AB276040,AB981169,GU911303,KP966108,MG747435,MT701591,ON170026,ON170032,ON170053,ON170059,ON170061,ON170067,ON170070,ON170073,ON170076,ON170078,ON170080,ON170081,ON170082,ON170084,ON170128,ON170130,ON170131,ON170135,ON170151,ON170173,ON170210,ON170213,ON170227,ON170230,ON170233,ON170235,ON170237,ON170239,ON170241,ON170243,ON170258,ON170261,ON170263,ON170266,ON170268,ON170270,ON170272,ON170276,ON170304,ON170305,ON170321,ON170325,ON170329,ON170344,ON170347,ON170348,ON170351,ON642070 |
| VC_10_0 | AP011955,AP011956,AY508486,CP062430,CP062436,CP062447,DQ530361,GQ398772,KC595278,KF831354,KJ452292,KJ596420,KT809368,KT809369,KT878766,KY653123,MG029510,MG029512,MH384259,MH401416,MW924889,OP038320,OP136135,OP328423,OP493558 |
| VC_100_0 | KC330679,KC330680,KC330681,KC330682,KC330683,KC330684,KF669648,KF669651,KF669659,KU836751,MG983742,MH884513,MN727882,MT422786,MT514532 |
| VC_1000_0 | MG945161,MK249131,MK249144,MK249156,MK249159,MK249160,MK249167,MK249179,MK249181,MZ364226,MZ364241,MZ364244,MZ364245,MZ364246,MZ364247,MZ364249,MZ364250,MZ364260,MZ364263,MZ364266,MZ364270 |
| VC_1002_0 | MT152698,MZ334527 |
| VC_1003_0 | MN698240,MN698245,MN698246,MN698248 |
| VC_1004_0 | MW202462,MW202544 |
| VC_1006_0 | KC595511,KT070866,MG967616,MG967617,MG967618,MK288022,MW392801,MW392802 |
| VC_1007_0 | MT074134,MT074135,MT074143,MT074144,MT074145,MT074147,MT074148,MT074149,MT074150,MT074151,MT074152,MT074153,MT074154,MT074155,MT074156,MT074157,MT074158,MT074159,MT074160 |
| VC_101_0 | KY565347,MH884514,OP341625 |
| VC_1011_0 | KC821606,KC821623,KC821628,KC821631,MT732445,MT732446,MT732447,MT732448,MT732449 |
| VC_1012_0 | KC821608,KC821620,KC821622,KC821633 |
| VC_1013_0 | MN812206,MN812207,MN812208,MN812209,MN812210,MN812212,MN812213,MN812214,MN812215,MN812216,MN812217,MN812218,MN812219,MN812220,MN812221,MN812222,MN812223,MN812224,MN812225,MN812226,MN812227,MN812228,MN812229,MN812230,MN812231,MN812232,MN812233,MN812234,MN812235,MN812236,MN812237,MN812238,MN812240,MN812241 |
| VC_1015_0 | MT497017,MT497065,MT497066,MT497067,MT497068,MT497069,MT497070,MT497071,MT497072,MT497073,MT497074,MT497075,MT497076,MT497077,MT497078,MT497079,MT497080,MT497081,MT497082,MT497083,MT497084,MT497085,MT497086,MT497087,MT497088,MT497089,MT497090,MT497091,MT497092,MT497093,MT497094,MT497095,MT497096,MT497097,MT497098,MT497099,MT497100,MT497101,MT497102,MT497103,MT497104,MT497105,MT497106,MT497107,MT497108,MT497109,MT497110,MT497111,MT497112,MT497113,MT497114,MT497115,MT497116,MT497117,MT497118,MT497119,MT497120,MT497121,MT497122,MT497123,MT497124,MT497125 |
| VC_1016_0 | MT732435,MT732436,MT732437,MT732438,MT732439,MT732440,MT732441,MT732442 |
| VC_1017_0 | KC821609,KC821632,KT962245,KT962246,KT962247 |
| VC_1018_0 | KJ003983,MT774389 |
| VC_1018_1 | MH675552,MT074136,MT074138,MT774378,MT774385,MT774387,MT774388,MT774390,MT774391,MT774392 |
| VC_1019_0 | KC821612,KC821614,MT732432,MT732433,MT732434 |
| VC_102_0 | KJ159566,KT001912 |
| VC_1020_0 | KC821613,KC821615,KC821617,KC821619,KC821627 |
| VC_1021_0 | MK764442,MK764445,MK764448,MN812203,MN812204,MN812205,MW421581,MW421582,MW421583,MW421584 |
| VC_1022_0 | KY206887,MK017819,MN417334,OP381444 |
| VC_1023_0 | KC959568,KU599887,KU599888,KU599889,MK764437,MK764440,MK764450,MW421576 |
| VC_1024_0 | KC960489,KF981876 |
| VC_1025_0 | KF005317,KF005318,KF005319,KF005320 |
| VC_1026_0 | KX257490,MW805362 |
| VC_1027_0 | KF322026,KF800937,KJ502657,KY290756,KY626176,KY626177,MF481197,MG018929,MG545917,MH113812,MH113813,MH113814,MH113815,MK455769,MK455770,MK820013,MN807240,MT941682,MW451247,MW865291 |
| VC_1028_0 | KU245542,OK638201,OK638202,OK638203,OL343755,OL473897,OL622073,ON857939 |
| VC_1029_0 | MW677511,MW677527,OP009274,OP009281 |
| VC_103_0 | MN176222,MN176223,MN176224,MN176225,MN176226,MN176229 |
| VC_1030_0 | MG596799,MK473373,MZ605292 |
| VC_1031_0 | MN096357,MN096358,MT818426,ON526969 |
| VC_1032_0 | KF669655,KF669657,KF669660,KM236247,KM236248,KP411017,KT001911 |
| VC_1034_0 | ON645346,OP580516 |
| VC_1034_1 | KF692088 |
| VC_1035_0 | MH834603,MH834605,MH834611,MH834614,MH834623,ON970561 |
| VC_1036_0 | MH834598,MH834612,MH834625,MH910036,MH910037,MH910041,MW712719,ON724017,ON970598,OP434442,OP434463 |
| VC_1037_0 | MH834621,MH834627,MK392368,MZ150784,MZ274308,ON392166 |
| VC_1039_0 | KF751793,KF751794,KF751795,KF751796,KF751797 |
| VC_104_0 | MH752385,MN013088,MN176219,MN176227,MN176228,X97918 |
| VC_1040_0 | KT876725,KU599880,KU599881,KU599882,KU599883,KU599884,KU599885,MK764436,MK764438,MK764439,MK764443,MK764444,MK764446,MK764447,OK268422,OK268423,OK268424,OK268425,OK268426,OK268427,OK340817,OK340818,OK340819 |
| VC_1041_0 | MG592391,MG592421,MG592436,MG592443,MG592445,MG592449,MG592661,OX241561,OX241569 |
| VC_1043_0 | MN183282,MT771347 |
| VC_1044_0 | OM913586,ON970564,ON970608 |
| VC_1045_0 | MN988468,MN988516 |
| VC_1046_0 | KJ510413,KM591905,MF155936,MG925349,MH001451,MK494089,MN735432 |
| VC_1047_0 | MW584157,MW584173,MW584189,MW584206,MW584209 |
| VC_1048_0 | KJ617393,LK392619,Z47794 |
| VC_1049_0 | KJ802832,KX987158,KY002061,MH586730,MZ726797,OL604152,ON602746,OP617744 |
| VC_1050_0 | MG592411,MG592538,MG592563,MG592564,MG592565,MG592571,MG592632,MG592633,MG592634,MG592635,MG592636,MG592637,MG592638,MG592639,MG592640,MG592642,MG592645,MG592646,MG592648,MG592649,MG592650,MG592651,MG592652,MG592653 |
| VC_1050_1 | MG592397,MG592406,MG592407,MG592408,MG592416,MG592417,MG592428,MG592463,MG592464,MG592471,MG592475,MG592476,MG592492,MG592493,MG592528,MG592601,MG592602,MG592617,MG592618,MG592655,MH925092,MW824418,MW824430,MW824432,MW824433,MW824434,OX241427,OX241556,OX241563 |
| VC_1051_0 | KM058087,MW900438 |
| VC_1052_0 | MT234342,MT234343 |
| VC_1052_1 | KP881232 |
| VC_1052_2 | MF403007 |
| VC_1052_3 | KM199770,MN549360,MN549361,MT778837,MT778839 |
| VC_1052_4 | MN988486,MN988509,MN988520,MN988532,MN988534,MN988553 |
| VC_1053_0 | MG945594 |
| VC_1054_0 | KM589498,KT264826,KT264827,MG945473,MG945703,MH931003,MK496723,MN379633,MN379651,OM522999,OM523001,OM869624 |
| VC_1056_0 | KT264807,KT264829,KX513869,MG945560 |
| VC_1058_0 | OM869532,OM869702 |
| VC_1059_0 | MG945835,MH992170,MH992193 |
| VC_1061_0 | KM589499,KM589507 |
| VC_1062_0 | KM589500,KM589503,KM589504,KM589505,KM589509,KM589510,KM589512,KM589513,KP087948,MG945178,MG945463,MH572367,MH572368,MH572377,MH572378,MH572379,MH572459,MH572460,MH572461,MH572462,MH572463,MH572465,MH992180,MK249133,MK249138,MK249191,MK249199,MK249207,MK249208,MK249228,MK249232,MK765579,MK765582,MK765639,MK765642,MK765643,MK765644,MK765646,MN311488,MN311489,MN311490,MN311491,MZ364242,OL617032,OL617033 |
| VC_1066_0 | KM589508,KT264808,KT264809,KT264810,MH992165,MK249127,MK249128,MK249129,MK249139,MK249149,MK249150,MK249153,MK249154,MK249155,MK249158,MK249162,MK249163,MK249166,MK249168,MK249174,MK249176,MK249178,MK249180,MK249188,MK249192,MK249194,MK249195,MK249196,MK249198,MK249204,MK249212,MK249214,MK249219,MN862338,MZ364222,MZ364223,MZ364232,MZ364248,MZ364254,MZ364255,MZ364261,MZ364262,MZ364264,MZ364265,MZ364268,MZ364269,MZ364271,MZ364272,MZ364273,MZ364275,MZ364276,MZ364278 |
| VC_1067_0 | MK249132,MK249205 |
| VC_1068_0 | KM589514,MK249222,MZ364231,MZ364280,MZ364284 |
| VC_1069_0 | KM589515,KM589516,MT135405,OM869497 |
| VC_107_0 | OP341624,OP341626,OP341627,OP341628,OP341630 |
| VC_1070_0 | MH153807,MH153812 |
| VC_1070_1 | KX774321 |
| VC_1071_0 | MH576964,MH576965,MH576968,MH590597,MH590599,MH727564,MK359332,MK359351,MK801722,MN369750,MN369757,MN428060,MN484599,MW291014,MW507134,ON260819,ON260828,ON970579,ON970590,OP021680 |
| VC_1071_1 | OL829978 |
| VC_1071_2 | MF155946,MT310865,MT684590,MT897905,ON970591 |
| VC_1071_3 | KM652554,MF347636,MF347637,MF347638,MF347639,MF358541,MF358542,MK460248,MK494112,MK977711,MN096371,MN369743,MW507136,MZ958750,OK310502,ON108650 |
| VC_1071_4 | MN444876 |
| VC_1071_5 | MT711976 |
| VC_1072_0 | MG757153,MK620896 |
| VC_1072_1 | MT024865,MT024872 |
| VC_1072_2 | MH590589,MH651172,MW365952,MZ820095,ON456343 |
| VC_1072_3 | MK061412,MT114162,MW435853,MZ820093,MZ820094,ON208830 |
| VC_1072_4 | MH536811,MH536814,MN369754,MT952852 |
| VC_1072_5 | MT684598,MW822145,ON645339 |
| VC_1073_0 | KT365401,KU160651,KU160656,KU160665,KU160668,KU647629,MF189176,MF189177,MF189178,MF324907,MF377441,MF377442,MH825703,MK919479,MN183279,MN183283,MT024868,MT114161,MT310895,MT776811,MT889388,MW862982,OK999980,ON970601 |
| VC_1074_0 | KM879463,KU160660,KX522650,KX670786,KX670787,KX855961,KX855962,MF185718,MF185721,MF185723,MF185724,MF185725,MF668274,MH744419,MK494107,MN204499,MN204503,MN234234,MN703411 |
| VC_1075_0 | MT498055,MW601223,MZ747522,OL742560 |
| VC_1077_0 | KP087938,KP087942,MG883726,MG945129,MG945206,MG945670,MG945836,MG945840,MG945846,MH572362,MH572410,MH572411,MH572412,MH572486,MH572521,MH931004,MH992163,MH992164,MH992192,MH992199,MH992202,MH992206,MH992208,MH992214,MH992215,MH992216,MH992217,MH992219,MH992220,MH992222,MK765645,MN862346,MW202445,MW202609,MW202760,MW202819,MZ089752,MZ364224,MZ364225,MZ364227,MZ364277,MZ364279,MZ364283,MZ364285,MZ364286,MZ364291,OL617030,OL617031 |
| VC_1079_0 | MH572360,MH572361,MH572409,MH572457,MH572482,MH572483 |
| VC_108_0 | AB712291,HE962497,JF731128,KF192053,KJ094032,KJ127303,KT932699,KY303907,KY472224,LC547238,MH618488,MH791415,MK570225,MK721189,MK721194,MK800154,MN103542,MN871443,MN939408,MW677132,MZ333457,OL505085,OL870612,ON086985,ON113171,ON113172,ON113173,ON113174,ON113175,ON113177,ON113178,ON113179,ON113180 |
| VC_1081_0 | MN379632 |
| VC_1082_0 | MH572366,MW202464,MW202671 |
| VC_1084_0 | MH572287,MH572288,MH572467,MH572468,MH572496,MH572497,MH572498,MH572499,MH572500 |
| VC_1085_0 | KT264812,MG945475 |
| VC_1086_0 | KP087943,KP087951,KP087957 |
| VC_1087_0 | KP087953,MH572371,MH572373,MH572380,MK249215 |
| VC_1088_0 | KP087955,MH572364,MH572458 |
| VC_1089_0 | MH572489,MW202618 |
| VC_109_0 | MZ398135,MZ398136,MZ399596 |
| VC_1090_0 | KP202971,KT001914,MF098558,ON323491 |
| VC_1091_0 | KP836355,KP836356 |
| VC_1092_0 | MW314851,MW314853,MW314857,MW584149,MW584151,MW584184,MW584193,MW584194,MW584198,MW584200,MW584205,MW584210 |
| VC_1093_0 | KU963250,MF919514,MG812487,MH153809,MH450129,MH479924,MH651166,MH651168,MH976506,MH976507,MH976508,MH976509,MH976510,MH976511,MH976512,MH976513,MH976514,MH976518,MH976519,MK937594,MK937601,MK967396,MK977702,MN329673,MN329677,MT498035,MT639640,MT723934,MT723937,MT723941,MW822146,MW924645,MZ322015,MZ681510,ON081332,ON456336,ON637761,ON724014,OP172877,OP434456 |
| VC_1095_0 | KU963253,MH669000,MK937607,MK937611,MN586026,MN586028,MN586047,MT498036,MW314847,MZ322009,MZ322020,ON392159,ON456337,ON456349,ON755186,OP068342 |
| VC_1096_0 | KR063280,MK279853,MN586022,MZ820089,ON755184 |
| VC_1098_0 | MK814760,MK894436,MN484601,OP297539 |
| VC_1098_1 | KR063281 |
| VC_1099_0 | MN234206,MN234211 |
| VC_1099_1 | OK040794 |
| VC_11_0 | AP019560,AP019561,AP019562,KX827368,KX827369,KX827370,MF428478,MF428479,MF428480,MF428481,MK075003,MK075006,MN028536,MT423823,MT423824,OM373551,OM373552,OM373553,OP168678 |
| VC_110_0 | FR687252,MZ333135,OM513679,OM513680 |
| VC_1100_0 | MN908685,MZ150783 |
| VC_1100_1 | OK040793,ON637770 |
| VC_1100_2 | OK040790 |
| VC_1100_3 | OP068343 |
| VC_1101_0 | KR136259,KR136260 |
| VC_1103_0 | KR704913,KR704915 |
| VC_1104_0 | KT264749,KT264750,KT264751,KT264752,KT264753,KT264754,KT264755,KT264756,KT264757,KT264758,KT264759,KT264760,KT264761,KT264762,KT264763,KT264764,KT264765,KT264766,KT264767,KT264768,KT264769,KT264779,KX513865,MG025518,MG641179,MG641180,MG641186,MG883717,MG945153,MG945158,MG945163,MG945190,MG945203,MG945205,MG945207,MG945213,MG945227,MG945237,MG945238,MG945318,MG945325,MG945330,MG945333,MG945346,MG945351,MG945355,MG945361,MG945363,MG945387,MG945393,MG945398,MG945402,MG945407,MG945415,MG945424,MG945429,MG945435,MG945438,MG945444,MG945459,MG945461,MG945467,MG945476,MG945486,MG945499,MG945508,MG945517,MG945526,MG945541,MG945548,MG945559,MG945565,MG945566,MG945570,MG945586,MG945587,MG945589,MG945596,MG945602,MG945646,MG945671,MG945673,MG945674,MG945681,MG945691,MG945699,MG945705,MG945708,MG945710,MG945728,MG945731,MG945744,MG945758,MG945761,MG945765,MG945768,MG945770,MG945786,MG945787,MG945793,MG945796,MG945801,MG945802,MG945820,MG945853,MG945855,MG945862,MG945864,MG945869,MK765620,MT135257,MT135263,MT135264,MT135272,MT135273,MT135278,MT135338,MZ089765,MZ089767,MZ089774,MZ089775,OM869524,OM869525,OM869526,OM869527,OM869528,OM869531,OM869533,OM869534,OM869535,OM869537,OM869538,OM869543,OM869546,OM869547,OM869550,OM869606,OM869635,OM869641,OM869642,OM869643,OM869644,OM869645,OM869664,OM869697,OM869698,OM869700,OM869701,OM869703,OM869704,OP549869,OP549872,OP549875,OP549876 |
| VC_1105_0 | MG945597,MK496743,MK496744 |
| VC_1108_0 | KT264775,MG883718,MG945386,MG945775 |
| VC_111_0 | AB716666,GQ413938,KF765493,KM576124,KP708985,KP708986,KT367886,KT964103,KU666550,KX211991,KX237514,KX587949,KX856662,KY000081,KY385423,KY684082,MF612071,MG972768,MH885472,MK135468,MK562503,MK598851,MK693005,MN101217,MN101219,MN101220,MN116494,MN176572,MN176573,MN380459,MN794000,MN794001,MN794002,MN794003,MT090077,MT197175,MT197176,MT360680,MT559526,MT559527,MT682065,MT701589,MT939252,MW590329,MZ208805,MZ424864,MZ571827,MZ602146,MZ634338,MZ634339,MZ634340,MZ634342,MZ634343,OK562429,OK583892,OK583893,OL362278,OL362279,OL474125,OL976437,OL979478,OL979479,OM864357,ON132389,ON146449,ON165415,ON602723,ON602724,ON602725,ON602726,ON602727,ON602728,ON602732,ON602739,ON602740,ON602743,ON602748,ON602752,ON602754,ON602756,ON602757,ON602761,ON602762,ON602767,ON809559,OP114731,OP413829,OU509533,OU509534,OX335379,OX335428 |
| VC_1110_0 | MG945181,MG945211,MG945232,MG945718,MG945783,MT135261,MT135334,MZ089771 |
| VC_1111_0 | MG945489,MT135282 |
| VC_1112_0 | MK496719,MK496738 |
| VC_1113_0 | MG945591 |
| VC_1114_0 | MG945131,MG945742 |
| VC_1116_0 | KT264776,MG883719,MG945139,MG945339,MG945394,MG945403,MG945542,MG945550,MG945645,MG945736,MG945740,MG945771,MG945819,OP209174 |
| VC_1117_0 | KT264781,MG945481,MG945807,MT135256,MT135266 |
| VC_1119_0 | KT264782,KT264783,KX513866,KX513870,MG945154,MG945472,MG945534,MG945613,MG945826,MK496754,MK496805,MK496812,MK496813,MK765559,MK765603,MK765621,MK765669,MZ089756,MZ089773,OM869503 |
| VC_112_0 | KF319020,KM819694,MG575418,MG575419,MK387869,MW358930,OL416096,OM234793 |
| VC_1120_0 | MG945704,MZ089755,MZ089759,MZ089761,MZ089764,OM869627,OM869629 |
| VC_1122_0 | KT264785,KX513873,MG945186,MG945569,MG945689,MG945797,MG945870,MN379630,MN379640,MT135351 |

| | |
|---|---|
| VC_1123_0 | KT264784,KX513864 |
| VC_1124_0 | KT264798,KT264799,KT264800,KT264801,MG945135,MG945136,MG945157,MG945391,MG945867,MT135258 |
| VC_1125_0 | KT264796,KT264797,MG883722,MG945286,MG945319,MG945338,MG945347,MG945385,MG945389,MG945410,MG945416,MG945437,MG945538,MG945635,MG945639,MG945746,MG945767,MG945858,OM869558,OM869663 |
| VC_1126_0 | KT264788,KT264789,KT264790,MG945138,MG945381,MG945382,MG945780,MT135259 |
| VC_1127_0 | KT264791,KT264792,KT264793,KX513868,KX513877,MG945162,MG945233,MG945334,MG945383,MG945388,MG945430,MG945447,MG945514,MG945642,MG945694,MG945753,MG945781,MT135277,MZ821031 |
| VC_1128_0 | KT264794,MG945231 |
| VC_113_0 | AB720063,AP008979,AY299121,DQ777876,EU717894,MW934263,MZ701794,OK258140,OM913894 |
| VC_1132_0 | KT264795,MG945738 |
| VC_1133_0 | MG641184,MZ089778,OM869517,OM869640 |
| VC_1134_0 | MK496714,MK496779,MK496792,MZ089777,OM869545 |
| VC_1136_0 | KT264803,MG945142 |
| VC_1137_0 | KT264802,MG945620,MG945741 |
| VC_1138_0 | KT264805,MG945519,MG945693,MG945809 |
| VC_1139_0 | KT264806,MG945320,MG945561,MG945721 |
| VC_1140_0 | KT264820,KT264821,MG945644,MK765602 |
| VC_1143_0 | KT264818,KT264819,KX513867,KX513872,KX513874,MG945573,MG945759,MG945760,MG945811 |
| VC_1144_0 | MG883714,MG945364,MG945525,MG945719,MG945805,MT135337,MT135424 |
| VC_1145_0 | OM869513,OM869516,OM869519,OM869634,OM869637 |
| VC_1146_0 | MG945234,MG945405,MG945827 |
| VC_1147_0 | KT264822,KT264830,MK496745 |
| VC_1148_0 | KT264823,KT264824,KT264825,MG945179,MG945357,MG945413,MG945506,MG945507,MG945609,MG945676,MG945706,MG945764,MG945817,MT135267,MT135271,MT135281 |
| VC_1149_0 | MG945441,MG945487,MG945505,MT135255,MT135275 |
| VC_115_0 | MG592523,OX241425 |
| VC_1151_0 | KT264828,MG945228,MG945426,MG945726,MG945754,MG945816,OP549870,OP549873,OP549877 |
| VC_1152_0 | MG945204,MG945727,MG945785,MG945810 |
| VC_1153_0 | KT264831,MN379641 |
| VC_1154_0 | KT264833,MG945418,MG945549,MG945747,OM869677 |
| VC_1155_0 | KT264834,MG945678 |
| VC_1156_0 | KT755656,MH431938,MH454076,MH454080,MH460824,MH460825,MH460826,MH460827 |
| VC_1157_0 | KT876724,KT876726,KU599877,KU599878,KU599879,KU599886,MK764441,MK764449,MW421577,MW421578,MW421579,MW421580,MW442825,MW442826,MW442827,MW442828,MW442829,MW442830,MW448539,MW448540,MW448541,OK340820,OK340821,OK340822 |
| VC_1158_0 | MH576962,MK620894,MW291021 |
| VC_1158_1 | MH744418,MT310898 |
| VC_1159_0 | MF668275,MZ955870,OV696619 |
| VC_116_0 | AB720064,LR743523,LR743528,LR743529,LR743531,MW802488,MW822535,MW822536,MW822537,MW822538,MW822539,OK913679,OK913680 |
| VC_1160_0 | KU160640,KU160643,KU160645,KU160648,KU160649,KU160652,KU160653,KU160659,KU160661,KU160662,KU160671,KU160672,KX576640,KX576641,KX621004,KX621005,KX688102,KX688103,MF140400,MF140404,MF140407,MF140424,MF140425,MF668285,MG198779,MG198781,MG198782,MG210949,MH450117,MH450118,MH450121,MH450131,MH744421,MK112526,MK112528,MK112529,MK112531,MK112535,MK112538,MK279844,MK279845,MK279847,MK279851,MK279854,MK279856,MK279860,MK279862,MK279863,MK279867,MK279868,MK279869,MK279872,MK279874,MK279875,MK279876,MK279880,MK279884,MK279886,MK279892,MK279896,MK279901,MK279907,MK660714,MK814749,MK919476,MK919484,MN586061,MN735429,MN813680,MT498045,MT498050,MW314848,OP434444 |
| VC_1161_0 | KX557278,MT771339 |
| VC_1162_0 | MK016495,MK016497,MK737942,MN369765,MT310855,MT498057,MZ150782,MZ747515,ON260821,ON970583 |
| VC_1163_0 | MG839027,MG925343,MG925345,MG925347,MG944216,MG944218,MG944226,MH513982,MH590604,MH590606,MK308637,MK620901,MK620904,MK814762,MK894432,MK937602,MN096367,MN096378,MN284901,MN310543,MT553340,MT657343,MT657344,MZ005670,OK318961,ON526972,ON970571,OP068339 |
| VC_1164_0 | KU160641,KU160646,MF038790,MF038791,MF140401,MF140423,MF140432,MH399789,MH450115,MH450125,MH479926,MH825711,MN234168,MN234194,MN234221,MN284902,MN703412,MW055912,MZ209303,MZ274309,OK310503,ON456335,ON456340,ON645338,OP434462 |
| VC_1166_0 | KU160644,MF140397,MH834595,MH834606,MH834608,MH834609,MH834616,MH834618,MH834620,MH834624,MN234214,MN484600,MN703409,MT889373,MT952853,MW507131,ON260822,ON970604,OP297550,OP297552 |
| VC_1167_0 | KU160650,MF668266,MF668273,MF668279,MH450114,ON337195 |
| VC_1168_0 | MW507126,MW590223 |
| VC_1169_0 | KU160654,KU160664,MF140418,MF140427,MF140434,MG099950,MN585972 |
| VC_117_0 | AB757800,AB757801,JX880072,KX365747,MH059635,MK574011,MW202665,MW202744,OL959999 |
| VC_1170_0 | MN586021,MT889394,OP434452 |
| VC_1170_1 | KU160669,KU160673,MW712735 |
| VC_1170_2 | MW862992,OM818331 |
| VC_1171_0 | MH153803,MH153813,MN183284,MT114166,MT639641,MT657333,MW365946,MW365949,MW365950,MW712724,MW712727,MW712728,MW924638,MZ322011,MZ322018,OM203157,OP068331,OP068332,OP068336,OP068337,OP068344 |
| VC_1173_0 | KU230356,MK605242,MK605243,MK605244,MK605245,MK605246 |
| VC_1174_0 | KU234532,KU234533,MT426122,MZ078141 |
| VC_1175_0 | KU963246,KU963247,MG099941,MH271295,MK279911,MK279912,MK359313,MK801734,MK875797,MN586055,MT771342,MT818424,MW055908,MW291013,MW507130,MW822147,MZ150788,MZ171371,MZ681509,OM203160,ON645343,OP297544 |
| VC_1176_0 | KU963251,KU963254,KU963259,KU998236,KU998248,KX557274,KX557275,MF919521,MH020241,MH153808,MH536818,MH576969,MK501729,MK501730,MK801721,MK801724,MK814757,MK864263,MK878896,MK919470,MK967389,MN010762,MN062704,MN096365,MN234161,MN234179,MN369740,MN585963,MN586031,MT553336,MT723935,MT723936,MT723942,MW291016,MW601225,MZ150785,MZ622164,ON724008,ON724011,ON724012,ON970567 |
| VC_1177_0 | KU963252,KU998235,KX557279,MH479915,MK061410,MK284521,MK878901,MK937599,MK967377,MN813694,MT498059,MT522006,MT771341,MW055901 |
| VC_1178_0 | KU998234,KX557286,MH479910,MH479917,MH669015,MK305889,MK814761,MK864264,MK864265,MK864266,MK864267,MK937603,MK967381,MN010760,MN284907,MT310872,MT521998,MT639651,MW507125,MZ005680,MZ150791,MZ622177,ON970602,ON970619 |
| VC_1179_0 | KU647626,KU647627,MN586053 |
| VC_118_0 | AB746912,LC545443,LC545444 |
| VC_1180_0 | MK814759,MZ209302,OM818327 |
| VC_1181_0 | KU998245,MK524501 |
| VC_1181_1 | KU998244,KU998246,KU998247,KX557276,MF919520,MF919537,MK376956,MK376959,MT498054,MZ388552 |
| VC_1182_0 | KX557272,MK977705,MN908691,MT657339 |
| VC_1182_1 | MH590603 |
| VC_1183_0 | MH479913,MH479925,MK919478,MN586014,MN813698,MW601220 |
| VC_1184_0 | KX557282,MK524505,MK814750,MW862986,ON724009 |
| VC_1186_0 | KX620750,KX620751,KX620753,KX620754 |
| VC_1187_0 | KX077892,OL774866,OL774876 |
| VC_1188_0 | MT344103,MT344104,MT344105 |
| VC_1189_0 | KX455876,KY290947,KY290949,KY290950,KY290952,MH179470,MH179471,MH179479,MT276994 |
| VC_119_0 | LC545445,LC545446 |
| VC_1190_0 | MG592484,MG592485 |
| VC_1191_0 | KX513871,MG945160,MG945185 |
| VC_1192_0 | MT135371,MT135422,MT135432 |
| VC_1193_0 | MG945352,MG945735,MG945752 |
| VC_1194_0 | MG945210,MG945327,MG945623,MT135440,MT135446 |
| VC_1196_0 | MG945748,MG945799,MG945800 |
| VC_1197_0 | KX513875,MG883712,MG945284,MG945309,MG945433,MG945502,MG945616,MG945794,MK496693,MK496774,MZ089789,OM869564,OM869565,OM869570,OM869709 |
| VC_1199_0 | MH727560,MH926061,MK937613,MK977706,MK977710,MK977712,MK977714,MK977715 |
| VC_12_0 | AP008954,AY954960,AY954961,CP062422,CP062462,CP071049,DQ530362,JQ973847,KJ140076,KJ452291,KP209285,KP893289,KT344895,MK290764,MK618716,MW238767,OM439663,OM439669,OM439672,OM439673,OM439674 |
| VC_12_1 | MG543995,MG564297 |
| VC_12_2 | AB370268,AP001553,AY954962,AY954963,AY954964,AY954965,AY954966,AY954967,AY954968,DQ908929,JX013863,JX094501,KC595279,KP893290,KR709302,KR709303,MK211557,MN904510,ON571632 |
| VC_1200_0 | MF197383,MH271317,MN234233,OP172878 |
| VC_1201_0 | KY817360,OL656105 |
| VC_1202_0 | LC035386,LC210520 |
| VC_1203_0 | MN988521,MN988522,MN988523,MN988524,MN988525,MN988559,MN988561 |
| VC_1203_1 | MN988560 |
| VC_1204_0 | LC644975,LC644976 |
| VC_1205_0 | MT740726,MT740743,MT740745 |
| VC_1206_0 | MH271298,MH271308,MH271320,MK880124,MT952845,MW862981,OK040779,ON970568 |
| VC_1207_0 | MH834610,MH834619,MH834629,MK061413,MN703413,MN813676,MN908689,MT024869,MT024871,MT639653,MT889366,MT889375,MT889376,MT897906,MW055910,MZ005682,MZ681506,MZ747516,MZ820087,OK040777,OL455896,OL549188,OL549189,OL549191,OL549192,ON208833,ON528933,ON970616,OP434455 |
| VC_1208_0 | LR535901,LR535903,LR535904,LR535905,LR535906,LR535907,LR535908,LR535911,LR535912,LR535913,LR535917 |
| VC_121_0 | JQ177061,JQ177064,KJ572844,MN379460 |
| VC_1210_0 | MK250021,MK250023 |
| VC_1210_1 | MK250020,MK250022,MK250024,MK250025,MK250026,MK250027,MK250028 |
| VC_1210_2 | MK250015,MK250016,MK250017,MK250018 |
| VC_1210_3 | MK250019,MK250029 |
| VC_1211_0 | LT986678,MG945141,MG945453,MG945455,MG945456,MG945498,MG945509,MG945564,MG945664,MG945680,MG945683,MG945713,MG945716,MG945730,MG945784,MG945790,MG945813,MG945845,MK765557,MK765655,MK765661,MK765667,MT135342,OM869561,OM869610 |
| VC_1212_0 | MF101922,MH015251 |
| VC_1213_0 | MF443783,MF443784 |
| VC_1215_0 | MF580955,MF580957,MF580958 |
| VC_1216_0 | MF580956,MF580959,MF580960 |
| VC_1217_0 | MF580961,MF580962,MF629150 |
| VC_1218_0 | MF979562,MK080312,MT074429,MT074440 |
| VC_122_0 | MG592400,MG592422,MG592425,MG592448,MG592486 |
| VC_122_1 | MG592434 |
| VC_1222_0 | MG214764,MW202596,OL617037 |
| VC_1223_0 | MG269973,MW202566 |
| VC_1224_0 | MG515223,MG518519,MG518520,MG663582,MG663583,MG663584,MG663585,MG663586,MH155868,MK112550,MN119376,MN284894,MN994275,MT684596,MZ622160,OL455897,ON108647,ON970569,OP021678 |
| VC_1225_0 | MG592394,MG592402,MG592427,MG592455 |
| VC_1226_0 | MG592533,MG592560 |
| VC_1227_0 | MG592536,MG592554,MG592555,MG592556,MG592591,MG592625 |
| VC_1228_0 | MK373772,OP136151 |
| VC_1229_0 | MG592596,MW824384,MW824385,MW824386,MW824395,MW824396,MW824397 |
| VC_123_0 | JX866719,KJ021043,KX237516,MF663761,MG197810,MG432137,MK521906,MK685667,MK798142,MN689778,MT559528,MT757392,MT894004,MZ374361,MZ398021,MZ398245,MZ504995,OK499978,ON086804,ON602758,ON807605,ON809560,OP114730 |
| VC_1230_0 | MG945523,MG945677,OM869552 |
| VC_1231_0 | MZ089785,OM869555 |
| VC_1232_0 | MG641185,MG945667 |
| VC_1233_0 | MG641188,MG945130,MG945256,MG945457,MG945557,MG945725,MG945757,MN379637,MN379647,MT135339,MT135348,MT135361,MT135384,MT135409,MT135435,OL617023 |
| VC_1234_0 | MG670586,MH045566,MH045567,MH045568,MH509447,MH834596,MH834599,MH834602,MH834604,MH834626,MK814753,MK937598,MK937610,MN062714,MN062717,MN183281,MN329675,MT024860,MT310853,MT310896,MT639649,MT818427,MT889365,MT889372,MT889389,MT889392,MW601221,MW601224,MW712721,MW924640,MW924647,MZ322012,MZ747513,MZ820083,OK318958,OK999979,OK999981,OL742562,OM818329,ON724018,OP172875,OP434439,OP434443,OP434445 |
| VC_124_0 | KF977490,MW960031 |
| VC_1240_0 | MG839015,MG839016,MG839017,MG839018,MG839020,MG839021,MG839022,MG839023,MG839026,MG839028,MG839029,MG920061,MG925352,MG944219,MG944222,MG962360,MG962367,MH045554,MH045558,MH045560,MH045562,MH045563,MH045564,MH045565,MH153802,MH153805,MH153806,MH153811,MH513981,MK620895,MK620902,MK620903,MK620905,MK757447,MK919471,MK967388,MK967391,MK967394,MK967400,MK977697,MN062702,MN062709,MN062713,MN234163,MN234177,MN234181,MN234204,MN234222,MN369737,MN617840,MN735430,MN735435,MN807246,MN892483,MT024864,MT310851,MT310856,MT310861,MT310866,MT310873,MT451985,MT498049,MT553339,MT553341,MT553343,MT771350,MT952846,MT986026,MT986027,MW055911,MW291026,MW435859,MZ005684,MZ958749,OK310506,ON260823,ON337194,ON456354,ON970581,ON970605,ON970606,OP021683 |
| VC_1245_0 | MG883710,MG945464 |
| VC_1246_0 | MG883713,MT135394 |
| VC_1247_0 | MG883720,MG945384,MG945482,MG945692,MG945697,MG945698,MG945734 |
| VC_125_0 | AB775548,GU903191,KM360178,KX231828,KY705409,MH252365,MT012302,MT161381,MT161382,MT161383,MT161384,MT375042,MT613935,MT701586,MT708550,MZ345003,ON853583 |
| VC_1250_0 | MG883724,MG945625,MG945629,MG945632,MG945633 |
| VC_1251_0 | MG945128,MG945159,MG945188,MG945229,MG945255,MG945323,MG945332,MG945401,MG945685,MG945745,MG945774,MG945803,MG945829,MT135336,MT135441 |
| VC_1252_0 | MG945544,MG945723,MT135439 |
| VC_1253_0 | MG945414,MG945445,MG945578,MG945611,MT135367,MT135433 |
| VC_1255_0 | MG945460,MG945496,MG945622,MG945830 |
| VC_1256_0 | MG945241,MG945276,MG945448,MG945643 |
| VC_1257_0 | MG945140,MG945257,MG945449,MG945458,MG945662,MG945815 |
| VC_1258_0 | MG945580,MG945804,MG945909 |
| VC_1259_0 | MG945166,MG945208,MG945329,MG945365,MG945512,MG945540,MG945687,MT135329 |
| VC_126_0 | EF057797,EU399241 |
| VC_1260_0 | MG945165,MT135354 |
| VC_1261_0 | MG945177,MG945349,MG945411,MG945462,MG945495 |
| VC_1262_0 | MN311487,MN311492 |
| VC_1263_0 | MG945183,MG945314 |
| VC_1264_0 | MG945261,MG945420,MG945470,MG945518,MK249148,MK496685 |
| VC_1266_0 | MG945189,MG945275,MG945408,MG945848,MG945852 |
| VC_1267_0 | MG945209,MG945263,MG945279,MG945404,MG945750,MG945776,MG945791 |
| VC_1268_0 | MG945212,MG945582,MT135345 |
| VC_127_0 | MF663786 |
| VC_127_1 | OL614104 |

| | |
|---|---|
| VC_127_2 | MT711888 |
| VC_1270_0 | MG945214,MT135380,MT135423 |
| VC_1271_0 | MG945230,MT135357 |
| VC_1272_0 | MG945235,MG945392,MT135388,MT135443,MT135462 |
| VC_1273_0 | MG945236,MG945822,MT135358,MT135377,MT135426,MT135427 |
| VC_1274_0 | MG945567,MG945631 |
| VC_1275_0 | MG945252,MG945584,MN379645,OM869494 |
| VC_1277_0 | MG945494,MG945563,MG945585,MK765590,MN379638,MN379648,MT135333,MT135379,MT135391,MT135401,MT135437,MT135445,OM869612,OM869668 |
| VC_1278_0 | MG945425,MG945789,MT135415 |
| VC_1279_0 | MG945258,MG945434,MT135434,MT135451 |
| VC_1280_0 | MK765572,OM869521 |
| VC_1281_0 | MG945278,MG945285,MG945492,MG945605,MG945751,MG945812,MG945825 |
| VC_1282_0 | MG945280,MK765649,OM869500 |
| VC_1283_0 | MK765552,MK765553,MK765556,MK765560,MK765562,MK765565,MK765591,MK765592,MK765593,MK765596,MK765606,MK765607,MK765609,MK765611,MK765612,MK765613,MK765615,MK765633,MK765648,MK765651,MK765657,MK765658,MK765660,MK765666,MT135276,MT135425 |
| VC_1285_0 | MG945316,MG945344,MG945468,MG945524,MG945556,MG945562,MG945579,MG945612,MG945640,MG945668,MG945712,MG945733,MG945806,MG945814,MK765571,MK765574,MK765576,MK765577,MK765580,MT135330,MT135406,MT135431,MT135458,OL617036,OM869623 |
| VC_1286_0 | MG945328 |
| VC_1287_0 | MG945331,MG945343,MG945490,MG945501,MG945665,MG945729,MG945782 |
| VC_1288_0 | MG945335,MG945778,MT135385 |
| VC_1289_0 | MG945336,MT135365 |
| VC_129_0 | AF493143,KJ507099,KR091952,KU064779,KY798120,KY798121,KY971611,LC730319,LN889756,LN889995,LN913017,MG250485,MG775258,MN729595,MN729596,MN729597,MN729598,MN729599,MN729600,MT104465,MT104466,MT711889,MT711890,MW812345,MZ501272,MZ598487,MZ826344,MZ8<br>26345,MZ826346,MZ826347,MZ826348,MZ826349,MZ826351,MZ826352,MZ826354,OK523998,OK523999,OL581611,OM256450,OM282085,OM471789,OM630590,ON189046 |
| VC_1290_0 | MG945337,MT135410 |
| VC_1291_0 | MG945345,MG945818 |
| VC_1292_0 | MG945348,MG945702,MK496732 |
| VC_1293_0 | MG945350,MG945432,MG945755,MT135447 |
| VC_1294_0 | MG945354,MG945576,MG945637,MK496765,OM869571,OP209176 |
| VC_1295_0 | MG945360,MG945478,MG945614 |
| VC_1296_0 | MG945362,MK496704,MK496729,MK496782 |
| VC_1297_0 | MG945395,MG945421,MG945480,MG945539,MG945696,MK496696,MK496711 |
| VC_1298_0 | MG945397,MT135331,MT135461 |
| VC_1299_0 | MG945409,OM869563 |
| VC_130_0 | AB775549,FR823298,GU583987,JX173487,KP025626,KX431888,KX449361,KX449362,KX449363,MH179472,MH179475,MH179478,MH268168,MK033137,MT094431,MT711887,OK094519,OM131411,ON109291 |
| VC_1300_0 | MG945412,MG945828 |
| VC_1301_0 | MG945417,MG945419 |
| VC_1302_0 | MG945427,MG945574,MT135411 |
| VC_1303_0 | MG945439,MG945628,MK496757,MK765601,MZ089768,OM869696 |
| VC_1304_0 | MZ089804,MZ089807,OM869574,OM869576,OM869577,OM869592,OM869653 |
| VC_1305_0 | MG945469,MG945824,MT135353,MT135400,MT135402 |
| VC_1306_0 | MG945483,MT135420 |
| VC_1307_0 | MT135397,MT135407 |
| VC_1308_0 | MG945500,MG945621,MH572369,MK765583,MT135326,OM869498,OM869502,OM869505,OM869587,OM869604,OM869633,OM869691 |
| VC_1309_0 | MT135328,OM869626 |
| VC_131_0 | HE600015,KJ716335,KM209228,KM209270,KM209320,KY942056,KY942057,MG266157,MH807812,MH807820,MN788369,MN813048,MN813049,MN813050,MN813051,MN813052,MN813053 |
| VC_131_1 | MW650886,OL656110,ON184125,ON184126,ON212266,ON862890 |
| VC_131_2 | FJ373894,JX181829,LC373201,MK509462,MT078988,OL615011,OL615013 |
| VC_131_3 | MT857002 |
| VC_1310_0 | MG945504,MG945617 |
| VC_1311_0 | MG945529,MG945568,MG945717,MG945739,MT135265 |
| VC_1312_0 | MG945533,MT135343,MT135416 |
| VC_1313_0 | MZ089798,MZ089799,MZ089800,MZ089801,MZ089803,MZ089805,OM869557,OM869648,OM869652,OM869654,OM869655,OM869666,OM869684 |
| VC_1314_0 | MZ089796,MZ089797,OM869566,OM869568 |
| VC_1315_0 | MT135349,MT135363,MT135392,MT135408 |
| VC_1319_0 | MG945554,MG945575 |
| VC_132_0 | JX878496,KX452697,MF285619,MH929319 |
| VC_132_1 | AB797215,KX147096,KY073123,MG428990,MG428991,MG770379,MK373783,MN045230,MN478483,MZ634341,ON202820,ON323462,ON399185,OX335402,OX335403,OX335405,OX335416 |
| VC_1321_0 | MZ089770,MZ089772,MZ089780,OM869514,OM869529,OM869540,OM869542,OM869544,OM869548,OM869553,OM869589,OM869590,OM869638,OM869707 |
| VC_1322_0 | MG945615,MT135372 |
| VC_1323_0 | MH572374,MH572375,MH572376,MH572414,MH572485,MH572522 |
| VC_1324_0 | MG945619,MH572524,MH992173,MZ364234 |
| VC_1325_0 | MK765555,MK765567,MK765600,MK765610,MK765617,MK765659,MK765664 |
| VC_1326_0 | MG945690,MK496692 |
| VC_1327_0 | MG945724,MT135383,MT135403,MT135456 |
| VC_1328_0 | MG945766,MG945823 |
| VC_1329_0 | MG945769,MG945792 |
| VC_133_0 | KF669653,KF669654 |
| VC_133_1 | MG428992 |
| VC_133_10 | MH709121,MN871504,MZ375357,MZ375358,OL964736,OL964737,OL964738,OL964746,OL964747,OL964753 |
| VC_133_11 | MW006476,MW006480 |
| VC_133_12 | MN871494,MN871495 |
| VC_133_13 | JN126049,LR800396 |
| VC_133_14 | KX438380,MN953776,OM681507 |
| VC_133_15 | KX765865,MW388005 |
| VC_133_16 | KJ174318 |
| VC_133_17 | MH709120 |
| VC_133_18 | KR296692 |
| VC_133_19 | MN871500,MT074475 |
| VC_133_2 | MT074472 |
| VC_133_20 | OM367911 |
| VC_133_21 | MK972690 |
| VC_133_22 | FQ312032,JX128257,JX181828,KJ174317,MH427377,MH688040,MH940212,MN066127,MN342150,MW021753,MW021757,OK322699,OP610151 |
| VC_133_23 | MT074470 |
| VC_133_24 | MF001362 |
| VC_133_25 | JN593240 |
| VC_133_26 | JN673056 |
| VC_133_27 | MG383452 |
| VC_133_28 | LR811967,LR811969,MW355478 |
| VC_133_29 | MN871475,MN871501 |
| VC_133_3 | MH370368,MH370371,MT074469 |
| VC_133_30 | KX245012,KY000003 |
| VC_133_31 | HQ259103,MH370370,MT074476,MW355450,MW355475,MW355477 |
| VC_133_32 | OM688977 |
| VC_133_33 | MT499896 |
| VC_133_34 | KY630163,KY787213,OK283619 |
| VC_133_35 | MZ156570 |
| VC_133_36 | MN871491,MN871492 |
| VC_133_37 | MN871493 |
| VC_133_38 | MN871502 |
| VC_133_39 | MT074478 |
| VC_133_4 | LR811968 |
| VC_133_40 | MH523359 |
| VC_133_41 | MT074474 |
| VC_133_42 | MN445184 |
| VC_133_43 | MN871496,MN871497,MN871498,MN871499 |
| VC_133_44 | MW021754 |
| VC_133_45 | MT074471,MT074480 |
| VC_133_46 | MW021760 |
| VC_133_47 | MT074479 |
| VC_133_48 | MK972698,MK972699 |
| VC_133_49 | ON169963 |
| VC_133_5 | MG748547 |
| VC_133_50 | ON169961 |
| VC_133_51 | ON169962 |
| VC_133_6 | MH791405 |
| VC_133_7 | MT974436 |
| VC_133_8 | MT074477 |
| VC_133_9 | KP797973,KX171211,MN384979 |
| VC_1330_0 | MG945788,MG945795 |
| VC_1331_0 | MG945798,MK765653,MT135350,MT135457 |
| VC_1332_0 | OM869639,OM869679 |
| VC_1333_0 | MG945808,MG945849 |
| VC_1334_0 | MG945821 |
| VC_1335_0 | MG945831,MT135370,MT135373,MT135418,MT135430,MT135455,MT135459 |
| VC_1336_0 | OP549864,OP549865,OP549866 |
| VC_1337_0 | MG945832,MT135421 |
| VC_1338_0 | MG945839,MH572484,MZ364252,MZ364293 |
| VC_1339_0 | MG945842,MH992159,MH992175,MH992200,MK249218,MK249229,MK249233 |
| VC_134_0 | MH363700,MK795384,OK428602 |
| VC_1340_0 | MG945850,MT135335,MT135340,MT135376,MT135393 |
| VC_1341_0 | MT135269,MT135429 |
| VC_1342_0 | MG945857,MG945859,MG945866 |
| VC_1343_0 | MH271292,MH271301,MK737941,MK801731,MK801732,MN497954,MT522001,MT522003,MT639643,MT684591,MT684592,MT818420,MW055903,MW291025,MZ681513,OK040788,OK318957,ON526977,ON637767,ON755183,OP580517 |
| VC_1344_0 | MH045557,MH045561,MH371111,MH371118,MH371124,MH399783,MH576963,MH651181,MH744416,MH779512,MH825700,MK524502,MK620897,MK801730,MK878899,MK894434,MN010755,MN010759,MN284892,MN329679,MN369745,MN428062,MN428064,MN585990,MN586008,MN586010,MT31086<br>4,MT316458,MT316459,MT316462,MT498038,MT498040,MT498046,MT498047,MT498051,MT498065,MT498067,MT521990,MT639642,MT639647,MT657332,MT776807,MT952850,MW055907,MW055909,MW291020,MW321655,MW507127,MW924642,MZ005676,MZ005678,MZ028628,MZ209304,MZ274<br>307,MZ322006,MZ322008,MZ322014,MZ322027,MZ622180,MZ648031,OK040781,OK040783,OK310504,ON081334,ON108642,ON108648,ON260812,ON260814,ON392161,ON456350,ON645341,ON645345,ON755182,ON755188,ON970573,ON970589,ON970613,OP580518 |
| VC_1345_0 | MN703407,MN703414 |
| VC_1345_1 | MH183162,MT639646 |
| VC_1346_0 | MW021755,MW021763,MW021766,OL539438,OL539439,OL539442,OL539455,OL539456,OL539464,OM135608 |
| VC_1348_0 | MH153799,MK737944,MZ456037,MZ622170,ON260825 |
| VC_1349_0 | MH271303,MK937591,MK937606,MK937608,MN813682,MN813684,OK040792 |
| VC_135_0 | MG592609,MG592621 |
| VC_135_1 | MW824370,MW824375 |
| VC_1350_0 | MT889371,ON970592 |
| VC_1351_0 | MH155870,MK240575 |
| VC_1352_0 | MH155873,MH271304,MH271312,MH271321,MH590600,MK524510,MK875798,MK977698,MN062719,MN175603,MN444877,MN585980,MN585986,MN586007,MN586042,MN586059,MT657336,MT818415,MW578837,MW601217,MZ150792,MZ322026,MZ622179,MZ648035,ON081330,ON260815,ON26082<br>0,ON970563,ON970612,OP068338,OP297543 |
| VC_1353_0 | MH229862,MH229863,MH229864,MH229865,MH536815,MH825706,MN224564,MN224565,ON456338,ON456348 |
| VC_1354_0 | MH447526,MK064562,MK064563 |
| VC_1355_0 | MH572274,MH572275,MH572276 |
| VC_1356_0 | MH572353,MH572354,MH572355,MH572357,MH572358,MH572407,MH572408,MH572516 |
| VC_1357_0 | MH572490,MH572491,MH572517,MH572518,MH572519,MH572520 |
| VC_1358_0 | MH707430,MW354669,MW354676,MW354677,MW354678 |
| VC_1359_0 | MH730557,MZ230992,MZ230993,MZ230994,MZ230995,MZ230996,MZ230997,MZ230998,MZ230999,MZ231000,MZ231001 |

| | |
|---|---|
| VC_136_0 | MH375644,MT366762 |
| VC_1360_0 | MK308638,MK757449,MK878904,MK894435,MK894438,MN310546,MN369747,MT310869,MT310875,MT310887,MT310888,MT310889,MT310890,MT310891,MT498064,MT657334,MT657337,MT684587,MT771344,MW055914,MW291018,MW365947,MW365948,MW507124,MW862991,MZ388554,OM9135 87,ON081328,ON456346,ON970584,OP297538 |
| VC_1361_0 | MH825698,MN062711,MT024863,ON260817,ON526980,OP068335 |
| VC_1362_0 | MH834607,MH834622,MK757448,MT024867,MT024870,MT889364 |
| VC_1363_0 | MH939157,MK071653 |
| VC_1364_0 | MH992154,MK249216,MK249217 |
| VC_1365_0 | MH992155,MH992196,MH992205 |
| VC_1367_0 | MH992167,MK496682 |
| VC_1368_0 | MH992171,MW202680 |
| VC_1369_0 | MK305890,MK620900,OP434453 |
| VC_137_0 | AJ609634,MK448910,OL774867 |
| VC_137_1 | MK448902 |
| VC_1370_0 | MK411746,MN586027 |
| VC_1371_0 | MK496679,MK496680,MK496776,OM869578,OM869711 |
| VC_1372_0 | MK496698,MK496772 |
| VC_1373_0 | MK496681,MK496760 |
| VC_1374_0 | MK496683,MK496761 |
| VC_1375_0 | MK496686,MK496764 |
| VC_1376_0 | MK496688,MK496690,MK496766 |
| VC_1377_0 | MK496689,MK496700,MK496784 |
| VC_1378_0 | MK496697,MK496780 |
| VC_1379_0 | MK496699,MK496785 |
| VC_138_0 | DQ340064,DQ840344 |
| VC_1380_0 | MK496701,MK496702 |
| VC_1381_0 | MK496710,MK496767,MK496768 |
| VC_1382_0 | MK496712,MK496787 |
| VC_1383_0 | MK496716,MK496790 |
| VC_1384_0 | MK496717,MK496804 |
| VC_1385_0 | MK496720,MK496796 |
| VC_1386_0 | MK496721,MK496781,MK496786 |
| VC_1387_0 | MK496725,MK496746,MK496811 |
| VC_1388_0 | MK496748,MK496815 |
| VC_1389_0 | MK496750,MK496816 |
| VC_139_0 | AY855346,LK985322 |
| VC_139_1 | DQ466086,HG531805,LN681541,LN681542 |
| VC_1390_0 | MK496753,MK496818 |
| VC_1391_0 | MK496756,MK496824 |
| VC_1392_0 | MK496758,MK496808 |
| VC_1393_0 | MK496770,MK496771 |
| VC_1394_0 | MK496820,OM869632 |
| VC_1395_0 | MK570053,MK570054,MK570055 |
| VC_1396_0 | MK719701,MK719702,MK719703,MK719704,MK719705,MK719706,MK719707,MK719708,MK719709,MK719710,MK719711,MK719712,MK719713,MK719714,MK719715,MK719716,MK719717,MK719718,MK719719,MK719720,MK719721,MK719722,MK719723,MK719724,MK719725,MK719726,MK7197 27,MK719728,MK719729,MK719730,MK719731,MK719732,MK719733,MK719734,MK719735,MK719736,MK719737,MK719738,MK719739,MK719740,MK719741,MK719742,MK719743,MK719744,MK719745,MK719746,MK719747,MK719748,MK719749,MK719750,MK719751,MK719752,MK719753,MK71 9754,MT497224,MT497225,MT497226,MT497227,MT497228,MT497229,MT497230,MT497231,MT497232,MT497233,MT497234,MT497235,MT497236,MT497237,MT497238,MT497239,MT497240,MT497241,MT497242,MT497243,MT497244,MT497245,MT497246,MT497247,MT497248,MT497249,MT4972 50,MT497251,MT497252,MT497253,MT497254,MT497255,MT497256,MT497257,MT497258,MT497259,MT497260,MT497261,MT497262,MT497263,MT497264,MT497265,MT497266,MT497267,MT497268,MT497269,MT497270,MT497271,MT497272,MT497273,MT497274,MT497275,MT497276,MT497277, MT497278,MT497279 |
| VC_1398_0 | MK765551,MK765650 |
| VC_1399_0 | MK765558,MK765581,MK765604,MK765622,MK765640,MK765654,MK765656,MK765663,MK765668 |
| VC_140_0 | DQ003640,MH341453 |
| VC_1400_0 | MK765573,MK765662 |
| VC_1401_0 | MK765575,MK765578 |
| VC_1402_0 | MK765587,MK765588,MK765598,MK765599,MK765623,MK765625,MK765628,MK765630,MK765631,MK765634,OL617020,OL617024,OL617026,OM782324 |
| VC_1403_0 | MK765589,MN988460,MN988467,MN988469,MN988471,MN988473,MN988474,MN988475,MN988478,MN988479,MN988480,MN988481,MN988489,MN988491,MN988494,MN988498,MN988499,MN988503,MN988504,MN988511,MN988518,MW960029,MW960035,MW980067,OL617021,OL617022,OL6170 25,OL617027 |
| VC_1404_0 | MK765626 |
| VC_1406_0 | MK765605,MK765641 |
| VC_1407_0 | MK929790,MW202545 |
| VC_1409_0 | MN379629,MN379636 |
| VC_141_0 | KC595514,KC595515,KC595517,KC595518,KT151956,KT151958 |
| VC_1410_0 | MN379639,MN379646 |
| VC_1411_0 | MN379643,MN379650 |
| VC_1413_0 | MN882551,MN882552 |
| VC_1414_0 | MN988461,MN988462,MN988463,MN988472 |
| VC_1415_0 | MN988485,MN988550 |
| VC_1416_0 | MN988515,MN988535,MN988540,MW980063,MW980064 |
| VC_1417_0 | MT732458,MT732459,MT732460,MT732461,MT732462,MT732463,MT732464,MT732465,MT732466,MT732467,MT732468,MT732469,MT732470,MT732471,MT732472 |
| VC_1418_0 | MT006234,MT006235 |
| VC_1419_0 | MT074137,MT074139,MT074140,MT074141,MT074142,MT074146 |
| VC_142_0 | KM983330,KM983333 |
| VC_1421_0 | MT135262,MT135274,MT135389,MT135413 |
| VC_1424_0 | MT135288,MT135290,MT135291,MT135301,MT135303,MT135307,MT135309,MT135311 |
| VC_1425_0 | MT135287,MT135294,MT135298,MT135299,MT135304,MT135306,MT135310 |
| VC_1426_0 | MT135292,MT135293,MT135296,MT135305 |
| VC_1427_0 | MT135332,MT135436 |
| VC_1428_0 | MT135346,MT135395 |
| VC_1429_0 | MT135359,MT135368,MT135374,MT135378,MT135399,MT135414,MT135419,MT135463 |
| VC_143_0 | KT626446,ON699007 |
| VC_1430_0 | MT135369,MT135396,MT135454,MT135460 |
| VC_1431_0 | MT135375,MT135438,MT135453 |
| VC_1433_0 | MT234670,ON014754 |
| VC_1434_0 | ON529863,ON529867 |
| VC_1438_0 | MT732450,MT732451,MT732452,MT732453,MT732454,MT732455 |
| VC_144_0 | MN604230,MN604698 |
| VC_1440_0 | MT952848,MZ150790,OL455892,OP068334 |
| VC_1441_0 | MT975991,OP433453 |
| VC_1442_0 | MW202613,MW202748 |
| VC_1443_0 | MW202637,MW202789 |
| VC_1444_0 | MW202486,MW202591,MW202766 |
| VC_1445_0 | MW202572,MW202631,MW202645,MW202742 |
| VC_1446_0 | MW202524,MW202718,MW202743 |
| VC_1447_0 | MW202601,MW202604 |
| VC_1448_0 | MW202608,MW202725,MW202812,MW202826,MW202842 |
| VC_1449_0 | MW202622,MW202658 |
| VC_1450_0 | MW202642,MW202843,MW202852 |
| VC_1451_0 | MW202657,MW202864 |
| VC_1452_0 | MW202753,MW202825 |
| VC_1453_0 | MW353166,MW353167,MW353168,MW353169,MW353170,MW353171,MW353172 |
| VC_1454_0 | MW462221,MZ047271 |
| VC_1457_0 | MZ089747,MZ089748 |
| VC_1458_0 | MZ089769,OM869588 |
| VC_1459_0 | MZ089783,OM869706 |
| VC_1460_0 | MZ089787,MZ089788,OM869708 |
| VC_1461_0 | MZ089790,MZ089792,MZ089794 |
| VC_1462_0 | MZ089811,OM869593,OM869669,OM869712 |
| VC_1463_0 | MZ364235,MZ364236,MZ364238,MZ364239,MZ364257,MZ364289 |
| VC_1464_0 | MZ364251,MZ364282,MZ364294 |
| VC_1465_0 | MZ447863,MZ570427 |
| VC_1466_0 | MZ474488,MZ474489 |
| VC_1467_0 | OL689226,OL689227,OL689230 |
| VC_1468_0 | OM869510,OM869512 |
| VC_1469_0 | OM869572,OM869573 |
| VC_1470_0 | OM869583,OM869672,OM869688 |
| VC_1471_0 | OM869594,OM869713 |
| VC_1472_0 | ON014753,ON014757 |
| VC_1474_0 | ON464759,ON464763,ON464764 |
| VC_1475_0 | ON529865,ON529866 |
| VC_1476_0 | OP549878,OP549879 |
| VC_1478_0 | PF3COMN,PF3COMNY |
| VC_149_0 | MT080592,MT080597,MT080598,MT080599,MT080603 |
| VC_149_1 | OM850391,OM850392,ON113344 |
| VC_149_10 | KY794641 |
| VC_149_11 | MT080582,MT080583,MT080586,MT080594 |
| VC_149_12 | MH107769,MN045228 |
| VC_149_13 | AB853330,JX878671,LR215719 |
| VC_149_14 | AB903967 |
| VC_149_15 | JQ686190,KM216423,KY779849,LC574321,LR215718,MF001365,MG557618,MH159197,MK250904,MN304941,MT786458,MW546073,MZ427930,MZ573923,OP235318,OP323059 |
| VC_149_16 | KJ888149 |
| VC_149_17 | MT080585,MT080589,MT080591,MT080601 |
| VC_149_18 | MT554104 |
| VC_149_19 | OP263968 |
| VC_149_2 | LR215720 |
| VC_149_20 | KP027446 |
| VC_149_21 | MG721208 |
| VC_149_22 | MT080593,MT080596 |
| VC_149_23 | AB853331 |
| VC_149_24 | MZ417322 |
| VC_149_25 | MZ417315,MZ417319,MZ417320,MZ417321,MZ417327,MZ417328,MZ417332,MZ417337,MZ417340,MZ417345,MZ417346,MZ417347,MZ417350,MZ417354 |
| VC_149_26 | MZ417331 |
| VC_149_27 | KX011028 |
| VC_149_28 | MN935200 |
| VC_149_29 | KY794642,KY794643,LR215721 |
| VC_149_3 | MT787017 |
| VC_149_4 | OL741057 |
| VC_149_5 | MN091625 |
| VC_149_6 | OP297178 |
| VC_149_7 | LC648442 |
| VC_149_8 | AY176327,AY954969,EU418428,FR852584,HQ163896,JX080300,JX080301,JX080302,JX080303,JX080304,JX080305,JX875065,KC012913,KF766114,KJ206559,KJ206560,KJ206561,KJ206562,KJ206563,KP687431,KP687432,KR902361,KR908644,KY581279,MF398190,MF405094,MG656408,MH136584,MH8445 28,MH844529,MK331930,MK331931,MK417514,MK417515,MK417516,MK584893,MK656892,MN047438,MN336261,MN336262,MN336263,MN539736,MN539737,MN539738,MT080600,MT080602,MT226657,MT338525,MW460250,MW528836,MW546064,MW546065,MW546066,MW546067,MW546068,M W546069,MW546070,MW546072,MW546074,OL362280,OL960567,OL960568,OL960569,OL960570,OM030343,OM373557 |
| VC_149_9 | AP018375,MT411892,OP263967 |
| VC_15_0 | KX827371,MK075002,OM373558 |

| | |
|---|---|
| VC_15_1 | AY954954,MK075001,MK075004,MK450538,OM373550,OM373554,OM373555,OM373556,OM373561 |
| VC_150_0 | KF021268,KP027447,MH321490,MH321491,MH542234,MT596501,MT596502,MT596503,MT596504,MT596505,MT596506,MZ417323,MZ417325 |
| VC_151_0 | KX171212,KX171213,MW430345,MZ417329,MZ417330,MZ417344 |
| VC_152_0 | LC647030 |
| VC_152_1 | MZ417326 |
| VC_153_0 | AP019522,JX194239,JX846612,JX846613,KP881332,KX532239,KY779848,LC492751,LC492752,MT080584,MT080587,MT080588,MT080590,MT080595,MW363798,MW546071,MW546075,MW546076,MW546077,MZ643272,ON814134,ON814135,ON814136,ON866946,OP263969 |
| VC_153_1 | LC576631 |
| VC_154_0 | AB854109,KY117485,MN844877,OL634959 |
| VC_155_0 | FN594518,MT104468,MT104469,MT104470,MT104471,MT104472,MT104476,MT104477,MT354570,MW627366,MZ868714,MZ868715,MZ868716,MZ868717 |
| VC_156_0 | FR751545,KM247287,MK629528,MN850568,MN850570,MN850583,MN850592,MN850608,MN850617,MN850624,MN850625,MW250275 |
| VC_157_0 | MK962628,MK962638,MK962640 |
| VC_159_0 | OM913597,OM913598 |
| VC_16_0 | AB012573,AB012574,AB572858,AF452449,AF453500,AY242528,CP017905,CP017906,D89074,FJ904927,MN536023,MN690600 |
| VC_160_0 | MF150911,MN996301 |
| VC_161_0 | AB863625,MT740737,MT740741 |
| VC_162_0 | AY349011,FJ937737,JN662425,JX104231,MH638294,MT361768,MT740727,MT740730,MT740731,MT740732,MT740733,MT740736,MT740738,MT740739,MT740740,MT740742,OM283127,OV121131 |
| VC_163_0 | MW202475,MW202867 |
| VC_164_0 | MK268344,MK773491,MT001829 |
| VC_164_1 | MT446416 |
| VC_164_10 | AB897757,JQ513383,MT708547,OP617745,OU509535 |
| VC_164_11 | KC295538,KM507819,LC494302,LT603033,MH383160,MH791411,MK327931,MK817115,MW822008,ON568341 |
| VC_164_2 | MH494197 |
| VC_164_3 | ON286972 |
| VC_164_4 | JN882285 |
| VC_164_5 | KU574722 |
| VC_164_6 | OP292654 |
| VC_164_7 | KY630187,MT939486,MT939487,MT939488,MW879342,MW879343,MW879344 |
| VC_164_8 | LR596615,LT960551,LT960552 |
| VC_164_9 | MG592671 |
| VC_165_0 | LC727697,LC727698,LR588166,MK797984 |
| VC_166_0 | AM749441,DQ163916,JX131330,KC758116,KT734862,KX198613,KX961385,KY000218,KY000219,KY000220,KY000221,MF490239,MH791403,MH791406,MN871456,MN871463,MN871469,MN871478,MN871482,MN871486,MT118302,MZ089734,MZ089741,OL763419,OM809160,ON165687 |
| VC_167_0 | AB910392,MK443266,MK803322 |
| VC_169_0 | JQ067089,JQ067093,KU204984,KX241618,LR743532,MG018224,MT664984,OK275492,OK275493,OL674541,ON005621,ON189045,ON189047,ON331942,ON529854,ON932079,ON932080,ON932084,OP361299,OP380269 |
| VC_17_0 | AB043678,AB043679,MN719123,MT188662,MT188663,MT188664,MT188665,MT188666,MT193890 |
| VC_170_0 | KT001913 |
| VC_170_1 | KY006853 |
| VC_171_0 | KP202969,KP202970,MH746817,MK962627,MK962633,MK962634,MT241605,MT241607 |
| VC_172_0 | KF626665,KF626668,LR743524,MZ326855,ON189048 |
| VC_173_0 | KT887558,MG707188 |
| VC_174_0 | AB910393,LC102729,OK094707 |
| VC_175_0 | MG592578,MG592660,MW865292,OX241572 |
| VC_176_0 | AF165214,HQ711985 |
| VC_176_1 | KT887557,ON857940 |
| VC_176_2 | OM141125 |
| VC_176_3 | KT968831 |
| VC_177_0 | AB910602,HG315669,KX181651,KY853667,MN335248 |
| VC_178_0 | MH206183,MH206184,MH218848,MK512531,MN263053 |
| VC_179_0 | AY394005,DQ631426,DQ873690,EU272036,EU272037,HE584812,HM624080,JN808773,JN811560,JQ067085,JQ762257,JX434030,JX434031,JX434032,JX434033,KF475786,KJ477077,KM233689,KT968832,KU199707,KU199708,KU199709,LN610590,LN907801,LT608331,LT999987,MH643777,MH643778,MK1<br>44667,MK511017,MK511018,MK511019,MK511020,MK511021,MK511022,MK511023,MK511024,MK511025,MK511026,MK511027,MK511028,MK511029,MK511030,MK511031,MK511032,MK511033,MK511034,MK511035,MK511036,MK511037,MK511038,MW390886,OM541307 |
| VC_18_0 | AB016282,HM072038,OM236514,ON528935 |
| VC_180_0 | AB916497,ON170148,ON170170,ON170354 |
| VC_182_0 | JF974307,JF974308,KT381864,KT381865,KX077179 |
| VC_183_0 | KF302035,MZ568827 |
| VC_184_0 | MH727593,MZ547449 |
| VC_185_0 | LR797314,MF959999,MK088078 |
| VC_187_0 | KX961630,MN150686,OK499977,OK499983 |
| VC_187_1 | JN797797,JN797798,KT187252,KT224359,KX987999,MH487649,MN013089 |
| VC_187_2 | KJ081346,KJ676859 |
| VC_187_3 | JX094431,KJ451625,KT995480,KU577463,KX495186,MG602477,MH638309,MH638312,OL964058 |
| VC_187_4 | MN364664 |
| VC_188_0 | AB971354,AF059242,AF059243,EF068134,FJ483840,FJ483841,FJ483842,FJ483843,FJ483844,FJ539132,FJ539133,FJ539134,FJ539135 |
| VC_189_0 | AJ604530,AJ604531 |
| VC_190_0 | KP143763,KX130863 |
| VC_190_1 | MK373792 |
| VC_190_10 | KR422353 |
| VC_190_11 | AC171169 |
| VC_190_12 | LR027384 |
| VC_190_13 | MH370377,MH370378,MN970211,MN994502 |
| VC_190_14 | LR597655,MN022787,MN316588 |
| VC_190_15 | KJ190157,KY677846,MN994496 |
| VC_190_16 | OK272480 |
| VC_190_17 | AP019559 |
| VC_190_18 | MF402939 |
| VC_190_19 | MZ501075 |
| VC_190_2 | MK373797,MK373799 |
| VC_190_20 | LN887948,MK972715,MK972716,MT843274 |
| VC_190_21 | KY114934 |
| VC_190_22 | KM979354,KM979355,MH203051 |
| VC_190_23 | HQ406778,KY963424,MH102285,MN026741,MT311645,MW149273,MW149275,OK108607,OK254197,OP394141 |
| VC_190_24 | HQ665011,LR597659,MZ398246,OK143207,ON185585 |
| VC_190_25 | MN956514 |
| VC_190_26 | MG387042 |
| VC_190_27 | KR233164 |
| VC_190_28 | OK665835,ON185581,ON185582,ON185583,ON185584 |
| VC_190_29 | MT080103 |
| VC_190_3 | MN994504 |
| VC_190_30 | MF774688 |
| VC_190_31 | MG065692 |
| VC_190_32 | MZ501058 |
| VC_190_33 | MZ501103 |
| VC_190_34 | MN445182 |
| VC_190_35 | OK272477 |
| VC_190_36 | MT653143 |
| VC_190_37 | MT653146 |
| VC_190_38 | AY543070,AY587007,AY692264 |
| VC_190_39 | MT233524 |
| VC_190_4 | MT577844 |
| VC_190_40 | MG969412,MW006479 |
| VC_190_5 | OM386657 |
| VC_190_6 | MZ501074 |
| VC_190_7 | MF431730,MF431731,MF431732,MF431733,MF431734,MF431735,MT074448 |
| VC_190_8 | OM386660 |
| VC_190_9 | MK373796 |
| VC_191_0 | MW006478,MW006481 |
| VC_191_1 | MK370036 |
| VC_191_10 | MN197465 |
| VC_191_11 | MF681663 |
| VC_191_12 | MT074464 |
| VC_191_13 | MK907267,MK907268,MK907269,MK907270,MK907271 |
| VC_191_14 | LR027389 |
| VC_191_15 | OL451946,OL502173 |
| VC_191_16 | KY787212,MF001363,MH370369,MK393882,MK947458,MK972717,MN218190,MN218191,MN379739,MN994501,MT074450,MT074458 |
| VC_191_17 | CP000917,MK947459,MZ501105,OL770310 |
| VC_191_18 | MT833283,MZ501072 |
| VC_191_19 | KM236244,KU927497,KX017521,LC659915,MG065642,MH631454,MK728824,MK728825,MN187969,MW357609,OK288022,OP045498 |
| VC_191_2 | MZ501109 |
| VC_191_20 | MK005300 |
| VC_191_21 | MZ501076 |
| VC_191_22 | MW567727 |
| VC_191_23 | MH370379 |
| VC_191_24 | MZ570151 |
| VC_191_25 | MW206381 |
| VC_191_26 | HE956708 |
| VC_191_27 | MK867835 |
| VC_191_28 | MZ501108 |
| VC_191_29 | MH370375 |
| VC_191_3 | MK972704,MK972707,MK972709,MT074453,MT074457 |
| VC_191_30 | MT074447,MT074461 |
| VC_191_31 | MK770410,MK972708 |
| VC_191_32 | MF431736,MF431737,MF431738,MF431739,MF431740,MF431741,MK050846,MK809530,MT074449,MT074465,MT653145,ON526840 |
| VC_191_33 | MW423798 |
| VC_191_4 | MH370366,MT074455 |
| VC_191_5 | MH370386,MT004791,MT074452,MT074456,MT074460,MT074463 |
| VC_191_6 | MT074468 |
| VC_191_7 | MT074467 |
| VC_191_8 | MH370376,MT074454 |
| VC_191_9 | MH370367,MH370380,MT074459,MT074466,MW411578 |
| VC_192_0 | KX845404,MG459987,MK521907,MK630230,MN101228,MN163280,MN434094,MN434096,MT380195,MW713578,MZ571828,MZ571830,MZ634344,ON637170,OP171943 |
| VC_192_1 | MZ079855 |
| VC_192_2 | MN478377 |
| VC_193_0 | KY363465,MW057857 |
| VC_195_0 | MG676224,MG676225 |
| VC_195_1 | MK318083 |
| VC_196_0 | MN095770,ON287373 |
| VC_197_0 | AE002163,CMU41758,MG641183,MN862345,MW202581 |
| VC_198_0 | AJ270057,AJ550635,AY769964,MG883723,MG945324,MG945546,MG945571,MG945669,MH572277,MH572280,MH572282,MH572290,MH572330,MH572334,MH572422,MH572438,MH572444,MH572448,MH572452,MH572454,MH992157,MH992174,MH992182,MH992210,MN862343,MW202732,MW202889<br>,OL617035 |
| VC_199_0 | KC131021,MH572341,MH572342,MH572343,MH572399,MH572400,MH572401,MH572402,MH572403,MH572453,MH572513,MH572514 |
| VC_2_0 | AB008550,MK034952,MK511066,Y13918 |
| VC_20_0 | KT372714,KT852578,KX190835,KX534340,MN065183 |
| VC_200_0 | MH992181 |
| VC_201_0 | MH572434,MH572475,MZ286294 |

| | |
|---|---|
| VC_202_0 | AF009630,AF011378,DQ054536,DQ227763,DQ227764,EU221285,FJ848881,FJ848882,FJ848883,FJ848884,FJ848885,GQ979703,KC182542,KC182543,KC182544,KC182545,KC182546,KC182547,KC182548,KC182549,KC182550,KC182551,KC182552,KC522412,KF676640,KM091442,KP793104,KP793105,KP793108,KP793109,KP793111,KP793113,KP793115,KP793116,KP793117,KP793118,KP793123,KP793124,KP793129,KP793134,KX346246,KX379667,KX379668,KX379670,KX379671,KY554760,KY554761,MF346933,MF448554,MF448555,MF448556,MF448557,MF448558,MF448559,MF448560,MF448561,MF448562,MF448563,MF448564,MF448565,MF448566,MF448567,MF448568,MF448569,MF448570,MF448571,MF448572,MF775703,MF775704,MF775705,MG253647,MG253648,MG253649,MG253650,MG253651,MG253652,MG253653,MG309717,MG309718,MK301437,MN552148,MN552149,MN552150,MN689514,MN689517,MN689518,MN689519,MN689520,MN689522,MN689523,MN689524,MN689525,MW032477,MW041632,MW041633,MW041634,MW041635,MW041636,MW041637,MW041638,MW041639,MW041640 |
| VC_209_0 | KY888143,KY926698,KY926699,MZ368140 |
| VC_21_0 | JF974314,KJ746502,MF614627,MF614628 |
| VC_211_0 | AP008983 |
| VC_211_1 | KP144388 |
| VC_211_2 | KT151959 |
| VC_211_3 | MT325768 |
| VC_213_0 | KT429160,MT880872 |
| VC_214_0 | ON210834,ON210835 |
| VC_214_1 | OM654379 |
| VC_215_0 | DQ398043,KF017927,KF416344,MG099947,MN585989 |
| VC_215_1 | AF022214,AP018476,AP018480,KC787102,KC787105,KC787106,KX574455,KX758538,KX897981,LC730318,MH513985,MK494087,MN234169,MT310876,MT310883,MT310884,MT310885,MW924646,OP297553 |
| VC_215_2 | JF946695,KT381276,MF919536,MH020237,Z18946 |
| VC_216_0 | MF185727,MF324899,MK801735,MN369748 |
| VC_216_1 | MK814748 |
| VC_216_2 | MN585968,MN586002 |
| VC_216_3 | GU339467,JN408461,KF024721,KU761558,KU761559,KX641263,MF140403,MH825709 |
| VC_216_4 | KJ959632,KM197169,KM677210,KU297783,KX683875,MF668269,MF919538,MH825701,MK061408,MK284522,MK967395,MZ322019,OM203159 |
| VC_216_5 | MW601216,MZ209305 |
| VC_216_6 | AP018479,JN408460,JX899358,KF981601,KJ410132,KJ410134,KM463009,KM591906,KT588442,KU985093,KU985095,KX576639,KX619650,KY965065,LC730320,MF472896,MF919490,MF919500,MF919531,MH051252,MH077576,MH338234,MH697591,MH825704,MK524491,MK919472,MN234203,MN585981,MN585983,MN585998,MT310877,MT310886,MT498041,MT498060,MT498066,MT658804,MW924643,MZ355728,MZ622171,MZ622182,MZ820090,OP297537 |
| VC_216_7 | MT310858 |
| VC_217_0 | EU744250,KU568494,KX758539,MK279857,MN703408 |
| VC_217_1 | MN062708 |
| VC_218_0 | KF861510,MN096376,MT639645 |
| VC_219_0 | JF957060,KC661275,KP273225,MG793454,MT553337 |
| VC_22_0 | AB036666,HQ906662,HQ906663,HQ906664,KX522565,MK976036,MN180248,MN180249,MN180250,MN180251,MN180252 |
| VC_220_0 | KR080193,MN310541,MT522004,OM203161 |
| VC_221_0 | KU695581,KY471460,MF140410,MH338236,MH338237,MH536828,MK494130,MK524507,MK524511,MK524513,MK524514,MK524519,MN310547,MN586006,MN703405,MN892487,MW055904,MZ747517,MZ747521,ON108640,OP297532,OP297549 |
| VC_222_0 | KX657793,MH513984,MK494113,MK524530,OM203158 |
| VC_223_0 | JF704097,JF937092,JF937094,JN049605,JN698999,JN699019,KF560332,KF560333,KF560334,KM401838,KU695582,KU985096,KX375815,KX576638,KX641261,MF668288,MG099945,MG099952,MH020240,MH051253,MH155876,MH779517,MK359352,MK359360,MK494090,MK894433,MN586039,MN728683,MT024861,MT657338,MW822143,MZ005685,MZ171376,MZ322007,MZ456034,MZ681516,OM913585,OP434447 |
| VC_224_0 | KU998249,KU998250,KU998251,KX557280,KX557283,KX557284,MF668282,MH001454,MH316566,MH513974,MK967382,MN284898,MN369756,MT522007,MZ028625,MZ622176,MZ681508,OM818328,OP021679 |
| VC_225_0 | JF704110,JN699005,KT285706,KU613353,KU716094,KU716095,KX585252,KX664447,KX685355,MG872835,MG872840,MG872842,MH371122,MK061409,MK468591,MK494108,MK524489,MK559428,MK977713,MN234167,MN234200,MN234209,MN234235,MN310542,MN369741,MN369763,MN369764,MN585974,MN585978,MN585979,MN585994,ON724016 |
| VC_227_0 | AF334111,HM066936 |
| VC_228_0 | AF063097,AY135486,AY135739,AY251033,B1U32222,HQ110084,KC139521,KC618326,KF322032,KR073660,KR073661,KT630644,KX833211,LR595869,LR595870,LR595873,LR595883,LR595885,LR595887,MF276773,MK416018,MK907236,MK907237,MK907238,MK907239,MK972710,MK972711,MN016939,MT239398,MT374855,MT374857,MT374858,MZ420230,MZ520833,OK490408,OK490461,OK490464,OK490467,OK490470,OK490473,OM046626,OM046627,OM046628,OM140653,OM475426,OM475429,OM475431,OM475433,OM475434,ON146439,ON146440,ON239131,ON470601,ON470606,ON470613,ON782130,OU784549,OU784746,OU784750,OV192280 |
| VC_229_0 | CP025874,CP025901,CP025923,CP052639,KP063541,KY271397,LR595891,MG589387,MK416015,OK490394,OK490396,OK490398,OK490400,OK490402,OK490404,OK490406,OK490412,OK490414,OK490422,OK490424,OK490426,OK490428,OK490432,OK490435,OK490437,OK490440,OK490443,OK490446,OK490449,OK490453,OK490456,OK490459 |
| VC_23_0 | AY133112,FN297812,JX556417 |
| VC_230_0 | JQ780327,MK524177,OL964755 |
| VC_230_1 | HM208537 |
| VC_231_0 | MT366580,MZ496293,OP121182 |
| VC_232_0 | AF065411,AF301375 |
| VC_233_0 | MH674343,MN737622 |
| VC_234_0 | AP018469,AP018470,AP018471,JN831653,KJ944841,KT361920,KX621007,KY087992,MF140402,MF140435,MH051258,MH926058,MK224497,MK524485,MK524488,MN369761,MN945904,MT310882,MW601218,ON526978,ON645342 |
| VC_236_0 | HM152764,HM152767,JF704105,JF704106,JN185608,JN643714,KJ567045,KM677211,KX585253,KX641264,KX657794,KX657795,KX657796,KY380102,MF140412,MF140428,MF140430,MF377440,MF472892,MF668267,MF919532,MG962364,MG962371,MH001453,MH051246,MH077577,MH371113,MH509444,MH513977,MH536821,MH536822,MH697578,MH727544,MK016503,MK112537,MK460247,MN234182,MN234197,MN234198,MN234205,MN234230,MN234231,MN369742,MN369746,MN908692,MW712722,MW712723,MW712730,MW712733,MW712734,MW862979,MW862984,MW862985,MW862990,MZ005674,MZ355721,MZ648037,OK040778,OK040782,OK040784,OK310505,OM818330,OM913584,ON081329,ON081331,ON260816,ON456339,ON456353,ON526984,ON687735,ON687736,ON724013 |
| VC_236_1 | MF140416,MN234165,MN234174,MN234201,MN234226 |
| VC_237_0 | KF713486,MF185722,MK310138,MK494119 |
| VC_237_1 | JF937104,JX042579,KP027199,KR080206,KX683428,KY087993,MF140398,MF140406,MF324915,MF373841,MN234199,MT723940,MW601222 |
| VC_237_2 | KY945355,MF324908,MF324909,MF324911,MF324912,MF324913,MF324914,MN234176,MN369762,MN945901,ON456351 |
| VC_237_3 | MH910038,MH910039,MH910042,MK376955,MK460246,MN284895,MT310859,MT310860 |
| VC_238_0 | JN243855,KJ567042,KM363596,KM923971,KX585251,KX636165,MF185717,MF185719,MF185720,MG962374,MH051255,MH576966,MH651171,MH697592,MW435854,MZ322016,MZ958747 |
| VC_24_0 | JX507079,MH142219,MH142220 |
| VC_240_0 | MW314859,MW584165,MW584169,MW584207 |
| VC_241_0 | MF975720,MT121966 |
| VC_242_0 | KT588074,MK340941 |
| VC_242_1 | KT630649,MK416007,MK416019,MK422455,MK433579,MK448230,MN098327,MN166823,MT135176,MT701587,MW057855,OK665842,OL791266,OM284015,OM621816,ON470579,ON470582,ON470588,ON470593,OP114732 |
| VC_243_0 | MF695815,MK416014 |
| VC_244_0 | KY271401,MK416017,OK490430,OK490433,OK490438,OK490441,OK490444,OK490447,OK490450,OK490451,OK490454,OK490458 |
| VC_245_0 | KY653119,LN997803,LR597642,ON470580,ON470583,ON470586,ON470600,ON470607,ON470612,ON470621 |
| VC_246_0 | AY583234,AY583235,AY583236 |
| VC_247_0 | AF234172,AF234173,AF503408,FO818745,KU760857,MH160767,MH422554,MH445380,MH445381,MZ188976,ON018986,ON470615,ON470617,OP279344 |
| VC_247_1 | MF356679 |
| VC_248_0 | HQ316581,MG592412,MG592507,MG592513,MG592524,MG592527,MG592588,MG592589,MG592669,MG592670,MZ568828 |
| VC_249_0 | ON170062,ON170209,ON170231,ON170302,ON170328 |
| VC_25_0 | ON170017,ON170042,ON170051,ON170056,ON170060,ON170066,ON170072,ON170166,ON170169,ON170274,ON170278,ON170357 |
| VC_251_0 | AF232233,JX495041,JX495042,JX495043,LN898172,MH719189,MH719190,MH719191,MH719192,MH719193,MH719194,MH719195,MK511057,MK511058,MK511059,MK511060,MK511061,MK511063,MK511064,MK511065,MK934841,MN536027 |
| VC_252_0 | AF085222,AF115102,AF115103,AF145054,FJ236310,KT717083,KT717084,KT717085,KY705251,KY705256,KY705257,KY705259,KY705260,KY705261,KY705262,KY705263,KY705264,KY705265,KY705266,KY705267,KY705270,KY705271,KY705272,KY705273,KY705274,KY705275,KY705276,KY705282,KY705283,KY705284,KY705287,KY705288,KY705289,KY705290,MF161328,MF580759,MF580760,MF580761,MF580762,MF580763,MF580764,MF580765,MF580766,MF580767,MF580768,MF580769,MF580770,MF580771,MF580772,MF580773,MF580774,MF580775,MG708273,MG708274,MG708275,MH000602,MH000603,MH000604,MH375594,MH375595,MH375596,MH375597,MH375598,MH375599,MH375600,MH892351,MH892352,MH892353,MH892354,MH892356,MH892357,MH892358,MH892359,MH892360,MH892361,MH892375,MH892377,MH892378,MH892379,MH892380,MH892381,MH892382,MH937458,MH937459,MH937461,MH937462,MH937465,MH937466,MH937467,MH937468,MH937469,MH937470,MH937471,MH937472,MH937474,MH937477,MH937480,MH937481,MH937482,MH937483,MH937485,MH937486,MH937487,MH937488,MH937489,MH937490,MH937491,MH937492,MH937493,MH937495,MH937496,MH937497,MH937499,MH937500,MH937501,MH937504,MH937506,MH937508,MH973661,MH973662,MZ090946 |
| VC_253_0 | MK448899 |
| VC_254_0 | AF158600,AY699705,EF529515,FJ226752,HE861935,HG380752,HG424323,KY705258,KY705268,KY705269,KY705277,KY705278,KY705279,KY705280,KY705281,KY705285,KY705286,MH375593,MH892362,MH892363,MH892364,MH892366,MH892376,MH892383,MH892384,MH892385,MH937457,MH937460,MH937463,MH937464,MH937473,MH937475,MH937476,MH937478,MH937479,MH937484,MH937494,MH937498,MH937502,MH937503,MH937505,MH937507,MH937509,MH937510,MH937511,U88974 |
| VC_256_0 | KU678389,KU678390,KU678391,KU678392,KX879641,KX879642,MH892350,MH892365,MH892368,MH892369,MH892370,MH892371,MH892372,MH892374 |
| VC_259_0 | MK448824,MK449002,MK449003 |
| VC_26_0 | AY714348,JN848801,MN200777,MN200778 |
| VC_260_0 | AF176025,AF323669,AF489521,AJ245616,LT985568,LT985571,LT993224,MN534316,MN534317,MN689509,MN689516,MN689521,MN689530 |
| VC_262_0 | KX160204,KX160219,KX160220,KX456211,MN534320 |
| VC_263_0 | AF323668,LT985569,LT985574,LT985578,LT985583,LT993229,LT996082 |
| VC_265_0 | DQ394807,DQ394808,DQ394809,DQ394810,KX456206,KX456208,KX456209,KX456210,KX456213,LT985575,LT985576,LT996081 |
| VC_267_0 | GQ452243,KT945994 |
| VC_268_0 | JX564242,JX567312,KX160206,KX160209,KX160210,KX160211,MN552144 |
| VC_269_0 | KJ608189,KU981050,KY744240 |
| VC_27_0 | AB063393,GQ403789 |
| VC_272_0 | AJ251789,AP018361,AY131267,AY605066,EU246945,KC171646,KC171647,MF999224,MH983004,MN830258,MT956952 |
| VC_274_0 | AJ421943,MH220877 |
| VC_276_0 | AF133242,AJ969242 |
| VC_277_0 | AF155037,MG966533,MK424903 |
| VC_278_0 | AY029185,AY526908,AY526909 |
| VC_279_0 | AY605181,MH128984 |
| VC_28_0 | MW722521,MW790496 |
| VC_28_1 | AB102868,AB362338,AB426868,AF217253,AF527608,AY052766,BK000583,CP051272,CP051278,CP051279,CP051285,CP051288,DQ003260,EU570103,FJ000341,GQ422450,GQ422451,GU573886,JF900176,JQ806763,KR296686,KR296687,KR296688,KU878967,KU927491,KU927492,KU927494,KU927495,KU927496,KU927498,KX776161,LC730322,LT714109,MF188997,MG407615,MG969415,MH370387,MK770414,MK770415,MK972685,MK972686,MK972687,MK972692,MK972693,MK972694,MN161573,MT774487,MT932212,MW013502,MW013503,MW355480,OP265889,OP377721 |
| VC_280_0 | GQ421471,MT084350 |
| VC_281_0 | MK733262,MK733263,MK733264 |
| VC_283_0 | MN508614,MN508618 |
| VC_283_1 | MZ502379 |
| VC_283_10 | AP011113 |
| VC_283_100 | MT515752,MT526905 |
| VC_283_101 | LR597660 |
| VC_283_102 | OK570375 |
| VC_283_103 | ON528735 |
| VC_283_104 | OL310488 |
| VC_283_105 | MZ234013 |
| VC_283_106 | MZ753803 |
| VC_283_107 | ON528739 |
| VC_283_108 | OK148441 |
| VC_283_109 | KY608967,OK275722 |
| VC_283_11 | ON528741 |
| VC_283_110 | MN508616,MN508617 |
| VC_283_111 | KY290975 |
| VC_283_112 | OL870317 |
| VC_283_113 | LR215724 |
| VC_283_114 | KX130861 |
| VC_283_115 | JN986846 |
| VC_283_116 | MZ501089 |
| VC_283_117 | MZ234031,MZ234035,MZ234038 |
| VC_283_118 | MZ501113,OL741432,OL870316 |
| VC_283_119 | MK327939 |
| VC_283_12 | MK373785 |
| VC_283_120 | MK962752 |
| VC_283_121 | MK327940,MK327945 |
| VC_283_122 | OK274152 |
| VC_283_123 | OL362041 |
| VC_283_124 | MK373779 |
| VC_283_125 | KX130862 |
| VC_283_126 | OM386666 |
| VC_283_127 | MK327929 |
| VC_283_128 | OK076929 |
| VC_283_129 | MH992510 |
| VC_283_13 | ON528745 |
| VC_283_130 | MN022785 |
| VC_283_131 | MN716856 |
| VC_283_132 | ON528740 |
| VC_283_133 | MK327936 |
| VC_283_134 | MK327944 |
| VC_283_135 | OP168898 |
| VC_283_136 | MT515753,MT515755 |
| VC_283_137 | LR597657 |
| VC_283_138 | MG867727 |
| VC_283_139 | KX009778 |
| VC_283_14 | MF001360 |

| | |
|---|---|
| VC_283_140 | OV876900,OV877085 |
| VC_283_141 | LR746311 |
| VC_283_142 | ON528726,ON528744 |
| VC_283_143 | KX452698 |
| VC_283_144 | MW341595 |
| VC_283_145 | KM501444 |
| VC_283_146 | ON528746 |
| VC_283_147 | MZ501096 |
| VC_283_148 | JN202312 |
| VC_283_149 | ON528736 |
| VC_283_15 | LC465543 |
| VC_283_150 | MZ065353 |
| VC_283_151 | MH051915 |
| VC_283_152 | OL960580 |
| VC_283_153 | MW749000 |
| VC_283_154 | HM035025 |
| VC_283_155 | MT630408,MZ681930 |
| VC_283_156 | MK327942 |
| VC_283_157 | OL960581 |
| VC_283_158 | MF327007 |
| VC_283_159 | MH837626 |
| VC_283_16 | MK327937 |
| VC_283_160 | ON229909 |
| VC_283_161 | MT179807 |
| VC_283_162 | ON720975 |
| VC_283_163 | KF925357,KR233165 |
| VC_283_164 | MK886800,MZ502380 |
| VC_283_165 | OK018184 |
| VC_283_166 | MZ189262 |
| VC_283_167 | OM135583 |
| VC_283_168 | MW822007 |
| VC_283_169 | MK962750 |
| VC_283_17 | MK327946 |
| VC_283_170 | MH051916 |
| VC_283_171 | MT682709 |
| VC_283_172 | MW883059 |
| VC_283_173 | MT968995 |
| VC_283_174 | MZ417521 |
| VC_283_175 | MW822011 |
| VC_283_176 | MZ150757 |
| VC_283_177 | OL770073 |
| VC_283_178 | ON645936 |
| VC_283_179 | MN580668 |
| VC_283_18 | MK327928 |
| VC_283_180 | MT515756 |
| VC_283_181 | MH243439 |
| VC_283_182 | LC348379 |
| VC_283_183 | MF001359 |
| VC_283_184 | MT884006,MT884007 |
| VC_283_185 | MN994497 |
| VC_283_186 | ON528742 |
| VC_283_187 | AP018932 |
| VC_283_188 | MH992121 |
| VC_283_189 | MF158046 |
| VC_283_19 | MF327008 |
| VC_283_190 | MK977694 |
| VC_283_2 | HM997020,KP869105 |
| VC_283_20 | MF327009 |
| VC_283_21 | MN781580 |
| VC_283_22 | OP197928 |
| VC_283_23 | MT828552 |
| VC_283_24 | MG775043 |
| VC_283_25 | KU867876 |
| VC_283_26 | OL615012 |
| VC_283_27 | MT176424 |
| VC_283_28 | MT176426 |
| VC_283_29 | ON528727 |
| VC_283_3 | FJ839693,KM607004,OK040907 |
| VC_283_30 | ON528728 |
| VC_283_31 | MN895437 |
| VC_283_32 | MZ436830 |
| VC_283_33 | LN881726,LN881729,LN881734,LN881737 |
| VC_283_34 | MH809535,MT515751,MT828551,MT828553 |
| VC_283_35 | MK962755 |
| VC_283_36 | MZ501065 |
| VC_283_37 | LN881732 |
| VC_283_38 | LR990704 |
| VC_283_39 | MN895434 |
| VC_283_4 | MZ291552,OP555980,OP555981,OP555982,OP572230 |
| VC_283_40 | MZ234018 |
| VC_283_41 | MZ501046 |
| VC_283_42 | MZ501050 |
| VC_283_43 | MK373786 |
| VC_283_44 | MN895438 |
| VC_283_45 | MH355584 |
| VC_283_46 | MZ501066 |
| VC_283_47 | MZ501067 |
| VC_283_48 | MN895436 |
| VC_283_49 | MT176425 |
| VC_283_5 | LC620505 |
| VC_283_50 | MZ501106 |
| VC_283_51 | MN895435 |
| VC_283_52 | MH553563 |
| VC_283_53 | MK373781 |
| VC_283_54 | MT682713 |
| VC_283_55 | ON528743 |
| VC_283_56 | MZ726795 |
| VC_283_57 | KT184311 |
| VC_283_58 | KX130864 |
| VC_283_59 | OK272473 |
| VC_283_6 | ON782582 |
| VC_283_60 | OK272476 |
| VC_283_61 | MT446387 |
| VC_283_62 | OK272484 |
| VC_283_63 | KT184312 |
| VC_283_64 | MF158045 |
| VC_283_65 | OL956808 |
| VC_283_66 | KT184308 |
| VC_283_67 | LR877332,MT515754 |
| VC_283_68 | MW749001 |
| VC_283_69 | HE956711 |
| VC_283_7 | MW286157 |
| VC_283_70 | MZ501098 |
| VC_283_71 | LR215723 |
| VC_283_72 | MT682710 |
| VC_283_73 | LN881728,LN881733,LN881736 |
| VC_283_74 | MT682714 |
| VC_283_75 | MZ170040 |
| VC_283_76 | MK373787 |
| VC_283_77 | MT682712,MZ170041 |
| VC_283_78 | MZ234027,MZ234030,MZ234034,MZ234036,MZ234037,MZ234039,MZ234040,MZ234041,MZ234042,MZ234043,MZ234051,MZ234052,MZ234053 |
| VC_283_79 | MF001354 |
| VC_283_8 | KU925172 |
| VC_283_80 | MT764206 |
| VC_283_81 | KR269718 |
| VC_283_82 | KM606994,KM606995,KM606996,KM606997,KM606998,KM606999 |
| VC_283_83 | KM607000 |
| VC_283_84 | FJ839692 |
| VC_283_85 | KX828711 |
| VC_283_86 | ON528734 |
| VC_283_87 | ON528738 |
| VC_283_88 | AP018814,MH550421 |
| VC_283_89 | MG781190,MG781191 |
| VC_283_9 | MZ501052 |
| VC_283_90 | KF208315 |
| VC_283_91 | AF158101,HM137666,KJ477684,KJ477685,KJ477686,KM607002,KM607003,MT984581,MZ375356,OL964735,OL964745 |
| VC_283_92 | AP018813,LC348380,MH751506,OM339528 |
| VC_283_93 | MT932213 |
| VC_283_94 | DQ904452,KM607001 |
| VC_283_95 | JX128259 |
| VC_283_96 | ON506924 |
| VC_283_97 | LR215722 |
| VC_283_98 | ON210139 |
| VC_283_99 | KJ668714 |
| VC_284_0 | KJ025957,MN095771,OM457001 |
| VC_286_0 | MW512264,MZ501047 |
| VC_286_1 | KR422352,LR027383,LR027386,LR027387,LR027390,LR027391,MH243438,MK327934,MK327943,MK373778,MK373782,MK373784,MK962754,MK962756,MK962757,MN850579,MN850590,MZ234025,OL960576,OL960577,OM960734 |
| VC_286_10 | AY303349,KF562341,KM407600,KY703222,MH992122,MT446390,MZ501056,OM982647,ON528737 |
| VC_286_11 | MH359124,MK524178,MW882907 |
| VC_286_12 | MG589383,MZ189261,OL770074 |
| VC_286_13 | KF582788,MF044457,MK639187,MT782071,MW630116,MZ311865,MZ560701,MZ868638,OV754623,OV877511 |
| VC_286_14 | JX536493 |

| | |
|---|---|
| VC_286_15 | MK047717,ON528725 |
| VC_286_16 | KX130865 |
| VC_286_17 | KP869101,KP869104,KP869111,KT184309,KT184310,MK047718,MN648445,MN850622 |
| VC_286_2 | MN850651 |
| VC_286_3 | MK373774,MK373775 |
| VC_286_4 | MK327933 |
| VC_286_5 | MT446423 |
| VC_286_6 | MN864145 |
| VC_286_7 | OM632671 |
| VC_286_8 | KU255730 |
| VC_286_9 | OM386662 |
| VC_287_0 | MT457552,MT457553 |
| VC_287_1 | KY971610 |
| VC_288_0 | MW358927,MW385300,OK499989 |
| VC_288_1 | KX078569 |
| VC_288_2 | MG696114,MK295204 |
| VC_288_3 | KP890823,MF001355,MW367898 |
| VC_288_4 | MW057858 |
| VC_29_0 | AF157835,EU794049,LR794146,LR794147,LR794148,LR794149,LR794150,LR794151,LR794152,LR797923 |
| VC_290_0 | MT129652,MT446396,MT446411,MT446412,MT446415,MT446421,MT611523,MW233709,MW250787,MW598459 |
| VC_290_1 | GQ981382,KP007360,MT446420,MT446422,MT533174,MW269952,OK254198,OL451225 |
| VC_290_2 | HM563683,KP007361 |
| VC_290_3 | KP007362,MT446392 |
| VC_290_4 | KP007359,OM386655 |
| VC_290_5 | HM071924,HQ829472,KT176190,KU878968,KU878969,MH051917,MK234886,MT150133,MT682711,MW149274,MW679410,MZ561532,OK335775,OL780484 |
| VC_290_6 | EF469154,EU863409,MH051913,MH791409,MK310184,MN850609,MN850621,MT129654,OL770107,OP292222,OV876907,OV877765 |
| VC_290_7 | MK295203 |
| VC_292_0 | MK448704,MK448707,MK448709,MK448710 |
| VC_293_0 | MK448750 |
| VC_293_1 | MK448995 |
| VC_293_2 | MK448740,MK448917 |
| VC_293_3 | MK448675,MK448676,MK448688,MK448690,MK448694,MK448772,MK448778,MK448782,MK448850,MK448859,MK448877,MK448931,MK448955,MK448962,MK448966,MK448973,MK448975,MK449004,OL774870 |
| VC_294_0 | MK448832,MK449008 |
| VC_295_0 | MK819239,ON857931 |
| VC_296_0 | MG592403,MG592404,MG592405,MG592551,MN592897,MW824406 |
| VC_297_0 | KM389210,KU199710,MW748993,MZ089728,ON778007 |
| VC_297_1 | HQ711984,JX403939,KU310943,MK511001,MK511002,MK511003,MK511004,MK511005,MK511006,MK511007,MK511008,MK511009,MK511010,MK511011,MK511012,MK511013,MK511014,MK511015,MK511016,MZ773939 |
| VC_298_0 | KU708004 |
| VC_298_1 | MW862109 |
| VC_299_0 | KX581090,KX581091,KX581092,KX581093,KX581094,KX581095,KX581096,KX581097,KX581098,KX581099,KX581100,KY658673,KY658674,KY658675,KY658676,KY658677,KY658678,KY658679,KY658680,MK672799,MK672800,MK672801,MK672803,MK672804 |
| VC_3_0 | CP000623,CP000624,CP008753,DQ087285,GU911304,HM461982,JX681814,LC528882,LC667452,MW072790,MZ375754,OM304382,OM638608,OM638609,OM638610,OM638611,ON135435 |
| VC_30_0 | JQ182733,JQ182734 |
| VC_30_1 | AF335538,AF547987,CP000711,JF974339,JN600960,KJ832078,MH717096,MN340231,MT813197,MT833387,MZ043897,OM022251,ON470589,ON470594,ON470605,OV696608 |
| VC_300_0 | HQ141411,MG788324,OP650937 |
| VC_301_0 | LT985589,LT993247 |
| VC_302_0 | MH837542,MH837543 |
| VC_303_0 | AF176027,AF176028,AF176029,AF176030,AF176031,AF176032,AF176033,AF176034,AF274751,AF299300,AF299301,AF299302,AF299303,AF299304,AF299305,AF299306,AF299307,AF299308,AF299309,AF299310,AF299311,AF299312,AF299313,AF299314,CP004084,DQ079880,DQ079881,DQ079882,DQ079883,DQ079884,DQ079885,DQ079886,DQ079887,DQ079888,DQ079889,DQ079890,DQ079891,DQ079892,DQ079893,DQ079894,DQ079895,EF380009,EF380010,EF380011,EF380012,EF380013,EF380014,EF380015,EF380016,EF380017,EF380018,EF380019,EF380020,EF380021,EF380022,EF380023,EF380024,EF380025,EF380026,EF380027,EF380028,EF380029,EF380030,EF380031,EF380032,FJ849058,GQ153912,GQ153913,GQ153914,GQ153915,GU385905,HM753658,HM753659,HM753660,HM753661,HM753662,HM753663,HM753664,HM753665,HM753666,HM753667,HM753668,HM753669,HM753670,HM753671,HM753672,HM753673,HM753674,HM753675,HM753676,HM753677,HM753678,HM753679,HM753680,HM753681,HM753682,HM753683,HM753684,HM753685,HM753686,HM753687,HM753688,HM753689,HM753690,HM753691,HM753692,HM753693,HM753694,HM753695,HM753696,HM753697,HM753698,HM753699,HM753700,HM753701,HM753702,HM753703,HM753704,HM753705,HM753706,HM753707,HM753708,HM753709,HM753710,HM753711,HM753712,HM753713,HM753714,HM753715,HM753716,HM753717,HM753718,HM775306,HM775307,HM775308,HM775309,HM775310,HM775311,JF719726,JF719727,JF719728,JQ764986,JQ764987,JQ764988,JQ764989,JQ764990,KJ997912,KY653237,MH378441,MH378442,MH378443,MN385565,MT947439,PX1CG,S13CG |
| VC_304_0 | AF189021,FJ867910,FJ867911,FJ867912,FJ867913,FJ867914,JQ809650,MF431615,MF431616,MF431617,MK613346 |
| VC_305_0 | GU557055,MK613343,MK613344,MK613345,MW514246,MW514247,MZ892987,MZ892988,MZ892989,MZ892990,MZ892991,MZ892992 |
| VC_306_0 | AF191797,MW557853 |
| VC_307_0 | AJ131519,MW476489 |
| VC_308_0 | AF195902,DQ320509,HE956704,KF767351 |
| VC_309_0 | MF041990,MN830256,X98106 |
| VC_31_0 | KY271395,KY271396,MK448231,MK448233,MK448235,MK714353,OK490410,OK490415,OK490416 |
| VC_31_1 | OK499971 |
| VC_31_2 | HQ201308 |
| VC_310_0 | AF222060,AY190604,KC292020,KC292022,MN901520,MN901521,MW450817,MZ334492,MZ334493,MZ334494,MZ334495,MZ334496,MZ334497,MZ334498,MZ334499,MZ334500,MZ334501,MZ334502,MZ334503,MZ334504,MZ334505,MZ334506,MZ334507,MZ334508,MZ334509,MZ334510,MZ334511,MZ334512,MZ334513,MZ334514,MZ334515,MZ334516,OK649958 |
| VC_311_0 | KC117376,KC292021,MZ334517,MZ334518,MZ334519,MZ334520 |
| VC_312_0 | KC292026,MZ334525 |
| VC_313_0 | KC292029,MZ334524 |
| VC_313_1 | KC117377,KC292027,MZ334523 |
| VC_314_0 | DQ836063,JX625144 |
| VC_315_0 | AF227250,EF108464,EF108465,EF204939,EF204940,FJ483837,FJ483838,FJ483839,JQ966307,LT898437,MK213795,MT830620 |
| VC_316_0 | MZ326866,MZ611865,ON711490 |
| VC_317_0 | MK448685,MK448763,MK448767,MK448771,MK448773,MK448774,MK448777,MK448786,MK448845,MK448853,MK448855,MK448942,MK448943,MK448963,MK448964,MK448969,MK448972,MK449012 |
| VC_317_1 | MK448706,MK448721 |
| VC_317_2 | AY007505,FR671408,FR671409,KC348600,KC348601,KC348602,KT337360,KX077894,KY065485,KY065503,KY065504,MK448814,MK448881,MK448882,MK448896,MK448909,MK448988,MK448989,OL774869,OL774877 |
| VC_317_3 | KP208803 |
| VC_317_4 | MK448900 |
| VC_318_0 | KY065466,KY065468,MK448982 |
| VC_32_0 | MH443100 |
| VC_323_0 | MK448821,MK448822 |
| VC_323_1 | KY065498,MK448735,MK448757,MK448758,MK448759,MK448893,MK448915,MK448939,OL774872 |
| VC_325_0 | MH513971,ON970586 |
| VC_325_1 | JN398369,MN617845 |
| VC_325_10 | MH744414,MK359331,MN119379 |
| VC_325_11 | MH001458 |
| VC_325_12 | MN813700 |
| VC_325_13 | EU744249,EU744252 |
| VC_325_14 | GU247132,KT818595,MT889384 |
| VC_325_15 | KJ174156 |
| VC_325_16 | MK310141,MK524523 |
| VC_325_17 | KC701493 |
| VC_325_18 | OL549190 |
| VC_325_19 | JN020140,MH338238,MK967387 |
| VC_325_2 | JF937099 |
| VC_325_20 | MN585970 |
| VC_325_21 | EU744248,KX522649,KX522943,KX574454,MK524497 |
| VC_325_22 | MK524499 |
| VC_325_23 | JF937103,KC661273,KT259047,MG920060,MG925351,MH020244,MH450113,MH450130,MH576959,MH576971,MH576974,MK359299,MK820638,MT639654,MW507123,MW712732,ON970627 |
| VC_325_24 | JF957057,MH230876,MN585976 |
| VC_325_25 | MZ648034 |
| VC_325_26 | MG944220 |
| VC_325_27 | MH536816,MH632118 |
| VC_325_28 | KX369584 |
| VC_325_29 | KP027196,KP027203,MW862983 |
| VC_325_3 | KF493880 |
| VC_325_30 | KJ409696,KT326768,MK494116,OK040791 |
| VC_325_31 | MK814754 |
| VC_325_32 | MW435858 |
| VC_325_33 | KF560331,MK112532,MK359316,OM203162 |
| VC_325_34 | ON677304 |
| VC_325_35 | EU826470,JN572689,JQ698665,JQ809703,KF024724,KF279415,KF279416,KM597531,KR080201,MF668283,MK359312,MK524506,MK524531,MN428054,MN586000,MT657342,MZ322004 |
| VC_325_36 | MH371110,MN369760 |
| VC_325_37 | MK061415,MN585985,MT776812 |
| VC_325_38 | KX683876,KX702320,MG099951,MH000606,MH651180,MT952847 |
| VC_325_39 | AF271693,MZ355726 |
| VC_325_4 | KX369586 |
| VC_325_40 | KP027205,MH697581 |
| VC_325_41 | JF937100,MK112530 |
| VC_325_42 | JN699016,JX307705,KT363731,MN119377 |
| VC_325_43 | KX712238 |
| VC_325_44 | EU744251,JN660814,MH669011 |
| VC_325_45 | MK524525 |
| VC_325_46 | MH576975,MK967401 |
| VC_325_47 | JN699000,MG962370 |
| VC_325_48 | KY213952,MK112540 |
| VC_325_49 | AP018477,AP018478,KF416340,MH632117 |
| VC_325_5 | AY500152,JF937108,JF937110,KJ194585,KT626047,MG872833,MG925344,MG962372,MK359354,MN585999,MT310897 |
| VC_325_50 | MG812486,MG812489,MG812495,MG872836 |
| VC_325_51 | KJ690250,MN585991 |
| VC_325_52 | JN153085,MH230878,MH697575,MN062710,MN234212 |
| VC_325_53 | KT388014,KX550443,MH020236,MH399782,MH590595,MK359300,MK359304 |
| VC_325_54 | KY224001 |
| VC_325_55 | MK305893,MZ355722 |
| VC_325_56 | MF919508,MN585997 |
| VC_325_57 | ON208829 |
| VC_325_58 | ON970624 |
| VC_325_59 | MT771340 |
| VC_325_6 | MN444869 |
| VC_325_60 | JN687951,MH338239 |
| VC_325_61 | JN153086,JN698998,MG925340,MG925342,MH450133,MH479919,MK310142,MT639644 |
| VC_325_62 | OL455898,OM105884,OP021677 |
| VC_325_7 | KR997929,KR997934,MG944221 |
| VC_325_8 | KF024726,KP027204 |
| VC_325_9 | AY500153,KT438500 |
| VC_326_0 | MW202895 |
| VC_327_0 | AF306496,MH992158,MH992160,MH992177,MH992178,MH992189,MH992191,MH992194,MH992197,MH992207,MH992218,MW202535 |
| VC_328_0 | EU710883,FQ482083,HQ728263,MG589384,MH426725,MK759884,MK770412,MN604407,OK483342,OM522317,ON706965 |
| VC_329_0 | MG646672,MH586731 |
| VC_329_1 | AF320576,JF461087,JX181822,KP143762,KY626162,MF370225,MG251391,MG251392,MG969405,MG969406,MK965969,MK965970,MW006474,MW006475,MW006482,MZ327261,MZ327262 |
| VC_329_10 | MN994498 |
| VC_329_11 | KP010413,MG646669,MG646670,MG646671,MN227145,MN481367,MN655999,MW311372,MW321605,MW362867,MZ150758,OL474141,OL800606,OL825705,ON239128 |
| VC_329_12 | MG065664,OK272472 |
| VC_329_13 | KM657822 |
| VC_329_14 | MG969404 |

| | |
|---|---|
| VC_329_15 | GU169904 |
| VC_329_16 | KF055347,KX405003,MT409176,MW423797 |
| VC_329_17 | KM092515,MT322327,OK040806,OK040807 |
| VC_329_18 | JX560968,MK524176,MN882542,MT833282,MT887289,MW296865,ON540300 |
| VC_329_19 | ON868915 |
| VC_329_2 | MN850631,MN850637 |
| VC_329_20 | LC739530,LC739531,LC739532,ON210138 |
| VC_329_21 | MH051914 |
| VC_329_22 | MZ311863,OM386661,OM386663,OM386664,OM386665 |
| VC_329_3 | KX377933,LR880803,MK962758,MN850564,MN850566,MN850577,MN850580,MN850585,MN850603,MN850605,MN850610,MN850611,MN850614,MN850619,MN850633,MN850635,MN850636,MN850639,MN850644,MN850647,MZ501086,OK272490,OM630585,OX016465 |
| VC_329_4 | JN225449,KR014248 |
| VC_329_5 | MN994499 |
| VC_329_6 | LR597658,MZ726791,MZ726792 |
| VC_329_7 | EU877232,KP869100,KP869106,KP869109,KP869110,KP869113,KT184313,KT184314,KT184315,KT184316,LC519489,LR699048,MH571750,MK482688,MK482689,MK482690,MT446388,MW481326 |
| VC_329_8 | LC473039,MN994503 |
| VC_329_9 | LR597663 |
| VC_33_0 | HQ201307,JQ182729,MH178096,MN688132 |
| VC_330_0 | KM236239,KT001916,KT363872,KT454805,KT454806,KY654690,MF158040,MF158041,MF158043,MF158044,MF327003,MF327004,MF327005,MH791400,MH791413,MH920362,MK562502,MK562504,MK562505,MN342151,MN850587,OK539913,OM953433,ON528729,ON528730,ON528731,ON528732 |
| VC_331_0 | MK290737,MK290738,MN270891,MN270892 |
| VC_332_0 | MW749004,MW749010,OK283618 |
| VC_333_0 | FN391954,KT337342,KY065453,KY065454,KY065458,KY065459,KY065460,KY065461,KY065462,KY065463,KY065469,KY065470,KY065471,KY065494,KY065495,KY065499,MK448732,MK448739,MK448901,MK448912,MK448914,MK448918,MK448920,MT900487,OL774868 |
| VC_334_0 | MK449000 |
| VC_335_0 | MK448804,MK448805,MK448807,MK448808,MK448815,MK448991 |
| VC_337_0 | MT980836,MT980837,MT980838,MT980839,MT980840,MT980841 |
| VC_339_0 | AF323671,LT985572,LT985581,LT985584,LT985586,LT985587,LT985593,LT993227,LT996080 |
| VC_34_0 | AY374448,CP038007,CP072507,KY271402 |
| VC_340_0 | AF338467,AP013538,AP013539,AY939843,EF372997,GU071093,GU071100,GU071102,GU071104,GU071107,GU943054,HM559717,HQ316584,HQ317388,HQ317389,HQ332137,HQ332140,HQ337022,HQ633062,HQ634152,HQ634188,KC310802,KC310803,KC310804,KC310805,KC310806,MH540083,MK016662,MT424636,MW015080,MZ803112 |
| VC_341_0 | AF396866,MW202489,OK085710 |
| VC_342_0 | MN988526,MN988556 |
| VC_344_0 | AF399011,AP019418,JX233784,KU521356,LC727695,LC727696,LC727699,LC727701,MF805716,MN563784,MN871480,MN871489,MT133560,MT995926,MT995927,MT995928,MW057919,MZ444140,ON631220,OP310983,OP310984 |
| VC_344_1 | MK599315,MN103543,OU343167 |
| VC_344_2 | MF063068 |
| VC_344_3 | MK552327 |
| VC_344_4 | EU197055 |
| VC_344_5 | MF042360 |
| VC_344_6 | HQ630627 |
| VC_345_0 | AP017924,AP017925 |
| VC_346_0 | MK308674,MK308675,MK308676,MK308677 |
| VC_346_1 | MT811961,OM319461 |
| VC_346_2 | OV032902 |
| VC_346_3 | MH595538 |
| VC_347_0 | LR881104,MN871444,MN871445 |
| VC_348_0 | AJ344259,AJ414696,AJ748296,AJ875026,FM164764,KP282675,KX712143,KY744228,KY744229,KY744230,KY744231,KY744232,KY744233,KY744234,KY744235,MK064565,MN876841,MN876842,MN876843,OM401712,OM401713,OM401714 |
| VC_349_0 | AF440571,AM087120,AM087121,AM087122,AM087123,EU545650,KX467643 |
| VC_35_0 | KY271399,MK416008,MK416013,MK416022,MK448234,MK448237 |
| VC_35_1 | OL617042 |
| VC_350_0 | AJ854042,MK064564 |
| VC_351_0 | AM419438,HQ332141,MW344765 |
| VC_352_0 | AF440695,MK002701,MK450543 |
| VC_353_0 | AF447491,AY453853,CP000625,OK095358,OM892879,OP293079 |
| VC_354_0 | MN512538,MN812722,MT227924,OP125534 |
| VC_355_0 | MT121967,MW651859 |
| VC_356_0 | HQ634187,MK105819 |
| VC_357_0 | MT740734,ON170024,ON170027,ON170031,ON170052,ON170057,ON170058,ON170068,ON170071,ON170075,ON170077,ON170079,ON170129,ON170133,ON170134,ON170136,ON170150,ON170172,ON170175,ON170179,ON170181,ON170182,ON170185,ON170187,ON170208,ON170214,ON170228,ON170229,ON170232,ON170234,ON170236,ON170238,ON170240,ON170242,ON170254,ON170257,ON170262,ON170264,ON170265,ON170267,ON170275,ON170279,ON170286,ON170287,ON170303,ON170317,ON170323,ON170324,ON170326,ON170327,ON170343,ON170355,ON170359 |
| VC_358_0 | ON170149,ON170171,ON170322 |
| VC_359_0 | JN585957,MF612073,MH426726,MK618715,MT251347,MT251348,MW722080 |
| VC_360_0 | AF454431,AY751298,DQ079896,DQ079897,DQ079899,DQ079900,DQ079901,DQ079902,DQ079903,DQ079904,DQ079906,DQ079908,DQ079909,GQ153920,GQ153922,GQ153923,JF719729,JF719730,JF719731,KX266300,KX266301,KX266302,KX266303,KX266304,KX266305,KX266306,KX266307,KX266308,KX266309,KX266310,KX266311,KX266312,KX266313,KX266314,KX266315,KX266316,KX266317,KX266318,KX266319,KX266320,KX266321,KX266322,KX266323,KX266324,KX266325,KX266326,KX266327,KX266328,KX266329,KX266330,KX266331,KX266332,KX266333,KX266334,KX266335,KX266336,KX266337,KX266338,KX266339,KX266340,KX266341,KX266342,KX266343,KX266344,KX266345,KX266346,KX266347,KX266348,KX266349,KX266350,KX266351,KX266352,KX266353,KX266354,KX266355,KX266356,KX266357,KX266358,KX266359,KX266360,KX266361,KX266362,KX266363,KX266364,KX266365,KX266366,KX266367,KX266368,KX266369,KX266370,KX266371,KX266372,KX266373,KX266374,KX266375,KX266376,KX266377,KX266378,KX266379,KX266380,KX266381,KX266382,KX266383,KX266384,KX266385,KX266386,KX266387,KX266388,KX266389,KX266390,KX266391,KX266392,KX266393,KX266394,KX266395,KX266396,KX266397,KX266398,KX266399,KX266400,KX266401,KX266402,KX266403,KX266404,KX266405,KX266406,KX266407,KX266408,KX266409,KX266410,KX266411,KX266412,KX266413,KX266414,KX266415,KX266416,KX266417,KX266418,KX266419,KX266420,KX266421,KX266422,KX266423,KX266424,KX266425,KX266426,KX266427,KX266428,KX266429,KX266430,KX266431,KX266432,KX266433,KX266434,KX266435,KX266436,KX266437,KX266438,KX266439,KX266440,KX266441,KX266442,KX266443,KX266444,KX266445,KX266446,KX266447,KX266448,KX266449,KX266450,KX266451,KX266452,KX266453,KX266454,KX266455,KX266456,KX266457,KX266458,KX266459,KX266460,KX266461,KX266462,KX266463,KX266464,KX266465,KX266466,KX266467,KX266468,KX266469,KX266470,KX266471,KX266472,KX266473,KX266474,KX266475,KX266476,KX266477,KX266478,KX266479,KX266480,KX266481,KX266482,KX266483,KX266484,KX266485,KX266486,KX266487,KX266488,KX266489,KX266490,KX266491,KX266492,KX266493,KX266494,KX266495,KX266496,KX266497,KX266498,KX266499,KX266500,KX266501,KX266502,KX266503,KX266504,KX266505,KX266506,KX266507,KX266508,KX266509,KX266510,KX266511,KX266512,KX266513,KX266514,KX266515,KX266516,KX266517,KX266518,KX266519,KX266520,KX266521,KX266522,KX266523,KX266524,KX266525,KX266526,KX266527,KX266528,KX266529,KX266530,KX266531,KX266532,KX266533,KX266534,KX266535,KX266536,KX266537,KX266538,KX266539,KX266540,KX266541,KX266542,KX266543,KX266544,KX266545,KX266546,KX266547,KX266548,KX266549,KX266550,KX266551,KX266552,KX266553,KX266554,KX266555,KX266556,KX266557,KX266558,KX266559,KX266560,KX266561,KX266562,KX266563,KX266564,KX266565,KX266566,KX266567,KX266568,KX266569,KX266570,KX266571,KX266572,KX266573,KX266574,KX266575,KX266576,KX266577,KX266578,KX266579,KX266580,KX266581,KX266582,KX266583,KX266584,KX266585,KX266586,KX266587,KX266588,KX266589,KX266590,KX266591,KX266592,KX266593,KX266594,KX266595,KX266596,KX266597,KX266598,KX266599,KX266600,KX266601,KX266602,KX266603,KX266604,KX266605,KX266606,KX266607,KX266608,KX266609,KX266610,KX266611,KX266612,KX266613,KX266614,KX266615,KX266616,KX266617,KX266618,KX266619,KX266620,KX266621,KX266622,KX266623,KX266624,KX266625,KX266626,KX266627,KX266628,KX266629,KX266630,KX266631,KX266632,KX266633,KX266634,KX266635,KX266636,KX266637,MG563770 |
| VC_362_0 | GQ153921,KC237308 |
| VC_363_0 | OK338687,OM055665 |
| VC_364_0 | AP008986,KY210139,MG944227,MG944228,MG944229,MG944230,MG944231,MG944232,MG944233,MG944234,MG944235,MG944236,MN461279,OK275494,ON996340,OP067662 |
| VC_369_0 | JQ177063,KY290953,KY290954,MH179474,MK804891,MK804892 |
| VC_37_0 | OM046621,OM046622,OM046623,OM046625,OM046629 |
| VC_37_1 | KU052037,MN013086,OL396571 |
| VC_371_0 | MT897903,ON456342 |
| VC_371_1 | KJ595576,KM347890,KR997967,KY006474,MH926060,MK279910,MK524512,MK524526,MN585967,MN586058,MW534377 |
| VC_371_10 | MH590594 |
| VC_371_11 | KX592589,MF155947,MF668276,MF919528,MF919539,MG757159,MH230874,MH513980,MH590588,MK112539,MK112546,MK112551,MK279843,MK279865,MK279870,MK279873,MK279878,MK279894,MK279895,MK279897,MN369744,MT310868,MT316460,MT316463,MW534381,MW712720,MW862980,MW862993,MZ648038,ON637769,OP434459 |
| VC_371_12 | MK112544,MT310867,MT952849,ON456332 |
| VC_371_13 | JN192463,KM363597,KR029086,KR029087,KX576645,KX576646,KX576647,KX702319,KY385380,KY385382,KY385383,MF919511,MG757164,MG757165,MG944223,MG962363,MH051251,MH155874,MH316568,MH371107,MH479916,MH479918,MH513973,MH576958,MK279850,MK279858,MK279864,MK279883,MK279890,MK279891,MN585965,MN945903,ON456330 |
| VC_371_14 | MN369738 |
| VC_371_15 | GQ303264,GQ303266,JF704103,JF704109,JF937097,JF957056,JN006063,JN638752,JN638753,JN699008,JN699010,KM408320,KP027208,KT599441,KU867907,KX578071,KX670813,KX683292,KX683293,KY676783,MG770212,MG925346,MG925350,MG944225,MH399775,MH399786,MH576970,MH651184,MH825702,MH825705,MK112527,MK112536,MK112555,MK279855,MK279859,MK279861,MK279866,MK279879,MK279881,MK279882,MK279885,MK279887,MK279888,MW712726 |
| VC_371_16 | AF547430,DQ398046,FJ174694,GU247133,GU247134,JF704102,JN699009,JX649096,JX649097,JX649100,KJ194580,KJ194583,KX369585,KX620786,MG757158,MG925348,MG962365,MH371114,MH371117,MH371125,MH399773,MH399780,MH450116,MH450122,MH479914,MH479921,MH651175,MH651186,MH727555,MH744417,MH779516,MK112542,MK112553,MK279846,MK279852,MK279871,MK494104,MN945899,MT897909,MW534379,MW534380,MZ622173 |
| VC_371_17 | JF937095,JN698990,JN699017 |
| VC_371_18 | JF704091,JF937109,JN698989,KJ194579,KP027197,KP027209,KR816508,KX576643,KY965066,MT897901 |
| VC_371_19 | MZ622172 |
| VC_371_2 | KC576784,KJ567044,MG925356,MG962375,MH077582,MK112552,MK279889,MN096366 |
| VC_371_20 | MN703415 |
| VC_371_21 | JN006064,KR781349 |
| VC_371_3 | GQ303259,JF704099,JX649098,JX649099,KC661274,KC691258,KF713485,KJ174157,KJ538723,KM236502,MF919503,MF919507,MF919523,MF919530,MG925339,MG962362,MH230875,MH779500,MK279877,MK279908,MN444868,MT897902 |
| VC_371_4 | KT364588,MH576954,MH576967,MH651177,MK279904 |
| VC_371_5 | MH051264,MK310139 |
| VC_371_6 | MF919519 |
| VC_371_7 | ON970594 |
| VC_371_8 | MZ648041 |
| VC_371_9 | MT776813,MT889369,MZ681520,ON970577 |
| VC_372_0 | DQ398044,EU770221,JF704094,JF704104,JN699011,KF279411,KF279412,KF279413,KF493881,KR080194,KR080200,KT270441,KX589269,MH513970,MK524486,MN234171,MN369759,MN428050,MT310870,MT639648,MT818419,MT897910,MW435857,ON970575 |
| VC_372_1 | JN699007,KF024727,KF416341,KR080199,KY224000,MH727561,MK524490,MN284893,MT889381 |
| VC_372_2 | MH744423,MK059749,MN617843,MW055906,OL455893 |
| VC_372_3 | KF024729,KJ433973,KJ433974,KJ433975,KJ433976 |
| VC_372_4 | MN813693,MZ150786,OP297541 |
| VC_373_0 | AJ006589,AJ550940,JX182372,MK433266,MK450421,MK450426,MK450431,MK450433,MK686068,MK686069 |
| VC_374_0 | GQ379227,MK937595 |
| VC_375_0 | MG593800,MG593803,MH001457 |
| VC_376_0 | MT701593,MT701595,MT701596 |
| VC_377_0 | KR905068,KU848187,LC625742 |
| VC_378_0 | AJ242593,DQ003637,KJ094022,KP399678,MH341452,MZ361825,MZ361826,MZ361827,ON456151 |
| VC_379_0 | AJ312240,DQ003639,JX120799,KJ094023,MT500540 |
| VC_38_0 | AB231700,KF356199,OP394178 |
| VC_380_0 | DQ003642,KP399677,MH341451 |
| VC_381_0 | GU323708,HQ141410 |
| VC_385_0 | AJ251805,AJ318471,EU547803,JX421753,KC960671,KM253764,KP313531,KP791805,KP791806,KP791807,KT008108,KU687347,KU687348,KX130726,KX452696,KX778611,LC730324,MF285616,MF285617,MF285620,MH717709,MK608335,MN013074,MN149904,MT419366,MT813142,MW296877,MW600722,MZ359670,MZ428229,OK149215,OL362281,OL362282,OL362283,OL691920,OL744209,OL744212,OL744217,OL800605,ON528720,ON528721,ON528722,ON528723,ON528724,ON950090,OP617747,OX335419,OX335420,OX335431,OX335434,OX335437 |
| VC_386_0 | AM183667,EU734170,EU734171,FJ194439,GQ468526,HQ333270,JQ965702,KF700371,KM366096,KP090453,KP090454,KY084243,LT960606,LT961836,LT961837,LT961838,LT961840,LT961841,LT961842,LT961843,LT961844,LT961845,LT961846,LT962379,LT962380,LT962475,LT962906,LT962907,MG471392,MH031343,MH809534,MK685668,MK907285,MN080711,MN252582,MT374859,MT515757,MT542512,MT625440,MW147603,MW367418,MW767996,MZ501055,MZ501064,OK663600,OL800603,OL856139,ON754979,ON922920 |
| VC_387_0 | HQ728265,MN450150,ON715521 |
| VC_388_0 | KT321314,ON602733,OX335378,OX335441 |
| VC_389_0 | MF979561,MW001769,ON995367 |
| VC_39_0 | AF125520,AP000363,AP000422,AP004402,AP005153,AP005154,AP012529,AP012531,AP012532,AP012535,EU311208,HM208303,KJ909655,KP682372,KP682373,KP682375,KP682376,KP682377,KP682378,KP682379,KP682380,KP682382,KP682383,KP682384,KP682385,KP682386,KP682387,KP682388,KP682389,KP682390,KP682391,KP682392,KU238068,KU238069,KU238070 |
| VC_390_0 | EU652770,KX078568,MW358931,OK499982 |
| VC_392_0 | AY247822,AY264774,AY264775,AY264776,AY264777,AY264778,EU734174,FR667955,GU071091,JQ957925,JQ965700,JQ965701,JQ965703,JX000007,JX872508,KM242061,KU927499,LC730325,LR745706,LR745707,LR745708,LR745709,LR745710,MG488277,MG832642,MG833025,MG966531,MH382198,MH717098,MH717099,MH779619,MK310182,MK542821,MN026740,MN098329,MN481365,MN481366,MT350293,MT543041,MT681669,MW074885,MW883060,MZ220766,MZ318361,MZ318362,MZ318363,MZ375233,MZ375234,MZ375235,MZ375236,MZ375237,MZ375238,MZ375239,MZ375240,MZ375241,MZ375242,MZ375243,MZ375244,MZ375245,MZ375246,MZ375247,MZ375248,MZ375249,MZ375250,MZ375251,MZ375252,MZ375253,MZ375254,MZ375255,MZ375256,MZ375257,MZ375258,MZ375259,MZ375260,MZ375261,MZ375262,MZ375263,MZ375264,MZ375265,MZ375266,MZ375267,MZ375268,MZ375269,MZ375270,MZ375271,MZ375272,MZ375273,MZ375274,MZ375275,MZ375276,MZ375277,MZ375278,MZ375279,MZ375280,MZ375281,MZ375282,MZ375283,MZ375284,MZ375285,MZ375286,MZ375287,MZ375288,MZ375289,MZ375290,MZ375291,MZ375292,MZ375293,MZ375294,MZ501078,MZ501081,MZ833439,OK042080,OL964739,OL964740,OL964741,OL964748,OL964749,OL964750,OM630586,ON528715,ON528716,ON528717,ON528719,ON568193,OP331230,V01146 |
| VC_393_0 | MH059639,MH460459,MN013082,MN013085,MN812693,MT822288,ON287378 |
| VC_394_0 | MH370477,MT465573 |
| VC_395_0 | MN184885,MT822284,MT822285,MT822286,MT822287 |
| VC_396_0 | MW057854,MW358928 |
| VC_398_0 | KP087939,KP087945,KT264811,MG945164,MG945682,MG945837,MH572269,MH572314,MH572315,MH572316,MH572317,MH572318,MH572319,MH572320,MH572321,MH572325,MH572326,MH572328,MH572329,MH572336,MH572351,MH572394,MH572395,MH572396,MH572397,MH572404,MH572419,MH572431,MH572439,MH572441,MH572443,MH572445,MH572446,MH572451,MH572473,MH572487,MH572492,MH572493,MH572494,MH572503,MH572510,MH572511,MH992195,MH992211,MK249143,MK249151,MK249223,MK249231 |
| VC_4_0 | DQ426904,DQ426905,JN255163,KP137432,KP137434,KP137436,KP137438 |
| VC_40_0 | LC567830,LC567838,LC567842,LC616079,ON416862 |
| VC_400_0 | MH572340 |
| VC_401_0 | MH572479 |
| VC_402_0 | MH572512 |
| VC_403_0 | KC131023,KC131025,MH572294,MH572295,MH572296,MH572384,MH572385,MH572386,MH572387,MH572388,MH572389,MH572390,MH572391,MH572430,MH572470,MH572471,MH572472,MH572505,MH572506,MH572507 |
| VC_404_0 | MH572279,MH572300,MH572301,MH572302,MH572303,MH572305,MH572306,MH572307,MH572308,MH572309,MH572310,MH572311,MH572312,MH572313,MH572345,MH572392,MH572393,MH572406,MH572435,MH572436,MH572437,MH572469,MH572480,MH572508,MH572509 |
| VC_406_0 | KP790011,KU998237,KU998238,KY471268,KY471269,MH651189,MH697580,MN010758,MT553344,MW132713,ON260826 |
| VC_407_0 | AJ278322,MG324353,MG324354 |
| VC_408_0 | KY566218,KY613597,KY624610,KY624611,KY624612,KY624613,KY624614,KY624615,KY624616 |
| VC_409_0 | KY203335,KY250482,MW582634 |
| VC_41_0 | AJ556162,JQ347801,LC727891,LC727892,LC727893,LC727894,LC727895,LC727896,LC727897,LC727898,LC727899,LC727900,LC727901,LC727902,ON470597,ON470625 |
| VC_41_1 | AP012530,AP012538,AP012539,AP012540,FJ184280,FJ188381,FM180578,LC645408,LC645425,LC645426 |

| | |
|---|---|
| VC_410_0 | MG198783,MG198784,MH271305,MH271318,MT521997,MZ681507,ON456352 |
| VC_411_0 | LC644972,LC644974,MT843275 |
| VC_412_0 | AJ302074,DQ113772,HF563658,JX879087,KC413988,KT337358,KT337364,KT388093,KX550065,KY065455,KY065479,KY065487,KY065490,KY065491,KY065492,KY065493,KY065501,KY065502,KY349816,MK448671,MK448708,MK448714,MK448831,MK448834,MK448861,MK448874,MK448875,MK448886,MK448887,MK448897,MK448898,MK448905,MK448925,MK448927,MK449006,MK449009,MT900488,MW495853,OL774873 |
| VC_413_0 | KY065465,MK448733,MK448904 |
| VC_414_0 | FR671405,FR671406,FR671407,FR671410,FR671411,HG799490,HG799496,HG799497,KJ417497,KT337339,KT337340,KT337341,KT337352,KT337354,KT337355,KT337356,KT337359,KT337362,KY065450,KY065451,KY065452,KY065456,KY065472,KY065473,KY065474,KY065475,KY065476,KY065477,KY065478,KY065480,KY065486,KY065488,KY065496,KY065497,KY065500,KY065505,OK173051 |
| VC_415_0 | KY065449 |
| VC_416_0 | MK448686,MK448762,MK448788,MK448791,MK448793,MK448862,MK448949,MK448951,MK448954,MK448958,MK448959,MK448960,MK448965,MK448977 |
| VC_419_0 | MK448680,MK448684,MK448698,MK448769,MK448895 |
| VC_42_0 | AP012534,HE664024,HG792102,HG792103,HG792104,HG792105,HG803182,HG803183,HG803184,HG803185,HQ424691,JQ011318,KF971864,KP682371,KP682374,KR781488,KU298437,KY914478,LC645409,LC645410,LC645411,LC645412,LC645413,LC645414,LC645415,LC645416,LC645417,LC645418,LC645419,LC645420,LC645421,LC645422,LC645423,LC645424,LC645427,LC645438,LC645442,LC645443,LC645444,LC645445,LC645446,LC645447,LC645448,LC645449,LC645450,LC645451,LC727903,MG710528,MN067333 |
| VC_422_0 | MF034659,MF166859,MT902335,MZ995503,MZ995506 |
| VC_423_0 | AJ505558,AM265638,AM910651,EU056923,EU236438,FN263372,JQ307386,JX501340,JX997978,KC969441,KM067278,KR054031,KU743887,KX711710,KY618819,LC727700,LN610574,LN610580,MH107770,MH979674,MN553585,MN553587,MN553588,MN553589,MN602045,MN615698,MN615699,MN692672,MN871455,MN871483,MN901924,MW117144,MW406975,MW406976,MZ553931,MZ687409,OK539824,OL754589,OL802210,ON857926,ON857928,ON857933,ON857935,OP292288,OP556577 |
| VC_424_0 | AJ536073,AY257527,AY701338,CP013282,EU408779,MZ089978,MZ089979 |
| VC_426_0 | AJ560763,GQ866233 |
| VC_426_1 | CP013974,MG575421,MZ424863 |
| VC_428_0 | KM366099,KX826077,KY608966,MF740800,MH181876,MK568062,MK673511,MK770411,MK770413,MN379740,MT012730,MT074434,MT074435,MT074436,MT074437,MT074438,MT074439,MT074441,MT074442,MT074443,MT074444,MT074445,MT074446,MT663719,MT764207,MT768059,MW413353,MW508890,MW602648,MZ127825,OM313460,OM313461,OP132241 |
| VC_43_0 | EU078592,JQ086376,JQ182735,KT232076,LAMCG,LC730321,LR595850,LR595859,LR595860,LR595861,LR595863,LR595864,LR595865,LR595866,LR597635,LR597636,LR597639,LR597643,LR597644,LR597648,LR597650,LR597651,LR597652,LR597653,MF374379,MH547045,OL657226,OL657227,OL657228,OM418625,OM418626,OM418627,OM475427,OM475428,OM475430,OM475432,OM475435,OV696612 |
| VC_431_0 | MK448692,MK448702,MK448711,MK448716,MK448724,MK448730,MK448744,MK448745,MK448923 |
| VC_432_0 | MN695334,MN695335 |
| VC_432_1 | MH412654 |
| VC_432_10 | GU071098,HQ317391 |
| VC_432_11 | HQ633071 |
| VC_432_12 | MW015081 |
| VC_432_13 | MK799832 |
| VC_432_14 | MN018232 |
| VC_432_15 | KU686204,KU686205,KU686206 |
| VC_432_16 | MH629685 |
| VC_432_2 | GU071095 |
| VC_432_3 | MF351863 |
| VC_432_4 | MT408532 |
| VC_432_5 | MK867354 |
| VC_432_6 | MH319749 |
| VC_432_7 | MH319754 |
| VC_432_8 | KM359505 |
| VC_432_9 | AY939844,GU071092,HQ632825 |
| VC_433_0 | HQ634177,KU686192,KU686193,KU686194,KU686195,KU686196 |
| VC_433_1 | KJ019054,KJ019069,KJ019082,KJ019089,KJ019094,KJ019156,KU686200,KU686201,KU686202 |
| VC_433_10 | KU686211 |
| VC_433_2 | KF156338 |
| VC_433_3 | KU686197,KU686198,KU686199 |
| VC_433_4 | GU071105,KU594607,KX349291,KX349292,KX349293,KX349294,KX349295,KX349296,KX349297 |
| VC_433_5 | AJ630128,LN828717 |
| VC_433_6 | KU594606 |
| VC_433_7 | MW147367 |
| VC_433_8 | FM207411 |
| VC_433_9 | MW117966 |
| VC_435_0 | MT162466,MT162467 |
| VC_435_1 | MK016664 |
| VC_436_0 | MZ577095,MZ577096,MZ577097,MZ577098 |
| VC_437_0 | KF381361,KR052480,KR052481,KR052482 |
| VC_438_0 | MH319720,MH319733,MH319742,MH319753,MH319758,MH327487 |
| VC_439_0 | MH319750,MH319759 |
| VC_440_0 | AJ635161,AY722806,MN876845,MT047590 |
| VC_442_0 | MK838116,MN102098,MN131137,MN159079 |
| VC_442_1 | MN642089,ON287370,ON287375 |
| VC_442_2 | JN627160,MK689364,MZ348422,MZ501267 |
| VC_442_3 | MN032614,MN453779,OL964756 |
| VC_442_4 | MK774614,OM810292 |
| VC_443_0 | MH791414,MW272540 |
| VC_443_1 | GU459069,KY290955 |
| VC_443_2 | JX123262,MF448340,MF498773,MF498775,MH791398,MH791404,MN871441,MN871442,MT713136,OM339718 |
| VC_444_0 | AJ783769,JQ287645 |
| VC_445_0 | MT732443,MT732444 |
| VC_445_1 | KC821607,KC821618 |
| VC_446_0 | HG322870,KP238129,KP282677,KP282678 |
| VC_447_0 | AJ888457,KX607101,KX607102 |
| VC_448_0 | KU878088,LC680884,MF360957,MW749003 |
| VC_448_1 | KT895374 |
| VC_449_0 | LC680885,MN091626,MW218148,MW248466,MW349128,MW349129,MZ779063 |
| VC_45_0 | AJ298298,AY055382,KC736978,KC814930,KU927493,MK733259,MK733260,MK733261,MZ501270,ON470584,ON470603,SFU82619 |
| VC_450_0 | AM076770,MN617773,MT366931,MT366932,MT366933,MT366934,MT366935,MT366936,MT366937,MT366938,MT366939,MT366940,MT366941,MT366942,MT366943,MT366944 |
| VC_451_0 | AM084414,DQ111067,EU734172,HG813241,HG818823,HG818824,KC109329,KJ748011,KT990215,KU687349,KU687350,KU687351,KX669227,KX689784,KY295894,KY962008,LT594300,MF630921,MG845865,MH400309,MH464253,MH491167,MK033136,MK301532,MK610268,MK903277,MK903279,MK903281,MN067430,MN218775,MN414250,MN510331,MN518893,MN518894,MN715150,MN812686,MT350292,MT446393,MT446394,MT496969,MT682707,MT808983,MT862763,MW258709,MW345254,MW416012,MW748991,MW876471,MZ223858,MZ234009,MZ234011,MZ234021,MZ234022,MZ234023,MZ234024,MZ234044,MZ234045,MZ234046,MZ234047,MZ311861,MZ311862,MZ398247,MZ648215,MZ726794,MZ923531,OK210074,OL362275,OL362276,OL539729,OL773676,OL773677,OM256482,OM355495,OM457002,OM867676,OM908436,OM937123,ON528733,ON548433,ON637250 |
| VC_452_0 | AM084415,AY288927,AY370673,AY370674,GU595417,JN986844,JQ837901,KM366097,KM370384,KM819695,KM819696,KP890822,KU946962,KX279892,KY295891,KY295892,KY295893,KY295897,KY295899,KY435490,KY744566,LC730323,MG004687,MG251390,MG775042,MH729818,MK373771,MK373773,MK380014,MN384978,MT496971,MT680615,MW394389,MZ031013,MZ234012,MZ234019,MZ234020,MZ234032,MZ234049,MZ234050,MZ398244,MZ573924,MZ747518,MZ997839,MZ997840,MZ997841,OK625527,OL362272,OL362273,OL744215,OL744218,ON148527,ON185580,ON210144,ON210145,ON461912,ON526839,ON602763,ON602764,ON602765,OP018999,OP038547,OP079918,OP413828,OP413830,OX335382 |
| VC_453_0 | EF160123,FQ482084,FQ482086,MG736918,OK570184,OM486945 |
| VC_455_0 | DQ121662,JF770475,JQ182736,KC333879,KC579452,KF771236,KF771237,KF771238,KF771239,KJ668713,KT825490,MF158039,MF158042,MW286156,OL539447,OL539451,OL539453,OL539467 |
| VC_456_0 | AM156909,JN986845,KX130960,KY398841,MG050172,MG065688,MK372342,MK373793,MK907226,MK907227,MK907228,MK907229,MK907230,MK907231,MK907232,MK907233,MK907234,MK907235,MK907240,MK907241,MK907242,MK907243,MK907244,MK907245,MK907246,MK907247,MK907248,MK907249,MK907250,MK907251,MK907252,MK907253,MK907254,MK907255,MK907256,MK907257,MK907258,MK907259,MK907260,MK907261,MK907262,MK907263,MK907264,MK907265,MK907266,MK907272,MK907273,MK907274,MK907275,MK907276,MN164484,MN840485,MT511058,MT682706,MW057918,MZ501051,MZ501080,MZ501087,OM258170,ON081052,OP087522,OP251154 |
| VC_458_0 | MH059632,MH807810,MH807813,MH807815,MH807817 |
| VC_459_0 | KC292023,MG550112,MZ334526 |
| VC_46_0 | KF766125,KJ603229 |
| VC_460_0 | JF974294,JF974312,MK327140,MT682064,MW119258,MW722082,MZ318367,MZ440881,MZ836210,OL362277,OL617041,ON602730,ON602737,ON602738,ON602744,ON602759,ON602760,OP373729,OP481875 |
| VC_461_0 | AM491472,KX017520,KX523699,KY860935,MF158038,MH023293,MH321493,MH424446,MH717097,MH791402,MH791410,MH791412,MK112901,MK599416,MK759854,MK947460,MN692673,MN737551,MN871490,MN871503,MT768060,MW427215,MW477799,MZ375232,MZ375295,MZ375296,MZ375297,MZ375298,MZ375299,MZ375300,MZ375301,MZ375302,MZ375303,MZ375304,MZ375305,MZ375306,MZ375307,MZ375308,MZ375309,MZ375310,MZ375311,MZ375312,MZ375313,MZ375314,MZ375315,MZ375316,MZ375317,MZ375318,MZ375319,MZ375320,MZ375321,MZ375322,MZ375323,MZ375324,MZ375325,MZ375326,MZ375327,MZ375328,MZ375329,MZ375330,MZ375331,MZ375332,MZ375333,MZ375334,MZ375335,MZ375336,MZ375337,MZ375338,MZ375339,MZ375340,MZ375341,MZ375342,MZ375343,MZ375344,MZ375345,MZ375346,MZ375347,MZ375348,MZ375349,MZ375350,MZ375351,MZ375352,MZ375353,MZ375354,MZ375355,MZ489634,MZ726796,OL362274,OL964734,OL964742,OL964743,OL964744,OL964751,OL964752,ON239132,ON637251 |
| VC_463_0 | AM749121,DQ386162,KR153145,MT430910 |
| VC_464_0 | JQ067087,KU356689,LN610577,LN610584,LN610585,MT108725,MT108731,MZ926748 |
| VC_465_0 | JQ067084,KX129925,KX898399,KX898400,MN504636,MW835180,ON911540 |
| VC_466_0 | MW091529,MW202777,MW462191 |
| VC_467_0 | MN862068,ON529856,ON529859,ON529862,ON529864 |
| VC_468_0 | MN988465,MN988541,MN988554,ON014755 |
| VC_469_0 | MT536174,OL960029 |
| VC_47_0 | LC616031,LC616032,LC616033,LC616034,LC616035,LC616036,LC616037,LC616038,LC616039,LC616040,LC616041,LC616042,LC616043,LC616044,LC616045,LC616046,LC616047,LC616048,LC616049,LC616050,LC616051,LC616052,LC616053,LC616054,LC616055,LC616056,LC616057,LC616058,LC616059,LC616060,LC616061,LC616062,LC616063,LC616064,LC616065,LC616066,LC616067,LC616068,LC616069,LC616070,LC616071,LC616072,LC616073,LC616074,LC616075,LC616076,LC616077,LC616078 |
| VC_470_0 | MN698239,MN698241,MW574966 |
| VC_471_0 | MW677525,OP009285 |
| VC_472_0 | AM910650,AY078382,HE983844,HF543949,HG518155,JN254801,JX863101,KC294142,KF856712,KP233880,KR054029,LC648443,LN610578,MF768469,MG432151,MG589385,MG589386,MG845684,MH973725,MK837011,MK837012,MN508619,MN553586,MN562749,MN871476,MN871484,MT108723,MT108724,MT118288,MT119362,MT119371,MT119373,MW329986,MW512831,MW526260,MZ089732,MZ089733,MZ089738,MZ089739,MZ089740,MZ089742,OL441337,ON156559,ON376263,ON857927,ON857936,OP342787 |
| VC_473_0 | CP067352 |
| VC_473_1 | CP069348 |
| VC_473_2 | CP011970,LN681537 |
| VC_473_3 | MF547662,MF547663 |
| VC_473_4 | CP067347 |
| VC_474_0 | HM029250,KF926093,KM677185,KY554768,KY554769,KY554770,KY554771,KY554772,KY554773,KY554774,KY554775,KY554776,KY554777,MN552145 |
| VC_475_0 | OM236516,OM634653,ON133270 |
| VC_475_1 | OK499972,OK499987,OK499994 |
| VC_475_10 | OK499992,ON548418 |
| VC_475_2 | KC481682,MT254578,ON548420 |
| VC_475_3 | ON881243 |
| VC_475_4 | MH884508 |
| VC_475_5 | ON548417,ON548419 |
| VC_475_6 | KT070867 |
| VC_475_7 | MW084976 |
| VC_475_8 | MK288021,OK499991 |
| VC_475_9 | ON548421 |
| VC_476_0 | AY150271,DQ126339,JN391180,JQ011317,JQ691610,KC911856,KC911857,KP869107,KP869108,KX231829,MK416010,MK422448,MN187550,MT261384,MT580116,MT580117,MW205203,MW722083,OK490417 |
| VC_477_0 | AP011616,AP011617,DQ997624,MK257744 |
| VC_478_0 | MG592415,OX241562 |
| VC_478_1 | MW073017 |
| VC_479_0 | OM416763,OM416764,OM416765,OM416766,OM416767,OM416768,OM416769,OM416770,OM416771,OM416772,OM416773,OM416774,OM416775,OM416776,OM416777,OM416778,OM416779 |
| VC_48_0 | LC645428,LC645429,LC645430,LC645431,LC645432,LC645433,LC645434,LC645435,LC645436,LC645437 |
| VC_480_0 | AP013541,AP013542,AP013545,KC465901,MH579717,MH598798,MH598800,MH598802,MH598803,MH598804,MH598805,MH598807,MT375520,MT375521,MT375526,MT375527,MT375528,MT375530,MT375531 |
| VC_481_0 | AP013544,JX483873,KM199771,KT997847,MT708545,MW023914,MW980070,MZ666938 |
| VC_482_0 | AP013546,AP013547,AP013548,MW822601 |
| VC_484_0 | KJ183191,KJ183192,KJ183193 |
| VC_485_0 | MN698242,MN698243,MN698247 |
| VC_486_0 | KC465898,MT375525,MT647605,MW273920,MW273922,MW273923,MW273924,MW273925,MW273926,OP131293,OP131294,OP131295,OP131296,OP131297,OP131299,OP131300,OP131301,OP131302 |
| VC_487_0 | MT375519,MT375529 |
| VC_488_0 | MN871446,MN871447,MN871448,MN871449,MN871451,MN871452,MN871453,MW394391 |
| VC_488_1 | MF285618,MK994515,MW021758,OK040170,OL539468,ON287372,ON287376,ON375847 |
| VC_488_2 | KF623294,MN334766 |
| VC_489_0 | MN445183,MW845760,MW879340 |
| VC_489_1 | KF806589,KU886222,KU886223,KU886224,KU886225,MG655267,MG655268,MG655269,MG655270,MK514281,MZ501268 |
| VC_489_2 | MT661596 |
| VC_49_0 | LC727840,LC727841,LC727842,LC727843,LC727844,LC727845,LC727846,LC727847,LC727848,LC727849,LC727850,LC727851,LC727852,LC727853,LC727854,LC727855,LC727856,LC727857,LC727858,LC727859,LC727860,LC727861,LC727862,LC727863,LC727864,LC727865,LC727866,LC727867,LC727868,LC727869,LC727870,LC727871,LC727872,LC727873,LC727874,LC727875 |
| VC_490_0 | AP014714,AP014715 |
| VC_490_1 | MN087708,MN882610,MZ348423,OL989991,ON630910 |
| VC_490_2 | KF835987 |
| VC_490_3 | HE956709,KY593455 |
| VC_490_4 | KP202158 |
| VC_490_5 | MH059636 |

| | |
|---|---|
| VC_490_6 | MT920315 |
| VC_491_0 | CP062992,KR269720,KT239446,KX258185,KY000080,LR877331,MG751100,MH333064,MH729874,MK421971,MN101226,MN101229,MN101230,MN434095,MT580897,MT701588,MW021752,MW239157,MZ560702,MZ602148,MZ890186,OK499980,OK499981,OK499996,OK499997,OK631811,OL539458,OL870318,OL870319,OM337876,OM337877,OM630219,ON550260,OP331213,OP617748,OU509536 |
| VC_491_1 | MN101225 |
| VC_492_0 | HQ698922,MK240351 |
| VC_492_1 | CAKLQC020000001,CAKLQD020000001,MH165274,MH460829,MH713599,MN648195,MN662249,MN709128,MN723850,MN732883,MN782535,MT385367,MT409116,MT741943,OL763420,OL770258,OL770259,OL770261,OL770262,OP585103 |
| VC_492_2 | MK278860,MN864865,MT949699,MW388001,MW388002,MW388003,MW388004,OL770260 |
| VC_492_3 | GU911519 |
| VC_492_4 | HM004124 |
| VC_492_5 | HM032710 |
| VC_492_6 | HM114315 |
| VC_493_0 | KT932418 |
| VC_493_1 | MG592562 |
| VC_493_10 | HQ634156 |
| VC_493_11 | KX507046 |
| VC_493_12 | MW824371,MW824372 |
| VC_493_13 | MW824376 |
| VC_493_14 | HQ316579 |
| VC_493_15 | HQ634195 |
| VC_493_16 | MG592553 |
| VC_493_2 | MG592529 |
| VC_493_3 | OX241437 |
| VC_493_4 | OX241455 |
| VC_493_5 | AP014858 |
| VC_493_6 | MG592495 |
| VC_493_7 | HQ634157 |
| VC_493_8 | MG592441 |
| VC_493_9 | ON922990 |
| VC_495_0 | KY883638,KY883639,KY883640,MH310933,MH310935,MN419153,MW794150,MW794151,MW794152,MW794153,MW794154,MW794155,MW794156,MW794157,MW794158,MW794159,MW794160,MW794161,MW794162,MW794163,MW794164,MW794165,MW794166,MW794167,MW794168,MW794169,MW794170,MW794171,MW794172,MW794173,MW794176,MW794177,MW794179,MW794180,MW794182,MW794183,MW794188,MW794190,MZ292654 |
| VC_495_1 | HQ641347,HQ641348,HQ641349,HQ641350,HQ641351,HQ641352,HQ641353,HQ641354,KY065147,KY883634,KY883635,KY883636,KY883637,MH310934,MH310936,MN402506,MW794140,MW794141,MW794142,MW794143,MW794144,MW794145,MW794146,MW794147,MW794148,MW794149,MW794174,MW794175,MW794178,MW794181,MW794184,MW794185,MW794186,MW794187,MW794189,MW794191,MW794192,MW911612,MW911613,MW911614,MW911615 |
| VC_496_0 | KF302034 |
| VC_496_1 | MF370964 |
| VC_496_2 | KR534323 |
| VC_496_3 | KX822733 |
| VC_497_0 | MG592459 |
| VC_497_1 | MG592473 |
| VC_498_0 | AP014887,AY328852,AY328853,KF854250,KX196154,KY622015,MG592395,OL658619 |
| VC_499_0 | KT160311,MG592392,MG592401,MG592414,MG592451,MG592461,MG592462,MG592467,MG592468,MG592472,MG592499,MG592526,MG592531,MG592541,MG592542,MG592547,MG592561,MG592572,MG592590,MG592592,MG592611,MW824380,MW824381,MW824382 |
| VC_50_0 | AF083977,GQ357916,KP010268,LR595867,MT374852,MT374853,MT374854,MT374856,MT591577 |
| VC_500_0 | OM287554,OM732336,OM732337 |
| VC_501_0 | AP014888,MG592509,MG592546,MG592620,MG592623,OX241426,OX241428,OX241430,OX241431,OX241432,OX241441,OX241448,OX241449,OX241494,OX241567,OX241568 |
| VC_505_0 | MW353175,MW353176,MW353177 |
| VC_506_0 | KR560069,MK580972,MN095772,MN098328,MZ042490,MZ326868,MZ398248,OL702939 |
| VC_506_1 | ON758385 |
| VC_507_0 | AP018462,AP018463,KY130461,MH001446,MH001450,MK494114,MK494131,MK494132 |
| VC_508_0 | AY129334,DQ398048,JN618996,JN698991,JN699004,KF024722,KM101117,KR997932,KT365402,KT880194,KX443696,MG757162,MG812490,MG812491,MH001452,MH051249,MH590591,MH697584,MH727546,MK061411,MK620898,MK814751,MN234225,MN428049,MN586045,MN586046,MN813678,MT639652,MW712731,MZ456036,ON970587,ON970593 |
| VC_509_0 | DQ398049,EU816589,FJ641182,JF704095,JN698992,JN699003,JN699006,JN699018,KF493879,KJ194581,KJ194584,KP027207,KR080203,KR080205,MG839014,MG920059,MG925338,MG925353,MH051265,MH316565,MH513976,MH576977,MK061414,MK359357,MN096359,MN096360,MN234223,MN444872,MN444873,MT310871,MT310892,MT952851,MW924644,MZ171375,MZ171381,OK318960,OP434441 |
| VC_51_0 | AB285204,JQ182728,LR595868,LR597641,MF807953,MW057859,ON470578,ON470598,ON470618,OV696610 |
| VC_510_0 | MH153810,MK801725,MN234196,MN369758,MN908693,MT657335,OK216879,OM105886,ON081333,ON108644,ON260813,OP434461 |
| VC_511_0 | JN698995,OP172873,OP172874 |
| VC_513_0 | AP018485,JF704111,JF957058,KC172839,KJ510415,KM592966,KT326767,KX640831,KX670828,KX683423,KY464936,LC730317,MF373840,MG812492,MH020248,MH047632,MH047633,MH051259,MH316563,MH976517,MK284519,MK494127,MK524496,MN062706,MN062712,MN586063,MT310878,MT310879,MT310880,MT310881,MW712718,ON970623 |
| VC_514_0 | AY129332,HM755814,JF704098,JF704101,JF704114,KC661272,KC661281,KF279418,KM101119,KM233454,KM233455,KP017310,KP017311,KP027202,KT359365,KT381277,KU255188,KU578077,KU984914,KU985091,KX458237,KX507362,KX523125,KX664448,KX670788,KX808129,KY965063,MF185729,MF185732,MF185733,MG757161,MG925341,MG925355,MH020238,MH077583,MH155864,MH338240,MH371121,MH399776,MH450119,MH450124,MH450126,MH450128,MH513975,MH536813,MH536825,MH576949,MH651179,MH727554,MK112543,MK279903,MK279913,MK284518,MK310140,MK359341,MK433273,MK494096,MK494128,MK875800,MK967399,MN062718,MN096368,MN284900,MN497953,MN585969,MN585984,MN586048,MN813699,MT657331,MT897907,MW507128,MZ681518,MZ747514,OK318959,OK999978 |
| VC_515_0 | JQ512844,JX307704,JX411619,KC661279,KJ156985,KM101120,MF773750,MH001459,MK494124,MN586038,MN807247,MT522002,MZ322017,MZ355723,MZ622159,ON337196 |
| VC_516_0 | MW314854,MW584159,MW584197 |
| VC_517_0 | JF704093,JN243857,KX550442,KX579975,KX641260,KY083059,MF668278,MK279898,MK359310,MK359322,MK494115,MK494117,MK494129,MK875799,MN284905,MN813697,MZ322023,ON970614 |
| VC_517_1 | GQ303263,HM152766,JF792674,JN243856,JN561150,JN699015,JQ809701,JQ896627,JX307702,JX307703,KC661271,KF024733,KF841475,KF841476,KF954506,KF959563,KJ510414,KP027198,KP057620,KT365397,KT716493,KU055616,KU867906,KU985092,KU985094,KX458236,KX550441,KX557234,KX619651,KX648373,KX808130,KX817174,KX817175,KY006473,KY083058,KY083060,KY083061,KY083062,KY083063,KY204244,KY204245,KY204246,KY204247,KY204248,KY204249,KY204250,KY204251,KY471266,KY549155,MF919496,MF919509,MG099942,MG944217,MG944224,MG962373,MH020249,MH051247,MH077581,MH155875,MH155879,MH230877,MH399777,MH399785,MH479909,MH479911,MH509443,MH536817,MH651170,MH697588,MH727563,MH825710,MK279902,MK310145,MK359301,MK359323,MK359325,MK359340,MK359356,MK494088,MK494093,MK494094,MK494097,MK494101,MK494125,MK494126,MK878895,MK878905,MN728684,MN813696,MT114158,MT490373,MT498056,MT553338,MT553346,MT723939,MT818417,MT889382,MT889386,MT889390,MW291019,MZ005673,MZ171380,MZ322025,MZ355725,MZ747509,ON637771,ON970565,OP297529 |
| VC_518_0 | AP018487,FJ168659,HM152765,JN699001,MN234195,MN234236 |
| VC_518_1 | AY129333,KP027201 |
| VC_519_0 | JN412591,JN572061,KC691256,KF841477,KP027200,KR080195,KT347313,KU985090,KX641262,MF133446,MF140399,MF281061,MF373843,MF472894,MF919492,MG198785,MK016499,MK494122,MK524508,MK814758,MK820639,MK919480,MK937604,MN010757,MN096356,MN585977,MN807248,MN807249,MN807250,MN892484,MN892486,MN908683,MT024866,MT498061,MW055902,ON637760,ON970574,OP172867,OP434454 |
| VC_52_0 | AB259123,AB434711,AB937974,AB981170,KX179905,MF716957,ON170020,ON170023,ON170029,ON170030,ON170034,ON170037,ON170041,ON170050,ON170055,ON170087,ON170089,ON170091,ON170093,ON170095,ON170097,ON170101,ON170103,ON170105,ON170107,ON170109,ON170111,ON170113,ON170115,ON170117,ON170119,ON170121,ON170125,ON170127,ON170132,ON170143,ON170145,ON170147,ON170153,ON170155,ON170157,ON170159,ON170161,ON170163,ON170165,ON170168,ON170193,ON170195,ON170197,ON170199,ON170201,ON170203,ON170205,ON170207,ON170212,ON170216,ON170218,ON170220,ON170223,ON170226,ON170245,ON170247,ON170249,ON170251,ON170253,ON170273,ON170277,ON170281,ON170283,ON170285,ON170290,ON170292,ON170294,ON170296,ON170299,ON170301,ON170308,ON170312,ON170314,ON170316,ON170331,ON170333,ON170335,ON170337,ON170339,ON170341,ON170353 |
| VC_520_0 | KT309034 |
| VC_520_1 | MT889374 |
| VC_520_10 | MN369752 |
| VC_520_11 | MN585995 |
| VC_520_12 | MN735431 |
| VC_520_13 | KT281793 |
| VC_520_14 | MT684595 |
| VC_520_15 | MF668270,MT684593 |
| VC_520_16 | KX808131 |
| VC_520_17 | MN369739 |
| VC_520_18 | MN586001 |
| VC_520_19 | KM652553,MN369749 |
| VC_520_2 | KR824843 |
| VC_520_20 | JN699012 |
| VC_520_21 | MH669001 |
| VC_520_22 | OP068340 |
| VC_520_23 | JN859129,KF024723 |
| VC_520_24 | EF536069 |
| VC_520_25 | MH590598 |
| VC_520_26 | HQ728524,JN698996,JX121091 |
| VC_520_27 | KT281792,KY471267 |
| VC_520_28 | JN020141,MT114167 |
| VC_520_29 | KM597530 |
| VC_520_3 | MF668287 |
| VC_520_30 | MH825707,ON526976 |
| VC_520_31 | FJ174690,GU060500,JF937093,KT281795,MH020235,MH077580,MK016494,MN585973,MW924649,MZ681519,OP297536 |
| VC_520_32 | MH651178 |
| VC_520_33 | MH371123,MN369755 |
| VC_520_34 | MH077578 |
| VC_520_35 | KR997930,KR997931,KR997968 |
| VC_520_36 | MH744415 |
| VC_520_37 | JF937102 |
| VC_520_38 | KJ025956,MH155865,MH155871,MH590602,MK524517,MT310893 |
| VC_520_39 | MH371120 |
| VC_520_4 | MH155866 |
| VC_520_40 | KU865302,KY965064,MH371115,MT553345 |
| VC_520_41 | MH651183,MN586011 |
| VC_520_42 | MH399771 |
| VC_520_43 | JN020143,ON456356,ON970562 |
| VC_520_44 | KY348865 |
| VC_520_45 | KT895281,MG872841,MH316564,MH399784,MH651169,MH834617,MK494110 |
| VC_520_46 | MZ274306 |
| VC_520_47 | MT684586 |
| VC_520_48 | MZ171378 |
| VC_520_49 | KM101122 |
| VC_520_5 | MH727548 |
| VC_520_50 | MT897904 |
| VC_520_51 | MH051257 |
| VC_520_52 | KC736071 |
| VC_520_53 | KY385384 |
| VC_520_54 | OM105885 |
| VC_520_55 | OP297542 |
| VC_520_56 | KM279937 |
| VC_520_57 | MK359343 |
| VC_520_58 | DQ398050,EU816590,ON970595 |
| VC_520_59 | JN542517,MH001448,MN585987,MN586005,MZ681512 |
| VC_520_6 | MN586004,MT684597,MT684601 |
| VC_520_60 | DQ398045,KT279576 |
| VC_520_61 | MW055915 |
| VC_520_62 | ON970566 |
| VC_520_63 | MF919501 |
| VC_520_64 | MG962361 |
| VC_520_65 | MZ747519 |
| VC_520_66 | MN586012 |
| VC_520_67 | ON970610 |
| VC_520_68 | KM347889 |
| VC_520_69 | MH669014 |
| VC_520_7 | MN310548 |
| VC_520_70 | KC661280 |
| VC_520_71 | FJ174693 |
| VC_520_72 | MK494118 |
| VC_520_73 | KR935214,MH669012 |
| VC_520_74 | JF704117,MK112556 |
| VC_520_75 | MN310544 |
| VC_520_76 | JN398368,MT114164 |
| VC_520_77 | JQ300538,KX576644,MK016496 |
| VC_520_78 | MH590596,OP297528 |
| VC_520_79 | KC661278 |

| | |
|---|---|
| VC_520_8 | MZ958744 |
| VC_520_80 | MH651188,MH669003,MK359315 |
| VC_520_81 | MF919526 |
| VC_520_82 | MN310545 |
| VC_520_83 | KX610764 |
| VC_520_84 | ON970585 |
| VC_520_85 | KT895280,MN369751 |
| VC_520_86 | MZ747512 |
| VC_520_87 | MK524503 |
| VC_520_88 | JN020142 |
| VC_520_89 | MN585975 |
| VC_520_9 | KF114874 |
| VC_520_90 | MK016504 |
| VC_520_91 | MZ209299 |
| VC_520_92 | KM402757,MK359307,MN428051,MT889396,MT889397 |
| VC_520_93 | JF937098,MH632119 |
| VC_520_94 | MK967397 |
| VC_520_95 | MK224592 |
| VC_520_96 | AY129330 |
| VC_520_97 | MG925354 |
| VC_520_98 | MG812488 |
| VC_521_0 | JN256079,JN624851,KC576783,KF986246,KM588359,KU935726,KU935727,KU935728,KU935729,KU935730,KU935731,KX756439,MG099936,MG099948,MH316570,MH399787,MH697576,MH697585,MH697593,MH926055,MK392367,MK524500,MK524509,MK524515,MK524518,MK524520,MK524522,MK<br>524528,MK977708,MT498042,MT498044,MT498048,MT684588,MT723932,MZ622162,OL742561,OP297540 |
| VC_522_0 | KU963256,MN175604,ON645344 |
| VC_522_1 | MN428053,MN585966,MT521995,OP297548,OP434457 |
| VC_523_0 | MK977709 |
| VC_524_0 | MW584176,MW584199 |
| VC_524_1 | MW584188 |
| VC_525_0 | KU963255,KU963257,KX557273,KX557287,MH479912,MH479927,MK016492,MK112547,MK801727,MK814755,MK820642,MK875793,MK875796,MK878898,MK919483,MT723938,MT889398,MT952844,MW712717,MW924648,MW924650,MZ171374,MZ274304,OP297546,OP434440 |
| VC_525_1 | KU963249,KU963263,MK919469,MK967384,MT639650 |
| VC_526_0 | MG557979,MG765277,MG765279,MH809529,MH809530,MH809531 |
| VC_526_1 | MF787246 |
| VC_526_2 | AY682195 |
| VC_526_3 | MK504444 |
| VC_527_0 | MW202473,MW202478,MW202738 |
| VC_529_0 | MW824391,MW824401 |
| VC_53_0 | HM064452,ON170025,ON170069,ON170139,ON170320 |
| VC_530_0 | AP019417,KJ936628,MF431726,MG592626,MK483107,MN062185,MW217515,MW451248,MW748992 |
| VC_531_0 | KC131129,KP671755,KT919972,KT919973,MK568540,MT612988,MT612989,ON000910,ON190025 |
| VC_531_1 | AY283928,JN849462,MN794232,MT135026,OK905446,ON190024 |
| VC_531_2 | HQ317393 |
| VC_531_3 | MW824377 |
| VC_531_4 | MG592456 |
| VC_531_5 | MT135025 |
| VC_531_6 | MT135024 |
| VC_532_0 | AP019524,AP019525 |
| VC_532_1 | LC168164 |
| VC_533_0 | LR756501,LR756503 |
| VC_533_1 | LR745206 |
| VC_533_2 | LR756502 |
| VC_533_3 | LR756504 |
| VC_534_0 | AP019527,FJ429185,KY554762,KY554763,KY554764,KY554765,KY554766,KY554767,MN528768 |
| VC_536_0 | EF153632,MT708549,MW460246 |
| VC_539_0 | AY048850,BK013337,BK013338 |
| VC_54_0 | AB259124,AB828698,AB931172,JQ408219,LC066596,MT495635 |
| VC_542_0 | MW478292,OK247671 |
| VC_543_0 | MG592399,MG592418,MG592419,MG592510,MG592511,MG592579 |
| VC_544_0 | MG592447,MG592453 |
| VC_546_0 | JN201525,MH399772 |
| VC_546_1 | MN586029,MW314846 |
| VC_546_10 | AY129338 |
| VC_546_11 | JF937101 |
| VC_546_2 | JN698997,KM400683,MF919534 |
| VC_546_3 | KM925136,MF072690,MF133445,MF668284 |
| VC_546_4 | KF114875,MH077579,MH669017,MH697582,MH697583,MH727551,MK524504,MK524521,MK524527,MK524529,MK967379,MK967402,MN062701,MN183286,MZ622169,MZ958748 |
| VC_546_5 | MF919504 |
| VC_546_6 | KF006818,MF919512 |
| VC_546_7 | JF937090,JF957059,KM101123,MK279849 |
| VC_546_8 | MW132714 |
| VC_546_9 | MK524516 |
| VC_55_0 | CP006891,CP023680,CP023686 |
| VC_550_0 | MW314855,MW314856,MW314858,MW584152,MW584158,MW584161,MW584164,MW584171,MW584179,MW584180,MW584196,MW584204 |
| VC_551_0 | JF937091,JF937106,MG872831,MG872832,MG872843,MH399774,MH399778,MH536820,MK016502,MK359320,MK801726,MK937593,MN096361,MN586013,MT723943,MW291023 |
| VC_551_1 | MF919541,MH576956,MN586043,MN586044 |
| VC_551_2 | EU816591,JF937096,JN006061,JN006062,KF188414,KR080204,KT222941,KT222942,KU865303,KX817173,KY319168,MH513972,MH513978,MN586032,MN586051,MT114165,MZ274305 |
| VC_551_3 | KF306380 |
| VC_551_4 | JN412590,KF416343 |
| VC_551_5 | MK016491,MK016498 |
| VC_551_6 | EU816588,KX611831,KX834009,MG757160,MG872837,MH371112,MH513983,MH536829,MH651174,MH669002,MH727557,MH779503,MK559429,MN428059,MN586034,MN586037,MT952854,MZ622157,MZ681505,OL742559,ON970597,ON970600 |
| VC_551_7 | AY129331,DQ398041,GQ303265,JN382248,JN391441,KC661277,KC691255,KC748971,KF279417,KT020852,KT373978,KY549152,MF668277,MH000607,MH536827,MH576953,MK359309,MK620893,MK757445,MN586017,MN586019,MN586035,MN586041,MN586050,MZ322024,MZ622158,MZ622163,ON45<br>6345 |
| VC_551_8 | KC748969,KF024732,KF493883,KF562099,KF562100,KF562101,KF562102,MF919506,MF919524,MF919525,MF919529,MF919535,MF919540,MG099953,MH020247,MH590587,MK433262,MN096364,MT522005 |
| VC_554_0 | JN698994,MPU46938 |
| VC_555_0 | MW314852,MW584148,MW584150,MW584154,MW584156,MW584162,MW584163,MW584181,MW584182,MW584192,MW584203,MW584208,MW584212 |
| VC_556_0 | AY129335,JN698993,KF017002,KF024730,KJ829260,KT281796,MG099943,MG872838,MH020246,MH590592,MH926062,MN428052,MN428058,MN585964,MT818418,MT818425,MW314845,MW712737,MZ209306,MZ747511,ON260829 |
| VC_557_0 | MK937597,MN096370,OP434458 |
| VC_557_1 | MG099939,MH651176,MH651185,MH779504,MK279906,MK376961,MK814752,MZ388553,MZ747510,ON456334,ON526981 |
| VC_557_2 | MH536812 |
| VC_557_3 | MW534375 |
| VC_558_0 | JN204348,JN412592 |
| VC_558_1 | GQ303260,MT553335 |
| VC_558_10 | MK112534 |
| VC_558_11 | KT281790,MT889395 |
| VC_558_12 | KC748970,MH825697,MK359348 |
| VC_558_13 | MK359321 |
| VC_558_14 | MG872839,MH316561,MK359303,MK359311,MK359358,MK450427,ON526982,ON637768 |
| VC_558_15 | MK359342 |
| VC_558_16 | ON755189 |
| VC_558_17 | MK359328 |
| VC_558_18 | MN183285 |
| VC_558_19 | ON637764 |
| VC_558_2 | EU826471,MH727543 |
| VC_558_20 | JF704096,KR080196,MF919493,MH697586,MH697590,MK359326,MK359335,MK359338,MK359339,MK359347,MK359362,MK450432,MN428066,MZ028626,MZ681511,ON081335,ON260824 |
| VC_558_21 | MT723945 |
| VC_558_22 | JF704092,MH590593,MK450423,MZ958745 |
| VC_558_23 | KX369583 |
| VC_558_24 | OP021681 |
| VC_558_25 | EU826468 |
| VC_558_26 | EU826469 |
| VC_558_27 | MK359361,MW712729 |
| VC_558_28 | MK359333 |
| VC_558_29 | MT818416 |
| VC_558_3 | MK359355 |
| VC_558_30 | EU826467,JN699625,MK359344 |
| VC_558_31 | DQ398053 |
| VC_558_32 | MK310143 |
| VC_558_33 | MH910040 |
| VC_558_34 | MK112549 |
| VC_558_35 | MT818423 |
| VC_558_36 | MK359336,MK359363,MK359366,MN234193 |
| VC_558_37 | KT365400 |
| VC_558_38 | MK878897,MN428065 |
| VC_558_39 | MK359364,MT723944,OP297534 |
| VC_558_4 | MK359353 |
| VC_558_40 | MN276192 |
| VC_558_41 | MK450424 |
| VC_558_42 | MZ856343 |
| VC_558_43 | MK359337 |
| VC_558_44 | MK359345 |
| VC_558_45 | MK359327 |
| VC_558_46 | MK359349 |
| VC_558_47 | MK112541,MK112545,MK112548 |
| VC_558_48 | KF024734,KX721256 |
| VC_558_49 | JN624850,KC748968,KX721255,MH779498,MK112554,MK359334,OP068341 |
| VC_558_5 | KC691257 |
| VC_558_50 | JQ911768,KJ595575,KM881426,KT321476,MF919499,MH051266,MH316567,MH669008,MH825713,MK359314,MK359365,MK450429,MK450434,OK040789,OP021684 |
| VC_558_51 | MK112533 |
| VC_558_52 | MH744422 |
| VC_558_53 | MK359306 |
| VC_558_54 | MH779502 |
| VC_558_55 | MF919494 |
| VC_558_56 | MH727558 |
| VC_558_57 | MK450425 |
| VC_558_58 | MZ388551 |
| VC_558_59 | MK359317 |
| VC_558_6 | JF704116,KT365398 |
| VC_558_60 | MN183280 |
| VC_558_61 | MH051260,MK359318 |
| VC_558_62 | MK359305 |
| VC_558_63 | JN699626 |
| VC_558_64 | MK359350 |
| VC_558_65 | MK450422,MW291012 |

| | |
|---|---|
| VC_558_66 | MK359330 |
| VC_558_67 | OP021682 |
| VC_558_68 | MG812494 |
| VC_558_69 | MK359329 |
| VC_558_7 | MH051261 |
| VC_558_70 | ON456344 |
| VC_558_71 | MK359346 |
| VC_558_72 | MG812493 |
| VC_558_73 | MH697579,MK359359 |
| VC_558_74 | MF919495 |
| VC_558_75 | ON551388 |
| VC_558_76 | MW822148 |
| VC_558_77 | MF919513,MK359308,MN062707 |
| VC_558_78 | JN699627 |
| VC_558_79 | MN735434 |
| VC_558_8 | AY129337,JN699013 |
| VC_558_80 | JN412588 |
| VC_558_81 | KX752698 |
| VC_558_82 | GQ303262,KX831080,MF919527 |
| VC_558_9 | KF006817,KX781992,MK359324 |
| VC_559_0 | EU826466,MF668280 |
| VC_559_1 | MZ820085 |
| VC_563_0 | KJ567043,MK016501 |
| VC_564_0 | MW584155,MW584175,MW584201 |
| VC_565_0 | DQ398047,DQ398051,EU676000,FJ168660,FJ168661,FJ168662,JF937107,JN699014,KJ194582,MH025888,MH316562,MH399788,MH834615,MK494102,MK524498,MK559427,MN119378,MN234227,MN284908,MT889385 |
| VC_566_0 | AY129339,MK279909 |
| VC_567_0 | EU770222,FJ174691,KF986247,KJ567046,MG962376,MH509445,MN010756,MT521996 |
| VC_568_0 | JF704112,KF416342,KT591076,KY969628,MH001447,MH926059,MK757446,MT684602 |
| VC_569_0 | JN412589,KR997933,ON108649 |
| VC_57_0 | ON470596,ON470604,ON470624,ON470627 |
| VC_570_0 | KX557285,MH025890,MH271313,MH576972,MT639639,MT771345,MT771348,MZ171372,ON970620 |
| VC_571_0 | MG757156,MH834613,MN585993,MT776806 |
| VC_572_0 | MN234162,MN234190,MN234207,MN234220,MN234232,MN586003,MZ005671,OL455887,OL455890,OL455895,ON755185,ON755187,ON970611,OP172868,OP172870,OP172876 |
| VC_573_0 | MN813687,ON108643 |
| VC_574_0 | JX901189,KF024728,MK279840,MW460248,MZ152914,OM793623,ON108645 |
| VC_575_0 | AY216660,HM035024,JX912252,KP085586,KU194205,KU194206,KX828710,KY985004,LC516895,MF093736,MF468274,MG049919,MH051911,MH051912,MH285980,MH917278,MK213796,MK335533,MK972831,MN153797,MN296515,MN337573,MN342247,MN393473,MT427400,MT682715,MZ501092,OL770108,OL800706,OL960578,OL960579,OL960582,ON185577,OP094641,OP455115 |
| VC_576_0 | MF996376,MH051918,MT023084,MZ501053 |
| VC_577_0 | AY236756,JN051154,JX486087,MG765275,MG765278,MH809528 |
| VC_578_0 | MG020111,MK504442 |
| VC_580_0 | KC954775,OM638103 |
| VC_581_0 | MW805361 |
| VC_581_1 | OM982621 |
| VC_582_0 | MN184887,MN732867,OP480062,OP480063 |
| VC_582_1 | OK136180,OK181858,OK181861 |
| VC_582_2 | KM190144 |
| VC_582_3 | KT934943,KX452695,KY652726,MG250486,MK024806,MT178275,MT438396,MT939253,MZ602147,ON602729,ON602736,OP296941 |
| VC_582_4 | JN882284,KF550303,MZ311864 |
| VC_582_5 | GU070616,JX181824,MW006477,MZ326168 |
| VC_582_6 | MG969411 |
| VC_584_0 | AY308796,KM236241,KX015770,KX015771,KY657202,KY694971,LR597656,MG241338,MH102284,MH491968,MH729819,MN994500,MT478992,MW021748,MW021750,MW598258,MZ501059,OK499973,OK499984,OL539443,OL539444,OL539461,OL539462,OL539465,OL539466,ON843697,OP292673,OP292674 |
| VC_585_0 | MN019128,MW032452,MZ501077,MZ501085 |
| VC_585_1 | MK778457,MN090155,MZ501099 |
| VC_585_2 | MW689258 |
| VC_585_3 | OM317910 |
| VC_586_0 | MN101222,OK019720,ON045001 |
| VC_587_0 | AY320035,KT221034 |
| VC_588_0 | KU958700,MH744420 |
| VC_589_0 | AY324828,MN710383 |
| VC_590_0 | AY343333,EF437941,EU863408,HE978309,KY683735,MF327006,MH560358,MK327932,MK327935,MK327941,MK327947,MK573636,MK962753,MN655998,MN709127,MN850574,MN850615,MN882349,MN894885,MT478991,MT764208,MW748999,MZ504876,MZ504877,MZ504878,ON286974,OX001802 |
| VC_591_0 | CP059058,KP063118,MG030346,MN062188,MN604053,MN840486,MW145139,OK499974,OK499975,OK499986,OK499998,OK500000,OL741431,OM962992 |
| VC_592_0 | JN871591,JQ340774,JX094499,JX296113,KC139512,KC139514,KC139515,KC139519,KC677662,KM366098,KM458633,KR296689,KR296691,KU927500,KX452699,KX452700,KX765862,KX765863,KX765864,KY014601,KY620117,LC553736,LR026998,MG280946,MH631453,MH791395,MH791396,MK249308,MK286578,MN871450,MT499897,MT499898,MT675124,MT675125,MT675126,MW057860,MW149272,MW355449,MW355460,MW355467,MW355470,MW355474,MW355476,MW355479,OL606627,OL699877,OM025231,OP453108,OP535471,OP535472 |
| VC_593_0 | KM581061,MH015249,MH015252,MH015254,MH015258 |
| VC_594_0 | KT321316,OK041469 |
| VC_597_0 | MK838113,MN317029,MT952005,MW978786,OL840901,OL964754 |
| VC_598_0 | HQ632859,KY249644,MF476925,MW269554 |
| VC_599_0 | AY370762,AY388628,AY423772,EU030938,EU030939,FJ870915,FJ870916,FJ870917,KC618393,KY563228,KY579375,MK054204,MK054205,MK054206,MK054207,MK054208,MK054209,MK054210,MK054211,MK054212,MK054213,MK054214,MK054215,MK054216,MK054217,MK054218,MK054219,MK054220,MK054221,MK054222,MK054223,MK054224,MK054225,MK054226,MK054227,MK054228,MK054229,MK054230,MK054231,MK054232,MK054233,MK054234,MK054235,MK054236,MK054237,MN496305,MN496306,MN496307,MN496308,X07234 |
| VC_6_0 | AY319521,CP019275,CP025900,HM770079,KT630647,KY271398,MK416009,MK416011,MK416012,MK416020,MK422449,MK422453,MK422454,MK433577,MK433578,MK433580,MK433581,MK433582,OK490395,OK490397,OK490399,OK490401,OK490403,OK490405,OK490407,OK490411,OK490413,OK490418,OK490423,OK490425,OK490427,OK490429,OK490431,OK490434,OK490436,OK490439,OK490442,OK490445,OK490448,OK490452,OK490455,OK490457,ON470620,OV696611 |
| VC_60_0 | OK490460,OK490463,OK490466,OK490469,OK490472 |
| VC_600_0 | MK577502,MN871507,ON042478 |
| VC_600_1 | AY375531,AY962392,KY290948,KY290951,KY290956,KY290957,KY290958 |
| VC_600_2 | DQ529280,HM452125,JN377894,JN377895,JX306041,MF479730,MH179476,MH179477,MW584871,OL840900 |
| VC_602_0 | JN190960,MW677528 |
| VC_603_0 | AY441783,AY848684,AY848685,AY848686,AY848687,AY848688,AY848689,OK041468,OM630588 |
| VC_604_0 | KR093634,KR093635,KR093636,KR093637,KR093639,KR093640,KR093641,KR093642,KR093643,KR093644,KR093645,KR093646,KR093647,KR093648,KR093649,KR093650,KR093651,KR093652,KR093653,KR093654,KR093655,KR093656 |
| VC_607_0 | AY505112,AY510084,JF712866,KM612259,KM612260,KM612261,KM612262,KM612263,KM612264,KM612265,KY883642,KY883647,KY883656,MH105773,MW824387,MW824404 |
| VC_608_0 | MG674163,MT992243,MW176032,MW176034,MW590698 |
| VC_609_0 | MF074189,MW202496,MW202711,MW202879 |
| VC_61_0 | AB334720,AB334721,F1CCG,GQ153916,GQ153917,GQ153918,GQ153919,JF719732,JF719733,JF719734,JX412914,PFDCG |
| VC_611_0 | AY569307,GU080336 |
| VC_612_0 | AY575774,DQ163914,KU884561,KU884562,KU884563,KU884564,KU884565,MK514283,ON857941 |
| VC_613_0 | JF412296,OM519319 |
| VC_614_0 | KX198614,MT795651,MW114771,ON457558,ON457559 |
| VC_615_0 | CAKLQA020000003,MZ326864 |
| VC_618_0 | JX128258,MH051335,MH816966,MK373776,MK373777,MK903282,MN038177,MN176217,MT446405,MT446408,OL856099,ON210137,OP352608 |
| VC_620_0 | KT361654,KT361655,KT361656,KT361657,OP494865,OP503543,OP503968,OP503969,OP503970,OP503971,OP503972,OP503973,OP503974,OP503975,OP503976,OP503977,OP503978,OP503979,OP503980,OP503981,OP503982,OP503983,OP503984,OP503985,OP503986,OP503987,OP503988 |
| VC_622_0 | MF176161,MH884512 |
| VC_623_0 | MK448818 |
| VC_625_0 | MK448890,MK448930 |
| VC_63_0 | AP009146,DQ785801 |
| VC_630_0 | AY625898,KC262634,KC900378,LT603684,MF490237,MK511039,MK511040,MK511041,MK511042,MK511043,MK511044,MK511045,MK511046,MK511047,MK511048,MK511049,MK511050,MK511051,MT118301,MZ285879 |
| VC_631_0 | MT152150,MT491207,MT491208 |
| VC_632_0 | FM864213,FN997652,KC348603,KC581799,KT336320,KT336321,KX077896,MK359990,MK448674,MK448741,MK448752,MK448811,MK448819,MK448828,MK448983,MN270258,MN270259,MN270260,MN270261,MN270262,MN270263,MN270264,MN270265,MN270266,MN270267,MN270268,MN270269,MN270270,MN270271,MN270272,MN270273,MN270274,MN270275,MN270276,MN270277,MN270278,MN270279 |
| VC_634_0 | KU052488,MG252693,MG765276 |
| VC_634_1 | JX486088,KX223815,KY381600,MG744354,MG765274,ON117106 |
| VC_636_0 | KM289195,MK448681,MK448683,MK448783,MK448790,MK448798,MK448833,MK448967,MK449010 |
| VC_637_0 | AY730274,EF177456,HE775250,HG934469,HG934470,JQ007353,JX202565,KC139511,KR270151,KT881477,KT962832,KU595434,KU951146,KU951147,KX245013,KX405002,KX649889,KY379853,KY626163,LC507823,MF001356,MF001366,MF431252,MG646668,MG969408,MG969409,MG969410,MH358359,MH370358,MH370359,MH370360,MH370361,MH370362,MH370363,MH370365,MH370372,MH370373,MH370374,MH370381,MH370384,MH370385,MH424443,MH424445,MH992509,MK249126,MK761195,MK761196,MK761197,MK761198,MK761199,MK770409,MK972691,MK972700,MK972701,MK972702,MK972703,MK972705,MK972706,MN336264,MN336265,MN336266,MN393078,MN722429,MN850616,MT074431,MT074432,MT074451,MT074462,MT074473,MT074481,MT074482,MT074483,MT074484,MT588083,MT632017,MT632018,MT632019,MT632020,MT653137,MT653144,MT677934,MT740291,MW201959,MW355463,MW355464,MW355465,MW715619,MW790495,MZ156567,MZ156568,MZ156569,MZ274225,MZ955866,OK108608,OK108609,OK143508,OL436139,OL519843,OL652658,OL770276,OM210019,OM291373,OM339548,ON167532,ON381768,ON409568,ON409569,ON409570,ON454038,ON526838,ON540301,ON720977,ON996339,OP484850,OP494211 |
| VC_637_1 | JX297445,KF562864,KF562865,KY176369,MK578530,MK759883,MK931446,MT074430,MT677933,MW355451,MW355452,MW355453,MW355454,MW355455,MW355456,MW355457,MW355458,MW355459,MW355461,MW355462,MW355466,MW355468,MW355469,MW355471,MW355472,MW355473,OK169616 |
| VC_637_2 | KF148055,KM272358 |
| VC_638_0 | GU196277,GU196278,GU196279,GU196280,GU196281,KY295895,KY295896,KY295898,MF153391,MG099933,MG252615,MG852086,MH051334,MK542015,MK737937,MK931438,MK931439,MK931440,MK931441,MN927225,MN927226,MT711523,MT711524,MT711525,MT711526,MT711527,MT711528,MW331437,MW722821,MZ234010,MZ234048,MZ576217,MZ576218,MZ832314,MZ997838,OK539620,OL539727,OM272052,OM632667,OM632668,OM810255,ON001686,ON391668,ON548432,ON556632 |
| VC_639_0 | MK813940,OM033133 |
| VC_64_0 | JN022534,MK798143,MK798144 |
| VC_640_0 | AY739900,EU340421,KF188410,KJ564036,KJ564037,KJ564038,MG913376 |
| VC_642_0 | HQ632860,MG592506,MG592545,MG592567,MG592568,MG592600,MG592630 |
| VC_642_1 | OX241574 |
| VC_642_2 | AY772740,JF974313,MG676223,MK097141,ON714422,OP375635 |
| VC_642_3 | KX507045,MG592494,MG592501,MG592508,MG592514,MG592583,MG592587,MG592595,MG592644 |
| VC_642_4 | MW824412,MW824413 |
| VC_642_5 | MG592544,MG592664,MG592665,OX241573 |
| VC_643_0 | JF974287,MK249872 |
| VC_644_0 | KC542353,KP994596,KU747973,MK387309,MN313256 |
| VC_645_0 | MG592390,MG592420,MG592423,MG592439,MG592444,MG592454,MG592465,MG592466 |
| VC_646_0 | MG592393,MG592477,MG592478,MG592518,MG592540,MG592552,MG592569,MG592570,MG592608,MG592614,MG592622,MG592627,MG592628,MG592641,MG592647,MG592656,MW824369,MW824373,MW824374,MW824378,MW824379,MW824383,OX241429,OX241438,OX241498,OX241557,OX241558,OX241575,OX241591 |
| VC_647_0 | CP069347,HG796225,HG798901,JX145342,LK985321,LN681538,LN681540,MK473382,MT193276,MW512571,MW512572 |
| VC_649_0 | KM983328,KM983331,KM983334 |
| VC_65_0 | MT366760,MT366761 |
| VC_652_0 | EU874396,GU229986,KX227758,KX227759 |
| VC_654_0 | KT725776,KX227757,KX227760,MW302330 |
| VC_655_0 | GU071099,MH512890 |
| VC_655_1 | GU071103 |
| VC_655_2 | AY940168,HQ337021,HQ634176 |
| VC_655_3 | HQ634175 |
| VC_655_4 | JN371769 |
| VC_655_5 | KU686207,KU686208,KU686209 |
| VC_655_6 | GU071106,HQ634190 |
| VC_655_7 | HQ634189,MH920639,MH920640 |
| VC_655_8 | GU071094 |
| VC_655_9 | GU071097 |
| VC_656_0 | HQ316583,HQ316603 |
| VC_656_1 | KJ019028,KJ019029,KJ019031,KJ019032,KJ019034,KJ019035,KJ019036,KJ019046,KJ019047,KJ019048,KJ019050,KJ019056,KJ019057,KJ019062,KJ019070,KJ019072,KJ019073,KJ019074,KJ019075,KJ019077,KJ019078,KJ019079,KJ019080,KJ019083,KJ019112,KJ019113,KJ019115,KJ019117,KJ019118,KJ019119,KJ019120,KJ019121,KJ019124,KJ019125,KJ019126,KJ019127,KJ019129,KJ019130,KJ019131,KJ019136,KJ019139,KJ019140,KJ019160,KJ019162,KJ019164,KJ019165 |
| VC_657_0 | KU686212,KU686213,MT586120 |
| VC_657_1 | KJ019037,KJ019045,KJ019052,KJ019053,KJ019058,KJ019059,KJ019066,KJ019085,KJ019086,KJ019090,KJ019091,KJ019092,KJ019093,KJ019095,KJ019096,KJ019097,KJ019098,KJ019099,KJ019100,KJ019101,KJ019102,KJ019103,KJ019105,KJ019106,KJ019107,KJ019111,KJ019123,KJ019141,KJ019142,KJ019143,KJ019144,KJ019145,KJ019146,KJ019147,KJ019148,KJ019149,KJ019150,KJ019151,KJ019152,KJ019155,KJ019159 |
| VC_659_0 | MH319737,MH319746 |
| VC_66_0 | MK552140,MK552141 |
| VC_66_1 | MN094788 |
| VC_66_2 | MW460245 |
| VC_66_3 | AB366653 |

| | |
|---|---|
| VC_662_0 | HQ317290,HQ317291,HQ317292,KX349226,KX349227,KX349228,KX349229,KX349230,KX349231,KX349232,KX349233,KX349234,KX349235,KX349236,KX349237,KX349238,KX349239,KX349240,KX349241,KX349242,KX349243,KX349244,KX349245,KX349246,KX349247,KX349248,KX349249,KX349250,KX349251,KX349252,KX349253,KX349254,KX349255,KX349256,KX349257,KX349258,KX349259,KX349260,KX349261,KX349262,KX349263,KX349264,KX349265,KX349266,KX349267,KX349268,KX349269,KX349270,KX349271,KX349272,KX349273,KX349274,KX349275,KX349276,KX349277,KX349278,KX349279,KX349280,KX349281,KX349282,KX349283,KX349284 |
| VC_664_0 | JQ245707,OP053360,OP053361,OP053362 |
| VC_666_0 | AY950802,JN968479,KC252997,KT809302 |
| VC_667_0 | KF929199,KJ804259,MT596499,MZ417336,MZ417339,MZ417343,MZ417348 |
| VC_667_1 | MH356730 |
| VC_667_2 | MK893987 |
| VC_668_0 | GU295964,HQ918180,KT001918,KT001919,KT321315,KX130727,LR746310,LT607758,MN101223,MT157285,MW448170,MZ322895,MZ612130,MZ707156,MZ707157,OK631812,OK631813,OM103621,OM971648,OM971649,ON515458,OP045497,OP684128,OX335396,OX335407 |
| VC_668_1 | OP617746 |
| VC_668_2 | MN101221 |
| VC_668_3 | MN101224 |
| VC_669_0 | JN882287,KT381880,KX431559,KX431560,KX443552,LC589952,MH823906,MN508621,MN508622,MN508623,MN508624,MT341500,MW021751,MW021756,MW250785,MW629017,MW767161,OL539470,OL539471,OL551674,ON157416 |
| VC_669_1 | AY967407,HE858210,HE981739,HM134276,KC801932,KM236237,KR869820,KX245890,MT334653,MW021749 |
| VC_669_2 | MN508817 |
| VC_669_3 | MN038175 |
| VC_669_4 | OK210075 |
| VC_669_5 | MT560058 |
| VC_67_0 | KY448244,MW366843 |
| VC_67_1 | MZ475891,MZ475892,MZ475893,MZ475894,MZ475895,MZ475896,MZ475897,MZ475898 |
| VC_67_2 | KY984068,MZ443769 |
| VC_67_3 | MH248138,MH460463,MZ443770,MZ443771,MZ443772,MZ443773,MZ443774,MZ443776,MZ443777,MZ443778,MZ443779,MZ443780,MZ443782,MZ443783,MZ443784,MZ443785,MZ443786,MZ443787,MZ443788,MZ443789 |
| VC_67_4 | MH389777,MH460460,MH460461,MH460462 |
| VC_67_5 | LT598654 |
| VC_670_0 | CAKLQA020000002,MW176033 |
| VC_671_0 | MN937349,MW393850,MZ326859,MZ398240,MZ398241 |
| VC_672_0 | BCP1,MG883715,MG883725,MG945215,MG945341,MG945520,MG945551,MG945686,MG945756,MT135344,MT135404,MT135412 |
| VC_673_0 | BK010471,MH552500,MH616963,MK069403,MK238400,MT006214,MT774381,MT774383,MT774384,MT774386,MT774394,MW067000,MW067001,MW067002,MW067003 |
| VC_68_0 | AP018399 |
| VC_68_1 | MN047793 |
| VC_681_0 | BK013345,BK013346,BK013347 |
| VC_682_0 | BK059895,OM373202 |
| VC_683_0 | BK061308,BK061309,BK061310,BK061311,BK061312 |
| VC_684_0 | MW353181,MW584168,MW584172,MW584178,MW584190 |
| VC_685_0 | MG872834,MH513979,MH651167,MK284520,MK433274,MK433275,MK820641,MN062715,MT521999,MW534378,MW601219,MZ322028,MZ622161,MZ622165,ON108651,OP434460 |
| VC_686_0 | MK494099,ON260818 |
| VC_686_1 | KU728633,MK977701 |
| VC_687_0 | KF114879,KF114880,KX656785 |
| VC_687_1 | KF114876,KF114877 |
| VC_688_0 | MF974396,MF974397 |
| VC_689_0 | DQ490056,MW791312 |
| VC_690_0 | C2PVCG,KX373685,KX373686,KX373687,KX373688,KX373689,KX373690,KX373691,KX373692,L67CG,MF443119,MF443120,MF443121,MF443122,MF443123,MF443124,MF443125,MF443126,MH779518,MH779519,MH779520,MH779521,MH779522,MH779523,MH779524,MH779525,MH779526,MH779527,MK301442,MK301443,MN337887,MN689503,MN689504,MN689505,MN689506,MN689507,MN689508,MN689510,MN689511,MN689512,MN689513,MN689515,MN689526,MN689527,MN689528,MN689529,MN689531,MN689532,MN689533,MZ955869 |
| VC_691_0 | MT939241,ON506926 |
| VC_691_1 | CAJCJZ010000002,LT546029,LT615366,MT119360,MT119361 |
| VC_692_0 | MF001358,MF001361 |
| VC_694_0 | MW672041 |
| VC_695_0 | CAJDJX010000002,CAJDKF010000002,JX193904,KF728385,KF733017,KJ127304,KR131750,KT932701,KX284704,LC623721,MF678788,MF678789,MF678790,MG264739,MG708276,MH193369,MH203383,MH203384,MH355583,MH375074,MK125140,MK360024,MK721186,MK721187,MK721190,MK721191,MK721199,MK721200,MK982307,MT119359,MT520979,MT627482,MT661597,MT661598,MT829326,MT857001,MZ182246,MZ272341,MZ326865,MZ333461,OL505084,OL539449,OL799256,OL799257,ON113176,ON113334,OP254195,OP559177 |
| VC_696_0 | KT588075,MZ514874 |
| VC_696_1 | CAKLQA020000001,JX403940,KP861230,KT588073,MH115576,MT361972,MZ675741,ON391949 |
| VC_697_0 | KJ507100,MT104473,MT104475,MZ348425,MZ868718 |
| VC_698_0 | HE806280,HM368260,JX976549,KJ817802,KP861229,KP861231,KU510289,KY670595,KY829116,MF346584,MH853786,MH853787,MH853788,MN166083,MN516421,MN516422,MZ593728,MZ618622,MZ712044,OL743187,OM471864,ON036882,ON036883,ON237674,ON480525,OP585104 |
| VC_699_0 | CAKLQB020000001,CAKLQG020000001,HQ186308,JX658790,KC311669,KR149290,KT339321,KT388102,KT388103,KT804908,KY082667,KY268295,KY268296,KY888680,MF033347,MF033348,MF033349,MF033350,MF033351,MG459218,MG599035,MH042230,MH763831,MH800200,MK089780,MK257719,MK257720,MK257721,MK257722,MK257723,MK278859,MN061582,MN212906,MN294712,MN395291,MN433707,MN604238,MN604239,MN614471,MN651570,MN807295,MT263719,MT633129,MT741944,MW056501,MW056502,MW056503,MW345241,MW366783,MW366784,MW459163,MW926912,MZ099557,MZ593174,MZ868724,MZ868725,MZ868726,MZ936314,MZ936315,MZ936316,MZ967493,OK546191,OL770263,OL964948,OM925528,ON210142,ON513429,OP019135,OP171942,OP291336,OP508218 |
| VC_7_0 | AB009866,AB044554,AB243556,AF424783,CP062426,CP062435,CP062445,CP062457,EF462197,EF462199,FJ713816,GQ915271,KC342645,KX232515,KY389063,KY389064,KY653125,KY695153,MH384260,MH401414,MH401415,MW147366,MW192778,MW627293,MW650841,OM439664,OP136133,OP493557 |
| VC_70_0 | AB920995,AB983711,GU393987,HG793132,HG796219,HG796220,HG796221,KC853746,KT381879,KX660669,KY316062,KY464836,KY981272,MF621978,MH684921,MN191861,MT740728,MZ326863,OK319016,OK335783,ON170174,ON170178,ON170184,ON170190,ON170318,ON170358,ON529868 |
| VC_700_0 | KJ628499,MH746814,MH800198,MH800199,MK170160,MT188223,MT623546,MW349133,OL960030,OP585105 |
| VC_700_1 | KJ473422 |
| VC_701_0 | MN276049,MZ477002 |
| VC_701_1 | MH133207,MZ043613 |
| VC_701_2 | CAKLQE020000001 |
| VC_702_0 | KM672662,KY000079,MW316731,MW316732,MW316733 |
| VC_704_0 | DQ150593,DQ222851,DQ222853,DQ222855,DQ289555,DQ289556,JQ062992,KY963369,KY963370,KY963371,MK085976,MT745954,MT745955,MT745956,OK499979,OK499999 |
| VC_705_0 | KF591601,MF614100,MH363708,MK105855,MK125141,MK651787,MN850581,MN850588,MN850623,MW258710,MW258711 |
| VC_706_0 | CP002495,CP091911 |
| VC_708_0 | KF010834,KP202972,KP296791,KP296793,KP296794,KP296795,KP296796,KT361649,KT361650,KT361652,MG727695,MG727696,MG727698,MG727699,MG727700,MG727701,MG727702,MH431931,MH431932,MH431933,MH431934,MH431935,MH431936,MH431937,MH454077,MH454078,MH454079,MH454082,MH454083,MH454084,MW927511,MW927513,MW927514,MW927515,MW927516,MW927517,MW927519,MW927520,MW927521,MW927522,MW927523,MW927524,MW927525,OP503992,OP503993,OP503994,OP503995 |
| VC_713_0 | ON464760,ON464761,ON464762 |
| VC_718_0 | KP137433,KP137435,KP137437,KP137439 |
| VC_72_0 | JN882298,MG201401 |
| VC_720_0 | KT151955,KT151957 |
| VC_720_1 | CP027117 |
| VC_721_0 | CP045566,MN890136,MN890137,MN890138,MN890139,MN890140,MN890141,MN890142,MN890143 |
| VC_722_0 | KJ419279,KP719132,KP719133,KP719134,KY926791,MK165087,MK373794,MK373795,MT682705,MZ501057,MZ501068,OM386652,ON922918 |
| VC_723_0 | CP053388,CP054387,CP058330,CP101890,CP101896,JQ965645,MK422451,MZ779062,OV696614 |
| VC_724_0 | MT770738,MT778838,MW960030 |
| VC_724_1 | JX483881,MN988505,MN988506,MN988543,MN988547,MN988548 |
| VC_725_0 | MG018927,MT354569,OM140839,OM223115 |
| VC_726_0 | JN116824,KR053195,MH153804 |
| VC_727_0 | JX182371,KY092479,KY092480,KY092481,KY092482,KY092483,KY092484,KY676784,MH825699,MK305888,MT711975 |
| VC_728_0 | MK279841,MT316461,MT723933 |
| VC_730_0 | CP060690,HQ377372,HQ377373,JF773396,KX879601,KX879602,KY661963,MT899443,MT899444 |
| VC_731_0 | KX190833,MK759918 |
| VC_733_0 | HM568888,KU057941,LN681535,LN681536,LN881738 |
| VC_739_0 | DQ003638,DQ004855,JQ797329,JX126918,KJ094029,KJ094030,KJ094031,KJ094033,KJ535721,KJ535722,KJ586794,KJ586795,KJ591604,KJ591605,KJ668715,KM373208,MK792752,MN128594,MN172529,MN939539,MN939540,MT178766,MT178767,MT438761,MT668623,MT668624 |
| VC_74_0 | KF626667,KU197013,KU595432,KU595433,LR778216,MF375456,MK903278,MN478375,OK258138,OK275495,ON911539,ON932078,ON932081,ON932083,ON960072 |
| VC_740_0 | DQ003641,EU855793,GQ403788 |
| VC_743_0 | DQ079870,DQ079871,DQ079872,DQ079873,DQ079874,DQ079875,DQ079876,DQ079878,DQ079879,DQ085810,GQ149088,MG945254,MK629527,X60322,X60323 |
| VC_745_0 | MT006233,MT006238,MT006239,MT006240 |
| VC_747_0 | DQ163912,KJ959591,KY707339,MK510962,MK510963,MK510964,MK510965,MK510968,MK510969,MK510970,MK510971,MK510974,MK510975,MK510976,MK510978,MK510984,MK510985,MK510986,MK510987,MK510988,MK510990,MK510992,MK510993 |
| VC_748_0 | MK510966,MK510967,MK510972,MK510973,MK510977,MK510979,MK510980,MK510981,MK510982,MK510983,MK510989,MK510991 |
| VC_749_0 | DQ163913,HG962376,JF939047,JQ067092,JQ307387,KM434185,KR537871,KR537872,KR935213,KR935215,KU197014,LC552830,LR743530,MG897800,MK165657,MK165658,MK574078,MN062187,MT118304,MT118305,MW512832,MW677514,MW677526,MZ501266,MZ927745,MZ927746,OK094665,OK330455,OK330456,OK539825,OL802211,OM234791,ON189042,ON189043,ON189044,ON911538,ON932082,ON932085,OP009282,OP422637 |
| VC_75_0 | MZ666965,MZ666966 |
| VC_750_0 | MG366115 |
| VC_751_0 | KP313532,MK962635,MK962636,MK962637 |
| VC_752_0 | MN734437,MN734438,MN734439 |
| VC_754_0 | HQ641345,HQ641346,KM224878,KM224879,KY883657,KY883658,MT066160,MT767883 |
| VC_755_0 | DQ163915,KU535860,LT986460,MF788075,MG676466,MW929175,MZ089731,MZ727333,ON857932 |
| VC_756_0 | KC751414,KF302037 |
| VC_757_0 | MG592599,MG592619 |
| VC_758_0 | MG592629,MW824407,MW824408,MW824409,MW865297,MW865299,MW865302,MW865307,OL676770,OX241435,OX241436,OX241445,OX241446,OX241447,OX241501,OX241504,OX241559,OX241560 |
| VC_759_0 | MN988507,MN988510,MN988527 |
| VC_76_0 | GU988610,HE983845,HQ832595,KC862297,KC862298,KC862300,KR052143,KT736033,KT804923,KU297675,KU497559,KU761955,KY000083,KY073228,LN610572,LN610575,LN610586,LN610587,MF974178,MH370478,MH725810,MH791399,MH791408,MH791416,MK138526,MK224498,MK340761,MK443264,MN013356,MN563785,MN871470,MN871481,MN871488,MW526259,OK539826,ON169972,ON361135 |
| VC_76_1 | MK443265 |
| VC_760_0 | MW202785,MW202796,OK172329 |
| VC_761_0 | DQ398042,EU568876,FJ973624,GQ303261,JN412593,JN699002,KC787103,KC787104,KC787107,KC787108,KC787109,KC787110,KC787111,KC787112,KJ725374,KM923970,KT347314,KT355472,KT355474,KT365399,KX443326,KX534004,KX588251,KX641265,KX641266,KX664455,MF668268,MF668272,MF919497,MG099946,MH001455,MH001456,MH045569,MH077584,MH371116,MH371119,MH450127,MH479920,MH590605,MH779505,MH779507,MH779509,MH779513,MH779514,MH779515,MK305884,MK310146,MK433277,MK433279,MK524493,MK524494,MK919475,MN444875,MT818422,MW119570,MW291028,MZ171377,MZ747508,ON392160,ON392162,ON637759,ON637765,ON637766,ON645337,ON970618,OP172872,OP294982 |
| VC_762_0 | KJ410133,KR080197,KR080198,KR080202,MF141540,MK494105,MN234183,MN234185,MN234219 |
| VC_763_0 | MW584160,MW584202 |
| VC_763_1 | JN699628,KC900379,KT591491,MH697587,MK937612,MN586024,MT658803,MW713383,OP297551 |
| VC_763_2 | JF937105,KY223999,KY783914,MF319184,MT657341,MT658802,MW578836 |
| VC_764_0 | DQ398052,KP027195,KY385381,MN585992 |
| VC_765_0 | DQ431235,FJ706171,FJ706172,JX262215,JX262216,JX262217,JX262218,JX262219,JX262220,JX262221,JX262222,JX262223,JX262224,JX262225,JX570702,JX570703,JX570704,JX570705,JX570706,JX570707,JX570708,JX570709,JX570710,JX570711,JX570712,JX570713,JX570714,KJ578758,KJ578759,KJ578760,KJ578761,KJ578762,KJ578763,KJ578764,KJ578765,KJ578766,KJ578767,KJ578768,KJ578769,KJ578770,KJ578771,KJ578772,KJ578773,KJ578774,KJ578775,KJ578776,KJ578777,KJ578778,KJ578779,KJ578780,KJ578781,KJ578782,KJ578783,KJ578784,KJ578785,KJ578786,KJ578787,KJ578788,KJ578789,KJ578790,KJ578791,KJ578792,KJ722067,KR337643,KR337644,KR337645,KR337646,KR337647,KR337648,KR337649,KR337650,KR337651,KR337652,KR337653,KR337654,KR347352,KR347353,KR347354,KR347355,KR902978,KR902979,KR902980,KR902981,KR902982,KR902983,KR902984,KR902985,KR902986,KR902987,KT934381,KY926792,KY926793,MF919491,MF919505,MF919515,MF919516,MF919522,MF919533,MG820632,MG820633,MG820634,MG820635,MG820636,MG820637,MG820638,MG820639,MG820640,MG820641,MG820642,MG820643,MG820644,MG820645,MN329678,MN813675,MN813677,MN813679,MN813683,MN813688,MN813689,MN813690,MN813691,MN813692,MT647607,MW161461,MW161462,MW161463,MW161464,MW161465,MW161466,MW161467,MW161468,MW505928 |
| VC_766_0 | LN681534,MN718463 |
| VC_767_0 | JF923796,JF923797,KR053201,MF919510,MF919542,MG757157,MG757166,MH536819,MH536824,MH576960,MH669007,MH669013,MH727547,MH727552,MH744424,MH779499,MH779501,MH779508,MH779510,MK376954,MK376960,MK376962,MK433272,MK977699,MN369753,MN369766,MN428063,MT889393,MT986025,MW507129,MZ322010,MZ322022,MZ622178,ON456333 |
| VC_768_0 | MG198776,MG198777,MG198778,MG198780,MH926056,MH926057,MH976516,MK977700 |
| VC_769_0 | MW452562,MW582531 |
| VC_77_0 | AB472900,KC862295,KC862296,KC862299,KC862301,KM434186,KY994101,LN610573,LN610576,LN610581,LN610582,LN610583,MK799650,MT108728,MT108730,MT118294,MT118295,MT118300,OM421595,ON653033 |
| VC_770_0 | DQ500118,MN988496,MW960032,OP484857,OP484858 |
| VC_771_0 | MN988490,MN988502 |
| VC_772_0 | MG065644,MG065662,MG065672,MG065674,MG065677,MG065690 |
| VC_772_1 | MK962749,ON286975 |
| VC_772_10 | MN850630,MT884008,MT884009,MT884010,MT884011,MZ501082,MZ501083,MZ501102 |
| VC_772_11 | LC739541,LC739542,MH491969,MH752840,MZ501048,MZ501094 |
| VC_772_12 | MG065659 |
| VC_772_13 | MG065653 |
| VC_772_14 | MG065654,MG065655,MG065666 |
| VC_772_15 | DQ832317,KC690136,KP869102,KP869103,KP869112 |
| VC_772_16 | KJ190158,LC739538,LC739540,MG065646,MG065647,MG065658,MG065661,MG065682,MG963916,MN850642 |
| VC_772_17 | MW748998,MZ501073 |
| VC_772_18 | MZ501061 |
| VC_772_19 | KT001917,LR699804,MG065635,MK327930,MK373780,MN850576,MN850578,MN850595,MN850597,MN850612,MN850626,MN850627,MZ501054,MZ501062,OK272482,OV101294 |
| VC_772_2 | MZ501060 |
| VC_772_20 | MN850593,MT682716 |
| VC_772_3 | MZ501093 |
| VC_772_4 | JQ031132,MK883717,MK883718,MW883061 |
| VC_772_5 | MT740314 |
| VC_772_6 | MN850646,MN850649,MZ501111 |
| VC_772_7 | LR694610,LR694611 |
| VC_772_8 | KR698074,ON286973 |
| VC_772_9 | KY683736,LN881727,LN881735,LR597647,LR694165,MG022439,MG022440 |

| | |
|---|---|
| VC_773_0 | DQ864624,MK448678,MK448689,MK448693,MK448696,MK448697,MK448699,MK448766,MK448779,MK448784,MK448789,MK448797,MK448852,MK448866,MK448867,MK448868,MK448871,MK448947,MK448952,MK448961,MK448968,MK448974,MK448976 |
| VC_774_0 | GQ478081,GQ478082,GQ478083,GQ478084,GQ478085 |
| VC_775_0 | MK448854,MK448933,MK448950 |
| VC_775_1 | MK448673,MK448765,MK448775,MK448794,MK448795,MK448858,MK448864,MK448941,MK448944,MK448945,MK448946,MK448953,MK448957,MK448970 |
| VC_777_0 | EF537008,KF598865,MT457475 |
| VC_778_0 | DQ875742,OM897575,ON677538,OP056300 |
| VC_779_0 | EF056009,HQ259105,JX415535,JX880034,KC206276,KF192075,KF562340,KF620435,KJ135004,LR999871,MG208881,MG770228,MG922974,MH358458,MH669274,MH733496,MH992123,MK931297,MT682708,MW175414,MW630115,MZ358387,MZ394712,MZ501049,OM632664,OM632665,OM632666,OM632669,OM632670,OM632672,OM982646,ON602749,ON602751,ON922919,OP312987,OV049961,OW991345 |
| VC_779_1 | KF787094,KF787095,KT321317,MG018928,MK962626,MK962629,MK962630,MK962631,MK962632,MK962639,MK962641 |
| VC_779_2 | KF806588,KX098389,KX098390,KX098391,MH160392 |
| VC_779_3 | MG983743,MK863032,MN062186,MZ326856,MZ326857,MZ326861,OM687511 |
| VC_78_0 | MG018930,MZ605293 |
| VC_78_1 | KU130126,KU130127,KU130128,KU130129,KU130130,KU130131 |
| VC_78_2 | OK040171 |
| VC_78_3 | KJ409772,MN905160,MT670417,MT670418,MT670419,MT670420,MT670421,MT670422,MW175491,MZ348426,MZ868713 |
| VC_78_4 | MN812694 |
| VC_78_5 | LN887844,MZ826350,MZ826353,OK258139,OK275491,OM982620 |
| VC_78_6 | MF893340 |
| VC_780_0 | KC139517,KC139520,KM236242,MH899585,MN163281,MW544066,OM858838,OM908766 |
| VC_781_0 | KU356690,KU356691 |
| VC_782_0 | MG592603,MG592604,MG592610 |
| VC_783_0 | MN228696,MW960036,MW960038 |
| VC_784_0 | KC438282,KC438283,KP280062,KY065148,KY612839,OP313112 |
| VC_786_0 | MT234338,MT234339,MT234340 |
| VC_79_0 | AB560486,AP018815,FM201282,JN254800,KP340287,KP340288,KR869157,KX171208,LC102730,LC105987,LC105988,LC105989,LC105990,LC472883,LC472884,LN610579,LN610588,MF490240,MG897799,MN871458,MN871459,MN871461,MN871462,MN871464,MN871465,MN871466,MN871473,MN871479,MN871485,MN871487,MN871508,MN871509,MT118291,MT118297,MT118298,MT118299,MT119363,MT119365,MT119368,MT119376,MT133561,MT133562,MT349888,MT413450,MT933737,MW512833,MW595221,MZ553930,OL362268,OL362269,OL754588,OM953790,ON324181,ON375838,ON375839,ON375840,ON529291,ON857930,ON857934,ON857942,OP310975,OP310977,OP314870,OP314871 |
| VC_79_1 | LC472881,MW406974 |
| VC_79_10 | AP019535 |
| VC_79_11 | KY000082 |
| VC_79_2 | FM201281 |
| VC_79_3 | OP310979 |
| VC_79_4 | MF623055,MT118289,MT118292 |
| VC_79_5 | MK837009,MW595220 |
| VC_79_6 | MK837010,MN871457,MN871460,MN871467,MT108729,MT491204,MT491205,MT491206,MW512830,MW512834,OP310974 |
| VC_79_7 | FM887021,FM897211,GU815091,KR054033,KU198331,MF490241,MF768470,MK318076,MN131141,MN131142,MN131143,MN615700,MN615701,MN615702,MT108727,MT118290,MT119366,MT119369,MZ285878,ON857925,ON857929,ON857937,ON857938,OP310970,OP310971,OP310972,OP310973,OP310976,OP310978,OP310980,OP310981,OP310982,OP314872 |
| VC_79_8 | KM434184 |
| VC_79_9 | MK340760,MN871468 |
| VC_791_0 | EF432053,KP282673,KP282674 |
| VC_792_0 | EF460875,HQ728264,KF534715,KY087898,MF044458,MH191398,MN045229,MN098326,MN117721,MN122072,MN136198,MN812688,MN850594,MT230534,MT446397,MT446398,MT446399,MT446400,MT446402,MT446403,MT446406,MT446409,MT446419,MW147598,MW147599,MW147600,MW147601,MW147602,MW349138,OK073976,OK129343,OK148439,OL741059,OL960574,ON778463 |
| VC_793_0 | KJ002054,KJ858521,KY709296,LR813619,MT623578,MW380983,MW380984,MW380985,MZ336020,OL770365,OM810291,OP056089 |
| VC_794_0 | EF506570,SVU28974,X51344 |
| VC_795_0 | KM236243,KP064094,KX534337,LN881725,LR025195,LR025196,LR025197,LR025198,LT907986,MH230177,MK672798,MZ501071,MZ501091,MZ501104,MZ501110,MZ501112,OL539454,OL960573,OL960575 |
| VC_796_0 | MK816297,MN029011,MN871471,MN871472,MN871474 |
| VC_799_0 | GU580941,GU580942 |
| VC_799_1 | MG962368,MG962369,MK376953,MK376957,MN010761,MN204501,MN284896,MN284904,MN585971,MN585982,MN585988,MN585996,MT310850,MT498039,MT498052,MT521994,MW291022,MW862987,MZ005675,MZ005683,MZ171370,MZ209301,MZ388556,MZ622168,ON526971,ON970572,ON970622 |
| VC_8_0 | AB045978,AF424782,AP009363,AP012341,AY954955,AY954956,AY954957,CP062434,CP062442,CP062444,CP062461,EF462198,EU861005,JQ779023,JQ957932,JX174275,KF598856,KT186243,KY056620,LC541428,MF580410,MG029509,MG029511,MG029513,MG029514,MG029515,MG029516,MG029517,MG029518,MG770897,MH384261,MK348510,MK801680,MK801681,MK940809,MT366568,MT899504,MW924497,OL690335,OL840290,OM304856,OM439665,OM439667,ON856257,ON856258,ON989481,OP038318,OP038319,OP136131,OP136132,OP136134,OP328420,OP328422,OP374154,OP374155,OP374156,OP584903,OP584904 |
| VC_80_0 | DQ163917,EU716414,KF981875,KR054028,KR054030,KT254130,KT254131,KT254132,KT254133,KT372690,KT372691,KT372692,KT372693,KT372694,KT372695,KT372696,KT372697,KT372698,LC472882,LN610589,MH536736,MK317959,MN508620,MN563783,MN564818,MN871454,MT108726,MT118293,MT118303,MT119364,MT119367,MT119370,MT119372,MT119374,MT119375,MT119377,MT119378,MT133563,MW557846,MZ089729,ON361134 |
| VC_800_0 | EU081845,MK779875,MN552143 |
| VC_801_0 | KJ489010,KJ489011,KJ489012,KJ489013 |
| VC_802_0 | KU160642,KU647628,MF189170,MF189171,MF189172,MF189173,MF189174,MF189175,MF324906,MH450120,MK919473,MK967398,MN284897,MZ209300,MZ209307,MZ747520,OP434451 |
| VC_803_0 | MH153800,MH153801,MK814756 |
| VC_805_0 | EU100883,EU100884,KX987127,MH992131 |
| VC_806_0 | KP792623,MW202557,MW202655 |
| VC_807_0 | EU124666,MG592549,MG592550,MG592581,MG592582,MK796797,MW824388,MW824389,MW824392,MW824393,MW824394,MW824398,MW824399,MW824402,MW824403,MW824405,OL762410 |
| VC_808_0 | MW202520,MW202674 |
| VC_809_0 | KJ028219,MG770216 |
| VC_810_0 | EU203571,KJ538722,KT222940,KT246485,KT454972,MF919517,MH697577,MH727549,MK494095,MK919482,MT114159,MT657330,MT684594,MT818421,MW924639,MZ005672,MZ622181,OP297533 |
| VC_812_0 | HM997019,MG873442,MW296032 |
| VC_812_1 | LC519601,MT028297,MT664722,MT740244 |
| VC_812_2 | JN882286,MW343794 |
| VC_812_3 | OL799327 |
| VC_813_0 | JX415536,LC553734,MK310183,MT446407,MT446413,MT446414,MZ234015,MZ234017,MZ342906,MZ398243,OK148440,OP293233 |
| VC_813_1 | EU330206,JX867715,KF981730,KM044272,KP308307,LC553735,MG673519,MG748548,MG976803,MG983840,MK373770,MN045231,MN148435,MN508615,MN715356,MT127620,MT129653,MT129655,MT446386,MT446410,MW741821,MW929182,MZ234028,MZ648214 |
| VC_814_0 | EU350361,KY296500,KY296501 |
| VC_815_0 | EU588980,GQ443085,JF767208,JF767209,JF767210,MH999280,MZ401006,MZ995505 |
| VC_817_0 | EU622808,EU771092,KU831549,MF407276,MT459794,MW354668,MW354670,MW477480,MZ570263,OL744111,OM249958,ON042750,PZACG,X99260 |
| VC_818_0 | KC685370,KX349899,KX349900,KX349901,KX961631,KY947509,MF156577,MF156578,MF156580,MH817022,MK084630,MK284526,MK284527,MK284528,MN082624,MT777452,MW012634,MW348917,MZ384014,OL744112 |
| VC_82_0 | DQ029335,FJ409640,HQ641340,HQ641341,HQ641342,HQ641343,HQ641344,JQ780163,KY883641,KY883643,KY883644,KY883645,KY883646,KY883648,KY883649,KY883650,KY883651,KY883652,KY883653,MF574151,MG957431,MT215167,ON464735,ON464736 |
| VC_823_0 | EU734173,GQ413937,HG994092,HM480846,KR149291,KT367885,KU183006,KX591654,KX712070,KX712071,KY389315,KY389316,KY450753,KY652723,KY652724,KY652725,LC413193,LC413194,LC413195,LN866626,MF285615,MG835568,MH172261,MH172262,MH172263,MH172264,MH587638,MK134560,MK203841,MK380015,MK380016,MK521905,MK618658,MK795385,MK903728,MN052874,MN101216,MN101218,MN101227,MN149903,MN176574,MN434093,MN585130,MT425185,MT446417,MT446418,MT542697,MT813140,MT813141,MT966872,MT966873,MW042787,MW042788,MW042789,MW042790,MW042791,MW042792,MW042793,MW042794,MW042795,MW042796,MW042797,MW042798,MW042799,MW042800,MW042801,MW042803,MW042804,MW042805,MW042806,MW042807,MW042808,MW042809,MW042810,MW042811,MW042812,MZ424865,MZ571832,OK138556,OL409034,OL799328,OM194188,OM194189,OM194190,ON148528,ON148529,ON571554,ON602742,ON602753,ON602755,ON880501,OP013026,OP045496,OP103917,OP103918,OP103919,OP103920,OP103921,OP103922,OP103923,OP256047,OU342754,OX335359,OX335360,OX335375,OX335376,OX335377,OX335389,OX335390,OX335392,OX335394,OX335398,OX335401,OX335408,OX335409,OX335413,OX335414,OX335418,OX335426,OX335427,OX335433,OX335438,OX335440,OX335443 |
| VC_826_0 | ON170019,ON170022,ON170028,ON170033,ON170036,ON170038,ON170040,ON170049,ON170054,ON170074,ON170086,ON170088,ON170090,ON170092,ON170094,ON170096,ON170098,ON170100,ON170102,ON170104,ON170106,ON170108,ON170110,ON170112,ON170114,ON170116,ON170118,ON170120,ON170122,ON170124,ON170126,ON170142,ON170144,ON170146,ON170152,ON170154,ON170156,ON170158,ON170160,ON170162,ON170164,ON170167,ON170176,ON170180,ON170186,ON170189,ON170192,ON170194,ON170196,ON170198,ON170200,ON170202,ON170204,ON170206,ON170211,ON170215,ON170217,ON170219,ON170221,ON170222,ON170225,ON170244,ON170246,ON170248,ON170250,ON170252,ON170271,ON170280,ON170282,ON170284,ON170289,ON170291,ON170293,ON170295,ON170297,ON170298,ON170300,ON170306,ON170307,ON170311,ON170313,ON170315,ON170330,ON170332,ON170334,ON170336,ON170338,ON170340,ON170352,ON170356,ON170360 |
| VC_827_0 | KJ018213,MG592431,MG592432,MG592435,MG592458,MG592666 |
| VC_828_0 | JQ619704,MG784342,MH598512 |
| VC_829_0 | EU887664,JX887877,KF296717,KX190834,MT136606 |
| VC_83_0 | AB597179,KP343639,OK274245,OK318991,OP313111,OP617560 |
| VC_830_0 | FJ008913,HE608841,MN078104,MT630433,MW314138,MW916539 |
| VC_833_0 | FJ591093,FJ591094,FR682616,FR719956,HQ317387,KJ621082,KJ803031,KU885988,KU885989,KU885990,MH015250,MH015253,MH015256,MH015257,MH015259,MZ773648 |
| VC_834_0 | FJ685651,GU321093,JN882264,JN882266,KF056323,MW344764,MW344766 |
| VC_835_0 | FJ750948,GQ502199,JN984867,JQ086375,JX865427,KM233151,KM896878,KX534335,KX534336,KX534338,KX534339,KX534341,LN881730,LN881731,LR597646,LR990702,LR999870,LT960609,MG065648,MG581355,MK138527,MK373789,MK373790,MK373791,MK728541,MN850575,MN850589,MN850628,MN850629,MN850650,MT331608,MT446389,MT446391,MT478993,MT884012,MT884013,MT884014,MT884015,MT951623,MW749005,MW822006,MW862804,MZ234008,MZ234014,MZ234016,MZ234026,MZ234029,MZ501070,MZ501088,MZ501095,MZ501097,MZ501107,OK272474,OK272488,OK310512,OK310513,OK310514,OK499985,OK499988,OK499993,OL615010,OL770278,OM262540,OM262541,ON184124,ON210136,OP046317 |
| VC_836_0 | KT367887,MN656993,MT075871,MW419910,OL539440,OM687892,ON602734,ON602747,ON602766 |
| VC_837_0 | FJ896200,LR794124,MF754111,MF754112,MK474470,MN974282,MT178448,MT501516,MW250641,MW331544,MZ020222,MZ182247,MZ516827,MZ592921,OK349506,OK585158,OK585159,OK585160,OM362522,OM799543,ON080940,ON080941,ON457554,ON457555,ON457556,ON457557,ON864052,ON872379,OP117450 |
| VC_838_0 | FN422399,HG962375,JX194238,KM411958,KM411959,KM411960,KM974184,KP994390,KR052142,KR054032,KU948710,KX171209,KX171210,MF490236,MF490238,MH356729,MN553583,MN553584,MN906996,MT080102,MZ955867,OK665488,OL519844,OM234792,OM937766,ON815901,ON815902,ON815903,ON857943 |
| VC_839_0 | FN422398,LC064302,MT416090,OL519842,OM782452 |
| VC_841_0 | KY083726,KY514264,KY514265,KY549659,KY769270,MH059638,MN812685,MN812687,MN812691,MZ348421 |
| VC_842_0 | MG775260,MH179480 |
| VC_843_0 | FN667788,FN667789,HE815464,MK408758,MT932329 |
| VC_844_0 | FR823450,GU296433,JN132397,JX569801,KF148616,KX229736,KX236333,KX879627,MH107028,MT863714,MT863715,MT863716,MT863717,MT863718,MT863719,MT863720,MT863721,MT863722,MT863723,MT863724,MT863725,MT863726,MT863727,MT863728,MT863729,MT863730,MT863731 |
| VC_845_0 | MK387337,MW451250,OM249648 |
| VC_846_0 | MK388689,OK570186 |
| VC_847_0 | LC739533,LC739534,LC739535,LC739537,LC739539 |
| VC_847_1 | JX561091,KX552041,KX664695,LR597654,LR746137,MG065650,MH051333,MH160766,MH252123,MK373788,MN850565,MN850571,MN850598,MN850618,MN850648,MT496970,MT944117,MW175890,MZ726793,OL505078,OL739525,OM362897,OM386656,OM386659,OM386667,OP562383 |
| VC_848_0 | FR775895,KU522583,LC739536,LR865361,MF805809,MG065639,MG065645,MG065656,MG065689,MH675927,MH816848,MN850573,MN850584,MN850601,MN850632,MN850645,MT932211,MZ501063,MZ501084,MZ501100,OK210077,OK272470,OM386653,OM386654,OM386658,OM401812,OP177727,OX090893 |
| VC_848_1 | OP617743 |
| VC_849_0 | MH533020,ON287369,ON287374 |
| VC_849_1 | MZ520832 |
| VC_849_2 | JQ015307,KY514263 |
| VC_849_3 | MF979560,MF979563 |
| VC_849_4 | MT552976,MT560059 |
| VC_849_5 | JQ691612,KC954774,KT353109 |
| VC_849_6 | JQ691611,MN013077,OL744211,OL744214,OL744216,OL744219,OL744220 |
| VC_849_7 | ON712643 |
| VC_849_8 | MW367417 |
| VC_849_9 | MW250786 |
| VC_85_0 | KJ010547,KJ010548,KM051843 |
| VC_85_1 | MW419084,MW419085,MW419086,MW419087 |
| VC_85_10 | MN043729 |
| VC_85_11 | MN940411 |
| VC_85_12 | MT151604 |
| VC_85_13 | KU640380 |
| VC_85_2 | KC699836,MW749002,MW749007 |
| VC_85_3 | AB930182,AP013029,KF669652,LT960608,MF422185,MG000860,MW354667,MZ501260,MZ501261,MZ501262,MZ501263,MZ501265,OM982668,OM982669,OM982670,OM982671,OM982672,OM982673,OM982674 |
| VC_85_4 | KY368640,MN043730,OK349510 |
| VC_85_5 | MH606185 |
| VC_85_6 | JX238501 |
| VC_85_7 | KM236246 |
| VC_85_8 | KU253712 |
| VC_85_9 | KM236245 |
| VC_850_0 | OK136181,OK181860,OK181862 |
| VC_851_0 | MH444512,MK932884 |
| VC_853_0 | MG941013,MW073100 |
| VC_855_0 | GQ241246,KF669650 |
| VC_856_0 | MH371108,MH371109,MH536823,MZ355724,MZ456035,ON260827,ON970603 |
| VC_856_1 | KJ173786,MH271300,MH271307,MH271310,MH271316,MH399779,MK376963,MK919474,MK937596,MN096374,MN908690,MZ622166,OP068333 |
| VC_857_0 | KU160638,KU160639,KU160647,KU160663,KU160666,MF185731,MN234164,MN234217,MN234224,MT310874,MT986028 |
| VC_859_0 | MH019215,MH019216,MK305891,MT498037,MT498053,MT897908,MT936332,MZ648033,MZ648036 |
| VC_860_0 | MN023176,MN023177,MN023178,MN023179,MN023180,MN023181,MN023182,MN023183,MN023184,MN023185,MN023186,MN023187,MN023188,MN023189,MN023190,MN023191 |
| VC_861_0 | JQ659259,KC013021,KC013022,KC013023,KC013024,KC013025,KX555527,MN552146,OV696621 |
| VC_862_0 | GQ451696,KC013026,KC013027,KC013028,KC013029,KM262191,KM262192,KX578042,KX578043,KX578044,MN552147 |
| VC_864_0 | GQ919031,MN234216 |
| VC_865_0 | MK804893,MK813939 |
| VC_865_1 | MK813938 |
| VC_865_2 | MK813941 |
| VC_865_3 | OM033134,OM033135 |
| VC_865_4 | OM033136 |
| VC_866_0 | JN035618,KR063278,MW534374,ON526973 |

| | |
|---|---|
| VC_866_1 | KU998240,MG099935,MH025889,MK450428,MK450430,MK450435,MK450436,MK494106,MK494109,MK494121,MK878903,MK919477,MK919481,ON724015,ON970582,ON970588,ON970596,ON970607,OP297547,OP434450 |
| VC_866_2 | KR011062,KR011063,KR011064 |
| VC_867_0 | KP790008,KP790009,KP790010,KR063279,KX557281,ON526975 |
| VC_868_0 | KU963245,KU998241,MG099940,MG757152,MG757155,MG770211,MG770214,MK359302,MK433261,MK433263,MK433265,MK433267,MK801728,MK801733,MK967390,MN444874,MN945905,MT114160,MT889368,MT889370,MT889379,MT986029,MW507132,MW507133,ON456331 |
| VC_869_0 | KU963258,KU963262,KU998242,KU998243,MH576976,MK494098,MN234166,OM203156,OP434448 |
| VC_87_0 | FJ230960,KF669649,KY368639,MT601270,MT601271,MW001214,MZ751040,OM236513 |
| VC_87_1 | KC595512,KC595513,KF554508 |
| VC_87_2 | MH884511 |
| VC_870_0 | KU998252,KU998253,KU998254,MH479923,MN096363 |
| VC_872_0 | KX670789,KX670790,MF467948,MF467949,MG757154,MH155877,MH155880,MH669006,MK433259,MK433260,MK433268,MK433269,MK967386,MN204492,MN204494,MN204495,MN204496,MN204497,MN204500,MN204504,MN586054,MN586056,MT310852,MT310854,MT657340,MT684589,MT684600,MW055905,MW145539,MW712736,ON108646 |
| VC_873_0 | GU936715,HM480106,MG271909 |
| VC_874_0 | KR935216,KR935217,KT253150 |
| VC_875_0 | GU942563,HQ224500,KJ619459,KP972568,KP972569,KR063267,KR063268,KT728930,KT728931,MF155889 |
| VC_877_0 | MN830254,MN830255 |
| VC_878_0 | HE956707,HE956710,JN651747,KC107834,KT184661,KT949345,KX756572,KY318515,LN878149,MF683623,MG878892,MH179473,MH992513,MK053931,MK330684,MK838107,MK838108,MK838109,MK838110,MK838111,MN509793,MT682386,MT682387,MT682388,MT682389,MT682390,MT682391,MW250276,MZ592920,OL362270,OM654404,ON287377 |
| VC_879_0 | HF569088,HF569089,HF569090,HF569091,HF569092,HG428758,JN939331,JN939332,KC556893,KC556894,KC556895,KC556896,KC556897,KC556898,KJ133688,KJ133689,KJ133690,KJ133691,KJ133692,KJ133693,KJ133694,KJ133695,KJ133696,KJ133697,KJ133698,KJ133699,KJ133700,KJ133701,KJ133702,KJ133703,KJ133704,KJ133705,KJ133706,KJ133707,KY056619,MT002975 |
| VC_88_0 | KT624200 |
| VC_88_1 | MF418016 |
| VC_880_0 | HG007973,HM150760 |
| VC_881_0 | KP282679,KP282680 |
| VC_882_0 | HG992588,JQ692107,JX556418,MF754113,MF754114,MF754115,MF754116,MG602476,MG893203,MH599087,MH925090,MN215888,MW451246,MW451251,MZ592922,OK483201,OK625546,OL314400,OP547477,OP595601 |
| VC_883_0 | JF966203,KJ489397,KJ489398,KJ489400,KU737346,MF288921,MF288922,MF498901,MH638310,MK215646 |
| VC_883_1 | KF208639,KF669647,KJ489402,KT307976,KU737345,KU737347,MF765814,MN038178,MW281503,OK499976,OK499990,OM350011 |
| VC_883_2 | JN790865,JN797796 |
| VC_883_3 | KF669662,KY888882 |
| VC_883_4 | MN153503 |
| VC_883_5 | MW787012 |
| VC_883_6 | OM743306 |
| VC_884_0 | KJ489399,KJ489401,KT207918,KU737344,KU737348,KU737349,KU737350,KU737351,KU737352,KX147229,KX156152,KX349902,KX349903,KX961629,KX961632,MF288917,MF288918,MF288919,MF288920,MG763894,MH538193,MH638311,MN038176,MN038179,MT584805,OK500001,OM350012,OM350013 |
| VC_884_1 | HM144387,JN654439,KC430106,KU847400,KX011169,MW392803,MW584228,MW596414 |
| VC_885_0 | MW677509,MW677510,MW677512,MW677515,MW677517,MW677520,OP009272,OP009275,OP009278,OP009280,OP009286 |
| VC_886_0 | MN988459,MN988558 |
| VC_886_1 | MW677529 |
| VC_887_0 | HM152763,JF704108,JF704113,JN600672,MF185726,MH744425,MK279905,MK310144,MK494092,MK494100,MN096382,MN586018,MN586023,MN617841,MT771349,MW435855,MW822149,MZ958746 |
| VC_888_0 | JF744988,JN680858,KC661276,KC691254,KF024731,KT591489,KU234099,KU997639,KX507361,KX580961,KX580962,MF140422,MF185728,MF185730,MF324910,MH779511,MH834600,MK937600,MN096380,MN234228,MN586030,MN586052,MN703406,MN703410,MT758688,MW578835,MZ171373,OL455899 |
| VC_888_1 | MH576973,MN234187,MZ005679 |
| VC_889_0 | KF024725,KT281791,KT281794,KT372003,MF140405,MF140413,MF140414,MF140421,MH727559,MK878900,MN096377,MT889383,MT889391,MW291024,MZ622183,ON755181 |
| VC_89_0 | MT601273,MT601274 |
| VC_89_1 | MT601272,OL580764 |
| VC_89_2 | KY030782,MT366945,MT366946,MT366947,MT366948 |
| VC_89_3 | AF020713 |
| VC_89_4 | ON107264 |
| VC_89_5 | MZ969646 |
| VC_89_6 | OM236515 |
| VC_890_0 | MH651182,MK967378,MT658805 |
| VC_890_1 | MH976515,MN444870,MT684599,OK040786 |
| VC_891_0 | HQ268735,KY629621 |
| VC_894_0 | HM588722,KF302032,KF302033 |
| VC_895_0 | KF302036,KX365748 |
| VC_896_0 | KR060090,KX912252,KY379511,MF370965,MT002875 |
| VC_897_0 | HM640230,JN800508,KY996523,MH999279,MW815121,OP022426 |
| VC_898_0 | JQ729990,JQ729991,JQ729992,MH248069,MW057920,MZ401005,MZ401007,MZ401008,MZ401009,OL547738 |
| VC_899_0 | MH393889,MK017820,OM638104,OP256049 |
| VC_9_0 | AB370205,AF424781,AP008953,AY954951,AY954952,AY954953,CP062427,CP062451,CP062454,DQ517338,DQ530359,DQ530360,EU861004,GU477322,HQ127381,JQ779024,JX274646,JX274647,KC677663,KJ716334,MG557619,MH321492,MK658834,MK801682,MK801683,MT013111,MW554488,OM439662,OM439666,OM439668,OM439670,OM439671,OM439675,ON911714,ON911715,ON911716,ON911717,ON911718 |
| VC_90_0 | LC597490 |
| VC_90_1 | OM634661 |
| VC_90_2 | EF583821 |
| VC_901_0 | KF623293,MW824400,MW824417,MW824421,MW824431 |
| VC_902_0 | HQ316582,MG592662,MG592672 |
| VC_903_0 | MW824414,MW824420,MW824427,MW824428,MW824429,MW865319,OX241434 |
| VC_904_0 | JF713456,MG592585,MG592586,MG720309,MK559459,MT735629 |
| VC_904_1 | HQ316604,KX198615,MT451873,ON015836 |
| VC_904_2 | KY022433,MH925091,MN871506,MW411580,OK483199,OP547478,OV032860 |
| VC_905_0 | MG189906,MT110073,MZ326862 |
| VC_907_0 | HQ317384,HQ332142,KM233261 |
| VC_908_0 | MN988533,MN988538,MW980068 |
| VC_909_0 | HQ317386,MG592522,MG640035,MT720689,OM837731 |
| VC_91_0 | AB609718,AP009390,AP018714,AP018715,CAJDJY010000002,CAJDJZ010000002,KJ801817,KR049063,LC596377,LC596378,LC596379,LR031359,MK268686,MK693030,MK721192,MN854830,MW633168,MZ420150,ON113167,ON113168,ON113169,ON286976 |
| VC_91_1 | OP559178 |
| VC_910_0 | HQ317390,MZ568826 |
| VC_911_0 | HQ317392,KC821610,KC821616,KC821629,KC821630,KC821634 |
| VC_912_0 | KM873719,KY951963,KY951964,KY979235,KY979236,KY979237,KY979238,KY979239,KY979240,KY979241,KY979242,KY979243,KY979244,KY979245,KY979246,KY979247,KY992519,KY992520,MK756083,MK756084,MK756085,MK756086,MK756087,MK756088,MK756089,MK756090,MK756091,MK756092,MK756093,MK756094,MK756095,MK756096,MT431535,MT585273,MT585274,MT585275,MT585276,MT585277,MT585278,MT585279,MT585280,MT585281,MT585282,MT585283,MT585284,MT585285,MT585286,MT585287,MT585288,MT585289,MT585290,MT585291,MT585292,MT585293,MT585294,MT585295,MT585296,MT585297,MT585298,MT585299,MT585300,MT585301,MT585302,MT585303,MT585304,MT585305,MT585306,MT585307,MT585308,MT585309,MT585310,MT585311,MT585312,MT585313,MT585314,MT585315 |
| VC_913_0 | HQ332136,MW452941 |
| VC_916_0 | HQ396194,MZ995504,ON922917,ON933637 |
| VC_919_0 | HQ403646,KU963260,KU998239,MG770215,MH271302,MH450132,MK279900,MK875791,MK875795,MK967383,MK967385,MN062716,MN234215,MN284906,MN310549,MN444871,MN444878,MN586015,MN586036,MN586049,MN586057,MT776808,MT776809,MT776810,MT889367,MT889378,MW507135,MW862988,MZ150793,MZ322013,MZ622167,OK040785,OL455888,OL829977,ON970576,ON970599,ON970609,ON970621,OP297531 |
| VC_92_0 | CAJDKA010000002,KP339049,KY549443,LR990835,MH791397,MH880817,MN027503,MN241318,MT856905,MT909815,MW004544,MW004545,MZ147816,MZ333462,OL799258,OL799260,OL870611,OM966901,ON113170,ON125307,ON506927,ON506928 |
| VC_920_0 | MN204493,MW822144 |
| VC_921_0 | HQ632857,HQ632858 |
| VC_922_0 | MK291441,MK291443 |
| VC_923_0 | MW677513,MW677516,MW677518,MW677519,MW677521,MW677522,MW677523,MW677524,OP009265,OP009266,OP009267,OP009268,OP009269,OP009270,OP009271,OP009273,OP009276,OP009277,OP009279,OP009283,OP009284,OP009287,OP009288 |
| VC_924_0 | MG592442,MG592450,MG592457,MG592460,MG592482,MG592483,MG592488,MG592489,MG592490,MG592502,MG592503,MG592504,MG592505,MG592516,MG592517,MG592525,MG592530,MG592532,MG592534,MG592535,MG592539,MG592543,MG592557,MG592558,MG592559,MG592576,MG592593,MG592594,MG592597,MG592598,MG592605,MG592606,MG592607,MG592631 |
| VC_925_0 | MN592896 |
| VC_926_0 | MG592520,OX241566,OX241570 |
| VC_927_0 | HQ698895,KJ410740 |
| VC_928_0 | JF974286,JF974321,MT490303 |
| VC_929_0 | HQ634192,KC821604,KC821621,KC821625 |
| VC_93_0 | FJ822135,MT758689 |
| VC_930_0 | KX889068,MG592409,MG592410,MG592470,MG592519,MG592521,MG592668 |
| VC_932_0 | HQ683709,MG209611 |
| VC_933_0 | KR093625,KR093626,KR093627,KR093628,KR093629,KR093630,KR093631,KR093633 |
| VC_934_0 | MT119766,MT210154 |
| VC_935_0 | MN988536,OP028995 |
| VC_936_0 | MN602266,MN602267 |
| VC_937_0 | JN672684,KR269719,KX237515,KY780482,MN013075,MN013076,MN013078,MN013079,MN013080,MN013083,MN013087,MT701592,MW021762,MW021764,MW021765,MW042802,MW417503,MW722081,MZ428221,MZ428224,MZ598515,MZ634348,MZ634350,MZ634351,OL539450,OL539472,OL865411,OM065837,OM065838,OM065839,OM065841,OM328157,OM735690,OP611406,OV877339,OV877387,OV877766,OX335361,OX335395,OX335410 |
| VC_938_0 | JF501022,KP658157,KR262148,KT001920,KY575286,LR757892,MF415412,MF612072,MG552615,MG894052,MH445453,MH633484,MH633485,MH633486,MH633487,MH688453,MH844531,MK170446,MK278861,MK521904,MK618657,MK931442,MK931443,MK931444,MK931445,MN379832,MN395285,MN395286,MN689779,MN783016,MT894005,MW655991,MW672037,MZ172460,MZ221764,MZ398242,MZ428222,MZ428223,MZ428225,MZ428226,MZ428227,MZ428228,MZ571831,MZ571833,MZ571834,MZ634345,MZ634346,OK138555,OK655936,OL539448,OL539460,OL674249,OL702938,OL744210,OL744213,OM032871,OM065840,OM065842,OM065843,OM687893,OM938991,OM938992,OM964875,ON489264,ON602735,ON602741,ON602750,ON881905,OP267563,OP413832,OP617742,OU342755,OW251746,OX335369,OX335370,OX335371,OX335380,OX335381,OX335383,OX335384,OX335385,OX335386,OX335387,OX335388,OX335391,OX335393,OX335400,OX335404,OX335411,OX335412,OX335417,OX335421,OX335422,OX335423,OX335429,OX335430,OX335432,OX335435,OX335439 |
| VC_939_0 | JF704100,KJ538721,KX159477,MG757167,MH727553,MH727562,MK061407,MK279842,MK305892,MK494123,MN329674,MN329676,MN428056,MN428061,ON637772 |
| VC_94_0 | HM242243,OM135992 |
| VC_940_0 | JF704107,JN083852,JN083853,JN408459,JQ684677,JX042578,KF017001,KF493882,KF560330,KM101118,KT004677,KT246486,KT438501,MG099934,MG099937,MG099938,MG099944,MG099949,MH020239,MH051250,MH051254,MH051256,MH338235,MH338241,MK305887,MK494091,MN096372,MN586016,MN586060,MN586062,MN617844,OL455889,ON970625,ON970626 |
| VC_941_0 | KC710998,MG065636,MG065640,MG065641,MG065643,MG065649,MG065657,MG065663,MG065665,MG065667,MG065669,MG065670,MG065671,MG065673,MG065675,MG065679,MG065680,MG065681,MG065687,MK373798,MN055691,MN432485,MN850567,MN850569,MN850572,MN850591,MN850600,MN850602,MN850607,MN850640,MT074433,MT127619,MT360681,MZ501079,MZ501090,OK272479,OK272487,OL539459,ON185589,ON528718 |
| VC_942_0 | LR027385,LR027388,LR596614,LT841304,LT961732,MF564201,MK962751,MN781674,MN850582,MN850586,MN850596,MN850604,MN850606,MN850613,MN850620,MN850634,MN850638,MN850641,MN850643,MZ234033,MZ501069,MZ501101,OK272471,OK272478,OK272481,OK272483,OK272485,OK272486,OK272489,OL539445,OL539446,ON185586,ON185587,ON185588,ON711395,OU734268 |
| VC_943_0 | JF917095,JQ010983 |
| VC_944_0 | MG592398,MG592496,MG592497,MG592584,MG592612,MG592613 |
| VC_946_0 | JX483876,JX483877,JX483878,MN988482,MN988500,MN988501,MN988508,MN988530,MN988545,MN988549,MW980073 |
| VC_948_0 | MG592429,MG592438,MG592487,MG592566,MG592654,MG592657,MG592658,MG592659,MZ369152 |
| VC_949_0 | JN116822,KR053197,KR053198,KU998255,KU998256,MG812496,MH271296,MK660712,MK801723,MN234213 |
| VC_95_0 | MK504443,MK504446,OM293948 |
| VC_950_0 | KT355471,KT355473,KT355475,KT783672,KU160655,KU160657,KU160658,KU160667,KU160670,KU160674,KX443695,KX576642,KX610765,KY434670,MF140409,MF140415,MF140417,MF140419,MF140420,MF140426,MF140429,MF140431,MF140433,MH576948,MH576950,MH576951,MH576952,MH576955,MH576957,MH576961,MH779506,MK494103,MK494111,MK737943,MK820640,MN617842,MN813695,ON970580 |
| VC_951_0 | MH834594,MH834597,MH834628 |
| VC_952_0 | JN116828,MW654182 |
| VC_953_0 | JN116823,MK967393 |
| VC_954_0 | KU963261,MH399781,MH509446,MK279893,MK305885,MK359319,MK376958,MK967392,MW291015,MW534376,MZ005677,ON637762,ON637763 |
| VC_955_0 | MZ322021,ON970578,OP297535 |
| VC_956_0 | JN116826,JN116827,KT372002,KT375356,KT959213,KT990218,KX550082,KX611788,KX712236,KX712237,KY549153,KY549154,MF189179,MF324898,MF324900,MF324901,MF324902,MF324903,MF324904,MF324905,MF373842,MF537628,MH271291,MH271293,MH271294,MH271297,MH271299,MH271306,MH271309,MH271311,MH271314,MH271315,MH271319,MH316569,MK524487,MK524495,MN945902,MW291027,MZ402608 |
| VC_957_0 | JN175269 |
| VC_957_1 | KU862660 |
| VC_958_0 | MW570842,MW584170,MW584191 |
| VC_959_0 | JX094500,KX397367,KX397372,MK514282,MW845759 |
| VC_959_1 | KX397368,KX397370,KX397371,MT710307,MW384823,MW812340,MW812342,MW812343 |
| VC_959_2 | KX397365,KX397366,MW760841,MW812338,MW812341 |
| VC_959_3 | KX397369,MH426724,MW845758 |
| VC_959_4 | JX316028,KX397364,KX397373 |
| VC_959_5 | JN641803,KU726251,LR535902,LR535909,LR535910,LR535914,LR535915,LR535916,LR535918,LR535919,LR535920,LR535921,MH221129,MN445185,MT679221,MT799840,MW812339,OK428535,OM456492 |
| VC_959_6 | MH221128 |
| VC_96_0 | AB626962,AB626963,AF513032,AF513033,AY954949,EU136189,HF937074,KJ210330,KM366100,KU992911,KY000084,KY000085,LC709172,MG766218,MG766219,MH107118,MH972260,MK764384,MK903033,MK922546,MK922547,MK922548,MK936475,MK936476,MN098325,MN150710,MT926124,MW286254,MW363799,MZ359091 |
| VC_960_0 | JN712910 |
| VC_960_1 | KX925554 |
| VC_961_0 | KX344510,KY264020,MW344767 |
| VC_962_0 | JN882267,OM621814 |
| VC_963_0 | KY965934,MG550110,MG550111,MG550113 |
| VC_965_0 | MG596800,OM982619 |
| VC_966_0 | MK608336,OK499995,OL539452 |
| VC_967_0 | JQ309827,KT932700,LT630001,MK721183,MK721184,MK721185,MK721188,MK721193,MK721195,MK721196,MK721197,MK721198,MT747434,MT757399,MW623430,ON352054 |

| | |
|---|---|
| VC_968_0 | KT945995,MK360025,OL799259,ON113181 |
| VC_969_0 | JQ312117,MK637516,MT234341 |
| VC_97_0 | AY212251,KJ528544,LC628082,LC644557,ON637248 |
| VC_970_0 | MN988470,MN988484,MN988495,MN988531,MN988537,MN988544,MN988546,MN988557,MW980069 |
| VC_971_0 | KP280063,KY883654,KY883655 |
| VC_971_1 | JQ340389,LC487410,MG649966,MG649967,MK796244,MK895508,MK907780,MN954399,MN958086,MN966730,MN966731,MN966732,MT448616,MT448617,MT459144,MT460514,MT460515,MT460516,MT460517,MT460518,MW344054,MW344055,MW344056,MW398869 |
| VC_971_2 | KT345706 |
| VC_972_0 | MH645904,MH925094,MN871505,MN871510,MN871511,MT442039 |
| VC_973_0 | MW824415,MW824416,MW824419,MW824422,MW824423,MW824424,MW824425,MW824426 |
| VC_974_0 | JQ823122,JQ823123 |
| VC_975_0 | KP876465,KP876466,KT989433,KU189325,KU530220,KY365739,MF975638,MH178381,MH178382,MK686071,ON526979 |
| VC_976_0 | ON529850,ON529857 |
| VC_976_1 | ON529851,ON529852,ON529853,ON529855,ON529860 |
| VC_976_2 | JX100810,MH588545 |
| VC_976_3 | JX100814,MH588544 |
| VC_976_4 | JX100809,JX100811,JX100812,JX100813,JX163858,KY555142,KY555143,KY555144,KY555145,KY555146,KY555147 |
| VC_976_5 | ON529858 |
| VC_977_0 | KT870145,MH015255 |
| VC_977_1 | MN602268,MT270409 |
| VC_977_2 | OP186293 |
| VC_978_0 | KY629563,OL955261 |
| VC_978_1 | OL963731 |
| VC_980_0 | JX126919,JX126920,JX442241,KJ094020,KJ094021,MH299806,MN114082,MN114083,MN128593,MN904502 |
| VC_981_0 | JX182369,ON526970,ON526974 |
| VC_981_1 | JX182370,JX262376,MF766044,MF766045,MF766046,MF766047,MF766048,MG593802,MH001460,MH590601,MH651190,MH669004,MH669016,MK392363,MK392364,MK392365,MK392366,MK460245,MK524524,MN234192,MN234208,MT310863,MT701594,MT701597,MT708548,OK412919 |
| VC_982_0 | JX889246,KC700556,KC700557,KC700558,KT124227,KT124228,KT124229,KT152029,KT184390,KT184391,KT186228,KT186229,KX344445,KX507343,KX507344,KX507345,KX815338,MF541403,MF541404,MF541405,MF541406,MF541407,MF541408,MF541409,MF541410,MG298964,MG757163,MH17109<br>3,MH171094,MH171095,MH171096,MH171097,MH171098,MH248947,MK433270,MK433271,MK433276,MK433278,MN096373,MN096375,MN096379,MN096381,MN204498,MN284899,MN586009,MT701598,MT711978,MT711979,MW365951,MW365953,MW435856,MZ648032,MZ648039,MZ648040,MZ820<br>088,MZ820091,MZ820092,OL455891,OL455894,OL742563,ON208832,ON526985 |
| VC_984_0 | JX290549,KR604693,KT240186,KX278418,KX278419,KY953156,LC063634,LC375533,MH807811,MH807814,MH807816,MH807818,MH807819,MK095192,MK095193,MK095194,MK095195,MK095196,MK095197,MK095198,MK095199,MK095200,MK095201,MK095202,MK095203,MK095204,MK095205,M<br>K095206,MK095207,MK095208,MK095209,MK095210,MK095211,MN270887,MN270888,MN271656,MN812690,MN812692 |
| VC_987_0 | JX483874,JX483875,JX483879,JX483880,MF403005,MF403006,MN988466,MN988488,MN988492,MN988493,MN988512,MN988513,MN988514,MN988528,MN988529,MT708544,MT708546,MW960034,MW980062,MW980065,MW980074,MZ462995 |
| VC_988_0 | MF278336,MW043865 |
| VC_989_0 | KY065149,MT227925 |
| VC_99_0 | AB647160,AB731695,JF734911,JQ617284,KX119174,KX119175,KX119176,KX119177,KX119182,KX119188,KX119189,KX119190,KX119191,KX119192,KX119193,KX119194,KX119195,KX119196,KX119197,KX119198,KX119199,KX119200,KX119201,KX119202,KX119203,KX119204,KX119205,KX119206<br>,MW247144,MW247145,MW247147,OM515228 |
| VC_990_0 | KF614509,MN988552,MT778840 |
| VC_991_0 | KC292024,KC292028 |
| VC_992_0 | KP087954,MH572347,MH572348,MH572349,MH572405,MH572418,MH572455,MH572515 |
| VC_994_0 | MK249135 |
| VC_995_0 | KC131022,KM589502,KM589511,KT264813,KT264814,KT264815,MG945202,MG945465,MG945477,MG945663,MG945838,MG945843,MH572283,MH572284,MH572285,MH572286,MH572291,MH572292,MH572293,MH572382,MH572383,MH572423,MH572424,MH572427,MH572428,MH572466,MH572495,<br>MH572504,MH992156,MH992161,MH992184,MH992185,MH992213,MK249141,MK249147,MK249165,MK249185,MK249190,MK249197,MK249201,MK249202,MK249203,MK249211,MK249213,MK249221,MK496825,MN862333,MN862335,MN862341,MW202412,MW202414,MW202466,MW202471,MW20<br>2549,MW202587,MW202847,MW202853,MW202883,MZ364228,MZ364233,MZ364240,MZ364243,MZ364256,MZ364258,MZ364259,MZ364267,MZ364274,OL617029,OM523002 |
| VC_996_0 | MN862339,MN862340 |
| VC_997_0 | MH992169 |
| VC_998_0 | MH572299,MH572425 |
| VC_999_0 | MH572426,MK249189 |

| VC_ME-vOTUs_ref | Members |
| --- | --- |
| VC_1009_0 | ON721384,ON721385,S7-wms_244866 |
| VC_1010_0 | MN732896,S11-wms_330479 |
| VC_1109_0 | KR816341,S7-wms-combined_235165 |
| VC_1243_0 | MG883709,MG945260,MG945282,MG945390,MG945396,MG945452,MG945485,MG945511,MG945515,MG945618,MG945634,MG945641,MG945647,MG945743,MH572525,MH572526,MK496694,MK496775,MK765561,MK765563,MK765564,MK765568,MK765569,MK765594,MK765638,MN311493,MT135347,MT135352,MT135355,MT135366,MT135381,MT135382,MT135387,MT135444,MW202703,MW202824,MZ089786,MZ089791,MZ089795,MZ089806,OL617028,OM869549,OM869559,OM869560,OM869562,OM869569,OM869650,OP549868,OP549871,OP549874,S1-wms-combined_357373,S9-wms_128232 |
| VC_1276_0 | MG945488,MG945572,MZ089802,MZ089808,MZ089809,OP549867,S17-wms_254872 |
| VC_1423_0 | MT135285,MT135286,MT135289,MT135297,MT135302,MT135308,MT135314,S9-wms_104169 |
| VC_181_0 | JF832915,KP137440,MK388859,S1-wms-combined_19031,S11-wms-combined_268967 |
| VC_19_0 | CP000315,S5-wms-combined_83412 |
| VC_250_0 | AY539836,CP000622,ON170188,S9-wms-combined_64277 |
| VC_255_0 | FJ965538,KX879643,KY705252,KY705253,KY705254,KY705255,MH892355,MH892367,MH892373,MH973663,MK202159,MK202160,MK202161 |
| VC_255_1 | S9-wms-combined_20485 |
| VC_427_0 | JF974295,MW202455,S1-wms-combined_324053 |
| VC_540_0 | KC139518,KT287080,MF158036,MF158037,MH424444,MK214385,MK479295,MN070121,MN393079,MT012729,MW725301,MZ004649,MZ501269,ON720976,S17-wms_266511 |
| VC_822_0 | LT600745,S7-wms-combined_225260 |

| VC_ME-vOTUs_only | Members |
| --- | --- |
| VC_1491_0 | S1-wms-combined_185336,S1-wms-combined_40115 |
| VC_1499_0 | S5-wms-combined_66017,S5-wms_210460 |
| VC_798_0 | S1-wms-combined_307323,S7-wms-combined_147158 |
| VC_798_1 | S1-wms-combined_85225 |
| VC_798_2 | S17-wms-combined_186314 |
| VC_914_0 | S5-wms_67169 |

| VC_SHHC_vOTUs_ref | Members |
|---|---|
| VC_1008_0 | MT635598,MT806185,MT806186,S4-wms_141607 |
| VC_1420_0 | MT121960,MT121964,S12-wms_401372,S2-wms-combined_197577,S2-wms_128831 |
| VC_1422_0 | MT135284,S2-wms-combined_132966 |
| VC_744_0 | DQ123818,MK016490,MK016500,MN988551,MZ333134,MZ333136,ON381767,ON724010,OV696617,S2-wms_79304 |

| VC_SHHC-vOTUs_only | Members |
|---|---|
| VC_1076_0 | S12-wms-combined_376326,S8-wms-combined_72911 |
| VC_1238_0 | S12-wms-combined_147628,S12-wms-combined_41160 |
| VC_1489_0 | S12-wms-combined_157167 |
| VC_1501_0 | S8-wms-combined_276204,S8-wms_57121 |
| VC_1511_0 | S12-wms_15700,S8-wms-combined_226076 |
| VC_674_0 | S8-wms_111056 |

| VC_shared_vOTUs_ref | Members |
| --- | --- |
| VC_1235_0 | MG711460,S1-wms_4277,S11-wms-combined_283220,S11-wms_3116,S12-wms-combined_299872,S12-wms_193565,S17-wms-combined_175924,S17-wms-combined_193958,S2-wms-combined_165833,S7-wms_113137,S7-wms_200936,S8-wms_30498 |
| VC_1235_1 | S2-wms-combined_157934 |
| VC_1477_0 | OP549880,S4-wms_161443,S7-wms_157264 |
| VC_417_0 | KC237729,MW478289,MZ503613,S2-wms_130687,S5-wms_49310 |
| VC_418_0 | LR596902,S1-wms-combined_224578,S2-wms-combined_246480,S2-wms_140933,S4-wms-combined_64632,S4-wms_62635 |
| VC_5_0 | AB374228,AF125163,AY027935,BHU24159,DQ114220,DQ674738,HG803181,HQ110083,KF361475,KT887559,KU504502,KX058879,KX774374,KY045851,LR597637,LR597638,LR597640,LR597649,MG592573,MK972712,MK972713,MK972714,MN563793,MT662115,OM095401,OP484843,S1-wms-combined_159544,S11-wms-combined_154845,S5-wms-combined_18356,S6-wms-combined_119017 |
| VC_606_0 | MG711465,S11-wms_55610,S2-wms_107329,S8-wms_82880 |
| VC_606_1 | S9-wms-combined_29611 |
| VC_621_0 | MG711464,S1-wms-combined_23248,S11-wms-combined_140009,S12-wms-combined_345948,S12-wms-combined_71235,S17-wms-combined_90378,S4-wms-combined_42647 |
| VC_626_0 | MF172979,MK448667,MK448700,MK448720,MK448839,MK448841,MK448843,MK448934,MK448997,MW387018,S1-wms-combined_332236,S11-wms-combined_194205,S12-wms-combined_368462,S12-wms-combined_7174,S12-wms-combined_99704,S17-wms-combined_9864,S2-wms-combined_174241,S2-wms-combined_244564,S5-wms-combined_206050,S5-wms-combined_29272,S8-wms-combined_108917,S8-wms-combined_186054,S9-wms-combined_25007 |
| VC_626_1 | S9-wms-combined_9931 |
| VC_626_2 | S12-wms-combined_98046 |
| VC_626_3 | S5-wms-combined_108794 |
| VC_677_0 | BK010646,S11-wms-combined_195425,S12-wms-combined_250551,S3-wms-combined_73262,S5-wms-combined_59064,S7-wms-combined_43950,S7-wms-combined_5625,S8-wms-combined_97848,S9-wms-combined_31456 |
| VC_707_0 | S1-wms-combined_132934,S11-wms-combined_304239,S12-wms-combined_17617,S12-wms-combined_18287,S17-wms-combined_199440,S6-wms-combined_8508 |
| VC_707_1 | MG711462,S12-wms-combined_129554,S17-wms-combined_127688,S2-wms-combined_180640,S2-wms-combined_85381,S6-wms-combined_117756,S7-wms_159432,S8-wms-combined_200168,S8-wms-combined_217866 |
| VC_707_2 | S12-wms-combined_129799 |
| VC_707_3 | S11-wms-combined_264050 |
| VC_710_0 | MG711466,ON470591,S1-wms_296850,S11-wms-combined_74983,S12-wms-combined_156234,S17-wms-combined_260343,S6-wms-combined_126236,S7-wms-combined_134458,S8-wms_152831,S8-wms_48935 |
| VC_710_1 | S2-wms-combined_137119 |
| VC_710_2 | S7-wms-combined_157260 |
| VC_710_3 | S6-wms-combined_118427 |
| VC_710_4 | S2-wms-combined_13246 |
| VC_711_0 | MG711467,S1-wms-combined_68998,S7-wms-combined_111004 |
| VC_711_1 | S12-wms-combined_323943 |
| VC_711_2 | S17-wms-combined_2234 |
| VC_711_3 | S2-wms-combined_140996 |
| VC_717_0 | LR596903,MG711461,S12-wms-combined_259783,S2-wms-combined_207832,S3-wms_22744,S5-wms-combined_134381,S7-wms_118607 |
| VC_717_1 | S17-wms-combined_248321 |
| VC_734_0 | HQ332138,KU517658,MF595878,S2-wms-combined_101826,S2-wms-combined_248329,S5-wms-combined_7292 |
| VC_804_0 | MG711463,S1-wms-combined_304377 |
| VC_804_1 | S17-wms-combined_259811,S17-wms-combined_47860 |
| VC_804_2 | S1-wms_212940,S12-wms-combined_107942,S2-wms-combined_62377,S2-wms_248315,S8-wms_127191,S9-wms-combined_26410 |
| VC_804_3 | S2-wms-combined_98888 |
| VC_804_4 | S5-wms-combined_72410 |
| VC_831_0 | MN929097,OK632025,S1-wms-combined_394574,S4-wms-combined_14411,S6-wms_54578 |
| VC_1198_0 | MT006237,S5-wms_94965,S7-wms-combined_189250 |
| VC_1219_0 | MG065691 |
| VC_1219_1 | S5-wms-combined_66662 |
| VC_1221_0 | MW660583 |
| VC_1221_1 | S7-wms-combined_62119 |
| VC_675_0 | MT774393,S7-wms_244425 |
| VC_502_0 | S2-wms_128962,S6-wms_138342 |
| VC_502_1 | MT774375,MT774376,MT774379,MT774380,MT774382,S4-wms_73990 |
| VC_502_2 | MT774401 |
| VC_502_3 | MT774399,MT774403 |
| VC_502_4 | MT774395,MT774397,MT774398 |
| VC_502_5 | MT774396,MT774405 |
| VC_502_6 | MT774404,MT774407 |
| VC_502_7 | MT774400 |
| VC_852_0 | GQ141189,S4-wms-combined_17284 |

| VC_shared_vOTUs_only | Members |
|---|---|
| VC_1209_0 | S1-wms-combined_41369,S1-wms_130214,S8-wms-combined_276211 |
| VC_1209_1 | S12-wms-combined_23517 |
| VC_1209_2 | S11-wms-combined_3507 |
| VC_1239_0 | S1-wms_185046,S11-wms_303398,S12-wms-combined_334806,S7-wms-combined_40779,S8-wms-combined_134098,S8-wms-combined_25607 |
| VC_1397_0 | S11-wms-combined_73204,S12-wms-combined_320727 |
| VC_1435_0 | S17-wms-combined_16309,S17-wms_124308,S2-wms_156439,S7-wms-combined_185016,S7-wms-combined_90393,S7-wms-combined_90728,S7-wms_123954,S9-wms-combined_33229,S9-wms-combined_33230 |
| VC_1435_1 | S6-wms-combined_82597 |
| VC_1436_0 | S12-wms-combined_351847,S12-wms_383851,S2-wms-combined_58121,S7-wms-combined_239784 |
| VC_1456_0 | S11-wms-combined_125176,S12-wms_386692,S6-wms-combined_91039,S7-wms-combined_14267 |
| VC_1473_0 | S1-wms_244836,S2-wms-combined_229529,S9-wms-combined_37587 |
| VC_1479_0 | S12-wms-combined_51076,S5-wms-combined_78434 |
| VC_1481_0 | S1-wms-combined_120533,S1-wms-combined_331896,S12-wms-combined_393703,S17-wms-combined_126600,S2-wms-combined_181095,S5-wms_52566,S8-wms-combined_234473,S8-wms-combined_264711,S9-wms-combined_56773 |
| VC_1482_0 | S17-wms-combined_30308,S4-wms-combined_143879 |
| VC_1483_0 | S7-wms-combined_169375 |
| VC_1483_1 | S8-wms-combined_40666 |
| VC_1484_0 | S1-wms-combined_151708,S1-wms-combined_164485,S12-wms-combined_329242,S17-wms-combined_126774,S17-wms-combined_146850,S17-wms-combined_157468,S2-wms-combined_4685,S2-wms-combined_6605,S6-wms-combined_41246,S7-wms-combined_84688 |
| VC_1487_0 | S1-wms-combined_156637,S12-wms-combined_210942 |
| VC_1492_0 | S1-wms_100010,S1-wms_225995,S12-wms-combined_264425,S12-wms-combined_299293,S12-wms_179388,S12-wms_401366,S12-wms_44817 |
| VC_1495_0 | S17-wms-combined_233761,S2-wms-combined_210822,S4-wms-combined_47565 |
| VC_1496_0 | S1-wms-combined_313851,S1-wms_313991,S2-wms_110761,S2-wms_23562,S6-wms_137591 |
| VC_1497_0 | S1-wms-combined_331544,S4-wms-combined_51079,S7-wms-combined_136642 |
| VC_1498_0 | S1-wms-combined_354899,S2-wms-combined_248318 |
| VC_1500_0 | S1-wms-combined_394628,S12-wms-combined_401379 |
| VC_1502_0 | S11-wms-combined_17029,S12-wms-combined_375343 |
| VC_1503_0 | S11-wms-combined_217790,S12-wms-combined_111524,S5-wms-combined_98202 |
| VC_1504_0 | S11-wms-combined_221310,S8-wms-combined_215102 |
| VC_1505_0 | S11-wms-combined_41367,S12-wms-combined_256299,S17-wms-combined_158354,S5-wms_85650 |
| VC_1506_0 | S5-wms-combined_178432,S6-wms-combined_72228 |
| VC_1508_0 | S17-wms-combined_203701,S4-wms-combined_57123,S6-wms-combined_135223 |
| VC_1510_0 | S12-wms-combined_401383,S9-wms-combined_3878 |
| VC_1513_0 | S17-wms-combined_225356,S4-wms_107182 |
| VC_212_0 | S12-wms-combined_263377,S6-wms_132588 |
| VC_212_1 | S2-wms-combined_31084,S7-wms-combined_7205 |
| VC_271_0 | S11-wms-combined_58538,S11-wms_14390,S2-wms-combined_93845,S8-wms-combined_146562,S8-wms-combined_68780 |
| VC_271_1 | S8-wms-combined_225114 |
| VC_271_2 | S7-wms-combined_238231 |
| VC_338_0 | S12-wms-combined_120133,S17-wms-combined_130215,S2-wms-combined_55495,S6-wms-combined_118484,S9-wms-combined_36459 |
| VC_420_0 | S1-wms-combined_72167,S2-wms-combined_227800,S7-wms-combined_28138,S7-wms_127801,S8-wms-combined_140059,S9-wms-combined_71647 |
| VC_535_0 | S4-wms_118690,S9-wms_19025 |
| VC_610_0 | S1-wms-combined_107073,S2-wms-combined_89742,S4-wms-combined_41490,S8-wms-combined_26087 |
| VC_627_0 | S1-wms_170115,S5-wms-combined_73301,S6-wms-combined_113986 |
| VC_629_0 | S2-wms-combined_196783,S7-wms_80255 |
| VC_650_0 | S1-wms-combined_353757,S11-wms-combined_119215,S11-wms-combined_299954,S11-wms-combined_49632,S17-wms-combined_136622,S17-wms-combined_247494,S17-wms-combined_79709,S2-wms-combined_126134,S5-wms-combined_145067,S7-wms-combined_31068 |
| VC_676_0 | S12-wms-combined_2450,S5-wms-combined_29447 |
| VC_676_1 | S11-wms-combined_83571,S6-wms-combined_50721,S7-wms-combined_107679_0,S9-wms-combined_28805 |
| VC_715_0 | S1-wms-combined_302166,S11-wms-combined_198512,S12-wms-combined_358010 |
| VC_732_0 | S12-wms-combined_191570,S2-wms-combined_238929 |
| VC_732_1 | S17-wms-combined_95360 |
| VC_736_0 | S12-wms-combined_168259,S12-wms-combined_282594,S5-wms-combined_52741 |
| VC_821_0 | S2-wms_53881,S7-wms_239833,S7-wms_53338,S8-wms_244525 |
| VC_825_0 | S1-wms-combined_118167,S1-wms-combined_169879,S11-wms-combined_200084,S2-wms-combined_176376,S2-wms_79944,S6-wms-combined_140731,S9-wms-combined_105462 |
| VC_825_1 | S11-wms-combined_267993 |
| VC_825_2 | S5-wms-combined_134743 |
| VC_832_0 | S2-wms-combined_113619,S9-wms-combined_5526 |
| VC_832_1 | S7-wms-combined_3697 |
| VC_876_0 | S12-wms-combined_245755,S5-wms-combined_26971,S7-wms-combined_139683 |
| VC_876_1 | S2-wms-combined_116221 |
| VC_893_0 | S11-wms-combined_46816,S4-wms-combined_113721 |
| VC_915_0 | S12-wms-combined_321174,S3-wms_23958,S8-wms-combined_169835 |
| VC_917_0 | S11-wms-combined_48246,S8-wms-combined_270362 |
| VC_917_1 | S5-wms-combined_90315 |
| VC_918_0 | S11-wms_320555,S12-wms_12658 |
| VC_918_1 | S7-wms-combined_26217 |
| VC_918_2 | S12-wms-combined_201125 |
| VC_947_0 | S2-wms-combined_135154,S9-wms-combined_53991,S9-wms-combined_89249 |
| VC_1014_0 | S11-wms-combined_92114,S17-wms-combined_199746,S7-wms-combined_163980 |
| VC_1408_0 | S1-wms-combined_197401,S17-wms-combined_162786 |
| VC_1408_1 | S1-wms-combined_220022 |
| VC_1437_0 | S1-wms-combined_263562,S5-wms-combined_207291,S7-wms-combined_104872 |
| VC_1494_0 | S1-wms-combined_248622,S17-wms-combined_70983 |
| VC_1509_0 | S17-wms-combined_179771,S7-wms-combined_72070 |
| VC_1512_0 | S17-wms-combined_223748,S9-wms_64472 |
| VC_1514_0 | S5-wms-combined_39793,S7-wms-combined_143218 |
| VC_616_0 | S11-wms-combined_193148,S11-wms-combined_204522 |
| VC_679_0 | S1-wms-combined_246923,S1-wms-combined_340558,S1-wms-combined_52643,S11-wms-combined_172889 |
| VC_680_0 | S5-wms-combined_83370,S9-wms-combined_44402 |
| VC_716_0 | S1-wms-combined_360285,S11-wms-combined_84326 |
| VC_719_0 | S1-wms_102342 |
| VC_719_1 | S7-wms-combined_20921 |
| VC_854_0 | S5-wms-combined_117213,S9-wms-combined_76274 |
| VC_1236_0 | S8-wms-combined_134284,S8-wms-combined_211119,S8-wms-combined_49369 |
| VC_1439_0 | S12-wms-combined_260036,S12-wms-combined_91231,S4-wms_45128 |
| VC_148_0 | S2-wms-combined_62519,S8-wms-combined_161181 |
| VC_148_1 | S2-wms-combined_218002,S6-wms-combined_119138,S6-wms-combined_32530 |
| VC_1485_0 | S2-wms-combined_233107 |
| VC_1507_0 | S12-wms-combined_264845,S12-wms_373775 |
| VC_324_0 | S2-wms-combined_166665,S6-wms-combined_49909 |
| VC_651_0 | S2-wms-combined_24549 |
| VC_678_0 | S12-wms-combined_358797 |
| VC_738_0 | S2-wms-combined_135139,S8-wms-combined_246185 |

After curation:

| **WMS-VCs:** | **Counts** |
|---|---|
| Total VCs | 1515 |
| VCs with "clustered" or "clustered/singleton" vOTUs and Ref genomes | 1301 |
| VCs having reference genomes only | 1181 |
| VCs having no reference genomes at all | 81 |
| VCs uniquely having ME-vOTUs with Ref. genomes | 13 |
| VCs uniquely having ME-vOTUs only (w/o Ref. genomes) | 4 |
| VCs uniquely having SHHC-vOTUs with Ref. genomes | 4 |
| VCs uniquely having SHHC-vOTUs only (w/o Ref. genomes) | 6 |
| VCs having shared vOTUs in both samples (with Ref. genomes) | 22 |
| VCs having shared vOTUs in both samples (w/o Ref. genomes) | 71 |

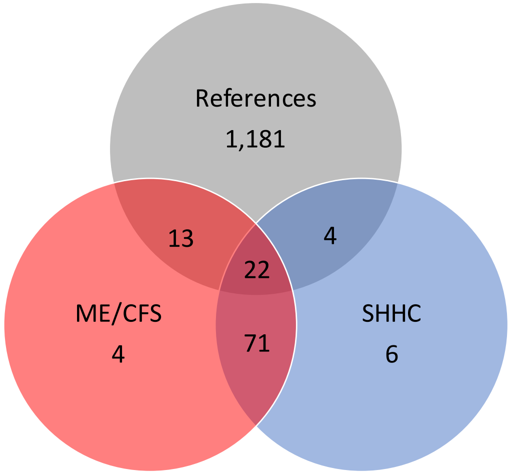


Before curation:

| **WMS-VCs:** | **Counts** |
|---|---|
| Total VCs | 1515 |
| VCs with "clustered" or "clustered/singleton" vOTUs and Ref genomes | 1301 |
| VCs having reference genomes only | 1181 |
| VCs having no reference genomes at all | 81 |
| VCs containing ME-vOTUs with Ref. genomes | 17 |
| VCs only having ME-vOTUs (w/o Ref. genomes) | 17 |
| VCs containing SHHC-vOTUs with Ref. genomes | 6 |
| VCs only having SHHC-vOTUs (w/o Ref. genomes) | 15 |
| VCs having both ME and SHHC-vOTUs with Ref. genomes | 16 |
| VCs having both ME and SHHC-vOTUs (w/o Ref. genomes) | 49 |

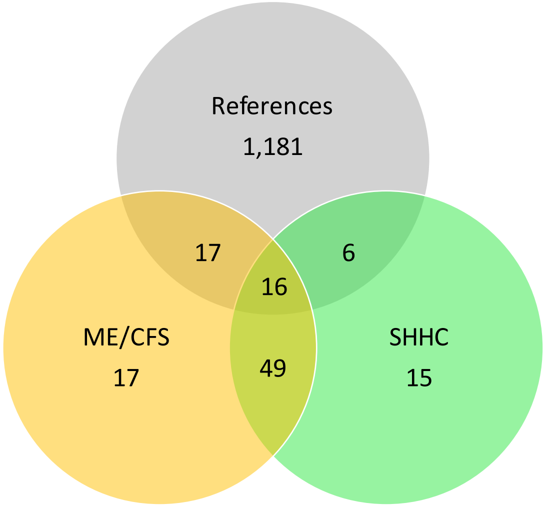
