## Supplementary material for "Comprehensive profiling of the human intestinal DNA virome and prediction of disease-associated bacterial hosts in severe Myalgic Encephalomyelitis/Chronic Fatigue Syndrome (ME/CFS)": S Table 11 Host predictions

| VLP_Virus | AAI to closest RaFAH reference | Host genus | Confidence score | List of methods |
|---|---|---|---|---|
| S10_vf_120738 | 22.73 | d__Bacteria;p__Firmicutes;c__Clostridia;o__Oscillospirales;f__Oscillospiraceae;g__Dysosmobacter | 95 | blast;96.50 iPHoP-RF;71.90 |
| S10_vf_120738 | 22.73 | d__Bacteria;p__Firmicutes;c__Clostridia;o__Oscillospirales;f__Oscillospiraceae;g__Intestinimonas | 93.5 | blast;95.40 iPHoP-RF;64.00 |
| S10_vf_18671 | 91.64 | d__Bacteria;p__Bacteroidota;c__Bacteroidia;o__Bacteroidales;f__Tannerellaceae;g__Parabacteroides | 91 | blast;93.40 iPHoP-RF;79.00 |
| S10_vf_24791 | 46.17 | d__Bacteria;p__Firmicutes;c__Clostridia;o__Oscillospirales;f__Ruminococcaceae;g__Faecalibacterium | 93.8 | blast;95.60 CRISPR;94.10 iPHoP-RF;82.20 |
| S10_vf_43964 | 91.64 | d__Bacteria;p__Bacteroidota;c__Bacteroidia;o__Bacteroidales;f__Bacteroidaceae;g__Bacteroides | 100 | RaFAH;99.20 CRISPR;97.80 iPHoP-RF;97.10 blast;77.70 |
| S10_vf_43964 | 91.64 | d__Bacteria;p__Bacteroidota;c__Bacteroidia;o__Bacteroidales;f__Bacteroidaceae;g__Phocaeicola | 98.2 | CRISPR;98.70 iPHoP-RF;97.10 blast;92.80 |
| S10_vf_46328 | 63.06 | d__Bacteria;p__Firmicutes;c__Clostridia;o__Lachnospirales;f__Lachnospiraceae;g__UBA9502 | 91.4 | CRISPR;93.70 blast;91.80 iPHoP-RF;89.50 |
| S10_vf_62230 | 25.76 | d__Bacteria;p__Firmicutes;c__Clostridia;o__Clostridiales;f__Clostridiaceae;g__Clostridium | 90.6 | blast;93.00 iPHoP-RF;61.40 |
| S10_vf_64371 | 86.67 | d__Bacteria;p__Firmicutes;c__Bacilli;o__Lactobacillales;f__Streptococcaceae;g__Streptococcus | 98.8 | RaFAH;98.90 iPHoP-RF;59.80 |
| S10_vf_80712 | 98.13 | d__Bacteria;p__Firmicutes;c__Clostridia;o__Lachnospirales;f__Lachnospiraceae;g__Blautia_A | 100 | RaFAH;99.20 blast;91.30 iPHoP-RF;86.30 |
| S10_vf_80712 | 98.13 | d__Bacteria;p__Firmicutes;c__Clostridia;o__Lachnospirales;f__Lachnospiraceae;g__Blautia | 99 | RaFAH;99.20 |
| S10_vs_106834 | 44.85 | d__Bacteria;p__Bacteroidota;c__Bacteroidia;o__Bacteroidales;f__Bacteroidaceae;g__Bacteroides | 92.3 | RaFAH;87.20 iPHoP-RF;78.70 |
| S10_vs_120489 | 7.64 | d__Bacteria;p__Firmicutes;c__Clostridia;o__Christensenellales;f__Borkfalkiaceae;g__QALS01 | 90.3 | blast;92.80 iPHoP-RF;56.50 |
| S10_vs_120541 | 14.85 | d__Bacteria;p__Firmicutes;c__Clostridia;o__Oscillospirales;f__Oscillospiraceae;g__CAG-110 | 94.7 | blast;96.30 iPHoP-RF;95.70 CRISPR;74.70 |
| S10_vs_120570 | 98.22 | d__Bacteria;p__Firmicutes;c__Clostridia;o__Oscillospirales;f__Ruminococcaceae;g__Ruminococcus_D | 99.7 | blast;98.90 iPHoP-RF;96.70 CRISPR;87.50 RaFAH;87.40 |
| S10_vs_14557 | 99.66 | d__Bacteria;p__Bacteroidota;c__Bacteroidia;o__Bacteroidales;f__Bacteroidaceae;g__Bacteroides | 100 | RaFAH;98.20 iPHoP-RF;97.10 blast;96.90 CRISPR;90.30 |
| S10_vs_2538 | 31.92 | d__Bacteria;p__Firmicutes;c__Clostridia;o__Peptostreptococcales;f__Anaerovoracaceae;g__CAG-145 | 94.3 | blast;96.00 iPHoP-RF;76.40 |
| S10_vs_35854 | 48.73 | d__Bacteria;p__Bacteroidota;c__Bacteroidia;o__Bacteroidales;f__Bacteroidaceae;g__Bacteroides | 99.3 | iPHoP-RF;97.10 blast;96.00 RaFAH;90.40 |
| S10_vs_35854 | 48.73 | d__Bacteria;p__Bacteroidota;c__Bacteroidia;o__Bacteroidales;f__Bacteroidaceae;g__Phocaeicola | 91.4 | iPHoP-RF;93.70 |
| S10_vs_36407 | 8.93 | d__Bacteria;p__Firmicutes;c__Clostridia;o__Oscillospirales;f__Acutalibacteraceae;g__UBA1691 | 94.5 | blast;96.10 CRISPR;90.70 iPHoP-RF;73.00 |
| S10_vs_36598 | 93.41 | d__Bacteria;p__Bacteroidota;c__Bacteroidia;o__Bacteroidales;f__Bacteroidaceae;g__Bacteroides | 100 | RaFAH;99.40 iPHoP-RF;96.70 |
| S10_vs_36598 | 93.41 | d__Bacteria;p__Bacteroidota;c__Bacteroidia;o__Bacteroidales;f__Bacteroidaceae;g__Phocaeicola | 95.9 | iPHoP-RF;97.10 blast;95.40 |
| S10_vs_37379 | 75.26 | d__Bacteria;p__Bacteroidota;c__Bacteroidia;o__Bacteroidales;f__Tannerellaceae;g__Parabacteroides | 100 | CRISPR;98.90 RaFAH;98.30 iPHoP-RF;97.10 blast;94.80 |
| S10_vs_37694 | 99.94 | d__Bacteria;p__Bacteroidota;c__Bacteroidia;o__Bacteroidales;f__Rikenellaceae;g__Alistipes | 99.1 | iPHoP-RF;97.10 blast;96.80 RaFAH;87.90 CRISPR;55.60 |
| S10_vs_46819 | 98.03 | d__Bacteria;p__Firmicutes;c__Clostridia;o__Oscillospirales;f__Ruminococcaceae;g__Faecalibacterium | 99.4 | blast;96.80 RaFAH;92.90 iPHoP-RF;79.00 |
| S10_vs_58014 | 85.85 | d__Bacteria;p__Bacteroidota;c__Bacteroidia;o__Bacteroidales;f__Bacteroidaceae;g__Bacteroides | 100 | RaFAH;99.50 blast;91.80 iPHoP-RF;64.00 |
| S10_vs_58014 | 85.85 | d__Bacteria;p__Bacteroidota;c__Bacteroidia;o__Bacteroidales;f__Bacteroidaceae;g__Phocaeicola | 94.6 | blast;96.20 iPHoP-RF;76.40 |
| S10_vs_62776 | 44.54 | d__Bacteria;p__Bacteroidota;c__Bacteroidia;o__Bacteroidales;f__Bacteroidaceae;g__Bacteroides | 99 | iPHoP-RF;96.70 blast;93.60 RaFAH;88.20 |
| S10_vs_62776 | 44.54 | d__Bacteria;p__Bacteroidota;c__Bacteroidia;o__Bacteroidales;f__Bacteroidaceae;g__Phocaeicola | 92.6 | iPHoP-RF;94.70 blast;84.90 |
| S10_vs_67471 | 98.13 | d__Bacteria;p__Bacteroidota;c__Bacteroidia;o__Bacteroidales;f__Rikenellaceae;g__Alistipes | 99.9 | RaFAH;97.30 blast;95.30 iPHoP-RF;52.20 |
| S10_vs_67471 | 98.13 | d__Bacteria;p__Bacteroidota;c__Bacteroidia;o__Bacteroidales;f__Rikenellaceae;g__Alistipes_A | 96.2 | RaFAH;97.30 |
| S10_vs_71900 | 97 | d__Bacteria;p__Bacteroidota;c__Bacteroidia;o__Bacteroidales;f__Bacteroidaceae;g__Bacteroides | 100 | RaFAH;99.30 iPHoP-RF;97.10 blast;90.90 |
| S10_vs_71900 | 97 | d__Bacteria;p__Bacteroidota;c__Bacteroidia;o__Bacteroidales;f__Bacteroidaceae;g__Phocaeicola | 95.9 | iPHoP-RF;97.10 blast;96.20 CRISPR;95.50 |
| S10_vs_71900 | 97 | d__Bacteria;p__Bacteroidota;c__Bacteroidia;o__Bacteroidales;f__Bacteroidaceae;g__OM05-12 | 91 | iPHoP-RF;93.40 |
| S10_vs_77004 | 10.42 | d__Bacteria;p__Firmicutes;c__Clostridia;o__Oscillospirales;f__Acutalibacteraceae;g__Pseudoruminococcus | 95.3 | blast;96.70 CRISPR;89.20 iPHoP-RF;87.60 |
| S11_vf_104033 | 40.07 | d__Bacteria;p__Bacteroidota;c__Bacteroidia;o__Bacteroidales;f__Bacteroidaceae;g__Bacteroides | 90.6 | RaFAH;93.00 |
| S11_vf_104034 | 21.95 | d__Bacteria;p__Firmicutes;c__Clostridia;o__Oscillospirales;f__Oscillospiraceae;g__ER4 | 90.3 | blast;92.80 iPHoP-RF;81.10 |
| S11_vf_104035 | 37.57 | d__Bacteria;p__Bacteroidota;c__Bacteroidia;o__Bacteroidales;f__Bacteroidaceae;g__Prevotella | 91.7 | RaFAH;94.00 |
| S11_vf_104035 | 37.57 | d__Bacteria;p__Spirochaetota;c__Spirochaetia;o__Sphaerochaetales;f__Sphaerochaetaceae;g__UBA9732 | 90 | blast;92.50 iPHoP-RF;70.20 |
| S11_vf_104052 | 38.91 | d__Bacteria;p__Firmicutes;c__Clostridia;o__Oscillospirales;f__Acutalibacteraceae;g__CAG-180 | 91.5 | blast;93.80 iPHoP-RF;55.40 |
| S11_vf_104071 | 20.29 | d__Bacteria;p__Bacteroidota;c__Bacteroidia;o__Bacteroidales;f__Rikenellaceae;g__Alistipes | 98.5 | CRISPR;98.90 iPHoP-RF;92.80 |
| S11_vf_10907 | 87.86 | d__Bacteria;p__Firmicutes;c__Clostridia;o__Lachnospirales;f__Lachnospiraceae;g__Bariatricus | 91.6 | blast;93.90 iPHoP-RF;82.80 |
| S11_vf_27845 | 53.78 | d__Bacteria;p__Firmicutes;c__Bacilli;o__Lactobacillales;f__Streptococcaceae;g__Streptococcus | 100 | CRISPR;98.80 RaFAH;97.70 iPHoP-RF;85.70 |
| S11_vf_27845 | 53.78 | d__Bacteria;p__Bacteroidota;c__Bacteroidia;o__Flavobacteriales;f__Flavobacteriaceae;g__Capnocytophaga | 91 | blast;93.40 iPHoP-RF;51.20 |
| S11_vf_28515 | 95.85 | d__Bacteria;p__Firmicutes;c__Clostridia;o__Lachnospirales;f__Lachnospiraceae;g__Blautia_A | 100 | RaFAH;98.80 blast;94.40 iPHoP-RF;90.50 CRISPR;68.00 |
| S11_vf_28515 | 95.85 | d__Bacteria;p__Firmicutes;c__Clostridia;o__Lachnospirales;f__Lachnospiraceae;g__Blautia | 98.4 | RaFAH;98.80 |
| S11_vf_28645 | 6.84 | d__Bacteria;p__Firmicutes;c__Clostridia;o__Lachnospirales;f__Lachnospiraceae;g__Blautia_A | 92.5 | blast;94.60 iPHoP-RF;51.70 |
| S11_vf_33117 | 96.79 | d__Bacteria;p__Firmicutes;c__Clostridia;o__Oscillospirales;f__Oscillospiraceae;g__CAG-83 | 97.3 | RaFAH;98.10 |
| S11_vf_33117 | 96.79 | d__Bacteria;p__Firmicutes;c__Clostridia;o__Oscillospirales;f__Oscillospiraceae;g__Dysosmobacter | 97.3 | RaFAH;98.10 |
| S11_vf_33117 | 96.79 | d__Bacteria;p__Firmicutes;c__Clostridia;o__Oscillospirales;f__Oscillospiraceae;g__Flavonifractor | 93.8 | blast;95.60 iPHoP-RF;71.50 |
| S11_vf_34008 | 44.72 | d__Bacteria;p__Firmicutes;c__Clostridia;o__Lachnospirales;f__Lachnospiraceae;g__ | 93.1 | iPHoP-RF;95.10 blast;93.90 |
| S11_vf_34008 | 44.72 | d__Bacteria;p__Firmicutes;c__Clostridia;o__Lachnospirales;f__Lachnospiraceae;g__UC5-1-2E3 | 91.6 | blast;93.90 iPHoP-RF;93.10 |
| S11_vf_43997 | 15.72 | d__Bacteria;p__Bacteroidota;c__Bacteroidia;o__Bacteroidales;f__Bacteroidaceae;g__Bacteroides | 91.4 | blast;93.70 iPHoP-RF;78.70 |
| S11_vf_58587 | 11.86 | d__Bacteria;p__Actinobacteriota;c__Actinomycetia;o__Actinomycetales;f__Bifidobacteriaceae;g__Bifidobacterium | 90.4 | blast;92.90 iPHoP-RF;65.60 |
| S11_vf_66522 | 97.65 | d__Bacteria;p__Actinobacteriota;c__Actinomycetia;o__Actinomycetales;f__Actinomycetaceae;g__Pauljensenia | 100 | RaFAH;99.20 CRISPR;99.10 blast;93.90 iPHoP-RF;90.50 |
| S11_vf_66522 | 97.65 | d__Bacteria;p__Actinobacteriota;c__Actinomycetia;o__Actinomycetales;f__Actinomycetaceae;g__Actinomyces | 99 | RaFAH;99.20 |
| S11_vf_68693 | 78.63 | d__Bacteria;p__Bacteroidota;c__Bacteroidia;o__Bacteroidales;f__Tannerellaceae;g__Parabacteroides | 99.3 | CRISPR;97.70 blast;97.30 iPHoP-RF;91.80 RaFAH;87.20 |
| S11_vf_70015 | 50.35 | d__Bacteria;p__Bacteroidota;c__Bacteroidia;o__Bacteroidales;f__Rikenellaceae;g__Alistipes | 97.8 | blast;96.10 RaFAH;79.90 |
| S11_vf_92391 | 5.33 | d__Bacteria;p__Firmicutes;c__Bacilli;o__Erysipelotrichales;f__Erysipelotrichaceae;g__Clostridium_AQ | 91 | CRISPR;93.40 |
| S11_vf_95067 | 39.9 | d__Bacteria;p__Firmicutes;c__Clostridia;o__Oscillospirales;f__Oscillospiraceae;g__Flavonifractor | 93.4 | blast;95.30 iPHoP-RF;58.20 |
| S11_vf_99751 | 8.04 | d__Bacteria;p__Firmicutes;c__Clostridia;o__Oscillospirales;f__Oscillospiraceae;g__CAG-83 | 94.9 | blast;96.40 iPHoP-RF;57.10 |
| S11_vs_104041 | 16.5 | d__Bacteria;p__Cyanobacteria;c__Vampirovibrionia;o__Gastranaerophilales;f__Gastranaerophilaceae;g__CAG-306 | 97.2 | blast;98.00 iPHoP-RF;56.00 |
| S11_vs_104042 | 75.46 | d__Bacteria;p__Firmicutes;c__Clostridia;o__Lachnospirales;f__Lachnospiraceae;g__Eubacterium_I | 98.1 | blast;98.60 iPHoP-RF;84.80 |
| S11_vs_104064 | 99.03 | d__Bacteria;p__Firmicutes;c__Clostridia;o__Oscillospirales;f__Ruminococcaceae;g__Negativibacillus | 92.1 | blast;94.30 |
| S11_vs_11994 | 21.97 | d__Bacteria;p__Firmicutes;c__Clostridia;o__Oscillospirales;f__Ruminococcaceae;g__Faecalibacterium | 91.7 | RaFAH;94.00 |
| S11_vs_13652 | 57.92 | d__Bacteria;p__Firmicutes;c__Clostridia;o__Oscillospirales;f__Ruminococcaceae;g__Massilioclostridium | 92.9 | blast;94.90 iPHoP-RF;66.50 |
| S11_vs_14882 | 11.87 | d__Bacteria;p__Firmicutes;c__Bacilli;o__Erysipelotrichales;f__Erysipelotrichaceae;g__Holdemania | 93.1 | blast;95.10 CRISPR;54.60 |
| S11_vs_1937 | 99.04 | d__Bacteria;p__Bacteroidota;c__Bacteroidia;o__Bacteroidales;f__Rikenellaceae;g__Alistipes | 100 | blast;98.60 RaFAH;96.60 iPHoP-RF;83.30 |
| S11_vs_1937 | 99.04 | d__Bacteria;p__Bacteroidota;c__Bacteroidia;o__Bacteroidales;f__Rikenellaceae;g__Alistipes_A | 95.2 | RaFAH;96.60 |
| S11_vs_19892 | 45.95 | d__Bacteria;p__Firmicutes;c__Clostridia;o__Oscillospirales;f__Oscillospiraceae;g__Lawsonibacter | 90.6 | blast;93.00 iPHoP-RF;67.50 |
| S11_vs_23188 | 66.52 | d__Bacteria;p__Bacteroidota;c__Bacteroidia;o__Bacteroidales;f__Bacteroidaceae;g__Bacteroides | 99.9 | iPHoP-RF;97.10 RaFAH;95.80 blast;93.80 CRISPR;83.30 |
| S11_vs_26767 | 40.78 | d__Bacteria;p__Firmicutes;c__Clostridia;o__Lachnospirales;f__Lachnospiraceae;g__Ventrimonas | 90.8 | blast;93.20 iPHoP-RF;72.20 |
| S11_vs_3006 | 44.36 | d__Bacteria;p__Firmicutes;c__Clostridia;o__Oscillospirales;f__Acutalibacteraceae;g__UMGS263 | 93.6 | blast;95.50 iPHoP-RF;67.00 |
| S11_vs_3277 | 99.63 | d__Bacteria;p__Bacteroidota;c__Bacteroidia;o__Bacteroidales;f__Bacteroidaceae;g__Bacteroides | 99.9 | RaFAH;98.80 iPHoP-RF;83.60 blast;80.70 |
| S11_vs_34137 | 17.51 | d__Bacteria;p__Firmicutes;c__Clostridia;o__Lachnospirales;f__Lachnospiraceae;g__Blautia_A | 91.4 | blast;93.70 iPHoP-RF;83.10 |
| S11_vs_36748 | 49.79 | d__Bacteria;p__Bacteroidota;c__Bacteroidia;o__Bacteroidales;f__Rikenellaceae;g__Alistipes | 98.5 | CRISPR;98.90 blast;96.70 iPHoP-RF;91.80 |

| | | | | |
|---|---|---|---|---|
| S11_vs_38860 | 39.67 | d__Bacteria;p__Firmicutes;c__Clostridia;o__Lachnospirales;f__Lachnospiraceae;g__Enterocloster | 95.2 | CRISPR;96.60 blast;95.90 iPHoP-RF;89.80 |
| S11_vs_41107 | 98.96 | d__Bacteria;p__Firmicutes;c__Clostridia;o__Clostridiales;f__Clostridiaceae;g__Clostridium | 93.6 | RaFAH;95.50 |
| S11_vs_41107 | 98.96 | d__Bacteria;p__Firmicutes;c__Clostridia;o__Clostridiales;f__Clostridiaceae;g__Clostridium_F | 93.6 | RaFAH;95.50 |
| S11_vs_41107 | 98.96 | d__Bacteria;p__Firmicutes;c__Clostridia;o__Clostridiales;f__Clostridiaceae;g__Clostridium_P | 93.6 | RaFAH;95.50 |
| S11_vs_45658 | 40.35 | d__Bacteria;p__Bacteroidota;c__Bacteroidia;o__Bacteroidales;f__Bacteroidaceae;g__Bacteroides | 95 | RaFAH;96.20 iPHoP-RF;53.60 |
| S11_vs_45658 | 40.35 | d__Bacteria;p__Bacteroidota;c__Bacteroidia;o__Bacteroidales;f__UBA11471;g__UBA11471 | 90 | blast;92.50 iPHoP-RF;77.50 |
| S11_vs_48109 | 63.66 | d__Bacteria;p__Firmicutes;c__Clostridia;o__Oscillospirales;f__Acutalibacteraceae;g__An172 | 93.4 | blast;95.30 iPHoP-RF;74.60 |
| S11_vs_5077 | 84.31 | d__Bacteria;p__Proteobacteria;c__Gammaproteobacteria;o__Enterobacterales;f__Enterobacteriaceae;g__Salmonella | 95.3 | blast;96.70 |
| S11_vs_5077 | 84.31 | d__Bacteria;p__Proteobacteria;c__Gammaproteobacteria;o__Enterobacterales;f__Enterobacteriaceae;g__Escherichia | 94.6 | blast;96.20 |
| S11_vs_5077 | 84.31 | d__Bacteria;p__Proteobacteria;c__Gammaproteobacteria;o__Enterobacterales;f__Enterobacteriaceae;g__Enterobacter | 93.9 | blast;95.70 iPHoP-RF;60.10 |
| S11_vs_52577 | 78.49 | d__Bacteria;p__Firmicutes;c__Clostridia;o__Oscillospirales;f__Oscillospiraceae;g__Dysosmobacter | 91 | blast;88.40 iPHoP-RF;73.50 RaFAH;71.60 |
| S11_vs_55144 | 12.8 | d__Bacteria;p__Firmicutes;c__Bacilli;o__Erysipelotrichales;f__Erysipelatoclostridiaceae;g__Massiliomicrobiota | 90.3 | blast;92.80 |
| S11_vs_58930 | 93.75 | d__Bacteria;p__Bacteroidota;c__Bacteroidia;o__Bacteroidales;f__Bacteroidaceae;g__Bacteroides | 98.1 | blast;92.60 RaFAH;90.60 iPHoP-RF;89.80 |
| S11_vs_58930 | 93.75 | d__Bacteria;p__Bacteroidota;c__Bacteroidia;o__Bacteroidales;f__Bacteroidaceae;g__Paraprevotella | 90 | blast;92.50 |
| S11_vs_6458 | 15.38 | d__Bacteria;p__Proteobacteria;c__Alphaproteobacteria;o__RF32;f__CAG-239;g__CAG-267 | 90.1 | blast;92.60 iPHoP-RF;81.90 |
| S11_vs_67573 | 11.32 | d__Bacteria;p__Firmicutes;c__Clostridia;o__Lachnospirales;f__Lachnospiraceae;g__Enterocloster | 92.9 | CRISPR;94.90 blast;92.10 iPHoP-RF;92.10 |
| S11_vs_68151 | 99.98 | d__Bacteria;p__Firmicutes;c__Clostridia;o__Lachnospirales;f__Lachnospiraceae;g__Mediterraneibacter | 95.3 | blast;96.70 |
| S11_vs_68151 | 99.98 | d__Bacteria;p__Firmicutes;c__Clostridia;o__Lachnospirales;f__Lachnospiraceae;g__Hungatella | 94.7 | blast;87.90 RaFAH;84.60 iPHoP-RF;53.30 |
| S11_vs_68348 | 49.51 | d__Bacteria;p__Firmicutes;c__Bacilli;o__Erysipelotrichales;f__Erysipelatoclostridiaceae;g__Erysipelatoclostridium | 95.2 | blast;96.60 iPHoP-RF;64.50 |
| S11_vs_6878 | 72.71 | d__Bacteria;p__Firmicutes;c__Clostridia;o__Lachnospirales;f__Lachnospiraceae;g__ | 91 | blast;93.40 iPHoP-RF;73.00 |
| S11_vs_76784 | 41.57 | d__Bacteria;p__Firmicutes;c__Clostridia;o__Oscillospirales;f__Ruminococcaceae;g__Ruthenibacterium | 93 | blast;95.00 iPHoP-RF;77.30 |
| S11_vs_79831 | 17.3 | d__Bacteria;p__Bacteroidota;c__Bacteroidia;o__Bacteroidales;f__UBA11471;g__UBA11471 | 90 | blast;92.50 iPHoP-RF;73.20 |
| S11_vs_81775 | 24.35 | d__Bacteria;p__Firmicutes;c__Clostridia;o__Eubacteriales;f__Eubacteriaceae;g__Eubacterium | 95 | CRISPR;96.50 blast;93.70 iPHoP-RF;81.40 |
| S11_vs_82331 | 98.94 | d__Bacteria;p__Cyanobacteria;c__Vampirovibrionia;o__Gastranaerophilales;f__Gastranaerophilaceae;g__QAMI01 | 93.1 | blast;95.10 |
| S11_vs_82368 | 62.18 | d__Bacteria;p__Firmicutes;c__Clostridia;o__Lachnospirales;f__Lachnospiraceae;g__Blautia_A | 98.1 | blast;92.60 RaFAH;90.70 iPHoP-RF;86.00 |
| S11_vs_83666 | 35.28 | d__Bacteria;p__Firmicutes;c__Clostridia;o__Lachnospirales;f__Lachnospiraceae;g__Dorea_A | 90.9 | blast;93.30 iPHoP-RF;67.70 |
| S11_vs_84627 | 6.74 | d__Bacteria;p__Proteobacteria;c__Alphaproteobacteria;o__RF32;f__CAG-239;g__51-20 | 95.6 | blast;96.90 iPHoP-RF;89.50 CRISPR;76.50 |
| S11_vs_84848 | 95.91 | d__Bacteria;p__Bacteroidota;c__Bacteroidia;o__Bacteroidales;f__Bacteroidaceae;g__Bacteroides | 95.9 | blast;91.20 RaFAH;83.50 iPHoP-RF;57.90 |
| S11_vs_84848 | 95.91 | d__Bacteria;p__Bacteroidota;c__Bacteroidia;o__Bacteroidales;f__Tannerellaceae;g__Parabacteroides | 93.2 | blast;92.10 CRISPR;86.70 iPHoP-RF;83.10 RaFAH;69.90 |
| S11_vs_98193 | 80.02 | d__Bacteria;p__Bacteroidota;c__Bacteroidia;o__Bacteroidales;f__Rikenellaceae;g__Alistipes | 96.2 | iPHoP-RF;92.10 blast;92.00 RaFAH;82.70 |
| S12_vf_105542 | 13.83 | d__Bacteria;p__Firmicutes;c__Bacilli;o__RF39;f__UBA660;g__CAG-460 | 94.6 | blast;96.20 iPHoP-RF;73.70 |
| S12_vf_10887 | 7.67 | d__Bacteria;p__Proteobacteria;c__Gammaproteobacteria;o__Enterobacterales;f__Pasteurellaceae;g__Glaesserella | 94.5 | iPHoP-RF;96.10 CRISPR;91.90 |
| S12_vf_10887 | 7.67 | d__Bacteria;p__Proteobacteria;c__Gammaproteobacteria;o__Enterobacterales;f__Pasteurellaceae;g__Bibersteinia | 93.9 | iPHoP-RF;95.70 |
| S12_vf_10887 | 7.67 | d__Bacteria;p__Proteobacteria;c__Gammaproteobacteria;o__Enterobacterales;f__Pasteurellaceae;g__Nicoletella | 93.9 | iPHoP-RF;95.70 |
| S12_vf_10887 | 7.67 | d__Bacteria;p__Proteobacteria;c__Gammaproteobacteria;o__Enterobacterales;f__Pasteurellaceae;g__Muribacter | 93.5 | iPHoP-RF;95.40 |
| S12_vf_10887 | 7.67 | d__Bacteria;p__Proteobacteria;c__Gammaproteobacteria;o__Enterobacterales;f__Pasteurellaceae;g__Actinobacillus | 92.2 | iPHoP-RF;94.40 |
| S12_vf_10887 | 7.67 | d__Bacteria;p__Proteobacteria;c__Gammaproteobacteria;o__Enterobacterales;f__Pasteurellaceae;g__Actinobacillus_C | 92.2 | iPHoP-RF;94.40 |
| S12_vf_10887 | 7.67 | d__Bacteria;p__Proteobacteria;c__Gammaproteobacteria;o__Enterobacterales;f__Pasteurellaceae;g__Mannheimia | 91.8 | iPHoP-RF;94.10 |
| S12_vf_145044 | 32.93 | d__Bacteria;p__Bacteroidota;c__Bacteroidia;o__Bacteroidales;f__Rikenellaceae;g__Alistipes_A | 91.6 | blast;93.90 iPHoP-RF;77.10 |
| S12_vf_16374 | 100 | d__Bacteria;p__Bacteroidota;c__Bacteroidia;o__Bacteroidales;f__Bacteroidaceae;g__Bacteroides | 100 | RaFAH;99.10 iPHoP-RF;97.10 blast;94.80 |
| S12_vf_16374 | 100 | d__Bacteria;p__Bacteroidota;c__Bacteroidia;o__Bacteroidales;f__Tannerellaceae;g__Parabacteroides | 92.9 | blast;94.90 |
| S12_vf_170614 | 7.03 | d__Bacteria;p__Actinobacteriota;c__Coriobacteriia;o__Coriobacteriales;f__Coriobacteriaceae;g__Collinsella | 96.4 | CRISPR;97.50 iPHoP-RF;95.10 |
| S12_vf_173208 | 10.29 | d__Bacteria;p__Firmicutes;c__Clostridia;o__Oscillospirales;f__Oscillospiraceae;g__CAG-170 | 93 | blast;95.00 iPHoP-RF;81.40 |
| S12_vf_173666 | 36.83 | d__Bacteria;p__Actinobacteriota;c__Coriobacteriia;o__Coriobacteriales;f__Eggerthellaceae;g__Adlercreutzia | 93.1 | blast;95.10 iPHoP-RF;76.40 |
| S12_vf_173666 | 36.83 | d__Bacteria;p__Actinobacteriota;c__Coriobacteriia;o__Coriobacteriales;f__Coriobacteriaceae;g__Collinsella | 92.6 | CRISPR;94.70 |
| S12_vf_174374 | 94.2 | d__Bacteria;p__Bacteroidota;c__Bacteroidia;o__Bacteroidales;f__Rikenellaceae;g__Alistipes | 96.2 | CRISPR;97.30 iPHoP-RF;97.10 blast;96.60 |
| S12_vf_174374 | 94.2 | d__Bacteria;p__Bacteroidota;c__Bacteroidia;o__Bacteroidales;f__Bacteroidaceae;g__Bacteroides | 91.6 | RaFAH;93.90 |
| S12_vf_176437 | 10.68 | d__Bacteria;p__Firmicutes;c__Clostridia;o__Lachnospirales;f__Lachnospiraceae;g__OF09-33XD | 95.3 | CRISPR;96.70 iPHoP-RF;82.20 |
| S12_vf_181024 | 98.48 | d__Bacteria;p__Firmicutes;c__Clostridia;o__Oscillospirales;f__Ruminococcaceae;g__Ruminococcus_D | 99.9 | CRISPR;97.30 RaFAH;97.20 blast;96.90 iPHoP-RF;95.70 |
| S12_vf_181024 | 98.48 | d__Bacteria;p__Firmicutes;c__Clostridia;o__Oscillospirales;f__Acutalibacteraceae;g__Ruminococcus_E | 96 | RaFAH;97.20 |
| S12_vf_188846 | | d__Bacteria;p__Bacteroidota;c__Bacteroidia;o__Bacteroidales;f__Barnesiellaceae;g__Barnesiella | 94.5 | blast;96.10 iPHoP-RF;68.60 |
| S12_vf_18902 | 80.1 | d__Bacteria;p__Firmicutes;c__Clostridia;o__Oscillospirales;f__Oscillospiraceae;g__ER4 | 90.6 | blast;93.00 iPHoP-RF;67.00 |
| S12_vf_60224 | 82.87 | d__Bacteria;p__Bacteroidota;c__Bacteroidia;o__Bacteroidales;f__Bacteroidaceae;g__Bacteroides | 100 | RaFAH;97.10 iPHoP-RF;96.70 blast;80.90 |
| S12_vf_60224 | 82.87 | d__Bacteria;p__Bacteroidota;c__Bacteroidia;o__Bacteroidales;f__Bacteroidaceae;g__Paraprevotella | 91.2 | blast;93.60 |
| S12_vf_61299 | 11.99 | d__Bacteria;p__Firmicutes;c__Clostridia;o__Lachnospirales;f__Lachnospiraceae;g__Muricomes | 96.9 | CRISPR;97.80 iPHoP-RF;59.20 |
| S12_vf_69136 | 51.25 | d__Bacteria;p__Bacteroidota;c__Bacteroidia;o__Bacteroidales;f__Bacteroidaceae;g__Prevotellamassilia | 98.4 | CRISPR;98.80 blast;82.30 iPHoP-RF;78.30 |
| S12_vf_69136 | 51.25 | d__Bacteria;p__Bacteroidota;c__Bacteroidia;o__Bacteroidales;f__Bacteroidaceae;g__Paraprevotella | 90.8 | blast;93.20 iPHoP-RF;67.50 |
| S12_vf_79666 | 99.55 | d__Bacteria;p__Firmicutes;c__Clostridia;o__Oscillospirales;f__Acutalibacteraceae;g__UBA1417 | 91.7 | blast;94.00 iPHoP-RF;58.20 |
| S12_vs_111286 | 44.45 | d__Bacteria;p__Bacteroidota;c__Bacteroidia;o__Bacteroidales;f__Muribaculaceae;g__UBA7173 | 91.6 | blast;93.90 iPHoP-RF;62.50 |
| S12_vs_121913 | 82.73 | d__Bacteria;p__Bacteroidota;c__Bacteroidia;o__Bacteroidales;f__Rikenellaceae;g__Alistipes | 98.5 | blast;96.70 RaFAH;83.70 |
| S12_vs_123620 | 10.35 | d__Bacteria;p__Firmicutes;c__Bacilli;o__RF39;f__UBA660;g__CAG-302 | 95.4 | blast;96.80 iPHoP-RF;64.80 |
| S12_vs_133070 | 14.52 | d__Bacteria;p__Firmicutes;c__Clostridia;o__Oscillospirales;f__Ruminococcaceae;g__CAG-353 | 91.1 | blast;93.50 iPHoP-RF;79.80 |
| S12_vs_138261 | 70.17 | d__Bacteria;p__Bacteroidota;c__Bacteroidia;o__Bacteroidales;f__Tannerellaceae;g__Parabacteroides | 98.4 | blast;98.80 CRISPR;97.40 iPHoP-RF;96.10 |
| S12_vs_141337 | 98.55 | d__Bacteria;p__Bacteroidota;c__Bacteroidia;o__Bacteroidales;f__Bacteroidaceae;g__Bacteroides | 100 | RaFAH;99.40 blast;92.10 iPHoP-RF;57.90 |
| S12_vs_142625 | 31.03 | d__Bacteria;p__Bacteroidota;c__Bacteroidia;o__Bacteroidales;f__Rikenellaceae;g__Alistipes | 95.9 | iPHoP-RF;97.10 blast;96.90 |
| S12_vs_151292 | 99.81 | d__Bacteria;p__Bacteroidota;c__Bacteroidia;o__Bacteroidales;f__Bacteroidaceae;g__Bacteroides | 100 | RaFAH;98.90 iPHoP-RF;97.10 blast;95.90 CRISPR;68.40 |
| S12_vs_151292 | 99.81 | d__Bacteria;p__Bacteroidota;c__Bacteroidia;o__Bacteroidales;f__Bacteroidaceae;g__Phocaeicola | 95.9 | iPHoP-RF;97.10 blast;93.30 CRISPR;74.50 |
| S12_vs_153780 | 97.5 | d__Bacteria;p__Proteobacteria;c__Alphaproteobacteria;o__RF32;f__CAG-239;g__CAG-495 | 90.9 | blast;93.30 iPHoP-RF;81.60 |
| S12_vs_163990 | 99.19 | d__Bacteria;p__Firmicutes;c__Clostridia;o__Lachnospirales;f__Lachnospiraceae;g__Dorea | 99.3 | RaFAH;99.40 |
| S12_vs_163990 | 99.19 | d__Bacteria;p__Firmicutes;c__Clostridia;o__Lachnospirales;f__Lachnospiraceae;g__Dorea_A | 99.3 | RaFAH;99.40 |
| S12_vs_163990 | 99.19 | d__Bacteria;p__Firmicutes;c__Clostridia;o__Oscillospirales;f__Oscillospiraceae;g__ER4 | 95 | blast;96.50 iPHoP-RF;60.30 |
| S12_vs_164056 | 11.8 | d__Bacteria;p__Firmicutes;c__Clostridia;o__Oscillospirales;f__Oscillospiraceae;g__CAG-83 | 95.9 | blast;97.10 iPHoP-RF;72.60 |
| S12_vs_172379 | 18.46 | d__Bacteria;p__Firmicutes;c__Bacilli;o__RF39;f__UBA660;g__CAG-877 | 90.8 | blast;93.20 |
| S12_vs_188820 | 7.83 | d__Bacteria;p__Firmicutes;c__Clostridia;o__Oscillospirales;f__CAG-272;g__QALR01 | 93 | blast;95.00 iPHoP-RF;87.00 |
| S12_vs_188828 | 8.93 | d__Bacteria;p__Firmicutes;c__Clostridia;o__Oscillospirales;f__UMGS1783;g__UMGS1783 | 92.3 | blast;94.50 |
| S12_vs_1980 | 94.44 | d__Bacteria;p__Bacteroidota;c__Bacteroidia;o__Bacteroidales;f__Bacteroidaceae;g__Bacteroides | 99.1 | iPHoP-RF;97.10 blast;93.60 RaFAH;87.80 CRISPR;58.50 |
| S12_vs_1980 | 94.44 | d__Bacteria;p__Bacteroidota;c__Bacteroidia;o__Bacteroidales;f__Bacteroidaceae;g__Phocaeicola | 94.5 | iPHoP-RF;96.10 blast;70.90 |

| | | | | |
|---|---|---|---|---|
| S12_vs_23796 | 72.14 | d__Bacteria;p__Bacteroidota;c__Bacteroidia;o__Bacteroidales;f__Rikenellaceae;g__Alistipes | 99.1 | iPHoP-RF;96.70 blast;96.60 RaFAH;89.00 |
| S12_vs_26336 | 100 | d__Bacteria;p__Bacteroidota;c__Bacteroidia;o__Bacteroidales;f__Bacteroidaceae;g__Bacteroides | 99.6 | RaFAH;99.60 |
| S12_vs_26336 | 100 | d__Bacteria;p__Bacteroidota;c__Bacteroidia;o__Bacteroidales;f__Rikenellaceae;g__Alistipes | 95.6 | blast;96.90 iPHoP-RF;96.70 |
| S12_vs_26847 | 68.45 | d__Bacteria;p__Firmicutes;c__Clostridia;o__Lachnospirales;f__Lachnospiraceae;g__Lachnospira | 99.3 | CRISPR;98.90 iPHoP-RF;95.70 blast;94.10 RaFAH;74.50 |
| S12_vs_32669 | 3.93 | d__Bacteria;p__Firmicutes;c__Clostridia;o__Christensenellales;f__CAG-917;g__CAG-475 | 94.7 | blast;96.30 iPHoP-RF;68.60 |
| S12_vs_48045 | 24.79 | d__Bacteria;p__Firmicutes;c__Clostridia;o__Christensenellales;f__CAG-74;g__UBA11524 | 94.9 | blast;96.40 iPHoP-RF;53.80 |
| S12_vs_48045 | 24.79 | d__Bacteria;p__Firmicutes;c__Clostridia;o__Christensenellales;f__CAG-74;g__UBA11524 | 92.7 | blast;94.80 iPHoP-RF;56.00 |
| S12_vs_6836 | 43.21 | d__Bacteria;p__Firmicutes;c__Clostridia;o__TANB77;f__CAG-508;g__CAG-269 | 92.7 | blast;94.80 iPHoP-RF;65.30 |
| S12_vs_71525 | 30.6 | d__Bacteria;p__Bacteroidota;c__Bacteroidia;o__Bacteroidales;f__Marinifilaceae;g__Butyricimonas | 93.8 | blast;95.60 iPHoP-RF;67.30 |
| S12_vs_71525 | 30.6 | d__Bacteria;p__Bacteroidota;c__Bacteroidia;o__Bacteroidales;f__Bacteroidaceae;g__Bacteroides | 92.1 | RaFAH;94.30 |
| S12_vs_74280 | 14.1 | d__Bacteria;p__Bacteroidota;c__Bacteroidia;o__Bacteroidales;f__Marinifilaceae;g__Butyricimonas | 94.3 | blast;96.00 iPHoP-RF;58.40 |
| S12_vs_81729 | 10.77 | d__Bacteria;p__Firmicutes;c__Clostridia;o__Oscillospirales;f__Ruminococcaceae;g__Faecalibacterium | 97.3 | CRISPR;98.10 iPHoP-RF;93.40 blast;92.10 |
| S12_vs_86928 | 18.68 | d__Bacteria;p__Verrucomicrobiota;c__Verrucomicrobiae;o__Verrucomicrobiales;f__Akkermansiaceae;g__Akkermansia | 100 | CRISPR;98.80 RaFAH;98.60 blast;96.90 iPHoP-RF;82.50 |
| S12_vs_87088 | 36.55 | d__Bacteria;p__Firmicutes;c__Clostridia;o__Oscillospirales;f__Ruminococcaceae;g__Faecalibacterium | 97.5 | blast;96.60 iPHoP-RF;92.40 RaFAH;73.80 CRISPR;53.50 |
| S12_vs_92868 | 65.38 | d__Bacteria;p__Firmicutes;c__Clostridia;o__Oscillospirales;f__Oscillospiraceae;g__Intestinimonas | 93 | blast;95.00 iPHoP-RF;58.70 |
| S12_vs_93357 | 40.56 | d__Bacteria;p__Proteobacteria;c__Gammaproteobacteria;o__Burkholderiales;f__Burkholderiaceae;g__Parasutterella | 97 | CRISPR;97.90 blast;96.90 iPHoP-RF;93.10 |
| S12_vs_94042 | 10.3 | d__Bacteria;p__Firmicutes;c__Clostridia;o__Christensenellales;f__CAG-74;g__UBA11524 | 95.6 | blast;96.90 iPHoP-RF;91.80 |
| S12_vs_94042 | 10.3 | d__Bacteria;p__Firmicutes;c__Clostridia;o__Christensenellales;f__CAG-74;g__UBA11524 | 95.6 | blast;96.90 CRISPR;96.60 iPHoP-RF;91.80 |
| S12_vs_98175 | 95.22 | d__Bacteria;p__Firmicutes;c__Clostridia;o__Lachnospirales;f__Lachnospiraceae;g__Bariatricus | 90.3 | iPHoP-RF;89.50 blast;81.30 RaFAH;65.60 |
| S12_vs_99092 | 78.56 | d__Bacteria;p__Firmicutes;c__Clostridia;o__Lachnospirales;f__Lachnospiraceae;g__Lachnospira | 99.6 | CRISPR;98.80 blast;98.80 iPHoP-RF;94.40 RaFAH;84.60 |
| S12_vs_99219 | 99.93 | d__Bacteria;p__Bacteroidota;c__Bacteroidia;o__Bacteroidales;f__Bacteroidaceae;g__Bacteroides | 100 | RaFAH;98.90 iPHoP-RF;96.70 blast;91.60 |
| S12_vs_99219 | 99.93 | d__Bacteria;p__Bacteroidota;c__Bacteroidia;o__Bacteroidales;f__Bacteroidaceae;g__Phocaeicola | 92.1 | blast;94.30 iPHoP-RF;94.10 |
| S13_vf_109235 | 72.39 | d__Bacteria;p__Firmicutes;c__Clostridia;o__Oscillospirales;f__Oscillospiraceae;g__CAG-83 | 90.1 | blast;92.60 iPHoP-RF;68.90 |
| S13_vf_11246 | 46.73 | d__Bacteria;p__Bacteroidota;c__Bacteroidia;o__Bacteroidales;f__Bacteroidaceae;g__Prevotella | 91.8 | RaFAH;94.10 |
| S13_vf_143422 | 75.96 | d__Bacteria;p__Bacteroidota;c__Bacteroidia;o__Bacteroidales;f__Tannerellaceae;g__Parabacteroides | 98.1 | blast;98.60 iPHoP-RF;83.30 |
| S13_vf_147589 | 6.46 | d__Bacteria;p__Firmicutes;c__Clostridia;o__Oscillospirales;f__Oscillospiraceae;g__Flavonifractor | 91.5 | blast;93.80 iPHoP-RF;78.50 |
| S13_vf_150407 | | d__Bacteria;p__Firmicutes;c__Clostridia;o__Oscillospirales;f__Oscillospiraceae;g__ER4 | 93.5 | CRISPR;95.40 |
| S13_vf_151045 | 98.31 | d__Bacteria;p__Proteobacteria;c__Gammaproteobacteria;o__Enterobacterales;f__Enterobacteriaceae;g__Escherichia | 95.7 | RaFAH;97.00 |
| S13_vf_28309 | 12.14 | d__Bacteria;p__Firmicutes;c__Clostridia;o__Oscillospirales;f__Acutalibacteraceae;g__Ruminococcus_E | 90.1 | blast;92.60 |
| S13_vf_54201 | 17.62 | d__Bacteria;p__Firmicutes;c__Clostridia;o__Oscillospirales;f__Ruminococcaceae;g__Faecalibacterium | 98.7 | RaFAH;93.20 blast;89.90 iPHoP-RF;62.20 |
| S13_vf_54201 | 17.62 | d__Bacteria;p__Firmicutes;c__Negativicutes;o__Acidaminococcales;f__Acidaminococcaceae;g__Phascolarctobacterium | 93.1 | blast;95.10 iPHoP-RF;68.20 |
| S13_vf_56074 | 91.56 | d__Bacteria;p__Bacteroidota;c__Bacteroidia;o__Bacteroidales;f__Rikenellaceae;g__Alistipes | 98.1 | RaFAH;98.60 |
| S13_vf_56074 | 91.56 | d__Bacteria;p__Bacteroidota;c__Bacteroidia;o__Bacteroidales;f__Rikenellaceae;g__Alistipes_A | 98.1 | RaFAH;98.60 |
| S13_vf_56074 | 91.56 | d__Bacteria;p__Bacteroidota;c__Bacteroidia;o__Bacteroidales;f__Bacteroidaceae;g__UBA6398 | 91.6 | blast;93.90 iPHoP-RF;74.30 |
| S13_vf_66508 | 88.17 | d__Bacteria;p__Bacteroidota;c__Bacteroidia;o__Bacteroidales;f__Bacteroidaceae;g__Bacteroides | 99.4 | iPHoP-RF;97.10 blast;96.60 RaFAH;91.90 |
| S13_vf_7670 | 6.84 | d__Bacteria;p__Verrucomicrobiota;c__Verrucomicrobiae;o__Verrucomicrobiales;f__Akkermansiaceae;g__Akkermansia | 100 | CRISPR;99.60 RaFAH;96.00 blast;93.60 iPHoP-RF;72.60 |
| S13_vf_87873 | 97.58 | d__Bacteria;p__Bacteroidota;c__Bacteroidia;o__Bacteroidales;f__Tannerellaceae;g__Marseille-P3160 | 98.4 | CRISPR;98.80 iPHoP-RF;72.60 |
| S13_vs_113593 | 40.01 | d__Bacteria;p__Actinobacteriota;c__Actinomycetia;o__Actinomycetales;f__Bifidobacteriaceae;g__Bifidobacterium | 91.5 | blast;93.80 iPHoP-RF;88.20 CRISPR;56.60 |
| S13_vs_121307 | 11.04 | d__Bacteria;p__Firmicutes;c__Clostridia;o__Lachnospirales;f__Lachnospiraceae;g__Blautia_A | 95.9 | CRISPR;89.80 RaFAH;85.80 iPHoP-RF;84.50 |
| S13_vs_13129 | 16.24 | d__Bacteria;p__Firmicutes;c__Clostridia;o__Oscillospirales;f__Oscillospiraceae;g__CAG-83 | 98.5 | CRISPR;98.90 blast;95.20 iPHoP-RF;87.90 |
| S13_vs_148294 | 57.95 | d__Bacteria;p__Firmicutes;c__Clostridia;o__Oscillospirales;f__Oscillospiraceae;g__ | 92.3 | blast;94.50 iPHoP-RF;80.60 |
| S13_vs_148294 | 57.95 | d__Bacteria;p__Firmicutes;c__Clostridia;o__Oscillospirales;f__Oscillospiraceae;g__HGM12998 | 91 | blast;93.40 iPHoP-RF;71.30 |
| S13_vs_148294 | 57.95 | d__Bacteria;p__Firmicutes;c__Clostridia;o__Oscillospirales;f__Oscillospiraceae;g__Flavonifractor | 90 | blast;92.50 iPHoP-RF;67.70 |
| S13_vs_149860 | 61.18 | d__Bacteria;p__Actinobacteriota;c__Actinomycetia;o__Actinomycetales;f__Bifidobacteriaceae;g__Bifidobacterium | 99.7 | CRISPR;99.70 iPHoP-RF;92.40 blast;92.00 |
| S13_vs_15645 | 92 | d__Bacteria;p__Bacteroidota;c__Bacteroidia;o__Bacteroidales;f__Tannerellaceae;g__Parabacteroides | 99.9 | CRISPR;99.20 iPHoP-RF;95.10 blast;93.30 RaFAH;90.50 |
| S13_vs_26756 | 9.7 | d__Bacteria;p__Firmicutes;c__Bacilli;o__RF39;f__UBA660;g__CAG-460 | 94 | blast;95.80 iPHoP-RF;53.30 |
| S13_vs_30101 | 54.46 | d__Bacteria;p__Firmicutes;c__Bacilli;o__ML615J-28;f__CAG-313;g__CAG-313 | 95.6 | blast;96.90 iPHoP-RF;81.60 |
| S13_vs_35816 | 99.57 | d__Bacteria;p__Firmicutes;c__Clostridia;o__Oscillospirales;f__Oscillospiraceae;g__CAG-83 | 99.7 | RaFAH;99.00 blast;82.80 |
| S13_vs_35816 | 99.57 | d__Bacteria;p__Firmicutes;c__Clostridia;o__Oscillospirales;f__Oscillospiraceae;g__Dysosmobacter | 98.7 | RaFAH;99.00 |
| S13_vs_45488 | 22.02 | d__Bacteria;p__Proteobacteria;c__Alphaproteobacteria;o__RF32;f__CAG-239;g__51-20 | 97.9 | CRISPR;98.50 blast;79.90 iPHoP-RF;74.60 |
| S13_vs_45488 | 22.02 | d__Bacteria;p__Proteobacteria;c__Alphaproteobacteria;o__RF32;f__CAG-239;g__CAG-267 | 95 | blast;96.50 iPHoP-RF;72.20 |
| S13_vs_57267 | 38.42 | d__Bacteria;p__Bacteroidota;c__Bacteroidia;o__Bacteroidales;f__Bacteroidaceae;g__Prevotella | 90.6 | RaFAH;93.00 |
| S13_vs_67429 | 14.26 | d__Bacteria;p__Firmicutes;c__Bacilli;o__RF39;f__UBA660;g__CAG-460 | 94.7 | blast;96.30 iPHoP-RF;53.80 |
| S13_vs_83447 | 97.87 | d__Bacteria;p__Bacteroidota;c__Bacteroidia;o__Bacteroidales;f__Rikenellaceae;g__Alistipes | 99.9 | blast;96.80 RaFAH;96.50 iPHoP-RF;82.80 CRISPR;82.60 |
| S13_vs_83447 | 97.87 | d__Bacteria;p__Bacteroidota;c__Bacteroidia;o__Bacteroidales;f__Rikenellaceae;g__Alistipes_A | 95 | RaFAH;96.50 |
| S13_vs_90967 | | d__Bacteria;p__Bacteroidota;c__Bacteroidia;o__Bacteroidales;f__Barnesiellaceae;g__Barnesiella | 90.3 | blast;92.80 iPHoP-RF;81.90 |
| S13_vs_9438 | 7.33 | d__Bacteria;p__Proteobacteria;c__Alphaproteobacteria;o__RF32;f__CAG-239;g__CAG-267 | 91.6 | blast;93.90 iPHoP-RF;83.30 |
| S14_vf_106156 | 21.19 | d__Bacteria;p__Firmicutes;c__Bacilli;o__Lactobacillales;f__Lactobacillaceae;g__Lactobacillus | 100 | RaFAH;98.90 CRISPR;96.80 iPHoP-RF;96.10 blast;95.50 |
| S14_vf_106156 | 21.19 | d__Bacteria;p__Firmicutes;c__Bacilli;o__Lactobacillales;f__Lactobacillaceae;g__Lacticaseibacillus | 98.5 | RaFAH;98.90 |
| S14_vf_106156 | 21.19 | d__Bacteria;p__Firmicutes;c__Bacilli;o__Lactobacillales;f__Lactobacillaceae;g__Ligilactobacillus | 98.5 | RaFAH;98.90 |
| S14_vf_106156 | 21.19 | d__Bacteria;p__Firmicutes;c__Bacilli;o__Lactobacillales;f__Lactobacillaceae;g__Limosilactobacillus | 98.5 | RaFAH;98.90 |
| S14_vf_109545 | 12.74 | d__Bacteria;p__Firmicutes;c__Clostridia;o__Oscillospirales;f__Oscillospiraceae;g__Intestinimonas | 90.8 | blast;93.20 iPHoP-RF;79.50 |
| S14_vf_115736 | 85.01 | d__Bacteria;p__Firmicutes;c__Clostridia;o__Oscillospirales;f__Oscillospiraceae;g__Flavonifractor | 91.2 | blast;93.60 iPHoP-RF;89.80 CRISPR;61.00 |
| S14_vf_115736 | 85.01 | d__Bacteria;p__Firmicutes;c__Clostridia;o__Oscillospirales;f__Oscillospiraceae;g__Intestinimonas | 91.2 | blast;93.60 |
| S14_vf_117147 | 30.35 | d__Bacteria;p__Firmicutes;c__Clostridia;o__Oscillospirales;f__Ruminococcaceae;g__Gemmiger | 95.3 | blast;96.70 CRISPR;94.60 iPHoP-RF;86.30 |
| S14_vf_117150 | 99.65 | d__Bacteria;p__Firmicutes;c__Clostridia;o__Lachnospirales;f__Lachnospiraceae;g__Lachnospira | 99.9 | CRISPR;98.90 RaFAH;91.00 blast;87.20 iPHoP-RF;83.10 |
| S14_vf_117150 | 99.65 | d__Bacteria;p__Firmicutes;c__Clostridia;o__Lachnospirales;f__Lachnospiraceae;g__Eubacterium_G | 91.8 | RaFAH;91.00 blast;67.20 |
| S14_vf_117155 | 63.17 | d__Bacteria;p__Actinobacteriota;c__Actinomycetia;o__Actinomycetales;f__Actinomycetaceae;g__Pauljensenia | 98.4 | RaFAH;94.80 CRISPR;83.10 iPHoP-RF;64.80 |
| S14_vf_117155 | 63.17 | d__Bacteria;p__Actinobacteriota;c__Actinomycetia;o__Actinomycetales;f__Actinomycetaceae;g__Actinomyces | 92.7 | RaFAH;94.80 |
| S14_vf_117157 | 99.57 | d__Bacteria;p__Firmicutes;c__Bacilli;o__Lactobacillales;f__Streptococcaceae;g__Streptococcus | 100 | RaFAH;99.40 CRISPR;98.80 iPHoP-RF;94.40 blast;93.60 |
| S14_vf_117162 | 23.32 | d__Bacteria;p__Bacteroidota;c__Bacteroidia;o__Bacteroidales;f__Bacteroidaceae;g__Bacteroides | 94 | RaFAH;90.00 iPHoP-RF;70.90 CRISPR;63.50 |
| S14_vf_37500 | 5.58 | d__Bacteria;p__Bacteroidota;c__Bacteroidia;o__Bacteroidales;f__Bacteroidaceae;g__Phocaeicola | 94.3 | CRISPR;96.00 iPHoP-RF;67.00 |
| S14_vf_51134 | 47.7 | d__Bacteria;p__Firmicutes;c__Clostridia;o__Oscillospirales;f__Ruminococcaceae;g__Ruminococcus_D | 97.2 | blast;97.90 iPHoP-RF;92.80 RaFAH;52.60 |
| S14_vf_56130 | 100 | d__Bacteria;p__Firmicutes;c__Clostridia;o__Clostridiales;f__Clostridiaceae;g__Clostridium | 98.8 | RaFAH;99.10 |
| S14_vf_56130 | 100 | d__Bacteria;p__Firmicutes;c__Clostridia;o__Clostridiales;f__Clostridiaceae;g__Clostridium_F | 98.8 | RaFAH;99.10 |
| S14_vf_56130 | 100 | d__Bacteria;p__Firmicutes;c__Clostridia;o__Clostridiales;f__Clostridiaceae;g__Clostridium_P | 98.8 | RaFAH;99.10 |
| S14_vf_57689 | 94.04 | d__Bacteria;p__Actinobacteriota;c__Actinomycetia;o__Actinomycetales;f__Actinomycetaceae;g__Actinomyces | 99 | blast;94.00 RaFAH;93.30 CRISPR;80.00 iPHoP-RF;75.40 |

| | | | | |
|---|---|---|---|---|
| S14_vf_57689 | 94.04 | d__Bacteria;p__Actinobacteriota;c__Actinomycetia;o__Actinomycetales;f__Actinomycetaceae;g__Pauljensenia | 90.9 | RaFAH;93.30 |
| S14_vf_58919 | 74.74 | d__Bacteria;p__Firmicutes;c__Clostridia;o__Lachnospirales;f__Lachnospiraceae;g__Roseburia | 95.3 | blast;90.40 iPHoP-RF;87.90 RaFAH;82.70 |
| S14_vf_58919 | 74.74 | d__Bacteria;p__Firmicutes;c__Clostridia;o__Lachnospirales;f__Lachnospiraceae;g__Agathobacter | 95.2 | blast;90.20 RaFAH;82.70 iPHoP-RF;78.30 |
| S14_vf_58919 | 74.74 | d__Bacteria;p__Firmicutes;c__Clostridia;o__Lachnospirales;f__Lachnospiraceae;g__COE1 | 90.4 | blast;92.90 iPHoP-RF;81.40 |
| S14_vf_72335 | 11.73 | d__Bacteria;p__Firmicutes;c__Bacilli;o__Erysipelotrichales;f__Erysipelotrichaceae;g__Holdemania | 92.1 | CRISPR;94.30 |
| S14_vf_72953 | 32.12 | d__Bacteria;p__Firmicutes;c__Bacilli;o__Lactobacillales;f__Lactobacillaceae;g__Lacticaseibacillus | 96.7 | RaFAH;97.70 |
| S14_vf_72953 | 32.12 | d__Bacteria;p__Firmicutes;c__Bacilli;o__Lactobacillales;f__Lactobacillaceae;g__Lactobacillus | 96.7 | RaFAH;97.70 |
| S14_vf_72953 | 32.12 | d__Bacteria;p__Firmicutes;c__Bacilli;o__Lactobacillales;f__Lactobacillaceae;g__Ligilactobacillus | 96.7 | RaFAH;97.70 |
| S14_vf_72953 | 32.12 | d__Bacteria;p__Firmicutes;c__Bacilli;o__Lactobacillales;f__Lactobacillaceae;g__Limosilactobacillus | 96.7 | RaFAH;97.70 |
| S14_vf_72953 | 32.12 | d__Bacteria;p__Firmicutes;c__Bacilli;o__Lactobacillales;f__Lactobacillaceae;g__Pediococcus | 91.8 | blast;94.10 iPHoP-RF;93.40 CRISPR;56.30 |
| S14_vf_78139 | 74.71 | d__Bacteria;p__Firmicutes;c__Bacilli;o__Lactobacillales;f__Lactobacillaceae;g__Lactobacillus | 100 | RaFAH;99.10 blast;96.80 CRISPR;96.50 iPHoP-RF;92.80 |
| S14_vf_78139 | 74.71 | d__Bacteria;p__Firmicutes;c__Bacilli;o__Lactobacillales;f__Lactobacillaceae;g__Lacticaseibacillus | 98.8 | RaFAH;99.10 |
| S14_vf_78139 | 74.71 | d__Bacteria;p__Firmicutes;c__Bacilli;o__Lactobacillales;f__Lactobacillaceae;g__Ligilactobacillus | 98.8 | RaFAH;99.10 |
| S14_vf_78139 | 74.71 | d__Bacteria;p__Firmicutes;c__Bacilli;o__Lactobacillales;f__Lactobacillaceae;g__Limosilactobacillus | 98.8 | RaFAH;99.10 |
| S14_vf_85122 | 16.16 | d__Bacteria;p__Firmicutes;c__Clostridia;o__Clostridiales;f__Clostridiaceae;g__Clostridium | 95 | blast;92.30 iPHoP-RF;86.00 RaFAH;77.50 |
| S14_vf_92240 | 99.79 | d__Bacteria;p__Firmicutes;c__Clostridia;o__Lachnospirales;f__Lachnospiraceae;g__Coprococcus | 93.2 | CRISPR;95.20 |
| S14_vf_93134 | 12.04 | d__Bacteria;p__Firmicutes;c__Clostridia;o__Lachnospirales;f__Lachnospiraceae;g__Muricomes | 96.7 | CRISPR;97.70 |
| S14_vf_99344 | 10.5 | d__Bacteria;p__Firmicutes;c__Clostridia;o__Lachnospirales;f__Lachnospiraceae;g__Eubacterium_F | 96.9 | CRISPR;97.80 |
| S14_vs_100227 | 81.21 | d__Bacteria;p__Firmicutes;c__Clostridia;o__Lachnospirales;f__Lachnospiraceae;g__Enterocloster | 94.2 | CRISPR;95.90 blast;94.00 iPHoP-RF;86.30 |
| S14_vs_100227 | 81.21 | d__Bacteria;p__Firmicutes;c__Clostridia;o__Lachnospirales;f__Lachnospiraceae;g__Hungatella_A | 91 | iPHoP-RF;93.40 |
| S14_vs_101482 | 6.74 | d__Bacteria;p__Firmicutes;c__Clostridia;o__Oscillospirales;f__Ruminococcaceae;g__QAMM01 | 92 | blast;94.20 iPHoP-RF;69.10 |
| S14_vs_102698 | 99.62 | d__Bacteria;p__Firmicutes;c__Clostridia;o__Acetivibrionales;f__Acetivibrionaceae;g__Ruminiclostridium | 90.1 | RaFAH;92.60 |
| S14_vs_112068 | 27.86 | d__Bacteria;p__Firmicutes;c__Clostridia;o__Oscillospirales;f__Butyricicoccaceae;g__Agathobaculum | 97.5 | blast;98.20 iPHoP-RF;66.00 |
| S14_vs_112345 | 71.03 | d__Bacteria;p__Bacteroidota;c__Bacteroidia;o__Bacteroidales;f__Bacteroidaceae;g__Bacteroides | 99.1 | iPHoP-RF;96.70 RaFAH;88.90 blast;75.90 |
| S14_vs_112345 | 71.03 | d__Bacteria;p__Bacteroidota;c__Bacteroidia;o__Bacteroidales;f__Bacteroidaceae;g__Phocaeicola | 95.6 | blast;96.90 iPHoP-RF;95.70 |
| S14_vs_11510 | 39.46 | d__Bacteria;p__Bacteroidota;c__Bacteroidia;o__Bacteroidales;f__Bacteroidaceae;g__Bacteroides | 96.9 | blast;91.70 RaFAH;87.00 iPHoP-RF;81.40 |
| S14_vs_115377 | 76.83 | d__Bacteria;p__Firmicutes;c__Clostridia;o__Lachnospirales;f__Lachnospiraceae;g__Blautia | 98.8 | CRISPR;97.10 RaFAH;83.90 iPHoP-RF;60.90 |
| S14_vs_115377 | 76.83 | d__Bacteria;p__Firmicutes;c__Clostridia;o__Lachnospirales;f__Lachnospiraceae;g__Hespellia | 94 | blast;95.80 iPHoP-RF;66.80 |
| S14_vs_115455 | 25.58 | d__Bacteria;p__Firmicutes;c__Clostridia;o__Eubacteriales;f__Eubacteriaceae;g__Eubacterium | 93 | blast;95.00 CRISPR;89.70 iPHoP-RF;84.50 |
| S14_vs_117153 | 96.78 | d__Bacteria;p__Firmicutes;c__Clostridia;o__Lachnospirales;f__Lachnospiraceae;g__Dorea_A | 94.2 | blast;95.90 iPHoP-RF;94.70 CRISPR;58.40 |
| S14_vs_117153 | 96.78 | d__Bacteria;p__Firmicutes;c__Clostridia;o__Lachnospirales;f__Lachnospiraceae;g__Dorea | 90.6 | blast;93.00 iPHoP-RF;68.60 |
| S14_vs_117166 | 22.09 | d__Bacteria;p__Firmicutes;c__Clostridia;o__Oscillospirales;f__Oscillospiraceae;g__Evtepia | 94.2 | blast;95.90 iPHoP-RF;66.30 |
| S14_vs_14737 | 33.52 | d__Bacteria;p__Firmicutes;c__Clostridia;o__Oscillospirales;f__Ruminococcaceae;g__Ruthenibacterium | 93.1 | blast;95.10 iPHoP-RF;68.40 |
| S14_vs_15228 | 86.47 | d__Bacteria;p__Bacteroidota;c__Bacteroidia;o__Bacteroidales;f__Tannerellaceae;g__Parabacteroides | 100 | CRISPR;99.00 blast;98.60 RaFAH;97.90 iPHoP-RF;96.10 |
| S14_vs_25852 | 26.78 | d__Bacteria;p__Bacteroidota;c__Bacteroidia;o__Bacteroidales;f__Bacteroidaceae;g__Bacteroides | 97.8 | blast;96.60 iPHoP-RF;95.10 CRISPR;93.00 RaFAH;77.10 |
| S14_vs_25852 | 26.78 | d__Bacteria;p__Bacteroidota;c__Bacteroidia;o__Bacteroidales;f__Bacteroidaceae;g__Phocaeicola | 90.1 | blast;92.60 CRISPR;86.70 |
| S14_vs_26320 | 65.93 | d__Bacteria;p__Firmicutes;c__Clostridia;o__Lachnospirales;f__Lachnospiraceae;g__Ruminococcus_B | 95.2 | blast;96.60 iPHoP-RF;82.20 |
| S14_vs_2870 | 56.13 | d__Bacteria;p__Proteobacteria;c__Gammaproteobacteria;o__Burkholderiales;f__Burkholderiaceae;g__Sutterella | 99 | blast;96.90 RaFAH;87.10 CRISPR;78.30 iPHoP-RF;69.50 |
| S14_vs_30108 | 19.83 | d__Bacteria;p__Firmicutes;c__Clostridia;o__Lachnospirales;f__Lachnospiraceae;g__Enterocloster | 92.3 | blast;94.50 CRISPR;82.40 iPHoP-RF;78.30 |
| S14_vs_32499 | 51.73 | d__Bacteria;p__Bacteroidota;c__Bacteroidia;o__Bacteroidales;f__Bacteroidaceae;g__Bacteroides | 99.1 | blast;98.10 iPHoP-RF;96.70 RaFAH;82.60 |
| S14_vs_3251 | 82.87 | d__Bacteria;p__Firmicutes;c__Clostridia;o__Oscillospirales;f__Oscillospiraceae;g__Flavonifractor | 95.6 | blast;96.90 iPHoP-RF;65.60 |
| S14_vs_38905 | 9.01 | d__Bacteria;p__Firmicutes;c__Clostridia;o__Oscillospirales;f__Ruminococcaceae;g__Ruthenibacterium | 93.8 | blast;95.60 iPHoP-RF;81.60 CRISPR;54.30 |
| S14_vs_39266 | 15.51 | d__Bacteria;p__Firmicutes;c__Clostridia;o__Oscillospirales;f__Acutalibacteraceae;g__Ruminococcus_E | 98.7 | CRISPR;99.00 blast;96.80 iPHoP-RF;93.70 |
| S14_vs_39797 | 47.42 | d__Bacteria;p__Firmicutes;c__Clostridia;o__Lachnospirales;f__Lachnospiraceae;g__Anaerostipes | 93.8 | blast;95.60 iPHoP-RF;67.00 |
| S14_vs_56251 | 20.56 | d__Bacteria;p__Firmicutes;c__Clostridia;o__Lachnospirales;f__Lachnospiraceae;g__Muricomes | 95.3 | iPHoP-RF;96.70 blast;93.30 CRISPR;68.70 |
| S14_vs_56410 | 80.58 | d__Bacteria;p__Firmicutes;c__Clostridia;o__Oscillospirales;f__Oscillospiraceae;g__Flavonifractor | 95.2 | blast;96.60 iPHoP-RF;95.40 CRISPR;94.00 |
| S14_vs_59347 | 29.43 | d__Bacteria;p__Firmicutes;c__Clostridia;o__Lachnospirales;f__Lachnospiraceae;g__Muricomes | 93.1 | iPHoP-RF;95.10 |
| S14_vs_59347 | 29.43 | d__Bacteria;p__Firmicutes;c__Clostridia;o__Lachnospirales;f__Lachnospiraceae;g__Ruminococcus_B | 90.7 | iPHoP-RF;93.10 |
| S14_vs_61530 | 22.32 | d__Bacteria;p__Firmicutes;c__Clostridia;o__Oscillospirales;f__Acutalibacteraceae;g__UBA1417 | 92 | blast;94.20 iPHoP-RF;82.80 |
| S14_vs_632 | 78.66 | d__Bacteria;p__Firmicutes;c__Clostridia;o__Clostridiales;f__Clostridiaceae;g__Clostridium | 91 | RaFAH;93.40 |
| S14_vs_632 | 78.66 | d__Bacteria;p__Firmicutes;c__Clostridia;o__Clostridiales;f__Clostridiaceae;g__Clostridium_F | 91 | RaFAH;93.40 |
| S14_vs_632 | 78.66 | d__Bacteria;p__Firmicutes;c__Clostridia;o__Clostridiales;f__Clostridiaceae;g__Clostridium_P | 91 | RaFAH;93.40 |
| S14_vs_66139 | 83.75 | d__Bacteria;p__Firmicutes;c__Clostridia;o__Oscillospirales;f__Acutalibacteraceae;g__Ruminococcus_E | 95.6 | blast;96.90 iPHoP-RF;85.70 |
| S14_vs_66208 | 100 | d__Bacteria;p__Bacteroidota;c__Bacteroidia;o__Bacteroidales;f__Bacteroidaceae;g__Bacteroides | 100 | RaFAH;98.30 iPHoP-RF;96.40 blast;93.70 |
| S14_vs_66841 | 15.26 | d__Bacteria;p__Firmicutes;c__Clostridia;o__Lachnospirales;f__Lachnospiraceae;g__Agathobacter | 90.4 | blast;92.90 iPHoP-RF;78.00 |
| S14_vs_67094 | 10 | d__Bacteria;p__Firmicutes;c__Clostridia;o__Oscillospirales;f__Oscillospiraceae;g__Lawsonibacter | 90.7 | blast;93.10 |
| S14_vs_70985 | 21.93 | d__Bacteria;p__Bacteroidota;c__Bacteroidia;o__Bacteroidales;f__Bacteroidaceae;g__Bacteroides | 99 | iPHoP-RF;96.70 blast;92.60 RaFAH;88.40 |
| S14_vs_73301 | 15.25 | d__Bacteria;p__Firmicutes;c__Clostridia;o__Oscillospirales;f__Ruminococcaceae;g__Massilimaliae | 94.2 | blast;95.90 iPHoP-RF;64.80 |
| S14_vs_74067 | 100 | d__Bacteria;p__Bacteroidota;c__Bacteroidia;o__Bacteroidales;f__Bacteroidaceae;g__Bacteroides | 99.6 | RaFAH;93.20 blast;92.00 iPHoP-RF;82.20 |
| S14_vs_78164 | 61.84 | d__Bacteria;p__Bacteroidota;c__Bacteroidia;o__Bacteroidales;f__Marinifilaceae;g__Odoribacter | 95.4 | blast;96.80 iPHoP-RF;84.80 CRISPR;56.80 |
| S14_vs_88666 | 32.05 | d__Bacteria;p__Firmicutes;c__Clostridia;o__Oscillospirales;f__Acutalibacteraceae;g__Fimivicinus | 95.4 | blast;96.80 iPHoP-RF;86.70 |
| S14_vs_88666 | 32.05 | d__Bacteria;p__Firmicutes;c__Clostridia;o__Oscillospirales;f__Acutalibacteraceae;g__UBA1691 | 95.4 | blast;96.80 CRISPR;90.80 iPHoP-RF;86.70 |
| S14_vs_90934 | 62.33 | d__Bacteria;p__Firmicutes;c__Clostridia;o__Oscillospirales;f__Ruminococcaceae;g__Faecalibacterium | 99.1 | blast;96.70 RaFAH;89.00 iPHoP-RF;81.10 |
| S14_vs_96286 | 99.54 | d__Bacteria;p__Firmicutes;c__Clostridia;o__Lachnospirales;f__Lachnospiraceae;g__Blautia | 96.3 | RaFAH;97.40 |
| S14_vs_96286 | 99.54 | d__Bacteria;p__Firmicutes;c__Clostridia;o__Lachnospirales;f__Lachnospiraceae;g__Blautia_A | 96.3 | RaFAH;97.40 |
| S14_vs_96286 | 99.54 | d__Bacteria;p__Firmicutes;c__Clostridia;o__Lachnospirales;f__Lachnospiraceae;g__Mediterraneibacter | 95.6 | blast;96.90 iPHoP-RF;55.70 |
| S14_vs_9676 | 24.59 | d__Bacteria;p__Firmicutes;c__Clostridia;o__Oscillospirales;f__Oscillospiraceae;g__Lawsonibacter | 90.7 | blast;93.10 iPHoP-RF;77.50 |
| S14_vs_97835 | | d__Bacteria;p__Firmicutes;c__Clostridia;o__Oscillospirales;f__Oscillospiraceae;g__CAG-83 | 90.6 | blast;93.00 iPHoP-RF;84.20 |
| S15_vf_2032 | 83.93 | d__Bacteria;p__Bacteroidota;c__Bacteroidia;o__Bacteroidales;f__Bacteroidaceae;g__Bacteroides | 99.3 | iPHoP-RF;97.10 blast;96.20 CRISPR;91.10 RaFAH;89.30 |
| S15_vf_2032 | 83.93 | d__Bacteria;p__Bacteroidota;c__Bacteroidia;o__Bacteroidales;f__Bacteroidaceae;g__Phocaeicola | 95.9 | iPHoP-RF;97.10 blast;89.90 CRISPR;76.20 |
| S15_vf_20644 | 83.74 | d__Bacteria;p__Bacteroidota;c__Bacteroidia;o__Bacteroidales;f__Rikenellaceae;g__Alistipes | 94.3 | RaFAH;96.00 |
| S15_vf_20644 | 83.74 | d__Bacteria;p__Bacteroidota;c__Bacteroidia;o__Bacteroidales;f__Rikenellaceae;g__Alistipes_A | 94.3 | RaFAH;96.00 |
| S15_vf_2513 | 25.17 | d__Bacteria;p__Firmicutes;c__Clostridia;o__Lachnospirales;f__Lachnospiraceae;g__Lachnospira | 94.5 | CRISPR;96.10 |
| S15_vf_75924 | 99.22 | d__Bacteria;p__Bacteroidota;c__Bacteroidia;o__Bacteroidales;f__Bacteroidaceae;g__Bacteroides | 99.6 | RaFAH;93.20 blast;91.90 iPHoP-RF;82.50 |
| S15_vs_18274 | 75.03 | d__Bacteria;p__Bacteroidota;c__Bacteroidia;o__Bacteroidales;f__Coprobacteraceae;g__Coprobacter | 98.7 | CRISPR;99.00 blast;93.10 iPHoP-RF;64.00 RaFAH;50.00 |
| S15_vs_27510 | 51.41 | d__Bacteria;p__Bacteroidota;c__Bacteroidia;o__Bacteroidales;f__Bacteroidaceae;g__Bacteroides | 99.1 | iPHoP-RF;96.10 blast;93.90 RaFAH;90.50 |
| S15_vs_27510 | 51.41 | d__Bacteria;p__Bacteroidota;c__Bacteroidia;o__Bacteroidales;f__Bacteroidaceae;g__Phocaeicola | 94.9 | iPHoP-RF;96.40 blast;68.30 |

| | | | |
|---|---|---|---|
| S15_vs_35445 | 27.45 | d__Bacteria;p__Bacteroidota;c__Bacteroidia;o__Bacteroidales;f__Bacteroidaceae;g__Bacteroides | 98.8 iPHoP-RF;96.70 blast;93.90 RaFAH;86.60 |
| S15_vs_3868 | 100 | d__Bacteria;p__Bacteroidota;c__Bacteroidia;o__Bacteroidales;f__Bacteroidaceae;g__Bacteroides | 100 RaFAH;99.10 blast;96.50 iPHoP-RF;61.40 |
| S15_vs_3868 | 100 | d__Bacteria;p__Bacteroidota;c__Bacteroidia;o__Bacteroidales;f__Tannerellaceae;g__Parabacteroides | 93 blast;95.00 iPHoP-RF;70.60 CRISPR;68.20 |
| S15_vs_42043 | 51.36 | d__Bacteria;p__Bacteroidota;c__Bacteroidia;o__Bacteroidales;f__Bacteroidaceae;g__Bacteroides | 97.6 blast;96.80 iPHoP-RF;95.10 RaFAH;72.80 |
| S15_vs_42043 | 51.36 | d__Bacteria;p__Bacteroidota;c__Bacteroidia;o__Bacteroidales;f__Bacteroidaceae;g__Phocaeicola | 94.9 iPHoP-RF;96.40 blast;88.20 |
| S15_vs_43913 | 99.8 | d__Bacteria;p__Bacteroidota;c__Bacteroidia;o__Bacteroidales;f__Bacteroidaceae;g__Bacteroides | 100 RaFAH;99.30 iPHoP-RF;97.10 blast;96.70 |
| S15_vs_71968 | 100 | d__Bacteria;p__Bacteroidota;c__Bacteroidia;o__Bacteroidales;f__Rikenellaceae;g__Alistipes | 100 RaFAH;98.90 blast;98.40 iPHoP-RF;95.40 |
| S15_vs_71968 | 100 | d__Bacteria;p__Bacteroidota;c__Bacteroidia;o__Bacteroidales;f__Rikenellaceae;g__Alistipes_A | 98.5 RaFAH;98.90 |
| S16_vf_100473 | 8.42 | d__Bacteria;p__Actinobacteriota;c__Actinomycetia;o__Actinomycetales;f__Bifidobacteriaceae;g__Bifidobacterium | 90.3 blast;92.80 |
| S16_vf_1037 | 73.36 | d__Bacteria;p__Firmicutes;c__Clostridia;o__Lachnospirales;f__Lachnospiraceae;g__Lachnospira | 99.7 CRISPR;99.70 iPHoP-RF;94.40 blast;93.60 |
| S16_vf_108554 | 18.88 | d__Bacteria;p__Firmicutes;c__Clostridia;o__Oscillospirales;f__Ruminococcaceae;g__Faecalibacterium | 97.9 CRISPR;98.50 blast;93.10 iPHoP-RF;92.10 |
| S16_vf_16182 | 64.12 | d__Bacteria;p__Actinobacteriota;c__Actinomycetia;o__Actinomycetales;f__Actinomycetaceae;g__Pauljensenia | 96.9 RaFAH;95.10 CRISPR;68.30 iPHoP-RF;64.80 |
| S16_vf_16182 | 64.12 | d__Bacteria;p__Actinobacteriota;c__Actinomycetia;o__Actinomycetales;f__Actinomycetaceae;g__Actinomyces | 93.1 RaFAH;95.10 |
| S16_vf_169895 | 11.84 | d__Bacteria;p__Firmicutes;c__Clostridia;o__Oscillospirales;f__Ruminococcaceae;g__Ruthenibacterium | 92.7 CRISPR;94.80 |
| S16_vf_172897 | 32.49 | d__Bacteria;p__Actinobacteriota;c__Actinomycetia;o__Actinomycetales;f__Bifidobacteriaceae;g__Bifidobacterium | 98.4 CRISPR;98.80 iPHoP-RF;97.10 blast;93.30 |
| S16_vf_173099 | 55.52 | d__Bacteria;p__Firmicutes;c__Clostridia;o__Oscillospirales;f__Oscillospiraceae;g__Flavonifractor | 93.8 blast;92.00 iPHoP-RF;86.00 RaFAH;72.50 |
| S16_vf_173109 | 7.33 | d__Bacteria;p__Actinobacteriota;c__Actinomycetia;o__Actinomycetales;f__Bifidobacteriaceae;g__Bifidobacterium | 97 CRISPR;97.90 iPHoP-RF;81.60 |
| S16_vf_36146 | 99.74 | d__Bacteria;p__Firmicutes;c__Clostridia;o__Clostridiales;f__Clostridiaceae;g__Clostridium | 96.2 RaFAH;97.30 |
| S16_vf_36146 | 99.74 | d__Bacteria;p__Firmicutes;c__Clostridia;o__Clostridiales;f__Clostridiaceae;g__Clostridium_F | 96.2 RaFAH;97.30 |
| S16_vf_36146 | 99.74 | d__Bacteria;p__Firmicutes;c__Clostridia;o__Clostridiales;f__Clostridiaceae;g__Clostridium_P | 96.2 RaFAH;97.30 |
| S16_vf_5809 | 87.43 | d__Bacteria;p__Firmicutes;c__Clostridia;o__Lachnospirales;f__Lachnospiraceae;g__Lachnospira | 99.9 CRISPR;99.00 iPHoP-RF;95.40 blast;93.40 RaFAH;91.70 |
| S16_vf_6102 | 38.14 | d__Bacteria;p__Firmicutes;c__Clostridia;o__Oscillospirales;f__Oscillospiraceae;g__ER4 | 91.1 blast;93.50 iPHoP-RF;66.80 |
| S16_vf_84462 | 31.67 | d__Bacteria;p__Firmicutes;c__Clostridia;o__Lachnospirales;f__Anaerotignaceae;g__Anaerotignum | 94.2 blast;95.90 iPHoP-RF;79.50 |
| S16_vf_87190 | 34.33 | d__Bacteria;p__Bacteroidota;c__Bacteroidia;o__Bacteroidales;f__Bacteroidaceae;g__Bacteroides | 95 RaFAH;87.80 iPHoP-RF;77.80 CRISPR;68.10 |
| S16_vf_90328 | 16.6 | d__Bacteria;p__Firmicutes;c__Clostridia;o__TANB77;f__CAG-508;g__CAG-269 | 91.4 blast;93.70 iPHoP-RF;53.00 |
| S16_vf_9270 | 8.58 | d__Bacteria;p__Bacteroidota;c__Bacteroidia;o__Bacteroidales;f__Rikenellaceae;g__Alistipes | 98.4 CRISPR;98.80 iPHoP-RF;95.10 |
| S16_vf_9270 | 8.58 | d__Bacteria;p__Bacteroidota;c__Bacteroidia;o__Bacteroidales;f__Bacteroidaceae;g__Bacteroides | 95.9 RaFAH;97.10 |
| S16_vf_96483 | 29.87 | d__Bacteria;p__Firmicutes;c__Clostridia;o__Lachnospirales;f__Lachnospiraceae;g__Blautia_A | 97.6 iPHoP-RF;93.40 blast;93.30 RaFAH;87.30 |
| S16_vf_97279 | 52.37 | d__Bacteria;p__Firmicutes;c__Clostridia;o__Oscillospirales;f__Oscillospiraceae;g__ER4 | 91 blast;93.40 iPHoP-RF;60.30 |
| S16_vs_101072 | 26.56 | d__Bacteria;p__Firmicutes;c__Clostridia;o__Lachnospirales;f__Lachnospiraceae;g__CAG-127 | 95 blast;96.50 iPHoP-RF;87.30 |
| S16_vs_103726 | 26.95 | d__Bacteria;p__Firmicutes;c__Clostridia;o__Oscillospirales;f__Ruminococcaceae;g__Ruthenibacterium | 94 blast;95.80 CRISPR;80.80 iPHoP-RF;79.80 |
| S16_vs_107235 | 98.85 | d__Bacteria;p__Firmicutes;c__Clostridia;o__Oscillospirales;f__Ruminococcaceae;g__Ruminococcus_D | 99.7 blast;99.00 iPHoP-RF;97.10 CRISPR;94.60 RaFAH;84.50 |
| S16_vs_116956 | 43.56 | d__Bacteria;p__Firmicutes;c__Clostridia;o__Lachnospirales;f__Lachnospiraceae;g__Acetatifactor | 91.6 blast;93.90 iPHoP-RF;75.20 |
| S16_vs_118029 | 43.23 | d__Bacteria;p__Firmicutes;c__Clostridia;o__Lachnospirales;f__Lachnospiraceae;g__Eubacterium_G | 94.6 blast;96.20 iPHoP-RF;85.70 |
| S16_vs_120149 | 12.05 | d__Bacteria;p__Firmicutes;c__Clostridia;o__Oscillospirales;f__Acutalibacteraceae;g__Eubacterium_R | 94.5 iPHoP-RF;96.10 blast;93.90 |
| S16_vs_120622 | 63.91 | d__Bacteria;p__Proteobacteria;c__Gammaproteobacteria;o__Burkholderiales;f__Burkholderiaceae;g__Parasutterella | 97.2 CRISPR;98.00 blast;96.90 iPHoP-RF;93.10 |
| S16_vs_127669 | 99.82 | d__Bacteria;p__Bacteroidota;c__Bacteroidia;o__Bacteroidales;f__Bacteroidaceae;g__Bacteroides | 100 RaFAH;99.00 CRISPR;97.60 iPHoP-RF;97.10 blast;90.90 |
| S16_vs_127669 | 99.82 | d__Bacteria;p__Bacteroidota;c__Bacteroidia;o__Bacteroidales;f__Bacteroidaceae;g__Phocaeicola | 96.6 CRISPR;97.60 iPHoP-RF;97.10 blast;94.60 |
| S16_vs_127905 | 55.65 | d__Bacteria;p__Bacteroidota;c__Bacteroidia;o__Bacteroidales;f__Bacteroidaceae;g__Bacteroides | 99.4 iPHoP-RF;97.10 blast;95.20 RaFAH;91.80 CRISPR;60.90 |
| S16_vs_127905 | 55.65 | d__Bacteria;p__Bacteroidota;c__Bacteroidia;o__Bacteroidales;f__Bacteroidaceae;g__Phocaeicola | 95.9 iPHoP-RF;97.10 blast;81.50 |
| S16_vs_132979 | 31.86 | d__Bacteria;p__Firmicutes;c__Clostridia;o__Oscillospirales;f__Ruminococcaceae;g__Faecalibacterium | 94.3 blast;96.00 iPHoP-RF;93.10 |
| S16_vs_136935 | 99.58 | d__Bacteria;p__Bacteroidota;c__Bacteroidia;o__Bacteroidales;f__Bacteroidaceae;g__Bacteroides | 100 RaFAH;98.20 iPHoP-RF;96.70 blast;89.50 |
| S16_vs_136935 | 99.58 | d__Bacteria;p__Bacteroidota;c__Bacteroidia;o__Bacteroidales;f__Bacteroidaceae;g__Phocaeicola | 97.6 CRISPR;98.30 blast;96.20 iPHoP-RF;95.70 |
| S16_vs_140709 | 84.15 | d__Bacteria;p__Firmicutes;c__Clostridia;o__Lachnospirales;f__Lachnospiraceae;g__CAG-317 | 94.5 iPHoP-RF;96.10 |
| S16_vs_140709 | 84.15 | d__Bacteria;p__Firmicutes;c__Clostridia;o__Lachnospirales;f__Lachnospiraceae;g__Bariatricus | 93.5 iPHoP-RF;95.40 blast;93.80 |
| S16_vs_140709 | 84.15 | d__Bacteria;p__Firmicutes;c__Clostridia;o__Lachnospirales;f__Lachnospiraceae;g__Dorea_B | 93.5 iPHoP-RF;95.40 |
| S16_vs_140709 | 84.15 | d__Bacteria;p__Firmicutes;c__Clostridia;o__Lachnospirales;f__Lachnospiraceae;g__Lachnoclostridium_B | 92.6 iPHoP-RF;94.70 |
| S16_vs_141218 | 10.25 | d__Bacteria;p__Firmicutes;c__Bacilli;o__ML615J-28;f__CAG-313;g__CAG-313 | 98.7 CRISPR;99.00 blast;96.10 |
| S16_vs_142561 | 99.95 | d__Bacteria;p__Bacteroidota;c__Bacteroidia;o__Bacteroidales;f__Rikenellaceae;g__Alistipes_A | 97.8 blast;98.20 RaFAH;56.20 iPHoP-RF;53.30 |
| S16_vs_142561 | 99.95 | d__Bacteria;p__Bacteroidota;c__Bacteroidia;o__Bacteroidales;f__Rikenellaceae;g__Alistipes | 96 blast;96.80 RaFAH;56.20 iPHoP-RF;50.90 |
| S16_vs_15093 | 19.11 | d__Bacteria;p__Firmicutes;c__Clostridia;o__Christensenellales;f__CAG-74;g__UBA11524 | 99.6 CRISPR;99.60 blast;96.60 iPHoP-RF;85.70 |
| S16_vs_165662 | 11.88 | d__Bacteria;p__Firmicutes;c__Bacilli;o__RF39;f__UBA660;g__CAG-302 | 92.7 blast;94.80 |
| S16_vs_173079 | 99.1 | d__Bacteria;p__Firmicutes;c__Clostridia;o__Lachnospirales;f__Lachnospiraceae;g__Coprococcus | 94 blast;95.80 iPHoP-RF;71.70 |
| S16_vs_173096 | 74.56 | d__Bacteria;p__Verrucomicrobiota;c__Verrucomicrobiae;o__Verrucomicrobiales;f__Akkermansiaceae;g__Akkermansia | 100 RaFAH;99.70 CRISPR;99.60 blast;96.80 iPHoP-RF;81.10 |
| S16_vs_173141 | | d__Bacteria;p__Firmicutes;c__Clostridia;o__Oscillospirales;f__Ruminococcaceae;g__Faecalibacterium | 96 CRISPR;97.20 iPHoP-RF;82.80 |
| S16_vs_17493 | 18.24 | d__Bacteria;p__Firmicutes;c__Clostridia;o__Oscillospirales;f__Ruminococcaceae;g__Massilimaliae | 93 blast;95.00 iPHoP-RF;58.40 |
| S16_vs_18440 | 67.51 | d__Bacteria;p__Firmicutes;c__Clostridia;o__Oscillospirales;f__Ruminococcaceae;g__Faecalibacterium | 100 RaFAH;99.30 CRISPR;98.90 blast;93.60 iPHoP-RF;85.70 |
| S16_vs_19968 | 93.01 | d__Bacteria;p__Bacteroidota;c__Bacteroidia;o__Bacteroidales;f__Rikenellaceae;g__Alistipes | 98.4 iPHoP-RF;97.10 blast;96.90 RaFAH;79.60 |
| S16_vs_20060 | 33.58 | d__Bacteria;p__Firmicutes;c__Clostridia;o__Oscillospirales;f__Oscillospiraceae;g__UMGS1766 | 91 blast;93.40 |
| S16_vs_32764 | 17.71 | d__Bacteria;p__Bacteroidota;c__Bacteroidia;o__Bacteroidales;f__Rikenellaceae;g__Alistipes | 95.9 iPHoP-RF;97.10 blast;96.90 |
| S16_vs_33281 | 7.09 | d__Bacteria;p__Bacteroidota;c__Bacteroidia;o__Bacteroidales;f__Rikenellaceae;g__Alistipes | 95.9 iPHoP-RF;97.10 blast;96.90 |
| S16_vs_33281 | 7.09 | d__Bacteria;p__Bacteroidota;c__Bacteroidia;o__Bacteroidales;f__Bacteroidaceae;g__Bacteroides | 92.9 RaFAH;94.90 |
| S16_vs_34021 | 99.81 | d__Bacteria;p__Firmicutes;c__Clostridia;o__Oscillospirales;f__Ruminococcaceae;g__Ruminococcus_D | 99 blast;96.90 iPHoP-RF;90.50 RaFAH;86.90 |
| S16_vs_38863 | 26.24 | d__Bacteria;p__Firmicutes;c__Clostridia;o__Monoglobales;f__Monoglobaceae;g__Monoglobus | 95 blast;96.50 iPHoP-RF;64.50 |
| S16_vs_4662 | 13.68 | d__Bacteria;p__Firmicutes;c__Clostridia;o__Christensenellales;f__CAG-138;g__UBA5394 | 92.6 CRISPR;94.70 blast;92.60 iPHoP-RF;74.30 |
| S16_vs_48714 | 5.9 | d__Bacteria;p__Firmicutes;c__Clostridia;o__Oscillospirales;f__Ruminococcaceae;g__Ruminococcus_D | 97 blast;97.90 iPHoP-RF;97.10 CRISPR;96.60 |
| S16_vs_51901 | 16.94 | d__Bacteria;p__Bacteroidota;c__Bacteroidia;o__Bacteroidales;f__Rikenellaceae;g__Alistipes | 97.6 blast;93.90 iPHoP-RF;90.10 RaFAH;85.80 |
| S16_vs_75108 | 32.65 | d__Bacteria;p__Firmicutes;c__Clostridia;o__Oscillospirales;f__Oscillospiraceae;g__UBA5446 | 95 blast;96.50 CRISPR;82.40 iPHoP-RF;65.30 |
| S16_vs_76823 | 20.02 | d__Bacteria;p__Firmicutes;c__Clostridia;o__Oscillospirales;f__Ruminococcaceae;g__Gemmiger | 98.5 CRISPR;98.90 blast;98.30 iPHoP-RF;94.40 |
| S16_vs_78361 | 75.01 | d__Bacteria;p__Actinobacteriota;c__Actinomycetia;o__Actinomycetales;f__Bifidobacteriaceae;g__Bifidobacterium | 98.5 CRISPR;98.90 iPHoP-RF;97.10 blast;96.90 |
| S16_vs_78429 | 22.27 | d__Bacteria;p__Firmicutes;c__Clostridia;o__Oscillospirales;f__Oscillospiraceae;g__ER4 | 91.5 blast;93.80 iPHoP-RF;52.80 |
| S16_vs_80244 | 99.97 | d__Bacteria;p__Bacteroidota;c__Bacteroidia;o__Bacteroidales;f__Bacteroidaceae;g__Bacteroides | 99.7 iPHoP-RF;97.10 RaFAH;95.20 blast;89.20 |
| S16_vs_80244 | 99.97 | d__Bacteria;p__Bacteroidota;c__Bacteroidia;o__Bacteroidales;f__Bacteroidaceae;g__Phocaeicola | 92.2 iPHoP-RF;94.40 blast;93.80 |
| S16_vs_81805 | 99.42 | d__Bacteria;p__Firmicutes;c__Clostridia;o__Oscillospirales;f__Ruminococcaceae;g__Ruthenibacterium | 94.7 blast;96.30 iPHoP-RF;89.50 CRISPR;57.90 |
| S16_vs_82116 | 98.74 | d__Bacteria;p__Firmicutes;c__Clostridia;o__Oscillospirales;f__Acutalibacteraceae;g__Ruminococcus_E | 99.6 CRISPR;98.90 blast;96.90 iPHoP-RF;94.10 RaFAH;81.90 |
| S17_vf_128147 | 3.97 | d__Bacteria;p__Bacteroidota;c__Bacteroidia;o__Bacteroidales;f__Bacteroidaceae;g__Bacteroides | 95.9 blast;97.10 iPHoP-RF;97.10 CRISPR;89.30 |
| S17_vf_128147 | 3.97 | d__Bacteria;p__Bacteroidota;c__Bacteroidia;o__Bacteroidales;f__Bacteroidaceae;g__Bacteroides_G | 94.5 iPHoP-RF;96.10 |

| | | | |
|---|---|---|---|
| S17_vf_128147 | 3.97 | d__Bacteria;p__Bacteroidota;c__Bacteroidia;o__Bacteroidales;f__Bacteroidaceae;g__Phocaeicola | 91 iPHoP-RF;93.40 blast;85.40 |
| S17_vf_140653 | 90.58 | d__Bacteria;p__Bacteroidota;c__Bacteroidia;o__Bacteroidales;f__Bacteroidaceae;g__Bacteroides | 99.7 blast;96.70 RaFAH;95.30 iPHoP-RF;85.70 |
| S17_vf_174033 | 68.79 | d__Bacteria;p__Firmicutes;c__Clostridia;o__Oscillospirales;f__Ruminococcaceae;g__Ruminococcus_D | 99.7 blast;97.00 iPHoP-RF;95.40 RaFAH;94.60 CRISPR;72.00 |
| S17_vf_174033 | 68.79 | d__Bacteria;p__Firmicutes;c__Clostridia;o__Oscillospirales;f__Acutalibacteraceae;g__Ruminococcus_E | 92.5 RaFAH;94.60 |
| S17_vf_35138 | 84.47 | d__Bacteria;p__Bacteroidota;c__Bacteroidia;o__Bacteroidales;f__Barnesiellaceae;g__Barnesiella | 99.1 blast;96.80 RaFAH;88.80 CRISPR;59.60 |
| S17_vf_36452 | 12 | d__Bacteria;p__Firmicutes;c__Clostridia;o__Lachnospirales;f__Lachnospiraceae;g__Muricomes | 97.8 CRISPR;98.40 iPHoP-RF;90.80 |
| S17_vf_46388 | 50.05 | d__Bacteria;p__Firmicutes;c__Clostridia;o__Lachnospirales;f__Lachnospiraceae;g__Blautia_A | 99.4 CRISPR;97.10 blast;94.50 iPHoP-RF;93.70 RaFAH;91.90 |
| S17_vf_81232 | 50.04 | d__Bacteria;p__Bacteroidota;c__Bacteroidia;o__Bacteroidales;f__Tannerellaceae;g__Parabacteroides | 97.8 blast;98.40 iPHoP-RF;95.10 CRISPR;83.70 |
| S17_vf_97671 | 81.31 | d__Bacteria;p__Firmicutes;c__Clostridia;o__Oscillospirales;f__Oscillospiraceae;g__Lawsonibacter | 90.2 blast;92.70 iPHoP-RF;73.20 |
| S17_vs_103009 | 22.09 | d__Bacteria;p__Firmicutes;c__Clostridia;o__Oscillospirales;f__Oscillospiraceae;g__ER4 | 96.7 blast;97.70 iPHoP-RF;65.10 |
| S17_vs_110666 | 98.39 | d__Bacteria;p__Firmicutes;c__Clostridia;o__Oscillospirales;f__Ruminococcaceae;g__Faecalibacterium | 100 RaFAH;99.70 blast;96.80 iPHoP-RF;93.40 CRISPR;67.80 |
| S17_vs_116640 | 83.12 | d__Bacteria;p__Bacteroidota;c__Bacteroidia;o__Bacteroidales;f__Bacteroidaceae;g__Bacteroides | 91 RaFAH;93.40 |
| S17_vs_117507 | 40.08 | d__Bacteria;p__Bacteroidota;c__Bacteroidia;o__Bacteroidales;f__Bacteroidaceae;g__Bacteroides | 97.5 blast;97.40 iPHoP-RF;96.40 RaFAH;66.20 |
| S17_vs_127519 | 61.22 | d__Bacteria;p__Firmicutes;c__Clostridia;o__Oscillospirales;f__Oscillospiraceae;g__Dysosmobacter | 91.2 blast;93.60 iPHoP-RF;86.70 |
| S17_vs_157209 | 25.09 | d__Bacteria;p__Bacteroidota;c__Bacteroidia;o__Bacteroidales;f__Rikenellaceae;g__Alistipes | 97.6 CRISPR;98.30 blast;95.20 iPHoP-RF;92.80 |
| S17_vs_160502 | 60.44 | d__Bacteria;p__Bacteroidota;c__Bacteroidia;o__Bacteroidales;f__Bacteroidaceae;g__Bacteroides | 99.9 iPHoP-RF;97.10 RaFAH;96.60 blast;92.60 CRISPR;56.20 |
| S17_vs_160502 | 60.44 | d__Bacteria;p__Bacteroidota;c__Bacteroidia;o__Bacteroidales;f__Bacteroidaceae;g__Phocaeicola | 95.9 iPHoP-RF;97.10 blast;84.10 |
| S17_vs_177196 | 18.1 | d__Bacteria;p__Bacteroidota;c__Bacteroidia;o__Bacteroidales;f__Rikenellaceae;g__Alistipes | 96.4 blast;91.90 iPHoP-RF;88.50 RaFAH;84.30 |
| S17_vs_177203 | 71.95 | d__Bacteria;p__Desulfobacterota;c__Desulfovibrionia;o__Desulfovibrionales;f__Desulfovibrionaceae;g__Bilophila | 93.9 blast;95.70 iPHoP-RF;90.10 |
| S17_vs_2079 | 67.97 | d__Bacteria;p__Bacteroidota;c__Bacteroidia;o__Bacteroidales;f__Bacteroidaceae;g__Bacteroides | 100 RaFAH;98.90 iPHoP-RF;94.40 blast;92.60 |
| S17_vs_31654 | 36.85 | d__Bacteria;p__Firmicutes;c__Bacilli;o__Erysipelotrichales;f__Erysipelatoclostridiaceae;g__Faecalibacillus | 94.6 blast;96.20 iPHoP-RF;81.60 CRISPR;54.30 |
| S17_vs_39515 | 82.52 | d__Bacteria;p__Bacteroidota;c__Bacteroidia;o__Bacteroidales;f__Bacteroidaceae;g__Bacteroides | 100 RaFAH;99.30 iPHoP-RF;97.10 blast;92.90 |
| S17_vs_39515 | 82.52 | d__Bacteria;p__Bacteroidota;c__Bacteroidia;o__Bacteroidales;f__Bacteroidaceae;g__Phocaeicola | 95.3 iPHoP-RF;96.70 |
| S17_vs_43307 | 72.14 | d__Bacteria;p__Bacteroidota;c__Bacteroidia;o__Bacteroidales;f__Tannerellaceae;g__Parabacteroides | 95.4 blast;96.80 CRISPR;96.70 iPHoP-RF;93.40 |
| S17_vs_8690 | 6.78 | d__Bacteria;p__Proteobacteria;c__Alphaproteobacteria;o__RF32;f__CAG-239;g__CAG-495 | 95.2 blast;96.60 |
| S17_vs_8690 | 6.78 | d__Bacteria;p__Proteobacteria;c__Alphaproteobacteria;o__RF32;f__CAG-239;g__Scatocola | 94.9 blast;96.40 iPHoP-RF;59.50 |
| S1_vf_136526 | 33.32 | d__Bacteria;p__Firmicutes;c__Clostridia;o__Oscillospirales;f__Oscillospiraceae;g__CAG-170 | 91.6 blast;93.90 iPHoP-RF;88.50 |
| S1_vf_158961 | 9.7 | d__Bacteria;p__Firmicutes;c__Clostridia;o__Christensenellales;f__CAG-74;g__UBA11524 | 95 blast;96.50 iPHoP-RF;77.30 |
| S1_vf_159951 | 22.14 | d__Bacteria;p__Bacteroidota;c__Bacteroidia;o__Bacteroidales;f__Rikenellaceae;g__Alistipes | 95.4 CRISPR;96.80 iPHoP-RF;72.20 |
| S1_vf_22149 | 43.65 | d__Bacteria;p__Firmicutes;c__Clostridia;o__Oscillospirales;f__Oscillospiraceae;g__ER4 | 90.8 blast;93.20 iPHoP-RF;88.90 |
| S1_vf_28398 | 12.4 | d__Bacteria;p__Firmicutes;c__Clostridia;o__Oscillospirales;f__Ruminococcaceae;g__Gemmiger | 91.6 blast;93.90 iPHoP-RF;75.40 |
| S1_vf_6272 | 25.47 | d__Bacteria;p__Firmicutes;c__Clostridia;o__Oscillospirales;f__Oscillospiraceae;g__CAG-170 | 92 CRISPR;94.20 blast;90.80 iPHoP-RF;78.00 |
| S1_vs_117317 | 21.23 | d__Bacteria;p__Firmicutes;c__Clostridia;o__Oscillospirales;f__Oscillospiraceae;g__CAG-170 | 93.4 blast;95.30 iPHoP-RF;89.50 CRISPR;71.10 |
| S1_vs_133577 | 37.09 | d__Bacteria;p__Firmicutes;c__Clostridia;o__Oscillospirales;f__Ruminococcaceae;g__Gemmiger | 96 blast;97.20 CRISPR;95.40 iPHoP-RF;94.40 |
| S1_vs_143929 | 42.31 | d__Bacteria;p__Firmicutes;c__Clostridia;o__Oscillospirales;f__Oscillospiraceae;g__ER4 | 91.6 blast;93.90 iPHoP-RF;75.00 |
| S1_vs_145792 | 13.59 | d__Bacteria;p__Firmicutes;c__Clostridia;o__Lachnospirales;f__Anaerotignaceae;g__Anaerotignum | 94.6 blast;96.20 iPHoP-RF;72.60 |
| S1_vs_155183 | 31.14 | d__Bacteria;p__Firmicutes;c__Clostridia;o__Oscillospirales;f__Oscillospiraceae;g__ER4 | 95.9 CRISPR;97.10 blast;96.20 iPHoP-RF;89.20 |
| S1_vs_23682 | 33.26 | d__Bacteria;p__Firmicutes;c__Clostridia;o__Oscillospirales;f__Ruminococcaceae;g__Faecalibacterium | 98.8 CRISPR;99.10 blast;96.20 iPHoP-RF;83.30 |
| S2_vf_16829 | 81.02 | d__Bacteria;p__Actinobacteriota;c__Coriobacteriia;o__Coriobacteriales;f__Eggerthellaceae;g__Eggerthella | 94.9 CRISPR;96.40 iPHoP-RF;53.80 RaFAH;50.20 |
| S2_vf_17850 | 11.78 | d__Bacteria;p__Firmicutes;c__Clostridia;o__Lachnospirales;f__Lachnospiraceae;g__Muricomes | 92.7 CRISPR;94.80 iPHoP-RF;78.70 |
| S2_vf_20910 | 5.26 | d__Bacteria;p__Firmicutes;c__Bacilli;o__Erysipelotrichales;f__Erysipelotrichaceae;g__Clostridium_AQ | 97.2 CRISPR;98.00 |
| S2_vs_10289 | 67.97 | d__Bacteria;p__Bacteroidota;c__Bacteroidia;o__Bacteroidales;f__Bacteroidaceae;g__Bacteroides | 100 RaFAH;98.90 blast;96.80 iPHoP-RF;87.90 |
| S2_vs_7306 | 96.01 | d__Bacteria;p__Bacteroidota;c__Bacteroidia;o__Bacteroidales;f__Tannerellaceae;g__Parabacteroides | 93 blast;95.00 iPHoP-RF;73.70 |
| S2_vs_7306 | 96.01 | d__Bacteria;p__Bacteroidota;c__Bacteroidia;o__Bacteroidales;f__Marinifilaceae;g__Odoribacter | 90 RaFAH;89.40 CRISPR;64.40 |
| S3_vf_100440 | 98.85 | d__Bacteria;p__Firmicutes;c__Clostridia;o__Clostridiales;f__Clostridiaceae;g__Clostridium | 94.7 RaFAH;96.30 |
| S3_vf_100440 | 98.85 | d__Bacteria;p__Firmicutes;c__Clostridia;o__Clostridiales;f__Clostridiaceae;g__Clostridium_F | 94.7 RaFAH;96.30 |
| S3_vf_100440 | 98.85 | d__Bacteria;p__Firmicutes;c__Clostridia;o__Clostridiales;f__Clostridiaceae;g__Clostridium_P | 94.7 RaFAH;96.30 |
| S3_vf_101611 | 23.54 | d__Bacteria;p__Firmicutes;c__Clostridia;o__Monoglobales;f__Monoglobaceae;g__Monoglobus | 95.2 blast;96.60 |
| S3_vf_106401 | 12.56 | d__Bacteria;p__Firmicutes;c__Bacilli;o__Haloplasmatales_A;f__Turicibacteraceae;g__Turicibacter | 92 blast;94.20 iPHoP-RF;66.30 |
| S3_vf_108959 | 42.95 | d__Bacteria;p__Firmicutes;c__Clostridia;o__Lachnospirales;f__Lachnospiraceae;g__Lachnospira | 99.3 CRISPR;99.30 blast;91.90 iPHoP-RF;88.90 RaFAH;55.00 |
| S3_vf_111893 | 41.65 | d__Bacteria;p__Firmicutes;c__Clostridia;o__Oscillospirales;f__Ruminococcaceae;g__Ruminococcus_D | 95.4 blast;96.80 CRISPR;96.10 iPHoP-RF;94.10 |
| S3_vf_11428 | 94.85 | d__Bacteria;p__Firmicutes;c__Clostridia;o__Lachnospirales;f__Lachnospiraceae;g__Blautia_A | 100 RaFAH;98.90 CRISPR;97.00 iPHoP-RF;93.40 blast;91.70 |
| S3_vf_11428 | 94.85 | d__Bacteria;p__Firmicutes;c__Clostridia;o__Lachnospirales;f__Lachnospiraceae;g__Blautia | 98.5 RaFAH;98.90 |
| S3_vf_115304 | 34.34 | d__Bacteria;p__Spirochaetota;c__Spirochaetia;o__Sphaerochaetales;f__Sphaerochaetaceae;g__UBA9732 | 91.2 blast;93.60 iPHoP-RF;81.90 |
| S3_vf_127392 | 52.43 | d__Bacteria;p__Firmicutes;c__Clostridia;o__Oscillospirales;f__CAG-272;g__CAG-272 | 92.1 blast;94.30 iPHoP-RF;52.20 |
| S3_vf_127399 | 23.09 | d__Bacteria;p__Bacteroidota;c__Bacteroidia;o__Bacteroidales;f__Bacteroidaceae;g__Bacteroides | 90.4 RaFAH;92.90 |
| S3_vf_127400 | 54.09 | d__Bacteria;p__Firmicutes;c__Clostridia;o__Oscillospirales;f__Oscillospiraceae;g__ER4 | 95 blast;96.50 iPHoP-RF;76.40 CRISPR;67.10 |
| S3_vf_127405 | 98.09 | d__Bacteria;p__Firmicutes;c__Clostridia;o__Lachnospirales;f__Lachnospiraceae;g__Lachnospira | 99.9 CRISPR;98.90 RaFAH;91.60 iPHoP-RF;90.80 blast;80.40 |
| S3_vf_127405 | 98.09 | d__Bacteria;p__Firmicutes;c__Clostridia;o__Lachnospirales;f__Lachnospiraceae;g__Eubacterium_G | 96.9 RaFAH;91.60 blast;87.20 |
| S3_vf_127406 | 89.48 | d__Bacteria;p__Bacteroidota;c__Bacteroidia;o__Bacteroidales;f__Rikenellaceae;g__Alistipes | 97.8 RaFAH;98.40 |
| S3_vf_127406 | 89.48 | d__Bacteria;p__Bacteroidota;c__Bacteroidia;o__Bacteroidales;f__Rikenellaceae;g__Alistipes_A | 97.8 RaFAH;98.40 |
| S3_vf_127406 | 89.48 | d__Bacteria;p__Bacteroidota;c__Bacteroidia;o__Bacteroidales;f__Bacteroidaceae;g__UBA6398 | 91.6 blast;93.90 iPHoP-RF;84.80 |
| S3_vf_127408 | 6.08 | d__Bacteria;p__Actinobacteriota;c__Actinomycetia;o__Actinomycetales;f__Bifidobacteriaceae;g__Bifidobacterium | 98.5 blast;98.90 CRISPR;97.60 iPHoP-RF;96.70 |
| S3_vf_127409 | 96.84 | d__Bacteria;p__Firmicutes;c__Clostridia;o__Oscillospirales;f__Ruminococcaceae;g__Faecalibacterium | 95.9 CRISPR;97.00 blast;95.70 iPHoP-RF;81.10 RaFAH;51.90 |
| S3_vf_127450 | 8.34 | d__Bacteria;p__Bacteroidota;c__Bacteroidia;o__Bacteroidales;f__Bacteroidaceae;g__Prevotella | 92.3 RaFAH;94.50 |
| S3_vf_18080 | 96.93 | d__Bacteria;p__Bacteroidota;c__Bacteroidia;o__Bacteroidales;f__Bacteroidaceae;g__Bacteroides | 99.7 iPHoP-RF;97.10 blast;96.10 RaFAH;94.50 CRISPR;81.40 |
| S3_vf_18080 | 96.93 | d__Bacteria;p__Bacteroidota;c__Bacteroidia;o__Bacteroidales;f__Bacteroidaceae;g__Phocaeicola | 91.8 iPHoP-RF;94.10 CRISPR;80.00 blast;78.90 |
| S3_vf_18829 | 63.94 | d__Bacteria;p__Bacteroidota;c__Bacteroidia;o__Bacteroidales;f__Bacteroidaceae;g__Bacteroides | 91.8 iPHoP-RF;94.10 CRISPR;73.10 |
| S3_vf_23042 | 83.69 | d__Bacteria;p__Firmicutes;c__Bacilli;o__Lactobacillales;f__Streptococcaceae;g__Lactococcus | 99.7 blast;98.70 iPHoP-RF;90.10 RaFAH;89.50 |
| S3_vf_44405 | 96.43 | d__Bacteria;p__Actinobacteriota;c__Actinomycetia;o__Actinomycetales;f__Actinomycetaceae;g__Actinomyces | 100 RaFAH;97.30 CRISPR;92.40 blast;92.10 iPHoP-RF;55.70 |
| S3_vf_44405 | 96.43 | d__Bacteria;p__Actinobacteriota;c__Actinomycetia;o__Actinomycetales;f__Actinomycetaceae;g__Pauljensenia | 96.2 RaFAH;97.30 |
| S3_vf_54353 | 46.9 | d__Bacteria;p__Actinobacteriota;c__Actinomycetia;o__Actinomycetales;f__Actinomycetaceae;g__Actinomyces | 97.8 RaFAH;97.30 iPHoP-RF;70.60 |
| S3_vf_54353 | 46.9 | d__Bacteria;p__Actinobacteriota;c__Actinomycetia;o__Actinomycetales;f__Actinomycetaceae;g__Pauljensenia | 96.2 RaFAH;97.30 |
| S3_vf_5525 | 41.37 | d__Bacteria;p__Actinobacteriota;c__Actinomycetia;o__Actinomycetales;f__Actinomycetaceae;g__Actinomyces | 92.6 RaFAH;85.90 iPHoP-RF;81.10 |
| S3_vf_56363 | 23.06 | d__Bacteria;p__Firmicutes;c__Clostridia;o__Peptostreptococcales;f__Peptostreptococcaceae;g__GCA-900066495 | 91.4 iPHoP-RF;93.70 |
| S3_vf_67333 | 76.26 | d__Bacteria;p__Firmicutes;c__Clostridia;o__Lachnospirales;f__Lachnospiraceae;g__Agathobacter | 99.9 CRISPR;98.90 RaFAH;90.80 blast;85.70 iPHoP-RF;81.90 |
| S3_vf_67333 | 76.26 | d__Bacteria;p__Firmicutes;c__Clostridia;o__Lachnospirales;f__Lachnospiraceae;g__Roseburia | 99.1 iPHoP-RF;96.10 RaFAH;90.80 blast;90.50 |

| | | | | |
|---|---|---|---|---|
| S3_vf_67333 | 76.26 | d__Bacteria;p__Firmicutes;c__Clostridia;o__Lachnospirales;f__Lachnospiraceae;g__RUG115 | 99 | RaFAH;90.80 blast;90.70 iPHoP-RF;77.50 |
| S3_vf_67333 | 76.26 | d__Bacteria;p__Firmicutes;c__Clostridia;o__Lachnospirales;f__Lachnospiraceae;g__Lachnospira | 98.7 | CRISPR;99.00 blast;90.70 iPHoP-RF;89.80 |
| S3_vf_67333 | 76.26 | d__Bacteria;p__Firmicutes;c__Clostridia;o__Lachnospirales;f__Lachnospiraceae;g__Eubacterium_F | 90.4 | blast;92.90 |
| S3_vf_67828 | | d__Bacteria;p__Bacteroidota;c__Bacteroidia;o__Bacteroidales;f__Rikenellaceae;g__Alistipes | 91.5 | blast;93.80 iPHoP-RF;64.50 |
| S3_vf_70746 | 87.64 | d__Bacteria;p__Actinobacteriota;c__Actinomycetia;o__Actinomycetales;f__Bifidobacteriaceae;g__Bifidobacterium | 95.3 | iPHoP-RF;96.70 blast;95.90 CRISPR;78.50 |
| S3_vf_7137 | 5.31 | d__Bacteria;p__Actinobacteriota;c__Actinomycetia;o__Actinomycetales;f__Bifidobacteriaceae;g__Bifidobacterium | 99.7 | CRISPR;99.70 iPHoP-RF;77.30 |
| S3_vf_73597 | 55.3 | d__Bacteria;p__Bacteroidota;c__Bacteroidia;o__Bacteroidales;f__Bacteroidaceae;g__Bacteroides | 99.7 | CRISPR;97.20 iPHoP-RF;97.10 RaFAH;93.80 blast;93.80 |
| S3_vf_73597 | 55.3 | d__Bacteria;p__Bacteroidota;c__Bacteroidia;o__Bacteroidales;f__Bacteroidaceae;g__Phocaeicola | 95.3 | iPHoP-RF;96.70 |
| S3_vf_80287 | 52.73 | d__Bacteria;p__Firmicutes;c__Clostridia;o__Oscillospirales;f__Ruminococcaceae;g__D5 | 97.2 | CRISPR;98.00 iPHoP-RF;52.20 |
| S3_vf_8058 | 98.11 | d__Bacteria;p__Firmicutes;c__Clostridia;o__Lachnospirales;f__Lachnospiraceae;g__Blautia_A | 98.1 | CRISPR;98.60 iPHoP-RF;81.60 |
| S3_vf_8164 | 10.61 | d__Bacteria;p__Firmicutes;c__Clostridia;o__Oscillospirales;f__Ruminococcaceae;g__Ruminococcus_D | 98.2 | CRISPR;98.70 blast;96.80 iPHoP-RF;95.10 |
| S3_vf_82571 | 21.78 | d__Bacteria;p__Firmicutes;c__Clostridia;o__Oscillospirales;f__Oscillospiraceae;g__CAG-170 | 93.1 | blast;95.10 iPHoP-RF;73.20 |
| S3_vf_99626 | 26.68 | d__Bacteria;p__Actinobacteriota;c__Coriobacteriia;o__Coriobacteriales;f__Coriobacteriaceae;g__Collinsella | 98.7 | CRISPR;99.00 iPHoP-RF;97.10 blast;92.10 |
| S3_vf_99734 | 99.18 | d__Bacteria;p__Firmicutes;c__Clostridia;o__Oscillospirales;f__Ruminococcaceae;g__Faecalibacterium | 95.2 | CRISPR;96.60 iPHoP-RF;64.80 blast;60.50 |
| S3_vf_99734 | 99.18 | d__Bacteria;p__Firmicutes;c__Clostridia;o__Oscillospirales;f__Oscillospiraceae;g__CAG-83 | 92 | RaFAH;94.20 |
| S3_vf_99734 | 99.18 | d__Bacteria;p__Firmicutes;c__Clostridia;o__Oscillospirales;f__Oscillospiraceae;g__Dysosmobacter | 92 | RaFAH;94.20 |
| S3_vs_114271 | 19.01 | d__Bacteria;p__Firmicutes;c__Clostridia;o__Oscillospirales;f__Acutalibacteraceae;g__Acutalibacter | 92.1 | blast;94.30 iPHoP-RF;56.80 |
| S3_vs_117894 | 81.25 | d__Bacteria;p__Bacteroidota;c__Bacteroidia;o__Bacteroidales;f__Marinifilaceae;g__Odoribacter | 99.9 | blast;98.40 iPHoP-RF;93.10 RaFAH;92.30 CRISPR;53.10 |
| S3_vs_119660 | 91.3 | d__Bacteria;p__Bacteroidota;c__Bacteroidia;o__Bacteroidales;f__Bacteroidaceae;g__Bacteroides | 100 | RaFAH;98.50 CRISPR;95.40 iPHoP-RF;95.40 blast;93.90 |
| S3_vs_119660 | 91.3 | d__Bacteria;p__Bacteroidota;c__Bacteroidia;o__Bacteroidales;f__Bacteroidaceae;g__CAG-462 | 96.9 | CRISPR;97.80 |
| S3_vs_119660 | 91.3 | d__Bacteria;p__Bacteroidota;c__Bacteroidia;o__Bacteroidales;f__Bacteroidaceae;g__Phocaeicola | 90.6 | CRISPR;93.00 blast;88.70 iPHoP-RF;75.70 |
| S3_vs_119771 | 27.39 | d__Bacteria;p__Firmicutes;c__Clostridia;o__Oscillospirales;f__Ruminococcaceae;g__Gemmiger | 95.9 | blast;97.10 iPHoP-RF;81.40 CRISPR;55.60 |
| S3_vs_122706 | 89.75 | d__Bacteria;p__Firmicutes;c__Clostridia;o__Oscillospirales;f__Acutalibacteraceae;g__CAG-217 | 95.3 | blast;96.70 iPHoP-RF;79.30 |
| S3_vs_123834 | 73.11 | d__Bacteria;p__Actinobacteriota;c__Actinomycetia;o__Actinomycetales;f__Bifidobacteriaceae;g__Bifidobacterium | 98.4 | CRISPR;98.40 blast;96.90 iPHoP-RF;96.70 RaFAH;61.70 |
| S3_vs_127391 | 18.32 | d__Bacteria;p__Firmicutes;c__Clostridia;o__Oscillospirales;f__Ruminococcaceae;g__Gemmiger | 95.6 | blast;96.90 CRISPR;96.30 iPHoP-RF;91.80 |
| S3_vs_127401 | 45.54 | d__Bacteria;p__Firmicutes;c__Clostridia;o__Lachnospirales;f__Lachnospiraceae;g__Lachnospira | 90.4 | blast;92.90 iPHoP-RF;70.60 |
| S3_vs_127411 | 9.3 | d__Bacteria;p__Firmicutes;c__Clostridia;o__Oscillospirales;f__Oscillospiraceae;g__ER4 | 95.2 | blast;96.60 iPHoP-RF;73.20 |
| S3_vs_127413 | 23.82 | d__Bacteria;p__Bacteroidota;c__Bacteroidia;o__Bacteroidales;f__Rikenellaceae;g__Alistipes | 97.9 | CRISPR;98.50 iPHoP-RF;80.00 |
| S3_vs_127421 | 95.6 | d__Bacteria;p__Firmicutes;c__Clostridia;o__Lachnospirales;f__Lachnospiraceae;g__Agathobacter | 100 | CRISPR;98.90 RaFAH;94.40 iPHoP-RF;92.10 |
| S3_vs_127421 | 95.6 | d__Bacteria;p__Firmicutes;c__Clostridia;o__Lachnospirales;f__Lachnospiraceae;g__Roseburia | 99.9 | CRISPR;98.50 iPHoP-RF;96.40 blast;95.20 RaFAH;94.40 |
| S3_vs_127421 | 95.6 | d__Bacteria;p__Firmicutes;c__Clostridia;o__Lachnospirales;f__Lachnospiraceae;g__RUG115 | 98.2 | RaFAH;94.40 iPHoP-RF;88.50 |
| S3_vs_127421 | 95.6 | d__Bacteria;p__Firmicutes;c__Clostridia;o__Lachnospirales;f__Lachnospiraceae;g__CAG-45 | 90.7 | iPHoP-RF;93.10 |
| S3_vs_127442 | | d__Bacteria;p__Bacteroidota;c__Bacteroidia;o__Bacteroidales;f__P3;g__Phil12 | 90.7 | iPHoP-RF;93.10 |
| S3_vs_15450 | 21.5 | d__Bacteria;p__Firmicutes;c__Clostridia;o__Oscillospirales;f__Ruminococcaceae;g__Gemmiger | 95.4 | blast;96.80 iPHoP-RF;83.90 CRISPR;68.80 |
| S3_vs_16825 | 80.45 | d__Bacteria;p__Firmicutes;c__Clostridia;o__Lachnospirales;f__Lachnospiraceae;g__Roseburia | 99.7 | CRISPR;98.90 iPHoP-RF;96.10 blast;92.80 RaFAH;88.00 |
| S3_vs_16825 | 80.45 | d__Bacteria;p__Firmicutes;c__Clostridia;o__Lachnospirales;f__Lachnospiraceae;g__Agathobacter | 99.4 | CRISPR;98.10 iPHoP-RF;93.40 RaFAH;88.00 |
| S3_vs_16825 | 80.45 | d__Bacteria;p__Firmicutes;c__Clostridia;o__Lachnospirales;f__Lachnospiraceae;g__RUG115 | 97.2 | iPHoP-RF;91.80 blast;90.60 RaFAH;88.00 |
| S3_vs_17168 | 100 | d__Bacteria;p__Bacteroidota;c__Bacteroidia;o__Bacteroidales;f__Rikenellaceae;g__Alistipes | 97.5 | blast;96.90 iPHoP-RF;93.10 RaFAH;71.20 |
| S3_vs_18858 | 82.07 | d__Bacteria;p__Firmicutes;c__Clostridia;o__Oscillospirales;f__Acutalibacteraceae;g__CAG-217 | 97.9 | blast;98.50 iPHoP-RF;86.30 CRISPR;68.10 |
| S3_vs_23194 | 42.15 | d__Bacteria;p__Bacteroidota;c__Bacteroidia;o__Bacteroidales;f__Bacteroidaceae;g__Bacteroides | 100 | RaFAH;98.70 iPHoP-RF;97.10 |
| S3_vs_23194 | 42.15 | d__Bacteria;p__Bacteroidota;c__Bacteroidia;o__Bacteroidales;f__Bacteroidaceae;g__Phocaeicola | 95.3 | iPHoP-RF;96.70 blast;93.10 CRISPR;75.20 |
| S3_vs_25896 | 11.85 | d__Bacteria;p__Firmicutes;c__Clostridia;o__Oscillospirales;f__Ruminococcaceae;g__Ruthenibacterium | 91.4 | blast;93.70 iPHoP-RF;78.30 |
| S3_vs_29077 | 34.15 | d__Bacteria;p__Firmicutes;c__Clostridia;o__Oscillospirales;f__Ruminococcaceae;g__Ruminococcus_C | 94.9 | blast;96.40 iPHoP-RF;80.00 CRISPR;51.40 |
| S3_vs_39774 | 100 | d__Bacteria;p__Bacteroidota;c__Bacteroidia;o__Bacteroidales;f__Bacteroidaceae;g__Bacteroides | 100 | RaFAH;99.00 blast;92.00 iPHoP-RF;53.80 |
| S3_vs_39940 | 5.41 | d__Bacteria;p__Actinobacteriota;c__Actinomycetia;o__Actinomycetales;f__Bifidobacteriaceae;g__Bifidobacterium | 95.9 | iPHoP-RF;97.10 blast;96.90 CRISPR;84.00 |
| S3_vs_40939 | 23.65 | d__Bacteria;p__Bacteroidota;c__Bacteroidia;o__Bacteroidales;f__Bacteroidaceae;g__Bacteroides | 99.3 | iPHoP-RF;97.10 blast;96.60 RaFAH;89.70 |
| S3_vs_40939 | 23.65 | d__Bacteria;p__Bacteroidota;c__Bacteroidia;o__Bacteroidales;f__Bacteroidaceae;g__Phocaeicola | 93.9 | iPHoP-RF;95.70 blast;86.80 |
| S3_vs_42569 | 73.75 | d__Bacteria;p__Firmicutes;c__Clostridia;o__Oscillospirales;f__Ruminococcaceae;g__Faecalibacterium | 99.7 | CRISPR;98.30 blast;96.90 RaFAH;90.40 iPHoP-RF;73.70 |
| S3_vs_45147 | 90.75 | d__Bacteria;p__Bacteroidota;c__Bacteroidia;o__Bacteroidales;f__Bacteroidaceae;g__Bacteroides | 99.4 | iPHoP-RF;97.10 RaFAH;91.20 |
| S3_vs_45147 | 90.75 | d__Bacteria;p__Bacteroidota;c__Bacteroidia;o__Bacteroidales;f__Bacteroidaceae;g__Phocaeicola | 95.9 | iPHoP-RF;97.10 CRISPR;60.90 |
| S3_vs_49835 | 13.1 | d__Bacteria;p__Firmicutes;c__Clostridia;o__Oscillospirales;f__Acutalibacteraceae;g__Ruminococcus_E | 99 | CRISPR;99.20 blast;98.60 iPHoP-RF;93.10 |
| S3_vs_50557 | 23.06 | d__Bacteria;p__Actinobacteriota;c__Coriobacteriia;o__Coriobacteriales;f__Eggerthellaceae;g__Gordonibacter | 93.4 | CRISPR;95.30 blast;93.90 iPHoP-RF;93.10 |
| S3_vs_50924 | 12.69 | d__Bacteria;p__Cyanobacteria;c__Vampirovibrionia;o__Gastranaerophilales;f__Gastranaerophilaceae;g__QAMI01 | 90.4 | blast;92.90 iPHoP-RF;57.10 |
| S3_vs_51931 | 21.18 | d__Bacteria;p__Proteobacteria;c__Gammaproteobacteria;o__Burkholderiales;f__Burkholderiaceae;g__Sutterella | 94.6 | CRISPR;96.20 iPHoP-RF;79.00 |
| S3_vs_54303 | 46.83 | d__Bacteria;p__Firmicutes;c__Clostridia;o__Oscillospirales;f__Oscillospiraceae;g__ | 91.6 | blast;93.90 iPHoP-RF;80.30 |
| S3_vs_56292 | 99.95 | d__Bacteria;p__Bacteroidota;c__Bacteroidia;o__Bacteroidales;f__Rikenellaceae;g__Alistipes | 99.9 | blast;96.90 RaFAH;96.10 iPHoP-RF;83.60 |
| S3_vs_56292 | 99.95 | d__Bacteria;p__Bacteroidota;c__Bacteroidia;o__Bacteroidales;f__Rikenellaceae;g__Alistipes_A | 94.5 | RaFAH;96.10 |
| S3_vs_64424 | 98.41 | d__Bacteria;p__Bacteroidota;c__Bacteroidia;o__Bacteroidales;f__Bacteroidaceae;g__Bacteroides | 100 | RaFAH;99.60 iPHoP-RF;97.10 blast;92.20 CRISPR;69.50 |
| S3_vs_64424 | 98.41 | d__Bacteria;p__Bacteroidota;c__Bacteroidia;o__Bacteroidales;f__Bacteroidaceae;g__Phocaeicola | 95.4 | blast;96.80 iPHoP-RF;96.40 CRISPR;92.60 |
| S3_vs_6604 | 75.2 | d__Bacteria;p__Bacteroidota;c__Bacteroidia;o__Bacteroidales;f__Marinifilaceae;g__Butyricimonas | 95.4 | blast;96.80 iPHoP-RF;65.60 |
| S3_vs_68031 | 27.92 | d__Bacteria;p__Firmicutes;c__Clostridia;o__Oscillospirales;f__Ruminococcaceae;g__Faecalibacterium | 95.6 | blast;96.90 iPHoP-RF;73.20 CRISPR;56.90 |
| S3_vs_70508 | 99.66 | d__Bacteria;p__Bacteroidota;c__Bacteroidia;o__Bacteroidales;f__Bacteroidaceae;g__Bacteroides | 100 | RaFAH;99.70 iPHoP-RF;93.10 blast;92.70 |
| S3_vs_73361 | 17.97 | d__Bacteria;p__Firmicutes;c__Clostridia;o__Christensenellales;f__CAG-138;g__SFEL01 | 96.6 | CRISPR;97.60 blast;96.40 iPHoP-RF;91.10 |
| S3_vs_77156 | | d__Bacteria;p__Bacteroidota;c__Bacteroidia;o__Bacteroidales;f__Rikenellaceae;g__Alistipes | 96.7 | CRISPR;97.70 iPHoP-RF;58.40 |
| S3_vs_80151 | 97.75 | d__Bacteria;p__Bacteroidota;c__Bacteroidia;o__Bacteroidales;f__Tannerellaceae;g__Parabacteroides | 100 | RaFAH;99.60 iPHoP-RF;94.40 blast;93.80 CRISPR;92.60 |
| S3_vs_82895 | 92.58 | d__Bacteria;p__Bacteroidota;c__Bacteroidia;o__Bacteroidales;f__Tannerellaceae;g__Parabacteroides | 100 | CRISPR;99.00 RaFAH;98.20 blast;93.90 iPHoP-RF;93.70 |
| S3_vs_87728 | 5.42 | d__Bacteria;p__Firmicutes;c__Clostridia;o__Oscillospirales;f__Ruminococcaceae;g__Faecalibacterium | 95.3 | blast;96.70 iPHoP-RF;67.00 |
| S3_vs_89485 | 73.42 | d__Bacteria;p__Bacteroidota;c__Bacteroidia;o__Bacteroidales;f__Rikenellaceae;g__Alistipes | 95 | blast;96.50 iPHoP-RF;93.40 |
| S3_vs_90331 | 99.17 | d__Bacteria;p__Firmicutes;c__Clostridia;o__Oscillospirales;f__Ruminococcaceae;g__Faecalibacterium | 100 | RaFAH;98.70 CRISPR;97.70 blast;92.00 iPHoP-RF;85.70 |
| S3_vs_90341 | 11.42 | d__Bacteria;p__Firmicutes;c__Clostridia;o__Christensenellales;f__CAG-74;g__UBA11524 | 95.2 | blast;96.60 CRISPR;94.50 iPHoP-RF;85.70 |
| S3_vs_90341 | 11.42 | d__Bacteria;p__Firmicutes;c__Clostridia;o__Christensenellales;f__CAG-74;g__UBA11524 | 94.9 | blast;96.40 iPHoP-RF;85.10 |
| S3_vs_9158 | 99.96 | d__Bacteria;p__Bacteroidota;c__Bacteroidia;o__Bacteroidales;f__Bacteroidaceae;g__Bacteroides | 100 | RaFAH;99.60 iPHoP-RF;97.10 blast;96.90 |
| S3_vs_9158 | 99.96 | d__Bacteria;p__Bacteroidota;c__Bacteroidia;o__Bacteroidales;f__Bacteroidaceae;g__Phocaeicola | 95.3 | iPHoP-RF;96.70 blast;90.50 |
| S3_vs_94577 | 45.75 | d__Bacteria;p__Firmicutes;c__Bacilli;o__Erysipelotrichales;f__Erysipelatoclostridiaceae;g__Faecalibacillus | 97.8 | blast;98.40 |
| S3_vs_95574 | 22.37 | d__Bacteria;p__Firmicutes;c__Clostridia;o__Oscillospirales;f__Acutalibacteraceae;g__UBA737 | 90.4 | blast;92.90 iPHoP-RF;70.60 |
| S3_vs_97827 | 10.17 | d__Bacteria;p__Bacteroidota;c__Bacteroidia;o__Bacteroidales;f__Marinifilaceae;g__Odoribacter | 97.9 | blast;98.50 iPHoP-RF;90.10 CRISPR;71.50 |
| S3_vs_98675 | 8.18 | d__Bacteria;p__Firmicutes;c__Clostridia;o__Clostridiales;f__Clostridiaceae;g__Clostridium | 98.5 | blast;96.40 RaFAH;85.10 iPHoP-RF;79.00 |

| | | | |
|---|---|---|---|
| S4_vf_10506 | 17.1 | d__Bacteria;p__Firmicutes;c__Bacilli;o__Erysipelotrichales;f__Erysipelotrichaceae;g__Holdemanella | 90.7 blast;93.10 |
| S4_vf_11308 | 96.94 | d__Bacteria;p__Firmicutes;c__Clostridia;o__Oscillospirales;f__Acutalibacteraceae;g__Ruminococcus_E | 100 CRISPR;98.90 blast;98.50 RaFAH;97.10 iPHoP-RF;93.70 |
| S4_vf_11308 | 96.94 | d__Bacteria;p__Firmicutes;c__Clostridia;o__Oscillospirales;f__Ruminococcaceae;g__Ruminococcus_D | 95.9 RaFAH;97.10 |
| S4_vf_12614 | 80.07 | d__Bacteria;p__Bacteroidota;c__Bacteroidia;o__Bacteroidales;f__Barnesiellaceae;g__Barnesiella | 98.7 blast;99.00 iPHoP-RF;76.10 CRISPR;67.90 |
| S4_vf_12624 | 24.52 | d__Bacteria;p__Actinobacteriota;c__Actinomycetia;o__Actinomycetales;f__Bifidobacteriaceae;g__Bifidobacterium | 97.5 CRISPR;98.20 blast;96.50 iPHoP-RF;91.80 |
| S4_vf_13353 | 93.68 | d__Bacteria;p__Firmicutes;c__Clostridia;o__Lachnospirales;f__Lachnospiraceae;g__Blautia_A | 99.9 CRISPR;97.90 RaFAH;95.20 blast;90.90 iPHoP-RF;89.20 |
| S4_vf_13353 | 93.68 | d__Bacteria;p__Firmicutes;c__Clostridia;o__Lachnospirales;f__Lachnospiraceae;g__Blautia | 93.2 RaFAH;95.20 |
| S4_vf_18803 | 47.59 | d__Bacteria;p__Bacteroidota;c__Bacteroidia;o__Bacteroidales;f__Bacteroidaceae;g__Prevotella | 98.5 CRISPR;98.90 iPHoP-RF;74.10 |
| S4_vf_20440 | 44.47 | d__Bacteria;p__Bacteroidota;c__Bacteroidia;o__Bacteroidales;f__Bacteroidaceae;g__Prevotella | 97.8 blast;98.40 iPHoP-RF;84.20 |
| S4_vf_30103 | 5.83 | d__Bacteria;p__Actinobacteriota;c__Actinomycetia;o__Actinomycetales;f__Bifidobacteriaceae;g__Bifidobacterium | 99.7 CRISPR;99.70 iPHoP-RF;92.10 |
| S4_vf_31973 | 23.92 | d__Bacteria;p__Patescibacteria;c__Paceibacteria;o__UBA9983_A;f__UBA1006;g__01-FULL-36-10b | 92.2 iPHoP-RF;94.40 |
| S4_vf_31978 | 62.52 | d__Bacteria;p__Firmicutes;c__Clostridia;o__Lachnospirales;f__Lachnospiraceae;g__Lachnospira | 99.6 CRISPR;99.60 blast;93.90 iPHoP-RF;92.40 |
| S4_vf_31979 | 24.1 | d__Bacteria;p__Firmicutes;c__Clostridia;o__Christensenellales;f__CAG-74;g__Ventricola | 92 blast;94.20 |
| S4_vf_31979 | 24.1 | d__Bacteria;p__Firmicutes;c__Clostridia;o__Christensenellales;f__CAG-74;g__SFFH01 | 91.5 blast;93.80 |
| S4_vf_31981 | 29.33 | d__Bacteria;p__Bacteroidota;c__Bacteroidia;o__Bacteroidales;f__Bacteroidaceae;g__Prevotella | 97.3 RaFAH;98.10 |
| S4_vf_4838 | 76.93 | d__Bacteria;p__Bacteroidota;c__Bacteroidia;o__Bacteroidales;f__Tannerellaceae;g__Parabacteroides | 97.6 blast;98.30 iPHoP-RF;96.10 CRISPR;71.50 |
| S4_vf_4891 | 28.59 | d__Bacteria;p__Bacteroidota;c__Bacteroidia;o__Bacteroidales;f__Bacteroidaceae;g__Prevotella | 94.6 blast;96.20 CRISPR;93.50 iPHoP-RF;92.10 |
| S4_vs_VIRSorter_12831-circular-cat_2 | 21.81 | d__Bacteria;p__Firmicutes;c__Clostridia;o__Lachnospirales;f__Lachnospiraceae;g__Blautia_A | 91.6 blast;93.90 iPHoP-RF;85.10 CRISPR;58.50 |
| S4_vs_VIRSorter_13190-cat_2 | 96.31 | d__Bacteria;p__Bacteroidota;c__Bacteroidia;o__Bacteroidales;f__Tannerellaceae;g__Parabacteroides | 100 RaFAH;99.80 iPHoP-RF;94.40 blast;91.20 CRISPR;66.50 |
| S4_vs_VIRSorter_15112-cat_2 | 16.47 | d__Bacteria;p__Firmicutes;c__Clostridia;o__Peptostreptococcales;f__Peptostreptococcaceae;g__GCA-900066495 | 91.6 blast;93.90 iPHoP-RF;75.70 |
| S4_vs_VIRSorter_15837-cat_2 | 96.3 | d__Bacteria;p__Firmicutes;c__Clostridia;o__Oscillospirales;f__Oscillospiraceae;g__Flavonifractor | 97.2 blast;98.00 CRISPR;89.70 iPHoP-RF;84.80 |
| S4_vs_VIRSorter_16493-cat_2 | 85.89 | d__Bacteria;p__Actinobacteriota;c__Actinomycetia;o__Actinomycetales;f__Bifidobacteriaceae;g__Bifidobacterium | 97.9 CRISPR;98.50 iPHoP-RF;96.40 blast;92.50 |
| S4_vs_VIRSorter_19324-cat_2 | 25.86 | d__Bacteria;p__Firmicutes;c__Clostridia;o__Lachnospirales;f__Lachnospiraceae;g__Mediterraneibacter | 91.4 blast;93.70 iPHoP-RF;76.40 |
| S4_vs_VIRSorter_19324-cat_2 | 25.86 | d__Bacteria;p__Firmicutes;c__Clostridia;o__Lachnospirales;f__Lachnospiraceae;g__Faecalimonas | 91 blast;93.40 iPHoP-RF;75.20 |
| S4_vs_VIRSorter_27316-cat_2 | 74.11 | d__Bacteria;p__Firmicutes;c__Clostridia;o__Oscillospirales;f__Ruminococcaceae;g__Faecalibacterium | 95.4 blast;96.80 iPHoP-RF;75.00 CRISPR;67.80 |
| S4_vs_VIRSorter_2744-cat_3 | 9.58 | d__Bacteria;p__Actinobacteriota;c__Actinomycetia;o__Actinomycetales;f__Bifidobacteriaceae;g__Bifidobacterium | 96 CRISPR;97.20 iPHoP-RF;97.10 blast;96.70 |
| S4_vs_VIRSorter_3006-cat_1 | 99.88 | d__Bacteria;p__Firmicutes;c__Clostridia;o__Lachnospirales;f__Lachnospiraceae;g__Mediterraneibacter | 91.6 blast;93.90 |
| S4_vs_VIRSorter_30222-cat_2 | 65.61 | d__Bacteria;p__Firmicutes;c__Clostridia;o__Oscillospirales;f__Oscillospiraceae;g__Intestinimonas | 91.5 blast;93.80 |
| S4_vs_VIRSorter_31171-circular-cat_2 | 46.28 | d__Bacteria;p__Firmicutes;c__Clostridia;o__Oscillospirales;f__Ruminococcaceae;g__QAMM01 | 90.2 blast;92.70 iPHoP-RF;54.90 |
| S4_vs_VIRSorter_31196-circular-cat_2 | 93.39 | d__Bacteria;p__Firmicutes;c__Clostridia;o__Oscillospirales;f__Oscillospiraceae;g__Flavonifractor | 99.4 CRISPR;97.90 blast;96.20 iPHoP-RF;88.90 RaFAH;87.70 |
| S4_vs_VIRSorter_31560-circular-cat_2 | 59.79 | d__Bacteria;p__Firmicutes;c__Clostridia;o__Oscillospirales;f__Oscillospiraceae;g__Flavonifractor | 90.3 blast;92.80 iPHoP-RF;86.30 CRISPR;64.80 |
| S4_vs_VIRSorter_31916-circular-cat_1 | 39.89 | d__Bacteria;p__Firmicutes;c__Clostridia;o__Lachnospirales;f__Lachnospiraceae;g__Mediterraneibacter | 90.7 blast;93.10 iPHoP-RF;68.20 |
| S4_vs_VIRSorter_31972-circular-cat_2 | 26.41 | d__Bacteria;p__Firmicutes;c__Clostridia;o__Oscillospirales;f__Ruminococcaceae;g__Ruminococcus_C | 97.3 blast;98.10 CRISPR;93.20 iPHoP-RF;90.50 |
| S4_vs_VIRSorter_31975-circular-cat_1 | 88.33 | d__Bacteria;p__Bacteroidota;c__Bacteroidia;o__Bacteroidales;f__Bacteroidaceae;g__Bacteroides | 99.6 iPHoP-RF;97.10 blast;96.60 CRISPR;94.50 RaFAH;92.70 |
| S4_vs_VIRSorter_31975-circular-cat_1 | 88.33 | d__Bacteria;p__Bacteroidota;c__Bacteroidia;o__Bacteroidales;f__Bacteroidaceae;g__Phocaeicola | 95.9 iPHoP-RF;97.10 blast;90.60 CRISPR;74.40 |
| S4_vs_VIRSorter_31976-circular-cat_2 | 17.17 | d__Bacteria;p__Firmicutes;c__Clostridia;o__Oscillospirales;f__Ruminococcaceae;g__Gemmiger | 97.2 CRISPR;98.00 blast;96.20 iPHoP-RF;93.70 |
| S4_vs_VIRSorter_31980-circular-cat_2 | 13.26 | d__Bacteria;p__Proteobacteria;c__Gammaproteobacteria;o__Burkholderiales;f__Burkholderiaceae;g__Duodenibacillus | 96.7 blast;97.70 CRISPR;83.90 iPHoP-RF;70.60 |
| S4_vs_VIRSorter_4746-cat_2 | 5.45 | d__Bacteria;p__Firmicutes;c__Clostridia;o__Lachnospirales;f__Lachnospiraceae;g__Roseburia | 90.3 iPHoP-RF;92.80 |
| S4_vs_VIRSorter_4839-cat_2 | 69.64 | d__Bacteria;p__Firmicutes;c__Clostridia;o__Oscillospirales;f__Ruminococcaceae;g__Massilimaliae | 91.4 blast;93.70 iPHoP-RF;75.70 |
| S4_vs_VIRSorter_4959-cat_2 | 50.34 | d__Bacteria;p__Firmicutes;c__Clostridia;o__Oscillospirales;f__Oscillospiraceae;g__ER4 | 94.2 blast;95.90 iPHoP-RF;82.50 |
| S4_vs_VIRSorter_5698-cat_2 | 87.05 | d__Bacteria;p__Actinobacteriota;c__Actinomycetia;o__Actinomycetales;f__Bifidobacteriaceae;g__Bifidobacterium | 99 CRISPR;98.50 blast;96.80 iPHoP-RF;96.10 RaFAH;71.90 |
| S4_vs_VIRSorter_6022-circular-cat_1 | 97.71 | d__Bacteria;p__Bacteroidota;c__Bacteroidia;o__Bacteroidales;f__Tannerellaceae;g__Marseille-P3160 | 98.4 CRISPR;98.80 iPHoP-RF;65.30 |
| S4_vs_VIRSorter_6022-circular-cat_1 | 97.71 | d__Bacteria;p__Bacteroidota;c__Bacteroidia;o__Bacteroidales;f__Marinifilaceae;g__Odoribacter | 97.3 RaFAH;98.10 |
| S4_vs_VIRSorter_9913-cat_1 | 98.3 | d__Bacteria;p__Bacteroidota;c__Bacteroidia;o__Bacteroidales;f__Marinifilaceae;g__Odoribacter | 99.7 CRISPR;97.90 RaFAH;92.60 |
| S5_vf_103206 | 3.75 | d__Bacteria;p__Firmicutes;c__Clostridia;o__Oscillospirales;f__Oscillospiraceae;g__Lawsonibacter | 90 blast;92.50 |
| S5_vf_105728 | 44.2 | d__Bacteria;p__Firmicutes;c__Clostridia;o__Lachnospirales;f__Lachnospiraceae;g__ | 94.5 iPHoP-RF;96.10 blast;93.40 |
| S5_vf_105728 | 44.2 | d__Bacteria;p__Firmicutes;c__Clostridia;o__Lachnospirales;f__Lachnospiraceae;g__UC5-1-2E3 | 93.1 iPHoP-RF;95.10 blast;91.90 |
| S5_vf_116300 | 13.55 | d__Bacteria;p__Firmicutes;c__Clostridia;o__Lachnospirales;f__Lachnospiraceae;g__Anaerostipes | 95.2 blast;96.60 iPHoP-RF;90.50 CRISPR;69.90 |
| S5_vf_134040 | 100 | d__Bacteria;p__Firmicutes;c__Clostridia;o__Oscillospirales;f__Ruminococcaceae;g__Faecalibacterium | 100 RaFAH;98.80 blast;96.50 iPHoP-RF;69.10 |
| S5_vf_136076 | 43.14 | d__Bacteria;p__Firmicutes;c__Clostridia;o__Oscillospirales;f__Oscillospiraceae;g__ER4 | 93 blast;95.00 iPHoP-RF;69.80 |
| S5_vf_137260 | 6.74 | d__Bacteria;p__Actinobacteriota;c__Coriobacteriia;o__Coriobacteriales;f__Coriobacteriaceae;g__Collinsella | 92.9 CRISPR;94.90 |
| S5_vf_145579 | 44.59 | d__Bacteria;p__Bacteroidota;c__Bacteroidia;o__Bacteroidales;f__Bacteroidaceae;g__Prevotella | 94.6 RaFAH;96.20 |
| S5_vf_145583 | 10.99 | d__Bacteria;p__Firmicutes;c__Clostridia;o__Clostridiales;f__Clostridiaceae;g__Clostridium | 95.2 blast;93.60 iPHoP-RF;80.80 RaFAH;73.30 CRISPR;62.00 |
| S5_vf_145585 | 9.85 | d__Bacteria;p__Firmicutes;c__Clostridia;o__Lachnospirales;f__Lachnospiraceae;g__Enterocloster | 91.2 blast;93.60 iPHoP-RF;91.40 CRISPR;53.80 |
| S5_vf_145588 | 34.8 | d__Bacteria;p__Firmicutes;c__Clostridia;o__Lachnospirales;f__Lachnospiraceae;g__Dorea_A | 97.2 CRISPR;98.00 iPHoP-RF;64.30 |
| S5_vf_145604 | 87.41 | d__Bacteria;p__Firmicutes;c__Clostridia;o__Lachnospirales;f__Lachnospiraceae;g__Blautia_A | 99.6 CRISPR;97.30 RaFAH;93.30 blast;92.90 iPHoP-RF;91.80 |
| S5_vf_145604 | 87.41 | d__Bacteria;p__Firmicutes;c__Clostridia;o__Lachnospirales;f__Lachnospiraceae;g__Blautia | 90.9 RaFAH;93.30 |
| S5_vf_18461 | 99.96 | d__Bacteria;p__Bacteroidota;c__Bacteroidia;o__Bacteroidales;f__Rikenellaceae;g__Alistipes | 92.7 RaFAH;92.00 blast;67.20 |
| S5_vf_21869 | 31.69 | d__Bacteria;p__Firmicutes;c__Clostridia;o__Oscillospirales;f__Oscillospiraceae;g__Flavonifractor | 93.1 blast;95.10 iPHoP-RF;73.90 |
| S5_vf_25423 | 81 | d__Bacteria;p__Firmicutes;c__Bacilli;o__Lactobacillales;f__Streptococcaceae;g__Lactococcus | 100 RaFAH;98.80 blast;98.50 iPHoP-RF;91.40 |
| S5_vf_28061 | 20.02 | d__Bacteria;p__Firmicutes;c__Bacilli;o__Haloplasmatales_A;f__Turicibacteraceae;g__Turicibacter | 91.5 blast;93.80 iPHoP-RF;68.60 |
| S5_vf_29332 | 12.73 | d__Bacteria;p__Actinobacteriota;c__Coriobacteriia;o__Coriobacteriales;f__Coriobacteriaceae;g__Collinsella | 97.3 CRISPR;98.10 blast;94.60 iPHoP-RF;79.50 |
| S5_vf_55687 | 91.19 | d__Bacteria;p__Firmicutes;c__Bacilli;o__Lactobacillales;f__Streptococcaceae;g__Lactococcus | 90.2 RaFAH;92.70 |
| S5_vf_60597 | 35.7 | d__Bacteria;p__Firmicutes;c__Clostridia;o__Lachnospirales;f__Anaerotignaceae;g__Anaerotignum | 93.9 blast;95.70 iPHoP-RF;84.20 |
| S5_vf_6506 | 20.08 | d__Bacteria;p__Firmicutes;c__Clostridia;o__Clostridiales;f__Clostridiaceae;g__Clostridium_J | 90.3 blast;92.80 iPHoP-RF;70.90 |
| S5_vf_74661 | 91.73 | d__Bacteria;p__Proteobacteria;c__Gammaproteobacteria;o__Enterobacterales;f__Enterobacteriaceae;g__Escherichia | 100 RaFAH;99.40 iPHoP-RF;97.10 blast;96.80 CRISPR;85.00 |
| S5_vf_8808 | 97.29 | d__Bacteria;p__Firmicutes;c__Negativicutes;o__Veillonellales;f__Veillonellaceae;g__Veillonella | 100 CRISPR;98.80 RaFAH;98.60 blast;93.70 iPHoP-RF;78.30 |
| S5_vf_8808 | 97.29 | d__Bacteria;p__Firmicutes;c__Negativicutes;o__Veillonellales;f__Veillonellaceae;g__F0422 | 98.1 RaFAH;98.60 |
| S5_vf_8808 | 97.29 | d__Bacteria;p__Firmicutes;c__Negativicutes;o__Veillonellales;f__Veillonellaceae;g__Veillonella_A | 98.1 RaFAH;98.60 |
| S5_vf_8965 | 100 | d__Bacteria;p__Bacteroidota;c__Bacteroidia;o__Bacteroidales;f__Bacteroidaceae;g__Bacteroides | 100 RaFAH;99.60 iPHoP-RF;97.10 blast;96.30 |
| S5_vf_8965 | 100 | d__Bacteria;p__Bacteroidota;c__Bacteroidia;o__Bacteroidales;f__Bacteroidaceae;g__Phocaeicola | 95.9 iPHoP-RF;97.10 |
| S5_vs_10428 | 10.64 | d__Bacteria;p__Firmicutes;c__Clostridia;o__Christensenellales;f__CAG-74;g__Alectryocaccomicrobium | 91.4 blast;93.70 |
| S5_vs_107105 | 21.85 | d__Bacteria;p__Firmicutes;c__Clostridia;o__Oscillospirales;f__Oscillospiraceae;g__CAG-110 | 94.7 blast;96.30 iPHoP-RF;67.00 |
| S5_vs_107105 | 21.85 | d__Bacteria;p__Firmicutes;c__Clostridia;o__Oscillospirales;f__Oscillospiraceae;g__Faecousia | 90.8 blast;93.20 iPHoP-RF;64.50 |
| S5_vs_109527 | 23.09 | d__Bacteria;p__Bacteroidota;c__Bacteroidia;o__Bacteroidales;f__Bacteroidaceae;g__Bacteroides | 99 iPHoP-RF;96.40 blast;92.90 RaFAH;89.10 |
| S5_vs_109527 | 23.09 | d__Bacteria;p__Bacteroidota;c__Bacteroidia;o__Bacteroidales;f__Bacteroidaceae;g__Phocaeicola | 92.6 iPHoP-RF;94.70 blast;89.10 |
| S5_vs_117319 | 51.09 | d__Bacteria;p__Firmicutes;c__Clostridia;o__Oscillospirales;f__Ruminococcaceae;g__Faecalibacterium | 98.4 blast;95.50 RaFAH;86.90 CRISPR;78.90 iPHoP-RF;76.80 |

| | | | | |
|---|---|---|---|---|
| S5_vs_117417 | 13.57 | d__Bacteria;p__Firmicutes;c__Clostridia;o__Oscillospirales;f__Oscillospiraceae;g__Enterenecus | 90.7 | blast;93.10 iPHoP-RF;70.90 |
| S5_vs_118168 | 30.01 | d__Bacteria;p__Firmicutes;c__Clostridia;o__Oscillospirales;f__Ruminococcaceae;g__Anaerotruncus | 91 | blast;93.40 iPHoP-RF;57.30 |
| S5_vs_120079 | 88.37 | d__Bacteria;p__Firmicutes;c__Clostridia;o__Lachnospirales;f__Lachnospiraceae;g__Roseburia | 95.3 | blast;96.70 iPHoP-RF;96.40 CRISPR;84.10 |
| S5_vs_120079 | 88.37 | d__Bacteria;p__Firmicutes;c__Clostridia;o__Lachnospirales;f__Lachnospiraceae;g__VUNI01 | 91 | iPHoP-RF;93.40 blast;89.80 |
| S5_vs_120584 | 98.83 | d__Bacteria;p__Firmicutes;c__Clostridia;o__Lachnospirales;f__Lachnospiraceae;g__Lachnospira | 97.5 | CRISPR;98.20 blast;93.50 iPHoP-RF;88.90 |
| S5_vs_121411 | 100 | d__Bacteria;p__Firmicutes;c__Clostridia;o__Oscillospirales;f__Ruminococcaceae;g__Ruthenibacterium | 95.3 | blast;96.70 iPHoP-RF;87.60 CRISPR;68.40 |
| S5_vs_122344 | 26.84 | d__Bacteria;p__Firmicutes;c__Clostridia;o__Lachnospirales;f__Lachnospiraceae;g__Roseburia | 97.6 | CRISPR;98.30 blast;95.70 iPHoP-RF;93.10 |
| S5_vs_122344 | 26.84 | d__Bacteria;p__Firmicutes;c__Clostridia;o__Lachnospirales;f__Lachnospiraceae;g__Agathobacter | 94 | CRISPR;95.80 iPHoP-RF;73.90 |
| S5_vs_122618 | 17.7 | d__Bacteria;p__Firmicutes;c__Bacilli;o__Erysipelotrichales;f__Erysipelotrichaceae;g__Clostridium_AQ | 95.4 | blast;96.80 iPHoP-RF;71.10 CRISPR;66.50 |
| S5_vs_123256 | 98.81 | d__Bacteria;p__Bacteroidota;c__Bacteroidia;o__Bacteroidales;f__Rikenellaceae;g__Alistipes | 100 | RaFAH;97.10 blast;96.90 iPHoP-RF;96.10 |
| S5_vs_123256 | 98.81 | d__Bacteria;p__Bacteroidota;c__Bacteroidia;o__Bacteroidales;f__Rikenellaceae;g__Alistipes_A | 95.9 | RaFAH;97.10 |
| S5_vs_124035 | 7.45 | d__Bacteria;p__Firmicutes;c__Clostridia;o__Oscillospirales;f__Oscillospiraceae;g__Intestinimonas | 91.2 | blast;93.60 iPHoP-RF;53.30 |
| S5_vs_124677 | 26.54 | d__Bacteria;p__Firmicutes;c__Clostridia;o__Oscillospirales;f__Ruminococcaceae;g__Faecalibacterium | 97 | CRISPR;97.90 blast;96.90 iPHoP-RF;90.80 |
| S5_vs_128398 | 22.85 | d__Bacteria;p__Firmicutes;c__Clostridia;o__Oscillospirales;f__Ruminococcaceae;g__Negativibacillus | 93.6 | blast;95.50 iPHoP-RF;82.20 |
| S5_vs_129970 | 93.93 | d__Bacteria;p__Bacteroidota;c__Bacteroidia;o__Bacteroidales;f__Bacteroidaceae;g__Bacteroides | 100 | RaFAH;99.60 blast;92.90 iPHoP-RF;92.80 |
| S5_vs_130032 | 98.82 | d__Bacteria;p__Proteobacteria;c__Gammaproteobacteria;o__Enterobacterales;f__Enterobacteriaceae;g__Escherichia | 100 | RaFAH;99.70 blast;96.90 iPHoP-RF;91.40 CRISPR;72.80 |
| S5_vs_130294 | 55.97 | d__Bacteria;p__Firmicutes;c__Clostridia;o__Oscillospirales;f__Ruminococcaceae;g__Faecalibacterium | 95.6 | blast;96.90 iPHoP-RF;81.60 |
| S5_vs_130958 | 18.11 | d__Bacteria;p__Firmicutes;c__Clostridia;o__Oscillospirales;f__Ruminococcaceae;g__Anaerotruncus | 94.2 | blast;95.90 iPHoP-RF;70.40 |
| S5_vs_131510 | 95 | d__Bacteria;p__Bacteroidota;c__Bacteroidia;o__Bacteroidales;f__Bacteroidaceae;g__Bacteroides | 100 | RaFAH;97.50 blast;96.90 iPHoP-RF;96.40 |
| S5_vs_13254 | 8.3 | d__Bacteria;p__Firmicutes;c__Clostridia;o__Oscillospirales;f__Ruminococcaceae;g__Ruthenibacterium | 95 | blast;96.50 iPHoP-RF;85.40 |
| S5_vs_1338 | 100 | d__Bacteria;p__Bacteroidota;c__Bacteroidia;o__Bacteroidales;f__Rikenellaceae;g__Alistipes | 100 | RaFAH;97.80 blast;97.50 iPHoP-RF;76.10 |
| S5_vs_1338 | 100 | d__Bacteria;p__Bacteroidota;c__Bacteroidia;o__Bacteroidales;f__Rikenellaceae;g__Alistipes_A | 99 | RaFAH;97.80 blast;81.20 |
| S5_vs_137154 | 10.76 | d__Bacteria;p__Firmicutes;c__Clostridia;o__Clostridiales;f__Clostridiaceae;g__Clostridium | 91.7 | blast;94.00 iPHoP-RF;67.30 |
| S5_vs_139401 | 12.88 | d__Bacteria;p__Firmicutes;c__Clostridia;o__Lachnospirales;f__Lachnospiraceae;g__Anaerostipes | 90.9 | blast;93.30 CRISPR;68.40 iPHoP-RF;67.50 |
| S5_vs_140946 | 9.68 | d__Bacteria;p__Firmicutes;c__Clostridia;o__Oscillospirales;f__Acutalibacteraceae;g__UMGS856 | 93.9 | blast;95.70 iPHoP-RF;76.80 |
| S5_vs_141621 | 97.16 | d__Bacteria;p__Firmicutes;c__Clostridia;o__Lachnospirales;f__Lachnospiraceae;g__Anaerostipes | 96.4 | blast;95.60 CRISPR;90.90 iPHoP-RF;88.90 RaFAH;72.10 |
| S5_vs_142195 | 100 | d__Bacteria;p__Bacteroidota;c__Bacteroidia;o__Bacteroidales;f__Bacteroidaceae;g__Bacteroides | 99.9 | RaFAH;99.80 |
| S5_vs_142195 | 100 | d__Bacteria;p__Bacteroidota;c__Bacteroidia;o__Bacteroidales;f__Rikenellaceae;g__Alistipes | 95.6 | blast;96.90 iPHoP-RF;94.10 |
| S5_vs_145578 | 39.67 | d__Bacteria;p__Firmicutes;c__Clostridia;o__Lachnospirales;f__Lachnospiraceae;g__Enterocloster | 95.2 | blast;96.60 iPHoP-RF;76.10 CRISPR;54.30 |
| S5_vs_145581 | 9.66 | d__Bacteria;p__Firmicutes;c__Clostridia;o__Christensenellales;f__Christensenellaceae;g__Christensenella | 90.4 | blast;92.90 iPHoP-RF;86.30 |
| S5_vs_145590 | 39.74 | d__Bacteria;p__Firmicutes;c__Clostridia;o__Lachnospirales;f__Lachnospiraceae;g__Clostridium_AP | 91.4 | iPHoP-RF;93.70 |
| S5_vs_145590 | 39.74 | d__Bacteria;p__Firmicutes;c__Clostridia;o__Lachnospirales;f__Lachnospiraceae;g__Lachnoclostridium_B | 91 | iPHoP-RF;93.40 blast;92.10 |
| S5_vs_145597 | 20.03 | d__Bacteria;p__Firmicutes;c__Clostridia;o__Christensenellales;f__NSJ-53;g__NSJ-53 | 94 | blast;95.80 iPHoP-RF;60.10 |
| S5_vs_145598 | 95.83 | d__Bacteria;p__Firmicutes;c__Clostridia;o__Lachnospirales;f__Lachnospiraceae;g__Roseburia | 92.1 | blast;94.30 iPHoP-RF;91.80 |
| S5_vs_145598 | 95.83 | d__Bacteria;p__Firmicutes;c__Clostridia;o__Lachnospirales;f__Lachnospiraceae;g__Agathobacter | 91 | iPHoP-RF;93.40 blast;91.60 CRISPR;61.60 |
| S5_vs_145602 | 37.74 | d__Bacteria;p__Firmicutes;c__Clostridia;o__Lachnospirales;f__Lachnospiraceae;g__Anaerobutyricum | 90.7 | blast;93.10 iPHoP-RF;65.30 |
| S5_vs_145605 | 17.51 | d__Bacteria;p__Firmicutes;c__Clostridia;o__Christensenellales;f__CAG-74;g__SFTH01 | 90.3 | blast;92.80 iPHoP-RF;68.20 |
| S5_vs_145611 | 45.88 | d__Bacteria;p__Firmicutes;c__Clostridia;o__Oscillospirales;f__Ruminococcaceae;g__Phocea | 90.7 | blast;93.10 iPHoP-RF;78.00 |
| S5_vs_1572 | 97.34 | d__Bacteria;p__Bacteroidota;c__Bacteroidia;o__Bacteroidales;f__Tannerellaceae;g__Parabacteroides | 91.6 | blast;93.90 iPHoP-RF;61.70 |
| S5_vs_17520 | 39.12 | d__Bacteria;p__Firmicutes;c__Clostridia;o__Lachnospirales;f__Lachnospiraceae;g__Hungatella | 95.9 | iPHoP-RF;97.10 CRISPR;88.40 |
| S5_vs_17520 | 39.12 | d__Bacteria;p__Firmicutes;c__Clostridia;o__Lachnospirales;f__Lachnospiraceae;g__Lacrimispora | 91.4 | iPHoP-RF;93.70 |
| S5_vs_20268 | 22.4 | d__Bacteria;p__Firmicutes;c__Clostridia;o__Oscillospirales;f__Acutalibacteraceae;g__Ruminococcus_E | 98.5 | CRISPR;98.90 blast;96.50 iPHoP-RF;89.50 |
| S5_vs_20500 | 99.42 | d__Bacteria;p__Bacteroidota;c__Bacteroidia;o__Bacteroidales;f__Rikenellaceae;g__Alistipes | 95.6 | blast;96.90 iPHoP-RF;96.70 |
| S5_vs_21778 | 11.27 | d__Bacteria;p__Firmicutes;c__Clostridia;o__Oscillospirales;f__Oscillospiraceae;g__Flavonifractor | 90.4 | blast;92.90 iPHoP-RF;59.20 |
| S5_vs_28263 | 16.17 | d__Bacteria;p__Firmicutes;c__Clostridia;o__Lachnospirales;f__Lachnospiraceae;g__Mediterraneibacter | 91.6 | blast;93.90 iPHoP-RF;86.30 CRISPR;58.00 |
| S5_vs_30490 | 42.23 | d__Bacteria;p__Firmicutes;c__Clostridia;o__Oscillospirales;f__Ruminococcaceae;g__Faecalibacterium | 99.3 | blast;97.10 RaFAH;90.30 iPHoP-RF;87.30 CRISPR;71.60 |
| S5_vs_32885 | 19.8 | d__Bacteria;p__Firmicutes;c__Clostridia;o__Oscillospirales;f__Ruminococcaceae;g__Anaerotruncus | 92.3 | blast;94.50 CRISPR;93.00 iPHoP-RF;81.90 |
| S5_vs_3501 | 32.19 | d__Bacteria;p__Firmicutes;c__Clostridia;o__Oscillospirales;f__Acutalibacteraceae;g__UBA1691 | 95.6 | blast;96.90 iPHoP-RF;90.10 CRISPR;85.90 |
| S5_vs_3501 | 32.19 | d__Bacteria;p__Firmicutes;c__Clostridia;o__Oscillospirales;f__Acutalibacteraceae;g__Fimivicinus | 94.5 | blast;96.10 iPHoP-RF;85.40 |
| S5_vs_40848 | 58.74 | d__Bacteria;p__Firmicutes;c__Clostridia;o__Oscillospirales;f__Acutalibacteraceae;g__Ruminococcus_E | 95.6 | blast;96.90 iPHoP-RF;92.80 |
| S5_vs_43207 | 30.09 | d__Bacteria;p__Firmicutes;c__Clostridia;o__Oscillospirales;f__Butyricicoccaceae;g__Agathobaculum | 91.1 | blast;93.50 iPHoP-RF;70.60 |
| S5_vs_46394 | 8.87 | d__Bacteria;p__Firmicutes;c__Clostridia;o__Christensenellales;f__QAND01;g__UMGS1975 | 90.3 | blast;92.80 iPHoP-RF;62.70 |
| S5_vs_4737 | 20.64 | d__Bacteria;p__Firmicutes;c__Clostridia;o__Christensenellales;f__CAG-74;g__Limiplasma | 93.9 | blast;95.70 iPHoP-RF;68.40 |
| S5_vs_4737 | 20.64 | d__Bacteria;p__Firmicutes;c__Clostridia;o__Christensenellales;f__CAG-74;g__Firm-11 | 91.5 | blast;93.80 iPHoP-RF;73.00 |
| S5_vs_51416 | 11.18 | d__Bacteria;p__Firmicutes;c__Bacilli;o__Erysipelotrichales;f__Erysipelotrichaceae;g__Holdemania | 90.3 | blast;92.80 CRISPR;73.80 iPHoP-RF;66.50 |
| S5_vs_58766 | 24.99 | d__Bacteria;p__Firmicutes;c__Clostridia;o__Lachnospirales;f__Lachnospiraceae;g__Frisingicoccus | 93.8 | blast;95.60 iPHoP-RF;76.60 |
| S5_vs_59798 | 40.34 | d__Bacteria;p__Firmicutes;c__Clostridia;o__Lachnospirales;f__Lachnospiraceae;g__Faecalimonas | 91.4 | blast;93.70 iPHoP-RF;91.10 CRISPR;78.90 |
| S5_vs_60507 | 96.47 | d__Bacteria;p__Firmicutes;c__Bacilli;o__Lactobacillales;f__Streptococcaceae;g__Streptococcus | 100 | RaFAH;99.70 blast;92.80 iPHoP-RF;91.10 CRISPR;52.30 |
| S5_vs_63924 | 98.89 | d__Bacteria;p__Actinobacteriota;c__Actinomycetia;o__Actinomycetales;f__Bifidobacteriaceae;g__Bifidobacterium | 99 | CRISPR;97.70 iPHoP-RF;96.70 blast;95.40 RaFAH;82.40 |
| S5_vs_6478 | 66.33 | d__Bacteria;p__Firmicutes;c__Bacilli;o__Lactobacillales;f__Lactobacillaceae;g__Lactobacillus | 100 | RaFAH;99.60 iPHoP-RF;96.70 blast;95.40 CRISPR;68.80 |
| S5_vs_6478 | 66.33 | d__Bacteria;p__Firmicutes;c__Bacilli;o__Lactobacillales;f__Lactobacillaceae;g__Lacticaseibacillus | 99.6 | RaFAH;99.60 |
| S5_vs_6478 | 66.33 | d__Bacteria;p__Firmicutes;c__Bacilli;o__Lactobacillales;f__Lactobacillaceae;g__Ligilactobacillus | 99.6 | RaFAH;99.60 |
| S5_vs_6478 | 66.33 | d__Bacteria;p__Firmicutes;c__Bacilli;o__Lactobacillales;f__Lactobacillaceae;g__Limosilactobacillus | 99.6 | RaFAH;99.60 |
| S5_vs_65490 | 3.66 | d__Bacteria;p__Firmicutes;c__Clostridia;o__Oscillospirales;f__Oscillospiraceae;g__Flavonifractor | 91.2 | blast;93.60 iPHoP-RF;56.20 |
| S5_vs_68254 | 58.14 | d__Bacteria;p__Firmicutes;c__Clostridia;o__Lachnospirales;f__Lachnospiraceae;g__Blautia_A | 93.8 | blast;93.80 iPHoP-RF;81.90 RaFAH;64.30 |
| S5_vs_69980 | 76.63 | d__Bacteria;p__Firmicutes;c__Clostridia;o__Oscillospirales;f__Oscillospiraceae;g__Flavonifractor | 95.3 | blast;96.70 iPHoP-RF;96.40 CRISPR;95.20 |
| S5_vs_72532 | 82.39 | d__Bacteria;p__Bacteroidota;c__Bacteroidia;o__Bacteroidales;f__Bacteroidaceae;g__Bacteroides | 99.4 | blast;96.90 iPHoP-RF;96.70 RaFAH;92.20 |
| S5_vs_74199 | 13.11 | d__Bacteria;p__Firmicutes;c__Clostridia;o__Oscillospirales;f__Oscillospiraceae;g__CAG-83 | 90.4 | blast;92.90 iPHoP-RF;92.10 |
| S5_vs_76500 | 80.28 | d__Bacteria;p__Bacteroidota;c__Bacteroidia;o__Bacteroidales;f__Rikenellaceae;g__Alistipes | 99 | blast;96.90 iPHoP-RF;94.40 RaFAH;87.40 |
| S5_vs_77593 | 41.43 | d__Bacteria;p__Firmicutes;c__Clostridia;o__Oscillospirales;f__Oscillospiraceae;g__CAG-103 | 90.6 | blast;93.00 iPHoP-RF;71.70 |
| S5_vs_79992 | 99.98 | d__Bacteria;p__Bacteroidota;c__Bacteroidia;o__Bacteroidales;f__Rikenellaceae;g__Alistipes | 100 | RaFAH;98.20 blast;97.10 iPHoP-RF;96.70 |
| S5_vs_79992 | 99.98 | d__Bacteria;p__Bacteroidota;c__Bacteroidia;o__Bacteroidales;f__Rikenellaceae;g__Alistipes_A | 97.5 | RaFAH;98.20 |
| S5_vs_80496 | 99.85 | d__Bacteria;p__Bacteroidota;c__Bacteroidia;o__Bacteroidales;f__Bacteroidaceae;g__Bacteroides | 99.7 | blast;96.90 iPHoP-RF;96.10 RaFAH;95.90 |
| S5_vs_80498 | 33.04 | d__Bacteria;p__Firmicutes;c__Clostridia;o__Lachnospirales;f__Lachnospiraceae;g__Muricomes | 93.2 | blast;95.20 iPHoP-RF;90.50 |
| S5_vs_8125 | 19.38 | d__Bacteria;p__Firmicutes;c__Clostridia;o__Lachnospirales;f__Lachnospiraceae;g__Eubacterium_G | 90.6 | blast;93.00 iPHoP-RF;53.30 |
| S5_vs_84574 | 8.38 | d__Bacteria;p__Firmicutes;c__Clostridia;o__Oscillospirales;f__Butyricicoccaceae;g__Agathobaculum | 91.6 | blast;93.90 iPHoP-RF;81.10 |
| S5_vs_84946 | 94.46 | d__Bacteria;p__Firmicutes;c__Clostridia;o__Lachnospirales;f__Lachnospiraceae;g__Dorea_A | 90.2 | blast;92.70 iPHoP-RF;83.60 |

| | | | | |
|---|---|---|---|---|
| S5_vs_84946 | 94.46 | d__Bacteria;p__Firmicutes;c__Clostridia;o__Lachnospirales;f__Lachnospiraceae;g__Dorea | 90.1 | blast;92.60 iPHoP-RF;75.20 |
| S5_vs_850 | 18.78 | d__Bacteria;p__Firmicutes;c__Clostridia;o__UBA1212;f__UBA1255;g__Egerieisoma | 92.7 | blast;94.80 iPHoP-RF;85.10 |
| S5_vs_850 | 18.78 | d__Bacteria;p__Firmicutes;c__Clostridia;o__UBA1212;f__UBA1255;g__UMGS1537 | 92.7 | blast;94.80 CRISPR;80.00 iPHoP-RF;78.30 |
| S5_vs_85065 | 67.47 | d__Bacteria;p__Firmicutes;c__Clostridia;o__Oscillospirales;f__Ruminococcaceae;g__Ruthenibacterium | 95.3 | blast;96.70 |
| S5_vs_86562 | 44.1 | d__Bacteria;p__Verrucomicrobiota;c__Verrucomicrobiae;o__Verrucomicrobiales;f__Akkermansiaceae;g__Akkermansia | 100 | RaFAH;99.10 CRISPR;98.80 blast;96.90 iPHoP-RF;89.20 |
| S5_vs_89540 | 26.84 | d__Bacteria;p__Firmicutes;c__Bacilli;o__Lactobacillales;f__Lactobacillaceae;g__Limosilactobacillus | 94.2 | blast;93.40 iPHoP-RF;81.90 RaFAH;68.80 |
| S5_vs_93839 | 98.83 | d__Bacteria;p__Bacteroidota;c__Bacteroidia;o__Bacteroidales;f__Rikenellaceae;g__Alistipes | 100 | RaFAH;98.90 iPHoP-RF;96.70 blast;95.50 |
| S5_vs_93839 | 98.83 | d__Bacteria;p__Bacteroidota;c__Bacteroidia;o__Bacteroidales;f__Rikenellaceae;g__Alistipes_A | 98.5 | RaFAH;98.90 |
| S5_vs_95307 | 57.8 | d__Bacteria;p__Firmicutes;c__Clostridia;o__Lachnospirales;f__Lachnospiraceae;g__Blautia | 92 | blast;91.60 iPHoP-RF;73.20 RaFAH;65.20 |
| S5_vs_9747 | 6.96 | d__Bacteria;p__Proteobacteria;c__Alphaproteobacteria;o__RF32;f__CAG-239;g__CAG-267 | 93 | blast;95.00 iPHoP-RF;69.80 |
| S5_vs_97574 | 99.69 | d__Bacteria;p__Firmicutes;c__Clostridia;o__Lachnospirales;f__Lachnospiraceae;g__Blautia_A | 100 | RaFAH;99.30 CRISPR;95.90 blast;93.60 iPHoP-RF;92.10 |
| S5_vs_97574 | 99.69 | d__Bacteria;p__Firmicutes;c__Clostridia;o__Lachnospirales;f__Lachnospiraceae;g__Blautia | 99.1 | RaFAH;99.30 |
| S6_vf_25710 | 89.8 | d__Bacteria;p__Verrucomicrobiota;c__Verrucomicrobiae;o__Verrucomicrobiales;f__Akkermansiaceae;g__Akkermansia | 99.7 | CRISPR;97.30 RaFAH;94.60 blast;93.60 iPHoP-RF;72.60 |
| S6_vf_56486 | 65.79 | d__Bacteria;p__Bacteroidota;c__Bacteroidia;o__Bacteroidales;f__Bacteroidaceae;g__Bacteroides | 99.6 | iPHoP-RF;96.10 RaFAH;94.70 |
| S6_vs_1146 | 50.09 | d__Bacteria;p__Firmicutes;c__Clostridia;o__Oscillospirales;f__Ruminococcaceae;g__Faecalibacterium | 98.4 | blast;96.90 iPHoP-RF;86.00 RaFAH;81.40 |
| S6_vs_12945 | 99.74 | d__Bacteria;p__Firmicutes;c__Clostridia;o__Lachnospirales;f__Lachnospiraceae;g__Roseburia | 99.3 | RaFAH;94.70 blast;88.50 iPHoP-RF;74.80 |
| S6_vs_12945 | 99.74 | d__Bacteria;p__Firmicutes;c__Clostridia;o__Lachnospirales;f__Lachnospiraceae;g__Agathobacter | 97.9 | RaFAH;94.70 blast;86.00 |
| S6_vs_12945 | 99.74 | d__Bacteria;p__Firmicutes;c__Clostridia;o__Lachnospirales;f__Lachnospiraceae;g__RUG115 | 96.4 | RaFAH;94.70 blast;76.10 |
| S6_vs_12945 | 99.74 | d__Bacteria;p__Firmicutes;c__Clostridia;o__Lachnospirales;f__Lachnospiraceae;g__CAG-81 | 90.9 | blast;93.30 |
| S6_vs_15390 | 33.39 | d__Bacteria;p__Bacteroidota;c__Bacteroidia;o__Bacteroidales;f__Tannerellaceae;g__Parabacteroides | 90.8 | blast;93.20 iPHoP-RF;92.10 |
| S6_vs_15813 | 90.87 | d__Bacteria;p__Firmicutes;c__Clostridia;o__Oscillospirales;f__Oscillospiraceae;g__Flavonifractor | 99.3 | blast;98.10 RaFAH;85.50 iPHoP-RF;85.10 CRISPR;56.80 |
| S6_vs_29152 | 23.31 | d__Bacteria;p__Firmicutes;c__Clostridia;o__Lachnospirales;f__Lachnospiraceae;g__Acetatifactor | 91.2 | blast;93.60 |
| S6_vs_32886 | 6.93 | d__Bacteria;p__Bacteroidota;c__Bacteroidia;o__Bacteroidales;f__Rikenellaceae;g__Alistipes_A | 94.2 | blast;95.90 iPHoP-RF;66.30 |
| S6_vs_33711 | 86.98 | d__Bacteria;p__Firmicutes;c__Clostridia;o__Oscillospirales;f__Ruminococcaceae;g__Ruthenibacterium | 94 | blast;95.80 iPHoP-RF;85.40 CRISPR;69.40 |
| S6_vs_34453 | 77 | d__Bacteria;p__Firmicutes;c__Clostridia;o__Oscillospirales;f__Ruminococcaceae;g__Faecalibacterium | 93.6 | blast;95.50 iPHoP-RF;64.30 |
| S6_vs_38250 | 15.25 | d__Bacteria;p__Firmicutes;c__Clostridia;o__Oscillospirales;f__Ruminococcaceae;g__UBA3818 | 94.3 | blast;96.00 iPHoP-RF;76.10 |
| S6_vs_39025 | 99.25 | d__Bacteria;p__Firmicutes;c__Clostridia;o__Lachnospirales;f__Lachnospiraceae;g__Enterocloster | 91.6 | blast;93.90 |
| S6_vs_39065 | 83.64 | d__Bacteria;p__Proteobacteria;c__Alphaproteobacteria;o__RF32;f__CAG-239;g__CAG-267 | 94.6 | blast;96.20 iPHoP-RF;75.20 |
| S6_vs_44070 | 14.4 | d__Bacteria;p__Proteobacteria;c__Alphaproteobacteria;o__RF32;f__CAG-239;g__51-20 | 94.7 | blast;96.30 iPHoP-RF;72.60 |
| S6_vs_46039 | 24.66 | d__Bacteria;p__Bacteroidota;c__Bacteroidia;o__Bacteroidales;f__Bacteroidaceae;g__Bacteroides | 93 | CRISPR;93.00 blast;89.60 iPHoP-RF;82.50 RaFAH;64.10 |
| S6_vs_46091 | 25.96 | d__Bacteria;p__Firmicutes;c__Clostridia;o__Oscillospirales;f__Ruminococcaceae;g__Negativibacillus | 96.6 | blast;97.60 iPHoP-RF;83.30 CRISPR;69.80 |
| S6_vs_53966 | 51.17 | d__Bacteria;p__Firmicutes;c__Clostridia;o__Oscillospirales;f__Ruminococcaceae;g__Faecalibacterium | 98.7 | blast;96.30 iPHoP-RF;90.50 RaFAH;86.50 |
| S6_vs_54606 | 99.52 | d__Bacteria;p__Firmicutes;c__Clostridia;o__Lachnospirales;f__Lachnospiraceae;g__Blautia | 97 | RaFAH;91.10 blast;88.40 |
| S6_vs_54606 | 99.52 | d__Bacteria;p__Firmicutes;c__Clostridia;o__Lachnospirales;f__Lachnospiraceae;g__Ruminococcus_B | 91.2 | blast;93.60 iPHoP-RF;61.70 |
| S6_vs_56485 | 14.92 | d__Bacteria;p__Firmicutes;c__Bacilli;o__Erysipelotrichales;f__Erysipelotrichaceae;g__Holdemania | 94.7 | CRISPR;96.30 blast;94.30 iPHoP-RF;79.80 |
| S6_vs_56490 | 26.23 | d__Bacteria;p__Firmicutes;c__Clostridia;o__Oscillospirales;f__Acutalibacteraceae;g__UMGS1071 | 95.9 | blast;97.10 iPHoP-RF;83.90 CRISPR;68.20 |
| S6_vs_56491 | 92.92 | d__Bacteria;p__Firmicutes;c__Clostridia;o__Lachnospirales;f__Lachnospiraceae;g__Sellimonas | 97.5 | blast;98.20 iPHoP-RF;75.20 |
| S6_vs_56492 | 27.91 | d__Bacteria;p__Firmicutes;c__Clostridia;o__Oscillospirales;f__Butyricicoccaceae;g__Agathobaculum | 98.2 | blast;98.70 iPHoP-RF;66.80 |
| S6_vs_56496 | 61.48 | d__Bacteria;p__Firmicutes;c__Clostridia;o__Lachnospirales;f__Lachnospiraceae;g__Marvinbryantia | 91.2 | blast;93.60 iPHoP-RF;65.10 |
| S6_vs_56498 | 78.58 | d__Bacteria;p__Firmicutes;c__Clostridia;o__Lachnospirales;f__Lachnospiraceae;g__Muricomes | 91 | blast;93.40 iPHoP-RF;69.80 |
| S6_vs_9784 | 91.55 | d__Bacteria;p__Bacteroidota;c__Bacteroidia;o__Bacteroidales;f__Bacteroidaceae;g__Bacteroides | 100 | RaFAH;97.10 iPHoP-RF;96.70 blast;93.90 |
| S6_vs_9784 | 91.55 | d__Bacteria;p__Bacteroidota;c__Bacteroidia;o__Bacteroidales;f__Bacteroidaceae;g__Phocaeicola | 94.9 | iPHoP-RF;96.40 |
| S7_vf_114476 | 27.19 | d__Bacteria;p__Firmicutes;c__Clostridia;o__Lachnospirales;f__Lachnospiraceae;g__Anaerobutyricum | 94.3 | blast;96.00 iPHoP-RF;74.30 |
| S7_vf_117697 | 98.64 | d__Bacteria;p__Bacteroidota;c__Bacteroidia;o__Bacteroidales;f__Rikenellaceae;g__Alistipes | 96.9 | CRISPR;97.80 iPHoP-RF;97.10 blast;96.80 |
| S7_vf_117697 | 98.64 | d__Bacteria;p__Bacteroidota;c__Bacteroidia;o__Bacteroidales;f__Bacteroidaceae;g__Bacteroides | 92.6 | RaFAH;94.70 |
| S7_vf_134023 | 12.46 | d__Bacteria;p__Bacteroidota;c__Bacteroidia;o__Bacteroidales;f__Rikenellaceae;g__Alistipes_A | 95 | blast;96.50 iPHoP-RF;54.40 |
| S7_vf_13614 | 39.23 | d__Bacteria;p__Firmicutes;c__Clostridia;o__Oscillospirales;f__Ruminococcaceae;g__Ruminococcus_D | 95.4 | blast;96.80 iPHoP-RF;95.40 CRISPR;50.20 |
| S7_vf_138019 | 8.23 | d__Bacteria;p__Firmicutes;c__Clostridia;o__Oscillospirales;f__Oscillospiraceae;g__Lawsonibacter | 91.1 | blast;93.50 iPHoP-RF;50.40 |
| S7_vf_138069 | 78.93 | d__Bacteria;p__Firmicutes;c__Bacilli;o__Lactobacillales;f__Streptococcaceae;g__Streptococcus | 99.6 | RaFAH;99.40 iPHoP-RF;67.30 |
| S7_vf_27309 | 18.88 | d__Bacteria;p__Bacteroidota;c__Bacteroidia;o__Bacteroidales;f__Bacteroidaceae;g__Bacteroides | 96.3 | CRISPR;97.40 iPHoP-RF;93.70 |
| S7_vf_28429 | 86.01 | d__Bacteria;p__Bacteroidota;c__Bacteroidia;o__Bacteroidales;f__Tannerellaceae;g__Parabacteroides | 100 | CRISPR;99.00 RaFAH;98.30 iPHoP-RF;94.10 blast;93.90 |
| S7_vf_29941 | 79.47 | d__Bacteria;p__Firmicutes;c__Bacilli;o__Lactobacillales;f__Streptococcaceae;g__Lactococcus | 99.4 | blast;97.20 RaFAH;91.90 iPHoP-RF;73.90 |
| S7_vf_37279 | 64.97 | d__Bacteria;p__Bacteroidota;c__Bacteroidia;o__Bacteroidales;f__Bacteroidaceae;g__Bacteroides | 99.7 | CRISPR;98.80 iPHoP-RF;97.10 blast;90.80 RaFAH;87.70 |
| S7_vf_37279 | 64.97 | d__Bacteria;p__Bacteroidota;c__Bacteroidia;o__Bacteroidales;f__Bacteroidaceae;g__Phocaeicola | 98.2 | CRISPR;98.70 iPHoP-RF;95.10 blast;91.00 |
| S7_vf_37293 | 98.54 | d__Bacteria;p__Firmicutes;c__Bacilli;o__Lactobacillales;f__Streptococcaceae;g__Streptococcus | 100 | RaFAH;99.60 CRISPR;99.00 iPHoP-RF;54.10 |
| S7_vf_37293 | 98.54 | d__Bacteria;p__Bacteroidota;c__Bacteroidia;o__Bacteroidales;f__Bacteroidaceae;g__Prevotella | 90.1 | blast;92.60 |
| S7_vf_38697 | 61.28 | d__Bacteria;p__Firmicutes;c__Bacilli;o__Erysipelotrichales;f__Erysipelatoclostridiaceae;g__Massiliomicrobiota | 94.2 | blast;95.90 iPHoP-RF;74.30 |
| S7_vf_39724 | 99.39 | d__Bacteria;p__Firmicutes;c__Clostridia;o__Oscillospirales;f__Ruminococcaceae;g__Faecalibacterium | 99.4 | RaFAH;99.50 |
| S7_vf_49007 | 85 | d__Bacteria;p__Firmicutes;c__Bacilli;o__Lactobacillales;f__Streptococcaceae;g__Streptococcus | 100 | CRISPR;98.70 RaFAH;97.80 iPHoP-RF;68.00 |
| S7_vf_49485 | 64.81 | d__Bacteria;p__Firmicutes;c__Clostridia;o__Lachnospirales;f__Lachnospiraceae;g__Anaerostipes | 98.5 | CRISPR;98.90 iPHoP-RF;56.20 |
| S7_vf_5801 | 93.48 | d__Bacteria;p__Bacteroidota;c__Bacteroidia;o__Bacteroidales;f__Bacteroidaceae;g__Bacteroides | 100 | RaFAH;96.60 iPHoP-RF;96.40 blast;91.00 |
| S7_vf_5801 | 93.48 | d__Bacteria;p__Bacteroidota;c__Bacteroidia;o__Bacteroidales;f__Bacteroidaceae;g__Phocaeicola | 95 | blast;96.50 iPHoP-RF;95.70 CRISPR;71.30 |
| S7_vf_5843 | 19.99 | d__Bacteria;p__Actinobacteriota;c__Actinomycetia;o__Actinomycetales;f__Bifidobacteriaceae;g__Bifidobacterium | 96.3 | blast;97.40 CRISPR;97.30 iPHoP-RF;93.40 |
| S7_vf_60968 | 71.38 | d__Bacteria;p__Firmicutes;c__Clostridia;o__Lachnospirales;f__Lachnospiraceae;g__Ruminococcus_B | 97.8 | blast;98.40 iPHoP-RF;81.40 CRISPR;66.90 |
| S7_vf_63894 | 77.84 | d__Bacteria;p__Firmicutes;c__Bacilli;o__Lactobacillales;f__Lactobacillaceae;g__Leuconostoc | 96.2 | iPHoP-RF;93.70 RaFAH;78.60 |
| S7_vf_67910 | 11.86 | d__Bacteria;p__Actinobacteriota;c__Coriobacteriia;o__Coriobacteriales;f__Eggerthellaceae;g__Eggerthella | 94.5 | blast;96.10 |
| S7_vf_68239 | 31.22 | d__Bacteria;p__Firmicutes;c__Bacilli;o__Lactobacillales;f__Streptococcaceae;g__Lactococcus | 93.1 | RaFAH;95.10 |
| S7_vf_68546 | 29.45 | d__Bacteria;p__Firmicutes;c__Clostridia;o__Lachnospirales;f__Lachnospiraceae;g__Faecalimonas | 94.5 | CRISPR;96.10 iPHoP-RF;76.10 |
| S7_vf_68546 | 29.45 | d__Bacteria;p__Firmicutes;c__Clostridia;o__Lachnospirales;f__Lachnospiraceae;g__Bariatricus | 94 | CRISPR;95.80 iPHoP-RF;84.80 |
| S7_vf_71527 | 3.4 | d__Bacteria;p__Firmicutes;c__Clostridia;o__Oscillospirales;f__Ruminococcaceae;g__Ruminococcus_D | 93.1 | iPHoP-RF;95.10 blast;94.30 CRISPR;92.20 |
| S7_vf_72493 | 13.24 | d__Bacteria;p__Firmicutes;c__Clostridia;o__Lachnospirales;f__Anaerotignaceae;g__UMGS1670 | 92.5 | blast;94.60 iPHoP-RF;65.10 |
| S7_vf_76463 | 42.93 | d__Bacteria;p__Firmicutes;c__Clostridia;o__Oscillospirales;f__Oscillospiraceae;g__CAG-103 | 95.3 | CRISPR;96.70 |
| S7_vf_8332 | 10.6 | d__Bacteria;p__Firmicutes;c__Clostridia;o__Oscillospirales;f__Ruminococcaceae;g__Ruminococcus_D | 98.5 | CRISPR;98.90 blast;96.20 iPHoP-RF;94.70 |
| S7_vf_84974 | 91.7 | d__Bacteria;p__Firmicutes;c__Clostridia;o__Oscillospirales;f__Ruminococcaceae;g__Faecalibacterium | 99.9 | RaFAH;98.40 blast;92.10 iPHoP-RF;63.30 CRISPR;56.10 |
| S7_vf_89197 | 89.48 | d__Bacteria;p__Firmicutes;c__Bacilli;o__Lactobacillales;f__Streptococcaceae;g__Lactococcus | 96.6 | RaFAH;95.80 iPHoP-RF;71.50 |
| S7_vs_101751 | 99.91 | d__Bacteria;p__Firmicutes;c__Clostridia;o__Lachnospirales;f__Lachnospiraceae;g__Blautia_A | 99.7 | RaFAH;94.40 blast;93.60 iPHoP-RF;76.80 |
| S7_vs_101751 | 99.91 | d__Bacteria;p__Firmicutes;c__Clostridia;o__Lachnospirales;f__Lachnospiraceae;g__Blautia | 94.5 | RaFAH;94.40 iPHoP-RF;64.80 |

| | | | | |
|---|---|---|---|---|
| S7_vs_103530 | 35.56 | d__Bacteria;p__Firmicutes;c__Clostridia;o__Oscillospirales;f__Butyricicoccaceae;g__Agathobaculum | 94.2 | blast;95.90 iPHoP-RF;60.30 |
| S7_vs_11081 | 95.36 | d__Bacteria;p__Bacteroidota;c__Bacteroidia;o__Bacteroidales;f__Bacteroidaceae;g__Bacteroides | 100 | RaFAH;98.70 iPHoP-RF;94.70 blast;91.00 |
| S7_vs_111831 | 38.83 | d__Bacteria;p__Firmicutes;c__Clostridia;o__Oscillospirales;f__Ruminococcaceae;g__Gemmiger | 97.2 | blast;98.00 iPHoP-RF;88.90 CRISPR;82.40 |
| S7_vs_118295 | 43.98 | d__Bacteria;p__Firmicutes;c__Clostridia;o__Oscillospirales;f__Oscillospiraceae;g__Intestinimonas | 91.7 | blast;94.00 iPHoP-RF;75.40 |
| S7_vs_118295 | 43.98 | d__Bacteria;p__Firmicutes;c__Clostridia;o__Oscillospirales;f__Oscillospiraceae;g__Flavonifractor | 91.4 | blast;93.70 iPHoP-RF;80.80 |
| S7_vs_119679 | 99.98 | d__Bacteria;p__Bacteroidota;c__Bacteroidia;o__Bacteroidales;f__Rikenellaceae;g__Alistipes | 100 | blast;98.70 RaFAH;98.60 iPHoP-RF;97.10 |
| S7_vs_119679 | 99.98 | d__Bacteria;p__Bacteroidota;c__Bacteroidia;o__Bacteroidales;f__Rikenellaceae;g__Alistipes_A | 98.1 | RaFAH;98.60 |
| S7_vs_129271 | 12.57 | d__Bacteria;p__Firmicutes;c__Clostridia;o__Oscillospirales;f__Acutalibacteraceae;g__UBA1691 | 98.5 | CRISPR;98.90 blast;97.30 iPHoP-RF;82.80 |
| S7_vs_129271 | 12.57 | d__Bacteria;p__Firmicutes;c__Clostridia;o__Oscillospirales;f__Acutalibacteraceae;g__Fimivicinus | 93.9 | blast;95.70 iPHoP-RF;80.30 |
| S7_vs_129858 | 10.46 | d__Bacteria;p__Firmicutes;c__Clostridia;o__Christensenellales;f__CAG-74;g__Firm-11 | 91.4 | blast;93.70 iPHoP-RF;85.40 CRISPR;50.00 |
| S7_vs_13041 | 17.74 | d__Bacteria;p__Firmicutes;c__Bacilli;o__Erysipelotrichales;f__Erysipelatoclostridiaceae;g__Erysipelatoclostridium | 94.5 | blast;96.10 iPHoP-RF;74.60 CRISPR;54.40 |
| S7_vs_13370 | 8.9 | d__Bacteria;p__Firmicutes;c__Clostridia;o__Oscillospirales;f__Ruminococcaceae;g__Faecalibacterium | 97 | blast;97.90 iPHoP-RF;90.10 CRISPR;77.30 |
| S7_vs_134391 | 17.86 | d__Bacteria;p__Firmicutes;c__Clostridia;o__Oscillospirales;f__UMGS1783;g__UMGS1783 | 94.9 | blast;96.40 iPHoP-RF;84.80 CRISPR;79.10 |
| S7_vs_134784 | 77.29 | d__Bacteria;p__Firmicutes;c__Negativicutes;o__Veillonellales;f__Dialisteraceae;g__Dialister | 97.3 | CRISPR;97.40 blast;96.60 iPHoP-RF;92.80 RaFAH;63.90 |
| S7_vs_135322 | 34.72 | d__Bacteria;p__Firmicutes;c__Clostridia;o__Oscillospirales;f__Ruminococcaceae;g__Faecalibacterium | 99.7 | CRISPR;98.30 blast;91.90 RaFAH;90.70 iPHoP-RF;90.50 |
| S7_vs_135679 | 23.81 | d__Bacteria;p__Firmicutes;c__Clostridia;o__Lachnospirales;f__Lachnospiraceae;g__Blautia_A | 99.6 | blast;96.50 RaFAH;94.00 iPHoP-RF;90.10 |
| S7_vs_135679 | 23.81 | d__Bacteria;p__Firmicutes;c__Clostridia;o__Lachnospirales;f__Lachnospiraceae;g__Blautia | 91.7 | RaFAH;94.00 |
| S7_vs_135828 | 3.2 | d__Bacteria;p__Bacteroidota;c__Bacteroidia;o__Bacteroidales;f__Bacteroidaceae;g__Bacteroides | 93 | blast;95.00 iPHoP-RF;86.00 |
| S7_vs_137611 | 29.3 | d__Bacteria;p__Firmicutes;c__Clostridia;o__Lachnospirales;f__Lachnospiraceae;g__Anaerobutyricum | 95 | blast;96.50 iPHoP-RF;87.60 |
| S7_vs_137915 | 55.05 | d__Bacteria;p__Bacteroidota;c__Bacteroidia;o__Bacteroidales;f__Tannerellaceae;g__Parabacteroides | 98.1 | CRISPR;98.30 blast;90.90 iPHoP-RF;85.40 RaFAH;59.50 |
| S7_vs_13799 | 38.3 | d__Bacteria;p__Firmicutes;c__Clostridia;o__Lachnospirales;f__Lachnospiraceae;g__Dorea_A | 93 | blast;95.00 iPHoP-RF;94.70 CRISPR;72.50 |
| S7_vs_137998 | 42.1 | d__Bacteria;p__Firmicutes;c__Clostridia;o__Lachnospirales;f__Lachnospiraceae;g__Blautia_A | 93.4 | blast;95.30 iPHoP-RF;78.00 |
| S7_vs_138109 | 31.21 | d__Bacteria;p__Firmicutes;c__Clostridia;o__Lachnospirales;f__Lachnospiraceae;g__Anaerobutyricum | 91 | blast;93.40 iPHoP-RF;75.90 |
| S7_vs_138116 | 5.66 | d__Bacteria;p__Firmicutes;c__Clostridia;o__Lachnospirales;f__Lachnospiraceae;g__Mediterraneibacter | 91.5 | blast;93.80 iPHoP-RF;89.50 |
| S7_vs_138173 | 100 | d__Bacteria;p__Bacteroidota;c__Bacteroidia;o__Bacteroidales;f__Rikenellaceae;g__Alistipes | 100 | RaFAH;98.50 blast;94.70 iPHoP-RF;74.10 |
| S7_vs_138173 | 100 | d__Bacteria;p__Bacteroidota;c__Bacteroidia;o__Bacteroidales;f__Rikenellaceae;g__Alistipes_A | 97.9 | RaFAH;98.50 |
| S7_vs_16344 | 12.56 | d__Bacteria;p__Firmicutes;c__Clostridia;o__Oscillospirales;f__Acutalibacteraceae;g__Ruminococcus_E | 98.8 | CRISPR;99.10 blast;96.90 iPHoP-RF;95.70 |
| S7_vs_21521 | 35.28 | d__Bacteria;p__Firmicutes;c__Clostridia;o__Oscillospirales;f__Ruminococcaceae;g__Ruminiclostridium_E | 90 | blast;92.50 iPHoP-RF;74.80 |
| S7_vs_2599 | 89.5 | d__Bacteria;p__Bacteroidota;c__Bacteroidia;o__Bacteroidales;f__Bacteroidaceae;g__Bacteroides | 100 | RaFAH;99.30 iPHoP-RF;94.40 blast;93.90 |
| S7_vs_26757 | | d__Bacteria;p__Firmicutes;c__Clostridia;o__Oscillospirales;f__Ruminococcaceae;g__Faecalibacterium | 94.6 | blast;96.20 iPHoP-RF;85.10 |
| S7_vs_28150 | 12.25 | d__Bacteria;p__Firmicutes;c__Clostridia;o__Oscillospirales;f__Oscillospiraceae;g__Dysosmobacter | 94.2 | blast;95.90 iPHoP-RF;74.10 |
| S7_vs_28733 | 56.54 | d__Bacteria;p__Firmicutes;c__Clostridia;o__Oscillospirales;f__Acutalibacteraceae;g__Hydrogeniiclostridium | 95.2 | blast;96.60 iPHoP-RF;62.20 |
| S7_vs_29595 | 82.91 | d__Bacteria;p__Verrucomicrobiota;c__Verrucomicrobiae;o__Verrucomicrobiales;f__Akkermansiaceae;g__Akkermansia | 99.4 | CRISPR;99.50 iPHoP-RF;57.30 |
| S7_vs_30640 | 25.04 | d__Bacteria;p__Firmicutes;c__Clostridia;o__Lachnospirales;f__Lachnospiraceae;g__Eubacterium_G | 94.3 | blast;96.00 iPHoP-RF;75.20 |
| S7_vs_30665 | 8.94 | d__Bacteria;p__Firmicutes;c__Clostridia;o__Christensenellales;f__CAG-74;g__UBA11524 | 93.8 | CRISPR;95.60 blast;92.80 iPHoP-RF;92.40 |
| S7_vs_38113 | 17.89 | d__Bacteria;p__Firmicutes;c__Clostridia;o__Oscillospirales;f__Acutalibacteraceae;g__Hydrogeniiclostridium | 91.7 | blast;94.00 iPHoP-RF;64.50 |
| S7_vs_38834 | 25.72 | d__Bacteria;p__Firmicutes;c__Clostridia;o__Lachnospirales;f__Lachnospiraceae;g__RUG115 | 91.5 | blast;93.80 |
| S7_vs_38834 | 25.72 | d__Bacteria;p__Firmicutes;c__Clostridia;o__Lachnospirales;f__Lachnospiraceae;g__Roseburia | 90.1 | blast;92.60 iPHoP-RF;69.50 |
| S7_vs_41098 | 97.6 | d__Bacteria;p__Firmicutes;c__Clostridia;o__Lachnospirales;f__Lachnospiraceae;g__Mediterraneibacter | 95.4 | blast;96.80 |
| S7_vs_42563 | 93.91 | d__Bacteria;p__Firmicutes;c__Clostridia;o__Oscillospirales;f__Ruminococcaceae;g__Gemmiger | 96.3 | blast;96.80 iPHoP-RF;75.40 RaFAH;59.00 |
| S7_vs_47050 | 16.97 | d__Bacteria;p__Firmicutes;c__Clostridia;o__Oscillospirales;f__Acutalibacteraceae;g__CAG-180 | 98.7 | CRISPR;99.00 blast;96.60 iPHoP-RF;92.40 |
| S7_vs_47050 | 16.97 | d__Bacteria;p__Firmicutes;c__Clostridia;o__Oscillospirales;f__Acutalibacteraceae;g__Fimenecus | 97.5 | blast;98.20 iPHoP-RF;92.10 |
| S7_vs_48523 | 66.97 | d__Bacteria;p__Proteobacteria;c__Gammaproteobacteria;o__Burkholderiales;f__Burkholderiaceae;g__Sutterella | 91.6 | blast;93.90 |
| S7_vs_48708 | 42.25 | d__Bacteria;p__Firmicutes;c__Clostridia;o__Lachnospirales;f__Lachnospiraceae;g__Blautia_A | 90.9 | blast;93.30 iPHoP-RF;69.50 |
| S7_vs_49426 | 18.87 | d__Bacteria;p__Firmicutes;c__Clostridia;o__Lachnospirales;f__Lachnospiraceae;g__Agathobacter | 91.5 | blast;93.80 iPHoP-RF;71.30 |
| S7_vs_4956 | 54.17 | d__Bacteria;p__Firmicutes;c__Clostridia;o__Oscillospirales;f__Oscillospiraceae;g__ | 91.6 | blast;93.90 iPHoP-RF;83.30 |
| S7_vs_50194 | 18.68 | d__Bacteria;p__Bacteroidota;c__Bacteroidia;o__Bacteroidales;f__Rikenellaceae;g__Alistipes_A | 96.9 | blast;90.90 RaFAH;87.90 iPHoP-RF;85.70 |
| S7_vs_50194 | 18.68 | d__Bacteria;p__Bacteroidota;c__Bacteroidia;o__Bacteroidales;f__Rikenellaceae;g__Alistipes | 96.6 | blast;89.90 RaFAH;87.90 |
| S7_vs_51332 | 13.21 | d__Bacteria;p__Bacteroidota;c__Bacteroidia;o__Bacteroidales;f__Coprobacteraceae;g__Coprobacter | 94.5 | blast;96.10 iPHoP-RF;53.30 |
| S7_vs_5236 | 93.85 | d__Bacteria;p__Firmicutes;c__Clostridia;o__Oscillospirales;f__Oscillospiraceae;g__CAG-103 | 93.5 | CRISPR;95.40 |
| S7_vs_54322 | 65.6 | d__Bacteria;p__Bacteroidota;c__Bacteroidia;o__Bacteroidales;f__Rikenellaceae;g__Alistipes | 98.7 | blast;96.80 CRISPR;95.60 iPHoP-RF;87.30 RaFAH;84.00 |
| S7_vs_55031 | 23.56 | d__Bacteria;p__Bacteroidota;c__Bacteroidia;o__Bacteroidales;f__Tannerellaceae;g__Parabacteroides | 98.1 | blast;98.60 CRISPR;97.70 iPHoP-RF;89.20 |
| S7_vs_55526 | 22.37 | d__Bacteria;p__Actinobacteriota;c__Coriobacteriia;o__Coriobacteriales;f__Coriobacteriaceae;g__Collinsella | 98.7 | CRISPR;99.00 blast;93.80 iPHoP-RF;84.50 |
| S7_vs_557 | 66.78 | d__Bacteria;p__Bacteroidota;c__Bacteroidia;o__Bacteroidales;f__Bacteroidaceae;g__Bacteroides | 99.3 | iPHoP-RF;97.10 blast;93.70 RaFAH;89.50 |
| S7_vs_557 | 66.78 | d__Bacteria;p__Bacteroidota;c__Bacteroidia;o__Bacteroidales;f__Bacteroidaceae;g__Phocaeicola | 94.9 | iPHoP-RF;96.40 blast;82.80 |
| S7_vs_60775 | 62.59 | d__Bacteria;p__Firmicutes;c__Clostridia;o__Lachnospirales;f__Lachnospiraceae;g__Eubacterium_F | 91.6 | blast;93.90 |
| S7_vs_65719 | 47.93 | d__Bacteria;p__Firmicutes;c__Clostridia;o__Lachnospirales;f__Lachnospiraceae;g__Enterocloster | 95.7 | CRISPR;97.00 blast;94.90 iPHoP-RF;88.90 |
| S7_vs_69630 | 35.61 | d__Bacteria;p__Firmicutes;c__Clostridia;o__Oscillospirales;f__Oscillospiraceae;g__Intestinimonas | 94.7 | blast;96.30 iPHoP-RF;65.10 |
| S7_vs_72737 | 3.91 | d__Bacteria;p__Firmicutes;c__Clostridia;o__Oscillospirales;f__Acutalibacteraceae;g__Ruminococcus_E | 97.9 | blast;98.50 iPHoP-RF;96.40 |
| S7_vs_73280 | 86.44 | d__Bacteria;p__Actinobacteriota;c__Actinomycetia;o__Actinomycetales;f__Bifidobacteriaceae;g__Bifidobacterium | 99.1 | CRISPR;99.30 iPHoP-RF;97.10 blast;96.90 |
| S7_vs_78507 | 36.34 | d__Bacteria;p__Firmicutes;c__Clostridia;o__Oscillospirales;f__Oscillospiraceae;g__Intestinimonas | 90.8 | blast;93.20 iPHoP-RF;70.90 |
| S7_vs_83100 | 97.77 | d__Bacteria;p__Firmicutes;c__Clostridia;o__Oscillospirales;f__Oscillospiraceae;g__Flavonifractor | 97.8 | blast;98.40 iPHoP-RF;85.10 CRISPR;74.00 |
| S7_vs_83474 | 31.73 | d__Bacteria;p__Firmicutes;c__Clostridia;o__Oscillospirales;f__Oscillospiraceae;g__Flavonifractor | 93.9 | blast;95.70 iPHoP-RF;90.80 |
| S7_vs_86377 | 91.04 | d__Bacteria;p__Firmicutes;c__Clostridia;o__Lachnospirales;f__Lachnospiraceae;g__Bariatricus | 92 | blast;94.20 iPHoP-RF;79.50 |
| S7_vs_86377 | 91.04 | d__Bacteria;p__Firmicutes;c__Clostridia;o__Lachnospirales;f__Lachnospiraceae;g__Lachnoclostridium_B | 91.2 | blast;93.60 iPHoP-RF;74.80 |
| S7_vs_87279 | 66.77 | d__Bacteria;p__Firmicutes;c__Clostridia;o__Oscillospirales;f__Ruminococcaceae;g__Faecalibacterium | 95.2 | blast;96.60 iPHoP-RF;62.50 |
| S7_vs_87439 | 31.68 | d__Bacteria;p__Verrucomicrobiota;c__Lentisphaeria;o__Victivallales;f__Victivallaceae;g__Victivallis | 90.2 | blast;92.70 iPHoP-RF;71.10 |
| S7_vs_944 | 66.43 | d__Bacteria;p__Bacteroidota;c__Bacteroidia;o__Bacteroidales;f__Bacteroidaceae;g__Bacteroides | 100 | RaFAH;98.90 iPHoP-RF;96.70 blast;91.80 |
| S7_vs_94738 | 58.62 | d__Bacteria;p__Firmicutes;c__Clostridia;o__Oscillospirales;f__Oscillospiraceae;g__Flavonifractor | 91.2 | blast;93.60 iPHoP-RF;71.30 |
| S7_vs_97737 | 56.07 | d__Bacteria;p__Firmicutes;c__Clostridia;o__Oscillospirales;f__Ruminococcaceae;g__Faecalibacterium | 95.6 | blast;96.90 iPHoP-RF;91.80 |
| S8_vf_101283 | 99.72 | d__Bacteria;p__Firmicutes;c__Clostridia;o__Oscillospirales;f__Ruminococcaceae;g__Faecalibacterium | 100 | RaFAH;99.60 CRISPR;98.20 blast;95.50 iPHoP-RF;84.80 |
| S8_vf_102508 | 92.39 | d__Bacteria;p__Actinobacteriota;c__Actinomycetia;o__Actinomycetales;f__Micrococcaceae;g__Rothia | 100 | RaFAH;99.40 CRISPR;98.40 iPHoP-RF;80.80 |
| S8_vf_111790 | 20.45 | d__Bacteria;p__Firmicutes;c__Clostridia;o__Oscillospirales;f__Oscillospiraceae;g__Flavonifractor | 91.6 | blast;93.90 iPHoP-RF;81.10 |
| S8_vf_117796 | 71.62 | d__Bacteria;p__Bacteroidota;c__Bacteroidia;o__Bacteroidales;f__Bacteroidaceae;g__Prevotella | 91.1 | blast;93.50 iPHoP-RF;88.50 |
| S8_vf_117796 | 71.62 | d__Bacteria;p__Bacteroidota;c__Bacteroidia;o__Bacteroidales;f__Bacteroidaceae;g__Bacteroides | 90.9 | RaFAH;93.30 |
| S8_vf_122206 | 16.65 | d__Bacteria;p__Bacteroidota;c__Bacteroidia;o__Bacteroidales;f__Bacteroidaceae;g__Prevotella | 94.7 | blast;96.30 iPHoP-RF;90.50 |
| S8_vf_129423 | 87.49 | d__Bacteria;p__Firmicutes;c__Bacilli;o__Lactobacillales;f__Lactobacillaceae;g__Leuconostoc | 97.5 | iPHoP-RF;93.70 RaFAH;86.10 |

| | | | | |
|---|---|---|---|---|
| S8_vf_131009 | 24.39 | d__Bacteria;p__Firmicutes;c__Clostridia;o__Lachnospirales;f__Lachnospiraceae;g__Eubacterium_F | 97.2 | CRISPR;98.00 blast;96.80 iPHoP-RF;95.40 |
| S8_vf_132697 | 84.84 | d__Bacteria;p__Firmicutes;c__Clostridia;o__Lachnospirales;f__Lachnospiraceae;g__Agathobacter | 99.9 | CRISPR;98.90 RaFAH;92.10 blast;90.20 iPHoP-RF;86.30 |
| S8_vf_132697 | 84.84 | d__Bacteria;p__Firmicutes;c__Clostridia;o__Lachnospirales;f__Lachnospiraceae;g__Roseburia | 99.3 | RaFAH;92.10 blast;87.10 iPHoP-RF;85.70 |
| S8_vf_132697 | 84.84 | d__Bacteria;p__Firmicutes;c__Clostridia;o__Lachnospirales;f__Lachnospiraceae;g__Lachnospira | 98.7 | CRISPR;99.00 blast;78.00 |
| S8_vf_132697 | 84.84 | d__Bacteria;p__Firmicutes;c__Clostridia;o__Lachnospirales;f__Lachnospiraceae;g__RUG115 | 96.6 | RaFAH;92.10 blast;85.00 |
| S8_vf_134943 | 81.72 | d__Bacteria;p__Firmicutes;c__Bacilli;o__Lactobacillales;f__Streptococcaceae;g__Streptococcus | 99.7 | RaFAH;99.40 iPHoP-RF;72.60 |
| S8_vf_13709 | 11.95 | d__Bacteria;p__Firmicutes;c__Clostridia;o__TANB77;f__CAG-508;g__CAG-354 | 90.2 | blast;92.70 iPHoP-RF;70.60 |
| S8_vf_13758 | 11.78 | d__Bacteria;p__Firmicutes;c__Clostridia;o__TANB77;f__CAG-508;g__CAG-354 | 93.9 | blast;95.70 iPHoP-RF;71.70 |
| S8_vf_142700 | 5.82 | d__Bacteria;p__Actinobacteriota;c__Coriobacteriia;o__Coriobacteriales;f__Coriobacteriaceae;g__Collinsella | 99.4 | CRISPR;99.50 iPHoP-RF;93.40 blast;91.90 |
| S8_vf_151384 | 47.18 | d__Bacteria;p__Firmicutes;c__Clostridia;o__Oscillospirales;f__Ruminococcaceae;g__Gemmiger | 96.3 | CRISPR;97.40 blast;92.00 iPHoP-RF;89.80 |
| S8_vf_153696 | 11.92 | d__Bacteria;p__Actinobacteriota;c__Actinomycetia;o__Actinomycetales;f__Bifidobacteriaceae;g__Bifidobacterium | 91.2 | blast;93.60 |
| S8_vf_158225 | 14.69 | d__Bacteria;p__Bacteroidota;c__Bacteroidia;o__Bacteroidales;f__Tannerellaceae;g__Parabacteroides | 94 | blast;95.80 |
| S8_vf_161703 | 49.06 | d__Bacteria;p__Firmicutes;c__Clostridia;o__Oscillospirales;f__Ruminococcaceae;g__Ruminococcus_D | 99.1 | CRISPR;97.90 blast;96.80 iPHoP-RF;96.40 RaFAH;83.30 |
| S8_vf_16172 | 45.4 | d__Bacteria;p__Bacteroidota;c__Bacteroidia;o__Bacteroidales;f__Tannerellaceae;g__Parabacteroides | 95.9 | iPHoP-RF;97.10 blast;96.90 CRISPR;92.40 |
| S8_vf_16172 | 45.4 | d__Bacteria;p__Bacteroidota;c__Bacteroidia;o__Bacteroidales;f__Bacteroidaceae;g__Bacteroides | 94.9 | RaFAH;96.40 |
| S8_vf_166403 | 69.48 | d__Bacteria;p__Firmicutes;c__Clostridia;o__Oscillospirales;f__Oscillospiraceae;g__Flavonifractor | 95.2 | blast;96.60 iPHoP-RF;72.20 |
| S8_vf_166677 | 20.94 | d__Bacteria;p__Bacteroidota;c__Bacteroidia;o__Bacteroidales;f__Bacteroidaceae;g__Prevotella | 99.9 | blast;98.70 CRISPR;96.60 iPHoP-RF;93.10 RaFAH;90.50 |
| S8_vf_170878 | 39.33 | d__Bacteria;p__Firmicutes;c__Clostridia;o__Oscillospirales;f__Oscillospiraceae;g__CAG-83 | 91.2 | blast;93.60 iPHoP-RF;65.60 |
| S8_vf_183045 | 35.16 | d__Bacteria;p__Bacteroidota;c__Bacteroidia;o__Bacteroidales;f__Bacteroidaceae;g__Prevotella | 98.5 | CRISPR;98.90 blast;93.00 iPHoP-RF;90.50 |
| S8_vf_187742 | 91.82 | d__Bacteria;p__Firmicutes;c__Clostridia;o__Oscillospirales;f__Ruminococcaceae;g__Faecalibacterium | 99.6 | blast;96.20 RaFAH;95.10 CRISPR;86.20 iPHoP-RF;65.30 |
| S8_vf_187742 | 91.82 | d__Bacteria;p__Firmicutes;c__Bacilli;o__Lactobacillales;f__Streptococcaceae;g__Streptococcus | 94.6 | blast;96.20 |
| S8_vf_195032 | 5.64 | d__Bacteria;p__Actinobacteriota;c__Coriobacteriia;o__Coriobacteriales;f__Eggerthellaceae;g__Gordonibacter | 94.9 | CRISPR;96.40 iPHoP-RF;75.70 |
| S8_vf_195032 | 5.64 | d__Bacteria;p__Actinobacteriota;c__Coriobacteriia;o__Coriobacteriales;f__Eggerthellaceae;g__Eggerthella | 91.6 | blast;93.90 iPHoP-RF;69.30 |
| S8_vf_197061 | 60.03 | d__Bacteria;p__Actinobacteriota;c__Actinomycetia;o__Actinomycetales;f__Bifidobacteriaceae;g__Bifidobacterium | 98.8 | CRISPR;99.00 iPHoP-RF;97.10 blast;96.80 RaFAH;52.60 |
| S8_vf_201695 | 30.61 | d__Bacteria;p__Firmicutes;c__Clostridia;o__Lachnospirales;f__Anaerotignaceae;g__Anaerotignum | 94.2 | blast;95.90 iPHoP-RF;76.40 |
| S8_vf_208110 | 89.6 | d__Bacteria;p__Firmicutes;c__Clostridia;o__Oscillospirales;f__Oscillospiraceae;g__Intestinimonas | 92 | blast;94.20 iPHoP-RF;50.20 |
| S8_vf_208111 | 7.17 | d__Bacteria;p__Desulfobacterota;c__Desulfovibrionia;o__Desulfovibrionales;f__Desulfovibrionaceae;g__Bilophila | 90.8 | CRISPR;93.20 iPHoP-RF;60.30 |
| S8_vf_208115 | | d__Bacteria;p__Bacteroidota;c__Bacteroidia;o__Bacteroidales;f__Barnesiellaceae;g__Barnesiella | 94.2 | blast;95.90 iPHoP-RF;72.80 |
| S8_vf_208119 | 18.24 | d__Bacteria;p__Firmicutes;c__Clostridia;o__Oscillospirales;f__Ruminococcaceae;g__Faecalibacterium | 97.2 | CRISPR;97.90 RaFAH;52.10 |
| S8_vf_208120 | 99.62 | d__Bacteria;p__Bacteroidota;c__Bacteroidia;o__Bacteroidales;f__Bacteroidaceae;g__Bacteroides | 98.2 | RaFAH;93.90 blast;88.30 iPHoP-RF;53.80 |
| S8_vf_208120 | 99.62 | d__Bacteria;p__Bacteroidota;c__Bacteroidia;o__Bacteroidales;f__Tannerellaceae;g__Parabacteroides | 95.4 | blast;96.80 iPHoP-RF;57.90 |
| S8_vf_208126 | 48.51 | d__Bacteria;p__Firmicutes;c__Clostridia;o__Oscillospirales;f__Ruminococcaceae;g__Gemmiger | 91.6 | blast;93.90 iPHoP-RF;60.90 |
| S8_vf_208146 | 25.58 | d__Bacteria;p__Firmicutes;c__Negativicutes;o__Acidaminococcales;f__Acidaminococcaceae;g__Phascolarctobacterium | 93.1 | CRISPR;95.10 |
| S8_vf_28956 | 91.89 | d__Bacteria;p__Bacteroidota;c__Bacteroidia;o__Bacteroidales;f__Bacteroidaceae;g__Bacteroides | 99.6 | iPHoP-RF;96.40 RaFAH;94.20 blast;93.80 |
| S8_vf_28956 | 91.89 | d__Bacteria;p__Bacteroidota;c__Bacteroidia;o__Bacteroidales;f__Bacteroidaceae;g__Phocaeicola | 94.9 | iPHoP-RF;96.40 blast;92.70 |
| S8_vf_45776 | 88.6 | d__Bacteria;p__Firmicutes;c__Bacilli;o__Lactobacillales;f__Streptococcaceae;g__Lactococcus | 100 | RaFAH;99.20 blast;98.30 iPHoP-RF;73.90 |
| S8_vf_55476 | 64.72 | d__Bacteria;p__Firmicutes;c__Clostridia;o__Oscillospirales;f__Oscillospiraceae;g__Intestinimonas | 91.6 | blast;93.90 iPHoP-RF;82.20 |
| S8_vf_65522 | 12.03 | d__Bacteria;p__Firmicutes;c__Clostridia;o__Christensenellales;f__CAG-74;g__UBA11524 | 97.2 | blast;98.00 iPHoP-RF;83.10 CRISPR;55.60 |
| S8_vf_7243 | 95.82 | d__Bacteria;p__Actinobacteriota;c__Actinomycetia;o__Actinomycetales;f__Micrococcaceae;g__Rothia | 99.4 | RaFAH;99.50 |
| S8_vf_79769 | 31.62 | d__Bacteria;p__Bacteroidota;c__Bacteroidia;o__Bacteroidales;f__Bacteroidaceae;g__Prevotella | 98.4 | CRISPR;98.80 blast;96.70 iPHoP-RF;92.40 |
| S8_vf_93976 | 9.53 | d__Bacteria;p__Firmicutes;c__Clostridia;o__Christensenellales;f__CAG-138;g__UBA1685 | 95.3 | blast;96.70 iPHoP-RF;66.50 CRISPR;59.10 |
| S8_vs_102608 | 20 | d__Bacteria;p__Firmicutes;c__Clostridia;o__Oscillospirales;f__Ruminococcaceae;g__Gemmiger | 95.7 | blast;97.00 CRISPR;96.10 iPHoP-RF;90.80 |
| S8_vs_115958 | 68.75 | d__Bacteria;p__Firmicutes;c__Clostridia;o__Oscillospirales;f__Acutalibacteraceae;g__Ruminococcus_E | 91.6 | blast;93.90 iPHoP-RF;67.00 |
| S8_vs_125717 | 17.94 | d__Bacteria;p__Firmicutes;c__Clostridia;o__Oscillospirales;f__Oscillospiraceae;g__CAG-83 | 95.6 | blast;96.90 iPHoP-RF;64.30 |
| S8_vs_126582 | 98.15 | d__Bacteria;p__Bacteroidota;c__Bacteroidia;o__Bacteroidales;f__Tannerellaceae;g__Parabacteroides | 100 | RaFAH;99.70 iPHoP-RF;89.20 blast;86.20 CRISPR;69.40 |
| S8_vs_134805 | 91.02 | d__Bacteria;p__Firmicutes;c__Clostridia;o__Oscillospirales;f__Oscillospiraceae;g__Lawsonibacter | 91.5 | blast;93.80 iPHoP-RF;73.20 |
| S8_vs_145173 | 54.47 | d__Bacteria;p__Firmicutes;c__Clostridia;o__Oscillospirales;f__Oscillospiraceae;g__UBA9475 | 91.2 | blast;93.60 iPHoP-RF;59.00 |
| S8_vs_147670 | 31.23 | d__Bacteria;p__Firmicutes;c__Negativicutes;o__Veillonellales;f__Megasphaeraceae;g__Caecibacter | 96.7 | CRISPR;97.70 blast;78.80 |
| S8_vs_147697 | 41.71 | d__Bacteria;p__Bacteroidota;c__Bacteroidia;o__Bacteroidales;f__Bacteroidaceae;g__Bacteroides | 98.8 | iPHoP-RF;95.70 blast;90.90 RaFAH;89.00 |
| S8_vs_148389 | 54.68 | d__Bacteria;p__Bacteroidota;c__Bacteroidia;o__Bacteroidales;f__Bacteroidaceae;g__Prevotella | 99.6 | blast;96.80 RaFAH;94.40 iPHoP-RF;91.80 |
| S8_vs_173961 | 95.47 | d__Bacteria;p__Firmicutes;c__Clostridia;o__Lachnospirales;f__Lachnospiraceae;g__Agathobacter | 99.6 | CRISPR;98.40 blast;90.90 RaFAH;89.00 iPHoP-RF;87.60 |
| S8_vs_173961 | 95.47 | d__Bacteria;p__Firmicutes;c__Clostridia;o__Lachnospirales;f__Lachnospiraceae;g__Roseburia | 99.4 | CRISPR;97.50 iPHoP-RF;96.40 blast;91.80 RaFAH;89.00 |
| S8_vs_173961 | 95.47 | d__Bacteria;p__Firmicutes;c__Clostridia;o__Lachnospirales;f__Lachnospiraceae;g__RUG115 | 97.9 | iPHoP-RF;93.40 blast;90.90 RaFAH;89.00 |
| S8_vs_176604 | 21.14 | d__Bacteria;p__Firmicutes;c__Clostridia;o__Christensenellales;f__CAG-138;g__SFEL01 | 94.9 | blast;96.40 iPHoP-RF;81.60 CRISPR;71.90 |
| S8_vs_176604 | 21.14 | d__Bacteria;p__Firmicutes;c__Clostridia;o__Christensenellales;f__CAG-138;g__SFEL01 | 94.2 | blast;95.90 iPHoP-RF;83.10 |
| S8_vs_179208 | 39.41 | d__Bacteria;p__Bacteroidota;c__Bacteroidia;o__Bacteroidales;f__Rikenellaceae;g__Alistipes | 95.4 | blast;96.80 iPHoP-RF;80.80 |
| S8_vs_179208 | 39.41 | d__Bacteria;p__Bacteroidota;c__Bacteroidia;o__Bacteroidales;f__Bacteroidaceae;g__Bacteroides | 91.7 | RaFAH;85.10 blast;79.80 |
| S8_vs_179529 | 98.83 | d__Bacteria;p__Firmicutes;c__Clostridia;o__Oscillospirales;f__Ruminococcaceae;g__Faecalibacterium | 100 | CRISPR;98.50 RaFAH;98.00 blast;95.30 iPHoP-RF;86.70 |
| S8_vs_182288 | 85.94 | d__Bacteria;p__Bacteroidota;c__Bacteroidia;o__Bacteroidales;f__Tannerellaceae;g__Parabacteroides | 100 | CRISPR;99.00 RaFAH;98.40 iPHoP-RF;96.40 blast;93.80 |
| S8_vs_206526 | 81.78 | d__Bacteria;p__Firmicutes;c__Clostridia;o__Lachnospirales;f__Lachnospiraceae;g__Roseburia | 97.9 | CRISPR;98.50 blast;92.20 iPHoP-RF;87.30 RaFAH;50.70 |
| S8_vs_206526 | 81.78 | d__Bacteria;p__Firmicutes;c__Clostridia;o__Lachnospirales;f__Lachnospiraceae;g__RUG115 | 97.3 | CRISPR;98.10 iPHoP-RF;56.20 RaFAH;50.70 |
| S8_vs_206526 | 81.78 | d__Bacteria;p__Firmicutes;c__Clostridia;o__Lachnospirales;f__Lachnospiraceae;g__UBA11774 | 97.3 | CRISPR;98.10 iPHoP-RF;66.30 |
| S8_vs_206526 | 81.78 | d__Bacteria;p__Firmicutes;c__Clostridia;o__Lachnospirales;f__Lachnospiraceae;g__Lachnospira | 95 | blast;96.50 iPHoP-RF;50.70 |
| S8_vs_208114 | 9.13 | d__Bacteria;p__Desulfobacterota;c__Desulfovibrionia;o__Desulfovibrionales;f__Desulfovibrionaceae;g__Mailhella | 90.1 | blast;92.60 iPHoP-RF;81.40 |
| S8_vs_28311 | 99.02 | d__Bacteria;p__Bacteroidota;c__Bacteroidia;o__Bacteroidales;f__Bacteroidaceae;g__Bacteroides | 100 | RaFAH;97.40 blast;97.40 iPHoP-RF;96.70 CRISPR;54.10 |
| S8_vs_40014 | 80.96 | d__Bacteria;p__Bacteroidota;c__Bacteroidia;o__Bacteroidales;f__Rikenellaceae;g__Alistipes | 94.6 | blast;96.20 iPHoP-RF;95.70 |
| S8_vs_4153 | 5.07 | d__Bacteria;p__Bacteroidota;c__Bacteroidia;o__Bacteroidales;f__Barnesiellaceae;g__Barnesiella | 97.9 | blast;98.50 iPHoP-RF;96.40 CRISPR;53.40 |
| S8_vs_50431 | 20.17 | d__Bacteria;p__Firmicutes;c__Clostridia;o__Oscillospirales;f__Oscillospiraceae;g__Dysosmobacter | 91.5 | blast;93.80 iPHoP-RF;50.70 |
| S8_vs_51368 | 31.95 | d__Bacteria;p__Firmicutes;c__Clostridia;o__Oscillospirales;f__Oscillospiraceae;g__CAG-170 | 91.5 | blast;93.80 iPHoP-RF;85.70 |
| S8_vs_5211 | 70.75 | d__Bacteria;p__Firmicutes;c__Clostridia;o__Lachnospirales;f__Lachnospiraceae;g__Massilistercora | 90.2 | blast;92.70 iPHoP-RF;58.40 |
| S8_vs_54183 | 20.38 | d__Bacteria;p__Firmicutes;c__Clostridia;o__Oscillospirales;f__Oscillospiraceae;g__CAG-170 | 91.8 | CRISPR;94.10 blast;90.90 iPHoP-RF;81.90 |
| S8_vs_59736 | 26.65 | d__Bacteria;p__Firmicutes;c__Bacilli;o__Erysipelotrichales;f__Erysipelatoclostridiaceae;g__Catenibacterium | 95.6 | blast;96.90 |
| S8_vs_63277 | 75.64 | d__Bacteria;p__Firmicutes;c__Clostridia;o__Oscillospirales;f__Ruminococcaceae;g__Faecalibacterium | 97.8 | blast;98.40 iPHoP-RF;88.20 |
| S8_vs_72117 | 19.43 | d__Bacteria;p__Bacteroidota;c__Bacteroidia;o__Bacteroidales;f__Bacteroidaceae;g__Bacteroides | 95.4 | iPHoP-RF;94.40 RaFAH;71.80 |
| S8_vs_9114 | 28.26 | d__Bacteria;p__Bacteroidota;c__Bacteroidia;o__Bacteroidales;f__Rikenellaceae;g__Alistipes_A | 91.1 | blast;93.50 iPHoP-RF;82.80 CRISPR;64.80 |
| S8_vs_99362 | 99.97 | d__Bacteria;p__Bacteroidota;c__Bacteroidia;o__Bacteroidales;f__Bacteroidaceae;g__Bacteroides | 100 | RaFAH;98.30 blast;96.50 iPHoP-RF;93.40 |
| S8_vs_99362 | 99.97 | d__Bacteria;p__Bacteroidota;c__Bacteroidia;o__Bacteroidales;f__Bacteroidaceae;g__Paraprevotella | 90.2 | blast;92.70 |

| | | | | |
|---|---|---|---|---|
| S8_vs_9987 | 99.87 | d__Bacteria;p__Bacteroidota;c__Bacteroidia;o__Bacteroidales;f__Rikenellaceae;g__Alistipes | 100 | blast;97.60 RaFAH;97.40 iPHoP-RF;93.40 |
| S8_vs_9987 | 99.87 | d__Bacteria;p__Bacteroidota;c__Bacteroidia;o__Bacteroidales;f__Rikenellaceae;g__Alistipes_A | 96.3 | RaFAH;97.40 |
| S9_vf_13507 | 66.3 | d__Bacteria;p__Bacteroidota;c__Bacteroidia;o__Bacteroidales;f__Bacteroidaceae;g__Prevotella | 95.6 | blast;96.90 iPHoP-RF;94.70 |
| S9_vf_15012 | 97.67 | d__Bacteria;p__Firmicutes;c__Bacilli;o__Lactobacillales;f__Streptococcaceae;g__Streptococcus | 100 | RaFAH;99.70 CRISPR;98.30 blast;94.40 iPHoP-RF;92.80 |
| S9_vf_19284 | 15.56 | d__Bacteria;p__Firmicutes;c__Bacilli;o__Lactobacillales;f__Streptococcaceae;g__Streptococcus | 93 | CRISPR;95.00 |
| S9_vf_4115 | 96.62 | d__Bacteria;p__Firmicutes;c__Bacilli;o__Lactobacillales;f__Streptococcaceae;g__Streptococcus | 100 | RaFAH;99.70 CRISPR;99.00 blast;96.80 iPHoP-RF;96.70 |
| S9_vf_51656 | 29.36 | d__Bacteria;p__Bacteroidota;c__Bacteroidia;o__Bacteroidales;f__Bacteroidaceae;g__Prevotella | 99 | CRISPR;99.00 blast;96.30 iPHoP-RF;96.10 RaFAH;59.50 |
| S9_vf_64844 | 89.68 | d__Bacteria;p__Firmicutes;c__Clostridia;o__Oscillospirales;f__Oscillospiraceae;g__CAG-83 | 93.4 | RaFAH;95.30 blast;50.00 |
| S9_vf_64844 | 89.68 | d__Bacteria;p__Firmicutes;c__Clostridia;o__Oscillospirales;f__Oscillospiraceae;g__Dysosmobacter | 93.4 | RaFAH;95.30 |
| S9_vf_73311 | 18.82 | d__Bacteria;p__Firmicutes;c__Clostridia;o__Lachnospirales;f__CAG-274;g__CAG-274 | 97.2 | CRISPR;98.00 blast;96.80 iPHoP-RF;78.70 |
| S9_vf_76769 | 53.32 | d__Bacteria;p__Bacteroidota;c__Bacteroidia;o__Bacteroidales;f__Bacteroidaceae;g__Prevotella | 92.6 | RaFAH;94.70 |
| S9_vf_90856 | 94.47 | d__Bacteria;p__Firmicutes;c__Bacilli;o__Lactobacillales;f__Streptococcaceae;g__Streptococcus | 100 | RaFAH;99.80 CRISPR;99.00 blast;95.80 iPHoP-RF;94.40 |
| S9_vf_90866 | 15.47 | d__Bacteria;p__Firmicutes;c__Bacilli;o__Lactobacillales;f__Streptococcaceae;g__Streptococcus | 99.3 | iPHoP-RF;97.10 blast;95.70 RaFAH;90.00 |
| S9_vf_934 | 17.76 | d__Bacteria;p__Actinobacteriota;c__Actinomycetia;o__Mycobacteriales;f__Mycobacteriaceae;g__Mycobacterium | 92.2 | iPHoP-RF;94.40 |
| S9_vs_14235 | 12.73 | d__Bacteria;p__Firmicutes;c__Clostridia;o__Oscillospirales;f__Oscillospiraceae;g__An92 | 91 | blast;93.40 iPHoP-RF;87.00 CRISPR;73.90 |
| S9_vs_14235 | 12.73 | d__Bacteria;p__Firmicutes;c__Clostridia;o__Oscillospirales;f__Oscillospiraceae;g__Flavonifractor | 90.4 | blast;92.90 iPHoP-RF;79.00 CRISPR;60.90 |
| S9_vs_1966 | 61.19 | d__Bacteria;p__Firmicutes;c__Clostridia;o__Lachnospirales;f__Lachnospiraceae;g__Bariatricus | 91.6 | blast;93.90 iPHoP-RF;77.30 |
| S9_vs_19689 | 9.42 | d__Bacteria;p__Firmicutes;c__Clostridia;o__Oscillospirales;f__Butyricicoccaceae;g__Agathobaculum | 91.6 | blast;93.90 iPHoP-RF;68.60 |
| S9_vs_21850 | 63.18 | d__Bacteria;p__Bacteroidota;c__Bacteroidia;o__Bacteroidales;f__Barnesiellaceae;g__Barnesiella | 97.2 | blast;96.90 iPHoP-RF;85.10 CRISPR;84.10 RaFAH;68.00 |
| S9_vs_26179 | 48.62 | d__Bacteria;p__Firmicutes;c__Clostridia;o__Oscillospirales;f__Acutalibacteraceae;g__UMGS1071 | 95.7 | blast;97.00 iPHoP-RF;83.30 CRISPR;81.20 |
| S9_vs_2799 | 10.92 | d__Bacteria;p__Bacteroidota;c__Bacteroidia;o__Bacteroidales;f__Rikenellaceae;g__Alistipes | 95.9 | iPHoP-RF;97.10 blast;96.90 |
| S9_vs_28197 | 87.18 | d__Bacteria;p__Bacteroidota;c__Bacteroidia;o__Bacteroidales;f__Bacteroidaceae;g__Bacteroides | 100 | RaFAH;99.40 iPHoP-RF;97.10 blast;96.30 |
| S9_vs_28197 | 87.18 | d__Bacteria;p__Bacteroidota;c__Bacteroidia;o__Bacteroidales;f__Bacteroidaceae;g__Phocaeicola | 95.3 | iPHoP-RF;96.70 |
| S9_vs_32588 | 67.17 | d__Bacteria;p__Firmicutes;c__Clostridia;o__Lachnospirales;f__Lachnospiraceae;g__Dorea | 95 | RaFAH;96.50 |
| S9_vs_32588 | 67.17 | d__Bacteria;p__Firmicutes;c__Clostridia;o__Lachnospirales;f__Lachnospiraceae;g__Dorea_A | 95 | RaFAH;96.50 |
| S9_vs_32588 | 67.17 | d__Bacteria;p__Firmicutes;c__Clostridia;o__Oscillospirales;f__Oscillospiraceae;g__Intestinimonas | 91.6 | blast;93.90 iPHoP-RF;73.70 |
| S9_vs_34898 | 91.82 | d__Bacteria;p__Firmicutes;c__Clostridia;o__Lachnospirales;f__Lachnospiraceae;g__Sellimonas | 93.9 | blast;95.70 iPHoP-RF;89.50 |
| S9_vs_37437 | 14.86 | d__Bacteria;p__Firmicutes;c__Bacilli;o__Erysipelotrichales;f__Erysipelotrichaceae;g__Holdemania | 91.6 | blast;93.90 CRISPR;91.60 iPHoP-RF;75.40 |
| S9_vs_41371 | 44.35 | d__Bacteria;p__Firmicutes;c__Clostridia;o__Lachnospirales;f__Lachnospiraceae;g__Enterocloster | 94 | blast;95.80 iPHoP-RF;85.10 |
| S9_vs_4343 | 24.4 | d__Bacteria;p__Firmicutes;c__Clostridia;o__Lachnospirales;f__Lachnospiraceae;g__Enterocloster | 96.6 | blast;97.60 CRISPR;96.60 iPHoP-RF;90.50 |
| S9_vs_44331 | 98.52 | d__Bacteria;p__Firmicutes;c__Clostridia;o__Lachnospirales;f__Lachnospiraceae;g__Ruminococcus_B | 98.1 | blast;98.60 iPHoP-RF;94.40 CRISPR;87.70 |
| S9_vs_4558 | 65.18 | d__Bacteria;p__Firmicutes;c__Clostridia;o__Oscillospirales;f__Oscillospiraceae;g__Flavonifractor | 90.2 | blast;92.70 iPHoP-RF;79.00 |
| S9_vs_46498 | 26.43 | d__Bacteria;p__Firmicutes;c__Bacilli;o__Erysipelotrichales;f__Erysipelatoclostridiaceae;g__Erysipelatoclostridium | 96.7 | blast;97.70 iPHoP-RF;74.10 |
| S9_vs_48240 | 63.98 | d__Bacteria;p__Firmicutes;c__Clostridia;o__Lachnospirales;f__Lachnospiraceae;g__Hungatella | 92.5 | blast;94.60 iPHoP-RF;94.40 |
| S9_vs_48347 | 25.89 | d__Bacteria;p__Firmicutes;c__Clostridia;o__Oscillospirales;f__Ruminococcaceae;g__Negativibacillus | 94.2 | blast;95.90 iPHoP-RF;85.40 CRISPR;59.60 |
| S9_vs_48424 | 86.27 | d__Bacteria;p__Bacteroidota;c__Bacteroidia;o__Bacteroidales;f__Rikenellaceae;g__Alistipes | 99.3 | blast;96.80 RaFAH;90.20 iPHoP-RF;54.90 |
| S9_vs_50439 | 93.5 | d__Bacteria;p__Firmicutes;c__Clostridia;o__Oscillospirales;f__Oscillospiraceae;g__Flavonifractor | 98.1 | blast;98.60 iPHoP-RF;74.30 |
| S9_vs_55738 | 76 | d__Bacteria;p__Firmicutes;c__Clostridia;o__Lachnospirales;f__Lachnospiraceae;g__Enterocloster | 94.6 | blast;96.20 iPHoP-RF;87.00 CRISPR;69.00 |
| S9_vs_5676 | 19.1 | d__Bacteria;p__Firmicutes;c__Clostridia;o__Oscillospirales;f__Oscillospiraceae;g__Intestinimonas | 92.6 | blast;94.70 iPHoP-RF;74.60 |
| S9_vs_58086 | 53.1 | d__Bacteria;p__Firmicutes;c__Clostridia;o__Lachnospirales;f__Lachnospiraceae;g__Dorea_A | 94.3 | blast;96.00 iPHoP-RF;80.60 |
| S9_vs_58086 | 53.1 | d__Bacteria;p__Firmicutes;c__Clostridia;o__Lachnospirales;f__Lachnospiraceae;g__Dorea | 91.5 | blast;93.80 iPHoP-RF;83.60 |
| S9_vs_62803 | 39.71 | d__Bacteria;p__Firmicutes;c__Clostridia;o__Lachnospirales;f__Lachnospiraceae;g__Sellimonas | 94.9 | blast;96.40 iPHoP-RF;84.50 |
| S9_vs_62803 | 39.71 | d__Bacteria;p__Firmicutes;c__Clostridia;o__Lachnospirales;f__Lachnospiraceae;g__Lachnoclostridium_B | 92.6 | iPHoP-RF;94.70 |
| S9_vs_66877 | 29.56 | d__Bacteria;p__Firmicutes;c__Clostridia;o__Oscillospirales;f__Ruminococcaceae;g__Ruthenibacterium | 95.3 | blast;96.70 CRISPR;95.50 iPHoP-RF;92.10 |
| S9_vs_67792 | 30.5 | d__Bacteria;p__Firmicutes;c__Negativicutes;o__Veillonellales;f__Dialisteraceae;g__Dialister | 96.6 | blast;97.60 iPHoP-RF;86.00 CRISPR;60.90 |
| S9_vs_69322 | 20.79 | d__Bacteria;p__Firmicutes;c__Clostridia;o__Oscillospirales;f__Oscillospiraceae;g__Dysosmobacter | 90.7 | blast;93.10 iPHoP-RF;82.20 |
| S9_vs_75050 | 26.71 | d__Bacteria;p__Firmicutes;c__Clostridia;o__Lachnospirales;f__Lachnospiraceae;g__CAG-127 | 96.4 | CRISPR;97.50 blast;96.80 iPHoP-RF;86.00 |
| S9_vs_83108 | 82.91 | d__Bacteria;p__Bacteroidota;c__Bacteroidia;o__Bacteroidales;f__Bacteroidaceae;g__Bacteroides | 99.4 | iPHoP-RF;97.10 blast;93.90 RaFAH;91.80 |
| S9_vs_83108 | 82.91 | d__Bacteria;p__Bacteroidota;c__Bacteroidia;o__Bacteroidales;f__Bacteroidaceae;g__Phocaeicola | 98.1 | CRISPR;98.60 iPHoP-RF;97.10 blast;91.90 |
| S9_vs_84046 | 20.91 | d__Bacteria;p__Firmicutes;c__Clostridia;o__Oscillospirales;f__Oscillospiraceae;g__An92 | 90.7 | blast;93.10 iPHoP-RF;70.60 |
| S9_vs_88286 | 10.88 | d__Bacteria;p__Firmicutes;c__Clostridia;o__Lachnospirales;f__Lachnospiraceae;g__Eisenbergiella | 98.1 | CRISPR;98.60 blast;95.70 iPHoP-RF;95.40 |
| S9_vs_90727 | 57.49 | d__Bacteria;p__Firmicutes;c__Clostridia;o__Oscillospirales;f__Acutalibacteraceae;g__Ruminococcus_E | 95.6 | blast;96.90 iPHoP-RF;88.20 |
| S9_vs_90846 | 37.71 | d__Bacteria;p__Firmicutes;c__Bacilli;o__Erysipelotrichales;f__Erysipelotrichaceae;g__Clostridium_AQ | 97.5 | blast;98.20 iPHoP-RF;62.70 |
| S9_vs_90848 | 51.41 | d__Bacteria;p__Firmicutes;c__Clostridia;o__Lachnospirales;f__Lachnospiraceae;g__Enterocloster | 98.7 | CRISPR;99.00 blast;93.90 iPHoP-RF;91.40 |
| S9_vs_90849 | 70.89 | d__Bacteria;p__Firmicutes;c__Clostridia;o__Lachnospirales;f__Anaerotignaceae;g__Anaerotignum | 95 | CRISPR;96.50 blast;93.70 iPHoP-RF;92.10 |
| S9_vs_90851 | 89.56 | d__Bacteria;p__Firmicutes;c__Clostridia;o__Clostridiales;f__Clostridiaceae;g__Clostridium | 95.6 | RaFAH;96.90 |
| S9_vs_90851 | 89.56 | d__Bacteria;p__Firmicutes;c__Clostridia;o__Clostridiales;f__Clostridiaceae;g__Clostridium_F | 95.6 | RaFAH;96.90 |
| S9_vs_90851 | 89.56 | d__Bacteria;p__Firmicutes;c__Clostridia;o__Clostridiales;f__Clostridiaceae;g__Clostridium_P | 95.6 | RaFAH;96.90 |
| S9_vs_90859 | 43.06 | d__Bacteria;p__Firmicutes;c__Clostridia;o__Lachnospirales;f__Lachnospiraceae;g__Blautia_A | 95.3 | blast;93.90 iPHoP-RF;75.90 RaFAH;73.20 |
| S9_vs_90860 | 20.66 | d__Bacteria;p__Firmicutes;c__Clostridia;o__Oscillospirales;f__Acutalibacteraceae;g__UMGS1591 | 93.4 | blast;95.30 iPHoP-RF;67.00 |

| WMS_Virus | AAI to closest RaFAH reference | Host genus | Confidence score | List of methods |
|---|---|---|---|---|
| S1-wms-combined_100430 | 99.71 | d__Bacteria;p__Firmicutes;c__Clostridia;o__Lachnospirales;f__Lachnospiraceae;g__Lachnospira | 99.3 | CRISPR;98.90 RaFAH;73.00 |
| S1-wms-combined_100430 | 99.71 | d__Bacteria;p__Firmicutes;c__Clostridia;o__Lachnospirales;f__Lachnospiraceae;g__Eubacterium_G | 95.4 | blast;94.10 RaFAH;73.00 |
| S1-wms-combined_100430 | 99.71 | d__Bacteria;p__Firmicutes;c__Clostridia;o__Lachnospirales;f__Lachnospiraceae;g__Roseburia | 94.3 | blast;96.00 iPHoP-RF;62.20 |
| S1-wms-combined_107766 | 92.99 | d__Bacteria;p__Bacteroidota;c__Bacteroidia;o__Bacteroidales;f__Rikenellaceae;g__Alistipes | 98.8 | iPHoP-RF;97.10 blast;96.90 RaFAH;83.70 |
| S1-wms-combined_118167 | 33.35 | d__Bacteria;p__Firmicutes;c__Clostridia;o__Oscillospirales;f__Ruminococcaceae;g__Faecalibacterium | 99.1 | CRISPR;96.40 blast;95.10 RaFAH;90.80 iPHoP-RF;89.50 |
| S1-wms-combined_131270 | 21.6 | d__Bacteria;p__Firmicutes;c__Clostridia;o__Oscillospirales;f__Ruminococcaceae;g__Faecalibacterium | 99 | CRISPR;99.20 blast;94.50 iPHoP-RF;84.50 |
| S1-wms-combined_132934 | 48.4 | d__Bacteria;p__Firmicutes;c__Clostridia;o__Oscillospirales;f__Oscillospiraceae;g__ER4 | 92.7 | blast;94.80 iPHoP-RF;64.80 |
| S1-wms-combined_136097 | 4.5 | d__Bacteria;p__Firmicutes;c__Clostridia;o__Oscillospirales;f__Oscillospiraceae;g__UBA738 | 92 | blast;94.20 iPHoP-RF;53.00 |
| S1-wms-combined_143459 | 18.79 | d__Bacteria;p__Desulfobacterota;c__Desulfovibrionia;o__Desulfovibrionales;f__Desulfovibrionaceae;g__Bilophila | 95.2 | blast;96.60 iPHoP-RF;65.80 CRISPR;58.60 |
| S1-wms-combined_143459 | 18.79 | d__Bacteria;p__Desulfobacterota;c__Desulfovibrionia;o__Desulfovibrionales;f__Desulfovibrionaceae;g__Bilophila | 90.8 | blast;93.20 iPHoP-RF;66.80 |
| S1-wms-combined_153670 | | d__Bacteria;p__Bacteroidota;c__Bacteroidia;o__Bacteroidales;f__Barnesiellaceae;g__Barnesiella | 94.2 | blast;95.90 iPHoP-RF;72.20 |
| S1-wms-combined_156637 | 11 | d__Bacteria;p__Firmicutes;c__Clostridia;o__Oscillospirales;f__Oscillospiraceae;g__ER4 | 93 | blast;95.00 iPHoP-RF;60.30 |
| S1-wms-combined_159544 | 36.49 | d__Bacteria;p__Desulfobacterota;c__Desulfovibrionia;o__Desulfovibrionales;f__Desulfovibrionaceae;g__Bilophila | 94 | blast;95.80 iPHoP-RF;79.30 CRISPR;57.40 |
| S1-wms-combined_164485 | 4.67 | d__Bacteria;p__Firmicutes;c__Clostridia;o__Oscillospirales;f__Oscillospiraceae;g__ER4 | 90.4 | blast;92.90 |
| S1-wms-combined_164894 | 72.02 | d__Bacteria;p__Firmicutes;c__Clostridia;o__Oscillospirales;f__Ruminococcaceae;g__Faecalibacterium | 91.8 | blast;94.10 iPHoP-RF;70.90 |
| S1-wms-combined_166392 | 37.1 | d__Bacteria;p__Firmicutes;c__Clostridia;o__Oscillospirales;f__Ruminococcaceae;g__Gemmiger | 95.7 | blast;97.00 iPHoP-RF;95.70 CRISPR;78.40 |
| S1-wms-combined_169788 | 5.96 | d__Bacteria;p__Firmicutes;c__Clostridia;o__Oscillospirales;f__Oscillospiraceae;g__CAG-83 | 93.4 | blast;95.30 iPHoP-RF;87.00 |
| S1-wms-combined_17651 | 9.21 | d__Bacteria;p__Firmicutes;c__Clostridia;o__Oscillospirales;f__Oscillospiraceae;g__ER4 | 90.6 | blast;93.00 iPHoP-RF;65.10 |
| S1-wms-combined_185336 | | d__Bacteria;p__Firmicutes;c__Clostridia;o__Oscillospirales;f__Oscillospiraceae;g__ER4 | 91.7 | blast;94.00 iPHoP-RF;63.50 |
| S1-wms-combined_186878 | 42.39 | d__Bacteria;p__Firmicutes;c__Clostridia;o__Oscillospirales;f__Oscillospiraceae;g__ER4 | 91.6 | blast;93.90 iPHoP-RF;76.40 |
| S1-wms-combined_19031 | 37.63 | d__Bacteria;p__Desulfobacterota;c__Desulfovibrionia;o__Desulfovibrionales;f__Desulfovibrionaceae;g__Bilophila | 98.2 | blast;98.70 iPHoP-RF;76.10 |
| S1-wms-combined_19031 | 37.63 | d__Bacteria;p__Desulfobacterota;c__Desulfovibrionia;o__Desulfovibrionales;f__Desulfovibrionaceae;g__Bilophila | 91.4 | blast;93.70 iPHoP-RF;78.30 |
| S1-wms-combined_212892 | 80.18 | d__Bacteria;p__Firmicutes;c__Clostridia;o__Lachnospirales;f__Lachnospiraceae;g__Anaerobutyricum | 98.1 | CRISPR;98.60 blast;91.50 iPHoP-RF;80.00 |
| S1-wms-combined_220022 | 6.93 | d__Bacteria;p__Firmicutes;c__Clostridia;o__Oscillospirales;f__Ruminococcaceae;g__Gemmiger | 93.2 | blast;95.20 iPHoP-RF;88.20 CRISPR;76.80 |
| S1-wms-combined_227003 | 41.88 | d__Bacteria;p__Firmicutes;c__Clostridia;o__Oscillospirales;f__Oscillospiraceae;g__CAG-110 | 94.2 | blast;95.90 iPHoP-RF;87.00 |
| S1-wms-combined_227917 | 27.55 | d__Bacteria;p__Firmicutes;c__Clostridia;o__Oscillospirales;f__Ruminococcaceae;g__Gemmiger | 95.2 | blast;96.60 iPHoP-RF;95.40 CRISPR;92.50 |
| S1-wms-combined_231411 | 3.46 | d__Bacteria;p__Desulfobacterota;c__Desulfovibrionia;o__Desulfovibrionales;f__Desulfovibrionaceae;g__Bilophila | 93 | blast;95.00 iPHoP-RF;73.20 CRISPR;51.00 |
| S1-wms-combined_231411 | 3.46 | d__Bacteria;p__Desulfobacterota;c__Desulfovibrionia;o__Desulfovibrionales;f__Desulfovibrionaceae;g__Bilophila | 90.2 | blast;92.70 iPHoP-RF;76.40 |
| S1-wms-combined_23248 | 61.01 | d__Bacteria;p__Firmicutes;c__Clostridia;o__Oscillospirales;f__Ruminococcaceae;g__Faecalibacterium | 95 | blast;96.50 iPHoP-RF;68.90 |
| S1-wms-combined_246923 | 43.06 | d__Bacteria;p__Bacteroidota;c__Bacteroidia;o__Bacteroidales;f__UBA11471;g__UBA11471 | 93.9 | blast;95.70 iPHoP-RF;68.60 |
| S1-wms-combined_248622 | 6.16 | d__Bacteria;p__Bacteroidota;c__Bacteroidia;o__Bacteroidales;f__Bacteroidaceae;g__Bacteroides | 95.9 | iPHoP-RF;97.10 blast;90.50 |
| S1-wms-combined_248622 | 6.16 | d__Bacteria;p__Bacteroidota;c__Bacteroidia;o__Bacteroidales;f__Bacteroidaceae;g__Phocaeicola | 94.5 | iPHoP-RF;96.10 blast;93.80 |
| S1-wms-combined_263562 | 99.98 | d__Bacteria;p__Bacteroidota;c__Bacteroidia;o__Bacteroidales;f__Bacteroidaceae;g__Bacteroides | 100 | RaFAH;99.80 blast;91.80 iPHoP-RF;82.20 |
| S1-wms-combined_279492 | 44.35 | d__Bacteria;p__Firmicutes;c__Clostridia;o__Oscillospirales;f__Ruminococcaceae;g__Gemmiger | 97.8 | blast;98.40 iPHoP-RF;96.40 CRISPR;58.00 |
| S1-wms-combined_285012 | 37.37 | d__Bacteria;p__Firmicutes;c__Clostridia;o__Oscillospirales;f__Ruminococcaceae;g__Faecalibacterium | 95 | CRISPR;96.50 iPHoP-RF;54.40 |
| S1-wms-combined_285012 | 37.37 | d__Bacteria;p__Firmicutes;c__Clostridia;o__Oscillospirales;f__Oscillospiraceae;g__Dysosmobacter | 90 | blast;92.50 |
| S1-wms-combined_302166 | 9.7 | d__Bacteria;p__Firmicutes;c__Clostridia;o__Christensenellales;f__CAG-74;g__UBA11524 | 95.6 | blast;96.90 iPHoP-RF;92.40 |
| S1-wms-combined_302166 | 9.7 | d__Bacteria;p__Firmicutes;c__Clostridia;o__Christensenellales;f__CAG-74;g__UBA11524 | 95.6 | blast;96.90 iPHoP-RF;84.50 |
| S1-wms-combined_304377 | 48.66 | d__Bacteria;p__Firmicutes;c__Clostridia;o__Oscillospirales;f__Ruminococcaceae;g__Faecalibacterium | 95.6 | blast;96.90 iPHoP-RF;78.50 |
| S1-wms-combined_307323 | 92.27 | d__Bacteria;p__Actinobacteriota;c__Actinomycetia;o__Actinomycetales;f__Bifidobacteriaceae;g__Bifidobacterium | 98.7 | CRISPR;99.00 iPHoP-RF;97.10 blast;96.90 |
| S1-wms-combined_308848 | 5.29 | d__Bacteria;p__Verrucomicrobiota;c__Verrucomicrobiae;o__Verrucomicrobiales;f__Akkermansiaceae;g__Akkermansia | 95.9 | CRISPR;97.10 blast;96.90 iPHoP-RF;93.10 |
| S1-wms-combined_32863 | 10.89 | d__Bacteria;p__Bacteroidota;c__Bacteroidia;o__Bacteroidales;f__Tannerellaceae;g__Parabacteroides | 95.6 | blast;96.90 iPHoP-RF;69.50 |
| S1-wms-combined_330329 | 86.77 | d__Bacteria;p__Bacteroidota;c__Bacteroidia;o__Bacteroidales;f__Bacteroidaceae;g__Bacteroides | 98.2 | RaFAH;97.70 iPHoP-RF;72.40 |
| S1-wms-combined_331544 | 89.75 | d__Bacteria;p__Actinobacteriota;c__Actinomycetia;o__Actinomycetales;f__Bifidobacteriaceae;g__Bifidobacterium | 99.9 | CRISPR;99.60 blast;96.90 iPHoP-RF;96.70 RaFAH;75.50 |
| S1-wms-combined_331896 | | d__Bacteria;p__Firmicutes;c__Clostridia;o__Oscillospirales;f__Oscillospiraceae;g__ER4 | 91.6 | blast;93.90 iPHoP-RF;58.40 |
| S1-wms-combined_340558 | 85.48 | d__Bacteria;p__Bacteroidota;c__Bacteroidia;o__Bacteroidales;f__Rikenellaceae;g__Alistipes | 99.7 | blast;96.80 iPHoP-RF;96.10 RaFAH;95.90 |
| S1-wms-combined_340558 | 85.48 | d__Bacteria;p__Bacteroidota;c__Bacteroidia;o__Bacteroidales;f__Rikenellaceae;g__Alistipes_A | 99.1 | RaFAH;95.90 blast;91.80 |
| S1-wms-combined_351889 | 29.41 | d__Bacteria;p__Firmicutes;c__Clostridia;o__Oscillospirales;f__Oscillospiraceae;g__ER4 | 96.2 | CRISPR;97.30 blast;96.20 iPHoP-RF;84.50 |
| S1-wms-combined_353757 | 21.95 | d__Bacteria;p__Firmicutes;c__Clostridia;o__Oscillospirales;f__Oscillospiraceae;g__ER4 | 93.2 | blast;95.20 iPHoP-RF;84.80 |
| S1-wms-combined_357373 | | d__Bacteria;p__Bacteroidota;c__Bacteroidia;o__Bacteroidales;f__Barnesiellaceae;g__Barnesiella | 91.8 | blast;94.10 iPHoP-RF;86.30 |
| S1-wms-combined_360285 | 17.11 | d__Bacteria;p__Desulfobacterota;c__Desulfovibrionia;o__Desulfovibrionales;f__Desulfovibrionaceae;g__Bilophila | 97.2 | CRISPR;98.00 blast;96.60 iPHoP-RF;89.80 |
| S1-wms-combined_360285 | 17.11 | d__Bacteria;p__Desulfobacterota;c__Desulfovibrionia;o__Desulfovibrionales;f__Desulfovibrionaceae;g__Bilophila | 94.2 | blast;95.90 iPHoP-RF;88.20 |
| S1-wms-combined_373308 | 6.25 | d__Bacteria;p__Firmicutes;c__Clostridia;o__Oscillospirales;f__Oscillospiraceae;g__CAG-83 | 97.9 | blast;98.50 iPHoP-RF;77.30 |
| S1-wms-combined_374219 | 10.38 | d__Bacteria;p__Firmicutes;c__Clostridia;o__Oscillospirales;f__Oscillospiraceae;g__Flavonifractor | 91.5 | blast;93.80 iPHoP-RF;79.80 |
| S1-wms-combined_394574 | 96.28 | d__Bacteria;p__Bacteroidota;c__Bacteroidia;o__Bacteroidales;f__Tannerellaceae;g__Parabacteroides | 91.6 | blast;93.90 iPHoP-RF;75.70 |
| S1-wms-combined_394574 | 96.28 | d__Bacteria;p__Bacteroidota;c__Bacteroidia;o__Bacteroidales;f__Marinifilaceae;g__Odoribacter | 90 | RaFAH;89.50 CRISPR;64.40 |
| S1-wms-combined_40115 | 17.83 | d__Bacteria;p__Firmicutes;c__Clostridia;o__Oscillospirales;f__Oscillospiraceae;g__ER4 | 97 | blast;97.90 iPHoP-RF;71.70 |
| S1-wms-combined_41369 | 37.96 | d__Bacteria;p__Firmicutes;c__Clostridia;o__Oscillospirales;f__Oscillospiraceae;g__ER4 | 94.6 | blast;96.20 iPHoP-RF;60.60 |
| S1-wms-combined_52708 | 55.79 | d__Bacteria;p__Firmicutes;c__Clostridia;o__Oscillospirales;f__Ruminococcaceae;g__Faecalibacterium | 96.3 | blast;97.10 iPHoP-RF;70.00 RaFAH;54.80 |
| S1-wms-combined_57370 | 50.73 | d__Bacteria;p__Firmicutes;c__Clostridia;o__Oscillospirales;f__Oscillospiraceae;g__CAG-83 | 91.7 | blast;89.00 RaFAH;72.80 |
| S1-wms-combined_68998 | 25.66 | d__Bacteria;p__Firmicutes;c__Clostridia;o__Oscillospirales;f__Ruminococcaceae;g__Faecalibacterium | 99.9 | CRISPR;98.60 blast;92.00 iPHoP-RF;91.40 RaFAH;91.30 |
| S1-wms-combined_72167 | 16.24 | d__Bacteria;p__Firmicutes;c__Clostridia;o__Oscillospirales;f__Oscillospiraceae;g__CAG-83 | 98.5 | CRISPR;98.90 blast;95.00 iPHoP-RF;90.80 |
| S1-wms-combined_85225 | 64.47 | d__Bacteria;p__Actinobacteriota;c__Actinomycetia;o__Actinomycetales;f__Bifidobacteriaceae;g__Bifidobacterium | 99.6 | CRISPR;99.60 iPHoP-RF;97.10 blast;96.90 |
| S1-wms-combined_92523 | 13.27 | d__Bacteria;p__Proteobacteria;c__Alphaproteobacteria;o__RF32;f__CAG-239;g__51-20 | 95 | blast;96.50 iPHoP-RF;71.30 |
| S1-wms-combined_94988 | 21.16 | d__Bacteria;p__Firmicutes;c__Clostridia;o__Oscillospirales;f__Oscillospiraceae;g__CAG-83 | 95.6 | blast;96.90 iPHoP-RF;94.10 |
| S1-wms_102342 | 14.71 | d__Bacteria;p__Firmicutes;c__Clostridia;o__Oscillospirales;f__Oscillospiraceae;g__Dysosmobacter | 95 | blast;96.50 iPHoP-RF;60.30 |
| S1-wms_102342 | 14.71 | d__Bacteria;p__Firmicutes;c__Clostridia;o__Oscillospirales;f__Oscillospiraceae;g__Intestinimonas | 94.9 | blast;96.40 iPHoP-RF;57.90 |
| S1-wms_130214 | 33.32 | d__Bacteria;p__Firmicutes;c__Clostridia;o__Oscillospirales;f__Oscillospiraceae;g__CAG-170 | 90.6 | blast;93.00 iPHoP-RF;90.50 CRISPR;82.90 |
| S1-wms_153615 | | d__Bacteria;p__Bacteroidota;c__Bacteroidia;o__Bacteroidales;f__Barnesiellaceae;g__Barnesiella | 93.8 | blast;95.60 iPHoP-RF;91.10 |
| S1-wms_185046 | 36.29 | d__Bacteria;p__Firmicutes;c__Clostridia;o__Oscillospirales;f__Oscillospiraceae;g__CAG-170 | 91.1 | blast;93.50 iPHoP-RF;82.20 |
| S1-wms_208875 | 95.4 | d__Bacteria;p__Bacteroidota;c__Bacteroidia;o__Bacteroidales;f__Bacteroidaceae;g__Bacteroides | 95.7 | blast;95.00 iPHoP-RF;94.40 RaFAH;70.30 |
| S1-wms_223620 | 12.07 | d__Bacteria;p__Firmicutes;c__Clostridia;o__Christensenellales;f__CAG-74;g__UBA11524 | 97.6 | blast;98.30 CRISPR;88.60 iPHoP-RF;86.00 |
| S1-wms_236105 | 71.3 | d__Bacteria;p__Firmicutes;c__Clostridia;o__Oscillospirales;f__Oscillospiraceae;g__ER4 | 92.1 | blast;94.30 iPHoP-RF;89.80 CRISPR;65.50 |
| S1-wms_244836 | 99.89 | d__Bacteria;p__Bacteroidota;c__Bacteroidia;o__Bacteroidales;f__Bacteroidaceae;g__Bacteroides | 100 | RaFAH;99.60 iPHoP-RF;96.40 blast;94.20 |
| S1-wms_244836 | 99.89 | d__Bacteria;p__Bacteroidota;c__Bacteroidia;o__Bacteroidales;f__Bacteroidaceae;g__Phocaeicola | 93.9 | iPHoP-RF;95.70 blast;92.10 |
| S1-wms_29288 | 41.26 | d__Bacteria;p__Firmicutes;c__Clostridia;o__Oscillospirales;f__Ruminococcaceae;g__Faecalibacterium | 91.6 | blast;93.90 CRISPR;78.00 iPHoP-RF;75.70 |

| | | | | |
|---|---|---|---|---|
| S1-wms_296850 | 38.32 | d__Bacteria;p__Firmicutes;c__Clostridia;o__Oscillospirales;f__Oscillospiraceae;g__ER4 | 93.1 | blast;95.10 iPHoP-RF;63.80 |
| S1-wms_394571 | 22.14 | d__Bacteria;p__Bacteroidota;c__Bacteroidia;o__Bacteroidales;f__Rikenellaceae;g__Alistipes | 95.4 | CRISPR;96.80 iPHoP-RF;71.90 |
| S1-wms_4277 | 93.43 | d__Bacteria;p__Firmicutes;c__Clostridia;o__Oscillospirales;f__Oscillospiraceae;g__Lawsonibacter | 91.4 | blast;93.70 |
| S11-wms-combined_101337 | 98.94 | d__Bacteria;p__Proteobacteria;c__Alphaproteobacteria;o__RF32;f__CAG-239;g__CAG-495 | 91.6 | blast;93.90 iPHoP-RF;86.30 CRISPR;51.40 |
| S11-wms-combined_112733 | 59.77 | d__Bacteria;p__Desulfobacterota;c__Desulfovibrionia;o__Desulfovibrionales;f__Desulfovibrionaceae;g__Desulfovibrio | 97.9 | blast;96.30 iPHoP-RF;88.90 RaFAH;79.80 CRISPR;67.90 |
| S11-wms-combined_116589 | 25.47 | d__Bacteria;p__Firmicutes;c__Clostridia;o__Oscillospirales;f__Oscillospiraceae;g__Dysosmobacter | 91.6 | blast;93.90 iPHoP-RF;88.90 |
| S11-wms-combined_119215 | 29.67 | d__Bacteria;p__Firmicutes;c__Clostridia;o__Oscillospirales;f__Oscillospiraceae;g__An92 | 94.9 | iPHoP-RF;96.40 blast;92.70 |
| S11-wms-combined_119215 | 29.67 | d__Bacteria;p__Firmicutes;c__Clostridia;o__Oscillospirales;f__Oscillospiraceae;g__Flavonifractor | 94.9 | iPHoP-RF;96.40 blast;92.00 CRISPR;55.60 |
| S11-wms-combined_119215 | 29.67 | d__Bacteria;p__Firmicutes;c__Clostridia;o__Oscillospirales;f__Oscillospiraceae;g__Pseudoflavonifractor | 91.4 | iPHoP-RF;93.70 |
| S11-wms-combined_120257 | 40.42 | d__Bacteria;p__Firmicutes;c__Clostridia;o__Oscillospirales;f__Oscillospiraceae;g__Flavonifractor | 91.1 | blast;93.50 iPHoP-RF;66.00 |
| S11-wms-combined_125176 | 94.89 | d__Bacteria;p__Bacteroidota;c__Bacteroidia;o__Bacteroidales;f__Rikenellaceae;g__Alistipes | 96.9 | CRISPR;97.80 iPHoP-RF;97.10 blast;96.90 |
| S11-wms-combined_140009 | 58.05 | d__Bacteria;p__Firmicutes;c__Clostridia;o__Oscillospirales;f__Ruminococcaceae;g__Faecalibacterium | 95.6 | blast;96.90 iPHoP-RF;71.90 |
| S11-wms-combined_154845 | 84.31 | d__Bacteria;p__Proteobacteria;c__Gammaproteobacteria;o__Enterobacterales;f__Enterobacteriaceae;g__Salmonella | 95.4 | blast;96.80 |
| S11-wms-combined_154845 | 84.31 | d__Bacteria;p__Proteobacteria;c__Gammaproteobacteria;o__Enterobacterales;f__Enterobacteriaceae;g__Escherichia | 95 | blast;96.50 |
| S11-wms-combined_154845 | 84.31 | d__Bacteria;p__Proteobacteria;c__Gammaproteobacteria;o__Enterobacterales;f__Enterobacteriaceae;g__Enterobacter | 93.8 | blast;95.60 iPHoP-RF;66.30 |
| S11-wms-combined_154845 | 84.31 | d__Bacteria;p__Proteobacteria;c__Gammaproteobacteria;o__Enterobacterales;f__Enterobacteriaceae;g__Citrobacter | 90 | blast;92.50 |
| S11-wms-combined_168048 | 95.9 | d__Bacteria;p__Firmicutes;c__Clostridia;o__Oscillospirales;f__Ruminococcaceae;g__Ruminiclostridium_E | 93.4 | blast;95.30 iPHoP-RF;66.80 |
| S11-wms-combined_168048 | 95.9 | d__Bacteria;p__Firmicutes;c__Clostridia;o__Acetivibrionales;f__Acetivibrionaceae;g__Ruminiclostridium | 93.1 | RaFAH;95.10 |
| S11-wms-combined_172889 | 100 | d__Bacteria;p__Bacteroidota;c__Bacteroidia;o__Bacteroidales;f__Rikenellaceae;g__Alistipes | 100 | RaFAH;99.10 blast;97.90 iPHoP-RF;94.70 |
| S11-wms-combined_172889 | 100 | d__Bacteria;p__Bacteroidota;c__Bacteroidia;o__Bacteroidales;f__Rikenellaceae;g__Alistipes_A | 98.8 | RaFAH;99.10 |
| S11-wms-combined_179605 | 37.98 | d__Bacteria;p__Firmicutes;c__Clostridia;o__Lachnospirales;f__Lachnospiraceae;g__UBA9502 | 91.2 | blast;93.60 iPHoP-RF;64.00 |
| S11-wms-combined_193148 | 97.83 | d__Bacteria;p__Proteobacteria;c__Alphaproteobacteria;o__RF32;f__CAG-239;g__CAG-267 | 94.9 | blast;96.40 iPHoP-RF;75.00 |
| S11-wms-combined_195425 | 89.45 | d__Bacteria;p__Bacteroidota;c__Bacteroidia;o__Bacteroidales;f__Bacteroidaceae;g__Bacteroides | 96.2 | blast;90.20 RaFAH;86.00 |
| S11-wms-combined_195425 | 89.45 | d__Bacteria;p__Bacteroidota;c__Bacteroidia;o__Bacteroidales;f__Tannerellaceae;g__Parabacteroides | 96.2 | blast;96.20 iPHoP-RF;78.00 RaFAH;64.50 CRISPR;58.70 |
| S11-wms-combined_200084 | 31.8 | d__Bacteria;p__Firmicutes;c__Clostridia;o__Lachnospirales;f__Lachnospiraceae;g__Blautia_A | 91.8 | blast;94.10 iPHoP-RF;53.60 |
| S11-wms-combined_204522 | 15.39 | d__Bacteria;p__Proteobacteria;c__Alphaproteobacteria;o__RF32;f__CAG-239;g__CAG-267 | 91.7 | blast;94.00 iPHoP-RF;72.60 |
| S11-wms-combined_215723 | 94.77 | d__Bacteria;p__Bacteroidota;c__Bacteroidia;o__Bacteroidales;f__Rikenellaceae;g__Alistipes | 99.4 | iPHoP-RF;97.10 blast;96.90 RaFAH;91.40 |
| S11-wms-combined_217790 | 99.99 | d__Bacteria;p__Bacteroidota;c__Bacteroidia;o__Bacteroidales;f__Bacteroidaceae;g__Bacteroides | 99.9 | RaFAH;99.80 |
| S11-wms-combined_217790 | 99.99 | d__Bacteria;p__Bacteroidota;c__Bacteroidia;o__Bacteroidales;f__Rikenellaceae;g__Alistipes | 97.3 | blast;98.10 iPHoP-RF;90.50 |
| S11-wms-combined_226165 | 80.02 | d__Bacteria;p__Bacteroidota;c__Bacteroidia;o__Bacteroidales;f__Rikenellaceae;g__Alistipes | 98.7 | iPHoP-RF;97.10 blast;93.80 RaFAH;82.70 |
| S11-wms-combined_236214 | 48.8 | d__Bacteria;p__Firmicutes;c__Clostridia;o__Oscillospirales;f__Ruminococcaceae;g__Faecalibacterium | 94.6 | blast;96.20 iPHoP-RF;57.60 |
| S11-wms-combined_264050 | 47.18 | d__Bacteria;p__Firmicutes;c__Clostridia;o__Oscillospirales;f__Ruminococcaceae;g__Gemmiger | 97.5 | blast;98.20 iPHoP-RF;93.40 CRISPR;91.60 |
| S11-wms-combined_267993 | 26.2 | d__Bacteria;p__Firmicutes;c__Clostridia;o__Christensenellales;f__CAG-74;g__Firm-11 | 91.5 | blast;93.80 iPHoP-RF;75.20 |
| S11-wms-combined_268967 | 35.57 | d__Bacteria;p__Desulfobacterota;c__Desulfovibrionia;o__Desulfovibrionales;f__Desulfovibrionaceae;g__Desulfovibrio | 93.8 | blast;95.10 iPHoP-RF;81.90 RaFAH;54.80 CRISPR;51.90 |
| S11-wms-combined_282227 | 60.6 | d__Bacteria;p__Firmicutes;c__Clostridia;o__Lachnospirales;f__Lachnospiraceae;g__Mediterraneibacter | 91.2 | blast;93.60 iPHoP-RF;73.00 |
| S11-wms-combined_283220 | 78.49 | d__Bacteria;p__Firmicutes;c__Clostridia;o__Oscillospirales;f__Oscillospiraceae;g__Dysosmobacter | 91.4 | blast;93.70 iPHoP-RF;83.60 |
| S11-wms-combined_299954 | 23.91 | d__Bacteria;p__Firmicutes;c__Clostridia;o__Oscillospirales;f__Oscillospiraceae;g__Intestinimonas | 90.2 | blast;92.70 iPHoP-RF;84.20 |
| S11-wms-combined_321938 | 99.59 | d__Bacteria;p__Bacteroidota;c__Bacteroidia;o__Bacteroidales;f__Bacteroidaceae;g__Bacteroides | 100 | RaFAH;98.70 iPHoP-RF;94.10 blast;80.70 |
| S11-wms-combined_329493 | 9.74 | d__Bacteria;p__Firmicutes;c__Clostridia;o__Oscillospirales;f__Oscillospiraceae;g__Faecousia | 96.7 | blast;97.70 iPHoP-RF;80.00 |
| S11-wms-combined_329493 | 9.74 | d__Bacteria;p__Firmicutes;c__Clostridia;o__Oscillospirales;f__Oscillospiraceae;g__CAG-110 | 93.6 | blast;95.50 iPHoP-RF;74.80 |
| S11-wms-combined_3507 | 24.17 | d__Bacteria;p__Firmicutes;c__Clostridia;o__Oscillospirales;f__Oscillospiraceae;g__CAG-83 | 98.1 | CRISPR;98.60 blast;96.90 iPHoP-RF;92.80 |
| S11-wms-combined_41367 | 40.35 | d__Bacteria;p__Bacteroidota;c__Bacteroidia;o__Bacteroidales;f__Bacteroidaceae;g__Bacteroides | 98.5 | RaFAH;97.90 iPHoP-RF;74.10 |
| S11-wms-combined_46816 | 20.69 | d__Bacteria;p__Firmicutes;c__Clostridia;o__Oscillospirales;f__Ruminococcaceae;g__Gemmiger | 95.4 | blast;96.80 iPHoP-RF;91.40 CRISPR;69.40 |
| S11-wms-combined_48246 | 99.82 | d__Bacteria;p__Bacteroidota;c__Bacteroidia;o__Bacteroidales;f__Rikenellaceae;g__Alistipes | 100 | blast;98.50 RaFAH;96.00 iPHoP-RF;95.40 |
| S11-wms-combined_48246 | 99.82 | d__Bacteria;p__Bacteroidota;c__Bacteroidia;o__Bacteroidales;f__Rikenellaceae;g__Alistipes_A | 94.3 | RaFAH;96.00 |
| S11-wms-combined_49632 | 14.66 | d__Bacteria;p__Firmicutes;c__Clostridia;o__Oscillospirales;f__Oscillospiraceae;g__Dysosmobacter | 91.5 | blast;93.80 iPHoP-RF;84.80 |
| S11-wms-combined_63818 | 50.37 | d__Bacteria;p__Bacteroidota;c__Bacteroidia;o__Bacteroidales;f__Rikenellaceae;g__Alistipes | 98.4 | iPHoP-RF;97.10 blast;96.90 RaFAH;78.80 |
| S11-wms-combined_64392 | 100 | d__Bacteria;p__Bacteroidota;c__Bacteroidia;o__Bacteroidales;f__Rikenellaceae;g__Alistipes | 99.6 | iPHoP-RF;97.10 blast;96.80 RaFAH;92.70 |
| S11-wms-combined_64392 | 100 | d__Bacteria;p__Bacteroidota;c__Bacteroidia;o__Bacteroidales;f__Rikenellaceae;g__Alistipes_A | 90.2 | RaFAH;92.70 |
| S11-wms-combined_73204 | 100 | d__Bacteria;p__Bacteroidota;c__Bacteroidia;o__Bacteroidales;f__Bacteroidaceae;g__Bacteroides | 99.4 | RaFAH;99.30 iPHoP-RF;66.50 |
| S11-wms-combined_83571 | 99.96 | d__Bacteria;p__Bacteroidota;c__Bacteroidia;o__Bacteroidales;f__Rikenellaceae;g__Alistipes | 100 | RaFAH;98.50 blast;98.40 iPHoP-RF;96.70 |
| S11-wms-combined_83571 | 99.96 | d__Bacteria;p__Bacteroidota;c__Bacteroidia;o__Bacteroidales;f__Rikenellaceae;g__Alistipes_A | 97.9 | RaFAH;98.50 |
| S11-wms-combined_84326 | 16.31 | d__Bacteria;p__Desulfobacterota;c__Desulfovibrionia;o__Desulfovibrionales;f__Desulfovibrionaceae;g__Desulfovibrio | 94.2 | blast;95.90 iPHoP-RF;89.50 CRISPR;83.30 |
| S11-wms-combined_92114 | 19.84 | d__Bacteria;p__Bacteroidota;c__Bacteroidia;o__Bacteroidales;f__Rikenellaceae;g__Alistipes | 97.2 | CRISPR;98.00 iPHoP-RF;97.10 blast;96.90 |
| S11-wms_14390 | 96.43 | d__Bacteria;p__Firmicutes;c__Clostridia;o__Oscillospirales;f__Oscillospiraceae;g__CAG-83 | 97.3 | RaFAH;98.10 |
| S11-wms_14390 | 96.43 | d__Bacteria;p__Firmicutes;c__Clostridia;o__Oscillospirales;f__Oscillospiraceae;g__Dysosmobacter | 97.3 | RaFAH;98.10 |
| S11-wms_14390 | 96.43 | d__Bacteria;p__Firmicutes;c__Clostridia;o__Oscillospirales;f__Oscillospiraceae;g__Flavonifractor | 93.5 | blast;95.40 iPHoP-RF;70.90 |
| S11-wms_3116 | 99.5 | d__Bacteria;p__Firmicutes;c__Clostridia;o__Oscillospirales;f__Ruminococcaceae;g__Faecalibacterium | 100 | RaFAH;99.60 blast;92.10 |
| S11-wms_320555 | 99.4 | d__Bacteria;p__Bacteroidota;c__Bacteroidia;o__Bacteroidales;f__Bacteroidaceae;g__Bacteroides | 99.7 | RaFAH;95.40 blast;92.70 iPHoP-RF;74.60 |
| S11-wms_32691 | 12 | d__Bacteria;p__Firmicutes;c__Clostridia;o__Oscillospirales;f__Oscillospiraceae;g__Pseudoflavonifractor | 94.9 | blast;96.40 iPHoP-RF;66.80 |
| S11-wms_330479 | 20.29 | d__Bacteria;p__Bacteroidota;c__Bacteroidia;o__Bacteroidales;f__Rikenellaceae;g__Alistipes | 98.5 | CRISPR;98.90 iPHoP-RF;78.50 |
| S12-wms-combined_111524 | 100 | d__Bacteria;p__Bacteroidota;c__Bacteroidia;o__Bacteroidales;f__Bacteroidaceae;g__Bacteroides | 99.6 | RaFAH;99.60 |
| S12-wms-combined_111524 | 100 | d__Bacteria;p__Bacteroidota;c__Bacteroidia;o__Bacteroidales;f__Rikenellaceae;g__Alistipes | 95.6 | blast;96.90 iPHoP-RF;96.70 CRISPR;80.20 |
| S12-wms-combined_117447 | | d__Bacteria;p__Firmicutes;c__Clostridia;o__Christensenellales;f__CAG-917;g__ | 91 | blast;93.40 iPHoP-RF;67.30 |
| S12-wms-combined_117628 | 32.91 | d__Bacteria;p__Firmicutes;c__Clostridia;o__Oscillospirales;f__Acutalibacteraceae;g__CAG-217 | 96.2 | blast;97.30 iPHoP-RF;73.20 |
| S12-wms-combined_120133 | 82.27 | d__Bacteria;p__Firmicutes;c__Clostridia;o__Lachnospirales;f__Lachnospiraceae;g__Lachnospira | 99.1 | CRISPR;97.80 blast;97.00 iPHoP-RF;96.40 RaFAH;84.70 |
| S12-wms-combined_129554 | 47.38 | d__Bacteria;p__Firmicutes;c__Clostridia;o__Oscillospirales;f__Oscillospiraceae;g__F23-B02 | 95 | blast;96.50 iPHoP-RF;75.90 |
| S12-wms-combined_129799 | 52.43 | d__Bacteria;p__Firmicutes;c__Clostridia;o__Oscillospirales;f__Oscillospiraceae;g__ER4 | 95.6 | blast;96.90 iPHoP-RF;63.00 |
| S12-wms-combined_147628 | 13.34 | d__Bacteria;p__Firmicutes;c__Clostridia;o__Christensenellales;f__CAG-138;g__SFEL01 | 96 | CRISPR;97.20 blast;93.30 iPHoP-RF;69.80 |
| S12-wms-combined_148741 | 20.82 | d__Bacteria;p__Firmicutes;c__Clostridia;o__Oscillospirales;f__Oscillospiraceae;g__CAG-83 | 93.9 | blast;95.70 iPHoP-RF;94.70 CRISPR;93.00 |
| S12-wms-combined_151843 | 24.49 | d__Bacteria;p__Firmicutes;c__Clostridia;o__Christensenellales;f__CAG-74;g__UBA11524 | 95.6 | blast;96.90 iPHoP-RF;92.80 |
| S12-wms-combined_151843 | 24.49 | d__Bacteria;p__Firmicutes;c__Clostridia;o__Christensenellales;f__CAG-74;g__UBA11524 | 95.6 | blast;96.90 iPHoP-RF;90.50 CRISPR;77.70 |
| S12-wms-combined_157167 | 8.42 | d__Bacteria;p__Firmicutes;c__Clostridia;o__Oscillospirales;f__Oscillospiraceae;g__CAG-83 | 92.5 | blast;94.60 iPHoP-RF;70.90 |
| S12-wms-combined_164656 | 3.93 | d__Bacteria;p__Firmicutes;c__Clostridia;o__Christensenellales;f__CAG-917;g__CAG-475 | 94.7 | blast;96.30 iPHoP-RF;68.60 |
| S12-wms-combined_168259 | 10.85 | d__Bacteria;p__Verrucomicrobiota;c__Verrucomicrobiae;o__Verrucomicrobiales;f__Akkermansiaceae;g__Akkermansia | 100 | RaFAH;98.70 blast;96.90 CRISPR;95.20 iPHoP-RF;87.60 |
| S12-wms-combined_184338 | 20.69 | d__Bacteria;p__Firmicutes;c__Clostridia;o__Oscillospirales;f__Oscillospiraceae;g__Dysosmobacter | 91.6 | blast;93.90 iPHoP-RF;83.90 |
| S12-wms-combined_189323 | 72.16 | d__Bacteria;p__Bacteroidota;c__Bacteroidia;o__Bacteroidales;f__Rikenellaceae;g__Alistipes | 99.4 | blast;97.60 iPHoP-RF;96.70 RaFAH;89.00 CRISPR;53.90 |

| | | | | |
|---|---|---|---|---|
| S12-wms-combined_190114 | 93.65 | d__Bacteria;p__Bacteroidota;c__Bacteroidia;o__Bacteroidales;f__Bacteroidaceae;g__Bacteroides | 100 | RaFAH;99.60 blast;96.70 iPHoP-RF;95.10 |
| S12-wms-combined_201125 | 83.81 | d__Bacteria;p__Bacteroidota;c__Bacteroidia;o__Bacteroidales;f__Bacteroidaceae;g__Bacteroides | 99.4 | iPHoP-RF;96.40 RaFAH;93.60 blast;77.30 |
| S12-wms-combined_201125 | 83.81 | d__Bacteria;p__Bacteroidota;c__Bacteroidia;o__Bacteroidales;f__Bacteroidaceae;g__Phocaeicola | 95.2 | blast;96.60 iPHoP-RF;80.60 |
| S12-wms-combined_210942 | 9.09 | d__Bacteria;p__Firmicutes;c__Clostridia;o__Oscillospirales;f__Oscillospiraceae;g__ER4 | 96.3 | blast;97.40 |
| S12-wms-combined_223043 | 99.11 | d__Bacteria;p__Firmicutes;c__Clostridia;o__Lachnospirales;f__Lachnospiraceae;g__Enterocloster | 91.5 | blast;93.80 iPHoP-RF;64.30 |
| S12-wms-combined_23517 | 28.19 | d__Bacteria;p__Firmicutes;c__Clostridia;o__Oscillospirales;f__Oscillospiraceae;g__CAG-83 | 98.5 | blast;98.90 CRISPR;98.40 iPHoP-RF;93.10 |
| S12-wms-combined_2450 | 57.23 | d__Bacteria;p__Bacteroidota;c__Bacteroidia;o__Bacteroidales;f__Rikenellaceae;g__Tidjanibacter | 90.2 | blast;92.70 iPHoP-RF;50.70 |
| S12-wms-combined_250551 | 98.55 | d__Bacteria;p__Bacteroidota;c__Bacteroidia;o__Bacteroidales;f__Bacteroidaceae;g__Bacteroides | 100 | RaFAH;99.40 blast;92.10 iPHoP-RF;83.90 |
| S12-wms-combined_250551 | 98.55 | d__Bacteria;p__Bacteroidota;c__Bacteroidia;o__Bacteroidales;f__Bacteroidaceae;g__Phocaeicola | 92.5 | blast;94.60 iPHoP-RF;75.00 |
| S12-wms-combined_25459 | 9.92 | d__Bacteria;p__Bacteroidota;c__Bacteroidia;o__Bacteroidales;f__Rikenellaceae;g__Alistipes | 95.6 | blast;96.90 iPHoP-RF;96.70 |
| S12-wms-combined_256299 | 44.45 | d__Bacteria;p__Bacteroidota;c__Bacteroidia;o__Bacteroidales;f__Muribaculaceae;g__UBA7173 | 90.1 | blast;92.60 iPHoP-RF;75.70 |
| S12-wms-combined_259783 | 81.56 | d__Bacteria;p__Firmicutes;c__Clostridia;o__Lachnospirales;f__Lachnospiraceae;g__Roseburia | 96.7 | CRISPR;97.70 blast;92.20 iPHoP-RF;70.60 |
| S12-wms-combined_259783 | 81.56 | d__Bacteria;p__Firmicutes;c__Clostridia;o__Lachnospirales;f__Lachnospiraceae;g__RUG115 | 96.6 | CRISPR;97.60 iPHoP-RF;50.40 |
| S12-wms-combined_259783 | 81.56 | d__Bacteria;p__Firmicutes;c__Clostridia;o__Lachnospirales;f__Lachnospiraceae;g__UBA11774 | 96.3 | CRISPR;97.40 iPHoP-RF;61.70 |
| S12-wms-combined_259783 | 81.56 | d__Bacteria;p__Firmicutes;c__Clostridia;o__Lachnospirales;f__Lachnospiraceae;g__Lachnospira | 95 | blast;96.50 iPHoP-RF;52.00 |
| S12-wms-combined_260036 | 44.16 | d__Bacteria;p__Bacteroidota;c__Bacteroidia;o__Bacteroidales;f__Barnesiellaceae;g__Barnesiella | 94.6 | blast;96.20 iPHoP-RF;94.10 CRISPR;67.50 |
| S12-wms-combined_263377 | 78.23 | d__Bacteria;p__Firmicutes;c__Clostridia;o__Oscillospirales;f__Acutalibacteraceae;g__UBA1417 | 91.7 | blast;94.00 iPHoP-RF;69.30 |
| S12-wms-combined_264077 | 19.02 | d__Bacteria;p__Firmicutes;c__Clostridia;o__Oscillospirales;f__Ruminococcaceae;g__Faecalibacterium | 90.4 | blast;92.90 iPHoP-RF;86.30 |
| S12-wms-combined_264425 | 99.56 | d__Bacteria;p__Firmicutes;c__Clostridia;o__Oscillospirales;f__Acutalibacteraceae;g__UBA1417 | 91.8 | blast;94.10 iPHoP-RF;57.30 |
| S12-wms-combined_264845 | 6.58 | d__Bacteria;p__Firmicutes;c__Clostridia;o__Oscillospirales;f__Oscillospiraceae;g__CAG-110 | 95 | blast;96.50 iPHoP-RF;90.10 |
| S12-wms-combined_270839 | 20.16 | d__Bacteria;p__Firmicutes;c__Clostridia;o__Oscillospirales;f__Oscillospiraceae;g__CAG-83 | 92.5 | blast;94.60 iPHoP-RF;79.80 |
| S12-wms-combined_282594 | 18.25 | d__Bacteria;p__Verrucomicrobiota;c__Verrucomicrobiae;o__Verrucomicrobiales;f__Akkermansiaceae;g__Akkermansia | 100 | CRISPR;98.70 RaFAH;98.50 blast;96.90 iPHoP-RF;78.50 |
| S12-wms-combined_293354 | 10.3 | d__Bacteria;p__Firmicutes;c__Clostridia;o__Christensenellales;f__CAG-74;g__UBA11524 | 95.6 | blast;96.90 iPHoP-RF;92.80 |
| S12-wms-combined_293354 | 10.3 | d__Bacteria;p__Firmicutes;c__Clostridia;o__Christensenellales;f__CAG-74;g__UBA11524 | 95.6 | blast;96.90 CRISPR;96.60 iPHoP-RF;93.10 |
| S12-wms-combined_297545 | 99.69 | d__Bacteria;p__Bacteroidota;c__Bacteroidia;o__Bacteroidales;f__Bacteroidaceae;g__Bacteroides | 99.9 | RaFAH;98.50 blast;90.90 iPHoP-RF;65.60 |
| S12-wms-combined_299293 | 62.93 | d__Bacteria;p__Firmicutes;c__Clostridia;o__Christensenellales;f__CAG-74;g__SFFH01 | 90.4 | blast;92.90 iPHoP-RF;75.40 |
| S12-wms-combined_302876 | 24.68 | d__Bacteria;p__Firmicutes;c__Clostridia;o__Lachnospirales;f__Lachnospiraceae;g__Lachnospira | 93.9 | blast;95.70 iPHoP-RF;73.50 |
| S12-wms-combined_302876 | 24.68 | d__Bacteria;p__Firmicutes;c__Clostridia;o__Lachnospirales;f__Lachnospiraceae;g__RUG115 | 93.9 | blast;95.70 iPHoP-RF;70.00 |
| S12-wms-combined_313953 | 97.5 | d__Bacteria;p__Proteobacteria;c__Alphaproteobacteria;o__RF32;f__CAG-239;g__CAG-495 | 90.9 | blast;93.30 iPHoP-RF;78.50 |
| S12-wms-combined_31761 | 54.85 | d__Bacteria;p__Firmicutes;c__Clostridia;o__Lachnospirales;f__Lachnospiraceae;g__COE1 | 93.6 | blast;95.50 iPHoP-RF;70.20 |
| S12-wms-combined_320262 | 15.45 | d__Bacteria;p__Bacteroidota;c__Bacteroidia;o__Bacteroidales;f__Rikenellaceae;g__Alistipes | 97.8 | blast;98.40 iPHoP-RF;97.10 |
| S12-wms-combined_320727 | 99.96 | d__Bacteria;p__Bacteroidota;c__Bacteroidia;o__Bacteroidales;f__Bacteroidaceae;g__Bacteroides | 93.8 | RaFAH;93.60 blast;61.60 iPHoP-RF;55.70 |
| S12-wms-combined_323943 | 18.94 | d__Bacteria;p__Firmicutes;c__Clostridia;o__Oscillospirales;f__Ruminococcaceae;g__Gemmiger | 94.6 | CRISPR;96.20 blast;93.90 iPHoP-RF;91.80 |
| S12-wms-combined_328554 | 31.03 | d__Bacteria;p__Bacteroidota;c__Bacteroidia;o__Bacteroidales;f__Rikenellaceae;g__Alistipes | 96.3 | iPHoP-RF;96.70 blast;95.80 RaFAH;60.10 |
| S12-wms-combined_329242 | 5.39 | d__Bacteria;p__Firmicutes;c__Clostridia;o__Oscillospirales;f__Oscillospiraceae;g__Dysosmobacter | 91.1 | blast;93.50 iPHoP-RF;56.00 |
| S12-wms-combined_334806 | 38.66 | d__Bacteria;p__Firmicutes;c__Clostridia;o__Oscillospirales;f__Oscillospiraceae;g__CAG-83 | 91.6 | blast;93.90 iPHoP-RF;61.90 |
| S12-wms-combined_345948 | 59.64 | d__Bacteria;p__Firmicutes;c__Clostridia;o__Oscillospirales;f__Ruminococcaceae;g__Faecalibacterium | 98.8 | blast;96.90 iPHoP-RF;92.10 RaFAH;84.90 CRISPR;63.30 |
| S12-wms-combined_351847 | 11.93 | d__Bacteria;p__Bacteroidota;c__Bacteroidia;o__Bacteroidales;f__Barnesiellaceae;g__Barnesiella | 95 | blast;96.50 iPHoP-RF;88.20 CRISPR;72.90 |
| S12-wms-combined_358797 | 99.42 | d__Bacteria;p__Bacteroidota;c__Bacteroidia;o__Bacteroidales;f__Rikenellaceae;g__Alistipes | 100 | blast;98.40 iPHoP-RF;96.10 RaFAH;95.40 |
| S12-wms-combined_358797 | 99.42 | d__Bacteria;p__Bacteroidota;c__Bacteroidia;o__Bacteroidales;f__Rikenellaceae;g__Alistipes_A | 93.5 | RaFAH;95.40 |
| S12-wms-combined_368190 | 100 | d__Bacteria;p__Bacteroidota;c__Bacteroidia;o__Bacteroidales;f__Bacteroidaceae;g__Bacteroides | 100 | RaFAH;98.90 iPHoP-RF;97.10 blast;91.30 |
| S12-wms-combined_368190 | 100 | d__Bacteria;p__Bacteroidota;c__Bacteroidia;o__Bacteroidales;f__Bacteroidaceae;g__Phocaeicola | 93.9 | blast;95.70 iPHoP-RF;95.70 CRISPR;79.60 |
| S12-wms-combined_368562 | 98.38 | d__Bacteria;p__Bacteroidota;c__Bacteroidia;o__Bacteroidales;f__Bacteroidaceae;g__Bacteroides | 100 | RaFAH;99.70 blast;93.90 iPHoP-RF;92.80 |
| S12-wms-combined_371541 | 14.41 | d__Bacteria;p__Bacteroidota;c__Bacteroidia;o__Bacteroidales;f__UBA11471;g__UBA11471 | 91.6 | blast;93.90 iPHoP-RF;90.10 |
| S12-wms-combined_371541 | 14.41 | d__Bacteria;p__Bacteroidota;c__Bacteroidia;o__Bacteroidales;f__Bacteroidaceae;g__Bacteroides | 90.7 | RaFAH;93.10 |
| S12-wms-combined_375343 | 20.72 | d__Bacteria;p__Bacteroidota;c__Bacteroidia;o__Bacteroidales;f__UBA11471;g__UBA11471 | 91.6 | blast;93.90 iPHoP-RF;88.50 |
| S12-wms-combined_393703 | 8.02 | d__Bacteria;p__Firmicutes;c__Clostridia;o__Oscillospirales;f__Oscillospiraceae;g__ER4 | 93.5 | blast;95.40 iPHoP-RF;50.20 |
| S12-wms-combined_48648 | 6.97 | d__Bacteria;p__Firmicutes;c__Clostridia;o__Oscillospirales;f__Oscillospiraceae;g__ER4 | 90.9 | blast;93.30 iPHoP-RF;72.40 |
| S12-wms-combined_52101 | 35.58 | d__Bacteria;p__Firmicutes;c__Clostridia;o__Oscillospirales;f__Ruminococcaceae;g__UBA3818 | 90.3 | blast;92.80 iPHoP-RF;62.20 |
| S12-wms-combined_63649 | 5.54 | d__Bacteria;p__Firmicutes;c__Clostridia;o__Oscillospirales;f__Oscillospiraceae;g__CAG-83 | 91.7 | blast;94.00 iPHoP-RF;90.10 |
| S12-wms-combined_71235 | 58.55 | d__Bacteria;p__Firmicutes;c__Clostridia;o__Oscillospirales;f__Ruminococcaceae;g__Faecalibacterium | 95.7 | blast;96.80 iPHoP-RF;84.20 RaFAH;53.60 |
| S12-wms-combined_75733 | | d__Bacteria;p__Bacteroidota;c__Bacteroidia;o__Bacteroidales;f__Barnesiellaceae;g__Barnesiella | 94.6 | blast;96.20 iPHoP-RF;69.80 |
| S12-wms-combined_91231 | 99.33 | d__Bacteria;p__Bacteroidota;c__Bacteroidia;o__Bacteroidales;f__Bacteroidaceae;g__Bacteroides | 100 | RaFAH;99.80 iPHoP-RF;96.10 blast;94.00 |
| S12-wms-combined_91231 | 99.33 | d__Bacteria;p__Bacteroidota;c__Bacteroidia;o__Bacteroidales;f__Bacteroidaceae;g__Phocaeicola | 91.6 | blast;93.90 iPHoP-RF;82.20 |
| S12-wms_12658 | 51.35 | d__Bacteria;p__Bacteroidota;c__Bacteroidia;o__Bacteroidales;f__Bacteroidaceae;g__Prevotellamassilia | 97.9 | CRISPR;98.50 blast;60.80 |
| S12-wms_12658 | 51.35 | d__Bacteria;p__Bacteroidota;c__Bacteroidia;o__Bacteroidales;f__Bacteroidaceae;g__Bacteroides | 92.7 | blast;90.60 RaFAH;72.60 |
| S12-wms_184276 | 36.83 | d__Bacteria;p__Actinobacteriota;c__Coriobacteriia;o__Coriobacteriales;f__Eggerthellaceae;g__Adlercreutzia | 93.1 | blast;95.10 iPHoP-RF;66.80 |
| S12-wms_184276 | 36.83 | d__Bacteria;p__Actinobacteriota;c__Coriobacteriia;o__Coriobacteriales;f__Coriobacteriaceae;g__Collinsella | 92.6 | CRISPR;94.70 |
| S12-wms_193565 | 96.64 | d__Bacteria;p__Firmicutes;c__Clostridia;o__Oscillospirales;f__Ruminococcaceae;g__Faecalibacterium | 90.2 | RaFAH;92.70 |
| S12-wms_223235 | | d__Bacteria;p__Firmicutes;c__Clostridia;o__Oscillospirales;f__Ruminococcaceae;g__Ruminococcus_D | 95.3 | blast;96.70 iPHoP-RF;74.80 CRISPR;67.70 |
| S12-wms_235125 | 20.48 | d__Bacteria;p__Firmicutes;c__Clostridia;o__Oscillospirales;f__Oscillospiraceae;g__ER4 | 90.8 | blast;93.20 iPHoP-RF;64.80 |
| S12-wms_336780 | 4.76 | d__Bacteria;p__Firmicutes;c__Clostridia;o__Oscillospirales;f__Ruminococcaceae;g__UMGS363 | 95.3 | blast;96.70 iPHoP-RF;93.10 |
| S12-wms_373775 | 11.82 | d__Bacteria;p__Firmicutes;c__Clostridia;o__Oscillospirales;f__Oscillospiraceae;g__CAG-83 | 91.5 | blast;93.80 iPHoP-RF;82.80 |
| S12-wms_386692 | 94.2 | d__Bacteria;p__Bacteroidota;c__Bacteroidia;o__Bacteroidales;f__Rikenellaceae;g__Alistipes | 96.2 | CRISPR;97.30 iPHoP-RF;97.10 blast;96.60 |
| S12-wms_386692 | 94.2 | d__Bacteria;p__Bacteroidota;c__Bacteroidia;o__Bacteroidales;f__Bacteroidaceae;g__Bacteroides | 91.6 | RaFAH;93.90 |
| S12-wms_401372 | 65.9 | d__Bacteria;p__Patescibacteria;c__Paceibacteria;o__Paceibacterales;f__UBA5633;g__MWCK01 | 91.1 | blast;93.50 |
| S17-wms-combined_106940 | 77.34 | d__Bacteria;p__Bacteroidota;c__Bacteroidia;o__Bacteroidales;f__Bacteroidaceae;g__Bacteroides | 99.6 | iPHoP-RF;97.10 blast;96.90 RaFAH;92.40 |
| S17-wms-combined_106940 | 77.34 | d__Bacteria;p__Bacteroidota;c__Bacteroidia;o__Bacteroidales;f__Bacteroidaceae;g__Phocaeicola | 94.9 | iPHoP-RF;96.40 blast;95.70 |
| S17-wms-combined_136622 | 24.07 | d__Bacteria;p__Firmicutes;c__Clostridia;o__Oscillospirales;f__Oscillospiraceae;g__Evtepia | 90.7 | blast;93.10 iPHoP-RF;81.60 |
| S17-wms-combined_146050 | 6.87 | d__Bacteria;p__Bacteroidota;c__Bacteroidia;o__Bacteroidales;f__Bacteroidaceae;g__Bacteroides | 100 | RaFAH;98.70 blast;96.80 iPHoP-RF;92.80 |
| S17-wms-combined_146850 | 3.97 | d__Bacteria;p__Firmicutes;c__Clostridia;o__Oscillospirales;f__Oscillospiraceae;g__Flavonifractor | 91.2 | blast;93.60 iPHoP-RF;74.30 |
| S17-wms-combined_157468 | 3.55 | d__Bacteria;p__Firmicutes;c__Clostridia;o__Oscillospirales;f__Oscillospiraceae;g__Flavonifractor | 93.4 | blast;95.30 iPHoP-RF;74.80 |
| S17-wms-combined_158354 | 99.65 | d__Bacteria;p__Bacteroidota;c__Bacteroidia;o__Bacteroidales;f__Bacteroidaceae;g__Bacteroides | 100 | RaFAH;99.30 iPHoP-RF;97.10 blast;92.60 |
| S17-wms-combined_158354 | 99.65 | d__Bacteria;p__Bacteroidota;c__Bacteroidia;o__Bacteroidales;f__Bacteroidaceae;g__Phocaeicola | 95.9 | iPHoP-RF;97.10 blast;96.20 CRISPR;86.70 |
| S17-wms-combined_162786 | 7.07 | d__Bacteria;p__Firmicutes;c__Clostridia;o__Oscillospirales;f__Oscillospiraceae;g__Marseille-P3106 | 94.7 | blast;96.30 iPHoP-RF;71.30 |
| S17-wms-combined_16309 | 27.66 | d__Bacteria;p__Bacteroidota;c__Bacteroidia;o__Bacteroidales;f__Bacteroidaceae;g__Bacteroides | 100 | RaFAH;99.10 iPHoP-RF;97.10 blast;90.90 |
| S17-wms-combined_16309 | 27.66 | d__Bacteria;p__Bacteroidota;c__Bacteroidia;o__Bacteroidales;f__Bacteroidaceae;g__Phocaeicola | 92.2 | iPHoP-RF;94.40 |

| | | | | |
|---|---|---|---|---|
| S17-wms-combined_171305 | 56.56 | d__Bacteria;p__Bacteroidota;c__Bacteroidia;o__Bacteroidales;f__Tannerellaceae;g__Parabacteroides | 98.7 | CRISPR;99.00 blast;98.50 iPHoP-RF;96.40 |
| S17-wms-combined_175924 | 99.75 | d__Bacteria;p__Firmicutes;c__Clostridia;o__Oscillospirales;f__Oscillospiraceae;g__Flavonifractor | 92.1 | blast;94.30 iPHoP-RF;78.00 |
| S17-wms-combined_179771 | 49.28 | d__Bacteria;p__Bacteroidota;c__Bacteroidia;o__Bacteroidales;f__Bacteroidaceae;g__Bacteroides | 97.9 | blast;98.30 iPHoP-RF;96.10 CRISPR;90.30 RaFAH;56.00 |
| S17-wms-combined_179771 | 49.28 | d__Bacteria;p__Bacteroidota;c__Bacteroidia;o__Bacteroidales;f__Bacteroidaceae;g__Phocaeicola | 90.3 | iPHoP-RF;92.80 |
| S17-wms-combined_185239 | 7.9 | d__Bacteria;p__Firmicutes;c__Clostridia;o__Oscillospirales;f__Oscillospiraceae;g__Flavonifractor | 91.4 | blast;93.70 iPHoP-RF;83.60 |
| S17-wms-combined_185239 | 7.9 | d__Bacteria;p__Firmicutes;c__Clostridia;o__Oscillospirales;f__Oscillospiraceae;g__Pseudoflavonifractor | 91.4 | blast;93.70 iPHoP-RF;52.20 |
| S17-wms-combined_186314 | 29.67 | d__Bacteria;p__Actinobacteriota;c__Actinomycetia;o__Actinomycetales;f__Bifidobacteriaceae;g__Bifidobacterium | 99.6 | CRISPR;99.60 iPHoP-RF;97.10 blast;96.90 |
| S17-wms-combined_193958 | 96.31 | d__Bacteria;p__Firmicutes;c__Clostridia;o__Oscillospirales;f__Ruminococcaceae;g__Faecalibacterium | 94.2 | blast;95.90 iPHoP-RF;53.80 |
| S17-wms-combined_199746 | 69.95 | d__Bacteria;p__Bacteroidota;c__Bacteroidia;o__Bacteroidales;f__Rikenellaceae;g__Alistipes | 97.6 | CRISPR;98.30 iPHoP-RF;97.10 blast;96.90 |
| S17-wms-combined_199746 | 69.95 | d__Bacteria;p__Bacteroidota;c__Bacteroidia;o__Bacteroidales;f__Bacteroidaceae;g__Bacteroides | 90.8 | RaFAH;93.20 |
| S17-wms-combined_223748 | 4.98 | d__Bacteria;p__Firmicutes;c__Clostridia;o__Lachnospirales;f__Lachnospiraceae;g__Enterocloster | 91.2 | blast;93.60 iPHoP-RF;75.20 |
| S17-wms-combined_225356 | 10.23 | d__Bacteria;p__Actinobacteriota;c__Coriobacteriia;o__Coriobacteriales;f__Eggerthellaceae;g__Eggerthella | 96.3 | CRISPR;97.40 blast;92.10 iPHoP-RF;87.00 |
| S17-wms-combined_225356 | 10.23 | d__Bacteria;p__Actinobacteriota;c__Coriobacteriia;o__Coriobacteriales;f__Eggerthellaceae;g__Arabia | 90.2 | CRISPR;92.70 iPHoP-RF;50.70 |
| S17-wms-combined_233761 | 88 | d__Bacteria;p__Bacteroidota;c__Bacteroidia;o__Bacteroidales;f__Bacteroidaceae;g__Bacteroides | 100 | RaFAH;99.00 blast;98.60 iPHoP-RF;96.40 CRISPR;80.00 |
| S17-wms-combined_247494 | 24.14 | d__Bacteria;p__Firmicutes;c__Clostridia;o__Oscillospirales;f__Oscillospiraceae;g__Flavonifractor | 91.5 | blast;93.80 iPHoP-RF;91.40 |
| S17-wms-combined_247494 | 24.14 | d__Bacteria;p__Firmicutes;c__Clostridia;o__Oscillospirales;f__Oscillospiraceae;g__Pseudoflavonifractor | 90.7 | iPHoP-RF;93.10 blast;89.90 |
| S17-wms-combined_253499 | 25.09 | d__Bacteria;p__Bacteroidota;c__Bacteroidia;o__Bacteroidales;f__Rikenellaceae;g__Alistipes | 97.2 | CRISPR;98.00 blast;95.10 iPHoP-RF;90.80 |
| S17-wms-combined_258087 | 97.18 | d__Bacteria;p__Firmicutes;c__Clostridia;o__Clostridiales;f__Clostridiaceae;g__Clostridium | 98.2 | RaFAH;98.70 |
| S17-wms-combined_258087 | 97.18 | d__Bacteria;p__Firmicutes;c__Clostridia;o__Clostridiales;f__Clostridiaceae;g__Clostridium_F | 98.2 | RaFAH;98.70 |
| S17-wms-combined_258087 | 97.18 | d__Bacteria;p__Firmicutes;c__Clostridia;o__Clostridiales;f__Clostridiaceae;g__Clostridium_P | 98.2 | RaFAH;98.70 |
| S17-wms-combined_259811 | 66.5 | d__Bacteria;p__Firmicutes;c__Clostridia;o__Oscillospirales;f__Oscillospiraceae;g__Lawsonibacter | 90.9 | blast;93.30 |
| S17-wms-combined_260343 | 38.59 | d__Bacteria;p__Firmicutes;c__Clostridia;o__Oscillospirales;f__Oscillospiraceae;g__Flavonifractor | 90.8 | blast;93.20 |
| S17-wms-combined_39687 | 83.01 | d__Bacteria;p__Firmicutes;c__Clostridia;o__Oscillospirales;f__Oscillospiraceae;g__Flavonifractor | 95.2 | blast;96.60 iPHoP-RF;89.20 |
| S17-wms-combined_56006 | 98.39 | d__Bacteria;p__Firmicutes;c__Clostridia;o__Oscillospirales;f__Ruminococcaceae;g__Faecalibacterium | 100 | RaFAH;98.90 blast;96.90 iPHoP-RF;87.00 CRISPR;67.80 |
| S17-wms-combined_56061 | 66.23 | d__Bacteria;p__Firmicutes;c__Clostridia;o__Oscillospirales;f__Ruminococcaceae;g__Faecalibacterium | 98.5 | blast;98.90 |
| S17-wms-combined_66123 | 97.65 | d__Bacteria;p__Firmicutes;c__Clostridia;o__Oscillospirales;f__Oscillospiraceae;g__Flavonifractor | 90.6 | blast;91.80 iPHoP-RF;85.40 CRISPR;73.10 RaFAH;57.20 |
| S17-wms-combined_70983 | 7.24 | d__Bacteria;p__Bacteroidota;c__Bacteroidia;o__Bacteroidales;f__Bacteroidaceae;g__Bacteroides | 95.9 | iPHoP-RF;97.10 blast;85.40 |
| S17-wms-combined_70983 | 7.24 | d__Bacteria;p__Bacteroidota;c__Bacteroidia;o__Bacteroidales;f__Bacteroidaceae;g__Phocaeicola | 94.5 | iPHoP-RF;96.10 blast;92.70 |
| S17-wms-combined_72837 | 25.38 | d__Bacteria;p__Firmicutes;c__Clostridia;o__Oscillospirales;f__Ruminococcaceae;g__Gemmiger | 95.2 | blast;96.60 iPHoP-RF;74.80 |
| S17-wms-combined_75892 | 53.86 | d__Bacteria;p__Firmicutes;c__Clostridia;o__Oscillospirales;f__Oscillospiraceae;g__HGM12998 | 91.4 | blast;93.70 iPHoP-RF;77.50 |
| S17-wms-combined_79709 | 19.94 | d__Bacteria;p__Firmicutes;c__Clostridia;o__Oscillospirales;f__Oscillospiraceae;g__An92 | 91.4 | blast;93.70 iPHoP-RF;92.10 |
| S17-wms-combined_90378 | 72.41 | d__Bacteria;p__Firmicutes;c__Clostridia;o__Oscillospirales;f__Ruminococcaceae;g__Faecalibacterium | 98.7 | blast;96.90 RaFAH;83.30 iPHoP-RF;76.80 CRISPR;69.70 |
| S17-wms-combined_95360 | 17.32 | d__Bacteria;p__Firmicutes;c__Negativicutes;o__Veillonellales;f__Dialisteraceae;g__Dialister | 97.2 | CRISPR;98.00 blast;96.90 iPHoP-RF;93.70 |
| S17-wms_124308 | 51.69 | d__Bacteria;p__Bacteroidota;c__Bacteroidia;o__Bacteroidales;f__Bacteroidaceae;g__Bacteroides | 99.1 | iPHoP-RF;96.40 RaFAH;90.70 |
| S17-wms_254872 | 5.1 | d__Bacteria;p__Bacteroidota;c__Bacteroidia;o__Bacteroidales;f__Bacteroidaceae;g__Bacteroides_G | 95.3 | iPHoP-RF;96.70 |
| S17-wms_254872 | 5.1 | d__Bacteria;p__Bacteroidota;c__Bacteroidia;o__Bacteroidales;f__Bacteroidaceae;g__Bacteroides | 94.9 | iPHoP-RF;96.40 CRISPR;89.30 |
| S17-wms_254872 | 5.1 | d__Bacteria;p__Proteobacteria;c__Gammaproteobacteria;o__Pseudomonadales;f__Saccharospirillaceae;g__Saccharospirillum | 90 | blast;92.50 |
| S2-wms-combined_101826 | 26.15 | d__Bacteria;p__Firmicutes;c__Clostridia;o__Oscillospirales;f__Ruminococcaceae;g__Ruminococcus_D | 98.8 | CRISPR;99.10 blast;96.90 iPHoP-RF;92.80 |
| S2-wms-combined_113619 | 97.74 | d__Bacteria;p__Bacteroidota;c__Bacteroidia;o__Bacteroidales;f__Rikenellaceae;g__Alistipes | 99 | blast;96.60 RaFAH;87.50 CRISPR;82.60 iPHoP-RF;77.80 |
| S2-wms-combined_116221 | 80.02 | d__Bacteria;p__Firmicutes;c__Clostridia;o__Lachnospirales;f__Anaerotignaceae;g__Anaerotignum | 95.6 | blast;96.90 iPHoP-RF;61.10 |
| S2-wms-combined_122457 | 52.93 | d__Bacteria;p__Firmicutes;c__Clostridia;o__Oscillospirales;f__Oscillospiraceae;g__Intestinimonas | 91.6 | blast;93.90 iPHoP-RF;56.50 |
| S2-wms-combined_126134 | 14.26 | d__Bacteria;p__Firmicutes;c__Clostridia;o__Oscillospirales;f__Oscillospiraceae;g__Lawsonibacter | 90.1 | blast;92.60 iPHoP-RF;75.70 |
| S2-wms-combined_126440 | 24 | d__Bacteria;p__Firmicutes;c__Clostridia;o__Lachnospirales;f__Lachnospiraceae;g__Acetatifactor | 92.7 | blast;94.80 iPHoP-RF;71.30 |
| S2-wms-combined_127080 | 8.57 | d__Bacteria;p__Firmicutes;c__Clostridia;o__Oscillospirales;f__Ruminococcaceae;g__Faecalibacterium | 98.8 | CRISPR;99.10 blast;93.50 iPHoP-RF;89.50 |
| S2-wms-combined_13246 | 32.69 | d__Bacteria;p__Firmicutes;c__Clostridia;o__Oscillospirales;f__Ruminococcaceae;g__Faecalibacterium | 99.6 | blast;96.90 RaFAH;93.60 iPHoP-RF;92.80 CRISPR;87.50 |
| S2-wms-combined_132966 | | d__Bacteria;p__Bacteroidota;c__Bacteroidia;o__Bacteroidales;f__Bacteroidaceae;g__Bacteroides | 93.1 | iPHoP-RF;95.10 |
| S2-wms-combined_135014 | 100 | d__Bacteria;p__Firmicutes;c__Negativicutes;o__Veillonellales;f__Dialisteraceae;g__Dialister | 100 | RaFAH;97.60 blast;96.90 CRISPR;96.10 iPHoP-RF;92.40 |
| S2-wms-combined_135139 | 38.63 | d__Bacteria;p__Verrucomicrobiota;c__Verrucomicrobiae;o__Verrucomicrobiales;f__Akkermansiaceae;g__Akkermansia | 97.8 | CRISPR;98.40 blast;96.90 iPHoP-RF;88.20 |
| S2-wms-combined_135154 | 34.29 | d__Bacteria;p__Firmicutes;c__Negativicutes;o__Veillonellales;f__Dialisteraceae;g__Dialister | 95.9 | blast;97.10 iPHoP-RF;97.10 CRISPR;75.50 |
| S2-wms-combined_137119 | 36.79 | d__Bacteria;p__Firmicutes;c__Clostridia;o__Oscillospirales;f__Oscillospiraceae;g__An92 | 93.5 | blast;95.40 iPHoP-RF;76.60 |
| S2-wms-combined_140996 | 59.11 | d__Bacteria;p__Firmicutes;c__Clostridia;o__Oscillospirales;f__Oscillospiraceae;g__ER4 | 95.6 | blast;96.90 iPHoP-RF;70.20 |
| S2-wms-combined_143737 | 99.75 | d__Bacteria;p__Bacteroidota;c__Bacteroidia;o__Bacteroidales;f__Bacteroidaceae;g__Bacteroides | 100 | blast;97.80 RaFAH;97.50 iPHoP-RF;78.50 |
| S2-wms-combined_147065 | 99.85 | d__Bacteria;p__Bacteroidota;c__Bacteroidia;o__Bacteroidales;f__Bacteroidaceae;g__Bacteroides | 100 | RaFAH;97.00 blast;96.90 iPHoP-RF;95.10 CRISPR;58.20 |
| S2-wms-combined_157934 | 99.51 | d__Bacteria;p__Firmicutes;c__Clostridia;o__Lachnospirales;f__Lachnospiraceae;g__Roseburia | 98.8 | CRISPR;98.90 iPHoP-RF;92.80 blast;89.30 RaFAH;59.00 |
| S2-wms-combined_157934 | 99.51 | d__Bacteria;p__Firmicutes;c__Clostridia;o__Lachnospirales;f__Lachnospiraceae;g__Agathobacter | 98.7 | CRISPR;98.80 iPHoP-RF;96.70 blast;96.30 RaFAH;59.00 |
| S2-wms-combined_157934 | 99.51 | d__Bacteria;p__Firmicutes;c__Clostridia;o__Lachnospirales;f__Lachnospiraceae;g__UBA11774 | 97.9 | CRISPR;98.50 iPHoP-RF;68.20 |
| S2-wms-combined_158821 | 99.16 | d__Bacteria;p__Firmicutes;c__Clostridia;o__Lachnospirales;f__Lachnospiraceae;g__Enterocloster | 91.5 | blast;93.80 iPHoP-RF;63.50 |
| S2-wms-combined_159670 | 16.22 | d__Bacteria;p__Firmicutes;c__Clostridia;o__Oscillospirales;f__Oscillospiraceae;g__CAG-83 | 91.6 | blast;93.90 iPHoP-RF;88.20 |
| S2-wms-combined_165833 | 74.42 | d__Bacteria;p__Firmicutes;c__Clostridia;o__Lachnospirales;f__Lachnospiraceae;g__Roseburia | 97.9 | blast;96.20 iPHoP-RF;92.80 RaFAH;79.80 |
| S2-wms-combined_166665 | 48.97 | d__Bacteria;p__Firmicutes;c__Clostridia;o__Oscillospirales;f__Ruminococcaceae;g__Faecalibacterium | 95.4 | blast;96.80 iPHoP-RF;96.10 |
| S2-wms-combined_176376 | 94.42 | d__Bacteria;p__Firmicutes;c__Clostridia;o__Lachnospirales;f__Lachnospiraceae;g__Blautia_A | 96.7 | blast;93.20 CRISPR;89.80 iPHoP-RF;87.90 RaFAH;83.30 |
| S2-wms-combined_181849 | 40.03 | d__Bacteria;p__Firmicutes;c__Clostridia;o__Oscillospirales;f__Ruminococcaceae;g__Faecalibacterium | 95.6 | blast;96.90 CRISPR;95.80 iPHoP-RF;87.30 |
| S2-wms-combined_185494 | 25.15 | d__Bacteria;p__Firmicutes;c__Negativicutes;o__Veillonellales;f__Dialisteraceae;g__Dialister | 96.3 | CRISPR;97.40 blast;96.90 iPHoP-RF;90.10 |
| S2-wms-combined_191613 | 13.4 | d__Bacteria;p__Firmicutes;c__Clostridia;o__Lachnospirales;f__Anaerotignaceae;g__Anaerotignum | 95.2 | blast;96.60 iPHoP-RF;70.60 |
| S2-wms-combined_196783 | 32.37 | d__Bacteria;p__Firmicutes;c__Clostridia;o__Oscillospirales;f__Oscillospiraceae;g__Intestinimonas | 95.9 | CRISPR;97.10 blast;91.40 iPHoP-RF;85.10 |
| S2-wms-combined_196783 | 32.37 | d__Bacteria;p__Firmicutes;c__Clostridia;o__Oscillospirales;f__Oscillospiraceae;g__Flavonifractor | 93.5 | CRISPR;95.40 blast;91.00 iPHoP-RF;83.30 |
| S2-wms-combined_197577 | 42.77 | d__Bacteria;p__Firmicutes;c__Negativicutes;o__Selenomonadales;f__Selenomonadaceae;g__Megamonas | 91.1 | blast;93.50 iPHoP-RF;53.30 |
| S2-wms-combined_199384 | 17.07 | d__Bacteria;p__Firmicutes;c__Clostridia;o__Oscillospirales;f__Butyricicoccaceae;g__Agathobaculum | 91.4 | blast;93.70 iPHoP-RF;73.50 |
| S2-wms-combined_199384 | 17.07 | d__Bacteria;p__Firmicutes;c__Clostridia;o__Oscillospirales;f__Butyricicoccaceae;g__Butyricicoccus | 91.1 | blast;93.50 iPHoP-RF;67.70 |
| S2-wms-combined_200390 | 14.49 | d__Bacteria;p__Firmicutes;c__Clostridia;o__Lachnospirales;f__Lachnospiraceae;g__Eisenbergiella | 92 | CRISPR;94.20 iPHoP-RF;67.50 |
| S2-wms-combined_201205 | 82.22 | d__Bacteria;p__Verrucomicrobiota;c__Verrucomicrobiae;o__Verrucomicrobiales;f__Akkermansiaceae;g__Akkermansia | 100 | CRISPR;99.30 blast;96.80 RaFAH;96.70 iPHoP-RF;68.60 |
| S2-wms-combined_204225 | 6.16 | d__Bacteria;p__Firmicutes;c__Clostridia;o__Lachnospirales;f__Lachnospiraceae;g__Lachnoclostridium_A | 91.6 | blast;93.90 iPHoP-RF;79.30 |
| S2-wms-combined_207000 | 92.52 | d__Bacteria;p__Firmicutes;c__Clostridia;o__Lachnospirales;f__Lachnospiraceae;g__Agathobacter | 93.4 | blast;95.30 iPHoP-RF;93.70 |
| S2-wms-combined_207000 | 92.52 | d__Bacteria;p__Firmicutes;c__Clostridia;o__Lachnospirales;f__Lachnospiraceae;g__Roseburia | 90.7 | iPHoP-RF;93.10 blast;92.00 |
| S2-wms-combined_208559 | 10.84 | d__Bacteria;p__Bacteroidota;c__Bacteroidia;o__Bacteroidales;f__Rikenellaceae;g__Alistipes | 95.6 | blast;96.90 iPHoP-RF;96.70 |
| S2-wms-combined_210822 | 37.52 | d__Bacteria;p__Bacteroidota;c__Bacteroidia;o__Bacteroidales;f__Bacteroidaceae;g__Bacteroides | 91.8 | iPHoP-RF;89.50 blast;81.20 RaFAH;71.80 |
| S2-wms-combined_218002 | 99.73 | d__Bacteria;p__Firmicutes;c__Clostridia;o__Lachnospirales;f__Lachnospiraceae;g__Roseburia | 99.9 | blast;98.50 iPHoP-RF;92.40 RaFAH;92.30 |

| | | | | |
|---|---|---|---|---|
| S2-wms-combined_218002 | 99.73 | d__Bacteria;p__Firmicutes;c__Clostridia;o__Lachnospirales;f__Lachnospiraceae;g__RUG115 | 99.3 | RaFAH;92.30 blast;87.70 iPHoP-RF;83.10 |
| S2-wms-combined_218002 | 99.73 | d__Bacteria;p__Firmicutes;c__Clostridia;o__Lachnospirales;f__Lachnospiraceae;g__Agathobacter | 99 | RaFAH;92.30 blast;89.90 iPHoP-RF;73.00 |
| S2-wms-combined_222535 | 7.99 | d__Bacteria;p__Firmicutes;c__Clostridia;o__Lachnospirales;f__Lachnospiraceae;g__Agathobacter | 92.1 | blast;94.30 iPHoP-RF;92.10 |
| S2-wms-combined_222535 | 7.99 | d__Bacteria;p__Firmicutes;c__Clostridia;o__Lachnospirales;f__Lachnospiraceae;g__Roseburia | 90.7 | iPHoP-RF;93.10 blast;91.00 |
| S2-wms-combined_225908 | 87.71 | d__Bacteria;p__Firmicutes;c__Clostridia;o__Oscillospirales;f__Acutalibacteraceae;g__CAG-217 | 95.6 | blast;96.90 CRISPR;93.30 iPHoP-RF;79.00 |
| S2-wms-combined_229529 | 100 | d__Bacteria;p__Bacteroidota;c__Bacteroidia;o__Bacteroidales;f__Bacteroidaceae;g__Bacteroides | 99.9 | blast;96.90 iPHoP-RF;96.70 RaFAH;96.10 |
| S2-wms-combined_238929 | 17.21 | d__Bacteria;p__Firmicutes;c__Clostridia;o__Oscillospirales;f__Oscillospiraceae;g__CAG-170 | 91.4 | blast;93.70 iPHoP-RF;78.30 |
| S2-wms-combined_24549 | 17.49 | d__Bacteria;p__Firmicutes;c__Clostridia;o__Oscillospirales;f__Oscillospiraceae;g__Intestinimonas | 90.9 | blast;93.30 |
| S2-wms-combined_25727 | 21.03 | d__Bacteria;p__Firmicutes;c__Clostridia;o__Christensenellales;f__CAG-74;g__Ventricola | 94.7 | blast;96.30 iPHoP-RF;74.60 |
| S2-wms-combined_25727 | 21.03 | d__Bacteria;p__Firmicutes;c__Clostridia;o__Christensenellales;f__CAG-74;g__SFFH01 | 94.7 | blast;96.30 iPHoP-RF;71.90 |
| S2-wms-combined_30750 | 11.82 | d__Bacteria;p__Firmicutes;c__Clostridia;o__Oscillospirales;f__Ruminococcaceae;g__Ruminococcus_D | 95.6 | blast;96.90 iPHoP-RF;82.50 |
| S2-wms-combined_31084 | 22.49 | d__Bacteria;p__Firmicutes;c__Clostridia;o__Lachnospirales;f__Lachnospiraceae;g__Acetivibrio_A | 90 | blast;92.50 iPHoP-RF;69.10 |
| S2-wms-combined_48661 | 11.3 | d__Bacteria;p__Firmicutes;c__Clostridia;o__Lachnospirales;f__Lachnospiraceae;g__Anaerosacchariphilus | 90.8 | blast;93.20 iPHoP-RF;77.30 |
| S2-wms-combined_52568 | 12.66 | d__Bacteria;p__Firmicutes;c__Clostridia;o__Oscillospirales;f__Oscillospiraceae;g__F23-B02 | 92.3 | blast;94.50 iPHoP-RF;80.30 |
| S2-wms-combined_57293 | 78.69 | d__Bacteria;p__Firmicutes;c__Clostridia;o__Oscillospirales;f__Ruminococcaceae;g__Faecalibacterium | 95.7 | blast;96.90 iPHoP-RF;88.90 RaFAH;51.90 |
| S2-wms-combined_5791 | 89.76 | d__Bacteria;p__Firmicutes;c__Clostridia;o__Lachnospirales;f__Lachnospiraceae;g__Agathobacter | 92.1 | CRISPR;94.30 |
| S2-wms-combined_5791 | 89.76 | d__Bacteria;p__Firmicutes;c__Clostridia;o__Lachnospirales;f__Lachnospiraceae;g__Roseburia | 91 | CRISPR;93.40 |
| S2-wms-combined_5791 | 89.76 | d__Bacteria;p__Firmicutes;c__Clostridia;o__Lachnospirales;f__Lachnospiraceae;g__Eisenbergiella | 90.8 | blast;93.20 |
| S2-wms-combined_58121 | 21.08 | d__Bacteria;p__Bacteroidota;c__Bacteroidia;o__Bacteroidales;f__Bacteroidaceae;g__Bacteroides | 95.9 | iPHoP-RF;97.10 blast;91.00 |
| S2-wms-combined_58121 | 21.08 | d__Bacteria;p__Bacteroidota;c__Bacteroidia;o__Bacteroidales;f__Bacteroidaceae;g__Phocaeicola | 95.2 | blast;96.60 iPHoP-RF;81.90 |
| S2-wms-combined_58563 | 88.99 | d__Bacteria;p__Firmicutes;c__Clostridia;o__Clostridiales;f__Clostridiaceae;g__Clostridium | 95.4 | RaFAH;96.80 |
| S2-wms-combined_58563 | 88.99 | d__Bacteria;p__Firmicutes;c__Clostridia;o__Clostridiales;f__Clostridiaceae;g__Clostridium_F | 95.4 | RaFAH;96.80 |
| S2-wms-combined_58563 | 88.99 | d__Bacteria;p__Firmicutes;c__Clostridia;o__Clostridiales;f__Clostridiaceae;g__Clostridium_P | 95.4 | RaFAH;96.80 |
| S2-wms-combined_58563 | 88.99 | d__Bacteria;p__Firmicutes;c__Clostridia;o__Oscillospirales;f__Oscillospiraceae;g__F23-B02 | 94.6 | blast;96.20 iPHoP-RF;73.70 |
| S2-wms-combined_62839 | 32.39 | d__Bacteria;p__Firmicutes;c__Clostridia;o__Oscillospirales;f__Ruminococcaceae;g__Gemmiger | 97.6 | blast;98.30 iPHoP-RF;87.30 CRISPR;73.60 |
| S2-wms-combined_64692 | 24.76 | d__Bacteria;p__Firmicutes;c__Clostridia;o__Oscillospirales;f__Oscillospiraceae;g__ | 90.8 | blast;93.20 iPHoP-RF;85.40 |
| S2-wms-combined_64692 | 24.76 | d__Bacteria;p__Firmicutes;c__Clostridia;o__Oscillospirales;f__Oscillospiraceae;g__Flavonifractor | 90.4 | blast;92.90 iPHoP-RF;82.20 CRISPR;53.30 |
| S2-wms-combined_6605 | 7.53 | d__Bacteria;p__Firmicutes;c__Clostridia;o__Oscillospirales;f__Oscillospiraceae;g__Flavonifractor | 91.4 | blast;93.70 iPHoP-RF;82.50 |
| S2-wms-combined_72725 | 19.31 | d__Bacteria;p__Firmicutes;c__Clostridia;o__Oscillospirales;f__Oscillospiraceae;g__Intestinimonas | 91.4 | blast;93.70 iPHoP-RF;75.40 |
| S2-wms-combined_73327 | 3.55 | d__Bacteria;p__Firmicutes;c__Clostridia;o__Lachnospirales;f__Lachnospiraceae;g__Faecalimonas | 91.6 | blast;93.90 iPHoP-RF;73.50 |
| S2-wms-combined_74387 | 38.64 | d__Bacteria;p__Firmicutes;c__Clostridia;o__Lachnospirales;f__Lachnospiraceae;g__Lachnoclostridium_A | 94.3 | blast;96.00 iPHoP-RF;82.50 |
| S2-wms-combined_75144 | 9.77 | d__Bacteria;p__Bacteroidota;c__Bacteroidia;o__Bacteroidales;f__Tannerellaceae;g__Parabacteroides | 95.4 | blast;96.80 iPHoP-RF;95.10 |
| S2-wms-combined_7756 | 9.67 | d__Bacteria;p__Verrucomicrobiota;c__Verrucomicrobiae;o__Verrucomicrobiales;f__Akkermansiaceae;g__Akkermansia | 96.9 | blast;96.70 iPHoP-RF;91.10 CRISPR;86.00 RaFAH;66.20 |
| S2-wms-combined_84608 | 21.12 | d__Bacteria;p__Proteobacteria;c__Gammaproteobacteria;o__Burkholderiales;f__Burkholderiaceae;g__Sutterella | 95.6 | blast;96.90 iPHoP-RF;90.80 CRISPR;79.90 |
| S2-wms-combined_85381 | 45.07 | d__Bacteria;p__Firmicutes;c__Clostridia;o__Lachnospirales;f__Lachnospiraceae;g__CAG-81 | 93.8 | blast;95.60 iPHoP-RF;80.60 |
| S2-wms-combined_93845 | 99.44 | d__Bacteria;p__Firmicutes;c__Clostridia;o__Oscillospirales;f__Oscillospiraceae;g__ER4 | 98.7 | CRISPR;99.00 blast;91.30 iPHoP-RF;81.90 |
| S2-wms-combined_93845 | 99.44 | d__Bacteria;p__Firmicutes;c__Clostridia;o__Oscillospirales;f__Oscillospiraceae;g__F23-B02 | 95.4 | blast;96.80 |
| S2-wms-combined_93845 | 99.44 | d__Bacteria;p__Firmicutes;c__Clostridia;o__Oscillospirales;f__Oscillospiraceae;g__Flavonifractor | 90.8 | RaFAH;93.20 |
| S2-wms-combined_95987 | 3.24 | d__Bacteria;p__Firmicutes;c__Clostridia;o__Lachnospirales;f__Lachnospiraceae;g__Lachnoclostridium_A | 93.2 | blast;95.20 iPHoP-RF;84.50 |
| S2-wms_124416 | 99.32 | d__Bacteria;p__Firmicutes;c__Clostridia;o__Oscillospirales;f__Ruminococcaceae;g__Faecalibacterium | 100 | RaFAH;99.60 blast;96.10 iPHoP-RF;79.30 CRISPR;55.70 |
| S2-wms_128831 | 98.35 | d__Bacteria;p__Proteobacteria;c__Gammaproteobacteria;o__Enterobacterales;f__Enterobacteriaceae;g__Escherichia | 95.6 | RaFAH;96.90 |
| S2-wms_128831 | 98.35 | d__Bacteria;p__Patescibacteria;c__Paceibacteria;o__Paceibacterales;f__UBA5633;g__MWCK01 | 92.3 | blast;94.50 iPHoP-RF;66.30 |
| S2-wms_128962 | 46.73 | d__Bacteria;p__Bacteroidota;c__Bacteroidia;o__Bacteroidales;f__Bacteroidaceae;g__Prevotella | 91.8 | RaFAH;94.10 |
| S2-wms_130687 | 95.89 | d__Bacteria;p__Firmicutes;c__Clostridia;o__Oscillospirales;f__Acutalibacteraceae;g__Ruminococcus_E | 99.7 | CRISPR;98.90 blast;96.90 iPHoP-RF;92.40 RaFAH;85.50 |
| S2-wms_140933 | 43.87 | d__Bacteria;p__Firmicutes;c__Clostridia;o__Lachnospirales;f__Lachnospiraceae;g__Blautia_A | 93.8 | blast;90.60 RaFAH;76.60 iPHoP-RF;62.50 |
| S2-wms_156439 | 96.43 | d__Bacteria;p__Bacteroidota;c__Bacteroidia;o__Bacteroidales;f__Bacteroidaceae;g__Bacteroides | 100 | RaFAH;99.60 blast;96.80 iPHoP-RF;95.70 |
| S2-wms_161986 | 99.14 | d__Bacteria;p__Firmicutes;c__Clostridia;o__Lachnospirales;f__Lachnospiraceae;g__Lachnospira | 99.9 | CRISPR;98.80 RaFAH;92.10 blast;84.30 iPHoP-RF;67.70 |
| S2-wms_161986 | 99.14 | d__Bacteria;p__Firmicutes;c__Clostridia;o__Lachnospirales;f__Lachnospiraceae;g__Eubacterium_G | 91 | RaFAH;92.10 blast;58.10 |
| S2-wms_192667 | 13.43 | d__Bacteria;p__Firmicutes;c__Clostridia;o__Lachnospirales;f__Lachnospiraceae;g__Eubacterium_I | 91.6 | blast;93.90 iPHoP-RF;62.20 |
| S2-wms_246296 | 98.04 | d__Bacteria;p__Firmicutes;c__Clostridia;o__Lachnospirales;f__Lachnospiraceae;g__Roseburia | 100 | CRISPR;98.90 blast;94.40 RaFAH;93.70 iPHoP-RF;92.80 |
| S2-wms_246296 | 98.04 | d__Bacteria;p__Firmicutes;c__Clostridia;o__Lachnospirales;f__Lachnospiraceae;g__RUG115 | 99.9 | CRISPR;98.20 RaFAH;93.70 blast;90.80 iPHoP-RF;84.80 |
| S2-wms_246296 | 98.04 | d__Bacteria;p__Firmicutes;c__Clostridia;o__Lachnospirales;f__Lachnospiraceae;g__Agathobacter | 93.6 | RaFAH;93.70 iPHoP-RF;64.00 |
| S2-wms_6424 | 8.31 | d__Bacteria;p__Bacteroidota;c__Bacteroidia;o__Bacteroidales;f__Bacteroidaceae;g__Bacteroides | 93.6 | RaFAH;95.50 |
| S2-wms_79304 | 81.02 | d__Bacteria;p__Actinobacteriota;c__Coriobacteriia;o__Coriobacteriales;f__Eggerthellaceae;g__Eggerthella | 94.9 | CRISPR;96.40 RaFAH;50.20 |
| S3-wms-combined_135428 | 11.75 | d__Bacteria;p__Actinobacteriota;c__Actinomycetia;o__Actinomycetales;f__Bifidobacteriaceae;g__Bifidobacterium | 95.3 | blast;96.70 iPHoP-RF;96.40 CRISPR;94.80 |
| S3-wms-combined_45305 | 27.39 | d__Bacteria;p__Firmicutes;c__Clostridia;o__Oscillospirales;f__Ruminococcaceae;g__Gemmiger | 94.2 | blast;95.90 iPHoP-RF;76.60 CRISPR;55.10 |
| S3-wms-combined_73262 | 99.96 | d__Bacteria;p__Bacteroidota;c__Bacteroidia;o__Bacteroidales;f__Bacteroidaceae;g__Bacteroides | 100 | RaFAH;99.80 blast;93.60 iPHoP-RF;79.80 CRISPR;67.80 |
| S3-wms_22744 | 99.21 | d__Bacteria;p__Firmicutes;c__Clostridia;o__Oscillospirales;f__Ruminococcaceae;g__Faecalibacterium | 97.2 | CRISPR;98.00 blast;60.70 iPHoP-RF;52.00 |
| S3-wms_22744 | 99.21 | d__Bacteria;p__Firmicutes;c__Clostridia;o__Oscillospirales;f__Oscillospiraceae;g__CAG-83 | 91.8 | RaFAH;94.10 |
| S3-wms_22744 | 99.21 | d__Bacteria;p__Firmicutes;c__Clostridia;o__Oscillospirales;f__Oscillospiraceae;g__Dysosmobacter | 91.8 | RaFAH;94.10 |
| S3-wms_23958 | 21.78 | d__Bacteria;p__Firmicutes;c__Clostridia;o__Oscillospirales;f__Oscillospiraceae;g__CAG-170 | 94.2 | blast;95.90 iPHoP-RF;58.70 |
| S3-wms_94494 | 94.44 | d__Bacteria;p__Actinobacteriota;c__Actinomycetia;o__Actinomycetales;f__Bifidobacteriaceae;g__Bifidobacterium | 99.3 | blast;98.30 iPHoP-RF;97.10 CRISPR;83.00 RaFAH;81.50 |
| S4-wms-combined_10758 | 88.87 | d__Bacteria;p__Bacteroidota;c__Bacteroidia;o__Bacteroidales;f__Tannerellaceae;g__Parabacteroides | 100 | RaFAH;99.50 blast;89.70 iPHoP-RF;87.00 |
| S4-wms-combined_113721 | 6.49 | d__Bacteria;p__Firmicutes;c__Clostridia;o__Oscillospirales;f__Ruminococcaceae;g__Gemmiger | 95.6 | blast;96.90 iPHoP-RF;92.80 CRISPR;57.80 |
| S4-wms-combined_118531 | 66.42 | d__Bacteria;p__Firmicutes;c__Negativicutes;o__Acidaminococcales;f__Acidaminococcaceae;g__Phascolarctobacterium_A | 98.2 | CRISPR;98.70 blast;96.80 iPHoP-RF;92.40 |
| S4-wms-combined_118531 | 66.42 | d__Bacteria;p__Firmicutes;c__Negativicutes;o__Acidaminococcales;f__Acidaminococcaceae;g__Phascolarctobacterium | 90.7 | blast;93.10 iPHoP-RF;84.20 |
| S4-wms-combined_121978 | 87.01 | d__Bacteria;p__Firmicutes;c__Clostridia;o__Lachnospirales;f__Lachnospiraceae;g__Mediterraneibacter | 95 | blast;96.50 iPHoP-RF;79.50 |
| S4-wms-combined_14411 | 97.82 | d__Bacteria;p__Bacteroidota;c__Bacteroidia;o__Bacteroidales;f__Tannerellaceae;g__Marseille-P3160 | 98.4 | CRISPR;98.80 iPHoP-RF;68.90 |
| S4-wms-combined_14411 | 97.82 | d__Bacteria;p__Bacteroidota;c__Bacteroidia;o__Bacteroidales;f__Marinifilaceae;g__Odoribacter | 97.5 | RaFAH;98.20 |
| S4-wms-combined_17284 | 85.97 | d__Bacteria;p__Actinobacteriota;c__Actinomycetia;o__Actinomycetales;f__Bifidobacteriaceae;g__Bifidobacterium | 95.3 | iPHoP-RF;96.70 CRISPR;96.00 blast;95.40 |
| S4-wms-combined_22012 | 25.83 | d__Bacteria;p__Actinobacteriota;c__Coriobacteriia;o__Coriobacteriales;f__Coriobacteriaceae;g__Collinsella | 98.7 | CRISPR;99.00 blast;96.80 iPHoP-RF;89.20 |
| S4-wms-combined_24659 | 6.25 | d__Bacteria;p__Firmicutes;c__Clostridia;o__Lachnospirales;f__Lachnospiraceae;g__Roseburia | 92.6 | iPHoP-RF;94.70 |
| S4-wms-combined_41490 | 99.65 | d__Bacteria;p__Bacteroidota;c__Bacteroidia;o__Bacteroidales;f__Rikenellaceae;g__Alistipes | 95.6 | blast;96.90 iPHoP-RF;96.10 |
| S4-wms-combined_42647 | 63.73 | d__Bacteria;p__Firmicutes;c__Clostridia;o__Oscillospirales;f__Ruminococcaceae;g__Faecalibacterium | 95.7 | blast;97.00 iPHoP-RF;78.00 CRISPR;67.90 |
| S4-wms-combined_47565 | 81.36 | d__Bacteria;p__Bacteroidota;c__Bacteroidia;o__Bacteroidales;f__Bacteroidaceae;g__Prevotella | 96.2 | blast;97.30 iPHoP-RF;90.80 |
| S4-wms-combined_51079 | 88.31 | d__Bacteria;p__Actinobacteriota;c__Actinomycetia;o__Actinomycetales;f__Bifidobacteriaceae;g__Bifidobacterium | 99.9 | CRISPR;99.60 iPHoP-RF;97.10 blast;96.80 RaFAH;71.10 |
| S4-wms-combined_57123 | 13.17 | d__Bacteria;p__Bacteroidota;c__Bacteroidia;o__Bacteroidales;f__Bacteroidaceae;g__Prevotella | 95.6 | blast;96.90 iPHoP-RF;73.50 |

| | | | | |
|---|---|---|---|---|
| S4-wms-combined_57963 | 74.8 | d__Bacteria;p__Firmicutes;c__Negativicutes;o__Acidaminococcales;f__Acidaminococcaceae;g__Phascolarctobacterium_A | 95.6 | blast;96.90 iPHoP-RF;64.80 |
| S4-wms-combined_64632 | 43.29 | d__Bacteria;p__Firmicutes;c__Clostridia;o__Lachnospirales;f__Lachnospiraceae;g__Dorea_A | 97.6 | CRISPR;98.30 iPHoP-RF;95.40 blast;92.00 |
| S4-wms-combined_64632 | 43.29 | d__Bacteria;p__Firmicutes;c__Clostridia;o__Lachnospirales;f__Lachnospiraceae;g__Lachnoclostridium_B | 91.4 | iPHoP-RF;93.70 blast;82.20 |
| S4-wms-combined_83492 | 86.97 | d__Bacteria;p__Bacteroidota;c__Bacteroidia;o__Bacteroidales;f__Bacteroidaceae;g__Bacteroides | 95.6 | blast;89.90 iPHoP-RF;86.30 RaFAH;84.50 |
| S4-wms-combined_83492 | 86.97 | d__Bacteria;p__Bacteroidota;c__Bacteroidia;o__Bacteroidales;f__Tannerellaceae;g__Parabacteroides | 93.9 | blast;95.70 iPHoP-RF;85.70 |
| S4-wms-combined_83492 | 86.97 | d__Bacteria;p__Bacteroidota;c__Bacteroidia;o__Bacteroidales;f__Bacteroidaceae;g__Phocaeicola | 91.5 | blast;93.80 iPHoP-RF;74.60 |
| S4-wms_107182 | 7.59 | d__Bacteria;p__Actinobacteriota;c__Coriobacteriia;o__Coriobacteriales;f__Eggerthellaceae;g__CAG-1427 | 96.9 | blast;97.80 |
| S4-wms_141607 | 37.81 | d__Bacteria;p__Bacteroidota;c__Bacteroidia;o__Bacteroidales;f__Bacteroidaceae;g__Prevotella | 98.4 | CRISPR;98.80 blast;96.40 iPHoP-RF;79.00 |
| S4-wms_161443 | 5.84 | d__Bacteria;p__Actinobacteriota;c__Actinomycetia;o__Actinomycetales;f__Bifidobacteriaceae;g__Bifidobacterium | 99.6 | CRISPR;99.60 iPHoP-RF;78.00 |
| S4-wms_45128 | 28.59 | d__Bacteria;p__Bacteroidota;c__Bacteroidia;o__Bacteroidales;f__Bacteroidaceae;g__Prevotella | 94.7 | blast;96.30 CRISPR;93.50 iPHoP-RF;91.40 |
| S4-wms_50475 | 70.49 | d__Bacteria;p__Bacteroidota;c__Bacteroidia;o__Bacteroidales;f__Bacteroidaceae;g__Prevotella | 97.3 | blast;96.90 iPHoP-RF;94.40 RaFAH;69.20 CRISPR;50.60 |
| S4-wms_62635 | 93.28 | d__Bacteria;p__Firmicutes;c__Clostridia;o__Lachnospirales;f__Lachnospiraceae;g__Blautia_A | 100 | CRISPR;98.90 RaFAH;95.20 iPHoP-RF;93.70 blast;90.40 |
| S4-wms_62635 | 93.28 | d__Bacteria;p__Firmicutes;c__Clostridia;o__Lachnospirales;f__Lachnospiraceae;g__Blautia | 93.2 | RaFAH;95.20 |
| S4-wms_73990 | 76.93 | d__Bacteria;p__Bacteroidota;c__Bacteroidia;o__Bacteroidales;f__Tannerellaceae;g__Parabacteroides | 97.6 | blast;98.30 iPHoP-RF;82.80 |
| S5-wms-combined_106597 | 25.93 | d__Bacteria;p__Campylobacterota;c__Campylobacteria;o__Campylobacterales;f__Campylobacteraceae;g__Campylobacter_D | 98.8 | RaFAH;99.10 |
| S5-wms-combined_106597 | 25.93 | d__Bacteria;p__Campylobacterota;c__Campylobacteria;o__Campylobacterales;f__Campylobacteraceae;g__Campylobacter_B | 91.6 | blast;93.90 |
| S5-wms-combined_108794 | 58.74 | d__Bacteria;p__Firmicutes;c__Clostridia;o__Oscillospirales;f__Acutalibacteraceae;g__Ruminococcus_E | 95.6 | blast;96.90 iPHoP-RF;89.50 |
| S5-wms-combined_117095 | 10.16 | d__Bacteria;p__Firmicutes;c__Clostridia;o__Oscillospirales;f__Acutalibacteraceae;g__UBA1417 | 91 | blast;93.40 iPHoP-RF;64.50 |
| S5-wms-combined_117213 | 97.16 | d__Bacteria;p__Actinobacteriota;c__Actinomycetia;o__Actinomycetales;f__Bifidobacteriaceae;g__Bifidobacterium | 98.8 | CRISPR;97.70 iPHoP-RF;97.10 blast;96.50 RaFAH;80.80 |
| S5-wms-combined_134381 | 97.01 | d__Bacteria;p__Firmicutes;c__Clostridia;o__Oscillospirales;f__Oscillospiraceae;g__CAG-83 | 98.4 | RaFAH;98.30 blast;64.70 |
| S5-wms-combined_134381 | 97.01 | d__Bacteria;p__Firmicutes;c__Clostridia;o__Oscillospirales;f__Oscillospiraceae;g__Dysosmobacter | 97.6 | RaFAH;98.30 |
| S5-wms-combined_134381 | 97.01 | d__Bacteria;p__Firmicutes;c__Clostridia;o__Oscillospirales;f__Oscillospiraceae;g__CAG-110 | 90.3 | blast;92.80 |
| S5-wms-combined_145067 | 82.3 | d__Bacteria;p__Firmicutes;c__Clostridia;o__Oscillospirales;f__Oscillospiraceae;g__Flavonifractor | 95.6 | blast;96.90 iPHoP-RF;95.70 CRISPR;53.80 |
| S5-wms-combined_145067 | 82.3 | d__Bacteria;p__Firmicutes;c__Clostridia;o__Oscillospirales;f__Oscillospiraceae;g__Intestinimonas | 93.1 | iPHoP-RF;95.10 |
| S5-wms-combined_145067 | 82.3 | d__Bacteria;p__Firmicutes;c__Clostridia;o__Oscillospirales;f__Oscillospiraceae;g__An92 | 92.2 | iPHoP-RF;94.40 |
| S5-wms-combined_145067 | 82.3 | d__Bacteria;p__Firmicutes;c__Clostridia;o__Oscillospirales;f__Oscillospiraceae;g__Pseudoflavonifractor | 92.2 | iPHoP-RF;94.40 |
| S5-wms-combined_156329 | 75.2 | d__Bacteria;p__Firmicutes;c__Bacilli;o__Erysipelotrichales;f__Erysipelotrichaceae;g__Clostridium_AQ | 95.9 | blast;97.10 CRISPR;90.20 iPHoP-RF;75.20 |
| S5-wms-combined_163165 | 3.66 | d__Bacteria;p__Firmicutes;c__Clostridia;o__Oscillospirales;f__Oscillospiraceae;g__Flavonifractor | 91.2 | blast;93.60 iPHoP-RF;59.50 |
| S5-wms-combined_178432 | 13.6 | d__Bacteria;p__Bacteroidota;c__Bacteroidia;o__Bacteroidales;f__Rikenellaceae;g__Alistipes | 95.9 | iPHoP-RF;97.10 blast;91.90 CRISPR;84.60 |
| S5-wms-combined_179241 | 13.11 | d__Bacteria;p__Firmicutes;c__Clostridia;o__Oscillospirales;f__Oscillospiraceae;g__CAG-83 | 91 | blast;93.40 iPHoP-RF;86.70 |
| S5-wms-combined_183113 | 11.29 | d__Bacteria;p__Bacteroidota;c__Bacteroidia;o__Bacteroidales;f__Rikenellaceae;g__Alistipes_A | 97 | blast;97.90 iPHoP-RF;94.70 |
| S5-wms-combined_18356 | 90.68 | d__Bacteria;p__Proteobacteria;c__Gammaproteobacteria;o__Enterobacterales;f__Enterobacteriaceae;g__Escherichia | 98.8 | iPHoP-RF;97.10 blast;96.40 RaFAH;85.20 CRISPR;70.30 |
| S5-wms-combined_18356 | 90.68 | d__Bacteria;p__Proteobacteria;c__Gammaproteobacteria;o__Enterobacterales;f__Enterobacteriaceae;g__Salmonella | 90.8 | blast;93.20 |
| S5-wms-combined_197186 | 3.6 | d__Bacteria;p__Firmicutes;c__Bacilli;o__Erysipelotrichales;f__Erysipelotrichaceae;g__Clostridium_AQ | 94.6 | blast;96.20 iPHoP-RF;73.70 |
| S5-wms-combined_199135 | 16.23 | d__Bacteria;p__Cyanobacteria;c__Vampirovibrionia;o__Gastranaerophilales;f__Gastranaerophilaceae;g__QAMI01 | 90.3 | blast;92.80 |
| S5-wms-combined_204782 | 6.93 | d__Bacteria;p__Bacteroidota;c__Bacteroidia;o__Bacteroidales;f__Rikenellaceae;g__Alistipes_A | 92.1 | blast;94.30 iPHoP-RF;69.10 |
| S5-wms-combined_205046 | 4.59 | d__Bacteria;p__Firmicutes;c__Clostridia;o__Oscillospirales;f__Oscillospiraceae;g__Dysosmobacter | 92.6 | iPHoP-RF;94.70 blast;93.60 |
| S5-wms-combined_207291 | 100 | d__Bacteria;p__Bacteroidota;c__Bacteroidia;o__Bacteroidales;f__Bacteroidaceae;g__Bacteroides | 100 | RaFAH;99.80 iPHoP-RF;95.70 blast;91.90 |
| S5-wms-combined_210959 | 81.3 | d__Bacteria;p__Bacteroidota;c__Bacteroidia;o__Bacteroidales;f__Rikenellaceae;g__Alistipes | 99.4 | blast;96.90 iPHoP-RF;96.70 RaFAH;91.40 CRISPR;57.80 |
| S5-wms-combined_26061 | 99.77 | d__Bacteria;p__Firmicutes;c__Clostridia;o__Lachnospirales;f__Lachnospiraceae;g__Blautia_A | 96.7 | blast;92.60 RaFAH;84.50 |
| S5-wms-combined_3201 | 23.69 | d__Bacteria;p__Proteobacteria;c__Gammaproteobacteria;o__Burkholderiales;f__Burkholderiaceae;g__Oxalobacter | 90.9 | blast;93.30 iPHoP-RF;69.80 |
| S5-wms-combined_39793 | | d__Bacteria;p__Actinobacteriota;c__Actinomycetia;o__Actinomycetales;f__Bifidobacteriaceae;g__Bifidobacterium | 91.6 | blast;93.90 iPHoP-RF;93.40 CRISPR;76.80 |
| S5-wms-combined_52741 | 50.99 | d__Bacteria;p__Verrucomicrobiota;c__Verrucomicrobiae;o__Verrucomicrobiales;f__Akkermansiaceae;g__Akkermansia | 100 | RaFAH;99.50 CRISPR;98.80 blast;96.90 iPHoP-RF;81.60 |
| S5-wms-combined_59064 | 95 | d__Bacteria;p__Bacteroidota;c__Bacteroidia;o__Bacteroidales;f__Bacteroidaceae;g__Bacteroides | 100 | RaFAH;97.60 blast;96.90 iPHoP-RF;96.10 |
| S5-wms-combined_66017 | 9.66 | d__Bacteria;p__Firmicutes;c__Clostridia;o__Christensenellales;f__Christensenellaceae;g__Christensenella | 90.7 | blast;93.10 iPHoP-RF;82.20 |
| S5-wms-combined_66662 | 100 | d__Bacteria;p__Firmicutes;c__Clostridia;o__Oscillospirales;f__Ruminococcaceae;g__Ruthenibacterium | 97.2 | blast;98.00 iPHoP-RF;88.50 |
| S5-wms-combined_67650 | 98.81 | d__Bacteria;p__Bacteroidota;c__Bacteroidia;o__Bacteroidales;f__Rikenellaceae;g__Alistipes | 100 | RaFAH;97.10 blast;96.90 iPHoP-RF;93.70 |
| S5-wms-combined_67650 | 98.81 | d__Bacteria;p__Bacteroidota;c__Bacteroidia;o__Bacteroidales;f__Rikenellaceae;g__Alistipes_A | 95.9 | RaFAH;97.10 |
| S5-wms-combined_72410 | 60.27 | d__Bacteria;p__Firmicutes;c__Clostridia;o__Oscillospirales;f__Ruminococcaceae;g__Ruthenibacterium | 90.8 | blast;93.20 iPHoP-RF;72.60 |
| S5-wms-combined_7292 | 22.4 | d__Bacteria;p__Firmicutes;c__Clostridia;o__Oscillospirales;f__Acutalibacteraceae;g__Ruminococcus_E | 98.5 | CRISPR;98.90 blast;96.60 iPHoP-RF;89.80 |
| S5-wms-combined_73301 | 96.75 | d__Bacteria;p__Firmicutes;c__Clostridia;o__Lachnospirales;f__Lachnospiraceae;g__Dorea_A | 94.6 | blast;96.20 iPHoP-RF;93.10 CRISPR;58.60 |
| S5-wms-combined_73301 | 96.75 | d__Bacteria;p__Firmicutes;c__Clostridia;o__Lachnospirales;f__Lachnospiraceae;g__Lachnoclostridium_B | 90.8 | blast;93.20 iPHoP-RF;77.80 |
| S5-wms-combined_83370 | 82.36 | d__Bacteria;p__Bacteroidota;c__Bacteroidia;o__Bacteroidales;f__Bacteroidaceae;g__Bacteroides | 99.4 | blast;96.90 iPHoP-RF;96.70 RaFAH;91.90 |
| S5-wms-combined_83412 | 25.58 | d__Bacteria;p__Firmicutes;c__Clostridia;o__Eubacteriales;f__Eubacteriaceae;g__Eubacterium | 92.6 | blast;94.70 iPHoP-RF;86.70 CRISPR;77.00 |
| S5-wms-combined_90315 | 99.97 | d__Bacteria;p__Bacteroidota;c__Bacteroidia;o__Bacteroidales;f__Rikenellaceae;g__Alistipes | 98.8 | RaFAH;96.10 blast;88.90 |
| S5-wms-combined_90315 | 99.97 | d__Bacteria;p__Bacteroidota;c__Bacteroidia;o__Bacteroidales;f__Rikenellaceae;g__Alistipes_A | 94.5 | RaFAH;96.10 |
| S5-wms-combined_94482 | 15.25 | d__Bacteria;p__Bacteroidota;c__Bacteroidia;o__Bacteroidales;f__Tannerellaceae;g__Parabacteroides | 94.9 | iPHoP-RF;96.40 blast;91.70 |
| S5-wms-combined_94482 | 15.25 | d__Bacteria;p__Bacteroidota;c__Bacteroidia;o__Bacteroidales;f__Tannerellaceae;g__Macellibacteroides | 91.8 | iPHoP-RF;94.10 blast;83.60 |
| S5-wms-combined_98202 | 100 | d__Bacteria;p__Bacteroidota;c__Bacteroidia;o__Bacteroidales;f__Bacteroidaceae;g__Bacteroides | 99.4 | RaFAH;99.50 |
| S5-wms-combined_98202 | 100 | d__Bacteria;p__Bacteroidota;c__Bacteroidia;o__Bacteroidales;f__Rikenellaceae;g__Alistipes | 95.6 | blast;96.90 iPHoP-RF;77.50 |
| S5-wms_10783 | 3.75 | d__Bacteria;p__Firmicutes;c__Clostridia;o__Oscillospirales;f__Oscillospiraceae;g__Lawsonibacter | 90.8 | blast;93.20 |
| S5-wms_212596 | 76.97 | d__Bacteria;p__Firmicutes;c__Bacilli;o__Lactobacillales;f__Lactobacillaceae;g__Lacticaseibacillus | 100 | blast;98.70 RaFAH;95.70 iPHoP-RF;77.10 |
| S5-wms_212596 | 76.97 | d__Bacteria;p__Firmicutes;c__Bacilli;o__Lactobacillales;f__Lactobacillaceae;g__Lactobacillus | 93.9 | RaFAH;95.70 |
| S5-wms_212596 | 76.97 | d__Bacteria;p__Firmicutes;c__Bacilli;o__Lactobacillales;f__Lactobacillaceae;g__Ligilactobacillus | 93.9 | RaFAH;95.70 |
| S5-wms_212596 | 76.97 | d__Bacteria;p__Firmicutes;c__Bacilli;o__Lactobacillales;f__Lactobacillaceae;g__Limosilactobacillus | 93.9 | RaFAH;95.70 |
| S5-wms_49310 | 96.19 | d__Bacteria;p__Firmicutes;c__Clostridia;o__Oscillospirales;f__Acutalibacteraceae;g__Ruminococcus_E | 99.7 | blast;98.00 CRISPR;97.80 RaFAH;93.50 iPHoP-RF;88.20 |
| S5-wms_49310 | 96.19 | d__Bacteria;p__Firmicutes;c__Clostridia;o__Oscillospirales;f__Ruminococcaceae;g__Ruminococcus_D | 91.1 | RaFAH;93.50 |
| S5-wms_52566 | 8.51 | d__Bacteria;p__Firmicutes;c__Clostridia;o__Oscillospirales;f__Oscillospiraceae;g__Intestinimonas | 91.1 | blast;93.50 iPHoP-RF;52.80 |
| S5-wms_62679 | 11.9 | d__Bacteria;p__Actinobacteriota;c__Coriobacteriia;o__Coriobacteriales;f__Eggerthellaceae;g__Eggerthella | 94.6 | blast;96.20 |
| S5-wms_85650 | 99.85 | d__Bacteria;p__Bacteroidota;c__Bacteroidia;o__Bacteroidales;f__Bacteroidaceae;g__Bacteroides | 100 | RaFAH;99.00 iPHoP-RF;97.10 blast;96.60 |
| S5-wms_85650 | 99.85 | d__Bacteria;p__Bacteroidota;c__Bacteroidia;o__Bacteroidales;f__Bacteroidaceae;g__Phocaeicola | 94.9 | iPHoP-RF;96.40 blast;90.30 |
| S5-wms_94965 | 12.7 | d__Bacteria;p__Actinobacteriota;c__Coriobacteriia;o__Coriobacteriales;f__Coriobacteriaceae;g__Collinsella | 97.5 | CRISPR;98.20 blast;95.40 iPHoP-RF;77.10 |
| S6-wms-combined_110950 | 42.23 | d__Bacteria;p__Firmicutes;c__Clostridia;o__Oscillospirales;f__Ruminococcaceae;g__Faecalibacterium | 99.6 | blast;96.80 RaFAH;93.40 iPHoP-RF;93.10 CRISPR;73.10 |
| S6-wms-combined_117756 | 50.35 | d__Bacteria;p__Firmicutes;c__Clostridia;o__Lachnospirales;f__Lachnospiraceae;g__Acetatifactor | 91.6 | blast;93.90 iPHoP-RF;77.50 |
| S6-wms-combined_118427 | 40.24 | d__Bacteria;p__Firmicutes;c__Clostridia;o__Oscillospirales;f__Oscillospiraceae;g__Dysosmobacter | 97.9 | iPHoP-RF;93.70 blast;91.90 RaFAH;88.40 |
| S6-wms-combined_118427 | 40.24 | d__Bacteria;p__Firmicutes;c__Clostridia;o__Oscillospirales;f__Oscillospiraceae;g__CAG-83 | 96.3 | blast;88.80 RaFAH;88.40 iPHoP-RF;78.50 |
| S6-wms-combined_119017 | 37.52 | d__Bacteria;p__Desulfobacterota;c__Desulfovibrionia;o__Desulfovibrionales;f__Desulfovibrionaceae;g__Bilophila | 94.2 | blast;95.90 iPHoP-RF;92.40 CRISPR;66.40 |

| | | | | |
|---|---|---|---|---|
| S6-wms-combined_119017 | 37.52 | d__Bacteria;p__Desulfobacterota;c__Desulfovibrionia;o__Desulfovibrionales;f__Desulfovibrionaceae;g__Bilophila | 92.7 | blast;94.80 iPHoP-RF;92.80 |
| S6-wms-combined_119138 | 99.25 | d__Bacteria;p__Firmicutes;c__Clostridia;o__Lachnospirales;f__Lachnospiraceae;g__Enterocloster | 91.6 | blast;93.90 |
| S6-wms-combined_126236 | 26.53 | d__Bacteria;p__Firmicutes;c__Clostridia;o__Oscillospirales;f__Ruminococcaceae;g__Faecalibacterium | 97.6 | blast;98.30 CRISPR;97.70 iPHoP-RF;86.70 |
| S6-wms-combined_135223 | 23.09 | d__Bacteria;p__Bacteroidota;c__Bacteroidia;o__Bacteroidales;f__Bacteroidaceae;g__Bacteroides | 99 | iPHoP-RF;96.40 blast;92.30 RaFAH;88.60 |
| S6-wms-combined_135223 | 23.09 | d__Bacteria;p__Bacteroidota;c__Bacteroidia;o__Bacteroidales;f__Bacteroidaceae;g__Phocaeicola | 94.9 | iPHoP-RF;96.40 blast;89.30 |
| S6-wms-combined_138710 | 55.97 | d__Bacteria;p__Firmicutes;c__Clostridia;o__Oscillospirales;f__Ruminococcaceae;g__Faecalibacterium | 96.2 | blast;96.90 iPHoP-RF;78.00 RaFAH;56.20 |
| S6-wms-combined_140731 | 27.76 | d__Bacteria;p__Firmicutes;c__Clostridia;o__Oscillospirales;f__Ruminococcaceae;g__Faecalibacterium | 98.4 | CRISPR;92.30 RaFAH;92.10 blast;92.10 iPHoP-RF;80.30 |
| S6-wms-combined_32530 | 99.74 | d__Bacteria;p__Firmicutes;c__Clostridia;o__Lachnospirales;f__Lachnospiraceae;g__Roseburia | 99.3 | RaFAH;94.70 blast;87.00 iPHoP-RF;77.10 |
| S6-wms-combined_32530 | 99.74 | d__Bacteria;p__Firmicutes;c__Clostridia;o__Lachnospirales;f__Lachnospiraceae;g__Agathobacter | 97.8 | RaFAH;94.70 blast;81.80 iPHoP-RF;59.80 |
| S6-wms-combined_32530 | 99.74 | d__Bacteria;p__Firmicutes;c__Clostridia;o__Lachnospirales;f__Lachnospiraceae;g__RUG115 | 96 | RaFAH;94.70 blast;71.80 iPHoP-RF;53.80 |
| S6-wms-combined_32530 | 99.74 | d__Bacteria;p__Firmicutes;c__Clostridia;o__Lachnospirales;f__Lachnospiraceae;g__Acetatifactor | 90.4 | blast;92.90 |
| S6-wms-combined_41246 | 21.08 | d__Bacteria;p__Firmicutes;c__Clostridia;o__Oscillospirales;f__Oscillospiraceae;g__Marseille-P3106 | 91.2 | blast;93.60 |
| S6-wms-combined_49909 | 50.09 | d__Bacteria;p__Firmicutes;c__Clostridia;o__Oscillospirales;f__Ruminococcaceae;g__Faecalibacterium | 91 | blast;93.40 iPHoP-RF;86.00 |
| S6-wms-combined_50721 | 99.98 | d__Bacteria;p__Bacteroidota;c__Bacteroidia;o__Bacteroidales;f__Rikenellaceae;g__Alistipes | 99.6 | blast;96.90 iPHoP-RF;96.70 RaFAH;94.30 |
| S6-wms-combined_50721 | 99.98 | d__Bacteria;p__Bacteroidota;c__Bacteroidia;o__Bacteroidales;f__Rikenellaceae;g__Alistipes_A | 92.1 | RaFAH;94.30 |
| S6-wms-combined_56464 | 51.17 | d__Bacteria;p__Firmicutes;c__Clostridia;o__Oscillospirales;f__Ruminococcaceae;g__Faecalibacterium | 99 | blast;96.90 iPHoP-RF;92.10 RaFAH;87.90 |
| S6-wms-combined_65225 | 23.74 | d__Bacteria;p__Firmicutes;c__Negativicutes;o__Veillonellales;f__Dialisteraceae;g__Dialister | 94.9 | blast;96.40 CRISPR;86.10 iPHoP-RF;77.30 |
| S6-wms-combined_72228 | 41.84 | d__Bacteria;p__Bacteroidota;c__Bacteroidia;o__Bacteroidales;f__Rikenellaceae;g__Alistipes | 99 | iPHoP-RF;97.10 blast;96.60 RaFAH;85.90 |
| S6-wms-combined_745 | 7.61 | d__Bacteria;p__Proteobacteria;c__Alphaproteobacteria;o__RF32;f__CAG-239;g__CAG-267 | 90.6 | blast;93.00 iPHoP-RF;84.50 |
| S6-wms-combined_82597 | 91.55 | d__Bacteria;p__Bacteroidota;c__Bacteroidia;o__Bacteroidales;f__Bacteroidaceae;g__Bacteroides | 99.7 | iPHoP-RF;97.10 blast;96.90 RaFAH;95.40 |
| S6-wms-combined_91039 | 100 | d__Bacteria;p__Bacteroidota;c__Bacteroidia;o__Bacteroidales;f__Rikenellaceae;g__Alistipes | 96.6 | CRISPR;97.60 iPHoP-RF;97.10 blast;96.70 |
| S6-wms-combined_91039 | 100 | d__Bacteria;p__Bacteroidota;c__Bacteroidia;o__Bacteroidales;f__Bacteroidaceae;g__Bacteroides | 91.5 | RaFAH;93.80 |
| S6-wms_132588 | 67.09 | d__Bacteria;p__Firmicutes;c__Clostridia;o__Oscillospirales;f__Ruminococcaceae;g__Faecalibacterium | 93.2 | CRISPR;95.20 blast;94.40 iPHoP-RF;92.80 |
| S6-wms_138342 | 37.57 | d__Bacteria;p__Bacteroidota;c__Bacteroidia;o__Bacteroidales;f__Bacteroidaceae;g__Prevotella | 91.7 | RaFAH;94.00 |
| S6-wms_138342 | 37.57 | d__Bacteria;p__Spirochaetota;c__Spirochaetia;o__Sphaerochaetales;f__Sphaerochaetaceae;g__UBA9732 | 90 | blast;92.50 iPHoP-RF;60.30 |
| S6-wms_140743 | 9.85 | d__Bacteria;p__Firmicutes;c__Clostridia;o__Lachnospirales;f__Lachnospiraceae;g__Enterocloster | 91.2 | blast;93.60 iPHoP-RF;92.40 CRISPR;53.80 |
| S6-wms_54578 | 97 | d__Bacteria;p__Bacteroidota;c__Bacteroidia;o__Bacteroidales;f__Tannerellaceae;g__Marseille-P3160 | 97.5 | CRISPR;98.20 iPHoP-RF;69.80 |
| S6-wms_54578 | 97 | d__Bacteria;p__Bacteroidota;c__Bacteroidia;o__Bacteroidales;f__Tannerellaceae;g__Parabacteroides | 92 | blast;94.20 iPHoP-RF;68.00 |
| S6-wms_54578 | 97 | d__Bacteria;p__Bacteroidota;c__Bacteroidia;o__Bacteroidales;f__Marinifilaceae;g__Odoribacter | 90.7 | RaFAH;93.10 |
| S7-wms-combined_104872 | 97.79 | d__Bacteria;p__Bacteroidota;c__Bacteroidia;o__Bacteroidales;f__Bacteroidaceae;g__Bacteroides | 100 | RaFAH;99.70 blast;92.10 iPHoP-RF;90.50 CRISPR;53.10 |
| S7-wms-combined_106253 | 71.87 | d__Bacteria;p__Firmicutes;c__Clostridia;o__Lachnospirales;f__Lachnospiraceae;g__Dorea_A | 95.3 | blast;96.70 iPHoP-RF;94.40 |
| S7-wms-combined_106253 | 71.87 | d__Bacteria;p__Firmicutes;c__Clostridia;o__Lachnospirales;f__Lachnospiraceae;g__Extibacter | 92.2 | iPHoP-RF;94.40 blast;79.50 |
| S7-wms-combined_107679_0 | 99.98 | d__Bacteria;p__Bacteroidota;c__Bacteroidia;o__Bacteroidales;f__Rikenellaceae;g__Alistipes | 100 | RaFAH;98.50 blast;97.20 iPHoP-RF;96.40 |
| S7-wms-combined_107679_0 | 99.98 | d__Bacteria;p__Bacteroidota;c__Bacteroidia;o__Bacteroidales;f__Rikenellaceae;g__Alistipes_A | 97.9 | RaFAH;98.50 |
| S7-wms-combined_107679_1 | 100 | d__Bacteria;p__Bacteroidota;c__Bacteroidia;o__Bacteroidales;f__Bacteroidaceae;g__Bacteroides | 99.9 | RaFAH;99.80 iPHoP-RF;57.60 |
| S7-wms-combined_107679_1 | 100 | d__Bacteria;p__Bacteroidota;c__Bacteroidia;o__Bacteroidales;f__Rikenellaceae;g__Alistipes | 98.1 | blast;98.60 iPHoP-RF;64.80 |
| S7-wms-combined_111004 | 34.72 | d__Bacteria;p__Firmicutes;c__Clostridia;o__Oscillospirales;f__Ruminococcaceae;g__Faecalibacterium | 99.7 | CRISPR;98.50 blast;96.60 RaFAH;90.40 iPHoP-RF;85.70 |
| S7-wms-combined_123085 | 3.26 | d__Bacteria;p__Firmicutes;c__Clostridia;o__Lachnospirales;f__Lachnospiraceae;g__Fusicatenibacter | 91.6 | blast;93.90 iPHoP-RF;83.90 |
| S7-wms-combined_134452 | 8.94 | d__Bacteria;p__Firmicutes;c__Clostridia;o__Christensenellales;f__CAG-74;g__UBA11524 | 95.6 | blast;96.90 iPHoP-RF;94.70 |
| S7-wms-combined_134452 | 8.94 | d__Bacteria;p__Firmicutes;c__Clostridia;o__Christensenellales;f__CAG-74;g__UBA11524 | 95.6 | blast;96.90 CRISPR;95.90 iPHoP-RF;92.80 |
| S7-wms-combined_134458 | 43.62 | d__Bacteria;p__Firmicutes;c__Clostridia;o__Oscillospirales;f__Ruminococcaceae;g__Gemmiger | 91.6 | blast;93.90 iPHoP-RF;88.90 CRISPR;69.70 |
| S7-wms-combined_135154 | 99.98 | d__Bacteria;p__Bacteroidota;c__Bacteroidia;o__Bacteroidales;f__Bacteroidaceae;g__Bacteroides | 100 | RaFAH;99.30 iPHoP-RF;97.10 blast;92.40 |
| S7-wms-combined_135154 | 99.98 | d__Bacteria;p__Bacteroidota;c__Bacteroidia;o__Bacteroidales;f__Bacteroidaceae;g__Phocaeicola | 93.9 | iPHoP-RF;95.70 blast;83.30 |
| S7-wms-combined_136642 | 81.83 | d__Bacteria;p__Actinobacteriota;c__Actinomycetia;o__Actinomycetales;f__Bifidobacteriaceae;g__Bifidobacterium | 99.7 | CRISPR;99.40 iPHoP-RF;97.10 blast;96.80 RaFAH;72.30 |
| S7-wms-combined_137858 | 55.93 | d__Bacteria;p__Firmicutes;c__Clostridia;o__Oscillospirales;f__Ruminococcaceae;g__Faecalibacterium | 95.6 | blast;96.90 iPHoP-RF;88.20 |
| S7-wms-combined_139683 | 93.85 | d__Bacteria;p__Firmicutes;c__Clostridia;o__Oscillospirales;f__Oscillospiraceae;g__CAG-103 | 93.5 | CRISPR;95.40 |
| S7-wms-combined_14267 | 96.95 | d__Bacteria;p__Bacteroidota;c__Bacteroidia;o__Bacteroidales;f__Rikenellaceae;g__Alistipes | 97.5 | CRISPR;98.20 blast;98.10 iPHoP-RF;97.10 |
| S7-wms-combined_143218 | 6.41 | d__Bacteria;p__Actinobacteriota;c__Actinomycetia;o__Actinomycetales;f__Bifidobacteriaceae;g__Bifidobacterium | 95.2 | blast;96.60 CRISPR;91.70 iPHoP-RF;81.90 |
| S7-wms-combined_14529 | 25.87 | d__Bacteria;p__Firmicutes;c__Clostridia;o__Christensenellales;f__CAG-74;g__UBA11524 | 95.6 | blast;96.90 iPHoP-RF;93.40 |
| S7-wms-combined_14529 | 25.87 | d__Bacteria;p__Firmicutes;c__Clostridia;o__Christensenellales;f__CAG-74;g__UBA11524 | 95.6 | blast;96.90 iPHoP-RF;92.80 CRISPR;76.00 |
| S7-wms-combined_147158 | 86.44 | d__Bacteria;p__Actinobacteriota;c__Actinomycetia;o__Actinomycetales;f__Bifidobacteriaceae;g__Bifidobacterium | 99 | CRISPR;99.20 blast;96.90 iPHoP-RF;95.70 RaFAH;50.40 |
| S7-wms-combined_156351 | 22 | d__Bacteria;p__Firmicutes;c__Clostridia;o__Lachnospirales;f__Lachnospiraceae;g__Blautia_A | 94.5 | blast;96.10 iPHoP-RF;94.10 |
| S7-wms-combined_157260 | 99.48 | d__Bacteria;p__Firmicutes;c__Clostridia;o__Lachnospirales;f__Lachnospiraceae;g__Lachnospira | 98.7 | CRISPR;99.00 |
| S7-wms-combined_157434 | 16.06 | d__Bacteria;p__Firmicutes;c__Clostridia;o__Lachnospirales;f__Lachnospiraceae;g__Mediterraneibacter | 91.4 | blast;93.70 iPHoP-RF;86.30 |
| S7-wms-combined_163980 | 78.4 | d__Bacteria;p__Bacteroidota;c__Bacteroidia;o__Bacteroidales;f__Rikenellaceae;g__Alistipes | 95.2 | blast;96.60 iPHoP-RF;93.70 |
| S7-wms-combined_167867 | 10.54 | d__Bacteria;p__Actinobacteriota;c__Actinomycetia;o__Actinomycetales;f__Bifidobacteriaceae;g__Bifidobacterium | 97.9 | CRISPR;98.50 iPHoP-RF;97.10 blast;96.70 |
| S7-wms-combined_169375 | 10.64 | d__Bacteria;p__Firmicutes;c__Bacilli;o__Erysipelotrichales;f__Erysipelatoclostridiaceae;g__Erysipelatoclostridium | 94.7 | blast;96.30 |
| S7-wms-combined_169375 | 10.64 | d__Bacteria;p__Firmicutes;c__Clostridia;o__Lachnospirales;f__Lachnospiraceae;g__Faecalimonas | 94.5 | blast;96.10 iPHoP-RF;79.00 |
| S7-wms-combined_174935 | 34.84 | d__Bacteria;p__Actinobacteriota;c__Actinomycetia;o__Actinomycetales;f__Bifidobacteriaceae;g__Bifidobacterium | 96.3 | CRISPR;97.40 blast;97.30 iPHoP-RF;97.10 |
| S7-wms-combined_185016 | 95.96 | d__Bacteria;p__Bacteroidota;c__Bacteroidia;o__Bacteroidales;f__Bacteroidaceae;g__Bacteroides | 100 | RaFAH;99.70 blast;95.30 iPHoP-RF;95.10 |
| S7-wms-combined_189250 | 22.37 | d__Bacteria;p__Actinobacteriota;c__Coriobacteriia;o__Coriobacteriales;f__Coriobacteriaceae;g__Collinsella | 98.7 | CRISPR;99.00 blast;91.90 iPHoP-RF;80.00 |
| S7-wms-combined_190281 | 13.36 | d__Bacteria;p__Actinobacteriota;c__Coriobacteriia;o__Coriobacteriales;f__Coriobacteriaceae;g__Collinsella | 96.2 | CRISPR;97.30 blast;96.90 iPHoP-RF;86.30 |
| S7-wms-combined_20921 | 29.3 | d__Bacteria;p__Firmicutes;c__Clostridia;o__Lachnospirales;f__Lachnospiraceae;g__Anaerobutyricum | 95 | blast;96.50 iPHoP-RF;77.50 |
| S7-wms-combined_217226 | 97.48 | d__Bacteria;p__Verrucomicrobiota;c__Verrucomicrobiae;o__Verrucomicrobiales;f__Akkermansiaceae;g__Akkermansia | 100 | CRISPR;99.50 RaFAH;97.70 iPHoP-RF;58.20 |
| S7-wms-combined_218379 | 16.97 | d__Bacteria;p__Firmicutes;c__Clostridia;o__Oscillospirales;f__Acutalibacteraceae;g__CAG-180 | 98.7 | CRISPR;99.00 blast;96.60 iPHoP-RF;86.30 |
| S7-wms-combined_218379 | 16.97 | d__Bacteria;p__Firmicutes;c__Clostridia;o__Oscillospirales;f__Acutalibacteraceae;g__Fimenecus | 91.7 | blast;94.00 iPHoP-RF;91.80 |
| S7-wms-combined_224523 | 99.68 | d__Bacteria;p__Firmicutes;c__Clostridia;o__Oscillospirales;f__Ruminococcaceae;g__Ruminococcus_D | 99.4 | blast;96.80 iPHoP-RF;93.70 RaFAH;91.90 |
| S7-wms-combined_235165 | 18.87 | d__Bacteria;p__Firmicutes;c__Clostridia;o__Lachnospirales;f__Lachnospiraceae;g__Agathobacter | 90.2 | blast;92.70 |
| S7-wms-combined_238231 | 39.27 | d__Bacteria;p__Firmicutes;c__Clostridia;o__Oscillospirales;f__Ruminococcaceae;g__Negativibacillus | 95.6 | blast;96.90 iPHoP-RF;77.50 CRISPR;68.00 |
| S7-wms-combined_239784 | 67.8 | d__Bacteria;p__Bacteroidota;c__Bacteroidia;o__Bacteroidales;f__Bacteroidaceae;g__Bacteroides | 99.3 | iPHoP-RF;97.10 RaFAH;89.00 blast;88.60 |
| S7-wms-combined_239784 | 67.8 | d__Bacteria;p__Bacteroidota;c__Bacteroidia;o__Bacteroidales;f__Bacteroidaceae;g__Phocaeicola | 93.5 | blast;95.40 iPHoP-RF;84.80 |
| S7-wms-combined_26217 | 100 | d__Bacteria;p__Bacteroidota;c__Bacteroidia;o__Bacteroidales;f__Rikenellaceae;g__Alistipes | 100 | RaFAH;98.00 blast;96.90 iPHoP-RF;88.90 |
| S7-wms-combined_26217 | 100 | d__Bacteria;p__Bacteroidota;c__Bacteroidia;o__Bacteroidales;f__Rikenellaceae;g__Alistipes_A | 97.2 | RaFAH;98.00 |
| S7-wms-combined_28138 | 12.56 | d__Bacteria;p__Firmicutes;c__Clostridia;o__Oscillospirales;f__Acutalibacteraceae;g__Ruminococcus_E | 98.7 | CRISPR;99.00 blast;96.80 iPHoP-RF;91.10 |
| S7-wms-combined_31068 | 9.44 | d__Bacteria;p__Firmicutes;c__Clostridia;o__Oscillospirales;f__Oscillospiraceae;g__Lawsonibacter | 91.5 | blast;93.80 iPHoP-RF;81.90 |
| S7-wms-combined_3669 | 8.7 | d__Bacteria;p__Bacteroidota;c__Bacteroidia;o__Bacteroidales;f__Bacteroidaceae;g__Bacteroides | 91.5 | blast;93.80 iPHoP-RF;87.60 |
| S7-wms-combined_3697 | 72.05 | d__Bacteria;p__Bacteroidota;c__Bacteroidia;o__Bacteroidales;f__Rikenellaceae;g__Alistipes | 98.8 | iPHoP-RF;97.10 blast;96.90 CRISPR;95.90 RaFAH;84.30 |

| | | | | |
|---|---|---|---|---|
| S7-wms-combined_43950 | 99.99 | d__Bacteria;p__Bacteroidota;c__Bacteroidia;o__Bacteroidales;f__Bacteroidaceae;g__Bacteroides | 100 | RaFAH;99.70 iPHoP-RF;95.10 blast;93.90 |
| S7-wms-combined_43950 | 99.99 | d__Bacteria;p__Bacteroidota;c__Bacteroidia;o__Bacteroidales;f__Bacteroidaceae;g__Phocaeicola | 92.9 | blast;94.90 iPHoP-RF;93.10 |
| S7-wms-combined_5625 | 95.36 | d__Bacteria;p__Bacteroidota;c__Bacteroidia;o__Bacteroidales;f__Bacteroidaceae;g__Bacteroides | 100 | RaFAH;98.60 iPHoP-RF;97.10 blast;96.90 |
| S7-wms-combined_62119 | 99.99 | d__Bacteria;p__Bacteroidota;c__Bacteroidia;o__Bacteroidales;f__Bacteroidaceae;g__Bacteroides | 98.8 | blast;96.90 iPHoP-RF;96.40 CRISPR;93.30 RaFAH;84.80 |
| S7-wms-combined_62119 | 99.99 | d__Bacteria;p__Bacteroidota;c__Bacteroidia;o__Bacteroidales;f__Bacteroidaceae;g__Phocaeicola | 95.7 | CRISPR;97.00 iPHoP-RF;96.40 blast;62.40 |
| S7-wms-combined_69024 | 58.17 | d__Bacteria;p__Firmicutes;c__Clostridia;o__Lachnospirales;f__Lachnospiraceae;g__Bariatricus | 91.6 | blast;93.90 iPHoP-RF;75.90 |
| S7-wms-combined_71592 | 39.23 | d__Bacteria;p__Firmicutes;c__Clostridia;o__Oscillospirales;f__Ruminococcaceae;g__Ruminococcus_D | 98.1 | blast;98.60 iPHoP-RF;92.40 CRISPR;61.00 |
| S7-wms-combined_72070 | 35.71 | d__Bacteria;p__Bacteroidota;c__Bacteroidia;o__Bacteroidales;f__Bacteroidaceae;g__Bacteroides | 96.2 | iPHoP-RF;97.10 blast;96.90 CRISPR;55.90 RaFAH;52.60 |
| S7-wms-combined_74829 | 77.29 | d__Bacteria;p__Firmicutes;c__Negativicutes;o__Veillonellales;f__Dialisteraceae;g__Dialister | 97.3 | CRISPR;97.40 blast;96.80 iPHoP-RF;94.70 RaFAH;63.90 |
| S7-wms-combined_84688 | 29.6 | d__Bacteria;p__Firmicutes;c__Clostridia;o__Oscillospirales;f__Oscillospiraceae;g__Marseille-P3106 | 91.6 | blast;93.90 iPHoP-RF;86.70 |
| S7-wms-combined_90393 | 32.36 | d__Bacteria;p__Bacteroidota;c__Bacteroidia;o__Bacteroidales;f__Bacteroidaceae;g__Bacteroides | 99.4 | iPHoP-RF;97.10 blast;91.80 RaFAH;91.30 |
| S7-wms-combined_90728 | 66.45 | d__Bacteria;p__Bacteroidota;c__Bacteroidia;o__Bacteroidales;f__Bacteroidaceae;g__Bacteroides | 100 | RaFAH;98.90 iPHoP-RF;96.70 blast;91.80 |
| S7-wms_113137 | 99.39 | d__Bacteria;p__Firmicutes;c__Clostridia;o__Oscillospirales;f__Ruminococcaceae;g__Faecalibacterium | 99.4 | RaFAH;99.50 |
| S7-wms_123954 | 100 | d__Bacteria;p__Bacteroidota;c__Bacteroidia;o__Bacteroidales;f__Tannerellaceae;g__Parabacteroides | 98.4 | blast;96.40 RaFAH;82.90 iPHoP-RF;60.30 |
| S7-wms_127801 | 10.6 | d__Bacteria;p__Firmicutes;c__Clostridia;o__Oscillospirales;f__Ruminococcaceae;g__Ruminococcus_D | 98.7 | CRISPR;99.00 blast;97.40 iPHoP-RF;93.70 |
| S7-wms_151112 | 20.08 | d__Bacteria;p__Firmicutes;c__Clostridia;o__Lachnospirales;f__Lachnospiraceae;g__Anaerobutyricum | 90.3 | blast;92.80 iPHoP-RF;86.00 |
| S7-wms_157264 | 5.31 | d__Bacteria;p__Actinobacteriota;c__Actinomycetia;o__Actinomycetales;f__Bifidobacteriaceae;g__Bifidobacterium | 99.7 | CRISPR;99.70 iPHoP-RF;73.50 |
| S7-wms_244866 | 18.88 | d__Bacteria;p__Bacteroidota;c__Bacteroidia;o__Bacteroidales;f__Bacteroidaceae;g__Bacteroides | 97.6 | CRISPR;98.30 iPHoP-RF;92.40 |
| S7-wms_80255 | 74.19 | d__Bacteria;p__Firmicutes;c__Clostridia;o__Oscillospirales;f__Ruminococcaceae;g__Ruminiclostridium_E | 91.5 | blast;93.80 iPHoP-RF;67.00 CRISPR;57.40 |
| S8-wms-combined_134098 | 81.54 | d__Bacteria;p__Firmicutes;c__Clostridia;o__Oscillospirales;f__Oscillospiraceae;g__ER4 | 95.2 | blast;96.60 iPHoP-RF;76.10 |
| S8-wms-combined_146562 | 66.24 | d__Bacteria;p__Firmicutes;c__Clostridia;o__Christensenellales;f__CAG-138;g__UBA1685 | 93 | blast;95.00 iPHoP-RF;82.80 CRISPR;70.40 |
| S8-wms-combined_164289 | 21.14 | d__Bacteria;p__Firmicutes;c__Clostridia;o__Christensenellales;f__CAG-138;g__SFEL01 | 94.5 | blast;96.10 iPHoP-RF;79.00 CRISPR;71.90 |
| S8-wms-combined_169835 | 17.94 | d__Bacteria;p__Firmicutes;c__Clostridia;o__Oscillospirales;f__Oscillospiraceae;g__CAG-83 | 95.6 | blast;96.90 iPHoP-RF;64.30 |
| S8-wms-combined_180957 | 99.92 | d__Bacteria;p__Bacteroidota;c__Bacteroidia;o__Bacteroidales;f__Tannerellaceae;g__Parabacteroides | 99.7 | blast;96.70 RaFAH;95.30 iPHoP-RF;84.50 CRISPR;57.90 |
| S8-wms-combined_185611 | 9.67 | d__Bacteria;p__Firmicutes;c__Clostridia;o__Christensenellales;f__CAG-74;g__Limiplasma | 94.2 | blast;95.90 iPHoP-RF;76.60 |
| S8-wms-combined_185611 | 9.67 | d__Bacteria;p__Firmicutes;c__Clostridia;o__Christensenellales;f__CAG-74;g__Firm-11 | 92 | blast;94.20 iPHoP-RF;85.10 |
| S8-wms-combined_211119 | 22.48 | d__Bacteria;p__Firmicutes;c__Clostridia;o__Christensenellales;f__CAG-138;g__UBA1685 | 95.9 | blast;97.10 iPHoP-RF;77.50 CRISPR;68.50 |
| S8-wms-combined_215102 | 12.51 | d__Bacteria;p__Firmicutes;c__Clostridia;o__Christensenellales;f__CAG-74;g__UBA11524 | 95.6 | blast;96.90 iPHoP-RF;94.70 |
| S8-wms-combined_215102 | 12.51 | d__Bacteria;p__Firmicutes;c__Clostridia;o__Christensenellales;f__CAG-74;g__UBA11524 | 95.6 | blast;96.90 iPHoP-RF;93.40 CRISPR;77.10 |
| S8-wms-combined_217866 | 98.95 | d__Bacteria;p__Firmicutes;c__Clostridia;o__Lachnospirales;f__Lachnospiraceae;g__Roseburia | 96.3 | CRISPR;97.40 blast;68.80 |
| S8-wms-combined_217866 | 98.95 | d__Bacteria;p__Firmicutes;c__Clostridia;o__Lachnospirales;f__Lachnospiraceae;g__Agathobacter | 95.7 | CRISPR;97.00 blast;61.30 |
| S8-wms-combined_217866 | 98.95 | d__Bacteria;p__Firmicutes;c__Clostridia;o__Lachnospirales;f__Lachnospiraceae;g__Eubacterium_F | 91.6 | blast;93.90 |
| S8-wms-combined_219796 | 14.07 | d__Bacteria;p__Firmicutes;c__Clostridia;o__Christensenellales;f__CAG-138;g__UBA1685 | 93.5 | blast;95.40 CRISPR;93.90 iPHoP-RF;88.20 |
| S8-wms-combined_225114 | 99.67 | d__Bacteria;p__Firmicutes;c__Clostridia;o__Oscillospirales;f__Ruminococcaceae;g__Faecalibacterium | 100 | CRISPR;98.40 RaFAH;98.00 blast;92.30 iPHoP-RF;88.50 |
| S8-wms-combined_23583 | 28.29 | d__Bacteria;p__Bacteroidota;c__Bacteroidia;o__Bacteroidales;f__Rikenellaceae;g__Alistipes_A | 90.3 | blast;92.80 iPHoP-RF;81.60 CRISPR;68.00 |
| S8-wms-combined_239791 | 19.46 | d__Bacteria;p__Bacteroidota;c__Bacteroidia;o__Bacteroidales;f__Bacteroidaceae;g__Bacteroides | 96.3 | iPHoP-RF;95.10 RaFAH;73.50 |
| S8-wms-combined_246185 | 33.37 | d__Bacteria;p__Verrucomicrobiota;c__Verrucomicrobiae;o__Verrucomicrobiales;f__Akkermansiaceae;g__Akkermansia | 98.4 | CRISPR;98.80 blast;96.90 iPHoP-RF;81.10 |
| S8-wms-combined_246389 | 21.76 | d__Bacteria;p__Firmicutes;c__Clostridia;o__Oscillospirales;f__Oscillospiraceae;g__ER4 | 91.6 | blast;93.90 iPHoP-RF;64.50 |
| S8-wms-combined_25607 | 54.47 | d__Bacteria;p__Firmicutes;c__Clostridia;o__Oscillospirales;f__Oscillospiraceae;g__UBA9475 | 91.2 | blast;93.60 iPHoP-RF;68.90 |
| S8-wms-combined_26087 | 100 | d__Bacteria;p__Bacteroidota;c__Bacteroidia;o__Bacteroidales;f__Rikenellaceae;g__Alistipes | 98.7 | blast;96.90 iPHoP-RF;91.40 RaFAH;83.40 |
| S8-wms-combined_264711 | 10.73 | d__Bacteria;p__Firmicutes;c__Clostridia;o__Christensenellales;f__CAG-138;g__SFEL01 | 90.9 | blast;93.30 iPHoP-RF;74.80 |
| S8-wms-combined_270362 | 99.95 | d__Bacteria;p__Bacteroidota;c__Bacteroidia;o__Bacteroidales;f__Rikenellaceae;g__Alistipes | 100 | RaFAH;97.30 blast;96.90 iPHoP-RF;90.10 |
| S8-wms-combined_270362 | 99.95 | d__Bacteria;p__Bacteroidota;c__Bacteroidia;o__Bacteroidales;f__Rikenellaceae;g__Alistipes_A | 96.2 | RaFAH;97.30 |
| S8-wms-combined_40666 | 26.57 | d__Bacteria;p__Firmicutes;c__Clostridia;o__Oscillospirales;f__Oscillospiraceae;g__CAG-103 | 97.2 | CRISPR;98.00 blast;94.30 iPHoP-RF;83.60 |
| S8-wms-combined_49369 | 96.98 | d__Bacteria;p__Firmicutes;c__Clostridia;o__Oscillospirales;f__Ruminococcaceae;g__Gemmiger | 94 | blast;95.20 iPHoP-RF;61.70 RaFAH;56.00 |
| S8-wms-combined_68780 | 51.97 | d__Bacteria;p__Firmicutes;c__Clostridia;o__Oscillospirales;f__Oscillospiraceae;g__F23-B02 | 94.6 | blast;96.20 iPHoP-RF;58.40 |
| S8-wms-combined_74275 | 54.68 | d__Bacteria;p__Bacteroidota;c__Bacteroidia;o__Bacteroidales;f__Bacteroidaceae;g__Prevotella | 99.3 | blast;96.90 iPHoP-RF;91.40 RaFAH;90.60 |
| S8-wms-combined_81597 | 99.93 | d__Bacteria;p__Bacteroidota;c__Bacteroidia;o__Bacteroidales;f__Bacteroidaceae;g__Bacteroides | 100 | RaFAH;99.50 blast;95.10 iPHoP-RF;85.40 |
| S8-wms-combined_97848 | 75.58 | d__Bacteria;p__Bacteroidota;c__Bacteroidia;o__Bacteroidales;f__Bacteroidaceae;g__Bacteroides | 98.8 | RaFAH;92.20 blast;92.00 iPHoP-RF;62.20 |
| S8-wms_152831 | 99.72 | d__Bacteria;p__Firmicutes;c__Clostridia;o__Oscillospirales;f__Ruminococcaceae;g__Faecalibacterium | 100 | RaFAH;99.50 CRISPR;97.80 blast;95.40 iPHoP-RF;77.30 |
| S8-wms_190781 | 93.96 | d__Bacteria;p__Firmicutes;c__Clostridia;o__Oscillospirales;f__Ruminococcaceae;g__Faecalibacterium | 92 | CRISPR;94.20 blast;94.00 iPHoP-RF;83.30 |
| S8-wms_20528 | 48.51 | d__Bacteria;p__Firmicutes;c__Clostridia;o__Oscillospirales;f__Ruminococcaceae;g__Gemmiger | 91.6 | blast;93.90 iPHoP-RF;84.50 |
| S8-wms_270 | | d__Bacteria;p__Bacteroidota;c__Bacteroidia;o__Bacteroidales;f__Barnesiellaceae;g__Barnesiella | 94.6 | blast;96.20 iPHoP-RF;68.20 |
| S8-wms_30498 | 86.77 | d__Bacteria;p__Firmicutes;c__Clostridia;o__Oscillospirales;f__Oscillospiraceae;g__CAG-83 | 92.7 | blast;86.00 RaFAH;81.50 iPHoP-RF;63.00 |
| S8-wms_48935 | 39.33 | d__Bacteria;p__Firmicutes;c__Clostridia;o__Oscillospirales;f__Oscillospiraceae;g__CAG-83 | 90.9 | blast;93.30 iPHoP-RF;77.30 |
| S8-wms_57121 | 9.54 | d__Bacteria;p__Firmicutes;c__Clostridia;o__Christensenellales;f__CAG-138;g__UBA1685 | 97.3 | blast;98.10 iPHoP-RF;76.60 CRISPR;55.40 |
| S8-wms_58515 | 74.23 | d__Bacteria;p__Bacteroidota;c__Bacteroidia;o__Bacteroidales;f__Rikenellaceae;g__Alistipes | 96.3 | iPHoP-RF;97.10 blast;96.70 CRISPR;91.70 RaFAH;55.50 |
| S8-wms_82880 | 30.76 | d__Bacteria;p__Firmicutes;c__Clostridia;o__Oscillospirales;f__Oscillospiraceae;g__An92 | 93 | blast;95.00 iPHoP-RF;60.10 |
| S8-wms_82880 | 30.76 | d__Bacteria;p__Firmicutes;c__Clostridia;o__Oscillospirales;f__Oscillospiraceae;g__Flavonifractor | 91 | blast;93.40 iPHoP-RF;75.90 |
| S9-wms-combined_105462 | 98 | d__Bacteria;p__Firmicutes;c__Clostridia;o__Lachnospirales;f__Lachnospiraceae;g__Anaerostipes | 95.4 | blast;96.80 CRISPR;91.30 iPHoP-RF;91.10 |
| S9-wms-combined_105822 | 44.89 | d__Bacteria;p__Firmicutes;c__Clostridia;o__Lachnospirales;f__Lachnospiraceae;g__Enterocloster | 95 | blast;96.50 iPHoP-RF;87.60 CRISPR;52.20 |
| S9-wms-combined_10990 | 10.09 | d__Bacteria;p__Firmicutes;c__Clostridia;o__Lachnospirales;f__Lachnospiraceae;g__Blautia_A | 91.4 | blast;93.70 iPHoP-RF;80.00 |
| S9-wms-combined_122742 | 8.62 | d__Bacteria;p__Firmicutes;c__Clostridia;o__Oscillospirales;f__Oscillospiraceae;g__Flavonifractor | 90.3 | blast;92.80 iPHoP-RF;72.80 |
| S9-wms-combined_124857 | 79.58 | d__Bacteria;p__Firmicutes;c__Clostridia;o__Oscillospirales;f__Oscillospiraceae;g__Flavonifractor | 90.7 | iPHoP-RF;93.10 blast;92.10 CRISPR;71.70 |
| S9-wms-combined_2042 | 10.88 | d__Bacteria;p__Firmicutes;c__Clostridia;o__Lachnospirales;f__Lachnospiraceae;g__Eisenbergiella | 95.2 | CRISPR;96.60 blast;95.10 iPHoP-RF;93.40 |
| S9-wms-combined_20485 | 94.46 | d__Bacteria;p__Firmicutes;c__Bacilli;o__Lactobacillales;f__Streptococcaceae;g__Streptococcus | 100 | RaFAH;99.70 CRISPR;99.00 iPHoP-RF;97.10 blast;96.90 |
| S9-wms-combined_26410 | 99.06 | d__Bacteria;p__Firmicutes;c__Clostridia;o__Lachnospirales;f__Lachnospiraceae;g__Mediterraneibacter | 95.4 | blast;96.80 |
| S9-wms-combined_26410 | 99.06 | d__Bacteria;p__Firmicutes;c__Clostridia;o__Lachnospirales;f__Lachnospiraceae;g__Hungatella | 95.2 | blast;88.20 RaFAH;85.60 |
| S9-wms-combined_27311 | 4.98 | d__Bacteria;p__Firmicutes;c__Clostridia;o__Oscillospirales;f__Acutalibacteraceae;g__Acutalibacter | 91.4 | blast;93.70 iPHoP-RF;76.10 |
| S9-wms-combined_28805 | 94.95 | d__Bacteria;p__Bacteroidota;c__Bacteroidia;o__Bacteroidales;f__Rikenellaceae;g__Alistipes | 99 | blast;96.10 RaFAH;89.50 iPHoP-RF;68.40 |
| S9-wms-combined_31456 | 63.18 | d__Bacteria;p__Bacteroidota;c__Bacteroidia;o__Bacteroidales;f__Barnesiellaceae;g__Barnesiella | 97.6 | CRISPR;97.40 blast;96.90 iPHoP-RF;83.60 RaFAH;68.00 |
| S9-wms-combined_33229 | 87.21 | d__Bacteria;p__Bacteroidota;c__Bacteroidia;o__Bacteroidales;f__Bacteroidaceae;g__Bacteroides | 99.1 | iPHoP-RF;97.10 blast;96.10 RaFAH;87.90 |
| S9-wms-combined_33229 | 87.21 | d__Bacteria;p__Bacteroidota;c__Bacteroidia;o__Bacteroidales;f__Bacteroidaceae;g__Phocaeicola | 94.5 | iPHoP-RF;96.10 |
| S9-wms-combined_33230 | 93.11 | d__Bacteria;p__Bacteroidota;c__Bacteroidia;o__Bacteroidales;f__Bacteroidaceae;g__Bacteroides | 100 | RaFAH;99.10 iPHoP-RF;96.40 blast;92.10 |
| S9-wms-combined_3461 | 69.22 | d__Bacteria;p__Firmicutes;c__Clostridia;o__Lachnospirales;f__Anaerotignaceae;g__Anaerotignum | 95 | CRISPR;96.50 blast;91.10 iPHoP-RF;88.90 |
| S9-wms-combined_36459 | 25.89 | d__Bacteria;p__Firmicutes;c__Clostridia;o__Oscillospirales;f__Ruminococcaceae;g__Negativibacillus | 96.2 | blast;97.30 iPHoP-RF;85.10 CRISPR;59.60 |
| S9-wms-combined_37587 | 92.98 | d__Bacteria;p__Bacteroidota;c__Bacteroidia;o__Bacteroidales;f__Bacteroidaceae;g__Bacteroides | 99.4 | blast;96.80 iPHoP-RF;96.40 RaFAH;92.80 |

| | | | | |
|---|---|---|---|---|
| S9-wms-combined_37587 | 92.98 | d__Bacteria;p__Bacteroidota;c__Bacteroidia;o__Bacteroidales;f__Bacteroidaceae;g__Phocaeicola | 93.1 | iPHoP-RF;95.10 blast;90.70 |
| S9-wms-combined_44402 | 80.74 | d__Bacteria;p__Bacteroidota;c__Bacteroidia;o__Bacteroidales;f__Bacteroidaceae;g__Bacteroides | 100 | RaFAH;97.80 blast;96.50 iPHoP-RF;96.40 |
| S9-wms-combined_53991 | 30.5 | d__Bacteria;p__Firmicutes;c__Negativicutes;o__Veillonellales;f__Dialisteraceae;g__Dialister | 95.4 | blast;96.80 iPHoP-RF;91.80 CRISPR;60.90 |
| S9-wms-combined_53991 | 30.5 | d__Bacteria;p__Firmicutes;c__Negativicutes;o__Veillonellales;f__Veillonellaceae;g__F0422 | 90.4 | RaFAH;92.90 |
| S9-wms-combined_53991 | 30.5 | d__Bacteria;p__Firmicutes;c__Negativicutes;o__Veillonellales;f__Veillonellaceae;g__Veillonella | 90.4 | RaFAH;92.90 |
| S9-wms-combined_53991 | 30.5 | d__Bacteria;p__Firmicutes;c__Negativicutes;o__Veillonellales;f__Veillonellaceae;g__Veillonella_A | 90.4 | RaFAH;92.90 |
| S9-wms-combined_5526 | 86.27 | d__Bacteria;p__Bacteroidota;c__Bacteroidia;o__Bacteroidales;f__Rikenellaceae;g__Alistipes | 99.3 | blast;96.80 iPHoP-RF;93.10 RaFAH;90.40 |
| S9-wms-combined_64277 | 98.37 | d__Bacteria;p__Proteobacteria;c__Gammaproteobacteria;o__Enterobacterales;f__Enterobacteriaceae;g__Escherichia | 100 | RaFAH;97.50 blast;96.90 iPHoP-RF;74.30 |
| S9-wms-combined_64277 | 98.37 | d__Bacteria;p__Proteobacteria;c__Gammaproteobacteria;o__Enterobacterales;f__Enterobacteriaceae;g__Escherichia_C | 93.8 | blast;95.60 |
| S9-wms-combined_64277 | 98.37 | d__Bacteria;p__Proteobacteria;c__Gammaproteobacteria;o__Enterobacterales;f__Enterobacteriaceae;g__Kluyvera | 93.5 | blast;95.40 |
| S9-wms-combined_64277 | 98.37 | d__Bacteria;p__Proteobacteria;c__Gammaproteobacteria;o__Enterobacterales;f__Enterobacteriaceae;g__Pseudocitrobacter | 91.7 | blast;94.00 |
| S9-wms-combined_76274 | 91.87 | d__Bacteria;p__Actinobacteriota;c__Actinomycetia;o__Actinomycetales;f__Bifidobacteriaceae;g__Bifidobacterium | 99.4 | CRISPR;98.60 iPHoP-RF;93.70 blast;92.70 RaFAH;81.60 |
| S9-wms-combined_93232 | 57.26 | d__Bacteria;p__Proteobacteria;c__Gammaproteobacteria;o__Enterobacterales;f__Enterobacteriaceae;g__Klebsiella | 98.2 | CRISPR;98.70 blast;96.90 iPHoP-RF;59.00 |
| S9-wms-combined_93232 | 57.26 | d__Bacteria;p__Proteobacteria;c__Gammaproteobacteria;o__Enterobacterales;f__Enterobacteriaceae;g__Klebsiella_A | 94.7 | blast;96.30 |
| S9-wms-combined_97755 | 40.71 | d__Bacteria;p__Firmicutes;c__Clostridia;o__Lachnospirales;f__Lachnospiraceae;g__Eisenbergiella | 91 | blast;93.40 iPHoP-RF;57.60 |
| S9-wms-combined_9931 | 57.49 | d__Bacteria;p__Firmicutes;c__Clostridia;o__Oscillospirales;f__Acutalibacteraceae;g__Ruminococcus_E | 95.4 | blast;96.80 iPHoP-RF;92.10 |
| S9-wms_104169 | | d__Bacteria;p__Firmicutes;c__Clostridia;o__Oscillospirales;f__Ruminococcaceae;g__Ruthenibacterium | 91.6 | blast;93.90 CRISPR;84.10 iPHoP-RF;79.50 |
| S9-wms_114971 | 60.7 | d__Bacteria;p__Firmicutes;c__Negativicutes;o__Veillonellales;f__Veillonellaceae;g__Veillonella | 98.4 | blast;95.00 RaFAH;88.20 iPHoP-RF;73.50 CRISPR;55.70 |
| S9-wms_19025 | 87.64 | d__Bacteria;p__Firmicutes;c__Bacilli;o__Lactobacillales;f__Streptococcaceae;g__Streptococcus | 98.4 | RaFAH;98.80 |
| S9-wms_64472 | 4.04 | d__Bacteria;p__Firmicutes;c__Clostridia;o__Lachnospirales;f__Lachnospiraceae;g__Enterocloster | 93.5 | blast;95.40 iPHoP-RF;85.10 |

VLP_STD+MAGs_tophits

| Virus | Status | Realm | Kingdom | Phylum | Class | Order | Family | Subfamily | Genus | AAI to closest RaFAH reference | Host genus | Confidence score | List of methods |
|---|---|---|---|---|---|---|---|---|---|---|---|---|---|
| S10_vf_120738 | Shared | n.a. | n.a. | n.a. | n.a. | n.a. | n.a. | n.a. | n.a. | 22.73 | d__Bacteria;p__Firmicutes;c__Clostridia;o__Oscillospirales;f__Oscillospiraceae;g__Dysosmobacter | 95 | blast;96.50 iPHoP-RF;71.90 |
| S10_vf_18671 | Shared | Duplodnaviria | Heunggongvirae | Uroviricota | Caudoviricetes | n.a. | Unclassified | Unclassified | Unclassified | 91.64 | d__Bacteria;p__Bacteroidota;c__Bacteroidia;o__Bacteroidales;f__Tannerellaceae;g__Parabacteroides | 91 | blast;93.40 iPHoP-RF;79.00 |
| S10_vf_24791 | SHHC | Duplodnaviria | Heunggongvirae | Uroviricota | Caudoviricetes | n.a. | Unclassified | Unclassified | Eponavirus | 46.17 | d__Bacteria;p__Firmicutes;c__Clostridia;o__Oscillospirales;f__Ruminococcaceae;g__Faecalibacterium | 93.8 | blast;95.60 CRISPR;94.10 iPHoP-RF;82.20 |
| S10_vf_43964 | SHHC | Duplodnaviria | Heunggongvirae | Uroviricota | Caudoviricetes | n.a. | Unclassified | O | O | 91.64 | d__Bacteria;p__Bacteroidota;c__Bacteroidia;o__Bacteroidales;f__Bacteroidaceae;g__Bacteroides | 100 | RaFAH;99.20 CRISPR;97.80 iPHoP-RF;97.10 blast;77.70 |
| S10_vf_64371 | Shared | n.a. | n.a. | n.a. | n.a. | n.a. | n.a. | n.a. | n.a. | 86.67 | d__Bacteria;p__Firmicutes;c__Bacilli;o__Lactobacillales;f__Streptococcaceae;g__Streptococcus | 98.8 | RaFAH;98.90 iPHoP-RF;59.80 |
| S10_vf_80712 | Shared | n.a. | n.a. | n.a. | n.a. | n.a. | n.a. | n.a. | n.a. | 98.13 | d__Bacteria;p__Firmicutes;c__Clostridia;o__Lachnospirales;f__Lachnospiraceae;g__Blautia_A | 100 | RaFAH;99.20 blast;91.30 iPHoP-RF;86.30 |
| S10_vs_120489 | SHHC | n.a. | n.a. | n.a. | n.a. | n.a. | n.a. | n.a. | n.a. | 7.64 | d__Bacteria;p__Firmicutes;c__Clostridia;o__Christensenellales;f__Borkfalkiaceae;g__QALS01 | 90.3 | blast;92.80 iPHoP-RF;56.50 |
| S10_vs_120541 | SHHC | n.a. | n.a. | n.a. | n.a. | n.a. | n.a. | n.a. | n.a. | 14.85 | d__Bacteria;p__Firmicutes;c__Clostridia;o__Oscillospirales;f__Oscillospiraceae;g__CAG-110 | 94.7 | blast;96.30 iPHoP-RF;95.70 CRISPR;74.70 |
| S10_vs_14557 | Shared | Duplodnaviria | Heunggongvirae | Uroviricota | Caudoviricetes | n.a. | Duneviridae | O | O | 99.66 | d__Bacteria;p__Bacteroidota;c__Bacteroidia;o__Bacteroidales;f__Bacteroidaceae;g__Bacteroides | 100 | RaFAH;98.20 iPHoP-RF;97.10 blast;96.90 CRISPR;90.30 |
| S10_vf_46328 | SHHC | Duplodnaviria | Heunggongvirae | Uroviricota | Caudoviricetes | n.a. | Unclassified | O | O | 63.06 | d__Bacteria;p__Firmicutes;c__Clostridia;o__Lachnospirales;f__Lachnospiraceae;g__UBA9502 | 91.4 | CRISPR;93.70 blast;91.80 iPHoP-RF;89.50 |
| S10_vs_2538 | Shared | Duplodnaviria | Heunggongvirae | Uroviricota | Caudoviricetes | n.a. | Unclassified | O | O | 31.92 | d__Bacteria;p__Firmicutes;c__Clostridia;o__Peptostreptococcales;f__Anaerovoracaceae;g__CAG-145 | 94.3 | blast;96.00 iPHoP-RF;76.40 |
| S10_vs_35854 | Shared | Duplodnaviria | Heunggongvirae | Uroviricota | Caudoviricetes | n.a. | Unclassified | O | O | 48.73 | d__Bacteria;p__Bacteroidota;c__Bacteroidia;o__Bacteroidales;f__Bacteroidaceae;g__Bacteroides | 99.3 | iPHoP-RF;97.10 blast;96.00 RaFAH;90.40 |
| S10_vs_36407 | SHHC | n.a. | n.a. | n.a. | n.a. | n.a. | n.a. | n.a. | n.a. | 8.93 | d__Bacteria;p__Firmicutes;c__Clostridia;o__Oscillospirales;f__Acutalibacteraceae;g__UBA1691 | 94.5 | blast;96.10 CRISPR;90.70 iPHoP-RF;73.00 |
| S10_vf_62230 | SHHC | Duplodnaviria | Heunggongvirae | Uroviricota | Caudoviricetes | n.a. | Unclassified | O | O | 25.76 | d__Bacteria;p__Firmicutes;c__Clostridia;o__Clostridiales;f__Clostridiaceae;g__Clostridium | 90.6 | blast;93.00 iPHoP-RF;61.40 |
| S10_vs_36598 | Shared | n.a. | n.a. | n.a. | n.a. | n.a. | n.a. | n.a. | n.a. | 93.41 | d__Bacteria;p__Bacteroidota;c__Bacteroidia;o__Bacteroidales;f__Bacteroidaceae;g__Bacteroides | 100 | RaFAH;99.40 iPHoP-RF;96.70 |
| S10_vs_37379 | Shared | n.a. | n.a. | n.a. | n.a. | n.a. | n.a. | n.a. | n.a. | 75.26 | d__Bacteria;p__Bacteroidota;c__Bacteroidia;o__Bacteroidales;f__Tannerellaceae;g__Parabacteroides | 100 | CRISPR;98.90 RaFAH;98.30 iPHoP-RF;97.10 blast;94.80 |
| S10_vs_58014 | Shared | Duplodnaviria | Heunggongvirae | Uroviricota | Caudoviricetes | n.a. | Unclassified | Unclassified | Unclassified | 85.85 | d__Bacteria;p__Bacteroidota;c__Bacteroidia;o__Bacteroidales;f__Bacteroidaceae;g__Bacteroides | 100 | RaFAH;99.50 blast;91.80 iPHoP-RF;64.00 |
| S10_vs_106834 | SHHC | Duplodnaviria | Heunggongvirae | Uroviricota | Caudoviricetes | n.a. | Unclassified | O | O | 44.85 | d__Bacteria;p__Bacteroidota;c__Bacteroidia;o__Bacteroidales;f__Bacteroidaceae;g__Bacteroides | 92.3 | RaFAH;87.20 iPHoP-RF;78.70 |
| S10_vs_67471 | SHHC | n.a. | n.a. | n.a. | n.a. | n.a. | n.a. | n.a. | n.a. | 98.13 | d__Bacteria;p__Bacteroidota;c__Bacteroidia;o__Bacteroidales;f__Rikenellaceae;g__Alistipes | 99.9 | RaFAH;97.30 blast;95.30 iPHoP-RF;52.20 |
| S10_vs_71900 | Shared | n.a. | n.a. | n.a. | n.a. | n.a. | n.a. | n.a. | n.a. | 97 | d__Bacteria;p__Bacteroidota;c__Bacteroidia;o__Bacteroidales;f__Bacteroidaceae;g__Bacteroides | 100 | RaFAH;99.30 iPHoP-RF;97.10 blast;90.90 |
| S11_vf_104035 | Shared | Duplodnaviria | Heunggongvirae | Uroviricota | Caudoviricetes | Crassvirales | Suoliviridae | O | O | 37.57 | d__Bacteria;p__Bacteroidota;c__Bacteroidia;o__Bacteroidales;f__Bacteroidaceae;g__Prevotella | 91.7 | RaFAH;94.00 |
| S10_vs_62776 | Shared | Duplodnaviria | Heunggongvirae | Uroviricota | Caudoviricetes | n.a. | Unclassified | O | O | 44.54 | d__Bacteria;p__Bacteroidota;c__Bacteroidia;o__Bacteroidales;f__Bacteroidaceae;g__Bacteroides | 99 | iPHoP-RF;96.70 blast;93.60 RaFAH;88.20 |
| S10_vs_120570 | Shared | n.a. | n.a. | n.a. | n.a. | n.a. | n.a. | n.a. | n.a. | 98.22 | d__Bacteria;p__Firmicutes;c__Clostridia;o__Oscillospirales;f__Ruminococcaceae;g__Ruminococcus_D | 99.7 | blast;98.90 iPHoP-RF;96.70 CRISPR;87.50 RaFAH;87.40 |
| S11_vf_104052 | ME | Duplodnaviria | Heunggongvirae | Uroviricota | Caudoviricetes | n.a. | Unclassified | O | O | 38.91 | d__Bacteria;p__Firmicutes;c__Clostridia;o__Oscillospirales;f__Acutalibacteraceae;g__CAG-180 | 91.5 | blast;93.80 iPHoP-RF;55.40 |
| S10_vs_77004 | Shared | n.a. | n.a. | n.a. | n.a. | n.a. | n.a. | n.a. | n.a. | 10.42 | d__Bacteria;p__Firmicutes;c__Clostridia;o__Oscillospirales;f__Acutalibacteraceae;g__Pseudoruminococcus | 95.3 | blast;96.70 CRISPR;89.20 iPHoP-RF;87.60 |
| S11_vf_104033 | ME | Duplodnaviria | Heunggongvirae | Uroviricota | Caudoviricetes | n.a. | Unclassified | O | O | 40.07 | d__Bacteria;p__Bacteroidota;c__Bacteroidia;o__Bacteroidales;f__Bacteroidaceae;g__Bacteroides | 90.6 | RaFAH;93.00 |
| S11_vf_104071 | ME | Duplodnaviria | Heunggongvirae | Uroviricota | Caudoviricetes | n.a. | Unclassified | O | O | 20.29 | d__Bacteria;p__Bacteroidota;c__Bacteroidia;o__Bacteroidales;f__Rikenellaceae;g__Alistipes | 98.5 | CRISPR;98.90 iPHoP-RF;92.80 |
| S11_vf_10907 | ME | Duplodnaviria | Heunggongvirae | Uroviricota | Caudoviricetes | n.a. | Unclassified | O | O | 87.86 | d__Bacteria;p__Firmicutes;c__Clostridia;o__Lachnospirales;f__Lachnospiraceae;g__Bariatricus | 91.6 | blast;93.90 iPHoP-RF;82.80 |
| S11_vf_27845 | Shared | n.a. | n.a. | n.a. | n.a. | n.a. | n.a. | n.a. | n.a. | 53.78 | d__Bacteria;p__Firmicutes;c__Bacilli;o__Lactobacillales;f__Streptococcaceae;g__Streptococcus | 100 | CRISPR;98.80 RaFAH;97.70 iPHoP-RF;85.70 |
| S11_vf_28515 | Shared | Duplodnaviria | Heunggongvirae | Uroviricota | Caudoviricetes | n.a. | Unclassified | O | O | 95.85 | d__Bacteria;p__Firmicutes;c__Clostridia;o__Lachnospirales;f__Lachnospiraceae;g__Blautia_A | 100 | RaFAH;98.80 blast;94.40 iPHoP-RF;90.50 CRISPR;68.00 |
| S11_vf_28645 | ME | n.a. | n.a. | n.a. | n.a. | n.a. | n.a. | n.a. | n.a. | 6.84 | d__Bacteria;p__Firmicutes;c__Clostridia;o__Lachnospirales;f__Lachnospiraceae;g__Blautia_A | 92.5 | blast;94.60 iPHoP-RF;51.70 |
| S11_vf_43997 | ME | Duplodnaviria | Heunggongvirae | Uroviricota | Caudoviricetes | Crassvirales | Unclassified | Unclassified | Unclassified | 15.72 | d__Bacteria;p__Bacteroidota;c__Bacteroidia;o__Bacteroidales;f__Bacteroidaceae;g__Bacteroides | 91.4 | blast;93.70 iPHoP-RF;78.70 |
| S11_vf_66522 | ME | Duplodnaviria | Heunggongvirae | Uroviricota | Caudoviricetes | n.a. | Unclassified | Unclassified | Unclassified | 97.65 | d__Bacteria;p__Actinobacteriota;c__Actinomycetia;o__Actinomycetales;f__Actinomycetaceae;g__Pauljensenia | 100 | RaFAH;99.20 CRISPR;99.10 blast;93.90 iPHoP-RF;90.50 |
| S10_vs_37694 | Shared | Duplodnaviria | Heunggongvirae | Uroviricota | Caudoviricetes | n.a. | Duneviridae | O | O | 99.94 | d__Bacteria;p__Bacteroidota;c__Bacteroidia;o__Bacteroidales;f__Rikenellaceae;g__Alistipes | 99.1 | iPHoP-RF;97.10 blast;96.80 RaFAH;87.90 CRISPR;55.60 |
| S11_vf_68693 | ME | n.a. | n.a. | n.a. | n.a. | n.a. | n.a. | n.a. | n.a. | 78.63 | d__Bacteria;p__Bacteroidota;c__Bacteroidia;o__Bacteroidales;f__Tannerellaceae;g__Parabacteroides | 99.3 | CRISPR;97.70 blast;97.30 iPHoP-RF;91.80 RaFAH;87.20 |
| S11_vf_92391 | Shared | n.a. | n.a. | n.a. | n.a. | n.a. | n.a. | n.a. | n.a. | 5.33 | d__Bacteria;p__Firmicutes;c__Bacilli;o__Erysipelotrichales;f__Erysipelotrichaceae;g__Clostridium_AQ | 91 | CRISPR;93.40 |
| S10_vs_46819 | SHHC | Duplodnaviria | Heunggongvirae | Uroviricota | Caudoviricetes | n.a. | Unclassified | O | O | 98.03 | d__Bacteria;p__Firmicutes;c__Clostridia;o__Oscillospirales;f__Ruminococcaceae;g__Faecalibacterium | 99.4 | blast;96.80 RaFAH;92.90 iPHoP-RF;79.00 |
| S11_vf_70015 | Shared | Duplodnaviria | Heunggongvirae | Uroviricota | Caudoviricetes | n.a. | Unclassified | O | O | 50.35 | d__Bacteria;p__Bacteroidota;c__Bacteroidia;o__Bacteroidales;f__Rikenellaceae;g__Alistipes | 97.8 | blast;96.10 RaFAH;79.90 |
| S11_vf_95067 | ME | n.a. | n.a. | n.a. | n.a. | n.a. | n.a. | n.a. | n.a. | 39.9 | d__Bacteria;p__Firmicutes;c__Clostridia;o__Oscillospirales;f__Oscillospiraceae;g__Flavonifractor | 93.4 | blast;95.30 iPHoP-RF;58.20 |
| S11_vs_104041 | ME | Duplodnaviria | Heunggongvirae | Uroviricota | Caudoviricetes | n.a. | Unclassified | O | O | 16.5 | d__Bacteria;p__Cyanobacteria;c__Vampirovibrionia;o__Gastranaerophilales;f__Gastranaerophilaceae;g__CAG-306 | 97.2 | blast;98.00 iPHoP-RF;56.00 |
| S11_vs_104064 | Shared | Duplodnaviria | Heunggongvirae | Uroviricota | Caudoviricetes | n.a. | Unclassified | O | O | 99.03 | d__Bacteria;p__Firmicutes;c__Clostridia;o__Oscillospirales;f__Ruminococcaceae;g__Negativibacillus | 92.1 | blast;94.30 |
| S11_vs_11994 | ME | Duplodnaviria | Heunggongvirae | Uroviricota | Caudoviricetes | n.a. | Unclassified | O | O | 21.97 | d__Bacteria;p__Firmicutes;c__Clostridia;o__Oscillospirales;f__Ruminococcaceae;g__Faecalibacterium | 91.7 | RaFAH;94.00 |
| S11_vs_14882 | ME | n.a. | n.a. | n.a. | n.a. | n.a. | n.a. | n.a. | n.a. | 11.87 | d__Bacteria;p__Firmicutes;c__Bacilli;o__Erysipelotrichales;f__Erysipelotrichaceae;g__Holdemania | 93.1 | blast;95.10 CRISPR;54.60 |
| S11_vs_1937 | Shared | Duplodnaviria | Heunggongvirae | Uroviricota | Caudoviricetes | n.a. | Unclassified | O | O | 99.04 | d__Bacteria;p__Bacteroidota;c__Bacteroidia;o__Bacteroidales;f__Rikenellaceae;g__Alistipes | 100 | blast;98.60 RaFAH;96.60 iPHoP-RF;83.30 |
| S11_vf_104034 | Shared | Duplodnaviria | Heunggongvirae | Uroviricota | Caudoviricetes | n.a. | Unclassified | Unclassified | O | 21.95 | d__Bacteria;p__Firmicutes;c__Clostridia;o__Oscillospirales;f__Oscillospiraceae;g__ER4 | 90.3 | blast;92.80 iPHoP-RF;81.10 |
| S11_vs_19892 | ME | Duplodnaviria | Heunggongvirae | Uroviricota | Caudoviricetes | n.a. | Unclassified | Unclassified | Eponavirus | 45.95 | d__Bacteria;p__Firmicutes;c__Clostridia;o__Oscillospirales;f__Oscillospiraceae;g__Lawsonibacter | 90.6 | blast;93.00 iPHoP-RF;67.50 |
| S11_vs_23188 | ME | n.a. | n.a. | n.a. | n.a. | n.a. | n.a. | n.a. | n.a. | 66.52 | d__Bacteria;p__Bacteroidota;c__Bacteroidia;o__Bacteroidales;f__Bacteroidaceae;g__Bacteroides | 99.9 | iPHoP-RF;97.10 RaFAH;95.80 blast;93.80 CRISPR;83.30 |
| S11_vs_26767 | ME | Duplodnaviria | Heunggongvirae | Uroviricota | Caudoviricetes | n.a. | Unclassified | O | O | 40.78 | d__Bacteria;p__Firmicutes;c__Clostridia;o__Lachnospirales;f__Lachnospiraceae;g__Ventrimonas | 90.8 | blast;93.20 iPHoP-RF;72.20 |
| S11_vs_3277 | Shared | Duplodnaviria | Heunggongvirae | Uroviricota | Caudoviricetes | n.a. | Unclassified | Unclassified | Unclassified | 99.63 | d__Bacteria;p__Bacteroidota;c__Bacteroidia;o__Bacteroidales;f__Bacteroidaceae;g__Bacteroides | 99.9 | RaFAH;98.80 iPHoP-RF;83.60 blast;80.70 |
| S11_vs_34137 | ME | Duplodnaviria | Heunggongvirae | Uroviricota | Caudoviricetes | n.a. | Unclassified | O | O | 17.51 | d__Bacteria;p__Firmicutes;c__Clostridia;o__Lachnospirales;f__Lachnospiraceae;g__Blautia_A | 91.4 | blast;93.70 iPHoP-RF;83.10 |
| S11_vs_41107 | ME | Duplodnaviria | Heunggongvirae | Uroviricota | Caudoviricetes | n.a. | Unclassified | O | O | 98.96 | d__Bacteria;p__Firmicutes;c__Clostridia;o__Clostridiales;f__Clostridiaceae;g__Clostridium | 93.6 | RaFAH;95.50 |
| S11_vs_5077 | ME | Duplodnaviria | Heunggongvirae | Uroviricota | Caudoviricetes | n.a. | Peduoviridae | Unclassified | Quadragintavirus | 84.31 | d__Bacteria;p__Proteobacteria;c__Gammaproteobacteria;o__Enterobacterales;f__Enterobacteriaceae;g__Salmonella | 95.3 | blast;96.70 |
| S11_vs_52577 | ME | Duplodnaviria | Heunggongvirae | Uroviricota | Caudoviricetes | n.a. | Unclassified | Unclassified | Mushuvirus | 78.49 | d__Bacteria;p__Firmicutes;c__Clostridia;o__Oscillospirales;f__Oscillospiraceae;g__Dysosmobacter | 91 | blast;88.40 iPHoP-RF;73.50 RaFAH;71.60 |
| S11_vs_58930 | Shared | Duplodnaviria | Heunggongvirae | Uroviricota | Caudoviricetes | n.a. | Unclassified | Unclassified | Unclassified | 93.75 | d__Bacteria;p__Bacteroidota;c__Bacteroidia;o__Bacteroidales;f__Bacteroidaceae;g__Bacteroides | 98.1 | blast;92.60 RaFAH;90.60 iPHoP-RF;89.80 |
| S11_vs_6458 | Shared | n.a. | n.a. | n.a. | n.a. | n.a. | n.a. | n.a. | n.a. | 15.38 | d__Bacteria;p__Proteobacteria;c__Alphaproteobacteria;o__RF32;f__CAG-239;g__CAG-267 | 90.1 | blast;92.60 iPHoP-RF;81.90 |
| S11_vs_68151 | Shared | Duplodnaviria | Heunggongvirae | Uroviricota | Caudoviricetes | n.a. | Unclassified | O | O | 99.98 | d__Bacteria;p__Firmicutes;c__Clostridia;o__Lachnospirales;f__Lachnospiraceae;g__Mediterraneibacter | 95.3 | blast;96.70 |
| S11_vs_76784 | Shared | Duplodnaviria | Heunggongvirae | Uroviricota | Caudoviricetes | n.a. | Unclassified | O | O | 41.57 | d__Bacteria;p__Firmicutes;c__Clostridia;o__Oscillospirales;f__Ruminococcaceae;g__Ruthenibacterium | 93 | blast;95.00 iPHoP-RF;77.30 |
| S11_vs_81775 | ME | Duplodnaviria | Heunggongvirae | Uroviricota | Caudoviricetes | n.a. | Unclassified | Unclassified | Unclassified | 24.35 | d__Bacteria;p__Firmicutes;c__Clostridia;o__Eubacteriales;f__Eubacteriaceae;g__Eubacterium | 95 | CRISPR;96.50 blast;93.70 iPHoP-RF;81.40 |
| S11_vs_84627 | ME | n.a. | n.a. | n.a. | n.a. | n.a. | n.a. | n.a. | n.a. | 6.74 | d__Bacteria;p__Proteobacteria;c__Alphaproteobacteria;o__RF32;f__CAG-239;g__51-20 | 95.6 | blast;96.90 iPHoP-RF;89.50 CRISPR;76.50 |
| S11_vs_84848 | ME | Duplodnaviria | Heunggongvirae | Uroviricota | Caudoviricetes | n.a. | Unclassified | Unclassified | Unclassified | 95.91 | d__Bacteria;p__Bacteroidota;c__Bacteroidia;o__Bacteroidales;f__Bacteroidaceae;g__Bacteroides | 95.9 | blast;91.20 RaFAH;83.50 iPHoP-RF;57.90 |
| S11_vf_33117 | ME | n.a. | n.a. | n.a. | n.a. | n.a. | n.a. | n.a. | n.a. | 96.79 | d__Bacteria;p__Firmicutes;c__Clostridia;o__Oscillospirales;f__Oscillospiraceae;g__CAG-83 | 97.3 | RaFAH;98.10 |
| S11_vs_98193 | ME | Duplodnaviria | Heunggongvirae | Uroviricota | Caudoviricetes | n.a. | Unclassified | Unclassified | Unclassified | 80.02 | d__Bacteria;p__Bacteroidota;c__Bacteroidia;o__Bacteroidales;f__Rikenellaceae;g__Alistipes | 96.2 | iPHoP-RF;92.10 blast;92.00 RaFAH;82.70 |
| S12_vf_10887 | SHHC | n.a. | n.a. | n.a. | n.a. | n.a. | n.a. | n.a. | n.a. | 7.67 | d__Bacteria;p__Proteobacteria;c__Gammaproteobacteria;o__Enterobacterales;f__Pasteurellaceae;g__Glaesserella | 94.5 | iPHoP-RF;96.10 CRISPR;91.90 |
| S11_vf_34008 | ME | Duplodnaviria | Heunggongvirae | Uroviricota | Caudoviricetes | n.a. | Unclassified | O | O | 44.72 | d__Bacteria;p__Firmicutes;c__Clostridia;o__Lachnospirales;f__Lachnospiraceae;g__ | 93.1 | iPHoP-RF;95.10 blast;93.90 |
| S12_vf_173208 | Shared | n.a. | n.a. | n.a. | n.a. | n.a. | n.a. | n.a. | n.a. | 10.29 | d__Bacteria;p__Firmicutes;c__Clostridia;o__Oscillospirales;f__Oscillospiraceae;g__CAG-170 | 93 | blast;95.00 iPHoP-RF;81.40 |
| S12_vf_174374 | Shared | n.a. | n.a. | n.a. | n.a. | n.a. | n.a. | n.a. | n.a. | 94.2 | d__Bacteria;p__Bacteroidota;c__Bacteroidia;o__Bacteroidales;f__Rikenellaceae;g__Alistipes | 96.2 | CRISPR;97.30 iPHoP-RF;97.10 blast;96.60 |
| S12_vf_176437 | SHHC | Duplodnaviria | Heunggongvirae | Uroviricota | Caudoviricetes | n.a. | Salasmaviridae | O | O | 10.68 | d__Bacteria;p__Firmicutes;c__Clostridia;o__Lachnospirales;f__Lachnospiraceae;g__OF09-33XD | 95.3 | CRISPR;96.70 iPHoP-RF;82.20 |
| S12_vf_181024 | SHHC | Duplodnaviria | Heunggongvirae | Uroviricota | Caudoviricetes | n.a. | Unclassified | O | O | 98.48 | d__Bacteria;p__Firmicutes;c__Clostridia;o__Oscillospirales;f__Ruminococcaceae;g__Ruminococcus_D | 99.9 | CRISPR;97.30 RaFAH;97.20 blast;96.90 iPHoP-RF;95.70 |
| S12_vf_60224 | SHHC | Duplodnaviria | Heunggongvirae | Uroviricota | Caudoviricetes | n.a. | Unclassified | O | O | 82.87 | d__Bacteria;p__Bacteroidota;c__Bacteroidia;o__Bacteroidales;f__Bacteroidaceae;g__Bacteroides | 100 | RaFAH;97.10 iPHoP-RF;96.70 blast;80.90 |
| S11_vf_58587 | ME | n.a. | n.a. | n.a. | n.a. | n.a. | n.a. | n.a. | n.a. | 11.86 | d__Bacteria;p__Actinobacteriota;c__Actinomycetia;o__Actinomycetales;f__Bifidobacteriaceae;g__Bifidobacterium | 90.4 | blast;92.90 iPHoP-RF;65.60 |
| S12_vf_69136 | SHHC | Duplodnaviria | Heunggongvirae | Uroviricota | Caudoviricetes | n.a. | Unclassified | O | O | 51.25 | d__Bacteria;p__Bacteroidota;c__Bacteroidia;o__Bacteroidales;f__Bacteroidaceae;g__Prevotellamassilia | 98.4 | CRISPR;98.80 blast;82.30 iPHoP-RF;78.30 |
| S12_vf_79666 | Shared | n.a. | n.a. | n.a. | n.a. | n.a. | n.a. | n.a. | n.a. | 99.55 | d__Bacteria;p__Firmicutes;c__Clostridia;o__Oscillospirales;f__Acutalibacteraceae;g__UBA1417 | 91.7 | blast;94.00 iPHoP-RF;58.20 |
| S12_vs_121913 | SHHC | n.a. | n.a. | n.a. | n.a. | n.a. | n.a. | n.a. | n.a. | 82.73 | d__Bacteria;p__Bacteroidota;c__Bacteroidia;o__Bacteroidales;f__Rikenellaceae;g__Alistipes | 98.5 | blast;96.70 RaFAH;83.70 |
| S12_vs_123620 | SHHC | n.a. | n.a. | n.a. | n.a. | n.a. | n.a. | n.a. | n.a. | 10.35 | d__Bacteria;p__Firmicutes;c__Bacilli;o__RF39;f__UBA660;g__CAG-302 | 95.4 | blast;96.80 iPHoP-RF;64.80 |
| S12_vs_138261 | Shared | n.a. | n.a. | n.a. | n.a. | n.a. | n.a. | n.a. | n.a. | 70.17 | d__Bacteria;p__Bacteroidota;c__Bacteroidia;o__Bacteroidales;f__Tannerellaceae;g__Parabacteroides | 98.4 | blast;98.80 CRISPR;97.40 iPHoP-RF;96.10 |
| S12_vs_141337 | Shared | Duplodnaviria | Heunggongvirae | Uroviricota | Caudoviricetes | n.a. | Unclassified | Unclassified | Unclassified | 98.55 | d__Bacteria;p__Bacteroidota;c__Bacteroidia;o__Bacteroidales;f__Bacteroidaceae;g__Bacteroides | 100 | RaFAH;99.40 blast;92.10 iPHoP-RF;57.90 |
| S12_vs_151292 | Shared | Duplodnaviria | Heunggongvirae | Uroviricota | Caudoviricetes | n.a. | Unclassified | Unclassified | Unclassified | 99.81 | d__Bacteria;p__Bacteroidota;c__Bacteroidia;o__Bacteroidales;f__Bacteroidaceae;g__Bacteroides | 100 | RaFAH;98.90 iPHoP-RF;97.10 blast;95.90 CRISPR;68.40 |
| S12_vs_163990 | Shared | Duplodnaviria | Heunggongvirae | Uroviricota | Caudoviricetes | n.a. | Unclassified | O | O | 99.19 | d__Bacteria;p__Firmicutes;c__Clostridia;o__Lachnospirales;f__Lachnospiraceae;g__Dorea | 99.3 | RaFAH;99.40 |
| S12_vs_172379 | SHHC | n.a. | n.a. | n.a. | n.a. | n.a. | n.a. | n.a. | n.a. | 18.46 | d__Bacteria;p__Firmicutes;c__Bacilli;o__RF39;f__UBA660;g__CAG-877 | 90.8 | blast;93.20 |
| S11_vf_99751 | ME | n.a. | n.a. | n.a. | n.a. | n.a. | n.a. | n.a. | n.a. | 8.04 | d__Bacteria;p__Firmicutes;c__Clostridia;o__Oscillospirales;f__Oscillospiraceae;g__CAG-83 | 94.9 | blast;96.40 iPHoP-RF;57.10 |
| S12_vs_188820 | SHHC | n.a. | n.a. | n.a. | n.a. | n.a. | n.a. | n.a. | n.a. | 7.83 | d__Bacteria;p__Firmicutes;c__Clostridia;o__Oscillospirales;f__CAG-272;g__QALR01 | 93 | blast;95.00 iPHoP-RF;87.00 |
| S12_vs_188828 | SHHC | n.a. | n.a. | n.a. | n.a. | n.a. | n.a. | n.a. | n.a. | 8.93 | d__Bacteria;p__Firmicutes;c__Clostridia;o__Oscillospirales;f__UMGS1783;g__UMGS1783 | 92.3 | blast;94.50 |
| S12_vs_23796 | SHHC | Duplodnaviria | Heunggongvirae | Uroviricota | Caudoviricetes | n.a. | Unclassified | Unclassified | Unclassified | 72.14 | d__Bacteria;p__Bacteroidota;c__Bacteroidia;o__Bacteroidales;f__Rikenellaceae;g__Alistipes | 99.1 | iPHoP-RF;96.70 blast;96.60 RaFAH;89.00 |
| S12_vs_26336 | Shared | n.a. | n.a. | n.a. | n.a. | n.a. | n.a. | n.a. | n.a. | 100 | d__Bacteria;p__Bacteroidota;c__Bacteroidia;o__Bacteroidales;f__Bacteroidaceae;g__Bacteroides | 99.6 | RaFAH;99.60 |
| S12_vs_48045 | SHHC | Duplodnaviria | Heunggongvirae | Uroviricota | Caudoviricetes | n.a. | Unclassified | Unclassified | Unclassified | 24.79 | d__Bacteria;p__Firmicutes;c__Clostridia;o__Christensenellales;f__CAG-74;g__UBA11524 | 94.9 | blast;96.40 iPHoP-RF;53.80 |
| S12_vs_6836 | SHHC | Duplodnaviria | Heunggongvirae | Uroviricota | Caudoviricetes | n.a. | Unclassified | O | O | 43.21 | d__Bacteria;p__Firmicutes;c__Clostridia;o__TANB77;f__CAG-508;g__CAG-269 | 92.7 | blast;94.80 iPHoP-RF;65.30 |
| S12_vs_71525 | Shared | n.a. | n.a. | n.a. | n.a. | n.a. | n.a. | n.a. | n.a. | 30.6 | d__Bacteria;p__Bacteroidota;c__Bacteroidia;o__Bacteroidales;f__Marinifilaceae;g__Butyricimonas | 93.8 | blast;95.60 iPHoP-RF;67.30 |
| S11_vs_104042 | ME | n.a. | n.a. | n.a. | n.a. | n.a. | n.a. | n.a. | n.a. | 75.46 | d__Bacteria;p__Firmicutes;c__Clostridia;o__Lachnospirales;f__Lachnospiraceae;g__Eubacterium_I | 98.1 | blast;98.60 iPHoP-RF;84.80 |
| S12_vs_86928 | SHHC | Duplodnaviria | Heunggongvirae | Uroviricota | Caudoviricetes | n.a. | Unclassified | O | O | 18.68 | d__Bacteria;p__Verrucomicrobiota;c__Verrucomicrobiae;o__Verrucomicrobiales;f__Akkermansiaceae;g__Akkermansia | 100 | CRISPR;98.80 RaFAH;98.60 blast;96.90 iPHoP-RF;82.50 |
| S12_vs_92868 | Shared | Duplodnaviria | Heunggongvirae | Uroviricota | Caudoviricetes | n.a. | Unclassified | O | O | 65.38 | d__Bacteria;p__Firmicutes;c__Clostridia;o__Oscillospirales;f__Oscillospiraceae;g__Intestinimonas | 93 | blast;95.00 iPHoP-RF;58.70 |
| S12_vs_93357 | SHHC | Duplodnaviria | Heunggongvirae | Uroviricota | Caudoviricetes | n.a. | Unclassified | O | O | 40.56 | d__Bacteria;p__Proteobacteria;c__Gammaproteobacteria;o__Burkholderiales;f__Burkholderiaceae;g__Parasutterella | 97 | CRISPR;97.90 blast;96.90 iPHoP-RF;93.10 |
| S12_vs_99092 | SHHC | Duplodnaviria | Heunggongvirae | Uroviricota | Caudoviricetes | n.a. | Unclassified | O | O | 78.56 | d__Bacteria;p__Firmicutes;c__Clostridia;o__Lachnospirales;f__Lachnospiraceae;g__Lachnospira | 99.6 | CRISPR;98.80 blast;98.80 iPHoP-RF;94.40 RaFAH;84.60 |
| S13_vf_109235 | ME | n.a. | n.a. | n.a. | n.a. | n.a. | n.a. | n.a. | n.a. | 72.39 | d__Bacteria;p__Firmicutes;c__Clostridia;o__Oscillospirales;f__Oscillospiraceae;g__CAG-83 | 90.1 | blast;92.60 iPHoP-RF;68.90 |
| S13_vf_11246 | Shared | Duplodnaviria | Heunggongvirae | Uroviricota | Caudoviricetes | Crassvirales | Suoliviridae | O | O | 46.73 | d__Bacteria;p__Bacteroidota;c__Bacteroidia;o__Bacteroidales;f__Bacteroidaceae;g__Prevotella | 91.8 | RaFAH;94.10 |
| S13_vf_143422 | Shared | Duplodnaviria | Heunggongvirae | Uroviricota | Caudoviricetes | Crassvirales | Suoliviridae | Boorivirinae | Canhaevirus | 75.96 | d__Bacteria;p__Bacteroidota;c__Bacteroidia;o__Bacteroidales;f__Tannerellaceae;g__Parabacteroides | 98.1 | blast;98.60 iPHoP-RF;83.30 |
| S13_vf_150407 | ME | Monodnaviria | Loebvirae | Hofneiviricota | Faserviricetes | Tubulavirales | Inoviridae | Unclassified | Inovirus | | d__Bacteria;p__Firmicutes;c__Clostridia;o__Oscillospirales;f__Oscillospiraceae;g__ER4 | 93.5 | CRISPR;95.40 |
| S13_vf_151045 | Shared | Duplodnaviria | Heunggongvirae | Uroviricota | Caudoviricetes | n.a. | Unclassified | O | O | 98.31 | d__Bacteria;p__Proteobacteria;c__Gammaproteobacteria;o__Enterobacterales;f__Enterobacteriaceae;g__Escherichia | 95.7 | RaFAH;97.00 |
| S11_vs_13652 | ME | Duplodnaviria | Heunggongvirae | Uroviricota | Caudoviricetes | n.a. | Unclassified | Unclassified | O | 57.92 | d__Bacteria;p__Firmicutes;c__Clostridia;o__Oscillospirales;f__Ruminococcaceae;g__Massilioclostridium | 92.9 | blast;94.90 iPHoP-RF;66.50 |
| S13_vf_28309 | ME | Duplodnaviria | Heunggongvirae | Uroviricota | Caudoviricetes | n.a. | Salasmaviridae | O | O | 12.14 | d__Bacteria;p__Firmicutes;c__Clostridia;o__Oscillospirales;f__Acutalibacteraceae;g__Ruminococcus_E | 90.1 | blast;92.60 |
| S13_vf_56074 | ME | Duplodnaviria | Heunggongvirae | Uroviricota | Caudoviricetes | n.a. | Unclassified | Unclassified | Unclassified | 91.56 | d__Bacteria;p__Bacteroidota;c__Bacteroidia;o__Bacteroidales;f__Rikenellaceae;g__Alistipes | 98.1 | RaFAH;98.60 |
| S13_vf_7670 | ME | n.a. | n.a. | n.a. | n.a. | n.a. | n.a. | n.a. | n.a. | 6.84 | d__Bacteria;p__Verrucomicrobiota;c__Verrucomicrobiae;o__Verrucomicrobiales;f__Akkermansiaceae;g__Akkermansia | 100 | CRISPR;99.60 RaFAH;96.00 blast;93.60 iPHoP-RF;72.60 |

| | | | | | | | | | | | | | |
|---|---|---|---|---|---|---|---|---|---|---|---|---|---|
| S13_vf_87873 | Shared | Duplodnaviria | Heunggongvirae | Uroviricota | Caudoviricetes | n.a. | Unclassified | Unclassified | Unclassified | 97.58 | d__Bacteria;p__Bacteroidota;c__Bacteroidia;o__Bacteroidales;f__Tannerellaceae;g__Marseille-P3160 | 98.4 | CRISPR;98.80 iPHoP-RF;72.60 |
| S13_vs_113593 | ME | Duplodnaviria | Heunggongvirae | Uroviricota | Caudoviricetes | n.a. | Unclassified | Unclassified | Unclassified | 40.01 | d__Bacteria;p__Actinobacteriota;c__Actinomycetia;o__Actinomycetales;f__Bifidobacteriaceae;g__Bifidobacterium | 91.5 | blast;93.80 iPHoP-RF;88.20 CRISPR;56.60 |
| S13_vs_121307 | ME | n.a. | n.a. | n.a. | n.a. | n.a. | n.a. | n.a. | n.a. | 11.04 | d__Bacteria;p__Firmicutes;c__Clostridia;o__Lachnospirales;f__Lachnospiraceae;g__Blautia_A | 95.9 | CRISPR;89.80 RaFAH;85.80 iPHoP-RF;84.50 |
| S13_vs_148294 | ME | Duplodnaviria | Heunggongvirae | Uroviricota | Caudoviricetes | n.a. | Unclassified | O | O | 57.95 | d__Bacteria;p__Firmicutes;c__Clostridia;o__Oscillospirales;f__Oscillospiraceae;g__ | 92.3 | blast;94.50 iPHoP-RF;80.60 |
| S13_vs_15645 | Shared | Duplodnaviria | Heunggongvirae | Uroviricota | Caudoviricetes | n.a. | Unclassified | O | O | 92 | d__Bacteria;p__Bacteroidota;c__Bacteroidia;o__Bacteroidales;f__Tannerellaceae;g__Parabacteroides | 99.9 | CRISPR;99.20 iPHoP-RF;95.10 blast;93.30 RaFAH;90.50 |
| S11_vs_3006 | ME | Duplodnaviria | Heunggongvirae | Uroviricota | Caudoviricetes | n.a. | Unclassified | Unclassified | O | 44.36 | d__Bacteria;p__Firmicutes;c__Clostridia;o__Oscillospirales;f__Acutalibacteraceae;g__UMGS263 | 93.6 | blast;95.50 iPHoP-RF;67.00 |
| S13_vs_30101 | Shared | Duplodnaviria | Heunggongvirae | Uroviricota | Caudoviricetes | n.a. | Unclassified | O | O | 54.46 | d__Bacteria;p__Firmicutes;c__Bacilli;o__ML615J-28;f__CAG-313;g__CAG-313 | 95.6 | blast;96.90 iPHoP-RF;81.60 |
| S13_vs_26756 | Shared | n.a. | n.a. | n.a. | n.a. | n.a. | n.a. | n.a. | n.a. | 9.7 | d__Bacteria;p__Firmicutes;c__Bacilli;o__RF39;f__UBA660;g__CAG-460 | 94 | blast;95.80 iPHoP-RF;53.30 |
| S13_vs_35816 | Shared | Duplodnaviria | Heunggongvirae | Uroviricota | Caudoviricetes | n.a. | Unclassified | Unclassified | Lagaffevirus | 99.57 | d__Bacteria;p__Firmicutes;c__Clostridia;o__Oscillospirales;f__Oscillospiraceae;g__CAG-83 | 99.7 | RaFAH;99.00 blast;82.80 |
| S13_vs_67429 | Shared | n.a. | n.a. | n.a. | n.a. | n.a. | n.a. | n.a. | n.a. | 14.26 | d__Bacteria;p__Firmicutes;c__Bacilli;o__RF39;f__UBA660;g__CAG-460 | 94.7 | blast;96.30 iPHoP-RF;53.80 |
| S11_vs_36748 | ME | n.a. | n.a. | n.a. | n.a. | n.a. | n.a. | n.a. | n.a. | 49.79 | d__Bacteria;p__Bacteroidota;c__Bacteroidia;o__Bacteroidales;f__Rikenellaceae;g__Alistipes | 98.5 | CRISPR;98.90 blast;96.70 iPHoP-RF;91.80 |
| S13_vs_83447 | Shared | n.a. | n.a. | n.a. | n.a. | n.a. | n.a. | n.a. | n.a. | 97.87 | d__Bacteria;p__Bacteroidota;c__Bacteroidia;o__Bacteroidales;f__Rikenellaceae;g__Alistipes | 99.9 | blast;96.80 RaFAH;96.50 iPHoP-RF;82.80 CRISPR;82.60 |
| S13_vs_90967 | Shared | Monodnaviria | Sangervirae | Phixviricota | Malgrandaviricetes | Petitvirales | Microviridae | O | O | | d__Bacteria;p__Bacteroidota;c__Bacteroidia;o__Bacteroidales;f__Barnesiellaceae;g__Barnesiella | 90.3 | blast;92.80 iPHoP-RF;81.90 |
| S11_vs_38860 | ME | n.a. | n.a. | n.a. | n.a. | n.a. | n.a. | n.a. | n.a. | 39.67 | d__Bacteria;p__Firmicutes;c__Clostridia;o__Lachnospirales;f__Lachnospiraceae;g__Enterocloster | 95.2 | CRISPR;96.60 blast;95.90 iPHoP-RF;89.80 |
| S13_vs_9438 | Shared | n.a. | n.a. | n.a. | n.a. | n.a. | n.a. | n.a. | n.a. | 7.33 | d__Bacteria;p__Proteobacteria;c__Alphaproteobacteria;o__RF32;f__CAG-239;g__CAG-267 | 91.6 | blast;93.90 iPHoP-RF;83.30 |
| S14_vf_106156 | ME | Duplodnaviria | Heunggongvirae | Uroviricota | Caudoviricetes | n.a. | Unclassified | Unclassified | Unclassified | 21.19 | d__Bacteria;p__Firmicutes;c__Bacilli;o__Lactobacillales;f__Lactobacillaceae;g__Lactobacillus | 100 | RaFAH;98.90 CRISPR;96.80 iPHoP-RF;96.10 blast;95.50 |
| S14_vf_109545 | Shared | n.a. | n.a. | n.a. | n.a. | n.a. | n.a. | n.a. | n.a. | 12.74 | d__Bacteria;p__Firmicutes;c__Clostridia;o__Oscillospirales;f__Oscillospiraceae;g__Intestinimonas | 90.8 | blast;93.20 iPHoP-RF;79.50 |
| S14_vf_117150 | ME | Duplodnaviria | Heunggongvirae | Uroviricota | Caudoviricetes | n.a. | Unclassified | O | O | 99.65 | d__Bacteria;p__Firmicutes;c__Clostridia;o__Lachnospirales;f__Lachnospiraceae;g__Lachnospira | 99.9 | CRISPR;98.90 RaFAH;91.00 blast;87.20 iPHoP-RF;83.10 |
| S11_vs_45658 | Shared | Duplodnaviria | Heunggongvirae | Uroviricota | Caudoviricetes | n.a. | Unclassified | O | O | 40.35 | d__Bacteria;p__Bacteroidota;c__Bacteroidia;o__Bacteroidales;f__Bacteroidaceae;g__Bacteroides | 95 | RaFAH;96.20 iPHoP-RF;53.60 |
| S14_vf_117157 | Shared | n.a. | n.a. | n.a. | n.a. | n.a. | n.a. | n.a. | n.a. | 99.57 | d__Bacteria;p__Firmicutes;c__Bacilli;o__Lactobacillales;f__Streptococcaceae;g__Streptococcus | 100 | RaFAH;99.40 CRISPR;98.80 iPHoP-RF;94.40 blast;93.60 |
| S11_vs_48109 | ME | Duplodnaviria | Heunggongvirae | Uroviricota | Caudoviricetes | n.a. | Unclassified | O | O | 63.66 | d__Bacteria;p__Firmicutes;c__Clostridia;o__Oscillospirales;f__Acutalibacteraceae;g__An172 | 93.4 | blast;95.30 iPHoP-RF;74.60 |
| S14_vf_37500 | ME | Monodnaviria | Sangervirae | Phixviricota | Malgrandaviricetes | Petitvirales | Microviridae | Unclassified | Unclassified | 5.58 | d__Bacteria;p__Bacteroidota;c__Bacteroidia;o__Bacteroidales;f__Bacteroidaceae;g__Phocaeicola | 94.3 | CRISPR;96.00 iPHoP-RF;67.00 |
| S14_vf_56130 | ME | Duplodnaviria | Heunggongvirae | Uroviricota | Caudoviricetes | n.a. | Unclassified | O | O | 100 | d__Bacteria;p__Firmicutes;c__Clostridia;o__Clostridiales;f__Clostridiaceae;g__Clostridium | 98.8 | RaFAH;99.10 |
| S14_vf_58919 | ME | Duplodnaviria | Heunggongvirae | Uroviricota | Caudoviricetes | n.a. | Unclassified | Unclassified | Mushuvirus | 74.74 | d__Bacteria;p__Firmicutes;c__Clostridia;o__Lachnospirales;f__Lachnospiraceae;g__Roseburia | 95.3 | blast;90.40 iPHoP-RF;87.90 RaFAH;82.70 |
| S14_vf_72335 | ME | Duplodnaviria | Heunggongvirae | Uroviricota | Caudoviricetes | n.a. | Unclassified | O | O | 11.73 | d__Bacteria;p__Firmicutes;c__Bacilli;o__Erysipelotrichales;f__Erysipelotrichaceae;g__Holdemania | 92.1 | CRISPR;94.30 |
| S14_vf_72953 | ME | Duplodnaviria | Heunggongvirae | Uroviricota | Caudoviricetes | n.a. | Unclassified | Unclassified | Unclassified | 32.12 | d__Bacteria;p__Firmicutes;c__Bacilli;o__Lactobacillales;f__Lactobacillaceae;g__Lacticaseibacillus | 96.7 | RaFAH;97.70 |
| S11_vs_55144 | ME | n.a. | n.a. | n.a. | n.a. | n.a. | n.a. | n.a. | n.a. | 12.8 | d__Bacteria;p__Firmicutes;c__Bacilli;o__Erysipelotrichales;f__Erysipelatoclostridiaceae;g__Massiliomicrobiota | 90.3 | blast;92.80 |
| S14_vf_78139 | ME | Duplodnaviria | Heunggongvirae | Uroviricota | Caudoviricetes | n.a. | Unclassified | Unclassified | Unclassified | 74.71 | d__Bacteria;p__Firmicutes;c__Bacilli;o__Lactobacillales;f__Lactobacillaceae;g__Lactobacillus | 100 | RaFAH;99.10 blast;96.80 CRISPR;96.50 iPHoP-RF;92.80 |
| S14_vf_92240 | Shared | Duplodnaviria | Heunggongvirae | Uroviricota | Caudoviricetes | n.a. | Unclassified | O | O | 99.79 | d__Bacteria;p__Firmicutes;c__Clostridia;o__Lachnospirales;f__Lachnospiraceae;g__Coprococcus | 93.2 | CRISPR;95.20 |
| S14_vf_99344 | Shared | n.a. | n.a. | n.a. | n.a. | n.a. | n.a. | n.a. | n.a. | 10.5 | d__Bacteria;p__Firmicutes;c__Clostridia;o__Lachnospirales;f__Lachnospiraceae;g__Eubacterium_F | 96.9 | CRISPR;97.80 |
| S14_vs_102698 | ME | Duplodnaviria | Heunggongvirae | Uroviricota | Caudoviricetes | n.a. | Unclassified | O | O | 99.62 | d__Bacteria;p__Firmicutes;c__Clostridia;o__Acetivibrionales;f__Acetivibrionaceae;g__Ruminiclostridium | 90.1 | RaFAH;92.60 |
| S14_vs_11510 | Shared | n.a. | n.a. | n.a. | n.a. | n.a. | n.a. | n.a. | n.a. | 39.46 | d__Bacteria;p__Bacteroidota;c__Bacteroidia;o__Bacteroidales;f__Bacteroidaceae;g__Bacteroides | 96.9 | blast;91.70 RaFAH;87.00 iPHoP-RF;81.40 |
| S14_vs_117166 | Shared | n.a. | n.a. | n.a. | n.a. | n.a. | n.a. | n.a. | n.a. | 22.09 | d__Bacteria;p__Firmicutes;c__Clostridia;o__Oscillospirales;f__Oscillospiraceae;g__Evtepia | 94.2 | blast;95.90 iPHoP-RF;66.30 |
| S11_vs_67573 | ME | n.a. | n.a. | n.a. | n.a. | n.a. | n.a. | n.a. | n.a. | 11.32 | d__Bacteria;p__Firmicutes;c__Clostridia;o__Lachnospirales;f__Lachnospiraceae;g__Enterocloster | 92.9 | CRISPR;94.90 blast;92.10 iPHoP-RF;92.10 |
| S14_vs_14737 | ME | Duplodnaviria | Heunggongvirae | Uroviricota | Caudoviricetes | n.a. | Unclassified | O | O | 33.52 | d__Bacteria;p__Firmicutes;c__Clostridia;o__Oscillospirales;f__Ruminococcaceae;g__Ruthenibacterium | 93.1 | blast;95.10 iPHoP-RF;68.40 |
| S14_vs_15228 | Shared | Duplodnaviria | Heunggongvirae | Uroviricota | Caudoviricetes | n.a. | Unclassified | O | O | 86.47 | d__Bacteria;p__Bacteroidota;c__Bacteroidia;o__Bacteroidales;f__Tannerellaceae;g__Parabacteroides | 100 | CRISPR;99.00 blast;98.60 RaFAH;97.90 iPHoP-RF;96.10 |
| S11_vs_68348 | ME | Duplodnaviria | Heunggongvirae | Uroviricota | Caudoviricetes | n.a. | Unclassified | O | O | 49.51 | d__Bacteria;p__Firmicutes;c__Bacilli;o__Erysipelotrichales;f__Erysipelatoclostridiaceae;g__Erysipelatoclostridium | 95.2 | blast;96.60 iPHoP-RF;64.50 |
| S14_vs_25852 | Shared | n.a. | n.a. | n.a. | n.a. | n.a. | n.a. | n.a. | n.a. | 26.78 | d__Bacteria;p__Bacteroidota;c__Bacteroidia;o__Bacteroidales;f__Bacteroidaceae;g__Bacteroides | 97.8 | blast;96.60 iPHoP-RF;95.10 CRISPR;93.00 RaFAH;77.10 |
| S11_vs_6878 | ME | Duplodnaviria | Heunggongvirae | Uroviricota | Caudoviricetes | n.a. | Unclassified | O | O | 72.71 | d__Bacteria;p__Firmicutes;c__Clostridia;o__Lachnospirales;f__Lachnospiraceae;g__ | 91 | blast;93.40 iPHoP-RF;73.00 |
| S14_vs_2870 | ME | Duplodnaviria | Heunggongvirae | Uroviricota | Caudoviricetes | n.a. | Unclassified | O | O | 56.13 | d__Bacteria;p__Proteobacteria;c__Gammaproteobacteria;o__Burkholderiales;f__Burkholderiaceae;g__Sutterella | 99 | blast;96.90 RaFAH;87.10 CRISPR;78.30 iPHoP-RF;69.50 |
| S14_vs_32499 | Shared | Duplodnaviria | Heunggongvirae | Uroviricota | Caudoviricetes | n.a. | Unclassified | O | O | 51.73 | d__Bacteria;p__Bacteroidota;c__Bacteroidia;o__Bacteroidales;f__Bacteroidaceae;g__Bacteroides | 99.1 | blast;98.10 iPHoP-RF;96.70 RaFAH;82.60 |
| S14_vs_3251 | ME | n.a. | n.a. | n.a. | n.a. | n.a. | n.a. | n.a. | n.a. | 82.87 | d__Bacteria;p__Firmicutes;c__Clostridia;o__Oscillospirales;f__Oscillospiraceae;g__Flavonifractor | 95.6 | blast;96.90 iPHoP-RF;65.60 |
| S14_vs_39797 | ME | Duplodnaviria | Heunggongvirae | Uroviricota | Caudoviricetes | n.a. | Unclassified | O | O | 47.42 | d__Bacteria;p__Firmicutes;c__Clostridia;o__Lachnospirales;f__Lachnospiraceae;g__Anaerostipes | 93.8 | blast;95.60 iPHoP-RF;67.00 |
| S11_vs_79831 | ME | n.a. | n.a. | n.a. | n.a. | n.a. | n.a. | n.a. | n.a. | 17.3 | d__Bacteria;p__Bacteroidota;c__Bacteroidia;o__Bacteroidales;f__UBA11471;g__UBA11471 | 90 | blast;92.50 iPHoP-RF;73.20 |
| S14_vs_59347 | ME | Duplodnaviria | Heunggongvirae | Uroviricota | Caudoviricetes | n.a. | Unclassified | O | O | 29.43 | d__Bacteria;p__Firmicutes;c__Clostridia;o__Lachnospirales;f__Lachnospiraceae;g__Muricomes | 93.1 | iPHoP-RF;95.10 |
| S14_vs_61530 | ME | n.a. | n.a. | n.a. | n.a. | n.a. | n.a. | n.a. | n.a. | 22.32 | d__Bacteria;p__Firmicutes;c__Clostridia;o__Oscillospirales;f__Acutalibacteraceae;g__UBA1417 | 92 | blast;94.20 iPHoP-RF;82.80 |
| S11_vs_82331 | ME | Duplodnaviria | Heunggongvirae | Uroviricota | Caudoviricetes | n.a. | Unclassified | O | O | 98.94 | d__Bacteria;p__Cyanobacteria;c__Vampirovibrionia;o__Gastranaerophilales;f__Gastranaerophilaceae;g__QAMI01 | 93.1 | blast;95.10 |
| S14_vs_632 | ME | Duplodnaviria | Heunggongvirae | Uroviricota | Caudoviricetes | n.a. | Unclassified | O | O | 78.66 | d__Bacteria;p__Firmicutes;c__Clostridia;o__Clostridiales;f__Clostridiaceae;g__Clostridium | 91 | RaFAH;93.40 |
| S11_vs_82368 | ME | Duplodnaviria | Heunggongvirae | Uroviricota | Caudoviricetes | n.a. | Unclassified | O | O | 62.18 | d__Bacteria;p__Firmicutes;c__Clostridia;o__Lachnospirales;f__Lachnospiraceae;g__Blautia_A | 98.1 | blast;92.60 RaFAH;90.70 iPHoP-RF;86.00 |
| S14_vs_66139 | Shared | Duplodnaviria | Heunggongvirae | Uroviricota | Caudoviricetes | n.a. | Unclassified | O | O | 83.75 | d__Bacteria;p__Firmicutes;c__Clostridia;o__Oscillospirales;f__Acutalibacteraceae;g__Ruminococcus_E | 95.6 | blast;96.90 iPHoP-RF;85.70 |
| S11_vs_83666 | ME | Duplodnaviria | Heunggongvirae | Uroviricota | Caudoviricetes | n.a. | Unclassified | O | O | 35.28 | d__Bacteria;p__Firmicutes;c__Clostridia;o__Lachnospirales;f__Lachnospiraceae;g__Dorea_A | 90.9 | blast;93.30 iPHoP-RF;67.70 |
| S14_vs_66208 | Shared | Duplodnaviria | Heunggongvirae | Uroviricota | Caudoviricetes | n.a. | Unclassified | O | O | 100 | d__Bacteria;p__Bacteroidota;c__Bacteroidia;o__Bacteroidales;f__Bacteroidaceae;g__Bacteroides | 100 | RaFAH;98.30 iPHoP-RF;96.40 blast;93.70 |
| S14_vs_66841 | Shared | Duplodnaviria | Heunggongvirae | Uroviricota | Caudoviricetes | n.a. | Unclassified | O | O | 15.26 | d__Bacteria;p__Firmicutes;c__Clostridia;o__Lachnospirales;f__Lachnospiraceae;g__Agathobacter | 90.4 | blast;92.90 iPHoP-RF;78.00 |
| S14_vs_67094 | ME | n.a. | n.a. | n.a. | n.a. | n.a. | n.a. | n.a. | n.a. | 10 | d__Bacteria;p__Firmicutes;c__Clostridia;o__Oscillospirales;f__Oscillospiraceae;g__Lawsonibacter | 90.7 | blast;93.10 |
| S14_vs_73301 | ME | Duplodnaviria | Heunggongvirae | Uroviricota | Caudoviricetes | n.a. | Unclassified | O | O | 15.25 | d__Bacteria;p__Firmicutes;c__Clostridia;o__Oscillospirales;f__Ruminococcaceae;g__Massilimaliae | 94.2 | blast;95.90 iPHoP-RF;64.80 |
| S14_vs_78164 | ME | Duplodnaviria | Heunggongvirae | Uroviricota | Caudoviricetes | n.a. | Winoviridae | O | O | 61.84 | d__Bacteria;p__Bacteroidota;c__Bacteroidia;o__Bacteroidales;f__Marinifilaceae;g__Odoribacter | 95.4 | blast;96.80 iPHoP-RF;84.80 CRISPR;56.80 |
| S14_vs_88666 | ME | Duplodnaviria | Heunggongvirae | Uroviricota | Caudoviricetes | n.a. | Unclassified | O | O | 32.05 | d__Bacteria;p__Firmicutes;c__Clostridia;o__Oscillospirales;f__Acutalibacteraceae;g__Fimivicinus | 95.4 | blast;96.80 iPHoP-RF;86.70 |
| S14_vs_9676 | ME | Duplodnaviria | Heunggongvirae | Uroviricota | Caudoviricetes | n.a. | Unclassified | O | O | 24.59 | d__Bacteria;p__Firmicutes;c__Clostridia;o__Oscillospirales;f__Oscillospiraceae;g__Lawsonibacter | 90.7 | blast;93.10 iPHoP-RF;77.50 |
| S12_vf_105542 | SHHC | Duplodnaviria | Heunggongvirae | Uroviricota | Caudoviricetes | n.a. | Unclassified | O | O | 13.83 | d__Bacteria;p__Firmicutes;c__Bacilli;o__RF39;f__UBA660;g__CAG-460 | 94.6 | blast;96.20 iPHoP-RF;73.70 |
| S15_vf_2032 | Shared | Duplodnaviria | Heunggongvirae | Uroviricota | Caudoviricetes | n.a. | Unclassified | O | O | 83.93 | d__Bacteria;p__Bacteroidota;c__Bacteroidia;o__Bacteroidales;f__Bacteroidaceae;g__Bacteroides | 99.3 | iPHoP-RF;97.10 blast;96.20 CRISPR;91.10 RaFAH;89.30 |
| S15_vf_20644 | SHHC | Duplodnaviria | Heunggongvirae | Uroviricota | Caudoviricetes | n.a. | Unclassified | Unclassified | Unclassified | 83.74 | d__Bacteria;p__Bacteroidota;c__Bacteroidia;o__Bacteroidales;f__Rikenellaceae;g__Alistipes | 94.3 | RaFAH;96.00 |
| S15_vf_2513 | SHHC | n.a. | n.a. | n.a. | n.a. | n.a. | n.a. | n.a. | n.a. | 25.17 | d__Bacteria;p__Firmicutes;c__Clostridia;o__Lachnospirales;f__Lachnospiraceae;g__Lachnospira | 94.5 | CRISPR;96.10 |
| S12_vf_145044 | Shared | n.a. | n.a. | n.a. | n.a. | n.a. | n.a. | n.a. | n.a. | 32.93 | d__Bacteria;p__Bacteroidota;c__Bacteroidia;o__Bacteroidales;f__Rikenellaceae;g__Alistipes_A | 91.6 | blast;93.90 iPHoP-RF;77.10 |
| S14_vs_70985 | Shared | n.a. | n.a. | n.a. | n.a. | n.a. | n.a. | n.a. | n.a. | 21.93 | d__Bacteria;p__Bacteroidota;c__Bacteroidia;o__Bacteroidales;f__Bacteroidaceae;g__Bacteroides | 99 | iPHoP-RF;96.70 blast;92.60 RaFAH;88.40 |
| S12_vf_16374 | Shared | Duplodnaviria | Heunggongvirae | Uroviricota | Caudoviricetes | n.a. | Unclassified | O | O | 100 | d__Bacteria;p__Bacteroidota;c__Bacteroidia;o__Bacteroidales;f__Bacteroidaceae;g__Bacteroides | 100 | RaFAH;99.10 iPHoP-RF;97.10 blast;94.80 |
| S15_vs_18274 | SHHC | n.a. | n.a. | n.a. | n.a. | n.a. | n.a. | n.a. | n.a. | 75.03 | d__Bacteria;p__Bacteroidota;c__Bacteroidia;o__Bacteroidales;f__Coprobacteraceae;g__Coprobacter | 98.7 | CRISPR;99.00 blast;93.10 iPHoP-RF;64.00 RaFAH;50.00 |
| S14_vs_74067 | Shared | n.a. | n.a. | n.a. | n.a. | n.a. | n.a. | n.a. | n.a. | 100 | d__Bacteria;p__Bacteroidota;c__Bacteroidia;o__Bacteroidales;f__Bacteroidaceae;g__Bacteroides | 99.6 | RaFAH;93.20 blast;92.00 iPHoP-RF;82.20 |
| S12_vf_170614 | SHHC | Duplodnaviria | Heunggongvirae | Uroviricota | Caudoviricetes | n.a. | Salasmaviridae | O | O | 7.03 | d__Bacteria;p__Actinobacteriota;c__Coriobacteriia;o__Coriobacteriales;f__Coriobacteriaceae;g__Collinsella | 96.4 | CRISPR;97.50 iPHoP-RF;95.10 |
| S15_vs_27510 | Shared | n.a. | n.a. | n.a. | n.a. | n.a. | n.a. | n.a. | n.a. | 51.41 | d__Bacteria;p__Bacteroidota;c__Bacteroidia;o__Bacteroidales;f__Bacteroidaceae;g__Bacteroides | 99.1 | iPHoP-RF;96.10 blast;93.90 RaFAH;90.50 |
| S15_vs_35445 | Shared | n.a. | n.a. | n.a. | n.a. | n.a. | n.a. | n.a. | n.a. | 27.45 | d__Bacteria;p__Bacteroidota;c__Bacteroidia;o__Bacteroidales;f__Bacteroidaceae;g__Bacteroides | 98.8 | iPHoP-RF;96.70 blast;93.90 RaFAH;86.60 |
| S12_vf_173666 | Shared | n.a. | n.a. | n.a. | n.a. | n.a. | n.a. | n.a. | n.a. | 36.83 | d__Bacteria;p__Actinobacteriota;c__Coriobacteriia;o__Coriobacteriales;f__Eggerthellaceae;g__Adlercreutzia | 93.1 | blast;95.10 iPHoP-RF;76.40 |
| S15_vs_71968 | Shared | Duplodnaviria | Heunggongvirae | Uroviricota | Caudoviricetes | n.a. | Unclassified | O | O | 100 | d__Bacteria;p__Bacteroidota;c__Bacteroidia;o__Bacteroidales;f__Rikenellaceae;g__Alistipes | 100 | RaFAH;98.90 blast;98.40 iPHoP-RF;95.40 |
| S16_vf_1037 | SHHC | n.a. | n.a. | n.a. | n.a. | n.a. | n.a. | n.a. | n.a. | 73.36 | d__Bacteria;p__Firmicutes;c__Clostridia;o__Lachnospirales;f__Lachnospiraceae;g__Lachnospira | 99.7 | CRISPR;99.70 iPHoP-RF;94.40 blast;93.60 |
| S15_vf_75924 | Shared | Duplodnaviria | Heunggongvirae | Uroviricota | Caudoviricetes | Crassvirales | Steigviridae | O | O | 99.22 | d__Bacteria;p__Bacteroidota;c__Bacteroidia;o__Bacteroidales;f__Bacteroidaceae;g__Bacteroides | 99.6 | RaFAH;93.20 blast;91.90 iPHoP-RF;82.50 |
| S16_vf_169895 | Shared | Duplodnaviria | Heunggongvirae | Uroviricota | Caudoviricetes | n.a. | Salasmaviridae | O | O | 11.84 | d__Bacteria;p__Firmicutes;c__Clostridia;o__Oscillospirales;f__Ruminococcaceae;g__Ruthenibacterium | 92.7 | CRISPR;94.80 |
| S16_vf_173109 | SHHC | Duplodnaviria | Heunggongvirae | Uroviricota | Caudoviricetes | n.a. | Unclassified | Unclassified | Unclassified | 7.33 | d__Bacteria;p__Actinobacteriota;c__Actinomycetia;o__Actinomycetales;f__Bifidobacteriaceae;g__Bifidobacterium | 97 | CRISPR;97.90 iPHoP-RF;81.60 |
| S16_vf_36146 | Shared | Duplodnaviria | Heunggongvirae | Uroviricota | Caudoviricetes | n.a. | Unclassified | O | O | 99.74 | d__Bacteria;p__Firmicutes;c__Clostridia;o__Clostridiales;f__Clostridiaceae;g__Clostridium | 96.2 | RaFAH;97.30 |
| S15_vs_42043 | Shared | n.a. | n.a. | n.a. | n.a. | n.a. | n.a. | n.a. | n.a. | 51.36 | d__Bacteria;p__Bacteroidota;c__Bacteroidia;o__Bacteroidales;f__Bacteroidaceae;g__Bacteroides | 97.6 | blast;96.80 iPHoP-RF;95.10 RaFAH;72.80 |
| S16_vf_6102 | Shared | Duplodnaviria | Heunggongvirae | Uroviricota | Caudoviricetes | n.a. | Unclassified | Unclassified | Toutatisvirus | 38.14 | d__Bacteria;p__Firmicutes;c__Clostridia;o__Oscillospirales;f__Oscillospiraceae;g__ER4 | 91.1 | blast;93.50 iPHoP-RF;66.80 |
| S16_vf_87190 | SHHC | Duplodnaviria | Heunggongvirae | Uroviricota | Caudoviricetes | n.a. | Unclassified | O | O | 34.33 | d__Bacteria;p__Bacteroidota;c__Bacteroidia;o__Bacteroidales;f__Bacteroidaceae;g__Bacteroides | 95 | RaFAH;87.80 iPHoP-RF;77.80 CRISPR;68.10 |
| S16_vf_90328 | SHHC | Duplodnaviria | Heunggongvirae | Uroviricota | Caudoviricetes | n.a. | Unclassified | O | O | 16.6 | d__Bacteria;p__Firmicutes;c__Clostridia;o__TANB77;f__CAG-508;g__CAG-269 | 91.4 | blast;93.70 iPHoP-RF;53.00 |
| S16_vf_97279 | Shared | Duplodnaviria | Heunggongvirae | Uroviricota | Caudoviricetes | n.a. | Unclassified | Unclassified | Eponavirus | 52.37 | d__Bacteria;p__Firmicutes;c__Clostridia;o__Oscillospirales;f__Oscillospiraceae;g__ER4 | 91 | blast;93.40 iPHoP-RF;60.30 |
| S12_vf_188846 | SHHC | Monodnaviria | Sangervirae | Phixviricota | Malgrandaviricetes | Petitvirales | Microviridae | O | O | | d__Bacteria;p__Bacteroidota;c__Bacteroidia;o__Bacteroidales;f__Barnesiellaceae;g__Barnesiella | 94.5 | blast;96.10 iPHoP-RF;68.60 |
| S16_vf_108554 | Shared | n.a. | n.a. | n.a. | n.a. | n.a. | n.a. | n.a. | n.a. | 18.88 | d__Bacteria;p__Firmicutes;c__Clostridia;o__Oscillospirales;f__Ruminococcaceae;g__Faecalibacterium | 97.9 | CRISPR;98.50 blast;93.10 iPHoP-RF;92.10 |
| S12_vf_18902 | Shared | Duplodnaviria | Heunggongvirae | Uroviricota | Caudoviricetes | n.a. | Unclassified | O | O | 80.1 | d__Bacteria;p__Firmicutes;c__Clostridia;o__Oscillospirales;f__Oscillospiraceae;g__ER4 | 90.6 | blast;93.00 iPHoP-RF;67.00 |
| S16_vs_103726 | SHHC | n.a. | n.a. | n.a. | n.a. | n.a. | n.a. | n.a. | n.a. | 26.95 | d__Bacteria;p__Firmicutes;c__Clostridia;o__Oscillospirales;f__Ruminococcaceae;g__Ruthenibacterium | 94 | blast;95.80 CRISPR;80.80 iPHoP-RF;79.80 |
| S16_vs_116956 | SHHC | n.a. | n.a. | n.a. | n.a. | n.a. | n.a. | n.a. | n.a. | 43.56 | d__Bacteria;p__Firmicutes;c__Clostridia;o__Lachnospirales;f__Lachnospiraceae;g__Acetatifactor | 91.6 | blast;93.90 iPHoP-RF;75.20 |
| S16_vs_120622 | SHHC | Duplodnaviria | Heunggongvirae | Uroviricota | Caudoviricetes | n.a. | Unclassified | O | O | 63.91 | d__Bacteria;p__Proteobacteria;c__Gammaproteobacteria;o__Burkholderiales;f__Burkholderiaceae;g__Parasutterella | 97.2 | CRISPR;98.00 blast;96.90 iPHoP-RF;93.10 |
| S16_vf_173099 | SHHC | n.a. | n.a. | n.a. | n.a. | n.a. | n.a. | n.a. | n.a. | 55.52 | d__Bacteria;p__Firmicutes;c__Clostridia;o__Oscillospirales;f__Oscillospiraceae;g__Flavonifractor | 93.8 | blast;92.00 iPHoP-RF;86.00 RaFAH;72.50 |
| S16_vs_127669 | Shared | Duplodnaviria | Heunggongvirae | Uroviricota | Caudoviricetes | n.a. | Unclassified | Unclassified | Unclassified | 99.82 | d__Bacteria;p__Bacteroidota;c__Bacteroidia;o__Bacteroidales;f__Bacteroidaceae;g__Bacteroides | 100 | RaFAH;99.00 CRISPR;97.60 iPHoP-RF;97.10 blast;90.90 |
| S12_vf_61299 | SHHC | Duplodnaviria | Heunggongvirae | Uroviricota | Caudoviricetes | n.a. | Salasmaviridae | O | O | 11.99 | d__Bacteria;p__Firmicutes;c__Clostridia;o__Lachnospirales;f__Lachnospiraceae;g__Muricomes | 96.9 | CRISPR;97.80 iPHoP-RF;59.20 |
| S16_vs_127905 | Shared | n.a. | n.a. | n.a. | n.a. | n.a. | n.a. | n.a. | n.a. | 55.65 | d__Bacteria;p__Bacteroidota;c__Bacteroidia;o__Bacteroidales;f__Bacteroidaceae;g__Bacteroides | 99.4 | iPHoP-RF;97.10 blast;95.20 RaFAH;91.80 CRISPR;60.90 |
| S16_vs_136935 | Shared | n.a. | n.a. | n.a. | n.a. | n.a. | n.a. | n.a. | n.a. | 99.58 | d__Bacteria;p__Bacteroidota;c__Bacteroidia;o__Bacteroidales;f__Bacteroidaceae;g__Bacteroides | 100 | RaFAH;98.20 iPHoP-RF;96.70 blast;89.50 |
| S16_vs_142561 | Shared | Duplodnaviria | Heunggongvirae | Uroviricota | Caudoviricetes | n.a. | Unclassified | O | O | 99.95 | d__Bacteria;p__Bacteroidota;c__Bacteroidia;o__Bacteroidales;f__Rikenellaceae;g__Alistipes_A | 97.8 | blast;98.20 RaFAH;56.20 iPHoP-RF;53.30 |
| S16_vs_173079 | SHHC | Duplodnaviria | Heunggongvirae | Uroviricota | Caudoviricetes | n.a. | Unclassified | O | O | 99.1 | d__Bacteria;p__Firmicutes;c__Clostridia;o__Lachnospirales;f__Lachnospiraceae;g__Coprococcus | 94 | blast;95.80 iPHoP-RF;71.70 |
| S16_vs_173096 | Shared | Duplodnaviria | Heunggongvirae | Uroviricota | Caudoviricetes | n.a. | Unclassified | Unclassified | Unclassified | 74.56 | d__Bacteria;p__Verrucomicrobiota;c__Verrucomicrobiae;o__Verrucomicrobiales;f__Akkermansiaceae;g__Akkermansia | 100 | RaFAH;99.70 CRISPR;99.60 blast;96.80 iPHoP-RF;81.10 |
| S16_vs_18440 | SHHC | Duplodnaviria | Heunggongvirae | Uroviricota | Caudoviricetes | n.a. | Unclassified | Unclassified | Toutatisvirus | 67.51 | d__Bacteria;p__Firmicutes;c__Clostridia;o__Oscillospirales;f__Ruminococcaceae;g__Faecalibacterium | 100 | RaFAH;99.30 CRISPR;98.90 blast;93.60 iPHoP-RF;85.70 |
| S12_vs_111286 | SHHC | n.a. | n.a. | n.a. | n.a. | n.a. | n.a. | n.a. | n.a. | 44.45 | d__Bacteria;p__Bacteroidota;c__Bacteroidia;o__Bacteroidales;f__Muribaculaceae;g__UBA7173 | 91.6 | blast;93.90 iPHoP-RF;62.50 |
| S16_vf_96483 | Shared | n.a. | n.a. | n.a. | n.a. | n.a. | n.a. | n.a. | n.a. | 29.87 | d__Bacteria;p__Firmicutes;c__Clostridia;o__Lachnospirales;f__Lachnospiraceae;g__Blautia_A | 97.6 | iPHoP-RF;93.40 blast;93.30 RaFAH;87.30 |
| S16_vs_19968 | Shared | n.a. | n.a. | n.a. | n.a. | n.a. | n.a. | n.a. | n.a. | 93.01 | d__Bacteria;p__Bacteroidota;c__Bacteroidia;o__Bacteroidales;f__Rikenellaceae;g__Alistipes | 98.4 | iPHoP-RF;97.10 blast;96.90 RaFAH;79.60 |
| S16_vs_32764 | Shared | n.a. | n.a. | n.a. | n.a. | n.a. | n.a. | n.a. | n.a. | 17.71 | d__Bacteria;p__Bacteroidota;c__Bacteroidia;o__Bacteroidales;f__Rikenellaceae;g__Alistipes | 95.9 | iPHoP-RF;97.10 blast;96.90 |
| S16_vs_33281 | Shared | n.a. | n.a. | n.a. | n.a. | n.a. | n.a. | n.a. | n.a. | 7.09 | d__Bacteria;p__Bacteroidota;c__Bacteroidia;o__Bacteroidales;f__Rikenellaceae;g__Alistipes | 95.9 | iPHoP-RF;97.10 blast;96.90 |
| S16_vs_4662 | SHHC | Duplodnaviria | Heunggongvirae | Uroviricota | Caudoviricetes | n.a. | Unclassified | Unclassified | Unclassified | 13.68 | d__Bacteria;p__Firmicutes;c__Clostridia;o__Christensenellales;f__CAG-138;g__UBA5394 | 92.6 | CRISPR;94.70 blast;92.60 iPHoP-RF;74.30 |

| | | | | | | | | | | | | | |
|---|---|---|---|---|---|---|---|---|---|---|---|---|---|
| S12_vs_133070 | Shared | Duplodnaviria | Heunggongvirae | Uroviricota | Caudoviricetes | n.a. | Unclassified | O | O | 14.52 | d__Bacteria;p__Firmicutes;c__Clostridia;o__Oscillospirales;f__Ruminococcaceae;g__CAG-353 | 91.1 | blast;93.50 iPHoP-RF;79.80 |
| S16_vs_48714 | SHHC | n.a. | n.a. | n.a. | n.a. | n.a. | n.a. | n.a. | n.a. | 5.9 | d__Bacteria;p__Firmicutes;c__Clostridia;o__Oscillospirales;f__Ruminococcaceae;g__Ruminococcus_D | 97 | blast;97.90 iPHoP-RF;97.10 CRISPR;96.60 |
| S16_vs_51901 | SHHC | n.a. | n.a. | n.a. | n.a. | n.a. | n.a. | n.a. | n.a. | 16.94 | d__Bacteria;p__Bacteroidota;c__Bacteroidia;o__Bacteroidales;f__Rikenellaceae;g__Alistipes | 97.6 | blast;93.90 iPHoP-RF;90.10 RaFAH;85.80 |
| S16_vs_75108 | Shared | n.a. | n.a. | n.a. | n.a. | n.a. | n.a. | n.a. | n.a. | 32.65 | d__Bacteria;p__Firmicutes;c__Clostridia;o__Oscillospirales;f__Oscillospiraceae;g__UBA5446 | 95 | blast;96.50 CRISPR;82.40 iPHoP-RF;65.30 |
| S16_vs_78361 | Shared | Duplodnaviria | Heunggongvirae | Uroviricota | Caudoviricetes | n.a. | Unclassified | O | O | 75.01 | d__Bacteria;p__Actinobacteriota;c__Actinomycetia;o__Actinomycetales;f__Bifidobacteriaceae;g__Bifidobacterium | 98.5 | CRISPR;98.90 iPHoP-RF;97.10 blast;96.90 |
| S12_vs_142625 | Shared | Duplodnaviria | Heunggongvirae | Uroviricota | Caudoviricetes | n.a. | Unclassified | O | O | 31.03 | d__Bacteria;p__Bacteroidota;c__Bacteroidia;o__Bacteroidales;f__Rikenellaceae;g__Alistipes | 95.9 | iPHoP-RF;97.10 blast;96.90 |
| S16_vs_80244 | Shared | n.a. | n.a. | n.a. | n.a. | n.a. | n.a. | n.a. | n.a. | 99.97 | d__Bacteria;p__Bacteroidota;c__Bacteroidia;o__Bacteroidales;f__Bacteroidaceae;g__Bacteroides | 99.7 | iPHoP-RF;97.10 RaFAH;95.20 blast;89.20 |
| S16_vs_15093 | SHHC | Duplodnaviria | Heunggongvirae | Uroviricota | Caudoviricetes | n.a. | Unclassified | O | O | 19.11 | d__Bacteria;p__Firmicutes;c__Clostridia;o__Christensenellales;f__CAG-74;g__UBA11524 | 99.6 | CRISPR;99.60 blast;96.60 iPHoP-RF;85.70 |
| S17_vf_174033 | Shared | Duplodnaviria | Heunggongvirae | Uroviricota | Caudoviricetes | n.a. | Unclassified | O | O | 68.79 | d__Bacteria;p__Firmicutes;c__Clostridia;o__Oscillospirales;f__Ruminococcaceae;g__Ruminococcus_D | 99.7 | blast;97.00 iPHoP-RF;95.40 RaFAH;94.60 CRISPR;72.00 |
| S17_vf_35138 | ME | Duplodnaviria | Heunggongvirae | Uroviricota | Caudoviricetes | n.a. | Unclassified | O | O | 84.47 | d__Bacteria;p__Bacteroidota;c__Bacteroidia;o__Bacteroidales;f__Barnesiellaceae;g__Barnesiella | 99.1 | blast;96.80 RaFAH;88.80 CRISPR;59.60 |
| S17_vf_81232 | Shared | Duplodnaviria | Heunggongvirae | Uroviricota | Caudoviricetes | Crassvirales | Suoliviridae | Boorivirinae | Canhaevirus | 50.04 | d__Bacteria;p__Bacteroidota;c__Bacteroidia;o__Bacteroidales;f__Tannerellaceae;g__Parabacteroides | 97.8 | blast;98.40 iPHoP-RF;95.10 CRISPR;83.70 |
| S12_vs_153780 | SHHC | Duplodnaviria | Heunggongvirae | Uroviricota | Caudoviricetes | n.a. | Unclassified | O | O | 97.5 | d__Bacteria;p__Proteobacteria;c__Alphaproteobacteria;o__RF32;f__CAG-239;g__CAG-495 | 90.9 | blast;93.30 iPHoP-RF;81.60 |
| S17_vf_97671 | ME | Duplodnaviria | Heunggongvirae | Uroviricota | Caudoviricetes | n.a. | Unclassified | Unclassified | Mushuvirus | 81.31 | d__Bacteria;p__Firmicutes;c__Clostridia;o__Oscillospirales;f__Oscillospiraceae;g__Lawsonibacter | 90.2 | blast;92.70 iPHoP-RF;73.20 |
| S17_vs_103009 | Shared | n.a. | n.a. | n.a. | n.a. | n.a. | n.a. | n.a. | n.a. | 22.09 | d__Bacteria;p__Firmicutes;c__Clostridia;o__Oscillospirales;f__Oscillospiraceae;g__ER4 | 96.7 | blast;97.70 iPHoP-RF;65.10 |
| S12_vs_164056 | Shared | n.a. | n.a. | n.a. | n.a. | n.a. | n.a. | n.a. | n.a. | 11.8 | d__Bacteria;p__Firmicutes;c__Clostridia;o__Oscillospirales;f__Oscillospiraceae;g__CAG-83 | 95.9 | blast;97.10 iPHoP-RF;72.60 |
| S17_vs_116640 | ME | Duplodnaviria | Heunggongvirae | Uroviricota | Caudoviricetes | n.a. | Unclassified | O | O | 83.12 | d__Bacteria;p__Bacteroidota;c__Bacteroidia;o__Bacteroidales;f__Bacteroidaceae;g__Bacteroides | 91 | RaFAH;93.40 |
| S17_vs_127519 | Shared | n.a. | n.a. | n.a. | n.a. | n.a. | n.a. | n.a. | n.a. | 61.22 | d__Bacteria;p__Firmicutes;c__Clostridia;o__Oscillospirales;f__Oscillospiraceae;g__Dysosmobacter | 91.2 | blast;93.60 iPHoP-RF;86.70 |
| S17_vs_160502 | ME | Duplodnaviria | Heunggongvirae | Uroviricota | Caudoviricetes | Crassvirales | Steigviridae | O | O | 60.44 | d__Bacteria;p__Bacteroidota;c__Bacteroidia;o__Bacteroidales;f__Bacteroidaceae;g__Bacteroides | 99.9 | iPHoP-RF;97.10 RaFAH;96.60 blast;92.60 CRISPR;56.20 |
| S17_vs_177196 | ME | n.a. | n.a. | n.a. | n.a. | n.a. | n.a. | n.a. | n.a. | 18.1 | d__Bacteria;p__Bacteroidota;c__Bacteroidia;o__Bacteroidales;f__Rikenellaceae;g__Alistipes | 96.4 | blast;91.90 iPHoP-RF;88.50 RaFAH;84.30 |
| S17_vs_2079 | Shared | Duplodnaviria | Heunggongvirae | Uroviricota | Caudoviricetes | n.a. | Unclassified | Unclassified | Unclassified | 67.97 | d__Bacteria;p__Bacteroidota;c__Bacteroidia;o__Bacteroidales;f__Bacteroidaceae;g__Bacteroides | 100 | RaFAH;98.90 iPHoP-RF;94.40 blast;92.60 |
| S17_vs_39515 | ME | Duplodnaviria | Heunggongvirae | Uroviricota | Caudoviricetes | Crassvirales | Intestiviridae | O | O | 82.52 | d__Bacteria;p__Bacteroidota;c__Bacteroidia;o__Bacteroidales;f__Bacteroidaceae;g__Bacteroides | 100 | RaFAH;99.30 iPHoP-RF;97.10 blast;92.90 |
| S17_vs_43307 | Shared | n.a. | n.a. | n.a. | n.a. | n.a. | n.a. | n.a. | n.a. | 72.14 | d__Bacteria;p__Bacteroidota;c__Bacteroidia;o__Bacteroidales;f__Tannerellaceae;g__Parabacteroides | 95.4 | blast;96.80 CRISPR;96.70 iPHoP-RF;93.40 |
| S1_vf_158961 | ME | Duplodnaviria | Heunggongvirae | Uroviricota | Caudoviricetes | n.a. | Unclassified | O | O | 9.7 | d__Bacteria;p__Firmicutes;c__Clostridia;o__Christensenellales;f__CAG-74;g__UBA11524 | 95 | blast;96.50 iPHoP-RF;77.30 |
| S1_vf_159951 | ME | Duplodnaviria | Heunggongvirae | Uroviricota | Caudoviricetes | n.a. | Unclassified | O | O | 22.14 | d__Bacteria;p__Bacteroidota;c__Bacteroidia;o__Bacteroidales;f__Rikenellaceae;g__Alistipes | 95.4 | CRISPR;96.80 iPHoP-RF;72.20 |
| S1_vf_22149 | ME | n.a. | n.a. | n.a. | n.a. | n.a. | n.a. | n.a. | n.a. | 43.65 | d__Bacteria;p__Firmicutes;c__Clostridia;o__Oscillospirales;f__Oscillospiraceae;g__ER4 | 90.8 | blast;93.20 iPHoP-RF;88.90 |
| S1_vs_143929 | ME | n.a. | n.a. | n.a. | n.a. | n.a. | n.a. | n.a. | n.a. | 42.31 | d__Bacteria;p__Firmicutes;c__Clostridia;o__Oscillospirales;f__Oscillospiraceae;g__ER4 | 91.6 | blast;93.90 iPHoP-RF;75.00 |
| S2_vf_20910 | SHHC | n.a. | n.a. | n.a. | n.a. | n.a. | n.a. | n.a. | n.a. | 5.26 | d__Bacteria;p__Firmicutes;c__Bacilli;o__Erysipelotrichales;f__Erysipelotrichaceae;g__Clostridium_AQ | 97.2 | CRISPR;98.00 |
| S12_vs_1980 | Shared | Duplodnaviria | Heunggongvirae | Uroviricota | Caudoviricetes | n.a. | Unclassified | O | O | 94.44 | d__Bacteria;p__Bacteroidota;c__Bacteroidia;o__Bacteroidales;f__Bacteroidaceae;g__Bacteroides | 99.1 | iPHoP-RF;97.10 blast;93.60 RaFAH;87.80 CRISPR;58.50 |
| S2_vs_10289 | Shared | Duplodnaviria | Heunggongvirae | Uroviricota | Caudoviricetes | n.a. | Unclassified | Unclassified | Unclassified | 67.97 | d__Bacteria;p__Bacteroidota;c__Bacteroidia;o__Bacteroidales;f__Bacteroidaceae;g__Bacteroides | 100 | RaFAH;98.90 blast;96.80 iPHoP-RF;87.90 |
| S2_vs_7306 | Shared | Duplodnaviria | Heunggongvirae | Uroviricota | Caudoviricetes | n.a. | Unclassified | Unclassified | Unclassified | 96.01 | d__Bacteria;p__Bacteroidota;c__Bacteroidia;o__Bacteroidales;f__Tannerellaceae;g__Parabacteroides | 93 | blast;95.00 iPHoP-RF;73.70 |
| S3_vf_100440 | ME | Duplodnaviria | Heunggongvirae | Uroviricota | Caudoviricetes | n.a. | Unclassified | O | O | 98.85 | d__Bacteria;p__Firmicutes;c__Clostridia;o__Clostridiales;f__Clostridiaceae;g__Clostridium | 94.7 | RaFAH;96.30 |
| S3_vf_106401 | ME | Duplodnaviria | Heunggongvirae | Uroviricota | Caudoviricetes | n.a. | Unclassified | O | O | 12.56 | d__Bacteria;p__Firmicutes;c__Bacilli;o__Haloplasmatales_A;f__Turicibacteraceae;g__Turicibacter | 92 | blast;94.20 iPHoP-RF;66.30 |
| S12_vs_26847 | SHHC | n.a. | n.a. | n.a. | n.a. | n.a. | n.a. | n.a. | n.a. | 68.45 | d__Bacteria;p__Firmicutes;c__Clostridia;o__Lachnospirales;f__Lachnospiraceae;g__Lachnospira | 99.3 | CRISPR;98.90 iPHoP-RF;95.70 blast;94.10 RaFAH;74.50 |
| S3_vf_115304 | Shared | Duplodnaviria | Heunggongvirae | Uroviricota | Caudoviricetes | Crassvirales | Suoliviridae | Uncouvirinae | Aurodevirus | 34.34 | d__Bacteria;p__Spirochaetota;c__Spirochaetia;o__Sphaerochaetales;f__Sphaerochaetaceae;g__UBA9732 | 91.2 | blast;93.60 iPHoP-RF;81.90 |
| S3_vf_127399 | Shared | Duplodnaviria | Heunggongvirae | Uroviricota | Caudoviricetes | n.a. | Unclassified | O | O | 23.09 | d__Bacteria;p__Bacteroidota;c__Bacteroidia;o__Bacteroidales;f__Bacteroidaceae;g__Bacteroides | 90.4 | RaFAH;92.90 |
| S12_vs_32669 | SHHC | n.a. | n.a. | n.a. | n.a. | n.a. | n.a. | n.a. | n.a. | 3.93 | d__Bacteria;p__Firmicutes;c__Clostridia;o__Christensenellales;f__CAG-917;g__CAG-475 | 94.7 | blast;96.30 iPHoP-RF;68.60 |
| S3_vf_127405 | ME | Duplodnaviria | Heunggongvirae | Uroviricota | Caudoviricetes | n.a. | Unclassified | O | O | 98.09 | d__Bacteria;p__Firmicutes;c__Clostridia;o__Lachnospirales;f__Lachnospiraceae;g__Lachnospira | 99.9 | CRISPR;98.90 RaFAH;91.60 iPHoP-RF;90.80 blast;80.40 |
| S3_vf_127406 | ME | Duplodnaviria | Heunggongvirae | Uroviricota | Caudoviricetes | n.a. | Unclassified | Unclassified | Unclassified | 89.48 | d__Bacteria;p__Bacteroidota;c__Bacteroidia;o__Bacteroidales;f__Rikenellaceae;g__Alistipes | 97.8 | RaFAH;98.40 |
| S3_vf_127409 | Shared | Duplodnaviria | Heunggongvirae | Uroviricota | Caudoviricetes | n.a. | Unclassified | O | O | 96.84 | d__Bacteria;p__Firmicutes;c__Clostridia;o__Oscillospirales;f__Ruminococcaceae;g__Faecalibacterium | 95.9 | CRISPR;97.00 blast;95.70 iPHoP-RF;81.10 RaFAH;51.90 |
| S3_vf_18080 | ME | n.a. | n.a. | n.a. | n.a. | n.a. | n.a. | n.a. | n.a. | 96.93 | d__Bacteria;p__Bacteroidota;c__Bacteroidia;o__Bacteroidales;f__Bacteroidaceae;g__Bacteroides | 99.7 | iPHoP-RF;97.10 blast;96.10 RaFAH;94.50 CRISPR;81.40 |
| S3_vf_18829 | ME | Duplodnaviria | Heunggongvirae | Uroviricota | Caudoviricetes | n.a. | Unclassified | O | O | 63.94 | d__Bacteria;p__Bacteroidota;c__Bacteroidia;o__Bacteroidales;f__Bacteroidaceae;g__Bacteroides | 91.8 | iPHoP-RF;94.10 CRISPR;73.10 |
| S3_vf_44405 | Shared | Duplodnaviria | Heunggongvirae | Uroviricota | Caudoviricetes | n.a. | Unclassified | Unclassified | Unclassified | 96.43 | d__Bacteria;p__Actinobacteriota;c__Actinomycetia;o__Actinomycetales;f__Actinomycetaceae;g__Actinomyces | 100 | RaFAH;97.30 CRISPR;92.40 blast;92.10 iPHoP-RF;55.70 |
| S3_vf_54353 | ME | Duplodnaviria | Heunggongvirae | Uroviricota | Caudoviricetes | n.a. | Unclassified | Unclassified | Cimpunavirus | 46.9 | d__Bacteria;p__Actinobacteriota;c__Actinomycetia;o__Actinomycetales;f__Actinomycetaceae;g__Actinomyces | 97.8 | RaFAH;97.30 iPHoP-RF;70.60 |
| S3_vf_67333 | ME | Duplodnaviria | Heunggongvirae | Uroviricota | Caudoviricetes | n.a. | Unclassified | O | O | 76.26 | d__Bacteria;p__Firmicutes;c__Clostridia;o__Lachnospirales;f__Lachnospiraceae;g__Agathobacter | 99.9 | CRISPR;98.90 RaFAH;90.80 blast;85.70 iPHoP-RF;81.90 |
| S12_vs_74280 | Shared | n.a. | n.a. | n.a. | n.a. | n.a. | n.a. | n.a. | n.a. | 14.1 | d__Bacteria;p__Bacteroidota;c__Bacteroidia;o__Bacteroidales;f__Marinifilaceae;g__Butyricimonas | 94.3 | blast;96.00 iPHoP-RF;58.40 |
| S3_vf_67828 | Shared | Monodnaviria | Sangervirae | Phixviricota | Malgrandaviricetes | Petitvirales | Microviridae | Unclassified | Unclassified | | d__Bacteria;p__Bacteroidota;c__Bacteroidia;o__Bacteroidales;f__Rikenellaceae;g__Alistipes | 91.5 | blast;93.80 iPHoP-RF;64.50 |
| S3_vf_108959 | ME | n.a. | n.a. | n.a. | n.a. | n.a. | n.a. | n.a. | n.a. | 42.95 | d__Bacteria;p__Firmicutes;c__Clostridia;o__Lachnospirales;f__Lachnospiraceae;g__Lachnospira | 99.3 | CRISPR;99.30 blast;91.90 iPHoP-RF;88.90 RaFAH;55.00 |
| S3_vf_70746 | Shared | Duplodnaviria | Heunggongvirae | Uroviricota | Caudoviricetes | n.a. | Unclassified | Unclassified | Unclassified | 87.64 | d__Bacteria;p__Actinobacteriota;c__Actinomycetia;o__Actinomycetales;f__Bifidobacteriaceae;g__Bifidobacterium | 95.3 | iPHoP-RF;96.70 blast;95.90 CRISPR;78.50 |
| S12_vs_81729 | SHHC | Duplodnaviria | Heunggongvirae | Uroviricota | Caudoviricetes | n.a. | Unclassified | O | O | 10.77 | d__Bacteria;p__Firmicutes;c__Clostridia;o__Oscillospirales;f__Ruminococcaceae;g__Faecalibacterium | 97.3 | CRISPR;98.10 iPHoP-RF;93.40 blast;92.10 |
| S3_vf_73597 | ME | Duplodnaviria | Heunggongvirae | Uroviricota | Caudoviricetes | n.a. | Unclassified | O | O | 55.3 | d__Bacteria;p__Bacteroidota;c__Bacteroidia;o__Bacteroidales;f__Bacteroidaceae;g__Bacteroides | 99.7 | CRISPR;97.20 iPHoP-RF;97.10 RaFAH;93.80 blast;93.80 |
| S3_vf_80287 | Shared | Duplodnaviria | Heunggongvirae | Uroviricota | Caudoviricetes | n.a. | Unclassified | O | O | 52.73 | d__Bacteria;p__Firmicutes;c__Clostridia;o__Oscillospirales;f__Ruminococcaceae;g__D5 | 97.2 | CRISPR;98.00 iPHoP-RF;52.20 |
| S3_vf_8058 | ME | Duplodnaviria | Heunggongvirae | Uroviricota | Caudoviricetes | n.a. | Unclassified | Unclassified | Eponavirus | 98.11 | d__Bacteria;p__Firmicutes;c__Clostridia;o__Lachnospirales;f__Lachnospiraceae;g__Blautia_A | 98.1 | CRISPR;98.60 iPHoP-RF;81.60 |
| S12_vs_87088 | Shared | Duplodnaviria | Heunggongvirae | Uroviricota | Caudoviricetes | n.a. | Unclassified | O | O | 36.55 | d__Bacteria;p__Firmicutes;c__Clostridia;o__Oscillospirales;f__Ruminococcaceae;g__Faecalibacterium | 97.5 | blast;96.60 iPHoP-RF;92.40 RaFAH;73.80 CRISPR;53.50 |
| S3_vf_82571 | Shared | Duplodnaviria | Heunggongvirae | Uroviricota | Caudoviricetes | n.a. | Unclassified | O | O | 21.78 | d__Bacteria;p__Firmicutes;c__Clostridia;o__Oscillospirales;f__Oscillospiraceae;g__CAG-170 | 93.1 | blast;95.10 iPHoP-RF;73.20 |
| S3_vf_99734 | Shared | Duplodnaviria | Heunggongvirae | Uroviricota | Caudoviricetes | n.a. | Unclassified | Unclassified | Lagaffevirus | 99.18 | d__Bacteria;p__Firmicutes;c__Clostridia;o__Oscillospirales;f__Ruminococcaceae;g__Faecalibacterium | 95.2 | CRISPR;96.60 iPHoP-RF;64.80 blast;60.50 |
| S3_vs_114271 | ME | n.a. | n.a. | n.a. | n.a. | n.a. | n.a. | n.a. | n.a. | 19.01 | d__Bacteria;p__Firmicutes;c__Clostridia;o__Oscillospirales;f__Acutalibacteraceae;g__Acutalibacter | 92.1 | blast;94.30 iPHoP-RF;56.80 |
| S12_vs_94042 | Shared | n.a. | n.a. | n.a. | n.a. | n.a. | n.a. | n.a. | n.a. | 10.3 | d__Bacteria;p__Firmicutes;c__Clostridia;o__Christensenellales;f__CAG-74;g__UBA11524 | 95.6 | blast;96.90 iPHoP-RF;91.80 |
| S3_vs_117894 | Shared | n.a. | n.a. | n.a. | n.a. | n.a. | n.a. | n.a. | n.a. | 81.25 | d__Bacteria;p__Bacteroidota;c__Bacteroidia;o__Bacteroidales;f__Marinifilaceae;g__Odoribacter | 99.9 | blast;98.40 iPHoP-RF;93.10 RaFAH;92.30 CRISPR;53.10 |
| S12_vs_98175 | SHHC | Duplodnaviria | Heunggongvirae | Uroviricota | Caudoviricetes | n.a. | Unclassified | O | O | 95.22 | d__Bacteria;p__Firmicutes;c__Clostridia;o__Lachnospirales;f__Lachnospiraceae;g__Bariatricus | 90.3 | iPHoP-RF;89.50 blast;81.30 RaFAH;65.60 |
| S3_vs_119660 | Shared | Duplodnaviria | Heunggongvirae | Uroviricota | Caudoviricetes | n.a. | Unclassified | Unclassified | Unclassified | 91.3 | d__Bacteria;p__Bacteroidota;c__Bacteroidia;o__Bacteroidales;f__Bacteroidaceae;g__Bacteroides | 100 | RaFAH;98.50 CRISPR;95.40 iPHoP-RF;95.40 blast;93.90 |
| S3_vs_123834 | Shared | Duplodnaviria | Heunggongvirae | Uroviricota | Caudoviricetes | n.a. | Unclassified | O | O | 73.11 | d__Bacteria;p__Actinobacteriota;c__Actinomycetia;o__Actinomycetales;f__Bifidobacteriaceae;g__Bifidobacterium | 98.4 | CRISPR;98.40 blast;96.90 iPHoP-RF;96.70 RaFAH;61.70 |
| S12_vs_99219 | Shared | n.a. | n.a. | n.a. | n.a. | n.a. | n.a. | n.a. | n.a. | 99.93 | d__Bacteria;p__Bacteroidota;c__Bacteroidia;o__Bacteroidales;f__Bacteroidaceae;g__Bacteroides | 100 | RaFAH;98.90 iPHoP-RF;96.70 blast;91.60 |
| S3_vs_127391 | ME | n.a. | n.a. | n.a. | n.a. | n.a. | n.a. | n.a. | n.a. | 18.32 | d__Bacteria;p__Firmicutes;c__Clostridia;o__Oscillospirales;f__Ruminococcaceae;g__Gemmiger | 95.6 | blast;96.90 CRISPR;96.30 iPHoP-RF;91.80 |
| S3_vs_127413 | ME | Duplodnaviria | Heunggongvirae | Uroviricota | Caudoviricetes | n.a. | Unclassified | O | O | 23.82 | d__Bacteria;p__Bacteroidota;c__Bacteroidia;o__Bacteroidales;f__Rikenellaceae;g__Alistipes | 97.9 | CRISPR;98.50 iPHoP-RF;80.00 |
| S3_vs_127442 | ME | Monodnaviria | Sangervirae | Phixviricota | Malgrandaviricetes | Petitvirales | Microviridae | Unclassified | Unclassified | | d__Bacteria;p__Bacteroidota;c__Bacteroidia;o__Bacteroidales;f__P3;g__Phil12 | 90.7 | iPHoP-RF;93.10 |
| S3_vs_17168 | Shared | Duplodnaviria | Heunggongvirae | Uroviricota | Caudoviricetes | n.a. | Unclassified | O | O | 100 | d__Bacteria;p__Bacteroidota;c__Bacteroidia;o__Bacteroidales;f__Rikenellaceae;g__Alistipes | 97.5 | blast;96.90 iPHoP-RF;93.10 RaFAH;71.20 |
| S3_vs_18858 | ME | n.a. | n.a. | n.a. | n.a. | n.a. | n.a. | n.a. | n.a. | 82.07 | d__Bacteria;p__Firmicutes;c__Clostridia;o__Oscillospirales;f__Acutalibacteraceae;g__CAG-217 | 97.9 | blast;98.50 iPHoP-RF;86.30 CRISPR;68.10 |
| S3_vs_29077 | ME | Duplodnaviria | Heunggongvirae | Uroviricota | Caudoviricetes | n.a. | Unclassified | O | O | 34.15 | d__Bacteria;p__Firmicutes;c__Clostridia;o__Oscillospirales;f__Ruminococcaceae;g__Ruminococcus_C | 94.9 | blast;96.40 iPHoP-RF;80.00 CRISPR;51.40 |
| S13_vf_147589 | ME | n.a. | n.a. | n.a. | n.a. | n.a. | n.a. | n.a. | n.a. | 6.46 | d__Bacteria;p__Firmicutes;c__Clostridia;o__Oscillospirales;f__Oscillospiraceae;g__Flavonifractor | 91.5 | blast;93.80 iPHoP-RF;78.50 |
| S3_vs_42569 | ME | Duplodnaviria | Heunggongvirae | Uroviricota | Caudoviricetes | n.a. | Unclassified | Unclassified | Toutatisvirus | 73.75 | d__Bacteria;p__Firmicutes;c__Clostridia;o__Oscillospirales;f__Ruminococcaceae;g__Faecalibacterium | 99.7 | CRISPR;98.30 blast;96.90 RaFAH;90.40 iPHoP-RF;73.70 |
| S3_vs_45147 | ME | n.a. | n.a. | n.a. | n.a. | n.a. | n.a. | n.a. | n.a. | 90.75 | d__Bacteria;p__Bacteroidota;c__Bacteroidia;o__Bacteroidales;f__Bacteroidaceae;g__Bacteroides | 99.4 | iPHoP-RF;97.10 RaFAH;91.20 |
| S3_vs_49835 | ME | n.a. | n.a. | n.a. | n.a. | n.a. | n.a. | n.a. | n.a. | 13.1 | d__Bacteria;p__Firmicutes;c__Clostridia;o__Oscillospirales;f__Acutalibacteraceae;g__Ruminococcus_E | 99 | CRISPR;99.20 blast;98.60 iPHoP-RF;93.10 |
| S3_vs_50924 | ME | n.a. | n.a. | n.a. | n.a. | n.a. | n.a. | n.a. | n.a. | 12.69 | d__Bacteria;p__Cyanobacteria;c__Vampirovibrionia;o__Gastranaerophilales;f__Gastranaerophilaceae;g__QAMI01 | 90.4 | blast;92.90 iPHoP-RF;57.10 |
| S3_vs_54303 | Shared | Duplodnaviria | Heunggongvirae | Uroviricota | Caudoviricetes | n.a. | Unclassified | Unclassified | Eponavirus | 46.83 | d__Bacteria;p__Firmicutes;c__Clostridia;o__Oscillospirales;f__Oscillospiraceae;g__ | 91.6 | blast;93.90 iPHoP-RF;80.30 |
| S3_vs_64424 | Shared | n.a. | n.a. | n.a. | n.a. | n.a. | n.a. | n.a. | n.a. | 98.41 | d__Bacteria;p__Bacteroidota;c__Bacteroidia;o__Bacteroidales;f__Bacteroidaceae;g__Bacteroides | 100 | RaFAH;99.60 iPHoP-RF;97.10 blast;92.20 CRISPR;69.50 |
| S3_vs_70508 | Shared | n.a. | n.a. | n.a. | n.a. | n.a. | n.a. | n.a. | n.a. | 99.66 | d__Bacteria;p__Bacteroidota;c__Bacteroidia;o__Bacteroidales;f__Bacteroidaceae;g__Bacteroides | 100 | RaFAH;99.70 iPHoP-RF;93.10 blast;92.70 |
| S3_vs_73361 | ME | n.a. | n.a. | n.a. | n.a. | n.a. | n.a. | n.a. | n.a. | 17.97 | d__Bacteria;p__Firmicutes;c__Clostridia;o__Christensenellales;f__CAG-138;g__SFEL01 | 96.6 | CRISPR;97.60 blast;96.40 iPHoP-RF;91.10 |
| S3_vs_77156 | ME | Monodnaviria | Sangervirae | Phixviricota | Malgrandaviricetes | Petitvirales | Microviridae | Unclassified | Unclassified | | d__Bacteria;p__Bacteroidota;c__Bacteroidia;o__Bacteroidales;f__Rikenellaceae;g__Alistipes | 96.7 | CRISPR;97.70 iPHoP-RF;58.40 |
| S3_vs_80151 | Shared | Duplodnaviria | Heunggongvirae | Uroviricota | Caudoviricetes | n.a. | Unclassified | O | O | 97.75 | d__Bacteria;p__Bacteroidota;c__Bacteroidia;o__Bacteroidales;f__Tannerellaceae;g__Parabacteroides | 100 | RaFAH;99.60 iPHoP-RF;94.40 blast;93.80 CRISPR;92.60 |
| S13_vf_54201 | ME | Duplodnaviria | Heunggongvirae | Uroviricota | Caudoviricetes | n.a. | Unclassified | O | O | 17.62 | d__Bacteria;p__Firmicutes;c__Clostridia;o__Oscillospirales;f__Ruminococcaceae;g__Faecalibacterium | 98.7 | RaFAH;93.20 blast;89.90 iPHoP-RF;62.20 |
| S3_vs_82895 | Shared | Duplodnaviria | Heunggongvirae | Uroviricota | Caudoviricetes | n.a. | Unclassified | O | O | 92.58 | d__Bacteria;p__Bacteroidota;c__Bacteroidia;o__Bacteroidales;f__Tannerellaceae;g__Parabacteroides | 100 | CRISPR;99.00 RaFAH;98.20 blast;93.90 iPHoP-RF;93.70 |
| S3_vs_90341 | Shared | n.a. | n.a. | n.a. | n.a. | n.a. | n.a. | n.a. | n.a. | 11.42 | d__Bacteria;p__Firmicutes;c__Clostridia;o__Christensenellales;f__CAG-74;g__UBA11524 | 95.2 | blast;96.60 CRISPR;94.50 iPHoP-RF;85.70 |
| S13_vf_66508 | ME | Duplodnaviria | Heunggongvirae | Uroviricota | Caudoviricetes | n.a. | Unclassified | O | O | 88.17 | d__Bacteria;p__Bacteroidota;c__Bacteroidia;o__Bacteroidales;f__Bacteroidaceae;g__Bacteroides | 99.4 | iPHoP-RF;97.10 blast;96.60 RaFAH;91.90 |
| S3_vs_16825 | Shared | Duplodnaviria | Heunggongvirae | Uroviricota | Caudoviricetes | n.a. | Unclassified | O | O | 80.45 | d__Bacteria;p__Firmicutes;c__Clostridia;o__Lachnospirales;f__Lachnospiraceae;g__Roseburia | 99.7 | CRISPR;98.90 iPHoP-RF;96.10 blast;92.80 RaFAH;88.00 |
| S3_vs_95574 | ME | n.a. | n.a. | n.a. | n.a. | n.a. | n.a. | n.a. | n.a. | 22.37 | d__Bacteria;p__Firmicutes;c__Clostridia;o__Oscillospirales;f__Acutalibacteraceae;g__UBA737 | 90.4 | blast;92.90 iPHoP-RF;70.60 |
| S3_vs_98675 | ME | Duplodnaviria | Heunggongvirae | Uroviricota | Caudoviricetes | n.a. | Unclassified | O | O | 8.18 | d__Bacteria;p__Firmicutes;c__Clostridia;o__Clostridiales;f__Clostridiaceae;g__Clostridium | 98.5 | blast;96.40 RaFAH;85.10 iPHoP-RF;79.00 |
| S4_vf_10506 | SHHC | n.a. | n.a. | n.a. | n.a. | n.a. | n.a. | n.a. | n.a. | 17.1 | d__Bacteria;p__Firmicutes;c__Bacilli;o__Erysipelotrichales;f__Erysipelotrichaceae;g__Holdemanella | 90.7 | blast;93.10 |
| S4_vf_12614 | SHHC | n.a. | n.a. | n.a. | n.a. | n.a. | n.a. | n.a. | n.a. | 80.07 | d__Bacteria;p__Bacteroidota;c__Bacteroidia;o__Bacteroidales;f__Barnesiellaceae;g__Barnesiella | 98.7 | blast;99.00 iPHoP-RF;76.10 CRISPR;67.90 |
| S4_vf_12624 | SHHC | Duplodnaviria | Heunggongvirae | Uroviricota | Caudoviricetes | n.a. | Unclassified | O | O | 24.52 | d__Bacteria;p__Actinobacteriota;c__Actinomycetia;o__Actinomycetales;f__Bifidobacteriaceae;g__Bifidobacterium | 97.5 | CRISPR;98.20 blast;96.50 iPHoP-RF;91.80 |
| S3_vs_39774 | Shared | n.a. | n.a. | n.a. | n.a. | n.a. | n.a. | n.a. | n.a. | 100 | d__Bacteria;p__Bacteroidota;c__Bacteroidia;o__Bacteroidales;f__Bacteroidaceae;g__Bacteroides | 100 | RaFAH;99.00 blast;92.00 iPHoP-RF;53.80 |
| S4_vf_18803 | SHHC | Duplodnaviria | Heunggongvirae | Uroviricota | Caudoviricetes | n.a. | Unclassified | Unclassified | Unclassified | 47.59 | d__Bacteria;p__Bacteroidota;c__Bacteroidia;o__Bacteroidales;f__Bacteroidaceae;g__Prevotella | 98.5 | CRISPR;98.90 iPHoP-RF;74.10 |
| S4_vf_31978 | SHHC | n.a. | n.a. | n.a. | n.a. | n.a. | n.a. | n.a. | n.a. | 62.52 | d__Bacteria;p__Firmicutes;c__Clostridia;o__Lachnospirales;f__Lachnospiraceae;g__Lachnospira | 99.6 | CRISPR;99.60 blast;93.90 iPHoP-RF;92.40 |
| S13_vs_13129 | ME | Duplodnaviria | Heunggongvirae | Uroviricota | Caudoviricetes | n.a. | Unclassified | O | O | 16.24 | d__Bacteria;p__Firmicutes;c__Clostridia;o__Oscillospirales;f__Oscillospiraceae;g__CAG-83 | 98.5 | CRISPR;98.90 blast;95.20 iPHoP-RF;87.90 |
| S4_vf_31981 | SHHC | Duplodnaviria | Heunggongvirae | Uroviricota | Caudoviricetes | Crassvirales | Suoliviridae | Uncouvirinae | Besingivirus | 29.33 | d__Bacteria;p__Bacteroidota;c__Bacteroidia;o__Bacteroidales;f__Bacteroidaceae;g__Prevotella | 97.3 | RaFAH;98.10 |
| S4_vf_4838 | Shared | Duplodnaviria | Heunggongvirae | Uroviricota | Caudoviricetes | Crassvirales | Suoliviridae | Boorivirinae | Cohcovirus | 76.93 | d__Bacteria;p__Bacteroidota;c__Bacteroidia;o__Bacteroidales;f__Tannerellaceae;g__Parabacteroides | 97.6 | blast;98.30 iPHoP-RF;96.10 CRISPR;71.50 |
| S4_vf_4891 | SHHC | n.a. | n.a. | n.a. | n.a. | n.a. | n.a. | n.a. | n.a. | 28.59 | d__Bacteria;p__Bacteroidota;c__Bacteroidia;o__Bacteroidales;f__Bacteroidaceae;g__Prevotella | 94.6 | blast;96.20 CRISPR;93.50 iPHoP-RF;92.10 |
| S13_vs_149860 | ME | Duplodnaviria | Heunggongvirae | Uroviricota | Caudoviricetes | n.a. | Unclassified | Unclassified | O | 61.18 | d__Bacteria;p__Actinobacteriota;c__Actinomycetia;o__Actinomycetales;f__Bifidobacteriaceae;g__Bifidobacterium | 99.7 | CRISPR;99.70 iPHoP-RF;92.40 blast;92.00 |
| S3_vs_68031 | ME | Duplodnaviria | Heunggongvirae | Uroviricota | Caudoviricetes | n.a. | Unclassified | O | O | 27.92 | d__Bacteria;p__Firmicutes;c__Clostridia;o__Oscillospirales;f__Ruminococcaceae;g__Faecalibacterium | 95.6 | blast;96.90 iPHoP-RF;73.20 CRISPR;56.90 |
| S4_vs_VIRSorter_12831-circular-cat_2 | SHHC | Duplodnaviria | Heunggongvirae | Uroviricota | Caudoviricetes | n.a. | Unclassified | O | O | 21.81 | d__Bacteria;p__Firmicutes;c__Clostridia;o__Lachnospirales;f__Lachnospiraceae;g__Blautia_A | 91.6 | blast;93.90 iPHoP-RF;85.10 CRISPR;58.50 |
| S4_vs_VIRSorter_13190-cat_2 | Shared | Duplodnaviria | Heunggongvirae | Uroviricota | Caudoviricetes | n.a. | Unclassified | O | O | 96.31 | d__Bacteria;p__Bacteroidota;c__Bacteroidia;o__Bacteroidales;f__Tannerellaceae;g__Parabacteroides | 100 | RaFAH;99.80 iPHoP-RF;94.40 blast;91.20 CRISPR;66.50 |
| S4_vs_VIRSorter_15112-cat_2 | SHHC | n.a. | n.a. | n.a. | n.a. | n.a. | n.a. | n.a. | n.a. | 16.47 | d__Bacteria;p__Firmicutes;c__Clostridia;o__Peptostreptococcales;f__Peptostreptococcaceae;g__GCA-900066495 | 91.6 | blast;93.90 iPHoP-RF;75.70 |
| S4_vs_VIRSorter_15837-cat_2 | Shared | Duplodnaviria | Heunggongvirae | Uroviricota | Caudoviricetes | n.a. | Unclassified | O | O | 96.3 | d__Bacteria;p__Firmicutes;c__Clostridia;o__Oscillospirales;f__Oscillospiraceae;g__Flavonifractor | 97.2 | blast;98.00 CRISPR;89.70 iPHoP-RF;84.80 |
| S4_vs_VIRSorter_16493-cat_2 | Shared | Duplodnaviria | Heunggongvirae | Uroviricota | Caudoviricetes | n.a. | Unclassified | Unclassified | Unclassified | 85.89 | d__Bacteria;p__Actinobacteriota;c__Actinomycetia;o__Actinomycetales;f__Bifidobacteriaceae;g__Bifidobacterium | 97.9 | CRISPR;98.50 iPHoP-RF;96.40 blast;92.50 |

| | | | | | | | | | | | | | |
|---|---|---|---|---|---|---|---|---|---|---|---|---|---|
| S4_vs_VIRSorter_19324-cat_2 | SHHC | Duplodnaviria | Heunggongvirae | Uroviricota | Caudoviricetes | n.a. | Unclassified | O | O | 25.86 | d__Bacteria;p__Firmicutes;c__Clostridia;o__Lachnospirales;f__Lachnospiraceae;g__Mediterraneibacter | 91.4 | blast;93.70 iPHoP-RF;76.40 |
| S4_vs_VIRSorter_3006-cat_1 | Shared | Duplodnaviria | Heunggongvirae | Uroviricota | Caudoviricetes | n.a. | Unclassified | O | O | 99.88 | d__Bacteria;p__Firmicutes;c__Clostridia;o__Lachnospirales;f__Lachnospiraceae;g__Mediterraneibacter | 91.6 | blast;93.90 |
| S4_vs_VIRSorter_30222-cat_2 | Shared | Duplodnaviria | Heunggongvirae | Uroviricota | Caudoviricetes | n.a. | Unclassified | O | O | 65.61 | d__Bacteria;p__Firmicutes;c__Clostridia;o__Oscillospirales;f__Oscillospiraceae;g__Intestinimonas | 91.5 | blast;93.80 |
| S4_vs_VIRSorter_31171-circular-cat_2 | SHHC | Duplodnaviria | Heunggongvirae | Uroviricota | Caudoviricetes | n.a. | Unclassified | O | O | 46.28 | d__Bacteria;p__Firmicutes;c__Clostridia;o__Oscillospirales;f__Ruminococcaceae;g__QAMM01 | 90.2 | blast;92.70 iPHoP-RF;54.90 |
| S4_vs_VIRSorter_31196-circular-cat_2 | Shared | n.a. | n.a. | n.a. | n.a. | n.a. | n.a. | n.a. | n.a. | 93.39 | d__Bacteria;p__Firmicutes;c__Clostridia;o__Oscillospirales;f__Oscillospiraceae;g__Flavonifractor | 99.4 | CRISPR;97.90 blast;96.20 iPHoP-RF;88.90 RaFAH;87.70 |
| S13_vs_45488 | Shared | Duplodnaviria | Heunggongvirae | Uroviricota | Caudoviricetes | n.a. | Unclassified | O | O | 22.02 | d__Bacteria;p__Proteobacteria;c__Alphaproteobacteria;o__RF32;f__CAG-239;g__51-20 | 97.9 | CRISPR;98.50 blast;79.90 iPHoP-RF;74.60 |
| S4_vs_VIRSorter_31560-circular-cat_2 | Shared | Duplodnaviria | Heunggongvirae | Uroviricota | Caudoviricetes | n.a. | Unclassified | O | O | 59.79 | d__Bacteria;p__Firmicutes;c__Clostridia;o__Oscillospirales;f__Oscillospiraceae;g__Flavonifractor | 90.3 | blast;92.80 iPHoP-RF;86.30 CRISPR;64.80 |
| S4_vs_VIRSorter_31976-circular-cat_2 | Shared | n.a. | n.a. | n.a. | n.a. | n.a. | n.a. | n.a. | n.a. | 17.17 | d__Bacteria;p__Firmicutes;c__Clostridia;o__Oscillospirales;f__Ruminococcaceae;g__Gemmiger | 97.2 | CRISPR;98.00 blast;96.20 iPHoP-RF;93.70 |
| S13_vs_57267 | Shared | Duplodnaviria | Heunggongvirae | Uroviricota | Caudoviricetes | n.a. | Unclassified | O | O | 38.42 | d__Bacteria;p__Bacteroidota;c__Bacteroidia;o__Bacteroidales;f__Bacteroidaceae;g__Prevotella | 90.6 | RaFAH;93.00 |
| S4_vs_VIRSorter_31980-circular-cat_2 | SHHC | Duplodnaviria | Heunggongvirae | Uroviricota | Caudoviricetes | n.a. | Unclassified | Unclassified | Unclassified | 13.26 | d__Bacteria;p__Proteobacteria;c__Gammaproteobacteria;o__Burkholderiales;f__Burkholderiaceae;g__Duodenibacillus | 96.7 | blast;97.70 CRISPR;83.90 iPHoP-RF;70.60 |
| S4_vs_VIRSorter_4746-cat_2 | Shared | Duplodnaviria | Heunggongvirae | Uroviricota | Caudoviricetes | n.a. | Salasmaviridae | O | O | 5.45 | d__Bacteria;p__Firmicutes;c__Clostridia;o__Lachnospirales;f__Lachnospiraceae;g__Roseburia | 90.3 | iPHoP-RF;92.80 |
| S4_vs_VIRSorter_4839-cat_2 | SHHC | Duplodnaviria | Heunggongvirae | Uroviricota | Caudoviricetes | n.a. | Unclassified | O | O | 69.64 | d__Bacteria;p__Firmicutes;c__Clostridia;o__Oscillospirales;f__Ruminococcaceae;g__Massilimaliae | 91.4 | blast;93.70 iPHoP-RF;75.70 |
| S4_vs_VIRSorter_4959-cat_2 | Shared | Duplodnaviria | Heunggongvirae | Uroviricota | Caudoviricetes | n.a. | Unclassified | O | O | 50.34 | d__Bacteria;p__Firmicutes;c__Clostridia;o__Oscillospirales;f__Oscillospiraceae;g__ER4 | 94.2 | blast;95.90 iPHoP-RF;82.50 |
| S4_vs_VIRSorter_6022-circular-cat_1 | Shared | Duplodnaviria | Heunggongvirae | Uroviricota | Caudoviricetes | n.a. | Unclassified | Unclassified | Unclassified | 97.71 | d__Bacteria;p__Bacteroidota;c__Bacteroidia;o__Bacteroidales;f__Tannerellaceae;g__Marseille-P3160 | 98.4 | CRISPR;98.80 iPHoP-RF;65.30 |
| S4_vs_VIRSorter_9913-cat_1 | Shared | Duplodnaviria | Heunggongvirae | Uroviricota | Caudoviricetes | n.a. | Unclassified | Unclassified | Unclassified | 98.3 | d__Bacteria;p__Bacteroidota;c__Bacteroidia;o__Bacteroidales;f__Marinifilaceae;g__Odoribacter | 99.7 | CRISPR;97.90 RaFAH;92.60 |
| S4_vf_31973 | Shared | n.a. | n.a. | n.a. | n.a. | n.a. | n.a. | n.a. | n.a. | 23.92 | d__Bacteria;p__Patescibacteria;c__Paceibacteria;o__UBA9983_A;f__UBA1006;g__01-FULL-36-10b | 92.2 | iPHoP-RF;94.40 |
| S5_vf_134040 | Shared | Duplodnaviria | Heunggongvirae | Uroviricota | Caudoviricetes | n.a. | Unclassified | Unclassified | Mushuvirus | 100 | d__Bacteria;p__Firmicutes;c__Clostridia;o__Oscillospirales;f__Ruminococcaceae;g__Faecalibacterium | 100 | RaFAH;98.80 blast;96.50 iPHoP-RF;69.10 |
| S5_vf_136076 | Shared | n.a. | n.a. | n.a. | n.a. | n.a. | n.a. | n.a. | n.a. | 43.14 | d__Bacteria;p__Firmicutes;c__Clostridia;o__Oscillospirales;f__Oscillospiraceae;g__ER4 | 93 | blast;95.00 iPHoP-RF;69.80 |
| S5_vf_137260 | Shared | Unclassified | n.a. | n.a. | n.a. | n.a. | Unclassified | O | O | 6.74 | d__Bacteria;p__Actinobacteriota;c__Coriobacteriia;o__Coriobacteriales;f__Coriobacteriaceae;g__Collinsella | 92.9 | CRISPR;94.90 |
| S5_vf_145585 | Shared | Duplodnaviria | Heunggongvirae | Uroviricota | Caudoviricetes | n.a. | Unclassified | Unclassified | Unclassified | 9.85 | d__Bacteria;p__Firmicutes;c__Clostridia;o__Lachnospirales;f__Lachnospiraceae;g__Enterocloster | 91.2 | blast;93.60 iPHoP-RF;91.40 CRISPR;53.80 |
| S14_vf_115736 | ME | Duplodnaviria | Heunggongvirae | Uroviricota | Caudoviricetes | n.a. | Unclassified | Unclassified | O | 85.01 | d__Bacteria;p__Firmicutes;c__Clostridia;o__Oscillospirales;f__Oscillospiraceae;g__Flavonifractor | 91.2 | blast;93.60 iPHoP-RF;89.80 CRISPR;61.00 |
| S5_vf_145588 | Shared | Duplodnaviria | Heunggongvirae | Uroviricota | Caudoviricetes | n.a. | Unclassified | O | O | 34.8 | d__Bacteria;p__Firmicutes;c__Clostridia;o__Lachnospirales;f__Lachnospiraceae;g__Dorea_A | 97.2 | CRISPR;98.00 iPHoP-RF;64.30 |
| S14_vf_117147 | ME | Duplodnaviria | Heunggongvirae | Uroviricota | Caudoviricetes | n.a. | Unclassified | O | O | 30.35 | d__Bacteria;p__Firmicutes;c__Clostridia;o__Oscillospirales;f__Ruminococcaceae;g__Gemmiger | 95.3 | blast;96.70 CRISPR;94.60 iPHoP-RF;86.30 |
| S5_vf_18461 | Shared | Duplodnaviria | Heunggongvirae | Uroviricota | Caudoviricetes | n.a. | Unclassified | O | O | 99.96 | d__Bacteria;p__Bacteroidota;c__Bacteroidia;o__Bacteroidales;f__Rikenellaceae;g__Alistipes | 92.7 | RaFAH;92.00 blast;67.20 |
| S5_vf_25423 | Shared | Duplodnaviria | Heunggongvirae | Uroviricota | Caudoviricetes | n.a. | Unclassified | Unclassified | Skunavirus | 81 | d__Bacteria;p__Firmicutes;c__Bacilli;o__Lactobacillales;f__Streptococcaceae;g__Lactococcus | 100 | RaFAH;98.80 blast;98.50 iPHoP-RF;91.40 |
| S14_vf_117155 | Shared | Duplodnaviria | Heunggongvirae | Uroviricota | Caudoviricetes | n.a. | Unclassified | O | O | 63.17 | d__Bacteria;p__Actinobacteriota;c__Actinomycetia;o__Actinomycetales;f__Actinomycetaceae;g__Pauljensenia | 98.4 | RaFAH;94.80 CRISPR;83.10 iPHoP-RF;64.80 |
| S5_vf_28061 | ME | n.a. | n.a. | n.a. | n.a. | n.a. | n.a. | n.a. | n.a. | 20.02 | d__Bacteria;p__Firmicutes;c__Bacilli;o__Haloplasmatales_A;f__Turicibacteraceae;g__Turicibacter | 91.5 | blast;93.80 iPHoP-RF;68.60 |
| S5_vf_6506 | ME | Duplodnaviria | Heunggongvirae | Uroviricota | Caudoviricetes | n.a. | Unclassified | O | O | 20.08 | d__Bacteria;p__Firmicutes;c__Clostridia;o__Clostridiales;f__Clostridiaceae;g__Clostridium_J | 90.3 | blast;92.80 iPHoP-RF;70.90 |
| S14_vf_117162 | ME | Duplodnaviria | Heunggongvirae | Uroviricota | Caudoviricetes | n.a. | Unclassified | Unclassified | O | 23.32 | d__Bacteria;p__Bacteroidota;c__Bacteroidia;o__Bacteroidales;f__Bacteroidaceae;g__Bacteroides | 94 | RaFAH;90.00 iPHoP-RF;70.90 CRISPR;63.50 |
| S5_vf_74661 | ME | Duplodnaviria | Heunggongvirae | Uroviricota | Caudoviricetes | n.a. | Unclassified | Unclassified | Unclassified | 91.73 | d__Bacteria;p__Proteobacteria;c__Gammaproteobacteria;o__Enterobacterales;f__Enterobacteriaceae;g__Escherichia | 100 | RaFAH;99.40 iPHoP-RF;97.10 blast;96.80 CRISPR;85.00 |
| S5_vf_8965 | ME | Duplodnaviria | Heunggongvirae | Uroviricota | Caudoviricetes | n.a. | Unclassified | O | O | 100 | d__Bacteria;p__Bacteroidota;c__Bacteroidia;o__Bacteroidales;f__Bacteroidaceae;g__Bacteroides | 100 | RaFAH;99.60 iPHoP-RF;97.10 blast;96.30 |
| S5_vs_107105 | ME | Duplodnaviria | Heunggongvirae | Uroviricota | Caudoviricetes | n.a. | Unclassified | O | O | 21.85 | d__Bacteria;p__Firmicutes;c__Clostridia;o__Oscillospirales;f__Oscillospiraceae;g__CAG-110 | 94.7 | blast;96.30 iPHoP-RF;67.00 |
| S5_vs_120079 | ME | Duplodnaviria | Heunggongvirae | Uroviricota | Caudoviricetes | n.a. | Unclassified | Unclassified | Unclassified | 88.37 | d__Bacteria;p__Firmicutes;c__Clostridia;o__Lachnospirales;f__Lachnospiraceae;g__Roseburia | 95.3 | blast;96.70 iPHoP-RF;96.40 CRISPR;84.10 |
| S4_vs_VIRSorter_31975-circular-cat_1 | Shared | Duplodnaviria | Heunggongvirae | Uroviricota | Caudoviricetes | n.a. | Unclassified | O | O | 88.33 | d__Bacteria;p__Bacteroidota;c__Bacteroidia;o__Bacteroidales;f__Bacteroidaceae;g__Bacteroides | 99.6 | iPHoP-RF;97.10 blast;96.60 CRISPR;94.50 RaFAH;92.70 |
| S14_vf_51134 | ME | Duplodnaviria | Heunggongvirae | Uroviricota | Caudoviricetes | n.a. | Unclassified | O | O | 47.7 | d__Bacteria;p__Firmicutes;c__Clostridia;o__Oscillospirales;f__Ruminococcaceae;g__Ruminococcus_D | 97.2 | blast;97.90 iPHoP-RF;92.80 RaFAH;52.60 |
| S14_vf_57689 | ME | Duplodnaviria | Heunggongvirae | Uroviricota | Caudoviricetes | n.a. | Unclassified | O | O | 94.04 | d__Bacteria;p__Actinobacteriota;c__Actinomycetia;o__Actinomycetales;f__Actinomycetaceae;g__Actinomyces | 99 | blast;94.00 RaFAH;93.30 CRISPR;80.00 iPHoP-RF;75.40 |
| S14_vf_85122 | ME | Duplodnaviria | Heunggongvirae | Uroviricota | Caudoviricetes | n.a. | Unclassified | Unclassified | O | 16.16 | d__Bacteria;p__Firmicutes;c__Clostridia;o__Clostridiales;f__Clostridiaceae;g__Clostridium | 95 | blast;92.30 iPHoP-RF;86.00 RaFAH;77.50 |
| S14_vf_93134 | ME | Duplodnaviria | Heunggongvirae | Uroviricota | Caudoviricetes | n.a. | Salasmaviridae | O | O | 12.04 | d__Bacteria;p__Firmicutes;c__Clostridia;o__Lachnospirales;f__Lachnospiraceae;g__Muricomes | 96.7 | CRISPR;97.70 |
| S5_vf_145579 | Shared | Duplodnaviria | Heunggongvirae | Uroviricota | Caudoviricetes | Crassvirales | Steigviridae | O | O | 44.59 | d__Bacteria;p__Bacteroidota;c__Bacteroidia;o__Bacteroidales;f__Bacteroidaceae;g__Prevotella | 94.6 | RaFAH;96.20 |
| S14_vs_100227 | ME | Duplodnaviria | Heunggongvirae | Uroviricota | Caudoviricetes | n.a. | Unclassified | O | O | 81.21 | d__Bacteria;p__Firmicutes;c__Clostridia;o__Lachnospirales;f__Lachnospiraceae;g__Enterocloster | 94.2 | CRISPR;95.90 blast;94.00 iPHoP-RF;86.30 |
| S14_vs_101482 | ME | n.a. | n.a. | n.a. | n.a. | n.a. | n.a. | n.a. | n.a. | 6.74 | d__Bacteria;p__Firmicutes;c__Clostridia;o__Oscillospirales;f__Ruminococcaceae;g__QAMM01 | 92 | blast;94.20 iPHoP-RF;69.10 |
| S14_vs_112068 | ME | n.a. | n.a. | n.a. | n.a. | n.a. | n.a. | n.a. | n.a. | 27.86 | d__Bacteria;p__Firmicutes;c__Clostridia;o__Oscillospirales;f__Butyricicoccaceae;g__Agathobaculum | 97.5 | blast;98.20 iPHoP-RF;66.00 |
| S14_vs_112345 | Shared | n.a. | n.a. | n.a. | n.a. | n.a. | n.a. | n.a. | n.a. | 71.03 | d__Bacteria;p__Bacteroidota;c__Bacteroidia;o__Bacteroidales;f__Bacteroidaceae;g__Bacteroides | 99.1 | iPHoP-RF;96.70 RaFAH;88.90 blast;75.90 |
| S14_vs_115377 | ME | Duplodnaviria | Heunggongvirae | Uroviricota | Caudoviricetes | n.a. | Unclassified | O | O | 76.83 | d__Bacteria;p__Firmicutes;c__Clostridia;o__Lachnospirales;f__Lachnospiraceae;g__Blautia | 98.8 | CRISPR;97.10 RaFAH;83.90 iPHoP-RF;60.90 |
| S14_vs_115455 | ME | Duplodnaviria | Heunggongvirae | Uroviricota | Caudoviricetes | n.a. | Unclassified | O | O | 25.58 | d__Bacteria;p__Firmicutes;c__Clostridia;o__Eubacteriales;f__Eubacteriaceae;g__Eubacterium | 93 | blast;95.00 CRISPR;89.70 iPHoP-RF;84.50 |
| S5_vs_109527 | Shared | n.a. | n.a. | n.a. | n.a. | n.a. | n.a. | n.a. | n.a. | 23.09 | d__Bacteria;p__Bacteroidota;c__Bacteroidia;o__Bacteroidales;f__Bacteroidaceae;g__Bacteroides | 99 | iPHoP-RF;96.40 blast;92.90 RaFAH;89.10 |
| S14_vs_117153 | ME | Duplodnaviria | Heunggongvirae | Uroviricota | Caudoviricetes | n.a. | Unclassified | O | O | 96.78 | d__Bacteria;p__Firmicutes;c__Clostridia;o__Lachnospirales;f__Lachnospiraceae;g__Dorea_A | 94.2 | blast;95.90 iPHoP-RF;94.70 CRISPR;58.40 |
| S5_vs_122344 | ME | Duplodnaviria | Heunggongvirae | Uroviricota | Caudoviricetes | n.a. | Unclassified | Unclassified | O | 26.84 | d__Bacteria;p__Firmicutes;c__Clostridia;o__Lachnospirales;f__Lachnospiraceae;g__Roseburia | 97.6 | CRISPR;98.30 blast;95.70 iPHoP-RF;93.10 |
| S14_vs_26320 | ME | Duplodnaviria | Heunggongvirae | Uroviricota | Caudoviricetes | n.a. | Unclassified | O | O | 65.93 | d__Bacteria;p__Firmicutes;c__Clostridia;o__Lachnospirales;f__Lachnospiraceae;g__Ruminococcus_B | 95.2 | blast;96.60 iPHoP-RF;82.20 |
| S14_vs_30108 | ME | n.a. | n.a. | n.a. | n.a. | n.a. | n.a. | n.a. | n.a. | 19.83 | d__Bacteria;p__Firmicutes;c__Clostridia;o__Lachnospirales;f__Lachnospiraceae;g__Enterocloster | 92.3 | blast;94.50 CRISPR;82.40 iPHoP-RF;78.30 |
| S14_vs_38905 | ME | n.a. | n.a. | n.a. | n.a. | n.a. | n.a. | n.a. | n.a. | 9.01 | d__Bacteria;p__Firmicutes;c__Clostridia;o__Oscillospirales;f__Ruminococcaceae;g__Ruthenibacterium | 93.8 | blast;95.60 iPHoP-RF;81.60 CRISPR;54.30 |
| S14_vs_39266 | ME | Duplodnaviria | Heunggongvirae | Uroviricota | Caudoviricetes | n.a. | Unclassified | Unclassified | O | 15.51 | d__Bacteria;p__Firmicutes;c__Clostridia;o__Oscillospirales;f__Acutalibacteraceae;g__Ruminococcus_E | 98.7 | CRISPR;99.00 blast;96.80 iPHoP-RF;93.70 |
| S14_vs_56251 | ME | Duplodnaviria | Heunggongvirae | Uroviricota | Caudoviricetes | n.a. | Unclassified | O | O | 20.56 | d__Bacteria;p__Firmicutes;c__Clostridia;o__Lachnospirales;f__Lachnospiraceae;g__Muricomes | 95.3 | iPHoP-RF;96.70 blast;93.30 CRISPR;68.70 |
| S14_vs_56410 | ME | Duplodnaviria | Heunggongvirae | Uroviricota | Caudoviricetes | n.a. | Unclassified | O | O | 80.58 | d__Bacteria;p__Firmicutes;c__Clostridia;o__Oscillospirales;f__Oscillospiraceae;g__Flavonifractor | 95.2 | blast;96.60 iPHoP-RF;95.40 CRISPR;94.00 |
| S14_vs_90934 | ME | n.a. | n.a. | n.a. | n.a. | n.a. | n.a. | n.a. | n.a. | 62.33 | d__Bacteria;p__Firmicutes;c__Clostridia;o__Oscillospirales;f__Ruminococcaceae;g__Faecalibacterium | 99.1 | blast;96.70 RaFAH;89.00 iPHoP-RF;81.10 |
| S14_vs_96286 | Shared | Duplodnaviria | Heunggongvirae | Uroviricota | Caudoviricetes | n.a. | Unclassified | O | O | 99.54 | d__Bacteria;p__Firmicutes;c__Clostridia;o__Lachnospirales;f__Lachnospiraceae;g__Blautia | 96.3 | RaFAH;97.40 |
| S14_vs_97835 | ME | n.a. | n.a. | n.a. | n.a. | n.a. | n.a. | n.a. | n.a. | | d__Bacteria;p__Firmicutes;c__Clostridia;o__Oscillospirales;f__Oscillospiraceae;g__CAG-83 | 90.6 | blast;93.00 iPHoP-RF;84.20 |
| S15_vs_3868 | Shared | Duplodnaviria | Heunggongvirae | Uroviricota | Caudoviricetes | n.a. | Unclassified | O | O | 100 | d__Bacteria;p__Bacteroidota;c__Bacteroidia;o__Bacteroidales;f__Bacteroidaceae;g__Bacteroides | 100 | RaFAH;99.10 blast;96.50 iPHoP-RF;61.40 |
| S15_vs_43913 | Shared | Duplodnaviria | Heunggongvirae | Uroviricota | Caudoviricetes | n.a. | Unclassified | O | O | 99.8 | d__Bacteria;p__Bacteroidota;c__Bacteroidia;o__Bacteroidales;f__Bacteroidaceae;g__Bacteroides | 100 | RaFAH;99.30 iPHoP-RF;97.10 blast;96.70 |
| S16_vf_100473 | Shared | n.a. | n.a. | n.a. | n.a. | n.a. | n.a. | n.a. | n.a. | 8.42 | d__Bacteria;p__Actinobacteriota;c__Actinomycetia;o__Actinomycetales;f__Bifidobacteriaceae;g__Bifidobacterium | 90.3 | blast;92.80 |
| S16_vf_16182 | Shared | Duplodnaviria | Heunggongvirae | Uroviricota | Caudoviricetes | n.a. | Unclassified | O | O | 64.12 | d__Bacteria;p__Actinobacteriota;c__Actinomycetia;o__Actinomycetales;f__Actinomycetaceae;g__Pauljensenia | 96.9 | RaFAH;95.10 CRISPR;68.30 iPHoP-RF;64.80 |
| S16_vf_172897 | SHHC | Duplodnaviria | Heunggongvirae | Uroviricota | Caudoviricetes | n.a. | Unclassified | O | O | 32.49 | d__Bacteria;p__Actinobacteriota;c__Actinomycetia;o__Actinomycetales;f__Bifidobacteriaceae;g__Bifidobacterium | 98.4 | CRISPR;98.80 iPHoP-RF;97.10 blast;93.30 |
| S16_vf_5809 | SHHC | n.a. | n.a. | n.a. | n.a. | n.a. | n.a. | n.a. | n.a. | 87.43 | d__Bacteria;p__Firmicutes;c__Clostridia;o__Lachnospirales;f__Lachnospiraceae;g__Lachnospira | 99.9 | CRISPR;99.00 iPHoP-RF;95.40 blast;93.40 RaFAH;91.70 |
| S16_vf_84462 | Shared | n.a. | n.a. | n.a. | n.a. | n.a. | n.a. | n.a. | n.a. | 31.67 | d__Bacteria;p__Firmicutes;c__Clostridia;o__Lachnospirales;f__Anaerotignaceae;g__Anaerotignum | 94.2 | blast;95.90 iPHoP-RF;79.50 |
| S16_vf_9270 | Shared | n.a. | n.a. | n.a. | n.a. | n.a. | n.a. | n.a. | n.a. | 8.58 | d__Bacteria;p__Bacteroidota;c__Bacteroidia;o__Bacteroidales;f__Rikenellaceae;g__Alistipes | 98.4 | CRISPR;98.80 iPHoP-RF;95.10 |
| S16_vs_101072 | SHHC | Duplodnaviria | Heunggongvirae | Uroviricota | Caudoviricetes | n.a. | Unclassified | O | O | 26.56 | d__Bacteria;p__Firmicutes;c__Clostridia;o__Lachnospirales;f__Lachnospiraceae;g__CAG-127 | 95 | blast;96.50 iPHoP-RF;87.30 |
| S16_vs_107235 | SHHC | n.a. | n.a. | n.a. | n.a. | n.a. | n.a. | n.a. | n.a. | 98.85 | d__Bacteria;p__Firmicutes;c__Clostridia;o__Oscillospirales;f__Ruminococcaceae;g__Ruminococcus_D | 99.7 | blast;99.00 iPHoP-RF;97.10 CRISPR;94.60 RaFAH;84.50 |
| S16_vs_118029 | SHHC | Duplodnaviria | Heunggongvirae | Uroviricota | Caudoviricetes | n.a. | Unclassified | O | O | 43.23 | d__Bacteria;p__Firmicutes;c__Clostridia;o__Lachnospirales;f__Lachnospiraceae;g__Eubacterium_G | 94.6 | blast;96.20 iPHoP-RF;85.70 |
| S16_vs_120149 | SHHC | n.a. | n.a. | n.a. | n.a. | n.a. | n.a. | n.a. | n.a. | 12.05 | d__Bacteria;p__Firmicutes;c__Clostridia;o__Oscillospirales;f__Acutalibacteraceae;g__Eubacterium_R | 94.5 | iPHoP-RF;96.10 blast;93.90 |
| S16_vs_132979 | Shared | Duplodnaviria | Heunggongvirae | Uroviricota | Caudoviricetes | n.a. | Unclassified | O | O | 31.86 | d__Bacteria;p__Firmicutes;c__Clostridia;o__Oscillospirales;f__Ruminococcaceae;g__Faecalibacterium | 94.3 | blast;96.00 iPHoP-RF;93.10 |
| S16_vs_140709 | Shared | n.a. | n.a. | n.a. | n.a. | n.a. | n.a. | n.a. | n.a. | 84.15 | d__Bacteria;p__Firmicutes;c__Clostridia;o__Lachnospirales;f__Lachnospiraceae;g__CAG-317 | 94.5 | iPHoP-RF;96.10 |
| S16_vs_141218 | Shared | n.a. | n.a. | n.a. | n.a. | n.a. | n.a. | n.a. | n.a. | 10.25 | d__Bacteria;p__Firmicutes;c__Bacilli;o__ML615J-28;f__CAG-313;g__CAG-313 | 98.7 | CRISPR;99.00 blast;96.10 |
| S16_vs_165662 | SHHC | Duplodnaviria | Heunggongvirae | Uroviricota | Caudoviricetes | n.a. | Unclassified | O | O | 11.88 | d__Bacteria;p__Firmicutes;c__Bacilli;o__RF39;f__UBA660;g__CAG-302 | 92.7 | blast;94.80 |
| S16_vs_173141 | Shared | n.a. | n.a. | n.a. | n.a. | n.a. | n.a. | n.a. | n.a. | | d__Bacteria;p__Firmicutes;c__Clostridia;o__Oscillospirales;f__Ruminococcaceae;g__Faecalibacterium | 96 | CRISPR;97.20 iPHoP-RF;82.80 |
| S16_vs_17493 | Shared | n.a. | n.a. | n.a. | n.a. | n.a. | n.a. | n.a. | n.a. | 18.24 | d__Bacteria;p__Firmicutes;c__Clostridia;o__Oscillospirales;f__Ruminococcaceae;g__Massilimaliae | 93 | blast;95.00 iPHoP-RF;58.40 |
| S16_vs_20060 | Shared | Duplodnaviria | Heunggongvirae | Uroviricota | Caudoviricetes | n.a. | Unclassified | Unclassified | O | 33.58 | d__Bacteria;p__Firmicutes;c__Clostridia;o__Oscillospirales;f__Oscillospiraceae;g__UMGS1766 | 91 | blast;93.40 |
| S16_vs_34021 | Shared | n.a. | n.a. | n.a. | n.a. | n.a. | n.a. | n.a. | n.a. | 99.81 | d__Bacteria;p__Firmicutes;c__Clostridia;o__Oscillospirales;f__Ruminococcaceae;g__Ruminococcus_D | 99 | blast;96.90 iPHoP-RF;90.50 RaFAH;86.90 |
| S16_vs_38863 | SHHC | Duplodnaviria | Heunggongvirae | Uroviricota | Caudoviricetes | n.a. | Unclassified | O | O | 26.24 | d__Bacteria;p__Firmicutes;c__Clostridia;o__Monoglobales;f__Monoglobaceae;g__Monoglobus | 95 | blast;96.50 iPHoP-RF;64.50 |
| S16_vs_76823 | SHHC | n.a. | n.a. | n.a. | n.a. | n.a. | n.a. | n.a. | n.a. | 20.02 | d__Bacteria;p__Firmicutes;c__Clostridia;o__Oscillospirales;f__Ruminococcaceae;g__Gemmiger | 98.5 | CRISPR;98.90 blast;98.30 iPHoP-RF;94.40 |
| S16_vs_78429 | SHHC | Duplodnaviria | Heunggongvirae | Uroviricota | Caudoviricetes | n.a. | Unclassified | Unclassified | O | 22.27 | d__Bacteria;p__Firmicutes;c__Clostridia;o__Oscillospirales;f__Oscillospiraceae;g__ER4 | 91.5 | blast;93.80 iPHoP-RF;52.80 |
| S16_vs_81805 | SHHC | Duplodnaviria | Heunggongvirae | Uroviricota | Caudoviricetes | n.a. | Unclassified | O | O | 99.42 | d__Bacteria;p__Firmicutes;c__Clostridia;o__Oscillospirales;f__Ruminococcaceae;g__Ruthenibacterium | 94.7 | blast;96.30 iPHoP-RF;89.50 CRISPR;57.90 |
| S16_vs_82116 | Shared | Duplodnaviria | Heunggongvirae | Uroviricota | Caudoviricetes | n.a. | Unclassified | Unclassified | O | 98.74 | d__Bacteria;p__Firmicutes;c__Clostridia;o__Oscillospirales;f__Acutalibacteraceae;g__Ruminococcus_E | 99.6 | CRISPR;98.90 blast;96.90 iPHoP-RF;94.10 RaFAH;81.90 |
| S17_vf_128147 | Shared | Monodnaviria | Sangervirae | Phixviricota | Malgrandaviricetes | Petitvirales | Microviridae | Unclassified | O | 3.97 | d__Bacteria;p__Bacteroidota;c__Bacteroidia;o__Bacteroidales;f__Bacteroidaceae;g__Bacteroides | 95.9 | blast;97.10 iPHoP-RF;97.10 CRISPR;89.30 |
| S17_vf_140653 | Shared | Duplodnaviria | Heunggongvirae | Uroviricota | Caudoviricetes | n.a. | Unclassified | O | O | 90.58 | d__Bacteria;p__Bacteroidota;c__Bacteroidia;o__Bacteroidales;f__Bacteroidaceae;g__Bacteroides | 99.7 | blast;96.70 RaFAH;95.30 iPHoP-RF;85.70 |
| S17_vf_36452 | ME | Duplodnaviria | Heunggongvirae | Uroviricota | Caudoviricetes | n.a. | Salasmaviridae | O | O | 12 | d__Bacteria;p__Firmicutes;c__Clostridia;o__Lachnospirales;f__Lachnospiraceae;g__Muricomes | 97.8 | CRISPR;98.40 iPHoP-RF;90.80 |
| S17_vf_46388 | Shared | Duplodnaviria | Heunggongvirae | Uroviricota | Caudoviricetes | n.a. | Unclassified | O | O | 50.05 | d__Bacteria;p__Firmicutes;c__Clostridia;o__Lachnospirales;f__Lachnospiraceae;g__Blautia_A | 99.4 | CRISPR;97.10 blast;94.50 iPHoP-RF;93.70 RaFAH;91.90 |
| S17_vs_110666 | ME | Duplodnaviria | Heunggongvirae | Uroviricota | Caudoviricetes | n.a. | Unclassified | O | O | 98.39 | d__Bacteria;p__Firmicutes;c__Clostridia;o__Oscillospirales;f__Ruminococcaceae;g__Faecalibacterium | 100 | RaFAH;99.70 blast;96.80 iPHoP-RF;93.40 CRISPR;67.80 |
| S17_vs_117507 | Shared | n.a. | n.a. | n.a. | n.a. | n.a. | n.a. | n.a. | n.a. | 40.08 | d__Bacteria;p__Bacteroidota;c__Bacteroidia;o__Bacteroidales;f__Bacteroidaceae;g__Bacteroides | 97.5 | blast;97.40 iPHoP-RF;96.40 RaFAH;66.20 |
| S17_vs_157209 | Shared | n.a. | n.a. | n.a. | n.a. | n.a. | n.a. | n.a. | n.a. | 25.09 | d__Bacteria;p__Bacteroidota;c__Bacteroidia;o__Bacteroidales;f__Rikenellaceae;g__Alistipes | 97.6 | CRISPR;98.30 blast;95.20 iPHoP-RF;92.80 |
| S17_vs_177203 | ME | Duplodnaviria | Heunggongvirae | Uroviricota | Caudoviricetes | n.a. | Unclassified | O | O | 71.95 | d__Bacteria;p__Desulfobacterota;c__Desulfovibrionia;o__Desulfovibrionales;f__Desulfovibrionaceae;g__Bilophila | 93.9 | blast;95.70 iPHoP-RF;90.10 |
| S17_vs_31654 | ME | Duplodnaviria | Heunggongvirae | Uroviricota | Caudoviricetes | n.a. | Unclassified | O | O | 36.85 | d__Bacteria;p__Firmicutes;c__Bacilli;o__Erysipelotrichales;f__Erysipelatoclostridiaceae;g__Faecalibacillus | 94.6 | blast;96.20 iPHoP-RF;81.60 CRISPR;54.30 |
| S17_vs_8690 | ME | n.a. | n.a. | n.a. | n.a. | n.a. | n.a. | n.a. | n.a. | 6.78 | d__Bacteria;p__Proteobacteria;c__Alphaproteobacteria;o__RF32;f__CAG-239;g__CAG-495 | 95.2 | blast;96.60 |
| S1_vf_136526 | ME | Duplodnaviria | Heunggongvirae | Uroviricota | Caudoviricetes | n.a. | Unclassified | Unclassified | O | 33.32 | d__Bacteria;p__Firmicutes;c__Clostridia;o__Oscillospirales;f__Oscillospiraceae;g__CAG-170 | 91.6 | blast;93.90 iPHoP-RF;88.50 |
| S1_vf_28398 | ME | n.a. | n.a. | n.a. | n.a. | n.a. | n.a. | n.a. | n.a. | 12.4 | d__Bacteria;p__Firmicutes;c__Clostridia;o__Oscillospirales;f__Ruminococcaceae;g__Gemmiger | 91.6 | blast;93.90 iPHoP-RF;75.40 |
| S1_vf_6272 | Shared | Duplodnaviria | Heunggongvirae | Uroviricota | Caudoviricetes | n.a. | Unclassified | Unclassified | O | 25.47 | d__Bacteria;p__Firmicutes;c__Clostridia;o__Oscillospirales;f__Oscillospiraceae;g__CAG-170 | 92 | CRISPR;94.20 blast;90.80 iPHoP-RF;78.00 |
| S1_vs_117317 | ME | Duplodnaviria | Heunggongvirae | Uroviricota | Caudoviricetes | n.a. | Unclassified | Unclassified | O | 21.23 | d__Bacteria;p__Firmicutes;c__Clostridia;o__Oscillospirales;f__Oscillospiraceae;g__CAG-170 | 93.4 | blast;95.30 iPHoP-RF;89.50 CRISPR;71.10 |
| S1_vs_133577 | ME | Duplodnaviria | Heunggongvirae | Uroviricota | Caudoviricetes | n.a. | Unclassified | O | O | 37.09 | d__Bacteria;p__Firmicutes;c__Clostridia;o__Oscillospirales;f__Ruminococcaceae;g__Gemmiger | 96 | blast;97.20 CRISPR;95.40 iPHoP-RF;94.40 |
| S1_vs_145792 | ME | n.a. | n.a. | n.a. | n.a. | n.a. | n.a. | n.a. | n.a. | 13.59 | d__Bacteria;p__Firmicutes;c__Clostridia;o__Lachnospirales;f__Anaerotignaceae;g__Anaerotignum | 94.6 | blast;96.20 iPHoP-RF;72.60 |
| S1_vs_155183 | Shared | Duplodnaviria | Heunggongvirae | Uroviricota | Caudoviricetes | n.a. | Unclassified | Unclassified | O | 31.14 | d__Bacteria;p__Firmicutes;c__Clostridia;o__Oscillospirales;f__Oscillospiraceae;g__ER4 | 95.9 | CRISPR;97.10 blast;96.20 iPHoP-RF;89.20 |
| S1_vs_23682 | ME | Duplodnaviria | Heunggongvirae | Uroviricota | Caudoviricetes | n.a. | Unclassified | O | O | 33.26 | d__Bacteria;p__Firmicutes;c__Clostridia;o__Oscillospirales;f__Ruminococcaceae;g__Faecalibacterium | 98.8 | CRISPR;99.10 blast;96.20 iPHoP-RF;83.30 |
| S2_vf_16829 | Shared | Duplodnaviria | Heunggongvirae | Uroviricota | Caudoviricetes | n.a. | Unclassified | Unclassified | O | 81.02 | d__Bacteria;p__Actinobacteriota;c__Coriobacteriia;o__Coriobacteriales;f__Eggerthellaceae;g__Eggerthella | 94.9 | CRISPR;96.40 iPHoP-RF;53.80 RaFAH;50.20 |
| S3_vf_101611 | Shared | Duplodnaviria | Heunggongvirae | Uroviricota | Caudoviricetes | n.a. | Unclassified | O | O | 23.54 | d__Bacteria;p__Firmicutes;c__Clostridia;o__Monoglobales;f__Monoglobaceae;g__Monoglobus | 95.2 | blast;96.60 |
| S2_vf_17850 | Shared | Duplodnaviria | Heunggongvirae | Uroviricota | Caudoviricetes | n.a. | Salasmaviridae | O | O | 11.78 | d__Bacteria;p__Firmicutes;c__Clostridia;o__Lachnospirales;f__Lachnospiraceae;g__Muricomes | 92.7 | CRISPR;94.80 iPHoP-RF;78.70 |

| | | | | | | | | | | | | | |
|---|---|---|---|---|---|---|---|---|---|---|---|---|---|
| S3_vf_111893 | Shared | Duplodnaviria | Heunggongvirae | Uroviricota | Caudoviricetes | n.a. | Unclassified | O | O | 41.65 | d__Bacteria;p__Firmicutes;c__Clostridia;o__Oscillospirales;f__Ruminococcaceae;g__Ruminococcus_D | 95.4 | blast;96.80 CRISPR;96.10 iPHoP-RF;94.10 |
| S3_vf_127392 | ME | Duplodnaviria | Heunggongvirae | Uroviricota | Caudoviricetes | n.a. | Unclassified | O | O | 52.43 | d__Bacteria;p__Firmicutes;c__Clostridia;o__Oscillospirales;f__CAG-272;g__CAG-272 | 92.1 | blast;94.30 iPHoP-RF;52.20 |
| S3_vf_127408 | ME | Duplodnaviria | Heunggongvirae | Uroviricota | Caudoviricetes | n.a. | Unclassified | O | O | 6.08 | d__Bacteria;p__Actinobacteriota;c__Actinomycetia;o__Actinomycetales;f__Bifidobacteriaceae;g__Bifidobacterium | 98.5 | blast;98.90 CRISPR;97.60 iPHoP-RF;96.70 |
| S3_vf_23042 | Shared | Duplodnaviria | Heunggongvirae | Uroviricota | Caudoviricetes | n.a. | Unclassified | O | O | 83.69 | d__Bacteria;p__Firmicutes;c__Bacilli;o__Lactobacillales;f__Streptococcaceae;g__Lactococcus | 99.7 | blast;98.70 iPHoP-RF;90.10 RaFAH;89.50 |
| S3_vf_5525 | ME | n.a. | n.a. | n.a. | n.a. | n.a. | n.a. | n.a. | n.a. | 41.37 | d__Bacteria;p__Actinobacteriota;c__Actinomycetia;o__Actinomycetales;f__Actinomycetaceae;g__Actinomyces | 92.6 | RaFAH;85.90 iPHoP-RF;81.10 |
| S3_vf_56363 | Shared | Duplodnaviria | Heunggongvirae | Uroviricota | Caudoviricetes | n.a. | Unclassified | O | O | 23.06 | d__Bacteria;p__Firmicutes;c__Clostridia;o__Peptostreptococcales;f__Peptostreptococcaceae;g__GCA-900066495 | 91.4 | iPHoP-RF;93.70 |
| S3_vf_99626 | Shared | Duplodnaviria | Heunggongvirae | Uroviricota | Caudoviricetes | n.a. | Unclassified | O | O | 26.68 | d__Bacteria;p__Actinobacteriota;c__Coriobacteriia;o__Coriobacteriales;f__Coriobacteriaceae;g__Collinsella | 98.7 | CRISPR;99.00 iPHoP-RF;97.10 blast;92.10 |
| S3_vs_119771 | ME | Duplodnaviria | Heunggongvirae | Uroviricota | Caudoviricetes | n.a. | Unclassified | O | O | 27.39 | d__Bacteria;p__Firmicutes;c__Clostridia;o__Oscillospirales;f__Ruminococcaceae;g__Gemmiger | 95.9 | blast;97.10 iPHoP-RF;81.40 CRISPR;55.60 |
| S3_vs_122706 | ME | Duplodnaviria | Heunggongvirae | Uroviricota | Caudoviricetes | n.a. | Unclassified | O | O | 89.75 | d__Bacteria;p__Firmicutes;c__Clostridia;o__Oscillospirales;f__Acutalibacteraceae;g__CAG-217 | 95.3 | blast;96.70 iPHoP-RF;79.30 |
| S3_vs_127401 | ME | Duplodnaviria | Heunggongvirae | Uroviricota | Caudoviricetes | n.a. | Unclassified | O | O | 45.54 | d__Bacteria;p__Firmicutes;c__Clostridia;o__Lachnospirales;f__Lachnospiraceae;g__Lachnospira | 90.4 | blast;92.90 iPHoP-RF;70.60 |
| S3_vs_15450 | ME | Duplodnaviria | Heunggongvirae | Uroviricota | Caudoviricetes | n.a. | Unclassified | O | O | 21.5 | d__Bacteria;p__Firmicutes;c__Clostridia;o__Oscillospirales;f__Ruminococcaceae;g__Gemmiger | 95.4 | blast;96.80 iPHoP-RF;83.90 CRISPR;68.80 |
| S3_vs_23194 | ME | Duplodnaviria | Heunggongvirae | Uroviricota | Caudoviricetes | n.a. | Duneviridae | O | O | 42.15 | d__Bacteria;p__Bacteroidota;c__Bacteroidia;o__Bacteroidales;f__Bacteroidaceae;g__Bacteroides | 100 | RaFAH;98.70 iPHoP-RF;97.10 |
| S3_vs_40939 | ME | n.a. | n.a. | n.a. | n.a. | n.a. | n.a. | n.a. | n.a. | 23.65 | d__Bacteria;p__Bacteroidota;c__Bacteroidia;o__Bacteroidales;f__Bacteroidaceae;g__Bacteroides | 99.3 | iPHoP-RF;97.10 blast;96.60 RaFAH;89.70 |
| S3_vs_56292 | Shared | Duplodnaviria | Heunggongvirae | Uroviricota | Caudoviricetes | n.a. | Unclassified | O | O | 99.95 | d__Bacteria;p__Bacteroidota;c__Bacteroidia;o__Bacteroidales;f__Rikenellaceae;g__Alistipes | 99.9 | blast;96.90 RaFAH;96.10 iPHoP-RF;83.60 |
| S3_vs_6604 | Shared | Duplodnaviria | Heunggongvirae | Uroviricota | Caudoviricetes | n.a. | Duneviridae | O | O | 75.2 | d__Bacteria;p__Bacteroidota;c__Bacteroidia;o__Bacteroidales;f__Marinifilaceae;g__Butyricimonas | 95.4 | blast;96.80 iPHoP-RF;65.60 |
| S3_vs_94577 | ME | Duplodnaviria | Heunggongvirae | Uroviricota | Caudoviricetes | n.a. | Unclassified | O | O | 45.75 | d__Bacteria;p__Firmicutes;c__Bacilli;o__Erysipelotrichales;f__Erysipelatoclostridiaceae;g__Faecalibacillus | 97.8 | blast;98.40 |
| S3_vs_97827 | ME | n.a. | n.a. | n.a. | n.a. | n.a. | n.a. | n.a. | n.a. | 10.17 | d__Bacteria;p__Bacteroidota;c__Bacteroidia;o__Bacteroidales;f__Marinifilaceae;g__Odoribacter | 97.9 | blast;98.50 iPHoP-RF;90.10 CRISPR;71.50 |
| S4_vf_20440 | SHHC | Duplodnaviria | Heunggongvirae | Uroviricota | Caudoviricetes | n.a. | Unclassified | O | O | 44.47 | d__Bacteria;p__Bacteroidota;c__Bacteroidia;o__Bacteroidales;f__Bacteroidaceae;g__Prevotella | 97.8 | blast;98.40 iPHoP-RF;84.20 |
| S3_vf_11428 | Shared | Duplodnaviria | Heunggongvirae | Uroviricota | Caudoviricetes | n.a. | Unclassified | O | O | 94.85 | d__Bacteria;p__Firmicutes;c__Clostridia;o__Lachnospirales;f__Lachnospiraceae;g__Blautia_A | 100 | RaFAH;98.90 CRISPR;97.00 iPHoP-RF;93.40 blast;91.70 |
| S4_vs_VIRSorter_2744-cat_3 | SHHC | Duplodnaviria | Heunggongvirae | Uroviricota | Caudoviricetes | n.a. | Unclassified | O | O | 9.58 | d__Bacteria;p__Actinobacteriota;c__Actinomycetia;o__Actinomycetales;f__Bifidobacteriaceae;g__Bifidobacterium | 96 | CRISPR;97.20 iPHoP-RF;97.10 blast;96.70 |
| S4_vs_VIRSorter_31972-circular-cat_2 | SHHC | Duplodnaviria | Heunggongvirae | Uroviricota | Caudoviricetes | n.a. | Unclassified | O | O | 26.41 | d__Bacteria;p__Firmicutes;c__Clostridia;o__Oscillospirales;f__Ruminococcaceae;g__Ruminococcus_C | 97.3 | blast;98.10 CRISPR;93.20 iPHoP-RF;90.50 |
| S4_vs_VIRSorter_5698-cat_2 | SHHC | Duplodnaviria | Heunggongvirae | Uroviricota | Caudoviricetes | n.a. | Unclassified | O | O | 87.05 | d__Bacteria;p__Actinobacteriota;c__Actinomycetia;o__Actinomycetales;f__Bifidobacteriaceae;g__Bifidobacterium | 99 | CRISPR;98.50 blast;96.80 iPHoP-RF;96.10 RaFAH;71.90 |
| S5_vf_103206 | Shared | n.a. | n.a. | n.a. | n.a. | n.a. | n.a. | n.a. | n.a. | 3.75 | d__Bacteria;p__Firmicutes;c__Clostridia;o__Oscillospirales;f__Oscillospiraceae;g__Lawsonibacter | 90 | blast;92.50 |
| S5_vf_116300 | ME | n.a. | n.a. | n.a. | n.a. | n.a. | n.a. | n.a. | n.a. | 13.55 | d__Bacteria;p__Firmicutes;c__Clostridia;o__Lachnospirales;f__Lachnospiraceae;g__Anaerostipes | 95.2 | blast;96.60 iPHoP-RF;90.50 CRISPR;69.90 |
| S3_vf_127400 | Shared | Duplodnaviria | Heunggongvirae | Uroviricota | Caudoviricetes | n.a. | Unclassified | Unclassified | O | 54.09 | d__Bacteria;p__Firmicutes;c__Clostridia;o__Oscillospirales;f__Oscillospiraceae;g__ER4 | 95 | blast;96.50 iPHoP-RF;76.40 CRISPR;67.10 |
| S5_vf_55687 | Shared | Duplodnaviria | Heunggongvirae | Uroviricota | Caudoviricetes | n.a. | Unclassified | O | O | 91.19 | d__Bacteria;p__Firmicutes;c__Bacilli;o__Lactobacillales;f__Streptococcaceae;g__Lactococcus | 90.2 | RaFAH;92.70 |
| S5_vf_8808 | ME | Duplodnaviria | Heunggongvirae | Uroviricota | Caudoviricetes | n.a. | Unclassified | O | O | 97.29 | d__Bacteria;p__Firmicutes;c__Negativicutes;o__Veillonellales;f__Veillonellaceae;g__Veillonella | 100 | CRISPR;98.80 RaFAH;98.60 blast;93.70 iPHoP-RF;78.30 |
| S5_vs_10428 | ME | Duplodnaviria | Heunggongvirae | Uroviricota | Caudoviricetes | n.a. | Unclassified | O | O | 10.64 | d__Bacteria;p__Firmicutes;c__Clostridia;o__Christensenellales;f__CAG-74;g__Alectryocaccomicrobium | 91.4 | blast;93.70 |
| S5_vs_118168 | ME | n.a. | n.a. | n.a. | n.a. | n.a. | n.a. | n.a. | n.a. | 30.01 | d__Bacteria;p__Firmicutes;c__Clostridia;o__Oscillospirales;f__Ruminococcaceae;g__Anaerotruncus | 91 | blast;93.40 iPHoP-RF;57.30 |
| S5_vs_120584 | Shared | Duplodnaviria | Heunggongvirae | Uroviricota | Caudoviricetes | n.a. | Unclassified | O | O | 98.83 | d__Bacteria;p__Firmicutes;c__Clostridia;o__Lachnospirales;f__Lachnospiraceae;g__Lachnospira | 97.5 | CRISPR;98.20 blast;93.50 iPHoP-RF;88.90 |
| S5_vs_121411 | ME | n.a. | n.a. | n.a. | n.a. | n.a. | n.a. | n.a. | n.a. | 100 | d__Bacteria;p__Firmicutes;c__Clostridia;o__Oscillospirales;f__Ruminococcaceae;g__Ruthenibacterium | 95.3 | blast;96.70 iPHoP-RF;87.60 CRISPR;68.40 |
| S3_vf_127450 | ME | n.a. | n.a. | n.a. | n.a. | n.a. | n.a. | n.a. | n.a. | 8.34 | d__Bacteria;p__Bacteroidota;c__Bacteroidia;o__Bacteroidales;f__Bacteroidaceae;g__Prevotella | 92.3 | RaFAH;94.50 |
| S3_vf_7137 | Shared | Unclassified | n.a. | n.a. | n.a. | n.a. | Unclassified | Unclassified | O | 5.31 | d__Bacteria;p__Actinobacteriota;c__Actinomycetia;o__Actinomycetales;f__Bifidobacteriaceae;g__Bifidobacterium | 99.7 | CRISPR;99.70 iPHoP-RF;77.30 |
| S3_vf_8164 | ME | Duplodnaviria | Heunggongvirae | Uroviricota | Caudoviricetes | n.a. | Unclassified | O | O | 10.61 | d__Bacteria;p__Firmicutes;c__Clostridia;o__Oscillospirales;f__Ruminococcaceae;g__Ruminococcus_D | 98.2 | CRISPR;98.70 blast;96.80 iPHoP-RF;95.10 |
| S5_vs_117319 | ME | Duplodnaviria | Heunggongvirae | Uroviricota | Caudoviricetes | n.a. | Unclassified | Unclassified | O | 51.09 | d__Bacteria;p__Firmicutes;c__Clostridia;o__Oscillospirales;f__Ruminococcaceae;g__Faecalibacterium | 98.4 | blast;95.50 RaFAH;86.90 CRISPR;78.90 iPHoP-RF;76.80 |
| S3_vs_127411 | ME | n.a. | n.a. | n.a. | n.a. | n.a. | n.a. | n.a. | n.a. | 9.3 | d__Bacteria;p__Firmicutes;c__Clostridia;o__Oscillospirales;f__Oscillospiraceae;g__ER4 | 95.2 | blast;96.60 iPHoP-RF;73.20 |
| S3_vs_51931 | ME | Monodnaviria | Sangervirae | Phixviricota | Malgrandaviricetes | Petitvirales | Microviridae | Gokushovirinae | O | 21.18 | d__Bacteria;p__Proteobacteria;c__Gammaproteobacteria;o__Burkholderiales;f__Burkholderiaceae;g__Sutterella | 94.6 | CRISPR;96.20 iPHoP-RF;79.00 |
| S3_vs_127421 | Shared | Duplodnaviria | Heunggongvirae | Uroviricota | Caudoviricetes | n.a. | Unclassified | O | O | 95.6 | d__Bacteria;p__Firmicutes;c__Clostridia;o__Lachnospirales;f__Lachnospiraceae;g__Agathobacter | 100 | CRISPR;98.90 RaFAH;94.40 iPHoP-RF;92.10 |
| S5_vf_60597 | Shared | n.a. | n.a. | n.a. | n.a. | n.a. | n.a. | n.a. | n.a. | 35.7 | d__Bacteria;p__Firmicutes;c__Clostridia;o__Lachnospirales;f__Anaerotignaceae;g__Anaerotignum | 93.9 | blast;95.70 iPHoP-RF;84.20 |
| S3_vs_25896 | ME | n.a. | n.a. | n.a. | n.a. | n.a. | n.a. | n.a. | n.a. | 11.85 | d__Bacteria;p__Firmicutes;c__Clostridia;o__Oscillospirales;f__Ruminococcaceae;g__Ruthenibacterium | 91.4 | blast;93.70 iPHoP-RF;78.30 |
| S3_vs_39940 | ME | n.a. | n.a. | n.a. | n.a. | n.a. | n.a. | n.a. | n.a. | 5.41 | d__Bacteria;p__Actinobacteriota;c__Actinomycetia;o__Actinomycetales;f__Bifidobacteriaceae;g__Bifidobacterium | 95.9 | iPHoP-RF;97.10 blast;96.90 CRISPR;84.00 |
| S3_vs_50557 | ME | Duplodnaviria | Heunggongvirae | Uroviricota | Caudoviricetes | n.a. | Unclassified | O | O | 23.06 | d__Bacteria;p__Actinobacteriota;c__Coriobacteriia;o__Coriobacteriales;f__Eggerthellaceae;g__Gordonibacter | 93.4 | CRISPR;95.30 blast;93.90 iPHoP-RF;93.10 |
| S3_vs_9158 | Shared | n.a. | n.a. | n.a. | n.a. | n.a. | n.a. | n.a. | n.a. | 99.96 | d__Bacteria;p__Bacteroidota;c__Bacteroidia;o__Bacteroidales;f__Bacteroidaceae;g__Bacteroides | 100 | RaFAH;99.60 iPHoP-RF;97.10 blast;96.90 |
| S3_vs_87728 | Shared | n.a. | n.a. | n.a. | n.a. | n.a. | n.a. | n.a. | n.a. | 5.42 | d__Bacteria;p__Firmicutes;c__Clostridia;o__Oscillospirales;f__Ruminococcaceae;g__Faecalibacterium | 95.3 | blast;96.70 iPHoP-RF;67.00 |
| S3_vs_89485 | Shared | n.a. | n.a. | n.a. | n.a. | n.a. | n.a. | n.a. | n.a. | 73.42 | d__Bacteria;p__Bacteroidota;c__Bacteroidia;o__Bacteroidales;f__Rikenellaceae;g__Alistipes | 95 | blast;96.50 iPHoP-RF;93.40 |
| S5_vs_117417 | ME | n.a. | n.a. | n.a. | n.a. | n.a. | n.a. | n.a. | n.a. | 13.57 | d__Bacteria;p__Firmicutes;c__Clostridia;o__Oscillospirales;f__Oscillospiraceae;g__Enterenecus | 90.7 | blast;93.10 iPHoP-RF;70.90 |
| S3_vs_90331 | Shared | Duplodnaviria | Heunggongvirae | Uroviricota | Caudoviricetes | n.a. | Unclassified | Unclassified | O | 99.17 | d__Bacteria;p__Firmicutes;c__Clostridia;o__Oscillospirales;f__Ruminococcaceae;g__Faecalibacterium | 100 | RaFAH;98.70 CRISPR;97.70 blast;92.00 iPHoP-RF;85.70 |
| S4_vf_30103 | Shared | Unclassified | n.a. | n.a. | n.a. | n.a. | Unclassified | Unclassified | O | 5.83 | d__Bacteria;p__Actinobacteriota;c__Actinomycetia;o__Actinomycetales;f__Bifidobacteriaceae;g__Bifidobacterium | 99.7 | CRISPR;99.70 iPHoP-RF;92.10 |
| S4_vf_11308 | Shared | Duplodnaviria | Heunggongvirae | Uroviricota | Caudoviricetes | n.a. | Unclassified | Unclassified | O | 96.94 | d__Bacteria;p__Firmicutes;c__Clostridia;o__Oscillospirales;f__Acutalibacteraceae;g__Ruminococcus_E | 100 | CRISPR;98.90 blast;98.50 RaFAH;97.10 iPHoP-RF;93.70 |
| S4_vf_13353 | Shared | Duplodnaviria | Heunggongvirae | Uroviricota | Caudoviricetes | n.a. | Unclassified | O | O | 93.68 | d__Bacteria;p__Firmicutes;c__Clostridia;o__Lachnospirales;f__Lachnospiraceae;g__Blautia_A | 99.9 | CRISPR;97.90 RaFAH;95.20 blast;90.90 iPHoP-RF;89.20 |
| S4_vs_VIRSorter_31916-circular-cat_1 | SHHC | Duplodnaviria | Heunggongvirae | Uroviricota | Caudoviricetes | n.a. | Unclassified | O | O | 39.89 | d__Bacteria;p__Firmicutes;c__Clostridia;o__Lachnospirales;f__Lachnospiraceae;g__Mediterraneibacter | 90.7 | blast;93.10 iPHoP-RF;68.20 |
| S4_vf_31979 | SHHC | Duplodnaviria | Heunggongvirae | Uroviricota | Caudoviricetes | n.a. | Unclassified | Unclassified | O | 24.1 | d__Bacteria;p__Firmicutes;c__Clostridia;o__Christensenellales;f__CAG-74;g__Ventricola | 92 | blast;94.20 |
| S4_vs_VIRSorter_27316-cat_2 | Shared | Duplodnaviria | Heunggongvirae | Uroviricota | Caudoviricetes | n.a. | Unclassified | O | O | 74.11 | d__Bacteria;p__Firmicutes;c__Clostridia;o__Oscillospirales;f__Ruminococcaceae;g__Faecalibacterium | 95.4 | blast;96.80 iPHoP-RF;75.00 CRISPR;67.80 |
| S5_vf_145583 | ME | Duplodnaviria | Heunggongvirae | Uroviricota | Caudoviricetes | n.a. | Unclassified | Unclassified | O | 10.99 | d__Bacteria;p__Firmicutes;c__Clostridia;o__Clostridiales;f__Clostridiaceae;g__Clostridium | 95.2 | blast;93.60 iPHoP-RF;80.80 RaFAH;73.30 CRISPR;62.00 |
| S5_vf_105728 | ME | Duplodnaviria | Heunggongvirae | Uroviricota | Caudoviricetes | n.a. | Unclassified | O | O | 44.2 | d__Bacteria;p__Firmicutes;c__Clostridia;o__Lachnospirales;f__Lachnospiraceae;g__ | 94.5 | iPHoP-RF;96.10 blast;93.40 |
| S5_vf_145604 | Shared | Duplodnaviria | Heunggongvirae | Uroviricota | Caudoviricetes | n.a. | Unclassified | O | O | 87.41 | d__Bacteria;p__Firmicutes;c__Clostridia;o__Lachnospirales;f__Lachnospiraceae;g__Blautia_A | 99.6 | CRISPR;97.30 RaFAH;93.30 blast;92.90 iPHoP-RF;91.80 |
| S5_vf_21869 | Shared | Duplodnaviria | Heunggongvirae | Uroviricota | Caudoviricetes | n.a. | Unclassified | Unclassified | O | 31.69 | d__Bacteria;p__Firmicutes;c__Clostridia;o__Oscillospirales;f__Oscillospiraceae;g__Flavonifractor | 93.1 | blast;95.10 iPHoP-RF;73.90 |
| S5_vf_29332 | ME | Duplodnaviria | Heunggongvirae | Uroviricota | Caudoviricetes | n.a. | Unclassified | O | O | 12.73 | d__Bacteria;p__Actinobacteriota;c__Coriobacteriia;o__Coriobacteriales;f__Coriobacteriaceae;g__Collinsella | 97.3 | CRISPR;98.10 blast;94.60 iPHoP-RF;79.50 |
| S5_vs_122618 | ME | n.a. | n.a. | n.a. | n.a. | n.a. | n.a. | n.a. | n.a. | 17.7 | d__Bacteria;p__Firmicutes;c__Bacilli;o__Erysipelotrichales;f__Erysipelotrichaceae;g__Clostridium_AQ | 95.4 | blast;96.80 iPHoP-RF;71.10 CRISPR;66.50 |
| S5_vs_123256 | Shared | Duplodnaviria | Heunggongvirae | Uroviricota | Caudoviricetes | n.a. | Unclassified | Unclassified | Unclassified | 98.81 | d__Bacteria;p__Bacteroidota;c__Bacteroidia;o__Bacteroidales;f__Rikenellaceae;g__Alistipes | 100 | RaFAH;97.10 blast;96.90 iPHoP-RF;96.10 |
| S5_vs_124035 | Shared | n.a. | n.a. | n.a. | n.a. | n.a. | n.a. | n.a. | n.a. | 7.45 | d__Bacteria;p__Firmicutes;c__Clostridia;o__Oscillospirales;f__Oscillospiraceae;g__Intestinimonas | 91.2 | blast;93.60 iPHoP-RF;53.30 |
| S5_vs_124677 | ME | Duplodnaviria | Heunggongvirae | Uroviricota | Caudoviricetes | n.a. | Unclassified | Unclassified | O | 26.54 | d__Bacteria;p__Firmicutes;c__Clostridia;o__Oscillospirales;f__Ruminococcaceae;g__Faecalibacterium | 97 | CRISPR;97.90 blast;96.90 iPHoP-RF;90.80 |
| S5_vs_128398 | ME | Duplodnaviria | Heunggongvirae | Uroviricota | Caudoviricetes | n.a. | Unclassified | O | O | 22.85 | d__Bacteria;p__Firmicutes;c__Clostridia;o__Oscillospirales;f__Ruminococcaceae;g__Negativibacillus | 93.6 | blast;95.50 iPHoP-RF;82.20 |
| S5_vs_129970 | Shared | n.a. | n.a. | n.a. | n.a. | n.a. | n.a. | n.a. | n.a. | 93.93 | d__Bacteria;p__Bacteroidota;c__Bacteroidia;o__Bacteroidales;f__Bacteroidaceae;g__Bacteroides | 100 | RaFAH;99.60 blast;92.90 iPHoP-RF;92.80 |
| S5_vs_130032 | ME | Duplodnaviria | Heunggongvirae | Uroviricota | Caudoviricetes | n.a. | Unclassified | O | O | 98.82 | d__Bacteria;p__Proteobacteria;c__Gammaproteobacteria;o__Enterobacterales;f__Enterobacteriaceae;g__Escherichia | 100 | RaFAH;99.70 blast;96.90 iPHoP-RF;91.40 CRISPR;72.80 |
| S5_vs_130294 | Shared | n.a. | n.a. | n.a. | n.a. | n.a. | n.a. | n.a. | n.a. | 55.97 | d__Bacteria;p__Firmicutes;c__Clostridia;o__Oscillospirales;f__Ruminococcaceae;g__Faecalibacterium | 95.6 | blast;96.90 iPHoP-RF;81.60 |
| S5_vs_130958 | ME | Duplodnaviria | Heunggongvirae | Uroviricota | Caudoviricetes | n.a. | Unclassified | O | O | 18.11 | d__Bacteria;p__Firmicutes;c__Clostridia;o__Oscillospirales;f__Ruminococcaceae;g__Anaerotruncus | 94.2 | blast;95.90 iPHoP-RF;70.40 |
| S5_vs_131510 | Shared | Duplodnaviria | Heunggongvirae | Uroviricota | Caudoviricetes | n.a. | Unclassified | Unclassified | O | 95 | d__Bacteria;p__Bacteroidota;c__Bacteroidia;o__Bacteroidales;f__Bacteroidaceae;g__Bacteroides | 100 | RaFAH;97.50 blast;96.90 iPHoP-RF;96.40 |
| S5_vs_13254 | ME | n.a. | n.a. | n.a. | n.a. | n.a. | n.a. | n.a. | n.a. | 8.3 | d__Bacteria;p__Firmicutes;c__Clostridia;o__Oscillospirales;f__Ruminococcaceae;g__Ruthenibacterium | 95 | blast;96.50 iPHoP-RF;85.40 |
| S5_vs_1338 | ME | n.a. | n.a. | n.a. | n.a. | n.a. | n.a. | n.a. | n.a. | 100 | d__Bacteria;p__Bacteroidota;c__Bacteroidia;o__Bacteroidales;f__Rikenellaceae;g__Alistipes | 100 | RaFAH;97.80 blast;97.50 iPHoP-RF;76.10 |
| S5_vs_137154 | ME | Duplodnaviria | Heunggongvirae | Uroviricota | Caudoviricetes | n.a. | Unclassified | O | O | 10.76 | d__Bacteria;p__Firmicutes;c__Clostridia;o__Clostridiales;f__Clostridiaceae;g__Clostridium | 91.7 | blast;94.00 iPHoP-RF;67.30 |
| S5_vs_139401 | ME | n.a. | n.a. | n.a. | n.a. | n.a. | n.a. | n.a. | n.a. | 12.88 | d__Bacteria;p__Firmicutes;c__Clostridia;o__Lachnospirales;f__Lachnospiraceae;g__Anaerostipes | 90.9 | blast;93.30 CRISPR;68.40 iPHoP-RF;67.50 |
| S5_vs_140946 | ME | n.a. | n.a. | n.a. | n.a. | n.a. | n.a. | n.a. | n.a. | 9.68 | d__Bacteria;p__Firmicutes;c__Clostridia;o__Oscillospirales;f__Acutalibacteraceae;g__UMGS856 | 93.9 | blast;95.70 iPHoP-RF;76.80 |
| S5_vs_141621 | ME | Duplodnaviria | Heunggongvirae | Uroviricota | Caudoviricetes | n.a. | Unclassified | O | O | 97.16 | d__Bacteria;p__Firmicutes;c__Clostridia;o__Lachnospirales;f__Lachnospiraceae;g__Anaerostipes | 96.4 | blast;95.60 CRISPR;90.90 iPHoP-RF;88.90 RaFAH;72.10 |
| S5_vs_142195 | Shared | n.a. | n.a. | n.a. | n.a. | n.a. | n.a. | n.a. | n.a. | 100 | d__Bacteria;p__Bacteroidota;c__Bacteroidia;o__Bacteroidales;f__Bacteroidaceae;g__Bacteroides | 99.9 | RaFAH;99.80 |
| S5_vs_145578 | ME | Duplodnaviria | Heunggongvirae | Uroviricota | Caudoviricetes | n.a. | Unclassified | O | O | 39.67 | d__Bacteria;p__Firmicutes;c__Clostridia;o__Lachnospirales;f__Lachnospiraceae;g__Enterocloster | 95.2 | blast;96.60 iPHoP-RF;76.10 CRISPR;54.30 |
| S5_vs_145581 | ME | n.a. | n.a. | n.a. | n.a. | n.a. | n.a. | n.a. | n.a. | 9.66 | d__Bacteria;p__Firmicutes;c__Clostridia;o__Christensenellales;f__Christensenellaceae;g__Christensenella | 90.4 | blast;92.90 iPHoP-RF;86.30 |
| S5_vs_145590 | ME | Duplodnaviria | Heunggongvirae | Uroviricota | Caudoviricetes | n.a. | Unclassified | O | O | 39.74 | d__Bacteria;p__Firmicutes;c__Clostridia;o__Lachnospirales;f__Lachnospiraceae;g__Clostridium_AP | 91.4 | iPHoP-RF;93.70 |
| S5_vs_145597 | ME | Duplodnaviria | Heunggongvirae | Uroviricota | Caudoviricetes | n.a. | Unclassified | O | O | 20.03 | d__Bacteria;p__Firmicutes;c__Clostridia;o__Christensenellales;f__NSJ-53;g__NSJ-53 | 94 | blast;95.80 iPHoP-RF;60.10 |
| S5_vs_145598 | ME | Duplodnaviria | Heunggongvirae | Uroviricota | Caudoviricetes | n.a. | Unclassified | O | O | 95.83 | d__Bacteria;p__Firmicutes;c__Clostridia;o__Lachnospirales;f__Lachnospiraceae;g__Roseburia | 92.1 | blast;94.30 iPHoP-RF;91.80 |
| S5_vs_145602 | ME | Duplodnaviria | Heunggongvirae | Uroviricota | Caudoviricetes | n.a. | Unclassified | O | O | 37.74 | d__Bacteria;p__Firmicutes;c__Clostridia;o__Lachnospirales;f__Lachnospiraceae;g__Anaerobutyricum | 90.7 | blast;93.10 iPHoP-RF;65.30 |
| S5_vs_145605 | ME | Duplodnaviria | Heunggongvirae | Uroviricota | Caudoviricetes | n.a. | Unclassified | O | O | 17.51 | d__Bacteria;p__Firmicutes;c__Clostridia;o__Christensenellales;f__CAG-74;g__SFTH01 | 90.3 | blast;92.80 iPHoP-RF;68.20 |
| S5_vs_145611 | ME | Duplodnaviria | Heunggongvirae | Uroviricota | Caudoviricetes | n.a. | Unclassified | O | O | 45.88 | d__Bacteria;p__Firmicutes;c__Clostridia;o__Oscillospirales;f__Ruminococcaceae;g__Phocea | 90.7 | blast;93.10 iPHoP-RF;78.00 |
| S5_vs_1572 | Shared | Duplodnaviria | Heunggongvirae | Uroviricota | Caudoviricetes | n.a. | Unclassified | Unclassified | Unclassified | 97.34 | d__Bacteria;p__Bacteroidota;c__Bacteroidia;o__Bacteroidales;f__Tannerellaceae;g__Parabacteroides | 91.6 | blast;93.90 iPHoP-RF;61.70 |
| S5_vs_17520 | ME | n.a. | n.a. | n.a. | n.a. | n.a. | n.a. | n.a. | n.a. | 39.12 | d__Bacteria;p__Firmicutes;c__Clostridia;o__Lachnospirales;f__Lachnospiraceae;g__Hungatella | 95.9 | iPHoP-RF;97.10 CRISPR;88.40 |
| S5_vs_20268 | ME | Duplodnaviria | Heunggongvirae | Uroviricota | Caudoviricetes | n.a. | Unclassified | Unclassified | Unclassified | 22.4 | d__Bacteria;p__Firmicutes;c__Clostridia;o__Oscillospirales;f__Acutalibacteraceae;g__Ruminococcus_E | 98.5 | CRISPR;98.90 blast;96.50 iPHoP-RF;89.50 |
| S5_vs_20500 | Shared | Duplodnaviria | Heunggongvirae | Uroviricota | Caudoviricetes | n.a. | Unclassified | O | O | 99.42 | d__Bacteria;p__Bacteroidota;c__Bacteroidia;o__Bacteroidales;f__Rikenellaceae;g__Alistipes | 95.6 | blast;96.90 iPHoP-RF;96.70 |
| S5_vs_21778 | ME | n.a. | n.a. | n.a. | n.a. | n.a. | n.a. | n.a. | n.a. | 11.27 | d__Bacteria;p__Firmicutes;c__Clostridia;o__Oscillospirales;f__Oscillospiraceae;g__Flavonifractor | 90.4 | blast;92.90 iPHoP-RF;59.20 |
| S5_vs_28263 | ME | n.a. | n.a. | n.a. | n.a. | n.a. | n.a. | n.a. | n.a. | 16.17 | d__Bacteria;p__Firmicutes;c__Clostridia;o__Lachnospirales;f__Lachnospiraceae;g__Mediterraneibacter | 91.6 | blast;93.90 iPHoP-RF;86.30 CRISPR;58.00 |
| S5_vs_30490 | ME | n.a. | n.a. | n.a. | n.a. | n.a. | n.a. | n.a. | n.a. | 42.23 | d__Bacteria;p__Firmicutes;c__Clostridia;o__Oscillospirales;f__Ruminococcaceae;g__Faecalibacterium | 99.3 | blast;97.10 RaFAH;90.30 iPHoP-RF;87.30 CRISPR;71.60 |
| S5_vs_32885 | ME | Duplodnaviria | Heunggongvirae | Uroviricota | Caudoviricetes | n.a. | Unclassified | O | O | 19.8 | d__Bacteria;p__Firmicutes;c__Clostridia;o__Oscillospirales;f__Ruminococcaceae;g__Anaerotruncus | 92.3 | blast;94.50 CRISPR;93.00 iPHoP-RF;81.90 |
| S5_vs_3501 | ME | Duplodnaviria | Heunggongvirae | Uroviricota | Caudoviricetes | n.a. | Unclassified | O | O | 32.19 | d__Bacteria;p__Firmicutes;c__Clostridia;o__Oscillospirales;f__Acutalibacteraceae;g__UBA1691 | 95.6 | blast;96.90 iPHoP-RF;90.10 CRISPR;85.90 |
| S5_vs_40848 | ME | Duplodnaviria | Heunggongvirae | Uroviricota | Caudoviricetes | n.a. | Unclassified | O | O | 58.74 | d__Bacteria;p__Firmicutes;c__Clostridia;o__Oscillospirales;f__Acutalibacteraceae;g__Ruminococcus_E | 95.6 | blast;96.90 iPHoP-RF;92.80 |
| S5_vs_43207 | ME | Duplodnaviria | Heunggongvirae | Uroviricota | Caudoviricetes | n.a. | Unclassified | O | O | 30.09 | d__Bacteria;p__Firmicutes;c__Clostridia;o__Oscillospirales;f__Butyricicoccaceae;g__Agathobaculum | 91.1 | blast;93.50 iPHoP-RF;70.60 |
| S5_vs_46394 | ME | n.a. | n.a. | n.a. | n.a. | n.a. | n.a. | n.a. | n.a. | 8.87 | d__Bacteria;p__Firmicutes;c__Clostridia;o__Christensenellales;f__QAND01;g__UMGS1975 | 90.3 | blast;92.80 iPHoP-RF;62.70 |
| S5_vs_4737 | ME | n.a. | n.a. | n.a. | n.a. | n.a. | n.a. | n.a. | n.a. | 20.64 | d__Bacteria;p__Firmicutes;c__Clostridia;o__Christensenellales;f__CAG-74;g__Limiplasma | 93.9 | blast;95.70 iPHoP-RF;68.40 |
| S5_vs_51416 | ME | Duplodnaviria | Heunggongvirae | Uroviricota | Caudoviricetes | n.a. | Unclassified | O | O | 11.18 | d__Bacteria;p__Firmicutes;c__Bacilli;o__Erysipelotrichales;f__Erysipelotrichaceae;g__Holdemania | 90.3 | blast;92.80 CRISPR;73.80 iPHoP-RF;66.50 |
| S5_vs_58766 | ME | Duplodnaviria | Heunggongvirae | Uroviricota | Caudoviricetes | n.a. | Unclassified | O | O | 24.99 | d__Bacteria;p__Firmicutes;c__Clostridia;o__Lachnospirales;f__Lachnospiraceae;g__Frisingicoccus | 93.8 | blast;95.60 iPHoP-RF;76.60 |
| S5_vs_59798 | ME | Duplodnaviria | Heunggongvirae | Uroviricota | Caudoviricetes | n.a. | Unclassified | O | O | 40.34 | d__Bacteria;p__Firmicutes;c__Clostridia;o__Lachnospirales;f__Lachnospiraceae;g__Faecalimonas | 91.4 | blast;93.70 iPHoP-RF;91.10 CRISPR;78.90 |
| S5_vs_60507 | ME | Duplodnaviria | Heunggongvirae | Uroviricota | Caudoviricetes | n.a. | Unclassified | Unclassified | O | 96.47 | d__Bacteria;p__Firmicutes;c__Bacilli;o__Lactobacillales;f__Streptococcaceae;g__Streptococcus | 100 | RaFAH;99.70 blast;92.80 iPHoP-RF;91.10 CRISPR;52.30 |
| S5_vs_63924 | Shared | Duplodnaviria | Heunggongvirae | Uroviricota | Caudoviricetes | n.a. | Unclassified | Unclassified | Unclassified | 98.89 | d__Bacteria;p__Actinobacteriota;c__Actinomycetia;o__Actinomycetales;f__Bifidobacteriaceae;g__Bifidobacterium | 99 | CRISPR;97.70 iPHoP-RF;96.70 blast;95.40 RaFAH;82.40 |
| S5_vs_6478 | ME | Duplodnaviria | Heunggongvirae | Uroviricota | Caudoviricetes | n.a. | Unclassified | Unclassified | O | 66.33 | d__Bacteria;p__Firmicutes;c__Bacilli;o__Lactobacillales;f__Lactobacillaceae;g__Lactobacillus | 100 | RaFAH;99.60 iPHoP-RF;96.70 blast;95.40 CRISPR;68.80 |
| S5_vs_65490 | ME | n.a. | n.a. | n.a. | n.a. | n.a. | n.a. | n.a. | n.a. | 3.66 | d__Bacteria;p__Firmicutes;c__Clostridia;o__Oscillospirales;f__Oscillospiraceae;g__Flavonifractor | 91.2 | blast;93.60 iPHoP-RF;56.20 |

| | | | | | | | | | | | | | |
|---|---|---|---|---|---|---|---|---|---|---|---|---|---|
| S5_vs_68254 | ME | Duplodnaviria | Heunggongvirae | Uroviricota | Caudoviricetes | n.a. | Unclassified | O | O | 58.14 | d__Bacteria;p__Firmicutes;c__Clostridia;o__Lachnospirales;f__Lachnospiraceae;g__Blautia_A | 93.8 | blast;93.80 iPHoP-RF;81.90 RaFAH;64.30 |
| S5_vs_69980 | ME | Duplodnaviria | Heunggongvirae | Uroviricota | Caudoviricetes | n.a. | Unclassified | O | O | 76.63 | d__Bacteria;p__Firmicutes;c__Clostridia;o__Oscillospirales;f__Oscillospiraceae;g__Flavonifractor | 95.3 | blast;96.70 iPHoP-RF;96.40 CRISPR;95.20 |
| S5_vs_72532 | ME | Duplodnaviria | Heunggongvirae | Uroviricota | Caudoviricetes | n.a. | Unclassified | O | O | 82.39 | d__Bacteria;p__Bacteroidota;c__Bacteroidia;o__Bacteroidales;f__Bacteroidaceae;g__Bacteroides | 99.4 | blast;96.90 iPHoP-RF;96.70 RaFAH;92.20 |
| S5_vs_74199 | ME | n.a. | n.a. | n.a. | n.a. | n.a. | n.a. | n.a. | n.a. | 13.11 | d__Bacteria;p__Firmicutes;c__Clostridia;o__Oscillospirales;f__Oscillospiraceae;g__CAG-83 | 90.4 | blast;92.90 iPHoP-RF;92.10 |
| S5_vs_76500 | Shared | Duplodnaviria | Heunggongvirae | Uroviricota | Caudoviricetes | n.a. | Unclassified | Unclassified | Unclassified | 80.28 | d__Bacteria;p__Bacteroidota;c__Bacteroidia;o__Bacteroidales;f__Rikenellaceae;g__Alistipes | 99 | blast;96.90 iPHoP-RF;94.40 RaFAH;87.40 |
| S5_vs_77593 | ME | Duplodnaviria | Heunggongvirae | Uroviricota | Caudoviricetes | n.a. | Unclassified | O | O | 41.43 | d__Bacteria;p__Firmicutes;c__Clostridia;o__Oscillospirales;f__Oscillospiraceae;g__CAG-103 | 90.6 | blast;93.00 iPHoP-RF;71.70 |
| S5_vs_79992 | Shared | Duplodnaviria | Heunggongvirae | Uroviricota | Caudoviricetes | n.a. | Winoviridae | O | O | 99.98 | d__Bacteria;p__Bacteroidota;c__Bacteroidia;o__Bacteroidales;f__Rikenellaceae;g__Alistipes | 100 | RaFAH;98.20 blast;97.10 iPHoP-RF;96.70 |
| S5_vs_80496 | Shared | n.a. | n.a. | n.a. | n.a. | n.a. | n.a. | n.a. | n.a. | 99.85 | d__Bacteria;p__Bacteroidota;c__Bacteroidia;o__Bacteroidales;f__Bacteroidaceae;g__Bacteroides | 99.7 | blast;96.90 iPHoP-RF;96.10 RaFAH;95.90 |
| S5_vs_80498 | ME | Duplodnaviria | Heunggongvirae | Uroviricota | Caudoviricetes | n.a. | Unclassified | O | O | 33.04 | d__Bacteria;p__Firmicutes;c__Clostridia;o__Lachnospirales;f__Lachnospiraceae;g__Muricomes | 93.2 | blast;95.20 iPHoP-RF;90.50 |
| S5_vs_8125 | ME | Duplodnaviria | Heunggongvirae | Uroviricota | Caudoviricetes | n.a. | Unclassified | O | O | 19.38 | d__Bacteria;p__Firmicutes;c__Clostridia;o__Lachnospirales;f__Lachnospiraceae;g__Eubacterium_G | 90.6 | blast;93.00 iPHoP-RF;53.30 |
| S5_vs_84574 | ME | n.a. | n.a. | n.a. | n.a. | n.a. | n.a. | n.a. | n.a. | 8.38 | d__Bacteria;p__Firmicutes;c__Clostridia;o__Oscillospirales;f__Butyricicoccaceae;g__Agathobaculum | 91.6 | blast;93.90 iPHoP-RF;81.10 |
| S5_vs_84946 | ME | Duplodnaviria | Heunggongvirae | Uroviricota | Caudoviricetes | n.a. | Unclassified | O | O | 94.46 | d__Bacteria;p__Firmicutes;c__Clostridia;o__Lachnospirales;f__Lachnospiraceae;g__Dorea_A | 90.2 | blast;92.70 iPHoP-RF;83.60 |
| S5_vs_850 | ME | n.a. | n.a. | n.a. | n.a. | n.a. | n.a. | n.a. | n.a. | 18.78 | d__Bacteria;p__Firmicutes;c__Clostridia;o__UBA1212;f__UBA1255;g__Egerieisoma | 92.7 | blast;94.80 iPHoP-RF;85.10 |
| S5_vs_85065 | Shared | Duplodnaviria | Heunggongvirae | Uroviricota | Caudoviricetes | n.a. | Unclassified | O | O | 67.47 | d__Bacteria;p__Firmicutes;c__Clostridia;o__Oscillospirales;f__Ruminococcaceae;g__Ruthenibacterium | 95.3 | blast;96.70 |
| S5_vs_86562 | ME | Duplodnaviria | Heunggongvirae | Uroviricota | Caudoviricetes | n.a. | Unclassified | O | O | 44.1 | d__Bacteria;p__Verrucomicrobiota;c__Verrucomicrobiae;o__Verrucomicrobiales;f__Akkermansiaceae;g__Akkermansia | 100 | RaFAH;99.10 CRISPR;98.80 blast;96.90 iPHoP-RF;89.20 |
| S5_vs_89540 | ME | Duplodnaviria | Heunggongvirae | Uroviricota | Caudoviricetes | n.a. | Unclassified | O | O | 26.84 | d__Bacteria;p__Firmicutes;c__Bacilli;o__Lactobacillales;f__Lactobacillaceae;g__Limosilactobacillus | 94.2 | blast;93.40 iPHoP-RF;81.90 RaFAH;68.80 |
| S5_vs_93839 | Shared | Duplodnaviria | Heunggongvirae | Uroviricota | Caudoviricetes | n.a. | Unclassified | O | O | 98.83 | d__Bacteria;p__Bacteroidota;c__Bacteroidia;o__Bacteroidales;f__Rikenellaceae;g__Alistipes | 100 | RaFAH;98.90 iPHoP-RF;96.70 blast;95.50 |
| S5_vs_95307 | ME | Duplodnaviria | Heunggongvirae | Uroviricota | Caudoviricetes | n.a. | Unclassified | Unclassified | O | 57.8 | d__Bacteria;p__Firmicutes;c__Clostridia;o__Lachnospirales;f__Lachnospiraceae;g__Blautia | 92 | blast;91.60 iPHoP-RF;73.20 RaFAH;65.20 |
| S5_vs_9747 | Shared | n.a. | n.a. | n.a. | n.a. | n.a. | n.a. | n.a. | n.a. | 6.96 | d__Bacteria;p__Proteobacteria;c__Alphaproteobacteria;o__RF32;f__CAG-239;g__CAG-267 | 93 | blast;95.00 iPHoP-RF;69.80 |
| S5_vs_97574 | Shared | Duplodnaviria | Heunggongvirae | Uroviricota | Caudoviricetes | n.a. | Unclassified | O | O | 99.69 | d__Bacteria;p__Firmicutes;c__Clostridia;o__Lachnospirales;f__Lachnospiraceae;g__Blautia_A | 100 | RaFAH;99.30 CRISPR;95.90 blast;93.60 iPHoP-RF;92.10 |
| S6_vf_25710 | Shared | n.a. | n.a. | n.a. | n.a. | n.a. | n.a. | n.a. | n.a. | 89.8 | d__Bacteria;p__Verrucomicrobiota;c__Verrucomicrobiae;o__Verrucomicrobiales;f__Akkermansiaceae;g__Akkermansia | 99.7 | CRISPR;97.30 RaFAH;94.60 blast;93.60 iPHoP-RF;72.60 |
| S6_vf_56486 | Shared | Duplodnaviria | Heunggongvirae | Uroviricota | Caudoviricetes | n.a. | Unclassified | O | O | 65.79 | d__Bacteria;p__Bacteroidota;c__Bacteroidia;o__Bacteroidales;f__Bacteroidaceae;g__Bacteroides | 99.6 | iPHoP-RF;96.10 RaFAH;94.70 |
| S6_vs_1146 | Shared | Duplodnaviria | Heunggongvirae | Uroviricota | Caudoviricetes | n.a. | Unclassified | O | O | 50.09 | d__Bacteria;p__Firmicutes;c__Clostridia;o__Oscillospirales;f__Ruminococcaceae;g__Faecalibacterium | 98.4 | blast;96.90 iPHoP-RF;86.00 RaFAH;81.40 |
| S6_vs_12945 | Shared | Duplodnaviria | Heunggongvirae | Uroviricota | Caudoviricetes | n.a. | Unclassified | O | O | 99.74 | d__Bacteria;p__Firmicutes;c__Clostridia;o__Lachnospirales;f__Lachnospiraceae;g__Roseburia | 99.3 | RaFAH;94.70 blast;88.50 iPHoP-RF;74.80 |
| S6_vs_15390 | Shared | Duplodnaviria | Heunggongvirae | Uroviricota | Caudoviricetes | n.a. | Unclassified | O | O | 33.39 | d__Bacteria;p__Bacteroidota;c__Bacteroidia;o__Bacteroidales;f__Tannerellaceae;g__Parabacteroides | 90.8 | blast;93.20 iPHoP-RF;92.10 |
| S6_vs_15813 | Shared | n.a. | n.a. | n.a. | n.a. | n.a. | n.a. | n.a. | n.a. | 90.87 | d__Bacteria;p__Firmicutes;c__Clostridia;o__Oscillospirales;f__Oscillospiraceae;g__Flavonifractor | 99.3 | blast;98.10 RaFAH;85.50 iPHoP-RF;85.10 CRISPR;56.80 |
| S6_vs_29152 | SHHC | Duplodnaviria | Heunggongvirae | Uroviricota | Caudoviricetes | n.a. | Unclassified | O | O | 23.31 | d__Bacteria;p__Firmicutes;c__Clostridia;o__Lachnospirales;f__Lachnospiraceae;g__Acetatifactor | 91.2 | blast;93.60 |
| S6_vs_32886 | Shared | n.a. | n.a. | n.a. | n.a. | n.a. | n.a. | n.a. | n.a. | 6.93 | d__Bacteria;p__Bacteroidota;c__Bacteroidia;o__Bacteroidales;f__Rikenellaceae;g__Alistipes_A | 94.2 | blast;95.90 iPHoP-RF;66.30 |
| S6_vs_33711 | SHHC | Duplodnaviria | Heunggongvirae | Uroviricota | Caudoviricetes | n.a. | Unclassified | Unclassified | Unclassified | 86.98 | d__Bacteria;p__Firmicutes;c__Clostridia;o__Oscillospirales;f__Ruminococcaceae;g__Ruthenibacterium | 94 | blast;95.80 iPHoP-RF;85.40 CRISPR;69.40 |
| S6_vs_34453 | Shared | Duplodnaviria | Heunggongvirae | Uroviricota | Caudoviricetes | n.a. | Unclassified | Unclassified | O | 77 | d__Bacteria;p__Firmicutes;c__Clostridia;o__Oscillospirales;f__Ruminococcaceae;g__Faecalibacterium | 93.6 | blast;95.50 iPHoP-RF;64.30 |
| S6_vs_38250 | SHHC | Duplodnaviria | Heunggongvirae | Uroviricota | Caudoviricetes | n.a. | Unclassified | Unclassified | Unclassified | 15.25 | d__Bacteria;p__Firmicutes;c__Clostridia;o__Oscillospirales;f__Ruminococcaceae;g__UBA3818 | 94.3 | blast;96.00 iPHoP-RF;76.10 |
| S6_vs_39025 | Shared | Duplodnaviria | Heunggongvirae | Uroviricota | Caudoviricetes | n.a. | Unclassified | O | O | 99.25 | d__Bacteria;p__Firmicutes;c__Clostridia;o__Lachnospirales;f__Lachnospiraceae;g__Enterocloster | 91.6 | blast;93.90 |
| S6_vs_39065 | Shared | n.a. | n.a. | n.a. | n.a. | n.a. | n.a. | n.a. | n.a. | 83.64 | d__Bacteria;p__Proteobacteria;c__Alphaproteobacteria;o__RF32;f__CAG-239;g__CAG-267 | 94.6 | blast;96.20 iPHoP-RF;75.20 |
| S6_vs_44070 | Shared | Unclassified | n.a. | n.a. | n.a. | n.a. | Unclassified | O | O | 14.4 | d__Bacteria;p__Proteobacteria;c__Alphaproteobacteria;o__RF32;f__CAG-239;g__51-20 | 94.7 | blast;96.30 iPHoP-RF;72.60 |
| S6_vs_46039 | Shared | n.a. | n.a. | n.a. | n.a. | n.a. | n.a. | n.a. | n.a. | 24.66 | d__Bacteria;p__Bacteroidota;c__Bacteroidia;o__Bacteroidales;f__Bacteroidaceae;g__Bacteroides | 93 | CRISPR;93.00 blast;89.60 iPHoP-RF;82.50 RaFAH;64.10 |
| S6_vs_46091 | Shared | Duplodnaviria | Heunggongvirae | Uroviricota | Caudoviricetes | n.a. | Unclassified | Unclassified | O | 25.96 | d__Bacteria;p__Firmicutes;c__Clostridia;o__Oscillospirales;f__Ruminococcaceae;g__Negativibacillus | 96.6 | blast;97.60 iPHoP-RF;83.30 CRISPR;69.80 |
| S6_vs_53966 | SHHC | Duplodnaviria | Heunggongvirae | Uroviricota | Caudoviricetes | n.a. | Unclassified | O | O | 51.17 | d__Bacteria;p__Firmicutes;c__Clostridia;o__Oscillospirales;f__Ruminococcaceae;g__Faecalibacterium | 98.7 | blast;96.30 iPHoP-RF;90.50 RaFAH;86.50 |
| S6_vs_54606 | Shared | Duplodnaviria | Heunggongvirae | Uroviricota | Caudoviricetes | n.a. | Unclassified | O | O | 99.52 | d__Bacteria;p__Firmicutes;c__Clostridia;o__Lachnospirales;f__Lachnospiraceae;g__Blautia | 97 | RaFAH;91.10 blast;88.40 |
| S6_vs_56485 | Shared | Duplodnaviria | Heunggongvirae | Uroviricota | Caudoviricetes | n.a. | Unclassified | O | O | 14.92 | d__Bacteria;p__Firmicutes;c__Bacilli;o__Erysipelotrichales;f__Erysipelotrichaceae;g__Holdemania | 94.7 | CRISPR;96.30 blast;94.30 iPHoP-RF;79.80 |
| S6_vs_56490 | Shared | Duplodnaviria | Heunggongvirae | Uroviricota | Caudoviricetes | n.a. | Unclassified | O | O | 26.23 | d__Bacteria;p__Firmicutes;c__Clostridia;o__Oscillospirales;f__Acutalibacteraceae;g__UMGS1071 | 95.9 | blast;97.10 iPHoP-RF;83.90 CRISPR;68.20 |
| S6_vs_56491 | Shared | Duplodnaviria | Heunggongvirae | Uroviricota | Caudoviricetes | n.a. | Unclassified | O | O | 92.92 | d__Bacteria;p__Firmicutes;c__Clostridia;o__Lachnospirales;f__Lachnospiraceae;g__Sellimonas | 97.5 | blast;98.20 iPHoP-RF;75.20 |
| S6_vs_56492 | Shared | n.a. | n.a. | n.a. | n.a. | n.a. | n.a. | n.a. | n.a. | 27.91 | d__Bacteria;p__Firmicutes;c__Clostridia;o__Oscillospirales;f__Butyricicoccaceae;g__Agathobaculum | 98.2 | blast;98.70 iPHoP-RF;66.80 |
| S6_vs_56496 | Shared | Duplodnaviria | Heunggongvirae | Uroviricota | Caudoviricetes | n.a. | Unclassified | Unclassified | Eponavirus | 61.48 | d__Bacteria;p__Firmicutes;c__Clostridia;o__Lachnospirales;f__Lachnospiraceae;g__Marvinbryantia | 91.2 | blast;93.60 iPHoP-RF;65.10 |
| S6_vs_56498 | Shared | Duplodnaviria | Heunggongvirae | Uroviricota | Caudoviricetes | n.a. | Unclassified | O | O | 78.58 | d__Bacteria;p__Firmicutes;c__Clostridia;o__Lachnospirales;f__Lachnospiraceae;g__Muricomes | 91 | blast;93.40 iPHoP-RF;69.80 |
| S6_vs_9784 | Shared | n.a. | n.a. | n.a. | n.a. | n.a. | n.a. | n.a. | n.a. | 91.55 | d__Bacteria;p__Bacteroidota;c__Bacteroidia;o__Bacteroidales;f__Bacteroidaceae;g__Bacteroides | 100 | RaFAH;97.10 iPHoP-RF;96.70 blast;93.90 |
| S7_vf_114476 | ME | Duplodnaviria | Heunggongvirae | Uroviricota | Caudoviricetes | n.a. | Unclassified | O | O | 27.19 | d__Bacteria;p__Firmicutes;c__Clostridia;o__Lachnospirales;f__Lachnospiraceae;g__Anaerobutyricum | 94.3 | blast;96.00 iPHoP-RF;74.30 |
| S7_vf_117697 | Shared | n.a. | n.a. | n.a. | n.a. | n.a. | n.a. | n.a. | n.a. | 98.64 | d__Bacteria;p__Bacteroidota;c__Bacteroidia;o__Bacteroidales;f__Rikenellaceae;g__Alistipes | 96.9 | CRISPR;97.80 iPHoP-RF;97.10 blast;96.80 |
| S7_vf_134023 | ME | n.a. | n.a. | n.a. | n.a. | n.a. | n.a. | n.a. | n.a. | 12.46 | d__Bacteria;p__Bacteroidota;c__Bacteroidia;o__Bacteroidales;f__Rikenellaceae;g__Alistipes_A | 95 | blast;96.50 iPHoP-RF;54.40 |
| S7_vf_13614 | Shared | Duplodnaviria | Heunggongvirae | Uroviricota | Caudoviricetes | n.a. | Unclassified | O | O | 39.23 | d__Bacteria;p__Firmicutes;c__Clostridia;o__Oscillospirales;f__Ruminococcaceae;g__Ruminococcus_D | 95.4 | blast;96.80 iPHoP-RF;95.40 CRISPR;50.20 |
| S7_vf_138019 | ME | n.a. | n.a. | n.a. | n.a. | n.a. | n.a. | n.a. | n.a. | 8.23 | d__Bacteria;p__Firmicutes;c__Clostridia;o__Oscillospirales;f__Oscillospiraceae;g__Lawsonibacter | 91.1 | blast;93.50 iPHoP-RF;50.40 |
| S7_vf_138069 | ME | n.a. | n.a. | n.a. | n.a. | n.a. | n.a. | n.a. | n.a. | 78.93 | d__Bacteria;p__Firmicutes;c__Bacilli;o__Lactobacillales;f__Streptococcaceae;g__Streptococcus | 99.6 | RaFAH;99.40 iPHoP-RF;67.30 |
| S7_vf_27309 | ME | Duplodnaviria | Heunggongvirae | Uroviricota | Caudoviricetes | n.a. | Unclassified | Unclassified | O | 18.88 | d__Bacteria;p__Bacteroidota;c__Bacteroidia;o__Bacteroidales;f__Bacteroidaceae;g__Bacteroides | 96.3 | CRISPR;97.40 iPHoP-RF;93.70 |
| S7_vf_28429 | Shared | Duplodnaviria | Heunggongvirae | Uroviricota | Caudoviricetes | n.a. | Unclassified | O | O | 86.01 | d__Bacteria;p__Bacteroidota;c__Bacteroidia;o__Bacteroidales;f__Tannerellaceae;g__Parabacteroides | 100 | CRISPR;99.00 RaFAH;98.30 iPHoP-RF;94.10 blast;93.90 |
| S7_vf_29941 | Shared | Duplodnaviria | Heunggongvirae | Uroviricota | Caudoviricetes | n.a. | Unclassified | O | O | 79.47 | d__Bacteria;p__Firmicutes;c__Bacilli;o__Lactobacillales;f__Streptococcaceae;g__Lactococcus | 99.4 | blast;97.20 RaFAH;91.90 iPHoP-RF;73.90 |
| S7_vf_37279 | Shared | Duplodnaviria | Heunggongvirae | Uroviricota | Caudoviricetes | n.a. | Unclassified | O | O | 64.97 | d__Bacteria;p__Bacteroidota;c__Bacteroidia;o__Bacteroidales;f__Bacteroidaceae;g__Bacteroides | 99.7 | CRISPR;98.80 iPHoP-RF;97.10 blast;90.80 RaFAH;87.70 |
| S7_vf_37293 | ME | Duplodnaviria | Heunggongvirae | Uroviricota | Caudoviricetes | n.a. | Unclassified | O | O | 98.54 | d__Bacteria;p__Firmicutes;c__Bacilli;o__Lactobacillales;f__Streptococcaceae;g__Streptococcus | 100 | RaFAH;99.60 CRISPR;99.00 iPHoP-RF;54.10 |
| S7_vf_38697 | ME | Duplodnaviria | Heunggongvirae | Uroviricota | Caudoviricetes | n.a. | Unclassified | O | O | 61.28 | d__Bacteria;p__Firmicutes;c__Bacilli;o__Erysipelotrichales;f__Erysipelatoclostridiaceae;g__Massiliomicrobiota | 94.2 | blast;95.90 iPHoP-RF;74.30 |
| S7_vf_39724 | Shared | Duplodnaviria | Heunggongvirae | Uroviricota | Caudoviricetes | n.a. | Unclassified | Unclassified | Mushuvirus | 99.39 | d__Bacteria;p__Firmicutes;c__Clostridia;o__Oscillospirales;f__Ruminococcaceae;g__Faecalibacterium | 99.4 | RaFAH;99.50 |
| S7_vf_49007 | ME | Duplodnaviria | Heunggongvirae | Uroviricota | Caudoviricetes | n.a. | Unclassified | O | O | 85 | d__Bacteria;p__Firmicutes;c__Bacilli;o__Lactobacillales;f__Streptococcaceae;g__Streptococcus | 100 | CRISPR;98.70 RaFAH;97.80 iPHoP-RF;68.00 |
| S7_vf_49485 | ME | Duplodnaviria | Heunggongvirae | Uroviricota | Caudoviricetes | n.a. | Unclassified | O | O | 64.81 | d__Bacteria;p__Firmicutes;c__Clostridia;o__Lachnospirales;f__Lachnospiraceae;g__Anaerostipes | 98.5 | CRISPR;98.90 iPHoP-RF;56.20 |
| S7_vf_5801 | Shared | Duplodnaviria | Heunggongvirae | Uroviricota | Caudoviricetes | n.a. | Unclassified | O | O | 93.48 | d__Bacteria;p__Bacteroidota;c__Bacteroidia;o__Bacteroidales;f__Bacteroidaceae;g__Bacteroides | 100 | RaFAH;96.60 iPHoP-RF;96.40 blast;91.00 |
| S7_vf_5843 | ME | Duplodnaviria | Heunggongvirae | Uroviricota | Caudoviricetes | n.a. | Unclassified | O | O | 19.99 | d__Bacteria;p__Actinobacteriota;c__Actinomycetia;o__Actinomycetales;f__Bifidobacteriaceae;g__Bifidobacterium | 96.3 | blast;97.40 CRISPR;97.30 iPHoP-RF;93.40 |
| S7_vf_60968 | ME | Duplodnaviria | Heunggongvirae | Uroviricota | Caudoviricetes | n.a. | Unclassified | O | O | 71.38 | d__Bacteria;p__Firmicutes;c__Clostridia;o__Lachnospirales;f__Lachnospiraceae;g__Ruminococcus_B | 97.8 | blast;98.40 iPHoP-RF;81.40 CRISPR;66.90 |
| S7_vf_63894 | ME | Duplodnaviria | Heunggongvirae | Uroviricota | Caudoviricetes | n.a. | Unclassified | Mccleskeyvirinae | Unaquatrovirus | 77.84 | d__Bacteria;p__Firmicutes;c__Bacilli;o__Lactobacillales;f__Lactobacillaceae;g__Leuconostoc | 96.2 | iPHoP-RF;93.70 RaFAH;78.60 |
| S7_vf_67910 | ME | n.a. | n.a. | n.a. | n.a. | n.a. | n.a. | n.a. | n.a. | 11.86 | d__Bacteria;p__Actinobacteriota;c__Coriobacteriia;o__Coriobacteriales;f__Eggerthellaceae;g__Eggerthella | 94.5 | blast;96.10 |
| S7_vf_68239 | ME | Duplodnaviria | Heunggongvirae | Uroviricota | Caudoviricetes | n.a. | Unclassified | O | O | 31.22 | d__Bacteria;p__Firmicutes;c__Bacilli;o__Lactobacillales;f__Streptococcaceae;g__Lactococcus | 93.1 | RaFAH;95.10 |
| S7_vf_68546 | ME | Duplodnaviria | Heunggongvirae | Uroviricota | Caudoviricetes | n.a. | Unclassified | O | O | 29.45 | d__Bacteria;p__Firmicutes;c__Clostridia;o__Lachnospirales;f__Lachnospiraceae;g__Faecalimonas | 94.5 | CRISPR;96.10 iPHoP-RF;76.10 |
| S7_vf_71527 | ME | n.a. | n.a. | n.a. | n.a. | n.a. | n.a. | n.a. | n.a. | 3.4 | d__Bacteria;p__Firmicutes;c__Clostridia;o__Oscillospirales;f__Ruminococcaceae;g__Ruminococcus_D | 93.1 | iPHoP-RF;95.10 blast;94.30 CRISPR;92.20 |
| S7_vf_72493 | ME | n.a. | n.a. | n.a. | n.a. | n.a. | n.a. | n.a. | n.a. | 13.24 | d__Bacteria;p__Firmicutes;c__Clostridia;o__Lachnospirales;f__Anaerotignaceae;g__UMGS1670 | 92.5 | blast;94.60 iPHoP-RF;65.10 |
| S7_vf_76463 | Shared | n.a. | n.a. | n.a. | n.a. | n.a. | n.a. | n.a. | n.a. | 42.93 | d__Bacteria;p__Firmicutes;c__Clostridia;o__Oscillospirales;f__Oscillospiraceae;g__CAG-103 | 95.3 | CRISPR;96.70 |
| S7_vf_8332 | ME | Duplodnaviria | Heunggongvirae | Uroviricota | Caudoviricetes | n.a. | Unclassified | O | O | 10.6 | d__Bacteria;p__Firmicutes;c__Clostridia;o__Oscillospirales;f__Ruminococcaceae;g__Ruminococcus_D | 98.5 | CRISPR;98.90 blast;96.20 iPHoP-RF;94.70 |
| S7_vf_84974 | ME | Duplodnaviria | Heunggongvirae | Uroviricota | Caudoviricetes | n.a. | Unclassified | O | O | 91.7 | d__Bacteria;p__Firmicutes;c__Clostridia;o__Oscillospirales;f__Ruminococcaceae;g__Faecalibacterium | 99.9 | RaFAH;98.40 blast;92.10 iPHoP-RF;63.30 CRISPR;56.10 |
| S7_vf_89197 | Shared | Duplodnaviria | Heunggongvirae | Uroviricota | Caudoviricetes | n.a. | Unclassified | Unclassified | Ceduovirus | 89.48 | d__Bacteria;p__Firmicutes;c__Bacilli;o__Lactobacillales;f__Streptococcaceae;g__Lactococcus | 96.6 | RaFAH;95.80 iPHoP-RF;71.50 |
| S7_vs_101751 | ME | n.a. | n.a. | n.a. | n.a. | n.a. | n.a. | n.a. | n.a. | 99.91 | d__Bacteria;p__Firmicutes;c__Clostridia;o__Lachnospirales;f__Lachnospiraceae;g__Blautia_A | 99.7 | RaFAH;94.40 blast;93.60 iPHoP-RF;76.80 |
| S7_vs_103530 | ME | n.a. | n.a. | n.a. | n.a. | n.a. | n.a. | n.a. | n.a. | 35.56 | d__Bacteria;p__Firmicutes;c__Clostridia;o__Oscillospirales;f__Butyricicoccaceae;g__Agathobaculum | 94.2 | blast;95.90 iPHoP-RF;60.30 |
| S7_vs_11081 | Shared | Duplodnaviria | Heunggongvirae | Uroviricota | Caudoviricetes | n.a. | Unclassified | Unclassified | Unclassified | 95.36 | d__Bacteria;p__Bacteroidota;c__Bacteroidia;o__Bacteroidales;f__Bacteroidaceae;g__Bacteroides | 100 | RaFAH;98.70 iPHoP-RF;94.70 blast;91.00 |
| S7_vs_111831 | ME | Duplodnaviria | Heunggongvirae | Uroviricota | Caudoviricetes | n.a. | Unclassified | O | O | 38.83 | d__Bacteria;p__Firmicutes;c__Clostridia;o__Oscillospirales;f__Ruminococcaceae;g__Gemmiger | 97.2 | blast;98.00 iPHoP-RF;88.90 CRISPR;82.40 |
| S7_vs_118295 | ME | n.a. | n.a. | n.a. | n.a. | n.a. | n.a. | n.a. | n.a. | 43.98 | d__Bacteria;p__Firmicutes;c__Clostridia;o__Oscillospirales;f__Oscillospiraceae;g__Intestinimonas | 91.7 | blast;94.00 iPHoP-RF;75.40 |
| S7_vs_119679 | Shared | Duplodnaviria | Heunggongvirae | Uroviricota | Caudoviricetes | n.a. | Winoviridae | O | O | 99.98 | d__Bacteria;p__Bacteroidota;c__Bacteroidia;o__Bacteroidales;f__Rikenellaceae;g__Alistipes | 100 | blast;98.70 RaFAH;98.60 iPHoP-RF;97.10 |
| S7_vs_129271 | ME | n.a. | n.a. | n.a. | n.a. | n.a. | n.a. | n.a. | n.a. | 12.57 | d__Bacteria;p__Firmicutes;c__Clostridia;o__Oscillospirales;f__Acutalibacteraceae;g__UBA1691 | 98.5 | CRISPR;98.90 blast;97.30 iPHoP-RF;82.80 |
| S7_vs_129858 | ME | n.a. | n.a. | n.a. | n.a. | n.a. | n.a. | n.a. | n.a. | 10.46 | d__Bacteria;p__Firmicutes;c__Clostridia;o__Christensenellales;f__CAG-74;g__Firm-11 | 91.4 | blast;93.70 iPHoP-RF;85.40 CRISPR;50.00 |
| S7_vs_13041 | ME | Duplodnaviria | Heunggongvirae | Uroviricota | Caudoviricetes | n.a. | Unclassified | O | O | 17.74 | d__Bacteria;p__Firmicutes;c__Bacilli;o__Erysipelotrichales;f__Erysipelatoclostridiaceae;g__Erysipelatoclostridium | 94.5 | blast;96.10 iPHoP-RF;74.60 CRISPR;54.40 |
| S7_vs_13370 | Shared | n.a. | n.a. | n.a. | n.a. | n.a. | n.a. | n.a. | n.a. | 8.9 | d__Bacteria;p__Firmicutes;c__Clostridia;o__Oscillospirales;f__Ruminococcaceae;g__Faecalibacterium | 97 | blast;97.90 iPHoP-RF;90.10 CRISPR;77.30 |
| S7_vs_134391 | ME | n.a. | n.a. | n.a. | n.a. | n.a. | n.a. | n.a. | n.a. | 17.86 | d__Bacteria;p__Firmicutes;c__Clostridia;o__Oscillospirales;f__UMGS1783;g__UMGS1783 | 94.9 | blast;96.40 iPHoP-RF;84.80 CRISPR;79.10 |
| S7_vs_134784 | ME | Duplodnaviria | Heunggongvirae | Uroviricota | Caudoviricetes | n.a. | Unclassified | O | O | 77.29 | d__Bacteria;p__Firmicutes;c__Negativicutes;o__Veillonellales;f__Dialisteraceae;g__Dialister | 97.3 | CRISPR;97.40 blast;96.60 iPHoP-RF;92.80 RaFAH;63.90 |
| S7_vs_135322 | Shared | Duplodnaviria | Heunggongvirae | Uroviricota | Caudoviricetes | n.a. | Unclassified | Unclassified | O | 34.72 | d__Bacteria;p__Firmicutes;c__Clostridia;o__Oscillospirales;f__Ruminococcaceae;g__Faecalibacterium | 99.7 | CRISPR;98.30 blast;91.90 RaFAH;90.70 iPHoP-RF;90.50 |
| S7_vs_135679 | ME | n.a. | n.a. | n.a. | n.a. | n.a. | n.a. | n.a. | n.a. | 23.81 | d__Bacteria;p__Firmicutes;c__Clostridia;o__Lachnospirales;f__Lachnospiraceae;g__Blautia_A | 99.6 | blast;96.50 RaFAH;94.00 iPHoP-RF;90.10 |
| S7_vs_135828 | ME | Monodnaviria | Sangervirae | Phixviricota | Malgrandaviricetes | Petitvirales | Microviridae | Unclassified | Unclassified | 3.2 | d__Bacteria;p__Bacteroidota;c__Bacteroidia;o__Bacteroidales;f__Bacteroidaceae;g__Bacteroides | 93 | blast;95.00 iPHoP-RF;86.00 |
| S7_vs_137611 | ME | Duplodnaviria | Heunggongvirae | Uroviricota | Caudoviricetes | n.a. | Unclassified | O | O | 29.3 | d__Bacteria;p__Firmicutes;c__Clostridia;o__Lachnospirales;f__Lachnospiraceae;g__Anaerobutyricum | 95 | blast;96.50 iPHoP-RF;87.60 |
| S7_vs_137915 | Shared | Duplodnaviria | Heunggongvirae | Uroviricota | Caudoviricetes | n.a. | Unclassified | O | O | 55.05 | d__Bacteria;p__Bacteroidota;c__Bacteroidia;o__Bacteroidales;f__Tannerellaceae;g__Parabacteroides | 98.1 | CRISPR;98.30 blast;90.90 iPHoP-RF;85.40 RaFAH;59.50 |
| S7_vs_13799 | ME | Duplodnaviria | Heunggongvirae | Uroviricota | Caudoviricetes | n.a. | Unclassified | O | O | 38.3 | d__Bacteria;p__Firmicutes;c__Clostridia;o__Lachnospirales;f__Lachnospiraceae;g__Dorea_A | 93 | blast;95.00 iPHoP-RF;94.70 CRISPR;72.50 |
| S7_vs_137998 | Shared | n.a. | n.a. | n.a. | n.a. | n.a. | n.a. | n.a. | n.a. | 42.1 | d__Bacteria;p__Firmicutes;c__Clostridia;o__Lachnospirales;f__Lachnospiraceae;g__Blautia_A | 93.4 | blast;95.30 iPHoP-RF;78.00 |
| S7_vs_138109 | ME | Duplodnaviria | Heunggongvirae | Uroviricota | Caudoviricetes | n.a. | Unclassified | O | O | 31.21 | d__Bacteria;p__Firmicutes;c__Clostridia;o__Lachnospirales;f__Lachnospiraceae;g__Anaerobutyricum | 91 | blast;93.40 iPHoP-RF;75.90 |
| S7_vs_138116 | ME | n.a. | n.a. | n.a. | n.a. | n.a. | n.a. | n.a. | n.a. | 5.66 | d__Bacteria;p__Firmicutes;c__Clostridia;o__Lachnospirales;f__Lachnospiraceae;g__Mediterraneibacter | 91.5 | blast;93.80 iPHoP-RF;89.50 |
| S7_vs_138173 | Shared | Duplodnaviria | Heunggongvirae | Uroviricota | Caudoviricetes | n.a. | Unclassified | O | O | 100 | d__Bacteria;p__Bacteroidota;c__Bacteroidia;o__Bacteroidales;f__Rikenellaceae;g__Alistipes | 100 | RaFAH;98.50 blast;94.70 iPHoP-RF;74.10 |
| S7_vs_16344 | Shared | Duplodnaviria | Heunggongvirae | Uroviricota | Caudoviricetes | n.a. | Unclassified | O | O | 12.56 | d__Bacteria;p__Firmicutes;c__Clostridia;o__Oscillospirales;f__Acutalibacteraceae;g__Ruminococcus_E | 98.8 | CRISPR;99.10 blast;96.90 iPHoP-RF;95.70 |
| S7_vs_21521 | Shared | n.a. | n.a. | n.a. | n.a. | n.a. | n.a. | n.a. | n.a. | 35.28 | d__Bacteria;p__Firmicutes;c__Clostridia;o__Oscillospirales;f__Ruminococcaceae;g__Ruminiclostridium_E | 90 | blast;92.50 iPHoP-RF;74.80 |
| S7_vs_2599 | ME | n.a. | n.a. | n.a. | n.a. | n.a. | n.a. | n.a. | n.a. | 89.5 | d__Bacteria;p__Bacteroidota;c__Bacteroidia;o__Bacteroidales;f__Bacteroidaceae;g__Bacteroides | 100 | RaFAH;99.30 iPHoP-RF;94.40 blast;93.90 |
| S7_vs_26757 | Shared | n.a. | n.a. | n.a. | n.a. | n.a. | n.a. | n.a. | n.a. | | d__Bacteria;p__Firmicutes;c__Clostridia;o__Oscillospirales;f__Ruminococcaceae;g__Faecalibacterium | 94.6 | blast;96.20 iPHoP-RF;85.10 |
| S7_vs_28150 | ME | n.a. | n.a. | n.a. | n.a. | n.a. | n.a. | n.a. | n.a. | 12.25 | d__Bacteria;p__Firmicutes;c__Clostridia;o__Oscillospirales;f__Oscillospiraceae;g__Dysosmobacter | 94.2 | blast;95.90 iPHoP-RF;74.10 |
| S7_vs_28733 | ME | Duplodnaviria | Heunggongvirae | Uroviricota | Caudoviricetes | n.a. | Unclassified | O | O | 56.54 | d__Bacteria;p__Firmicutes;c__Clostridia;o__Oscillospirales;f__Acutalibacteraceae;g__Hydrogeniiclostridium | 95.2 | blast;96.60 iPHoP-RF;62.20 |
| S7_vs_29595 | ME | Duplodnaviria | Heunggongvirae | Uroviricota | Caudoviricetes | n.a. | Unclassified | O | O | 82.91 | d__Bacteria;p__Verrucomicrobiota;c__Verrucomicrobiae;o__Verrucomicrobiales;f__Akkermansiaceae;g__Akkermansia | 99.4 | CRISPR;99.50 iPHoP-RF;57.30 |
| S7_vs_30640 | ME | Duplodnaviria | Heunggongvirae | Uroviricota | Caudoviricetes | n.a. | Unclassified | O | O | 25.04 | d__Bacteria;p__Firmicutes;c__Clostridia;o__Lachnospirales;f__Lachnospiraceae;g__Eubacterium_G | 94.3 | blast;96.00 iPHoP-RF;75.20 |

| | | | | | | | | | | | | | |
|---|---|---|---|---|---|---|---|---|---|---|---|---|---|
| S7_vs_30665 | ME | n.a. | n.a. | n.a. | n.a. | n.a. | n.a. | n.a. | n.a. | 8.94 | d__Bacteria;p__Firmicutes;c__Clostridia;o__Christensenellales;f__CAG-74;g__UBA11524 | 93.8 | CRISPR;95.60 blast;92.80 iPHoP-RF;92.40 |
| S7_vs_38113 | ME | Duplodnaviria | Heunggongvirae | Uroviricota | Caudoviricetes | n.a. | Unclassified | O | O | 17.89 | d__Bacteria;p__Firmicutes;c__Clostridia;o__Oscillospirales;f__Acutalibacteraceae;g__Hydrogeniiclostridium | 91.7 | blast;94.00 iPHoP-RF;64.50 |
| S7_vs_38834 | ME | Duplodnaviria | Heunggongvirae | Uroviricota | Caudoviricetes | n.a. | Unclassified | O | O | 25.72 | d__Bacteria;p__Firmicutes;c__Clostridia;o__Lachnospirales;f__Lachnospiraceae;g__RUG115 | 91.5 | blast;93.80 |
| S7_vs_41098 | Shared | Duplodnaviria | Heunggongvirae | Uroviricota | Caudoviricetes | n.a. | Unclassified | O | O | 97.6 | d__Bacteria;p__Firmicutes;c__Clostridia;o__Lachnospirales;f__Lachnospiraceae;g__Mediterraneibacter | 95.4 | blast;96.80 |
| S7_vs_42563 | Shared | n.a. | n.a. | n.a. | n.a. | n.a. | n.a. | n.a. | n.a. | 93.91 | d__Bacteria;p__Firmicutes;c__Clostridia;o__Oscillospirales;f__Ruminococcaceae;g__Gemmiger | 96.3 | blast;96.80 iPHoP-RF;75.40 RaFAH;59.00 |
| S7_vs_47050 | ME | Duplodnaviria | Heunggongvirae | Uroviricota | Caudoviricetes | n.a. | Unclassified | O | O | 16.97 | d__Bacteria;p__Firmicutes;c__Clostridia;o__Oscillospirales;f__Acutalibacteraceae;g__CAG-180 | 98.7 | CRISPR;99.00 blast;96.60 iPHoP-RF;92.40 |
| S7_vs_48523 | ME | Duplodnaviria | Heunggongvirae | Uroviricota | Caudoviricetes | n.a. | Unclassified | O | O | 66.97 | d__Bacteria;p__Proteobacteria;c__Gammaproteobacteria;o__Burkholderiales;f__Burkholderiaceae;g__Sutterella | 91.6 | blast;93.90 |
| S7_vs_48708 | ME | Duplodnaviria | Heunggongvirae | Uroviricota | Caudoviricetes | n.a. | Unclassified | O | O | 42.25 | d__Bacteria;p__Firmicutes;c__Clostridia;o__Lachnospirales;f__Lachnospiraceae;g__Blautia_A | 90.9 | blast;93.30 iPHoP-RF;69.50 |
| S7_vs_49426 | Shared | Duplodnaviria | Heunggongvirae | Uroviricota | Caudoviricetes | n.a. | Unclassified | O | O | 18.87 | d__Bacteria;p__Firmicutes;c__Clostridia;o__Lachnospirales;f__Lachnospiraceae;g__Agathobacter | 91.5 | blast;93.80 iPHoP-RF;71.30 |
| S7_vs_4956 | Shared | Duplodnaviria | Heunggongvirae | Uroviricota | Caudoviricetes | n.a. | Unclassified | Unclassified | Eponavirus | 54.17 | d__Bacteria;p__Firmicutes;c__Clostridia;o__Oscillospirales;f__Oscillospiraceae;g__ | 91.6 | blast;93.90 iPHoP-RF;83.30 |
| S7_vs_50194 | Shared | n.a. | n.a. | n.a. | n.a. | n.a. | n.a. | n.a. | n.a. | 18.68 | d__Bacteria;p__Bacteroidota;c__Bacteroidia;o__Bacteroidales;f__Rikenellaceae;g__Alistipes_A | 96.9 | blast;90.90 RaFAH;87.90 iPHoP-RF;85.70 |
| S7_vs_51332 | ME | n.a. | n.a. | n.a. | n.a. | n.a. | n.a. | n.a. | n.a. | 13.21 | d__Bacteria;p__Bacteroidota;c__Bacteroidia;o__Bacteroidales;f__Coprobacteraceae;g__Coprobacter | 94.5 | blast;96.10 iPHoP-RF;53.30 |
| S7_vs_5236 | ME | Duplodnaviria | Heunggongvirae | Uroviricota | Caudoviricetes | n.a. | Unclassified | O | O | 93.85 | d__Bacteria;p__Firmicutes;c__Clostridia;o__Oscillospirales;f__Oscillospiraceae;g__CAG-103 | 93.5 | CRISPR;95.40 |
| S7_vs_54322 | ME | n.a. | n.a. | n.a. | n.a. | n.a. | n.a. | n.a. | n.a. | 65.6 | d__Bacteria;p__Bacteroidota;c__Bacteroidia;o__Bacteroidales;f__Rikenellaceae;g__Alistipes | 98.7 | blast;96.80 CRISPR;95.60 iPHoP-RF;87.30 RaFAH;84.00 |
| S7_vs_55031 | ME | n.a. | n.a. | n.a. | n.a. | n.a. | n.a. | n.a. | n.a. | 23.56 | d__Bacteria;p__Bacteroidota;c__Bacteroidia;o__Bacteroidales;f__Tannerellaceae;g__Parabacteroides | 98.1 | blast;98.60 CRISPR;97.70 iPHoP-RF;89.20 |
| S7_vs_55526 | ME | n.a. | n.a. | n.a. | n.a. | n.a. | n.a. | n.a. | n.a. | 22.37 | d__Bacteria;p__Actinobacteriota;c__Coriobacteriia;o__Coriobacteriales;f__Coriobacteriaceae;g__Collinsella | 98.7 | CRISPR;99.00 blast;93.80 iPHoP-RF;84.50 |
| S7_vs_557 | ME | Duplodnaviria | Heunggongvirae | Uroviricota | Caudoviricetes | n.a. | Unclassified | O | O | 66.78 | d__Bacteria;p__Bacteroidota;c__Bacteroidia;o__Bacteroidales;f__Bacteroidaceae;g__Bacteroides | 99.3 | iPHoP-RF;97.10 blast;93.70 RaFAH;89.50 |
| S7_vs_60775 | ME | Duplodnaviria | Heunggongvirae | Uroviricota | Caudoviricetes | n.a. | Unclassified | O | O | 62.59 | d__Bacteria;p__Firmicutes;c__Clostridia;o__Lachnospirales;f__Lachnospiraceae;g__Eubacterium_F | 91.6 | blast;93.90 |
| S7_vs_65719 | Shared | Duplodnaviria | Heunggongvirae | Uroviricota | Caudoviricetes | n.a. | Unclassified | O | O | 47.93 | d__Bacteria;p__Firmicutes;c__Clostridia;o__Lachnospirales;f__Lachnospiraceae;g__Enterocloster | 95.7 | CRISPR;97.00 blast;94.90 iPHoP-RF;88.90 |
| S7_vs_69630 | ME | Duplodnaviria | Heunggongvirae | Uroviricota | Caudoviricetes | n.a. | Unclassified | O | O | 35.61 | d__Bacteria;p__Firmicutes;c__Clostridia;o__Oscillospirales;f__Oscillospiraceae;g__Intestinimonas | 94.7 | blast;96.30 iPHoP-RF;65.10 |
| S7_vs_72737 | ME | n.a. | n.a. | n.a. | n.a. | n.a. | n.a. | n.a. | n.a. | 3.91 | d__Bacteria;p__Firmicutes;c__Clostridia;o__Oscillospirales;f__Acutalibacteraceae;g__Ruminococcus_E | 97.9 | blast;98.50 iPHoP-RF;96.40 |
| S7_vs_73280 | ME | Duplodnaviria | Heunggongvirae | Uroviricota | Caudoviricetes | n.a. | Unclassified | Unclassified | Minunavirus | 86.44 | d__Bacteria;p__Actinobacteriota;c__Actinomycetia;o__Actinomycetales;f__Bifidobacteriaceae;g__Bifidobacterium | 99.1 | CRISPR;99.30 iPHoP-RF;97.10 blast;96.90 |
| S7_vs_78507 | Shared | n.a. | n.a. | n.a. | n.a. | n.a. | n.a. | n.a. | n.a. | 36.34 | d__Bacteria;p__Firmicutes;c__Clostridia;o__Oscillospirales;f__Oscillospiraceae;g__Intestinimonas | 90.8 | blast;93.20 iPHoP-RF;70.90 |
| S7_vs_83100 | ME | Duplodnaviria | Heunggongvirae | Uroviricota | Caudoviricetes | n.a. | Unclassified | O | O | 97.77 | d__Bacteria;p__Firmicutes;c__Clostridia;o__Oscillospirales;f__Oscillospiraceae;g__Flavonifractor | 97.8 | blast;98.40 iPHoP-RF;85.10 CRISPR;74.00 |
| S7_vs_83474 | ME | Duplodnaviria | Heunggongvirae | Uroviricota | Caudoviricetes | n.a. | Unclassified | Unclassified | O | 31.73 | d__Bacteria;p__Firmicutes;c__Clostridia;o__Oscillospirales;f__Oscillospiraceae;g__Flavonifractor | 93.9 | blast;95.70 iPHoP-RF;90.80 |
| S7_vs_86377 | ME | Duplodnaviria | Heunggongvirae | Uroviricota | Caudoviricetes | n.a. | Unclassified | O | O | 91.04 | d__Bacteria;p__Firmicutes;c__Clostridia;o__Lachnospirales;f__Lachnospiraceae;g__Bariatricus | 92 | blast;94.20 iPHoP-RF;79.50 |
| S7_vs_87279 | ME | Duplodnaviria | Heunggongvirae | Uroviricota | Caudoviricetes | n.a. | Unclassified | Unclassified | O | 66.77 | d__Bacteria;p__Firmicutes;c__Clostridia;o__Oscillospirales;f__Ruminococcaceae;g__Faecalibacterium | 95.2 | blast;96.60 iPHoP-RF;62.50 |
| S7_vs_87439 | ME | n.a. | n.a. | n.a. | n.a. | n.a. | n.a. | n.a. | n.a. | 31.68 | d__Bacteria;p__Verrucomicrobiota;c__Lentisphaeria;o__Victivallales;f__Victivallaceae;g__Victivallis | 90.2 | blast;92.70 iPHoP-RF;71.10 |
| S7_vs_944 | ME | n.a. | n.a. | n.a. | n.a. | n.a. | n.a. | n.a. | n.a. | 66.43 | d__Bacteria;p__Bacteroidota;c__Bacteroidia;o__Bacteroidales;f__Bacteroidaceae;g__Bacteroides | 100 | RaFAH;98.90 iPHoP-RF;96.70 blast;91.80 |
| S7_vs_94738 | ME | Duplodnaviria | Heunggongvirae | Uroviricota | Caudoviricetes | n.a. | Unclassified | Unclassified | O | 58.62 | d__Bacteria;p__Firmicutes;c__Clostridia;o__Oscillospirales;f__Oscillospiraceae;g__Flavonifractor | 91.2 | blast;93.60 iPHoP-RF;71.30 |
| S7_vs_97737 | ME | n.a. | n.a. | n.a. | n.a. | n.a. | n.a. | n.a. | n.a. | 56.07 | d__Bacteria;p__Firmicutes;c__Clostridia;o__Oscillospirales;f__Ruminococcaceae;g__Faecalibacterium | 95.6 | blast;96.90 iPHoP-RF;91.80 |
| S8_vf_101283 | SHHC | Duplodnaviria | Heunggongvirae | Uroviricota | Caudoviricetes | n.a. | Unclassified | Unclassified | Toutatisvirus | 99.72 | d__Bacteria;p__Firmicutes;c__Clostridia;o__Oscillospirales;f__Ruminococcaceae;g__Faecalibacterium | 100 | RaFAH;99.60 CRISPR;98.20 blast;95.50 iPHoP-RF;84.80 |
| S8_vf_102508 | SHHC | n.a. | n.a. | n.a. | n.a. | n.a. | n.a. | n.a. | n.a. | 92.39 | d__Bacteria;p__Actinobacteriota;c__Actinomycetia;o__Actinomycetales;f__Micrococcaceae;g__Rothia | 100 | RaFAH;99.40 CRISPR;98.40 iPHoP-RF;80.80 |
| S8_vf_111790 | Shared | Duplodnaviria | Heunggongvirae | Uroviricota | Caudoviricetes | n.a. | Unclassified | O | O | 20.45 | d__Bacteria;p__Firmicutes;c__Clostridia;o__Oscillospirales;f__Oscillospiraceae;g__Flavonifractor | 91.6 | blast;93.90 iPHoP-RF;81.10 |
| S8_vf_117796 | Shared | Duplodnaviria | Heunggongvirae | Uroviricota | Caudoviricetes | n.a. | Unclassified | Unclassified | Unclassified | 71.62 | d__Bacteria;p__Bacteroidota;c__Bacteroidia;o__Bacteroidales;f__Bacteroidaceae;g__Prevotella | 91.1 | blast;93.50 iPHoP-RF;88.50 |
| S8_vf_122206 | Shared | Duplodnaviria | Heunggongvirae | Uroviricota | Caudoviricetes | n.a. | Unclassified | O | O | 16.65 | d__Bacteria;p__Bacteroidota;c__Bacteroidia;o__Bacteroidales;f__Bacteroidaceae;g__Prevotella | 94.7 | blast;96.30 iPHoP-RF;90.50 |
| S8_vf_129423 | SHHC | Duplodnaviria | Heunggongvirae | Uroviricota | Caudoviricetes | n.a. | Unclassified | Mccleskeyvirinae | Unaquatrovirus | 87.49 | d__Bacteria;p__Firmicutes;c__Bacilli;o__Lactobacillales;f__Lactobacillaceae;g__Leuconostoc | 97.5 | iPHoP-RF;93.70 RaFAH;86.10 |
| S8_vf_131009 | SHHC | Duplodnaviria | Heunggongvirae | Uroviricota | Caudoviricetes | n.a. | Unclassified | O | O | 24.39 | d__Bacteria;p__Firmicutes;c__Clostridia;o__Lachnospirales;f__Lachnospiraceae;g__Eubacterium_F | 97.2 | CRISPR;98.00 blast;96.80 iPHoP-RF;95.40 |
| S8_vf_132697 | Shared | Duplodnaviria | Heunggongvirae | Uroviricota | Caudoviricetes | n.a. | Unclassified | O | O | 84.84 | d__Bacteria;p__Firmicutes;c__Clostridia;o__Lachnospirales;f__Lachnospiraceae;g__Agathobacter | 99.9 | CRISPR;98.90 RaFAH;92.10 blast;90.20 iPHoP-RF;86.30 |
| S8_vf_134943 | SHHC | n.a. | n.a. | n.a. | n.a. | n.a. | n.a. | n.a. | n.a. | 81.72 | d__Bacteria;p__Firmicutes;c__Bacilli;o__Lactobacillales;f__Streptococcaceae;g__Streptococcus | 99.7 | RaFAH;99.40 iPHoP-RF;72.60 |
| S8_vf_13709 | SHHC | Duplodnaviria | Heunggongvirae | Uroviricota | Caudoviricetes | n.a. | Unclassified | O | O | 11.95 | d__Bacteria;p__Firmicutes;c__Clostridia;o__TANB77;f__CAG-508;g__CAG-354 | 90.2 | blast;92.70 iPHoP-RF;70.60 |
| S8_vf_13758 | SHHC | Duplodnaviria | Heunggongvirae | Uroviricota | Caudoviricetes | n.a. | Unclassified | O | O | 11.78 | d__Bacteria;p__Firmicutes;c__Clostridia;o__TANB77;f__CAG-508;g__CAG-354 | 93.9 | blast;95.70 iPHoP-RF;71.70 |
| S8_vf_142700 | SHHC | Unclassified | n.a. | n.a. | n.a. | n.a. | Unclassified | O | O | 5.82 | d__Bacteria;p__Actinobacteriota;c__Coriobacteriia;o__Coriobacteriales;f__Coriobacteriaceae;g__Collinsella | 99.4 | CRISPR;99.50 iPHoP-RF;93.40 blast;91.90 |
| S8_vf_151384 | SHHC | Duplodnaviria | Heunggongvirae | Uroviricota | Caudoviricetes | n.a. | Unclassified | Unclassified | Eponavirus | 47.18 | d__Bacteria;p__Firmicutes;c__Clostridia;o__Oscillospirales;f__Ruminococcaceae;g__Gemmiger | 96.3 | CRISPR;97.40 blast;92.00 iPHoP-RF;89.80 |
| S8_vf_153696 | Shared | n.a. | n.a. | n.a. | n.a. | n.a. | n.a. | n.a. | n.a. | 11.92 | d__Bacteria;p__Actinobacteriota;c__Actinomycetia;o__Actinomycetales;f__Bifidobacteriaceae;g__Bifidobacterium | 91.2 | blast;93.60 |
| S8_vf_158225 | Shared | Duplodnaviria | Heunggongvirae | Uroviricota | Caudoviricetes | Crassvirales | Intestiviridae | Crudevirinae | Diorhovirus | 14.69 | d__Bacteria;p__Bacteroidota;c__Bacteroidia;o__Bacteroidales;f__Tannerellaceae;g__Parabacteroides | 94 | blast;95.80 |
| S8_vf_161703 | SHHC | Duplodnaviria | Heunggongvirae | Uroviricota | Caudoviricetes | n.a. | Unclassified | O | O | 49.06 | d__Bacteria;p__Firmicutes;c__Clostridia;o__Oscillospirales;f__Ruminococcaceae;g__Ruminococcus_D | 99.1 | CRISPR;97.90 blast;96.80 iPHoP-RF;96.40 RaFAH;83.30 |
| S8_vf_16172 | SHHC | n.a. | n.a. | n.a. | n.a. | n.a. | n.a. | n.a. | n.a. | 45.4 | d__Bacteria;p__Bacteroidota;c__Bacteroidia;o__Bacteroidales;f__Tannerellaceae;g__Parabacteroides | 95.9 | iPHoP-RF;97.10 blast;96.90 CRISPR;92.40 |
| S8_vf_166403 | Shared | n.a. | n.a. | n.a. | n.a. | n.a. | n.a. | n.a. | n.a. | 69.48 | d__Bacteria;p__Firmicutes;c__Clostridia;o__Oscillospirales;f__Oscillospiraceae;g__Flavonifractor | 95.2 | blast;96.60 iPHoP-RF;72.20 |
| S8_vf_166677 | Shared | Duplodnaviria | Heunggongvirae | Uroviricota | Caudoviricetes | Crassvirales | Intestiviridae | Obtuvirinae | Fohxhuevirus | 20.94 | d__Bacteria;p__Bacteroidota;c__Bacteroidia;o__Bacteroidales;f__Bacteroidaceae;g__Prevotella | 99.9 | blast;98.70 CRISPR;96.60 iPHoP-RF;93.10 RaFAH;90.50 |
| S8_vf_170878 | Shared | Duplodnaviria | Heunggongvirae | Uroviricota | Caudoviricetes | n.a. | Unclassified | Unclassified | Toutatisvirus | 39.33 | d__Bacteria;p__Firmicutes;c__Clostridia;o__Oscillospirales;f__Oscillospiraceae;g__CAG-83 | 91.2 | blast;93.60 iPHoP-RF;65.60 |
| S8_vf_183045 | Shared | Duplodnaviria | Heunggongvirae | Uroviricota | Caudoviricetes | n.a. | Unclassified | Unclassified | Unclassified | 35.16 | d__Bacteria;p__Bacteroidota;c__Bacteroidia;o__Bacteroidales;f__Bacteroidaceae;g__Prevotella | 98.5 | CRISPR;98.90 blast;93.00 iPHoP-RF;90.50 |
| S8_vf_187742 | SHHC | Duplodnaviria | Heunggongvirae | Uroviricota | Caudoviricetes | n.a. | Unclassified | O | O | 91.82 | d__Bacteria;p__Firmicutes;c__Clostridia;o__Oscillospirales;f__Ruminococcaceae;g__Faecalibacterium | 99.6 | blast;96.20 RaFAH;95.10 CRISPR;86.20 iPHoP-RF;65.30 |
| S8_vf_195032 | SHHC | Duplodnaviria | Heunggongvirae | Uroviricota | Caudoviricetes | n.a. | Unclassified | O | O | 5.64 | d__Bacteria;p__Actinobacteriota;c__Coriobacteriia;o__Coriobacteriales;f__Eggerthellaceae;g__Gordonibacter | 94.9 | CRISPR;96.40 iPHoP-RF;75.70 |
| S8_vf_197061 | Shared | Duplodnaviria | Heunggongvirae | Uroviricota | Caudoviricetes | n.a. | Unclassified | Unclassified | Unclassified | 60.03 | d__Bacteria;p__Actinobacteriota;c__Actinomycetia;o__Actinomycetales;f__Bifidobacteriaceae;g__Bifidobacterium | 98.8 | CRISPR;99.00 iPHoP-RF;97.10 blast;96.80 RaFAH;52.60 |
| S8_vf_201695 | Shared | n.a. | n.a. | n.a. | n.a. | n.a. | n.a. | n.a. | n.a. | 30.61 | d__Bacteria;p__Firmicutes;c__Clostridia;o__Lachnospirales;f__Anaerotignaceae;g__Anaerotignum | 94.2 | blast;95.90 iPHoP-RF;76.40 |
| S8_vf_208110 | Shared | Duplodnaviria | Heunggongvirae | Uroviricota | Caudoviricetes | n.a. | Unclassified | O | O | 89.6 | d__Bacteria;p__Firmicutes;c__Clostridia;o__Oscillospirales;f__Oscillospiraceae;g__Intestinimonas | 92 | blast;94.20 iPHoP-RF;50.20 |
| S8_vf_208111 | Shared | n.a. | n.a. | n.a. | n.a. | n.a. | n.a. | n.a. | n.a. | 7.17 | d__Bacteria;p__Desulfobacterota;c__Desulfovibrionia;o__Desulfovibrionales;f__Desulfovibrionaceae;g__Bilophila | 90.8 | CRISPR;93.20 iPHoP-RF;60.30 |
| S8_vf_208115 | SHHC | Monodnaviria | Sangervirae | Phixviricota | Malgrandaviricetes | Petitvirales | Microviridae | O | O | | d__Bacteria;p__Bacteroidota;c__Bacteroidia;o__Bacteroidales;f__Barnesiellaceae;g__Barnesiella | 94.2 | blast;95.90 iPHoP-RF;72.80 |
| S8_vf_208119 | SHHC | Monodnaviria | Sangervirae | Phixviricota | Malgrandaviricetes | Petitvirales | Microviridae | Gokushovirinae | Gokushovirus | 18.24 | d__Bacteria;p__Firmicutes;c__Clostridia;o__Oscillospirales;f__Ruminococcaceae;g__Faecalibacterium | 97.2 | CRISPR;97.90 RaFAH;52.10 |
| S8_vf_208120 | Shared | Duplodnaviria | Heunggongvirae | Uroviricota | Caudoviricetes | n.a. | Unclassified | O | O | 99.62 | d__Bacteria;p__Bacteroidota;c__Bacteroidia;o__Bacteroidales;f__Bacteroidaceae;g__Bacteroides | 98.2 | RaFAH;93.90 blast;88.30 iPHoP-RF;53.80 |
| S8_vf_208126 | SHHC | Duplodnaviria | Heunggongvirae | Uroviricota | Caudoviricetes | n.a. | Unclassified | O | O | 48.51 | d__Bacteria;p__Firmicutes;c__Clostridia;o__Oscillospirales;f__Ruminococcaceae;g__Gemmiger | 91.6 | blast;93.90 iPHoP-RF;60.90 |
| S8_vf_208146 | SHHC | Monodnaviria | Sangervirae | Phixviricota | Malgrandaviricetes | Petitvirales | Microviridae | Gokushovirinae | Unclassified | 25.58 | d__Bacteria;p__Firmicutes;c__Negativicutes;o__Acidaminococcales;f__Acidaminococcaceae;g__Phascolarctobacterium | 93.1 | CRISPR;95.10 |
| S8_vf_28956 | Shared | n.a. | n.a. | n.a. | n.a. | n.a. | n.a. | n.a. | n.a. | 91.89 | d__Bacteria;p__Bacteroidota;c__Bacteroidia;o__Bacteroidales;f__Bacteroidaceae;g__Bacteroides | 99.6 | iPHoP-RF;96.40 RaFAH;94.20 blast;93.80 |
| S8_vf_45776 | Shared | Duplodnaviria | Heunggongvirae | Uroviricota | Caudoviricetes | n.a. | Unclassified | Unclassified | Skunavirus | 88.6 | d__Bacteria;p__Firmicutes;c__Bacilli;o__Lactobacillales;f__Streptococcaceae;g__Lactococcus | 100 | RaFAH;99.20 blast;98.30 iPHoP-RF;73.90 |
| S8_vf_55476 | Shared | n.a. | n.a. | n.a. | n.a. | n.a. | n.a. | n.a. | n.a. | 64.72 | d__Bacteria;p__Firmicutes;c__Clostridia;o__Oscillospirales;f__Oscillospiraceae;g__Intestinimonas | 91.6 | blast;93.90 iPHoP-RF;82.20 |
| S8_vf_65522 | Shared | n.a. | n.a. | n.a. | n.a. | n.a. | n.a. | n.a. | n.a. | 12.03 | d__Bacteria;p__Firmicutes;c__Clostridia;o__Christensenellales;f__CAG-74;g__UBA11524 | 97.2 | blast;98.00 iPHoP-RF;83.10 CRISPR;55.60 |
| S8_vf_7243 | SHHC | n.a. | n.a. | n.a. | n.a. | n.a. | n.a. | n.a. | n.a. | 95.82 | d__Bacteria;p__Actinobacteriota;c__Actinomycetia;o__Actinomycetales;f__Micrococcaceae;g__Rothia | 99.4 | RaFAH;99.50 |
| S8_vf_79769 | Shared | n.a. | n.a. | n.a. | n.a. | n.a. | n.a. | n.a. | n.a. | 31.62 | d__Bacteria;p__Bacteroidota;c__Bacteroidia;o__Bacteroidales;f__Bacteroidaceae;g__Prevotella | 98.4 | CRISPR;98.80 blast;96.70 iPHoP-RF;92.40 |
| S8_vf_93976 | SHHC | n.a. | n.a. | n.a. | n.a. | n.a. | n.a. | n.a. | n.a. | 9.53 | d__Bacteria;p__Firmicutes;c__Clostridia;o__Christensenellales;f__CAG-138;g__UBA1685 | 95.3 | blast;96.70 iPHoP-RF;66.50 CRISPR;59.10 |
| S8_vs_102608 | SHHC | n.a. | n.a. | n.a. | n.a. | n.a. | n.a. | n.a. | n.a. | 20 | d__Bacteria;p__Firmicutes;c__Clostridia;o__Oscillospirales;f__Ruminococcaceae;g__Gemmiger | 95.7 | blast;97.00 CRISPR;96.10 iPHoP-RF;90.80 |
| S8_vs_115958 | Shared | Duplodnaviria | Heunggongvirae | Uroviricota | Caudoviricetes | n.a. | Unclassified | Unclassified | O | 68.75 | d__Bacteria;p__Firmicutes;c__Clostridia;o__Oscillospirales;f__Acutalibacteraceae;g__Ruminococcus_E | 91.6 | blast;93.90 iPHoP-RF;67.00 |
| S8_vs_125717 | Shared | n.a. | n.a. | n.a. | n.a. | n.a. | n.a. | n.a. | n.a. | 17.94 | d__Bacteria;p__Firmicutes;c__Clostridia;o__Oscillospirales;f__Oscillospiraceae;g__CAG-83 | 95.6 | blast;96.90 iPHoP-RF;64.30 |
| S8_vs_126582 | SHHC | Duplodnaviria | Heunggongvirae | Uroviricota | Caudoviricetes | n.a. | Unclassified | O | O | 98.15 | d__Bacteria;p__Bacteroidota;c__Bacteroidia;o__Bacteroidales;f__Tannerellaceae;g__Parabacteroides | 100 | RaFAH;99.70 iPHoP-RF;89.20 blast;86.20 CRISPR;69.40 |
| S8_vs_134805 | Shared | Duplodnaviria | Heunggongvirae | Uroviricota | Caudoviricetes | n.a. | Unclassified | Unclassified | Mushuvirus | 91.02 | d__Bacteria;p__Firmicutes;c__Clostridia;o__Oscillospirales;f__Oscillospiraceae;g__Lawsonibacter | 91.5 | blast;93.80 iPHoP-RF;73.20 |
| S8_vs_145173 | SHHC | Duplodnaviria | Heunggongvirae | Uroviricota | Caudoviricetes | n.a. | Unclassified | O | O | 54.47 | d__Bacteria;p__Firmicutes;c__Clostridia;o__Oscillospirales;f__Oscillospiraceae;g__UBA9475 | 91.2 | blast;93.60 iPHoP-RF;59.00 |
| S8_vs_147670 | SHHC | Duplodnaviria | Heunggongvirae | Uroviricota | Caudoviricetes | n.a. | Unclassified | O | O | 31.23 | d__Bacteria;p__Firmicutes;c__Negativicutes;o__Veillonellales;f__Megasphaeraceae;g__Caecibacter | 96.7 | CRISPR;97.70 blast;78.80 |
| S8_vs_147697 | Shared | n.a. | n.a. | n.a. | n.a. | n.a. | n.a. | n.a. | n.a. | 41.71 | d__Bacteria;p__Bacteroidota;c__Bacteroidia;o__Bacteroidales;f__Bacteroidaceae;g__Bacteroides | 98.8 | iPHoP-RF;95.70 blast;90.90 RaFAH;89.00 |
| S8_vs_148389 | SHHC | Duplodnaviria | Heunggongvirae | Uroviricota | Caudoviricetes | n.a. | Unclassified | O | O | 54.68 | d__Bacteria;p__Bacteroidota;c__Bacteroidia;o__Bacteroidales;f__Bacteroidaceae;g__Prevotella | 99.6 | blast;96.80 RaFAH;94.40 iPHoP-RF;91.80 |
| S8_vs_173961 | Shared | Duplodnaviria | Heunggongvirae | Uroviricota | Caudoviricetes | n.a. | Unclassified | O | O | 95.47 | d__Bacteria;p__Firmicutes;c__Clostridia;o__Lachnospirales;f__Lachnospiraceae;g__Agathobacter | 99.6 | CRISPR;98.40 blast;90.90 RaFAH;89.00 iPHoP-RF;87.60 |
| S8_vs_176604 | SHHC | n.a. | n.a. | n.a. | n.a. | n.a. | n.a. | n.a. | n.a. | 21.14 | d__Bacteria;p__Firmicutes;c__Clostridia;o__Christensenellales;f__CAG-138;g__SFEL01 | 94.9 | blast;96.40 iPHoP-RF;81.60 CRISPR;71.90 |
| S8_vs_179208 | Shared | n.a. | n.a. | n.a. | n.a. | n.a. | n.a. | n.a. | n.a. | 39.41 | d__Bacteria;p__Bacteroidota;c__Bacteroidia;o__Bacteroidales;f__Rikenellaceae;g__Alistipes | 95.4 | blast;96.80 iPHoP-RF;80.80 |
| S8_vs_179529 | SHHC | Duplodnaviria | Heunggongvirae | Uroviricota | Caudoviricetes | n.a. | Unclassified | Unclassified | O | 98.83 | d__Bacteria;p__Firmicutes;c__Clostridia;o__Oscillospirales;f__Ruminococcaceae;g__Faecalibacterium | 100 | CRISPR;98.50 RaFAH;98.00 blast;95.30 iPHoP-RF;86.70 |
| S8_vs_182288 | Shared | Duplodnaviria | Heunggongvirae | Uroviricota | Caudoviricetes | n.a. | Unclassified | O | O | 85.94 | d__Bacteria;p__Bacteroidota;c__Bacteroidia;o__Bacteroidales;f__Tannerellaceae;g__Parabacteroides | 100 | CRISPR;99.00 RaFAH;98.40 iPHoP-RF;96.40 blast;93.80 |
| S8_vs_206526 | Shared | Duplodnaviria | Heunggongvirae | Uroviricota | Caudoviricetes | n.a. | Unclassified | Unclassified | Unclassified | 81.78 | d__Bacteria;p__Firmicutes;c__Clostridia;o__Lachnospirales;f__Lachnospiraceae;g__Roseburia | 97.9 | CRISPR;98.50 blast;92.20 iPHoP-RF;87.30 RaFAH;50.70 |
| S8_vs_208114 | SHHC | Duplodnaviria | Heunggongvirae | Uroviricota | Caudoviricetes | n.a. | Unclassified | O | O | 9.13 | d__Bacteria;p__Desulfobacterota;c__Desulfovibrionia;o__Desulfovibrionales;f__Desulfovibrionaceae;g__Mailhella | 90.1 | blast;92.60 iPHoP-RF;81.40 |
| S8_vs_28311 | Shared | Duplodnaviria | Heunggongvirae | Uroviricota | Caudoviricetes | n.a. | Duneviridae | O | O | 99.02 | d__Bacteria;p__Bacteroidota;c__Bacteroidia;o__Bacteroidales;f__Bacteroidaceae;g__Bacteroides | 100 | RaFAH;97.40 blast;97.40 iPHoP-RF;96.70 CRISPR;54.10 |
| S8_vs_40014 | Shared | n.a. | n.a. | n.a. | n.a. | n.a. | n.a. | n.a. | n.a. | 80.96 | d__Bacteria;p__Bacteroidota;c__Bacteroidia;o__Bacteroidales;f__Rikenellaceae;g__Alistipes | 94.6 | blast;96.20 iPHoP-RF;95.70 |
| S8_vs_4153 | Shared | n.a. | n.a. | n.a. | n.a. | n.a. | n.a. | n.a. | n.a. | 5.07 | d__Bacteria;p__Bacteroidota;c__Bacteroidia;o__Bacteroidales;f__Barnesiellaceae;g__Barnesiella | 97.9 | blast;98.50 iPHoP-RF;96.40 CRISPR;53.40 |
| S8_vs_50431 | SHHC | n.a. | n.a. | n.a. | n.a. | n.a. | n.a. | n.a. | n.a. | 20.17 | d__Bacteria;p__Firmicutes;c__Clostridia;o__Oscillospirales;f__Oscillospiraceae;g__Dysosmobacter | 91.5 | blast;93.80 iPHoP-RF;50.70 |
| S8_vs_51368 | SHHC | n.a. | n.a. | n.a. | n.a. | n.a. | n.a. | n.a. | n.a. | 31.95 | d__Bacteria;p__Firmicutes;c__Clostridia;o__Oscillospirales;f__Oscillospiraceae;g__CAG-170 | 91.5 | blast;93.80 iPHoP-RF;85.70 |
| S8_vs_5211 | Shared | n.a. | n.a. | n.a. | n.a. | n.a. | n.a. | n.a. | n.a. | 70.75 | d__Bacteria;p__Firmicutes;c__Clostridia;o__Lachnospirales;f__Lachnospiraceae;g__Massilistercora | 90.2 | blast;92.70 iPHoP-RF;58.40 |
| S8_vs_54183 | SHHC | Duplodnaviria | Heunggongvirae | Uroviricota | Caudoviricetes | n.a. | Unclassified | Unclassified | O | 20.38 | d__Bacteria;p__Firmicutes;c__Clostridia;o__Oscillospirales;f__Oscillospiraceae;g__CAG-170 | 91.8 | CRISPR;94.10 blast;90.90 iPHoP-RF;81.90 |
| S8_vs_59736 | Shared | Duplodnaviria | Heunggongvirae | Uroviricota | Caudoviricetes | n.a. | Unclassified | O | O | 26.65 | d__Bacteria;p__Firmicutes;c__Bacilli;o__Erysipelotrichales;f__Erysipelatoclostridiaceae;g__Catenibacterium | 95.6 | blast;96.90 |
| S8_vs_63277 | SHHC | Duplodnaviria | Heunggongvirae | Uroviricota | Caudoviricetes | n.a. | Unclassified | O | O | 75.64 | d__Bacteria;p__Firmicutes;c__Clostridia;o__Oscillospirales;f__Ruminococcaceae;g__Faecalibacterium | 97.8 | blast;98.40 iPHoP-RF;88.20 |
| S8_vs_72117 | Shared | n.a. | n.a. | n.a. | n.a. | n.a. | n.a. | n.a. | n.a. | 19.43 | d__Bacteria;p__Bacteroidota;c__Bacteroidia;o__Bacteroidales;f__Bacteroidaceae;g__Bacteroides | 95.4 | iPHoP-RF;94.40 RaFAH;71.80 |
| S8_vs_9114 | SHHC | n.a. | n.a. | n.a. | n.a. | n.a. | n.a. | n.a. | n.a. | 28.26 | d__Bacteria;p__Bacteroidota;c__Bacteroidia;o__Bacteroidales;f__Rikenellaceae;g__Alistipes_A | 91.1 | blast;93.50 iPHoP-RF;82.80 CRISPR;64.80 |
| S8_vs_99362 | Shared | Duplodnaviria | Heunggongvirae | Uroviricota | Caudoviricetes | n.a. | Unclassified | Unclassified | Unclassified | 99.97 | d__Bacteria;p__Bacteroidota;c__Bacteroidia;o__Bacteroidales;f__Bacteroidaceae;g__Bacteroides | 100 | RaFAH;98.30 blast;96.50 iPHoP-RF;93.40 |
| S8_vs_9987 | Shared | Duplodnaviria | Heunggongvirae | Uroviricota | Caudoviricetes | n.a. | Unclassified | O | O | 99.87 | d__Bacteria;p__Bacteroidota;c__Bacteroidia;o__Bacteroidales;f__Rikenellaceae;g__Alistipes | 100 | blast;97.60 RaFAH;97.40 iPHoP-RF;93.40 |
| S9_vf_13507 | Shared | n.a. | n.a. | n.a. | n.a. | n.a. | n.a. | n.a. | n.a. | 66.3 | d__Bacteria;p__Bacteroidota;c__Bacteroidia;o__Bacteroidales;f__Bacteroidaceae;g__Prevotella | 95.6 | blast;96.90 iPHoP-RF;94.70 |
| S9_vf_15012 | ME | Duplodnaviria | Heunggongvirae | Uroviricota | Caudoviricetes | n.a. | Unclassified | Unclassified | Unclassified | 97.67 | d__Bacteria;p__Firmicutes;c__Bacilli;o__Lactobacillales;f__Streptococcaceae;g__Streptococcus | 100 | RaFAH;99.70 CRISPR;98.30 blast;94.40 iPHoP-RF;92.80 |
| S9_vf_19284 | ME | n.a. | n.a. | n.a. | n.a. | n.a. | n.a. | n.a. | n.a. | 15.56 | d__Bacteria;p__Firmicutes;c__Bacilli;o__Lactobacillales;f__Streptococcaceae;g__Streptococcus | 93 | CRISPR;95.00 |
| S9_vf_4115 | ME | Duplodnaviria | Heunggongvirae | Uroviricota | Caudoviricetes | n.a. | Unclassified | Unclassified | Unclassified | 96.62 | d__Bacteria;p__Firmicutes;c__Bacilli;o__Lactobacillales;f__Streptococcaceae;g__Streptococcus | 100 | RaFAH;99.70 CRISPR;99.00 blast;96.80 iPHoP-RF;96.70 |

| | | | | | | | | | | | | | |
|---|---|---|---|---|---|---|---|---|---|---|---|---|---|
| S9_vf_51656 | Shared | Duplodnaviria | Heunggongvirae | Uroviricota | Caudoviricetes | Crassvirales | Intestiviridae | Obtuvirinae | Fohxhuevirus | 29.36 | d__Bacteria;p__Bacteroidota;c__Bacteroidia;o__Bacteroidales;f__Bacteroidaceae;g__Prevotella | 99 | CRISPR;99.00 blast;96.30 iPHoP-RF;96.10 RaFAH;59.50 |
| S9_vf_64844 | Shared | Duplodnaviria | Heunggongvirae | Uroviricota | Caudoviricetes | n.a. | Unclassified | Unclassified | Lagaffevirus | 89.68 | d__Bacteria;p__Firmicutes;c__Clostridia;o__Oscillospirales;f__Oscillospiraceae;g__CAG-83 | 93.4 | RaFAH;95.30 blast;50.00 |
| S9_vf_73311 | ME | Duplodnaviria | Heunggongvirae | Uroviricota | Caudoviricetes | n.a. | Unclassified | Unclassified | Unclassified | 18.82 | d__Bacteria;p__Firmicutes;c__Clostridia;o__Lachnospirales;f__CAG-274;g__CAG-274 | 97.2 | CRISPR;98.00 blast;96.80 iPHoP-RF;78.70 |
| S9_vf_76769 | Shared | Duplodnaviria | Heunggongvirae | Uroviricota | Caudoviricetes | Crassvirales | Suoliviridae | Loutivirinae | Buchavirus | 53.32 | d__Bacteria;p__Bacteroidota;c__Bacteroidia;o__Bacteroidales;f__Bacteroidaceae;g__Prevotella | 92.6 | RaFAH;94.70 |
| S9_vf_90856 | ME | Duplodnaviria | Heunggongvirae | Uroviricota | Caudoviricetes | n.a. | Unclassified | Unclassified | O | 94.47 | d__Bacteria;p__Firmicutes;c__Bacilli;o__Lactobacillales;f__Streptococcaceae;g__Streptococcus | 100 | RaFAH;99.80 CRISPR;99.00 blast;95.80 iPHoP-RF;94.40 |
| S9_vf_90866 | ME | Unclassified | n.a. | n.a. | n.a. | n.a. | Unclassified | Unclassified | Unclassified | 15.47 | d__Bacteria;p__Firmicutes;c__Bacilli;o__Lactobacillales;f__Streptococcaceae;g__Streptococcus | 99.3 | iPHoP-RF;97.10 blast;95.70 RaFAH;90.00 |
| S9_vf_934 | ME | Duplodnaviria | Heunggongvirae | Uroviricota | Caudoviricetes | n.a. | Unclassified | O | O | 17.76 | d__Bacteria;p__Actinobacteriota;c__Actinomycetia;o__Mycobacteriales;f__Mycobacteriaceae;g__Mycobacterium | 92.2 | iPHoP-RF;94.40 |
| S9_vs_14235 | ME | n.a. | n.a. | n.a. | n.a. | n.a. | n.a. | n.a. | n.a. | 12.73 | d__Bacteria;p__Firmicutes;c__Clostridia;o__Oscillospirales;f__Oscillospiraceae;g__An92 | 91 | blast;93.40 iPHoP-RF;87.00 CRISPR;73.90 |
| S9_vs_1966 | Shared | Duplodnaviria | Heunggongvirae | Uroviricota | Caudoviricetes | n.a. | Unclassified | O | O | 61.19 | d__Bacteria;p__Firmicutes;c__Clostridia;o__Lachnospirales;f__Lachnospiraceae;g__Bariatricus | 91.6 | blast;93.90 iPHoP-RF;77.30 |
| S9_vs_19689 | ME | n.a. | n.a. | n.a. | n.a. | n.a. | n.a. | n.a. | n.a. | 9.42 | d__Bacteria;p__Firmicutes;c__Clostridia;o__Oscillospirales;f__Butyricicoccaceae;g__Agathobaculum | 91.6 | blast;93.90 iPHoP-RF;68.60 |
| S9_vs_21850 | ME | Duplodnaviria | Heunggongvirae | Uroviricota | Caudoviricetes | n.a. | Unclassified | Unclassified | Unclassified | 63.18 | d__Bacteria;p__Bacteroidota;c__Bacteroidia;o__Bacteroidales;f__Barnesiellaceae;g__Barnesiella | 97.2 | blast;96.90 iPHoP-RF;85.10 CRISPR;84.10 RaFAH;68.00 |
| S9_vs_26179 | ME | n.a. | n.a. | n.a. | n.a. | n.a. | n.a. | n.a. | n.a. | 48.62 | d__Bacteria;p__Firmicutes;c__Clostridia;o__Oscillospirales;f__Acutalibacteraceae;g__UMGS1071 | 95.7 | blast;97.00 iPHoP-RF;83.30 CRISPR;81.20 |
| S9_vs_2799 | Shared | n.a. | n.a. | n.a. | n.a. | n.a. | n.a. | n.a. | n.a. | 10.92 | d__Bacteria;p__Bacteroidota;c__Bacteroidia;o__Bacteroidales;f__Rikenellaceae;g__Alistipes | 95.9 | iPHoP-RF;97.10 blast;96.90 |
| S9_vs_28197 | Shared | Duplodnaviria | Heunggongvirae | Uroviricota | Caudoviricetes | n.a. | Unclassified | O | O | 87.18 | d__Bacteria;p__Bacteroidota;c__Bacteroidia;o__Bacteroidales;f__Bacteroidaceae;g__Bacteroides | 100 | RaFAH;99.40 iPHoP-RF;97.10 blast;96.30 |
| S9_vs_32588 | ME | Duplodnaviria | Heunggongvirae | Uroviricota | Caudoviricetes | n.a. | Unclassified | O | O | 67.17 | d__Bacteria;p__Firmicutes;c__Clostridia;o__Lachnospirales;f__Lachnospiraceae;g__Dorea | 95 | RaFAH;96.50 |
| S9_vs_34898 | ME | Duplodnaviria | Heunggongvirae | Uroviricota | Caudoviricetes | n.a. | Unclassified | O | O | 91.82 | d__Bacteria;p__Firmicutes;c__Clostridia;o__Lachnospirales;f__Lachnospiraceae;g__Sellimonas | 93.9 | blast;95.70 iPHoP-RF;89.50 |
| S9_vs_37437 | ME | Duplodnaviria | Heunggongvirae | Uroviricota | Caudoviricetes | n.a. | Unclassified | O | O | 14.86 | d__Bacteria;p__Firmicutes;c__Bacilli;o__Erysipelotrichales;f__Erysipelotrichaceae;g__Holdemania | 91.6 | blast;93.90 CRISPR;91.60 iPHoP-RF;75.40 |
| S9_vs_41371 | ME | Duplodnaviria | Heunggongvirae | Uroviricota | Caudoviricetes | n.a. | Unclassified | O | O | 44.35 | d__Bacteria;p__Firmicutes;c__Clostridia;o__Lachnospirales;f__Lachnospiraceae;g__Enterocloster | 94 | blast;95.80 iPHoP-RF;85.10 |
| S9_vs_4343 | ME | n.a. | n.a. | n.a. | n.a. | n.a. | n.a. | n.a. | n.a. | 24.4 | d__Bacteria;p__Firmicutes;c__Clostridia;o__Lachnospirales;f__Lachnospiraceae;g__Enterocloster | 96.6 | blast;97.60 CRISPR;96.60 iPHoP-RF;90.50 |
| S9_vs_44331 | ME | Duplodnaviria | Heunggongvirae | Uroviricota | Caudoviricetes | n.a. | Unclassified | O | O | 98.52 | d__Bacteria;p__Firmicutes;c__Clostridia;o__Lachnospirales;f__Lachnospiraceae;g__Ruminococcus_B | 98.1 | blast;98.60 iPHoP-RF;94.40 CRISPR;87.70 |
| S9_vs_4558 | ME | Duplodnaviria | Heunggongvirae | Uroviricota | Caudoviricetes | n.a. | Unclassified | O | O | 65.18 | d__Bacteria;p__Firmicutes;c__Clostridia;o__Oscillospirales;f__Oscillospiraceae;g__Flavonifractor | 90.2 | blast;92.70 iPHoP-RF;79.00 |
| S9_vs_46498 | ME | Duplodnaviria | Heunggongvirae | Uroviricota | Caudoviricetes | n.a. | Unclassified | O | O | 26.43 | d__Bacteria;p__Firmicutes;c__Bacilli;o__Erysipelotrichales;f__Erysipelatoclostridiaceae;g__Erysipelatoclostridium | 96.7 | blast;97.70 iPHoP-RF;74.10 |
| S9_vs_48240 | ME | Duplodnaviria | Heunggongvirae | Uroviricota | Caudoviricetes | n.a. | Unclassified | Unclassified | O | 63.98 | d__Bacteria;p__Firmicutes;c__Clostridia;o__Lachnospirales;f__Lachnospiraceae;g__Hungatella | 92.5 | blast;94.60 iPHoP-RF;94.40 |
| S9_vs_48347 | ME | Duplodnaviria | Heunggongvirae | Uroviricota | Caudoviricetes | n.a. | Unclassified | Unclassified | O | 25.89 | d__Bacteria;p__Firmicutes;c__Clostridia;o__Oscillospirales;f__Ruminococcaceae;g__Negativibacillus | 94.2 | blast;95.90 iPHoP-RF;85.40 CRISPR;59.60 |
| S9_vs_48424 | Shared | n.a. | n.a. | n.a. | n.a. | n.a. | n.a. | n.a. | n.a. | 86.27 | d__Bacteria;p__Bacteroidota;c__Bacteroidia;o__Bacteroidales;f__Rikenellaceae;g__Alistipes | 99.3 | blast;96.80 RaFAH;90.20 iPHoP-RF;54.90 |
| S9_vs_50439 | ME | n.a. | n.a. | n.a. | n.a. | n.a. | n.a. | n.a. | n.a. | 93.5 | d__Bacteria;p__Firmicutes;c__Clostridia;o__Oscillospirales;f__Oscillospiraceae;g__Flavonifractor | 98.1 | blast;98.60 iPHoP-RF;74.30 |
| S9_vs_55738 | ME | Duplodnaviria | Heunggongvirae | Uroviricota | Caudoviricetes | n.a. | Unclassified | O | O | 76 | d__Bacteria;p__Firmicutes;c__Clostridia;o__Lachnospirales;f__Lachnospiraceae;g__Enterocloster | 94.6 | blast;96.20 iPHoP-RF;87.00 CRISPR;69.00 |
| S9_vs_5676 | ME | Duplodnaviria | Heunggongvirae | Uroviricota | Caudoviricetes | n.a. | Unclassified | O | O | 19.1 | d__Bacteria;p__Firmicutes;c__Clostridia;o__Oscillospirales;f__Oscillospiraceae;g__Intestinimonas | 92.6 | blast;94.70 iPHoP-RF;74.60 |
| S9_vs_58086 | ME | Duplodnaviria | Heunggongvirae | Uroviricota | Caudoviricetes | n.a. | Unclassified | O | O | 53.1 | d__Bacteria;p__Firmicutes;c__Clostridia;o__Lachnospirales;f__Lachnospiraceae;g__Dorea_A | 94.3 | blast;96.00 iPHoP-RF;80.60 |
| S9_vs_62803 | ME | Duplodnaviria | Heunggongvirae | Uroviricota | Caudoviricetes | n.a. | Unclassified | O | O | 39.71 | d__Bacteria;p__Firmicutes;c__Clostridia;o__Lachnospirales;f__Lachnospiraceae;g__Sellimonas | 94.9 | blast;96.40 iPHoP-RF;84.50 |
| S9_vs_66877 | ME | Duplodnaviria | Heunggongvirae | Uroviricota | Caudoviricetes | n.a. | Unclassified | O | O | 29.56 | d__Bacteria;p__Firmicutes;c__Clostridia;o__Oscillospirales;f__Ruminococcaceae;g__Ruthenibacterium | 95.3 | blast;96.70 CRISPR;95.50 iPHoP-RF;92.10 |
| S9_vs_67792 | ME | Duplodnaviria | Heunggongvirae | Uroviricota | Caudoviricetes | n.a. | Unclassified | O | O | 30.5 | d__Bacteria;p__Firmicutes;c__Negativicutes;o__Veillonellales;f__Dialisteraceae;g__Dialister | 96.6 | blast;97.60 iPHoP-RF;86.00 CRISPR;60.90 |
| S9_vs_69322 | ME | n.a. | n.a. | n.a. | n.a. | n.a. | n.a. | n.a. | n.a. | 20.79 | d__Bacteria;p__Firmicutes;c__Clostridia;o__Oscillospirales;f__Oscillospiraceae;g__Dysosmobacter | 90.7 | blast;93.10 iPHoP-RF;82.20 |
| S9_vs_75050 | ME | Duplodnaviria | Heunggongvirae | Uroviricota | Caudoviricetes | n.a. | Unclassified | O | O | 26.71 | d__Bacteria;p__Firmicutes;c__Clostridia;o__Lachnospirales;f__Lachnospiraceae;g__CAG-127 | 96.4 | CRISPR;97.50 blast;96.80 iPHoP-RF;86.00 |
| S9_vs_83108 | ME | n.a. | n.a. | n.a. | n.a. | n.a. | n.a. | n.a. | n.a. | 82.91 | d__Bacteria;p__Bacteroidota;c__Bacteroidia;o__Bacteroidales;f__Bacteroidaceae;g__Bacteroides | 99.4 | iPHoP-RF;97.10 blast;93.90 RaFAH;91.80 |
| S9_vs_84046 | ME | Duplodnaviria | Heunggongvirae | Uroviricota | Caudoviricetes | n.a. | Unclassified | O | O | 20.91 | d__Bacteria;p__Firmicutes;c__Clostridia;o__Oscillospirales;f__Oscillospiraceae;g__An92 | 90.7 | blast;93.10 iPHoP-RF;70.60 |
| S9_vs_88286 | ME | n.a. | n.a. | n.a. | n.a. | n.a. | n.a. | n.a. | n.a. | 10.88 | d__Bacteria;p__Firmicutes;c__Clostridia;o__Lachnospirales;f__Lachnospiraceae;g__Eisenbergiella | 98.1 | CRISPR;98.60 blast;95.70 iPHoP-RF;95.40 |
| S9_vs_90727 | Shared | Duplodnaviria | Heunggongvirae | Uroviricota | Caudoviricetes | n.a. | Unclassified | Unclassified | O | 57.49 | d__Bacteria;p__Firmicutes;c__Clostridia;o__Oscillospirales;f__Acutalibacteraceae;g__Ruminococcus_E | 95.6 | blast;96.90 iPHoP-RF;88.20 |
| S9_vs_90846 | ME | Duplodnaviria | Heunggongvirae | Uroviricota | Caudoviricetes | n.a. | Unclassified | O | O | 37.71 | d__Bacteria;p__Firmicutes;c__Bacilli;o__Erysipelotrichales;f__Erysipelotrichaceae;g__Clostridium_AQ | 97.5 | blast;98.20 iPHoP-RF;62.70 |
| S9_vs_90848 | Shared | Duplodnaviria | Heunggongvirae | Uroviricota | Caudoviricetes | n.a. | Unclassified | O | O | 51.41 | d__Bacteria;p__Firmicutes;c__Clostridia;o__Lachnospirales;f__Lachnospiraceae;g__Enterocloster | 98.7 | CRISPR;99.00 blast;93.90 iPHoP-RF;91.40 |
| S9_vs_90849 | ME | n.a. | n.a. | n.a. | n.a. | n.a. | n.a. | n.a. | n.a. | 70.89 | d__Bacteria;p__Firmicutes;c__Clostridia;o__Lachnospirales;f__Anaerotignaceae;g__Anaerotignum | 95 | CRISPR;96.50 blast;93.70 iPHoP-RF;92.10 |
| S9_vs_90851 | ME | Duplodnaviria | Heunggongvirae | Uroviricota | Caudoviricetes | n.a. | Unclassified | O | O | 89.56 | d__Bacteria;p__Firmicutes;c__Clostridia;o__Clostridiales;f__Clostridiaceae;g__Clostridium | 95.6 | RaFAH;96.90 |
| S9_vs_90859 | ME | Unclassified | n.a. | n.a. | n.a. | n.a. | Unclassified | O | O | 43.06 | d__Bacteria;p__Firmicutes;c__Clostridia;o__Lachnospirales;f__Lachnospiraceae;g__Blautia_A | 95.3 | blast;93.90 iPHoP-RF;75.90 RaFAH;73.20 |
| S9_vs_90860 | ME | Duplodnaviria | Heunggongvirae | Uroviricota | Caudoviricetes | n.a. | Unclassified | O | O | 20.66 | d__Bacteria;p__Firmicutes;c__Clostridia;o__Oscillospirales;f__Acutalibacteraceae;g__UMGS1591 | 93.4 | blast;95.30 iPHoP-RF;67.00 |

4.WMS_STD+MAGs_tophits

| Virus | Status | Realm | Kingdom | Phylum | Class | Order | Family | Subfamily | Genus | AAI to closest RaFAH reference | Host genus | Confidence score | List of methods |
|---|---|---|---|---|---|---|---|---|---|---|---|---|---|
| S1-wms-combined_118167 | Shared | Duplodnaviria | Heunggongvirae | Uroviricota | Caudoviricetes | n.a. | Unclassified | O | O | 33.35 | d__Bacteria;p__Firmicutes;c__Clostridia;o__Oscillospirales;f__Ruminococcaceae;g__Faecalibacterium | 99.1 | CRISPR;96.40 blast;95.10 RaFAH;90.80 iPHoP-RF;89.50 |
| S1-wms-combined_100430 | ME | Duplodnaviria | Heunggongvirae | Uroviricota | Caudoviricetes | n.a. | Unclassified | Unclassified | O | 99.71 | d__Bacteria;p__Firmicutes;c__Clostridia;o__Lachnospirales;f__Lachnospiraceae;g__Lachnospira | 99.3 | CRISPR;98.90 RaFAH;73.00 |
| S1-wms-combined_131270 | ME | Duplodnaviria | Heunggongvirae | Uroviricota | Caudoviricetes | n.a. | Unclassified | O | O | 21.6 | d__Bacteria;p__Firmicutes;c__Clostridia;o__Oscillospirales;f__Ruminococcaceae;g__Faecalibacterium | 99 | CRISPR;99.20 blast;94.50 iPHoP-RF;84.50 |
| S1-wms-combined_132934 | Shared | Duplodnaviria | Heunggongvirae | Uroviricota | Caudoviricetes | n.a. | Unclassified | O | O | 48.4 | d__Bacteria;p__Firmicutes;c__Clostridia;o__Oscillospirales;f__Oscillospiraceae;g__ER4 | 92.7 | blast;94.80 iPHoP-RF;64.80 |
| S1-wms-combined_136097 | Shared | n.a. | n.a. | n.a. | n.a. | n.a. | n.a. | n.a. | n.a. | 4.5 | d__Bacteria;p__Firmicutes;c__Clostridia;o__Oscillospirales;f__Oscillospiraceae;g__UBA738 | 92 | blast;94.20 iPHoP-RF;53.00 |
| S1-wms-combined_107766 | Shared | n.a. | n.a. | n.a. | n.a. | n.a. | n.a. | n.a. | n.a. | 92.99 | d__Bacteria;p__Bacteroidota;c__Bacteroidia;o__Bacteroidales;f__Rikenellaceae;g__Alistipes | 98.8 | iPHoP-RF;97.10 blast;96.90 RaFAH;83.70 |
| S1-wms-combined_156637 | Shared | n.a. | n.a. | n.a. | n.a. | n.a. | n.a. | n.a. | n.a. | 11 | d__Bacteria;p__Firmicutes;c__Clostridia;o__Oscillospirales;f__Oscillospiraceae;g__ER4 | 93 | blast;95.00 iPHoP-RF;60.30 |
| S1-wms-combined_143459 | ME | Duplodnaviria | Heunggongvirae | Uroviricota | Caudoviricetes | n.a. | Unclassified | O | O | 18.79 | d__Bacteria;p__Desulfobacterota;c__Desulfovibrionia;o__Desulfovibrionales;f__Desulfovibrionaceae;g__Bilophila | 95.2 | blast;96.60 iPHoP-RF;65.80 CRISPR;58.60 |
| S1-wms-combined_159544 | ME | Duplodnaviria | Heunggongvirae | Uroviricota | Caudoviricetes | n.a. | Peduoviridae | Unclassified | Phitrevirus | 36.49 | d__Bacteria;p__Desulfobacterota;c__Desulfovibrionia;o__Desulfovibrionales;f__Desulfovibrionaceae;g__Bilophila | 94 | blast;95.80 iPHoP-RF;79.30 CRISPR;57.40 |
| S1-wms-combined_164485 | Shared | n.a. | n.a. | n.a. | n.a. | n.a. | n.a. | n.a. | n.a. | 4.67 | d__Bacteria;p__Firmicutes;c__Clostridia;o__Oscillospirales;f__Oscillospiraceae;g__ER4 | 90.4 | blast;92.90 |
| S1-wms-combined_185336 | ME | n.a. | n.a. | n.a. | n.a. | n.a. | n.a. | n.a. | n.a. | | d__Bacteria;p__Firmicutes;c__Clostridia;o__Oscillospirales;f__Oscillospiraceae;g__ER4 | 91.7 | blast;94.00 iPHoP-RF;63.50 |
| S1-wms-combined_19031 | ME | Duplodnaviria | Heunggongvirae | Uroviricota | Caudoviricetes | n.a. | Unclassified | Unclassified | Unclassified | 37.63 | d__Bacteria;p__Desulfobacterota;c__Desulfovibrionia;o__Desulfovibrionales;f__Desulfovibrionaceae;g__Bilophila | 98.2 | blast;98.70 iPHoP-RF;76.10 |
| S1-wms-combined_164894 | ME | Duplodnaviria | Heunggongvirae | Uroviricota | Caudoviricetes | n.a. | Unclassified | O | O | 72.02 | d__Bacteria;p__Firmicutes;c__Clostridia;o__Oscillospirales;f__Ruminococcaceae;g__Faecalibacterium | 91.8 | blast;94.10 iPHoP-RF;70.90 |
| S1-wms-combined_23248 | Shared | Duplodnaviria | Heunggongvirae | Uroviricota | Caudoviricetes | n.a. | Unclassified | Unclassified | Lughvirus | 61.01 | d__Bacteria;p__Firmicutes;c__Clostridia;o__Oscillospirales;f__Ruminococcaceae;g__Faecalibacterium | 95 | blast;96.50 iPHoP-RF;68.90 |
| S1-wms-combined_246923 | ME | Duplodnaviria | Heunggongvirae | Uroviricota | Caudoviricetes | n.a. | Unclassified | O | O | 43.06 | d__Bacteria;p__Bacteroidota;c__Bacteroidia;o__Bacteroidales;f__UBA11471;g__UBA11471 | 93.9 | blast;95.70 iPHoP-RF;68.60 |
| S1-wms-combined_153670 | ME | n.a. | n.a. | n.a. | n.a. | n.a. | n.a. | n.a. | n.a. | | d__Bacteria;p__Bacteroidota;c__Bacteroidia;o__Bacteroidales;f__Barnesiellaceae;g__Barnesiella | 94.2 | blast;95.90 iPHoP-RF;72.20 |
| S1-wms-combined_248622 | Shared | n.a. | n.a. | n.a. | n.a. | n.a. | n.a. | n.a. | n.a. | 6.16 | d__Bacteria;p__Bacteroidota;c__Bacteroidia;o__Bacteroidales;f__Bacteroidaceae;g__Bacteroides | 95.9 | iPHoP-RF;97.10 blast;90.50 |
| S1-wms-combined_186878 | ME | n.a. | n.a. | n.a. | n.a. | n.a. | n.a. | n.a. | n.a. | 42.39 | d__Bacteria;p__Firmicutes;c__Clostridia;o__Oscillospirales;f__Oscillospiraceae;g__ER4 | 91.6 | blast;93.90 iPHoP-RF;76.40 |
| S1-wms-combined_263562 | Shared | n.a. | n.a. | n.a. | n.a. | n.a. | n.a. | n.a. | n.a. | 99.98 | d__Bacteria;p__Bacteroidota;c__Bacteroidia;o__Bacteroidales;f__Bacteroidaceae;g__Bacteroides | 100 | RaFAH;99.80 blast;91.80 iPHoP-RF;82.20 |
| S1-wms-combined_302166 | Shared | Duplodnaviria | Heunggongvirae | Uroviricota | Caudoviricetes | n.a. | Unclassified | O | O | 9.7 | d__Bacteria;p__Firmicutes;c__Clostridia;o__Christensenellales;f__CAG-74;g__UBA11524 | 95.6 | blast;96.90 iPHoP-RF;92.40 |
| S1-wms-combined_212892 | ME | Duplodnaviria | Heunggongvirae | Uroviricota | Caudoviricetes | n.a. | Unclassified | O | O | 80.18 | d__Bacteria;p__Firmicutes;c__Clostridia;o__Lachnospirales;f__Lachnospiraceae;g__Anaerobutyricum | 98.1 | CRISPR;98.60 blast;91.50 iPHoP-RF;80.00 |
| S1-wms-combined_166392 | ME | Duplodnaviria | Heunggongvirae | Uroviricota | Caudoviricetes | n.a. | Unclassified | O | O | 37.1 | d__Bacteria;p__Firmicutes;c__Clostridia;o__Oscillospirales;f__Ruminococcaceae;g__Gemmiger | 95.7 | blast;97.00 iPHoP-RF;95.70 CRISPR;78.40 |
| S1-wms-combined_304377 | Shared | Duplodnaviria | Heunggongvirae | Uroviricota | Caudoviricetes | n.a. | Unclassified | Unclassified | Oengusvirus | 48.66 | d__Bacteria;p__Firmicutes;c__Clostridia;o__Oscillospirales;f__Ruminococcaceae;g__Faecalibacterium | 95.6 | blast;96.90 iPHoP-RF;78.50 |
| S1-wms-combined_169788 | Shared | n.a. | n.a. | n.a. | n.a. | n.a. | n.a. | n.a. | n.a. | 5.96 | d__Bacteria;p__Firmicutes;c__Clostridia;o__Oscillospirales;f__Oscillospiraceae;g__CAG-83 | 93.4 | blast;95.30 iPHoP-RF;87.00 |
| S1-wms-combined_220022 | ME | n.a. | n.a. | n.a. | n.a. | n.a. | n.a. | n.a. | n.a. | 6.93 | d__Bacteria;p__Firmicutes;c__Clostridia;o__Oscillospirales;f__Ruminococcaceae;g__Gemmiger | 93.2 | blast;95.20 iPHoP-RF;88.20 CRISPR;76.80 |
| S1-wms-combined_307323 | ME | Duplodnaviria | Heunggongvirae | Uroviricota | Caudoviricetes | n.a. | Unclassified | O | O | 92.27 | d__Bacteria;p__Actinobacteriota;c__Actinomycetia;o__Actinomycetales;f__Bifidobacteriaceae;g__Bifidobacterium | 98.7 | CRISPR;99.00 iPHoP-RF;97.10 blast;96.90 |
| S1-wms-combined_17651 | Shared | n.a. | n.a. | n.a. | n.a. | n.a. | n.a. | n.a. | n.a. | 9.21 | d__Bacteria;p__Firmicutes;c__Clostridia;o__Oscillospirales;f__Oscillospiraceae;g__ER4 | 90.6 | blast;93.00 iPHoP-RF;65.10 |
| S1-wms-combined_227003 | ME | n.a. | n.a. | n.a. | n.a. | n.a. | n.a. | n.a. | n.a. | 41.88 | d__Bacteria;p__Firmicutes;c__Clostridia;o__Oscillospirales;f__Oscillospiraceae;g__CAG-110 | 94.2 | blast;95.90 iPHoP-RF;87.00 |
| S1-wms-combined_227917 | ME | Duplodnaviria | Heunggongvirae | Uroviricota | Caudoviricetes | n.a. | Unclassified | O | O | 27.55 | d__Bacteria;p__Firmicutes;c__Clostridia;o__Oscillospirales;f__Ruminococcaceae;g__Gemmiger | 95.2 | blast;96.60 iPHoP-RF;95.40 CRISPR;92.50 |
| S1-wms-combined_331544 | ME | n.a. | n.a. | n.a. | n.a. | n.a. | n.a. | n.a. | n.a. | 89.75 | d__Bacteria;p__Actinobacteriota;c__Actinomycetia;o__Actinomycetales;f__Bifidobacteriaceae;g__Bifidobacterium | 99.9 | CRISPR;99.60 blast;96.90 iPHoP-RF;96.70 RaFAH;75.50 |
| S1-wms-combined_331896 | Shared | n.a. | n.a. | n.a. | n.a. | n.a. | n.a. | n.a. | n.a. | | d__Bacteria;p__Firmicutes;c__Clostridia;o__Oscillospirales;f__Oscillospiraceae;g__ER4 | 91.6 | blast;93.90 iPHoP-RF;58.40 |
| S1-wms-combined_340558 | Shared | Duplodnaviria | Heunggongvirae | Uroviricota | Caudoviricetes | n.a. | Unclassified | O | O | 85.48 | d__Bacteria;p__Bacteroidota;c__Bacteroidia;o__Bacteroidales;f__Rikenellaceae;g__Alistipes | 99.7 | blast;96.80 iPHoP-RF;96.10 RaFAH;95.90 |
| S1-wms-combined_353757 | ME | Duplodnaviria | Heunggongvirae | Uroviricota | Caudoviricetes | n.a. | Unclassified | O | O | 21.95 | d__Bacteria;p__Firmicutes;c__Clostridia;o__Oscillospirales;f__Oscillospiraceae;g__ER4 | 93.2 | blast;95.20 iPHoP-RF;84.80 |
| S1-wms-combined_279492 | Shared | Duplodnaviria | Heunggongvirae | Uroviricota | Caudoviricetes | n.a. | Unclassified | O | O | 44.35 | d__Bacteria;p__Firmicutes;c__Clostridia;o__Oscillospirales;f__Ruminococcaceae;g__Gemmiger | 97.8 | blast;98.40 iPHoP-RF;96.40 CRISPR;58.00 |
| S1-wms-combined_357373 | ME | Monodnaviria | Sangervirae | Phixviricota | Malgrandaviricetes | Petitvirales | Microviridae | Unclassified | Unclassified | | d__Bacteria;p__Bacteroidota;c__Bacteroidia;o__Bacteroidales;f__Barnesiellaceae;g__Barnesiella | 91.8 | blast;94.10 iPHoP-RF;86.30 |
| S1-wms-combined_360285 | Shared | Duplodnaviria | Heunggongvirae | Uroviricota | Caudoviricetes | n.a. | Unclassified | O | O | 17.11 | d__Bacteria;p__Desulfobacterota;c__Desulfovibrionia;o__Desulfovibrionales;f__Desulfovibrionaceae;g__Bilophila | 97.2 | CRISPR;98.00 blast;96.60 iPHoP-RF;89.80 |
| S1-wms-combined_394574 | Shared | Duplodnaviria | Heunggongvirae | Uroviricota | Caudoviricetes | n.a. | Unclassified | Unclassified | Unclassified | 96.28 | d__Bacteria;p__Bacteroidota;c__Bacteroidia;o__Bacteroidales;f__Tannerellaceae;g__Parabacteroides | 91.6 | blast;93.90 iPHoP-RF;75.70 |
| S1-wms-combined_308848 | Shared | n.a. | n.a. | n.a. | n.a. | n.a. | n.a. | n.a. | n.a. | 5.29 | d__Bacteria;p__Verrucomicrobiota;c__Verrucomicrobiae;o__Verrucomicrobiales;f__Akkermansiaceae;g__Akkermansia | 95.9 | CRISPR;97.10 blast;96.90 iPHoP-RF;93.10 |
| S1-wms-combined_231411 | ME | n.a. | n.a. | n.a. | n.a. | n.a. | n.a. | n.a. | n.a. | 3.46 | d__Bacteria;p__Desulfobacterota;c__Desulfovibrionia;o__Desulfovibrionales;f__Desulfovibrionaceae;g__Bilophila | 93 | blast;95.00 iPHoP-RF;73.20 CRISPR;51.00 |
| S1-wms-combined_40115 | ME | n.a. | n.a. | n.a. | n.a. | n.a. | n.a. | n.a. | n.a. | 17.83 | d__Bacteria;p__Firmicutes;c__Clostridia;o__Oscillospirales;f__Oscillospiraceae;g__ER4 | 97 | blast;97.90 iPHoP-RF;71.70 |
| S1-wms-combined_41369 | ME | Duplodnaviria | Heunggongvirae | Uroviricota | Caudoviricetes | n.a. | Unclassified | O | O | 37.96 | d__Bacteria;p__Firmicutes;c__Clostridia;o__Oscillospirales;f__Oscillospiraceae;g__ER4 | 94.6 | blast;96.20 iPHoP-RF;60.60 |
| S1-wms-combined_68998 | ME | Duplodnaviria | Heunggongvirae | Uroviricota | Caudoviricetes | n.a. | Unclassified | Unclassified | Taranisvirus | 25.66 | d__Bacteria;p__Firmicutes;c__Clostridia;o__Oscillospirales;f__Ruminococcaceae;g__Faecalibacterium | 99.9 | CRISPR;98.60 blast;92.00 iPHoP-RF;91.40 RaFAH;91.30 |
| S1-wms-combined_72167 | ME | Duplodnaviria | Heunggongvirae | Uroviricota | Caudoviricetes | n.a. | Unclassified | O | O | 16.24 | d__Bacteria;p__Firmicutes;c__Clostridia;o__Oscillospirales;f__Oscillospiraceae;g__CAG-83 | 98.5 | CRISPR;98.90 blast;95.00 iPHoP-RF;90.80 |
| S1-wms_130214 | ME | Duplodnaviria | Heunggongvirae | Uroviricota | Caudoviricetes | n.a. | Unclassified | O | O | 33.32 | d__Bacteria;p__Firmicutes;c__Clostridia;o__Oscillospirales;f__Oscillospiraceae;g__CAG-170 | 90.6 | blast;93.00 iPHoP-RF;90.50 CRISPR;82.90 |
| S1-wms_185046 | Shared | n.a. | n.a. | n.a. | n.a. | n.a. | n.a. | n.a. | n.a. | 36.29 | d__Bacteria;p__Firmicutes;c__Clostridia;o__Oscillospirales;f__Oscillospiraceae;g__CAG-170 | 91.1 | blast;93.50 iPHoP-RF;82.20 |
| S1-wms_244836 | Shared | n.a. | n.a. | n.a. | n.a. | n.a. | n.a. | n.a. | n.a. | 99.89 | d__Bacteria;p__Bacteroidota;c__Bacteroidia;o__Bacteroidales;f__Bacteroidaceae;g__Bacteroides | 100 | RaFAH;99.60 iPHoP-RF;96.40 blast;94.20 |
| S1-wms-combined_285012 | Shared | Duplodnaviria | Heunggongvirae | Uroviricota | Caudoviricetes | n.a. | Unclassified | Unclassified | O | 37.37 | d__Bacteria;p__Firmicutes;c__Clostridia;o__Oscillospirales;f__Ruminococcaceae;g__Faecalibacterium | 95 | CRISPR;96.50 iPHoP-RF;54.40 |
| S1-wms_296850 | Shared | Duplodnaviria | Heunggongvirae | Uroviricota | Caudoviricetes | n.a. | Unclassified | Unclassified | Toutatisvirus | 38.32 | d__Bacteria;p__Firmicutes;c__Clostridia;o__Oscillospirales;f__Oscillospiraceae;g__ER4 | 93.1 | blast;95.10 iPHoP-RF;63.80 |
| S1-wms_4277 | Shared | Duplodnaviria | Heunggongvirae | Uroviricota | Caudoviricetes | n.a. | Unclassified | Unclassified | Mushuvirus | 93.43 | d__Bacteria;p__Firmicutes;c__Clostridia;o__Oscillospirales;f__Oscillospiraceae;g__Lawsonibacter | 91.4 | blast;93.70 |
| S11-wms-combined_119215 | ME | Duplodnaviria | Heunggongvirae | Uroviricota | Caudoviricetes | n.a. | Unclassified | O | O | 29.67 | d__Bacteria;p__Firmicutes;c__Clostridia;o__Oscillospirales;f__Oscillospiraceae;g__An92 | 94.9 | iPHoP-RF;96.40 blast;92.70 |
| S1-wms-combined_57370 | Shared | Duplodnaviria | Heunggongvirae | Uroviricota | Caudoviricetes | n.a. | Unclassified | O | O | 50.73 | d__Bacteria;p__Firmicutes;c__Clostridia;o__Oscillospirales;f__Oscillospiraceae;g__CAG-83 | 91.7 | blast;89.00 RaFAH;72.80 |
| S11-wms-combined_125176 | Shared | Duplodnaviria | Heunggongvirae | Uroviricota | Caudoviricetes | n.a. | Unclassified | O | O | 94.89 | d__Bacteria;p__Bacteroidota;c__Bacteroidia;o__Bacteroidales;f__Rikenellaceae;g__Alistipes | 96.9 | CRISPR;97.80 iPHoP-RF;97.10 blast;96.90 |
| S11-wms-combined_140009 | Shared | Duplodnaviria | Heunggongvirae | Uroviricota | Caudoviricetes | n.a. | Unclassified | Unclassified | Lughvirus | 58.05 | d__Bacteria;p__Firmicutes;c__Clostridia;o__Oscillospirales;f__Ruminococcaceae;g__Faecalibacterium | 95.6 | blast;96.90 iPHoP-RF;71.90 |
| S11-wms-combined_154845 | ME | Duplodnaviria | Heunggongvirae | Uroviricota | Caudoviricetes | n.a. | Peduoviridae | Unclassified | Quadragintavirus | 84.31 | d__Bacteria;p__Proteobacteria;c__Gammaproteobacteria;o__Enterobacterales;f__Enterobacteriaceae;g__Salmonella | 95.4 | blast;96.80 |
| S1-wms-combined_32863 | Shared | n.a. | n.a. | n.a. | n.a. | n.a. | n.a. | n.a. | n.a. | 10.89 | d__Bacteria;p__Bacteroidota;c__Bacteroidia;o__Bacteroidales;f__Tannerellaceae;g__Parabacteroides | 95.6 | blast;96.90 iPHoP-RF;69.50 |
| S1-wms-combined_85225 | ME | Duplodnaviria | Heunggongvirae | Uroviricota | Caudoviricetes | n.a. | Unclassified | O | O | 64.47 | d__Bacteria;p__Actinobacteriota;c__Actinomycetia;o__Actinomycetales;f__Bifidobacteriaceae;g__Bifidobacterium | 99.6 | CRISPR;99.60 iPHoP-RF;97.10 blast;96.90 |
| S1-wms-combined_330329 | ME | Duplodnaviria | Heunggongvirae | Uroviricota | Caudoviricetes | n.a. | Unclassified | O | O | 86.77 | d__Bacteria;p__Bacteroidota;c__Bacteroidia;o__Bacteroidales;f__Bacteroidaceae;g__Bacteroides | 98.2 | RaFAH;97.70 iPHoP-RF;72.40 |
| S1-wms-combined_92523 | ME | Unclassified | n.a. | n.a. | n.a. | n.a. | Unclassified | O | O | 13.27 | d__Bacteria;p__Proteobacteria;c__Alphaproteobacteria;o__RF32;f__CAG-239;g__51-20 | 95 | blast;96.50 iPHoP-RF;71.30 |
| S11-wms-combined_172889 | ME | Duplodnaviria | Heunggongvirae | Uroviricota | Caudoviricetes | n.a. | Unclassified | O | O | 100 | d__Bacteria;p__Bacteroidota;c__Bacteroidia;o__Bacteroidales;f__Rikenellaceae;g__Alistipes | 100 | RaFAH;99.10 blast;97.90 iPHoP-RF;94.70 |
| S1-wms-combined_94988 | Shared | n.a. | n.a. | n.a. | n.a. | n.a. | n.a. | n.a. | n.a. | 21.16 | d__Bacteria;p__Firmicutes;c__Clostridia;o__Oscillospirales;f__Oscillospiraceae;g__CAG-83 | 95.6 | blast;96.90 iPHoP-RF;94.10 |
| S11-wms-combined_193148 | Shared | n.a. | n.a. | n.a. | n.a. | n.a. | n.a. | n.a. | n.a. | 97.83 | d__Bacteria;p__Proteobacteria;c__Alphaproteobacteria;o__RF32;f__CAG-239;g__CAG-267 | 94.9 | blast;96.40 iPHoP-RF;75.00 |
| S1-wms_102342 | Shared | Duplodnaviria | Heunggongvirae | Uroviricota | Caudoviricetes | n.a. | Unclassified | O | O | 14.71 | d__Bacteria;p__Firmicutes;c__Clostridia;o__Oscillospirales;f__Oscillospiraceae;g__Dysosmobacter | 95 | blast;96.50 iPHoP-RF;60.30 |
| S1-wms-combined_351889 | Shared | Duplodnaviria | Heunggongvirae | Uroviricota | Caudoviricetes | n.a. | Unclassified | Unclassified | O | 29.41 | d__Bacteria;p__Firmicutes;c__Clostridia;o__Oscillospirales;f__Oscillospiraceae;g__ER4 | 96.2 | CRISPR;97.30 blast;96.20 iPHoP-RF;84.50 |
| S11-wms-combined_195425 | ME | Duplodnaviria | Heunggongvirae | Uroviricota | Caudoviricetes | n.a. | Unclassified | Unclassified | Unclassified | 89.45 | d__Bacteria;p__Bacteroidota;c__Bacteroidia;o__Bacteroidales;f__Bacteroidaceae;g__Bacteroides | 96.2 | blast;90.20 RaFAH;86.00 |
| S11-wms-combined_200084 | ME | Duplodnaviria | Heunggongvirae | Uroviricota | Caudoviricetes | n.a. | Unclassified | O | O | 31.8 | d__Bacteria;p__Firmicutes;c__Clostridia;o__Lachnospirales;f__Lachnospiraceae;g__Blautia_A | 91.8 | blast;94.10 iPHoP-RF;53.60 |
| S1-wms_208875 | ME | Duplodnaviria | Heunggongvirae | Uroviricota | Caudoviricetes | n.a. | Unclassified | O | O | 95.4 | d__Bacteria;p__Bacteroidota;c__Bacteroidia;o__Bacteroidales;f__Bacteroidaceae;g__Bacteroides | 95.7 | blast;95.00 iPHoP-RF;94.40 RaFAH;70.30 |
| S11-wms-combined_204522 | ME | Duplodnaviria | Heunggongvirae | Uroviricota | Caudoviricetes | n.a. | Unclassified | O | O | 15.39 | d__Bacteria;p__Proteobacteria;c__Alphaproteobacteria;o__RF32;f__CAG-239;g__CAG-267 | 91.7 | blast;94.00 iPHoP-RF;72.60 |
| S11-wms-combined_217790 | Shared | n.a. | n.a. | n.a. | n.a. | n.a. | n.a. | n.a. | n.a. | 99.99 | d__Bacteria;p__Bacteroidota;c__Bacteroidia;o__Bacteroidales;f__Bacteroidaceae;g__Bacteroides | 99.9 | RaFAH;99.80 |
| S11-wms-combined_373308 | Shared | n.a. | n.a. | n.a. | n.a. | n.a. | n.a. | n.a. | n.a. | 6.25 | d__Bacteria;p__Firmicutes;c__Clostridia;o__Oscillospirales;f__Oscillospiraceae;g__CAG-83 | 97.9 | blast;98.50 iPHoP-RF;77.30 |
| S1-wms_236105 | Shared | Duplodnaviria | Heunggongvirae | Uroviricota | Caudoviricetes | n.a. | Unclassified | O | O | 71.3 | d__Bacteria;p__Firmicutes;c__Clostridia;o__Oscillospirales;f__Oscillospiraceae;g__ER4 | 92.1 | blast;94.30 iPHoP-RF;89.80 CRISPR;65.50 |
| S1-wms-combined_374219 | ME | n.a. | n.a. | n.a. | n.a. | n.a. | n.a. | n.a. | n.a. | 10.38 | d__Bacteria;p__Firmicutes;c__Clostridia;o__Oscillospirales;f__Oscillospiraceae;g__Flavonifractor | 91.5 | blast;93.80 iPHoP-RF;79.80 |
| S11-wms-combined_268967 | ME | Duplodnaviria | Heunggongvirae | Uroviricota | Caudoviricetes | n.a. | Unclassified | Unclassified | Unclassified | 35.57 | d__Bacteria;p__Desulfobacterota;c__Desulfovibrionia;o__Desulfovibrionales;f__Desulfovibrionaceae;g__Desulfovibrio | 93.8 | blast;95.10 iPHoP-RF;81.90 RaFAH;54.80 CRISPR;51.90 |
| S1-wms_29288 | ME | Duplodnaviria | Heunggongvirae | Uroviricota | Caudoviricetes | n.a. | Unclassified | O | O | 41.26 | d__Bacteria;p__Firmicutes;c__Clostridia;o__Oscillospirales;f__Ruminococcaceae;g__Faecalibacterium | 91.6 | blast;93.90 CRISPR;78.00 iPHoP-RF;75.70 |
| S11-wms-combined_283220 | ME | Duplodnaviria | Heunggongvirae | Uroviricota | Caudoviricetes | n.a. | Unclassified | Unclassified | Mushuvirus | 78.49 | d__Bacteria;p__Firmicutes;c__Clostridia;o__Oscillospirales;f__Oscillospiraceae;g__Dysosmobacter | 91.4 | blast;93.70 iPHoP-RF;83.60 |
| S11-wms-combined_299954 | ME | n.a. | n.a. | n.a. | n.a. | n.a. | n.a. | n.a. | n.a. | 23.91 | d__Bacteria;p__Firmicutes;c__Clostridia;o__Oscillospirales;f__Oscillospiraceae;g__Intestinimonas | 90.2 | blast;92.70 iPHoP-RF;84.20 |
| S1-wms_394571 | Shared | Duplodnaviria | Heunggongvirae | Uroviricota | Caudoviricetes | n.a. | Unclassified | O | O | 22.14 | d__Bacteria;p__Bacteroidota;c__Bacteroidia;o__Bacteroidales;f__Rikenellaceae;g__Alistipes | 95.4 | CRISPR;96.80 iPHoP-RF;71.90 |
| S11-wms-combined_41367 | ME | n.a. | n.a. | n.a. | n.a. | n.a. | n.a. | n.a. | n.a. | 40.35 | d__Bacteria;p__Bacteroidota;c__Bacteroidia;o__Bacteroidales;f__Bacteroidaceae;g__Bacteroides | 98.5 | RaFAH;97.90 iPHoP-RF;74.10 |
| S1-wms-combined_52708 | Shared | n.a. | n.a. | n.a. | n.a. | n.a. | n.a. | n.a. | n.a. | 55.79 | d__Bacteria;p__Firmicutes;c__Clostridia;o__Oscillospirales;f__Ruminococcaceae;g__Faecalibacterium | 96.3 | blast;97.10 iPHoP-RF;70.00 RaFAH;54.80 |
| S11-wms-combined_101337 | Shared | Duplodnaviria | Heunggongvirae | Uroviricota | Caudoviricetes | n.a. | Unclassified | O | O | 98.94 | d__Bacteria;p__Proteobacteria;c__Alphaproteobacteria;o__RF32;f__CAG-239;g__CAG-495 | 91.6 | blast;93.90 iPHoP-RF;86.30 CRISPR;51.40 |
| S11-wms-combined_46816 | Shared | n.a. | n.a. | n.a. | n.a. | n.a. | n.a. | n.a. | n.a. | 20.69 | d__Bacteria;p__Firmicutes;c__Clostridia;o__Oscillospirales;f__Ruminococcaceae;g__Gemmiger | 95.4 | blast;96.80 iPHoP-RF;91.40 CRISPR;69.40 |
| S11-wms-combined_112733 | Shared | Duplodnaviria | Heunggongvirae | Uroviricota | Caudoviricetes | n.a. | Unclassified | O | O | 59.77 | d__Bacteria;p__Desulfobacterota;c__Desulfovibrionia;o__Desulfovibrionales;f__Desulfovibrionaceae;g__Desulfovibrio | 97.9 | blast;96.30 iPHoP-RF;88.90 RaFAH;79.80 CRISPR;67.90 |
| S11-wms-combined_48246 | Shared | Duplodnaviria | Heunggongvirae | Uroviricota | Caudoviricetes | n.a. | Unclassified | O | O | 99.82 | d__Bacteria;p__Bacteroidota;c__Bacteroidia;o__Bacteroidales;f__Rikenellaceae;g__Alistipes | 100 | blast;98.50 RaFAH;96.00 iPHoP-RF;95.40 |
| S11-wms-combined_116589 | Shared | n.a. | n.a. | n.a. | n.a. | n.a. | n.a. | n.a. | n.a. | 25.47 | d__Bacteria;p__Firmicutes;c__Clostridia;o__Oscillospirales;f__Oscillospiraceae;g__Dysosmobacter | 91.6 | blast;93.90 iPHoP-RF;88.90 |
| S11-wms-combined_49632 | ME | Duplodnaviria | Heunggongvirae | Uroviricota | Caudoviricetes | n.a. | Unclassified | O | O | 14.66 | d__Bacteria;p__Firmicutes;c__Clostridia;o__Oscillospirales;f__Oscillospiraceae;g__Dysosmobacter | 91.5 | blast;93.80 iPHoP-RF;84.80 |
| S11-wms-combined_73204 | Shared | n.a. | n.a. | n.a. | n.a. | n.a. | n.a. | n.a. | n.a. | 100 | d__Bacteria;p__Bacteroidota;c__Bacteroidia;o__Bacteroidales;f__Bacteroidaceae;g__Bacteroides | 99.4 | RaFAH;99.30 iPHoP-RF;66.50 |
| S11-wms-combined_83571 | Shared | Duplodnaviria | Heunggongvirae | Uroviricota | Caudoviricetes | n.a. | Winoviridae | O | O | 99.96 | d__Bacteria;p__Bacteroidota;c__Bacteroidia;o__Bacteroidales;f__Rikenellaceae;g__Alistipes | 100 | RaFAH;98.50 blast;98.40 iPHoP-RF;96.70 |
| S11-wms-combined_84326 | ME | Duplodnaviria | Heunggongvirae | Uroviricota | Caudoviricetes | n.a. | Unclassified | O | O | 16.31 | d__Bacteria;p__Desulfobacterota;c__Desulfovibrionia;o__Desulfovibrionales;f__Desulfovibrionaceae;g__Desulfovibrio | 94.2 | blast;95.90 iPHoP-RF;89.50 CRISPR;83.30 |
| S11-wms-combined_168048 | Shared | Duplodnaviria | Heunggongvirae | Uroviricota | Caudoviricetes | n.a. | Unclassified | O | O | 95.9 | d__Bacteria;p__Firmicutes;c__Clostridia;o__Oscillospirales;f__Ruminococcaceae;g__Ruminiclostridium_E | 93.4 | blast;95.30 iPHoP-RF;66.80 |
| S11-wms-combined_92114 | Shared | n.a. | n.a. | n.a. | n.a. | n.a. | n.a. | n.a. | n.a. | 19.84 | d__Bacteria;p__Bacteroidota;c__Bacteroidia;o__Bacteroidales;f__Rikenellaceae;g__Alistipes | 97.2 | CRISPR;98.00 iPHoP-RF;97.10 blast;96.90 |
| S1-wms_153615 | ME | n.a. | n.a. | n.a. | n.a. | n.a. | n.a. | n.a. | n.a. | | d__Bacteria;p__Bacteroidota;c__Bacteroidia;o__Bacteroidales;f__Barnesiellaceae;g__Barnesiella | 93.8 | blast;95.60 iPHoP-RF;91.10 |
| S11-wms_14390 | ME | n.a. | n.a. | n.a. | n.a. | n.a. | n.a. | n.a. | n.a. | 96.43 | d__Bacteria;p__Firmicutes;c__Clostridia;o__Oscillospirales;f__Oscillospiraceae;g__CAG-83 | 97.3 | RaFAH;98.10 |
| S11-wms_3116 | Shared | Duplodnaviria | Heunggongvirae | Uroviricota | Caudoviricetes | n.a. | Unclassified | Unclassified | Mushuvirus | 99.5 | d__Bacteria;p__Firmicutes;c__Clostridia;o__Oscillospirales;f__Ruminococcaceae;g__Faecalibacterium | 100 | RaFAH;99.60 blast;92.10 |
| S11-wms_320555 | ME | Duplodnaviria | Heunggongvirae | Uroviricota | Caudoviricetes | n.a. | Unclassified | O | O | 99.4 | d__Bacteria;p__Bacteroidota;c__Bacteroidia;o__Bacteroidales;f__Bacteroidaceae;g__Bacteroides | 99.7 | RaFAH;95.40 blast;92.70 iPHoP-RF;74.60 |
| S11-wms_330479 | ME | Unclassified | n.a. | n.a. | n.a. | n.a. | Unclassified | Unclassified | Unclassified | 20.29 | d__Bacteria;p__Bacteroidota;c__Bacteroidia;o__Bacteroidales;f__Rikenellaceae;g__Alistipes | 98.5 | CRISPR;98.90 iPHoP-RF;78.50 |
| S11-wms-combined_215723 | Shared | Duplodnaviria | Heunggongvirae | Uroviricota | Caudoviricetes | n.a. | Duneviridae | O | O | 94.77 | d__Bacteria;p__Bacteroidota;c__Bacteroidia;o__Bacteroidales;f__Rikenellaceae;g__Alistipes | 99.4 | iPHoP-RF;97.10 blast;96.90 RaFAH;91.40 |
| S1-wms_223620 | Shared | Duplodnaviria | Heunggongvirae | Uroviricota | Caudoviricetes | n.a. | Unclassified | O | O | 12.07 | d__Bacteria;p__Firmicutes;c__Clostridia;o__Christensenellales;f__CAG-74;g__UBA11524 | 97.6 | blast;98.30 CRISPR;88.60 iPHoP-RF;86.00 |
| S12-wms-combined_111524 | Shared | n.a. | n.a. | n.a. | n.a. | n.a. | n.a. | n.a. | n.a. | 100 | d__Bacteria;p__Bacteroidota;c__Bacteroidia;o__Bacteroidales;f__Bacteroidaceae;g__Bacteroides | 99.6 | RaFAH;99.60 |
| S12-wms-combined_120133 | Shared | Duplodnaviria | Heunggongvirae | Uroviricota | Caudoviricetes | n.a. | Unclassified | O | O | 82.27 | d__Bacteria;p__Firmicutes;c__Clostridia;o__Lachnospirales;f__Lachnospiraceae;g__Lachnospira | 99.1 | CRISPR;97.80 blast;97.00 iPHoP-RF;96.40 RaFAH;84.70 |
| S11-wms-combined_236214 | Shared | Duplodnaviria | Heunggongvirae | Uroviricota | Caudoviricetes | n.a. | Unclassified | O | O | 48.8 | d__Bacteria;p__Firmicutes;c__Clostridia;o__Oscillospirales;f__Ruminococcaceae;g__Faecalibacterium | 94.6 | blast;96.20 iPHoP-RF;57.60 |
| S12-wms-combined_129554 | SHHC | Duplodnaviria | Heunggongvirae | Uroviricota | Caudoviricetes | n.a. | Unclassified | Unclassified | Eponavirus | 47.38 | d__Bacteria;p__Firmicutes;c__Clostridia;o__Oscillospirales;f__Oscillospiraceae;g__F23-B02 | 95 | blast;96.50 iPHoP-RF;75.90 |
| S11-wms-combined_264050 | Shared | Duplodnaviria | Heunggongvirae | Uroviricota | Caudoviricetes | n.a. | Unclassified | Unclassified | O | 47.18 | d__Bacteria;p__Firmicutes;c__Clostridia;o__Oscillospirales;f__Ruminococcaceae;g__Gemmiger | 97.5 | blast;98.20 iPHoP-RF;93.40 CRISPR;91.60 |

| | | | | | | | | | | | | | |
|---|---|---|---|---|---|---|---|---|---|---|---|---|---|
| S12-wms-combined_147628 | SHHC | Duplodnaviria | Heunggongvirae | Uroviricota | Caudoviricetes | n.a. | Unclassified | O | O | 13.34 | d__Bacteria;p__Firmicutes;c__Clostridia;o__Christensenellales;f__CAG-138;g__SFEL01 | 96 | CRISPR;97.20 blast;93.30 iPHoP-RF;69.80 |
| S11-wms-combined_267993 | ME | Duplodnaviria | Heunggongvirae | Uroviricota | Caudoviricetes | n.a. | Unclassified | O | O | 26.2 | d__Bacteria;p__Firmicutes;c__Clostridia;o__Christensenellales;f__CAG-74;g__Firm-11 | 91.5 | blast;93.80 iPHoP-RF;75.20 |
| S12-wms-combined_168259 | Shared | Duplodnaviria | Heunggongvirae | Uroviricota | Caudoviricetes | n.a. | Unclassified | O | O | 10.85 | d__Bacteria;p__Verrucomicrobiota;c__Verrucomicrobiae;o__Verrucomicrobiales;f__Akkermansiaceae;g__Akkermansia | 100 | RaFAH;98.70 blast;96.90 CRISPR;95.20 iPHoP-RF;87.60 |
| S11-wms-combined_282227 | ME | Duplodnaviria | Heunggongvirae | Uroviricota | Caudoviricetes | n.a. | Unclassified | O | O | 60.6 | d__Bacteria;p__Firmicutes;c__Clostridia;o__Lachnospirales;f__Lachnospiraceae;g__Mediterraneibacter | 91.2 | blast;93.60 iPHoP-RF;73.00 |
| S12-wms-combined_210942 | Shared | n.a. | n.a. | n.a. | n.a. | n.a. | n.a. | n.a. | n.a. | 9.09 | d__Bacteria;p__Firmicutes;c__Clostridia;o__Oscillospirales;f__Oscillospiraceae;g__ER4 | 96.3 | blast;97.40 |
| S11-wms-combined_329493 | Shared | n.a. | n.a. | n.a. | n.a. | n.a. | n.a. | n.a. | n.a. | 9.74 | d__Bacteria;p__Firmicutes;c__Clostridia;o__Oscillospirales;f__Oscillospiraceae;g__Faecousia | 96.7 | blast;97.70 iPHoP-RF;80.00 |
| S12-wms-combined_2450 | Shared | Duplodnaviria | Heunggongvirae | Uroviricota | Caudoviricetes | n.a. | Winoviridae | O | O | 57.23 | d__Bacteria;p__Bacteroidota;c__Bacteroidia;o__Bacteroidales;f__Rikenellaceae;g__Tidjanibacter | 90.2 | blast;92.70 iPHoP-RF;50.70 |
| S11-wms-combined_3507 | Shared | Duplodnaviria | Heunggongvirae | Uroviricota | Caudoviricetes | n.a. | Unclassified | O | O | 24.17 | d__Bacteria;p__Firmicutes;c__Clostridia;o__Oscillospirales;f__Oscillospiraceae;g__CAG-83 | 98.1 | CRISPR;98.60 blast;96.90 iPHoP-RF;92.80 |
| S12-wms-combined_250551 | Shared | Duplodnaviria | Heunggongvirae | Uroviricota | Caudoviricetes | n.a. | Unclassified | Unclassified | Unclassified | 98.55 | d__Bacteria;p__Bacteroidota;c__Bacteroidia;o__Bacteroidales;f__Bacteroidaceae;g__Bacteroides | 100 | RaFAH;99.40 blast;92.10 iPHoP-RF;83.90 |
| S12-wms-combined_256299 | SHHC | n.a. | n.a. | n.a. | n.a. | n.a. | n.a. | n.a. | n.a. | 44.45 | d__Bacteria;p__Bacteroidota;c__Bacteroidia;o__Bacteroidales;f__Muribaculaceae;g__UBA7173 | 90.1 | blast;92.60 iPHoP-RF;75.70 |
| S12-wms-combined_259783 | Shared | Duplodnaviria | Heunggongvirae | Uroviricota | Caudoviricetes | n.a. | Unclassified | Unclassified | Unclassified | 81.56 | d__Bacteria;p__Firmicutes;c__Clostridia;o__Lachnospirales;f__Lachnospiraceae;g__Roseburia | 96.7 | CRISPR;97.70 blast;92.20 iPHoP-RF;70.60 |
| S12-wms-combined_260036 | Shared | Duplodnaviria | Heunggongvirae | Uroviricota | Caudoviricetes | Crassvirales | Steigviridae | O | O | 44.16 | d__Bacteria;p__Bacteroidota;c__Bacteroidia;o__Bacteroidales;f__Barnesiellaceae;g__Barnesiella | 94.6 | blast;96.20 iPHoP-RF;94.10 CRISPR;67.50 |
| S11-wms-combined_63818 | Shared | Duplodnaviria | Heunggongvirae | Uroviricota | Caudoviricetes | n.a. | Unclassified | O | O | 50.37 | d__Bacteria;p__Bacteroidota;c__Bacteroidia;o__Bacteroidales;f__Rikenellaceae;g__Alistipes | 98.4 | iPHoP-RF;97.10 blast;96.90 RaFAH;78.80 |
| S11-wms-combined_120257 | Shared | Duplodnaviria | Heunggongvirae | Uroviricota | Caudoviricetes | n.a. | Unclassified | Unclassified | O | 40.42 | d__Bacteria;p__Firmicutes;c__Clostridia;o__Oscillospirales;f__Oscillospiraceae;g__Flavonifractor | 91.1 | blast;93.50 iPHoP-RF;66.00 |
| S12-wms-combined_263377 | SHHC | Duplodnaviria | Heunggongvirae | Uroviricota | Caudoviricetes | n.a. | Unclassified | O | O | 78.23 | d__Bacteria;p__Firmicutes;c__Clostridia;o__Oscillospirales;f__Acutalibacteraceae;g__UBA1417 | 91.7 | blast;94.00 iPHoP-RF;69.30 |
| S12-wms-combined_264425 | SHHC | n.a. | n.a. | n.a. | n.a. | n.a. | n.a. | n.a. | n.a. | 99.56 | d__Bacteria;p__Firmicutes;c__Clostridia;o__Oscillospirales;f__Acutalibacteraceae;g__UBA1417 | 91.8 | blast;94.10 iPHoP-RF;57.30 |
| S12-wms-combined_264845 | Shared | n.a. | n.a. | n.a. | n.a. | n.a. | n.a. | n.a. | n.a. | 6.58 | d__Bacteria;p__Firmicutes;c__Clostridia;o__Oscillospirales;f__Oscillospiraceae;g__CAG-110 | 95 | blast;96.50 iPHoP-RF;90.10 |
| S12-wms-combined_282594 | Shared | Duplodnaviria | Heunggongvirae | Uroviricota | Caudoviricetes | n.a. | Unclassified | O | O | 18.25 | d__Bacteria;p__Verrucomicrobiota;c__Verrucomicrobiae;o__Verrucomicrobiales;f__Akkermansiaceae;g__Akkermansia | 100 | CRISPR;98.70 RaFAH;98.50 blast;96.90 iPHoP-RF;78.50 |
| S12-wms-combined_299293 | Shared | n.a. | n.a. | n.a. | n.a. | n.a. | n.a. | n.a. | n.a. | 62.93 | d__Bacteria;p__Firmicutes;c__Clostridia;o__Christensenellales;f__CAG-74;g__SFFH01 | 90.4 | blast;92.90 iPHoP-RF;75.40 |
| S12-wms-combined_320727 | Shared | Duplodnaviria | Heunggongvirae | Uroviricota | Caudoviricetes | n.a. | Unclassified | O | O | 99.96 | d__Bacteria;p__Bacteroidota;c__Bacteroidia;o__Bacteroidales;f__Bacteroidaceae;g__Bacteroides | 93.8 | RaFAH;93.60 blast;61.60 iPHoP-RF;55.70 |
| S12-wms-combined_329242 | Shared | n.a. | n.a. | n.a. | n.a. | n.a. | n.a. | n.a. | n.a. | 5.39 | d__Bacteria;p__Firmicutes;c__Clostridia;o__Oscillospirales;f__Oscillospiraceae;g__Dysosmobacter | 91.1 | blast;93.50 iPHoP-RF;56.00 |
| S12-wms-combined_334806 | Shared | n.a. | n.a. | n.a. | n.a. | n.a. | n.a. | n.a. | n.a. | 38.66 | d__Bacteria;p__Firmicutes;c__Clostridia;o__Oscillospirales;f__Oscillospiraceae;g__CAG-83 | 91.6 | blast;93.90 iPHoP-RF;61.90 |
| S11-wms-combined_179605 | Shared | n.a. | n.a. | n.a. | n.a. | n.a. | n.a. | n.a. | n.a. | 37.98 | d__Bacteria;p__Firmicutes;c__Clostridia;o__Lachnospirales;f__Lachnospiraceae;g__UBA9502 | 91.2 | blast;93.60 iPHoP-RF;64.00 |
| S12-wms-combined_345948 | SHHC | Duplodnaviria | Heunggongvirae | Uroviricota | Caudoviricetes | n.a. | Unclassified | Unclassified | Lughvirus | 59.64 | d__Bacteria;p__Firmicutes;c__Clostridia;o__Oscillospirales;f__Ruminococcaceae;g__Faecalibacterium | 98.8 | blast;96.90 iPHoP-RF;92.10 RaFAH;84.90 CRISPR;63.30 |
| S12-wms-combined_351847 | Shared | n.a. | n.a. | n.a. | n.a. | n.a. | n.a. | n.a. | n.a. | 11.93 | d__Bacteria;p__Bacteroidota;c__Bacteroidia;o__Bacteroidales;f__Barnesiellaceae;g__Barnesiella | 95 | blast;96.50 iPHoP-RF;88.20 CRISPR;72.90 |
| S12-wms-combined_375343 | Shared | n.a. | n.a. | n.a. | n.a. | n.a. | n.a. | n.a. | n.a. | 20.72 | d__Bacteria;p__Bacteroidota;c__Bacteroidia;o__Bacteroidales;f__UBA11471;g__UBA11471 | 91.6 | blast;93.90 iPHoP-RF;88.50 |
| S12-wms-combined_129799 | Shared | Duplodnaviria | Heunggongvirae | Uroviricota | Caudoviricetes | n.a. | Unclassified | Unclassified | O | 52.43 | d__Bacteria;p__Firmicutes;c__Clostridia;o__Oscillospirales;f__Oscillospiraceae;g__ER4 | 95.6 | blast;96.90 iPHoP-RF;63.00 |
| S12-wms-combined_393703 | Shared | n.a. | n.a. | n.a. | n.a. | n.a. | n.a. | n.a. | n.a. | 8.02 | d__Bacteria;p__Firmicutes;c__Clostridia;o__Oscillospirales;f__Oscillospiraceae;g__ER4 | 93.5 | blast;95.40 iPHoP-RF;50.20 |
| S12-wms-combined_151843 | Shared | Duplodnaviria | Heunggongvirae | Uroviricota | Caudoviricetes | n.a. | Unclassified | O | O | 24.49 | d__Bacteria;p__Firmicutes;c__Clostridia;o__Christensenellales;f__CAG-74;g__UBA11524 | 95.6 | blast;96.90 iPHoP-RF;92.80 |
| S12-wms-combined_157167 | SHHC | n.a. | n.a. | n.a. | n.a. | n.a. | n.a. | n.a. | n.a. | 8.42 | d__Bacteria;p__Firmicutes;c__Clostridia;o__Oscillospirales;f__Oscillospiraceae;g__CAG-83 | 92.5 | blast;94.60 iPHoP-RF;70.90 |
| S12-wms-combined_164656 | SHHC | n.a. | n.a. | n.a. | n.a. | n.a. | n.a. | n.a. | n.a. | 3.93 | d__Bacteria;p__Firmicutes;c__Clostridia;o__Christensenellales;f__CAG-917;g__CAG-475 | 94.7 | blast;96.30 iPHoP-RF;68.60 |
| S12-wms-combined_71235 | SHHC | Duplodnaviria | Heunggongvirae | Uroviricota | Caudoviricetes | n.a. | Unclassified | Unclassified | Lughvirus | 58.55 | d__Bacteria;p__Firmicutes;c__Clostridia;o__Oscillospirales;f__Ruminococcaceae;g__Faecalibacterium | 95.7 | blast;96.80 iPHoP-RF;84.20 RaFAH;53.60 |
| S12-wms-combined_91231 | SHHC | n.a. | n.a. | n.a. | n.a. | n.a. | n.a. | n.a. | n.a. | 99.33 | d__Bacteria;p__Bacteroidota;c__Bacteroidia;o__Bacteroidales;f__Bacteroidaceae;g__Bacteroides | 100 | RaFAH;99.80 iPHoP-RF;96.10 blast;94.00 |
| S11-wms-combined_226165 | Shared | Duplodnaviria | Heunggongvirae | Uroviricota | Caudoviricetes | n.a. | Unclassified | O | O | 80.02 | d__Bacteria;p__Bacteroidota;c__Bacteroidia;o__Bacteroidales;f__Rikenellaceae;g__Alistipes | 98.7 | iPHoP-RF;97.10 blast;93.80 RaFAH;82.70 |
| S12-wms-combined_190114 | Shared | Duplodnaviria | Heunggongvirae | Uroviricota | Caudoviricetes | n.a. | Unclassified | O | O | 93.65 | d__Bacteria;p__Bacteroidota;c__Bacteroidia;o__Bacteroidales;f__Bacteroidaceae;g__Bacteroides | 100 | RaFAH;99.60 blast;96.70 iPHoP-RF;95.10 |
| S12-wms_12658 | Shared | Duplodnaviria | Heunggongvirae | Uroviricota | Caudoviricetes | n.a. | Unclassified | O | O | 51.35 | d__Bacteria;p__Bacteroidota;c__Bacteroidia;o__Bacteroidales;f__Bacteroidaceae;g__Prevotellamassilia | 97.9 | CRISPR;98.50 blast;60.80 |
| S12-wms-combined_201125 | Shared | Duplodnaviria | Heunggongvirae | Uroviricota | Caudoviricetes | n.a. | Unclassified | O | O | 83.81 | d__Bacteria;p__Bacteroidota;c__Bacteroidia;o__Bacteroidales;f__Bacteroidaceae;g__Bacteroides | 99.4 | iPHoP-RF;96.40 RaFAH;93.60 blast;77.30 |
| S12-wms-combined_223043 | SHHC | Duplodnaviria | Heunggongvirae | Uroviricota | Caudoviricetes | n.a. | Unclassified | O | O | 99.11 | d__Bacteria;p__Firmicutes;c__Clostridia;o__Lachnospirales;f__Lachnospiraceae;g__Enterocloster | 91.5 | blast;93.80 iPHoP-RF;64.30 |
| S12-wms_193565 | SHHC | Duplodnaviria | Heunggongvirae | Uroviricota | Caudoviricetes | n.a. | Unclassified | Unclassified | Mushuvirus | 96.64 | d__Bacteria;p__Firmicutes;c__Clostridia;o__Oscillospirales;f__Ruminococcaceae;g__Faecalibacterium | 90.2 | RaFAH;92.70 |
| S12-wms-combined_23517 | Shared | Duplodnaviria | Heunggongvirae | Uroviricota | Caudoviricetes | n.a. | Unclassified | O | O | 28.19 | d__Bacteria;p__Firmicutes;c__Clostridia;o__Oscillospirales;f__Oscillospiraceae;g__CAG-83 | 98.5 | blast;98.90 CRISPR;98.40 iPHoP-RF;93.10 |
| S12-wms_373775 | Shared | n.a. | n.a. | n.a. | n.a. | n.a. | n.a. | n.a. | n.a. | 11.82 | d__Bacteria;p__Firmicutes;c__Clostridia;o__Oscillospirales;f__Oscillospiraceae;g__CAG-83 | 91.5 | blast;93.80 iPHoP-RF;82.80 |
| S12-wms_386692 | Shared | n.a. | n.a. | n.a. | n.a. | n.a. | n.a. | n.a. | n.a. | 94.2 | d__Bacteria;p__Bacteroidota;c__Bacteroidia;o__Bacteroidales;f__Rikenellaceae;g__Alistipes | 96.2 | CRISPR;97.30 iPHoP-RF;97.10 blast;96.60 |
| S12-wms_401372 | SHHC | Duplodnaviria | Heunggongvirae | Uroviricota | Caudoviricetes | n.a. | Unclassified | Unclassified | Unclassified | 65.9 | d__Bacteria;p__Patescibacteria;c__Paceibacteria;o__Paceibacterales;f__UBA5633;g__MWCK01 | 91.1 | blast;93.50 |
| S12-wms-combined_264077 | SHHC | Duplodnaviria | Heunggongvirae | Uroviricota | Caudoviricetes | n.a. | Unclassified | O | O | 19.02 | d__Bacteria;p__Firmicutes;c__Clostridia;o__Oscillospirales;f__Ruminococcaceae;g__Faecalibacterium | 90.4 | blast;92.90 iPHoP-RF;86.30 |
| S17-wms-combined_136622 | ME | n.a. | n.a. | n.a. | n.a. | n.a. | n.a. | n.a. | n.a. | 24.07 | d__Bacteria;p__Firmicutes;c__Clostridia;o__Oscillospirales;f__Oscillospiraceae;g__Evtepia | 90.7 | blast;93.10 iPHoP-RF;81.60 |
| S11-wms-combined_321938 | Shared | Duplodnaviria | Heunggongvirae | Uroviricota | Caudoviricetes | n.a. | Unclassified | O | O | 99.59 | d__Bacteria;p__Bacteroidota;c__Bacteroidia;o__Bacteroidales;f__Bacteroidaceae;g__Bacteroides | 100 | RaFAH;98.70 iPHoP-RF;94.10 blast;80.70 |
| S17-wms-combined_146850 | ME | n.a. | n.a. | n.a. | n.a. | n.a. | n.a. | n.a. | n.a. | 3.97 | d__Bacteria;p__Firmicutes;c__Clostridia;o__Oscillospirales;f__Oscillospiraceae;g__Flavonifractor | 91.2 | blast;93.60 iPHoP-RF;74.30 |
| S17-wms-combined_157468 | ME | n.a. | n.a. | n.a. | n.a. | n.a. | n.a. | n.a. | n.a. | 3.55 | d__Bacteria;p__Firmicutes;c__Clostridia;o__Oscillospirales;f__Oscillospiraceae;g__Flavonifractor | 93.4 | blast;95.30 iPHoP-RF;74.80 |
| S12-wms-combined_293354 | Shared | Duplodnaviria | Heunggongvirae | Uroviricota | Caudoviricetes | n.a. | Unclassified | O | O | 10.3 | d__Bacteria;p__Firmicutes;c__Clostridia;o__Christensenellales;f__CAG-74;g__UBA11524 | 95.6 | blast;96.90 iPHoP-RF;92.80 |
| S17-wms-combined_158354 | ME | n.a. | n.a. | n.a. | n.a. | n.a. | n.a. | n.a. | n.a. | 99.65 | d__Bacteria;p__Bacteroidota;c__Bacteroidia;o__Bacteroidales;f__Bacteroidaceae;g__Bacteroides | 100 | RaFAH;99.30 iPHoP-RF;97.10 blast;92.60 |
| S17-wms-combined_162786 | ME | n.a. | n.a. | n.a. | n.a. | n.a. | n.a. | n.a. | n.a. | 7.07 | d__Bacteria;p__Firmicutes;c__Clostridia;o__Oscillospirales;f__Oscillospiraceae;g__Marseille-P3106 | 94.7 | blast;96.30 iPHoP-RF;71.30 |
| S17-wms-combined_16309 | ME | Duplodnaviria | Heunggongvirae | Uroviricota | Caudoviricetes | n.a. | Unclassified | O | O | 27.66 | d__Bacteria;p__Bacteroidota;c__Bacteroidia;o__Bacteroidales;f__Bacteroidaceae;g__Bacteroides | 100 | RaFAH;99.10 iPHoP-RF;97.10 blast;90.90 |
| S12-wms-combined_302876 | SHHC | Duplodnaviria | Heunggongvirae | Uroviricota | Caudoviricetes | n.a. | Unclassified | O | O | 24.68 | d__Bacteria;p__Firmicutes;c__Clostridia;o__Lachnospirales;f__Lachnospiraceae;g__Lachnospira | 93.9 | blast;95.70 iPHoP-RF;73.50 |
| S17-wms-combined_175924 | ME | Duplodnaviria | Heunggongvirae | Uroviricota | Caudoviricetes | n.a. | Unclassified | Unclassified | Mushuvirus | 99.75 | d__Bacteria;p__Firmicutes;c__Clostridia;o__Oscillospirales;f__Oscillospiraceae;g__Flavonifractor | 92.1 | blast;94.30 iPHoP-RF;78.00 |
| S12-wms-combined_313953 | SHHC | Duplodnaviria | Heunggongvirae | Uroviricota | Caudoviricetes | n.a. | Unclassified | O | O | 97.5 | d__Bacteria;p__Proteobacteria;c__Alphaproteobacteria;o__RF32;f__CAG-239;g__CAG-495 | 90.9 | blast;93.30 iPHoP-RF;78.50 |
| S11-wms-combined_64392 | Shared | Duplodnaviria | Heunggongvirae | Uroviricota | Caudoviricetes | n.a. | Unclassified | O | O | 100 | d__Bacteria;p__Bacteroidota;c__Bacteroidia;o__Bacteroidales;f__Rikenellaceae;g__Alistipes | 99.6 | iPHoP-RF;97.10 blast;96.80 RaFAH;92.70 |
| S17-wms-combined_179771 | Shared | n.a. | n.a. | n.a. | n.a. | n.a. | n.a. | n.a. | n.a. | 49.28 | d__Bacteria;p__Bacteroidota;c__Bacteroidia;o__Bacteroidales;f__Bacteroidaceae;g__Bacteroides | 97.9 | blast;98.30 iPHoP-RF;96.10 CRISPR;90.30 RaFAH;56.00 |
| S17-wms-combined_193958 | Shared | Duplodnaviria | Heunggongvirae | Uroviricota | Caudoviricetes | n.a. | Unclassified | Unclassified | Mushuvirus | 96.31 | d__Bacteria;p__Firmicutes;c__Clostridia;o__Oscillospirales;f__Ruminococcaceae;g__Faecalibacterium | 94.2 | blast;95.90 iPHoP-RF;53.80 |
| S17-wms-combined_199746 | Shared | Duplodnaviria | Heunggongvirae | Uroviricota | Caudoviricetes | n.a. | Unclassified | O | O | 69.95 | d__Bacteria;p__Bacteroidota;c__Bacteroidia;o__Bacteroidales;f__Rikenellaceae;g__Alistipes | 97.6 | CRISPR;98.30 iPHoP-RF;97.10 blast;96.90 |
| S12-wms-combined_323943 | SHHC | Duplodnaviria | Heunggongvirae | Uroviricota | Caudoviricetes | n.a. | Unclassified | Unclassified | O | 18.94 | d__Bacteria;p__Firmicutes;c__Clostridia;o__Oscillospirales;f__Ruminococcaceae;g__Gemmiger | 94.6 | CRISPR;96.20 blast;93.90 iPHoP-RF;91.80 |
| S12-wms-combined_328554 | Shared | Duplodnaviria | Heunggongvirae | Uroviricota | Caudoviricetes | n.a. | Unclassified | O | O | 31.03 | d__Bacteria;p__Bacteroidota;c__Bacteroidia;o__Bacteroidales;f__Rikenellaceae;g__Alistipes | 96.3 | iPHoP-RF;96.70 blast;95.80 RaFAH;60.10 |
| S17-wms-combined_223748 | ME | n.a. | n.a. | n.a. | n.a. | n.a. | n.a. | n.a. | n.a. | 4.98 | d__Bacteria;p__Firmicutes;c__Clostridia;o__Lachnospirales;f__Lachnospiraceae;g__Enterocloster | 91.2 | blast;93.60 iPHoP-RF;75.20 |
| S17-wms-combined_225356 | ME | n.a. | n.a. | n.a. | n.a. | n.a. | n.a. | n.a. | n.a. | 10.23 | d__Bacteria;p__Actinobacteriota;c__Coriobacteriia;o__Coriobacteriales;f__Eggerthellaceae;g__Eggerthella | 96.3 | CRISPR;97.40 blast;92.10 iPHoP-RF;87.00 |
| S17-wms-combined_233761 | Shared | n.a. | n.a. | n.a. | n.a. | n.a. | n.a. | n.a. | n.a. | 88 | d__Bacteria;p__Bacteroidota;c__Bacteroidia;o__Bacteroidales;f__Bacteroidaceae;g__Bacteroides | 100 | RaFAH;99.00 blast;98.60 iPHoP-RF;96.40 CRISPR;80.00 |
| S17-wms-combined_247494 | Shared | Duplodnaviria | Heunggongvirae | Uroviricota | Caudoviricetes | n.a. | Unclassified | O | O | 24.14 | d__Bacteria;p__Firmicutes;c__Clostridia;o__Oscillospirales;f__Oscillospiraceae;g__Flavonifractor | 91.5 | blast;93.80 iPHoP-RF;91.40 |
| S17-wms-combined_259811 | Shared | Duplodnaviria | Heunggongvirae | Uroviricota | Caudoviricetes | n.a. | Unclassified | O | O | 66.5 | d__Bacteria;p__Firmicutes;c__Clostridia;o__Oscillospirales;f__Oscillospiraceae;g__Lawsonibacter | 90.9 | blast;93.30 |
| S11-wms_32691 | ME | n.a. | n.a. | n.a. | n.a. | n.a. | n.a. | n.a. | n.a. | 12 | d__Bacteria;p__Firmicutes;c__Clostridia;o__Oscillospirales;f__Oscillospiraceae;g__Pseudoflavonifractor | 94.9 | blast;96.40 iPHoP-RF;66.80 |
| S12-wms-combined_358797 | Shared | Duplodnaviria | Heunggongvirae | Uroviricota | Caudoviricetes | n.a. | Unclassified | O | O | 99.42 | d__Bacteria;p__Bacteroidota;c__Bacteroidia;o__Bacteroidales;f__Rikenellaceae;g__Alistipes | 100 | blast;98.40 iPHoP-RF;96.10 RaFAH;95.40 |
| S17-wms-combined_260343 | ME | Duplodnaviria | Heunggongvirae | Uroviricota | Caudoviricetes | n.a. | Unclassified | Unclassified | Toutatisvirus | 38.59 | d__Bacteria;p__Firmicutes;c__Clostridia;o__Oscillospirales;f__Oscillospiraceae;g__Flavonifractor | 90.8 | blast;93.20 |
| S12-wms-combined_368562 | Shared | Duplodnaviria | Heunggongvirae | Uroviricota | Caudoviricetes | n.a. | Unclassified | O | O | 98.38 | d__Bacteria;p__Bacteroidota;c__Bacteroidia;o__Bacteroidales;f__Bacteroidaceae;g__Bacteroides | 100 | RaFAH;99.70 blast;93.90 iPHoP-RF;92.80 |
| S12-wms-combined_371541 | Shared | n.a. | n.a. | n.a. | n.a. | n.a. | n.a. | n.a. | n.a. | 14.41 | d__Bacteria;p__Bacteroidota;c__Bacteroidia;o__Bacteroidales;f__UBA11471;g__UBA11471 | 91.6 | blast;93.90 iPHoP-RF;90.10 |
| S17-wms-combined_70983 | Shared | n.a. | n.a. | n.a. | n.a. | n.a. | n.a. | n.a. | n.a. | 7.24 | d__Bacteria;p__Bacteroidota;c__Bacteroidia;o__Bacteroidales;f__Bacteroidaceae;g__Bacteroides | 95.9 | iPHoP-RF;97.10 blast;85.40 |
| S17-wms-combined_79709 | ME | Duplodnaviria | Heunggongvirae | Uroviricota | Caudoviricetes | n.a. | Unclassified | O | O | 19.94 | d__Bacteria;p__Firmicutes;c__Clostridia;o__Oscillospirales;f__Oscillospiraceae;g__An92 | 91.4 | blast;93.70 iPHoP-RF;92.10 |
| S17-wms-combined_90378 | ME | Duplodnaviria | Heunggongvirae | Uroviricota | Caudoviricetes | n.a. | Unclassified | Unclassified | Lughvirus | 72.41 | d__Bacteria;p__Firmicutes;c__Clostridia;o__Oscillospirales;f__Ruminococcaceae;g__Faecalibacterium | 98.7 | blast;96.90 RaFAH;83.30 iPHoP-RF;76.80 CRISPR;69.70 |
| S12-wms-combined_117447 | SHHC | n.a. | n.a. | n.a. | n.a. | n.a. | n.a. | n.a. | n.a. | | d__Bacteria;p__Firmicutes;c__Clostridia;o__Christensenellales;f__CAG-917;g__ | 91 | blast;93.40 iPHoP-RF;67.30 |
| S12-wms-combined_48648 | Shared | n.a. | n.a. | n.a. | n.a. | n.a. | n.a. | n.a. | n.a. | 6.97 | d__Bacteria;p__Firmicutes;c__Clostridia;o__Oscillospirales;f__Oscillospiraceae;g__ER4 | 90.9 | blast;93.30 iPHoP-RF;72.40 |
| S12-wms-combined_117628 | Shared | Duplodnaviria | Heunggongvirae | Uroviricota | Caudoviricetes | n.a. | Unclassified | Unclassified | O | 32.91 | d__Bacteria;p__Firmicutes;c__Clostridia;o__Oscillospirales;f__Acutalibacteraceae;g__CAG-217 | 96.2 | blast;97.30 iPHoP-RF;73.20 |
| S17-wms_124308 | ME | Duplodnaviria | Heunggongvirae | Uroviricota | Caudoviricetes | n.a. | Unclassified | O | O | 51.69 | d__Bacteria;p__Bacteroidota;c__Bacteroidia;o__Bacteroidales;f__Bacteroidaceae;g__Bacteroides | 99.1 | iPHoP-RF;96.40 RaFAH;90.70 |
| S12-wms-combined_63649 | Shared | n.a. | n.a. | n.a. | n.a. | n.a. | n.a. | n.a. | n.a. | 5.54 | d__Bacteria;p__Firmicutes;c__Clostridia;o__Oscillospirales;f__Oscillospiraceae;g__CAG-83 | 91.7 | blast;94.00 iPHoP-RF;90.10 |
| S17-wms_254872 | ME | Monodnaviria | Sangervirae | Phixviricota | Malgrandaviricetes | Petitvirales | Microviridae | Unclassified | Microvirus | 5.1 | d__Bacteria;p__Bacteroidota;c__Bacteroidia;o__Bacteroidales;f__Bacteroidaceae;g__Bacteroides_G | 95.3 | iPHoP-RF;96.70 |
| S2-wms-combined_101826 | Shared | Duplodnaviria | Heunggongvirae | Uroviricota | Caudoviricetes | n.a. | Unclassified | Unclassified | Unclassified | 26.15 | d__Bacteria;p__Firmicutes;c__Clostridia;o__Oscillospirales;f__Ruminococcaceae;g__Ruminococcus_D | 98.8 | CRISPR;99.10 blast;96.90 iPHoP-RF;92.80 |
| S2-wms-combined_113619 | Shared | Duplodnaviria | Heunggongvirae | Uroviricota | Caudoviricetes | n.a. | Unclassified | O | O | 97.74 | d__Bacteria;p__Bacteroidota;c__Bacteroidia;o__Bacteroidales;f__Rikenellaceae;g__Alistipes | 99 | blast;96.60 RaFAH;87.50 CRISPR;82.60 iPHoP-RF;77.80 |
| S2-wms-combined_126134 | Shared | n.a. | n.a. | n.a. | n.a. | n.a. | n.a. | n.a. | n.a. | 14.26 | d__Bacteria;p__Firmicutes;c__Clostridia;o__Oscillospirales;f__Oscillospiraceae;g__Lawsonibacter | 90.1 | blast;92.60 iPHoP-RF;75.70 |
| S2-wms-combined_132966 | SHHC | Monodnaviria | Loebvirae | Hofneiviricota | Faserviricetes | Tubulavirales | Inoviridae | Unclassified | Inovirus | | d__Bacteria;p__Bacteroidota;c__Bacteroidia;o__Bacteroidales;f__Bacteroidaceae;g__Bacteroides | 93.1 | iPHoP-RF;95.10 |
| S12-wms-combined_148741 | Shared | n.a. | n.a. | n.a. | n.a. | n.a. | n.a. | n.a. | n.a. | 20.82 | d__Bacteria;p__Firmicutes;c__Clostridia;o__Oscillospirales;f__Oscillospiraceae;g__CAG-83 | 93.9 | blast;95.70 iPHoP-RF;94.70 CRISPR;93.00 |
| S2-wms-combined_135139 | Shared | n.a. | n.a. | n.a. | n.a. | n.a. | n.a. | n.a. | n.a. | 38.63 | d__Bacteria;p__Verrucomicrobiota;c__Verrucomicrobiae;o__Verrucomicrobiales;f__Akkermansiaceae;g__Akkermansia | 97.8 | CRISPR;98.40 blast;96.90 iPHoP-RF;88.20 |
| S2-wms-combined_135154 | Shared | n.a. | n.a. | n.a. | n.a. | n.a. | n.a. | n.a. | n.a. | 34.29 | d__Bacteria;p__Firmicutes;c__Negativicutes;o__Veillonellales;f__Dialisteraceae;g__Dialister | 95.9 | blast;97.10 iPHoP-RF;97.10 CRISPR;75.50 |
| S12-wms_235125 | SHHC | n.a. | n.a. | n.a. | n.a. | n.a. | n.a. | n.a. | n.a. | 20.48 | d__Bacteria;p__Firmicutes;c__Clostridia;o__Oscillospirales;f__Oscillospiraceae;g__ER4 | 90.8 | blast;93.20 iPHoP-RF;64.80 |
| S2-wms-combined_165833 | SHHC | Duplodnaviria | Heunggongvirae | Uroviricota | Caudoviricetes | n.a. | Unclassified | Unclassified | Mushuvirus | 74.42 | d__Bacteria;p__Firmicutes;c__Clostridia;o__Lachnospirales;f__Lachnospiraceae;g__Roseburia | 97.9 | blast;96.20 iPHoP-RF;92.80 RaFAH;79.80 |
| S2-wms-combined_166665 | Shared | Duplodnaviria | Heunggongvirae | Uroviricota | Caudoviricetes | n.a. | Unclassified | O | O | 48.97 | d__Bacteria;p__Firmicutes;c__Clostridia;o__Oscillospirales;f__Ruminococcaceae;g__Faecalibacterium | 95.4 | blast;96.80 iPHoP-RF;96.10 |
| S2-wms-combined_176376 | SHHC | Duplodnaviria | Heunggongvirae | Uroviricota | Caudoviricetes | n.a. | Unclassified | O | O | 94.42 | d__Bacteria;p__Firmicutes;c__Clostridia;o__Lachnospirales;f__Lachnospiraceae;g__Blautia_A | 96.7 | blast;93.20 CRISPR;89.80 iPHoP-RF;87.90 RaFAH;83.30 |
| S17-wms-combined_106940 | Shared | Duplodnaviria | Heunggongvirae | Uroviricota | Caudoviricetes | n.a. | Unclassified | O | O | 77.34 | d__Bacteria;p__Bacteroidota;c__Bacteroidia;o__Bacteroidales;f__Bacteroidaceae;g__Bacteroides | 99.6 | iPHoP-RF;97.10 blast;96.90 RaFAH;92.40 |
| S2-wms-combined_196783 | Shared | Duplodnaviria | Heunggongvirae | Uroviricota | Caudoviricetes | n.a. | Unclassified | O | O | 32.37 | d__Bacteria;p__Firmicutes;c__Clostridia;o__Oscillospirales;f__Oscillospiraceae;g__Intestinimonas | 95.9 | CRISPR;97.10 blast;91.40 iPHoP-RF;85.10 |
| S12-wms-combined_184338 | Shared | Duplodnaviria | Heunggongvirae | Uroviricota | Caudoviricetes | n.a. | Unclassified | O | O | 20.69 | d__Bacteria;p__Firmicutes;c__Clostridia;o__Oscillospirales;f__Oscillospiraceae;g__Dysosmobacter | 91.6 | blast;93.90 iPHoP-RF;83.90 |
| S17-wms-combined_146050 | Shared | n.a. | n.a. | n.a. | n.a. | n.a. | n.a. | n.a. | n.a. | 6.87 | d__Bacteria;p__Bacteroidota;c__Bacteroidia;o__Bacteroidales;f__Bacteroidaceae;g__Bacteroides | 100 | RaFAH;98.70 blast;96.80 iPHoP-RF;92.80 |
| S12-wms-combined_189323 | Shared | Duplodnaviria | Heunggongvirae | Uroviricota | Caudoviricetes | n.a. | Unclassified | O | O | 72.16 | d__Bacteria;p__Bacteroidota;c__Bacteroidia;o__Bacteroidales;f__Rikenellaceae;g__Alistipes | 99.4 | blast;97.60 iPHoP-RF;96.70 RaFAH;89.00 CRISPR;53.90 |
| S2-wms-combined_197577 | SHHC | Duplodnaviria | Heunggongvirae | Uroviricota | Caudoviricetes | n.a. | Unclassified | Unclassified | Unclassified | 42.77 | d__Bacteria;p__Firmicutes;c__Negativicutes;o__Selenomonadales;f__Selenomonadaceae;g__Megamonas | 91.1 | blast;93.50 iPHoP-RF;53.30 |
| S2-wms-combined_210822 | Shared | n.a. | n.a. | n.a. | n.a. | n.a. | n.a. | n.a. | n.a. | 37.52 | d__Bacteria;p__Bacteroidota;c__Bacteroidia;o__Bacteroidales;f__Bacteroidaceae;g__Bacteroides | 91.8 | iPHoP-RF;89.50 blast;81.20 RaFAH;71.80 |
| S2-wms-combined_218002 | Shared | Duplodnaviria | Heunggongvirae | Uroviricota | Caudoviricetes | n.a. | Unclassified | O | O | 99.73 | d__Bacteria;p__Firmicutes;c__Clostridia;o__Lachnospirales;f__Lachnospiraceae;g__Roseburia | 99.9 | blast;98.50 iPHoP-RF;92.40 RaFAH;92.30 |
| S2-wms-combined_229529 | Shared | Duplodnaviria | Heunggongvirae | Uroviricota | Caudoviricetes | n.a. | Unclassified | O | O | 100 | d__Bacteria;p__Bacteroidota;c__Bacteroidia;o__Bacteroidales;f__Bacteroidaceae;g__Bacteroides | 99.9 | blast;96.90 iPHoP-RF;96.70 RaFAH;96.10 |
| S17-wms-combined_171305 | ME | Duplodnaviria | Heunggongvirae | Uroviricota | Caudoviricetes | n.a. | Unclassified | O | O | 56.56 | d__Bacteria;p__Bacteroidota;c__Bacteroidia;o__Bacteroidales;f__Tannerellaceae;g__Parabacteroides | 98.7 | CRISPR;99.00 blast;98.50 iPHoP-RF;96.40 |
| S2-wms-combined_238929 | Shared | Duplodnaviria | Heunggongvirae | Uroviricota | Caudoviricetes | n.a. | Unclassified | O | O | 17.21 | d__Bacteria;p__Firmicutes;c__Clostridia;o__Oscillospirales;f__Oscillospiraceae;g__CAG-170 | 91.4 | blast;93.70 iPHoP-RF;78.30 |
| S17-wms-combined_185239 | Shared | n.a. | n.a. | n.a. | n.a. | n.a. | n.a. | n.a. | n.a. | 7.9 | d__Bacteria;p__Firmicutes;c__Clostridia;o__Oscillospirales;f__Oscillospiraceae;g__Flavonifractor | 91.4 | blast;93.70 iPHoP-RF;83.60 |

| | | | | | | | | | | | | | |
|---|---|---|---|---|---|---|---|---|---|---|---|---|---|
| S17-wms-combined_186314 | ME | Duplodnaviria | Heunggongvirae | Uroviricota | Caudoviricetes | n.a. | Unclassified | O | O | 29.67 | d__Bacteria;p__Actinobacteriota;c__Actinomycetia;o__Actinomycetales;f__Bifidobacteriaceae;g__Bifidobacterium | 99.6 | CRISPR;99.60 iPHoP-RF;97.10 blast;96.90 |
| S2-wms-combined_31084 | SHHC | Duplodnaviria | Heunggongvirae | Uroviricota | Caudoviricetes | n.a. | Unclassified | O | O | 22.49 | d__Bacteria;p__Firmicutes;c__Clostridia;o__Lachnospirales;f__Lachnospiraceae;g__Acetivibrio_A | 90 | blast;92.50 iPHoP-RF;69.10 |
| S2-wms-combined_58121 | Shared | n.a. | n.a. | n.a. | n.a. | n.a. | n.a. | n.a. | n.a. | 21.08 | d__Bacteria;p__Bacteroidota;c__Bacteroidia;o__Bacteroidales;f__Bacteroidaceae;g__Bacteroides | 95.9 | iPHoP-RF;97.10 blast;91.00 |
| S2-wms-combined_6605 | Shared | n.a. | n.a. | n.a. | n.a. | n.a. | n.a. | n.a. | n.a. | 7.53 | d__Bacteria;p__Firmicutes;c__Clostridia;o__Oscillospirales;f__Oscillospiraceae;g__Flavonifractor | 91.4 | blast;93.70 iPHoP-RF;82.50 |
| S12-wms-combined_25459 | Shared | n.a. | n.a. | n.a. | n.a. | n.a. | n.a. | n.a. | n.a. | 9.92 | d__Bacteria;p__Bacteroidota;c__Bacteroidia;o__Bacteroidales;f__Rikenellaceae;g__Alistipes | 95.6 | blast;96.90 iPHoP-RF;96.70 |
| S2-wms-combined_85381 | SHHC | Duplodnaviria | Heunggongvirae | Uroviricota | Caudoviricetes | n.a. | Unclassified | Unclassified | Eponavirus | 45.07 | d__Bacteria;p__Firmicutes;c__Clostridia;o__Lachnospirales;f__Lachnospiraceae;g__CAG-81 | 93.8 | blast;95.60 iPHoP-RF;80.60 |
| S2-wms-combined_93845 | Shared | Duplodnaviria | Heunggongvirae | Uroviricota | Caudoviricetes | n.a. | Unclassified | O | O | 99.44 | d__Bacteria;p__Firmicutes;c__Clostridia;o__Oscillospirales;f__Oscillospiraceae;g__ER4 | 98.7 | CRISPR;99.00 blast;91.30 iPHoP-RF;81.90 |
| S2-wms_128831 | SHHC | Duplodnaviria | Heunggongvirae | Uroviricota | Caudoviricetes | n.a. | Unclassified | Unclassified | Unclassified | 98.35 | d__Bacteria;p__Proteobacteria;c__Gammaproteobacteria;o__Enterobacterales;f__Enterobacteriaceae;g__Escherichia | 95.6 | RaFAH;96.90 |
| S2-wms_128962 | Shared | Duplodnaviria | Heunggongvirae | Uroviricota | Caudoviricetes | Crassvirales | Suoliviridae | O | O | 46.73 | d__Bacteria;p__Bacteroidota;c__Bacteroidia;o__Bacteroidales;f__Bacteroidaceae;g__Prevotella | 91.8 | RaFAH;94.10 |
| S2-wms_130687 | Shared | Duplodnaviria | Heunggongvirae | Uroviricota | Caudoviricetes | n.a. | Unclassified | Unclassified | Unclassified | 95.89 | d__Bacteria;p__Firmicutes;c__Clostridia;o__Oscillospirales;f__Acutalibacteraceae;g__Ruminococcus_E | 99.7 | CRISPR;98.90 blast;96.90 iPHoP-RF;92.40 RaFAH;85.50 |
| S17-wms-combined_72837 | ME | Duplodnaviria | Heunggongvirae | Uroviricota | Caudoviricetes | n.a. | Unclassified | O | O | 25.38 | d__Bacteria;p__Firmicutes;c__Clostridia;o__Oscillospirales;f__Ruminococcaceae;g__Gemmiger | 95.2 | blast;96.60 iPHoP-RF;74.80 |
| S2-wms_140933 | Shared | Duplodnaviria | Heunggongvirae | Uroviricota | Caudoviricetes | n.a. | Unclassified | Unclassified | Unclassified | 43.87 | d__Bacteria;p__Firmicutes;c__Clostridia;o__Lachnospirales;f__Lachnospiraceae;g__Blautia_A | 93.8 | blast;90.60 RaFAH;76.60 iPHoP-RF;62.50 |
| S2-wms_156439 | Shared | Duplodnaviria | Heunggongvirae | Uroviricota | Caudoviricetes | n.a. | Unclassified | O | O | 96.43 | d__Bacteria;p__Bacteroidota;c__Bacteroidia;o__Bacteroidales;f__Bacteroidaceae;g__Bacteroides | 100 | RaFAH;99.60 blast;96.80 iPHoP-RF;95.70 |
| S12-wms-combined_270839 | Shared | n.a. | n.a. | n.a. | n.a. | n.a. | n.a. | n.a. | n.a. | 20.16 | d__Bacteria;p__Firmicutes;c__Clostridia;o__Oscillospirales;f__Oscillospiraceae;g__CAG-83 | 92.5 | blast;94.60 iPHoP-RF;79.80 |
| S17-wms-combined_95360 | Shared | Duplodnaviria | Heunggongvirae | Uroviricota | Caudoviricetes | n.a. | Unclassified | O | O | 17.32 | d__Bacteria;p__Firmicutes;c__Negativicutes;o__Veillonellales;f__Dialisteraceae;g__Dialister | 97.2 | CRISPR;98.00 blast;96.90 iPHoP-RF;93.70 |
| S2-wms_79304 | SHHC | Duplodnaviria | Heunggongvirae | Uroviricota | Caudoviricetes | n.a. | Unclassified | Unclassified | Dybvigvirus | 81.02 | d__Bacteria;p__Actinobacteriota;c__Coriobacteriia;o__Coriobacteriales;f__Eggerthellaceae;g__Eggerthella | 94.9 | CRISPR;96.40 RaFAH;50.20 |
| S3-wms-combined_73262 | Shared | Duplodnaviria | Heunggongvirae | Uroviricota | Caudoviricetes | n.a. | Unclassified | Unclassified | Unclassified | 99.96 | d__Bacteria;p__Bacteroidota;c__Bacteroidia;o__Bacteroidales;f__Bacteroidaceae;g__Bacteroides | 100 | RaFAH;99.80 blast;93.60 iPHoP-RF;79.80 CRISPR;67.80 |
| S12-wms-combined_297545 | Shared | Duplodnaviria | Heunggongvirae | Uroviricota | Caudoviricetes | n.a. | Unclassified | O | O | 99.69 | d__Bacteria;p__Bacteroidota;c__Bacteroidia;o__Bacteroidales;f__Bacteroidaceae;g__Bacteroides | 99.9 | RaFAH;98.50 blast;90.90 iPHoP-RF;65.60 |
| S3-wms_22744 | Shared | Duplodnaviria | Heunggongvirae | Uroviricota | Caudoviricetes | n.a. | Unclassified | Unclassified | Lagaffevirus | 99.21 | d__Bacteria;p__Firmicutes;c__Clostridia;o__Oscillospirales;f__Ruminococcaceae;g__Faecalibacterium | 97.2 | CRISPR;98.00 blast;60.70 iPHoP-RF;52.00 |
| S2-wms-combined_116221 | Shared | Duplodnaviria | Heunggongvirae | Uroviricota | Caudoviricetes | n.a. | Unclassified | O | O | 80.02 | d__Bacteria;p__Firmicutes;c__Clostridia;o__Lachnospirales;f__Anaerotignaceae;g__Anaerotignum | 95.6 | blast;96.90 iPHoP-RF;61.10 |
| S3-wms_23958 | Shared | Duplodnaviria | Heunggongvirae | Uroviricota | Caudoviricetes | n.a. | Unclassified | O | O | 21.78 | d__Bacteria;p__Firmicutes;c__Clostridia;o__Oscillospirales;f__Oscillospiraceae;g__CAG-170 | 94.2 | blast;95.90 iPHoP-RF;58.70 |
| S4-wms-combined_113721 | Shared | n.a. | n.a. | n.a. | n.a. | n.a. | n.a. | n.a. | n.a. | 6.49 | d__Bacteria;p__Firmicutes;c__Clostridia;o__Oscillospirales;f__Ruminococcaceae;g__Gemmiger | 95.6 | blast;96.90 iPHoP-RF;92.80 CRISPR;57.80 |
| S2-wms-combined_126440 | SHHC | Duplodnaviria | Heunggongvirae | Uroviricota | Caudoviricetes | n.a. | Unclassified | O | O | 24 | d__Bacteria;p__Firmicutes;c__Clostridia;o__Lachnospirales;f__Lachnospiraceae;g__Acetatifactor | 92.7 | blast;94.80 iPHoP-RF;71.30 |
| S12-wms-combined_31761 | SHHC | Duplodnaviria | Heunggongvirae | Uroviricota | Caudoviricetes | n.a. | Unclassified | Unclassified | O | 54.85 | d__Bacteria;p__Firmicutes;c__Clostridia;o__Lachnospirales;f__Lachnospiraceae;g__COE1 | 93.6 | blast;95.50 iPHoP-RF;70.20 |
| S4-wms-combined_14411 | SHHC | Duplodnaviria | Heunggongvirae | Uroviricota | Caudoviricetes | n.a. | Unclassified | Unclassified | Unclassified | 97.82 | d__Bacteria;p__Bacteroidota;c__Bacteroidia;o__Bacteroidales;f__Tannerellaceae;g__Marseille-P3160 | 98.4 | CRISPR;98.80 iPHoP-RF;68.90 |
| S12-wms-combined_320262 | Shared | n.a. | n.a. | n.a. | n.a. | n.a. | n.a. | n.a. | n.a. | 15.45 | d__Bacteria;p__Bacteroidota;c__Bacteroidia;o__Bacteroidales;f__Rikenellaceae;g__Alistipes | 97.8 | blast;98.40 iPHoP-RF;97.10 |
| S2-wms-combined_127080 | SHHC | n.a. | n.a. | n.a. | n.a. | n.a. | n.a. | n.a. | n.a. | 8.57 | d__Bacteria;p__Firmicutes;c__Clostridia;o__Oscillospirales;f__Ruminococcaceae;g__Faecalibacterium | 98.8 | CRISPR;99.10 blast;93.50 iPHoP-RF;89.50 |
| S4-wms-combined_17284 | Shared | Duplodnaviria | Heunggongvirae | Uroviricota | Caudoviricetes | n.a. | Unclassified | Unclassified | Unclassified | 85.97 | d__Bacteria;p__Actinobacteriota;c__Actinomycetia;o__Actinomycetales;f__Bifidobacteriaceae;g__Bifidobacterium | 95.3 | iPHoP-RF;96.70 CRISPR;96.00 blast;95.40 |
| S2-wms-combined_13246 | Shared | Duplodnaviria | Heunggongvirae | Uroviricota | Caudoviricetes | n.a. | Unclassified | Unclassified | O | 32.69 | d__Bacteria;p__Firmicutes;c__Clostridia;o__Oscillospirales;f__Ruminococcaceae;g__Faecalibacterium | 99.6 | blast;96.90 RaFAH;93.60 iPHoP-RF;92.80 CRISPR;87.50 |
| S4-wms-combined_41490 | Shared | Duplodnaviria | Heunggongvirae | Uroviricota | Caudoviricetes | n.a. | Unclassified | O | O | 99.65 | d__Bacteria;p__Bacteroidota;c__Bacteroidia;o__Bacteroidales;f__Rikenellaceae;g__Alistipes | 95.6 | blast;96.90 iPHoP-RF;96.10 |
| S4-wms-combined_42647 | Shared | Duplodnaviria | Heunggongvirae | Uroviricota | Caudoviricetes | n.a. | Unclassified | Unclassified | Lughvirus | 63.73 | d__Bacteria;p__Firmicutes;c__Clostridia;o__Oscillospirales;f__Ruminococcaceae;g__Faecalibacterium | 95.7 | blast;97.00 iPHoP-RF;78.00 CRISPR;67.90 |
| S4-wms-combined_47565 | SHHC | n.a. | n.a. | n.a. | n.a. | n.a. | n.a. | n.a. | n.a. | 81.36 | d__Bacteria;p__Bacteroidota;c__Bacteroidia;o__Bacteroidales;f__Bacteroidaceae;g__Prevotella | 96.2 | blast;97.30 iPHoP-RF;90.80 |
| S4-wms-combined_51079 | Shared | n.a. | n.a. | n.a. | n.a. | n.a. | n.a. | n.a. | n.a. | 88.31 | d__Bacteria;p__Actinobacteriota;c__Actinomycetia;o__Actinomycetales;f__Bifidobacteriaceae;g__Bifidobacterium | 99.9 | CRISPR;99.60 iPHoP-RF;97.10 blast;96.80 RaFAH;71.10 |
| S4-wms-combined_57123 | SHHC | n.a. | n.a. | n.a. | n.a. | n.a. | n.a. | n.a. | n.a. | 13.17 | d__Bacteria;p__Bacteroidota;c__Bacteroidia;o__Bacteroidales;f__Bacteroidaceae;g__Prevotella | 95.6 | blast;96.90 iPHoP-RF;73.50 |
| S2-wms-combined_137119 | Shared | Duplodnaviria | Heunggongvirae | Uroviricota | Caudoviricetes | n.a. | Unclassified | Unclassified | O | 36.79 | d__Bacteria;p__Firmicutes;c__Clostridia;o__Oscillospirales;f__Oscillospiraceae;g__An92 | 93.5 | blast;95.40 iPHoP-RF;76.60 |
| S4-wms-combined_64632 | SHHC | Duplodnaviria | Heunggongvirae | Uroviricota | Caudoviricetes | n.a. | Unclassified | Unclassified | Unclassified | 43.29 | d__Bacteria;p__Firmicutes;c__Clostridia;o__Lachnospirales;f__Lachnospiraceae;g__Dorea_A | 97.6 | CRISPR;98.30 iPHoP-RF;95.40 blast;92.00 |
| S2-wms-combined_140996 | Shared | Duplodnaviria | Heunggongvirae | Uroviricota | Caudoviricetes | n.a. | Unclassified | Unclassified | O | 59.11 | d__Bacteria;p__Firmicutes;c__Clostridia;o__Oscillospirales;f__Oscillospiraceae;g__ER4 | 95.6 | blast;96.90 iPHoP-RF;70.20 |
| S4-wms_107182 | SHHC | n.a. | n.a. | n.a. | n.a. | n.a. | n.a. | n.a. | n.a. | 7.59 | d__Bacteria;p__Actinobacteriota;c__Coriobacteriia;o__Coriobacteriales;f__Eggerthellaceae;g__CAG-1427 | 96.9 | blast;97.80 |
| S2-wms-combined_143737 | Shared | Duplodnaviria | Heunggongvirae | Uroviricota | Caudoviricetes | n.a. | Duneviridae | O | O | 99.75 | d__Bacteria;p__Bacteroidota;c__Bacteroidia;o__Bacteroidales;f__Bacteroidaceae;g__Bacteroides | 100 | blast;97.80 RaFAH;97.50 iPHoP-RF;78.50 |
| S12-wms-combined_368190 | Shared | Duplodnaviria | Heunggongvirae | Uroviricota | Caudoviricetes | n.a. | Unclassified | O | O | 100 | d__Bacteria;p__Bacteroidota;c__Bacteroidia;o__Bacteroidales;f__Bacteroidaceae;g__Bacteroides | 100 | RaFAH;98.90 iPHoP-RF;97.10 blast;91.30 |
| S4-wms_141607 | SHHC | Duplodnaviria | Heunggongvirae | Uroviricota | Caudoviricetes | n.a. | Unclassified | Unclassified | Unclassified | 37.81 | d__Bacteria;p__Bacteroidota;c__Bacteroidia;o__Bacteroidales;f__Bacteroidaceae;g__Prevotella | 98.4 | CRISPR;98.80 blast;96.40 iPHoP-RF;79.00 |
| S2-wms-combined_157934 | Shared | Duplodnaviria | Heunggongvirae | Uroviricota | Caudoviricetes | n.a. | Unclassified | Unclassified | O | 99.51 | d__Bacteria;p__Firmicutes;c__Clostridia;o__Lachnospirales;f__Lachnospiraceae;g__Roseburia | 98.8 | CRISPR;98.90 iPHoP-RF;92.80 blast;89.30 RaFAH;59.00 |
| S4-wms_161443 | SHHC | Unclassified | n.a. | n.a. | n.a. | n.a. | Unclassified | Unclassified | Unclassified | 5.84 | d__Bacteria;p__Actinobacteriota;c__Actinomycetia;o__Actinomycetales;f__Bifidobacteriaceae;g__Bifidobacterium | 99.6 | CRISPR;99.60 iPHoP-RF;78.00 |
| S4-wms_45128 | SHHC | n.a. | n.a. | n.a. | n.a. | n.a. | n.a. | n.a. | n.a. | 28.59 | d__Bacteria;p__Bacteroidota;c__Bacteroidia;o__Bacteroidales;f__Bacteroidaceae;g__Prevotella | 94.7 | blast;96.30 CRISPR;93.50 iPHoP-RF;91.40 |
| S2-wms-combined_159670 | Shared | n.a. | n.a. | n.a. | n.a. | n.a. | n.a. | n.a. | n.a. | 16.22 | d__Bacteria;p__Firmicutes;c__Clostridia;o__Oscillospirales;f__Oscillospiraceae;g__CAG-83 | 91.6 | blast;93.90 iPHoP-RF;88.20 |
| S4-wms_62635 | SHHC | Duplodnaviria | Heunggongvirae | Uroviricota | Caudoviricetes | n.a. | Unclassified | Unclassified | Unclassified | 93.28 | d__Bacteria;p__Firmicutes;c__Clostridia;o__Lachnospirales;f__Lachnospiraceae;g__Blautia_A | 100 | CRISPR;98.90 RaFAH;95.20 iPHoP-RF;93.70 blast;90.40 |
| S4-wms_73990 | SHHC | Duplodnaviria | Heunggongvirae | Uroviricota | Caudoviricetes | Crassvirales | Suoliviridae | Boorivirinae | Cohcovirus | 76.93 | d__Bacteria;p__Bacteroidota;c__Bacteroidia;o__Bacteroidales;f__Tannerellaceae;g__Parabacteroides | 97.6 | blast;98.30 iPHoP-RF;82.80 |
| S5-wms-combined_117213 | Shared | Duplodnaviria | Heunggongvirae | Uroviricota | Caudoviricetes | n.a. | Unclassified | O | O | 97.16 | d__Bacteria;p__Actinobacteriota;c__Actinomycetia;o__Actinomycetales;f__Bifidobacteriaceae;g__Bifidobacterium | 98.8 | CRISPR;97.70 iPHoP-RF;97.10 blast;96.50 RaFAH;80.80 |
| S5-wms-combined_134381 | Shared | Duplodnaviria | Heunggongvirae | Uroviricota | Caudoviricetes | n.a. | Unclassified | Unclassified | Lagaffevirus | 97.01 | d__Bacteria;p__Firmicutes;c__Clostridia;o__Oscillospirales;f__Oscillospiraceae;g__CAG-83 | 98.4 | RaFAH;98.30 blast;64.70 |
| S5-wms-combined_145067 | Shared | Duplodnaviria | Heunggongvirae | Uroviricota | Caudoviricetes | n.a. | Unclassified | O | O | 82.3 | d__Bacteria;p__Firmicutes;c__Clostridia;o__Oscillospirales;f__Oscillospiraceae;g__Flavonifractor | 95.6 | blast;96.90 iPHoP-RF;95.70 CRISPR;53.80 |
| S2-wms-combined_185494 | Shared | Duplodnaviria | Heunggongvirae | Uroviricota | Caudoviricetes | n.a. | Autographiviridae | O | O | 25.15 | d__Bacteria;p__Firmicutes;c__Negativicutes;o__Veillonellales;f__Dialisteraceae;g__Dialister | 96.3 | CRISPR;97.40 blast;96.90 iPHoP-RF;90.10 |
| S5-wms-combined_178432 | Shared | n.a. | n.a. | n.a. | n.a. | n.a. | n.a. | n.a. | n.a. | 13.6 | d__Bacteria;p__Bacteroidota;c__Bacteroidia;o__Bacteroidales;f__Rikenellaceae;g__Alistipes | 95.9 | iPHoP-RF;97.10 blast;91.90 CRISPR;84.60 |
| S2-wms-combined_191613 | Shared | n.a. | n.a. | n.a. | n.a. | n.a. | n.a. | n.a. | n.a. | 13.4 | d__Bacteria;p__Firmicutes;c__Clostridia;o__Lachnospirales;f__Anaerotignaceae;g__Anaerotignum | 95.2 | blast;96.60 iPHoP-RF;70.60 |
| S5-wms-combined_18356 | ME | Duplodnaviria | Heunggongvirae | Uroviricota | Caudoviricetes | n.a. | Peduoviridae | Unclassified | Evevirus | 90.68 | d__Bacteria;p__Proteobacteria;c__Gammaproteobacteria;o__Enterobacterales;f__Enterobacteriaceae;g__Escherichia | 98.8 | iPHoP-RF;97.10 blast;96.40 RaFAH;85.20 CRISPR;70.30 |
| S12-wms-combined_52101 | SHHC | Duplodnaviria | Heunggongvirae | Uroviricota | Caudoviricetes | n.a. | Unclassified | Unclassified | O | 35.58 | d__Bacteria;p__Firmicutes;c__Clostridia;o__Oscillospirales;f__Ruminococcaceae;g__UBA3818 | 90.3 | blast;92.80 iPHoP-RF;62.20 |
| S5-wms-combined_207291 | Shared | n.a. | n.a. | n.a. | n.a. | n.a. | n.a. | n.a. | n.a. | 100 | d__Bacteria;p__Bacteroidota;c__Bacteroidia;o__Bacteroidales;f__Bacteroidaceae;g__Bacteroides | 100 | RaFAH;99.80 iPHoP-RF;95.70 blast;91.90 |
| S2-wms-combined_199384 | SHHC | Duplodnaviria | Heunggongvirae | Uroviricota | Caudoviricetes | n.a. | Unclassified | O | O | 17.07 | d__Bacteria;p__Firmicutes;c__Clostridia;o__Oscillospirales;f__Butyricicoccaceae;g__Agathobaculum | 91.4 | blast;93.70 iPHoP-RF;73.50 |
| S2-wms-combined_200390 | Shared | Duplodnaviria | Heunggongvirae | Uroviricota | Caudoviricetes | n.a. | Unclassified | O | O | 14.49 | d__Bacteria;p__Firmicutes;c__Clostridia;o__Lachnospirales;f__Lachnospiraceae;g__Eisenbergiella | 92 | CRISPR;94.20 iPHoP-RF;67.50 |
| S5-wms-combined_39793 | Shared | n.a. | n.a. | n.a. | n.a. | n.a. | n.a. | n.a. | n.a. | | d__Bacteria;p__Actinobacteriota;c__Actinomycetia;o__Actinomycetales;f__Bifidobacteriaceae;g__Bifidobacterium | 91.6 | blast;93.90 iPHoP-RF;93.40 CRISPR;76.80 |
| S5-wms-combined_52741 | Shared | Duplodnaviria | Heunggongvirae | Uroviricota | Caudoviricetes | n.a. | Unclassified | O | O | 50.99 | d__Bacteria;p__Verrucomicrobiota;c__Verrucomicrobiae;o__Verrucomicrobiales;f__Akkermansiaceae;g__Akkermansia | 100 | RaFAH;99.50 CRISPR;98.80 blast;96.90 iPHoP-RF;81.60 |
| S12-wms-combined_75733 | SHHC | Monodnaviria | Sangervirae | Phixviricota | Malgrandaviricetes | Petitvirales | Microviridae | O | O | | d__Bacteria;p__Bacteroidota;c__Bacteroidia;o__Bacteroidales;f__Barnesiellaceae;g__Barnesiella | 94.6 | blast;96.20 iPHoP-RF;69.80 |
| S5-wms-combined_59064 | Shared | Duplodnaviria | Heunggongvirae | Uroviricota | Caudoviricetes | n.a. | Unclassified | Unclassified | Unclassified | 95 | d__Bacteria;p__Bacteroidota;c__Bacteroidia;o__Bacteroidales;f__Bacteroidaceae;g__Bacteroides | 100 | RaFAH;97.60 blast;96.90 iPHoP-RF;96.10 |
| S5-wms-combined_66017 | ME | n.a. | n.a. | n.a. | n.a. | n.a. | n.a. | n.a. | n.a. | 9.66 | d__Bacteria;p__Firmicutes;c__Clostridia;o__Christensenellales;f__Christensenellaceae;g__Christensenella | 90.7 | blast;93.10 iPHoP-RF;82.20 |
| S5-wms-combined_7292 | Shared | Duplodnaviria | Heunggongvirae | Uroviricota | Caudoviricetes | n.a. | Unclassified | Unclassified | Unclassified | 22.4 | d__Bacteria;p__Firmicutes;c__Clostridia;o__Oscillospirales;f__Acutalibacteraceae;g__Ruminococcus_E | 98.5 | CRISPR;98.90 blast;96.60 iPHoP-RF;89.80 |
| S5-wms-combined_73301 | ME | Duplodnaviria | Heunggongvirae | Uroviricota | Caudoviricetes | n.a. | Unclassified | O | O | 96.75 | d__Bacteria;p__Firmicutes;c__Clostridia;o__Lachnospirales;f__Lachnospiraceae;g__Dorea_A | 94.6 | blast;96.20 iPHoP-RF;93.10 CRISPR;58.60 |
| S2-wms-combined_225908 | Shared | Unclassified | n.a. | n.a. | n.a. | n.a. | Unclassified | O | O | 87.71 | d__Bacteria;p__Firmicutes;c__Clostridia;o__Oscillospirales;f__Acutalibacteraceae;g__CAG-217 | 95.6 | blast;96.90 CRISPR;93.30 iPHoP-RF;79.00 |
| S5-wms-combined_83370 | Shared | Duplodnaviria | Heunggongvirae | Uroviricota | Caudoviricetes | n.a. | Unclassified | O | O | 82.36 | d__Bacteria;p__Bacteroidota;c__Bacteroidia;o__Bacteroidales;f__Bacteroidaceae;g__Bacteroides | 99.4 | blast;96.90 iPHoP-RF;96.70 RaFAH;91.90 |
| S5-wms-combined_83412 | ME | Duplodnaviria | Heunggongvirae | Uroviricota | Caudoviricetes | n.a. | Unclassified | Unclassified | Unclassified | 25.58 | d__Bacteria;p__Firmicutes;c__Clostridia;o__Eubacteriales;f__Eubacteriaceae;g__Eubacterium | 92.6 | blast;94.70 iPHoP-RF;86.70 CRISPR;77.00 |
| S2-wms-combined_24549 | Shared | Duplodnaviria | Heunggongvirae | Uroviricota | Caudoviricetes | n.a. | Unclassified | O | O | 17.49 | d__Bacteria;p__Firmicutes;c__Clostridia;o__Oscillospirales;f__Oscillospiraceae;g__Intestinimonas | 90.9 | blast;93.30 |
| S5-wms-combined_98202 | Shared | n.a. | n.a. | n.a. | n.a. | n.a. | n.a. | n.a. | n.a. | 100 | d__Bacteria;p__Bacteroidota;c__Bacteroidia;o__Bacteroidales;f__Bacteroidaceae;g__Bacteroides | 99.4 | RaFAH;99.50 |
| S2-wms-combined_25727 | Shared | Duplodnaviria | Heunggongvirae | Uroviricota | Caudoviricetes | n.a. | Unclassified | O | O | 21.03 | d__Bacteria;p__Firmicutes;c__Clostridia;o__Christensenellales;f__CAG-74;g__Ventricola | 94.7 | blast;96.30 iPHoP-RF;74.60 |
| S12-wms_184276 | SHHC | Duplodnaviria | Heunggongvirae | Uroviricota | Caudoviricetes | n.a. | Unclassified | O | O | 36.83 | d__Bacteria;p__Actinobacteriota;c__Coriobacteriia;o__Coriobacteriales;f__Eggerthellaceae;g__Adlercreutzia | 93.1 | blast;95.10 iPHoP-RF;66.80 |
| S12-wms_223235 | SHHC | n.a. | n.a. | n.a. | n.a. | n.a. | n.a. | n.a. | n.a. | | d__Bacteria;p__Firmicutes;c__Clostridia;o__Oscillospirales;f__Ruminococcaceae;g__Ruminococcus_D | 95.3 | blast;96.70 iPHoP-RF;74.80 CRISPR;67.70 |
| S2-wms-combined_52568 | Shared | n.a. | n.a. | n.a. | n.a. | n.a. | n.a. | n.a. | n.a. | 12.66 | d__Bacteria;p__Firmicutes;c__Clostridia;o__Oscillospirales;f__Oscillospiraceae;g__F23-B02 | 92.3 | blast;94.50 iPHoP-RF;80.30 |
| S12-wms_336780 | Shared | n.a. | n.a. | n.a. | n.a. | n.a. | n.a. | n.a. | n.a. | 4.76 | d__Bacteria;p__Firmicutes;c__Clostridia;o__Oscillospirales;f__Ruminococcaceae;g__UMGS363 | 95.3 | blast;96.70 iPHoP-RF;93.10 |
| S2-wms-combined_5791 | Shared | Duplodnaviria | Heunggongvirae | Uroviricota | Caudoviricetes | n.a. | Unclassified | O | O | 89.76 | d__Bacteria;p__Firmicutes;c__Clostridia;o__Lachnospirales;f__Lachnospiraceae;g__Agathobacter | 92.1 | CRISPR;94.30 |
| S2-wms-combined_62839 | Shared | Duplodnaviria | Heunggongvirae | Uroviricota | Caudoviricetes | n.a. | Unclassified | O | O | 32.39 | d__Bacteria;p__Firmicutes;c__Clostridia;o__Oscillospirales;f__Ruminococcaceae;g__Gemmiger | 97.6 | blast;98.30 iPHoP-RF;87.30 CRISPR;73.60 |
| S2-wms-combined_72725 | Shared | n.a. | n.a. | n.a. | n.a. | n.a. | n.a. | n.a. | n.a. | 19.31 | d__Bacteria;p__Firmicutes;c__Clostridia;o__Oscillospirales;f__Oscillospiraceae;g__Intestinimonas | 91.4 | blast;93.70 iPHoP-RF;75.40 |
| S2-wms-combined_74387 | SHHC | Duplodnaviria | Heunggongvirae | Uroviricota | Caudoviricetes | n.a. | Unclassified | O | O | 38.64 | d__Bacteria;p__Firmicutes;c__Clostridia;o__Lachnospirales;f__Lachnospiraceae;g__Lachnoclostridium_A | 94.3 | blast;96.00 iPHoP-RF;82.50 |
| S2-wms-combined_75144 | Shared | n.a. | n.a. | n.a. | n.a. | n.a. | n.a. | n.a. | n.a. | 9.77 | d__Bacteria;p__Bacteroidota;c__Bacteroidia;o__Bacteroidales;f__Tannerellaceae;g__Parabacteroides | 95.4 | blast;96.80 iPHoP-RF;95.10 |
| S2-wms-combined_7756 | Shared | Duplodnaviria | Heunggongvirae | Uroviricota | Caudoviricetes | n.a. | Unclassified | O | O | 9.67 | d__Bacteria;p__Verrucomicrobiota;c__Verrucomicrobiae;o__Verrucomicrobiales;f__Akkermansiaceae;g__Akkermansia | 96.9 | blast;96.70 iPHoP-RF;91.10 CRISPR;86.00 RaFAH;66.20 |
| S17-wms-combined_253499 | ME | n.a. | n.a. | n.a. | n.a. | n.a. | n.a. | n.a. | n.a. | 25.09 | d__Bacteria;p__Bacteroidota;c__Bacteroidia;o__Bacteroidales;f__Rikenellaceae;g__Alistipes | 97.2 | CRISPR;98.00 blast;95.10 iPHoP-RF;90.80 |
| S17-wms-combined_258087 | Shared | Duplodnaviria | Heunggongvirae | Uroviricota | Caudoviricetes | n.a. | Unclassified | Unclassified | O | 97.18 | d__Bacteria;p__Firmicutes;c__Clostridia;o__Clostridiales;f__Clostridiaceae;g__Clostridium | 98.2 | RaFAH;98.70 |
| S17-wms-combined_39687 | ME | n.a. | n.a. | n.a. | n.a. | n.a. | n.a. | n.a. | n.a. | 83.01 | d__Bacteria;p__Firmicutes;c__Clostridia;o__Oscillospirales;f__Oscillospiraceae;g__Flavonifractor | 95.2 | blast;96.60 iPHoP-RF;89.20 |
| S17-wms-combined_56006 | Shared | Duplodnaviria | Heunggongvirae | Uroviricota | Caudoviricetes | n.a. | Unclassified | O | O | 98.39 | d__Bacteria;p__Firmicutes;c__Clostridia;o__Oscillospirales;f__Ruminococcaceae;g__Faecalibacterium | 100 | RaFAH;98.90 blast;96.90 iPHoP-RF;87.00 CRISPR;67.80 |
| S17-wms-combined_56061 | Shared | n.a. | n.a. | n.a. | n.a. | n.a. | n.a. | n.a. | n.a. | 66.23 | d__Bacteria;p__Firmicutes;c__Clostridia;o__Oscillospirales;f__Ruminococcaceae;g__Faecalibacterium | 98.5 | blast;98.90 |
| S17-wms-combined_66123 | ME | Duplodnaviria | Heunggongvirae | Uroviricota | Caudoviricetes | n.a. | Unclassified | Unclassified | O | 97.65 | d__Bacteria;p__Firmicutes;c__Clostridia;o__Oscillospirales;f__Oscillospiraceae;g__Flavonifractor | 90.6 | blast;91.80 iPHoP-RF;85.40 CRISPR;73.10 RaFAH;57.20 |
| S5-wms-combined_108794 | Shared | Duplodnaviria | Heunggongvirae | Uroviricota | Caudoviricetes | n.a. | Unclassified | Unclassified | O | 58.74 | d__Bacteria;p__Firmicutes;c__Clostridia;o__Oscillospirales;f__Acutalibacteraceae;g__Ruminococcus_E | 95.6 | blast;96.90 iPHoP-RF;89.50 |
| S17-wms-combined_75892 | ME | Duplodnaviria | Heunggongvirae | Uroviricota | Caudoviricetes | n.a. | Unclassified | Unclassified | O | 53.86 | d__Bacteria;p__Firmicutes;c__Clostridia;o__Oscillospirales;f__Oscillospiraceae;g__HGM12998 | 91.4 | blast;93.70 iPHoP-RF;77.50 |
| S5-wms-combined_66662 | Shared | Unclassified | n.a. | n.a. | n.a. | n.a. | Unclassified | Unclassified | O | 100 | d__Bacteria;p__Firmicutes;c__Clostridia;o__Oscillospirales;f__Ruminococcaceae;g__Ruthenibacterium | 97.2 | blast;98.00 iPHoP-RF;88.50 |
| S5-wms-combined_72410 | Shared | Duplodnaviria | Heunggongvirae | Uroviricota | Caudoviricetes | n.a. | Unclassified | Unclassified | O | 60.27 | d__Bacteria;p__Firmicutes;c__Clostridia;o__Oscillospirales;f__Ruminococcaceae;g__Ruthenibacterium | 90.8 | blast;93.20 iPHoP-RF;72.60 |
| S2-wms-combined_122457 | Shared | Duplodnaviria | Heunggongvirae | Uroviricota | Caudoviricetes | n.a. | Unclassified | Unclassified | O | 52.93 | d__Bacteria;p__Firmicutes;c__Clostridia;o__Oscillospirales;f__Oscillospiraceae;g__Intestinimonas | 91.6 | blast;93.90 iPHoP-RF;56.50 |
| S5-wms-combined_90315 | Shared | Duplodnaviria | Heunggongvirae | Uroviricota | Caudoviricetes | n.a. | Unclassified | O | O | 99.97 | d__Bacteria;p__Bacteroidota;c__Bacteroidia;o__Bacteroidales;f__Rikenellaceae;g__Alistipes | 98.8 | RaFAH;96.10 blast;88.90 |
| S2-wms-combined_135014 | Shared | Duplodnaviria | Heunggongvirae | Uroviricota | Caudoviricetes | n.a. | Unclassified | Unclassified | O | 100 | d__Bacteria;p__Firmicutes;c__Negativicutes;o__Veillonellales;f__Dialisteraceae;g__Dialister | 100 | RaFAH;97.60 blast;96.90 CRISPR;96.10 iPHoP-RF;92.40 |
| S2-wms-combined_147065 | Shared | Duplodnaviria | Heunggongvirae | Uroviricota | Caudoviricetes | n.a. | Unclassified | O | O | 99.85 | d__Bacteria;p__Bacteroidota;c__Bacteroidia;o__Bacteroidales;f__Bacteroidaceae;g__Bacteroides | 100 | RaFAH;97.00 blast;96.90 iPHoP-RF;95.10 CRISPR;58.20 |
| S2-wms-combined_158821 | Shared | Duplodnaviria | Heunggongvirae | Uroviricota | Caudoviricetes | n.a. | Unclassified | Unclassified | O | 99.16 | d__Bacteria;p__Firmicutes;c__Clostridia;o__Lachnospirales;f__Lachnospiraceae;g__Enterocloster | 91.5 | blast;93.80 iPHoP-RF;63.50 |
| S2-wms-combined_181849 | Shared | n.a. | n.a. | n.a. | n.a. | n.a. | n.a. | n.a. | n.a. | 40.03 | d__Bacteria;p__Firmicutes;c__Clostridia;o__Oscillospirales;f__Ruminococcaceae;g__Faecalibacterium | 95.6 | blast;96.90 CRISPR;95.80 iPHoP-RF;87.30 |
| S2-wms-combined_201205 | Shared | Duplodnaviria | Heunggongvirae | Uroviricota | Caudoviricetes | n.a. | Unclassified | Unclassified | O | 82.22 | d__Bacteria;p__Verrucomicrobiota;c__Verrucomicrobiae;o__Verrucomicrobiales;f__Akkermansiaceae;g__Akkermansia | 100 | CRISPR;99.30 blast;96.80 RaFAH;96.70 iPHoP-RF;68.60 |
| S2-wms-combined_204225 | SHHC | n.a. | n.a. | n.a. | n.a. | n.a. | n.a. | n.a. | n.a. | 6.16 | d__Bacteria;p__Firmicutes;c__Clostridia;o__Lachnospirales;f__Lachnospiraceae;g__Lachnoclostridium_A | 91.6 | blast;93.90 iPHoP-RF;79.30 |
| S2-wms-combined_207000 | SHHC | Duplodnaviria | Heunggongvirae | Uroviricota | Caudoviricetes | n.a. | Unclassified | Unclassified | O | 92.52 | d__Bacteria;p__Firmicutes;c__Clostridia;o__Lachnospirales;f__Lachnospiraceae;g__Agathobacter | 93.4 | blast;95.30 iPHoP-RF;93.70 |
| S2-wms-combined_84608 | Shared | Duplodnaviria | Heunggongvirae | Uroviricota | Caudoviricetes | n.a. | Unclassified | O | O | 21.12 | d__Bacteria;p__Proteobacteria;c__Gammaproteobacteria;o__Burkholderiales;f__Burkholderiaceae;g__Sutterella | 95.6 | blast;96.90 iPHoP-RF;90.80 CRISPR;79.90 |
| S2-wms_246296 | Shared | n.a. | n.a. | n.a. | n.a. | n.a. | n.a. | n.a. | n.a. | 98.04 | d__Bacteria;p__Firmicutes;c__Clostridia;o__Lachnospirales;f__Lachnospiraceae;g__Roseburia | 100 | CRISPR;98.90 blast;94.40 RaFAH;93.70 iPHoP-RF;92.80 |
| S2-wms-combined_208559 | Shared | n.a. | n.a. | n.a. | n.a. | n.a. | n.a. | n.a. | n.a. | 10.84 | d__Bacteria;p__Bacteroidota;c__Bacteroidia;o__Bacteroidales;f__Rikenellaceae;g__Alistipes | 95.6 | blast;96.90 iPHoP-RF;96.70 |

| | | | | | | | | | | | | | |
|---|---|---|---|---|---|---|---|---|---|---|---|---|---|
| S2-wms_6424 | SHHC | Duplodnaviria | Heunggongvirae | Uroviricota | Caudoviricetes | n.a. | Pachyviridae | O | O | 8.31 | d__Bacteria;p__Bacteroidota;c__Bacteroidia;o__Bacteroidales;f__Bacteroidaceae;g__Bacteroides | 93.6 | RaFAH;95.50 |
| S3-wms-combined_135428 | ME | Duplodnaviria | Heunggongvirae | Uroviricota | Caudoviricetes | n.a. | Unclassified | O | O | 11.75 | d__Bacteria;p__Actinobacteriota;c__Actinomycetia;o__Actinomycetales;f__Bifidobacteriaceae;g__Bifidobacterium | 95.3 | blast;96.70 iPHoP-RF;96.40 CRISPR;94.80 |
| S3-wms-combined_45305 | Shared | Duplodnaviria | Heunggongvirae | Uroviricota | Caudoviricetes | n.a. | Unclassified | O | O | 27.39 | d__Bacteria;p__Firmicutes;c__Clostridia;o__Oscillospirales;f__Ruminococcaceae;g__Gemmiger | 94.2 | blast;95.90 iPHoP-RF;76.60 CRISPR;55.10 |
| S2-wms-combined_222535 | SHHC | n.a. | n.a. | n.a. | n.a. | n.a. | n.a. | n.a. | n.a. | 7.99 | d__Bacteria;p__Firmicutes;c__Clostridia;o__Lachnospirales;f__Lachnospiraceae;g__Agathobacter | 92.1 | blast;94.30 iPHoP-RF;92.10 |
| S3-wms_94494 | ME | Duplodnaviria | Heunggongvirae | Uroviricota | Caudoviricetes | n.a. | Unclassified | O | O | 94.44 | d__Bacteria;p__Actinobacteriota;c__Actinomycetia;o__Actinomycetales;f__Bifidobacteriaceae;g__Bifidobacterium | 99.3 | blast;98.30 iPHoP-RF;97.10 CRISPR;83.00 RaFAH;81.50 |
| S4-wms-combined_10758 | SHHC | Duplodnaviria | Heunggongvirae | Uroviricota | Caudoviricetes | n.a. | Unclassified | O | O | 88.87 | d__Bacteria;p__Bacteroidota;c__Bacteroidia;o__Bacteroidales;f__Tannerellaceae;g__Parabacteroides | 100 | RaFAH;99.50 blast;89.70 iPHoP-RF;87.00 |
| S4-wms-combined_118531 | SHHC | Duplodnaviria | Heunggongvirae | Uroviricota | Caudoviricetes | n.a. | Unclassified | O | O | 66.42 | d__Bacteria;p__Firmicutes;c__Negativicutes;o__Acidaminococcales;f__Acidaminococcaceae;g__Phascolarctobacterium_A | 98.2 | CRISPR;98.70 blast;96.80 iPHoP-RF;92.40 |
| S4-wms-combined_121978 | Shared | Duplodnaviria | Heunggongvirae | Uroviricota | Caudoviricetes | n.a. | Unclassified | O | O | 87.01 | d__Bacteria;p__Firmicutes;c__Clostridia;o__Lachnospirales;f__Lachnospiraceae;g__Mediterraneibacter | 95 | blast;96.50 iPHoP-RF;79.50 |
| S4-wms-combined_22012 | SHHC | Duplodnaviria | Heunggongvirae | Uroviricota | Caudoviricetes | n.a. | Unclassified | O | O | 25.83 | d__Bacteria;p__Actinobacteriota;c__Coriobacteriia;o__Coriobacteriales;f__Coriobacteriaceae;g__Collinsella | 98.7 | CRISPR;99.00 blast;96.80 iPHoP-RF;89.20 |
| S4-wms-combined_83492 | Shared | Duplodnaviria | Heunggongvirae | Uroviricota | Caudoviricetes | n.a. | Unclassified | O | O | 86.97 | d__Bacteria;p__Bacteroidota;c__Bacteroidia;o__Bacteroidales;f__Bacteroidaceae;g__Bacteroides | 95.6 | blast;89.90 iPHoP-RF;86.30 RaFAH;84.50 |
| S4-wms_50475 | SHHC | Duplodnaviria | Heunggongvirae | Uroviricota | Caudoviricetes | n.a. | Unclassified | O | O | 70.49 | d__Bacteria;p__Bacteroidota;c__Bacteroidia;o__Bacteroidales;f__Bacteroidaceae;g__Prevotella | 97.3 | blast;96.90 iPHoP-RF;94.40 RaFAH;69.20 CRISPR;50.60 |
| S5-wms-combined_117095 | Shared | n.a. | n.a. | n.a. | n.a. | n.a. | n.a. | n.a. | n.a. | 10.16 | d__Bacteria;p__Firmicutes;c__Clostridia;o__Oscillospirales;f__Acutalibacteraceae;g__UBA1417 | 91 | blast;93.40 iPHoP-RF;64.50 |
| S5-wms-combined_156329 | ME | Duplodnaviria | Heunggongvirae | Uroviricota | Caudoviricetes | n.a. | Unclassified | O | O | 75.2 | d__Bacteria;p__Firmicutes;c__Bacilli;o__Erysipelotrichales;f__Erysipelotrichaceae;g__Clostridium_AQ | 95.9 | blast;97.10 CRISPR;90.20 iPHoP-RF;75.20 |
| S5-wms-combined_179241 | Shared | n.a. | n.a. | n.a. | n.a. | n.a. | n.a. | n.a. | n.a. | 13.11 | d__Bacteria;p__Firmicutes;c__Clostridia;o__Oscillospirales;f__Oscillospiraceae;g__CAG-83 | 91 | blast;93.40 iPHoP-RF;86.70 |
| S5-wms-combined_204782 | ME | n.a. | n.a. | n.a. | n.a. | n.a. | n.a. | n.a. | n.a. | 6.93 | d__Bacteria;p__Bacteroidota;c__Bacteroidia;o__Bacteroidales;f__Rikenellaceae;g__Alistipes_A | 92.1 | blast;94.30 iPHoP-RF;69.10 |
| S5-wms-combined_3201 | ME | Duplodnaviria | Heunggongvirae | Uroviricota | Caudoviricetes | n.a. | Unclassified | O | O | 23.69 | d__Bacteria;p__Proteobacteria;c__Gammaproteobacteria;o__Burkholderiales;f__Burkholderiaceae;g__Oxalobacter | 90.9 | blast;93.30 iPHoP-RF;69.80 |
| S5-wms-combined_94482 | ME | n.a. | n.a. | n.a. | n.a. | n.a. | n.a. | n.a. | n.a. | 15.25 | d__Bacteria;p__Bacteroidota;c__Bacteroidia;o__Bacteroidales;f__Tannerellaceae;g__Parabacteroides | 94.9 | iPHoP-RF;96.40 blast;91.70 |
| S2-wms-combined_30750 | SHHC | n.a. | n.a. | n.a. | n.a. | n.a. | n.a. | n.a. | n.a. | 11.82 | d__Bacteria;p__Firmicutes;c__Clostridia;o__Oscillospirales;f__Ruminococcaceae;g__Ruminococcus_D | 95.6 | blast;96.90 iPHoP-RF;82.50 |
| S2-wms-combined_48661 | SHHC | n.a. | n.a. | n.a. | n.a. | n.a. | n.a. | n.a. | n.a. | 11.3 | d__Bacteria;p__Firmicutes;c__Clostridia;o__Lachnospirales;f__Lachnospiraceae;g__Anaerosacchariphilus | 90.8 | blast;93.20 iPHoP-RF;77.30 |
| S2-wms-combined_57293 | Shared | Unclassified | n.a. | n.a. | n.a. | n.a. | Unclassified | O | O | 78.69 | d__Bacteria;p__Firmicutes;c__Clostridia;o__Oscillospirales;f__Ruminococcaceae;g__Faecalibacterium | 95.7 | blast;96.90 iPHoP-RF;88.90 RaFAH;51.90 |
| S2-wms-combined_58563 | Shared | Duplodnaviria | Heunggongvirae | Uroviricota | Caudoviricetes | n.a. | Unclassified | Unclassified | O | 88.99 | d__Bacteria;p__Firmicutes;c__Clostridia;o__Clostridiales;f__Clostridiaceae;g__Clostridium | 95.4 | RaFAH;96.80 |
| S2-wms-combined_64692 | SHHC | Duplodnaviria | Heunggongvirae | Uroviricota | Caudoviricetes | n.a. | Unclassified | Unclassified | O | 24.76 | d__Bacteria;p__Firmicutes;c__Clostridia;o__Oscillospirales;f__Oscillospiraceae;g__ | 90.8 | blast;93.20 iPHoP-RF;85.40 |
| S2-wms-combined_73327 | SHHC | n.a. | n.a. | n.a. | n.a. | n.a. | n.a. | n.a. | n.a. | 3.55 | d__Bacteria;p__Firmicutes;c__Clostridia;o__Lachnospirales;f__Lachnospiraceae;g__Faecalimonas | 91.6 | blast;93.90 iPHoP-RF;73.50 |
| S2-wms-combined_95987 | SHHC | n.a. | n.a. | n.a. | n.a. | n.a. | n.a. | n.a. | n.a. | 3.24 | d__Bacteria;p__Firmicutes;c__Clostridia;o__Lachnospirales;f__Lachnospiraceae;g__Lachnoclostridium_A | 93.2 | blast;95.20 iPHoP-RF;84.50 |
| S2-wms_124416 | Shared | Duplodnaviria | Heunggongvirae | Uroviricota | Caudoviricetes | n.a. | Unclassified | O | O | 99.32 | d__Bacteria;p__Firmicutes;c__Clostridia;o__Oscillospirales;f__Ruminococcaceae;g__Faecalibacterium | 100 | RaFAH;99.60 blast;96.10 iPHoP-RF;79.30 CRISPR;55.70 |
| S2-wms_161986 | Shared | Duplodnaviria | Heunggongvirae | Uroviricota | Caudoviricetes | n.a. | Unclassified | Unclassified | O | 99.14 | d__Bacteria;p__Firmicutes;c__Clostridia;o__Lachnospirales;f__Lachnospiraceae;g__Lachnospira | 99.9 | CRISPR;98.80 RaFAH;92.10 blast;84.30 iPHoP-RF;67.70 |
| S2-wms_192667 | SHHC | Duplodnaviria | Heunggongvirae | Uroviricota | Caudoviricetes | n.a. | Unclassified | Unclassified | O | 13.43 | d__Bacteria;p__Firmicutes;c__Clostridia;o__Lachnospirales;f__Lachnospiraceae;g__Eubacterium_I | 91.6 | blast;93.90 iPHoP-RF;62.20 |
| S4-wms-combined_57963 | SHHC | Duplodnaviria | Heunggongvirae | Uroviricota | Caudoviricetes | n.a. | Unclassified | Unclassified | O | 74.8 | d__Bacteria;p__Firmicutes;c__Negativicutes;o__Acidaminococcales;f__Acidaminococcaceae;g__Phascolarctobacterium_A | 95.6 | blast;96.90 iPHoP-RF;64.80 |
| S5-wms-combined_163165 | ME | n.a. | n.a. | n.a. | n.a. | n.a. | n.a. | n.a. | n.a. | 3.66 | d__Bacteria;p__Firmicutes;c__Clostridia;o__Oscillospirales;f__Oscillospiraceae;g__Flavonifractor | 91.2 | blast;93.60 iPHoP-RF;59.50 |
| S4-wms-combined_24659 | SHHC | n.a. | n.a. | n.a. | n.a. | n.a. | n.a. | n.a. | n.a. | 6.25 | d__Bacteria;p__Firmicutes;c__Clostridia;o__Lachnospirales;f__Lachnospiraceae;g__Roseburia | 92.6 | iPHoP-RF;94.70 |
| S5-wms-combined_183113 | ME | n.a. | n.a. | n.a. | n.a. | n.a. | n.a. | n.a. | n.a. | 11.29 | d__Bacteria;p__Bacteroidota;c__Bacteroidia;o__Bacteroidales;f__Rikenellaceae;g__Alistipes_A | 97 | blast;97.90 iPHoP-RF;94.70 |
| S5-wms-combined_210959 | Shared | Duplodnaviria | Heunggongvirae | Uroviricota | Caudoviricetes | n.a. | Unclassified | O | O | 81.3 | d__Bacteria;p__Bacteroidota;c__Bacteroidia;o__Bacteroidales;f__Rikenellaceae;g__Alistipes | 99.4 | blast;96.90 iPHoP-RF;96.70 RaFAH;91.40 CRISPR;57.80 |
| S5-wms-combined_67650 | Shared | Duplodnaviria | Heunggongvirae | Uroviricota | Caudoviricetes | n.a. | Unclassified | O | O | 98.81 | d__Bacteria;p__Bacteroidota;c__Bacteroidia;o__Bacteroidales;f__Rikenellaceae;g__Alistipes | 100 | RaFAH;97.10 blast;96.90 iPHoP-RF;93.70 |
| S5-wms-combined_106597 | ME | n.a. | n.a. | n.a. | n.a. | n.a. | n.a. | n.a. | n.a. | 25.93 | d__Bacteria;p__Campylobacterota;c__Campylobacteria;o__Campylobacterales;f__Campylobacteraceae;g__Campylobacter_D | 98.8 | RaFAH;99.10 |
| S5-wms-combined_199135 | ME | n.a. | n.a. | n.a. | n.a. | n.a. | n.a. | n.a. | n.a. | 16.23 | d__Bacteria;p__Cyanobacteria;c__Vampirovibrionia;o__Gastranaerophilales;f__Gastranaerophilaceae;g__QAMI01 | 90.3 | blast;92.80 |
| S5-wms-combined_26061 | Shared | Duplodnaviria | Heunggongvirae | Uroviricota | Caudoviricetes | n.a. | Unclassified | Unclassified | O | 99.77 | d__Bacteria;p__Firmicutes;c__Clostridia;o__Lachnospirales;f__Lachnospiraceae;g__Blautia_A | 96.7 | blast;92.60 RaFAH;84.50 |
| S5-wms-combined_197186 | ME | n.a. | n.a. | n.a. | n.a. | n.a. | n.a. | n.a. | n.a. | 3.6 | d__Bacteria;p__Firmicutes;c__Bacilli;o__Erysipelotrichales;f__Erysipelotrichaceae;g__Clostridium_AQ | 94.6 | blast;96.20 iPHoP-RF;73.70 |
| S5-wms-combined_205046 | Shared | n.a. | n.a. | n.a. | n.a. | n.a. | n.a. | n.a. | n.a. | 4.59 | d__Bacteria;p__Firmicutes;c__Clostridia;o__Oscillospirales;f__Oscillospiraceae;g__Dysosmobacter | 92.6 | iPHoP-RF;94.70 blast;93.60 |
| S5-wms_10783 | Shared | n.a. | n.a. | n.a. | n.a. | n.a. | n.a. | n.a. | n.a. | 3.75 | d__Bacteria;p__Firmicutes;c__Clostridia;o__Oscillospirales;f__Oscillospiraceae;g__Lawsonibacter | 90.8 | blast;93.20 |
| S5-wms_212596 | ME | n.a. | n.a. | n.a. | n.a. | n.a. | n.a. | n.a. | n.a. | 76.97 | d__Bacteria;p__Firmicutes;c__Bacilli;o__Lactobacillales;f__Lactobacillaceae;g__Lacticaseibacillus | 100 | blast;98.70 RaFAH;95.70 iPHoP-RF;77.10 |
| S5-wms_49310 | Shared | Duplodnaviria | Heunggongvirae | Uroviricota | Caudoviricetes | n.a. | Unclassified | Unclassified | Unclassified | 96.19 | d__Bacteria;p__Firmicutes;c__Clostridia;o__Oscillospirales;f__Acutalibacteraceae;g__Ruminococcus_E | 99.7 | blast;98.00 CRISPR;97.80 RaFAH;93.50 iPHoP-RF;88.20 |
| S5-wms_52566 | Shared | n.a. | n.a. | n.a. | n.a. | n.a. | n.a. | n.a. | n.a. | 8.51 | d__Bacteria;p__Firmicutes;c__Clostridia;o__Oscillospirales;f__Oscillospiraceae;g__Intestinimonas | 91.1 | blast;93.50 iPHoP-RF;52.80 |
| S5-wms_62679 | ME | Duplodnaviria | Heunggongvirae | Uroviricota | Caudoviricetes | n.a. | Unclassified | O | O | 11.9 | d__Bacteria;p__Actinobacteriota;c__Coriobacteriia;o__Coriobacteriales;f__Eggerthellaceae;g__Eggerthella | 94.6 | blast;96.20 |
| S5-wms_85650 | Shared | n.a. | n.a. | n.a. | n.a. | n.a. | n.a. | n.a. | n.a. | 99.85 | d__Bacteria;p__Bacteroidota;c__Bacteroidia;o__Bacteroidales;f__Bacteroidaceae;g__Bacteroides | 100 | RaFAH;99.00 iPHoP-RF;97.10 blast;96.60 |
| S5-wms_94965 | ME | Duplodnaviria | Heunggongvirae | Uroviricota | Caudoviricetes | n.a. | Unclassified | Unclassified | Unclassified | 12.7 | d__Bacteria;p__Actinobacteriota;c__Coriobacteriia;o__Coriobacteriales;f__Coriobacteriaceae;g__Collinsella | 97.5 | CRISPR;98.20 blast;95.40 iPHoP-RF;77.10 |
| S6-wms-combined_110950 | Shared | n.a. | n.a. | n.a. | n.a. | n.a. | n.a. | n.a. | n.a. | 42.23 | d__Bacteria;p__Firmicutes;c__Clostridia;o__Oscillospirales;f__Ruminococcaceae;g__Faecalibacterium | 99.6 | blast;96.80 RaFAH;93.40 iPHoP-RF;93.10 CRISPR;73.10 |
| S6-wms-combined_117756 | SHHC | Duplodnaviria | Heunggongvirae | Uroviricota | Caudoviricetes | n.a. | Unclassified | Unclassified | Eponavirus | 50.35 | d__Bacteria;p__Firmicutes;c__Clostridia;o__Lachnospirales;f__Lachnospiraceae;g__Acetatifactor | 91.6 | blast;93.90 iPHoP-RF;77.50 |
| S6-wms-combined_118427 | Shared | Duplodnaviria | Heunggongvirae | Uroviricota | Caudoviricetes | n.a. | Unclassified | Unclassified | O | 40.24 | d__Bacteria;p__Firmicutes;c__Clostridia;o__Oscillospirales;f__Oscillospiraceae;g__Dysosmobacter | 97.9 | iPHoP-RF;93.70 blast;91.90 RaFAH;88.40 |
| S6-wms-combined_119017 | SHHC | Duplodnaviria | Heunggongvirae | Uroviricota | Caudoviricetes | n.a. | Peduoviridae | Unclassified | Phitrevirus | 37.52 | d__Bacteria;p__Desulfobacterota;c__Desulfovibrionia;o__Desulfovibrionales;f__Desulfovibrionaceae;g__Bilophila | 94.2 | blast;95.90 iPHoP-RF;92.40 CRISPR;66.40 |
| S6-wms-combined_119138 | Shared | Duplodnaviria | Heunggongvirae | Uroviricota | Caudoviricetes | n.a. | Unclassified | O | O | 99.25 | d__Bacteria;p__Firmicutes;c__Clostridia;o__Lachnospirales;f__Lachnospiraceae;g__Enterocloster | 91.6 | blast;93.90 |
| S6-wms-combined_126236 | Shared | Duplodnaviria | Heunggongvirae | Uroviricota | Caudoviricetes | n.a. | Unclassified | Unclassified | Toutatisvirus | 26.53 | d__Bacteria;p__Firmicutes;c__Clostridia;o__Oscillospirales;f__Ruminococcaceae;g__Faecalibacterium | 97.6 | blast;98.30 CRISPR;97.70 iPHoP-RF;86.70 |
| S6-wms-combined_135223 | Shared | n.a. | n.a. | n.a. | n.a. | n.a. | n.a. | n.a. | n.a. | 23.09 | d__Bacteria;p__Bacteroidota;c__Bacteroidia;o__Bacteroidales;f__Bacteroidaceae;g__Bacteroides | 99 | iPHoP-RF;96.40 blast;92.30 RaFAH;88.60 |
| S6-wms-combined_138710 | Shared | n.a. | n.a. | n.a. | n.a. | n.a. | n.a. | n.a. | n.a. | 55.97 | d__Bacteria;p__Firmicutes;c__Clostridia;o__Oscillospirales;f__Ruminococcaceae;g__Faecalibacterium | 96.2 | blast;96.90 iPHoP-RF;78.00 RaFAH;56.20 |
| S6-wms-combined_140731 | SHHC | n.a. | n.a. | n.a. | n.a. | n.a. | n.a. | n.a. | n.a. | 27.76 | d__Bacteria;p__Firmicutes;c__Clostridia;o__Oscillospirales;f__Ruminococcaceae;g__Faecalibacterium | 98.4 | CRISPR;92.30 RaFAH;92.10 blast;92.10 iPHoP-RF;80.30 |
| S6-wms-combined_32530 | Shared | Duplodnaviria | Heunggongvirae | Uroviricota | Caudoviricetes | n.a. | Unclassified | O | O | 99.74 | d__Bacteria;p__Firmicutes;c__Clostridia;o__Lachnospirales;f__Lachnospiraceae;g__Roseburia | 99.3 | RaFAH;94.70 blast;87.00 iPHoP-RF;77.10 |
| S6-wms-combined_41246 | Shared | n.a. | n.a. | n.a. | n.a. | n.a. | n.a. | n.a. | n.a. | 21.08 | d__Bacteria;p__Firmicutes;c__Clostridia;o__Oscillospirales;f__Oscillospiraceae;g__Marseille-P3106 | 91.2 | blast;93.60 |
| S6-wms-combined_49909 | SHHC | Duplodnaviria | Heunggongvirae | Uroviricota | Caudoviricetes | n.a. | Unclassified | O | O | 50.09 | d__Bacteria;p__Firmicutes;c__Clostridia;o__Oscillospirales;f__Ruminococcaceae;g__Faecalibacterium | 91 | blast;93.40 iPHoP-RF;86.00 |
| S6-wms-combined_50721 | Shared | Duplodnaviria | Heunggongvirae | Uroviricota | Caudoviricetes | n.a. | Winoviridae | O | O | 99.98 | d__Bacteria;p__Bacteroidota;c__Bacteroidia;o__Bacteroidales;f__Rikenellaceae;g__Alistipes | 99.6 | blast;96.90 iPHoP-RF;96.70 RaFAH;94.30 |
| S6-wms-combined_56464 | Shared | Duplodnaviria | Heunggongvirae | Uroviricota | Caudoviricetes | n.a. | Unclassified | O | O | 51.17 | d__Bacteria;p__Firmicutes;c__Clostridia;o__Oscillospirales;f__Ruminococcaceae;g__Faecalibacterium | 99 | blast;96.90 iPHoP-RF;92.10 RaFAH;87.90 |
| S6-wms-combined_65225 | SHHC | n.a. | n.a. | n.a. | n.a. | n.a. | n.a. | n.a. | n.a. | 23.74 | d__Bacteria;p__Firmicutes;c__Negativicutes;o__Veillonellales;f__Dialisteraceae;g__Dialister | 94.9 | blast;96.40 CRISPR;86.10 iPHoP-RF;77.30 |
| S6-wms-combined_72228 | Shared | n.a. | n.a. | n.a. | n.a. | n.a. | n.a. | n.a. | n.a. | 41.84 | d__Bacteria;p__Bacteroidota;c__Bacteroidia;o__Bacteroidales;f__Rikenellaceae;g__Alistipes | 99 | iPHoP-RF;97.10 blast;96.60 RaFAH;85.90 |
| S6-wms-combined_745 | SHHC | n.a. | n.a. | n.a. | n.a. | n.a. | n.a. | n.a. | n.a. | 7.61 | d__Bacteria;p__Proteobacteria;c__Alphaproteobacteria;o__RF32;f__CAG-239;g__CAG-267 | 90.6 | blast;93.00 iPHoP-RF;84.50 |
| S6-wms-combined_82597 | Shared | n.a. | n.a. | n.a. | n.a. | n.a. | n.a. | n.a. | n.a. | 91.55 | d__Bacteria;p__Bacteroidota;c__Bacteroidia;o__Bacteroidales;f__Bacteroidaceae;g__Bacteroides | 99.7 | iPHoP-RF;97.10 blast;96.90 RaFAH;95.40 |
| S6-wms-combined_91039 | Shared | n.a. | n.a. | n.a. | n.a. | n.a. | n.a. | n.a. | n.a. | 100 | d__Bacteria;p__Bacteroidota;c__Bacteroidia;o__Bacteroidales;f__Rikenellaceae;g__Alistipes | 96.6 | CRISPR;97.60 iPHoP-RF;97.10 blast;96.70 |
| S6-wms_132588 | Shared | Duplodnaviria | Heunggongvirae | Uroviricota | Caudoviricetes | n.a. | Unclassified | O | O | 67.09 | d__Bacteria;p__Firmicutes;c__Clostridia;o__Oscillospirales;f__Ruminococcaceae;g__Faecalibacterium | 93.2 | CRISPR;95.20 blast;94.40 iPHoP-RF;92.80 |
| S6-wms_138342 | SHHC | Duplodnaviria | Heunggongvirae | Uroviricota | Caudoviricetes | Crassvirales | Suoliviridae | O | O | 37.57 | d__Bacteria;p__Bacteroidota;c__Bacteroidia;o__Bacteroidales;f__Bacteroidaceae;g__Prevotella | 91.7 | RaFAH;94.00 |
| S6-wms_140743 | SHHC | Duplodnaviria | Heunggongvirae | Uroviricota | Caudoviricetes | n.a. | Unclassified | O | O | 9.85 | d__Bacteria;p__Firmicutes;c__Clostridia;o__Lachnospirales;f__Lachnospiraceae;g__Enterocloster | 91.2 | blast;93.60 iPHoP-RF;92.40 CRISPR;53.80 |
| S6-wms_54578 | SHHC | Duplodnaviria | Heunggongvirae | Uroviricota | Caudoviricetes | n.a. | Unclassified | Unclassified | Unclassified | 97 | d__Bacteria;p__Bacteroidota;c__Bacteroidia;o__Bacteroidales;f__Tannerellaceae;g__Marseille-P3160 | 97.5 | CRISPR;98.20 iPHoP-RF;69.80 |
| S7-wms-combined_104872 | Shared | n.a. | n.a. | n.a. | n.a. | n.a. | n.a. | n.a. | n.a. | 97.79 | d__Bacteria;p__Bacteroidota;c__Bacteroidia;o__Bacteroidales;f__Bacteroidaceae;g__Bacteroides | 100 | RaFAH;99.70 blast;92.10 iPHoP-RF;90.50 CRISPR;53.10 |
| S7-wms-combined_106253 | Shared | Duplodnaviria | Heunggongvirae | Uroviricota | Caudoviricetes | n.a. | Unclassified | O | O | 71.87 | d__Bacteria;p__Firmicutes;c__Clostridia;o__Lachnospirales;f__Lachnospiraceae;g__Dorea_A | 95.3 | blast;96.70 iPHoP-RF;94.40 |
| S7-wms-combined_107679_0 | Shared | Duplodnaviria | Heunggongvirae | Uroviricota | Caudoviricetes | n.a. | Winoviridae | O | O | 99.98 | d__Bacteria;p__Bacteroidota;c__Bacteroidia;o__Bacteroidales;f__Rikenellaceae;g__Alistipes | 100 | RaFAH;98.50 blast;97.20 iPHoP-RF;96.40 |
| S7-wms-combined_107679_1 | Shared | Duplodnaviria | Heunggongvirae | Uroviricota | Caudoviricetes | n.a. | Unclassified | O | O | 100 | d__Bacteria;p__Bacteroidota;c__Bacteroidia;o__Bacteroidales;f__Bacteroidaceae;g__Bacteroides | 99.9 | RaFAH;99.80 iPHoP-RF;57.60 |
| S7-wms-combined_111004 | Shared | Duplodnaviria | Heunggongvirae | Uroviricota | Caudoviricetes | n.a. | Unclassified | Unclassified | Taranisvirus | 34.72 | d__Bacteria;p__Firmicutes;c__Clostridia;o__Oscillospirales;f__Ruminococcaceae;g__Faecalibacterium | 99.7 | CRISPR;98.50 blast;96.60 RaFAH;90.40 iPHoP-RF;85.70 |
| S7-wms-combined_123085 | Shared | n.a. | n.a. | n.a. | n.a. | n.a. | n.a. | n.a. | n.a. | 3.26 | d__Bacteria;p__Firmicutes;c__Clostridia;o__Lachnospirales;f__Lachnospiraceae;g__Fusicatenibacter | 91.6 | blast;93.90 iPHoP-RF;83.90 |
| S7-wms-combined_134452 | Shared | n.a. | n.a. | n.a. | n.a. | n.a. | n.a. | n.a. | n.a. | 8.94 | d__Bacteria;p__Firmicutes;c__Clostridia;o__Christensenellales;f__CAG-74;g__UBA11524 | 95.6 | blast;96.90 iPHoP-RF;94.70 |
| S7-wms-combined_134458 | Shared | Duplodnaviria | Heunggongvirae | Uroviricota | Caudoviricetes | n.a. | Unclassified | Unclassified | Toutatisvirus | 43.62 | d__Bacteria;p__Firmicutes;c__Clostridia;o__Oscillospirales;f__Ruminococcaceae;g__Gemmiger | 91.6 | blast;93.90 iPHoP-RF;88.90 CRISPR;69.70 |
| S7-wms-combined_135154 | ME | n.a. | n.a. | n.a. | n.a. | n.a. | n.a. | n.a. | n.a. | 99.98 | d__Bacteria;p__Bacteroidota;c__Bacteroidia;o__Bacteroidales;f__Bacteroidaceae;g__Bacteroides | 100 | RaFAH;99.30 iPHoP-RF;97.10 blast;92.40 |
| S7-wms-combined_136642 | ME | n.a. | n.a. | n.a. | n.a. | n.a. | n.a. | n.a. | n.a. | 81.83 | d__Bacteria;p__Actinobacteriota;c__Actinomycetia;o__Actinomycetales;f__Bifidobacteriaceae;g__Bifidobacterium | 99.7 | CRISPR;99.40 iPHoP-RF;97.10 blast;96.80 RaFAH;72.30 |
| S7-wms-combined_137858 | Shared | n.a. | n.a. | n.a. | n.a. | n.a. | n.a. | n.a. | n.a. | 55.93 | d__Bacteria;p__Firmicutes;c__Clostridia;o__Oscillospirales;f__Ruminococcaceae;g__Faecalibacterium | 95.6 | blast;96.90 iPHoP-RF;88.20 |
| S7-wms-combined_139683 | Shared | Duplodnaviria | Heunggongvirae | Uroviricota | Caudoviricetes | n.a. | Unclassified | O | O | 93.85 | d__Bacteria;p__Firmicutes;c__Clostridia;o__Oscillospirales;f__Oscillospiraceae;g__CAG-103 | 93.5 | CRISPR;95.40 |
| S7-wms-combined_14267 | Shared | n.a. | n.a. | n.a. | n.a. | n.a. | n.a. | n.a. | n.a. | 96.95 | d__Bacteria;p__Bacteroidota;c__Bacteroidia;o__Bacteroidales;f__Rikenellaceae;g__Alistipes | 97.5 | CRISPR;98.20 blast;98.10 iPHoP-RF;97.10 |
| S7-wms-combined_143218 | ME | n.a. | n.a. | n.a. | n.a. | n.a. | n.a. | n.a. | n.a. | 6.41 | d__Bacteria;p__Actinobacteriota;c__Actinomycetia;o__Actinomycetales;f__Bifidobacteriaceae;g__Bifidobacterium | 95.2 | blast;96.60 CRISPR;91.70 iPHoP-RF;81.90 |
| S7-wms-combined_14529 | Shared | Duplodnaviria | Heunggongvirae | Uroviricota | Caudoviricetes | n.a. | Unclassified | O | O | 25.87 | d__Bacteria;p__Firmicutes;c__Clostridia;o__Christensenellales;f__CAG-74;g__UBA11524 | 95.6 | blast;96.90 iPHoP-RF;93.40 |
| S7-wms-combined_147158 | ME | Duplodnaviria | Heunggongvirae | Uroviricota | Caudoviricetes | n.a. | Unclassified | O | O | 86.44 | d__Bacteria;p__Actinobacteriota;c__Actinomycetia;o__Actinomycetales;f__Bifidobacteriaceae;g__Bifidobacterium | 99 | CRISPR;99.20 blast;96.90 iPHoP-RF;95.70 RaFAH;50.40 |
| S7-wms-combined_156351 | ME | n.a. | n.a. | n.a. | n.a. | n.a. | n.a. | n.a. | n.a. | 22 | d__Bacteria;p__Firmicutes;c__Clostridia;o__Lachnospirales;f__Lachnospiraceae;g__Blautia_A | 94.5 | blast;96.10 iPHoP-RF;94.10 |
| S7-wms-combined_157260 | Shared | Duplodnaviria | Heunggongvirae | Uroviricota | Caudoviricetes | n.a. | Unclassified | Unclassified | O | 99.48 | d__Bacteria;p__Firmicutes;c__Clostridia;o__Lachnospirales;f__Lachnospiraceae;g__Lachnospira | 98.7 | CRISPR;99.00 |
| S7-wms-combined_157434 | Shared | n.a. | n.a. | n.a. | n.a. | n.a. | n.a. | n.a. | n.a. | 16.06 | d__Bacteria;p__Firmicutes;c__Clostridia;o__Lachnospirales;f__Lachnospiraceae;g__Mediterraneibacter | 91.4 | blast;93.70 iPHoP-RF;86.30 |
| S7-wms-combined_163980 | ME | n.a. | n.a. | n.a. | n.a. | n.a. | n.a. | n.a. | n.a. | 78.4 | d__Bacteria;p__Bacteroidota;c__Bacteroidia;o__Bacteroidales;f__Rikenellaceae;g__Alistipes | 95.2 | blast;96.60 iPHoP-RF;93.70 |
| S7-wms-combined_167867 | ME | n.a. | n.a. | n.a. | n.a. | n.a. | n.a. | n.a. | n.a. | 10.54 | d__Bacteria;p__Actinobacteriota;c__Actinomycetia;o__Actinomycetales;f__Bifidobacteriaceae;g__Bifidobacterium | 97.9 | CRISPR;98.50 iPHoP-RF;97.10 blast;96.70 |
| S7-wms-combined_169375 | ME | n.a. | n.a. | n.a. | n.a. | n.a. | n.a. | n.a. | n.a. | 10.64 | d__Bacteria;p__Firmicutes;c__Bacilli;o__Erysipelotrichales;f__Erysipelatoclostridiaceae;g__Erysipelatoclostridium | 94.7 | blast;96.30 |
| S7-wms-combined_174935 | Shared | Duplodnaviria | Heunggongvirae | Uroviricota | Caudoviricetes | n.a. | Unclassified | O | O | 34.84 | d__Bacteria;p__Actinobacteriota;c__Actinomycetia;o__Actinomycetales;f__Bifidobacteriaceae;g__Bifidobacterium | 96.3 | CRISPR;97.40 blast;97.30 iPHoP-RF;97.10 |
| S7-wms-combined_185016 | Shared | Duplodnaviria | Heunggongvirae | Uroviricota | Caudoviricetes | n.a. | Unclassified | O | O | 95.96 | d__Bacteria;p__Bacteroidota;c__Bacteroidia;o__Bacteroidales;f__Bacteroidaceae;g__Bacteroides | 100 | RaFAH;99.70 blast;95.30 iPHoP-RF;95.10 |
| S7-wms-combined_189250 | Shared | Duplodnaviria | Heunggongvirae | Uroviricota | Caudoviricetes | n.a. | Unclassified | Unclassified | Unclassified | 22.37 | d__Bacteria;p__Actinobacteriota;c__Coriobacteriia;o__Coriobacteriales;f__Coriobacteriaceae;g__Collinsella | 98.7 | CRISPR;99.00 blast;91.90 iPHoP-RF;80.00 |
| S7-wms-combined_190281 | Shared | n.a. | n.a. | n.a. | n.a. | n.a. | n.a. | n.a. | n.a. | 13.36 | d__Bacteria;p__Actinobacteriota;c__Coriobacteriia;o__Coriobacteriales;f__Coriobacteriaceae;g__Collinsella | 96.2 | CRISPR;97.30 blast;96.90 iPHoP-RF;86.30 |
| S7-wms-combined_20921 | ME | Duplodnaviria | Heunggongvirae | Uroviricota | Caudoviricetes | n.a. | Unclassified | O | O | 29.3 | d__Bacteria;p__Firmicutes;c__Clostridia;o__Lachnospirales;f__Lachnospiraceae;g__Anaerobutyricum | 95 | blast;96.50 iPHoP-RF;77.50 |
| S7-wms-combined_217226 | ME | Duplodnaviria | Heunggongvirae | Uroviricota | Caudoviricetes | n.a. | Unclassified | O | O | 97.48 | d__Bacteria;p__Verrucomicrobiota;c__Verrucomicrobiae;o__Verrucomicrobiales;f__Akkermansiaceae;g__Akkermansia | 100 | CRISPR;99.50 RaFAH;97.70 iPHoP-RF;58.20 |
| S7-wms-combined_218379 | Shared | Duplodnaviria | Heunggongvirae | Uroviricota | Caudoviricetes | n.a. | Unclassified | O | O | 16.97 | d__Bacteria;p__Firmicutes;c__Clostridia;o__Oscillospirales;f__Acutalibacteraceae;g__CAG-180 | 98.7 | CRISPR;99.00 blast;96.60 iPHoP-RF;86.30 |
| S7-wms-combined_224523 | Shared | Duplodnaviria | Heunggongvirae | Uroviricota | Caudoviricetes | n.a. | Unclassified | O | O | 99.68 | d__Bacteria;p__Firmicutes;c__Clostridia;o__Oscillospirales;f__Ruminococcaceae;g__Ruminococcus_D | 99.4 | blast;96.80 iPHoP-RF;93.70 RaFAH;91.90 |
| S7-wms-combined_235165 | ME | Duplodnaviria | Heunggongvirae | Uroviricota | Caudoviricetes | n.a. | Unclassified | Unclassified | Unclassified | 18.87 | d__Bacteria;p__Firmicutes;c__Clostridia;o__Lachnospirales;f__Lachnospiraceae;g__Agathobacter | 90.2 | blast;92.70 |
| S7-wms-combined_238231 | Shared | n.a. | n.a. | n.a. | n.a. | n.a. | n.a. | n.a. | n.a. | 39.27 | d__Bacteria;p__Firmicutes;c__Clostridia;o__Oscillospirales;f__Ruminococcaceae;g__Negativibacillus | 95.6 | blast;96.90 iPHoP-RF;77.50 CRISPR;68.00 |
| S7-wms-combined_239784 | Shared | n.a. | n.a. | n.a. | n.a. | n.a. | n.a. | n.a. | n.a. | 67.8 | d__Bacteria;p__Bacteroidota;c__Bacteroidia;o__Bacteroidales;f__Bacteroidaceae;g__Bacteroides | 99.3 | iPHoP-RF;97.10 RaFAH;89.00 blast;88.60 |
| S7-wms-combined_26217 | Shared | Duplodnaviria | Heunggongvirae | Uroviricota | Caudoviricetes | n.a. | Unclassified | O | O | 100 | d__Bacteria;p__Bacteroidota;c__Bacteroidia;o__Bacteroidales;f__Rikenellaceae;g__Alistipes | 100 | RaFAH;98.00 blast;96.90 iPHoP-RF;88.90 |
| S7-wms-combined_28138 | ME | Duplodnaviria | Heunggongvirae | Uroviricota | Caudoviricetes | n.a. | Unclassified | O | O | 12.56 | d__Bacteria;p__Firmicutes;c__Clostridia;o__Oscillospirales;f__Acutalibacteraceae;g__Ruminococcus_E | 98.7 | CRISPR;99.00 blast;96.80 iPHoP-RF;91.10 |

| | | | | | | | | | | | | | |
|---|---|---|---|---|---|---|---|---|---|---|---|---|---|
| S7-wms-combined_31068 | Shared | n.a. | n.a. | n.a. | n.a. | n.a. | n.a. | n.a. | n.a. | 9.44 | d__Bacteria;p__Firmicutes;c__Clostridia;o__Oscillospirales;f__Oscillospiraceae;g__Lawsonibacter | 91.5 | blast;93.80 iPHoP-RF;81.90 |
| S7-wms-combined_3669 | ME | Monodnaviria | Sangervirae | Phixviricota | Malgrandaviricetes | Petitvirales | Microviridae | O | O | 8.7 | d__Bacteria;p__Bacteroidota;c__Bacteroidia;o__Bacteroidales;f__Bacteroidaceae;g__Bacteroides | 91.5 | blast;93.80 iPHoP-RF;87.60 |
| S7-wms-combined_3697 | Shared | Unclassified | n.a. | n.a. | n.a. | n.a. | Unclassified | O | O | 72.05 | d__Bacteria;p__Bacteroidota;c__Bacteroidia;o__Bacteroidales;f__Rikenellaceae;g__Alistipes | 98.8 | iPHoP-RF;97.10 blast;96.90 CRISPR;95.90 RaFAH;84.30 |
| S7-wms-combined_43950 | Shared | Duplodnaviria | Heunggongvirae | Uroviricota | Caudoviricetes | n.a. | Unclassified | Unclassified | Unclassified | 99.99 | d__Bacteria;p__Bacteroidota;c__Bacteroidia;o__Bacteroidales;f__Bacteroidaceae;g__Bacteroides | 100 | RaFAH;99.70 iPHoP-RF;95.10 blast;93.90 |
| S7-wms-combined_5625 | Shared | Duplodnaviria | Heunggongvirae | Uroviricota | Caudoviricetes | n.a. | Unclassified | Unclassified | Unclassified | 95.36 | d__Bacteria;p__Bacteroidota;c__Bacteroidia;o__Bacteroidales;f__Bacteroidaceae;g__Bacteroides | 100 | RaFAH;98.60 iPHoP-RF;97.10 blast;96.90 |
| S7-wms-combined_62119 | Shared | Duplodnaviria | Heunggongvirae | Uroviricota | Caudoviricetes | n.a. | Unclassified | Unclassified | O | 99.99 | d__Bacteria;p__Bacteroidota;c__Bacteroidia;o__Bacteroidales;f__Bacteroidaceae;g__Bacteroides | 98.8 | blast;96.90 iPHoP-RF;96.40 CRISPR;93.30 RaFAH;84.80 |
| S7-wms-combined_69024 | ME | Duplodnaviria | Heunggongvirae | Uroviricota | Caudoviricetes | n.a. | Unclassified | O | O | 58.17 | d__Bacteria;p__Firmicutes;c__Clostridia;o__Lachnospirales;f__Lachnospiraceae;g__Bariatricus | 91.6 | blast;93.90 iPHoP-RF;75.90 |
| S7-wms-combined_71592 | Shared | Duplodnaviria | Heunggongvirae | Uroviricota | Caudoviricetes | n.a. | Unclassified | O | O | 39.23 | d__Bacteria;p__Firmicutes;c__Clostridia;o__Oscillospirales;f__Ruminococcaceae;g__Ruminococcus_D | 98.1 | blast;98.60 iPHoP-RF;92.40 CRISPR;61.00 |
| S7-wms-combined_72070 | Shared | n.a. | n.a. | n.a. | n.a. | n.a. | n.a. | n.a. | n.a. | 35.71 | d__Bacteria;p__Bacteroidota;c__Bacteroidia;o__Bacteroidales;f__Bacteroidaceae;g__Bacteroides | 96.2 | iPHoP-RF;97.10 blast;96.90 CRISPR;55.90 RaFAH;52.60 |
| S7-wms-combined_74829 | Shared | Duplodnaviria | Heunggongvirae | Uroviricota | Caudoviricetes | n.a. | Unclassified | O | O | 77.29 | d__Bacteria;p__Firmicutes;c__Negativicutes;o__Veillonellales;f__Dialisteraceae;g__Dialister | 97.3 | CRISPR;97.40 blast;96.80 iPHoP-RF;94.70 RaFAH;63.90 |
| S7-wms-combined_84688 | Shared | n.a. | n.a. | n.a. | n.a. | n.a. | n.a. | n.a. | n.a. | 29.6 | d__Bacteria;p__Firmicutes;c__Clostridia;o__Oscillospirales;f__Oscillospiraceae;g__Marseille-P3106 | 91.6 | blast;93.90 iPHoP-RF;86.70 |
| S7-wms-combined_90393 | ME | Duplodnaviria | Heunggongvirae | Uroviricota | Caudoviricetes | n.a. | Unclassified | O | O | 32.36 | d__Bacteria;p__Bacteroidota;c__Bacteroidia;o__Bacteroidales;f__Bacteroidaceae;g__Bacteroides | 99.4 | iPHoP-RF;97.10 blast;91.80 RaFAH;91.30 |
| S7-wms-combined_90728 | ME | Duplodnaviria | Heunggongvirae | Uroviricota | Caudoviricetes | n.a. | Unclassified | O | O | 66.45 | d__Bacteria;p__Bacteroidota;c__Bacteroidia;o__Bacteroidales;f__Bacteroidaceae;g__Bacteroides | 100 | RaFAH;98.90 iPHoP-RF;96.70 blast;91.80 |
| S7-wms_113137 | Shared | Duplodnaviria | Heunggongvirae | Uroviricota | Caudoviricetes | n.a. | Unclassified | Unclassified | Mushuvirus | 99.39 | d__Bacteria;p__Firmicutes;c__Clostridia;o__Oscillospirales;f__Ruminococcaceae;g__Faecalibacterium | 99.4 | RaFAH;99.50 |
| S7-wms_123954 | ME | n.a. | n.a. | n.a. | n.a. | n.a. | n.a. | n.a. | n.a. | 100 | d__Bacteria;p__Bacteroidota;c__Bacteroidia;o__Bacteroidales;f__Tannerellaceae;g__Parabacteroides | 98.4 | blast;96.40 RaFAH;82.90 iPHoP-RF;60.30 |
| S7-wms_127801 | Shared | Duplodnaviria | Heunggongvirae | Uroviricota | Caudoviricetes | n.a. | Unclassified | O | O | 10.6 | d__Bacteria;p__Firmicutes;c__Clostridia;o__Oscillospirales;f__Ruminococcaceae;g__Ruminococcus_D | 98.7 | CRISPR;99.00 blast;97.40 iPHoP-RF;93.70 |
| S7-wms_151112 | ME | Duplodnaviria | Heunggongvirae | Uroviricota | Caudoviricetes | n.a. | Unclassified | O | O | 20.08 | d__Bacteria;p__Firmicutes;c__Clostridia;o__Lachnospirales;f__Lachnospiraceae;g__Anaerobutyricum | 90.3 | blast;92.80 iPHoP-RF;86.00 |
| S7-wms_157264 | ME | Unclassified | n.a. | n.a. | n.a. | n.a. | Unclassified | Unclassified | Unclassified | 5.31 | d__Bacteria;p__Actinobacteriota;c__Actinomycetia;o__Actinomycetales;f__Bifidobacteriaceae;g__Bifidobacterium | 99.7 | CRISPR;99.70 iPHoP-RF;73.50 |
| S7-wms_244866 | ME | Duplodnaviria | Heunggongvirae | Uroviricota | Caudoviricetes | n.a. | Unclassified | Unclassified | Unclassified | 18.88 | d__Bacteria;p__Bacteroidota;c__Bacteroidia;o__Bacteroidales;f__Bacteroidaceae;g__Bacteroides | 97.6 | CRISPR;98.30 iPHoP-RF;92.40 |
| S7-wms_80255 | ME | Duplodnaviria | Heunggongvirae | Uroviricota | Caudoviricetes | n.a. | Unclassified | O | O | 74.19 | d__Bacteria;p__Firmicutes;c__Clostridia;o__Oscillospirales;f__Ruminococcaceae;g__Ruminiclostridium_E | 91.5 | blast;93.80 iPHoP-RF;67.00 CRISPR;57.40 |
| S8-wms-combined_134098 | SHHC | n.a. | n.a. | n.a. | n.a. | n.a. | n.a. | n.a. | n.a. | 81.54 | d__Bacteria;p__Firmicutes;c__Clostridia;o__Oscillospirales;f__Oscillospiraceae;g__ER4 | 95.2 | blast;96.60 iPHoP-RF;76.10 |
| S8-wms-combined_146562 | SHHC | Duplodnaviria | Heunggongvirae | Uroviricota | Caudoviricetes | n.a. | Unclassified | O | O | 66.24 | d__Bacteria;p__Firmicutes;c__Clostridia;o__Christensenellales;f__CAG-138;g__UBA1685 | 93 | blast;95.00 iPHoP-RF;82.80 CRISPR;70.40 |
| S8-wms-combined_164289 | SHHC | n.a. | n.a. | n.a. | n.a. | n.a. | n.a. | n.a. | n.a. | 21.14 | d__Bacteria;p__Firmicutes;c__Clostridia;o__Christensenellales;f__CAG-138;g__SFEL01 | 94.5 | blast;96.10 iPHoP-RF;79.00 CRISPR;71.90 |
| S8-wms-combined_169835 | Shared | n.a. | n.a. | n.a. | n.a. | n.a. | n.a. | n.a. | n.a. | 17.94 | d__Bacteria;p__Firmicutes;c__Clostridia;o__Oscillospirales;f__Oscillospiraceae;g__CAG-83 | 95.6 | blast;96.90 iPHoP-RF;64.30 |
| S8-wms-combined_180957 | SHHC | Duplodnaviria | Heunggongvirae | Uroviricota | Caudoviricetes | n.a. | Unclassified | O | O | 99.92 | d__Bacteria;p__Bacteroidota;c__Bacteroidia;o__Bacteroidales;f__Tannerellaceae;g__Parabacteroides | 99.7 | blast;96.70 RaFAH;95.30 iPHoP-RF;84.50 CRISPR;57.90 |
| S8-wms-combined_185611 | Shared | n.a. | n.a. | n.a. | n.a. | n.a. | n.a. | n.a. | n.a. | 9.67 | d__Bacteria;p__Firmicutes;c__Clostridia;o__Christensenellales;f__CAG-74;g__Limiplasma | 94.2 | blast;95.90 iPHoP-RF;76.60 |
| S8-wms-combined_211119 | SHHC | n.a. | n.a. | n.a. | n.a. | n.a. | n.a. | n.a. | n.a. | 22.48 | d__Bacteria;p__Firmicutes;c__Clostridia;o__Christensenellales;f__CAG-138;g__UBA1685 | 95.9 | blast;97.10 iPHoP-RF;77.50 CRISPR;68.50 |
| S8-wms-combined_215102 | Shared | n.a. | n.a. | n.a. | n.a. | n.a. | n.a. | n.a. | n.a. | 12.51 | d__Bacteria;p__Firmicutes;c__Clostridia;o__Christensenellales;f__CAG-74;g__UBA11524 | 95.6 | blast;96.90 iPHoP-RF;94.70 |
| S8-wms-combined_217866 | SHHC | Duplodnaviria | Heunggongvirae | Uroviricota | Caudoviricetes | n.a. | Unclassified | Unclassified | Eponavirus | 98.95 | d__Bacteria;p__Firmicutes;c__Clostridia;o__Lachnospirales;f__Lachnospiraceae;g__Roseburia | 96.3 | CRISPR;97.40 blast;68.80 |
| S8-wms-combined_219796 | SHHC | Duplodnaviria | Heunggongvirae | Uroviricota | Caudoviricetes | n.a. | Unclassified | Unclassified | O | 14.07 | d__Bacteria;p__Firmicutes;c__Clostridia;o__Christensenellales;f__CAG-138;g__UBA1685 | 93.5 | blast;95.40 CRISPR;93.90 iPHoP-RF;88.20 |
| S8-wms-combined_225114 | SHHC | Duplodnaviria | Heunggongvirae | Uroviricota | Caudoviricetes | n.a. | Unclassified | O | O | 99.67 | d__Bacteria;p__Firmicutes;c__Clostridia;o__Oscillospirales;f__Ruminococcaceae;g__Faecalibacterium | 100 | CRISPR;98.40 RaFAH;98.00 blast;92.30 iPHoP-RF;88.50 |
| S8-wms-combined_23583 | Shared | n.a. | n.a. | n.a. | n.a. | n.a. | n.a. | n.a. | n.a. | 28.29 | d__Bacteria;p__Bacteroidota;c__Bacteroidia;o__Bacteroidales;f__Rikenellaceae;g__Alistipes_A | 90.3 | blast;92.80 iPHoP-RF;81.60 CRISPR;68.00 |
| S8-wms-combined_239791 | SHHC | Duplodnaviria | Heunggongvirae | Uroviricota | Caudoviricetes | n.a. | Aggregaviridae | O | O | 19.46 | d__Bacteria;p__Bacteroidota;c__Bacteroidia;o__Bacteroidales;f__Bacteroidaceae;g__Bacteroides | 96.3 | iPHoP-RF;95.10 RaFAH;73.50 |
| S8-wms-combined_246185 | Shared | Duplodnaviria | Heunggongvirae | Uroviricota | Caudoviricetes | n.a. | Unclassified | O | O | 33.37 | d__Bacteria;p__Verrucomicrobiota;c__Verrucomicrobiae;o__Verrucomicrobiales;f__Akkermansiaceae;g__Akkermansia | 98.4 | CRISPR;98.80 blast;96.90 iPHoP-RF;81.10 |
| S8-wms-combined_246389 | SHHC | Duplodnaviria | Heunggongvirae | Uroviricota | Caudoviricetes | n.a. | Unclassified | Unclassified | O | 21.76 | d__Bacteria;p__Firmicutes;c__Clostridia;o__Oscillospirales;f__Oscillospiraceae;g__ER4 | 91.6 | blast;93.90 iPHoP-RF;64.50 |
| S8-wms-combined_25607 | SHHC | Duplodnaviria | Heunggongvirae | Uroviricota | Caudoviricetes | n.a. | Unclassified | O | O | 54.47 | d__Bacteria;p__Firmicutes;c__Clostridia;o__Oscillospirales;f__Oscillospiraceae;g__UBA9475 | 91.2 | blast;93.60 iPHoP-RF;68.90 |
| S8-wms-combined_26087 | Shared | Duplodnaviria | Heunggongvirae | Uroviricota | Caudoviricetes | n.a. | Unclassified | O | O | 100 | d__Bacteria;p__Bacteroidota;c__Bacteroidia;o__Bacteroidales;f__Rikenellaceae;g__Alistipes | 98.7 | blast;96.90 iPHoP-RF;91.40 RaFAH;83.40 |
| S8-wms-combined_264711 | SHHC | n.a. | n.a. | n.a. | n.a. | n.a. | n.a. | n.a. | n.a. | 10.73 | d__Bacteria;p__Firmicutes;c__Clostridia;o__Christensenellales;f__CAG-138;g__SFEL01 | 90.9 | blast;93.30 iPHoP-RF;74.80 |
| S8-wms-combined_270362 | Shared | Duplodnaviria | Heunggongvirae | Uroviricota | Caudoviricetes | n.a. | Unclassified | O | O | 99.95 | d__Bacteria;p__Bacteroidota;c__Bacteroidia;o__Bacteroidales;f__Rikenellaceae;g__Alistipes | 100 | RaFAH;97.30 blast;96.90 iPHoP-RF;90.10 |
| S8-wms-combined_40666 | Shared | n.a. | n.a. | n.a. | n.a. | n.a. | n.a. | n.a. | n.a. | 26.57 | d__Bacteria;p__Firmicutes;c__Clostridia;o__Oscillospirales;f__Oscillospiraceae;g__CAG-103 | 97.2 | CRISPR;98.00 blast;94.30 iPHoP-RF;83.60 |
| S8-wms-combined_49369 | Shared | n.a. | n.a. | n.a. | n.a. | n.a. | n.a. | n.a. | n.a. | 96.98 | d__Bacteria;p__Firmicutes;c__Clostridia;o__Oscillospirales;f__Ruminococcaceae;g__Gemmiger | 94 | blast;95.20 iPHoP-RF;61.70 RaFAH;56.00 |
| S8-wms-combined_68780 | Shared | n.a. | n.a. | n.a. | n.a. | n.a. | n.a. | n.a. | n.a. | 51.97 | d__Bacteria;p__Firmicutes;c__Clostridia;o__Oscillospirales;f__Oscillospiraceae;g__F23-B02 | 94.6 | blast;96.20 iPHoP-RF;58.40 |
| S8-wms-combined_74275 | SHHC | Duplodnaviria | Heunggongvirae | Uroviricota | Caudoviricetes | n.a. | Unclassified | O | O | 54.68 | d__Bacteria;p__Bacteroidota;c__Bacteroidia;o__Bacteroidales;f__Bacteroidaceae;g__Prevotella | 99.3 | blast;96.90 iPHoP-RF;91.40 RaFAH;90.60 |
| S8-wms-combined_81597 | Shared | Duplodnaviria | Heunggongvirae | Uroviricota | Caudoviricetes | n.a. | Unclassified | O | O | 99.93 | d__Bacteria;p__Bacteroidota;c__Bacteroidia;o__Bacteroidales;f__Bacteroidaceae;g__Bacteroides | 100 | RaFAH;99.50 blast;95.10 iPHoP-RF;85.40 |
| S8-wms-combined_97848 | SHHC | Duplodnaviria | Heunggongvirae | Uroviricota | Caudoviricetes | n.a. | Unclassified | Unclassified | Unclassified | 75.58 | d__Bacteria;p__Bacteroidota;c__Bacteroidia;o__Bacteroidales;f__Bacteroidaceae;g__Bacteroides | 98.8 | RaFAH;92.20 blast;92.00 iPHoP-RF;62.20 |
| S8-wms_152831 | SHHC | Duplodnaviria | Heunggongvirae | Uroviricota | Caudoviricetes | n.a. | Unclassified | Unclassified | Toutatisvirus | 99.72 | d__Bacteria;p__Firmicutes;c__Clostridia;o__Oscillospirales;f__Ruminococcaceae;g__Faecalibacterium | 100 | RaFAH;99.50 CRISPR;97.80 blast;95.40 iPHoP-RF;77.30 |
| S8-wms_190781 | SHHC | Duplodnaviria | Heunggongvirae | Uroviricota | Caudoviricetes | n.a. | Unclassified | O | O | 93.96 | d__Bacteria;p__Firmicutes;c__Clostridia;o__Oscillospirales;f__Ruminococcaceae;g__Faecalibacterium | 92 | CRISPR;94.20 blast;94.00 iPHoP-RF;83.30 |
| S8-wms_20528 | SHHC | Duplodnaviria | Heunggongvirae | Uroviricota | Caudoviricetes | n.a. | Unclassified | Unclassified | O | 48.51 | d__Bacteria;p__Firmicutes;c__Clostridia;o__Oscillospirales;f__Ruminococcaceae;g__Gemmiger | 91.6 | blast;93.90 iPHoP-RF;84.50 |
| S8-wms_270 | SHHC | Monodnaviria | Sangervirae | Phixviricota | Malgrandaviricetes | Petitvirales | Microviridae | O | O | | d__Bacteria;p__Bacteroidota;c__Bacteroidia;o__Bacteroidales;f__Barnesiellaceae;g__Barnesiella | 94.6 | blast;96.20 iPHoP-RF;68.20 |
| S8-wms_30498 | Shared | Duplodnaviria | Heunggongvirae | Uroviricota | Caudoviricetes | n.a. | Unclassified | Unclassified | Mushuvirus | 86.77 | d__Bacteria;p__Firmicutes;c__Clostridia;o__Oscillospirales;f__Oscillospiraceae;g__CAG-83 | 92.7 | blast;86.00 RaFAH;81.50 iPHoP-RF;63.00 |
| S8-wms_48935 | Shared | Duplodnaviria | Heunggongvirae | Uroviricota | Caudoviricetes | n.a. | Unclassified | Unclassified | Toutatisvirus | 39.33 | d__Bacteria;p__Firmicutes;c__Clostridia;o__Oscillospirales;f__Oscillospiraceae;g__CAG-83 | 90.9 | blast;93.30 iPHoP-RF;77.30 |
| S8-wms_57121 | SHHC | n.a. | n.a. | n.a. | n.a. | n.a. | n.a. | n.a. | n.a. | 9.54 | d__Bacteria;p__Firmicutes;c__Clostridia;o__Christensenellales;f__CAG-138;g__UBA1685 | 97.3 | blast;98.10 iPHoP-RF;76.60 CRISPR;55.40 |
| S8-wms_58515 | Shared | n.a. | n.a. | n.a. | n.a. | n.a. | n.a. | n.a. | n.a. | 74.23 | d__Bacteria;p__Bacteroidota;c__Bacteroidia;o__Bacteroidales;f__Rikenellaceae;g__Alistipes | 96.3 | iPHoP-RF;97.10 blast;96.70 CRISPR;91.70 RaFAH;55.50 |
| S8-wms_82880 | Shared | Duplodnaviria | Heunggongvirae | Uroviricota | Caudoviricetes | n.a. | Unclassified | Unclassified | Brigitvirus | 30.76 | d__Bacteria;p__Firmicutes;c__Clostridia;o__Oscillospirales;f__Oscillospiraceae;g__An92 | 93 | blast;95.00 iPHoP-RF;60.10 |
| S9-wms-combined_105462 | ME | Duplodnaviria | Heunggongvirae | Uroviricota | Caudoviricetes | n.a. | Unclassified | O | O | 98 | d__Bacteria;p__Firmicutes;c__Clostridia;o__Lachnospirales;f__Lachnospiraceae;g__Anaerostipes | 95.4 | blast;96.80 CRISPR;91.30 iPHoP-RF;91.10 |
| S9-wms-combined_105822 | ME | Duplodnaviria | Heunggongvirae | Uroviricota | Caudoviricetes | n.a. | Unclassified | Unclassified | O | 44.89 | d__Bacteria;p__Firmicutes;c__Clostridia;o__Lachnospirales;f__Lachnospiraceae;g__Enterocloster | 95 | blast;96.50 iPHoP-RF;87.60 CRISPR;52.20 |
| S9-wms-combined_10990 | Shared | n.a. | n.a. | n.a. | n.a. | n.a. | n.a. | n.a. | n.a. | 10.09 | d__Bacteria;p__Firmicutes;c__Clostridia;o__Lachnospirales;f__Lachnospiraceae;g__Blautia_A | 91.4 | blast;93.70 iPHoP-RF;80.00 |
| S9-wms-combined_122742 | Shared | n.a. | n.a. | n.a. | n.a. | n.a. | n.a. | n.a. | n.a. | 8.62 | d__Bacteria;p__Firmicutes;c__Clostridia;o__Oscillospirales;f__Oscillospiraceae;g__Flavonifractor | 90.3 | blast;92.80 iPHoP-RF;72.80 |
| S9-wms-combined_124857 | Shared | n.a. | n.a. | n.a. | n.a. | n.a. | n.a. | n.a. | n.a. | 79.58 | d__Bacteria;p__Firmicutes;c__Clostridia;o__Oscillospirales;f__Oscillospiraceae;g__Flavonifractor | 90.7 | iPHoP-RF;93.10 blast;92.10 CRISPR;71.70 |
| S9-wms-combined_2042 | ME | n.a. | n.a. | n.a. | n.a. | n.a. | n.a. | n.a. | n.a. | 10.88 | d__Bacteria;p__Firmicutes;c__Clostridia;o__Lachnospirales;f__Lachnospiraceae;g__Eisenbergiella | 95.2 | CRISPR;96.60 blast;95.10 iPHoP-RF;93.40 |
| S9-wms-combined_20485 | ME | Duplodnaviria | Heunggongvirae | Uroviricota | Caudoviricetes | n.a. | Unclassified | Unclassified | O | 94.46 | d__Bacteria;p__Firmicutes;c__Bacilli;o__Lactobacillales;f__Streptococcaceae;g__Streptococcus | 100 | RaFAH;99.70 CRISPR;99.00 iPHoP-RF;97.10 blast;96.90 |
| S9-wms-combined_26410 | ME | Duplodnaviria | Heunggongvirae | Uroviricota | Caudoviricetes | n.a. | Unclassified | O | O | 99.06 | d__Bacteria;p__Firmicutes;c__Clostridia;o__Lachnospirales;f__Lachnospiraceae;g__Mediterraneibacter | 95.4 | blast;96.80 |
| S9-wms-combined_27311 | ME | n.a. | n.a. | n.a. | n.a. | n.a. | n.a. | n.a. | n.a. | 4.98 | d__Bacteria;p__Firmicutes;c__Clostridia;o__Oscillospirales;f__Acutalibacteraceae;g__Acutalibacter | 91.4 | blast;93.70 iPHoP-RF;76.10 |
| S9-wms-combined_28805 | ME | Duplodnaviria | Heunggongvirae | Uroviricota | Caudoviricetes | n.a. | Winoviridae | O | O | 94.95 | d__Bacteria;p__Bacteroidota;c__Bacteroidia;o__Bacteroidales;f__Rikenellaceae;g__Alistipes | 99 | blast;96.10 RaFAH;89.50 iPHoP-RF;68.40 |
| S9-wms-combined_31456 | Shared | Duplodnaviria | Heunggongvirae | Uroviricota | Caudoviricetes | n.a. | Unclassified | Unclassified | Unclassified | 63.18 | d__Bacteria;p__Bacteroidota;c__Bacteroidia;o__Bacteroidales;f__Barnesiellaceae;g__Barnesiella | 97.6 | CRISPR;97.40 blast;96.90 iPHoP-RF;83.60 RaFAH;68.00 |
| S9-wms-combined_33229 | ME | Duplodnaviria | Heunggongvirae | Uroviricota | Caudoviricetes | n.a. | Unclassified | O | O | 87.21 | d__Bacteria;p__Bacteroidota;c__Bacteroidia;o__Bacteroidales;f__Bacteroidaceae;g__Bacteroides | 99.1 | iPHoP-RF;97.10 blast;96.10 RaFAH;87.90 |
| S9-wms-combined_33230 | Shared | Duplodnaviria | Heunggongvirae | Uroviricota | Caudoviricetes | n.a. | Unclassified | O | O | 93.11 | d__Bacteria;p__Bacteroidota;c__Bacteroidia;o__Bacteroidales;f__Bacteroidaceae;g__Bacteroides | 100 | RaFAH;99.10 iPHoP-RF;96.40 blast;92.10 |
| S9-wms-combined_3461 | ME | n.a. | n.a. | n.a. | n.a. | n.a. | n.a. | n.a. | n.a. | 69.22 | d__Bacteria;p__Firmicutes;c__Clostridia;o__Lachnospirales;f__Anaerotignaceae;g__Anaerotignum | 95 | CRISPR;96.50 blast;91.10 iPHoP-RF;88.90 |
| S9-wms-combined_36459 | ME | Duplodnaviria | Heunggongvirae | Uroviricota | Caudoviricetes | n.a. | Unclassified | O | O | 25.89 | d__Bacteria;p__Firmicutes;c__Clostridia;o__Oscillospirales;f__Ruminococcaceae;g__Negativibacillus | 96.2 | blast;97.30 iPHoP-RF;85.10 CRISPR;59.60 |
| S9-wms-combined_37587 | Shared | Duplodnaviria | Heunggongvirae | Uroviricota | Caudoviricetes | n.a. | Unclassified | O | O | 92.98 | d__Bacteria;p__Bacteroidota;c__Bacteroidia;o__Bacteroidales;f__Bacteroidaceae;g__Bacteroides | 99.4 | blast;96.80 iPHoP-RF;96.40 RaFAH;92.80 |
| S9-wms-combined_44402 | Shared | Duplodnaviria | Heunggongvirae | Uroviricota | Caudoviricetes | n.a. | Unclassified | O | O | 80.74 | d__Bacteria;p__Bacteroidota;c__Bacteroidia;o__Bacteroidales;f__Bacteroidaceae;g__Bacteroides | 100 | RaFAH;97.80 blast;96.50 iPHoP-RF;96.40 |
| S9-wms-combined_53991 | Shared | Duplodnaviria | Heunggongvirae | Uroviricota | Caudoviricetes | n.a. | Unclassified | O | O | 30.5 | d__Bacteria;p__Firmicutes;c__Negativicutes;o__Veillonellales;f__Dialisteraceae;g__Dialister | 95.4 | blast;96.80 iPHoP-RF;91.80 CRISPR;60.90 |
| S9-wms-combined_5526 | Shared | Duplodnaviria | Heunggongvirae | Uroviricota | Caudoviricetes | n.a. | Unclassified | O | O | 86.27 | d__Bacteria;p__Bacteroidota;c__Bacteroidia;o__Bacteroidales;f__Rikenellaceae;g__Alistipes | 99.3 | blast;96.80 iPHoP-RF;93.10 RaFAH;90.40 |
| S9-wms-combined_64277 | ME | Duplodnaviria | Heunggongvirae | Uroviricota | Caudoviricetes | n.a. | Unclassified | Unclassified | Bcepmuvirus | 98.37 | d__Bacteria;p__Proteobacteria;c__Gammaproteobacteria;o__Enterobacterales;f__Enterobacteriaceae;g__Escherichia | 100 | RaFAH;97.50 blast;96.90 iPHoP-RF;74.30 |
| S9-wms-combined_76274 | Shared | Duplodnaviria | Heunggongvirae | Uroviricota | Caudoviricetes | n.a. | Unclassified | O | O | 91.87 | d__Bacteria;p__Actinobacteriota;c__Actinomycetia;o__Actinomycetales;f__Bifidobacteriaceae;g__Bifidobacterium | 99.4 | CRISPR;98.60 iPHoP-RF;93.70 blast;92.70 RaFAH;81.60 |
| S9-wms-combined_93232 | ME | Duplodnaviria | Heunggongvirae | Uroviricota | Caudoviricetes | n.a. | Unclassified | O | O | 57.26 | d__Bacteria;p__Proteobacteria;c__Gammaproteobacteria;o__Enterobacterales;f__Enterobacteriaceae;g__Klebsiella | 98.2 | CRISPR;98.70 blast;96.90 iPHoP-RF;59.00 |
| S9-wms-combined_97755 | ME | Duplodnaviria | Heunggongvirae | Uroviricota | Caudoviricetes | n.a. | Unclassified | Unclassified | O | 40.71 | d__Bacteria;p__Firmicutes;c__Clostridia;o__Lachnospirales;f__Lachnospiraceae;g__Eisenbergiella | 91 | blast;93.40 iPHoP-RF;57.60 |
| S9-wms-combined_9931 | Shared | Duplodnaviria | Heunggongvirae | Uroviricota | Caudoviricetes | n.a. | Unclassified | Unclassified | O | 57.49 | d__Bacteria;p__Firmicutes;c__Clostridia;o__Oscillospirales;f__Acutalibacteraceae;g__Ruminococcus_E | 95.4 | blast;96.80 iPHoP-RF;92.10 |
| S9-wms_104169 | ME | Monodnaviria | Loebvirae | Hofneiviricota | Faserviricetes | Tubulavirales | Inoviridae | Unclassified | Inovirus | | d__Bacteria;p__Firmicutes;c__Clostridia;o__Oscillospirales;f__Ruminococcaceae;g__Ruthenibacterium | 91.6 | blast;93.90 CRISPR;84.10 iPHoP-RF;79.50 |
| S9-wms_114971 | ME | Duplodnaviria | Heunggongvirae | Uroviricota | Caudoviricetes | n.a. | Unclassified | O | O | 60.7 | d__Bacteria;p__Firmicutes;c__Negativicutes;o__Veillonellales;f__Veillonellaceae;g__Veillonella | 98.4 | blast;95.00 RaFAH;88.20 iPHoP-RF;73.50 CRISPR;55.70 |
| S9-wms_19025 | ME | Duplodnaviria | Heunggongvirae | Uroviricota | Caudoviricetes | n.a. | Unclassified | O | O | 87.64 | d__Bacteria;p__Firmicutes;c__Bacilli;o__Lactobacillales;f__Streptococcaceae;g__Streptococcus | 98.4 | RaFAH;98.80 |
| S9-wms_64472 | Shared | n.a. | n.a. | n.a. | n.a. | n.a. | n.a. | n.a. | n.a. | 4.04 | d__Bacteria;p__Firmicutes;c__Clostridia;o__Lachnospirales;f__Lachnospiraceae;g__Enterocloster | 93.5 | blast;95.40 iPHoP-RF;85.10 |

| Host interactions with: | # Hosts |
| --- | --- |
| ME-viruses-only | 59 |
| SHHC-viruses-only | 26 |
| Shared/communal viruses | 90 |

**VLP_Hosts**

**d__Bacteria;p__Actinobacteriota;c__Actinomycetia;o__Actinomycetales;f__Actinomycetaceae;g__Actinomyces**

**ME**

S14_vf_57689

**Caudoviricetes**

Unclassified

**O**

O

S3_vf_54353

**Caudoviricetes**

Unclassified

**Unclassified**

Cimpunavirus

S3_vf_5525

**n.a.**

n.a.

**n.a.**

n.a.

**Shared**

S3_vf_44405

**Caudoviricetes**

Unclassified

**Unclassified**

Unclassified

**d__Bacteria;p__Actinobacteriota;c__Actinomycetia;o__Actinomycetales;f__Actinomycetaceae;g__Pauljensenia**

**ME**

S11_vf_66522

**Caudoviricetes**

Unclassified

**Unclassified**

Unclassified

**Shared**

S14_vf_117155

**Caudoviricetes**

Unclassified

**O**

O

S16_vf_16182

**Caudoviricetes**

Unclassified

**O**

O

**d__Bacteria;p__Actinobacteriota;c__Actinomycetia;o__Actinomycetales;f__Bifidobacteriaceae;g__Bifidobacterium**

**ME**

S11_vf_58587

**n.a.**

n.a.

**n.a.**

n.a.

S13_vs_113593

**Caudoviricetes**

Unclassified
**Unclassified**
Unclassified
S13_vs_149860
**Caudoviricetes**
Unclassified
**Unclassified**
O
S3_vf_127408
**Caudoviricetes**
Unclassified
**O**
O
S3_vs_39940
**n.a.**
n.a.
**n.a.**
n.a.
S7_vf_5843
**Caudoviricetes**
Unclassified
**O**
O
S7_vs_73280
**Caudoviricetes**
Unclassified
**Unclassified**
Minunavirus
**Shared**
S16_vf_100473
**n.a.**
n.a.
**n.a.**
n.a.
S16_vs_78361
**Caudoviricetes**
Unclassified
**O**
O
S3_vf_70746
**Caudoviricetes**
Unclassified
**Unclassified**
Unclassified
S3_vf_7137
**n.a.**
Unclassified
**Unclassified**
O
S3_vs_123834
**Caudoviricetes**

Unclassified
**O**
O
S4_vf_30103
**n.a.**
Unclassified
**Unclassified**
O
S4_vs_VIRSorter_16493-cat_2
**Caudoviricetes**
Unclassified
**Unclassified**
Unclassified
S5_vs_63924
**Caudoviricetes**
Unclassified
**Unclassified**
Unclassified
S8_vf_153696
**n.a.**
n.a.
**n.a.**
n.a.
S8_vf_197061
**Caudoviricetes**
Unclassified
**Unclassified**
Unclassified
**SHHC**
S16_vf_172897
**Caudoviricetes**
Unclassified
**O**
O
S16_vf_173109
**Caudoviricetes**
Unclassified
**Unclassified**
Unclassified
S4_vf_12624
**Caudoviricetes**
Unclassified
**O**
O
S4_vs_VIRSorter_2744-cat_3
**Caudoviricetes**
Unclassified
**O**
O
S4_vs_VIRSorter_5698-cat_2
**Caudoviricetes**

Unclassified
**O**
O
**d__Bacteria;p__Actinobacteriota;c__Actinomycetia;o__Actinomycetales;f__Micrococcaceae;g__Rothia**
**SHHC**
S8_vf_102508
**n.a.**
n.a.
**n.a.**
n.a.
S8_vf_7243
**n.a.**
n.a.
**n.a.**
n.a.
**d__Bacteria;p__Actinobacteriota;c__Actinomycetia;o__Mycobacteriales;f__Mycobacteriaceae;g__Mycobacterium**
**ME**
S9_vf_934
**Caudoviricetes**
Unclassified
**O**
O
**d__Bacteria;p__Actinobacteriota;c__Coriobacteriia;o__Coriobacteriales;f__Coriobacteriaceae;g__Collinsella**
**ME**
S5_vf_29332
**Caudoviricetes**
Unclassified
**O**
O
S7_vs_55526
**n.a.**
n.a.
**n.a.**
n.a.
**Shared**
S3_vf_99626
**Caudoviricetes**
Unclassified
**O**
O
S5_vf_137260
**n.a.**
Unclassified
**O**
O
**SHHC**
S12_vf_170614
**Caudoviricetes**
Salasmaviridae
**O**
O

S8_vf_142700
**n.a.**
Unclassified
**O**
O
**d__Bacteria;p__Actinobacteriota;c__Coriobacteriia;o__Coriobacteriales;f__Eggerthellaceae;g__Adlercreutzia**
**Shared**
S12_vf_173666
**n.a.**
n.a.
**n.a.**
n.a.
**d__Bacteria;p__Actinobacteriota;c__Coriobacteriia;o__Coriobacteriales;f__Eggerthellaceae;g__Eggerthella**
**ME**
S7_vf_67910
**n.a.**
n.a.
**n.a.**
n.a.
**Shared**
S2_vf_16829
**Caudoviricetes**
Unclassified
**Unclassified**
O
**d__Bacteria;p__Actinobacteriota;c__Coriobacteriia;o__Coriobacteriales;f__Eggerthellaceae;g__Gordonibacter**
**ME**
S3_vs_50557
**Caudoviricetes**
Unclassified
**O**
O
**SHHC**
S8_vf_195032
**Caudoviricetes**
Unclassified
**O**
O
**d__Bacteria;p__Bacteroidota;c__Bacteroidia;o__Bacteroidales;f__Bacteroidaceae;g__Bacteroides**
**ME**
S11_vf_104033
**Caudoviricetes**
Unclassified
**O**
O
S11_vf_43997
**Caudoviricetes**
Unclassified
**Unclassified**
Unclassified
S11_vs_23188

**n.a.**
n.a.
**n.a.**
n.a.
S11_vs_84848
**Caudoviricetes**
Unclassified
**Unclassified**
Unclassified
S13_vf_66508
**Caudoviricetes**
Unclassified
**O**
O
S14_vf_117162
**Caudoviricetes**
Unclassified
**Unclassified**
O
S17_vs_116640
**Caudoviricetes**
Unclassified
**O**
O
S17_vs_160502
**Caudoviricetes**
Steigviridae
**O**
O
S17_vs_39515
**Caudoviricetes**
Intestiviridae
**O**
O
S3_vf_18080
**n.a.**
n.a.
**n.a.**
n.a.
S3_vf_18829
**Caudoviricetes**
Unclassified
**O**
O
S3_vf_73597
**Caudoviricetes**
Unclassified
**O**
O
S3_vs_23194
**Caudoviricetes**

Duneviridae
**O**
O
S3_vs_40939
**n.a.**
n.a.
**n.a.**
n.a.
S3_vs_45147
**n.a.**
n.a.
**n.a.**
n.a.
S5_vf_8965
**Caudoviricetes**
Unclassified
**O**
O
S5_vs_72532
**Caudoviricetes**
Unclassified
**O**
O
S7_vf_27309
**Caudoviricetes**
Unclassified
**Unclassified**
O
S7_vs_135828
**Malgrandaviricetes**
Microviridae
**Unclassified**
Unclassified
S7_vs_2599
**n.a.**
n.a.
**n.a.**
n.a.
S7_vs_557
**Caudoviricetes**
Unclassified
**O**
O
S7_vs_944
**n.a.**
n.a.
**n.a.**
n.a.
S9_vs_83108
**n.a.**
n.a.

**n.a.**

n.a.

- **Shared**
  - S10_vs_14557
    - **Caudoviricetes**
      - Duneviridae
        - **O**
          - O
  - S10_vs_35854
    - **Caudoviricetes**
      - Unclassified
        - **O**
          - O
  - S10_vs_36598
    - **n.a.**
      - n.a.
        - **n.a.**
          - n.a.
  - S10_vs_58014
    - **Caudoviricetes**
      - Unclassified
        - **Unclassified**
          - Unclassified
  - S10_vs_62776
    - **Caudoviricetes**
      - Unclassified
        - **O**
          - O
  - S10_vs_71900
    - **n.a.**
      - n.a.
        - **n.a.**
          - n.a.
  - S11_vs_3277
    - **Caudoviricetes**
      - Unclassified
        - **Unclassified**
          - Unclassified
  - S11_vs_45658
    - **Caudoviricetes**
      - Unclassified
        - **O**
          - O
  - S11_vs_58930
    - **Caudoviricetes**
      - Unclassified
        - **Unclassified**
          - Unclassified
  - S12_vf_16374
    - **Caudoviricetes**
      - Unclassified

**O**
O
S12_vs_141337
**Caudoviricetes**
Unclassified
**Unclassified**
Unclassified
S12_vs_151292
**Caudoviricetes**
Unclassified
**Unclassified**
Unclassified
S12_vs_1980
**Caudoviricetes**
Unclassified
**O**
O
S12_vs_26336
**n.a.**
n.a.
**n.a.**
n.a.
S12_vs_99219
**n.a.**
n.a.
**n.a.**
n.a.
S14_vs_112345
**n.a.**
n.a.
**n.a.**
n.a.
S14_vs_11510
**n.a.**
n.a.
**n.a.**
n.a.
S14_vs_25852
**n.a.**
n.a.
**n.a.**
n.a.
S14_vs_32499
**Caudoviricetes**
Unclassified
**O**
O
S14_vs_66208
**Caudoviricetes**
Unclassified
**O**

O
S14_vs_70985
**n.a.**
n.a.
**n.a.**
n.a.
S14_vs_74067
**n.a.**
n.a.
**n.a.**
n.a.
S15_vf_2032
**Caudoviricetes**
Unclassified
**O**
O
S15_vf_75924
**Caudoviricetes**
Steigviridae
**O**
O
S15_vs_27510
**n.a.**
n.a.
**n.a.**
n.a.
S15_vs_35445
**n.a.**
n.a.
**n.a.**
n.a.
S15_vs_3868
**Caudoviricetes**
Unclassified
**O**
O
S15_vs_42043
**n.a.**
n.a.
**n.a.**
n.a.
S15_vs_43913
**Caudoviricetes**
Unclassified
**O**
O
S16_vs_127669
**Caudoviricetes**
Unclassified
**Unclassified**
Unclassified

S16_vs_127905
**n.a.**
n.a.
**n.a.**
n.a.
S16_vs_136935
**n.a.**
n.a.
**n.a.**
n.a.
S16_vs_80244
**n.a.**
n.a.
**n.a.**
n.a.
S17_vf_128147
**Malgrandaviricetes**
Microviridae
**Unclassified**
O
S17_vf_140653
**Caudoviricetes**
Unclassified
**O**
O
S17_vs_117507
**n.a.**
n.a.
**n.a.**
n.a.
S17_vs_2079
**Caudoviricetes**
Unclassified
**Unclassified**
Unclassified
S2_vs_10289
**Caudoviricetes**
Unclassified
**Unclassified**
Unclassified
S3_vf_127399
**Caudoviricetes**
Unclassified
**O**
O
S3_vs_119660
**Caudoviricetes**
Unclassified
**Unclassified**
Unclassified
S3_vs_39774

**n.a.**
n.a.
**n.a.**
n.a.
S3_vs_64424
**n.a.**
n.a.
**n.a.**
n.a.
S3_vs_70508
**n.a.**
n.a.
**n.a.**
n.a.
S3_vs_9158
**n.a.**
n.a.
**n.a.**
n.a.
S4_vs_VIRSorter_31975-circular-cat_1
**Caudoviricetes**
Unclassified
**O**
O
S5_vs_109527
**n.a.**
n.a.
**n.a.**
n.a.
S5_vs_129970
**n.a.**
n.a.
**n.a.**
n.a.
S5_vs_131510
**Caudoviricetes**
Unclassified
**Unclassified**
O
S5_vs_142195
**n.a.**
n.a.
**n.a.**
n.a.
S5_vs_80496
**n.a.**
n.a.
**n.a.**
n.a.
S6_vf_56486
**Caudoviricetes**

Unclassified
**O**
O
S6_vs_46039
**n.a.**
n.a.
**n.a.**
n.a.
S6_vs_9784
**n.a.**
n.a.
**n.a.**
n.a.
S7_vf_37279
**Caudoviricetes**
Unclassified
**O**
O
S7_vf_5801
**Caudoviricetes**
Unclassified
**O**
O
S7_vs_11081
**Caudoviricetes**
Unclassified
**Unclassified**
Unclassified
S8_vf_208120
**Caudoviricetes**
Unclassified
**O**
O
S8_vf_28956
**n.a.**
n.a.
**n.a.**
n.a.
S8_vs_147697
**n.a.**
n.a.
**n.a.**
n.a.
S8_vs_28311
**Caudoviricetes**
Duneviridae
**O**
O
S8_vs_72117
**n.a.**
n.a.

**n.a.**
n.a.
S8_vs_99362
**Caudoviricetes**
Unclassified
**Unclassified**
Unclassified
S9_vs_28197
**Caudoviricetes**
Unclassified
**O**
O
**SHHC**
S10_vf_43964
**Caudoviricetes**
Unclassified
**O**
O
S10_vs_106834
**Caudoviricetes**
Unclassified
**O**
O
S12_vf_60224
**Caudoviricetes**
Unclassified
**O**
O
S16_vf_87190
**Caudoviricetes**
Unclassified
**O**
O
**d__Bacteria;p__Bacteroidota;c__Bacteroidia;o__Bacteroidales;f__Bacteroidaceae;g__Phocaeicola**
**ME**
S14_vf_37500
**Malgrandaviricetes**
Microviridae
**Unclassified**
Unclassified
**d__Bacteria;p__Bacteroidota;c__Bacteroidia;o__Bacteroidales;f__Bacteroidaceae;g__Prevotella**
**ME**
S3_vf_127450
**n.a.**
n.a.
**n.a.**
n.a.
**Shared**
S11_vf_104035
**Caudoviricetes**
Suoliviridae

**O**
O
S13_vf_11246
**Caudoviricetes**
Suoliviridae
**O**
O
S13_vs_57267
**Caudoviricetes**
Unclassified
**O**
O
S5_vf_145579
**Caudoviricetes**
Steigviridae
**O**
O
S8_vf_117796
**Caudoviricetes**
Unclassified
**Unclassified**
Unclassified
S8_vf_122206
**Caudoviricetes**
Unclassified
**O**
O
S8_vf_166677
**Caudoviricetes**
Intestiviridae
**Obtuvirinae**
Fohxhuevirus
S8_vf_183045
**Caudoviricetes**
Unclassified
**Unclassified**
Unclassified
S8_vf_79769
**n.a.**
n.a.
**n.a.**
n.a.
S9_vf_13507
**n.a.**
n.a.
**n.a.**
n.a.
S9_vf_51656
**Caudoviricetes**
Intestiviridae
**Obtuvirinae**

Fohxhuevirus
S9_vf_76769
**Caudoviricetes**
Suoliviridae
**Loutivirinae**
Buchavirus
**SHHC**
S4_vf_18803
**Caudoviricetes**
Unclassified
**Unclassified**
Unclassified
S4_vf_20440
**Caudoviricetes**
Unclassified
**O**
O
S4_vf_31981
**Caudoviricetes**
Suoliviridae
**Uncouvirinae**
Besingivirus
S4_vf_4891
**n.a.**
n.a.
**n.a.**
n.a.
S8_vs_148389
**Caudoviricetes**
Unclassified
**O**
O
**d__Bacteria;p__Bacteroidota;c__Bacteroidia;o__Bacteroidales;f__Bacteroidaceae;g__Prevotellamassilia**
**SHHC**
S12_vf_69136
**Caudoviricetes**
Unclassified
**O**
O
**d__Bacteria;p__Bacteroidota;c__Bacteroidia;o__Bacteroidales;f__Barnesiellaceae;g__Barnesiella**
**ME**
S17_vf_35138
**Caudoviricetes**
Unclassified
**O**
O
S9_vs_21850
**Caudoviricetes**
Unclassified
**Unclassified**
Unclassified

**Shared**
S13_vs_90967
**Malgrandaviricetes**
Microviridae
**O**
O
S8_vs_4153
**n.a.**
n.a.
**n.a.**
n.a.
**SHHC**
S12_vf_188846
**Malgrandaviricetes**
Microviridae
**O**
O
S4_vf_12614
**n.a.**
n.a.
**n.a.**
n.a.
S8_vf_208115
**Malgrandaviricetes**
Microviridae
**O**
O

**d__Bacteria;p__Bacteroidota;c__Bacteroidia;o__Bacteroidales;f__Coprobacteraceae;g__Coprobacter**

**ME**
S7_vs_51332
**n.a.**
n.a.
**n.a.**
n.a.
**SHHC**
S15_vs_18274
**n.a.**
n.a.
**n.a.**
n.a.

**d__Bacteria;p__Bacteroidota;c__Bacteroidia;o__Bacteroidales;f__Marinifilaceae;g__Butyricimonas**

**Shared**
S12_vs_71525
**n.a.**
n.a.
**n.a.**
n.a.
S12_vs_74280
**n.a.**
n.a.
**n.a.**

n.a.
S3_vs_6604
**Caudoviricetes**
Duneviridae
**O**
O
**d__Bacteria;p__Bacteroidota;c__Bacteroidia;o__Bacteroidales;f__Marinifilaceae;g__Odoribacter**
**ME**
S14_vs_78164
**Caudoviricetes**
Winoviridae
**O**
O
S3_vs_97827
**n.a.**
n.a.
**n.a.**
n.a.
**Shared**
S3_vs_117894
**n.a.**
n.a.
**n.a.**
n.a.
S4_vs_VIRSorter_9913-cat_1
**Caudoviricetes**
Unclassified
**Unclassified**
Unclassified
**d__Bacteria;p__Bacteroidota;c__Bacteroidia;o__Bacteroidales;f__Muribaculaceae;g__UBA7173**
**SHHC**
S12_vs_111286
**n.a.**
n.a.
**n.a.**
n.a.
**d__Bacteria;p__Bacteroidota;c__Bacteroidia;o__Bacteroidales;f__P3;g__Phil12**
**ME**
S3_vs_127442
**Malgrandaviricetes**
Microviridae
**Unclassified**
Unclassified
**d__Bacteria;p__Bacteroidota;c__Bacteroidia;o__Bacteroidales;f__Rikenellaceae;g__Alistipes**
**ME**
S1_vf_159951
**Caudoviricetes**
Unclassified
**O**
O
S11_vf_104071

**Caudoviricetes**
Unclassified
**O**
O
S11_vs_36748
**n.a.**
n.a.
**n.a.**
n.a.
S11_vs_98193
**Caudoviricetes**
Unclassified
**Unclassified**
Unclassified
S13_vf_56074
**Caudoviricetes**
Unclassified
**Unclassified**
Unclassified
S17_vs_177196
**n.a.**
n.a.
**n.a.**
n.a.
S3_vf_127406
**Caudoviricetes**
Unclassified
**Unclassified**
Unclassified
S3_vs_127413
**Caudoviricetes**
Unclassified
**O**
O
S3_vs_77156
**Malgrandaviricetes**
Microviridae
**Unclassified**
Unclassified
S5_vs_1338
**n.a.**
n.a.
**n.a.**
n.a.
S7_vs_54322
**n.a.**
n.a.
**n.a.**
n.a.
**Shared**
S10_vs_37694

**Caudoviricetes**
Duneviridae
**O**
O
S11_vf_70015
**Caudoviricetes**
Unclassified
**O**
O
S11_vs_1937
**Caudoviricetes**
Unclassified
**O**
O
S12_vf_174374
**n.a.**
n.a.
**n.a.**
n.a.
S12_vs_142625
**Caudoviricetes**
Unclassified
**O**
O
S13_vs_83447
**n.a.**
n.a.
**n.a.**
n.a.
S15_vs_71968
**Caudoviricetes**
Unclassified
**O**
O
S16_vf_9270
**n.a.**
n.a.
**n.a.**
n.a.
S16_vs_19968
**n.a.**
n.a.
**n.a.**
n.a.
S16_vs_32764
**n.a.**
n.a.
**n.a.**
n.a.
S16_vs_33281
**n.a.**

n.a.
**n.a.**
n.a.
S17_vs_157209
**n.a.**
n.a.
**n.a.**
n.a.
S3_vf_67828
**Malgrandaviricetes**
Microviridae
**Unclassified**
Unclassified
S3_vs_17168
**Caudoviricetes**
Unclassified
**O**
O
S3_vs_56292
**Caudoviricetes**
Unclassified
**O**
O
S3_vs_89485
**n.a.**
n.a.
**n.a.**
n.a.
S5_vf_18461
**Caudoviricetes**
Unclassified
**O**
O
S5_vs_123256
**Caudoviricetes**
Unclassified
**Unclassified**
Unclassified
S5_vs_20500
**Caudoviricetes**
Unclassified
**O**
O
S5_vs_76500
**Caudoviricetes**
Unclassified
**Unclassified**
Unclassified
S5_vs_79992
**Caudoviricetes**
Winoviridae

**O**
O
S5_vs_93839
**Caudoviricetes**
Unclassified
**O**
O
S7_vf_117697
**n.a.**
n.a.
**n.a.**
n.a.
S7_vs_119679
**Caudoviricetes**
Winoviridae
**O**
O
S7_vs_138173
**Caudoviricetes**
Unclassified
**O**
O
S8_vs_179208
**n.a.**
n.a.
**n.a.**
n.a.
S8_vs_40014
**n.a.**
n.a.
**n.a.**
n.a.
S8_vs_9987
**Caudoviricetes**
Unclassified
**O**
O
S9_vs_2799
**n.a.**
n.a.
**n.a.**
n.a.
S9_vs_48424
**n.a.**
n.a.
**n.a.**
n.a.
**SHHC**
S10_vs_67471
**n.a.**
n.a.

**n.a.**
n.a.
S12_vs_121913
**n.a.**
n.a.
**n.a.**
n.a.
S12_vs_23796
**Caudoviricetes**
Unclassified
**Unclassified**
Unclassified
S15_vf_20644
**Caudoviricetes**
Unclassified
**Unclassified**
Unclassified
S16_vs_51901
**n.a.**
n.a.
**n.a.**
n.a.
**d__Bacteria;p__Bacteroidota;c__Bacteroidia;o__Bacteroidales;f__Rikenellaceae;g__Alistipes_A**
**ME**
S7_vf_134023
**n.a.**
n.a.
**n.a.**
n.a.
**Shared**
S12_vf_145044
**n.a.**
n.a.
**n.a.**
n.a.
S16_vs_142561
**Caudoviricetes**
Unclassified
**O**
O
S6_vs_32886
**n.a.**
n.a.
**n.a.**
n.a.
S7_vs_50194
**n.a.**
n.a.
**n.a.**
n.a.
**SHHC**

S8_vs_9114
**n.a.**
n.a.
**n.a.**
n.a.
**d__Bacteria;p__Bacteroidota;c__Bacteroidia;o__Bacteroidales;f__Tannerellaceae;g__Marseille-P3160**
**Shared**
S13_vf_87873
**Caudoviricetes**
Unclassified
**Unclassified**
Unclassified
S4_vs_VIRSorter_6022-circular-cat_1
**Caudoviricetes**
Unclassified
**Unclassified**
Unclassified
**d__Bacteria;p__Bacteroidota;c__Bacteroidia;o__Bacteroidales;f__Tannerellaceae;g__Parabacteroides**
**ME**
S11_vf_68693
**n.a.**
n.a.
**n.a.**
n.a.
S7_vs_55031
**n.a.**
n.a.
**n.a.**
n.a.
**Shared**
S10_vf_18671
**Caudoviricetes**
Unclassified
**Unclassified**
Unclassified
S10_vs_37379
**n.a.**
n.a.
**n.a.**
n.a.
S12_vs_138261
**n.a.**
n.a.
**n.a.**
n.a.
S13_vf_143422
**Caudoviricetes**
Suoliviridae
**Boorivirinae**
Canhaevirus
S13_vs_15645

**Caudoviricetes**
Unclassified
**O**
O
S14_vs_15228
**Caudoviricetes**
Unclassified
**O**
O
S17_vf_81232
**Caudoviricetes**
Suoliviridae
**Boorivirinae**
Canhaevirus
S17_vs_43307
**n.a.**
n.a.
**n.a.**
n.a.
S2_vs_7306
**Caudoviricetes**
Unclassified
**Unclassified**
Unclassified
S3_vs_80151
**Caudoviricetes**
Unclassified
**O**
O
S3_vs_82895
**Caudoviricetes**
Unclassified
**O**
O
S4_vf_4838
**Caudoviricetes**
Suoliviridae
**Boorivirinae**
Cohcovirus
S4_vs_VIRSorter_13190-cat_2
**Caudoviricetes**
Unclassified
**O**
O
S5_vs_1572
**Caudoviricetes**
Unclassified
**Unclassified**
Unclassified
S6_vs_15390
**Caudoviricetes**

Unclassified
**O**
O
S7_vf_28429
**Caudoviricetes**
Unclassified
**O**
O
S7_vs_137915
**Caudoviricetes**
Unclassified
**O**
O
S8_vf_158225
**Caudoviricetes**
Intestiviridae
**Crudevirinae**
Diorhovirus
S8_vs_182288
**Caudoviricetes**
Unclassified
**O**
O
**SHHC**
S8_vf_16172
**n.a.**
n.a.
**n.a.**
n.a.
S8_vs_126582
**Caudoviricetes**
Unclassified
**O**
O
**d__Bacteria;p__Bacteroidota;c__Bacteroidia;o__Bacteroidales;f__UBA11471;g__UBA11471**
**ME**
S11_vs_79831
**n.a.**
n.a.
**n.a.**
n.a.
**d__Bacteria;p__Cyanobacteria;c__Vampirovibrionia;o__Gastranaerophilales;f__Gastranaerophilaceae;g__CAG-306**
**ME**
S11_vs_104041
**Caudoviricetes**
Unclassified
**O**
O
**d__Bacteria;p__Cyanobacteria;c__Vampirovibrionia;o__Gastranaerophilales;f__Gastranaerophilaceae;g__QAMI01**
**ME**
S11_vs_82331

**Caudoviricetes**
Unclassified
**O**
O
S3_vs_50924
**n.a.**
n.a.
**n.a.**
n.a.
**d__Bacteria;p__Desulfobacterota;c__Desulfovibrionia;o__Desulfovibrionales;f__Desulfovibrionaceae;g__Bilophila**
**ME**
S17_vs_177203
**Caudoviricetes**
Unclassified
**O**
O
**Shared**
S8_vf_208111
**n.a.**
n.a.
**n.a.**
n.a.
**d__Bacteria;p__Desulfobacterota;c__Desulfovibrionia;o__Desulfovibrionales;f__Desulfovibrionaceae;g__Mailhella**
**SHHC**
S8_vs_208114
**Caudoviricetes**
Unclassified
**O**
O
**d__Bacteria;p__Firmicutes;c__Bacilli;o__Erysipelotrichales;f__Erysipelatoclostridiaceae;g__Catenibacterium**
**Shared**
S8_vs_59736
**Caudoviricetes**
Unclassified
**O**
O
**d__Bacteria;p__Firmicutes;c__Bacilli;o__Erysipelotrichales;f__Erysipelatoclostridiaceae;g__Erysipelatoclostridium**
**ME**
S11_vs_68348
**Caudoviricetes**
Unclassified
**O**
O
S7_vs_13041
**Caudoviricetes**
Unclassified
**O**
O
S9_vs_46498
**Caudoviricetes**
Unclassified

**O**
O
**d__Bacteria;p__Firmicutes;c__Bacilli;o__Erysipelotrichales;f__Erysipelatoclostridiaceae;g__Faecalibacillus**
**ME**
S17_vs_31654
**Caudoviricetes**
Unclassified
**O**
O
S3_vs_94577
**Caudoviricetes**
Unclassified
**O**
O
**d__Bacteria;p__Firmicutes;c__Bacilli;o__Erysipelotrichales;f__Erysipelatoclostridiaceae;g__Massiliomicrobiota**
**ME**
S11_vs_55144
**n.a.**
n.a.
**n.a.**
n.a.
S7_vf_38697
**Caudoviricetes**
Unclassified
**O**
O
**d__Bacteria;p__Firmicutes;c__Bacilli;o__Erysipelotrichales;f__Erysipelotrichaceae;g__Clostridium_AQ**
**ME**
S5_vs_122618
**n.a.**
n.a.
**n.a.**
n.a.
S9_vs_90846
**Caudoviricetes**
Unclassified
**O**
O
**Shared**
S11_vf_92391
**n.a.**
n.a.
**n.a.**
n.a.
**SHHC**
S2_vf_20910
**n.a.**
n.a.
**n.a.**
n.a.
**d__Bacteria;p__Firmicutes;c__Bacilli;o__Erysipelotrichales;f__Erysipelotrichaceae;g__Holdemanella**

**SHHC**
S4_vf_10506
**n.a.**
n.a.
**n.a.**
n.a.
**d__Bacteria;p__Firmicutes;c__Bacilli;o__Erysipelotrichales;f__Erysipelotrichaceae;g__Holdemania**
**ME**
S11_vs_14882
**n.a.**
n.a.
**n.a.**
n.a.
S14_vf_72335
**Caudoviricetes**
Unclassified
**O**
O
S5_vs_51416
**Caudoviricetes**
Unclassified
**O**
O
S9_vs_37437
**Caudoviricetes**
Unclassified
**O**
O
**Shared**
S6_vs_56485
**Caudoviricetes**
Unclassified
**O**
O
**d__Bacteria;p__Firmicutes;c__Bacilli;o__Haloplasmatales_A;f__Turicibacteraceae;g__Turicibacter**
**ME**
S3_vf_106401
**Caudoviricetes**
Unclassified
**O**
O
S5_vf_28061
**n.a.**
n.a.
**n.a.**
n.a.
**d__Bacteria;p__Firmicutes;c__Bacilli;o__Lactobacillales;f__Lactobacillaceae;g__Lacticaseibacillus**
**ME**
S14_vf_72953
**Caudoviricetes**
Unclassified

**Unclassified**
Unclassified
**d__Bacteria;p__Firmicutes;c__Bacilli;o__Lactobacillales;f__Lactobacillaceae;g__Lactobacillus**
**ME**
S14_vf_106156
**Caudoviricetes**
Unclassified
**Unclassified**
Unclassified
S14_vf_78139
**Caudoviricetes**
Unclassified
**Unclassified**
Unclassified
S5_vs_6478
**Caudoviricetes**
Unclassified
**Unclassified**
O
**d__Bacteria;p__Firmicutes;c__Bacilli;o__Lactobacillales;f__Lactobacillaceae;g__Leuconostoc**
**ME**
S7_vf_63894
**Caudoviricetes**
Unclassified
**Mccleskeyvirinae**
Unaquatrovirus
**SHHC**
S8_vf_129423
**Caudoviricetes**
Unclassified
**Mccleskeyvirinae**
Unaquatrovirus
**d__Bacteria;p__Firmicutes;c__Bacilli;o__Lactobacillales;f__Lactobacillaceae;g__Limosilactobacillus**
**ME**
S5_vs_89540
**Caudoviricetes**
Unclassified
**O**
O
**d__Bacteria;p__Firmicutes;c__Bacilli;o__Lactobacillales;f__Streptococcaceae;g__Lactococcus**
**ME**
S7_vf_68239
**Caudoviricetes**
Unclassified
**O**
O
**Shared**
S3_vf_23042
**Caudoviricetes**
Unclassified
**O**

O
S5_vf_25423
**Caudoviricetes**
Unclassified
**Unclassified**
Skunavirus
S5_vf_55687
**Caudoviricetes**
Unclassified
**O**
O
S7_vf_29941
**Caudoviricetes**
Unclassified
**O**
O
S7_vf_89197
**Caudoviricetes**
Unclassified
**Unclassified**
Ceduovirus
S8_vf_45776
**Caudoviricetes**
Unclassified
**Unclassified**
Skunavirus

**d__Bacteria;p__Firmicutes;c__Bacilli;o__Lactobacillales;f__Streptococcaceae;g__Streptococcus**

**ME**
S5_vs_60507
**Caudoviricetes**
Unclassified
**Unclassified**
O
S7_vf_138069
**n.a.**
n.a.
**n.a.**
n.a.
S7_vf_37293
**Caudoviricetes**
Unclassified
**O**
O
S7_vf_49007
**Caudoviricetes**
Unclassified
**O**
O
S9_vf_15012
**Caudoviricetes**
Unclassified

**Unclassified**
Unclassified
S9_vf_19284
**n.a.**
n.a.
**n.a.**
n.a.
S9_vf_4115
**Caudoviricetes**
Unclassified
**Unclassified**
Unclassified
S9_vf_90856
**Caudoviricetes**
Unclassified
**Unclassified**
O
S9_vf_90866
**n.a.**
Unclassified
**Unclassified**
Unclassified
**Shared**
S10_vf_64371
**n.a.**
n.a.
**n.a.**
n.a.
S11_vf_27845
**n.a.**
n.a.
**n.a.**
n.a.
S14_vf_117157
**n.a.**
n.a.
**n.a.**
n.a.
**SHHC**
S8_vf_134943
**n.a.**
n.a.
**n.a.**
n.a.
**d__Bacteria;p__Firmicutes;c__Bacilli;o__ML615J-28;f__CAG-313;g__CAG-313**
**Shared**
S13_vs_30101
**Caudoviricetes**
Unclassified
**O**
O

S16_vs_141218
**n.a.**
n.a.
**n.a.**
n.a.
**d__Bacteria;p__Firmicutes;c__Bacilli;o__RF39;f__UBA660;g__CAG-302**
**SHHC**
S12_vs_123620
**n.a.**
n.a.
**n.a.**
n.a.
S16_vs_165662
**Caudoviricetes**
Unclassified
**O**
O
**d__Bacteria;p__Firmicutes;c__Bacilli;o__RF39;f__UBA660;g__CAG-460**
**Shared**
S13_vs_26756
**n.a.**
n.a.
**n.a.**
n.a.
S13_vs_67429
**n.a.**
n.a.
**n.a.**
n.a.
**SHHC**
S12_vf_105542
**Caudoviricetes**
Unclassified
**O**
O
**d__Bacteria;p__Firmicutes;c__Bacilli;o__RF39;f__UBA660;g__CAG-877**
**SHHC**
S12_vs_172379
**n.a.**
n.a.
**n.a.**
n.a.
**d__Bacteria;p__Firmicutes;c__Clostridia;o__Acetivibrionales;f__Acetivibrionaceae;g__Ruminiclostridium**
**ME**
S14_vs_102698
**Caudoviricetes**
Unclassified
**O**
O
**d__Bacteria;p__Firmicutes;c__Clostridia;o__Christensenellales;f__Borkfalkiaceae;g__QALS01**
**SHHC**

S10_vs_120489
**n.a.**
n.a.
**n.a.**
n.a.
**d__Bacteria;p__Firmicutes;c__Clostridia;o__Christensenellales;f__CAG-138;g__SFEL01**
**ME**
S3_vs_73361
**n.a.**
n.a.
**n.a.**
n.a.
**SHHC**
S8_vs_176604
**n.a.**
n.a.
**n.a.**
n.a.
**d__Bacteria;p__Firmicutes;c__Clostridia;o__Christensenellales;f__CAG-138;g__UBA1685**
**SHHC**
S8_vf_93976
**n.a.**
n.a.
**n.a.**
n.a.
**d__Bacteria;p__Firmicutes;c__Clostridia;o__Christensenellales;f__CAG-138;g__UBA5394**
**SHHC**
S16_vs_4662
**Caudoviricetes**
Unclassified
**Unclassified**
Unclassified
**d__Bacteria;p__Firmicutes;c__Clostridia;o__Christensenellales;f__CAG-74;g__Alectryocaccomicrobium**
**ME**
S5_vs_10428
**Caudoviricetes**
Unclassified
**O**
O
**d__Bacteria;p__Firmicutes;c__Clostridia;o__Christensenellales;f__CAG-74;g__Firm-11**
**ME**
S7_vs_129858
**n.a.**
n.a.
**n.a.**
n.a.
**d__Bacteria;p__Firmicutes;c__Clostridia;o__Christensenellales;f__CAG-74;g__Limiplasma**
**ME**
S5_vs_4737
**n.a.**
n.a.

**n.a.**
n.a.
**d__Bacteria;p__Firmicutes;c__Clostridia;o__Christensenellales;f__CAG-74;g__SFTH01**
**ME**
S5_vs_145605
**Caudoviricetes**
Unclassified
**O**
O
**d__Bacteria;p__Firmicutes;c__Clostridia;o__Christensenellales;f__CAG-74;g__UBA11524**
**ME**
S1_vf_158961
**Caudoviricetes**
Unclassified
**O**
O
S7_vs_30665
**n.a.**
n.a.
**n.a.**
n.a.
**Shared**
S12_vs_94042
**n.a.**
n.a.
**n.a.**
n.a.
S3_vs_90341
**n.a.**
n.a.
**n.a.**
n.a.
S8_vf_65522
**n.a.**
n.a.
**n.a.**
n.a.
**SHHC**
S12_vs_48045
**Caudoviricetes**
Unclassified
**Unclassified**
Unclassified
S16_vs_15093
**Caudoviricetes**
Unclassified
**O**
O
**d__Bacteria;p__Firmicutes;c__Clostridia;o__Christensenellales;f__CAG-74;g__Ventricola**
**SHHC**
S4_vf_31979

Caudoviricetes
Unclassified
Unclassified
O
**d__Bacteria;p__Firmicutes;c__Clostridia;o__Christensenellales;f__CAG-917;g__CAG-475**
**SHHC**
S12_vs_32669
**n.a.**
n.a.
**n.a.**
n.a.
**d__Bacteria;p__Firmicutes;c__Clostridia;o__Christensenellales;f__Christensenellaceae;g__Christensenella**
**ME**
S5_vs_145581
**n.a.**
n.a.
**n.a.**
n.a.
**d__Bacteria;p__Firmicutes;c__Clostridia;o__Christensenellales;f__NSJ-53;g__NSJ-53**
**ME**
S5_vs_145597
**Caudoviricetes**
Unclassified
**O**
O
**d__Bacteria;p__Firmicutes;c__Clostridia;o__Christensenellales;f__QAND01;g__UMGS1975**
**ME**
S5_vs_46394
**n.a.**
n.a.
**n.a.**
n.a.
**d__Bacteria;p__Firmicutes;c__Clostridia;o__Clostridiales;f__Clostridiaceae;g__Clostridium**
**ME**
S11_vs_41107
**Caudoviricetes**
Unclassified
**O**
O
S14_vf_56130
**Caudoviricetes**
Unclassified
**O**
O
S14_vf_85122
**Caudoviricetes**
Unclassified
**Unclassified**
O
S14_vs_632
**Caudoviricetes**

Unclassified
**O**
O
S3_vf_100440
**Caudoviricetes**
Unclassified
**O**
O
S3_vs_98675
**Caudoviricetes**
Unclassified
**O**
O
S5_vf_145583
**Caudoviricetes**
Unclassified
**Unclassified**
O
S5_vs_137154
**Caudoviricetes**
Unclassified
**O**
O
S9_vs_90851
**Caudoviricetes**
Unclassified
**O**
O
**Shared**
S16_vf_36146
**Caudoviricetes**
Unclassified
**O**
O
**SHHC**
S10_vf_62230
**Caudoviricetes**
Unclassified
**O**
O
**d__Bacteria;p__Firmicutes;c__Clostridia;o__Clostridiales;f__Clostridiaceae;g__Clostridium_J**
**ME**
S5_vf_6506
**Caudoviricetes**
Unclassified
**O**
O
**d__Bacteria;p__Firmicutes;c__Clostridia;o__Eubacteriales;f__Eubacteriaceae;g__Eubacterium**
**ME**
S11_vs_81775
**Caudoviricetes**

Unclassified
**Unclassified**
Unclassified
S14_vs_115455
**Caudoviricetes**
Unclassified
**O**
O
**d__Bacteria;p__Firmicutes;c__Clostridia;o__Lachnospirales;f__Anaerotignaceae;g__Anaerotignum**
**ME**
S1_vs_145792
**n.a.**
n.a.
**n.a.**
n.a.
S9_vs_90849
**n.a.**
n.a.
**n.a.**
n.a.
**Shared**
S16_vf_84462
**n.a.**
n.a.
**n.a.**
n.a.
S5_vf_60597
**n.a.**
n.a.
**n.a.**
n.a.
S8_vf_201695
**n.a.**
n.a.
**n.a.**
n.a.
**d__Bacteria;p__Firmicutes;c__Clostridia;o__Lachnospirales;f__Anaerotignaceae;g__UMGS1670**
**ME**
S7_vf_72493
**n.a.**
n.a.
**n.a.**
n.a.
**d__Bacteria;p__Firmicutes;c__Clostridia;o__Lachnospirales;f__CAG-274;g__CAG-274**
**ME**
S9_vf_73311
**Caudoviricetes**
Unclassified
**Unclassified**
Unclassified
**d__Bacteria;p__Firmicutes;c__Clostridia;o__Lachnospirales;f__Lachnospiraceae;g__**

**ME**
S11_vf_34008
**Caudoviricetes**
Unclassified
**O**
O
S11_vs_6878
**Caudoviricetes**
Unclassified
**O**
O
S5_vf_105728
**Caudoviricetes**
Unclassified
**O**
O
**d__Bacteria;p__Firmicutes;c__Clostridia;o__Lachnospirales;f__Lachnospiraceae;g__Acetatifactor**
**SHHC**
S16_vs_116956
**n.a.**
n.a.
**n.a.**
n.a.
S6_vs_29152
**Caudoviricetes**
Unclassified
**O**
O
**d__Bacteria;p__Firmicutes;c__Clostridia;o__Lachnospirales;f__Lachnospiraceae;g__Agathobacter**
**ME**
S3_vf_67333
**Caudoviricetes**
Unclassified
**O**
O
**Shared**
S14_vs_66841
**Caudoviricetes**
Unclassified
**O**
O
S3_vs_127421
**Caudoviricetes**
Unclassified
**O**
O
S7_vs_49426
**Caudoviricetes**
Unclassified
**O**
O

S8_vf_132697
**Caudoviricetes**
Unclassified
**O**
O
S8_vs_173961
**Caudoviricetes**
Unclassified
**O**
O
**d__Bacteria;p__Firmicutes;c__Clostridia;o__Lachnospirales;f__Lachnospiraceae;g__Anaerobutyricum**
**ME**
S5_vs_145602
**Caudoviricetes**
Unclassified
**O**
O
S7_vf_114476
**Caudoviricetes**
Unclassified
**O**
O
S7_vs_137611
**Caudoviricetes**
Unclassified
**O**
O
S7_vs_138109
**Caudoviricetes**
Unclassified
**O**
O
**d__Bacteria;p__Firmicutes;c__Clostridia;o__Lachnospirales;f__Lachnospiraceae;g__Anaerostipes**
**ME**
S14_vs_39797
**Caudoviricetes**
Unclassified
**O**
O
S5_vf_116300
**n.a.**
n.a.
**n.a.**
n.a.
S5_vs_139401
**n.a.**
n.a.
**n.a.**
n.a.
S5_vs_141621
**Caudoviricetes**

Unclassified
**O**
O
S7_vf_49485
**Caudoviricetes**
Unclassified
**O**
O

**d__Bacteria;p__Firmicutes;c__Clostridia;o__Lachnospirales;f__Lachnospiraceae;g__Bariatricus**

**ME**
S11_vf_10907
**Caudoviricetes**
Unclassified
**O**
O
S7_vs_86377
**Caudoviricetes**
Unclassified
**O**
O
**Shared**
S9_vs_1966
**Caudoviricetes**
Unclassified
**O**
O
**SHHC**
S12_vs_98175
**Caudoviricetes**
Unclassified
**O**
O

**d__Bacteria;p__Firmicutes;c__Clostridia;o__Lachnospirales;f__Lachnospiraceae;g__Blautia**

**ME**
S14_vs_115377
**Caudoviricetes**
Unclassified
**O**
O
S5_vs_95307
**Caudoviricetes**
Unclassified
**Unclassified**
O
**Shared**
S14_vs_96286
**Caudoviricetes**
Unclassified
**O**
O
S6_vs_54606

**Caudoviricetes**
Unclassified
**O**
O
**d__Bacteria;p__Firmicutes;c__Clostridia;o__Lachnospirales;f__Lachnospiraceae;g__Blautia_A**
**ME**
S11_vf_28645
**n.a.**
n.a.
**n.a.**
n.a.
S11_vs_34137
**Caudoviricetes**
Unclassified
**O**
O
S11_vs_82368
**Caudoviricetes**
Unclassified
**O**
O
S13_vs_121307
**n.a.**
n.a.
**n.a.**
n.a.
S3_vf_8058
**Caudoviricetes**
Unclassified
**Unclassified**
Eponavirus
S5_vs_68254
**Caudoviricetes**
Unclassified
**O**
O
S7_vs_101751
**n.a.**
n.a.
**n.a.**
n.a.
S7_vs_135679
**n.a.**
n.a.
**n.a.**
n.a.
S7_vs_48708
**Caudoviricetes**
Unclassified
**O**
O

S9_vs_90859
**n.a.**
Unclassified
**O**
O
**Shared**
S10_vf_80712
**n.a.**
n.a.
**n.a.**
n.a.
S11_vf_28515
**Caudoviricetes**
Unclassified
**O**
O
S16_vf_96483
**n.a.**
n.a.
**n.a.**
n.a.
S17_vf_46388
**Caudoviricetes**
Unclassified
**O**
O
S3_vf_11428
**Caudoviricetes**
Unclassified
**O**
O
S4_vf_13353
**Caudoviricetes**
Unclassified
**O**
O
S5_vf_145604
**Caudoviricetes**
Unclassified
**O**
O
S5_vs_97574
**Caudoviricetes**
Unclassified
**O**
O
S7_vs_137998
**n.a.**
n.a.
**n.a.**
n.a.

**SHHC**
S4_vs_VIRSorter_12831-circular-cat_2
**Caudoviricetes**
Unclassified
**O**
O
**d__Bacteria;p__Firmicutes;c__Clostridia;o__Lachnospirales;f__Lachnospiraceae;g__CAG-127**
**ME**
S9_vs_75050
**Caudoviricetes**
Unclassified
**O**
O
**SHHC**
S16_vs_101072
**Caudoviricetes**
Unclassified
**O**
O
**d__Bacteria;p__Firmicutes;c__Clostridia;o__Lachnospirales;f__Lachnospiraceae;g__CAG-317**
**Shared**
S16_vs_140709
**n.a.**
n.a.
**n.a.**
n.a.
**d__Bacteria;p__Firmicutes;c__Clostridia;o__Lachnospirales;f__Lachnospiraceae;g__Clostridium_AP**
**ME**
S5_vs_145590
**Caudoviricetes**
Unclassified
**O**
O
**d__Bacteria;p__Firmicutes;c__Clostridia;o__Lachnospirales;f__Lachnospiraceae;g__Coprococcus**
**Shared**
S14_vf_92240
**Caudoviricetes**
Unclassified
**O**
O
**SHHC**
S16_vs_173079
**Caudoviricetes**
Unclassified
**O**
O
**d__Bacteria;p__Firmicutes;c__Clostridia;o__Lachnospirales;f__Lachnospiraceae;g__Dorea**
**ME**
S9_vs_32588
**Caudoviricetes**
Unclassified

**O**

O

**Shared**

S12_vs_163990

**Caudoviricetes**

Unclassified

**O**

O

**d__Bacteria;p__Firmicutes;c__Clostridia;o__Lachnospirales;f__Lachnospiraceae;g__Dorea_A**

**ME**

S11_vs_83666

**Caudoviricetes**

Unclassified

**O**

O

S14_vs_117153

**Caudoviricetes**

Unclassified

**O**

O

S5_vs_84946

**Caudoviricetes**

Unclassified

**O**

O

S7_vs_13799

**Caudoviricetes**

Unclassified

**O**

O

S9_vs_58086

**Caudoviricetes**

Unclassified

**O**

O

**Shared**

S5_vf_145588

**Caudoviricetes**

Unclassified

**O**

O

**d__Bacteria;p__Firmicutes;c__Clostridia;o__Lachnospirales;f__Lachnospiraceae;g__Eisenbergiella**

**ME**

S9_vs_88286

**n.a.**

n.a.

**n.a.**

n.a.

**d__Bacteria;p__Firmicutes;c__Clostridia;o__Lachnospirales;f__Lachnospiraceae;g__Enterocloster**

**ME**

S11_vs_38860

- **n.a.**
  - n.a.
    - **n.a.**
      - n.a.
- S11_vs_67573
  - **n.a.**
    - n.a.
      - **n.a.**
        - n.a.
- S14_vs_100227
  - **Caudoviricetes**
    - Unclassified
      - **O**
        - O
- S14_vs_30108
  - **n.a.**
    - n.a.
      - **n.a.**
        - n.a.
- S5_vs_145578
  - **Caudoviricetes**
    - Unclassified
      - **O**
        - O
- S9_vs_41371
  - **Caudoviricetes**
    - Unclassified
      - **O**
        - O
- S9_vs_4343
  - **n.a.**
    - n.a.
      - **n.a.**
        - n.a.
- S9_vs_55738
  - **Caudoviricetes**
    - Unclassified
      - **O**
        - O

**Shared**

- S5_vf_145585
  - **Caudoviricetes**
    - Unclassified
      - **Unclassified**
        - Unclassified
- S6_vs_39025
  - **Caudoviricetes**
    - Unclassified
      - **O**
        - O
- S7_vs_65719

**Caudoviricetes**
Unclassified
**O**
O
S9_vs_90848
**Caudoviricetes**
Unclassified
**O**
O
**d__Bacteria;p__Firmicutes;c__Clostridia;o__Lachnospirales;f__Lachnospiraceae;g__Eubacterium_F**
**ME**
S7_vs_60775
**Caudoviricetes**
Unclassified
**O**
O
**Shared**
S14_vf_99344
**n.a.**
n.a.
**n.a.**
n.a.
**SHHC**
S8_vf_131009
**Caudoviricetes**
Unclassified
**O**
O
**d__Bacteria;p__Firmicutes;c__Clostridia;o__Lachnospirales;f__Lachnospiraceae;g__Eubacterium_G**
**ME**
S5_vs_8125
**Caudoviricetes**
Unclassified
**O**
O
S7_vs_30640
**Caudoviricetes**
Unclassified
**O**
O
**SHHC**
S16_vs_118029
**Caudoviricetes**
Unclassified
**O**
O
**d__Bacteria;p__Firmicutes;c__Clostridia;o__Lachnospirales;f__Lachnospiraceae;g__Eubacterium_I**
**ME**
S11_vs_104042
**n.a.**
n.a.

**n.a.**
n.a.
**d__Bacteria;p__Firmicutes;c__Clostridia;o__Lachnospirales;f__Lachnospiraceae;g__Faecalimonas**
**ME**
S5_vs_59798
**Caudoviricetes**
Unclassified
**O**
O
S7_vf_68546
**Caudoviricetes**
Unclassified
**O**
O
**d__Bacteria;p__Firmicutes;c__Clostridia;o__Lachnospirales;f__Lachnospiraceae;g__Frisingicoccus**
**ME**
S5_vs_58766
**Caudoviricetes**
Unclassified
**O**
O
**d__Bacteria;p__Firmicutes;c__Clostridia;o__Lachnospirales;f__Lachnospiraceae;g__Hungatella**
**ME**
S5_vs_17520
**n.a.**
n.a.
**n.a.**
n.a.
S9_vs_48240
**Caudoviricetes**
Unclassified
**Unclassified**
O
**d__Bacteria;p__Firmicutes;c__Clostridia;o__Lachnospirales;f__Lachnospiraceae;g__Lachnospira**
**ME**
S14_vf_117150
**Caudoviricetes**
Unclassified
**O**
O
S3_vf_108959
**n.a.**
n.a.
**n.a.**
n.a.
S3_vf_127405
**Caudoviricetes**
Unclassified
**O**
O
S3_vs_127401

**Caudoviricetes**
Unclassified
**O**
O
**Shared**
S5_vs_120584
**Caudoviricetes**
Unclassified
**O**
O
**SHHC**
S12_vs_26847
**n.a.**
n.a.
**n.a.**
n.a.
S12_vs_99092
**Caudoviricetes**
Unclassified
**O**
O
S15_vf_2513
**n.a.**
n.a.
**n.a.**
n.a.
S16_vf_1037
**n.a.**
n.a.
**n.a.**
n.a.
S16_vf_5809
**n.a.**
n.a.
**n.a.**
n.a.
S4_vf_31978
**n.a.**
n.a.
**n.a.**
n.a.

**d__Bacteria;p__Firmicutes;c__Clostridia;o__Lachnospirales;f__Lachnospiraceae;g__Marvinbryantia**

**Shared**
S6_vs_56496
**Caudoviricetes**
Unclassified
**Unclassified**
Eponavirus

**d__Bacteria;p__Firmicutes;c__Clostridia;o__Lachnospirales;f__Lachnospiraceae;g__Massilistercora**

**Shared**
S8_vs_5211

**n.a.**
n.a.
**n.a.**
n.a.
**d__Bacteria;p__Firmicutes;c__Clostridia;o__Lachnospirales;f__Lachnospiraceae;g__Mediterraneibacter**
**ME**
S5_vs_28263
**n.a.**
n.a.
**n.a.**
n.a.
S7_vs_138116
**n.a.**
n.a.
**n.a.**
n.a.
**Shared**
S11_vs_68151
**Caudoviricetes**
Unclassified
**O**
O
S4_vs_VIRSorter_3006-cat_1
**Caudoviricetes**
Unclassified
**O**
O
S7_vs_41098
**Caudoviricetes**
Unclassified
**O**
O
**SHHC**
S4_vs_VIRSorter_19324-cat_2
**Caudoviricetes**
Unclassified
**O**
O
S4_vs_VIRSorter_31916-circular-cat_1
**Caudoviricetes**
Unclassified
**O**
O
**d__Bacteria;p__Firmicutes;c__Clostridia;o__Lachnospirales;f__Lachnospiraceae;g__Muricomes**
**ME**
S14_vf_93134
**Caudoviricetes**
Salasmaviridae
**O**
O
S14_vs_56251

**Caudoviricetes**
Unclassified
**O**
O
S14_vs_59347
**Caudoviricetes**
Unclassified
**O**
O
S17_vf_36452
**Caudoviricetes**
Salasmaviridae
**O**
O
S5_vs_80498
**Caudoviricetes**
Unclassified
**O**
O
**Shared**
S2_vf_17850
**Caudoviricetes**
Salasmaviridae
**O**
O
S6_vs_56498
**Caudoviricetes**
Unclassified
**O**
O
**SHHC**
S12_vf_61299
**Caudoviricetes**
Salasmaviridae
**O**
O
**d__Bacteria;p__Firmicutes;c__Clostridia;o__Lachnospirales;f__Lachnospiraceae;g__OF09-33XD**
**SHHC**
S12_vf_176437
**Caudoviricetes**
Salasmaviridae
**O**
O
**d__Bacteria;p__Firmicutes;c__Clostridia;o__Lachnospirales;f__Lachnospiraceae;g__Roseburia**
**ME**
S14_vf_58919
**Caudoviricetes**
Unclassified
**Unclassified**
Mushuvirus
S5_vs_120079

**Caudoviricetes**
Unclassified
**Unclassified**
Unclassified
S5_vs_122344
**Caudoviricetes**
Unclassified
**Unclassified**
O
S5_vs_145598
**Caudoviricetes**
Unclassified
**O**
O
**Shared**
S3_vs_16825
**Caudoviricetes**
Unclassified
**O**
O
S4_vs_VIRSorter_4746-cat_2
**Caudoviricetes**
Salasmaviridae
**O**
O
S6_vs_12945
**Caudoviricetes**
Unclassified
**O**
O
S8_vs_206526
**Caudoviricetes**
Unclassified
**Unclassified**
Unclassified
**d__Bacteria;p__Firmicutes;c__Clostridia;o__Lachnospirales;f__Lachnospiraceae;g__RUG115**
**ME**
S7_vs_38834
**Caudoviricetes**
Unclassified
**O**
O
**d__Bacteria;p__Firmicutes;c__Clostridia;o__Lachnospirales;f__Lachnospiraceae;g__Ruminococcus_B**
**ME**
S14_vs_26320
**Caudoviricetes**
Unclassified
**O**
O
S7_vf_60968
**Caudoviricetes**

Unclassified
**O**
O
S9_vs_44331
**Caudoviricetes**
Unclassified
**O**
O
**d__Bacteria;p__Firmicutes;c__Clostridia;o__Lachnospirales;f__Lachnospiraceae;g__Sellimonas**
**ME**
S9_vs_34898
**Caudoviricetes**
Unclassified
**O**
O
S9_vs_62803
**Caudoviricetes**
Unclassified
**O**
O
**Shared**
S6_vs_56491
**Caudoviricetes**
Unclassified
**O**
O
**d__Bacteria;p__Firmicutes;c__Clostridia;o__Lachnospirales;f__Lachnospiraceae;g__UBA9502**
**SHHC**
S10_vf_46328
**Caudoviricetes**
Unclassified
**O**
O
**d__Bacteria;p__Firmicutes;c__Clostridia;o__Lachnospirales;f__Lachnospiraceae;g__Ventrimonas**
**ME**
S11_vs_26767
**Caudoviricetes**
Unclassified
**O**
O
**d__Bacteria;p__Firmicutes;c__Clostridia;o__Monoglobales;f__Monoglobaceae;g__Monoglobus**
**Shared**
S3_vf_101611
**Caudoviricetes**
Unclassified
**O**
O
**SHHC**
S16_vs_38863
**Caudoviricetes**
Unclassified

**O**
O
**d__Bacteria;p__Firmicutes;c__Clostridia;o__Oscillospirales;f__Acutalibacteraceae;g__Acutalibacter**
**ME**
S3_vs_114271
**n.a.**
n.a.
**n.a.**
n.a.
**d__Bacteria;p__Firmicutes;c__Clostridia;o__Oscillospirales;f__Acutalibacteraceae;g__An172**
**ME**
S11_vs_48109
**Caudoviricetes**
Unclassified
**O**
O
**d__Bacteria;p__Firmicutes;c__Clostridia;o__Oscillospirales;f__Acutalibacteraceae;g__CAG-180**
**ME**
S11_vf_104052
**Caudoviricetes**
Unclassified
**O**
O
S7_vs_47050
**Caudoviricetes**
Unclassified
**O**
O
**d__Bacteria;p__Firmicutes;c__Clostridia;o__Oscillospirales;f__Acutalibacteraceae;g__CAG-217**
**ME**
S3_vs_122706
**Caudoviricetes**
Unclassified
**O**
O
S3_vs_18858
**n.a.**
n.a.
**n.a.**
n.a.
**d__Bacteria;p__Firmicutes;c__Clostridia;o__Oscillospirales;f__Acutalibacteraceae;g__Eubacterium_R**
**SHHC**
S16_vs_120149
**n.a.**
n.a.
**n.a.**
n.a.
**d__Bacteria;p__Firmicutes;c__Clostridia;o__Oscillospirales;f__Acutalibacteraceae;g__Fimivicinus**
**ME**
S14_vs_88666
**Caudoviricetes**

Unclassified
**O**
O
**d__Bacteria;p__Firmicutes;c__Clostridia;o__Oscillospirales;f__Acutalibacteraceae;g__Hydrogeniiclostridium**
**ME**
S7_vs_28733
**Caudoviricetes**
Unclassified
**O**
O
S7_vs_38113
**Caudoviricetes**
Unclassified
**O**
O
**d__Bacteria;p__Firmicutes;c__Clostridia;o__Oscillospirales;f__Acutalibacteraceae;g__Pseudoruminococcus**
**Shared**
S10_vs_77004
**n.a.**
n.a.
**n.a.**
n.a.
**d__Bacteria;p__Firmicutes;c__Clostridia;o__Oscillospirales;f__Acutalibacteraceae;g__Ruminococcus_E**
**ME**
S13_vf_28309
**Caudoviricetes**
Salasmaviridae
**O**
O
S14_vs_39266
**Caudoviricetes**
Unclassified
**Unclassified**
O
S3_vs_49835
**n.a.**
n.a.
**n.a.**
n.a.
S5_vs_20268
**Caudoviricetes**
Unclassified
**Unclassified**
Unclassified
S5_vs_40848
**Caudoviricetes**
Unclassified
**O**
O
S7_vs_72737
**n.a.**

n.a.
**n.a.**
n.a.
**Shared**
S14_vs_66139
**Caudoviricetes**
Unclassified
**O**
O
S16_vs_82116
**Caudoviricetes**
Unclassified
**Unclassified**
O
S4_vf_11308
**Caudoviricetes**
Unclassified
**Unclassified**
O
S7_vs_16344
**Caudoviricetes**
Unclassified
**O**
O
S8_vs_115958
**Caudoviricetes**
Unclassified
**Unclassified**
O
S9_vs_90727
**Caudoviricetes**
Unclassified
**Unclassified**
O
**d__Bacteria;p__Firmicutes;c__Clostridia;o__Oscillospirales;f__Acutalibacteraceae;g__UBA1417**
**ME**
S14_vs_61530
**n.a.**
n.a.
**n.a.**
n.a.
**Shared**
S12_vf_79666
**n.a.**
n.a.
**n.a.**
n.a.
**d__Bacteria;p__Firmicutes;c__Clostridia;o__Oscillospirales;f__Acutalibacteraceae;g__UBA1691**
**ME**
S5_vs_3501
**Caudoviricetes**

Unclassified
**O**
O
S7_vs_129271
**n.a.**
n.a.
**n.a.**
n.a.
**SHHC**
S10_vs_36407
**n.a.**
n.a.
**n.a.**
n.a.
**d__Bacteria;p__Firmicutes;c__Clostridia;o__Oscillospirales;f__Acutalibacteraceae;g__UBA737**
**ME**
S3_vs_95574
**n.a.**
n.a.
**n.a.**
n.a.
**d__Bacteria;p__Firmicutes;c__Clostridia;o__Oscillospirales;f__Acutalibacteraceae;g__UMGS1071**
**ME**
S9_vs_26179
**n.a.**
n.a.
**n.a.**
n.a.
**Shared**
S6_vs_56490
**Caudoviricetes**
Unclassified
**O**
O
**d__Bacteria;p__Firmicutes;c__Clostridia;o__Oscillospirales;f__Acutalibacteraceae;g__UMGS1591**
**ME**
S9_vs_90860
**Caudoviricetes**
Unclassified
**O**
O
**d__Bacteria;p__Firmicutes;c__Clostridia;o__Oscillospirales;f__Acutalibacteraceae;g__UMGS263**
**ME**
S11_vs_3006
**Caudoviricetes**
Unclassified
**Unclassified**
O
**d__Bacteria;p__Firmicutes;c__Clostridia;o__Oscillospirales;f__Acutalibacteraceae;g__UMGS856**
**ME**
S5_vs_140946

**n.a.**
n.a.
**n.a.**
n.a.
**d__Bacteria;p__Firmicutes;c__Clostridia;o__Oscillospirales;f__Butyricicoccaceae;g__Agathobaculum**
**ME**
S14_vs_112068
**n.a.**
n.a.
**n.a.**
n.a.
S5_vs_43207
**Caudoviricetes**
Unclassified
**O**
O
S5_vs_84574
**n.a.**
n.a.
**n.a.**
n.a.
S7_vs_103530
**n.a.**
n.a.
**n.a.**
n.a.
S9_vs_19689
**n.a.**
n.a.
**n.a.**
n.a.
**Shared**
S6_vs_56492
**n.a.**
n.a.
**n.a.**
n.a.
**d__Bacteria;p__Firmicutes;c__Clostridia;o__Oscillospirales;f__CAG-272;g__CAG-272**
**ME**
S3_vf_127392
**Caudoviricetes**
Unclassified
**O**
O
**d__Bacteria;p__Firmicutes;c__Clostridia;o__Oscillospirales;f__CAG-272;g__QALR01**
**SHHC**
S12_vs_188820
**n.a.**
n.a.
**n.a.**
n.a.

**d__Bacteria;p__Firmicutes;c__Clostridia;o__Oscillospirales;f__Oscillospiraceae;g__**

**ME**

S13_vs_148294

**Caudoviricetes**

Unclassified

**O**

O

**Shared**

S3_vs_54303

**Caudoviricetes**

Unclassified

**Unclassified**

Eponavirus

S7_vs_4956

**Caudoviricetes**

Unclassified

**Unclassified**

Eponavirus

**d__Bacteria;p__Firmicutes;c__Clostridia;o__Oscillospirales;f__Oscillospiraceae;g__An92**

**ME**

S9_vs_14235

**n.a.**

n.a.

**n.a.**

n.a.

S9_vs_84046

**Caudoviricetes**

Unclassified

**O**

O

**d__Bacteria;p__Firmicutes;c__Clostridia;o__Oscillospirales;f__Oscillospiraceae;g__CAG-103**

**ME**

S5_vs_77593

**Caudoviricetes**

Unclassified

**O**

O

S7_vs_5236

**Caudoviricetes**

Unclassified

**O**

O

**Shared**

S7_vf_76463

**n.a.**

n.a.

**n.a.**

n.a.

**d__Bacteria;p__Firmicutes;c__Clostridia;o__Oscillospirales;f__Oscillospiraceae;g__CAG-110**

**ME**

S5_vs_107105

**Caudoviricetes**
Unclassified
**O**
O
**SHHC**
S10_vs_120541
**n.a.**
n.a.
**n.a.**
n.a.
**d__Bacteria;p__Firmicutes;c__Clostridia;o__Oscillospirales;f__Oscillospiraceae;g__CAG-170**
**ME**
S1_vf_136526
**Caudoviricetes**
Unclassified
**Unclassified**
O
S1_vs_117317
**Caudoviricetes**
Unclassified
**Unclassified**
O
**Shared**
S1_vf_6272
**Caudoviricetes**
Unclassified
**Unclassified**
O
S12_vf_173208
**n.a.**
n.a.
**n.a.**
n.a.
S3_vf_82571
**Caudoviricetes**
Unclassified
**O**
O
**SHHC**
S8_vs_51368
**n.a.**
n.a.
**n.a.**
n.a.
S8_vs_54183
**Caudoviricetes**
Unclassified
**Unclassified**
O
**d__Bacteria;p__Firmicutes;c__Clostridia;o__Oscillospirales;f__Oscillospiraceae;g__CAG-83**
**ME**

S11_vf_33117
**n.a.**
n.a.
**n.a.**
n.a.
S11_vf_99751
**n.a.**
n.a.
**n.a.**
n.a.
S13_vf_109235
**n.a.**
n.a.
**n.a.**
n.a.
S13_vs_13129
**Caudoviricetes**
Unclassified
**O**
O
S14_vs_97835
**n.a.**
n.a.
**n.a.**
n.a.
S5_vs_74199
**n.a.**
n.a.
**n.a.**
n.a.
**Shared**
S12_vs_164056
**n.a.**
n.a.
**n.a.**
n.a.
S13_vs_35816
**Caudoviricetes**
Unclassified
**Unclassified**
Lagaffevirus
S8_vf_170878
**Caudoviricetes**
Unclassified
**Unclassified**
Toutatisvirus
S8_vs_125717
**n.a.**
n.a.
**n.a.**
n.a.

S9_vf_64844
**Caudoviricetes**
Unclassified
**Unclassified**
Lagaffevirus
**d__Bacteria;p__Firmicutes;c__Clostridia;o__Oscillospirales;f__Oscillospiraceae;g__Dysosmobacter**
**ME**
S11_vs_52577
**Caudoviricetes**
Unclassified
**Unclassified**
Mushuvirus
S7_vs_28150
**n.a.**
n.a.
**n.a.**
n.a.
S9_vs_69322
**n.a.**
n.a.
**n.a.**
n.a.
**Shared**
S10_vf_120738
**n.a.**
n.a.
**n.a.**
n.a.
S17_vs_127519
**n.a.**
n.a.
**n.a.**
n.a.
**SHHC**
S8_vs_50431
**n.a.**
n.a.
**n.a.**
n.a.
**d__Bacteria;p__Firmicutes;c__Clostridia;o__Oscillospirales;f__Oscillospiraceae;g__Enterenecus**
**ME**
S5_vs_117417
**n.a.**
n.a.
**n.a.**
n.a.
**d__Bacteria;p__Firmicutes;c__Clostridia;o__Oscillospirales;f__Oscillospiraceae;g__ER4**
**ME**
S1_vf_22149
**n.a.**
n.a.

**n.a.**
n.a.
S1_vs_143929
**n.a.**
n.a.
**n.a.**
n.a.
S13_vf_150407
**Faserviricetes**
Inoviridae
**Unclassified**
Inovirus
S3_vs_127411
**n.a.**
n.a.
**n.a.**
n.a.
**Shared**
S1_vs_155183
**Caudoviricetes**
Unclassified
**Unclassified**
O
S11_vf_104034
**Caudoviricetes**
Unclassified
**Unclassified**
O
S12_vf_18902
**Caudoviricetes**
Unclassified
**O**
O
S16_vf_6102
**Caudoviricetes**
Unclassified
**Unclassified**
Toutatisvirus
S16_vf_97279
**Caudoviricetes**
Unclassified
**Unclassified**
Eponavirus
S17_vs_103009
**n.a.**
n.a.
**n.a.**
n.a.
S3_vf_127400
**Caudoviricetes**
Unclassified

**Unclassified**
O
S4_vs_VIRSorter_4959-cat_2
**Caudoviricetes**
Unclassified
**O**
O
S5_vf_136076
**n.a.**
n.a.
**n.a.**
n.a.
**SHHC**
S16_vs_78429
**Caudoviricetes**
Unclassified
**Unclassified**
O

**d__Bacteria;p__Firmicutes;c__Clostridia;o__Oscillospirales;f__Oscillospiraceae;g__Evtepia**

**Shared**
S14_vs_117166
**n.a.**
n.a.
**n.a.**
n.a.

**d__Bacteria;p__Firmicutes;c__Clostridia;o__Oscillospirales;f__Oscillospiraceae;g__Flavonifractor**

**ME**
S11_vf_95067
**n.a.**
n.a.
**n.a.**
n.a.
S13_vf_147589
**n.a.**
n.a.
**n.a.**
n.a.
S14_vf_115736
**Caudoviricetes**
Unclassified
**Unclassified**
O
S14_vs_3251
**n.a.**
n.a.
**n.a.**
n.a.
S14_vs_56410
**Caudoviricetes**
Unclassified
**O**

O
S5_vs_21778
**n.a.**
n.a.
**n.a.**
n.a.
S5_vs_65490
**n.a.**
n.a.
**n.a.**
n.a.
S5_vs_69980
**Caudoviricetes**
Unclassified
**O**
O
S7_vs_83100
**Caudoviricetes**
Unclassified
**O**
O
S7_vs_83474
**Caudoviricetes**
Unclassified
**Unclassified**
O
S7_vs_94738
**Caudoviricetes**
Unclassified
**Unclassified**
O
S9_vs_4558
**Caudoviricetes**
Unclassified
**O**
O
S9_vs_50439
**n.a.**
n.a.
**n.a.**
n.a.
**Shared**
S4_vs_VIRSorter_15837-cat_2
**Caudoviricetes**
Unclassified
**O**
O
S4_vs_VIRSorter_31196-circular-cat_2
**n.a.**
n.a.
**n.a.**

n.a.
S4_vs_VIRSorter_31560-circular-cat_2
**Caudoviricetes**
Unclassified
**O**
O
S5_vf_21869
**Caudoviricetes**
Unclassified
**Unclassified**
O
S6_vs_15813
**n.a.**
n.a.
**n.a.**
n.a.
S8_vf_111790
**Caudoviricetes**
Unclassified
**O**
O
S8_vf_166403
**n.a.**
n.a.
**n.a.**
n.a.
**SHHC**
S16_vf_173099
**n.a.**
n.a.
**n.a.**
n.a.
**d__Bacteria;p__Firmicutes;c__Clostridia;o__Oscillospirales;f__Oscillospiraceae;g__Intestinimonas**
**ME**
S7_vs_118295
**n.a.**
n.a.
**n.a.**
n.a.
S7_vs_69630
**Caudoviricetes**
Unclassified
**O**
O
S9_vs_5676
**Caudoviricetes**
Unclassified
**O**
O
**Shared**
S12_vs_92868

**Caudoviricetes**
Unclassified
**O**
O
S14_vf_109545
**n.a.**
n.a.
**n.a.**
n.a.
S4_vs_VIRSorter_30222-cat_2
**Caudoviricetes**
Unclassified
**O**
O
S5_vs_124035
**n.a.**
n.a.
**n.a.**
n.a.
S7_vs_78507
**n.a.**
n.a.
**n.a.**
n.a.
S8_vf_208110
**Caudoviricetes**
Unclassified
**O**
O
S8_vf_55476
**n.a.**
n.a.
**n.a.**
n.a.
**d__Bacteria;p__Firmicutes;c__Clostridia;o__Oscillospirales;f__Oscillospiraceae;g__Lawsonibacter**
**ME**
S11_vs_19892
**Caudoviricetes**
Unclassified
**Unclassified**
Eponavirus
S14_vs_67094
**n.a.**
n.a.
**n.a.**
n.a.
S14_vs_9676
**Caudoviricetes**
Unclassified
**O**
O

S17_vf_97671
**Caudoviricetes**
Unclassified
**Unclassified**
Mushuvirus
S7_vf_138019
**n.a.**
n.a.
**n.a.**
n.a.
**Shared**
S5_vf_103206
**n.a.**
n.a.
**n.a.**
n.a.
S8_vs_134805
**Caudoviricetes**
Unclassified
**Unclassified**
Mushuvirus
**d__Bacteria;p__Firmicutes;c__Clostridia;o__Oscillospirales;f__Oscillospiraceae;g__UBA5446**
**Shared**
S16_vs_75108
**n.a.**
n.a.
**n.a.**
n.a.
**d__Bacteria;p__Firmicutes;c__Clostridia;o__Oscillospirales;f__Oscillospiraceae;g__UBA9475**
**SHHC**
S8_vs_145173
**Caudoviricetes**
Unclassified
**O**
O
**d__Bacteria;p__Firmicutes;c__Clostridia;o__Oscillospirales;f__Oscillospiraceae;g__UMGS1766**
**Shared**
S16_vs_20060
**Caudoviricetes**
Unclassified
**Unclassified**
O
**d__Bacteria;p__Firmicutes;c__Clostridia;o__Oscillospirales;f__Ruminococcaceae;g__Anaerotruncus**
**ME**
S5_vs_118168
**n.a.**
n.a.
**n.a.**
n.a.
S5_vs_130958
**Caudoviricetes**

Unclassified
**O**
O
S5_vs_32885
**Caudoviricetes**
Unclassified
**O**
O
**d__Bacteria;p__Firmicutes;c__Clostridia;o__Oscillospirales;f__Ruminococcaceae;g__CAG-353**
**Shared**
S12_vs_133070
**Caudoviricetes**
Unclassified
**O**
O
**d__Bacteria;p__Firmicutes;c__Clostridia;o__Oscillospirales;f__Ruminococcaceae;g__D5**
**Shared**
S3_vf_80287
**Caudoviricetes**
Unclassified
**O**
O
**d__Bacteria;p__Firmicutes;c__Clostridia;o__Oscillospirales;f__Ruminococcaceae;g__Faecalibacterium**
**ME**
S1_vs_23682
**Caudoviricetes**
Unclassified
**O**
O
S11_vs_11994
**Caudoviricetes**
Unclassified
**O**
O
S13_vf_54201
**Caudoviricetes**
Unclassified
**O**
O
S14_vs_90934
**n.a.**
n.a.
**n.a.**
n.a.
S17_vs_110666
**Caudoviricetes**
Unclassified
**O**
O
S3_vs_42569
**Caudoviricetes**

Unclassified
**Unclassified**
Toutatisvirus
S3_vs_68031
**Caudoviricetes**
Unclassified
**O**
O
S5_vs_117319
**Caudoviricetes**
Unclassified
**Unclassified**
O
S5_vs_124677
**Caudoviricetes**
Unclassified
**Unclassified**
O
S5_vs_30490
**n.a.**
n.a.
**n.a.**
n.a.
S7_vf_84974
**Caudoviricetes**
Unclassified
**O**
O
S7_vs_87279
**Caudoviricetes**
Unclassified
**Unclassified**
O
S7_vs_97737
**n.a.**
n.a.
**n.a.**
n.a.
**Shared**
S12_vs_87088
**Caudoviricetes**
Unclassified
**O**
O
S16_vf_108554
**n.a.**
n.a.
**n.a.**
n.a.
S16_vs_132979
**Caudoviricetes**

Unclassified
**O**
O
S16_vs_173141
**n.a.**
n.a.
**n.a.**
n.a.
S3_vf_127409
**Caudoviricetes**
Unclassified
**O**
O
S3_vf_99734
**Caudoviricetes**
Unclassified
**Unclassified**
Lagaffevirus
S3_vs_87728
**n.a.**
n.a.
**n.a.**
n.a.
S3_vs_90331
**Caudoviricetes**
Unclassified
**Unclassified**
O
S4_vs_VIRSorter_27316-cat_2
**Caudoviricetes**
Unclassified
**O**
O
S5_vf_134040
**Caudoviricetes**
Unclassified
**Unclassified**
Mushuvirus
S5_vs_130294
**n.a.**
n.a.
**n.a.**
n.a.
S6_vs_1146
**Caudoviricetes**
Unclassified
**O**
O
S6_vs_34453
**Caudoviricetes**
Unclassified

**Unclassified**
O
S7_vf_39724
**Caudoviricetes**
Unclassified
**Unclassified**
Mushuvirus
S7_vs_13370
**n.a.**
n.a.
**n.a.**
n.a.
S7_vs_135322
**Caudoviricetes**
Unclassified
**Unclassified**
O
S7_vs_26757
**n.a.**
n.a.
**n.a.**
n.a.
**SHHC**
S10_vf_24791
**Caudoviricetes**
Unclassified
**Unclassified**
Eponavirus
S10_vs_46819
**Caudoviricetes**
Unclassified
**O**
O
S12_vs_81729
**Caudoviricetes**
Unclassified
**O**
O
S16_vs_18440
**Caudoviricetes**
Unclassified
**Unclassified**
Toutatisvirus
S6_vs_53966
**Caudoviricetes**
Unclassified
**O**
O
S8_vf_101283
**Caudoviricetes**
Unclassified

**Unclassified**
Toutatisvirus
S8_vf_187742
**Caudoviricetes**
Unclassified
**O**
O
S8_vf_208119
**Malgrandaviricetes**
Microviridae
**Gokushovirinae**
Gokushovirus
S8_vs_179529
**Caudoviricetes**
Unclassified
**Unclassified**
O
S8_vs_63277
**Caudoviricetes**
Unclassified
**O**
O
**d__Bacteria;p__Firmicutes;c__Clostridia;o__Oscillospirales;f__Ruminococcaceae;g__Gemmiger**
**ME**
S1_vf_28398
**n.a.**
n.a.
**n.a.**
n.a.
S1_vs_133577
**Caudoviricetes**
Unclassified
**O**
O
S14_vf_117147
**Caudoviricetes**
Unclassified
**O**
O
S3_vs_119771
**Caudoviricetes**
Unclassified
**O**
O
S3_vs_127391
**n.a.**
n.a.
**n.a.**
n.a.
S3_vs_15450
**Caudoviricetes**

Unclassified
**O**
O
S7_vs_111831
**Caudoviricetes**
Unclassified
**O**
O
**Shared**
S4_vs_VIRSorter_31976-circular-cat_2
**n.a.**
n.a.
**n.a.**
n.a.
S7_vs_42563
**n.a.**
n.a.
**n.a.**
n.a.
**SHHC**
S16_vs_76823
**n.a.**
n.a.
**n.a.**
n.a.
S8_vf_151384
**Caudoviricetes**
Unclassified
**Unclassified**
Eponavirus
S8_vf_208126
**Caudoviricetes**
Unclassified
**O**
O
S8_vs_102608
**n.a.**
n.a.
**n.a.**
n.a.
**d__Bacteria;p__Firmicutes;c__Clostridia;o__Oscillospirales;f__Ruminococcaceae;g__Massilimaliae**
**ME**
S14_vs_73301
**Caudoviricetes**
Unclassified
**O**
O
**Shared**
S16_vs_17493
**n.a.**
n.a.

**n.a.**
n.a.
**SHHC**
S4_vs_VIRSorter_4839-cat_2
**Caudoviricetes**
Unclassified
**O**
O
**d__Bacteria;p__Firmicutes;c__Clostridia;o__Oscillospirales;f__Ruminococcaceae;g__Massilioclostridium**
**ME**
S11_vs_13652
**Caudoviricetes**
Unclassified
**Unclassified**
O
**d__Bacteria;p__Firmicutes;c__Clostridia;o__Oscillospirales;f__Ruminococcaceae;g__Negativibacillus**
**ME**
S5_vs_128398
**Caudoviricetes**
Unclassified
**O**
O
S9_vs_48347
**Caudoviricetes**
Unclassified
**Unclassified**
O
**Shared**
S11_vs_104064
**Caudoviricetes**
Unclassified
**O**
O
S6_vs_46091
**Caudoviricetes**
Unclassified
**Unclassified**
O
**d__Bacteria;p__Firmicutes;c__Clostridia;o__Oscillospirales;f__Ruminococcaceae;g__Phocea**
**ME**
S5_vs_145611
**Caudoviricetes**
Unclassified
**O**
O
**d__Bacteria;p__Firmicutes;c__Clostridia;o__Oscillospirales;f__Ruminococcaceae;g__QAMM01**
**ME**
S14_vs_101482
**n.a.**
n.a.
**n.a.**

n.a.
**SHHC**
S4_vs_VIRSorter_31171-circular-cat_2
**Caudoviricetes**
Unclassified
**O**
O
**d__Bacteria;p__Firmicutes;c__Clostridia;o__Oscillospirales;f__Ruminococcaceae;g__Ruminiclostridium_E**
**Shared**
S7_vs_21521
**n.a.**
n.a.
**n.a.**
n.a.
**d__Bacteria;p__Firmicutes;c__Clostridia;o__Oscillospirales;f__Ruminococcaceae;g__Ruminococcus_C**
**ME**
S3_vs_29077
**Caudoviricetes**
Unclassified
**O**
O
**SHHC**
S4_vs_VIRSorter_31972-circular-cat_2
**Caudoviricetes**
Unclassified
**O**
O
**d__Bacteria;p__Firmicutes;c__Clostridia;o__Oscillospirales;f__Ruminococcaceae;g__Ruminococcus_D**
**ME**
S14_vf_51134
**Caudoviricetes**
Unclassified
**O**
O
S3_vf_8164
**Caudoviricetes**
Unclassified
**O**
O
S7_vf_71527
**n.a.**
n.a.
**n.a.**
n.a.
S7_vf_8332
**Caudoviricetes**
Unclassified
**O**
O
**Shared**
S10_vs_120570

**n.a.**
n.a.
**n.a.**
n.a.
S16_vs_34021
**n.a.**
n.a.
**n.a.**
n.a.
S17_vf_174033
**Caudoviricetes**
Unclassified
**O**
O
S3_vf_111893
**Caudoviricetes**
Unclassified
**O**
O
S7_vf_13614
**Caudoviricetes**
Unclassified
**O**
O
**SHHC**
S12_vf_181024
**Caudoviricetes**
Unclassified
**O**
O
S16_vs_107235
**n.a.**
n.a.
**n.a.**
n.a.
S16_vs_48714
**n.a.**
n.a.
**n.a.**
n.a.
S8_vf_161703
**Caudoviricetes**
Unclassified
**O**
O
**d__Bacteria;p__Firmicutes;c__Clostridia;o__Oscillospirales;f__Ruminococcaceae;g__Ruthenibacterium**
**ME**
S14_vs_14737
**Caudoviricetes**
Unclassified
**O**

O
S14_vs_38905
**n.a.**
n.a.
**n.a.**
n.a.
S3_vs_25896
**n.a.**
n.a.
**n.a.**
n.a.
S5_vs_121411
**n.a.**
n.a.
**n.a.**
n.a.
S5_vs_13254
**n.a.**
n.a.
**n.a.**
n.a.
S9_vs_66877
**Caudoviricetes**
Unclassified
**O**
O
**Shared**
S11_vs_76784
**Caudoviricetes**
Unclassified
**O**
O
S16_vf_169895
**Caudoviricetes**
Salasmaviridae
**O**
O
S5_vs_85065
**Caudoviricetes**
Unclassified
**O**
O
**SHHC**
S16_vs_103726
**n.a.**
n.a.
**n.a.**
n.a.
S16_vs_81805
**Caudoviricetes**
Unclassified

**O**
O
S6_vs_33711
**Caudoviricetes**
Unclassified
**Unclassified**
Unclassified
**d__Bacteria;p__Firmicutes;c__Clostridia;o__Oscillospirales;f__Ruminococcaceae;g__UBA3818**
**SHHC**
S6_vs_38250
**Caudoviricetes**
Unclassified
**Unclassified**
Unclassified
**d__Bacteria;p__Firmicutes;c__Clostridia;o__Oscillospirales;f__UMGS1783;g__UMGS1783**
**ME**
S7_vs_134391
**n.a.**
n.a.
**n.a.**
n.a.
**SHHC**
S12_vs_188828
**n.a.**
n.a.
**n.a.**
n.a.
**d__Bacteria;p__Firmicutes;c__Clostridia;o__Peptostreptococcales;f__Anaerovoracaceae;g__CAG-145**
**Shared**
S10_vs_2538
**Caudoviricetes**
Unclassified
**O**
O
**d__Bacteria;p__Firmicutes;c__Clostridia;o__Peptostreptococcales;f__Peptostreptococcaceae;g__GCA-900066495**
**Shared**
S3_vf_56363
**Caudoviricetes**
Unclassified
**O**
O
**SHHC**
S4_vs_VIRSorter_15112-cat_2
**n.a.**
n.a.
**n.a.**
n.a.
**d__Bacteria;p__Firmicutes;c__Clostridia;o__TANB77;f__CAG-508;g__CAG-269**
**SHHC**
S12_vs_6836
**Caudoviricetes**

Unclassified
**O**
O
S16_vf_90328
**Caudoviricetes**
Unclassified
**O**
O
**d__Bacteria;p__Firmicutes;c__Clostridia;o__TANB77;f__CAG-508;g__CAG-354**
**SHHC**
S8_vf_13709
**Caudoviricetes**
Unclassified
**O**
O
S8_vf_13758
**Caudoviricetes**
Unclassified
**O**
O
**d__Bacteria;p__Firmicutes;c__Clostridia;o__UBA1212;f__UBA1255;g__Egerieisoma**
**ME**
S5_vs_850
**n.a.**
n.a.
**n.a.**
n.a.
**d__Bacteria;p__Firmicutes;c__Negativicutes;o__Acidaminococcales;f__Acidaminococcaceae;g__Phascolarctobacterium**
**SHHC**
S8_vf_208146
**Malgrandaviricetes**
Microviridae
**Gokushovirinae**
Unclassified
**d__Bacteria;p__Firmicutes;c__Negativicutes;o__Veillonellales;f__Dialisteraceae;g__Dialister**
**ME**
S7_vs_134784
**Caudoviricetes**
Unclassified
**O**
O
S9_vs_67792
**Caudoviricetes**
Unclassified
**O**
O
**d__Bacteria;p__Firmicutes;c__Negativicutes;o__Veillonellales;f__Megasphaeraceae;g__Caecibacter**
**SHHC**
S8_vs_147670
**Caudoviricetes**
Unclassified

**O**
O
**d__Bacteria;p__Firmicutes;c__Negativicutes;o__Veillonellales;f__Veillonellaceae;g__Veillonella**
**ME**
S5_vf_8808
**Caudoviricetes**
Unclassified
**O**
O
**d__Bacteria;p__Patescibacteria;c__Paceibacteria;o__UBA9983_A;f__UBA1006;g__01-FULL-36-10b**
**Shared**
S4_vf_31973
**n.a.**
n.a.
**n.a.**
n.a.
**d__Bacteria;p__Proteobacteria;c__Alphaproteobacteria;o__RF32;f__CAG-239;g__51-20**
**ME**
S11_vs_84627
**n.a.**
n.a.
**n.a.**
n.a.
**Shared**
S13_vs_45488
**Caudoviricetes**
Unclassified
**O**
O
S6_vs_44070
**n.a.**
Unclassified
**O**
O
**d__Bacteria;p__Proteobacteria;c__Alphaproteobacteria;o__RF32;f__CAG-239;g__CAG-267**
**Shared**
S11_vs_6458
**n.a.**
n.a.
**n.a.**
n.a.
S13_vs_9438
**n.a.**
n.a.
**n.a.**
n.a.
S5_vs_9747
**n.a.**
n.a.
**n.a.**
n.a.

S6_vs_39065
**n.a.**
n.a.
**n.a.**
n.a.
**d__Bacteria;p__Proteobacteria;c__Alphaproteobacteria;o__RF32;f__CAG-239;g__CAG-495**
**ME**
S17_vs_8690
**n.a.**
n.a.
**n.a.**
n.a.
**SHHC**
S12_vs_153780
**Caudoviricetes**
Unclassified
**O**
O
**d__Bacteria;p__Proteobacteria;c__Gammaproteobacteria;o__Burkholderiales;f__Burkholderiaceae;g__Duodenibacillus**
**SHHC**
S4_vs_VIRSorter_31980-circular-cat_2
**Caudoviricetes**
Unclassified
**Unclassified**
Unclassified
**d__Bacteria;p__Proteobacteria;c__Gammaproteobacteria;o__Burkholderiales;f__Burkholderiaceae;g__Parasutterella**
**SHHC**
S12_vs_93357
**Caudoviricetes**
Unclassified
**O**
O
S16_vs_120622
**Caudoviricetes**
Unclassified
**O**
O
**d__Bacteria;p__Proteobacteria;c__Gammaproteobacteria;o__Burkholderiales;f__Burkholderiaceae;g__Sutterella**
**ME**
S14_vs_2870
**Caudoviricetes**
Unclassified
**O**
O
S3_vs_51931
**Malgrandaviricetes**
Microviridae
**Gokushovirinae**
O
S7_vs_48523
**Caudoviricetes**

Unclassified
**O**
O
**d__Bacteria;p__Proteobacteria;c__Gammaproteobacteria;o__Enterobacterales;f__Enterobacteriaceae;g__Escherichia**
**ME**
S5_vf_74661
**Caudoviricetes**
Unclassified
**Unclassified**
Unclassified
S5_vs_130032
**Caudoviricetes**
Unclassified
**O**
O
**Shared**
S13_vf_151045
**Caudoviricetes**
Unclassified
**O**
O
**d__Bacteria;p__Proteobacteria;c__Gammaproteobacteria;o__Enterobacterales;f__Enterobacteriaceae;g__Salmonella**
**ME**
S11_vs_5077
**Caudoviricetes**
Peduoviridae
**Unclassified**
Quadragintavirus
**d__Bacteria;p__Proteobacteria;c__Gammaproteobacteria;o__Enterobacterales;f__Pasteurellaceae;g__Glaesserella**
**SHHC**
S12_vf_10887
**n.a.**
n.a.
**n.a.**
n.a.
**d__Bacteria;p__Spirochaetota;c__Spirochaetia;o__Sphaerochaetales;f__Sphaerochaetaceae;g__UBA9732**
**Shared**
S3_vf_115304
**Caudoviricetes**
Suoliviridae
**Uncouvirinae**
Aurodevirus
**d__Bacteria;p__Verrucomicrobiota;c__Lentisphaeria;o__Victivallales;f__Victivallaceae;g__Victivallis**
**ME**
S7_vs_87439
**n.a.**
n.a.
**n.a.**
n.a.
**d__Bacteria;p__Verrucomicrobiota;c__Verrucomicrobiae;o__Verrucomicrobiales;f__Akkermansiaceae;g__Akkermansia**
**ME**

S13_vf_7670
**n.a.**
n.a.
**n.a.**
n.a.
S5_vs_86562
**Caudoviricetes**
Unclassified
**O**
O
S7_vs_29595
**Caudoviricetes**
Unclassified
**O**
O
**Shared**
S16_vs_173096
**Caudoviricetes**
Unclassified
**Unclassified**
Unclassified
S6_vf_25710
**n.a.**
n.a.
**n.a.**
n.a.
**SHHC**
S12_vs_86928
**Caudoviricetes**
Unclassified
**O**
O

| Host interactions with: | # Hosts |
|---|---|
| ME-viruses-only | 22 |
| SHHC-viruses-only | 23 |
| Shared/communal viruses | 61 |

**WMS_Hosts**

- **d__Bacteria;p__Actinobacteriota;c__Actinomycetia;o__Actinomycetales;f__Bifidobacteriaceae;g__Bifidobacterium**
  - **ME**
    - S17-wms-combined_186314
      - **Caudoviricetes**
        - Unclassified
          - **O**
            - O
    - S1-wms-combined_307323
      - **Caudoviricetes**
        - Unclassified
          - **O**
            - O
    - S1-wms-combined_331544
      - **n.a.**
        - n.a.
          - **n.a.**
            - n.a.
    - S1-wms-combined_85225
      - **Caudoviricetes**
        - Unclassified
          - **O**
            - O
    - S3-wms_94494
      - **Caudoviricetes**
        - Unclassified
          - **O**
            - O
    - S3-wms-combined_135428
      - **Caudoviricetes**
        - Unclassified
          - **O**
            - O
    - S7-wms_157264
      - **n.a.**
        - Unclassified
          - **Unclassified**
            - Unclassified
    - S7-wms-combined_136642
      - **n.a.**
        - n.a.
          - **n.a.**
            - n.a.
    - S7-wms-combined_143218
      - **n.a.**
        - n.a.
          - **n.a.**
            - n.a.
    - S7-wms-combined_147158
      - **Caudoviricetes**
        - Unclassified

**O**
O
S7-wms-combined_167867
**n.a.**
n.a.
**n.a.**
n.a.
**Shared**
S4-wms-combined_17284
**Caudoviricetes**
Unclassified
**Unclassified**
Unclassified
S4-wms-combined_51079
**n.a.**
n.a.
**n.a.**
n.a.
S5-wms-combined_117213
**Caudoviricetes**
Unclassified
**O**
O
S5-wms-combined_39793
**n.a.**
n.a.
**n.a.**
n.a.
S7-wms-combined_174935
**Caudoviricetes**
Unclassified
**O**
O
S9-wms-combined_76274
**Caudoviricetes**
Unclassified
**O**
O
**SHHC**
S4-wms_161443
**n.a.**
Unclassified
**Unclassified**
Unclassified
**d__Bacteria;p__Actinobacteriota;c__Coriobacteriia;o__Coriobacteriales;f__Coriobacteriaceae;g__Collinsella**
**ME**
S5-wms_94965
**Caudoviricetes**
Unclassified
**Unclassified**
Unclassified

**Shared**
S7-wms-combined_189250
**Caudoviricetes**
Unclassified
**Unclassified**
Unclassified
S7-wms-combined_190281
**n.a.**
n.a.
**n.a.**
n.a.
**SHHC**
S4-wms-combined_22012
**Caudoviricetes**
Unclassified
**O**
O
**d__Bacteria;p__Actinobacteriota;c__Coriobacteriia;o__Coriobacteriales;f__Eggerthellaceae;g__Adlercreutzia**
**SHHC**
S12-wms_184276
**Caudoviricetes**
Unclassified
**O**
O
**d__Bacteria;p__Actinobacteriota;c__Coriobacteriia;o__Coriobacteriales;f__Eggerthellaceae;g__CAG-1427**
**SHHC**
S4-wms_107182
**n.a.**
n.a.
**n.a.**
n.a.
**d__Bacteria;p__Actinobacteriota;c__Coriobacteriia;o__Coriobacteriales;f__Eggerthellaceae;g__Eggerthella**
**ME**
S17-wms-combined_225356
**n.a.**
n.a.
**n.a.**
n.a.
S5-wms_62679
**Caudoviricetes**
Unclassified
**O**
O
**SHHC**
S2-wms_79304
**Caudoviricetes**
Unclassified
**Unclassified**
Dybvigvirus
**d__Bacteria;p__Bacteroidota;c__Bacteroidia;o__Bacteroidales;f__Bacteroidaceae;g__Bacteroides**
**ME**

S11-wms_320555
    **Caudoviricetes**
        Unclassified
            **O**
                O
S11-wms-combined_195425
    **Caudoviricetes**
        Unclassified
            **Unclassified**
                Unclassified
S11-wms-combined_41367
    **n.a.**
        n.a.
            **n.a.**
                n.a.
S17-wms_124308
    **Caudoviricetes**
        Unclassified
            **O**
                O
S17-wms-combined_158354
    **n.a.**
        n.a.
            **n.a.**
                n.a.
S17-wms-combined_16309
    **Caudoviricetes**
        Unclassified
            **O**
                O
S1-wms_208875
    **Caudoviricetes**
        Unclassified
            **O**
                O
S1-wms-combined_330329
    **Caudoviricetes**
        Unclassified
            **O**
                O
S7-wms_244866
    **Caudoviricetes**
        Unclassified
            **Unclassified**
                Unclassified
S7-wms-combined_135154
    **n.a.**
        n.a.
            **n.a.**
                n.a.
S7-wms-combined_3669

**Malgrandaviricetes**
Microviridae
**O**
O
S7-wms-combined_90393
**Caudoviricetes**
Unclassified
**O**
O
S7-wms-combined_90728
**Caudoviricetes**
Unclassified
**O**
O
S9-wms-combined_33229
**Caudoviricetes**
Unclassified
**O**
O
**Shared**
S11-wms-combined_217790
**n.a.**
n.a.
**n.a.**
n.a.
S11-wms-combined_321938
**Caudoviricetes**
Unclassified
**O**
O
S11-wms-combined_73204
**n.a.**
n.a.
**n.a.**
n.a.
S12-wms-combined_111524
**n.a.**
n.a.
**n.a.**
n.a.
S12-wms-combined_190114
**Caudoviricetes**
Unclassified
**O**
O
S12-wms-combined_201125
**Caudoviricetes**
Unclassified
**O**
O
S12-wms-combined_250551

**Caudoviricetes**
Unclassified
**Unclassified**
Unclassified
S12-wms-combined_297545
**Caudoviricetes**
Unclassified
**O**
O
S12-wms-combined_320727
**Caudoviricetes**
Unclassified
**O**
O
S12-wms-combined_368190
**Caudoviricetes**
Unclassified
**O**
O
S12-wms-combined_368562
**Caudoviricetes**
Unclassified
**O**
O
S17-wms-combined_106940
**Caudoviricetes**
Unclassified
**O**
O
S17-wms-combined_146050
**n.a.**
n.a.
**n.a.**
n.a.
S17-wms-combined_179771
**n.a.**
n.a.
**n.a.**
n.a.
S17-wms-combined_233761
**n.a.**
n.a.
**n.a.**
n.a.
S17-wms-combined_70983
**n.a.**
n.a.
**n.a.**
n.a.
S1-wms_244836
**n.a.**

n.a.
**n.a.**
n.a.
S1-wms-combined_248622
**n.a.**
n.a.
**n.a.**
n.a.
S1-wms-combined_263562
**n.a.**
n.a.
**n.a.**
n.a.
S2-wms_156439
**Caudoviricetes**
Unclassified
**O**
O
S2-wms-combined_143737
**Caudoviricetes**
Duneviridae
**O**
O
S2-wms-combined_147065
**Caudoviricetes**
Unclassified
**O**
O
S2-wms-combined_210822
**n.a.**
n.a.
**n.a.**
n.a.
S2-wms-combined_229529
**Caudoviricetes**
Unclassified
**O**
O
S2-wms-combined_58121
**n.a.**
n.a.
**n.a.**
n.a.
S3-wms-combined_73262
**Caudoviricetes**
Unclassified
**Unclassified**
Unclassified
S4-wms-combined_83492
**Caudoviricetes**
Unclassified

**O**
O
S5-wms_85650
**n.a.**
n.a.
**n.a.**
n.a.
S5-wms-combined_207291
**n.a.**
n.a.
**n.a.**
n.a.
S5-wms-combined_59064
**Caudoviricetes**
Unclassified
**Unclassified**
Unclassified
S5-wms-combined_83370
**Caudoviricetes**
Unclassified
**O**
O
S5-wms-combined_98202
**n.a.**
n.a.
**n.a.**
n.a.
S6-wms-combined_135223
**n.a.**
n.a.
**n.a.**
n.a.
S6-wms-combined_82597
**n.a.**
n.a.
**n.a.**
n.a.
S7-wms-combined_104872
**n.a.**
n.a.
**n.a.**
n.a.
S7-wms-combined_107679_1
**Caudoviricetes**
Unclassified
**O**
O
S7-wms-combined_185016
**Caudoviricetes**
Unclassified
**O**

O
S7-wms-combined_239784
**n.a.**
n.a.
**n.a.**
n.a.
S7-wms-combined_43950
**Caudoviricetes**
Unclassified
**Unclassified**
Unclassified
S7-wms-combined_5625
**Caudoviricetes**
Unclassified
**Unclassified**
Unclassified
S7-wms-combined_62119
**Caudoviricetes**
Unclassified
**Unclassified**
O
S7-wms-combined_72070
**n.a.**
n.a.
**n.a.**
n.a.
S8-wms-combined_81597
**Caudoviricetes**
Unclassified
**O**
O
S9-wms-combined_33230
**Caudoviricetes**
Unclassified
**O**
O
S9-wms-combined_37587
**Caudoviricetes**
Unclassified
**O**
O
S9-wms-combined_44402
**Caudoviricetes**
Unclassified
**O**
O
**SHHC**
S12-wms-combined_91231
**n.a.**
n.a.
**n.a.**

n.a.
S2-wms_6424
**Caudoviricetes**
Pachyviridae
**O**
O
S2-wms-combined_132966
**Faserviricetes**
Inoviridae
**Unclassified**
Inovirus
S8-wms-combined_239791
**Caudoviricetes**
Aggregaviridae
**O**
O
S8-wms-combined_97848
**Caudoviricetes**
Unclassified
**Unclassified**
Unclassified
**d__Bacteria;p__Bacteroidota;c__Bacteroidia;o__Bacteroidales;f__Bacteroidaceae;g__Bacteroides_G**
**ME**
S17-wms_254872
**Malgrandaviricetes**
Microviridae
**Unclassified**
Microvirus
**d__Bacteria;p__Bacteroidota;c__Bacteroidia;o__Bacteroidales;f__Bacteroidaceae;g__Prevotella**
**Shared**
S2-wms_128962
**Caudoviricetes**
Suoliviridae
**O**
O
**SHHC**
S4-wms_141607
**Caudoviricetes**
Unclassified
**Unclassified**
Unclassified
S4-wms_45128
**n.a.**
n.a.
**n.a.**
n.a.
S4-wms_50475
**Caudoviricetes**
Unclassified
**O**
O

S4-wms-combined_47565
**n.a.**
n.a.
**n.a.**
n.a.
S4-wms-combined_57123
**n.a.**
n.a.
**n.a.**
n.a.
S6-wms_138342
**Caudoviricetes**
Suoliviridae
**O**
O
S8-wms-combined_74275
**Caudoviricetes**
Unclassified
**O**
O

**d__Bacteria;p__Bacteroidota;c__Bacteroidia;o__Bacteroidales;f__Bacteroidaceae;g__Prevotellamassilia**

**Shared**
S12-wms_12658
**Caudoviricetes**
Unclassified
**O**
O

**d__Bacteria;p__Bacteroidota;c__Bacteroidia;o__Bacteroidales;f__Barnesiellaceae;g__Barnesiella**

**ME**
S1-wms_153615
**n.a.**
n.a.
**n.a.**
n.a.
S1-wms-combined_153670
**n.a.**
n.a.
**n.a.**
n.a.
S1-wms-combined_357373
**Malgrandaviricetes**
Microviridae
**Unclassified**
Unclassified
**Shared**
S12-wms-combined_260036
**Caudoviricetes**
Steigviridae
**O**
O
S12-wms-combined_351847

**n.a.**
n.a.
**n.a.**
n.a.
S9-wms-combined_31456
**Caudoviricetes**
Unclassified
**Unclassified**
Unclassified
**SHHC**
S12-wms-combined_75733
**Malgrandaviricetes**
Microviridae
**O**
O
S8-wms_270
**Malgrandaviricetes**
Microviridae
**O**
O
**d__Bacteria;p__Bacteroidota;c__Bacteroidia;o__Bacteroidales;f__Muribaculaceae;g__UBA7173**
**SHHC**
S12-wms-combined_256299
**n.a.**
n.a.
**n.a.**
n.a.
**d__Bacteria;p__Bacteroidota;c__Bacteroidia;o__Bacteroidales;f__Rikenellaceae;g__Alistipes**
**ME**
S11-wms_330479
**n.a.**
Unclassified
**Unclassified**
Unclassified
S11-wms-combined_172889
**Caudoviricetes**
Unclassified
**O**
O
S17-wms-combined_253499
**n.a.**
n.a.
**n.a.**
n.a.
S7-wms-combined_163980
**n.a.**
n.a.
**n.a.**
n.a.
S9-wms-combined_28805
**Caudoviricetes**

Winoviridae
**O**
O
**Shared**
S11-wms-combined_125176
**Caudoviricetes**
Unclassified
**O**
O
S11-wms-combined_215723
**Caudoviricetes**
Duneviridae
**O**
O
S11-wms-combined_226165
**Caudoviricetes**
Unclassified
**O**
O
S11-wms-combined_48246
**Caudoviricetes**
Unclassified
**O**
O
S11-wms-combined_63818
**Caudoviricetes**
Unclassified
**O**
O
S11-wms-combined_64392
**Caudoviricetes**
Unclassified
**O**
O
S11-wms-combined_83571
**Caudoviricetes**
Winoviridae
**O**
O
S11-wms-combined_92114
**n.a.**
n.a.
**n.a.**
n.a.
S12-wms_386692
**n.a.**
n.a.
**n.a.**
n.a.
S12-wms-combined_189323
**Caudoviricetes**

Unclassified
**O**
O
S12-wms-combined_25459
**n.a.**
n.a.
**n.a.**
n.a.
S12-wms-combined_320262
**n.a.**
n.a.
**n.a.**
n.a.
S12-wms-combined_328554
**Caudoviricetes**
Unclassified
**O**
O
S12-wms-combined_358797
**Caudoviricetes**
Unclassified
**O**
O
S17-wms-combined_199746
**Caudoviricetes**
Unclassified
**O**
O
S1-wms_394571
**Caudoviricetes**
Unclassified
**O**
O
S1-wms-combined_107766
**n.a.**
n.a.
**n.a.**
n.a.
S1-wms-combined_340558
**Caudoviricetes**
Unclassified
**O**
O
S2-wms-combined_113619
**Caudoviricetes**
Unclassified
**O**
O
S2-wms-combined_208559
**n.a.**
n.a.

**n.a.**
n.a.
S4-wms-combined_41490
**Caudoviricetes**
Unclassified
**O**
O
S5-wms-combined_178432
**n.a.**
n.a.
**n.a.**
n.a.
S5-wms-combined_210959
**Caudoviricetes**
Unclassified
**O**
O
S5-wms-combined_67650
**Caudoviricetes**
Unclassified
**O**
O
S5-wms-combined_90315
**Caudoviricetes**
Unclassified
**O**
O
S6-wms-combined_50721
**Caudoviricetes**
Winoviridae
**O**
O
S6-wms-combined_72228
**n.a.**
n.a.
**n.a.**
n.a.
S6-wms-combined_91039
**n.a.**
n.a.
**n.a.**
n.a.
S7-wms-combined_107679_0
**Caudoviricetes**
Winoviridae
**O**
O
S7-wms-combined_14267
**n.a.**
n.a.
**n.a.**

n.a.
S7-wms-combined_26217
**Caudoviricetes**
Unclassified
**O**
O
S7-wms-combined_3697
**n.a.**
Unclassified
**O**
O
S8-wms_58515
**n.a.**
n.a.
**n.a.**
n.a.
S8-wms-combined_26087
**Caudoviricetes**
Unclassified
**O**
O
S8-wms-combined_270362
**Caudoviricetes**
Unclassified
**O**
O
S9-wms-combined_5526
**Caudoviricetes**
Unclassified
**O**
O
**d__Bacteria;p__Bacteroidota;c__Bacteroidia;o__Bacteroidales;f__Rikenellaceae;g__Alistipes_A**
**ME**
S5-wms-combined_183113
**n.a.**
n.a.
**n.a.**
n.a.
S5-wms-combined_204782
**n.a.**
n.a.
**n.a.**
n.a.
**Shared**
S8-wms-combined_23583
**n.a.**
n.a.
**n.a.**
n.a.
**d__Bacteria;p__Bacteroidota;c__Bacteroidia;o__Bacteroidales;f__Rikenellaceae;g__Tidjanibacter**
**Shared**

S12-wms-combined_2450
**Caudoviricetes**
Winoviridae
**O**
O
**d__Bacteria;p__Bacteroidota;c__Bacteroidia;o__Bacteroidales;f__Tannerellaceae;g__Marseille-P3160**
**SHHC**
S4-wms-combined_14411
**Caudoviricetes**
Unclassified
**Unclassified**
Unclassified
S6-wms_54578
**Caudoviricetes**
Unclassified
**Unclassified**
Unclassified
**d__Bacteria;p__Bacteroidota;c__Bacteroidia;o__Bacteroidales;f__Tannerellaceae;g__Parabacteroides**
**ME**
S17-wms-combined_171305
**Caudoviricetes**
Unclassified
**O**
O
S5-wms-combined_94482
**n.a.**
n.a.
**n.a.**
n.a.
S7-wms_123954
**n.a.**
n.a.
**n.a.**
n.a.
**Shared**
S1-wms-combined_32863
**n.a.**
n.a.
**n.a.**
n.a.
S1-wms-combined_394574
**Caudoviricetes**
Unclassified
**Unclassified**
Unclassified
S2-wms-combined_75144
**n.a.**
n.a.
**n.a.**
n.a.
**SHHC**

S4-wms_73990
**Caudoviricetes**
Suoliviridae
**Boorivirinae**
Cohcovirus
S4-wms-combined_10758
**Caudoviricetes**
Unclassified
**O**
O
S8-wms-combined_180957
**Caudoviricetes**
Unclassified
**O**
O

**d__Bacteria;p__Bacteroidota;c__Bacteroidia;o__Bacteroidales;f__UBA11471;g__UBA11471**

**ME**
S1-wms-combined_246923
**Caudoviricetes**
Unclassified
**O**
O
**Shared**
S12-wms-combined_371541
**n.a.**
n.a.
**n.a.**
n.a.
S12-wms-combined_375343
**n.a.**
n.a.
**n.a.**
n.a.

**d__Bacteria;p__Campylobacterota;c__Campylobacteria;o__Campylobacterales;f__Campylobacteraceae;g__Campylobacter_D**

**ME**
S5-wms-combined_106597
**n.a.**
n.a.
**n.a.**
n.a.

**d__Bacteria;p__Cyanobacteria;c__Vampirovibrionia;o__Gastranaerophilales;f__Gastranaerophilaceae;g__QAMI01**

**ME**
S5-wms-combined_199135
**n.a.**
n.a.
**n.a.**
n.a.

**d__Bacteria;p__Desulfobacterota;c__Desulfovibrionia;o__Desulfovibrionales;f__Desulfovibrionaceae;g__Bilophila**

**ME**
S1-wms-combined_143459
**Caudoviricetes**

Unclassified
**O**
O
S1-wms-combined_159544
**Caudoviricetes**
Peduoviridae
**Unclassified**
Phitrevirus
S1-wms-combined_19031
**Caudoviricetes**
Unclassified
**Unclassified**
Unclassified
S1-wms-combined_231411
**n.a.**
n.a.
**n.a.**
n.a.
**Shared**
S1-wms-combined_360285
**Caudoviricetes**
Unclassified
**O**
O
**SHHC**
S6-wms-combined_119017
**Caudoviricetes**
Peduoviridae
**Unclassified**
Phitrevirus
**d__Bacteria;p__Desulfobacterota;c__Desulfovibrionia;o__Desulfovibrionales;f__Desulfovibrionaceae;g__Desulfovibrio**
**ME**
S11-wms-combined_268967
**Caudoviricetes**
Unclassified
**Unclassified**
Unclassified
S11-wms-combined_84326
**Caudoviricetes**
Unclassified
**O**
O
**Shared**
S11-wms-combined_112733
**Caudoviricetes**
Unclassified
**O**
O
**d__Bacteria;p__Firmicutes;c__Bacilli;o__Erysipelotrichales;f__Erysipelatoclostridiaceae;g__Erysipelatoclostridium**
**ME**
S7-wms-combined_169375

**n.a.**
n.a.
**n.a.**
n.a.
**d__Bacteria;p__Firmicutes;c__Bacilli;o__Erysipelotrichales;f__Erysipelotrichaceae;g__Clostridium_AQ**
**ME**
S5-wms-combined_156329
**Caudoviricetes**
Unclassified
**O**
O
S5-wms-combined_197186
**n.a.**
n.a.
**n.a.**
n.a.
**d__Bacteria;p__Firmicutes;c__Bacilli;o__Lactobacillales;f__Lactobacillaceae;g__Lacticaseibacillus**
**ME**
S5-wms_212596
**n.a.**
n.a.
**n.a.**
n.a.
**d__Bacteria;p__Firmicutes;c__Bacilli;o__Lactobacillales;f__Streptococcaceae;g__Streptococcus**
**ME**
S9-wms_19025
**Caudoviricetes**
Unclassified
**O**
O
S9-wms-combined_20485
**Caudoviricetes**
Unclassified
**Unclassified**
O
**d__Bacteria;p__Firmicutes;c__Clostridia;o__Christensenellales;f__CAG-138;g__SFEL01**
**SHHC**
S12-wms-combined_147628
**Caudoviricetes**
Unclassified
**O**
O
S8-wms-combined_164289
**n.a.**
n.a.
**n.a.**
n.a.
S8-wms-combined_264711
**n.a.**
n.a.
**n.a.**

n.a.

**d__Bacteria;p__Firmicutes;c__Clostridia;o__Christensenellales;f__CAG-138;g__UBA1685**

**SHHC**

S8-wms_57121

**n.a.**

n.a.

**n.a.**

n.a.

S8-wms-combined_146562

**Caudoviricetes**

Unclassified

**O**

O

S8-wms-combined_211119

**n.a.**

n.a.

**n.a.**

n.a.

S8-wms-combined_219796

**Caudoviricetes**

Unclassified

**Unclassified**

O

**d__Bacteria;p__Firmicutes;c__Clostridia;o__Christensenellales;f__CAG-74;g__Firm-11**

**ME**

S11-wms-combined_267993

**Caudoviricetes**

Unclassified

**O**

O

**d__Bacteria;p__Firmicutes;c__Clostridia;o__Christensenellales;f__CAG-74;g__Limiplasma**

**Shared**

S8-wms-combined_185611

**n.a.**

n.a.

**n.a.**

n.a.

**d__Bacteria;p__Firmicutes;c__Clostridia;o__Christensenellales;f__CAG-74;g__SFFH01**

**Shared**

S12-wms-combined_299293

**n.a.**

n.a.

**n.a.**

n.a.

**d__Bacteria;p__Firmicutes;c__Clostridia;o__Christensenellales;f__CAG-74;g__UBA11524**

**Shared**

S12-wms-combined_151843

**Caudoviricetes**

Unclassified

**O**

O

S12-wms-combined_293354
**Caudoviricetes**
Unclassified
**O**
O
S1-wms_223620
**Caudoviricetes**
Unclassified
**O**
O
S1-wms-combined_302166
**Caudoviricetes**
Unclassified
**O**
O
S7-wms-combined_134452
**n.a.**
n.a.
**n.a.**
n.a.
S7-wms-combined_14529
**Caudoviricetes**
Unclassified
**O**
O
S8-wms-combined_215102
**n.a.**
n.a.
**n.a.**
n.a.
**d__Bacteria;p__Firmicutes;c__Clostridia;o__Christensenellales;f__CAG-74;g__Ventricola**
**Shared**
S2-wms-combined_25727
**Caudoviricetes**
Unclassified
**O**
O
**d__Bacteria;p__Firmicutes;c__Clostridia;o__Christensenellales;f__CAG-917;g__**
**SHHC**
S12-wms-combined_117447
**n.a.**
n.a.
**n.a.**
n.a.
**d__Bacteria;p__Firmicutes;c__Clostridia;o__Christensenellales;f__CAG-917;g__CAG-475**
**SHHC**
S12-wms-combined_164656
**n.a.**
n.a.
**n.a.**
n.a.

**d__Bacteria;p__Firmicutes;c__Clostridia;o__Christensenellales;f__Christensenellaceae;g__Christensenella**
**ME**
S5-wms-combined_66017
**n.a.**
n.a.
**n.a.**
n.a.
**d__Bacteria;p__Firmicutes;c__Clostridia;o__Clostridiales;f__Clostridiaceae;g__Clostridium**
**Shared**
S17-wms-combined_258087
**Caudoviricetes**
Unclassified
**Unclassified**
O
S2-wms-combined_58563
**Caudoviricetes**
Unclassified
**Unclassified**
O
**d__Bacteria;p__Firmicutes;c__Clostridia;o__Eubacteriales;f__Eubacteriaceae;g__Eubacterium**
**ME**
S5-wms-combined_83412
**Caudoviricetes**
Unclassified
**Unclassified**
Unclassified
**d__Bacteria;p__Firmicutes;c__Clostridia;o__Lachnospirales;f__Anaerotignaceae;g__Anaerotignum**
**ME**
S9-wms-combined_3461
**n.a.**
n.a.
**n.a.**
n.a.
**Shared**
S2-wms-combined_116221
**Caudoviricetes**
Unclassified
**O**
O
S2-wms-combined_191613
**n.a.**
n.a.
**n.a.**
n.a.
**d__Bacteria;p__Firmicutes;c__Clostridia;o__Lachnospirales;f__Lachnospiraceae;g__Acetatifactor**
**SHHC**
S2-wms-combined_126440
**Caudoviricetes**
Unclassified
**O**
O

S6-wms-combined_117756
**Caudoviricetes**
Unclassified
**Unclassified**
Eponavirus
**d__Bacteria;p__Firmicutes;c__Clostridia;o__Lachnospirales;f__Lachnospiraceae;g__Acetivibrio_A**
**SHHC**
S2-wms-combined_31084
**Caudoviricetes**
Unclassified
**O**
O
**d__Bacteria;p__Firmicutes;c__Clostridia;o__Lachnospirales;f__Lachnospiraceae;g__Agathobacter**
**ME**
S7-wms-combined_235165
**Caudoviricetes**
Unclassified
**Unclassified**
Unclassified
**Shared**
S2-wms-combined_5791
**Caudoviricetes**
Unclassified
**O**
O
**SHHC**
S2-wms-combined_207000
**Caudoviricetes**
Unclassified
**Unclassified**
O
S2-wms-combined_222535
**n.a.**
n.a.
**n.a.**
n.a.
**d__Bacteria;p__Firmicutes;c__Clostridia;o__Lachnospirales;f__Lachnospiraceae;g__Anaerobutyricum**
**ME**
S1-wms-combined_212892
**Caudoviricetes**
Unclassified
**O**
O
S7-wms_151112
**Caudoviricetes**
Unclassified
**O**
O
S7-wms-combined_20921
**Caudoviricetes**
Unclassified

**O**
O
**d__Bacteria;p__Firmicutes;c__Clostridia;o__Lachnospirales;f__Lachnospiraceae;g__Anaerosacchariphilus**
**SHHC**
S2-wms-combined_48661
**n.a.**
n.a.
**n.a.**
n.a.
**d__Bacteria;p__Firmicutes;c__Clostridia;o__Lachnospirales;f__Lachnospiraceae;g__Anaerostipes**
**ME**
S9-wms-combined_105462
**Caudoviricetes**
Unclassified
**O**
O
**d__Bacteria;p__Firmicutes;c__Clostridia;o__Lachnospirales;f__Lachnospiraceae;g__Bariatricus**
**ME**
S7-wms-combined_69024
**Caudoviricetes**
Unclassified
**O**
O
**d__Bacteria;p__Firmicutes;c__Clostridia;o__Lachnospirales;f__Lachnospiraceae;g__Blautia_A**
**ME**
S11-wms-combined_200084
**Caudoviricetes**
Unclassified
**O**
O
S7-wms-combined_156351
**n.a.**
n.a.
**n.a.**
n.a.
**Shared**
S2-wms_140933
**Caudoviricetes**
Unclassified
**Unclassified**
Unclassified
S5-wms-combined_26061
**Caudoviricetes**
Unclassified
**Unclassified**
O
S9-wms-combined_10990
**n.a.**
n.a.
**n.a.**
n.a.

**SHHC**
S2-wms-combined_176376
**Caudoviricetes**
Unclassified
**O**
O
S4-wms_62635
**Caudoviricetes**
Unclassified
**Unclassified**
Unclassified

**d__Bacteria;p__Firmicutes;c__Clostridia;o__Lachnospirales;f__Lachnospiraceae;g__CAG-81**

**SHHC**
S2-wms-combined_85381
**Caudoviricetes**
Unclassified
**Unclassified**
Eponavirus

**d__Bacteria;p__Firmicutes;c__Clostridia;o__Lachnospirales;f__Lachnospiraceae;g__COE1**

**SHHC**
S12-wms-combined_31761
**Caudoviricetes**
Unclassified
**Unclassified**
O

**d__Bacteria;p__Firmicutes;c__Clostridia;o__Lachnospirales;f__Lachnospiraceae;g__Dorea_A**

**ME**
S5-wms-combined_73301
**Caudoviricetes**
Unclassified
**O**
O
**Shared**
S7-wms-combined_106253
**Caudoviricetes**
Unclassified
**O**
O
**SHHC**
S4-wms-combined_64632
**Caudoviricetes**
Unclassified
**Unclassified**
Unclassified

**d__Bacteria;p__Firmicutes;c__Clostridia;o__Lachnospirales;f__Lachnospiraceae;g__Eisenbergiella**

**ME**
S9-wms-combined_2042
**n.a.**
n.a.
**n.a.**
n.a.

S9-wms-combined_97755
**Caudoviricetes**
Unclassified
**Unclassified**
O
**Shared**
S2-wms-combined_200390
**Caudoviricetes**
Unclassified
**O**
O
**d__Bacteria;p__Firmicutes;c__Clostridia;o__Lachnospirales;f__Lachnospiraceae;g__Enterocloster**
**ME**
S17-wms-combined_223748
**n.a.**
n.a.
**n.a.**
n.a.
S9-wms-combined_105822
**Caudoviricetes**
Unclassified
**Unclassified**
O
**Shared**
S2-wms-combined_158821
**Caudoviricetes**
Unclassified
**Unclassified**
O
S6-wms-combined_119138
**Caudoviricetes**
Unclassified
**O**
O
S9-wms_64472
**n.a.**
n.a.
**n.a.**
n.a.
**SHHC**
S12-wms-combined_223043
**Caudoviricetes**
Unclassified
**O**
O
S6-wms_140743
**Caudoviricetes**
Unclassified
**O**
O
**d__Bacteria;p__Firmicutes;c__Clostridia;o__Lachnospirales;f__Lachnospiraceae;g__Eubacterium_I**

**SHHC**
S2-wms_192667
**Caudoviricetes**
Unclassified
**Unclassified**
O
**d__Bacteria;p__Firmicutes;c__Clostridia;o__Lachnospirales;f__Lachnospiraceae;g__Faecalimonas**
**SHHC**
S2-wms-combined_73327
**n.a.**
n.a.
**n.a.**
n.a.
**d__Bacteria;p__Firmicutes;c__Clostridia;o__Lachnospirales;f__Lachnospiraceae;g__Fusicatenibacter**
**Shared**
S7-wms-combined_123085
**n.a.**
n.a.
**n.a.**
n.a.
**d__Bacteria;p__Firmicutes;c__Clostridia;o__Lachnospirales;f__Lachnospiraceae;g__Lachnoclostridium_A**
**SHHC**
S2-wms-combined_204225
**n.a.**
n.a.
**n.a.**
n.a.
S2-wms-combined_74387
**Caudoviricetes**
Unclassified
**O**
O
S2-wms-combined_95987
**n.a.**
n.a.
**n.a.**
n.a.
**d__Bacteria;p__Firmicutes;c__Clostridia;o__Lachnospirales;f__Lachnospiraceae;g__Lachnospira**
**ME**
S1-wms-combined_100430
**Caudoviricetes**
Unclassified
**Unclassified**
O
**Shared**
S12-wms-combined_120133
**Caudoviricetes**
Unclassified
**O**
O
S2-wms_161986

**Caudoviricetes**
Unclassified
**Unclassified**
O
S7-wms-combined_157260
**Caudoviricetes**
Unclassified
**Unclassified**
O
**SHHC**
S12-wms-combined_302876
**Caudoviricetes**
Unclassified
**O**
O
**d__Bacteria;p__Firmicutes;c__Clostridia;o__Lachnospirales;f__Lachnospiraceae;g__Mediterraneibacter**
**ME**
S11-wms-combined_282227
**Caudoviricetes**
Unclassified
**O**
O
S9-wms-combined_26410
**Caudoviricetes**
Unclassified
**O**
O
**Shared**
S4-wms-combined_121978
**Caudoviricetes**
Unclassified
**O**
O
S7-wms-combined_157434
**n.a.**
n.a.
**n.a.**
n.a.
**d__Bacteria;p__Firmicutes;c__Clostridia;o__Lachnospirales;f__Lachnospiraceae;g__Roseburia**
**Shared**
S12-wms-combined_259783
**Caudoviricetes**
Unclassified
**Unclassified**
Unclassified
S2-wms_246296
**n.a.**
n.a.
**n.a.**
n.a.
S2-wms-combined_157934

**Caudoviricetes**
Unclassified
**Unclassified**
O
S2-wms-combined_218002
**Caudoviricetes**
Unclassified
**O**
O
S6-wms-combined_32530
**Caudoviricetes**
Unclassified
**O**
O
**SHHC**
S2-wms-combined_165833
**Caudoviricetes**
Unclassified
**Unclassified**
Mushuvirus
S4-wms-combined_24659
**n.a.**
n.a.
**n.a.**
n.a.
S8-wms-combined_217866
**Caudoviricetes**
Unclassified
**Unclassified**
Eponavirus
**d__Bacteria;p__Firmicutes;c__Clostridia;o__Lachnospirales;f__Lachnospiraceae;g__UBA9502**
**Shared**
S11-wms-combined_179605
**n.a.**
n.a.
**n.a.**
n.a.
**d__Bacteria;p__Firmicutes;c__Clostridia;o__Oscillospirales;f__Acutalibacteraceae;g__Acutalibacter**
**ME**
S9-wms-combined_27311
**n.a.**
n.a.
**n.a.**
n.a.
**d__Bacteria;p__Firmicutes;c__Clostridia;o__Oscillospirales;f__Acutalibacteraceae;g__CAG-180**
**Shared**
S7-wms-combined_218379
**Caudoviricetes**
Unclassified
**O**
O

**d__Bacteria;p__Firmicutes;c__Clostridia;o__Oscillospirales;f__Acutalibacteraceae;g__CAG-217**

- **Shared**
  - S12-wms-combined_117628
    - **Caudoviricetes**
      - Unclassified
        - **Unclassified**
          - O
  - S2-wms-combined_225908
    - **n.a.**
      - Unclassified
        - **O**
          - O

**d__Bacteria;p__Firmicutes;c__Clostridia;o__Oscillospirales;f__Acutalibacteraceae;g__Ruminococcus_E**

- **ME**
  - S7-wms-combined_28138
    - **Caudoviricetes**
      - Unclassified
        - **O**
          - O
- **Shared**
  - S2-wms_130687
    - **Caudoviricetes**
      - Unclassified
        - **Unclassified**
          - Unclassified
  - S5-wms_49310
    - **Caudoviricetes**
      - Unclassified
        - **Unclassified**
          - Unclassified
  - S5-wms-combined_108794
    - **Caudoviricetes**
      - Unclassified
        - **Unclassified**
          - O
  - S5-wms-combined_7292
    - **Caudoviricetes**
      - Unclassified
        - **Unclassified**
          - Unclassified
  - S9-wms-combined_9931
    - **Caudoviricetes**
      - Unclassified
        - **Unclassified**
          - O

**d__Bacteria;p__Firmicutes;c__Clostridia;o__Oscillospirales;f__Acutalibacteraceae;g__UBA1417**

- **Shared**
  - S5-wms-combined_117095
    - **n.a.**
      - n.a.
        - **n.a.**

n.a.
**SHHC**
S12-wms-combined_263377
**Caudoviricetes**
Unclassified
**O**
O
S12-wms-combined_264425
**n.a.**
n.a.
**n.a.**
n.a.
**d__Bacteria;p__Firmicutes;c__Clostridia;o__Oscillospirales;f__Butyricicoccaceae;g__Agathobaculum**
**SHHC**
S2-wms-combined_199384
**Caudoviricetes**
Unclassified
**O**
O
**d__Bacteria;p__Firmicutes;c__Clostridia;o__Oscillospirales;f__Oscillospiraceae;g__**
**SHHC**
S2-wms-combined_64692
**Caudoviricetes**
Unclassified
**Unclassified**
O
**d__Bacteria;p__Firmicutes;c__Clostridia;o__Oscillospirales;f__Oscillospiraceae;g__An92**
**ME**
S11-wms-combined_119215
**Caudoviricetes**
Unclassified
**O**
O
S17-wms-combined_79709
**Caudoviricetes**
Unclassified
**O**
O
**Shared**
S2-wms-combined_137119
**Caudoviricetes**
Unclassified
**Unclassified**
O
S8-wms_82880
**Caudoviricetes**
Unclassified
**Unclassified**
Brigitvirus
**d__Bacteria;p__Firmicutes;c__Clostridia;o__Oscillospirales;f__Oscillospiraceae;g__CAG-103**
**Shared**

S7-wms-combined_139683
**Caudoviricetes**
Unclassified
**O**
O
S8-wms-combined_40666
**n.a.**
n.a.
**n.a.**
n.a.

**d__Bacteria;p__Firmicutes;c__Clostridia;o__Oscillospirales;f__Oscillospiraceae;g__CAG-110**

**ME**
S1-wms-combined_227003
**n.a.**
n.a.
**n.a.**
n.a.
**Shared**
S12-wms-combined_264845
**n.a.**
n.a.
**n.a.**
n.a.

**d__Bacteria;p__Firmicutes;c__Clostridia;o__Oscillospirales;f__Oscillospiraceae;g__CAG-170**

**ME**
S1-wms_130214
**Caudoviricetes**
Unclassified
**O**
O
**Shared**
S1-wms_185046
**n.a.**
n.a.
**n.a.**
n.a.
S2-wms-combined_238929
**Caudoviricetes**
Unclassified
**O**
O
S3-wms_23958
**Caudoviricetes**
Unclassified
**O**
O

**d__Bacteria;p__Firmicutes;c__Clostridia;o__Oscillospirales;f__Oscillospiraceae;g__CAG-83**

**ME**
S11-wms_14390
**n.a.**
n.a.

**n.a.**
n.a.
S1-wms-combined_72167
**Caudoviricetes**
Unclassified
**O**
O
**Shared**
S11-wms-combined_3507
**Caudoviricetes**
Unclassified
**O**
O
S12-wms_373775
**n.a.**
n.a.
**n.a.**
n.a.
S12-wms-combined_148741
**n.a.**
n.a.
**n.a.**
n.a.
S12-wms-combined_23517
**Caudoviricetes**
Unclassified
**O**
O
S12-wms-combined_270839
**n.a.**
n.a.
**n.a.**
n.a.
S12-wms-combined_334806
**n.a.**
n.a.
**n.a.**
n.a.
S12-wms-combined_63649
**n.a.**
n.a.
**n.a.**
n.a.
S1-wms-combined_169788
**n.a.**
n.a.
**n.a.**
n.a.
S1-wms-combined_373308
**n.a.**
n.a.

**n.a.**
n.a.
S1-wms-combined_57370
**Caudoviricetes**
Unclassified
**O**
O
S1-wms-combined_94988
**n.a.**
n.a.
**n.a.**
n.a.
S2-wms-combined_159670
**n.a.**
n.a.
**n.a.**
n.a.
S5-wms-combined_134381
**Caudoviricetes**
Unclassified
**Unclassified**
Lagaffevirus
S5-wms-combined_179241
**n.a.**
n.a.
**n.a.**
n.a.
S8-wms_30498
**Caudoviricetes**
Unclassified
**Unclassified**
Mushuvirus
S8-wms_48935
**Caudoviricetes**
Unclassified
**Unclassified**
Toutatisvirus
S8-wms-combined_169835
**n.a.**
n.a.
**n.a.**
n.a.
**SHHC**
S12-wms-combined_157167
**n.a.**
n.a.
**n.a.**
n.a.
**d__Bacteria;p__Firmicutes;c__Clostridia;o__Oscillospirales;f__Oscillospiraceae;g__Dysosmobacter**
**ME**
S11-wms-combined_283220

**Caudoviricetes**
Unclassified
**Unclassified**
Mushuvirus
S11-wms-combined_49632
**Caudoviricetes**
Unclassified
**O**
O
**Shared**
S11-wms-combined_116589
**n.a.**
n.a.
**n.a.**
n.a.
S12-wms-combined_184338
**Caudoviricetes**
Unclassified
**O**
O
S12-wms-combined_329242
**n.a.**
n.a.
**n.a.**
n.a.
S1-wms_102342
**Caudoviricetes**
Unclassified
**O**
O
S5-wms-combined_205046
**n.a.**
n.a.
**n.a.**
n.a.
S6-wms-combined_118427
**Caudoviricetes**
Unclassified
**Unclassified**
O
**d__Bacteria;p__Firmicutes;c__Clostridia;o__Oscillospirales;f__Oscillospiraceae;g__ER4**
**ME**
S1-wms-combined_185336
**n.a.**
n.a.
**n.a.**
n.a.
S1-wms-combined_186878
**n.a.**
n.a.
**n.a.**

n.a.
S1-wms-combined_353757
**Caudoviricetes**
Unclassified
**O**
O
S1-wms-combined_40115
**n.a.**
n.a.
**n.a.**
n.a.
S1-wms-combined_41369
**Caudoviricetes**
Unclassified
**O**
O
**Shared**
S12-wms-combined_129799
**Caudoviricetes**
Unclassified
**Unclassified**
O
S12-wms-combined_210942
**n.a.**
n.a.
**n.a.**
n.a.
S12-wms-combined_393703
**n.a.**
n.a.
**n.a.**
n.a.
S12-wms-combined_48648
**n.a.**
n.a.
**n.a.**
n.a.
S1-wms_236105
**Caudoviricetes**
Unclassified
**O**
O
S1-wms_296850
**Caudoviricetes**
Unclassified
**Unclassified**
Toutatisvirus
S1-wms-combined_132934
**Caudoviricetes**
Unclassified
**O**

O
S1-wms-combined_156637
**n.a.**
n.a.
**n.a.**
n.a.
S1-wms-combined_164485
**n.a.**
n.a.
**n.a.**
n.a.
S1-wms-combined_17651
**n.a.**
n.a.
**n.a.**
n.a.
S1-wms-combined_331896
**n.a.**
n.a.
**n.a.**
n.a.
S1-wms-combined_351889
**Caudoviricetes**
Unclassified
**Unclassified**
O
S2-wms-combined_140996
**Caudoviricetes**
Unclassified
**Unclassified**
O
S2-wms-combined_93845
**Caudoviricetes**
Unclassified
**O**
O
**SHHC**
S12-wms_235125
**n.a.**
n.a.
**n.a.**
n.a.
S8-wms-combined_134098
**n.a.**
n.a.
**n.a.**
n.a.
S8-wms-combined_246389
**Caudoviricetes**
Unclassified
**Unclassified**

O

**d__Bacteria;p__Firmicutes;c__Clostridia;o__Oscillospirales;f__Oscillospiraceae;g__Evtepia**

**ME**

S17-wms-combined_136622

**n.a.**

n.a.

**n.a.**

n.a.

**d__Bacteria;p__Firmicutes;c__Clostridia;o__Oscillospirales;f__Oscillospiraceae;g__F23-B02**

**Shared**

S2-wms-combined_52568

**n.a.**

n.a.

**n.a.**

n.a.

S8-wms-combined_68780

**n.a.**

n.a.

**n.a.**

n.a.

**SHHC**

S12-wms-combined_129554

**Caudoviricetes**

Unclassified

**Unclassified**

Eponavirus

**d__Bacteria;p__Firmicutes;c__Clostridia;o__Oscillospirales;f__Oscillospiraceae;g__Faecousia**

**Shared**

S11-wms-combined_329493

**n.a.**

n.a.

**n.a.**

n.a.

**d__Bacteria;p__Firmicutes;c__Clostridia;o__Oscillospirales;f__Oscillospiraceae;g__Flavonifractor**

**ME**

S17-wms-combined_146850

**n.a.**

n.a.

**n.a.**

n.a.

S17-wms-combined_157468

**n.a.**

n.a.

**n.a.**

n.a.

S17-wms-combined_175924

**Caudoviricetes**

Unclassified

**Unclassified**

Mushuvirus

S17-wms-combined_260343

**Caudoviricetes**
Unclassified
**Unclassified**
Toutatisvirus
S17-wms-combined_39687
**n.a.**
n.a.
**n.a.**
n.a.
S17-wms-combined_66123
**Caudoviricetes**
Unclassified
**Unclassified**
O
S1-wms-combined_374219
**n.a.**
n.a.
**n.a.**
n.a.
S5-wms-combined_163165
**n.a.**
n.a.
**n.a.**
n.a.
**Shared**
S11-wms-combined_120257
**Caudoviricetes**
Unclassified
**Unclassified**
O
S17-wms-combined_185239
**n.a.**
n.a.
**n.a.**
n.a.
S17-wms-combined_247494
**Caudoviricetes**
Unclassified
**O**
O
S2-wms-combined_6605
**n.a.**
n.a.
**n.a.**
n.a.
S5-wms-combined_145067
**Caudoviricetes**
Unclassified
**O**
O
S9-wms-combined_122742

**n.a.**
n.a.
**n.a.**
n.a.
S9-wms-combined_124857
**n.a.**
n.a.
**n.a.**
n.a.
**d__Bacteria;p__Firmicutes;c__Clostridia;o__Oscillospirales;f__Oscillospiraceae;g__HGM12998**
**ME**
S17-wms-combined_75892
**Caudoviricetes**
Unclassified
**Unclassified**
O
**d__Bacteria;p__Firmicutes;c__Clostridia;o__Oscillospirales;f__Oscillospiraceae;g__Intestinimonas**
**ME**
S11-wms-combined_299954
**n.a.**
n.a.
**n.a.**
n.a.
**Shared**
S2-wms-combined_122457
**Caudoviricetes**
Unclassified
**Unclassified**
O
S2-wms-combined_196783
**Caudoviricetes**
Unclassified
**O**
O
S2-wms-combined_24549
**Caudoviricetes**
Unclassified
**O**
O
S2-wms-combined_72725
**n.a.**
n.a.
**n.a.**
n.a.
S5-wms_52566
**n.a.**
n.a.
**n.a.**
n.a.
**d__Bacteria;p__Firmicutes;c__Clostridia;o__Oscillospirales;f__Oscillospiraceae;g__Lawsonibacter**
**Shared**

S17-wms-combined_259811
**Caudoviricetes**
Unclassified
**O**
O
S1-wms_4277
**Caudoviricetes**
Unclassified
**Unclassified**
Mushuvirus
S2-wms-combined_126134
**n.a.**
n.a.
**n.a.**
n.a.
S5-wms_10783
**n.a.**
n.a.
**n.a.**
n.a.
S7-wms-combined_31068
**n.a.**
n.a.
**n.a.**
n.a.
**d__Bacteria;p__Firmicutes;c__Clostridia;o__Oscillospirales;f__Oscillospiraceae;g__Marseille-P3106**
**ME**
S17-wms-combined_162786
**n.a.**
n.a.
**n.a.**
n.a.
**Shared**
S6-wms-combined_41246
**n.a.**
n.a.
**n.a.**
n.a.
S7-wms-combined_84688
**n.a.**
n.a.
**n.a.**
n.a.
**d__Bacteria;p__Firmicutes;c__Clostridia;o__Oscillospirales;f__Oscillospiraceae;g__Pseudoflavonifractor**
**ME**
S11-wms_32691
**n.a.**
n.a.
**n.a.**
n.a.
**d__Bacteria;p__Firmicutes;c__Clostridia;o__Oscillospirales;f__Oscillospiraceae;g__UBA738**

**Shared**
S1-wms-combined_136097
**n.a.**
n.a.
**n.a.**
n.a.
**d__Bacteria;p__Firmicutes;c__Clostridia;o__Oscillospirales;f__Oscillospiraceae;g__UBA9475**
**SHHC**
S8-wms-combined_25607
**Caudoviricetes**
Unclassified
**O**
O
**d__Bacteria;p__Firmicutes;c__Clostridia;o__Oscillospirales;f__Ruminococcaceae;g__Faecalibacterium**
**ME**
S17-wms-combined_90378
**Caudoviricetes**
Unclassified
**Unclassified**
Lughvirus
S1-wms_29288
**Caudoviricetes**
Unclassified
**O**
O
S1-wms-combined_131270
**Caudoviricetes**
Unclassified
**O**
O
S1-wms-combined_164894
**Caudoviricetes**
Unclassified
**O**
O
S1-wms-combined_68998
**Caudoviricetes**
Unclassified
**Unclassified**
Taranisvirus
**Shared**
S11-wms_3116
**Caudoviricetes**
Unclassified
**Unclassified**
Mushuvirus
S11-wms-combined_140009
**Caudoviricetes**
Unclassified
**Unclassified**
Lughvirus

S11-wms-combined_236214
**Caudoviricetes**
Unclassified
**O**
O
S17-wms-combined_193958
**Caudoviricetes**
Unclassified
**Unclassified**
Mushuvirus
S17-wms-combined_56006
**Caudoviricetes**
Unclassified
**O**
O
S17-wms-combined_56061
**n.a.**
n.a.
**n.a.**
n.a.
S1-wms-combined_118167
**Caudoviricetes**
Unclassified
**O**
O
S1-wms-combined_23248
**Caudoviricetes**
Unclassified
**Unclassified**
Lughvirus
S1-wms-combined_285012
**Caudoviricetes**
Unclassified
**Unclassified**
O
S1-wms-combined_304377
**Caudoviricetes**
Unclassified
**Unclassified**
Oengusvirus
S1-wms-combined_52708
**n.a.**
n.a.
**n.a.**
n.a.
S2-wms_124416
**Caudoviricetes**
Unclassified
**O**
O
S2-wms-combined_13246

**Caudoviricetes**
Unclassified
**Unclassified**
O
S2-wms-combined_166665
**Caudoviricetes**
Unclassified
**O**
O
S2-wms-combined_181849
**n.a.**
n.a.
**n.a.**
n.a.
S2-wms-combined_57293
**n.a.**
Unclassified
**O**
O
S3-wms_22744
**Caudoviricetes**
Unclassified
**Unclassified**
Lagaffevirus
S4-wms-combined_42647
**Caudoviricetes**
Unclassified
**Unclassified**
Lughvirus
S6-wms_132588
**Caudoviricetes**
Unclassified
**O**
O
S6-wms-combined_110950
**n.a.**
n.a.
**n.a.**
n.a.
S6-wms-combined_126236
**Caudoviricetes**
Unclassified
**Unclassified**
Toutatisvirus
S6-wms-combined_138710
**n.a.**
n.a.
**n.a.**
n.a.
S6-wms-combined_56464
**Caudoviricetes**

Unclassified
**O**
O
S7-wms_113137
**Caudoviricetes**
Unclassified
**Unclassified**
Mushuvirus
S7-wms-combined_111004
**Caudoviricetes**
Unclassified
**Unclassified**
Taranisvirus
S7-wms-combined_137858
**n.a.**
n.a.
**n.a.**
n.a.
**SHHC**
S12-wms_193565
**Caudoviricetes**
Unclassified
**Unclassified**
Mushuvirus
S12-wms-combined_264077
**Caudoviricetes**
Unclassified
**O**
O
S12-wms-combined_345948
**Caudoviricetes**
Unclassified
**Unclassified**
Lughvirus
S12-wms-combined_71235
**Caudoviricetes**
Unclassified
**Unclassified**
Lughvirus
S2-wms-combined_127080
**n.a.**
n.a.
**n.a.**
n.a.
S6-wms-combined_140731
**n.a.**
n.a.
**n.a.**
n.a.
S6-wms-combined_49909
**Caudoviricetes**

Unclassified
**O**
O
S8-wms_152831
**Caudoviricetes**
Unclassified
**Unclassified**
Toutatisvirus
S8-wms_190781
**Caudoviricetes**
Unclassified
**O**
O
S8-wms-combined_225114
**Caudoviricetes**
Unclassified
**O**
O
**d__Bacteria;p__Firmicutes;c__Clostridia;o__Oscillospirales;f__Ruminococcaceae;g__Gemmiger**
**ME**
S17-wms-combined_72837
**Caudoviricetes**
Unclassified
**O**
O
S1-wms-combined_166392
**Caudoviricetes**
Unclassified
**O**
O
S1-wms-combined_220022
**n.a.**
n.a.
**n.a.**
n.a.
S1-wms-combined_227917
**Caudoviricetes**
Unclassified
**O**
O
**Shared**
S11-wms-combined_264050
**Caudoviricetes**
Unclassified
**Unclassified**
O
S11-wms-combined_46816
**n.a.**
n.a.
**n.a.**
n.a.

S1-wms-combined_279492
**Caudoviricetes**
Unclassified
**O**
O
S2-wms-combined_62839
**Caudoviricetes**
Unclassified
**O**
O
S3-wms-combined_45305
**Caudoviricetes**
Unclassified
**O**
O
S4-wms-combined_113721
**n.a.**
n.a.
**n.a.**
n.a.
S7-wms-combined_134458
**Caudoviricetes**
Unclassified
**Unclassified**
Toutatisvirus
S8-wms-combined_49369
**n.a.**
n.a.
**n.a.**
n.a.
**SHHC**
S12-wms-combined_323943
**Caudoviricetes**
Unclassified
**Unclassified**
O
S8-wms_20528
**Caudoviricetes**
Unclassified
**Unclassified**
O
**d__Bacteria;p__Firmicutes;c__Clostridia;o__Oscillospirales;f__Ruminococcaceae;g__Negativibacillus**
**ME**
S9-wms-combined_36459
**Caudoviricetes**
Unclassified
**O**
O
**Shared**
S7-wms-combined_238231
**n.a.**

n.a.
**n.a.**
n.a.
**d__Bacteria;p__Firmicutes;c__Clostridia;o__Oscillospirales;f__Ruminococcaceae;g__Ruminiclostridium_E**
**ME**
S7-wms_80255
**Caudoviricetes**
Unclassified
**O**
O
**Shared**
S11-wms-combined_168048
**Caudoviricetes**
Unclassified
**O**
O
**d__Bacteria;p__Firmicutes;c__Clostridia;o__Oscillospirales;f__Ruminococcaceae;g__Ruminococcus_D**
**Shared**
S2-wms-combined_101826
**Caudoviricetes**
Unclassified
**Unclassified**
Unclassified
S7-wms_127801
**Caudoviricetes**
Unclassified
**O**
O
S7-wms-combined_224523
**Caudoviricetes**
Unclassified
**O**
O
S7-wms-combined_71592
**Caudoviricetes**
Unclassified
**O**
O
**SHHC**
S12-wms_223235
**n.a.**
n.a.
**n.a.**
n.a.
S2-wms-combined_30750
**n.a.**
n.a.
**n.a.**
n.a.
**d__Bacteria;p__Firmicutes;c__Clostridia;o__Oscillospirales;f__Ruminococcaceae;g__Ruthenibacterium**
**ME**

S9-wms_104169
**Faserviricetes**
Inoviridae
**Unclassified**
Inovirus
**Shared**
S5-wms-combined_66662
**n.a.**
Unclassified
**Unclassified**
O
S5-wms-combined_72410
**Caudoviricetes**
Unclassified
**Unclassified**
O
**d__Bacteria;p__Firmicutes;c__Clostridia;o__Oscillospirales;f__Ruminococcaceae;g__UBA3818**
**SHHC**
S12-wms-combined_52101
**Caudoviricetes**
Unclassified
**Unclassified**
O
**d__Bacteria;p__Firmicutes;c__Clostridia;o__Oscillospirales;f__Ruminococcaceae;g__UMGS363**
**Shared**
S12-wms_336780
**n.a.**
n.a.
**n.a.**
n.a.
**d__Bacteria;p__Firmicutes;c__Negativicutes;o__Acidaminococcales;f__Acidaminococcaceae;g__Phascolarctobacterium_A**
**SHHC**
S4-wms-combined_118531
**Caudoviricetes**
Unclassified
**O**
O
S4-wms-combined_57963
**Caudoviricetes**
Unclassified
**Unclassified**
O
**d__Bacteria;p__Firmicutes;c__Negativicutes;o__Selenomonadales;f__Selenomonadaceae;g__Megamonas**
**SHHC**
S2-wms-combined_197577
**Caudoviricetes**
Unclassified
**Unclassified**
Unclassified
**d__Bacteria;p__Firmicutes;c__Negativicutes;o__Veillonellales;f__Dialisteraceae;g__Dialister**
**Shared**

S17-wms-combined_95360
**Caudoviricetes**
Unclassified
**O**
O
S2-wms-combined_135014
**Caudoviricetes**
Unclassified
**Unclassified**
O
S2-wms-combined_135154
**n.a.**
n.a.
**n.a.**
n.a.
S2-wms-combined_185494
**Caudoviricetes**
Autographiviridae
**O**
O
S7-wms-combined_74829
**Caudoviricetes**
Unclassified
**O**
O
S9-wms-combined_53991
**Caudoviricetes**
Unclassified
**O**
O
**SHHC**
S6-wms-combined_65225
**n.a.**
n.a.
**n.a.**
n.a.
**d__Bacteria;p__Firmicutes;c__Negativicutes;o__Veillonellales;f__Veillonellaceae;g__Veillonella**
**ME**
S9-wms_114971
**Caudoviricetes**
Unclassified
**O**
O
**d__Bacteria;p__Patescibacteria;c__Paceibacteria;o__Paceibacterales;f__UBA5633;g__MWCK01**
**SHHC**
S12-wms_401372
**Caudoviricetes**
Unclassified
**Unclassified**
Unclassified
**d__Bacteria;p__Proteobacteria;c__Alphaproteobacteria;o__RF32;f__CAG-239;g__51-20**

**ME**
S1-wms-combined_92523
**n.a.**
Unclassified
**O**
O
**d__Bacteria;p__Proteobacteria;c__Alphaproteobacteria;o__RF32;f__CAG-239;g__CAG-267**
**ME**
S11-wms-combined_204522
**Caudoviricetes**
Unclassified
**O**
O
**Shared**
S11-wms-combined_193148
**n.a.**
n.a.
**n.a.**
n.a.
**SHHC**
S6-wms-combined_745
**n.a.**
n.a.
**n.a.**
n.a.
**d__Bacteria;p__Proteobacteria;c__Alphaproteobacteria;o__RF32;f__CAG-239;g__CAG-495**
**Shared**
S11-wms-combined_101337
**Caudoviricetes**
Unclassified
**O**
O
**SHHC**
S12-wms-combined_313953
**Caudoviricetes**
Unclassified
**O**
O
**d__Bacteria;p__Proteobacteria;c__Gammaproteobacteria;o__Burkholderiales;f__Burkholderiaceae;g__Oxalobacter**
**ME**
S5-wms-combined_3201
**Caudoviricetes**
Unclassified
**O**
O
**d__Bacteria;p__Proteobacteria;c__Gammaproteobacteria;o__Burkholderiales;f__Burkholderiaceae;g__Sutterella**
**Shared**
S2-wms-combined_84608
**Caudoviricetes**
Unclassified
**O**

O

**d__Bacteria;p__Proteobacteria;c__Gammaproteobacteria;o__Enterobacterales;f__Enterobacteriaceae;g__Escherichia**

**ME**

S5-wms-combined_18356

**Caudoviricetes**

Peduoviridae

**Unclassified**

Evevirus

S9-wms-combined_64277

**Caudoviricetes**

Unclassified

**Unclassified**

Bcepmuvirus

**SHHC**

S2-wms_128831

**Caudoviricetes**

Unclassified

**Unclassified**

Unclassified

**d__Bacteria;p__Proteobacteria;c__Gammaproteobacteria;o__Enterobacterales;f__Enterobacteriaceae;g__Klebsiella**

**ME**

S9-wms-combined_93232

**Caudoviricetes**

Unclassified

**O**

O

**d__Bacteria;p__Proteobacteria;c__Gammaproteobacteria;o__Enterobacterales;f__Enterobacteriaceae;g__Salmonella**

**ME**

S11-wms-combined_154845

**Caudoviricetes**

Peduoviridae

**Unclassified**

Quadragintavirus

**d__Bacteria;p__Verrucomicrobiota;c__Verrucomicrobiae;o__Verrucomicrobiales;f__Akkermansiaceae;g__Akkermansia**

**ME**

S7-wms-combined_217226

**Caudoviricetes**

Unclassified

**O**

O

**Shared**

S12-wms-combined_168259

**Caudoviricetes**

Unclassified

**O**

O

S12-wms-combined_282594

**Caudoviricetes**

Unclassified

**O**

O

S1-wms-combined_308848
**n.a.**
n.a.
**n.a.**
n.a.
S2-wms-combined_135139
**n.a.**
n.a.
**n.a.**
n.a.
S2-wms-combined_201205
**Caudoviricetes**
Unclassified
**Unclassified**
O
S2-wms-combined_7756
**Caudoviricetes**
Unclassified
**O**
O
S5-wms-combined_52741
**Caudoviricetes**
Unclassified
**O**
O
S8-wms-combined_246185
**Caudoviricetes**
Unclassified
**O**
O

| Shared_hosts_seen_in_both_datasets | Unique_hosts_in_VLP | Unique_hosts_in_WMS |
| --- | --- | --- |
| d__Bacteria;p__Actinobacteriota;c__Actinomycetia;o__Actinomycetales;f__Bifidobacteriaceae;g__Bifidobacterium | d__Bacteria;p__Actinobacteriota;c__Actinomycetia;o__Actinomycetales;f__Actinomycetaceae;g__Actinomyces | d__Bacteria;p__Actinobacteriota;c__Coriobacteriia;o__Coriobacteriales;f__Eggerthellaceae;g__CAG-1427 |
| d__Bacteria;p__Actinobacteriota;c__Coriobacteriia;o__Coriobacteriales;f__Coriobacteriaceae;g__Collinsella | d__Bacteria;p__Actinobacteriota;c__Actinomycetia;o__Actinomycetales;f__Actinomycetaceae;g__Pauljensenia | d__Bacteria;p__Bacteroidota;c__Bacteroidia;o__Bacteroidales;f__Bacteroidaceae;g__Bacteroides_G |
| d__Bacteria;p__Actinobacteriota;c__Coriobacteriia;o__Coriobacteriales;f__Eggerthellaceae;g__Adlercreutzia | d__Bacteria;p__Actinobacteriota;c__Actinomycetia;o__Actinomycetales;f__Micrococcaceae;g__Rothia | d__Bacteria;p__Bacteroidota;c__Bacteroidia;o__Bacteroidales;f__Rikenellaceae;g__Tidjanibacter |
| d__Bacteria;p__Actinobacteriota;c__Coriobacteriia;o__Coriobacteriales;f__Eggerthellaceae;g__Eggerthella | d__Bacteria;p__Actinobacteriota;c__Actinomycetia;o__Mycobacteriales;f__Mycobacteriaceae;g__Mycobacterium | d__Bacteria;p__Campylobacterota;c__Campylobacteria;o__Campylobacterales;f__Campylobacteraceae;g__Campylobacter_D |
| d__Bacteria;p__Bacteroidota;c__Bacteroidia;o__Bacteroidales;f__Bacteroidaceae;g__Bacteroides | d__Bacteria;p__Actinobacteriota;c__Coriobacteriia;o__Coriobacteriales;f__Eggerthellaceae;g__Gordonibacter | d__Bacteria;p__Desulfobacterota;c__Desulfovibrionia;o__Desulfovibrionales;f__Desulfovibrionaceae;g__Desulfovibrio |
| d__Bacteria;p__Bacteroidota;c__Bacteroidia;o__Bacteroidales;f__Bacteroidaceae;g__Prevotella | d__Bacteria;p__Bacteroidota;c__Bacteroidia;o__Bacteroidales;f__Bacteroidaceae;g__Phocaeicola | d__Bacteria;p__Firmicutes;c__Clostridia;o__Christensenellales;f__CAG-74;g__SFFH01 |
| d__Bacteria;p__Bacteroidota;c__Bacteroidia;o__Bacteroidales;f__Bacteroidaceae;g__Prevotellamassilia | d__Bacteria;p__Bacteroidota;c__Bacteroidia;o__Bacteroidales;f__Coprobacteraceae;g__Coprobacter | d__Bacteria;p__Firmicutes;c__Clostridia;o__Christensenellales;f__CAG-917;g__ |
| d__Bacteria;p__Bacteroidota;c__Bacteroidia;o__Bacteroidales;f__Barnesiellaceae;g__Barnesiella | d__Bacteria;p__Bacteroidota;c__Bacteroidia;o__Bacteroidales;f__Marinifilaceae;g__Butyricimonas | d__Bacteria;p__Firmicutes;c__Clostridia;o__Lachnospirales;f__Lachnospiraceae;g__Acetivibrio_A |
| d__Bacteria;p__Bacteroidota;c__Bacteroidia;o__Bacteroidales;f__Muribaculaceae;g__UBA7173 | d__Bacteria;p__Bacteroidota;c__Bacteroidia;o__Bacteroidales;f__Marinifilaceae;g__Odoribacter | d__Bacteria;p__Firmicutes;c__Clostridia;o__Lachnospirales;f__Lachnospiraceae;g__Anaerosacchariphilus |
| d__Bacteria;p__Bacteroidota;c__Bacteroidia;o__Bacteroidales;f__Rikenellaceae;g__Alistipes | d__Bacteria;p__Bacteroidota;c__Bacteroidia;o__Bacteroidales;f__P3;g__Phil12 | d__Bacteria;p__Firmicutes;c__Clostridia;o__Lachnospirales;f__Lachnospiraceae;g__CAG-81 |
| d__Bacteria;p__Bacteroidota;c__Bacteroidia;o__Bacteroidales;f__Rikenellaceae;g__Alistipes_A | d__Bacteria;p__Cyanobacteria;c__Vampirovibrionia;o__Gastranaerophilales;f__Gastranaerophilaceae;g__CAG-306 | d__Bacteria;p__Firmicutes;c__Clostridia;o__Lachnospirales;f__Lachnospiraceae;g__COE1 |
| d__Bacteria;p__Bacteroidota;c__Bacteroidia;o__Bacteroidales;f__Tannerellaceae;g__Marseille-P3160 | d__Bacteria;p__Desulfobacterota;c__Desulfovibrionia;o__Desulfovibrionales;f__Desulfovibrionaceae;g__Mailhella | d__Bacteria;p__Firmicutes;c__Clostridia;o__Lachnospirales;f__Lachnospiraceae;g__Fusicatenibacter |
| d__Bacteria;p__Bacteroidota;c__Bacteroidia;o__Bacteroidales;f__Tannerellaceae;g__Parabacteroides | d__Bacteria;p__Firmicutes;c__Bacilli;o__Erysipelotrichales;f__Erysipelatoclostridiaceae;g__Catenibacterium | d__Bacteria;p__Firmicutes;c__Clostridia;o__Lachnospirales;f__Lachnospiraceae;g__Lachnoclostridium_A |
| d__Bacteria;p__Bacteroidota;c__Bacteroidia;o__Bacteroidales;f__UBA11471;g__UBA11471 | d__Bacteria;p__Firmicutes;c__Bacilli;o__Erysipelotrichales;f__Erysipelatoclostridiaceae;g__Faecalibacillus | d__Bacteria;p__Firmicutes;c__Clostridia;o__Oscillospirales;f__Oscillospiraceae;g__F23-B02 |
| d__Bacteria;p__Cyanobacteria;c__Vampirovibrionia;o__Gastranaerophilales;f__Gastranaerophilaceae;g__QAMI01 | d__Bacteria;p__Firmicutes;c__Bacilli;o__Erysipelotrichales;f__Erysipelatoclostridiaceae;g__Massiliomicrobiota | d__Bacteria;p__Firmicutes;c__Clostridia;o__Oscillospirales;f__Oscillospiraceae;g__Faecousia |
| d__Bacteria;p__Desulfobacterota;c__Desulfovibrionia;o__Desulfovibrionales;f__Desulfovibrionaceae;g__Bilophila | d__Bacteria;p__Firmicutes;c__Bacilli;o__Erysipelotrichales;f__Erysipelotrichaceae;g__Holdemanella | d__Bacteria;p__Firmicutes;c__Clostridia;o__Oscillospirales;f__Oscillospiraceae;g__HGM12998 |
| d__Bacteria;p__Firmicutes;c__Bacilli;o__Erysipelotrichales;f__Erysipelatoclostridiaceae;g__Erysipelatoclostridium | d__Bacteria;p__Firmicutes;c__Bacilli;o__Erysipelotrichales;f__Erysipelotrichaceae;g__Holdemania | d__Bacteria;p__Firmicutes;c__Clostridia;o__Oscillospirales;f__Oscillospiraceae;g__Marseille-P3106 |
| d__Bacteria;p__Firmicutes;c__Bacilli;o__Erysipelotrichales;f__Erysipelotrichaceae;g__Clostridium_AQ | d__Bacteria;p__Firmicutes;c__Bacilli;o__Haloplasmatales_A;f__Turicibacteraceae;g__Turicibacter | d__Bacteria;p__Firmicutes;c__Clostridia;o__Oscillospirales;f__Oscillospiraceae;g__Pseudoflavonifractor |
| d__Bacteria;p__Firmicutes;c__Bacilli;o__Lactobacillales;f__Lactobacillaceae;g__Lacticaseibacillus | d__Bacteria;p__Firmicutes;c__Bacilli;o__Lactobacillales;f__Lactobacillaceae;g__Lactobacillus | d__Bacteria;p__Firmicutes;c__Clostridia;o__Oscillospirales;f__Oscillospiraceae;g__UBA738 |
| d__Bacteria;p__Firmicutes;c__Bacilli;o__Lactobacillales;f__Streptococcaceae;g__Streptococcus | d__Bacteria;p__Firmicutes;c__Bacilli;o__Lactobacillales;f__Lactobacillaceae;g__Leuconostoc | d__Bacteria;p__Firmicutes;c__Clostridia;o__Oscillospirales;f__Ruminococcaceae;g__UMGS363 |
| d__Bacteria;p__Firmicutes;c__Clostridia;o__Christensenellales;f__CAG-138;g__SFEL01 | d__Bacteria;p__Firmicutes;c__Bacilli;o__Lactobacillales;f__Lactobacillaceae;g__Limosilactobacillus | d__Bacteria;p__Firmicutes;c__Negativicutes;o__Acidaminococcales;f__Acidaminococcaceae;g__Phascolarctobacterium_A |
| d__Bacteria;p__Firmicutes;c__Clostridia;o__Christensenellales;f__CAG-138;g__UBA1685 | d__Bacteria;p__Firmicutes;c__Bacilli;o__Lactobacillales;f__Streptococcaceae;g__Lactococcus | d__Bacteria;p__Firmicutes;c__Negativicutes;o__Selenomonadales;f__Selenomonadaceae;g__Megamonas |
| d__Bacteria;p__Firmicutes;c__Clostridia;o__Christensenellales;f__CAG-74;g__Firm-11 | d__Bacteria;p__Firmicutes;c__Bacilli;o__ML615J-28;f__CAG-313;g__CAG-313 | d__Bacteria;p__Patescibacteria;c__Paceibacteria;o__Paceibacterales;f__UBA5633;g__MWCK01 |
| d__Bacteria;p__Firmicutes;c__Clostridia;o__Christensenellales;f__CAG-74;g__Limiplasma | d__Bacteria;p__Firmicutes;c__Bacilli;o__RF39;f__UBA660;g__CAG-302 | d__Bacteria;p__Proteobacteria;c__Gammaproteobacteria;o__Burkholderiales;f__Burkholderiaceae;g__Oxalobacter |
| d__Bacteria;p__Firmicutes;c__Clostridia;o__Christensenellales;f__CAG-74;g__UBA11524 | d__Bacteria;p__Firmicutes;c__Bacilli;o__RF39;f__UBA660;g__CAG-460 | d__Bacteria;p__Proteobacteria;c__Gammaproteobacteria;o__Enterobacterales;f__Enterobacteriaceae;g__Klebsiella |
| d__Bacteria;p__Firmicutes;c__Clostridia;o__Christensenellales;f__CAG-74;g__Ventricola | d__Bacteria;p__Firmicutes;c__Bacilli;o__RF39;f__UBA660;g__CAG-877 | |
| d__Bacteria;p__Firmicutes;c__Clostridia;o__Christensenellales;f__CAG-917;g__CAG-475 | d__Bacteria;p__Firmicutes;c__Clostridia;o__Acetivibrionales;f__Acetivibrionaceae;g__Ruminiclostridium | **Stats:** |
| d__Bacteria;p__Firmicutes;c__Clostridia;o__Christensenellales;f__Christensenellaceae;g__Christensenella | d__Bacteria;p__Firmicutes;c__Clostridia;o__Christensenellales;f__Borkfalkiaceae;g__QALS01 | Hosts interact with ME-vOTUs uniquely: 6/25 (24%) |
| d__Bacteria;p__Firmicutes;c__Clostridia;o__Clostridiales;f__Clostridiaceae;g__Clostridium | d__Bacteria;p__Firmicutes;c__Clostridia;o__Christensenellales;f__CAG-138;g__UBA5394 | Hosts interact with SHHC-vOTUs uniquely: 10/25 (40%) |
| d__Bacteria;p__Firmicutes;c__Clostridia;o__Eubacteriales;f__Eubacteriaceae;g__Eubacterium | d__Bacteria;p__Firmicutes;c__Clostridia;o__Christensenellales;f__CAG-74;g__Alectryocaccomicrobium | Hosts interact with common viruses (vOTUs) shared in both ME and SHHC: 9/25 (36%) |
| d__Bacteria;p__Firmicutes;c__Clostridia;o__Lachnospirales;f__Anaerotignaceae;g__Anaerotignum | d__Bacteria;p__Firmicutes;c__Clostridia;o__Christensenellales;f__CAG-74;g__SFTH01 | |
| d__Bacteria;p__Firmicutes;c__Clostridia;o__Lachnospirales;f__Lachnospiraceae;g__Acetatifactor | d__Bacteria;p__Firmicutes;c__Clostridia;o__Christensenellales;f__NSJ-53;g__NSJ-53 | |
| d__Bacteria;p__Firmicutes;c__Clostridia;o__Lachnospirales;f__Lachnospiraceae;g__Agathobacter | d__Bacteria;p__Firmicutes;c__Clostridia;o__Christensenellales;f__QAND01;g__UMGS1975 | |
| d__Bacteria;p__Firmicutes;c__Clostridia;o__Lachnospirales;f__Lachnospiraceae;g__Anaerobutyricum | d__Bacteria;p__Firmicutes;c__Clostridia;o__Clostridiales;f__Clostridiaceae;g__Clostridium_J | |
| d__Bacteria;p__Firmicutes;c__Clostridia;o__Lachnospirales;f__Lachnospiraceae;g__Anaerostipes | d__Bacteria;p__Firmicutes;c__Clostridia;o__Lachnospirales;f__Anaerotignaceae;g__UMGS1670 | |
| d__Bacteria;p__Firmicutes;c__Clostridia;o__Lachnospirales;f__Lachnospiraceae;g__Bariatricus | d__Bacteria;p__Firmicutes;c__Clostridia;o__Lachnospirales;f__CAG-274;g__CAG-274 | |
| d__Bacteria;p__Firmicutes;c__Clostridia;o__Lachnospirales;f__Lachnospiraceae;g__Blautia_A | d__Bacteria;p__Firmicutes;c__Clostridia;o__Lachnospirales;f__Lachnospiraceae;g__ | |
| d__Bacteria;p__Firmicutes;c__Clostridia;o__Lachnospirales;f__Lachnospiraceae;g__Dorea_A | d__Bacteria;p__Firmicutes;c__Clostridia;o__Lachnospirales;f__Lachnospiraceae;g__Blautia | |
| d__Bacteria;p__Firmicutes;c__Clostridia;o__Lachnospirales;f__Lachnospiraceae;g__Eisenbergiella | d__Bacteria;p__Firmicutes;c__Clostridia;o__Lachnospirales;f__Lachnospiraceae;g__CAG-127 | |
| d__Bacteria;p__Firmicutes;c__Clostridia;o__Lachnospirales;f__Lachnospiraceae;g__Enterocloster | d__Bacteria;p__Firmicutes;c__Clostridia;o__Lachnospirales;f__Lachnospiraceae;g__CAG-317 | |
| d__Bacteria;p__Firmicutes;c__Clostridia;o__Lachnospirales;f__Lachnospiraceae;g__Eubacterium_I | d__Bacteria;p__Firmicutes;c__Clostridia;o__Lachnospirales;f__Lachnospiraceae;g__Clostridium_AP | |
| d__Bacteria;p__Firmicutes;c__Clostridia;o__Lachnospirales;f__Lachnospiraceae;g__Faecalimonas | d__Bacteria;p__Firmicutes;c__Clostridia;o__Lachnospirales;f__Lachnospiraceae;g__Coprococcus | |
| d__Bacteria;p__Firmicutes;c__Clostridia;o__Lachnospirales;f__Anaerotignaceae;g__Anaerotignum | d__Bacteria;p__Firmicutes;c__Clostridia;o__Lachnospirales;f__Lachnospiraceae;g__Dorea | |
| d__Bacteria;p__Firmicutes;c__Clostridia;o__Lachnospirales;f__Lachnospiraceae;g__Mediterraneibacter | d__Bacteria;p__Firmicutes;c__Clostridia;o__Lachnospirales;f__Lachnospiraceae;g__Eubacterium_F | |
| d__Bacteria;p__Firmicutes;c__Clostridia;o__Lachnospirales;f__Lachnospiraceae;g__Roseburia | d__Bacteria;p__Firmicutes;c__Clostridia;o__Lachnospirales;f__Lachnospiraceae;g__Eubacterium_G | |
| d__Bacteria;p__Firmicutes;c__Clostridia;o__Lachnospirales;f__Lachnospiraceae;g__UBA9502 | d__Bacteria;p__Firmicutes;c__Clostridia;o__Lachnospirales;f__Lachnospiraceae;g__Frisingicoccus | |
| d__Bacteria;p__Firmicutes;c__Clostridia;o__Oscillospirales;f__Acutalibacteraceae;g__Acutalibacter | d__Bacteria;p__Firmicutes;c__Clostridia;o__Lachnospirales;f__Lachnospiraceae;g__Hungatella | |
| d__Bacteria;p__Firmicutes;c__Clostridia;o__Oscillospirales;f__Acutalibacteraceae;g__CAG-180 | d__Bacteria;p__Firmicutes;c__Clostridia;o__Lachnospirales;f__Lachnospiraceae;g__Marvinbryantia | |
| d__Bacteria;p__Firmicutes;c__Clostridia;o__Oscillospirales;f__Acutalibacteraceae;g__CAG-217 | d__Bacteria;p__Firmicutes;c__Clostridia;o__Lachnospirales;f__Lachnospiraceae;g__Massilistercora | |
| d__Bacteria;p__Firmicutes;c__Clostridia;o__Oscillospirales;f__Acutalibacteraceae;g__Ruminococcus_E | d__Bacteria;p__Firmicutes;c__Clostridia;o__Lachnospirales;f__Lachnospiraceae;g__Muricomes | |
| d__Bacteria;p__Firmicutes;c__Clostridia;o__Oscillospirales;f__Acutalibacteraceae;g__UBA1417 | d__Bacteria;p__Firmicutes;c__Clostridia;o__Lachnospirales;f__Lachnospiraceae;g__OF09-33XD | |
| d__Bacteria;p__Firmicutes;c__Clostridia;o__Oscillospirales;f__Butyricicoccaceae;g__Agathobaculum | d__Bacteria;p__Firmicutes;c__Clostridia;o__Lachnospirales;f__Lachnospiraceae;g__RUG115 | |
| d__Bacteria;p__Firmicutes;c__Clostridia;o__Oscillospirales;f__Oscillospiraceae;g__ | d__Bacteria;p__Firmicutes;c__Clostridia;o__Lachnospirales;f__Lachnospiraceae;g__Ruminococcus_B | |
| d__Bacteria;p__Firmicutes;c__Clostridia;o__Oscillospirales;f__Oscillospiraceae;g__An92 | d__Bacteria;p__Firmicutes;c__Clostridia;o__Lachnospirales;f__Lachnospiraceae;g__Sellimonas | |
| d__Bacteria;p__Firmicutes;c__Clostridia;o__Oscillospirales;f__Oscillospiraceae;g__CAG-103 | d__Bacteria;p__Firmicutes;c__Clostridia;o__Lachnospirales;f__Lachnospiraceae;g__Ventrimonas | |
| d__Bacteria;p__Firmicutes;c__Clostridia;o__Oscillospirales;f__Oscillospiraceae;g__CAG-110 | d__Bacteria;p__Firmicutes;c__Clostridia;o__Monoglobales;f__Monoglobaceae;g__Monoglobus | |
| d__Bacteria;p__Firmicutes;c__Clostridia;o__Oscillospirales;f__Oscillospiraceae;g__CAG-170 | d__Bacteria;p__Firmicutes;c__Clostridia;o__Oscillospirales;f__Acutalibacteraceae;g__An172 | |
| d__Bacteria;p__Firmicutes;c__Clostridia;o__Oscillospirales;f__Oscillospiraceae;g__CAG-83 | d__Bacteria;p__Firmicutes;c__Clostridia;o__Oscillospirales;f__Acutalibacteraceae;g__Eubacterium_R | |
| d__Bacteria;p__Firmicutes;c__Clostridia;o__Oscillospirales;f__Oscillospiraceae;g__Dysosmobacter | d__Bacteria;p__Firmicutes;c__Clostridia;o__Oscillospirales;f__Acutalibacteraceae;g__Fimivicinus | |
| d__Bacteria;p__Firmicutes;c__Clostridia;o__Oscillospirales;f__Oscillospiraceae;g__ER4 | d__Bacteria;p__Firmicutes;c__Clostridia;o__Oscillospirales;f__Acutalibacteraceae;g__Hydrogeniiclostridium | |
| d__Bacteria;p__Firmicutes;c__Clostridia;o__Oscillospirales;f__Oscillospiraceae;g__Evtepia | d__Bacteria;p__Firmicutes;c__Clostridia;o__Oscillospirales;f__Acutalibacteraceae;g__Pseudoruminococcus | |
| d__Bacteria;p__Firmicutes;c__Clostridia;o__Oscillospirales;f__Oscillospiraceae;g__Flavonifractor | d__Bacteria;p__Firmicutes;c__Clostridia;o__Oscillospirales;f__Acutalibacteraceae;g__UBA1691 | |
| d__Bacteria;p__Firmicutes;c__Clostridia;o__Oscillospirales;f__Oscillospiraceae;g__Intestinimonas | d__Bacteria;p__Firmicutes;c__Clostridia;o__Oscillospirales;f__Acutalibacteraceae;g__UBA737 | |
| d__Bacteria;p__Firmicutes;c__Clostridia;o__Oscillospirales;f__Oscillospiraceae;g__Lawsonibacter | d__Bacteria;p__Firmicutes;c__Clostridia;o__Oscillospirales;f__Acutalibacteraceae;g__UMGS1071 | |
| d__Bacteria;p__Firmicutes;c__Clostridia;o__Oscillospirales;f__Oscillospiraceae;g__UBA9475 | d__Bacteria;p__Firmicutes;c__Clostridia;o__Oscillospirales;f__Acutalibacteraceae;g__UMGS1591 | |
| d__Bacteria;p__Firmicutes;c__Clostridia;o__Oscillospirales;f__Ruminococcaceae;g__Faecalibacterium | d__Bacteria;p__Firmicutes;c__Clostridia;o__Oscillospirales;f__Acutalibacteraceae;g__UMGS263 | |
| d__Bacteria;p__Firmicutes;c__Clostridia;o__Oscillospirales;f__Ruminococcaceae;g__Gemmiger | d__Bacteria;p__Firmicutes;c__Clostridia;o__Oscillospirales;f__Acutalibacteraceae;g__UMGS856 | |
| d__Bacteria;p__Firmicutes;c__Clostridia;o__Oscillospirales;f__Ruminococcaceae;g__Negativibacillus | d__Bacteria;p__Firmicutes;c__Clostridia;o__Oscillospirales;f__CAG-272;g__CAG-272 | |
| d__Bacteria;p__Firmicutes;c__Clostridia;o__Oscillospirales;f__Ruminococcaceae;g__Ruminiclostridium_E | d__Bacteria;p__Firmicutes;c__Clostridia;o__Oscillospirales;f__CAG-272;g__QALR01 | |
| d__Bacteria;p__Firmicutes;c__Clostridia;o__Oscillospirales;f__Ruminococcaceae;g__Ruminococcus_D | d__Bacteria;p__Firmicutes;c__Clostridia;o__Oscillospirales;f__Oscillospiraceae;g__Enterenecus | |
| d__Bacteria;p__Firmicutes;c__Clostridia;o__Oscillospirales;f__Ruminococcaceae;g__Ruthenibacterium | d__Bacteria;p__Firmicutes;c__Clostridia;o__Oscillospirales;f__Oscillospiraceae;g__UBA5446 | |
| d__Bacteria;p__Firmicutes;c__Clostridia;o__Oscillospirales;f__Ruminococcaceae;g__UBA3818 | d__Bacteria;p__Firmicutes;c__Clostridia;o__Oscillospirales;f__Oscillospiraceae;g__UMGS1766 | |

d__Bacteria;p__Firmicutes;c__Negativicutes;o__Veillonellales;f__Dialisteraceae;g__Dialister
d__Bacteria;p__Firmicutes;c__Negativicutes;o__Veillonellales;f__Veillonellaceae;g__Veillonella
d__Bacteria;p__Proteobacteria;c__Alphaproteobacteria;o__RF32;f__CAG-239;g__51-20
d__Bacteria;p__Proteobacteria;c__Alphaproteobacteria;o__RF32;f__CAG-239;g__CAG-267
d__Bacteria;p__Proteobacteria;c__Alphaproteobacteria;o__RF32;f__CAG-239;g__CAG-495
d__Bacteria;p__Proteobacteria;c__Gammaproteobacteria;o__Burkholderiales;f__Burkholderiaceae;g__Sutterella
d__Bacteria;p__Proteobacteria;c__Gammaproteobacteria;o__Enterobacterales;f__Enterobacteriaceae;g__Escherichia
d__Bacteria;p__Proteobacteria;c__Gammaproteobacteria;o__Enterobacterales;f__Enterobacteriaceae;g__Salmonella
d__Bacteria;p__Verrucomicrobiota;c__Verrucomicrobiae;o__Verrucomicrobiales;f__Akkermansiaceae;g__Akkermansia

**Stats:**
Hosts interact with ME-vOTUs uniquely: 11/81 (13.6%)
Hosts interact with SHHC-vOTUs uniquely: 6/81 (7.4%)
Hosts interact with communal viruses shared in both ME and SHHC: 64/81 (79%)

d__Bacteria;p__Firmicutes;c__Clostridia;o__Oscillospirales;f__Ruminococcaceae;g__Anaerotruncus
d__Bacteria;p__Firmicutes;c__Clostridia;o__Oscillospirales;f__Ruminococcaceae;g__CAG-353
d__Bacteria;p__Firmicutes;c__Clostridia;o__Oscillospirales;f__Ruminococcaceae;g__D5
d__Bacteria;p__Firmicutes;c__Clostridia;o__Oscillospirales;f__Ruminococcaceae;g__Massilimaliae
d__Bacteria;p__Firmicutes;c__Clostridia;o__Oscillospirales;f__Ruminococcaceae;g__Massilioclostridium
d__Bacteria;p__Firmicutes;c__Clostridia;o__Oscillospirales;f__Ruminococcaceae;g__Phocea
d__Bacteria;p__Firmicutes;c__Clostridia;o__Oscillospirales;f__Ruminococcaceae;g__QAMM01
d__Bacteria;p__Firmicutes;c__Clostridia;o__Oscillospirales;f__Ruminococcaceae;g__Ruminococcus_C
d__Bacteria;p__Firmicutes;c__Clostridia;o__Oscillospirales;f__UMGS1783;g__UMGS1783
d__Bacteria;p__Firmicutes;c__Clostridia;o__Peptostreptococcales;f__Anaerovoracaceae;g__CAG-145
d__Bacteria;p__Firmicutes;c__Clostridia;o__Peptostreptococcales;f__Peptostreptococcaceae;g__GCA-900066495
d__Bacteria;p__Firmicutes;c__Clostridia;o__TANB77;f__CAG-508;g__CAG-269
d__Bacteria;p__Firmicutes;c__Clostridia;o__TANB77;f__CAG-508;g__CAG-354
d__Bacteria;p__Firmicutes;c__Clostridia;o__UBA1212;f__UBA1255;g__Egerieisoma
d__Bacteria;p__Firmicutes;c__Negativicutes;o__Acidaminococcales;f__Acidaminococcaceae;g__Phascolarctobacterium
d__Bacteria;p__Firmicutes;c__Negativicutes;o__Veillonellales;f__Megasphaeraceae;g__Caecibacter
d__Bacteria;p__Patescibacteria;c__Paceibacteria;o__UBA9983_A;f__UBA1006;g__01-FULL-36-10b
d__Bacteria;p__Proteobacteria;c__Gammaproteobacteria;o__Burkholderiales;f__Burkholderiaceae;g__Duodenibacillus
d__Bacteria;p__Proteobacteria;c__Gammaproteobacteria;o__Burkholderiales;f__Burkholderiaceae;g__Parasutterella
d__Bacteria;p__Proteobacteria;c__Gammaproteobacteria;o__Enterobacterales;f__Pasteurellaceae;g__Glaesserella
d__Bacteria;p__Spirochaetota;c__Spirochaetia;o__Sphaerochaetales;f__Sphaerochaetaceae;g__UBA9732
d__Bacteria;p__Verrucomicrobiota;c__Lentisphaeria;o__Victivallales;f__Victivallaceae;g__Victivallis
