## Supplementary material for "Comprehensive profiling of the human intestinal DNA virome and prediction of disease-associated bacterial hosts in severe Myalgic Encephalomyelitis/Chronic Fatigue Syndrome (ME/CFS)": S Table Cohort and sequence stats

| Sample IDs | Health status | Gender | Age range at time of collection | # Household-matched pair | Collection of clinical metadata | Sample type | Sample source | Bristol stool scale | Sample dry weight (g) | VLP/mL | VLP/g feces | Total DNA (ng) | DNA yield (ng/g feces) | VLP-enriched DNA virome | Bulk whole metagenome (WMS) |
|---|---|---|---|---|---|---|---|---|---|---|---|---|---|---|---|
| S1 | Severe ME/CFS | F | 56-60 | 1M | Y | Frozen faeces | Homo sapiens | 2 | 22.25 | $1.1 \times 10^{8}$ | $7.1 \times 10^{8}$ | 2000.0 | 89.9 | Y | Y |
| S2 | SHHC | M | 61-65 | 1C | NA | Frozen faeces | Homo sapiens | 7 | 21.92 | $1.4 \times 10^{8}$ | $8.8 \times 10^{8}$ | 6120.0 | 279.2 | Y | Y |
| S3 | Severe ME/CFS | F | 36-40 | 2M | Y | Frozen faeces | Homo sapiens | 6 | 44.83 | $2.7 \times 10^{8}$ | $1.7 \times 10^{9}$ | 2745.0 | 61.2 | Y | Y |
| S4 | SHHC | M | 36-40 | 2C | NA | Frozen faeces | Homo sapiens | 5 | 37.29 | $2.1 \times 10^{8}$ | $1.3 \times 10^{9}$ | 3852.0 | 103.3 | Y | Y |
| S5 | Severe ME/CFS | F | 21-25 | 3M | Y | Frozen faeces | Homo sapiens | 1 | 20.77 | $1.2 \times 10^{8}$ | $7.7 \times 10^{8}$ | 383.5 | 18.5 | Y | Y |
| S6 | SHHC | F | 56-60 | 3C | NA | Frozen faeces | Homo sapiens | 6 | 19.42 | $6.2 \times 10^{7}$ | $3.9 \times 10^{8}$ | 3657.0 | 188.3 | Y | Y |
| S7 | Severe ME/CFS | F | 21-25 | 4M | Y | Frozen faeces | Homo sapiens | 5 | 20.00 | $4.0 \times 10^{8}$ | $2.5 \times 10^{9}$ | 615.0 | 30.8 | Y | Y |
| S8 | SHHC | M | 21-25 | 4C | NA | Frozen faeces | Homo sapiens | 2 | 20.29 | $3.2 \times 10^{8}$ | $2.0 \times 10^{9}$ | 3733.2 | 184.0 | Y | Y |
| S9 | Severe ME/CFS | F | 36-40 | 5M | | Frozen faeces | Homo sapiens | | 50.00 | $2.1 \times 10^{8}$ | $1.3 \times 10^{9}$ | 600.0 | 12.0 | Y | Y |
| S10 | SHHC | F | 61-65 | 5C | | Frozen faeces | Homo sapiens | | 50.00 | $2.2 \times 10^{8}$ | $1.4 \times 10^{9}$ | 1842.0 | 36.8 | Y | N |
| S11 | Severe ME/CFS | F | 21-25 | 6M | | Frozen faeces | Homo sapiens | | 41.21 | $4.6 \times 10^{8}$ | $2.9 \times 10^{9}$ | 1027.0 | 24.9 | Y | Y |
| S12 | SHHC | F | 56-60 | 6C | | Frozen faeces | Homo sapiens | | 38.17 | $3.3 \times 10^{8}$ | $2.1 \times 10^{9}$ | 945.0 | 24.8 | Y | Y |
| S13 | Severe ME/CFS | F | 36-40 | 9M (Unmatched) | Unrecorded | Frozen faeces | Homo sapiens | Unrecorded | 33.62 | $7.2 \times 10^{7}$ | $4.5 \times 10^{8}$ | 4670.0 | 138.9 | Y | N |
| S14 | Severe ME/CFS | F | 16-20 | 7M | | Frozen faeces | Homo sapiens | | 16.37 | $2.1 \times 10^{8}$ | $1.3 \times 10^{9}$ | 488.0 | 29.8 | Y | N |
| S15 | SHHC | F | 51-55 | 7C | | Frozen faeces | Homo sapiens | | 17.42 | $4.1 \times 10^{8}$ | $2.6 \times 10^{9}$ | 5592.0 | 321.0 | Y | N |
| S16 | SHHC | F | 66-70 | 8C | | Frozen faeces | Homo sapiens | | 31.65 | $1.9 \times 10^{8}$ | $1.2 \times 10^{9}$ | 2806.0 | 88.7 | Y | N |
| S17 | Severe ME/CFS | F | 41-45 | 8M | | Frozen faeces | Homo sapiens | | 40.16 | $4.6 \times 10^{8}$ | $2.8 \times 10^{9}$ | 1738.0 | 43.3 | Y | Y |
| NC1 | Blank | NA | NA | NA | NA | Buffer only | NA | NA | NA | 0 | 0 | ND | ND | Y | NA |
| NC2 | Blank | NA | NA | NA | NA | Buffer only | NA | NA | NA | 0 | 0 | ND | ND | Y | NA |
| NC3 | Blank | NA | NA | NA | NA | Buffer only | NA | NA | NA | 0 | 0 | ND | ND | Y | NA |

ME/CFS: Myalgic Encephalomyelitis and chronic fatigue syndrome
SHHC: same household healthy control
NC: negative/processing control sample (buffer only)
M: ME/CFS sample
C: SHHC sample
NA: not applicable
ND: not detectable

**enriched-vlp-reads and contigs**

| Sample_IDs | # Raw reads (PE) | # Cleaned reads (PE) | # Total raw contigs | N50 (bp) | Longest contig length (bp) |
|---|---|---|---|---|---|
| S1 | 72345890 | 35113882 | 160117 | 1328 | 101988 |
| S2 | 66466528 | 56868494 | 21603 | 1903 | 269575 |
| S3 | 210025926 | 174177994 | 126065 | 7721 | 303660 |
| S4 | 49280544 | 47904786 | 32063 | 7049 | 118697 |
| S5 | 80290420 | 65733438 | 143928 | 10731 | 474479 |
| S6 | 81370828 | 68149392 | 56204 | 15510 | 501646 |
| S7 | 97834806 | 81075886 | 138310 | 3884 | 405769 |
| S8 | 223264658 | 183020194 | 205876 | 2915 | 522585 |
| S9 | 84139802 | 71023532 | 90503 | 2228 | 321418 |
| S10 | 87299754 | 73760336 | 121264 | 4484 | 699342 |
| S11 | 80553640 | 67000402 | 103564 | 3234 | 251440 |
| S12 | 100680974 | 85558566 | 187492 | 3525 | 621377 |
| S13 | 72438224 | 60096768 | 154404 | 1978 | 289301 |
| S14 | 75299764 | 61978780 | 112646 | 4931 | 361320 |
| S15 | 40202586 | 32235928 | 77515 | 4759 | 317833 |
| S16 | 89664968 | 74204032 | 173237 | 4480 | 530177 |
| S17 | 75436218 | 58989106 | 174502 | 3202 | 335129 |
| Sum | 1586595530 | 1296891516 | 2079293 | 83862 | 6425736 |
| Mean | 93329148.8 | 76287736.0 | 122311.4 | 4933.0 | 377984.0 |
| SD | 48857207.3 | 41056515.8 | 53541.4 | 3618.0 | 163066.2 |

**enriched-vlp-viral genomes**

| | Merged_UViGs | UViGs_50-100%_completeness_checkv | nr_vOTUs | vOTUs_size_filtration_1kb_cut_off | HQ-vOTUs_50-100%_completeness_checkv |
|---|---|---|---|---|---|
| # Viral genomes | 311208 | 2267 | 192146 | 54779 | 1231 |
| # Viral genomes ≥ 1 kb | 88434 | 2267 | 54779 | 54779 | 1231 |
| # Viral genomes ≥ 5 kb | 13160 | 2240 | 8038 | 8038 | 1210 |
| # Viral genomes ≥ 10 kb | 5686 | 2180 | 3278 | 2378 | 1172 |
| N50 (bp) | 4064 | NA | 3694 | 6580 | 49258 |
| Longest representative (bp) | 289184 | 289184 | 289184 | 289184 | 289184 |
| Shortest representative (bp) | 167 | 2070 | 180 | 1000 | 2070 |

nr: non-redundancy
UViG: uncultivated virus genome
vOTU: virus  operational taxonomic unit
HQ: high quality
#: quantity or number
NA: not applicable

**wms-reads and contigs**

| Sample_IDs | # Raw reads (PE) | # Cleaned reads (PE) | # Total raw contigs | N50 (bp) | Longest contig length (bp) |
|---|---|---|---|---|---|
| S1 | 144016818 | 144000416 | 368621 | 3209 | 593488 |
| S2 | 148221396 | 148218256 | 238143 | 7203 | 593489 |
| S3 | 12026774 | 12026424 | 135476 | 1492 | 99031 |
| S4 | 53446050 | 53444298 | 164089 | 2217 | 415342 |
| S5 | 186302984 | 186273684 | 210113 | 9964 | 587364 |
| S6 | 57739770 | 57739286 | 139652 | 3307 | 563736 |
| S7 | 137959804 | 137959384 | 240000 | 4991 | 606105 |
| S8 | 90881904 | 90880674 | 266924 | 2693 | 525861 |
| S9 | 87344710 | 87343694 | 126134 | 8430 | 680403 |
| S11 | 84507078 | 84502272 | 321191 | 2940 | 302351 |
| S12 | 140009544 | 140008922 | 386774 | 3041 | 431188 |
| S17 | 110272558 | 110271798 | 257744 | 3366 | 502130 |
| Sum | 1252729390 | 1252669108 | 2854861 | 52853 | 5900488 |
| Mean | 104394115.8 | 104389092.3 | 237905.1 | 4404.4 | 491707.3 |
| SD | 49361687.0 | 49356246.2 | 88566.7 | 2683.3 | 160841.5 |

**wms-viral genomes**

| | Merged_UViGs | UViGs_50-100%_completeness_checkv | nr_vOTUs | Size_filtration_1kb_cut_off | HQ-vOTUs_50-100%_completeness_checkv |
|---|---|---|---|---|---|
| # Viral genomes | 261744 | 961 | 184317 | 59753 | 688 |
| # Viral genomes ≥ 1 kb | 82029 | 961 | 59753 | 59753 | 688 |
| # Viral genomes ≥ 5 kb | 11871 | 923 | 8708 | 8708 | 659 |
| # Viral genomes ≥ 10 kb | 4417 | 891 | 3227 | 3227 | 636 |
| N50 (bp) | 3491 | NA | 3616 | 5509 | 62201 |
| Longest representative (bp) | 413955 | 413955 | 413955 | 413955 | 413955 |
| Shortest representative (bp) | 193 | 1992 | 196 | 1000 | 2384 |

nr: non-redundancy
UViG: uncultivated virus genome
vOTU: virus operational taxonomic unit
HQ: high quality
#: quantity or number
NA: not applicable

**WMS-coassembly for bacteriome analysis**

| | # Contigs | Total length (bp) | Mean of length (bp) | N50 (bp) | Longest length (bp) | Shortest length (bp) |
|---|---|---|---|---|---|---|
| Quantity | 774,887 | 591,013,764 | 762.71 | 890 | 135,997 | 212 |

BLAST_WMS_against_VLP

| qseqid | sseqid | pident | qcovs | length | mismatch | gapopen | qstart | send | sstart | send | evalue | bitscore | Sorting: |
|---|---|---|---|---|---|---|---|---|---|---|---|---|---|
| S2-wms-combined_95987 | S14_vs_9162 | 100 | 0 | 29 | 0 | 0 | 10311 | 10339 | 47371 | 47399 | 2.05E-05 | 54.7 | 1. evalue: < 1E-05 |
| S2-wms_248327 | S8_vs_50431 | 100 | 0 | 29 | 0 | 0 | 5176 | 5204 | 23296 | 23324 | 1.87E-05 | 54.7 | 2. bitscore: set score > 1000 |
| S2-wms-combined_185494 | S15_vf_33505 | 100 | 0 | 33 | 0 | 0 | 28928 | 28960 | 5866 | 5834 | 2.68E-06 | 62.1 | 3. pident: percentage of identical matches: > 95% (similarity) |
| S1-wms-combined_353757 | S14_vs_66139 | 97.297 | 0 | 37 | 1 | 0 | 18939 | 18975 | 156473 | 156437 | 1.14E-07 | 63.9 | 4. qcovs: query coverage per subject: > 85% (coverage) |
| S9-wms-combined_93232 | S5_vf_74661 | 90 | 0 | 50 | 5 | 0 | 13333 | 13382 | 20063 | 20014 | 4.47E-08 | 65.8 | 5. Result: 193/600 hits of WMS-HQ-vOTUs (688) are highly identical against 1231 VLP-HQ-vC |
| S2-wms-combined_190624 | S16_vs_17493 | 81.818 | 0 | 77 | 14 | 0 | 14724 | 14800 | 12955 | 12879 | 2.35E-08 | 65.8 | |
| S6-wms-combined_41246 | S5_vs_65490 | 81.176 | 0 | 85 | 14 | 2 | 34140 | 34223 | 27618 | 27701 | 9.95E-09 | 67.6 | |
| S2-wms-combined_135139 | S16_vs_173096 | 95.349 | 0 | 43 | 2 | 0 | 4942 | 4984 | 15558 | 15516 | 7.94E-09 | 69.4 | |
| S1-wms_244836 | S10_vs_36598 | 74.375 | 1 | 160 | 41 | 0 | 3739 | 3898 | 23033 | 22874 | 2.14E-09 | 69.4 | |
| S4-wms-combined_118531 | S9_vs_90850 | 77.778 | 0 | 135 | 28 | 2 | 31257 | 31390 | 2797 | 2664 | 6.07E-13 | 82.4 | |
| S2-wms-combined_99446 | S11_vs_8456 | 94.444 | 0 | 54 | 1 | 1 | 19347 | 19398 | 22391 | 22338 | 3.77E-13 | 82.4 | |
| S2-wms-combined_7756 | S5_vs_86562 | 73.953 | 0 | 215 | 52 | 4 | 1587 | 1799 | 24722 | 24934 | 1.54E-13 | 84.2 | |
| S2-wms-combined_48661 | S9_vf_19284 | 100 | 1 | 43 | 0 | 0 | 944 | 986 | 28724 | 28766 | 1.20E-13 | 80.5 | |
| S17-wms-combined_162786 | S16_vf_173099 | 85.542 | 0 | 83 | 11 | 1 | 62725 | 62806 | 23501 | 23583 | 1.03E-13 | 86.1 | |
| S4-wms-combined_113721 | S7_vs_111831 | 93.103 | 0 | 58 | 3 | 1 | 18807 | 18863 | 3057 | 3000 | 7.22E-14 | 84.2 | |
| S2-wms_192667 | S5_vs_8125 | 83.168 | 0 | 101 | 11 | 5 | 4216 | 4313 | 22124 | 22027 | 5.05E-15 | 87.9 | |
| S8-wms-combined_185611 | S1_vs_21439 | 76.536 | 0 | 179 | 34 | 8 | 32101 | 32275 | 21956 | 22130 | 5.24E-16 | 91.6 | |
| S2-wms-combined_52568 | S1_vs_155183 | 93.75 | 0 | 64 | 4 | 0 | 40704 | 40767 | 1581 | 1518 | 1.54E-17 | 97.1 | |
| S9-wms-combined_71647 | S14_vs_96286 | 95.161 | 0 | 62 | 3 | 0 | 13236 | 13297 | 7771 | 7710 | 1.94E-18 | 99 | |
| S7-wms-combined_143218 | S12_vf_18902 | 89.474 | 1 | 76 | 8 | 0 | 483 | 558 | 7058 | 7133 | 1.80E-18 | 97.1 | |
| S11-wms_32691 | S5_vf_103206 | 90.909 | 0 | 77 | 7 | 0 | 114865 | 114941 | 18187 | 18263 | 2.91E-19 | 104 | |
| S1-wms-combined_197401 | S16_vs_26078 | 83.761 | 0 | 117 | 15 | 3 | 51590 | 51706 | 19779 | 19667 | 7.72E-21 | 108 | |
| S2-wms_140933 | S4_vf_13353 | 85.577 | 0 | 104 | 14 | 1 | 23828 | 23930 | 44241 | 44138 | 6.61E-21 | 108 | |
| S17-wms-combined_179363 | S12_vs_75098 | 71.007 | 3 | 576 | 137 | 26 | 2605 | 3168 | 12004 | 11447 | 2.05E-22 | 111 | |
| S1-wms-combined_40115 | S5_vs_77593 | 92.308 | 3 | 78 | 6 | 0 | 62 | 139 | 98222 | 98145 | 2.63E-23 | 111 | |
| S9-wms-combined_89249 | S9_vs_90859 | 91.011 | 0 | 89 | 4 | 3 | 20484 | 20569 | 24510 | 24597 | 6.02E-24 | 117 | |
| S8-wms-combined_40666 | S5_vs_46394 | 89.216 | 0 | 102 | 5 | 5 | 44263 | 44360 | 8663 | 8762 | 3.69E-25 | 122 | |
| S17-wms_106671 | S7_vs_40259 | 72.235 | 2 | 425 | 108 | 9 | 6592 | 7011 | 2702 | 3121 | 1.22E-25 | 122 | |
| S2-wms-combined_248318 | S5_vf_105728 | 76.754 | 1 | 228 | 49 | 4 | 822 | 1047 | 33418 | 33643 | 3.57E-26 | 124 | |
| S2-wms-combined_180640 | S3_vs_16825 | 91.753 | 1 | 97 | 6 | 2 | 3254 | 3349 | 47899 | 47994 | 7.24E-29 | 134 | |
| S2-wms-combined_89742 | S11_vs_36748 | 86.154 | 1 | 130 | 10 | 5 | 21057 | 21184 | 4352 | 4229 | 5.81E-29 | 134 | |
| S1-wms-combined_143459 | S17_vs_177203 | 91.919 | 1 | 99 | 5 | 1 | 706 | 804 | 19956 | 19861 | 2.38E-29 | 135 | |
| S4-wms_107182 | S5_vf_29332 | 80.628 | 1 | 191 | 33 | 4 | 3577 | 3765 | 22074 | 22262 | 1.90E-32 | 145 | |
| S5-wms-combined_94482 | S16_vs_32764 | 86.62 | 0 | 142 | 12 | 6 | 54562 | 54700 | 42685 | 42822 | 1.42E-33 | 150 | |
| S17-wms-combined_146050 | S16_vs_32764 | 79.654 | 1 | 231 | 34 | 11 | 4341 | 4563 | 42908 | 42683 | 7.44E-35 | 154 | |
| S12-wms-combined_147628 | S16_vs_26078 | 91.379 | 0 | 116 | 6 | 3 | 10620 | 10735 | 19779 | 19668 | 4.04E-35 | 156 | |
| S9-wms_64472 | S17_vs_133231 | 85.276 | 3 | 163 | 13 | 11 | 144 | 300 | 8959 | 8802 | 7.67E-37 | 158 | |
| S5-wms-combined_94786 | S3_vs_50557 | 74.783 | 1 | 460 | 98 | 15 | 29109 | 29559 | 4001 | 4451 | 1.01E-45 | 191 | |
| S17-wms-combined_95360 | S7_vs_134784 | 76.574 | 0 | 397 | 66 | 16 | 42179 | 42551 | 3186 | 2793 | 4.97E-46 | 193 | |
| S8-wms_82880 | S17_vs_110666 | 76.923 | 1 | 351 | 77 | 4 | 23882 | 24230 | 37120 | 36772 | 1.43E-47 | 196 | |
| S1-wms-combined_159544 | S17_vs_177203 | 98.23 | 1 | 113 | 2 | 0 | 254 | 366 | 20342 | 20230 | 1.76E-48 | 198 | |
| S2-wms-combined_135154 | S7_vs_134784 | 90 | 0 | 160 | 10 | 5 | 33499 | 33655 | 594 | 750 | 4.28E-49 | 202 | |
| S5-wms-combined_205046 | S14_vs_9162 | 76.903 | 1 | 381 | 74 | 11 | 14516 | 14889 | 43334 | 43707 | 6.10E-50 | 204 | |
| S1-wms-combined_32863 | S15_vs_42043 | 93.706 | 1 | 143 | 7 | 1 | 8139 | 8281 | 193486 | 193346 | 4.73E-53 | 213 | |
| S4-wms-combined_24659 | S3_vf_67333 | 75.67 | 7 | 448 | 101 | 6 | 3021 | 3464 | 10612 | 11055 | 2.13E-54 | 217 | |
| S2-wms_246296 | S3_vf_108959 | 88.205 | 0 | 195 | 23 | 0 | 31598 | 31792 | 53743 | 53937 | 1.24E-58 | 233 | |
| S2-wms-combined_238929 | S8_vf_208126 | 72.355 | 2 | 879 | 213 | 29 | 18441 | 19304 | 43437 | 42574 | 9.55E-64 | 250 | |
| S12-wms-combined_299872 | S14_vf_58919 | 71.527 | 4 | 1166 | 277 | 49 | 12153 | 13292 | 13146 | 14282 | 9.01E-68 | 263 | |
| S8-wms-combined_211119 | S8_vf_93976 | 81.176 | 1 | 340 | 52 | 11 | 6911 | 7241 | 24527 | 24863 | 7.46E-68 | 263 | |
| S4-wms-combined_64632 | S8_vs_59736 | 78.883 | 1 | 412 | 79 | 6 | 31134 | 31540 | 23239 | 22831 | 2.76E-70 | 272 | |
| S12-wms-combined_263377 | S16_vs_172749 | 76.392 | 1 | 521 | 117 | 5 | 51866 | 52383 | 1363 | 846 | 2.21E-71 | 276 | |
| S12-wms_373775 | S5_vs_145593 | 78.641 | 3 | 412 | 88 | 0 | 9724 | 10135 | 14381 | 13970 | 1.91E-71 | 274 | |
| S17-wms-combined_106262 | S14_vs_117161 | 80.585 | 1 | 376 | 63 | 9 | 6766 | 7136 | 8510 | 8140 | 4.61E-73 | 281 | |
| S7-wms-combined_136642 | S4_vf_12624 | 75.916 | 5 | 573 | 124 | 13 | 6145 | 6710 | 2408 | 2973 | 1.99E-73 | 281 | |
| S5-wms-combined_183113 | S14_vs_74067 | 72.756 | 3 | 947 | 236 | 18 | 27317 | 28252 | 74941 | 75876 | 6.49E-79 | 300 | |
| S11-wms_320555 | S12_vf_69136 | 72.246 | 7 | 1189 | 259 | 43 | 29883 | 31036 | 27457 | 28609 | 1.92E-79 | 302 | |
| S11-wms-combined_46816 | S3_vs_15450 | 79.47 | 2 | 453 | 85 | 7 | 15653 | 16101 | 26767 | 26319 | 2.35E-83 | 315 | |
| S2-wms-combined_58121 | S3_vs_64424 | 89.328 | 1 | 253 | 27 | 0 | 20668 | 20920 | 34002 | 33750 | 1.67E-84 | 318 | |
| S12-wms-combined_334806 | S6_vs_1146 | 73.326 | 1 | 911 | 227 | 13 | 42974 | 43876 | 51895 | 52797 | 3.92E-85 | 322 | |
| S2-wms-combined_176376 | S16_vs_173079 | 80.134 | 1 | 448 | 78 | 11 | 19202 | 19643 | 14178 | 14620 | 5.52E-86 | 324 | |
| S2-wms-combined_143737 | S10_vs_62776 | 83.196 | 1 | 363 | 61 | 0 | 30942 | 31304 | 14020 | 13658 | 1.15E-88 | 333 | |
| S7-wms-combined_157434 | S16_vs_140709 | 78.794 | 1 | 514 | 100 | 9 | 29926 | 30435 | 19561 | 20069 | 6.02E-90 | 337 | |
| S12-wms-combined_270839 | S9_vs_84046 | 73.313 | 2 | 1008 | 235 | 29 | 4381 | 5371 | 28239 | 27249 | 7.16E-91 | 340 | |
| S12-wms-combined_148741 | S9_vs_84046 | 73.42 | 2 | 997 | 231 | 28 | 39287 | 40266 | 27257 | 28236 | 1.50E-91 | 342 | |

| | | | | | | | | | | | | |
|---|---|---|---|---|---|---|---|---|---|---|---|---|
| S4-wms-combined_51079 | S4_vf_12624 | 78.276 | 3 | 580 | 108 | 14 | 11993 | 12563 | 2973 | 2403 | 3.20E-96 | 357 |
| S2-wms-combined_5791 | S7_vs_16344 | 72.511 | 1 | 1175 | 301 | 19 | 5921 | 7084 | 75136 | 73973 | 1.44E-96 | 361 |
| S1-wms-combined_331544 | S4_vf_12624 | 78.448 | 2 | 580 | 107 | 14 | 14954 | 15524 | 2973 | 2403 | 7.32E-98 | 363 |
| S2-wms-combined_93845 | S5_vs_138980 | 77.424 | 1 | 629 | 132 | 9 | 29374 | 29997 | 35421 | 34798 | 1.22E-98 | 366 |
| S12-wms-combined_376326 | S12_vs_106329 | 74.734 | 2 | 847 | 201 | 13 | 44098 | 44938 | 4417 | 5256 | 1.00E-98 | 366 |
| S4-wms-combined_83492 | S3_vs_80151 | 78.415 | 2 | 593 | 114 | 14 | 33415 | 34000 | 144122 | 144707 | 4.92E-101 | 374 |
| S12-wms-combined_264845 | S5_vs_124677 | 77.607 | 6 | 652 | 136 | 9 | 2626 | 3272 | 57017 | 56371 | 3.27E-105 | 387 |
| S7-wms-combined_14529 | S16_vs_15093 | 78.737 | 1 | 649 | 120 | 12 | 36814 | 37449 | 726 | 83 | 3.69E-114 | 418 |
| S12-wms-combined_264077 | S5_vs_124677 | 79.868 | 2 | 606 | 99 | 21 | 19690 | 20285 | 8948 | 9540 | 1.46E-115 | 422 |
| S7-wms_80255 | S3_vf_111893 | 73.715 | 6 | 1187 | 271 | 35 | 10141 | 11306 | 22000 | 23166 | 7.37E-117 | 425 |
| S17-wms-combined_225356 | S3_vf_99626 | 75.276 | 4 | 1088 | 228 | 36 | 327 | 1394 | 25763 | 24697 | 1.98E-133 | 481 |
| S12-wms-combined_52101 | S5_vs_112452 | 74.513 | 9 | 1181 | 277 | 20 | 13598 | 14766 | 2052 | 884 | 8.28E-137 | 492 |
| S5-wms-combined_178432 | S11_vs_98193 | 78.396 | 2 | 773 | 161 | 4 | 2925 | 3694 | 279 | 1048 | 3.70E-138 | 497 |
| S17-wms-combined_126774 | S1_vf_138157 | 86.266 | 1 | 466 | 64 | 0 | 64011 | 64476 | 20571 | 21036 | 8.12E-141 | 507 |
| S8-wms-combined_117281 | S16_vs_18293 | 75.729 | 4 | 1063 | 236 | 21 | 5979 | 7030 | 27940 | 28991 | 2.00E-143 | 514 |
| S2-wms-combined_246480 | S3_vs_127401 | 82.295 | 2 | 610 | 97 | 8 | 24054 | 24655 | 24927 | 24321 | 1.71E-144 | 518 |
| S12-wms-combined_223043 | S14_vf_58919 | 72.439 | 10 | 1923 | 452 | 64 | 3522 | 5406 | 32739 | 30857 | 1.87E-152 | 544 |
| S11-wms-combined_58538 | S11_vs_26767 | 83.533 | 3 | 583 | 96 | 0 | 33450 | 34032 | 3832 | 3250 | 9.21E-153 | 545 |
| S11-wms_55610 | S4_vs_VIRSorter_4959-cat_2 | 80.16 | 1 | 751 | 140 | 6 | 44063 | 44807 | 2577 | 1830 | 9.02E-155 | 553 |
| S12-wms-combined_168259 | S12_vs_86928 | 84.22 | 6 | 583 | 76 | 12 | 14402 | 14976 | 1198 | 1772 | 6.18E-155 | 553 |
| S2-wms-combined_101826 | S3_vf_8164 | 84.764 | 1 | 571 | 85 | 2 | 22938 | 23507 | 12660 | 13229 | 2.65E-160 | 571 |
| S1-wms-combined_120533 | S5_vs_124035 | 80.816 | 8 | 735 | 133 | 7 | 6115 | 6845 | 7955 | 8685 | 1.33E-160 | 569 |
| S4-wms-combined_57123 | S6_vs_46039 | 74.981 | 7 | 1335 | 293 | 35 | 16159 | 17471 | 75449 | 76764 | 4.39E-162 | 577 |
| S12-wms_223235 | S16_vs_48714 | 90.805 | 11 | 435 | 40 | 0 | 2326 | 2760 | 16647 | 17081 | 7.74E-165 | 582 |
| S12-wms-combined_48648 | S8_vs_149700 | 95.418 | 3 | 371 | 16 | 1 | 10825 | 11195 | 67882 | 68251 | 1.31E-166 | 590 |
| S6-wms-combined_65225 | S7_vs_134784 | 83.645 | 3 | 642 | 101 | 3 | 7032 | 7672 | 24267 | 23629 | 1.26E-169 | 601 |
| S2-wms-combined_25727 | S4_vf_31979 | 87.199 | 2 | 539 | 58 | 7 | 33482 | 34013 | 19953 | 20487 | 8.75E-170 | 603 |
| S8-wms_113693 | S4_vf_31979 | 87.546 | 3 | 538 | 56 | 7 | 16119 | 16649 | 19953 | 20486 | 4.93E-173 | 612 |
| S5-wms_67169 | S8_vf_195032 | 70.787 | 9 | 3584 | 843 | 175 | 33149 | 36636 | 2326 | 5801 | 2.75E-178 | 630 |
| S1-wms-combined_340558 | S8_vs_9114 | 91.485 | 2 | 458 | 39 | 0 | 26876 | 27333 | 4427 | 3970 | 1.92E-178 | 630 |
| S9-wms-combined_53991 | S9_vs_67792 | 99.992 | 100 | 61938 | 4 | 1 | 1 | 61938 | 61937 | 1 | 0 | 1.14E+05 |
| S9-wms-combined_97755 | S9_vs_7881 | 99.95 | 57 | 26114 | 13 | 0 | 1 | 26114 | 2152 | 28265 | 0 | 48152 |
| S9-wms-combined_33230 | S5_vf_8965 | 95.799 | 35 | 6284 | 259 | 4 | 7261 | 13541 | 8044 | 1763 | 0 | 10137 |
| S9-wms-combined_2042 | S9_vs_88286 | 100 | 81 | 31342 | 0 | 0 | 1 | 31342 | 44732 | 13391 | 0 | 57878 |
| S9-wms-combined_124857 | S9_vs_50439 | 100 | 28 | 30128 | 0 | 0 | 50939 | 81066 | 1 | 30128 | 0 | 55636 |
| S9-wms-combined_28805 | S7_vs_119679 | 92.087 | 47 | 13535 | 987 | 54 | 30565 | 44043 | 44863 | 31357 | 0 | 18984 |
| S9-wms-combined_109322 | S7_vs_48832 | 83.583 | 2 | 2199 | 340 | 19 | 19556 | 21744 | 35780 | 33593 | 0 | 2041 |
| S9-wms-combined_3461 | S9_vs_90849 | 99.995 | 95 | 36619 | 2 | 0 | 1 | 36619 | 36619 | 1 | 0 | 67612 |
| S9-wms-combined_29611 | S17_vs_127519 | 89.171 | 0 | 868 | 89 | 4 | 345751 | 346617 | 18567 | 17704 | 0 | 1077 |
| S9-wms-combined_25007 | S13_vs_23314 | 74.995 | 20 | 4587 | 990 | 107 | 11442 | 15966 | 22553 | 18062 | 0 | 1971 |
| S9-wms-combined_3878 | S9_vs_90863 | 100 | 91 | 4891 | 0 | 0 | 665 | 5555 | 1 | 4891 | 0 | 9033 |
| S9-wms-combined_105462 | S5_vf_116300 | 84.985 | 2 | 686 | 82 | 16 | 30046 | 30714 | 17730 | 17049 | 0 | 676 |
| S9-wms-combined_20485 | S9_vf_90856 | 100 | 42 | 24912 | 0 | 0 | 49881 | 74792 | 24912 | 1 | 0 | 46004 |
| S9-wms-combined_9931 | S9_vs_90727 | 100 | 100 | 90630 | 0 | 0 | 1 | 90630 | 66187 | 156816 | 0 | 1.67E+05 |
| S9-wms-combined_36459 | S9_vs_48347 | 100 | 99 | 27978 | 0 | 0 | 1 | 27978 | 27978 | 1 | 0 | 51666 |
| S9-wms-combined_105822 | S9_vs_41371 | 79.467 | 35 | 4466 | 862 | 46 | 21501 | 25934 | 4677 | 235 | 0 | 3116 |
| S9-wms-combined_26410 | S11_vs_68151 | 96.563 | 96 | 26502 | 868 | 22 | 427 | 26896 | 34866 | 8376 | 0 | 43853 |
| S9-wms-combined_31456 | S9_vs_21850 | 99.995 | 76 | 36645 | 1 | 1 | 9789 | 46433 | 1 | 36644 | 0 | 67658 |
| S9-wms-combined_37587 | S7_vs_557 | 81.25 | 1 | 1040 | 181 | 11 | 4689 | 5721 | 22885 | 21853 | 0 | 828 |
| S9-wms-combined_5526 | S9_vs_48424 | 100 | 74 | 38027 | 0 | 0 | 1 | 38027 | 16762 | 54788 | 0 | 70223 |
| S9-wms-combined_33229 | S9_vs_28197 | 99.996 | 100 | 23889 | 1 | 0 | 1 | 23889 | 23889 | 1 | 0 | 44110 |
| S9-wms-combined_76274 | S5_vs_63924 | 99.978 | 51 | 27377 | 4 | 2 | 10684 | 38060 | 35637 | 8263 | 0 | 50521 |
| S9-wms-combined_44402 | S5_vs_72532 | 79.495 | 29 | 4394 | 836 | 48 | 2491 | 6844 | 45184 | 40816 | 0 | 3062 |
| S9-wms_19025 | S10_vf_64371 | 99.935 | 100 | 24556 | 16 | 0 | 26716 | 51271 | 59185 | 34630 | 0 | 45258 |
| S9-wms_128232 | S9_vf_23901 | 99.934 | 100 | 4540 | 3 | 0 | 1444 | 5983 | 1 | 4540 | 0 | 8368 |
| S11-wms-combined_41367 | S11_vs_45658 | 99.997 | 99 | 39169 | 1 | 0 | 13 | 39181 | 8205 | 47373 | 0 | 72327 |
| S11-wms-combined_193148 | S6_vs_39065 | 98.05 | 95 | 7743 | 69 | 20 | 3727 | 11429 | 9993 | 17693 | 0 | 13387 |
| S11-wms-combined_179605 | S11_vs_26767 | 89.152 | 3 | 719 | 74 | 2 | 21129 | 21847 | 1 | 715 | 0 | 893 |
| S11-wms-combined_221310 | S7_vs_129858 | 97.802 | 59 | 10375 | 226 | 2 | 22134 | 32507 | 21195 | 10822 | 0 | 17895 |
| S11-wms-combined_14636 | S11_vs_20225 | 99.994 | 99 | 35471 | 2 | 0 | 468 | 35938 | 1 | 35471 | 0 | 65492 |
| S11-wms-combined_83571 | S5_vs_79992 | 99.958 | 76 | 30845 | 1 | 1 | 9777 | 40621 | 1 | 30833 | 0 | 56877 |
| S11-wms-combined_125176 | S7_vf_117697 | 99.374 | 37 | 24610 | 151 | 3 | 1 | 24608 | 883 | 25491 | 0 | 44590 |
| S11-wms-combined_304239 | S11_vs_40945 | 99.983 | 99 | 29259 | 5 | 0 | 15448 | 44706 | 44575 | 15317 | 0 | 54004 |
| S11-wms-combined_194205 | S8_vs_149700 | 92.66 | 39 | 5940 | 430 | 5 | 140 | 6076 | 56070 | 50134 | 0 | 8549 |
| S11-wms-combined_236214 | S7_vs_87279 | 98.021 | 23 | 5912 | 93 | 19 | 75906 | 81805 | 40875 | 46774 | 0 | 10248 |
| S11-wms-combined_264050 | S8_vf_151384 | 97.92 | 51 | 29368 | 591 | 12 | 46455 | 75808 | 1192 | 30553 | 0 | 50830 |

| | | | | | | | | | | | | |
|---|---|---|---|---|---|---|---|---|---|---|---|---|
| S11-wms-combined_140009 | S3_vs_119771 | 78.529 | 12 | 4541 | 823 | 116 | 40919 | 45412 | 9454 | 13889 | 0 | 2844 |
| S11-wms-combined_168048 | S7_vs_21521 | 84.655 | 3 | 1349 | 174 | 20 | 3509 | 4854 | 3115 | 1797 | 0 | 1314 |
| S11-wms-combined_195425 | S11_vs_84848 | 99.976 | 100 | 29097 | 7 | 0 | 1 | 29097 | 2550 | 31646 | 0 | 53694 |
| S11-wms-combined_154845 | S11_vs_5077 | 99.996 | 100 | 25037 | 1 | 0 | 1 | 25037 | 27863 | 2827 | 0 | 46230 |
| S11-wms-combined_17029 | S11_vs_79831 | 99.97 | 100 | 30219 | 9 | 0 | 1 | 30219 | 30219 | 1 | 0 | 55755 |
| S11-wms-combined_3507 | S1_vf_138157 | 90.858 | 6 | 2855 | 243 | 3 | 42288 | 45142 | 14189 | 11353 | 0 | 3810 |
| S11-wms-combined_320710 | S11_vs_104069 | 99.981 | 100 | 26195 | 0 | 3 | 18122 | 44314 | 1 | 26192 | 0 | 48340 |
| S11-wms-combined_101337 | S11_vs_82331 | 99.999 | 100 | 78390 | 1 | 0 | 1 | 78390 | 14307 | 92696 | 0 | 1.45E+05 |
| S11-wms-combined_73204 | S14_vs_74067 | 99.303 | 95 | 64838 | 398 | 48 | 1 | 64812 | 125374 | 190183 | 0 | 1.17E+05 |
| S11-wms-combined_226165 | S11_vs_98193 | 100 | 99 | 33032 | 0 | 0 | 1 | 33032 | 245 | 33276 | 0 | 60999 |
| S11-wms-combined_321938 | S11_vs_3277 | 99.924 | 84 | 35683 | 27 | 0 | 4328 | 40010 | 36230 | 548 | 0 | 65745 |
| S11-wms-combined_198512 | S11_vf_66160 | 99.931 | 86 | 34798 | 24 | 0 | 5708 | 40505 | 34800 | 3 | 0 | 64127 |
| S11-wms-combined_282227 | S5_vs_8847 | 81.746 | 42 | 3380 | 567 | 37 | 5223 | 8590 | 23672 | 20331 | 0 | 2778 |
| S11-wms-combined_267993 | S1_vf_158961 | 76.142 | 4 | 3370 | 692 | 78 | 51281 | 54576 | 10311 | 6980 | 0 | 1666 |
| S11-wms-combined_74983 | S7_vs_134583 | 98.78 | 93 | 24091 | 289 | 3 | 18563 | 42649 | 1044 | 25133 | 0 | 42854 |
| S11-wms-combined_116589 | S7_vs_28150 | 80.844 | 11 | 2892 | 502 | 43 | 18891 | 21759 | 4754 | 1892 | 0 | 2224 |
| S11-wms-combined_48246 | S11_vs_1937 | 99.994 | 71 | 34922 | 2 | 0 | 8306 | 43227 | 1 | 34922 | 0 | 64478 |
| S11-wms-combined_283220 | S11_vs_52577 | 99.943 | 88 | 29970 | 17 | 0 | 4 | 29973 | 29970 | 1 | 0 | 55251 |
| S11-wms-combined_172889 | S7_vs_119679 | 88.612 | 13 | 3776 | 375 | 45 | 25479 | 29228 | 42686 | 46432 | 0 | 4540 |
| S11-wms-combined_117756 | S11_vs_104037 | 100 | 100 | 25717 | 0 | 0 | 1 | 25717 | 18094 | 43810 | 0 | 47491 |
| S11-wms-combined_49632 | S5_vs_69980 | 74.662 | 18 | 3919 | 844 | 124 | 5148 | 8994 | 3846 | 7687 | 0 | 1598 |
| S11-wms-combined_63818 | S11_vf_70015 | 99.964 | 53 | 53033 | 19 | 0 | 10357 | 63389 | 796 | 53828 | 0 | 97829 |
| S11-wms-combined_215723 | S10_vs_37694 | 98.172 | 35 | 16031 | 262 | 18 | 11601 | 27621 | 20356 | 4347 | 0 | 27952 |
| S11-wms-combined_64392 | S12_vs_23796 | 77.726 | 16 | 5841 | 1162 | 123 | 22260 | 28030 | 18578 | 24349 | 0 | 3450 |
| S11-wms-combined_92114 | S11_vs_36748 | 99.993 | 55 | 27110 | 1 | 1 | 1 | 27109 | 32562 | 5453 | 0 | 50050 |
| S11-wms-combined_119215 | S5_vs_69980 | 79.881 | 37 | 6377 | 1101 | 146 | 11419 | 17682 | 7938 | 1631 | 0 | 4501 |
| S11-wms-combined_299954 | S11_vs_104049 | 99.988 | 97 | 16934 | 2 | 0 | 9379 | 26312 | 1 | 16934 | 0 | 31261 |
| S11-wms-combined_217790 | S12_vs_26336 | 99.995 | 74 | 40062 | 2 | 0 | 1 | 40062 | 27031 | 67092 | 0 | 73970 |
| S11-wms-combined_204522 | S11_vs_6458 | 99.974 | 100 | 30685 | 8 | 0 | 1 | 30685 | 35244 | 4560 | 0 | 56621 |
| S11-wms_303398 | S5_vf_21869 | 89.593 | 17 | 1768 | 169 | 13 | 10493 | 12256 | 23361 | 21605 | 0 | 2231 |
| S11-wms_14390 | S11_vf_33117 | 99.997 | 100 | 38385 | 1 | 0 | 1 | 38385 | 14291 | 52675 | 0 | 70879 |
| S11-wms_3116 | S7_vf_39724 | 96.431 | 81 | 31210 | 1004 | 69 | 7366 | 38537 | 36206 | 5069 | 0 | 51361 |
| S11-wms_87293 | S11_vf_104048 | 100 | 100 | 26833 | 0 | 0 | 1 | 26833 | 26833 | 1 | 0 | 49552 |
| S11-wms_6203 | S11_vf_89309 | 99.998 | 100 | 45102 | 1 | 0 | 1 | 45102 | 1 | 45102 | 0 | 83283 |
| S11-wms_330479 | S11_vf_104071 | 99.924 | 100 | 31533 | 0 | 1 | 1 | 31509 | 31533 | 1 | 0 | 58076 |
| S1-wms-combined_324053 | S13_vs_98591 | 99.924 | 90 | 35476 | 6 | 1 | 37 | 35491 | 1341 | 36816 | 0 | 65342 |
| S1-wms-combined_394574 | S2_vs_7306 | 99.967 | 100 | 41917 | 11 | 1 | 4917 | 46833 | 1 | 41914 | 0 | 77325 |
| S1-wms-combined_394628 | S12_vs_137069 | 88.457 | 91 | 5371 | 542 | 69 | 3793 | 9122 | 5574 | 10907 | 0 | 6412 |
| S1-wms-combined_246923 | S5_vs_69083 | 78.779 | 12 | 2686 | 507 | 53 | 4 | 2652 | 1955 | 4614 | 0 | 1744 |
| S1-wms-combined_263562 | S3_vs_70508 | 98.849 | 97 | 14948 | 145 | 19 | 13188 | 28128 | 28635 | 13708 | 0 | 26626 |
| S1-wms-combined_164894 | S14_vs_9162 | 97.934 | 36 | 7502 | 134 | 16 | 6365 | 13857 | 85064 | 92553 | 0 | 12975 |
| S1-wms-combined_231234 | S13_vs_14794 | 99.807 | 84 | 19668 | 29 | 1 | 1 | 19668 | 19659 | 1 | 0 | 36101 |
| S1-wms-combined_132934 | S11_vs_19892 | 88.986 | 59 | 23615 | 2412 | 144 | 15106 | 38628 | 1 | 23518 | 0 | 29024 |
| S1-wms-combined_17651 | S12_vs_164056 | 76.08 | 24 | 1806 | 394 | 31 | 5712 | 7506 | 1849 | 71 | 0 | 907 |
| S1-wms-combined_330329 | S11_vs_3277 | 89.217 | 92 | 26172 | 2692 | 105 | 5646 | 31762 | 7537 | 33633 | 0 | 32577 |
| S1-wms-combined_116938 | S7_vs_111831 | 74.418 | 39 | 4636 | 1043 | 119 | 9880 | 14446 | 28761 | 33322 | 0 | 1858 |
| S1-wms-combined_166392 | S1_vs_133577 | 99.907 | 100 | 19346 | 17 | 1 | 1 | 19346 | 1 | 19345 | 0 | 35624 |
| S1-wms-combined_118167 | S16_vs_18440 | 91.491 | 4 | 2045 | 162 | 11 | 48211 | 50249 | 29851 | 27813 | 0 | 2802 |
| S1-wms-combined_331896 | S5_vs_124035 | 74.938 | 80 | 2019 | 427 | 61 | 468 | 2445 | 7404 | 5424 | 0 | 852 |
| S1-wms-combined_332236 | S13_vs_30167 | 94.773 | 97 | 14139 | 705 | 21 | 703 | 14822 | 53140 | 39017 | 0 | 21985 |
| S1-wms-combined_151708 | S8_vs_132472 | 76.895 | 23 | 3246 | 635 | 85 | 10361 | 13540 | 1 | 3197 | 0 | 1733 |
| S1-wms-combined_52708 | S7_vs_97737 | 78.577 | 23 | 2796 | 548 | 46 | 16339 | 19107 | 53143 | 50372 | 0 | 1797 |
| S1-wms-combined_68998 | S7_vs_135322 | 89.374 | 62 | 11067 | 1113 | 42 | 16349 | 27383 | 22258 | 11223 | 0 | 13863 |
| S1-wms-combined_252439 | S1_vs_91270 | 99.988 | 97 | 16857 | 2 | 0 | 1 | 16857 | 16857 | 1 | 0 | 31119 |
| S1-wms-combined_302166 | S1_vf_158961 | 99.905 | 57 | 42117 | 39 | 1 | 17428 | 59543 | 42248 | 132 | 0 | 77553 |
| S1-wms-combined_285012 | S8_vf_170878 | 85.079 | 1 | 630 | 92 | 2 | 76117 | 76745 | 3477 | 4105 | 0 | 641 |
| S1-wms-combined_186878 | S1_vs_143929 | 99.982 | 100 | 27678 | 5 | 0 | 99 | 27776 | 39654 | 11977 | 0 | 51084 |
| S1-wms-combined_153670 | S13_vs_90967 | 97.139 | 92 | 2552 | 69 | 4 | 1 | 2550 | 1761 | 4310 | 0 | 4305 |
| S1-wms-combined_351889 | S1_vs_155183 | 99.951 | 96 | 30748 | 3 | 1 | 1279 | 32026 | 1 | 30736 | 0 | 56687 |
| S1-wms-combined_23248 | S3_vs_119771 | 78.475 | 6 | 4539 | 829 | 110 | 38167 | 42660 | 13889 | 9454 | 0 | 2833 |
| S1-wms-combined_72167 | S13_vs_13129 | 99.973 | 99 | 18770 | 5 | 0 | 1 | 18770 | 15307 | 34076 | 0 | 34635 |
| S1-wms-combined_304377 | S7_vs_87279 | 97.235 | 43 | 10089 | 274 | 4 | 13057 | 23144 | 20423 | 10339 | 0 | 17080 |
| S1-wms-combined_57370 | S14_vs_117161 | 96.542 | 73 | 9340 | 313 | 10 | 10442 | 19776 | 1302 | 10636 | 0 | 15450 |
| S1-wms-combined_41369 | S1_vf_138157 | 80.771 | 6 | 2231 | 395 | 29 | 16270 | 18486 | 10858 | 8648 | 0 | 1712 |
| S1-wms-combined_156637 | S3_vs_127411 | 93.517 | 10 | 833 | 54 | 0 | 2705 | 3537 | 11335 | 12167 | 0 | 1240 |
| S1-wms-combined_107766 | S16_vs_19968 | 99.82 | 99 | 39354 | 71 | 0 | 166 | 39519 | 54417 | 15064 | 0 | 72280 |

| | | | | | | | | | | | | |
|---|---|---|---|---|---|---|---|---|---|---|---|---|
| S1-wms-combined_92523 | S6_vs_44070 | 88.065 | 16 | 1349 | 157 | 2 | 27795 | 29141 | 16641 | 17987 | 0 | 1596 |
| S1-wms-combined_224578 | S11_vf_34008 | 78.275 | 6 | 1565 | 301 | 32 | 11163 | 12710 | 27010 | 25468 | 0 | 970 |
| S1-wms-combined_357373 | S1_vs_100019 | 99.865 | 78 | 2226 | 2 | 1 | 741 | 2966 | 1 | 2225 | 0 | 4093 |
| S1-wms-combined_373308 | S12_vs_164056 | 90.88 | 35 | 4309 | 365 | 23 | 9116 | 13405 | 4300 | 1 | 0 | 5755 |
| S1-wms-combined_374219 | S5_vs_117417 | 96.517 | 82 | 12374 | 426 | 5 | 4556 | 16926 | 15211 | 2840 | 0 | 20458 |
| S1-wms-combined_227003 | S7_vf_76463 | 83.178 | 10 | 2152 | 335 | 22 | 6598 | 8740 | 34307 | 36440 | 0 | 1943 |
| S1-wms-combined_227917 | S1_vs_133577 | 99.806 | 81 | 14986 | 27 | 2 | 2035 | 17019 | 37683 | 22699 | 0 | 27512 |
| S1-wms-combined_212892 | S5_vf_145604 | 84.853 | 3 | 647 | 85 | 10 | 580 | 1219 | 27596 | 26956 | 0 | 640 |
| S1-wms-combined_131270 | S1_vs_23682 | 99.91 | 99 | 36800 | 9 | 1 | 1 | 36800 | 7677 | 44452 | 0 | 67752 |
| S1-wms-combined_279492 | S6_vs_1146 | 93.135 | 17 | 2389 | 152 | 10 | 5501 | 7884 | 13399 | 15780 | 0 | 3493 |
| S1-wms-combined_164485 | S8_vs_149700 | 92.014 | 8 | 1152 | 85 | 5 | 12560 | 13711 | 67882 | 69026 | 0 | 1611 |
| S1-wms-combined_85225 | S7_vs_73280 | 84.375 | 9 | 2016 | 286 | 24 | 27145 | 29145 | 26605 | 24604 | 0 | 1951 |
| S1-wms-combined_307323 | S3_vs_123834 | 93.732 | 22 | 4403 | 244 | 18 | 6445 | 10824 | 11639 | 7246 | 0 | 6573 |
| S1-wms-combined_94988 | S5_vs_74199 | 78.085 | 13 | 2318 | 397 | 84 | 15923 | 18174 | 20855 | 23127 | 0 | 1363 |
| S1-wms_296850 | S16_vf_6102 | 98.914 | 96 | 34893 | 364 | 5 | 1776 | 36666 | 34880 | 1 | 0 | 62321 |
| S1-wms_313991 | S1_vf_125650 | 99.92 | 57 | 4994 | 4 | 0 | 6014 | 11007 | 6751 | 1758 | 0 | 9201 |
| S1-wms_100010 | S1_vf_22149 | 99.84 | 92 | 22487 | 33 | 2 | 267 | 22751 | 22486 | 1 | 0 | 41323 |
| S1-wms_265285 | S9_vf_90871 | 87.154 | 86 | 15242 | 1829 | 99 | 17945 | 33100 | 15615 | 30813 | 0 | 17180 |
| S1-wms_185046 | S12_vf_55175 | 79.294 | 52 | 3907 | 683 | 94 | 2722 | 6590 | 22432 | 18614 | 0 | 2617 |
| S1-wms_102342 | S10_vf_120738 | 98.645 | 93 | 17051 | 230 | 1 | 18212 | 35261 | 35815 | 18765 | 0 | 30206 |
| S1-wms_4277 | S8_vs_134805 | 87.005 | 83 | 21170 | 2552 | 157 | 7374 | 28453 | 34530 | 13470 | 0 | 23669 |
| S1-wms_153615 | S1_vs_100019 | 99.963 | 98 | 2717 | 1 | 0 | 67 | 2783 | 4858 | 2142 | 0 | 5012 |
| S1-wms_236105 | S1_vf_138157 | 99.948 | 86 | 19146 | 10 | 0 | 3216 | 22361 | 1 | 19146 | 0 | 35301 |
| S1-wms_170115 | S4_vf_31682 | 98.584 | 89 | 15257 | 215 | 1 | 15058 | 30313 | 18014 | 2758 | 0 | 26976 |
| S1-wms_106115 | S13_vf_147396 | 99.975 | 100 | 16296 | 4 | 0 | 10610 | 26905 | 35175 | 18880 | 0 | 30071 |
| S1-wms_223620 | S8_vf_65522 | 98.859 | 91 | 29009 | 283 | 31 | 1102 | 30078 | 1 | 28993 | 0 | 51692 |
| S1-wms_208875 | S13_vf_66508 | 100 | 100 | 18901 | 0 | 0 | 1 | 18901 | 33222 | 14322 | 0 | 34904 |
| S1-wms_225995 | S13_vf_109235 | 99.907 | 96 | 34569 | 31 | 1 | 11727 | 46295 | 39090 | 4523 | 0 | 63658 |
| S1-wms_29288 | S7_vs_135322 | 93.961 | 30 | 2484 | 148 | 2 | 32341 | 34823 | 50304 | 47822 | 0 | 3755 |
| S1-wms_130214 | S1_vf_136526 | 99.994 | 77 | 31148 | 1 | 1 | 1 | 31148 | 1 | 31147 | 0 | 57507 |
| S1-wms_212940 | S11_vf_13033 | 96.799 | 98 | 13994 | 434 | 6 | 11596 | 25578 | 26063 | 12073 | 0 | 23348 |
| S1-wms_394571 | S1_vf_159951 | 99.99 | 100 | 101244 | 10 | 0 | 1 | 101244 | 745 | 101988 | 0 | 1.87E+05 |
| S3-wms-combined_135428 | S3_vs_39940 | 99.972 | 38 | 10828 | 3 | 0 | 17503 | 28330 | 1 | 10828 | 0 | 19980 |
| S3-wms-combined_73262 | S10_vs_58014 | 82.72 | 66 | 8947 | 1407 | 120 | 21376 | 30263 | 23818 | 32684 | 0 | 7829 |
| S3-wms-combined_45305 | S3_vs_119771 | 99.99 | 100 | 19171 | 2 | 0 | 1 | 19171 | 265 | 19435 | 0 | 35392 |
| S3-wms_23958 | S3_vf_82571 | 99.892 | 72 | 25955 | 25 | 3 | 9515 | 35467 | 1 | 25954 | 0 | 47772 |
| S3-wms_94494 | S4_vs_VIRSorter_5698-cat_2 | 96.64 | 5 | 1012 | 30 | 4 | 19338 | 20347 | 15959 | 16968 | 0 | 1677 |
| S3-wms_22744 | S3_vf_99734 | 99.886 | 99 | 46297 | 50 | 2 | 1562 | 47857 | 46295 | 1 | 0 | 85198 |
| S5-wms-combined_26061 | S14_vf_92240 | 98.169 | 94 | 12288 | 213 | 9 | 7330 | 19610 | 23924 | 11642 | 0 | 21435 |
| S5-wms-combined_161191 | S14_vs_117166 | 99.997 | 21 | 30789 | 1 | 0 | 145128 | 175916 | 1 | 30789 | 0 | 56852 |
| S5-wms-combined_134381 | S13_vs_35816 | 97.052 | 52 | 13296 | 346 | 27 | 61694 | 74956 | 24969 | 11687 | 0 | 22340 |
| S5-wms-combined_90315 | S5_vf_18461 | 99.99 | 29 | 29821 | 3 | 0 | 29093 | 58913 | 29821 | 1 | 0 | 55053 |
| S5-wms-combined_134743 | S5_vs_41663 | 100 | 59 | 57172 | 0 | 0 | 40772 | 97943 | 57172 | 1 | 0 | 1.06E+05 |
| S5-wms-combined_72410 | S5_vs_85065 | 100 | 24 | 33638 | 0 | 0 | 71400 | 105037 | 24195 | 57832 | 0 | 62118 |
| S5-wms-combined_206050 | S14_vs_96111 | 99.986 | 51 | 29345 | 4 | 0 | 10578 | 39922 | 29345 | 1 | 0 | 54168 |
| S5-wms-combined_73301 | S14_vs_117153 | 99.995 | 100 | 20824 | 1 | 0 | 1 | 20824 | 13941 | 34764 | 0 | 38450 |
| S5-wms-combined_108794 | S5_vs_40848 | 99.999 | 100 | 177204 | 1 | 0 | 1 | 177204 | 3467 | 180670 | 0 | 3.27E+05 |
| S5-wms-combined_145067 | S5_vs_69980 | 100 | 72 | 22516 | 0 | 0 | 3039 | 25554 | 22516 | 1 | 0 | 41580 |
| S5-wms-combined_207291 | S5_vs_129970 | 99.997 | 56 | 32526 | 0 | 1 | 12401 | 44926 | 32525 | 1 | 0 | 60057 |
| S5-wms-combined_29272 | S8_vs_149700 | 93.628 | 23 | 5901 | 361 | 9 | 6013 | 11902 | 46073 | 40177 | 0 | 8800 |
| S5-wms-combined_29447 | S5_vs_69083 | 100 | 28 | 32116 | 0 | 0 | 220 | 32335 | 32116 | 1 | 0 | 59308 |
| S5-wms-combined_199135 | S5_vs_36605 | 99.996 | 100 | 26478 | 1 | 0 | 1 | 26478 | 26478 | 1 | 0 | 48891 |
| S5-wms-combined_83370 | S5_vs_72532 | 99.92 | 81 | 41058 | 33 | 0 | 4090 | 45147 | 12526 | 53583 | 0 | 75638 |
| S5-wms-combined_83412 | S14_vs_115455 | 99.996 | 99 | 24313 | 1 | 0 | 5079 | 29391 | 1 | 24313 | 0 | 44893 |
| S5-wms-combined_66017 | S5_vs_145581 | 100 | 97 | 23587 | 0 | 0 | 8165 | 31751 | 1 | 23587 | 0 | 43558 |
| S5-wms-combined_66662 | S5_vs_121411 | 100 | 39 | 36545 | 0 | 0 | 57695 | 94239 | 1 | 36545 | 0 | 67486 |
| S5-wms-combined_147993 | S5_vs_59798 | 100 | 99 | 16692 | 0 | 0 | 1392 | 18083 | 1 | 16692 | 0 | 30825 |
| S5-wms-combined_156329 | S5_vs_122618 | 99.068 | 2 | 429 | 4 | 0 | 1 | 429 | 17277 | 17705 | 0 | 771 |
| S5-wms-combined_7292 | S5_vs_20268 | 100 | 100 | 14771 | 0 | 0 | 1 | 14771 | 1 | 14771 | 0 | 27277 |
| S5-wms-combined_59064 | S5_vs_131510 | 99.985 | 100 | 117646 | 18 | 0 | 1 | 117646 | 169132 | 51487 | 0 | 2.17E+05 |
| S5-wms-combined_175145 | S5_vs_109085 | 99.989 | 99 | 26577 | 0 | 2 | 1 | 26576 | 26575 | 1 | 0 | 49059 |
| S5-wms-combined_67650 | S5_vs_123256 | 99.997 | 81 | 86610 | 3 | 0 | 6355 | 92964 | 1 | 86610 | 0 | 1.60E+05 |
| S5-wms-combined_210959 | S5_vs_76500 | 99.998 | 33 | 64407 | 1 | 0 | 122519 | 186925 | 1 | 64407 | 0 | 1.19E+05 |
| S5-wms-combined_78434 | S5_vs_21778 | 99.996 | 99 | 144358 | 6 | 0 | 2087 | 146444 | 157096 | 12739 | 0 | 2.67E+05 |
| S5-wms-combined_204782 | S6_vs_32886 | 99.941 | 99 | 23879 | 11 | 3 | 9302 | 33178 | 14736 | 38613 | 0 | 44016 |
| S5-wms-combined_98202 | S5_vs_142195 | 100 | 73 | 50930 | 0 | 0 | 1 | 50930 | 50930 | 1 | 0 | 94050 |

| | | | | | | | | | | | | |
|---|---|---|---|---|---|---|---|---|---|---|---|---|
| S5-wms-combined_26971 | S11_vf_47632 | 94.477 | 33 | 8076 | 401 | 31 | 14024 | 22071 | 6 | 8064 | 0 | 12401 |
| S5-wms-combined_179241 | S5_vs_74199 | 100 | 85 | 30526 | 0 | 0 | 3467 | 33992 | 30526 | 1 | 0 | 56371 |
| S5-wms-combined_117213 | S5_vs_63924 | 99.989 | 49 | 27314 | 2 | 1 | 1 | 27314 | 8325 | 35637 | 0 | 50422 |
| S5-wms-combined_163165 | S5_vs_65490 | 100 | 94 | 30653 | 0 | 0 | 1 | 30653 | 4908 | 35560 | 0 | 56606 |
| S5-wms-combined_52741 | S5_vs_86562 | 100 | 100 | 23075 | 0 | 0 | 1 | 23075 | 3687 | 26761 | 0 | 42612 |
| S5-wms_62679 | S7_vf_67910 | 99.911 | 96 | 15743 | 12 | 2 | 1 | 15742 | 15742 | 1 | 0 | 28993 |
| S5-wms_10783 | S5_vf_103206 | 99.995 | 98 | 18577 | 1 | 0 | 1 | 18577 | 1 | 18577 | 0 | 34300 |
| S5-wms_164032 | S5_vf_55687 | 95.161 | 100 | 11283 | 513 | 25 | 1 | 11275 | 14211 | 2954 | 0 | 17780 |
| S5-wms_49310 | S4_vf_11308 | 97.519 | 87 | 8264 | 184 | 7 | 920 | 9163 | 604 | 8866 | 0 | 14105 |
| S5-wms_85650 | S5_vs_80496 | 99.979 | 100 | 61026 | 13 | 0 | 1 | 61026 | 60938 | 121963 | 0 | 1.13E+05 |
| S5-wms_210460 | S5_vs_145581 | 98.301 | 4 | 412 | 7 | 0 | 673 | 1084 | 22912 | 22501 | 0 | 723 |
| S5-wms_94965 | S5_vf_29332 | 99.88 | 90 | 19974 | 8 | 2 | 3989 | 23947 | 17638 | 37610 | 0 | 36738 |
| S5-wms_52566 | S5_vs_124035 | 99.49 | 100 | 6468 | 33 | 0 | 1 | 6468 | 6992 | 525 | 0 | 11762 |
| S7-wms-combined_217226 | S7_vs_29595 | 99.97 | 83 | 64326 | 18 | 1 | 1 | 64326 | 64325 | 1 | 0 | 1.19E+05 |
| S7-wms-combined_139683 | S7_vs_5236 | 99.983 | 100 | 24118 | 4 | 0 | 8605 | 32722 | 1 | 24118 | 0 | 44516 |
| S7-wms-combined_71592 | S7_vf_13614 | 100 | 77 | 29799 | 0 | 0 | 8953 | 38751 | 5020 | 34818 | 0 | 55029 |
| S7-wms-combined_235165 | S7_vs_49426 | 99.997 | 100 | 31876 | 1 | 0 | 1 | 31876 | 56890 | 25015 | 0 | 58859 |
| S7-wms-combined_225260 | S7_vs_13903 | 100 | 100 | 15940 | 0 | 0 | 1 | 15940 | 1 | 15940 | 0 | 29436 |
| S7-wms-combined_31068 | S5_vs_69980 | 79.463 | 8 | 2196 | 383 | 54 | 1 | 2167 | 5636 | 3480 | 0 | 1495 |
| S7-wms-combined_20921 | S7_vs_137611 | 99.998 | 80 | 90273 | 2 | 0 | 51998 | 142270 | 1 | 90273 | 0 | 1.67E+05 |
| S7-wms-combined_62119 | S16_vs_127669 | 99.376 | 23 | 22259 | 129 | 8 | 76099 | 98351 | 24833 | 47087 | 0 | 40326 |
| S7-wms-combined_185016 | S5_vs_80496 | 98.562 | 25 | 10431 | 150 | 0 | 1 | 10431 | 27299 | 37729 | 0 | 18432 |
| S7-wms-combined_174935 | S4_vs_VIRSorter_5698-cat_2 | 97.746 | 19 | 8297 | 184 | 3 | 1 | 8295 | 198 | 8493 | 0 | 14283 |
| S7-wms-combined_134452 | S7_vs_30665 | 99.99 | 15 | 9811 | 1 | 0 | 42397 | 52207 | 1 | 9811 | 0 | 18113 |
| S7-wms-combined_134458 | S16_vf_6102 | 74.964 | 15 | 6926 | 1469 | 199 | 22348 | 29143 | 8485 | 1695 | 0 | 2939 |
| S7-wms-combined_21693 | S8_vs_185041 | 98.306 | 78 | 23264 | 385 | 8 | 10483 | 33741 | 49913 | 26654 | 0 | 40769 |
| S7-wms-combined_2122 | S7_vs_138109 | 100 | 76 | 29629 | 0 | 0 | 12049 | 41677 | 29629 | 1 | 0 | 54715 |
| S7-wms-combined_156351 | S7_vs_135679 | 99.964 | 100 | 14020 | 5 | 0 | 1 | 14020 | 14020 | 1 | 0 | 25863 |
| S7-wms-combined_104872 | S3_vs_70508 | 97.47 | 91 | 19839 | 496 | 6 | 8821 | 28657 | 37180 | 57014 | 0 | 33850 |
| S7-wms-combined_238231 | S11_vf_33117 | 78.715 | 1 | 1043 | 206 | 14 | 65821 | 66855 | 36792 | 37826 | 0 | 682 |
| S7-wms-combined_43950 | S10_vs_58014 | 82.625 | 31 | 8944 | 1421 | 114 | 31064 | 39951 | 32684 | 23818 | 0 | 7784 |
| S7-wms-combined_3697 | S7_vs_54322 | 100 | 25 | 49782 | 0 | 0 | 74473 | 124254 | 56805 | 7024 | 0 | 91931 |
| S7-wms-combined_157260 | S6_vs_39025 | 82.11 | 2 | 2152 | 365 | 17 | 112278 | 114421 | 54371 | 52232 | 0 | 1823 |
| S7-wms-combined_74829 | S7_vs_134784 | 100 | 82 | 33154 | 0 | 0 | 4293 | 37446 | 35044 | 1891 | 0 | 61224 |
| S7-wms-combined_14267 | S7_vf_117697 | 100 | 40 | 25491 | 0 | 0 | 26553 | 52043 | 1 | 25491 | 0 | 47074 |
| S7-wms-combined_106253 | S11_vs_104042 | 77.93 | 1 | 1237 | 242 | 20 | 2198 | 3407 | 23479 | 24711 | 0 | 743 |
| S7-wms-combined_239784 | S16_vs_136935 | 86.943 | 4 | 942 | 87 | 29 | 20580 | 21497 | 26788 | 27717 | 0 | 1026 |
| S7-wms-combined_189250 | S7_vs_55526 | 99.995 | 67 | 19404 | 1 | 0 | 15530 | 34933 | 1 | 19404 | 0 | 35827 |
| S7-wms-combined_137858 | S7_vs_97737 | 99.938 | 98 | 50000 | 29 | 2 | 1 | 50000 | 24630 | 74627 | 0 | 92160 |
| S7-wms-combined_5625 | S7_vs_11081 | 99.965 | 34 | 40232 | 14 | 0 | 27422 | 67653 | 1 | 40232 | 0 | 74217 |
| S7-wms-combined_190281 | S7_vs_55526 | 83.12 | 1 | 936 | 144 | 11 | 62131 | 63064 | 18481 | 19404 | 0 | 841 |
| S7-wms-combined_26217 | S7_vs_138173 | 99.969 | 44 | 42156 | 12 | 1 | 37843 | 79998 | 42155 | 1 | 0 | 77774 |
| S7-wms-combined_69024 | S14_vs_117153 | 82.082 | 21 | 4370 | 737 | 38 | 2074 | 6422 | 5255 | 9599 | 0 | 3690 |
| S7-wms-combined_7205 | S7_vs_48708 | 99.969 | 88 | 62174 | 19 | 0 | 600 | 62773 | 62174 | 1 | 0 | 1.15E+05 |
| S7-wms-combined_121062 | S7_vf_9334 | 99.983 | 100 | 23648 | 4 | 0 | 5355 | 29002 | 40566 | 16919 | 0 | 43648 |
| S7-wms-combined_90393 | S7_vs_557 | 100 | 100 | 19692 | 0 | 0 | 1 | 19692 | 5333 | 25024 | 0 | 36365 |
| S7-wms-combined_111004 | S7_vs_135322 | 99.927 | 81 | 35801 | 14 | 10 | 5071 | 40862 | 35798 | 1 | 0 | 65957 |
| S7-wms-combined_90728 | S7_vs_944 | 99.974 | 99 | 23257 | 6 | 0 | 195 | 23451 | 23257 | 1 | 0 | 42915 |
| S7-wms-combined_40779 | S17_vs_127519 | 98.01 | 69 | 5076 | 89 | 10 | 14114 | 19183 | 23904 | 18835 | 0 | 8804 |
| S7-wms-combined_28138 | S7_vs_16344 | 99.982 | 92 | 28325 | 5 | 0 | 2605 | 30929 | 28325 | 1 | 0 | 52279 |
| S7-wms-combined_224523 | S16_vs_34021 | 99.325 | 100 | 19119 | 111 | 1 | 1 | 19119 | 12384 | 31484 | 0 | 34575 |
| S7-wms-combined_163980 | S3_vs_89485 | 99.985 | 89 | 20218 | 3 | 0 | 1677 | 21894 | 11996 | 32213 | 0 | 37320 |
| S7-wms-combined_72070 | S17_vs_117507 | 93.003 | 27 | 6417 | 415 | 25 | 22600 | 28999 | 1239 | 7638 | 0 | 9332 |
| S7-wms-combined_147158 | S7_vs_73280 | 99.937 | 48 | 19161 | 11 | 1 | 3 | 19163 | 19884 | 39043 | 0 | 35316 |
| S7-wms-combined_218379 | S7_vs_47050 | 99.997 | 39 | 36988 | 1 | 0 | 1 | 36988 | 36988 | 1 | 0 | 68299 |
| S7-wms-combined_107679_0 | S7_vs_119679 | 99.823 | 59 | 40003 | 67 | 4 | 1 | 40001 | 11155 | 51155 | 0 | 73475 |
| S7-wms-combined_107679_1 | S10_vs_106834 | 79.276 | 6 | 1935 | 375 | 21 | 17125 | 19045 | 14754 | 12832 | 0 | 1328 |
| S7-wms_113137 | S7_vf_39724 | 100 | 99 | 36889 | 0 | 0 | 3 | 36891 | 1 | 36889 | 0 | 68122 |
| S7-wms_123954 | S17_vf_140653 | 92.053 | 49 | 9664 | 725 | 37 | 2000 | 11646 | 20708 | 11071 | 0 | 13551 |
| S7-wms_53338 | S7_vf_89668 | 99.988 | 99 | 16883 | 2 | 0 | 1 | 16883 | 23 | 16905 | 0 | 31167 |
| S7-wms_157264 | S3_vf_7137 | 99.981 | 100 | 15928 | 3 | 0 | 1 | 15928 | 1 | 15928 | 0 | 29397 |
| S7-wms_239833 | S7_vf_19866 | 99.994 | 99 | 16468 | 1 | 0 | 1 | 16468 | 23 | 16490 | 0 | 30406 |
| S7-wms_127801 | S7_vf_8332 | 100 | 46 | 31505 | 0 | 0 | 36900 | 68404 | 1 | 31505 | 0 | 58179 |
| S7-wms_118607 | S16_vs_86116 | 78.201 | 55 | 7404 | 1399 | 170 | 17996 | 25286 | 16 | 7317 | 0 | 4532 |
| S7-wms_159432 | S7_vf_5439 | 99.997 | 100 | 33004 | 0 | 1 | 1 | 33004 | 33142 | 140 | 0 | 60940 |
| S7-wms_200936 | S8_vs_134805 | 93.064 | 96 | 16364 | 1084 | 41 | 13221 | 29561 | 18185 | 34520 | 0 | 23883 |

| | | | | | | | | | | | | |
|---|---|---|---|---|---|---|---|---|---|---|---|---|
| S7-wms_151112 | S7_vf_114476 | 100 | 38 | 10873 | 0 | 0 | 1 | 10873 | 10873 | 1 | 0 | 20079 |
| S7-wms_244425 | S10_vf_36880 | 97.256 | 97 | 27695 | 731 | 27 | 38974 | 66659 | 78667 | 50993 | 0 | 46906 |
| S7-wms_244866 | S7_vf_27309 | 99.955 | 100 | 26772 | 5 | 4 | 1 | 26772 | 26765 | 1 | 0 | 49365 |
| S17-wms-combined_157468 | S5_vs_65490 | 72.956 | 11 | 2507 | 605 | 61 | 19550 | 22015 | 25978 | 28452 | 0 | 806 |
| S17-wms-combined_233761 | S10_vs_14557 | 93.302 | 11 | 1896 | 127 | 0 | 1 | 1896 | 61788 | 59893 | 0 | 2798 |
| S17-wms-combined_56006 | S17_vs_110666 | 99.943 | 73 | 70350 | 37 | 2 | 12005 | 82353 | 1 | 70348 | 0 | 1.30E+05 |
| S17-wms-combined_56061 | S5_vs_130294 | 76.48 | 12 | 4898 | 975 | 138 | 101121 | 105907 | 26371 | 31202 | 0 | 2499 |
| S17-wms-combined_223748 | S17_vs_133231 | 99.979 | 100 | 4859 | 1 | 0 | 1 | 4859 | 8659 | 3801 | 0 | 8968 |
| S17-wms-combined_79709 | S5_vs_69980 | 79.583 | 19 | 3737 | 658 | 86 | 2955 | 6641 | 3544 | 7225 | 0 | 2577 |
| S17-wms-combined_90378 | S4_vs_VIRSorter_27316-cat_2 | 83.569 | 33 | 4382 | 637 | 69 | 17148 | 21485 | 50311 | 54653 | 0 | 4026 |
| S17-wms-combined_179771 | S17_vs_117507 | 100 | 60 | 25659 | 0 | 0 | 738 | 26396 | 25659 | 1 | 0 | 47384 |
| S17-wms-combined_2234 | S5_vf_122568 | 92.568 | 51 | 15044 | 998 | 90 | 23746 | 38738 | 39202 | 24228 | 0 | 21477 |
| S17-wms-combined_258087 | S16_vf_36146 | 96.296 | 75 | 15495 | 520 | 43 | 3824 | 19293 | 17210 | 1745 | 0 | 25385 |
| S17-wms-combined_136622 | S3_vs_81793 | 95.034 | 8 | 745 | 37 | 0 | 25800 | 26544 | 52631 | 53375 | 0 | 1171 |
| S17-wms-combined_203701 | S8_vs_179208 | 99.821 | 81 | 57584 | 99 | 4 | 1 | 57582 | 57582 | 1 | 0 | 1.06E+05 |
| S17-wms-combined_158354 | S16_vs_136935 | 92.101 | 5 | 2228 | 169 | 7 | 13137 | 15360 | 5816 | 8040 | 0 | 3133 |
| S17-wms-combined_247494 | S5_vs_69980 | 76.493 | 4 | 1842 | 363 | 56 | 16453 | 18256 | 3440 | 1631 | 0 | 939 |
| S17-wms-combined_126600 | S5_vs_7444 | 82.056 | 44 | 3093 | 506 | 45 | 5149 | 8219 | 4266 | 1201 | 0 | 2591 |
| S17-wms-combined_171305 | S17_vs_43307 | 100 | 50 | 19701 | 0 | 0 | 1 | 19701 | 22799 | 42499 | 0 | 36381 |
| S17-wms-combined_248321 | S13_vs_35816 | 98.629 | 35 | 18094 | 242 | 3 | 98275 | 116362 | 24960 | 43053 | 0 | 32034 |
| S17-wms-combined_193958 | S17_vf_97671 | 100 | 18 | 18558 | 0 | 0 | 82525 | 101082 | 19897 | 1340 | 0 | 34271 |
| S17-wms-combined_47860 | S9_vs_4558 | 80.256 | 82 | 6964 | 1286 | 74 | 48108 | 55026 | 44902 | 37983 | 0 | 5160 |
| S17-wms-combined_259811 | S4_vs_VIRSorter_30222-cat_2 | 99.338 | 64 | 16308 | 94 | 13 | 1031 | 17332 | 87349 | 71050 | 0 | 29505 |
| S17-wms-combined_127688 | S7_vf_5439 | 98.554 | 95 | 16534 | 222 | 4 | 29029 | 45560 | 34248 | 50766 | 0 | 29192 |
| S17-wms-combined_260343 | S8_vf_170878 | 81.982 | 40 | 4268 | 736 | 30 | 22384 | 26636 | 26023 | 30272 | 0 | 3591 |
| S17-wms-combined_72837 | S14_vf_117147 | 97.194 | 88 | 12294 | 322 | 13 | 16277 | 28559 | 15918 | 3637 | 0 | 20770 |
| S17-wms-combined_16309 | S16_vs_127905 | 92.096 | 4 | 544 | 43 | 0 | 18098 | 18641 | 18595 | 18052 | 0 | 767 |
| S17-wms-combined_185239 | S8_vs_125717 | 79.349 | 7 | 1167 | 227 | 14 | 30472 | 31631 | 21770 | 22929 | 0 | 808 |
| S17-wms-combined_39687 | S14_vs_3251 | 99.948 | 100 | 19402 | 10 | 0 | 1 | 19402 | 14745 | 34146 | 0 | 35774 |
| S17-wms-combined_140517 | S8_vs_185041 | 74.877 | 15 | 1624 | 370 | 33 | 15158 | 16762 | 21996 | 23600 | 0 | 704 |
| S17-wms-combined_106940 | S7_vf_5801 | 89.4 | 29 | 2783 | 281 | 13 | 19119 | 21894 | 10857 | 8082 | 0 | 3493 |
| S17-wms-combined_186314 | S4_vs_VIRSorter_2744-cat_3 | 84.932 | 5 | 1825 | 218 | 33 | 63551 | 65351 | 12090 | 10299 | 0 | 1794 |
| S17-wms-combined_130215 | S14_vs_30439 | 99.491 | 86 | 18873 | 89 | 5 | 6696 | 25565 | 34044 | 15176 | 0 | 34313 |
| S17-wms-combined_75892 | S13_vs_148294 | 81.511 | 15 | 2396 | 392 | 39 | 20421 | 22775 | 29230 | 31615 | 0 | 1923 |
| S17-wms-combined_175924 | S11_vs_52577 | 75.836 | 34 | 5682 | 1203 | 136 | 13419 | 19005 | 7373 | 12979 | 0 | 2730 |
| S17-wms-combined_199440 | S7_vs_6626 | 97.629 | 80 | 9741 | 186 | 24 | 13840 | 23569 | 44403 | 34697 | 0 | 16667 |
| S17-wms-combined_199746 | S17_vs_116640 | 99.976 | 35 | 20806 | 5 | 0 | 6812 | 27617 | 2 | 20807 | 0 | 38394 |
| S17-wms-combined_66123 | S7_vs_83100 | 99.417 | 84 | 10471 | 61 | 0 | 8814 | 19284 | 1050 | 11520 | 0 | 18999 |
| S17-wms-combined_9864 | S8_vs_149700 | 79.804 | 31 | 4793 | 909 | 52 | 35602 | 40365 | 32563 | 27801 | 0 | 3434 |
| S17-wms-combined_253499 | S17_vs_157209 | 99.974 | 97 | 42973 | 2 | 1 | 576 | 43548 | 43342 | 379 | 0 | 79287 |
| S17-wms_124308 | S14_vs_32499 | 96.585 | 94 | 14552 | 480 | 13 | 1 | 14543 | 40325 | 25782 | 0 | 24103 |
| S17-wms_235536 | S17_vf_21718 | 99.997 | 100 | 30415 | 1 | 0 | 1 | 30415 | 949 | 31363 | 0 | 56161 |
| S17-wms_116248 | S11_vf_104070 | 78.573 | 58 | 10216 | 1951 | 200 | 4676 | 14776 | 12921 | 23013 | 0 | 6519 |
| S17-wms_29082 | S11_vf_34224 | 98.401 | 74 | 29840 | 473 | 3 | 11787 | 41624 | 29838 | 1 | 0 | 52458 |
| S17-wms_254872 | S17_vf_128147 | 99.92 | 99 | 6243 | 5 | 0 | 1 | 6243 | 561 | 6803 | 0 | 11502 |
| S17-wms_266511 | S17_vf_177199 | 99.997 | 100 | 35820 | 0 | 1 | 1 | 35820 | 35819 | 1 | 0 | 66140 |
| S12-wms-combined_321174 | S12_vs_188833 | 99.898 | 100 | 16622 | 17 | 0 | 15500 | 32121 | 1 | 16622 | 0 | 30601 |
| S12-wms-combined_401379 | S12_vs_137069 | 99.99 | 100 | 10338 | 1 | 0 | 1 | 10338 | 10338 | 1 | 0 | 19086 |
| S12-wms-combined_401383 | S12_vs_183891 | 99.975 | 95 | 4005 | 1 | 0 | 1 | 4005 | 4005 | 1 | 0 | 7391 |
| S12-wms-combined_184338 | S8_vs_50431 | 84.462 | 28 | 7929 | 1198 | 28 | 37132 | 45044 | 10404 | 2494 | 0 | 7786 |
| S12-wms-combined_368462 | S8_vs_149700 | 79.304 | 31 | 6610 | 1266 | 92 | 35649 | 42200 | 34366 | 27801 | 0 | 4534 |
| S12-wms-combined_385270 | S12_vs_75098 | 100 | 99 | 41466 | 0 | 0 | 1 | 41466 | 41466 | 1 | 0 | 76574 |
| S12-wms-combined_201125 | S12_vf_60224 | 99.996 | 24 | 54526 | 2 | 0 | 165792 | 220317 | 1 | 54526 | 0 | 1.01E+05 |
| S12-wms-combined_368562 | S5_vs_131510 | 92.463 | 77 | 16798 | 1206 | 51 | 5231 | 21994 | 96448 | 113219 | 0 | 23952 |
| S12-wms-combined_351847 | S8_vs_4153 | 98.193 | 98 | 12067 | 206 | 5 | 876 | 12931 | 1019 | 13084 | 0 | 21065 |
| S12-wms-combined_117628 | S3_vs_122706 | 90.112 | 1 | 627 | 57 | 3 | 57937 | 58559 | 3332 | 2707 | 0 | 809 |
| S12-wms-combined_17617 | S7_vs_6626 | 96.359 | 66 | 15106 | 499 | 39 | 36517 | 51591 | 33990 | 18905 | 0 | 24801 |
| S12-wms-combined_51076 | S5_vs_21778 | 80.346 | 8 | 2310 | 432 | 20 | 67764 | 70066 | 14549 | 12255 | 0 | 1731 |
| S12-wms-combined_151843 | S12_vs_48045 | 99.994 | 26 | 32825 | 2 | 0 | 21039 | 53863 | 32825 | 1 | 0 | 60606 |
| S12-wms-combined_18287 | S7_vs_4956 | 96.39 | 43 | 26818 | 902 | 49 | 45437 | 72211 | 35149 | 8355 | 0 | 44100 |
| S12-wms-combined_320262 | S9_vs_2799 | 97.867 | 73 | 18327 | 385 | 5 | 1 | 18325 | 54267 | 35945 | 0 | 31673 |
| S12-wms-combined_2450 | S5_vs_69083 | 99.981 | 38 | 31347 | 5 | 1 | 50592 | 81938 | 32120 | 775 | 0 | 57852 |
| S12-wms-combined_320727 | S14_vs_74067 | 99.682 | 75 | 52155 | 161 | 5 | 94741 | 146894 | 200667 | 252817 | 0 | 95387 |
| S12-wms-combined_371541 | S11_vs_79831 | 95.928 | 12 | 4273 | 156 | 10 | 29418 | 33678 | 8904 | 4638 | 0 | 6911 |
| S12-wms-combined_120133 | S12_vs_99092 | 100 | 69 | 29908 | 0 | 0 | 1 | 29908 | 5354 | 35261 | 0 | 55230 |
| S12-wms-combined_71235 | S4_vs_VIRSorter_27316-cat_2 | 76.386 | 26 | 3985 | 826 | 94 | 14465 | 18387 | 50326 | 54257 | 0 | 2039 |

| | | | | | | | | | | | | |
|---|---|---|---|---|---|---|---|---|---|---|---|---|
| S12-wms-combined_189323 | S12_vs_23796 | 99.997 | 76 | 63354 | 2 | 0 | 1 | 63354 | 1 | 63354 | 0 | 1.17E+05 |
| S12-wms-combined_190114 | S2_vs_10289 | 79.04 | 29 | 3769 | 719 | 56 | 9589 | 13328 | 11073 | 14799 | 0 | 2518 |
| S12-wms-combined_256299 | S12_vs_111286 | 99.959 | 92 | 49063 | 10 | 3 | 4356 | 53411 | 49194 | 135 | 0 | 90483 |
| S12-wms-combined_156234 | S7_vs_134583 | 98.778 | 65 | 23903 | 285 | 5 | 1 | 23898 | 1233 | 25133 | 0 | 42516 |
| S12-wms-combined_323943 | S7_vs_135322 | 80.742 | 8 | 1698 | 308 | 13 | 29525 | 31214 | 1985 | 299 | 0 | 1306 |
| S12-wms-combined_191570 | S12_vs_150812 | 99.982 | 85 | 22142 | 2 | 2 | 3986 | 26127 | 36712 | 14573 | 0 | 40865 |
| S12-wms-combined_23517 | S1_vs_21439 | 78.102 | 4 | 3046 | 620 | 40 | 21038 | 24061 | 23404 | 26424 | 0 | 1886 |
| S12-wms-combined_375343 | S11_vs_79831 | 98.254 | 17 | 4182 | 73 | 0 | 21169 | 25350 | 18895 | 23076 | 0 | 7319 |
| S12-wms-combined_325418 | S8_vs_115958 | 93.474 | 50 | 12029 | 773 | 12 | 22681 | 34704 | 81002 | 93023 | 0 | 17854 |
| S12-wms-combined_41160 | S12_vs_28661 | 99.777 | 95 | 21498 | 44 | 4 | 483 | 21978 | 27374 | 5879 | 0 | 39430 |
| S12-wms-combined_7174 | S13_vs_30167 | 87.921 | 62 | 9256 | 1050 | 53 | 1723 | 10935 | 36149 | 26919 | 0 | 10837 |
| S12-wms-combined_358010 | S12_vs_167155 | 82.911 | 2 | 749 | 107 | 16 | 49810 | 50551 | 21451 | 22185 | 0 | 654 |
| S12-wms-combined_91231 | S12_vs_99219 | 99.996 | 100 | 49609 | 2 | 0 | 1 | 49609 | 51797 | 2189 | 0 | 91600 |
| S12-wms-combined_107942 | S11_vs_104064 | 96.568 | 99 | 30917 | 1012 | 30 | 2 | 30890 | 26657 | 57552 | 0 | 51169 |
| S12-wms-combined_293354 | S12_vs_94042 | 99.97 | 92 | 36599 | 10 | 1 | 3468 | 40065 | 2435 | 39033 | 0 | 67523 |
| S12-wms-combined_75733 | S12_vf_188846 | 99.966 | 100 | 2940 | 1 | 0 | 1 | 2940 | 2722 | 5661 | 0 | 5424 |
| S12-wms-combined_393703 | S14_vs_67094 | 80.254 | 16 | 1889 | 340 | 29 | 4196 | 6068 | 3379 | 1508 | 0 | 1391 |
| S12-wms-combined_358797 | S3_vs_56292 | 98.859 | 62 | 15689 | 175 | 4 | 300 | 15986 | 52533 | 36847 | 0 | 27977 |
| S12-wms-combined_25459 | S9_vs_2799 | 81.271 | 2 | 1180 | 202 | 17 | 35581 | 36749 | 47629 | 46458 | 0 | 937 |
| S12-wms-combined_359190 | S8_vs_185041 | 91.792 | 45 | 16338 | 1289 | 42 | 36389 | 52695 | 9711 | 26027 | 0 | 22694 |
| S12-wms-combined_259783 | S8_vs_206526 | 99.107 | 98 | 33822 | 294 | 5 | 11597 | 45415 | 1 | 33817 | 0 | 60778 |
| S12-wms-combined_260036 | S4_vf_12614 | 92.864 | 3 | 1107 | 74 | 5 | 2382 | 3485 | 1110 | 6 | 0 | 1602 |
| S12-wms-combined_210942 | S3_vs_127411 | 91.155 | 60 | 3663 | 293 | 20 | 8095 | 11750 | 15600 | 11962 | 0 | 4940 |
| S12-wms-combined_328554 | S12_vs_142625 | 100 | 100 | 43415 | 0 | 0 | 1 | 43415 | 80692 | 37278 | 0 | 80173 |
| S12-wms-combined_329242 | S5_vs_65490 | 79.266 | 11 | 3188 | 594 | 52 | 46056 | 49208 | 25978 | 29133 | 0 | 2163 |
| S12-wms-combined_345948 | S4_vs_VIRSorter_27316-cat_2 | 76.242 | 37 | 4268 | 894 | 104 | 14581 | 18785 | 54536 | 50326 | 0 | 2154 |
| S12-wms-combined_313953 | S12_vs_153780 | 99.998 | 100 | 61118 | 1 | 0 | 1 | 61118 | 61281 | 164 | 0 | 1.13E+05 |
| S12-wms-combined_245755 | S14_vs_117161 | 83.032 | 72 | 8522 | 1262 | 144 | 11240 | 19670 | 23993 | 32421 | 0 | 7557 |
| S12-wms-combined_111524 | S12_vs_26336 | 99.994 | 60 | 48766 | 3 | 0 | 1 | 48766 | 67092 | 18327 | 0 | 90038 |
| S12-wms-combined_297545 | S15_vs_3868 | 99.157 | 39 | 23716 | 195 | 2 | 29802 | 53514 | 33905 | 57618 | 0 | 42682 |
| S12-wms-combined_129554 | S7_vs_6626 | 75.138 | 17 | 4887 | 1100 | 94 | 5629 | 10441 | 42769 | 37924 | 0 | 2187 |
| S12-wms-combined_129799 | S16_vf_97279 | 97.633 | 55 | 27075 | 625 | 11 | 40343 | 67411 | 1 | 27065 | 0 | 46433 |
| S12-wms-combined_282594 | S12_vs_86928 | 99.996 | 44 | 26839 | 1 | 0 | 15499 | 42337 | 41629 | 14791 | 0 | 49557 |
| S12-wms-combined_264425 | S12_vf_79666 | 99.866 | 99 | 41164 | 42 | 1 | 5374 | 46524 | 46218 | 5055 | 0 | 75698 |
| S12-wms-combined_299293 | S11_vf_84303 | 74.794 | 15 | 4360 | 949 | 117 | 52642 | 56920 | 29921 | 25631 | 0 | 1825 |
| S12-wms-combined_98046 | S8_vs_149700 | 78.923 | 2 | 1504 | 285 | 26 | 68293 | 69780 | 44578 | 46065 | 0 | 992 |
| S12-wms-combined_31761 | S6_vs_54606 | 72.725 | 11 | 4462 | 972 | 194 | 20175 | 24533 | 11678 | 7359 | 0 | 1271 |
| S12-wms-combined_99704 | S14_vs_96111 | 96.081 | 51 | 16278 | 628 | 9 | 15873 | 32142 | 1 | 16276 | 0 | 26517 |
| S12-wms-combined_132907 | S12_vs_79785 | 99.565 | 98 | 40727 | 154 | 7 | 626 | 41330 | 1398 | 42123 | 0 | 74206 |
| S12-wms-combined_250551 | S12_vs_141337 | 99.998 | 97 | 40473 | 1 | 0 | 1269 | 41741 | 44369 | 3897 | 0 | 74735 |
| S12-wms-combined_368190 | S12_vf_16374 | 99.997 | 53 | 59768 | 2 | 0 | 3482 | 63249 | 59768 | 1 | 0 | 1.10E+05 |
| S12-wms-combined_157167 | S12_vs_164056 | 99.971 | 88 | 3403 | 1 | 0 | 481 | 3883 | 6664 | 3262 | 0 | 6279 |
| S12-wms-combined_164656 | S12_vs_32669 | 100 | 100 | 24358 | 0 | 0 | 1 | 24358 | 1 | 24358 | 0 | 44981 |
| S12-wms_184276 | S12_vf_173666 | 99.935 | 99 | 30650 | 20 | 0 | 20 | 30669 | 1 | 30650 | 0 | 56490 |
| S12-wms_235125 | S12_vf_55175 | 99.6 | 100 | 12497 | 50 | 0 | 11093 | 23589 | 1 | 12497 | 0 | 22801 |
| S12-wms_17950 | S15_vs_1407 | 98.983 | 92 | 7372 | 74 | 1 | 4499 | 11870 | 1706 | 9076 | 0 | 13197 |
| S12-wms_352846 | S12_vf_188830 | 100 | 100 | 38917 | 0 | 0 | 28954 | 67870 | 1 | 38917 | 0 | 71867 |
| S12-wms_386692 | S12_vf_174374 | 99.945 | 100 | 34590 | 19 | 0 | 1 | 34590 | 34590 | 1 | 0 | 63771 |
| S12-wms_202803 | S8_vf_72773 | 98.54 | 94 | 18358 | 238 | 12 | 10336 | 28663 | 23930 | 42287 | 0 | 32389 |
| S12-wms_336780 | S12_vs_133070 | 98.186 | 8 | 441 | 8 | 0 | 8957 | 9397 | 131663 | 132103 | 0 | 771 |
| S12-wms_193565 | S17_vs_110666 | 79.622 | 31 | 16493 | 2986 | 298 | 1 | 16323 | 33876 | 50163 | 0 | 11490 |
| S12-wms_161239 | S11_vf_104070 | 81.863 | 43 | 4433 | 745 | 51 | 14637 | 19043 | 36162 | 31763 | 0 | 3677 |
| S12-wms_44817 | S5_vf_110171 | 93.802 | 94 | 11036 | 601 | 65 | 23980 | 34968 | 36290 | 47289 | 0 | 16513 |
| S12-wms_179388 | S11_vf_84303 | 97.227 | 96 | 21781 | 577 | 11 | 23677 | 45451 | 34626 | 12867 | 0 | 36851 |
| S12-wms_12658 | S12_vf_69136 | 99.903 | 55 | 32030 | 30 | 1 | 26593 | 58621 | 137 | 32166 | 0 | 58975 |
| S12-wms_15700 | S3_vf_2186 | 97.923 | 99 | 26005 | 519 | 17 | 1 | 25994 | 38190 | 12196 | 0 | 45011 |
| S12-wms_401366 | S12_vf_128794 | 99.96 | 100 | 40062 | 16 | 0 | 5029 | 45090 | 1 | 40062 | 0 | 73892 |
| S12-wms_401372 | S12_vf_188824 | 100 | 100 | 72979 | 0 | 0 | 14932 | 87910 | 87909 | 14931 | 0 | 1.35E+05 |
| S2-wms-combined_207832 | S3_vf_99734 | 97.993 | 92 | 13603 | 268 | 3 | 29467 | 43068 | 23646 | 10048 | 0 | 23603 |
| S2-wms-combined_248329 | S2_vs_4368 | 99.99 | 66 | 20545 | 2 | 0 | 11106 | 31650 | 20983 | 439 | 0 | 37929 |
| S2-wms-combined_31084 | S7_vs_48708 | 77.385 | 31 | 7190 | 1520 | 84 | 11859 | 18997 | 57086 | 49952 | 0 | 4172 |
| S2-wms-combined_196783 | S7_vs_69630 | 83.914 | 11 | 1579 | 246 | 7 | 28042 | 29616 | 6841 | 8415 | 0 | 1502 |
| S2-wms-combined_165833 | S14_vf_58919 | 86.515 | 68 | 12621 | 1602 | 76 | 6614 | 19164 | 20753 | 8163 | 0 | 13786 |
| S2-wms-combined_62377 | S13_vs_152223 | 99.964 | 99 | 60900 | 10 | 1 | 4 | 60903 | 60888 | 1 | 0 | 1.12E+05 |
| S2-wms-combined_72725 | S11_vs_104049 | 94.444 | 46 | 6750 | 346 | 27 | 22333 | 29074 | 6778 | 13506 | 0 | 10360 |
| S2-wms-combined_62519 | S4_vs_VIRSorter_4959-cat_2 | 99.816 | 41 | 34265 | 62 | 1 | 7917 | 42180 | 34360 | 96 | 0 | 62925 |

| | | | | | | | | | | | | |
|---|---|---|---|---|---|---|---|---|---|---|---|---|
| S2-wms-combined_218002 | S6_vs_12945 | 97.878 | 46 | 34587 | 702 | 27 | 41654 | 76226 | 37406 | 2838 | 0 | 59775 |
| S2-wms-combined_166665 | S6_vs_1146 | 98.199 | 89 | 13603 | 230 | 11 | 32492 | 46086 | 36531 | 50126 | 0 | 23749 |
| S2-wms-combined_62839 | S8_vf_187742 | 72.201 | 3 | 5047 | 1238 | 131 | 16568 | 21533 | 57166 | 62128 | 0 | 1395 |
| S2-wms-combined_197577 | S3_vs_127445 | 98.536 | 98 | 43028 | 594 | 28 | 21370 | 64386 | 27698 | 70700 | 0 | 75935 |
| S2-wms-combined_208559 | S9_vs_2799 | 78.521 | 8 | 2542 | 506 | 37 | 33281 | 35801 | 15884 | 18406 | 0 | 1633 |
| S2-wms-combined_229529 | S13_vf_66508 | 94.27 | 1 | 1431 | 70 | 8 | 7979 | 9401 | 35724 | 34298 | 0 | 2178 |
| S2-wms-combined_126134 | S11_vs_104049 | 80.893 | 12 | 2240 | 412 | 14 | 6040 | 8271 | 9655 | 7424 | 0 | 1751 |
| S2-wms-combined_74387 | S16_vs_92150 | 74.1 | 5 | 3278 | 757 | 80 | 36461 | 39694 | 29660 | 26431 | 0 | 1266 |
| S2-wms-combined_147065 | S11_vs_3277 | 98.944 | 21 | 26693 | 282 | 0 | 103892 | 130584 | 30777 | 4085 | 0 | 47731 |
| S2-wms-combined_137119 | S7_vs_134583 | 77.564 | 25 | 7898 | 1614 | 147 | 81703 | 89519 | 17313 | 25133 | 0 | 4623 |
| S2-wms-combined_116221 | S11_vf_47632 | 98.516 | 34 | 17180 | 252 | 2 | 21664 | 38840 | 6 | 17185 | 0 | 30310 |
| S2-wms-combined_157934 | S6_vs_10314 | 74.751 | 2 | 2210 | 474 | 69 | 73807 | 75965 | 7306 | 9482 | 0 | 913 |
| S2-wms-combined_64692 | S13_vs_148294 | 97.681 | 20 | 6424 | 130 | 15 | 37911 | 44330 | 11277 | 17685 | 0 | 11020 |
| S2-wms-combined_127080 | S12_vs_81729 | 92.694 | 64 | 4038 | 258 | 20 | 12280 | 16307 | 23582 | 27592 | 0 | 5788 |
| S2-wms-combined_13246 | S5_vs_30490 | 81.887 | 6 | 8530 | 1382 | 119 | 218362 | 226805 | 16635 | 8183 | 0 | 7042 |
| S2-wms-combined_199384 | S12_vs_14467 | 92.81 | 19 | 1808 | 123 | 7 | 15223 | 17027 | 15899 | 14096 | 0 | 2612 |
| S2-wms-combined_55495 | S1_vs_62409 | 97.533 | 82 | 8997 | 190 | 25 | 2846 | 11826 | 1 | 8981 | 0 | 15356 |
| S2-wms-combined_158821 | S4_vs_VIRSorter_13488-cat_2 | 75.842 | 52 | 5936 | 1220 | 170 | 11107 | 16934 | 537 | 6366 | 0 | 2820 |
| S2-wms-combined_210822 | S5_vs_109527 | 93.541 | 11 | 5063 | 315 | 10 | 40354 | 45409 | 24761 | 29818 | 0 | 7528 |
| S2-wms-combined_24549 | S11_vs_104049 | 89.92 | 9 | 2252 | 221 | 3 | 51988 | 54235 | 29939 | 27690 | 0 | 2896 |
| S2-wms-combined_170845 | S8_vs_185041 | 88.87 | 36 | 5867 | 628 | 21 | 6928 | 12776 | 31885 | 37744 | 0 | 7193 |
| S2-wms-combined_159670 | S1_vs_143929 | 94.342 | 6 | 813 | 44 | 2 | 35637 | 36448 | 15383 | 14572 | 0 | 1245 |
| S2-wms-combined_181391 | S14_vs_3251 | 95.111 | 9 | 3968 | 186 | 5 | 114724 | 118687 | 30183 | 34146 | 0 | 6246 |
| S2-wms-combined_200390 | S9_vs_88286 | 86.188 | 11 | 7689 | 1030 | 21 | 30584 | 38255 | 796 | 8469 | 0 | 8287 |
| S2-wms-combined_233107 | S5_vs_65490 | 98.184 | 72 | 10020 | 163 | 10 | 6176 | 16186 | 20563 | 10554 | 0 | 17477 |
| S2-wms-combined_57293 | S5_vs_130294 | 77.139 | 39 | 4908 | 941 | 144 | 14230 | 19027 | 31207 | 26371 | 0 | 2680 |
| S2-wms-combined_191613 | S1_vs_145792 | 91.144 | 25 | 5375 | 414 | 34 | 14602 | 19921 | 1 | 5368 | 0 | 7232 |
| S2-wms-combined_98888 | S4_vs_VIRSorter_3006-cat_1 | 95.807 | 38 | 13499 | 559 | 4 | 86150 | 99646 | 49541 | 36048 | 0 | 21786 |
| S2-wms-combined_181849 | S7_vs_87279 | 89.372 | 3 | 1289 | 117 | 14 | 6693 | 7968 | 28322 | 29603 | 0 | 1604 |
| S2-wms-combined_201205 | S16_vs_173096 | 96.286 | 66 | 11200 | 390 | 18 | 18021 | 29207 | 30126 | 18940 | 0 | 18354 |
| S2-wms-combined_78670 | S14_vf_56130 | 86.325 | 41 | 5031 | 643 | 35 | 26230 | 31237 | 9963 | 4955 | 0 | 5437 |
| S2-wms-combined_140996 | S5_vf_122568 | 97.59 | 20 | 25930 | 600 | 21 | 113454 | 139370 | 15141 | 41058 | 0 | 44398 |
| S2-wms-combined_58563 | S9_vs_90851 | 82.246 | 43 | 10730 | 1707 | 160 | 34324 | 44958 | 19154 | 8528 | 0 | 9079 |
| S2-wms-combined_174241 | S3_vs_119605 | 93.61 | 33 | 13553 | 834 | 28 | 37436 | 50970 | 28472 | 14934 | 0 | 20201 |
| S2-wms-combined_110215 | S14_vs_9162 | 79.159 | 2 | 1569 | 301 | 21 | 3536 | 5088 | 53481 | 51923 | 0 | 1062 |
| S2-wms-combined_122457 | S11_vs_104049 | 88.67 | 5 | 2639 | 288 | 9 | 22737 | 25367 | 27194 | 29829 | 0 | 3206 |
| S2-wms-combined_244564 | S11_vs_42927 | 72.591 | 18 | 4681 | 1154 | 112 | 47032 | 51648 | 40109 | 44724 | 0 | 1417 |
| S2-wms-combined_6605 | S5_vs_65490 | 79.266 | 6 | 3188 | 594 | 52 | 18271 | 21423 | 25978 | 29133 | 0 | 2163 |
| S2-wms-combined_225908 | S3_vs_122706 | 95.209 | 11 | 12105 | 557 | 19 | 99704 | 111799 | 9501 | 21591 | 0 | 19119 |
| S2-wms-combined_113619 | S13_vs_83447 | 99.941 | 100 | 35571 | 3 | 3 | 1 | 35571 | 21098 | 56650 | 0 | 65555 |
| S2-wms-combined_227800 | S16_vs_34751 | 86.224 | 3 | 980 | 126 | 8 | 1953 | 2926 | 1476 | 500 | 0 | 1053 |
| S2-wms-combined_52006 | S14_vs_97835 | 79.167 | 60 | 1632 | 292 | 32 | 1097 | 2711 | 11 | 1611 | 0 | 1086 |
| S2-wms-combined_4685 | S8_vs_132472 | 92.297 | 25 | 3726 | 265 | 16 | 665 | 4374 | 3720 | 1 | 0 | 5271 |
| S2-wms-combined_207000 | S5_vs_145598 | 94.742 | 45 | 5382 | 277 | 6 | 22354 | 27732 | 38361 | 32983 | 0 | 8366 |
| S2-wms_124416 | S17_vs_110666 | 80.22 | 74 | 20020 | 3563 | 326 | 38430 | 58267 | 30851 | 50655 | 0 | 14665 |
| S2-wms_156439 | S5_vf_8965 | 96.85 | 60 | 18033 | 537 | 18 | 211 | 18231 | 1784 | 19797 | 0 | 30125 |
| S2-wms_53881 | S14_vf_114491 | 99.975 | 97 | 15913 | 3 | 1 | 478 | 16389 | 15913 | 1 | 0 | 29362 |
| S2-wms_23562 | S2_vf_3356 | 100 | 99 | 13546 | 0 | 0 | 1 | 13546 | 13546 | 1 | 0 | 25015 |
| S2-wms_128831 | S13_vf_151045 | 99.112 | 99 | 52952 | 410 | 45 | 33370 | 86290 | 1 | 52923 | 0 | 95125 |
| S2-wms_128962 | S13_vf_11246 | 99.974 | 100 | 72445 | 19 | 0 | 1 | 72445 | 72445 | 1 | 0 | 1.34E+05 |
| S2-wms_79304 | S2_vf_16829 | 100 | 100 | 17153 | 0 | 0 | 5 | 17157 | 17189 | 37 | 0 | 31676 |
| S2-wms_161986 | S3_vf_127405 | 95.487 | 90 | 26346 | 1155 | 30 | 2969 | 29296 | 36208 | 9879 | 0 | 42034 |
| S2-wms_130687 | S16_vs_82116 | 98.576 | 77 | 29631 | 392 | 12 | 1786 | 31389 | 66695 | 37068 | 0 | 52353 |
| S2-wms_110761 | S1_vf_125650 | 99.925 | 99 | 6707 | 5 | 0 | 1 | 6707 | 45 | 6751 | 0 | 12358 |
| S2-wms_6424 | S11_vf_33433 | 97.724 | 94 | 19773 | 428 | 17 | 13740 | 33500 | 75021 | 55259 | 0 | 34001 |
| S2-wms_245232 | S11_vf_10907 | 78.964 | 16 | 2163 | 390 | 47 | 34908 | 37045 | 20907 | 18785 | 0 | 1413 |
| S2-wms_248315 | S2_vf_21511 | 99.996 | 99 | 47745 | 1 | 1 | 16505 | 64248 | 64083 | 16339 | 0 | 88156 |
| S2-wms_248324 | S2_vf_21630 | 100 | 100 | 27232 | 0 | 0 | 1 | 27232 | 27232 | 1 | 0 | 50289 |
| S4-wms-combined_14411 | S4_vs_VIRSorter_6022-circular-cat_[illegible] | 99.995 | 100 | 40647 | 2 | 0 | 2863 | 43509 | 1 | 40647 | 0 | 75050 |
| S4-wms-combined_112093 | S4_vs_VIRSorter_13488-cat_2 | 99.987 | 99 | 15391 | 2 | 0 | 10467 | 25857 | 1 | 15391 | 0 | 28411 |
| S4-wms-combined_42647 | S4_vs_VIRSorter_27316-cat_2 | 77.973 | 27 | 4390 | 861 | 91 | 11786 | 16122 | 54662 | 50326 | 0 | 2652 |
| S4-wms-combined_22012 | S3_vf_99626 | 99.86 | 87 | 23615 | 32 | 1 | 3592 | 27205 | 141 | 23755 | 0 | 43425 |
| S4-wms-combined_57963 | S11_vf_34224 | 98.468 | 22 | 15864 | 238 | 4 | 65681 | 81539 | 29838 | 13975 | 0 | 27944 |
| S4-wms-combined_10758 | S4_vs_VIRSorter_13190-cat_2 | 99.975 | 71 | 59082 | 12 | 3 | 24455 | 83535 | 1 | 59080 | 0 | 1.09E+05 |
| S4-wms-combined_17284 | S4_vs_VIRSorter_16493-cat_2 | 99.974 | 54 | 22682 | 5 | 1 | 32844 | 55524 | 40429 | 17748 | 0 | 41851 |
| S4-wms-combined_121978 | S8_vs_185041 | 91.31 | 24 | 20414 | 1628 | 102 | 59998 | 80312 | 5857 | 26223 | 0 | 27735 |

| | | | | | | | | | | | | |
|---|---|---|---|---|---|---|---|---|---|---|---|---|
| S4-wms-combined_25201 | S7_vs_86377 | 84.896 | 23 | 7018 | 1011 | 44 | 11812 | 18802 | 6996 | 1 | 0 | 7042 |
| S4-wms-combined_47565 | S8_vs_148389 | 97.442 | 5 | 782 | 19 | 1 | 3816 | 4596 | 10885 | 10104 | 0 | 1332 |
| S4-wms-combined_41490 | S5_vs_20500 | 98.984 | 75 | 48236 | 399 | 42 | 6363 | 54558 | 1 | 48185 | 0 | 86276 |
| S4-wms_118690 | S4_vf_10506 | 100 | 100 | 22121 | 0 | 0 | 1 | 22121 | 22121 | 1 | 0 | 40850 |
| S4-wms_161443 | S4_vf_30103 | 99.949 | 100 | 15635 | 1 | 4 | 48 | 15678 | 1 | 15632 | 0 | 28821 |
| S4-wms_50475 | S4_vf_20440 | 99.973 | 63 | 63000 | 14 | 1 | 24181 | 87180 | 77 | 63073 | 0 | 1.16E+05 |
| S4-wms_141607 | S4_vf_18803 | 99.905 | 81 | 28337 | 22 | 2 | 120 | 28451 | 28337 | 1 | 0 | 52174 |
| S4-wms_45128 | S4_vf_4891 | 99.998 | 98 | 40481 | 1 | 0 | 1 | 40481 | 2650 | 43130 | 0 | 74749 |
| S4-wms_73990 | S4_vf_4838 | 100 | 100 | 67580 | 0 | 0 | 1 | 67580 | 26145 | 93724 | 0 | 1.25E+05 |
| S4-wms_62635 | S4_vf_13353 | 100 | 100 | 28687 | 0 | 0 | 15609 | 44295 | 184 | 28870 | 0 | 52975 |
| S6-wms-combined_117756 | S6_vs_56496 | 100 | 97 | 27718 | 0 | 0 | 277 | 27994 | 27718 | 1 | 0 | 51186 |
| S6-wms-combined_135223 | S5_vs_109527 | 99.999 | 96 | 142385 | 1 | 1 | 5529 | 147913 | 1 | 142384 | 0 | 2.63E+05 |
| S6-wms-combined_88414 | S6_vs_10314 | 99.928 | 83 | 83767 | 28 | 16 | 1 | 83750 | 1 | 83752 | 0 | 1.54E+05 |
| S6-wms-combined_745 | S13_vs_9438 | 99.996 | 70 | 25922 | 1 | 0 | 10893 | 36814 | 1 | 25922 | 0 | 47864 |
| S6-wms-combined_82597 | S6_vs_9784 | 99.948 | 28 | 59906 | 25 | 2 | 155325 | 215227 | 1 | 59903 | 0 | 1.10E+05 |
| S6-wms-combined_118427 | S8_vf_170878 | 97.052 | 26 | 21810 | 636 | 5 | 124794 | 146597 | 46644 | 24836 | 0 | 36706 |
| S6-wms-combined_118484 | S14_vs_30439 | 97.081 | 80 | 14630 | 417 | 8 | 49 | 14670 | 33781 | 19154 | 0 | 24642 |
| S6-wms-combined_72228 | S3_vs_17168 | 77.771 | 10 | 3392 | 658 | 73 | 113 | 3439 | 51889 | 55249 | 0 | 1999 |
| S6-wms-combined_119138 | S6_vs_39025 | 99.998 | 93 | 57387 | 1 | 0 | 4197 | 61583 | 1 | 57387 | 0 | 1.06E+05 |
| S6-wms-combined_49909 | S6_vs_1146 | 99.986 | 100 | 28186 | 4 | 0 | 1 | 28186 | 32607 | 4422 | 0 | 52028 |
| S6-wms-combined_113986 | S6_vs_29152 | 100 | 100 | 34147 | 0 | 0 | 1 | 34147 | 46560 | 12414 | 0 | 63058 |
| S6-wms-combined_8508 | S5_vs_38949 | 99.943 | 50 | 27835 | 16 | 0 | 9269 | 37103 | 16281 | 44115 | 0 | 51313 |
| S6-wms-combined_32530 | S6_vs_12945 | 99.989 | 82 | 36605 | 4 | 0 | 8194 | 44798 | 1 | 36605 | 0 | 67575 |
| S6-wms-combined_126236 | S5_vs_124677 | 99.999 | 100 | 69368 | 1 | 0 | 1 | 69368 | 73006 | 3639 | 0 | 1.28E+05 |
| S6-wms-combined_50721 | S5_vs_79992 | 99.996 | 41 | 54138 | 2 | 0 | 77965 | 132102 | 1 | 54138 | 0 | 99963 |
| S6-wms-combined_138710 | S5_vs_130294 | 99.997 | 97 | 38590 | 1 | 0 | 1 | 38590 | 38590 | 1 | 0 | 71257 |
| S6-wms-combined_56464 | S6_vs_53966 | 100 | 62 | 21784 | 0 | 0 | 1 | 21784 | 21784 | 1 | 0 | 40228 |
| S6-wms-combined_110950 | S5_vs_30490 | 99.976 | 99 | 33429 | 6 | 2 | 16 | 33443 | 56501 | 23074 | 0 | 61686 |
| S6-wms-combined_128972 | S5_vs_117417 | 99.584 | 89 | 19695 | 82 | 0 | 2181 | 21875 | 19695 | 1 | 0 | 35916 |
| S6-wms-combined_91039 | S7_vf_117697 | 99.197 | 42 | 25160 | 199 | 3 | 5767 | 30924 | 25491 | 333 | 0 | 45340 |
| S6-wms_54578 | S4_vs_VIRSorter_6022-circular-cat_[illegible] | 94.848 | 62 | 5648 | 257 | 24 | 7344 | 12972 | 25477 | 19845 | 0 | 8787 |
| S6-wms_137591 | S6_vf_6314 | 99.961 | 56 | 7600 | 3 | 0 | 335 | 7934 | 1 | 7600 | 0 | 14019 |
| S6-wms_138342 | S11_vf_104035 | 100 | 100 | 89495 | 0 | 0 | 1 | 89495 | 89495 | 1 | 0 | 1.65E+05 |
| S6-wms_140743 | S5_vf_145585 | 99.99 | 100 | 47998 | 5 | 0 | 1 | 47998 | 47998 | 1 | 0 | 88608 |
| S8-wms-combined_246389 | S8_vs_35308 | 100 | 99 | 38635 | 0 | 0 | 2571 | 41205 | 41153 | 2519 | 0 | 71346 |
| S8-wms-combined_215102 | S3_vs_90341 | 97.755 | 49 | 11091 | 247 | 2 | 42375 | 53464 | 33587 | 22498 | 0 | 19101 |
| S8-wms-combined_276204 | S8_vs_71212 | 100 | 100 | 21132 | 0 | 0 | 1 | 21132 | 19049 | 40180 | 0 | 39024 |
| S8-wms-combined_276211 | S8_vs_88013 | 99.972 | 100 | 24611 | 0 | 1 | 17853 | 42456 | 42463 | 17853 | 0 | 45402 |
| S8-wms-combined_23583 | S8_vs_9114 | 100 | 38 | 40264 | 0 | 0 | 43816 | 84079 | 40264 | 1 | 0 | 74354 |
| S8-wms-combined_81597 | S8_vs_99362 | 99.864 | 100 | 24282 | 30 | 2 | 1 | 24279 | 2538 | 26819 | 0 | 44654 |
| S8-wms-combined_140059 | S7_vs_56529 | 99.991 | 99 | 21727 | 2 | 0 | 1 | 21727 | 10539 | 32265 | 0 | 40112 |
| S8-wms-combined_186054 | S8_vs_149700 | 99.968 | 100 | 53551 | 17 | 0 | 1 | 53551 | 15806 | 69356 | 0 | 98796 |
| S8-wms-combined_25607 | S8_vs_145173 | 99.991 | 100 | 22179 | 2 | 0 | 20576 | 42754 | 1 | 22179 | 0 | 40946 |
| S8-wms-combined_26087 | S15_vs_71968 | 99.404 | 48 | 26856 | 154 | 4 | 28179 | 55030 | 39396 | 12543 | 0 | 48702 |
| S8-wms-combined_164289 | S8_vs_176604 | 99.964 | 100 | 27875 | 9 | 1 | 1 | 27875 | 7709 | 35582 | 0 | 51419 |
| S8-wms-combined_72911 | S5_vs_10428 | 94.32 | 1 | 581 | 33 | 0 | 30990 | 31570 | 23705 | 23125 | 0 | 891 |
| S8-wms-combined_49369 | S7_vs_42563 | 99.488 | 63 | 18957 | 80 | 2 | 10181 | 29120 | 9868 | 28824 | 0 | 34454 |
| S8-wms-combined_234473 | S5_vs_124035 | 99.045 | 99 | 3038 | 19 | 8 | 5 | 3037 | 8442 | 5410 | 0 | 5441 |
| S8-wms-combined_74275 | S8_vs_148389 | 100 | 65 | 26113 | 0 | 0 | 1 | 26113 | 11619 | 37731 | 0 | 48222 |
| S8-wms-combined_270362 | S8_vs_9987 | 99.919 | 84 | 49188 | 36 | 4 | 10377 | 59562 | 49186 | 1 | 0 | 90608 |
| S8-wms-combined_200168 | S7_vs_6626 | 75.956 | 6 | 4891 | 1066 | 91 | 46503 | 51322 | 37917 | 42768 | 0 | 2416 |
| S8-wms-combined_97848 | S10_vs_58014 | 87.952 | 34 | 6134 | 713 | 20 | 793 | 6910 | 6854 | 12977 | 0 | 7210 |
| S8-wms-combined_108917 | S8_vs_115958 | 100 | 99 | 69791 | 0 | 0 | 667 | 70457 | 137050 | 67260 | 0 | 1.29E+05 |
| S8-wms-combined_225114 | S8_vs_179529 | 99.934 | 100 | 51282 | 28 | 3 | 1 | 51279 | 52599 | 1321 | 0 | 94507 |
| S8-wms-combined_134098 | S8_vs_145173 | 76.628 | 13 | 2088 | 415 | 59 | 7680 | 9738 | 36098 | 34055 | 0 | 1085 |
| S8-wms-combined_134284 | S8_vs_208113 | 99.996 | 100 | 26767 | 1 | 0 | 212 | 26978 | 41644 | 14878 | 0 | 49424 |
| S8-wms-combined_226076 | S8_vf_208107 | 99.99 | 100 | 30625 | 2 | 1 | 1 | 30625 | 30624 | 1 | 0 | 56536 |
| S8-wms-combined_169835 | S8_vs_125717 | 99.962 | 100 | 52956 | 20 | 0 | 1 | 52956 | 1 | 52956 | 0 | 97681 |
| S8-wms-combined_239791 | S8_vs_72117 | 99.892 | 100 | 21293 | 6 | 4 | 2 | 21277 | 11812 | 33104 | 0 | 39177 |
| S8-wms-combined_217866 | S8_vs_47265 | 99.996 | 100 | 50262 | 1 | 1 | 42 | 50303 | 18417 | 68677 | 0 | 92804 |
| S8-wms-combined_68780 | S8_vs_208127 | 99.971 | 65 | 20685 | 6 | 0 | 51979 | 72663 | 11808 | 32492 | 0 | 38165 |
| S8-wms-combined_161181 | S4_vs_VIRSorter_4959-cat_2 | 92.664 | 66 | 23173 | 1661 | 31 | 8567 | 31714 | 97 | 23255 | 0 | 33338 |
| S8-wms-combined_246185 | S5_vs_86562 | 85.968 | 2 | 1012 | 126 | 12 | 2 | 1005 | 24638 | 23635 | 0 | 1068 |
| S8-wms_270 | S8_vf_208115 | 100 | 99 | 3507 | 0 | 0 | 1894 | 5400 | 1 | 3507 | 0 | 6477 |
| S8-wms_127191 | S8_vf_163760 | 99.992 | 100 | 38123 | 3 | 0 | 21865 | 59987 | 1 | 38123 | 0 | 70384 |
| S8-wms_58515 | S12_vf_174374 | 98.718 | 33 | 3900 | 46 | 4 | 15400 | 19297 | 33777 | 29880 | 0 | 6922 |

| | | | | | | | | | | | | |
|---|---|---|---|---|---|---|---|---|---|---|---|---|
| S8-wms_244525 | S8_vf_203044 | 100 | 100 | 17376 | 0 | 0 | 1 | 17376 | 17388 | 13 | 0 | 32088 |
| S8-wms_152831 | S8_vf_101283 | 99.99 | 100 | 28619 | 3 | 0 | 1 | 28619 | 29450 | 832 | 0 | 52833 |
| S8-wms_48935 | S8_vf_170878 | 99.98 | 99 | 58755 | 12 | 0 | 1 | 58755 | 58755 | 1 | 0 | 1.08E+05 |
| S8-wms_190781 | S3_vf_127409 | 98.147 | 99 | 14681 | 264 | 6 | 1 | 14674 | 23623 | 38302 | 0 | 25597 |
| S8-wms_30498 | S8_vs_134805 | 99.864 | 80 | 24933 | 34 | 0 | 6294 | 31226 | 37347 | 12415 | 0 | 45855 |
| S8-wms_111056 | S13_vf_3728 | 97.743 | 87 | 22729 | 499 | 12 | 20250 | 42972 | 19557 | 42277 | 0 | 39118 |
| S8-wms_20528 | S8_vf_208126 | 99.977 | 100 | 26226 | 1 | 2 | 1 | 26226 | 26221 | 1 | 0 | 48392 |
| S8-wms_57121 | S8_vf_93976 | 99.993 | 76 | 26682 | 2 | 0 | 7746 | 34427 | 26682 | 1 | 0 | 49262 |

Read_mapping_rates

Reference genome: VLP-HQ-nr-vOTUs: 1231

| Sample | # Cleaned reads (PE) | # Reads mapped to reference nr-vOTUs | Mapping rate (%) | # Unmapped reads | Unmapping rate (%) |
|---|---|---|---|---|---|
| S1-vlp | 35113882 | 12741090 | 36.29% | 22372792 | 63.71% |
| S2-vlp | 56868494 | 53293863 | 93.71% | 3574631 | 6.29% |
| S3-vlp | 174177994 | 120181758 | 69.00% | 53996236 | 31.00% |
| S4-vlp | 47904786 | 38373316 | 80.10% | 9531470 | 19.90% |
| S5-vlp | 65733438 | 31012444 | 47.18% | 34720994 | 52.82% |
| S6-vlp | 68149392 | 55096198 | 80.85% | 13053194 | 19.15% |
| S7-vlp | 81075886 | 65132881 | 80.34% | 15943005 | 19.66% |
| S8-vlp | 183020194 | 121979288 | 66.65% | 61040906 | 33.35% |
| S9-vlp | 71023532 | 59039474 | 83.13% | 11984058 | 16.87% |
| S10-vlp | 73760336 | 22080498 | 29.94% | 51679838 | 70.06% |
| S11-vlp | 67000402 | 34020860 | 50.78% | 32979542 | 49.22% |
| S12-vlp | 85558566 | 39625666 | 46.31% | 45932900 | 53.69% |
| S13-vlp | 60096768 | 31551452 | 52.50% | 28545316 | 47.50% |
| S14-vlp | 61978780 | 24937001 | 40.23% | 37041779 | 59.77% |
| S15-vlp | 32235928 | 11812670 | 36.64% | 20423258 | 63.36% |
| S16-vlp | 74204032 | 41380932 | 55.77% | 32823100 | 44.23% |
| S17-vlp | 58989106 | 7611856 | 12.90% | 51377250 | 87.10% |
| Mean | 76287736.24 | 45286543.94 | 59.36% | 31001192.29 | 40.63% |
| SD | | | 22.34% | | 77.66% |
| NC1 | 360240 | 23180 | 6.43% | 337060 | 93.57% |
| NC2 | 18592 | 2610 | 14.04% | 15982 | 85.96% |
| NC3 | 111664 | 16079 | 14.40% | 95585 | 85.60% |

Reference genome: WMS-HQ-nr-vOTUs: 688

| Sample | # Cleaned reads (PE) | # Reads mapped to reference nr-vOTUs | Mapping rate (%) | # Unmapped reads | Unmapping rate (%) |
|---|---|---|---|---|---|
| S1-wms | 144000416 | 16319573 | 11.33% | 127680843 | 88.67% |
| S2-wms | 148218256 | 17114196 | 11.55% | 131104060 | 88.45% |
| S3-wms | 12026424 | 1244734 | 10.35% | 10781690 | 89.65% |
| S4-wms | 53444298 | 3980159 | 7.45% | 49464139 | 92.55% |
| S5-wms | 186273684 | 21308808 | 11.44% | 164964876 | 88.56% |
| S6-wms | 57739286 | 7700678 | 13.34% | 50038608 | 86.66% |
| S7-wms | 137959384 | 16360994 | 11.86% | 121598390 | 88.14% |
| S8-wms | 90880674 | 11462097 | 12.61% | 79418577 | 87.39% |
| S9-wms | 87343694 | 6561914 | 7.51% | 80781780 | 92.49% |
| S11-wms | 84502272 | 9987117 | 11.82% | 74515155 | 88.18% |
| S12-wms | 140008922 | 14878767 | 10.63% | 125130155 | 89.37% |
| S17-wms | 110271798 | 11923131 | 10.81% | 98348667 | 89.19% |
| Mean | 104389092.33 | 11570180.67 | 11.08% | 92818911.67 | 88.92% |
| SD | | | 1.79% | | 98.21% |

VLP_avg_cov

| vOTUs | NC1_vlp | NC2_vlp | NC3_vlp | S1_vlp_ME | S2_vlp_SHHC | S3_vlp_ME | S4_vlp_SHHC | S5_vlp_ME | S6_vlp_SHHC | S7_vlp_ME | S8_vlp_SHHC | S9_vlp_ME | S10_vlp_SHHC | S11_vlp_ME | S12_vlp_SHHC | S13_vlp_ME | S14_vlp_ME | S15_vlp_SHHC | S16_vlp_SHHC | S17_vlp_ME | Status | Note |
|---|---|---|---|---|---|---|---|---|---|---|---|---|---|---|---|---|---|---|---|---|---|---|
| S17_vf_140653 | 0 | 0 | 0.01 | 1.05 | 2.42 | 12.02 | 0.08 | 3.36 | 1.32 | 3.32 | 3.68 | 0.1 | 15.18 | 2.07 | 4.57 | 3.36 | 8.91 | 6.12 | 9.62 | 81.3 | Shared | 1. The average coverage large than 10 is highlighted in red |
| S17_vf_148303 | 0 | 0 | 0 | 0.39 | 0.31 | 0.84 | 0.11 | 0.91 | 1.06 | 693.64 | 0.88 | 0.13 | 0.93 | 0.34 | 2.62 | 0.31 | 0.36 | 0.22 | 0.92 | 312.48 | ME | 2. By setting a threshold value at 10, those having values > 10 in both ME and SHHC samples are considered as "sha |
| S17_vf_67042 | 0 | 0 | 0 | 0.34 | 0.42 | 0.36 | 0.06 | 0.11 | 0.13 | 0.12 | 0.55 | 0.06 | 0.38 | 0.06 | 0.28 | 0.14 | 0.12 | 0.43 | 5.37 | 192.46 | ME | |
| S17_vf_111931 | 0.03 | 0 | 0 | 0.57 | 0.02 | 0.42 | 0.18 | 0.22 | 0.06 | 0.27 | 0.91 | 0.07 | 0.25 | 1.34 | 0.99 | 0.37 | 14.25 | 0.24 | 3.34 | 80.79 | ME | |
| S17_vf_97671 | 0.01 | 0 | 0.01 | 2.15 | 0.02 | 0.52 | 0.55 | 3.81 | 0.59 | 2.91 | 8.34 | 2.22 | 1.48 | 3.88 | 3.35 | 1.29 | 2.7 | 3.31 | 4.67 | 33.52 | ME | |
| S17_vf_46388 | 0.01 | 0 | 0 | 0.87 | 0.04 | 0.63 | 8.31 | 0.16 | 0.16 | 0.17 | 0.58 | 0.12 | 0.58 | 0.66 | 0.43 | 0.71 | 0.28 | 0.45 | 106.98 | 222.67 | Shared | |
| S17_vf_172132 | 0 | 0 | 0 | 0.3 | 0.04 | 0.36 | 0.01 | 0.15 | 0.22 | 619.23 | 0.97 | 0.08 | 0.29 | 0.08 | 1.42 | 0.2 | 0.11 | 0.16 | 1.05 | 288.85 | ME | |
| S17_vf_128147 | 0 | 0 | 0 | 1.65 | 4.55 | 24.48 | 0.08 | 9.02 | 1.95 | 11.6 | 3.91 | 0.3 | 33.05 | 4.5 | 8.63 | 8.03 | 24.31 | 14.07 | 20.81 | 50.82 | Shared | |
| S17_vf_77057 | 0 | 0 | 0.01 | 0.37 | 0.68 | 0.69 | 0.13 | 0.39 | 0.16 | 395.03 | 0.77 | 0.12 | 0.84 | 0.39 | 0.89 | 0.44 | 0.35 | 0.26 | 0.63 | 154.25 | ME | |
| S17_vf_174033 | 0 | 0 | 0 | 1.68 | 0.04 | 0.64 | 0.14 | 0.19 | 0.12 | 0.18 | 0.82 | 0.13 | 2.66 | 0.28 | 4.86 | 0.63 | 0.4 | 0.47 | 23.94 | 55.63 | Shared | |
| S17_vf_174203 | 0 | 0 | 0 | 0.41 | 0.19 | 0.87 | 0.12 | 0.28 | 0.27 | 481.8 | 1.3 | 0.13 | 0.66 | 0.28 | 2.03 | 0.49 | 0.34 | 0.18 | 0.73 | 343.07 | ME | |
| S17_vf_35138 | 0 | 0 | 0 | 0.85 | 0.01 | 0.47 | 18.34 | 0.24 | 0.88 | 0.21 | 0.48 | 0.27 | 0.91 | 0.46 | 1.17 | 0.23 | 0.31 | 0.27 | 12.9 | 45.31 | ME | |
| S17_vf_79151 | 0 | 0 | 0 | 0.26 | 0.02 | 0.53 | 0.17 | 0.27 | 0.09 | 107.18 | 0.8 | 0.09 | 1.58 | 0.1 | 0.63 | 0.46 | 0.31 | 0.26 | 0.49 | 61.23 | ME | |
| S17_vf_36452 | 0 | 0 | 0 | 6.07 | 1.45 | 0.37 | 0.05 | 0.2 | 0.06 | 0.57 | 0.78 | 0.16 | 0.36 | 0.31 | 1.16 | 0.44 | 116.2 | 0.28 | 15.14 | 563.74 | ME | |
| S17_vf_21718 | 0 | 0 | 0 | 3.24 | 0.4 | 19.18 | 0.88 | 1.56 | 1.16 | 2.82 | 3.54 | 7.88 | 21.12 | 5.41 | 7.68 | 1.36 | 58.28 | 1.35 | 17.12 | 141.82 | ME | |
| S17_vf_81232 | 0 | 0 | 0 | 0.95 | 0.08 | 0.41 | 36.61 | 0.25 | 0.16 | 134.28 | 157.96 | 0.09 | 1.69 | 0.43 | 1.09 | 0.58 | 0.34 | 0.26 | 0.81 | 61.74 | Shared | |
| S17_vf_177197 | 0.01 | 0 | 0 | 2.32 | 2.91 | 4.75 | 29.77 | 2.81 | 2.63 | 3.22 | 21725.43 | 0.09 | 0.73 | 0.38 | 0.55 | 506.01 | 0.33 | 0.37 | 1.21 | 29.14 | Shared | |
| S17_vf_177199 | 0 | 0 | 0 | 0.61 | 0.01 | 0.21 | 0.09 | 0.09 | 0.06 | 0.06 | 0.97 | 0.19 | 0.26 | 0.51 | 0.33 | 0.15 | 0.22 | 0.16 | 15.66 | 734.13 | ME | |
| S17_vf_177200 | 0.06 | 0.04 | 0 | 312.8 | 0.06 | 0.55 | 0.08 | 0.22 | 0.1 | 0.13 | 0.55 | 0.1 | 0.64 | 0.13 | 0.25 | 34.93 | 0.19 | 0.74 | 107.46 | 214.04 | Shared | |
| S17_vs_138036 | 0 | 0 | 0 | 0.66 | 0.03 | 0.75 | 0.13 | 0.25 | 0.17 | 0.19 | 1.07 | 0.22 | 0.66 | 0.84 | 0.69 | 0.38 | 0.38 | 0.23 | 6.61 | 343.02 | ME | |
| S17_vs_103009 | 0.18 | 0.04 | 0 | 711.72 | 0.15 | 1.25 | 0.1 | 0.39 | 0.23 | 0.36 | 162.59 | 0.6 | 0.43 | 0.29 | 1.26 | 233.34 | 0.42 | 1.15 | 343.22 | 686.05 | Shared | |
| S17_vs_177196 | 0.01 | 0 | 0 | 0.4 | 0.01 | 0.24 | 0.3 | 0.2 | 0.27 | 0.1 | 1.58 | 0.12 | 0.37 | 0.09 | 0.29 | 0.2 | 0.14 | 0.24 | 56.29 | 110.08 | ME | |
| S17_vs_177202 | 0 | 0 | 0 | 0.96 | 0.24 | 0.3 | 0.07 | 0.16 | 0.09 | 1.17 | 0.51 | 0.11 | 1.11 | 0.36 | 0.26 | 0.24 | 0.36 | 4.18 | 1.74 | 42.94 | ME | |
| S17_vs_177203 | 0 | 0 | 0 | 0.86 | 0.05 | 0.43 | 0.06 | 0.26 | 0.17 | 126.22 | 0.41 | 0.06 | 0.28 | 0.35 | 0.49 | 0.28 | 0.79 | 0.56 | 0.68 | 31.24 | ME | |
| S17_vs_177210 | 0.02 | 0 | 0 | 0.92 | 0.02 | 10.97 | 0.01 | 0.08 | 0.13 | 0.13 | 0.32 | 0.09 | 1.01 | 0.6 | 0.27 | 0.4 | 0.3 | 0.21 | 1.81 | 22.37 | ME | |
| S17_vs_133231 | 0 | 0 | 0 | 0.58 | 0.04 | 0.37 | 0.02 | 0.29 | 0.41 | 0.18 | 0.63 | 0.19 | 0.62 | 2 | 1.92 | 0.71 | 0.2 | 0.35 | 0.88 | 30.36 | ME | |
| S17_vs_52890 | 0.12 | 0.01 | 0 | 82 | 1.21 | 46.76 | 0.35 | 8.06 | 24.86 | 19.73 | 54.65 | 14.73 | 48.2 | 16.34 | 44.7 | 73.29 | 26.18 | 30 | 74.95 | 328.2 | Shared | |
| S17_vs_157209 | 0.01 | 0 | 0 | 0.64 | 0.01 | 0.67 | 0.14 | 0.34 | 0.23 | 0.1 | 2.08 | 0.58 | 0.51 | 0.51 | 0.73 | 0.19 | 0.43 | 0.51 | 54.44 | 1741.89 | Shared | |
| S17_vs_31654 | 0.01 | 0 | 0 | 0.46 | 0.04 | 2.48 | 0.18 | 0.57 | 0.19 | 0.21 | 1.09 | 0.12 | 0.71 | 0.18 | 0.76 | 0.61 | 0.39 | 0.77 | 1.09 | 32.83 | ME | |
| S17_vs_2079 | 0 | 0 | 0 | 31.29 | 0.01 | 39.28 | 4.91 | 0.92 | 1.19 | 1.49 | 0.92 | 0.33 | 3.08 | 11.2 | 1.92 | 5.02 | 1.48 | 45.36 | 11.37 | 174.85 | Shared | |
| S17_vs_39515 | 0 | 0 | 0 | 0.33 | 0.03 | 1.48 | 0.54 | 0.28 | 0.18 | 4.57 | 0.59 | 0.1 | 1.79 | 67.41 | 0.52 | 0.35 | 10.96 | 0.33 | 5.24 | 59.22 | ME | |
| S17_vs_127519 | 0.02 | 0 | 0 | 1.63 | 0.08 | 1.72 | 40.17 | 10.8 | 2.89 | 2.11 | 0.93 | 0.23 | 0.35 | 1.11 | 0.7 | 1.53 | 33.4 | 0.51 | 15.82 | 97.3 | Shared | |
| S17_vs_121041 | 0 | 0 | 0 | 43.06 | 0.32 | 0.87 | 1.17 | 4.32 | 3.85 | 4.25 | 2.73 | 0.43 | 1.04 | 0.98 | 0.35 | 4.77 | 4.92 | 0.48 | 35.98 | 55.87 | Shared | |
| S17_vs_41845 | 0 | 0 | 0 | 0.56 | 0.05 | 0.38 | 0.06 | 0.3 | 0.17 | 3.16 | 0.98 | 0.12 | 0.28 | 0.95 | 0.28 | 0.44 | 1.45 | 0.27 | 2.45 | 44.5 | ME | |
| S17_vs_160502 | 0 | 0 | 0 | 0.87 | 0.06 | 1.28 | 0.35 | 0.19 | 0.15 | 0.91 | 0.49 | 0.24 | 0.75 | 1.2 | 0.88 | 0.55 | 0.32 | 0.27 | 4.1 | 122.46 | ME | |
| S17_vs_116640 | 0 | 0 | 0 | 0.58 | 0.02 | 0.23 | 0.05 | 0.16 | 0.12 | 0.11 | 0.86 | 0.05 | 0.16 | 0.09 | 0.26 | 0.16 | 0.28 | 0.4 | 0.48 | 19.9 | ME | |
| S17_vs_43307 | 0 | 0 | 0 | 0.37 | 0.06 | 0.38 | 0.06 | 0.18 | 0.06 | 2.59 | 3.45 | 0.06 | 0.33 | 3.99 | 1.79 | 0.62 | 0.19 | 0.19 | 12.38 | 79.43 | Shared | |
| S17_vs_58025 | 0 | 0 | 0 | 1.08 | 1.48 | 2.54 | 0.08 | 1.37 | 1.1 | 1.6 | 10406.98 | 0.11 | 0.26 | 0.37 | 0.51 | 0.79 | 0.25 | 0.18 | 0.34 | 14.16 | Shared | |
| S17_vs_117507 | 0.01 | 0 | 0 | 1.12 | 0.06 | 3.79 | 0.13 | 5.03 | 1.76 | 15.28 | 6.77 | 0.11 | 36.52 | 1.88 | 11.9 | 19.31 | 20.69 | 0.31 | 7.68 | 40.45 | Shared | |
| S17_vs_110666 | 0.01 | 0.01 | 0 | 1.54 | 0.1 | 3.74 | 1.12 | 5.72 | 3.76 | 3.54 | 3.89 | 0.4 | 0.81 | 0.69 | 1.18 | 0.93 | 0.86 | 0.91 | 3.77 | 43.31 | ME | |
| S17_vs_73830 | 0 | 0 | 0 | 1.29 | 0.01 | 0.5 | 0.17 | 0.36 | 0.19 | 0.2 | 0.98 | 0.12 | 0.74 | 0.31 | 0.7 | 0.36 | 0.39 | 0.43 | 4.6 | 114.81 | ME | |
| S17_vs_8690 | 0 | 0 | 0 | 1.01 | 0.02 | 0.34 | 1 | 0.38 | 0.45 | 1.87 | 1.98 | 0.06 | 0.49 | 17.49 | 2.3 | 0.29 | 0.27 | 0.28 | 1.3 | 55.34 | ME | |
| S9_vf_4115 | 0 | 0 | 0.02 | 0.36 | 0.06 | 0.59 | 0.08 | 1.66 | 0.11 | 0.13 | 1.46 | 114.53 | 0.46 | 0.27 | 0.49 | 0.22 | 0.46 | 0.2 | 0.43 | 0.71 | ME | |
| S9_vf_26805 | 0 | 0 | 0 | 1.25 | 0.09 | 2.14 | 0.1 | 0.31 | 0.88 | 0.06 | 0.49 | 19.85 | 0.48 | 0.17 | 0.26 | 0.09 | 1.85 | 0.22 | 0.51 | 0.87 | ME | |
| S9_vf_19284 | 0 | 0 | 0.01 | 10.81 | 0.03 | 2.96 | 0.06 | 0.75 | 0.22 | 0.36 | 2.08 | 145.81 | 0.9 | 1.69 | 4.26 | 2.04 | 1.28 | 0.38 | 1.29 | 2.98 | ME | |
| S9_vf_79919 | 0 | 0 | 0.02 | 0.39 | 0.04 | 0.79 | 0.1 | 0.39 | 0.14 | 0.37 | 0.95 | 225.2 | 0.78 | 0.3 | 0.48 | 0.36 | 0.82 | 0.31 | 0.81 | 1.31 | ME | |
| S9_vf_64844 | 0.02 | 0 | 0.03 | 91.44 | 0.16 | 34.81 | 113.08 | 7.57 | 1.25 | 16.99 | 43.46 | 440.57 | 2.03 | 0.72 | 1.45 | 25.1 | 33.44 | 0.66 | 11.31 | 6.39 | Shared | |
| S9_vf_57277 | 0 | 0 | 0.01 | 10.88 | 0.16 | 7.74 | 0.41 | 1.79 | 0.12 | 1.43 | 18.19 | 198.8 | 0.25 | 10.07 | 1.05 | 2.62 | 6.2 | 1.78 | 15.79 | 3.33 | Shared | |
| S9_vf_34736 | 0 | 0 | 0 | 0.35 | 0.42 | 0.39 | 0.01 | 0.27 | 0.12 | 0.09 | 0.2 | 16.44 | 0.69 | 0.53 | 0.26 | 0.13 | 0.24 | 0.14 | 0.52 | 0.71 | ME | |
| S9_vf_934 | 0 | 0 | 0 | 0.2 | 0 | 0.37 | 0 | 0.14 | 0.12 | 0.04 | 0.23 | 25.87 | 0.22 | 0.02 | 0.18 | 0.12 | 0.25 | 0.28 | 0.24 | 0.41 | ME | |
| S9_vf_73311 | 0 | 0 | 0.01 | 0.48 | 0.06 | 0.97 | 0.16 | 0.37 | 0.2 | 0.2 | 0.6 | 226.33 | 0.8 | 0.64 | 0.9 | 0.57 | 0.4 | 0.55 | 0.53 | 1.34 | ME | |
| S9_vf_23901 | 0 | 0 | 0 | 1.4 | 0.44 | 1.74 | 0.3 | 0.18 | 0.11 | 0.26 | 0.41 | 134.27 | 1.04 | 0.45 | 1.32 | 0.41 | 0.26 | 0.73 | 0.98 | 1.11 | ME | |
| S9_vf_76769 | 0 | 0 | 0.02 | 0.41 | 0.02 | 0.79 | 0.1 | 0.26 | 0.13 | 0.13 | 1.01 | 384.53 | 0.89 | 0.43 | 3.57 | 0.33 | 0.38 | 18.17 | 0.49 | 0.94 | Shared | |
| S9_vf_70236 | 0 | 0 | 0 | 0.61 | 0.04 | 0.2 | 0.02 | 0.18 | 0.14 | 0.14 | 0.26 | 175.55 | 0.23 | 0.27 | 0.3 | 0.31 | 0.22 | 0.29 | 0.27 | 0.5 | ME | |
| S9_vf_13507 | 0 | 0 | 0.01 | 1.83 | 0.19 | 1.42 | 92.81 | 1 | 0.51 | 0.45 | 61.57 | 50.59 | 9.36 | 0.62 | 0.99 | 6.08 | 1.47 | 1.23 | 1.44 | 3.03 | Shared | |
| S9_vf_51656 | 0 | 0 | 2.04 | 0.63 | 0.05 | 26.98 | 1825.52 | 0.39 | 0.21 | 0.26 | 4.8 | 33667.86 | 5.02 | 5.42 | 5.46 | 0.42 | 4.52 | 2.69 | 4.37 | 9.22 | Shared | |
| S9_vf_75363 | 0 | 0 | 0 | 0.55 | 5.25 | 0.55 | 0.05 | 0.49 | 0.19 | 0.22 | 0.66 | 20.04 | 0.98 | 0.22 | 0.97 | 0.24 | 0.36 | 0.29 | 0.65 | 1.03 | ME | |
| S9_vf_15012 | 0 | 0 | 0 | 0.67 | 0.02 | 0.87 | 0.16 | 0.31 | 0.16 | 0.2 | 0.78 | 23.24 | 0.56 | 0.46 | 0.9 | 0.57 | 0.59 | 0.36 | 0.53 | 0.78 | ME | |
| S9_vf_90847 | 0 | 0 | 0.04 | 0.62 | 0.04 | 1.2 | 0.19 | 0.3 | 0.18 | 0.21 | 1.11 | 629.09 | 1.01 | 0.39 | 0.84 | 0.63 | 0.53 | 0.49 | 5.54 | 1.2 | ME | |
| S9_vf_90852 | 0 | 0 | 0 | 0.26 | 0.01 | 0.35 | 0 | 0.19 | 0.15 | 0.09 | 0.11 | 29.39 | 0.46 | 0.2 | 0.28 | 0.01 | 0.17 | 0.24 | 0.32 | 0.6 | ME | |
| S9_vf_90856 | 0 | 0 | 0 | 0.56 | 0.03 | 2.63 | 0.11 | 0.73 | 0.09 | 0.59 | 1.14 | 79.66 | 0.48 | 2.03 | 0.35 | 0.36 | 0.32 | 0.21 | 1.2 | 2.4 | ME | |
| S9_vf_90866 | 0 | 0 | 0.09 | 0.87 | 0.05 | 3.89 | 0.1 | 1.84 | 0.29 | 0.22 | 0.57 | 554.23 | 0.61 | 0.23 | 0.62 | 0.84 | 0.38 | 0.46 | 0.71 | 1.92 | ME | |
| S9_vf_90871 | 0 | 0 | 0.01 | 5.7 | 0.17 | 0.24 | 0.02 | 0.78 | 0.09 | 0.69 | 3.35 | 24.66 | 0.33 | 4.08 | 1.87 | 2.63 | 0.31 | 1.47 | 4.36 | 1.12 | ME | |
| S9_vs_45812 | 0.01 | 0 | 0 | 0.83 | 0.03 | 0.74 | 0.08 | 0.65 | 0.36 | 0.44 | 0.66 | 137.76 | 0.55 | 0.27 | 0.45 | 0.51 | 1.09 | 0.44 | 0.44 | 1.16 | ME | |
| S9_vs_88286 | 0 | 0 | 0.1 | 1.62 | 0.02 | 2.77 | 0.12 | 0.5 | 0.15 | 0.19 | 2.46 | 1860.08 | 0.65 | 0.67 | 0.74 | 0.63 | 2.56 | 0.44 | 0.53 | 1.19 | ME | |
| S9_vs_50439 | 0 | 0 | 0.03 | 6.17 | 0.29 | 5.11 | 1.05 | 0.6 | 0.27 | 8.38 | 9.22 | 936.33 | 0.43 | 2.8 | 2.81 | 1.58 | 35.92 | 0.25 | 1.86 | 3.77 | ME | |
| S9_vs_5676 | 0 | 0 | 0 | 0.87 | 0.06 | 0.62 | 0.01 | 0.21 | 0.13 | 0.17 | 0.45 | 21.91 | 0.53 | 0.1 | 0.34 | 0.35 | 0.34 | 0.23 | 0.24 | 0.48 | ME | |
| S9_vs_66877 | 0 | 0 | 0 | 0.81 | 0.01 | 0.55 | 0.14 | 1.32 | 1 | 0.56 | 0.8 | 17.82 | 0.29 | 0.92 | 0.28 | 0.19 | 0.9 | 0.23 | 0.34 | 0.43 | ME | |
| S9_vs_75050 | 0 | 0 | 0.01 | 0.71 | 0.04 | 6.28 | 0.06 | 0.33 | 0.17 | 0.18 | 0.84 | 36.63 | 0.52 | 0.22 | 0.35 | 0.31 | 0.36 | 0.34 | 2.03 | 0.96 | ME | |
| S9_vs_48424 | 0 | 0 | 0.17 | 22.71 | 3.86 | 3.65 | 0.06 | 0.95 | 1.18 | 0.8 | 46.65 | 3226.36 | 1.31 | 0.76 | 1.99 | 8.46 | 0.56 | 9.14 | 2.97 | 3 | Shared | |
| S9_vs_90846 | 0 | 0 | 0.01 | 0.83 | 0.03 | 0.47 | 0.06 | 10.24 | 0.46 | 0.24 | 0.91 | 75.25 | 0.5 | 0.91 | 0.35 | 0.25 | 0.39 | 0.39 | 1.57 | 0.84 | ME | |
| S9_vs_90848 | 0 | 0 | 0 | 1.2 | 0.02 | 1.56 | 0.18 | 15.04 | 15.05 | 0.81 | 0.58 | 206.53 | 0.94 | 0.92 | 1.32 | 0.54 | 7.85 | 0.56 | 6.32 | 3.2 | Shared | |
| S9_vs_90849 | 0 | 0 | 0.11 | 1.03 | 0.03 | 1.74 | 0.06 | 2.4 | 0.21 | 0.25 | 0.54 | 1905.28 | 0.86 | 0.52 | 0.76 | 0.46 | 0.51 | 0.61 | 0.62 | 1.2 | ME | |
| S9_vs_90850 | 0.01 | 0 | 0 | 0.79 | 0.03 | 0.45 | 0.14 | 0.42 | 0.2 | 0.28 | 0.67 | 19.14 | 0.8 | 0.4 | 0.25 | 0.48 | 0.4 | 0.43 | 0.58 | 0.96 | ME | |
| S9_vs_90851 | 0 | 0 | 0.01 | 2.72 | 0.12 | 2.19 | 0.41 | 3.77 | 0.1 | 1.19 | 1.04 | 245.81 | 0.93 | 1.05 | 2.53 | 0.96 | 1.2 | 7.47 | 1.05 | 1.81 | ME | |
| S9_vs_90857 | 0.01 | 0 | 0 | 1.05 | 0.01 | 0.29 | 0.09 | 0.65 | 0.08 | 0.28 | 0.71 | 18.33 | 0.54 | 0.18 | 0.45 | 0.48 | 0.6 | 0.19 | 0.3 | 0.6 | ME | |
| S9_vs_90859 | 0 | 0 | 0.05 | 0.45 | 0.04 | 1.49 | 0.11 | 2.64 | 0.22 | 2.06 | 1.73 | 1293.28 | 0.83 | 1.73 | 1.56 | 0.42 | 1.52 | 0.23 | 0.53 | 1.23 | ME | |
| S9_vs_90860 | 0.01 | 0 | 0 | 0.65 | 0.11 | 0.46 | 0.1 | 0.48 | 0.12 | 0.96 | 0.81 | 21.71 | 0.47 | 0.26 | 0.43 | 0.43 | 1.15 | 0.26 | 0.99 | 0.77 | ME | |
| S9_vs_90863 | 0 | 0 | 0 | 0.09 | 0 | 0.59 | 0 | 0.15 | 0.11 | 0.35 | 0.81 | 140.2 | 0.36 | 0.33 | 0.19 | 0.09 | 0.21 | 0.14 | 0.24 | 0.51 | ME | |
| S9_vs_7881 | 0 | 0 | 0 | 1.21 | 0.05 | 1.07 | 0.11 | 0.5 | 0.18 | 0.6 | 0.89 | 190.11 | 2.12 | 0.78 | 0.32 | 0.61 | 0.9 | 0.51 | 0.78 | 1.73 | ME | |
| S9_vs_4343 | 0 | 0 | 0 | 30.66 | 0.01 | 1.33 | 0.06 | 1.77 | 0.2 | 0.63 | 0.55 | 19.89 | 1.43 | 0.39 | 0.69 | 3.02 | 0.69 | 1.09 | 5.71 | 12.28 | ME | |
| S9_vs_84046 | 0.01 | 0 | 0.01 | 0.67 | 0 | 0.35 | 0.09 | 0.96 | 0.09 | 0.25 | 0.48 | 73.7 | 0.36 | 0.14 | 0.28 | 0.5 | 0.92 | 0.26 | 0.45 | 0.55 | ME | |

| | | | | | | | | | | | | | | | | | | | | | |
|---|---|---|---|---|---|---|---|---|---|---|---|---|---|---|---|---|---|---|---|---|---|
| S9_vs_19689 | 0.01 | 0 | 0 | 1.86 | 0.03 | 0.48 | 0.06 | 0.23 | 0.2 | 1.09 | 0.47 | 15.12 | 0.33 | 0.53 | 1.17 | 0.39 | 0.35 | 0.16 | 0.46 | 1.24 | ME |
| S9_vs_34898 | 0 | 0 | 0 | 0.45 | 0.18 | 0.41 | 0.05 | 1.1 | 1.9 | 0.88 | 0.59 | 17.33 | 0.62 | 1.92 | 0.35 | 0.25 | 0.84 | 0.56 | 0.27 | 0.8 | ME |
| S9_vs_4558 | 0 | 0 | 0.01 | 0.57 | 0.09 | 0.37 | 0.03 | 1.42 | 1.81 | 0.21 | 0.4 | 33.09 | 0.51 | 0.22 | 0.29 | 0.28 | 0.33 | 0.31 | 0.39 | 4.82 | ME |
| S9_vs_46498 | 0 | 0 | 0 | 1.25 | 0.04 | 0.73 | 0.11 | 0.37 | 0.2 | 0.73 | 1.51 | 61.14 | 1.07 | 0.64 | 0.5 | 0.35 | 3.66 | 0.54 | 0.61 | 1.4 | ME |
| S9_vs_27813 | 0 | 0 | 0.02 | 1.28 | 2.96 | 0.58 | 1.52 | 7.49 | 6.63 | 11.07 | 1.06 | 282.15 | 0.57 | 2.17 | 1.31 | 20.94 | 2.87 | 0.34 | 4.37 | 0.95 | ME |
| S9_vs_69322 | 0 | 0 | 0 | 0.97 | 0.02 | 0.84 | 0.16 | 0.97 | 0.12 | 3.36 | 1.65 | 15.45 | 0.6 | 0.75 | 0.71 | 0.57 | 0.45 | 0.35 | 0.38 | 0.9 | ME |
| S9_vs_58086 | 0 | 0 | 0 | 3.04 | 0.02 | 0.51 | 0.09 | 2.66 | 0.46 | 0.31 | 0.7 | 22.39 | 1.59 | 0.52 | 0.32 | 0.38 | 1.18 | 0.33 | 0.55 | 1.63 | ME |
| S9_vs_28197 | 0 | 0 | 0 | 0.66 | 0.03 | 0.42 | 0.07 | 0.21 | 0.21 | 0.13 | 0.28 | 23.53 | 0.57 | 13.24 | 0.44 | 0.33 | 0.22 | 0.27 | 12.03 | 0.91 | Shared |
| S9_vs_1966 | 0 | 0 | 0 | 2.03 | 0.03 | 0.49 | 0.01 | 2.19 | 0.12 | 2.03 | 0.43 | 16.73 | 0.51 | 0.91 | 11.06 | 0.63 | 1.17 | 0.73 | 0.96 | 2.43 | Shared |
| S9_vs_58402 | 0 | 0.01 | 0.01 | 2.65 | 0.09 | 6.17 | 0.02 | 1.88 | 0.17 | 2.24 | 1.17 | 15.62 | 0.45 | 0.21 | 1.59 | 0.72 | 10.04 | 0.34 | 0.98 | 0.68 | ME |
| S9_vs_73764 | 0 | 0 | 0 | 0.37 | 0.01 | 0.33 | 0.02 | 0.22 | 0.06 | 0.29 | 0.61 | 15.58 | 0.22 | 0.15 | 0.25 | 0.41 | 0.29 | 0.15 | 0.86 | 0.6 | ME |
| S9_vs_88847 | 0 | 0 | 0.02 | 0.84 | 0.29 | 201.2 | 1.26 | 0.23 | 0.15 | 0.23 | 0.51 | 19.73 | 0.33 | 0.1 | 0.76 | 0.34 | 2.88 | 1.06 | 0.25 | 0.34 | ME |
| S9_vs_32588 | 0.01 | 0 | 0.01 | 0.62 | 0.06 | 2.14 | 0.06 | 3.06 | 0.17 | 11.27 | 0.32 | 94.74 | 0.54 | 1.11 | 0.26 | 0.23 | 0.68 | 0.27 | 1.17 | 5.03 | ME |
| S9_vs_62803 | 0 | 0 | 0 | 0.98 | 0.05 | 0.46 | 0.15 | 0.38 | 0.19 | 0.61 | 0.48 | 39.43 | 0.4 | 0.42 | 0.4 | 0.36 | 0.98 | 8 | 0.37 | 0.9 | ME |
| S9_vs_44331 | 0.01 | 0 | 0 | 1.07 | 0.03 | 0.58 | 0.44 | 0.48 | 0.35 | 0.24 | 0.51 | 23.36 | 0.73 | 18.72 | 0.3 | 0.34 | 1.16 | 0.35 | 0.66 | 2.62 | ME |
| S9_vs_55738 | 0 | 0 | 0.01 | 0.76 | 0.05 | 0.47 | 0.05 | 0.5 | 0.26 | 0.51 | 0.44 | 19.83 | 0.67 | 1.76 | 0.33 | 0.2 | 34.18 | 0.16 | 0.34 | 0.79 | ME |
| S9_vs_2799 | 0.01 | 0 | 0.01 | 17.83 | 10.31 | 13.56 | 0.41 | 29.86 | 19.66 | 13.79 | 134.6 | 19.68 | 9.03 | 7.26 | 71.37 | 6.8 | 1.43 | 40.41 | 80.52 | 11.49 | Shared |
| S9_vs_21850 | 0 | 0 | 0 | 1.13 | 0.06 | 0.96 | 4.32 | 0.88 | 1.83 | 0.62 | 1.59 | 22.64 | 2.21 | 0.33 | 0.36 | 1.11 | 0.92 | 2.57 | 0.69 | 1.31 | ME |
| S9_vs_10424 | 0 | 0 | 0 | 3.08 | 0.08 | 3.03 | 0.64 | 1.47 | 1.65 | 0.74 | 5.51 | 15.68 | 0.41 | 0.76 | 0.35 | 0.49 | 23.22 | 7.3 | 8.88 | 3.61 | ME |
| S9_vs_63485 | 0 | 0 | 0 | 0.55 | 0.01 | 7.12 | 0.09 | 0.2 | 0.11 | 0.12 | 0.48 | 42.94 | 0.61 | 0.21 | 0.34 | 0.29 | 0.32 | 0.21 | 0.38 | 0.69 | ME |
| S9_vs_48240 | 0 | 0 | 0 | 0.68 | 0.03 | 0.34 | 0.05 | 0.4 | 0.21 | 0.47 | 0.63 | 16.97 | 0.26 | 0.56 | 0.27 | 0.27 | 0.59 | 0.23 | 1.07 | 0.76 | ME |
| S9_vs_37437 | 0 | 0 | 0 | 0.65 | 0.01 | 3.02 | 0.06 | 0.92 | 0.44 | 1.21 | 0.48 | 14.75 | 0.38 | 0.24 | 0.27 | 0.25 | 2.19 | 0.23 | 0.47 | 0.92 | ME |
| S9_vs_48347 | 0 | 0 | 0.32 | 0.74 | 0.01 | 5.22 | 0.07 | 1.29 | 1.67 | 0.12 | 0.58 | 5976.89 | 1.15 | 1.32 | 1.39 | 0.21 | 0.96 | 0.58 | 0.96 | 1.86 | ME |
| S9_vs_14235 | 0.01 | 0 | 0.01 | 0.69 | 0.01 | 0.49 | 0.03 | 0.68 | 0.09 | 1.59 | 0.5 | 15.67 | 0.38 | 1.08 | 0.33 | 0.28 | 1.15 | 0.24 | 0.45 | 1.13 | ME |
| S9_vs_56354 | 0.01 | 0 | 0.01 | 0.8 | 0.5 | 0.29 | 0.33 | 5.1 | 0.1 | 0.41 | 0.88 | 159.95 | 0.33 | 0.71 | 3.43 | 0.42 | 0.32 | 0.13 | 0.31 | 0.44 | ME |
| S9_vs_41371 | 0 | 0 | 0 | 0.7 | 0.04 | 0.96 | 0.06 | 0.46 | 0.16 | 0.37 | 1.8 | 13.88 | 0.6 | 0.21 | 0.27 | 0.37 | 0.44 | 0.39 | 1.4 | 0.88 | ME |
| S9_vs_26179 | 0 | 0 | 0 | 1.85 | 0.05 | 3.29 | 0.09 | 15.46 | 7.85 | 2.91 | 0.88 | 12.81 | 1.3 | 1.23 | 0.67 | 0.55 | 3.32 | 0.49 | 0.89 | 1.73 | ME |
| S9_vs_75290 | 0 | 0 | 0 | 0.42 | 0.04 | 0.29 | 0.19 | 0.14 | 0.12 | 0.07 | 0.4 | 12.47 | 0.25 | 0.2 | 0.23 | 0.17 | 0.23 | 0.21 | 0.13 | 0.4 | ME |
| S9_vs_67792 | 0.01 | 0 | 0 | 1.16 | 0.02 | 0.49 | 0.08 | 2.6 | 0.41 | 0.95 | 0.49 | 25.46 | 0.78 | 0.31 | 0.51 | 0.32 | 5.26 | 0.36 | 0.81 | 5.98 | ME |
| S9_vs_83108 | 0 | 0 | 0.01 | 0.49 | 0.02 | 8.19 | 0.38 | 0.23 | 0.12 | 0.13 | 0.32 | 172.97 | 1.13 | 0.39 | 0.5 | 0.42 | 0.56 | 1.41 | 0.39 | 0.82 | ME |
| S9_vs_90727 | 0.01 | 0 | 0 | 1.46 | 0.16 | 1.86 | 0.19 | 6.01 | 0.79 | 3.08 | 80.01 | 45.39 | 1.62 | 19.12 | 1 | 1.12 | 4.04 | 0.72 | 1.61 | 3.33 | Shared |
| S9_vs_42609 | 0 | 0 | 0 | 0.46 | 0.14 | 2.22 | 0.11 | 0.41 | 0.29 | 0.36 | 2.91 | 10.91 | 2.67 | 2.09 | 2.12 | 0.56 | 1.38 | 1.03 | 1.86 | 2.12 | ME |
| S11_vf_58587 | 0 | 0 | 0 | 75.99 | 4.08 | 1.13 | 0.01 | 0.08 | 0.16 | 3.51 | 0.1 | 0.39 | 0.2 | 201.73 | 0.15 | 1.48 | 0.03 | 0.2 | 1.18 | 3.1 | ME |
| S11_vf_13033 | 0 | 0.05 | 0.06 | 407.99 | 0.23 | 904.28 | 71.32 | 2.09 | 2.47 | 53.45 | 55.96 | 1.3 | 0.71 | 62.87 | 204.54 | 100.64 | 0.85 | 22.66 | 7.9 | 16.75 | Shared |
| S11_vf_33117 | 0.01 | 0 | 0.01 | 0.7 | 0.02 | 0.3 | 0.15 | 1.15 | 0.61 | 1.24 | 0.85 | 0.25 | 1.32 | 35.13 | 1.39 | 0.38 | 1.17 | 0.22 | 0.92 | 0.73 | ME |
| S11_vf_33433 | 0.01 | 0 | 0 | 0.53 | 72.15 | 0.35 | 0.26 | 0.19 | 0.48 | 0.29 | 0.43 | 0.21 | 0.54 | 495.64 | 163.28 | 0.31 | 0.33 | 0.27 | 0.34 | 0.83 | Shared |
| S11_vf_66160 | 0 | 0 | 0 | 1.27 | 0.03 | 0.82 | 0.41 | 3.44 | 0.44 | 3.66 | 3 | 1.51 | 0.74 | 50.13 | 6.14 | 1.86 | 4.7 | 2.08 | 11.29 | 1.79 | Shared |
| S11_vf_92391 | 0 | 0 | 0 | 0.79 | 0.07 | 0.45 | 0.08 | 0.37 | 3.32 | 0.26 | 0.47 | 1.37 | 1.46 | 3997.25 | 109.94 | 0.3 | 1.09 | 0.45 | 0.82 | 2.15 | Shared |
| S11_vf_66522 | 0 | 0 | 0 | 0.64 | 0.02 | 0.15 | 0.05 | 0.2 | 0.09 | 0.12 | 0.96 | 0.07 | 0.29 | 20.56 | 0.5 | 0.32 | 0.14 | 0.14 | 1.72 | 0.4 | ME |
| S11_vf_27845 | 0 | 0 | 0 | 0.55 | 0.02 | 0.24 | 0.03 | 0.14 | 0.1 | 9.38 | 0.41 | 0.05 | 0.29 | 48.98 | 13.78 | 0.17 | 0.13 | 0.14 | 9.01 | 0.4 | Shared |
| S11_vf_34008 | 0 | 0 | 0 | 0.64 | 0.04 | 2.61 | 0.04 | 5.96 | 0.31 | 0.17 | 1.1 | 0.56 | 0.42 | 94.8 | 0.35 | 0.32 | 1.47 | 0.19 | 0.26 | 0.62 | ME |
| S11_vf_34224 | 0 | 0 | 0 | 11.83 | 0.05 | 129.09 | 18.42 | 0.36 | 0.49 | 51.23 | 0.83 | 0.26 | 3.24 | 95.97 | 2.27 | 1.94 | 0.17 | 2.55 | 0.55 | 8.75 | Shared |
| S11_vf_99751 | 0 | 0 | 0 | 0.52 | 0.19 | 0.17 | 0.05 | 0.17 | 0.11 | 0.07 | 0.24 | 0.04 | 0.32 | 17.62 | 0.17 | 0.88 | 0.21 | 0.13 | 0.17 | 0.4 | ME |
| S11_vf_47632 | 0.01 | 0 | 0 | 26.75 | 0.05 | 5.52 | 213.94 | 45.74 | 15.79 | 3.06 | 135.99 | 0.31 | 1.1 | 49.63 | 60.45 | 18.99 | 1.03 | 93.98 | 10.84 | 0.73 | Shared |
| S11_vf_28515 | 0 | 0 | 0 | 3.45 | 0.04 | 13.78 | 0.09 | 4.59 | 0.62 | 0.2 | 31.43 | 0.38 | 33.06 | 618 | 0.65 | 9.77 | 4.03 | 1.41 | 0.57 | 1.08 | Shared |
| S11_vf_28645 | 0 | 0 | 0 | 0.94 | 0 | 1.54 | 0.13 | 0.21 | 0.29 | 0.34 | 2.78 | 0.1 | 8.8 | 285.18 | 0.4 | 2.09 | 0.35 | 0.27 | 0.46 | 2.75 | ME |
| S11_vf_3790 | 0 | 0 | 0 | 0.74 | 0.03 | 0.95 | 0.2 | 0.41 | 0.26 | 0.24 | 1 | 0.14 | 1.5 | 63.95 | 0.87 | 0.53 | 0.75 | 0.51 | 0.81 | 1.38 | ME |
| S11_vf_95067 | 0 | 0 | 0 | 0.71 | 0.04 | 0.27 | 0.24 | 0.24 | 0.41 | 0.33 | 0.44 | 0.15 | 0.94 | 289.81 | 0.45 | 0.34 | 0.21 | 0.29 | 0.57 | 0.58 | ME |
| S11_vf_68693 | 0 | 0 | 0 | 0.67 | 0.02 | 0.33 | 0.01 | 2.19 | 6.24 | 0.48 | 4.24 | 0.09 | 0.54 | 95.97 | 0.75 | 0.28 | 0.31 | 0.28 | 1.09 | 3.93 | ME |
| S11_vf_10907 | 0 | 0 | 0 | 1.1 | 0.21 | 11.92 | 0.1 | 0.28 | 0.28 | 0.21 | 0.95 | 0.87 | 0.84 | 98.05 | 2.93 | 0.58 | 0.64 | 1 | 0.96 | 1.95 | ME |
| S11_vf_89309 | 0 | 0 | 0 | 0.37 | 0.04 | 0.83 | 0.1 | 0.87 | 0.68 | 0.39 | 1.34 | 5.62 | 0.99 | 446.47 | 0.66 | 0.64 | 0.66 | 0.41 | 1.31 | 1.13 | ME |
| S11_vf_43997 | 0 | 0 | 0 | 0.73 | 0.03 | 0.56 | 0.18 | 0.36 | 0.26 | 0.37 | 0.79 | 0.14 | 1.14 | 89.06 | 0.79 | 0.6 | 0.51 | 0.44 | 0.72 | 1.11 | ME |
| S11_vf_70015 | 0 | 0 | 0 | 1.07 | 0.05 | 6.31 | 0.08 | 0.75 | 0.29 | 0.35 | 1.09 | 0.16 | 11.97 | 21.89 | 1.07 | 0.63 | 0.28 | 0.83 | 0.8 | 0.82 | Shared |
| S11_vf_84303 | 0 | 0 | 0 | 1.41 | 0.02 | 0.18 | 2.51 | 0.12 | 0.1 | 0.11 | 0.46 | 0.06 | 1.62 | 21.68 | 6.75 | 0.84 | 0.2 | 0.44 | 0.42 | 0.57 | ME |
| S11_vf_104033 | 0 | 0 | 0 | 0.53 | 0.01 | 0.3 | 0.13 | 0.17 | 0.13 | 0.08 | 0.43 | 0.1 | 0.38 | 37.56 | 0.3 | 0.16 | 0.19 | 0.21 | 0.23 | 0.45 | ME |
| S11_vf_104034 | 0 | 0 | 0 | 1.25 | 0.03 | 0.23 | 0.05 | 0.28 | 0.24 | 0.15 | 16.87 | 0.08 | 0.83 | 15.06 | 0.62 | 0.43 | 0.23 | 0.22 | 0.29 | 0.47 | Shared |
| S11_vf_104035 | 0.07 | 0 | 0 | 25.78 | 449.65 | 25.08 | 0.08 | 24634.69 | 64096.44 | 14.48 | 23.84 | 0.11 | 31.94 | 90.58 | 60.25 | 12.29 | 4.5 | 0.38 | 0.36 | 0.61 | Shared |
| S11_vf_104044 | 0 | 0 | 0 | 1.49 | 0.02 | 0.36 | 1098.23 | 0.2 | 0.64 | 0.35 | 1.31 | 0.29 | 0.85 | 635.58 | 274.01 | 0.23 | 0.43 | 0.37 | 0.44 | 0.74 | Shared |
| S11_vf_104046 | 0 | 0 | 0 | 0.39 | 0.02 | 0.31 | 0.05 | 0.21 | 0.15 | 0.14 | 0.51 | 0.07 | 0.61 | 37.31 | 0.44 | 0.25 | 0.31 | 0.26 | 0.35 | 0.6 | ME |
| S11_vf_104048 | 0.01 | 0 | 0 | 0.35 | 0 | 0.24 | 0.05 | 0.19 | 32.01 | 0.09 | 0.26 | 11.8 | 8.2 | 40124.46 | 7.67 | 0.19 | 5.53 | 3.04 | 4.64 | 11.07 | Shared |
| S11_vf_104052 | 0 | 0 | 0 | 1.13 | 0.13 | 0.87 | 0.09 | 0.29 | 0.28 | 0.25 | 0.43 | 0.11 | 0.65 | 63.2 | 0.51 | 0.26 | 0.46 | 0.42 | 0.47 | 1.32 | ME |
| S11_vf_104059 | 0 | 0 | 0 | 0.75 | 0.08 | 0.74 | 0.2 | 0.36 | 3.16 | 4.89 | 100.86 | 1.54 | 2.25 | 3486.56 | 2311.55 | 0.37 | 1.48 | 1.07 | 1.34 | 2.85 | Shared |
| S11_vf_104062 | 0 | 0 | 0 | 23.39 | 0.51 | 0.42 | 0.13 | 0.1 | 0.13 | 198.94 | 0.31 | 0.07 | 0.71 | 25.7 | 0.4 | 0.18 | 0.17 | 0.21 | 0.21 | 0.42 | ME |
| S11_vf_104070 | 0.01 | 0 | 0.04 | 0.63 | 0.09 | 470.25 | 0.07 | 0.22 | 0.21 | 0.71 | 0.74 | 0.54 | 0.49 | 80.86 | 0.44 | 0.3 | 0.14 | 0.23 | 0.28 | 1.23 | ME |
| S11_vf_104071 | 0 | 0 | 0 | 7.08 | 0.02 | 6.56 | 0.02 | 0.14 | 2.18 | 0.06 | 0.44 | 0.79 | 0.8 | 2705.27 | 0.77 | 0.16 | 0.56 | 0.41 | 0.89 | 1.02 | ME |
| S11_vs_84627 | 0.01 | 0 | 0 | 2.15 | 0.03 | 0.5 | 0.15 | 2.7 | 4.41 | 0.39 | 0.89 | 0.11 | 0.64 | 87.4 | 0.79 | 0.77 | 0.55 | 0.36 | 0.61 | 1.02 | ME |
| S11_vs_40945 | 0 | 0 | 0 | 0.53 | 0.13 | 0.29 | 0.11 | 0.36 | 0.34 | 0.56 | 0.63 | 0.09 | 0.56 | 92.93 | 0.63 | 0.6 | 0.19 | 0.26 | 0.37 | 0.65 | ME |
| S11_vs_93101 | 0 | 0 | 0.01 | 0.37 | 0.03 | 0.72 | 0.23 | 0.29 | 0.26 | 0.17 | 0.98 | 0.16 | 2.02 | 114.51 | 0.75 | 0.43 | 0.36 | 0.37 | 0.64 | 1.19 | ME |
| S11_vs_3006 | 0 | 0 | 0 | 0.69 | 0.02 | 0.31 | 0.07 | 0.24 | 0.14 | 0.15 | 0.48 | 0.1 | 0.48 | 24.55 | 0.23 | 0.2 | 0.3 | 0.21 | 0.29 | 0.72 | ME |
| S11_vs_75554 | 0 | 0 | 0 | 0.86 | 0 | 0.41 | 0.61 | 0.12 | 0.99 | 0.18 | 0.79 | 0.22 | 0.69 | 923.6 | 0.55 | 0.12 | 0.28 | 0.37 | 0.4 | 0.66 | ME |
| S11_vs_49071 | 0 | 0 | 0 | 0.79 | 0.04 | 0.65 | 0.15 | 1.08 | 0.22 | 0.28 | 1 | 0.17 | 0.79 | 51.86 | 0.59 | 0.69 | 0.63 | 0.38 | 0.59 | 2.67 | ME |
| S11_vs_36748 | 0.01 | 0 | 0 | 0.75 | 0.04 | 2.91 | 0.37 | 0.68 | 0.96 | 0.44 | 0.92 | 0.1 | 0.49 | 48.96 | 1.06 | 0.99 | 0.49 | 1.06 | 4.72 | 8.17 | ME |
| S11_vs_82368 | 0 | 0 | 0 | 0.65 | 0.01 | 0.52 | 0.43 | 0.54 | 0.52 | 0.51 | 0.64 | 0.47 | 0.92 | 350.43 | 0.7 | 0.35 | 1.26 | 0.62 | 0.64 | 1.6 | ME |
| S11_vs_104032 | 0 | 0 | 0 | 0.61 | 0.02 | 0.53 | 0.05 | 0.26 | 1.47 | 0.18 | 0.86 | 1.14 | 1.2 | 1546.22 | 0.93 | 0.72 | 0.6 | 0.36 | 0.65 | 1.53 | ME |
| S11_vs_104036 | 0 | 0 | 0 | 0.59 | 0.02 | 0.59 | 0.12 | 0.59 | 1.52 | 0.11 | 1.92 | 0.28 | 0.87 | 685.55 | 0.87 | 0.36 | 0.37 | 0.39 | 0.56 | 0.98 | ME |
| S11_vs_104037 | 0.01 | 0 | 0 | 0.92 | 0.02 | 3.06 | 0.7 | 0.25 | 0.31 | 0.12 | 0.41 | 0.11 | 1.55 | 203.09 | 0.44 | 0.31 | 0.26 | 0.32 | 0.33 | 0.54 | ME |
| S11_vs_104038 | 0 | 0 | 0 | 0.87 | 0.02 | 0.86 | 0.28 | 0.54 | 0.91 | 0.6 | 0.85 | 0.38 | 1.32 | 679.32 | 1.47 | 0.58 | 0.61 | 0.53 | 0.83 | 1.49 | ME |
| S11_vs_104041 | 0 | 0 | 0 | 2.12 | 0.05 | 0.63 | 0.13 | 0.37 | 0.19 | 0.22 | 0.59 | 0.16 | 2.37 | 80.68 | 0.68 | 1.6 | 0.3 | 0.4 | 0.5 | 0.93 | ME |
| S11_vs_104042 | 0 | 0 | 0 | 1.18 | 0.01 | 0.88 | 0.49 | 0.31 | 0.26 | 0.29 | 1.07 | 0.27 | 0.66 | 79.12 | 0.38 | 0.42 | 0.44 | 0.64 | 2.84 | 1.01 | ME |
| S11_vs_104047 | 0 | 0 | 0 | 0.7 | 0.06 | 0.26 | 0.06 | 0.4 | 0.15 | 1.56 | 0.62 | 0.16 | 0.22 | 42.71 | 0.29 | 1.29 | 0.37 | 0.13 | 0.26 | 0.45 | ME |
| S11_vs_104049 | 0 | 0 | 0.01 | 0.82 | 0.21 | 0.71 | 0.11 | 1.13 | 0.2 | 1.6 | 0.81 | 0.33 | 0.72 | 32.32 | 0.5 | 0.66 | 1.29 | 0.22 | 0.42 | 0.97 | ME |
| S11_vs_104053 | 0 | 0 | 0 | 0.39 | 0.03 | 1.54 | 0.12 | 0.22 | 0.23 | 68.04 | 16.97 | 0.1 | 2.4 | 41.8 | 10.42 | 0.32 | 0.85 | 0.39 | 0.89 | 0.92 | Shared |
| S11_vs_104054 | 0 | 0 | 0 | 0.41 | 0.05 | 0.41 | 0.05 | 0.28 | 0.16 | 0.16 | 0.67 | 0.05 | 0.53 | 21.77 | 1.48 | 0.25 | 0.34 | 0.94 | 0.3 | 0.72 | ME |
| S11_vs_104064 | 0 | 0 | 0 | 1.29 | 0.03 | 0.4 | 0.14 | 0.35 | 0.19 | 5.24 | 4.94 | 1.1 | 16.35 | 70.19 | 2362 | 0.68 | 1.09 | 0.44 | 1.03 | 6.27 | Shared |

| | | | | | | | | | | | | | | | | | | | | | |
|---|---|---|---|---|---|---|---|---|---|---|---|---|---|---|---|---|---|---|---|---|---|
| S11_vs_104069 | 0.01 | 0 | 0 | 0.4 | 0.02 | 4.47 | 0.28 | 0.17 | 1.63 | 0.26 | 0.35 | 0.52 | 0.59 | 1710.2 | 0.51 | 0.16 | 0.43 | 0.33 | 0.46 | 0.75 | ME |
| S11_vs_58702 | 0 | 0 | 0 | 0.4 | 0.04 | 0.44 | 0.06 | 0.51 | 0.15 | 1.1 | 0.77 | 0.12 | 0.43 | 13.17 | 0.62 | 0.25 | 0.42 | 0.51 | 0.75 | 1.43 | ME |
| S11_vs_45658 | 0.01 | 0 | 0 | 7.27 | 0.69 | 5.09 | 1.08 | 5.35 | 5.11 | 2.32 | 4.66 | 0.76 | 11.71 | 113.75 | 15.65 | 3.42 | 6.47 | 6 | 8.09 | 19.49 | Shared |
| S11_vs_84848 | 0 | 0 | 0 | 5.94 | 0.1 | 1.66 | 1.08 | 0.89 | 1.04 | 0.71 | 1.01 | 0.31 | 3.04 | 51.26 | 0.51 | 10.42 | 1.05 | 3.26 | 1.89 | 36.75 | ME |
| S11_vs_19892 | 0.01 | 0 | 0 | 1.13 | 9.39 | 0.23 | 3.98 | 0.65 | 0.38 | 0.92 | 0.78 | 2.81 | 0.32 | 20.79 | 1.72 | 0.4 | 0.24 | 0.32 | 0.39 | 0.25 | ME |
| S11_vs_6878 | 0 | 0 | 0 | 0.43 | 0.03 | 0.28 | 0.04 | 0.45 | 0.31 | 0.15 | 0.4 | 0.25 | 0.27 | 204.29 | 0.33 | 0.26 | 0.4 | 0.26 | 0.3 | 0.72 | ME |
| S11_vs_58930 | 0 | 0.01 | 0 | 134.63 | 0.07 | 6.19 | 3.05 | 0.3 | 0.4 | 1.98 | 0.54 | 0.28 | 0.67 | 119.09 | 18.16 | 17.66 | 0.32 | 111.26 | 11.05 | 233.63 | Shared |
| S11_vs_52577 | 0.02 | 0 | 0 | 1.06 | 0.06 | 0.49 | 0.1 | 0.65 | 0.21 | 0.42 | 0.91 | 0.1 | 0.39 | 45.87 | 0.9 | 0.53 | 0.42 | 0.67 | 1.69 | 1.69 | ME |
| S11_vs_98193 | 0.01 | 0 | 0 | 0.63 | 0.13 | 0.46 | 0.03 | 12.07 | 2.4 | 0.23 | 2.39 | 0.23 | 0.74 | 19.31 | 4.82 | 0.38 | 0.98 | 0.57 | 1.05 | 1.18 | ME |
| S11_vs_20225 | 0.01 | 0 | 0 | 0.79 | 0.07 | 0.56 | 0.13 | 1.32 | 0.28 | 3.94 | 2.37 | 0.18 | 0.4 | 206.5 | 2.17 | 2.58 | 0.56 | 0.21 | 3.19 | 0.53 | ME |
| S11_vs_26767 | 0 | 0 | 0 | 0.71 | 0.04 | 0.37 | 0.18 | 0.28 | 0.17 | 0.2 | 0.47 | 0.36 | 0.56 | 17.51 | 0.22 | 0.19 | 0.52 | 0.39 | 0.41 | 0.62 | ME |
| S11_vs_79275 | 0.01 | 0 | 0.01 | 0.82 | 0.05 | 0.34 | 0.04 | 0.39 | 0.14 | 0.16 | 0.54 | 0.13 | 0.26 | 31.1 | 0.6 | 0.49 | 0.21 | 0.2 | 0.34 | 0.47 | ME |
| S11_vs_13652 | 0 | 0 | 0 | 0.84 | 0.03 | 2.14 | 0.17 | 0.36 | 0.64 | 0.57 | 0.6 | 1.47 | 0.52 | 261.61 | 0.44 | 0.34 | 0.59 | 0.42 | 0.66 | 0.99 | ME |
| S11_vs_53675 | 0 | 0 | 0 | 0.91 | 0.01 | 0.47 | 0.09 | 1.27 | 0.2 | 0.15 | 0.65 | 0.29 | 0.62 | 19.99 | 0.98 | 0.17 | 0.54 | 0.4 | 0.41 | 1 | ME |
| S11_vs_79831 | 0.01 | 0 | 0 | 0.48 | 0.03 | 0.56 | 0.08 | 0.39 | 0.36 | 0.14 | 0.93 | 0.09 | 0.84 | 79.36 | 6.13 | 0.38 | 0.44 | 0.32 | 0.81 | 1.28 | ME |
| S11_vs_1937 | 0.01 | 0 | 0 | 1.48 | 0.03 | 27.31 | 0.07 | 0.16 | 0.1 | 0.08 | 23.52 | 0.06 | 24.24 | 13.59 | 0.25 | 0.77 | 0.16 | 0.14 | 4.31 | 9.11 | Shared |
| S11_vs_8456 | 0 | 0 | 0 | 0.63 | 0.01 | 0.52 | 0.06 | 0.36 | 0.22 | 0.25 | 0.68 | 0.1 | 0.75 | 38.55 | 0.76 | 0.51 | 0.65 | 0.28 | 0.45 | 1.12 | ME |
| S11_vs_34137 | 0.01 | 0 | 0 | 0.68 | 0.03 | 0.67 | 0.09 | 0.34 | 0.12 | 1.2 | 0.53 | 0.38 | 0.54 | 22.22 | 0.41 | 0.47 | 0.48 | 0.22 | 1.15 | 0.76 | ME |
| S11_vs_14882 | 0 | 0 | 0 | 0.57 | 0.02 | 0.35 | 0.05 | 0.15 | 0.1 | 0.09 | 0.39 | 0.1 | 0.58 | 14.03 | 0.4 | 0.24 | 0.59 | 0.39 | 0.34 | 0.48 | ME |
| S11_vs_41107 | 0 | 0 | 0 | 3.79 | 0.08 | 1.74 | 1.98 | 8.33 | 0.09 | 0.54 | 1.9 | 0.11 | 1.04 | 16.15 | 1.51 | 1.18 | 2.46 | 1.4 | 1.8 | 2.15 | ME |
| S11_vs_67573 | 0 | 0 | 0 | 0.94 | 0.02 | 2.18 | 0.72 | 1.09 | 1.96 | 1.25 | 1.08 | 2.03 | 0.8 | 49.97 | 0.48 | 0.61 | 0.52 | 0.48 | 0.9 | 1.68 | ME |
| S11_vs_48109 | 0 | 0 | 0 | 0.77 | 0.02 | 0.78 | 0.14 | 0.3 | 0.95 | 0.41 | 1.1 | 0.42 | 0.86 | 899.64 | 0.89 | 0.67 | 0.5 | 0.37 | 0.69 | 1.18 | ME |
| S11_vs_3277 | 0 | 0 | 0.01 | 16.69 | 1.18 | 24.07 | 0.17 | 0.95 | 0.39 | 3.14 | 76.54 | 0.21 | 2.44 | 77.96 | 1.15 | 8.02 | 6.05 | 1.77 | 6.04 | 7.16 | Shared |
| S11_vs_55144 | 0 | 0 | 0 | 0.45 | 0.02 | 0.37 | 0.05 | 0.29 | 0.25 | 1.53 | 0.72 | 0.11 | 1.04 | 50.56 | 0.54 | 0.37 | 0.32 | 0.35 | 0.47 | 1.06 | ME |
| S11_vs_23188 | 0 | 0 | 0 | 0.42 | 0.1 | 1.07 | 0.21 | 0.24 | 1.17 | 0.23 | 0.74 | 0.39 | 0.89 | 1066.6 | 0.51 | 0.24 | 10.22 | 0.34 | 0.71 | 1.33 | ME |
| S11_vs_23357 | 0.01 | 0 | 0.01 | 1.04 | 0.26 | 0.16 | 0.16 | 3.65 | 4.81 | 4.92 | 0.34 | 3.39 | 0.5 | 50.16 | 0.21 | 0.46 | 0.56 | 0.3 | 0.81 | 0.64 | ME |
| S11_vs_10172 | 0 | 0 | 0.01 | 0.82 | 23.54 | 0.19 | 10.21 | 3.85 | 0.34 | 0.7 | 0.55 | 0.18 | 0.55 | 21.96 | 1.1 | 0.46 | 0.31 | 0.84 | 0.6 | 0.44 | Shared |
| S11_vs_68151 | 0 | 0 | 0 | 1.22 | 0.17 | 4.71 | 0.05 | 0.4 | 0.67 | 0.31 | 4.4 | 213.71 | 49.89 | 74.29 | 2.16 | 9.17 | 13.49 | 3.29 | 450.57 | 1.36 | Shared |
| S11_vs_81775 | 0 | 0 | 0 | 0.48 | 0.02 | 0.43 | 0.08 | 0.24 | 0.14 | 3.38 | 0.42 | 0.1 | 0.56 | 22.65 | 0.61 | 0.27 | 0.27 | 0.28 | 0.33 | 0.59 | ME |
| S11_vs_68348 | 0 | 0 | 0 | 0.79 | 0.03 | 0.6 | 0.26 | 0.43 | 0.24 | 0.28 | 0.59 | 2.18 | 1.08 | 21.88 | 0.54 | 0.46 | 1.12 | 0.43 | 0.61 | 1.22 | ME |
| S11_vs_42927 | 0 | 0 | 0 | 1.53 | 0.05 | 1.53 | 0.29 | 8.5 | 0.53 | 2.93 | 2.37 | 5.81 | 1 | 24.23 | 1.45 | 2.01 | 17.76 | 0.48 | 2.74 | 1.86 | ME |
| S11_vs_62573 | 0 | 0 | 0 | 0.79 | 0.02 | 0.34 | 0.05 | 0.31 | 0.13 | 0.62 | 0.6 | 0.08 | 0.25 | 45.11 | 0.43 | 8.12 | 0.43 | 0.24 | 1.73 | 0.47 | ME |
| S11_vs_56762 | 0 | 0 | 0 | 0.78 | 0.02 | 0.6 | 0.02 | 2.92 | 0.15 | 0.16 | 0.48 | 0.23 | 0.51 | 16.96 | 0.48 | 0.21 | 1.62 | 0.31 | 0.36 | 0.78 | ME |
| S11_vs_82331 | 0 | 0 | 0 | 2.02 | 0.03 | 0.45 | 0.09 | 1.03 | 1.45 | 1.97 | 1.9 | 0.12 | 0.57 | 43.74 | 3.52 | 1.09 | 0.4 | 0.65 | 1.05 | 32.63 | ME |
| S11_vs_11267 | 0 | 0 | 0 | 6.23 | 13.14 | 0.21 | 0.06 | 3.88 | 7.12 | 2.41 | 1.31 | 0.69 | 0.68 | 111.39 | 148.85 | 0.71 | 0.2 | 0.24 | 0.44 | 0.65 | Shared |
| S11_vs_76784 | 0.01 | 0 | 0 | 1.77 | 0 | 0.23 | 6.28 | 0.38 | 0.3 | 7.44 | 1.31 | 0.17 | 0.27 | 35.74 | 15.9 | 0.38 | 0.44 | 2.56 | 0.4 | 3.09 | Shared |
| S11_vs_5077 | 0.01 | 0 | 0.01 | 0.36 | 0.01 | 0.11 | 0.1 | 0.17 | 0.19 | 0.08 | 0.3 | 0.06 | 0.13 | 86.16 | 0.28 | 0.14 | 0.17 | 0.1 | 0.17 | 0.27 | ME |
| S11_vs_30739 | 0 | 0 | 0 | 1.1 | 0.04 | 6.28 | 1.19 | 2.45 | 0.42 | 1.01 | 1.21 | 0.07 | 0.3 | 15.61 | 1.35 | 0.39 | 0.52 | 0.16 | 0.42 | 0.69 | ME |
| S11_vs_37666 | 0.01 | 0 | 0 | 0.51 | 0.05 | 0.57 | 0.03 | 0.33 | 0.22 | 0.13 | 0.48 | 0.12 | 0.85 | 115.53 | 0.46 | 0.21 | 0.99 | 0.24 | 0.24 | 1.41 | ME |
| S11_vs_89911 | 0 | 0 | 0.02 | 0.78 | 0.03 | 0.33 | 0.12 | 0.96 | 0.14 | 0.72 | 0.32 | 0.05 | 0.24 | 14.6 | 0.2 | 0.18 | 0.22 | 0.17 | 0.31 | 0.68 | ME |
| S11_vs_11994 | 0.01 | 0 | 0 | 0.91 | 0.02 | 0.3 | 0.04 | 3.63 | 0.11 | 0.19 | 4.17 | 0.42 | 0.2 | 29.71 | 0.42 | 0.37 | 0.71 | 0.2 | 0.33 | 2.01 | ME |
| S11_vs_83666 | 0 | 0 | 0 | 1.2 | 0.02 | 0.72 | 0.16 | 0.61 | 0.21 | 0.49 | 0.5 | 0.32 | 1.1 | 12.56 | 0.43 | 0.24 | 0.37 | 0.29 | 0.64 | 1.02 | ME |
| S11_vs_38860 | 0 | 0 | 0 | 0.56 | 0.04 | 0.28 | 0.08 | 22.36 | 0.4 | 1.05 | 0.57 | 0.66 | 1.08 | 14.9 | 0.18 | 0.27 | 8.39 | 0.22 | 0.24 | 0.96 | ME |
| S11_vs_6458 | 0 | 0 | 0 | 10.34 | 0.02 | 0.45 | 0.19 | 20.18 | 33.65 | 0.18 | 1.01 | 0.12 | 0.55 | 21.64 | 0.59 | 6.75 | 0.3 | 0.38 | 0.34 | 0.8 | Shared |
| S13_vf_1770 | 0 | 0.05 | 0 | 120.75 | 0 | 0.19 | 0.02 | 0.09 | 0.1 | 0.1 | 0.84 | 0.06 | 0.18 | 0.22 | 0.31 | 60.47 | 0.03 | 0.25 | 0.16 | 0.19 | ME |
| S13_vf_3728 | 0.03 | 0.01 | 0 | 50.14 | 41.5 | 2.21 | 0.19 | 1.2 | 0.98 | 1.47 | 7827.32 | 0.06 | 1.09 | 0.31 | 0.81 | 189.79 | 0.35 | 0.76 | 0.75 | 10.84 | Shared |
| S13_vf_4901 | 0 | 0 | 0 | 7.15 | 0.03 | 0.4 | 0.15 | 0.41 | 0.52 | 0.22 | 2.74 | 0.72 | 0.95 | 290.64 | 3.28 | 227.33 | 0.46 | 0.32 | 9.49 | 0.82 | ME |
| S13_vf_7670 | 0.02 | 0 | 0 | 86.37 | 0.08 | 0.26 | 0.03 | 0.16 | 0.1 | 0.1 | 0.28 | 0.04 | 0.28 | 0.1 | 0.18 | 43.63 | 0.12 | 0.42 | 0.26 | 0.42 | ME |
| S13_vf_11246 | 6.5 | 0.12 | 0.01 | 830.44 | 63863.92 | 11.12 | 0.08 | 233.67 | 552.7 | 7.78 | 14 | 0.17 | 1.25 | 1.29 | 21.23 | 338.41 | 65.71 | 1.67 | 0.74 | 0.58 | Shared |
| S13_vf_22175 | 0 | 0 | 0 | 0.64 | 0.02 | 0.25 | 0.06 | 0.2 | 0.09 | 0.15 | 0.44 | 0.06 | 0.34 | 0.33 | 0.32 | 152.91 | 0.2 | 0.17 | 0.56 | 0.45 | ME |
| S13_vf_28309 | 0 | 0 | 0 | 48.94 | 0.07 | 0.76 | 0.49 | 0.38 | 0.28 | 0.28 | 0.95 | 0.09 | 1.12 | 0.35 | 1.4 | 157.09 | 0.47 | 0.44 | 0.56 | 1.15 | ME |
| S13_vf_38708 | 0 | 0 | 0 | 1.11 | 0.04 | 0.23 | 0.09 | 0.33 | 0.09 | 0.22 | 0.39 | 0.03 | 0.25 | 0.35 | 0.23 | 61.18 | 0.32 | 0.17 | 1.84 | 0.42 | ME |
| S13_vf_54201 | 0.01 | 0 | 0 | 0.72 | 0.02 | 0.17 | 0.14 | 0.33 | 0.1 | 0.12 | 0.54 | 0.07 | 0.23 | 0.42 | 0.25 | 56.72 | 0.32 | 0.21 | 1.16 | 0.44 | ME |
| S13_vf_56074 | 0 | 0 | 0.01 | 2.7 | 0.12 | 4.49 | 0.08 | 0.26 | 0.18 | 0.21 | 0.65 | 0.06 | 8.2 | 0.18 | 0.33 | 1129.59 | 0.22 | 1.63 | 3.41 | 3.97 | ME |
| S13_vf_66508 | 0 | 0.01 | 0 | 87.55 | 2.56 | 28.96 | 0.15 | 0.55 | 0.22 | 1 | 1.77 | 1.36 | 2.84 | 1.19 | 0.75 | 35.77 | 0.56 | 0.64 | 1.84 | 1.21 | ME |
| S13_vf_87873 | 0.04 | 0.01 | 0 | 23.71 | 36.16 | 2.85 | 5.17 | 29.52 | 51.16 | 141.83 | 1.17 | 0.07 | 1.81 | 16.31 | 0.35 | 366.63 | 0.35 | 0.18 | 25.72 | 0.42 | Shared |
| S13_vf_91189 | 0 | 0 | 0 | 0.29 | 0.07 | 0.35 | 0.01 | 0.12 | 0.3 | 0.11 | 0.7 | 0.08 | 0.85 | 0.38 | 0.56 | 582.7 | 0.43 | 0.24 | 1.38 | 0.55 | ME |
| S13_vf_97702 | 0.02 | 0 | 0 | 116.3 | 0.02 | 0.45 | 0.01 | 0.17 | 0.16 | 13.78 | 0.71 | 0.18 | 0.44 | 0.32 | 0.49 | 72.43 | 0.23 | 0.64 | 75.06 | 170.13 | Shared |
| S13_vf_109235 | 0.03 | 0.08 | 0 | 760.94 | 0.17 | 0.47 | 0.09 | 0.35 | 0.28 | 0.54 | 0.7 | 0.09 | 0.26 | 2.58 | 0.72 | 342.22 | 1.62 | 2.38 | 0.73 | 0.56 | ME |
| S13_vf_131208 | 0 | 0 | 0 | 14.07 | 0.04 | 0.59 | 0.45 | 0.21 | 0.09 | 0.13 | 0.53 | 0.13 | 0.55 | 0.25 | 1.34 | 24.47 | 0.48 | 0.39 | 2.24 | 5.58 | ME |
| S13_vf_143422 | 0 | 0.01 | 0 | 2.47 | 0.14 | 0.61 | 244.14 | 0.38 | 0.19 | 0.92 | 930.39 | 0.06 | 0.55 | 0.53 | 0.65 | 35.62 | 0.29 | 0.31 | 0.49 | 2.4 | Shared |
| S13_vf_147396 | 0.03 | 0.03 | 0 | 265.85 | 0.08 | 0.68 | 0.19 | 0.4 | 0.32 | 0.24 | 0.95 | 0.15 | 0.61 | 0.33 | 0.6 | 53.81 | 0.43 | 0.78 | 0.53 | 0.93 | ME |
| S13_vf_147589 | 0 | 0 | 0 | 0.68 | 0.01 | 0.19 | 0.04 | 0.89 | 0.11 | 0.08 | 0.39 | 0.02 | 0.5 | 0.06 | 0.93 | 49.25 | 0.74 | 0.21 | 0.23 | 0.43 | ME |
| S13_vf_150407 | 0 | 0 | 0 | 30.88 | 0.02 | 0.75 | 0.29 | 0.12 | 0.14 | 0.24 | 0.34 | 0.08 | 0.18 | 0.08 | 0.36 | 40.31 | 0.08 | 0.28 | 0.32 | 0.2 | ME |
| S13_vf_151045 | 0.01 | 0.06 | 0 | 577.5 | 66.99 | 0.85 | 0.08 | 0.44 | 0.31 | 0.32 | 0.71 | 0.08 | 2.04 | 0.28 | 0.62 | 267.24 | 1.42 | 1.37 | 1.13 | 1.72 | Shared |
| S13_vf_151755 | 0 | 0 | 0 | 0.72 | 0.12 | 0.4 | 0.13 | 0.22 | 0.19 | 0.2 | 791.16 | 0.05 | 0.4 | 0.12 | 0.47 | 59.04 | 0.1 | 0.16 | 0.23 | 1.61 | Shared |
| S13_vs_12957 | 0.02 | 0 | 0 | 260.16 | 1.6 | 0.31 | 0.07 | 0.19 | 0.09 | 0.1 | 0.43 | 0.1 | 0.09 | 0.22 | 0.18 | 27.19 | 0.03 | 0.28 | 0.16 | 0.33 | ME |
| S13_vs_15645 | 0 | 0.01 | 0 | 36.07 | 0.5 | 9.56 | 16.68 | 10.99 | 1.71 | 23.27 | 9 | 0.11 | 3.36 | 198.94 | 0.29 | 4022.16 | 46.66 | 0.26 | 5.58 | 0.52 | Shared |
| S13_vs_27462 | 0 | 0.01 | 0 | 0.74 | 0.07 | 0.76 | 0.19 | 15.68 | 0.2 | 0.23 | 1 | 10.19 | 18941.24 | 3.11 | 3.25 | 230.12 | 2.9 | 335.83 | 5.26 | 5.25 | Shared |
| S13_vs_148294 | 0.01 | 0.01 | 0 | 153.07 | 4.85 | 1.35 | 0.08 | 1.32 | 2.12 | 1.03 | 2.63 | 4.94 | 0.35 | 2.99 | 0.34 | 44.11 | 1.53 | 0.41 | 1.53 | 0.95 | ME |
| S13_vs_149860 | 0 | 0.02 | 0 | 83.95 | 0.04 | 0.32 | 0.03 | 0.23 | 0.1 | 0.11 | 0.55 | 0.03 | 0.43 | 0.09 | 0.23 | 61.46 | 0.18 | 0.39 | 0.22 | 0.9 | ME |
| S13_vs_151510 | 0.01 | 0 | 0.01 | 0.52 | 0.07 | 0.76 | 1.55 | 3.63 | 0.17 | 0.25 | 1.76 | 2.63 | 4266.81 | 0.8 | 1.23 | 64.17 | 1.03 | 0.68 | 1.33 | 1.58 | Shared |
| S13_vs_9438 | 0.04 | 0.03 | 0 | 120.15 | 0.07 | 0.59 | 0.1 | 194.41 | 296.71 | 0.33 | 0.91 | 0.1 | 1.01 | 3.03 | 0.38 | 68.58 | 0.29 | 0.46 | 0.4 | 0.75 | Shared |
| S13_vs_13129 | 0.01 | 0.02 | 0.01 | 494.99 | 0.09 | 0.36 | 0.09 | 0.57 | 0.13 | 0.29 | 0.95 | 0.21 | 0.34 | 0.16 | 0.48 | 98.91 | 0.32 | 0.79 | 0.38 | 0.47 | ME |
| S13_vs_14794 | 0.02 | 0.09 | 0 | 734.08 | 0.14 | 0.44 | 0.07 | 0.23 | 0.19 | 0.18 | 0.52 | 0.06 | 0.31 | 0.08 | 0.18 | 74.81 | 0.18 | 1.18 | 0.22 | 0.33 | ME |
| S13_vs_23314 | 0 | 0 | 0 | 1.64 | 0.1 | 1.29 | 0.07 | 5.33 | 0.37 | 4.27 | 3.36 | 2.07 | 1.94 | 2.35 | 1.51 | 230.41 | 11.21 | 0.43 | 5.13 | 1.34 | ME |
| S13_vs_26756 | 0.07 | 0 | 0 | 14.2 | 0.08 | 1.49 | 0.16 | 0.57 | 0.26 | 0.36 | 1.78 | 0.19 | 2.1 | 0.59 | 24.12 | 113.68 | 0.82 | 0.74 | 1.28 | 1.69 | Shared |
| S13_vs_30101 | 0 | 0.01 | 0 | 10.56 | 0.08 | 1.52 | 0.19 | 0.56 | 0.28 | 0.45 | 57.74 | 0.64 | 4.04 | 1.86 | 8.93 | 78.16 | 1.36 | 0.55 | 25.93 | 1.58 | Shared |
| S13_vs_30167 | 0 | 0 | 0 | 13.36 | 0.57 | 4.54 | 0.27 | 27.14 | 1.5 | 15.54 | 8.83 | 3.3 | 6.09 | 9.61 | 5.05 | 181.07 | 34.54 | 1.67 | 15.97 | 4.98 | Shared |
| S13_vs_33947 | 0.02 | 0.01 | 0.01 | 1.71 | 0.05 | 1.18 | 1.05 | 1.67 | 0.28 | 1.18 | 5.37 | 0.34 | 1.09 | 48.95 | 2.13 | 100.45 | 1.25 | 0.72 | 2.02 | 1.04 | ME |
| S13_vs_35816 | 0.02 | 0.08 | 0.01 | 778.44 | 0.26 | 41.56 | 194.79 | 19.15 | 2.6 | 23.61 | 90.92 | 54.11 | 4.82 | 2.02 | 4.31 | 280.1 | 65.66 | 2.67 | 21.54 | 10.15 | Shared |
| S13_vs_45488 | 0 | 0 | 0 | 60.81 | 0.1 | 1.44 | 0.16 | 27.53 | 50.38 | 0.32 | 2.69 | 0.23 | 1.12 | 13.14 | 1.31 | 60.82 | 0.46 | 0.65 | 0.67 | 1.65 | Shared |
| S13_vs_49889 | 0 | 0.03 | 0 | 73.87 | 0.04 | 0.26 | 0.04 | 0.23 | 0.14 | 0.13 | 1.77 | 0.06 | 0.43 | 0.24 | 0.5 | 32.82 | 0.29 | 0.32 | 0.73 | 1.58 | ME |
| S13_vs_57267 | 0.22 | 0.02 | 0 | 72.1 | 0.15 | 1.04 | 0.22 | 0.42 | 0.25 | 0.25 | 1.26 | 0.58 | 1.46 | 0.6 | 0.88 | 239.75 | 0.59 | 0.62 | 56.71 | 170.58 | Shared |

| | | | | | | | | | | | | | | | | | | | | | |
|---|---|---|---|---|---|---|---|---|---|---|---|---|---|---|---|---|---|---|---|---|---|
| S13_vs_67429 | 0 | 0 | 0 | 12.95 | 0.31 | 1.89 | 0.17 | 0.6 | 0.36 | 0.36 | 17.34 | 0.2 | 31.67 | 0.74 | 52.51 | 113.38 | 0.81 | 0.75 | 5.57 | 1.81 | Shared |
| S13_vs_68102 | 0 | 0.09 | 0 | 473.7 | 0.06 | 0.54 | 0.07 | 0.42 | 0.21 | 0.24 | 0.76 | 0.08 | 0.61 | 0.23 | 0.56 | 192.67 | 0.39 | 0.95 | 0.49 | 0.9 | ME |
| S13_vs_83447 | 0.04 | 0.07 | 0.02 | 345.72 | 67.67 | 2.6 | 0.52 | 6.81 | 0.61 | 3.68 | 79.74 | 875.16 | 4.7 | 1.89 | 0.62 | 171.77 | 1.58 | 22.5 | 3.92 | 1.41 | Shared |
| S13_vs_87108 | 0 | 0 | 0 | 7.77 | 1.17 | 17.11 | 0.63 | 0.19 | 0.1 | 0.07 | 0.27 | 0.05 | 0.15 | 0.06 | 0.31 | 28.85 | 0.11 | 0.29 | 0.23 | 0.56 | ME |
| S13_vs_90967 | 0 | 0 | 0 | 56.85 | 0.07 | 0.11 | 0 | 0.16 | 0.09 | 0.01 | 0.19 | 0.09 | 0.27 | 0.01 | 0.13 | 53.07 | 0.02 | 0.23 | 2.18 | 10.5 | Shared |
| S13_vs_98591 | 0.01 | 0.26 | 0 | 2726.02 | 0.65 | 1.66 | 0.08 | 536.63 | 794.31 | 0.97 | 1.79 | 0.07 | 1.19 | 1.7 | 0.44 | 631.9 | 0.18 | 4.66 | 1.1 | 0.65 | Shared |
| S13_vs_113593 | 0 | 0.11 | 0 | 1062.8 | 0.2 | 0.68 | 0.02 | 0.3 | 0.25 | 0.33 | 0.67 | 0.05 | 0.36 | 0.49 | 0.31 | 333.41 | 0.28 | 1.75 | 0.68 | 0.59 | ME |
| S13_vs_114213 | 0.01 | 0 | 0.01 | 1.56 | 0.07 | 0.69 | 0.3 | 3.74 | 0.69 | 1.71 | 3.05 | 1.26 | 0.74 | 19.9 | 1.18 | 59.21 | 2.23 | 0.85 | 1.55 | 1.82 | ME |
| S13_vs_121307 | 0 | 0 | 0 | 47.08 | 0.02 | 0.25 | 0.09 | 0.65 | 0.14 | 0.14 | 0.61 | 0.15 | 0.31 | 0.12 | 0.39 | 25.92 | 0.37 | 0.38 | 0.25 | 0.65 | ME |
| S13_vs_152223 | 0.06 | 0.35 | 0 | 3438.14 | 758.35 | 1.42 | 0.05 | 0.9 | 0.64 | 0.82 | 1.56 | 0.08 | 0.44 | 0.2 | 0.32 | 522.1 | 0.87 | 4.82 | 1.06 | 0.51 | Shared |
| S14_vf_58919 | 0 | 0 | 0 | 0.86 | 0.13 | 0.51 | 0.17 | 1.24 | 0.18 | 0.27 | 0.86 | 0.61 | 1.35 | 0.34 | 0.68 | 0.4 | 34.05 | 0.37 | 0.63 | 13.41 | ME |
| S14_vf_44452 | 0 | 0 | 0 | 0.35 | 0.09 | 0.1 | 0.04 | 0.11 | 0.07 | 1.31 | 10.4 | 0.06 | 0.16 | 0.04 | 0.08 | 0.33 | 62.68 | 2.06 | 0.25 | 1.92 | Shared |
| S14_vf_37500 | 0 | 0 | 0 | 2.87 | 0.35 | 1.08 | 0.1 | 0.28 | 0.93 | 0.07 | 0.18 | 0.19 | 0.52 | 0.35 | 0.38 | 0.85 | 533.38 | 0.19 | 0.48 | 0.36 | ME |
| S14_vf_106156 | 0 | 0 | 0 | 0.84 | 0.03 | 0.37 | 0.03 | 0.24 | 0.09 | 0.12 | 0.43 | 0.08 | 1.18 | 0.34 | 0.43 | 0.41 | 39.06 | 0.28 | 0.39 | 1.21 | ME |
| S14_vf_92240 | 0.01 | 0 | 0 | 2.79 | 0.21 | 62.91 | 31.15 | 103.93 | 11.11 | 16.69 | 8.59 | 0.24 | 2.11 | 2.94 | 8.76 | 1.23 | 156.96 | 1.59 | 15.4 | 10.45 | Shared |
| S14_vf_85122 | 0 | 0 | 0 | 0.52 | 0.03 | 1.38 | 0.3 | 6.85 | 0.18 | 0.22 | 0.79 | 0.22 | 0.81 | 0.21 | 0.88 | 0.6 | 21.95 | 0.32 | 0.66 | 0.82 | ME |
| S14_vf_99344 | 0.17 | 0 | 0 | 0.45 | 1.8 | 0.77 | 0.09 | 0.27 | 0.17 | 0.21 | 14.71 | 0.59 | 1.1 | 0.47 | 1.18 | 0.24 | 2505.39 | 0.72 | 0.56 | 1.32 | Shared |
| S14_vf_114491 | 2.39 | 0 | 0 | 4.4 | 27996.65 | 5.29 | 0.09 | 2.73 | 2.81 | 3.25 | 6.23 | 0.08 | 0.6 | 0.42 | 0.59 | 2.57 | 29.76 | 0.33 | 0.5 | 0.71 | Shared |
| S14_vf_78139 | 0 | 0 | 0 | 0.38 | 0.04 | 0.23 | 0.11 | 3.61 | 0.07 | 0.11 | 0.36 | 0.07 | 0.25 | 0.08 | 0.19 | 0.18 | 31.75 | 0.17 | 0.2 | 0.48 | ME |
| S14_vf_56130 | 0 | 0 | 0 | 3.12 | 0.12 | 1.08 | 4.47 | 14.69 | 0.11 | 0.81 | 1.69 | 0.07 | 0.58 | 3.06 | 1.82 | 1.58 | 24.08 | 4.81 | 8.13 | 1.15 | ME |
| S14_vf_93134 | 0.16 | 0 | 0 | 4.01 | 1.9 | 0.32 | 0.02 | 0.22 | 0.18 | 0.34 | 0.54 | 0.55 | 0.85 | 0.47 | 0.96 | 0.48 | 3146.77 | 0.56 | 1.29 | 23.02 | ME |
| S14_vf_48775 | 0 | 0 | 0 | 0.4 | 0.12 | 0.22 | 0.04 | 0.13 | 0.07 | 0.05 | 0.21 | 0.07 | 0.38 | 0.1 | 0.43 | 0.1 | 187.15 | 0.81 | 0.25 | 0.4 | ME |
| S14_vf_115736 | 0.02 | 0 | 0 | 7.45 | 0.4 | 1.42 | 0.18 | 1.13 | 0.42 | 1.68 | 1.61 | 0.42 | 1.72 | 1.31 | 0.37 | 2.54 | 182.26 | 0.92 | 1.37 | 0.8 | ME |
| S14_vf_43340 | 0 | 0 | 0 | 1.68 | 3.97 | 0.46 | 3.12 | 0.71 | 0.91 | 0.1 | 0.27 | 0.09 | 0.36 | 1.51 | 0.98 | 1.72 | 64.69 | 1.82 | 0.5 | 1.12 | ME |
| S14_vf_57689 | 0.04 | 0 | 0.01 | 0.57 | 0.03 | 0.17 | 0.15 | 0.24 | 0.13 | 0.12 | 0.92 | 0.1 | 0.18 | 0.09 | 0.25 | 0.33 | 24.65 | 0.15 | 0.25 | 0.45 | ME |
| S14_vf_109545 | 0.01 | 0 | 0 | 1.04 | 0.01 | 1.55 | 0.49 | 1.95 | 3.32 | 0.37 | 5.56 | 12.96 | 0.17 | 3.03 | 0.64 | 0.39 | 26.76 | 14.44 | 3.24 | 1.78 | Shared |
| S14_vf_72335 | 0 | 0 | 0 | 0.69 | 0.07 | 0.45 | 0.02 | 4.08 | 0.08 | 0.14 | 0.44 | 0.07 | 0.48 | 0.42 | 0.36 | 0.31 | 31.27 | 0.24 | 0.28 | 0.58 | ME |
| S14_vf_117147 | 0 | 0 | 0 | 0.87 | 0.08 | 0.34 | 0.1 | 0.22 | 0.11 | 0.2 | 1.59 | 0.2 | 0.23 | 0.39 | 0.32 | 0.46 | 196.08 | 0.4 | 0.46 | 3.2 | ME |
| S14_vf_117150 | 0 | 0.01 | 0 | 2.02 | 0.25 | 3.24 | 0.63 | 0.23 | 0.18 | 0.37 | 0.6 | 0.07 | 3.47 | 0.6 | 0.93 | 0.75 | 89.41 | 5.27 | 4.83 | 1.1 | ME |
| S14_vf_72953 | 0 | 0 | 0 | 0.61 | 0.05 | 0.54 | 0.05 | 1.36 | 0.13 | 0.11 | 0.43 | 0.08 | 0.81 | 0.13 | 0.39 | 0.35 | 33.23 | 0.31 | 0.31 | 0.81 | ME |
| S14_vf_117155 | 0.1 | 0.03 | 0 | 0.89 | 1.13 | 0.18 | 0.16 | 0.14 | 0.11 | 0.04 | 0.2 | 0.48 | 0.39 | 0.32 | 0.47 | 0.19 | 2051.33 | 501.52 | 0.5 | 0.95 | Shared |
| S14_vf_117157 | 0.03 | 0 | 0 | 0.57 | 0.27 | 0.31 | 0.03 | 0.13 | 0.12 | 0.16 | 0.41 | 0.21 | 0.74 | 58.81 | 81.6 | 0.21 | 394.45 | 7.22 | 4.81 | 0.6 | Shared |
| S14_vf_117162 | 0.17 | 0 | 0 | 0.72 | 1.45 | 0.28 | 0.06 | 0.27 | 0.08 | 0.09 | 0.23 | 0.57 | 0.51 | 0.53 | 0.6 | 0.12 | 2756.02 | 0.45 | 0.59 | 0.82 | ME |
| S14_vf_117164 | 0.01 | 0 | 0 | 0.64 | 0.15 | 0.42 | 0.06 | 0.78 | 0.17 | 0.24 | 27.52 | 0.13 | 0.37 | 0.17 | 0.57 | 1.12 | 304.75 | 0.34 | 0.34 | 0.52 | Shared |
| S14_vf_51134 | 0.04 | 0 | 0 | 0.84 | 0.74 | 1.97 | 0.07 | 0.18 | 0.05 | 1.48 | 2.66 | 0.27 | 0.64 | 0.42 | 0.43 | 0.23 | 1271.13 | 9.28 | 3.86 | 0.76 | ME |
| S14_vs_15228 | 0.28 | 0 | 0 | 22.18 | 3.51 | 10.79 | 130.56 | 1222.83 | 118.36 | 60.75 | 4.02 | 0.98 | 11.58 | 97.48 | 1.02 | 40.86 | 5596.85 | 0.65 | 0.69 | 1.51 | Shared |
| S14_vs_88666 | 0 | 0 | 0 | 0.48 | 0.07 | 0.21 | 0.02 | 99.56 | 0.5 | 1.68 | 0.44 | 0.11 | 1.02 | 0.26 | 0.23 | 1.2 | 137.87 | 0.24 | 0.2 | 0.36 | ME |
| S14_vs_30439 | 0 | 0 | 0 | 0.74 | 0.17 | 0.79 | 2.98 | 61.71 | 23.95 | 11.58 | 0.59 | 0.28 | 0.29 | 1.44 | 0.73 | 0.28 | 96.93 | 0.27 | 14.17 | 5.24 | Shared |
| S14_vs_96286 | 0 | 0 | 0 | 1.31 | 0.06 | 1.26 | 0.99 | 14.53 | 0.36 | 4.33 | 0.52 | 0.25 | 14.23 | 0.89 | 2.48 | 9.42 | 26.7 | 1.4 | 3.24 | 1.14 | Shared |
| S14_vs_9676 | 0.06 | 0.01 | 0 | 91.56 | 0.53 | 1.19 | 0.2 | 0.4 | 0.2 | 0.33 | 5.68 | 0.56 | 2.36 | 0.33 | 0.75 | 38.67 | 1086.22 | 0.5 | 0.7 | 1.44 | ME |
| S14_vs_3251 | 0.02 | 0 | 0.01 | 0.63 | 0.17 | 0.87 | 0.04 | 2.75 | 0.65 | 1.36 | 1.9 | 0.38 | 0.3 | 0.66 | 0.38 | 0.41 | 28.35 | 0.15 | 2.39 | 0.88 | ME |
| S14_vs_26320 | 0 | 0 | 0 | 1.14 | 0.17 | 1.83 | 0.19 | 0.48 | 0.31 | 0.71 | 1.21 | 0.26 | 1.39 | 0.9 | 1.21 | 0.7 | 203.27 | 0.72 | 1.05 | 2.48 | ME |
| S14_vs_4887 | 0 | 0 | 0 | 0.71 | 0.29 | 0.77 | 0.07 | 3.37 | 0.35 | 2.56 | 0.59 | 0.11 | 0.25 | 0.19 | 0.34 | 0.2 | 276.96 | 0.15 | 0.37 | 1.11 | ME |
| S14_vs_117153 | 0 | 0 | 0 | 1.34 | 0.14 | 0.86 | 0.14 | 246.29 | 0.16 | 0.55 | 0.56 | 0.14 | 1.12 | 0.5 | 0.41 | 0.53 | 70.13 | 0.39 | 0.84 | 1.62 | ME |
| S14_vs_117158 | 0 | 0 | 0 | 0.68 | 0.07 | 0.42 | 0.04 | 48.07 | 0.11 | 0.14 | 0.59 | 0.12 | 0.49 | 0.23 | 0.34 | 0.39 | 40.17 | 0.31 | 28.21 | 0.56 | Shared |
| S14_vs_117160 | 0.05 | 0 | 0 | 0.52 | 0.96 | 0.75 | 0.07 | 0.29 | 0.2 | 0.21 | 0.88 | 0.32 | 1.28 | 0.61 | 0.78 | 0.51 | 1188.97 | 0.49 | 0.99 | 1.98 | ME |
| S14_vs_117161 | 0.01 | 0 | 0 | 1.32 | 0.09 | 0.26 | 0.14 | 3.4 | 5.1 | 18.97 | 0.38 | 0.11 | 0.77 | 0.66 | 5.94 | 0.29 | 97.82 | 0.42 | 0.51 | 5.47 | ME |
| S14_vs_117165 | 0.01 | 0 | 0 | 5.39 | 0.35 | 6.44 | 1.02 | 0.4 | 0.51 | 42.43 | 1.49 | 1.36 | 0.89 | 4.61 | 0.6 | 1.27 | 94.74 | 0.46 | 4.19 | 1.09 | ME |
| S14_vs_117166 | 0.05 | 0 | 0 | 2.72 | 0.33 | 0.6 | 0.05 | 1028.2 | 59.88 | 2 | 0.95 | 0.23 | 3.02 | 0.92 | 0.34 | 0.9 | 279.88 | 0.19 | 0.95 | 0.54 | Shared |
| S14_vs_73301 | 0 | 0 | 0 | 0.6 | 0.04 | 0.47 | 0.18 | 7.75 | 0.17 | 0.4 | 0.63 | 0.08 | 1.41 | 0.36 | 0.24 | 0.34 | 31.18 | 0.38 | 0.36 | 0.88 | ME |
| S14_vs_14737 | 0 | 0 | 0 | 0.74 | 0.05 | 0.34 | 3 | 10.35 | 0.16 | 2.1 | 1.51 | 0.09 | 0.23 | 0.32 | 0.36 | 0.28 | 26.87 | 0.94 | 0.34 | 1.55 | ME |
| S14_vs_102698 | 0 | 0 | 0 | 3.03 | 4.16 | 6.74 | 0.16 | 24.97 | 1.42 | 9.72 | 2.54 | 3.65 | 0.36 | 0.48 | 0.5 | 0.47 | 116.8 | 0.3 | 6.41 | 2.38 | ME |
| S14_vs_66139 | 0 | 0 | 0 | 3.76 | 0.89 | 7.87 | 0.64 | 4.09 | 0.5 | 16.04 | 13.13 | 4.04 | 3.08 | 11.32 | 1.64 | 1.98 | 33.47 | 0.48 | 4.4 | 1.89 | Shared |
| S14_vs_66208 | 0 | 0 | 0 | 26.2 | 3.33 | 15.18 | 0.62 | 20.6 | 10.57 | 6.2 | 5.2 | 2.18 | 285.56 | 3.33 | 4.68 | 45.18 | 60.59 | 259.56 | 18.5 | 27.08 | Shared |
| S14_vs_632 | 0.02 | 0 | 0 | 0.78 | 0.06 | 0.34 | 0.16 | 73.56 | 0.15 | 0.54 | 0.51 | 3.52 | 0.42 | 0.37 | 0.45 | 0.35 | 30.04 | 0.46 | 0.3 | 0.55 | ME |
| S14_vs_110290 | 0 | 0 | 0 | 0.88 | 0.11 | 0.65 | 0.07 | 0.69 | 0.11 | 0.19 | 0.39 | 2.55 | 1.03 | 0.39 | 13.21 | 0.27 | 23.82 | 0.17 | 0.53 | 0.59 | Shared |
| S14_vs_59347 | 0 | 0 | 0 | 0.51 | 0.04 | 0.42 | 0.16 | 7.49 | 0.14 | 0.16 | 0.42 | 0.08 | 0.6 | 0.66 | 0.53 | 0.41 | 32.89 | 0.25 | 0.41 | 0.7 | ME |
| S14_vs_30108 | 0 | 0 | 0 | 0.46 | 0.08 | 0.49 | 0.06 | 4.3 | 4.78 | 13.28 | 0.55 | 0.62 | 0.63 | 5 | 0.55 | 0.23 | 39.79 | 0.26 | 1.32 | 0.87 | ME |
| S14_vs_96111 | 0.01 | 0 | 0 | 1.21 | 0.1 | 0.73 | 0.04 | 270.39 | 0.2 | 0.72 | 2.03 | 0.13 | 1.14 | 0.64 | 1.6 | 0.83 | 58.03 | 0.29 | 1.56 | 2 | ME |
| S14_vs_74067 | 0.02 | 0.02 | 0.01 | 18.15 | 16.83 | 99.95 | 0.55 | 77.49 | 11.79 | 20.57 | 18.49 | 1.28 | 249.01 | 19.35 | 64.86 | 37.15 | 157.33 | 172.77 | 132.97 | 62.64 | Shared |
| S14_vs_30566 | 0 | 0 | 0 | 0.86 | 0.05 | 0.74 | 0.07 | 0.49 | 0.2 | 0.32 | 1.04 | 0.95 | 0.96 | 0.33 | 1.62 | 0.53 | 31.94 | 0.47 | 0.55 | 1.28 | ME |
| S14_vs_66841 | 0 | 0 | 0 | 0.56 | 0.03 | 0.26 | 0.03 | 42.44 | 0.17 | 4.69 | 0.91 | 0.07 | 0.31 | 0.44 | 0.27 | 0.2 | 16.53 | 0.2 | 13.8 | 0.44 | Shared |
| S14_vs_112068 | 0.01 | 0 | 0 | 2.8 | 0.12 | 0.48 | 0.09 | 0.6 | 0.6 | 1.23 | 0.55 | 0.63 | 0.35 | 0.74 | 5.77 | 0.39 | 36.39 | 0.24 | 1.14 | 1.64 | ME |
| S14_vs_38905 | 0.01 | 0 | 0.01 | 0.6 | 0.03 | 0.37 | 0.03 | 35.96 | 0.18 | 0.5 | 0.66 | 0.12 | 0.69 | 1.19 | 0.21 | 0.3 | 32.07 | 0.18 | 0.27 | 0.62 | ME |
| S14_vs_9162 | 0.1 | 0 | 0.04 | 9.3 | 0.43 | 47.4 | 7.62 | 30.28 | 3.78 | 26.76 | 20.84 | 5.97 | 10.07 | 12.88 | 12.04 | 14.03 | 69.12 | 6.62 | 11.85 | 44.32 | Shared |
| S14_vs_112252 | 0.01 | 0 | 0 | 0.86 | 0.03 | 0.41 | 0.03 | 0.33 | 0.17 | 0.24 | 0.42 | 0.08 | 0.78 | 0.37 | 0.48 | 0.44 | 24.23 | 0.3 | 0.66 | 0.74 | ME |
| S14_vs_112345 | 0.03 | 0.01 | 0 | 19.3 | 2.57 | 51.1 | 1.71 | 75.98 | 13.29 | 71.56 | 31.65 | 9.09 | 282.17 | 11.32 | 31.88 | 20.48 | 390.02 | 62.25 | 112.75 | 302.28 | Shared |
| S14_vs_39266 | 0.01 | 0 | 0 | 3.18 | 0.42 | 4.78 | 0.67 | 17.68 | 1.73 | 19.96 | 1.71 | 8.38 | 3.19 | 14.42 | 1.3 | 1.35 | 45.11 | 0.2 | 9.21 | 1.31 | ME |
| S14_vs_90934 | 0.03 | 0 | 0.02 | 1.6 | 0.34 | 15.75 | 1.11 | 0.54 | 0.17 | 0.78 | 1.01 | 373.69 | 1.15 | 0.2 | 4.85 | 0.67 | 46.16 | 2.17 | 1.01 | 1.7 | ME |
| S14_vs_39797 | 0 | 0 | 0 | 1.38 | 0.07 | 3.64 | 0.29 | 43.22 | 0.56 | 1 | 2.53 | 0.23 | 1.05 | 0.8 | 0.69 | 0.4 | 42.97 | 0.3 | 0.67 | 1.02 | ME |
| S14_vs_97835 | 0 | 0 | 0 | 0.44 | 0.06 | 0 | 0 | 17.4 | 0.04 | 2.59 | 0.39 | 0 | 0.58 | 0.15 | 0.42 | 0.07 | 33.98 | 0.37 | 0.11 | 0.13 | ME |
| S14_vs_61530 | 0.01 | 0 | 0 | 0.66 | 0.04 | 5.97 | 0.04 | 6.26 | 0.06 | 0.35 | 0.54 | 0.14 | 0.77 | 0.38 | 0.49 | 0.3 | 43.3 | 0.5 | 2.61 | 0.41 | ME |
| S14_vs_2870 | 0.04 | 0 | 0 | 3.56 | 2.11 | 1.84 | 0.17 | 0.22 | 0.08 | 1.03 | 1.82 | 0.56 | 0.53 | 0.27 | 0.28 | 2.85 | 73.43 | 1.58 | 0.31 | 0.62 | ME |
| S14_vs_32499 | 0.01 | 0 | 0.01 | 12.43 | 16.36 | 70.17 | 0.77 | 22.7 | 8.81 | 12.94 | 13.9 | 1.03 | 109.04 | 11.65 | 21.54 | 26.39 | 122.88 | 67.07 | 57.45 | 96.19 | Shared |
| S14_vs_62178 | 0 | 0 | 0 | 0.84 | 0.09 | 1.25 | 0.07 | 0.4 | 0.18 | 0.21 | 0.72 | 0.22 | 0.55 | 0.55 | 0.53 | 0.28 | 82.04 | 0.34 | 0.55 | 0.67 | ME |
| S14_vs_25852 | 0.01 | 0 | 0 | 0.63 | 0.48 | 1.36 | 1.61 | 32.02 | 23.61 | 5.55 | 7.39 | 0.17 | 5.58 | 6.7 | 2.09 | 1.35 | 43.76 | 7.49 | 23.38 | 13.12 | Shared |
| S14_vs_11510 | 0.01 | 0.02 | 0 | 4.97 | 1.03 | 32.83 | 1.51 | 72.15 | 49.06 | 10.17 | 27 | 1.21 | 38.42 | 9 | 47.14 | 7.38 | 75.33 | 40.67 | 70.26 | 13.05 | Shared |
| S14_vs_78164 | 0 | 0 | 0 | 1.67 | 0.24 | 3.33 | 0.31 | 0.55 | 0.74 | 1.55 | 2.24 | 0.6 | 2.22 | 0.52 | 2.48 | 0.59 | 32.53 | 0.66 | 1.83 | 4.27 | ME |
| S14_vs_56251 | 0 | 0 | 0 | 0.64 | 0.1 | 0.72 | 0.15 | 2.25 | 0.29 | 1.37 | 0.68 | 0.21 | 0.7 | 0.81 | 0.33 | 0.21 | 121.87 | 0.64 | 0.51 | 1.13 | ME |
| S14_vs_70985 | 0.01 | 0 | 0 | 12.86 | 0.74 | 9.75 | 0.64 | 10.97 | 2.41 | 3.84 | 4.67 | 0.94 | 18.53 | 2.04 | 33.32 | 2.45 | 48.43 | 5.32 | 6.46 | 5.08 | Shared |
| S14_vs_56410 | 0.01 | 0 | 0 | 0.87 | 0.15 | 0.87 | 0.34 | 7.08 | 0.83 | 4.64 | 3.81 | 0.15 | 0.3 | 0.28 | 0.23 | 0.31 | 20.89 | 0.21 | 0.58 | 2.02 | ME |
| S14_vs_100227 | 0.03 | 0 | 0 | 1.27 | 0.09 | 0.48 | 0.12 | 4.79 | 3.83 | 0.7 | 0.49 | 0.65 | 0.67 | 6.1 | 0.35 | 0.31 | 55.77 | 0.35 | 1.04 | 0.96 | ME |
| S14_vs_115377 | 0 | 0 | 0 | 0.28 | 0.03 | 0.42 | 0.08 | 120.96 | 0.52 | 0.35 | 0.26 | 0.87 | 0.55 | 1.1 | 0.23 | 0.23 | 24.26 | 0.32 | 0.61 | 0.91 | ME |
| S14_vs_93565 | 0 | 0 | 0 | 0.82 | 0.07 | 0.74 | 0.01 | 1.38 | 0.17 | 1.01 | 0.94 | 0.21 | 0.32 | 0.43 | 0.5 | 2.1 | 19.48 | 0.31 | 0.81 | 1.04 | ME |

| | | | | | | | | | | | | | | | | | | | | | |
|---|---|---|---|---|---|---|---|---|---|---|---|---|---|---|---|---|---|---|---|---|---|
| S14_vs_20193 | 0.02 | 0 | 0 | 0.62 | 0.15 | 0.57 | 0.08 | 0.37 | 0.39 | 0.57 | 0.76 | 0.12 | 0.72 | 0.73 | 0.4 | 0.28 | 130.77 | 0.25 | 0.41 | 0.59 | ME |
| S14_vs_115455 | 0.01 | 0 | 0 | 1.08 | 0.08 | 0.32 | 0.18 | 97.57 | 0.38 | 0.19 | 0.74 | 0.09 | 0.71 | 1.04 | 0.4 | 0.34 | 55.69 | 0.3 | 0.41 | 0.76 | ME |
| S14_vs_108376 | 0 | 0 | 0 | 1.21 | 0.03 | 1.62 | 0.16 | 24.68 | 0.15 | 0.7 | 0.46 | 0.04 | 0.33 | 0.26 | 1.58 | 0.33 | 14.71 | 0.15 | 0.32 | 0.43 | ME |
| S14_vs_79505 | 0 | 0 | 0 | 0.63 | 0.06 | 0.28 | 0.16 | 0.16 | 0.14 | 0.09 | 0.33 | 0.09 | 0.31 | 0.14 | 0.32 | 0.17 | 30.74 | 0.21 | 0.21 | 0.49 | ME |
| S14_vs_101482 | 0 | 0 | 0.01 | 1.08 | 0.09 | 1.14 | 0.56 | 58.06 | 0.11 | 0.31 | 0.74 | 0.12 | 0.32 | 1.84 | 0.72 | 0.48 | 57.54 | 0.31 | 0.29 | 0.82 | ME |
| S14_vs_67094 | 0.01 | 0 | 0 | 1.31 | 0.02 | 1.24 | 0.17 | 24.32 | 0.22 | 1.92 | 1.68 | 0.61 | 0.69 | 0.52 | 0.66 | 1.36 | 14.19 | 0.63 | 1.19 | 1.9 | ME |
| S1_vf_140259 | 0 | 0 | 0 | 12.11 | 0 | 0.19 | 0 | 0.02 | 0.07 | 0.06 | 0.25 | 0.02 | 0.05 | 0.06 | 0.89 | 7.51 | 0 | 0.11 | 0.12 | 0.45 | ME |
| S1_vf_28398 | 0 | 0.04 | 0 | 773.56 | 0.11 | 0.38 | 0.07 | 0.52 | 0.21 | 0.25 | 0.74 | 0.08 | 0.31 | 0.09 | 0.33 | 5.57 | 0.26 | 1.27 | 2.87 | 6.15 | ME |
| S1_vf_22149 | 0 | 0.01 | 0 | 39 | 0.28 | 0.26 | 0.23 | 0.26 | 0.1 | 0.18 | 0.53 | 0.07 | 0.4 | 0.23 | 0.31 | 12.07 | 0.22 | 0.34 | 0.31 | 0.45 | ME |
| S1_vf_136526 | 0.01 | 0.01 | 0 | 382.86 | 0.07 | 0.29 | 0.13 | 0.23 | 0.11 | 0.35 | 0.83 | 0.1 | 0.42 | 0.13 | 0.35 | 19.44 | 0.12 | 1.18 | 0.27 | 0.46 | ME |
| S1_vf_50591 | 0 | 0 | 0 | 71.75 | 0.05 | 0.35 | 0.04 | 0.68 | 0.09 | 0.09 | 0.17 | 0.06 | 0.2 | 0.09 | 0.17 | 29.13 | 0.24 | 0.24 | 0.24 | 0.37 | ME |
| S1_vf_138157 | 0 | 0.01 | 0 | 87.61 | 0.16 | 1.89 | 0.13 | 4.45 | 0.86 | 1.21 | 1188.7 | 0.21 | 0.86 | 0.67 | 0.77 | 4.59 | 4.61 | 1.66 | 0.72 | 4.65 | Shared |
| S1_vf_125650 | 0.02 | 0 | 0 | 205.81 | 25.37 | 0.3 | 0 | 0.15 | 0.13 | 0.15 | 0.69 | 0.04 | 0.17 | 0.14 | 0.26 | 13.72 | 0.09 | 0.31 | 0.3 | 0.15 | Shared |
| S1_vf_6272 | 0.01 | 0 | 0 | 71.71 | 0.02 | 0.39 | 0.01 | 0.29 | 0.14 | 0.15 | 49.87 | 0.08 | 0.28 | 0.38 | 0.29 | 14.3 | 0.25 | 1.9 | 7.32 | 0.48 | Shared |
| S1_vf_158961 | 0.01 | 0.01 | 0 | 51.54 | 0.04 | 0.35 | 0.04 | 0.28 | 0.11 | 0.18 | 0.51 | 0.07 | 0.32 | 0.25 | 0.37 | 17.66 | 0.27 | 0.33 | 0.45 | 0.69 | ME |
| S1_vf_159951 | 0.01 | 0.01 | 0 | 159.71 | 3.04 | 0.23 | 0.04 | 0.14 | 0.13 | 0.13 | 4.44 | 0.03 | 0.28 | 0.2 | 0.26 | 42.67 | 0.15 | 0.4 | 0.26 | 0.38 | ME |
| S1_vs_143929 | 0 | 0.01 | 0 | 58.83 | 0.09 | 0.38 | 0.33 | 0.34 | 0.11 | 0.17 | 4.09 | 0.18 | 4.7 | 0.21 | 0.39 | 5.1 | 0.24 | 0.74 | 0.34 | 0.58 | ME |
| S1_vs_133577 | 0 | 0.01 | 0 | 23.23 | 0.04 | 1.83 | 0.42 | 0.81 | 0.18 | 0.86 | 0.92 | 0.32 | 0.44 | 0.22 | 0.55 | 1.94 | 0.32 | 0.31 | 2.68 | 2.26 | ME |
| S1_vs_21439 | 0.01 | 0.01 | 0 | 60.41 | 0.06 | 0.31 | 0.02 | 0.39 | 0.35 | 0.27 | 44.41 | 0.12 | 0.66 | 0.41 | 0.43 | 3.44 | 0.24 | 0.36 | 0.32 | 0.7 | Shared |
| S1_vs_155183 | 0 | 0 | 0 | 233.25 | 0.11 | 0.36 | 0.06 | 0.27 | 0.17 | 0.21 | 265.56 | 0.12 | 0.86 | 0.25 | 2.17 | 20.94 | 0.44 | 0.82 | 1.14 | 0.79 | Shared |
| S1_vs_62409 | 0 | 0 | 0 | 29.05 | 30.55 | 0.32 | 0.02 | 2.09 | 2.88 | 0.44 | 0.29 | 0.08 | 0.77 | 15.79 | 192.65 | 5.19 | 0.31 | 0.18 | 0.21 | 0.52 | Shared |
| S1_vs_117317 | 0.01 | 0.01 | 0 | 48.58 | 0.04 | 0.31 | 0.04 | 0.25 | 0.13 | 0.21 | 1.34 | 0.05 | 0.37 | 0.29 | 0.41 | 2.15 | 0.21 | 0.48 | 0.49 | 0.54 | ME |
| S1_vs_23682 | 0 | 0 | 0 | 116.29 | 0.1 | 1.18 | 0.08 | 0.5 | 0.13 | 0.54 | 0.69 | 0.1 | 0.65 | 0.26 | 0.34 | 11.3 | 1.3 | 0.59 | 0.42 | 0.78 | ME |
| S1_vs_3267 | 0 | 0 | 0 | 22.36 | 0 | 20.22 | 0.01 | 0.12 | 0.09 | 0.03 | 0.08 | 0.02 | 0.2 | 0.05 | 0.13 | 12 | 0.18 | 0.06 | 3.05 | 0.36 | ME |
| S1_vs_3435 | 0 | 0 | 0 | 25.84 | 0 | 0.27 | 0 | 0.28 | 0.11 | 0.09 | 0.23 | 0.04 | 1.18 | 0.17 | 0.62 | 5.51 | 0.37 | 0.34 | 0.18 | 0.74 | ME |
| S1_vs_91270 | 0 | 0 | 0 | 82.45 | 0.17 | 0.22 | 0.05 | 0.21 | 0.14 | 2.1 | 5.34 | 0.03 | 0.28 | 0.15 | 0.34 | 0.33 | 0.18 | 0.28 | 0.21 | 0.42 | ME |
| S1_vs_145792 | 0.01 | 0.01 | 0 | 51.7 | 0.06 | 0.32 | 0.1 | 0.26 | 0.1 | 0.17 | 0.68 | 0.08 | 0.53 | 0.26 | 0.33 | 3 | 0.24 | 0.39 | 1.32 | 2.14 | ME |
| S1_vs_100019 | 0 | 0 | 0 | 75.44 | 0.02 | 0.37 | 0.02 | 0.18 | 0.07 | 0.15 | 0.4 | 0.02 | 0.93 | 0.16 | 0.18 | 21.02 | 0.24 | 0.33 | 3.03 | 7.17 | ME |
| S3_vf_8058 | 0 | 0 | 0.02 | 0.44 | 0.05 | 160.96 | 0.49 | 0.22 | 0.08 | 0.1 | 0.28 | 0.27 | 0.36 | 0.17 | 0.26 | 0.21 | 0.23 | 0.21 | 0.33 | 0.59 | ME |
| S3_vf_8164 | 0 | 0 | 0.01 | 0.97 | 0.01 | 35.95 | 0.04 | 0.18 | 0.09 | 1.76 | 0.38 | 0.05 | 0.45 | 0.37 | 1.08 | 0.2 | 0.23 | 2.85 | 0.56 | 0.83 | ME |
| S3_vf_111776 | 0 | 0 | 0.33 | 0.39 | 0.39 | 3273.02 | 0.11 | 0.52 | 0.45 | 0.61 | 1.46 | 4.48 | 0.24 | 0.19 | 0.29 | 0.73 | 0.29 | 0.26 | 0.29 | 0.26 | ME |
| S3_vf_87896 | 0 | 0 | 0 | 0.53 | 0.01 | 32.36 | 0.03 | 0.38 | 0.1 | 0.17 | 0.51 | 0.11 | 0.59 | 0.5 | 0.67 | 0.37 | 1.71 | 0.26 | 0.39 | 0.98 | ME |
| S3_vf_111893 | 0.19 | 0 | 0.07 | 9.53 | 0.18 | 1033.62 | 0.04 | 0.22 | 0.22 | 0.42 | 0.67 | 1.63 | 0.31 | 0.25 | 44.52 | 1.51 | 2.7 | 7.5 | 9.33 | 1.5 | Shared |
| S3_vf_96222 | 0.32 | 0 | 0.03 | 0.81 | 2.79 | 394.21 | 114.11 | 9.33 | 3.2 | 5.23 | 19.69 | 6.75 | 1.13 | 1.05 | 1.01 | 0.39 | 4242.9 | 0.71 | 6.67 | 355.21 | Shared |
| S3_vf_80287 | 0.01 | 0 | 0.01 | 0.68 | 0.01 | 104.72 | 0.31 | 0.17 | 0.12 | 0.09 | 1.84 | 0.24 | 1.81 | 2.24 | 1.9 | 0.42 | 0.23 | 16.14 | 0.31 | 0.52 | Shared |
| S3_vf_56363 | 0 | 0 | 0.05 | 0.88 | 0.27 | 1175.93 | 12.07 | 107.94 | 0.46 | 0.81 | 1.55 | 3.49 | 2.13 | 0.98 | 1.56 | 1.25 | 2.49 | 1.22 | 1.46 | 1.32 | Shared |
| S3_vf_33389 | 0.13 | 0 | 0.04 | 2.19 | 0.39 | 432.12 | 0.04 | 0.11 | 0.09 | 0.23 | 0.35 | 648.36 | 0.39 | 1.23 | 0.62 | 1.62 | 0.34 | 0.46 | 1708.36 | 143.32 | Shared |
| S3_vf_73597 | 0 | 0 | 0.01 | 0.72 | 0.15 | 29.14 | 0.2 | 0.21 | 0.31 | 0.27 | 0.71 | 0.18 | 1.07 | 39.36 | 0.48 | 0.75 | 13.02 | 1.11 | 1.33 | 7.03 | ME |
| S3_vf_25163 | 0.02 | 0 | 0.08 | 0.35 | 0.15 | 760.09 | 0.08 | 0.36 | 0.19 | 0.27 | 6.03 | 0.97 | 19.11 | 0.15 | 26.24 | 344.95 | 0.3 | 0.5 | 0.7 | 0.59 | Shared |
| S3_vf_57422 | 0.02 | 0 | 0.13 | 3.69 | 0.1 | 1786.39 | 11.26 | 0.28 | 0.19 | 0.31 | 0.48 | 2.28 | 0.19 | 0.41 | 0.09 | 0.96 | 0.15 | 0.1 | 0.23 | 0.36 | Shared |
| S3_vf_17748 | 2.3 | 0 | 2.16 | 5.6 | 4.29 | 24202.18 | 0.62 | 6.39 | 5.72 | 2553.43 | 51.27 | 32.15 | 1473.23 | 0.83 | 4.68 | 8970.63 | 0.56 | 0.51 | 12.24 | 1.51 | Shared |
| S3_vf_2186 | 0 | 0 | 0.04 | 0.57 | 0.08 | 316.36 | 0.13 | 0.25 | 0.73 | 0.72 | 0.76 | 0.84 | 1.06 | 760.28 | 554.78 | 0.45 | 0.41 | 0.42 | 0.53 | 1.1 | Shared |
| S3_vf_11428 | 0 | 0 | 0.01 | 0.89 | 0.04 | 48.13 | 29.85 | 0.33 | 0.26 | 0.35 | 0.53 | 0.22 | 0.62 | 2.67 | 0.55 | 0.33 | 1.33 | 3.64 | 14.61 | 30.54 | Shared |
| S3_vf_18080 | 0 | 0 | 0.02 | 10.76 | 4.19 | 165.31 | 2.79 | 0.63 | 0.32 | 1.84 | 3.05 | 0.61 | 3.96 | 11.07 | 6.31 | 14.19 | 97.44 | 6.01 | 8.24 | 18.44 | ME |
| S3_vf_106401 | 0.01 | 0 | 0 | 0.75 | 0.12 | 31.5 | 2.8 | 3.39 | 0.13 | 1.96 | 0.66 | 0.28 | 2.06 | 0.86 | 0.56 | 0.32 | 86.78 | 0.38 | 1.64 | 0.99 | ME |
| S3_vf_59078 | 0 | 0 | 0 | 1.53 | 3.21 | 20.01 | 2.76 | 0.32 | 1.64 | 0.09 | 0.84 | 0.05 | 1.18 | 0.29 | 0.74 | 3.15 | 0.5 | 1.14 | 0.51 | 0.77 | ME |
| S3_vf_82571 | 0 | 0 | 0.03 | 0.65 | 0.03 | 463.07 | 0.26 | 0.14 | 0.08 | 0.13 | 6.98 | 0.55 | 8.9 | 0.07 | 17.74 | 0.22 | 0.15 | 0.2 | 1.45 | 0.59 | Shared |
| S3_vf_35073 | 0 | 0 | 0.02 | 1.02 | 0.03 | 110.94 | 0.11 | 0.34 | 0.24 | 0.14 | 0.77 | 0.23 | 1.04 | 0.18 | 0.55 | 0.5 | 0.25 | 0.38 | 0.6 | 0.79 | ME |
| S3_vf_83536 | 0 | 0 | 0.04 | 0.73 | 0.04 | 262.43 | 0.09 | 0.3 | 0.19 | 0.1 | 0.64 | 0.43 | 0.37 | 0.11 | 0.39 | 0.26 | 0.12 | 0.3 | 0.37 | 0.73 | ME |
| S3_vf_18829 | 0.02 | 0 | 0.02 | 0.4 | 0.05 | 372.84 | 10.39 | 0.18 | 0.17 | 0.12 | 0.46 | 1.2 | 6.03 | 32.06 | 0.38 | 0.24 | 0.28 | 0.24 | 0.35 | 0.64 | ME |
| S3_vf_115304 | 0.37 | 0 | 1.18 | 2.05 | 1.37 | 12099.04 | 3647.6 | 1.55 | 1.72 | 12.79 | 3.71 | 18.38 | 2.8 | 2.07 | 14583.94 | 1.75 | 2.78 | 1.78 | 1.64 | 4.02 | Shared |
| S3_vf_67333 | 0.15 | 0 | 0.1 | 3.76 | 0.66 | 1541.92 | 0.1 | 0.42 | 0.3 | 0.39 | 5.64 | 2.04 | 0.46 | 0.31 | 0.65 | 1.2 | 0.26 | 0.33 | 0.57 | 0.79 | ME |
| S3_vf_99626 | 0 | 0 | 0.05 | 0.92 | 0.12 | 821.15 | 254.23 | 0.32 | 0.24 | 0.18 | 0.78 | 1.04 | 0.38 | 0.22 | 0.37 | 0.54 | 0.25 | 0.21 | 0.35 | 0.67 | Shared |
| S3_vf_44405 | 0 | 0 | 0.02 | 0.54 | 0.05 | 167.94 | 0.19 | 0.23 | 0.39 | 1.22 | 31.27 | 0.38 | 0.24 | 3.3 | 5.03 | 1.96 | 6.35 | 1.55 | 1.82 | 0.39 | Shared |
| S3_vf_99734 | 0.12 | 0 | 0.15 | 8.71 | 13.41 | 2005.64 | 11.08 | 1.74 | 0.61 | 4.23 | 6.71 | 7.11 | 2.03 | 0.31 | 105.01 | 9.7 | 10.15 | 0.28 | 2.73 | 1.76 | Shared |
| S3_vf_67828 | 0 | 0 | 6.63 | 10.25 | 8.57 | 56345.14 | 0.03 | 9.33 | 9.31 | 9.73 | 21.23 | 119.17 | 0.23 | 0.14 | 2.35 | 7.03 | 0.07 | 0.41 | 0.35 | 0.62 | Shared |
| S3_vf_84832 | 0 | 0 | 0 | 0.62 | 0.04 | 24.96 | 0.02 | 0.24 | 0.23 | 0.21 | 0.41 | 0.12 | 0.5 | 0.24 | 0.92 | 0.37 | 0.29 | 0.47 | 0.49 | 0.92 | ME |
| S3_vf_61860 | 0 | 0 | 0.09 | 1.07 | 0.33 | 930.09 | 1.07 | 6.31 | 0.42 | 0.49 | 1.73 | 1.51 | 3.58 | 0.98 | 1.56 | 1.82 | 16.13 | 2.92 | 1.06 | 1.5 | ME |
| S3_vf_100440 | 0 | 0 | 0 | 10.41 | 0.05 | 19.34 | 3.53 | 12.09 | 0.1 | 0.32 | 3.91 | 0.14 | 2.84 | 4.3 | 6.21 | 2.62 | 1.24 | 1.49 | 5.52 | 4.79 | ME |
| S3_vf_108959 | 0 | 0 | 0.04 | 1.76 | 0.17 | 324.47 | 0.14 | 0.35 | 0.26 | 0.28 | 0.86 | 0.53 | 1.33 | 0.6 | 2.26 | 0.66 | 0.56 | 0.5 | 2.7 | 1.27 | ME |
| S3_vf_109277 | 0 | 0 | 0 | 0.64 | 0 | 19.79 | 0.03 | 0.14 | 0.09 | 0.09 | 2.73 | 0.1 | 30.59 | 3.32 | 0.25 | 4.98 | 0.35 | 0.2 | 0.39 | 1.54 | Shared |
| S3_vf_5525 | 0.01 | 0 | 0 | 0.55 | 0 | 26.8 | 0.27 | 0.14 | 0.06 | 0.08 | 0.3 | 0.11 | 0.1 | 0.05 | 0.15 | 0.22 | 0.09 | 0.13 | 0.15 | 0.24 | ME |
| S3_vf_54353 | 0 | 0 | 0 | 0.27 | 0.02 | 91.16 | 0.24 | 0.19 | 0.05 | 0.06 | 1.56 | 0.18 | 0.19 | 0.15 | 0.23 | 0.19 | 0.21 | 0.12 | 0.13 | 0.54 | ME |
| S3_vf_54498 | 0.05 | 0 | 0 | 1.36 | 0 | 26.69 | 0.17 | 0.37 | 0.2 | 0.23 | 0.35 | 0.13 | 0.72 | 0.11 | 0.29 | 0.22 | 0.25 | 0.15 | 0.31 | 0.6 | ME |
| S3_vf_101591 | 0 | 0 | 0 | 0.22 | 0.02 | 20.46 | 4.26 | 0.3 | 0.36 | 0.15 | 0.78 | 0.11 | 0.69 | 0.11 | 0.21 | 0.38 | 0.42 | 0.55 | 0.33 | 1 | ME |
| S3_vf_101611 | 0 | 0 | 0 | 0.54 | 0.07 | 59.39 | 0.06 | 0.27 | 0.15 | 3.19 | 0.53 | 0.15 | 4.44 | 0.43 | 3.26 | 0.31 | 0.26 | 2.9 | 15.52 | 0.76 | Shared |
| S3_vf_7137 | 0 | 0 | 0 | 6.82 | 10.11 | 14.73 | 0.02 | 9.43 | 8.14 | 70748.48 | 20.55 | 0.04 | 0.27 | 0.08 | 85.33 | 6.52 | 0.11 | 0.17 | 0.14 | 0.4 | Shared |
| S3_vf_22968 | 0 | 0 | 0 | 1.01 | 0 | 62.02 | 0.21 | 0.27 | 0.18 | 1 | 0.7 | 0.15 | 1.05 | 0.31 | 0.74 | 0.48 | 0.65 | 0.31 | 0.84 | 1.86 | ME |
| S3_vf_23042 | 0.27 | 0 | 0.04 | 0.32 | 3.9 | 241.41 | 121.03 | 12.11 | 3.17 | 3.25 | 16.01 | 6.5 | 1.52 | 1.24 | 1.57 | 0.66 | 5731.35 | 0.71 | 4.39 | 206.65 | Shared |
| S3_vf_70746 | 0.01 | 0 | 0.02 | 0.99 | 0.03 | 106.38 | 0.56 | 0.23 | 0.1 | 0.09 | 0.54 | 0.24 | 0.29 | 0.1 | 0.42 | 0.43 | 0.2 | 0.23 | 10.27 | 0.75 | Shared |
| S3_vf_127168 | 0 | 0 | 0.02 | 0.31 | 0.19 | 787.16 | 0.02 | 406.81 | 585.49 | 0.3 | 60.8 | 0.91 | 0.74 | 1.11 | 0.31 | 0.47 | 0.29 | 0.23 | 0.62 | 0.78 | Shared |
| S3_vf_127392 | 0.1 | 0 | 0.02 | 0.61 | 0.03 | 85.42 | 0.13 | 0.26 | 0.13 | 0.13 | 0.51 | 0.23 | 0.33 | 0.23 | 0.35 | 0.22 | 27.17 | 1.25 | 0.22 | 0.6 | ME |
| S3_vf_127399 | 0 | 0 | 0 | 0.66 | 0.03 | 48.54 | 0.07 | 0.16 | 0.09 | 0.08 | 32.24 | 0.11 | 0.39 | 0.31 | 0.18 | 2.74 | 0.13 | 0.23 | 0.16 | 0.38 | Shared |
| S3_vf_127400 | 0.01 | 0.01 | 0.01 | 18.47 | 0.09 | 50.19 | 0.06 | 0.5 | 0.3 | 0.19 | 407.57 | 0.27 | 1.77 | 0.36 | 1.03 | 2.91 | 0.22 | 0.92 | 0.44 | 1.14 | Shared |
| S3_vf_127405 | 0 | 0 | 0 | 2.47 | 0.06 | 41.17 | 0.05 | 0.23 | 0.27 | 0.26 | 0.55 | 0.14 | 5.07 | 0.66 | 0.76 | 1.29 | 3.99 | 2.62 | 5.46 | 0.8 | ME |
| S3_vf_127406 | 0.02 | 0.01 | 0.03 | 2.63 | 0.07 | 509.53 | 0.09 | 0.29 | 0.16 | 0.14 | 0.84 | 0.79 | 0.42 | 0.19 | 0.42 | 13.81 | 0.23 | 0.63 | 2.35 | 5.18 | ME |
| S3_vf_127408 | 0.01 | 0 | 0.03 | 0.48 | 0.01 | 174.14 | 0.02 | 0.16 | 0.08 | 0.1 | 0.25 | 0.2 | 0.18 | 0.16 | 0.21 | 0.15 | 0.15 | 0.1 | 0.19 | 0.33 | ME |
| S3_vf_127409 | 0.01 | 0 | 0 | 1.2 | 0.05 | 52.77 | 0.15 | 0.28 | 0.13 | 0.16 | 90.06 | 0.13 | 0.48 | 0.47 | 0.17 | 0.34 | 0.35 | 0.35 | 0.3 | 0.82 | Shared |
| S3_vf_127423 | 0 | 0 | 0.26 | 0.74 | 0.34 | 2637.65 | 0.38 | 0.86 | 0.54 | 0.62 | 1.56 | 3.81 | 1.16 | 0.32 | 1.24 | 0.82 | 0.51 | 0.52 | 0.81 | 1.12 | ME |
| S3_vf_127426 | 0 | 0 | 0 | 0.52 | 0.02 | 23.12 | 0.22 | 0.31 | 0.37 | 1.08 | 0.9 | 0.19 | 0.39 | 0.08 | 0.24 | 1.03 | 0.19 | 0.27 | 0.54 | 2.15 | ME |
| S3_vf_127431 | 0 | 0 | 0 | 0.26 | 0 | 76.75 | 0.03 | 0.16 | 0.12 | 0.19 | 0.2 | 0.07 | 0.73 | 0.73 | 0.43 | 0.24 | 0.76 | 0.17 | 0.63 | 0.89 | ME |
| S3_vf_127447 | 0.01 | 0 | 0.03 | 0.41 | 0.07 | 378.85 | 0.1 | 0.33 | 0.22 | 0.18 | 51.46 | 0.59 | 1.03 | 0.38 | 0.55 | 0.39 | 0.32 | 0.55 | 0.45 | 0.76 | Shared |
| S3_vf_127450 | 0 | 0 | 0.53 | 0.81 | 0.63 | 5254.48 | 0.13 | 1.03 | 0.76 | 0.99 | 2.04 | 6.63 | 2.11 | 0.45 | 1.25 | 1.23 | 0.44 | 0.33 | 0.85 | 1.1 | ME |
| S3_vs_119605 | 0 | 0 | 0.01 | 3.72 | 0.23 | 27.51 | 0.16 | 10.14 | 0.54 | 7.05 | 3.94 | 4.37 | 1.64 | 1.69 | 2.42 | 10.55 | 13.87 | 0.48 | 6.15 | 1.32 | ME |

| | | | | | | | | | | | | | | | | | | | | | |
|---|---|---|---|---|---|---|---|---|---|---|---|---|---|---|---|---|---|---|---|---|---|
| S3_vs_80151 | 0.9 | 0.03 | 1.41 | 15.19 | 1.85 | 17429.87 | 3079.55 | 2.17 | 2.31 | 2.39 | 4.59 | 22.4 | 0.74 | 0.23 | 0.43 | 1.93 | 0.42 | 0.32 | 0.46 | 1.28 | Shared |
| S3_vs_16825 | 0.19 | 0 | 0.18 | 79.83 | 35.71 | 2451.15 | 0.1 | 2 | 0.57 | 0.59 | 5.66 | 3.2 | 0.64 | 3.18 | 0.67 | 7.69 | 0.57 | 0.4 | 0.62 | 1.15 | Shared |
| S3_vs_49835 | 0.06 | 0 | 0.22 | 73.53 | 0.3 | 3104.54 | 0.11 | 1.2 | 0.55 | 1.04 | 1.37 | 5 | 0.53 | 0.38 | 0.44 | 26.02 | 0.59 | 0.47 | 0.57 | 1.13 | ME |
| S3_vs_89485 | 0 | 0 | 0 | 0.77 | 0.28 | 28.23 | 0.58 | 0.57 | 0.51 | 2.2 | 1085 | 0.12 | 1.59 | 0.3 | 0.48 | 0.26 | 0.25 | 0.51 | 1.69 | 2.56 | Shared |
| S3_vs_81793 | 0.01 | 0 | 0.01 | 1.41 | 0.02 | 86.69 | 0.09 | 1.64 | 0.24 | 0.77 | 3.47 | 2.81 | 1.84 | 0.44 | 0.5 | 1.5 | 0.57 | 0.45 | 0.97 | 1.86 | ME |
| S3_vs_90331 | 0.02 | 0 | 0.01 | 1.18 | 0.03 | 47.56 | 0.1 | 0.39 | 0.12 | 1.07 | 3.05 | 0.29 | 2.65 | 0.17 | 0.54 | 0.91 | 0.31 | 6.67 | 12.91 | 0.97 | Shared |
| S3_vs_90341 | 0.01 | 0 | 0.01 | 1.31 | 0.02 | 108.88 | 0.07 | 0.26 | 0.09 | 0.19 | 12.7 | 0.21 | 0.38 | 0.12 | 0.73 | 0.97 | 0.26 | 0.25 | 0.87 | 0.51 | Shared |
| S3_vs_50924 | 0.02 | 0 | 0.03 | 2.09 | 0.09 | 473.77 | 0.06 | 0.51 | 0.78 | 1.09 | 0.65 | 0.84 | 0.32 | 0.09 | 0.48 | 0.33 | 0.42 | 0.19 | 0.29 | 1.29 | ME |
| S3_vs_98675 | 0 | 0 | 0.04 | 0.33 | 0.14 | 727.66 | 0.86 | 2.92 | 0.32 | 0.46 | 0.91 | 1.22 | 2.22 | 0.49 | 0.75 | 0.98 | 0.86 | 0.89 | 0.95 | 1.57 | ME |
| S3_vs_82895 | 0.11 | 0 | 0.36 | 21.71 | 0.53 | 4629.97 | 133.2 | 3.13 | 0.92 | 17.33 | 36.15 | 5.9 | 18.17 | 65.25 | 0.21 | 9.29 | 10.44 | 0.17 | 0.2 | 0.39 | Shared |
| S3_vs_108076 | 0 | 0 | 0.04 | 0.6 | 0.13 | 115.56 | 0.07 | 0.35 | 0.13 | 0.55 | 1.14 | 0.45 | 1.07 | 0.41 | 0.48 | 0.53 | 0.74 | 0.23 | 1.66 | 2.72 | ME |
| S3_vs_28402 | 0 | 0 | 0.02 | 1.28 | 0.15 | 268.83 | 0.05 | 1.28 | 0.19 | 1.43 | 1.04 | 3.13 | 0.69 | 0.89 | 1.78 | 0.36 | 7.29 | 0.35 | 2.48 | 0.68 | ME |
| S3_vs_127391 | 0.02 | 0 | 0 | 0.75 | 0.05 | 227.16 | 0.15 | 0.41 | 0.09 | 0.16 | 0.62 | 0.37 | 0.3 | 0.12 | 0.23 | 0.33 | 1.12 | 0.16 | 5.52 | 0.45 | ME |
| S3_vs_127395 | 0.01 | 0 | 0 | 0.74 | 0.02 | 23.89 | 0.03 | 0.26 | 0.14 | 0.25 | 0.54 | 0.16 | 0.45 | 0.49 | 0.39 | 0.28 | 0.34 | 0.31 | 0.6 | 0.9 | ME |
| S3_vs_127397 | 0 | 0 | 0 | 0.8 | 0.03 | 71.5 | 0.4 | 0.25 | 0.15 | 0.2 | 5.77 | 0.18 | 0.38 | 0.26 | 0.46 | 0.18 | 0.28 | 0.38 | 0.27 | 0.64 | ME |
| S3_vs_127401 | 0.01 | 0 | 0.01 | 1.36 | 0.09 | 101.21 | 0.13 | 0.32 | 0.18 | 0.18 | 1.02 | 0.32 | 0.63 | 0.22 | 0.32 | 0.58 | 2.04 | 0.53 | 0.61 | 4.2 | ME |
| S3_vs_127404 | 0.25 | 0 | 0.1 | 0.51 | 0.09 | 928.71 | 0.02 | 0.48 | 0.32 | 0.27 | 0.66 | 1.29 | 0.69 | 0.13 | 15.26 | 0.37 | 0.33 | 0.27 | 0.49 | 0.64 | Shared |
| S3_vs_127410 | 0.01 | 0 | 0.03 | 0.74 | 0.02 | 271.59 | 0.09 | 0.51 | 0.24 | 0.27 | 0.46 | 0.46 | 3.57 | 0.35 | 40.8 | 0.64 | 1.81 | 0.24 | 0.35 | 0.65 | Shared |
| S3_vs_127411 | 0 | 0 | 0 | 4.27 | 0 | 49.1 | 0.23 | 0.26 | 0.16 | 0.17 | 2.41 | 0.17 | 1.46 | 0.37 | 1.08 | 1.22 | 0.24 | 1.45 | 0.45 | 0.93 | ME |
| S3_vs_127413 | 0.08 | 0 | 0.06 | 4.07 | 0.14 | 1471.15 | 0.08 | 0.29 | 0.24 | 0.29 | 0.55 | 1.73 | 0.19 | 0.66 | 0.15 | 0.25 | 0.06 | 0.15 | 0.12 | 0.33 | ME |
| S3_vs_127421 | 0 | 0 | 0.03 | 1.32 | 0.24 | 159.6 | 0.28 | 0.58 | 0.21 | 0.18 | 35.77 | 0.35 | 0.75 | 0.31 | 0.77 | 0.33 | 0.57 | 0.35 | 0.46 | 0.88 | Shared |
| S3_vs_127442 | 0.26 | 0 | 0 | 1.11 | 0.04 | 189.54 | 0.23 | 0.36 | 0.19 | 0.14 | 0.53 | 0.22 | 0.26 | 0.25 | 0.45 | 0.37 | 0.26 | 0.3 | 0.16 | 0.67 | ME |
| S3_vs_127445 | 0.16 | 0.01 | 0.12 | 33.26 | 48.78 | 1089.34 | 0.22 | 0.65 | 0.68 | 0.43 | 1.19 | 1.4 | 1.05 | 0.44 | 1.03 | 33.16 | 0.49 | 0.6 | 0.74 | 1.22 | Shared |
| S3_vs_39940 | 0.03 | 0 | 0 | 0.77 | 0.06 | 46.73 | 0.01 | 0.15 | 0.09 | 0.19 | 0.49 | 0.19 | 0.39 | 0.1 | 0.2 | 0.25 | 13.33 | 0.16 | 0.22 | 0.34 | ME |
| S3_vs_119660 | 0 | 0 | 0.02 | 10.25 | 0.06 | 69.41 | 5.67 | 5.3 | 2.07 | 2.48 | 1.56 | 0.54 | 1.5 | 3.7 | 1.31 | 2.11 | 4.77 | 15.64 | 4.48 | 34.1 | Shared |
| S3_vs_119771 | 0 | 0 | 0.01 | 6.76 | 0.1 | 65.61 | 1.19 | 19.94 | 0.5 | 4.88 | 4.34 | 3.56 | 2.78 | 0.61 | 1.33 | 1.77 | 0.85 | 0.61 | 7.12 | 5.73 | ME |
| S3_vs_72118 | 0 | 0 | 0.01 | 0.59 | 0.07 | 42.82 | 0.02 | 0.36 | 0.16 | 0.54 | 0.58 | 0.14 | 0.24 | 2.04 | 0.43 | 0.27 | 0.35 | 0.2 | 0.27 | 0.65 | ME |
| S3_vs_56292 | 0 | 0 | 0 | 10.73 | 8.35 | 117.66 | 1.72 | 63.66 | 33.72 | 20.56 | 29.7 | 2.69 | 55.65 | 13.88 | 50.18 | 10.56 | 75.91 | 92.85 | 69.35 | 23.37 | Shared |
| S3_vs_9158 | 0.01 | 0.01 | 0.01 | 47.88 | 7.77 | 61.45 | 1.67 | 63.65 | 25.61 | 15.23 | 18.83 | 3.99 | 32.22 | 8.24 | 11.01 | 7.43 | 46.86 | 25.34 | 31.23 | 60.96 | Shared |
| S3_vs_64424 | 0.05 | 0 | 0 | 12.78 | 1.01 | 65.79 | 3.83 | 52.81 | 7.51 | 7.07 | 15.81 | 2.07 | 78.92 | 3.76 | 16.36 | 12.57 | 344.79 | 39.34 | 55.27 | 94.93 | Shared |
| S3_vs_1031 | 0 | 0 | 0 | 0.87 | 0.08 | 157.31 | 0 | 0.18 | 0.09 | 0.74 | 0.27 | 0.4 | 0.21 | 0.12 | 0.82 | 0.25 | 0.21 | 0.2 | 0.22 | 0.21 | ME |
| S3_vs_73361 | 0 | 0 | 0 | 4.31 | 0.02 | 26.35 | 0.09 | 0.8 | 0.12 | 0.12 | 5.55 | 0.09 | 1.4 | 0.08 | 1.53 | 4.08 | 0.25 | 0.24 | 1.21 | 0.38 | ME |
| S3_vs_1143 | 0 | 0 | 0.02 | 0.42 | 0.04 | 184.89 | 0.1 | 0.3 | 0.15 | 0.15 | 0.45 | 0.36 | 0.57 | 0.28 | 0.33 | 0.25 | 0.28 | 0.29 | 0.3 | 0.8 | ME |
| S3_vs_17168 | 0.02 | 0 | 0 | 6.09 | 0.59 | 52.98 | 11.4 | 10.8 | 4.88 | 7 | 26.21 | 1.66 | 2.12 | 8.28 | 12.29 | 2.63 | 1.33 | 39.84 | 9.42 | 5.22 | Shared |
| S3_vs_25896 | 0.01 | 0 | 0 | 1.17 | 0.12 | 32.3 | 0.16 | 17.71 | 0.32 | 1.28 | 0.9 | 2.83 | 1.03 | 0.97 | 0.64 | 1.25 | 13.7 | 0.61 | 1.86 | 1.23 | ME |
| S3_vs_97827 | 0 | 0 | 0.01 | 1.33 | 0.18 | 30.12 | 0.42 | 0.92 | 1.21 | 2.91 | 2.95 | 1.98 | 3.31 | 1.79 | 6.36 | 0.59 | 4.69 | 1.44 | 3.1 | 3.87 | ME |
| S3_vs_89898 | 0.37 | 0 | 0.42 | 2.02 | 0.61 | 5505.16 | 7 | 23.98 | 36.18 | 12.65 | 1.95 | 7.5 | 0.77 | 1.59 | 0.6 | 0.93 | 0.39 | 0.37 | 0.47 | 0.98 | Shared |
| S3_vs_50557 | 0.01 | 0 | 0.03 | 0.58 | 0.07 | 313.07 | 0.04 | 0.29 | 0.12 | 0.19 | 0.51 | 0.55 | 0.23 | 0.22 | 0.38 | 0.33 | 0.18 | 0.23 | 0.27 | 0.66 | ME |
| S3_vs_42569 | 0.01 | 0 | 0.01 | 1.63 | 0.1 | 93.03 | 7.81 | 5.43 | 1.21 | 6.11 | 3.49 | 1.31 | 1.58 | 0.77 | 1.91 | 1.4 | 2.98 | 1.52 | 2.43 | 12.19 | ME |
| S3_vs_114271 | 0.01 | 0 | 0 | 0.55 | 0.01 | 25.51 | 0.31 | 2.46 | 0.09 | 0.12 | 1.8 | 0.12 | 0.22 | 0.16 | 0.62 | 0.45 | 0.33 | 0.17 | 0.23 | 0.32 | ME |
| S3_vs_51005 | 0.01 | 0 | 0 | 1.07 | 0 | 65.28 | 0.01 | 0.08 | 0.15 | 0.06 | 0.08 | 0.05 | 0.05 | 0.01 | 0.48 | 0.47 | 0.03 | 0 | 2.37 | 1.12 | ME |
| S3_vs_51931 | 0 | 0 | 0 | 0.5 | 0.11 | 19.85 | 0 | 0.23 | 0.01 | 0.34 | 0.1 | 0.26 | 0.08 | 0.02 | 0.08 | 0.15 | 2.6 | 0.04 | 0.1 | 0.06 | ME |
| S3_vs_122706 | 0 | 0 | 0.01 | 6.91 | 0.09 | 64.11 | 0.04 | 0.23 | 0.08 | 0.19 | 0.52 | 0.16 | 0.43 | 0.6 | 1.52 | 0.45 | 0.36 | 0.27 | 1.11 | 1.34 | ME |
| S3_vs_12973 | 0 | 0 | 0 | 1.44 | 0.03 | 27.16 | 0.08 | 0.44 | 0.48 | 0.17 | 1.32 | 0.17 | 0.57 | 1.4 | 0.42 | 0.64 | 0.34 | 0.64 | 0.45 | 1.1 | ME |
| S3_vs_12985 | 0.01 | 0 | 0 | 1.55 | 0.02 | 17.97 | 1.3 | 4.61 | 0.17 | 1.13 | 2.1 | 0.12 | 0.1 | 6.11 | 1.09 | 0.38 | 1.12 | 0.68 | 0.19 | 0.5 | ME |
| S3_vs_18858 | 0.01 | 0 | 0 | 1.7 | 0.06 | 29.55 | 0.07 | 0.3 | 0.16 | 0.19 | 0.49 | 0.21 | 0.37 | 0.19 | 0.4 | 0.64 | 0.27 | 7.49 | 1.24 | 0.97 | ME |
| S3_vs_77156 | 0 | 0 | 0.05 | 0.82 | 0.01 | 19.98 | 0.03 | 0.15 | 0.32 | 0.13 | 0.29 | 0.12 | 0.57 | 0.07 | 0.42 | 0.04 | 0.38 | 0.34 | 1.66 | 0.51 | ME |
| S3_vs_123834 | 0.01 | 0 | 0.02 | 11.31 | 0.02 | 44.86 | 0.02 | 0.16 | 0.09 | 0.09 | 1.14 | 0.09 | 0.15 | 0.11 | 0.24 | 0.45 | 0.11 | 0.15 | 11.21 | 1.89 | Shared |
| S3_vs_20315 | 0 | 0 | 0.6 | 1.38 | 0.75 | 6057.75 | 1.87 | 10.91 | 0.87 | 1.17 | 2.89 | 8.33 | 19.52 | 0.24 | 1.68 | 2.78 | 0.85 | 20.4 | 2.97 | 3.68 | Shared |
| S3_vs_68031 | 0.03 | 0 | 0.02 | 2.82 | 0.14 | 127.71 | 4.81 | 123.54 | 1.48 | 58.46 | 4.4 | 1.31 | 4.06 | 1.83 | 2.97 | 2.08 | 2.29 | 5.2 | 7.04 | 14.1 | ME |
| S3_vs_29077 | 0 | 0 | 0 | 1.05 | 0.02 | 42.81 | 0.22 | 0.29 | 0.22 | 0.5 | 0.51 | 0.19 | 1.29 | 0.49 | 0.4 | 0.41 | 0.38 | 0.28 | 0.34 | 0.84 | ME |
| S3_vs_45147 | 0.06 | 0 | 0.05 | 0.54 | 0.19 | 564.44 | 0.1 | 0.27 | 0.18 | 0.15 | 0.5 | 4.73 | 0.41 | 0.75 | 0.29 | 0.32 | 0.41 | 5.83 | 0.47 | 0.7 | ME |
| S3_vs_62410 | 0 | 0 | 0 | 0.78 | 0.03 | 35.98 | 0.05 | 0.37 | 0.14 | 4.18 | 0.64 | 0.16 | 0.9 | 0.57 | 0.56 | 0.45 | 0.44 | 0.31 | 0.72 | 0.65 | ME |
| S3_vs_100785 | 0 | 0 | 0 | 0.83 | 0.11 | 52.51 | 0.11 | 1.83 | 0.19 | 0.18 | 0.84 | 0.27 | 0.51 | 1.83 | 0.39 | 0.22 | 0.38 | 25.16 | 0.43 | 0.92 | Shared |
| S3_vs_15450 | 0.01 | 0 | 0 | 0.62 | 0.02 | 39.93 | 0.08 | 0.3 | 0.08 | 0.14 | 4.53 | 0.12 | 0.27 | 0.23 | 0.41 | 0.55 | 0.52 | 0.16 | 0.25 | 0.52 | ME |
| S3_vs_54303 | 0.02 | 0 | 0.02 | 0.79 | 2.06 | 45.96 | 1.21 | 14.89 | 0.19 | 2.67 | 2.18 | 0.14 | 1.12 | 34.19 | 21.49 | 2.8 | 0.74 | 0.35 | 0.79 | 0.39 | Shared |
| S3_vs_30318 | 0 | 0 | 0.01 | 0.86 | 0.03 | 23.8 | 0.14 | 0.33 | 0.18 | 0.2 | 0.64 | 0.11 | 1.14 | 0.49 | 0.5 | 0.47 | 0.48 | 0.36 | 0.52 | 1.16 | ME |
| S3_vs_94577 | 0.01 | 0 | 0.03 | 1.13 | 0.13 | 202.06 | 0.81 | 0.57 | 0.72 | 0.38 | 1.8 | 0.37 | 3.49 | 0.5 | 2.89 | 0.93 | 0.65 | 1.35 | 1.52 | 16.91 | ME |
| S3_vs_6604 | 0 | 0 | 0.01 | 3.2 | 0.45 | 58.42 | 0.61 | 0.95 | 1 | 1.47 | 11.34 | 0.63 | 2.86 | 1.27 | 9.48 | 0.55 | 1.36 | 3.18 | 6.96 | 11.03 | Shared |
| S3_vs_70093 | 0.01 | 0 | 0 | 5.76 | 0.07 | 18.2 | 0.44 | 0.2 | 0.2 | 91.85 | 0.65 | 0.79 | 1.15 | 5.87 | 0.53 | 1.48 | 9.91 | 0.09 | 5.34 | 0.73 | ME |
| S3_vs_70508 | 0.02 | 0 | 0 | 34.8 | 26.74 | 51.55 | 0.27 | 26.23 | 9.87 | 53.89 | 203.4 | 1.51 | 5.93 | 2.99 | 7.35 | 10.54 | 18.13 | 6.37 | 9 | 35.79 | Shared |
| S3_vs_39774 | 0.02 | 0.01 | 0.01 | 7.75 | 1.11 | 53.34 | 0.12 | 55.68 | 2.38 | 8.51 | 17.42 | 0.36 | 259.82 | 8 | 62.67 | 42.86 | 85.59 | 211.1 | 112.13 | 13.66 | Shared |
| S3_vs_95574 | 0 | 0 | 0.03 | 0.43 | 0.03 | 18.92 | 0.07 | 0.42 | 0.13 | 0.15 | 1.36 | 0.11 | 1.39 | 0.71 | 0.34 | 0.27 | 0.27 | 0.28 | 0.36 | 0.66 | ME |
| S3_vs_23194 | 0.01 | 0 | 0 | 1.44 | 0.07 | 48.79 | 1.49 | 0.55 | 0.17 | 0.25 | 1.04 | 17.78 | 1.16 | 0.48 | 0.49 | 0.76 | 22.69 | 0.38 | 0.81 | 3.1 | ME |
| S3_vs_127387 | 0 | 0 | 0 | 0.87 | 0.38 | 63.38 | 0.48 | 0.51 | 0.28 | 1.19 | 0.9 | 0.22 | 1.73 | 19.43 | 0.95 | 1.42 | 0.66 | 0.58 | 1.14 | 1.29 | ME |
| S3_vs_87728 | 0.02 | 0 | 0.02 | 4.26 | 0.09 | 72.84 | 13.48 | 2.75 | 4.09 | 7.92 | 3.56 | 1.54 | 1.3 | 1.27 | 3.5 | 1.65 | 14.9 | 1.92 | 5.67 | 13.1 | Shared |
| S3_vs_40939 | 0 | 0.01 | 0.01 | 2.41 | 0.82 | 39.61 | 0.2 | 10.82 | 5.95 | 8.18 | 3.12 | 0.31 | 3.09 | 2.07 | 2.02 | 1.46 | 2.23 | 4.6 | 7.75 | 15.52 | ME |
| S3_vs_117894 | 0 | 0 | 0.01 | 2.22 | 0.41 | 44.48 | 0.81 | 0.97 | 1.3 | 8.12 | 14.12 | 3.33 | 5.16 | 1.2 | 6.63 | 0.88 | 7.08 | 1.39 | 3.6 | 7.53 | Shared |
| S5_vf_18461 | 0 | 0 | 0 | 0.6 | 0.04 | 0.42 | 0.28 | 12.44 | 0.48 | 0.18 | 0.63 | 0.1 | 13.67 | 0.43 | 7.91 | 0.17 | 0.62 | 0.19 | 0.28 | 0.53 | Shared |
| S5_vf_6506 | 0 | 0 | 0 | 0.64 | 0.01 | 4.79 | 0.7 | 35.39 | 0.25 | 0.21 | 0.81 | 0.13 | 1.08 | 0.58 | 0.91 | 0.43 | 1.12 | 0.34 | 0.56 | 0.84 | ME |
| S5_vf_122568 | 0.01 | 0 | 0 | 3.29 | 0.36 | 4.87 | 2.77 | 799.91 | 0.39 | 0.35 | 22.35 | 2.33 | 1.58 | 0.36 | 16.29 | 1.78 | 20.3 | 2.47 | 0.68 | 2.92 | Shared |
| S5_vf_134040 | 0.02 | 0.01 | 0 | 9.52 | 0.31 | 58.41 | 6.08 | 73.16 | 3.28 | 26.43 | 22.97 | 3.32 | 12.65 | 8.22 | 19.21 | 9 | 8.7 | 9.59 | 11.86 | 27.02 | Shared |
| S5_vf_116300 | 0 | 0 | 0 | 4.28 | 0.32 | 0.71 | 0.33 | 40.86 | 1.36 | 0.43 | 1.63 | 1.32 | 0.97 | 0.96 | 0.7 | 2.13 | 1.93 | 0.38 | 0.8 | 2.67 | ME |
| S5_vf_67622 | 0 | 0 | 0 | 0.5 | 0.08 | 0.74 | 0.14 | 66.98 | 128.64 | 0.44 | 23.84 | 0.1 | 1.13 | 0.56 | 1.87 | 0.5 | 0.43 | 0.33 | 0.48 | 0.91 | Shared |
| S5_vf_110171 | 0 | 0 | 0 | 0.57 | 0.03 | 0.33 | 0.37 | 21.83 | 0.09 | 0.42 | 0.84 | 0.06 | 4.38 | 0.18 | 4.92 | 1.04 | 1.76 | 0.25 | 0.51 | 0.45 | ME |
| S5_vf_55415 | 0 | 0 | 0 | 0.76 | 0.39 | 2.29 | 0.6 | 1420.76 | 1514.05 | 0.83 | 8.83 | 0.07 | 2.58 | 2.23 | 0.73 | 0.59 | 0.31 | 0.3 | 0.38 | 0.85 | Shared |
| S5_vf_38011 | 0.01 | 0 | 0 | 3.22 | 0.08 | 0.26 | 0.01 | 287.06 | 251.71 | 0.51 | 0.28 | 0.04 | 0.72 | 0.82 | 0.08 | 1.38 | 0.13 | 0.15 | 0.18 | 1.03 | Shared |
| S5_vf_55687 | 0 | 0 | 0.11 | 0.44 | 0.26 | 1016.37 | 0.05 | 348.36 | 17.73 | 246.4 | 10.05 | 1.34 | 0.72 | 0.72 | 0.63 | 0.3 | 1.99 | 0.29 | 0.32 | 12.3 | Shared |
| S5_vf_25423 | 0.37 | 0 | 0 | 0.37 | 4.78 | 180.73 | 242.15 | 295.25 | 8.83 | 18.34 | 48.76 | 41.97 | 2.87 | 2.88 | 2.23 | 1.13 | 6860.37 | 0.93 | 16.58 | 951.76 | Shared |
| S5_vf_123990 | 0 | 0 | 0 | 0.53 | 6.37 | 0.29 | 0.05 | 56.93 | 82.49 | 0.14 | 0.68 | 0.03 | 0.47 | 0.31 | 0.49 | 0.41 | 0.26 | 0.17 | 0.23 | 0.65 | Shared |
| S5_vf_105728 | 0 | 0 | 0 | 0.63 | 0.03 | 0.33 | 0.06 | 13.73 | 0.44 | 0.39 | 1.57 | 0.21 | 0.48 | 65.11 | 0.36 | 0.19 | 4.26 | 0.22 | 0.28 | 0.52 | ME |
| S5_vf_8808 | 0 | 0 | 0 | 0.18 | 0.02 | 0.36 | 0.14 | 14.19 | 0.19 | 0.16 | 0.47 | 0.08 | 0.3 | 0.66 | 0.46 | 0.31 | 4.2 | 0.18 | 0.3 | 0.7 | ME |
| S5_vf_8965 | 0 | 0 | 0 | 3.66 | 1.86 | 3.41 | 0.57 | 51.24 | 0.63 | 1.85 | 4.59 | 38.56 | 4.02 | 0.98 | 6.39 | 0.65 | 4.6 | 26.38 | 4.38 | 2.54 | ME |
| S5_vf_74661 | 0.02 | 0 | 0 | 0.45 | 0.04 | 0.3 | 0.02 | 108.37 | 0.08 | 0.11 | 0.54 | 4.51 | 0.37 | 0.13 | 0.6 | 0.17 | 0.17 | 0.2 | 0.23 | 0.5 | ME |

| | | | | | | | | | | | | | | | | | | | | | |
|---|---|---|---|---|---|---|---|---|---|---|---|---|---|---|---|---|---|---|---|---|---|
| S5_vf_136076 | 0 | 0 | 0 | 0.65 | 0.03 | 0.48 | 2.4 | 19.19 | 36.18 | 0.24 | 1.74 | 0.1 | 1.32 | 0.87 | 0.36 | 0.29 | 0.49 | 0.31 | 0.33 | 0.48 | Shared |
| S5_vf_21869 | 0 | 0 | 0 | 14.42 | 7.58 | 0.73 | 0.23 | 63.83 | 102.34 | 2.93 | 2.21 | 0.53 | 1.13 | 82.02 | 70.77 | 4.04 | 2.8 | 0.24 | 2.04 | 0.74 | Shared |
| S5_vf_6032 | 0 | 0 | 0 | 0.32 | 0.03 | 0.12 | 0.08 | 64.45 | 1.92 | 1.93 | 6.09 | 0.02 | 0.19 | 0.03 | 0.04 | 0.3 | 8.46 | 0.08 | 0.21 | 3.21 | ME |
| S5_vf_137260 | 0 | 0 | 0 | 0.51 | 0.04 | 0.25 | 0.01 | 49.37 | 52.39 | 0.09 | 0.27 | 0.04 | 0.21 | 0.15 | 0.24 | 0.32 | 0.12 | 0.16 | 0.27 | 0.34 | Shared |
| S5_vf_11450 | 0.01 | 0 | 0 | 0.47 | 0.19 | 0.29 | 0.02 | 83.61 | 0.09 | 0.23 | 0.49 | 0.06 | 0.37 | 0.15 | 0.66 | 0.2 | 0.15 | 0.13 | 0.44 | 0.41 | ME |
| S5_vf_28061 | 0 | 0 | 0 | 0.52 | 0.06 | 1.33 | 0.75 | 370.71 | 0.21 | 0.52 | 0.55 | 0.16 | 1 | 0.63 | 0.68 | 0.33 | 10.64 | 0.27 | 0.55 | 0.66 | ME |
| S5_vf_77701 | 0 | 0 | 0 | 0.46 | 0.01 | 0.6 | 0.01 | 29.26 | 0.12 | 0.06 | 0.26 | 0.02 | 0.3 | 0.21 | 0.28 | 0.19 | 0.19 | 0.23 | 0.18 | 0.45 | ME |
| S5_vf_145579 | 0 | 0 | 0 | 0.85 | 0.32 | 1.2 | 0.18 | 547.38 | 1778.42 | 0.54 | 1.28 | 0.09 | 1.75 | 2.75 | 1.99 | 0.62 | 0.4 | 0.36 | 0.46 | 0.86 | Shared |
| S5_vf_145583 | 0 | 0 | 0 | 0.55 | 0.04 | 0.81 | 0.46 | 198.81 | 0.18 | 0.24 | 0.61 | 0.12 | 1.79 | 0.28 | 0.63 | 0.56 | 2.08 | 0.38 | 0.8 | 1.06 | ME |
| S5_vf_29332 | 0.02 | 0 | 0 | 0.94 | 0.03 | 0.44 | 0.07 | 40.01 | 0.12 | 0.18 | 0.53 | 0.07 | 0.36 | 0.08 | 0.36 | 0.31 | 0.27 | 0.29 | 0.32 | 0.42 | ME |
| S5_vf_145585 | 0 | 0 | 0 | 0.46 | 0.02 | 0.38 | 0.09 | 44.5 | 96.5 | 0.14 | 0.37 | 0.08 | 0.53 | 0.36 | 0.26 | 0.23 | 0.25 | 0.27 | 0.28 | 0.65 | Shared |
| S5_vf_145586 | 0 | 0 | 0 | 0.58 | 0.08 | 0.34 | 0.08 | 114.11 | 0.18 | 0.12 | 0.41 | 0.29 | 0.85 | 0.24 | 0.67 | 0.23 | 3.34 | 0.22 | 0.29 | 0.48 | ME |
| S5_vf_145588 | 0 | 0 | 0 | 1.55 | 0.28 | 1.41 | 0.19 | 1139.5 | 1002.38 | 0.76 | 1.29 | 0.16 | 3.13 | 1.89 | 0.65 | 0.79 | 0.67 | 0.46 | 0.61 | 1.21 | Shared |
| S5_vf_145604 | 0 | 0 | 0 | 1.42 | 0.08 | 1.07 | 14.96 | 534.39 | 0.25 | 0.27 | 0.87 | 0.08 | 1.31 | 0.88 | 0.62 | 0.39 | 27.22 | 2.28 | 8.02 | 16.6 | Shared |
| S5_vf_103206 | 0 | 0 | 0 | 3.84 | 0.09 | 3.51 | 0.58 | 45.06 | 0.61 | 4.05 | 12.03 | 2.1 | 2.36 | 32.08 | 7.76 | 6.71 | 20.63 | 1.21 | 3.58 | 4.39 | Shared |
| S5_vf_60597 | 0 | 0 | 0 | 4.87 | 0.05 | 1.85 | 0.05 | 32.45 | 57.81 | 0.18 | 5.95 | 1.37 | 0.72 | 0.44 | 0.62 | 1.57 | 0.36 | 0.32 | 1.9 | 25.36 | Shared |
| S5_vs_36605 | 0 | 0 | 0 | 0.67 | 0.17 | 1.18 | 0.07 | 362.28 | 0.61 | 1 | 0.5 | 0.09 | 0.7 | 0.09 | 0.17 | 0.16 | 34.44 | 0.14 | 0.35 | 0.48 | ME |
| S5_vs_122344 | 0.02 | 0 | 0 | 3.62 | 0.91 | 47.06 | 0.06 | 166.98 | 5.86 | 0.21 | 1.68 | 0.22 | 0.69 | 0.54 | 0.41 | 0.62 | 24.45 | 0.29 | 0.77 | 1.1 | ME |
| S5_vs_122618 | 0 | 0 | 0 | 0.57 | 0.15 | 0.72 | 0.1 | 794.79 | 0.44 | 2.61 | 0.67 | 0.92 | 1.65 | 0.65 | 0.56 | 0.32 | 0.42 | 0.35 | 0.79 | 1.18 | ME |
| S5_vs_97574 | 0.03 | 0 | 0 | 1.51 | 0.32 | 5.97 | 0.12 | 500.54 | 0.34 | 0.42 | 15.05 | 0.25 | 5.12 | 21.42 | 0.82 | 2.14 | 377.05 | 3.55 | 1.01 | 1.64 | Shared |
| S5_vs_37659 | 0 | 0 | 0 | 0.56 | 0.02 | 0.37 | 0.11 | 93.17 | 0.12 | 0.17 | 0.4 | 1.31 | 0.61 | 0.38 | 0.35 | 0.14 | 5.92 | 0.17 | 0.2 | 0.36 | ME |
| S5_vs_67958 | 0 | 0 | 0 | 2.62 | 0.02 | 0.64 | 0.11 | 68.15 | 0.15 | 0.6 | 0.39 | 0.08 | 0.4 | 0.23 | 0.32 | 1.28 | 14.81 | 0.19 | 0.35 | 0.57 | ME |
| S5_vs_68254 | 0.01 | 0 | 0 | 0.85 | 0.13 | 0.61 | 0.54 | 548.47 | 0.29 | 1.03 | 0.77 | 0.13 | 1.32 | 0.8 | 0.47 | 0.35 | 0.82 | 0.43 | 0.38 | 0.86 | ME |
| S5_vs_38949 | 0 | 0 | 0 | 12.57 | 1.37 | 0.33 | 0.1 | 292.43 | 296.52 | 21.7 | 1.69 | 0.11 | 2.99 | 1.07 | 0.55 | 5.43 | 9.95 | 0.21 | 5.12 | 11.58 | Shared |
| S5_vs_120079 | 0.02 | 0 | 0 | 8.37 | 3.93 | 6.41 | 0.13 | 160.73 | 0.87 | 0.5 | 1.83 | 0.19 | 0.91 | 0.42 | 0.45 | 0.95 | 100.46 | 0.58 | 0.42 | 1.01 | ME |
| S5_vs_10428 | 0.01 | 0 | 0 | 0.41 | 0.02 | 0.21 | 0.16 | 45.76 | 0.11 | 0.23 | 0.94 | 0.05 | 0.37 | 0.3 | 0.51 | 0.28 | 1.23 | 0.2 | 0.5 | 0.46 | ME |
| S5_vs_136679 | 0 | 0 | 0 | 0.57 | 0.03 | 0.38 | 0.08 | 18.81 | 0.16 | 0.19 | 0.33 | 0.18 | 0.42 | 0.28 | 0.37 | 0.2 | 0.4 | 0.29 | 0.32 | 1.18 | ME |
| S5_vs_120584 | 0 | 0 | 0 | 8.2 | 0.02 | 2.5 | 0.19 | 44.75 | 0.28 | 4.07 | 33.52 | 0.18 | 2.54 | 0.42 | 1.47 | 1.24 | 0.75 | 2.87 | 1.92 | 2.71 | Shared |
| S5_vs_109085 | 0.01 | 0 | 0 | 0.93 | 0.09 | 0.5 | 0.13 | 879.44 | 0.12 | 0.31 | 1.01 | 0.18 | 1.32 | 0.65 | 0.28 | 0.27 | 7.23 | 0.21 | 0.36 | 2.67 | ME |
| S5_vs_121411 | 0 | 0 | 0 | 1.32 | 0.03 | 0.73 | 0.06 | 114.52 | 0.38 | 0.2 | 0.62 | 0.52 | 0.55 | 0.41 | 0.37 | 0.39 | 29.1 | 0.25 | 0.32 | 0.79 | ME |
| S5_vs_121419 | 0 | 0 | 0 | 0.69 | 0.04 | 0.75 | 0.13 | 95.41 | 0.14 | 0.24 | 0.89 | 0.1 | 0.38 | 0.52 | 0.55 | 0.23 | 15.44 | 0.34 | 0.32 | 0.81 | ME |
| S5_vs_27918 | 0 | 0 | 0 | 0.52 | 0.04 | 0.29 | 0.04 | 37.52 | 0.13 | 0.35 | 0.23 | 0.08 | 0.25 | 0.84 | 0.81 | 0.19 | 0.16 | 0.37 | 0.22 | 0.44 | ME |
| S5_vs_145578 | 0.01 | 0 | 0 | 0.36 | 0.07 | 0.42 | 0.04 | 79.22 | 0.3 | 0.94 | 1.16 | 0.59 | 0.65 | 6.03 | 0.25 | 0.32 | 18.72 | 0.23 | 0.46 | 1.4 | ME |
| S5_vs_145581 | 0 | 0 | 0 | 0.9 | 0.01 | 0.51 | 0.11 | 55 | 0.14 | 0.1 | 0.53 | 0.08 | 0.42 | 0.29 | 0.4 | 0.34 | 5.47 | 0.38 | 0.36 | 0.51 | ME |
| S5_vs_145582 | 0 | 0 | 0 | 1.12 | 0.04 | 0.55 | 0.08 | 47.59 | 0.24 | 0.86 | 1.17 | 0.19 | 0.6 | 0.33 | 0.51 | 1.31 | 0.62 | 0.39 | 0.5 | 1.16 | ME |
| S5_vs_145590 | 0 | 0 | 0 | 0.94 | 0.03 | 0.39 | 0.06 | 31.05 | 0.12 | 0.32 | 0.42 | 0.16 | 0.89 | 0.17 | 0.29 | 0.31 | 0.87 | 0.21 | 0.46 | 0.66 | ME |
| S5_vs_145591 | 0 | 0 | 0 | 0.54 | 0.03 | 0.23 | 0.06 | 22.59 | 0.13 | 2.22 | 0.45 | 0.07 | 0.37 | 1.17 | 0.28 | 0.19 | 21.42 | 0.2 | 0.19 | 0.44 | ME |
| S5_vs_145592 | 0 | 0 | 0 | 0.94 | 0.02 | 0.81 | 0.13 | 57.99 | 0.24 | 0.53 | 0.36 | 0.16 | 0.43 | 0.19 | 1.2 | 0.2 | 0.29 | 0.39 | 0.3 | 0.43 | ME |
| S5_vs_145593 | 0 | 0 | 0 | 1.23 | 0.03 | 0.33 | 0.07 | 154.2 | 0.05 | 0.34 | 0.39 | 0.05 | 0.8 | 0.18 | 0.36 | 1.6 | 0.51 | 0.17 | 0.9 | 0.79 | ME |
| S5_vs_145597 | 0 | 0 | 0 | 0.82 | 0.01 | 0.21 | 0.01 | 40.04 | 0.4 | 0.18 | 0.59 | 0.04 | 0.28 | 0.4 | 0.19 | 0.18 | 2.72 | 0.28 | 0.2 | 0.36 | ME |
| S5_vs_145598 | 0 | 0 | 0 | 0.72 | 0.16 | 0.49 | 0.25 | 31.45 | 0.19 | 0.2 | 5.81 | 0.16 | 0.66 | 0.23 | 0.45 | 0.25 | 0.46 | 0.4 | 0.55 | 1.24 | ME |
| S5_vs_145599 | 0 | 0 | 0 | 0.83 | 0.03 | 6.36 | 0.09 | 142.64 | 0.54 | 0.73 | 3.65 | 0.34 | 4.07 | 0.84 | 6.02 | 1.55 | 1.33 | 0.58 | 4.29 | 10.91 | ME |
| S5_vs_145601 | 0 | 0 | 0 | 0.75 | 0.02 | 0.61 | 0.05 | 68.03 | 0.6 | 0.17 | 0.89 | 0.16 | 0.85 | 0.2 | 0.26 | 0.37 | 9.46 | 0.2 | 0.42 | 0.74 | ME |
| S5_vs_145602 | 0.01 | 0 | 0 | 0.87 | 0.06 | 0.7 | 0.09 | 41.27 | 0.18 | 0.47 | 0.5 | 0.15 | 0.84 | 0.18 | 0.24 | 0.25 | 0.43 | 0.42 | 0.52 | 0.72 | ME |
| S5_vs_145605 | 0.01 | 0 | 0 | 0.9 | 0.01 | 0.54 | 0.01 | 25.19 | 0.19 | 0.8 | 0.91 | 0.04 | 0.24 | 0.97 | 0.64 | 1.46 | 0.8 | 0.25 | 0.34 | 0.33 | ME |
| S5_vs_145611 | 0 | 0 | 0 | 0.94 | 0.07 | 1.52 | 0.06 | 57.07 | 0.09 | 0.2 | 0.97 | 0.32 | 0.59 | 0.54 | 0.34 | 0.29 | 0.27 | 0.21 | 0.32 | 0.69 | ME |
| S5_vs_145633 | 0 | 0 | 0 | 1.8 | 0.02 | 0.23 | 0.01 | 30.23 | 0.1 | 0.16 | 1.02 | 0.06 | 0.1 | 0.25 | 0.37 | 0.25 | 0.11 | 0.2 | 0.28 | 0.79 | ME |
| S5_vs_85065 | 0 | 0 | 0 | 0.75 | 14.13 | 2.06 | 0.08 | 100.97 | 0.91 | 0.49 | 3.27 | 0.23 | 0.77 | 11.37 | 0.46 | 0.44 | 12.24 | 1.01 | 0.56 | 0.79 | Shared |
| S5_vs_30490 | 0.01 | 0 | 0.01 | 18.47 | 0.29 | 8.04 | 0.8 | 152.73 | 4.93 | 12.77 | 2.37 | 0.54 | 4.27 | 0.9 | 1.5 | 1.41 | 0.8 | 7.66 | 2.45 | 5.88 | ME |
| S5_vs_6478 | 0.01 | 0 | 0 | 0.49 | 0.03 | 0.58 | 0.2 | 54.02 | 0.12 | 0.16 | 0.64 | 0.11 | 0.96 | 0.25 | 0.57 | 0.46 | 0.37 | 0.26 | 0.43 | 0.99 | ME |
| S5_vs_109527 | 0 | 0 | 0 | 1.69 | 0.9 | 10.06 | 1.11 | 83.64 | 14.78 | 8.28 | 5.68 | 0.43 | 18.73 | 2.68 | 22.19 | 2.45 | 16.96 | 7.54 | 8.71 | 8.02 | Shared |
| S5_vs_49011 | 0.01 | 0 | 0 | 0.45 | 0.02 | 0.44 | 0.08 | 34.49 | 0.11 | 0.15 | 0.49 | 0.99 | 0.63 | 0.17 | 0.33 | 0.34 | 1.38 | 0.23 | 0.36 | 0.68 | ME |
| S5_vs_43207 | 0 | 0 | 0 | 0.84 | 0.02 | 0.3 | 0.08 | 15.39 | 0.14 | 0.6 | 0.51 | 0.26 | 0.24 | 0.37 | 0.33 | 0.28 | 0.73 | 0.24 | 0.32 | 0.63 | ME |
| S5_vs_850 | 0 | 0 | 0 | 0.74 | 0.07 | 0.71 | 0.07 | 33.11 | 0.23 | 0.36 | 7.26 | 0.12 | 1.36 | 0.25 | 1.1 | 0.44 | 0.74 | 0.45 | 0.7 | 1.32 | ME |
| S5_vs_116235 | 0 | 0 | 0 | 8.66 | 0.01 | 0.41 | 0.03 | 11.4 | 0.12 | 0.14 | 0.33 | 0.08 | 0.24 | 0.17 | 0.46 | 3.2 | 0.16 | 0.2 | 0.26 | 0.59 | ME |
| S5_vs_1338 | 0 | 0 | 0 | 1.97 | 0.02 | 0.69 | 0.05 | 16.14 | 8.72 | 0.11 | 0.47 | 0.07 | 0.67 | 1.29 | 0.94 | 2.07 | 1.32 | 0.64 | 0.58 | 0.65 | ME |
| S5_vs_49480 | 0.01 | 0 | 0 | 1.31 | 0.01 | 0.31 | 0.08 | 22.25 | 0.11 | 0.27 | 31.42 | 0.1 | 0.31 | 0.65 | 6.04 | 0.5 | 0.37 | 0.25 | 0.87 | 0.6 | Shared |
| S5_vs_13090 | 0 | 0 | 0 | 0.72 | 0.02 | 0.73 | 0.09 | 14.11 | 0.16 | 0.35 | 1.42 | 0.44 | 0.51 | 0.56 | 0.34 | 0.22 | 2.74 | 0.24 | 0.38 | 0.72 | ME |
| S5_vs_128398 | 0 | 0 | 0 | 0.99 | 0.09 | 0.31 | 0.06 | 83.21 | 8.29 | 0.2 | 0.57 | 0.27 | 0.37 | 0.26 | 0.34 | 0.33 | 53.96 | 0.19 | 0.28 | 0.58 | ME |
| S5_vs_123256 | 0.03 | 0 | 0 | 9.72 | 0.62 | 24.11 | 0.83 | 34.58 | 12.8 | 7.1 | 27.1 | 1.33 | 3.86 | 8.11 | 15.07 | 3.53 | 4.2 | 28.21 | 12.85 | 7.46 | Shared |
| S5_vs_13254 | 0 | 0 | 0 | 0.66 | 0.08 | 0.4 | 0.04 | 39.52 | 0.17 | 0.67 | 0.73 | 0.08 | 1.03 | 1.21 | 0.36 | 0.42 | 12.8 | 0.23 | 0.34 | 0.78 | ME |
| S5_vs_1572 | 0 | 0.01 | 0 | 18.52 | 13.01 | 2.23 | 1 | 36.73 | 57.93 | 206.69 | 1.94 | 0.06 | 2.15 | 21.69 | 0.56 | 23.45 | 0.25 | 0.19 | 35.92 | 0.54 | Shared |
| S5_vs_117319 | 0.02 | 0 | 0 | 3.05 | 0.26 | 32.91 | 4.06 | 54.3 | 2.19 | 19.28 | 5.22 | 1.79 | 4 | 1.39 | 4.98 | 1.46 | 6.29 | 4.16 | 5.91 | 19.76 | ME |
| S5_vs_117417 | 0 | 0 | 0 | 1.08 | 0.07 | 1.99 | 0.12 | 35.92 | 4.46 | 6.39 | 0.79 | 0.12 | 0.43 | 0.42 | 0.45 | 1.09 | 11 | 0.27 | 1.3 | 1.92 | ME |
| S5_vs_128881 | 0.01 | 0 | 0 | 3.15 | 0.24 | 1.57 | 1.56 | 58.06 | 0.26 | 1.35 | 2.49 | 0.81 | 0.77 | 23.95 | 0.63 | 0.9 | 34.71 | 0.77 | 26.82 | 3.66 | Shared |
| S5_vs_86562 | 0 | 0 | 0 | 0.89 | 0.1 | 1.49 | 0.08 | 192.6 | 0.23 | 1.08 | 1.76 | 0.13 | 0.52 | 0.17 | 0.28 | 0.29 | 4.14 | 0.62 | 1.2 | 0.61 | ME |
| S5_vs_140946 | 0 | 0.02 | 0 | 110.82 | 0.08 | 2.18 | 0.65 | 48.22 | 1.86 | 0.6 | 2.11 | 0.13 | 0.63 | 0.46 | 0.21 | 32.87 | 45.64 | 1.99 | 4.4 | 0.7 | ME |
| S5_vs_8125 | 0.01 | 0 | 0.01 | 2.1 | 0.08 | 5.45 | 0.54 | 24.18 | 0.19 | 0.24 | 0.77 | 0.28 | 1.73 | 0.44 | 0.57 | 0.53 | 22.93 | 0.76 | 1.06 | 4.19 | ME |
| S5_vs_124035 | 0.02 | 0 | 0.06 | 5.82 | 0.25 | 9.71 | 1.9 | 36.28 | 1.73 | 12.4 | 25.64 | 4.58 | 4.32 | 14.02 | 7.9 | 23.18 | 20.09 | 2.58 | 5.13 | 10.3 | Shared |
| S5_vs_20268 | 0 | 0 | 0 | 0.76 | 0.17 | 0.39 | 0.11 | 81.81 | 7.57 | 1.26 | 0.41 | 0.03 | 0.68 | 0.23 | 6.81 | 0.36 | 9.03 | 0.11 | 0.46 | 0.43 | ME |
| S5_vs_79992 | 0.02 | 0 | 0 | 9.53 | 0.94 | 14.65 | 0.54 | 51.76 | 13.56 | 13.55 | 28.24 | 2.6 | 3.36 | 5.55 | 17.17 | 2.37 | 6.79 | 22.05 | 14.6 | 7.44 | Shared |
| S5_vs_118168 | 0 | 0 | 0 | 0.63 | 0.03 | 0.27 | 0.13 | 52.6 | 0.18 | 0.32 | 0.33 | 0.68 | 0.36 | 0.58 | 0.22 | 0.37 | 4.91 | 0.16 | 0.25 | 0.34 | ME |
| S5_vs_32885 | 0 | 0 | 0 | 0.46 | 0.17 | 0.95 | 0.1 | 93.15 | 0.1 | 0.2 | 0.49 | 0.06 | 0.36 | 0.2 | 0.6 | 0.2 | 3.64 | 0.2 | 0.36 | 0.7 | ME |
| S5_vs_2876 | 0 | 0 | 0 | 1.16 | 0.03 | 0.19 | 0.12 | 106.91 | 0.18 | 0.14 | 0.51 | 0.07 | 0.38 | 0.52 | 0.25 | 0.7 | 17.34 | 0.13 | 0.49 | 0.87 | ME |
| S5_vs_56083 | 0 | 0 | 0 | 26.84 | 0.04 | 18.79 | 0.2 | 20.92 | 4.44 | 0.18 | 271.66 | 0.17 | 14.75 | 4.69 | 10.92 | 11.39 | 1.34 | 27.92 | 16.96 | 0.76 | Shared |
| S5_vs_20500 | 0.01 | 0 | 0 | 5.43 | 0.34 | 4.72 | 126.01 | 41.21 | 9.15 | 4.59 | 13.31 | 0.86 | 1.43 | 7.29 | 6.39 | 1.71 | 3.81 | 7.16 | 10.95 | 3.92 | Shared |
| S5_vs_3183 | 0 | 0 | 0 | 6.23 | 0.03 | 0.8 | 0.03 | 21.34 | 0.11 | 0.82 | 1.87 | 0.53 | 0.69 | 0.28 | 0.53 | 4.64 | 2.12 | 0.41 | 7.74 | 1.61 | ME |
| S5_vs_141621 | 0 | 0 | 0 | 6.22 | 0.06 | 0.52 | 0.32 | 57.99 | 2.21 | 0.18 | 3.31 | 1.74 | 1.26 | 0.68 | 0.8 | 2.15 | 4.1 | 0.33 | 1.45 | 4.16 | ME |
| S5_vs_69083 | 0.01 | 0 | 0 | 1.14 | 0.08 | 0.26 | 0.09 | 42.48 | 0.38 | 0.13 | 0.76 | 0.06 | 0.91 | 0.14 | 5.74 | 0.31 | 5.64 | 0.28 | 0.48 | 0.74 | ME |
| S5_vs_51416 | 0 | 0 | 0 | 0.64 | 0.02 | 0.65 | 0.07 | 37.61 | 0.09 | 1.22 | 0.4 | 0.27 | 0.36 | 0.34 | 0.65 | 0.27 | 2.92 | 0.18 | 0.24 | 0.74 | ME |
| S5_vs_8847 | 0 | 0 | 0 | 0.99 | 0.03 | 0.69 | 0.05 | 28.62 | 0.25 | 2.75 | 1.32 | 1.42 | 0.81 | 1.56 | 1.15 | 0.89 | 4.43 | 0.25 | 1.44 | 0.78 | ME |
| S5_vs_93839 | 0 | 0 | 0.01 | 0.51 | 0.04 | 6.9 | 0.07 | 16.49 | 5.9 | 0.38 | 2.12 | 1.44 | 1.1 | 3.23 | 0.7 | 0.58 | 2.54 | 0.26 | 37.58 | 12.38 | Shared |
| S5_vs_3501 | 0.03 | 0 | 0 | 0.7 | 0.11 | 1.24 | 0.05 | 68.65 | 0.78 | 2.06 | 1.58 | 0.28 | 1.5 | 3 | 0.78 | 1.99 | 89.76 | 0.25 | 0.51 | 0.97 | ME |
| S5_vs_129970 | 0.02 | 0 | 0 | 13.03 | 13.45 | 2.2 | 0.08 | 23.34 | 1.04 | 1.9 | 97.37 | 0.21 | 2.26 | 0.91 | 5.43 | 4.97 | 5.37 | 1.15 | 2.29 | 5.02 | Shared |

| | | | | | | | | | | | | | | | | | | | | | | |
|---|---|---|---|---|---|---|---|---|---|---|---|---|---|---|---|---|---|---|---|---|---|---|
| S5_vs_124677 | 0.03 | 0 | 0.02 | 4.58 | 0.27 | 60.23 | 8.73 | 91.22 | 6.62 | 33.18 | 9.49 | 2.01 | 7.57 | 1.53 | 5.56 | 3.38 | 6.76 | 7.06 | 8.53 | 30.86 | ME | |
| S5_vs_130032 | 2.16 | 0.29 | 0 | 0.51 | 0.01 | 0.24 | 0.46 | 16.84 | 0.1 | 0.11 | 0.25 | 0.42 | 0.27 | 0.28 | 0.26 | 0.19 | 0.27 | 0.3 | 0.21 | 0.63 | ME | |
| S5_vs_130294 | 0.02 | 0 | 0 | 6.75 | 0.27 | 72.49 | 8.87 | 88.77 | 2.76 | 15.16 | 11.21 | 3.04 | 8.4 | 3.9 | 6.69 | 4.42 | 7.97 | 6.16 | 10.16 | 34.56 | Shared | |
| S5_vs_80496 | 0.01 | 0.01 | 0 | 38.3 | 3.97 | 30.75 | 1.12 | 77.36 | 38.2 | 12.96 | 9.61 | 4.05 | 17.1 | 5.32 | 9.86 | 7.02 | 40.47 | 24.13 | 24.5 | 42.04 | Shared | |
| S5_vs_142195 | 0 | 0 | 0 | 6.52 | 0.17 | 13.28 | 0.75 | 17.27 | 11.16 | 4.07 | 19.64 | 0.42 | 1.48 | 6.65 | 16.63 | 1.42 | 1.32 | 33.83 | 5.63 | 3.45 | Shared | |
| S5_vs_80498 | 0 | 0 | 0 | 0.61 | 0.01 | 0.48 | 0.05 | 175.7 | 0.36 | 0.66 | 1.88 | 0.09 | 0.62 | 1.88 | 0.51 | 0.19 | 4.61 | 0.93 | 0.38 | 0.66 | ME | |
| S5_vs_130476 | 0 | 0 | 0 | 1.12 | 0.04 | 0.31 | 0.03 | 9.93 | 0.18 | 0.46 | 0.29 | 0.08 | 0.42 | 0.45 | 0.24 | 0.32 | 0.39 | 0.35 | 0.72 | 1.32 | ME | All values are below 10 |
| S5_vs_107105 | 0 | 0 | 0 | 0.42 | 0.01 | 0.18 | 0.12 | 14.69 | 0.85 | 0.21 | 0.56 | 0.06 | 0.22 | 0.27 | 0.35 | 0.17 | 2.21 | 0.18 | 0.12 | 0.43 | ME | |
| S5_vs_142450 | 0 | 0 | 0 | 0.36 | 0.64 | 1.23 | 0.1 | 572.97 | 2672.03 | 6.18 | 1.17 | 0.04 | 1.19 | 5.13 | 0.24 | 0.57 | 0.19 | 0.15 | 0.64 | 0.73 | Shared | |
| S5_vs_112452 | 0 | 0 | 0 | 1.65 | 0.04 | 1.75 | 0.12 | 26.44 | 0.14 | 0.91 | 0.42 | 0.1 | 0.3 | 0.11 | 1.39 | 0.49 | 15.34 | 0.22 | 0.52 | 0.46 | ME | |
| S5_vs_63924 | 0.02 | 0.01 | 0 | 0.53 | 0.04 | 9.97 | 0.74 | 31.91 | 13.61 | 0.1 | 1.88 | 1.68 | 0.27 | 0.07 | 0.31 | 0.4 | 29.72 | 0.19 | 0.29 | 0.56 | Shared | |
| S5_vs_130958 | 0 | 0 | 0 | 0.57 | 0.02 | 0.2 | 0.05 | 13.55 | 0.08 | 0.13 | 0.5 | 3.39 | 0.29 | 0.46 | 0.22 | 0.27 | 5.98 | 0.19 | 0.24 | 0.79 | ME | |
| S5_vs_9747 | 0.02 | 0.01 | 0 | 30.09 | 0.13 | 0.63 | 0.03 | 270.98 | 446.34 | 0.37 | 2.05 | 0.07 | 0.88 | 7.59 | 0.69 | 20.93 | 0.33 | 0.41 | 0.42 | 0.81 | Shared | |
| S5_vs_4737 | 0.03 | 0 | 0 | 0.92 | 0.01 | 0.25 | 0.06 | 37.29 | 0.14 | 0.34 | 1.3 | 0.07 | 0.35 | 2.81 | 0.74 | 0.55 | 4.33 | 0.16 | 0.51 | 0.45 | ME | |
| S5_vs_80911 | 0 | 0 | 0 | 0.67 | 0.11 | 1.96 | 1.59 | 96.76 | 0.15 | 2.07 | 0.61 | 0.15 | 1.6 | 1.69 | 0.84 | 0.56 | 9.21 | 0.76 | 0.83 | 0.9 | ME | |
| S5_vs_69980 | 0.01 | 0 | 0 | 0.88 | 0.08 | 2.2 | 0.89 | 18.29 | 1.85 | 7.15 | 1.96 | 0.16 | 0.54 | 0.95 | 0.31 | 0.52 | 13.77 | 0.38 | 0.98 | 2.8 | ME | |
| S5_vs_21778 | 0 | 0 | 0 | 0.57 | 0.03 | 0.54 | 0.06 | 22.24 | 0.15 | 0.29 | 0.72 | 0.16 | 0.41 | 0.56 | 0.49 | 1.46 | 0.92 | 0.31 | 0.98 | 0.75 | ME | |
| S5_vs_113386 | 0.02 | 0 | 0 | 0.86 | 0.05 | 0.39 | 0.26 | 10.7 | 0.15 | 0.14 | 0.64 | 0.26 | 0.58 | 0.29 | 2.65 | 0.21 | 0.27 | 0.27 | 0.43 | 0.73 | ME | |
| S5_vs_95307 | 0 | 0 | 0 | 0.48 | 0.03 | 0.45 | 0.06 | 23.67 | 0.22 | 0.63 | 0.4 | 1.21 | 1.45 | 2.07 | 0.4 | 0.38 | 8.45 | 0.21 | 0.47 | 1.2 | ME | |
| S5_vs_131510 | 0.02 | 0.01 | 0 | 63.14 | 4.27 | 34.55 | 4.43 | 64.14 | 13.8 | 13.05 | 13.08 | 3.84 | 23.86 | 6.87 | 12.73 | 4.99 | 38.44 | 25.84 | 18.57 | 70.22 | Shared | |
| S5_vs_46394 | 0.01 | 0 | 0.01 | 1.53 | 0.08 | 1.9 | 0.26 | 18.54 | 0.44 | 5.02 | 2.79 | 0.48 | 1.15 | 1.11 | 1.12 | 2.31 | 14 | 0.81 | 1 | 1.73 | ME | |
| S5_vs_89540 | 0 | 0 | 0 | 0.68 | 0.04 | 0.38 | 0.05 | 43.2 | 0.13 | 0.16 | 0.32 | 0.08 | 0.36 | 0.54 | 0.52 | 0.29 | 0.82 | 0.24 | 0.22 | 0.73 | ME | |
| S5_vs_17520 | 0 | 0 | 0 | 0.58 | 0.01 | 0.56 | 0.06 | 17.71 | 0.14 | 0.58 | 0.6 | 0.79 | 0.49 | 0.32 | 0.24 | 0.17 | 1.84 | 0.26 | 0.27 | 0.89 | ME | |
| S5_vs_137154 | 0.01 | 0 | 0 | 0.33 | 0.05 | 1.05 | 0.24 | 45.76 | 0.28 | 0.3 | 1.11 | 0.1 | 1.72 | 0.32 | 0.63 | 0.72 | 0.93 | 0.48 | 0.92 | 0.95 | ME | |
| S5_vs_40848 | 0 | 0 | 0 | 4.04 | 1.03 | 10.62 | 0.86 | 50.29 | 4.08 | 23.77 | 9.33 | 4.27 | 4.09 | 12.35 | 1.85 | 2.22 | 8.31 | 0.49 | 6.12 | 1.62 | ME | |
| S5_vs_28263 | 0 | 0 | 0 | 0.81 | 0.07 | 0.46 | 0.2 | 20.75 | 0.09 | 0.64 | 0.76 | 0.13 | 0.46 | 0.32 | 0.56 | 0.33 | 3.39 | 0.22 | 0.45 | 0.94 | ME | |
| S5_vs_72532 | 0.01 | 0 | 0 | 16.28 | 1.45 | 12.18 | 0.32 | 33.6 | 5.89 | 4.12 | 4.52 | 5.58 | 6.14 | 2.51 | 3.78 | 1.73 | 14.28 | 5.89 | 8.27 | 18.62 | ME | |
| S5_vs_41663 | 0 | 0 | 0 | 0.61 | 0.02 | 0.32 | 0.04 | 43.3 | 0.11 | 0.1 | 0.41 | 0.06 | 0.4 | 0.85 | 0.37 | 0.29 | 0.33 | 0.17 | 0.22 | 0.51 | ME | |
| S5_vs_145026 | 0 | 0 | 0 | 0.71 | 1.23 | 0.34 | 0.08 | 58.63 | 0.08 | 0.14 | 0.46 | 0.06 | 0.39 | 0.17 | 0.16 | 0.23 | 5.78 | 0.16 | 0.25 | 0.44 | ME | |
| S5_vs_77593 | 0.04 | 0 | 0.01 | 2.55 | 0.09 | 1.44 | 0.25 | 35.03 | 0.32 | 1.72 | 5.15 | 0.37 | 1.61 | 1.11 | 2.98 | 2.5 | 2.61 | 1.47 | 1.47 | 1.46 | ME | |
| S5_vs_58766 | 0 | 0 | 0 | 0.41 | 0.02 | 0.4 | 0.16 | 11.51 | 0.09 | 0.25 | 0.63 | 0.19 | 0.62 | 0.35 | 0.36 | 0.23 | 2.31 | 0.17 | 0.24 | 0.84 | ME | |
| S5_vs_58859 | 0.01 | 0 | 0 | 0.59 | 0.04 | 0.6 | 0.01 | 25.01 | 0.16 | 1.53 | 0.92 | 0.09 | 0.42 | 0.25 | 0.49 | 0.2 | 22.58 | 0.23 | 1.08 | 0.71 | ME | |
| S5_vs_83886 | 0 | 0 | 0 | 0.7 | 0.05 | 0.48 | 0.08 | 46.53 | 0.47 | 10.53 | 9.14 | 0.23 | 2.43 | 3.22 | 0.4 | 1.17 | 11.57 | 0.54 | 9.36 | 9.16 | ME | |
| S5_vs_138980 | 0.01 | 0 | 0 | 1.15 | 0.03 | 0.89 | 0.12 | 16.09 | 0.24 | 0.85 | 2.83 | 0.45 | 1.13 | 2.46 | 1.98 | 1.9 | 3.38 | 0.52 | 0.67 | 1.07 | ME | |
| S5_vs_59798 | 0.03 | 0 | 0 | 4.15 | 0.09 | 0.56 | 0.09 | 114.49 | 0.15 | 0.68 | 0.81 | 0.24 | 1.1 | 0.47 | 0.53 | 0.35 | 127.16 | 0.28 | 0.45 | 1.27 | ME | |
| S5_vs_84574 | 0.02 | 0 | 0 | 1.2 | 0.07 | 11.35 | 0.36 | 25.91 | 0.16 | 0.32 | 1 | 2.98 | 0.38 | 0.53 | 0.48 | 0.41 | 8.42 | 0.49 | 0.42 | 0.65 | ME | |
| S5_vs_84946 | 0 | 0 | 0 | 3.08 | 0.09 | 4 | 0.07 | 60.91 | 0.12 | 1.13 | 0.93 | 0.95 | 0.9 | 0.48 | 7.93 | 0.31 | 37.18 | 0.79 | 0.57 | 1.16 | ME | |
| S5_vs_139401 | 0 | 0 | 0 | 1.25 | 0.05 | 0.36 | 0.19 | 43.59 | 0.2 | 0.59 | 0.82 | 0.24 | 0.44 | 0.29 | 0.5 | 0.48 | 0.31 | 0.25 | 0.29 | 0.73 | ME | |
| S5_vs_60507 | 0 | 0 | 0 | 0.43 | 0.03 | 0.46 | 0.06 | 19.6 | 0.13 | 0.28 | 0.54 | 0.07 | 0.41 | 0.37 | 0.41 | 0.34 | 2.55 | 0.25 | 0.4 | 0.98 | ME | |
| S5_vs_7444 | 0 | 0 | 0 | 0.91 | 0.01 | 1.42 | 0.13 | 27.64 | 0.09 | 0.36 | 1.33 | 0.08 | 0.15 | 0.67 | 1.06 | 1.18 | 14.99 | 0.14 | 0.71 | 0.56 | ME | |
| S5_vs_74199 | 0.01 | 0 | 0 | 3.57 | 0.1 | 1.64 | 0.47 | 33.91 | 0.48 | 5.24 | 8.29 | 0.19 | 2.76 | 0.65 | 3.45 | 2.13 | 22.33 | 1.6 | 0.6 | 1.53 | ME | |
| S5_vs_65490 | 0 | 0 | 0 | 0.74 | 0.21 | 0.31 | 0.03 | 19.44 | 0.19 | 0.15 | 4.58 | 0.11 | 0.44 | 3.46 | 0.61 | 0.65 | 0.2 | 0.2 | 0.3 | 0.53 | ME | |
| S5_vs_76500 | 0 | 0 | 0 | 1.14 | 0.19 | 5 | 0.09 | 93.72 | 21.82 | 1.17 | 2.97 | 0.5 | 4.47 | 2.85 | 8.08 | 1.05 | 9.65 | 4.51 | 3.84 | 6.25 | Shared | |
| S7_vf_5439 | 0.02 | 0 | 0.06 | 3.52 | 0.38 | 500.25 | 7.47 | 0.89 | 0.36 | 1141.77 | 7.39 | 2.24 | 16.63 | 0.16 | 3.43 | 4.26 | 0.28 | 24.11 | 72.7 | 33.9 | Shared | |
| S7_vf_5801 | 0.06 | 0.01 | 0 | 28.98 | 7.19 | 18.91 | 77.67 | 2.68 | 0.72 | 138.92 | 14.64 | 2.44 | 63.57 | 8.59 | 7.78 | 27.93 | 1126.64 | 31.79 | 52.14 | 61.77 | Shared | |
| S7_vf_5843 | 0.01 | 0 | 0.01 | 0.67 | 0 | 0.29 | 7.73 | 0.14 | 0.06 | 10.69 | 0.36 | 0.03 | 0.1 | 0.12 | 0.21 | 0.22 | 1.03 | 0.14 | 0.18 | 0.4 | ME | |
| S7_vf_8332 | 0.01 | 0 | 0 | 1.48 | 0.09 | 1.33 | 0.17 | 1.39 | 0.31 | 28.36 | 1.99 | 0.5 | 2.51 | 3.3 | 2.96 | 2.15 | 1.29 | 3.05 | 1.67 | 1.73 | ME | |
| S7_vf_9334 | 0 | 0 | 0.02 | 0.52 | 0.42 | 192.04 | 0.07 | 0.54 | 0.48 | 1998.96 | 0.89 | 0.39 | 0.63 | 0.31 | 2.75 | 0.33 | 0.22 | 0.15 | 0.19 | 0.52 | ME | |
| S7_vf_11744 | 0 | 0 | 0 | 0.28 | 0.07 | 0.69 | 0.11 | 0.28 | 0.13 | 22.37 | 2.49 | 0.11 | 1.55 | 0.75 | 1.33 | 0.64 | 0.44 | 0.27 | 0.44 | 0.85 | ME | |
| S7_vf_13614 | 0 | 0 | 0 | 3.14 | 0.26 | 0.62 | 0.05 | 0.35 | 0.3 | 1397.29 | 0.84 | 0.08 | 0.28 | 0.06 | 1.79 | 0.85 | 1.43 | 12.99 | 2.26 | 0.58 | Shared | |
| S7_vf_19866 | 0 | 0 | 0 | 1.81 | 3.17 | 3.58 | 0.11 | 2.78 | 2.07 | 16931.24 | 5.5 | 0.08 | 0.21 | 0.28 | 21.87 | 1.98 | 4.59 | 0.18 | 0.27 | 0.46 | Shared | |
| S7_vf_20129 | 0 | 0 | 0 | 0.74 | 0.88 | 1.44 | 0 | 0.79 | 1.83 | 7039.56 | 2.3 | 0.16 | 0.44 | 910.65 | 8.93 | 0.75 | 0.38 | 0.31 | 0.32 | 0.73 | ME | |
| S7_vf_27309 | 0 | 0.01 | 0 | 21.4 | 0.8 | 0.75 | 0.06 | 0.37 | 0.33 | 1840.74 | 0.93 | 0.05 | 0.36 | 0.11 | 2.63 | 0.36 | 0.13 | 0.26 | 0.18 | 0.32 | ME | |
| S7_vf_27437 | 0 | 0 | 0 | 4.22 | 9.85 | 0.3 | 0.18 | 0.2 | 0.09 | 120.54 | 0.67 | 0.08 | 0.72 | 13.01 | 0.7 | 0.21 | 206.01 | 0.35 | 0.29 | 1.04 | ME | |
| S7_vf_28429 | 0 | 0 | 0 | 16.2 | 0.74 | 21.53 | 11.9 | 13 | 2.92 | 4020.95 | 13.98 | 0.26 | 5.57 | 548.46 | 5.53 | 27.36 | 48.29 | 0.24 | 0.29 | 0.54 | Shared | |
| S7_vf_29196 | 0 | 0 | 0 | 7.57 | 0.19 | 3.15 | 0.06 | 0.19 | 0.1 | 16.34 | 3.26 | 249.4 | 0.27 | 0.28 | 0.33 | 2.52 | 9.68 | 0.19 | 0.27 | 3.63 | ME | |
| S7_vf_29941 | 0.23 | 0 | 0.01 | 0.39 | 2.69 | 80.75 | 65.6 | 10.68 | 3.5 | 39.21 | 32.53 | 14.16 | 1.72 | 0.92 | 1.69 | 1.83 | 4082.48 | 0.73 | 9.63 | 554.71 | Shared | |
| S7_vf_32396 | 0 | 0 | 0 | 0.39 | 0.02 | 0.16 | 0.05 | 0.11 | 0.09 | 99.68 | 0.38 | 2.87 | 0.19 | 31.44 | 0.47 | 0.95 | 0.21 | 0.16 | 1.66 | 0.71 | ME | |
| S7_vf_32559 | 0 | 0 | 0 | 0.31 | 0.07 | 0.4 | 0.05 | 0.18 | 0.13 | 661.84 | 4.22 | 0.04 | 0.12 | 0.05 | 1.25 | 0.19 | 0.2 | 0.04 | 0.18 | 0.31 | ME | |
| S7_vf_36627 | 0 | 0 | 0 | 1.74 | 0.03 | 0.39 | 0.16 | 0.19 | 0.13 | 48.44 | 0.71 | 0.1 | 0.9 | 0.19 | 0.46 | 0.83 | 0.32 | 0.31 | 0.56 | 0.84 | ME | |
| S7_vf_37279 | 0.02 | 0 | 0.01 | 1.69 | 0.49 | 70.4 | 12.51 | 1.14 | 0.25 | 145.67 | 0.89 | 0.34 | 1.3 | 16.82 | 5.73 | 1.27 | 29.56 | 5.31 | 1.11 | 53.51 | Shared | |
| S7_vf_37293 | 0 | 0 | 0 | 0.39 | 0.03 | 1.14 | 0.06 | 0.22 | 0.18 | 35.88 | 0.65 | 0.08 | 0.4 | 5.69 | 0.36 | 0.38 | 0.27 | 0.28 | 0.28 | 0.66 | ME | |
| S7_vf_38697 | 0 | 0 | 0 | 1.37 | 1.12 | 1.87 | 0.08 | 1.61 | 0.94 | 5690.73 | 2.57 | 0.13 | 1.25 | 0.22 | 8.16 | 1.01 | 0.54 | 0.44 | 0.87 | 1 | ME | |
| S7_vf_39724 | 0.02 | 0 | 0.01 | 4.09 | 0.05 | 40.87 | 1.14 | 18.14 | 0.71 | 153.34 | 14.82 | 0.48 | 6.89 | 5.36 | 11.14 | 3.3 | 0.69 | 6.13 | 4.25 | 1.4 | Shared | |
| S7_vf_49007 | 0 | 0 | 0 | 0.46 | 0.01 | 0.29 | 0.05 | 0.14 | 0.1 | 23.56 | 9.8 | 0.04 | 0.54 | 3.54 | 0.41 | 0.22 | 0.25 | 0.23 | 0.3 | 0.51 | ME | |
| S7_vf_49485 | 0.01 | 0 | 0 | 8.96 | 0.04 | 0.51 | 0.1 | 0.3 | 0.16 | 39.56 | 0.77 | 0.11 | 0.81 | 0.32 | 0.5 | 4.1 | 0.45 | 0.4 | 5.41 | 11.8 | ME | |
| S7_vf_60968 | 0 | 0 | 0 | 0.63 | 0.09 | 0.56 | 0.15 | 0.8 | 1.15 | 11.49 | 0.53 | 0.24 | 0.68 | 1.11 | 0.45 | 0.41 | 1.46 | 0.38 | 0.46 | 5.93 | ME | |
| S7_vf_63894 | 0 | 0 | 0 | 0.41 | 0.01 | 0.28 | 0.05 | 0.33 | 0.15 | 13.38 | 0.48 | 0.04 | 0.4 | 2.81 | 0.44 | 0.22 | 1.22 | 0.1 | 0.53 | 0.44 | ME | |
| S7_vf_67910 | 0.01 | 0 | 0.01 | 0.78 | 0.03 | 0.44 | 0.03 | 2.13 | 0.13 | 20.88 | 2.65 | 0.05 | 0.24 | 5.03 | 0.28 | 0.3 | 0.14 | 0.22 | 0.25 | 0.55 | ME | |
| S7_vf_68239 | 0 | 0 | 0 | 0.38 | 0.04 | 0.65 | 3.01 | 0.42 | 0.15 | 50.21 | 1.13 | 0.28 | 0.92 | 0.35 | 0.84 | 0.47 | 38.07 | 0.24 | 0.82 | 26.24 | ME | |
| S7_vf_68546 | 0 | 0 | 0 | 0.75 | 0.04 | 3.55 | 0.22 | 0.23 | 0.21 | 19.5 | 1.28 | 0.12 | 0.55 | 0.78 | 0.73 | 0.34 | 0.28 | 0.42 | 0.45 | 0.75 | ME | |
| S7_vf_69348 | 0 | 0 | 0 | 0.46 | 0 | 0.36 | 0.02 | 0.15 | 0.08 | 33.71 | 0.37 | 0.07 | 0.56 | 0.42 | 0.52 | 0.3 | 0.27 | 0.15 | 0.21 | 15.55 | ME | |
| S7_vf_71527 | 0 | 0 | 0 | 1.91 | 0 | 1.94 | 0.36 | 0.56 | 0.33 | 15.5 | 3.44 | 1.58 | 2.24 | 9.79 | 1.71 | 1.97 | 1.65 | 1.27 | 4.04 | 1.52 | ME | |
| S7_vf_72493 | 0 | 0 | 0 | 0.49 | 0.04 | 0.5 | 0.03 | 0.59 | 0.19 | 14.4 | 0.63 | 0.05 | 4.06 | 0.3 | 0.51 | 0.74 | 0.33 | 0.35 | 0.39 | 6.67 | ME | |
| S7_vf_76382 | 0 | 0 | 0 | 0.33 | 0.1 | 0.36 | 0.04 | 0.61 | 0.63 | 29.71 | 537.55 | 0.03 | 4.53 | 0.19 | 0.34 | 0.79 | 0.13 | 0.08 | 0.21 | 1.23 | Shared | |
| S7_vf_76463 | 0 | 0 | 0 | 1.2 | 0.05 | 0.46 | 64.69 | 0.34 | 0.1 | 75.82 | 28.24 | 0.07 | 8.94 | 0.3 | 0.45 | 0.33 | 0.34 | 0.6 | 0.81 | 0.4 | Shared | |
| S7_vf_83624 | 0 | 0 | 0 | 0.43 | 0.08 | 0.35 | 0.15 | 0.29 | 0.2 | 233.03 | 0.93 | 0.08 | 1.07 | 0.24 | 0.68 | 0.36 | 5.83 | 0.26 | 1.42 | 1.06 | ME | |
| S7_vf_84974 | 0 | 0 | 0 | 0.82 | 0.03 | 1.15 | 0.29 | 1.07 | 0.21 | 18.41 | 1.18 | 0.18 | 0.53 | 0.24 | 0.58 | 0.31 | 0.48 | 0.29 | 0.54 | 7.49 | ME | |
| S7_vf_89197 | 0 | 0 | 0.1 | 0.94 | 0.35 | 1054.06 | 0.25 | 137.03 | 10.87 | 794.43 | 12.79 | 1.5 | 0.72 | 0.29 | 1.54 | 0.53 | 2.2 | 0.4 | 0.33 | 15.86 | Shared | |
| S7_vf_89668 | 0 | 0 | 0 | 0.6 | 0.86 | 1.34 | 0.12 | 0.93 | 0.82 | 5858.05 | 2.19 | 0.09 | 0.2 | 0.07 | 6.98 | 0.76 | 0.13 | 0.08 | 0.13 | 0.35 | ME | |
| S7_vf_114476 | 0.01 | 0 | 0 | 0.72 | 0.03 | 0.57 | 0.03 | 0.2 | 0.17 | 37.11 | 0.31 | 0.05 | 0.47 | 0.09 | 0.5 | 0.34 | 0.24 | 0.16 | 0.3 | 0.67 | ME | |
| S7_vf_117697 | 0 | 0 | 0.03 | 41.24 | 8.67 | 578.02 | 5.44 | 446.26 | 20.72 | 74.77 | 4.51 | 0.83 | 20.63 | 17.49 | 1.42 | 14.85 | 16.15 | 0.79 | 9.79 | 17.97 | Shared | |
| S7_vf_134023 | 0 | 0 | 0 | 0.78 | 0.03 | 4.35 | 0.14 | 0.78 | 0.23 | 32.63 | 0.73 | 0.09 | 1.26 | 1.05 | 6.66 | 0.32 | 0.78 | 0.3 | 0.48 | 0.54 | ME | |
| S7_vf_135778 | 0 | 0 | 0 | 0.32 | 0 | 0.24 | 0.07 | 0.15 | 0.07 | 69.47 | 0.5 | 0.06 | 0.23 | 0.34 | 0.35 | 0.23 | 0.11 | 0.14 | 0.21 | 0.49 | ME | |

| | | | | | | | | | | | | | | | | | | | | | |
|---|---|---|---|---|---|---|---|---|---|---|---|---|---|---|---|---|---|---|---|---|---|
| S7_vf_138019 | 0 | 0 | 0 | 1.1 | 0.01 | 6.96 | 0.04 | 0.61 | 0.2 | 101.28 | 1.15 | 0.11 | 1 | 0.54 | 0.29 | 0.51 | 0.41 | 0.24 | 3 | 0.85 | ME |
| S7_vf_138069 | 0 | 0 | 0 | 0.46 | 0.1 | 1.1 | 0.08 | 41.72 | 0.13 | 97.63 | 0.48 | 0.1 | 0.78 | 0.3 | 0.86 | 0.36 | 0.32 | 0.27 | 7.34 | 0.71 | ME |
| S7_vf_138225 | 0 | 0 | 0 | 0.44 | 0.03 | 0.27 | 0.09 | 0.17 | 0.11 | 13.95 | 0.35 | 0.09 | 0.31 | 2.3 | 0.24 | 0.15 | 0.31 | 0.31 | 0.26 | 0.74 | ME |
| S7_vs_557 | 0 | 0 | 0 | 1.02 | 0.23 | 1.17 | 0.13 | 0.63 | 0.66 | 1443.35 | 0.91 | 0.37 | 1.51 | 4.07 | 2.3 | 1.02 | 0.62 | 2.42 | 1.16 | 5.98 | ME |
| S7_vs_2599 | 0 | 0 | 0 | 1.27 | 0.54 | 44.38 | 1.05 | 0.4 | 0.23 | 177.66 | 3.47 | 0.19 | 1.39 | 1.12 | 0.7 | 0.76 | 20.78 | 0.4 | 0.45 | 2.7 | ME |
| S7_vs_4956 | 0 | 0 | 0 | 3.6 | 7.32 | 0.75 | 2.77 | 111.37 | 0.59 | 2589.19 | 2.8 | 0.25 | 2.03 | 3.81 | 52.99 | 6.46 | 5.03 | 0.21 | 0.87 | 0.42 | Shared |
| S7_vs_5236 | 0.01 | 0 | 0 | 1.02 | 0.03 | 3.24 | 0.77 | 2.24 | 0.22 | 118.14 | 1.61 | 0.38 | 0.25 | 2.68 | 3.54 | 8.43 | 0.28 | 1.16 | 0.48 | 2.45 | ME |
| S7_vs_27530 | 0 | 0 | 0 | 0.63 | 0.06 | 0.63 | 0.15 | 0.64 | 0.21 | 25.45 | 1.83 | 0.07 | 0.5 | 1.63 | 1.26 | 3.61 | 0.47 | 0.24 | 2.93 | 0.56 | ME |
| S7_vs_36656 | 0 | 0 | 0 | 0.58 | 0.06 | 0.32 | 0.26 | 0.16 | 0.09 | 23.35 | 0.44 | 0.05 | 0.43 | 0.22 | 0.4 | 0.29 | 0.17 | 0.17 | 0.19 | 0.58 | ME |
| S7_vs_42563 | 0.01 | 0 | 0.02 | 1.11 | 0.07 | 0.51 | 0.05 | 0.45 | 0.14 | 30.45 | 10.31 | 0.15 | 1.19 | 0.42 | 0.71 | 0.52 | 0.23 | 0.34 | 8.07 | 0.62 | Shared |
| S7_vs_54322 | 0 | 0 | 0 | 1.21 | 0.16 | 0.53 | 0.15 | 0.33 | 0.21 | 24.73 | 6.66 | 36.15 | 2.55 | 0.26 | 0.74 | 0.36 | 0.36 | 0.99 | 1.07 | 1.84 | ME |
| S7_vs_60775 | 0 | 0 | 0 | 1.19 | 0.07 | 2.46 | 0.02 | 0.18 | 0.12 | 37.18 | 0.36 | 0.09 | 0.4 | 0.56 | 0.79 | 0.37 | 0.32 | 0.33 | 0.36 | 0.76 | ME |
| S7_vs_65719 | 0 | 0 | 0 | 0.73 | 0.02 | 1.08 | 0.04 | 13.67 | 14.78 | 63.57 | 0.26 | 0.45 | 0.32 | 11.66 | 0.37 | 0.17 | 30.26 | 0.26 | 4.4 | 1.88 | Shared |
| S7_vs_94738 | 0 | 0 | 0 | 7.01 | 1.37 | 1.41 | 0.84 | 0.72 | 0.3 | 44.98 | 1.96 | 0.35 | 1.9 | 1.4 | 0.5 | 2.83 | 2.88 | 0.31 | 2.56 | 2.06 | ME |
| S7_vs_95276 | 0 | 0 | 0 | 0.67 | 0.72 | 3.11 | 0.07 | 3.42 | 0.2 | 39.87 | 6.06 | 0.05 | 0.26 | 0.11 | 0.24 | 0.21 | 13.34 | 0.14 | 0.27 | 1.47 | ME |
| S7_vs_103530 | 0 | 0 | 0 | 5.8 | 0.08 | 76.93 | 7.73 | 0.29 | 0.15 | 28.17 | 1.01 | 14.15 | 0.43 | 0.42 | 0.63 | 1.15 | 0.49 | 0.44 | 3.92 | 14.01 | ME |
| S7_vs_129271 | 0 | 0 | 0 | 0.68 | 0.04 | 0.23 | 0.05 | 2.11 | 0.15 | 84.05 | 0.35 | 0.08 | 3.13 | 10.09 | 0.33 | 1.58 | 1.11 | 0.23 | 0.23 | 0.51 | ME |
| S7_vs_134391 | 0 | 0 | 0 | 5.31 | 0.01 | 0.15 | 0 | 0.15 | 0.09 | 36.08 | 0.55 | 0.04 | 0.13 | 0.22 | 0.81 | 1.93 | 0.16 | 0.16 | 0.15 | 0.31 | ME |
| S7_vs_134784 | 0 | 0 | 0 | 0.65 | 0.06 | 0.4 | 0.17 | 0.43 | 0.26 | 45.31 | 0.5 | 0.06 | 0.41 | 0.28 | 0.39 | 0.15 | 0.6 | 0.33 | 0.38 | 0.91 | ME |
| S7_vs_135322 | 0.01 | 0 | 0 | 1.79 | 0.02 | 5.57 | 0.18 | 0.25 | 0.11 | 58.32 | 1.1 | 0.31 | 3.58 | 0.13 | 0.77 | 0.57 | 0.16 | 0.68 | 42.35 | 0.84 | Shared |
| S7_vs_135679 | 0 | 0 | 0 | 1.49 | 0.02 | 0.68 | 0.19 | 0.43 | 0.2 | 36.7 | 1.5 | 0.52 | 1.07 | 0.13 | 0.73 | 0.25 | 0.32 | 0.64 | 0.66 | 0.94 | ME |
| S7_vs_135828 | 0 | 0 | 0 | 0.59 | 0.19 | 0.84 | 0.52 | 0.43 | 0.4 | 59.62 | 0.72 | 0.17 | 1.64 | 127.2 | 0.76 | 0.4 | 0.73 | 0.52 | 0.49 | 1.27 | ME |
| S7_vs_136164 | 0 | 0 | 0 | 0.87 | 0.02 | 0.25 | 0.05 | 0.44 | 0.1 | 18.85 | 0.57 | 0.1 | 0.22 | 0.52 | 0.43 | 0.24 | 0.43 | 0.25 | 0.3 | 0.56 | ME |
| S7_vs_137611 | 0 | 0 | 0 | 0.74 | 0.07 | 1.39 | 0.21 | 0.35 | 0.21 | 73.63 | 0.65 | 0.12 | 1.03 | 0.43 | 0.9 | 0.45 | 0.51 | 0.47 | 0.56 | 1.06 | ME |
| S7_vs_137915 | 0 | 0 | 0 | 0.98 | 0.09 | 0.75 | 0.14 | 0.35 | 0.22 | 444.06 | 11.57 | 0.29 | 0.92 | 0.28 | 1.16 | 0.51 | 0.46 | 0.45 | 0.68 | 1.01 | Shared |
| S7_vs_137998 | 0 | 0 | 0 | 4.9 | 0.08 | 1.07 | 0.22 | 0.43 | 0.19 | 295.05 | 0.93 | 0.15 | 0.78 | 0.59 | 2.29 | 1 | 0.49 | 0.86 | 28.92 | 14.8 | Shared |
| S7_vs_138096 | 0 | 0 | 0 | 0.59 | 0.08 | 4.32 | 0.07 | 0.25 | 0.21 | 315.06 | 0.49 | 0.1 | 0.35 | 0.28 | 1.25 | 0.42 | 0.24 | 0.45 | 0.25 | 0.54 | ME |
| S7_vs_138109 | 0 | 0 | 0 | 0.98 | 0.05 | 0.97 | 0.04 | 0.9 | 0.2 | 38.54 | 0.62 | 0.11 | 0.91 | 0.47 | 1.61 | 0.44 | 1.11 | 0.45 | 0.47 | 1.14 | ME |
| S7_vs_138116 | 0 | 0 | 0 | 0.41 | 0.04 | 0.54 | 0.02 | 0.22 | 0.15 | 37.91 | 0.33 | 0.1 | 0.75 | 0.67 | 0.59 | 0.29 | 0.65 | 0.37 | 0.36 | 1.87 | ME |
| S7_vs_138140 | 0 | 0 | 0 | 0.9 | 0.02 | 3.67 | 0.02 | 0.42 | 0.51 | 51.84 | 0.64 | 0.15 | 0.38 | 2.72 | 0.57 | 0.3 | 0.29 | 0.24 | 0.38 | 0.86 | ME |
| S7_vs_138173 | 0 | 0 | 0 | 0.46 | 0.02 | 12.01 | 31.4 | 0.38 | 1.19 | 63.17 | 7.22 | 0.08 | 6.23 | 0.25 | 0.31 | 0.23 | 0.41 | 71.73 | 13.86 | 16.61 | Shared |
| S7_vs_944 | 0.01 | 0 | 0 | 0.73 | 0.02 | 0.34 | 0.05 | 0.17 | 0.12 | 12.64 | 0.87 | 0.08 | 0.68 | 0.2 | 0.3 | 0.25 | 0.31 | 0.44 | 0.36 | 5.48 | ME |
| S7_vs_4114 | 0 | 0 | 0 | 2.19 | 0.53 | 3.07 | 0.33 | 0.79 | 0.1 | 42.51 | 34.28 | 0.75 | 0.33 | 3.32 | 1.5 | 1.26 | 3.12 | 1.1 | 1.83 | 1.05 | Shared |
| S7_vs_6626 | 0.02 | 0 | 0 | 14.11 | 0.45 | 3.1 | 13.52 | 28.29 | 8.2 | 454.41 | 6.42 | 4.87 | 55.93 | 0.39 | 1.47 | 4.86 | 2.23 | 7.22 | 2.65 | 7.22 | Shared |
| S7_vs_11081 | 0.01 | 0.01 | 0.01 | 14.35 | 0.09 | 12.69 | 61.95 | 0.82 | 0.32 | 337.04 | 2.03 | 0.2 | 101.57 | 7.92 | 2.96 | 5.66 | 0.46 | 97.69 | 10.35 | 10.13 | Shared |
| S7_vs_13041 | 0 | 0 | 0 | 0.56 | 0.09 | 1.17 | 0.18 | 0.42 | 0.3 | 138.06 | 1.24 | 0.16 | 1.42 | 1.03 | 1.18 | 0.57 | 0.69 | 0.47 | 0.7 | 1.38 | ME |
| S7_vs_13370 | 0.04 | 0 | 0.01 | 7 | 0.38 | 74.41 | 12.32 | 68.51 | 4.04 | 62.23 | 11.89 | 3.87 | 12.23 | 2.27 | 9.68 | 3.64 | 12.42 | 11.3 | 13.25 | 53.52 | Shared |
| S7_vs_13799 | 0 | 0 | 0 | 1.28 | 0.02 | 0.44 | 0.18 | 2.87 | 0.21 | 15.4 | 0.49 | 0.3 | 0.71 | 0.8 | 0.58 | 0.27 | 1.25 | 0.35 | 0.56 | 7.16 | ME |
| S7_vs_13903 | 0 | 0 | 0 | 1.32 | 1.02 | 2.33 | 0.12 | 1.18 | 0.83 | 5498.38 | 2.47 | 0.31 | 1.07 | 0.53 | 7.11 | 0.92 | 0.47 | 0.15 | 0.36 | 0.92 | ME |
| S7_vs_14275 | 0 | 0 | 0 | 1.35 | 0.03 | 0.36 | 0.14 | 0.18 | 0.13 | 54.87 | 12.18 | 0.08 | 1.18 | 0.19 | 0.76 | 0.96 | 0.22 | 0.25 | 0.46 | 0.86 | Shared |
| S7_vs_16344 | 0.02 | 0 | 0 | 11.9 | 2.77 | 16.93 | 2 | 49.57 | 4.32 | 53.76 | 30.99 | 10.46 | 8.39 | 52.66 | 4.84 | 5.94 | 33.08 | 1.37 | 13.28 | 6.42 | Shared |
| S7_vs_20200 | 0 | 0 | 0 | 0.98 | 0.11 | 0.49 | 0.05 | 1 | 0.22 | 25.37 | 4.01 | 0.2 | 0.25 | 0.47 | 0.29 | 0.39 | 1.98 | 0.35 | 0.51 | 2.19 | ME |
| S7_vs_21521 | 0 | 0 | 0 | 1.43 | 0.06 | 3.57 | 0.27 | 2.6 | 0.36 | 18.82 | 10.49 | 2.02 | 1.98 | 42.2 | 6.71 | 9.78 | 1.48 | 1.05 | 7.34 | 8.62 | Shared |
| S7_vs_23770 | 0 | 0 | 0 | 1.73 | 2.98 | 4.28 | 1.31 | 118.16 | 4.75 | 59.79 | 17.36 | 0.62 | 0.59 | 0.76 | 1.76 | 0.51 | 185.93 | 0.35 | 7.34 | 1.18 | Shared |
| S7_vs_26235 | 0 | 0 | 0 | 0.54 | 0.07 | 0.48 | 0.1 | 0.62 | 0.93 | 19.03 | 0.75 | 0.13 | 0.27 | 0.33 | 0.29 | 0.35 | 1.04 | 0.22 | 0.43 | 8.78 | ME |
| S7_vs_26757 | 0 | 0 | 0 | 5.11 | 0.53 | 52.68 | 8.68 | 39.96 | 0.92 | 48.8 | 9.3 | 0.76 | 9.5 | 2.53 | 8.34 | 2.35 | 13.52 | 5.47 | 12.23 | 69.11 | Shared |
| S7_vs_28150 | 0.01 | 0 | 0 | 1.23 | 0.03 | 1.72 | 0.13 | 1.81 | 0.24 | 14.57 | 0.91 | 0.79 | 0.46 | 0.82 | 1.54 | 0.4 | 0.97 | 0.33 | 0.78 | 1.2 | ME |
| S7_vs_28733 | 0 | 0 | 0 | 0.69 | 0.26 | 0.71 | 0.05 | 0.85 | 0.19 | 218.26 | 0.75 | 30.39 | 0.52 | 0.32 | 0.73 | 0.27 | 18.68 | 0.28 | 3.55 | 3.35 | ME |
| S7_vs_29595 | 0 | 0 | 0 | 0.75 | 0.11 | 0.36 | 0.08 | 0.28 | 0.22 | 603.3 | 0.53 | 0.08 | 0.35 | 0.28 | 0.94 | 0.32 | 0.16 | 0.24 | 0.23 | 0.57 | ME |
| S7_vs_30640 | 0 | 0 | 0 | 0.45 | 0.04 | 1.01 | 0.16 | 0.31 | 0.19 | 39.51 | 0.52 | 0.31 | 0.82 | 0.3 | 0.91 | 0.36 | 0.32 | 0.32 | 5.38 | 23.84 | ME |
| S7_vs_30665 | 0 | 0 | 0 | 0.64 | 0.01 | 0.14 | 0.12 | 0.16 | 0.08 | 8.01 | 0.29 | 0.04 | 0.11 | 0.2 | 0.39 | 0.12 | 0.14 | 0.11 | 0.15 | 0.43 | ME |
| S7_vs_38113 | 0.01 | 0 | 0.01 | 1 | 0.11 | 1.2 | 0.03 | 0.54 | 0.17 | 317.08 | 0.8 | 4.09 | 0.2 | 0.3 | 0.53 | 0.61 | 0.34 | 0.31 | 0.59 | 0.75 | ME |
| S7_vs_38834 | 0 | 0 | 0 | 1.77 | 0.04 | 1.36 | 0.03 | 2.03 | 0.97 | 21.45 | 3.51 | 0.73 | 2.62 | 19.63 | 0.65 | 1.26 | 0.85 | 1.49 | 2.64 | 5.44 | ME |
| S7_vs_40259 | 0 | 0 | 0 | 0.5 | 0.01 | 0.31 | 0.06 | 0.26 | 0.12 | 17.16 | 0.51 | 0.09 | 0.4 | 0.38 | 0.39 | 0.39 | 0.42 | 0.3 | 0.34 | 0.7 | ME |
| S7_vs_41098 | 0 | 0 | 0.01 | 1.65 | 0.04 | 11.83 | 0.07 | 0.51 | 0.17 | 30.73 | 0.44 | 14.09 | 1.13 | 0.61 | 7.96 | 1.41 | 0.86 | 2.82 | 58.68 | 8.5 | Shared |
| S7_vs_41534 | 0 | 0 | 0 | 0.63 | 0.02 | 0.37 | 0.16 | 0.2 | 0.12 | 218.38 | 0.31 | 0.02 | 0.61 | 0.44 | 0.41 | 0.19 | 0.16 | 0.16 | 0.18 | 0.59 | ME |
| S7_vs_43529 | 0 | 0 | 0 | 1.77 | 0.08 | 0.55 | 0.05 | 0.24 | 0.22 | 20.01 | 0.54 | 0.92 | 0.21 | 0.58 | 1.58 | 0.39 | 0.66 | 0.18 | 0.56 | 0.75 | ME |
| S7_vs_47050 | 0 | 0 | 0.02 | 1.33 | 0.06 | 173.49 | 0.22 | 0.94 | 0.32 | 109.84 | 2.73 | 0.43 | 1.49 | 1.14 | 2.11 | 1.5 | 0.56 | 0.48 | 1.06 | 1.99 | ME |
| S7_vs_48523 | 0 | 0 | 0 | 8.47 | 0.26 | 6.93 | 2.4 | 1.51 | 3.82 | 36.57 | 4.78 | 5.21 | 1.04 | 1.16 | 0.43 | 2.26 | 2.32 | 0.48 | 2.5 | 1.53 | ME |
| S7_vs_48708 | 0 | 0 | 0 | 2.05 | 0.07 | 0.77 | 0.04 | 0.46 | 0.29 | 40.27 | 1.61 | 0.23 | 2.34 | 0.54 | 0.56 | 2.09 | 0.45 | 0.5 | 1.97 | 3.52 | ME |
| S7_vs_48832 | 0 | 0 | 0 | 0.84 | 0.06 | 1.34 | 0.14 | 0.36 | 0.2 | 49.75 | 1.86 | 0.12 | 0.46 | 0.43 | 0.35 | 0.38 | 4.75 | 0.47 | 0.84 | 1.78 | ME |
| S7_vs_49426 | 0 | 0 | 0 | 3.31 | 0.1 | 11.75 | 0.13 | 13.08 | 0.83 | 28.84 | 9.17 | 0.76 | 7.11 | 2.86 | 2.07 | 2.67 | 7.77 | 2.75 | 15.87 | 6.5 | Shared |
| S7_vs_50194 | 0 | 0 | 0 | 0.48 | 0.05 | 0.37 | 0.03 | 17.91 | 32.42 | 92.51 | 1.13 | 0.08 | 0.29 | 0.14 | 0.71 | 0.22 | 0.22 | 0.23 | 0.36 | 25.03 | Shared |
| S7_vs_51332 | 0 | 0 | 0 | 5.97 | 0.89 | 3.27 | 0.24 | 2.3 | 3.45 | 15.2 | 2.59 | 0.45 | 7.06 | 3.06 | 2.91 | 1.84 | 4.94 | 2.87 | 2.87 | 7.81 | ME |
| S7_vs_55031 | 0 | 0 | 0 | 0.62 | 0.01 | 0.21 | 0.31 | 0.13 | 0.07 | 22.38 | 1.3 | 0.04 | 0.57 | 0.31 | 0.42 | 0.14 | 0.25 | 0.11 | 0.47 | 5.92 | ME |
| S7_vs_55409 | 0.01 | 0 | 0 | 0.57 | 0.04 | 0.91 | 0.29 | 1.28 | 0.69 | 40.85 | 0.95 | 0.62 | 0.44 | 0.66 | 1.02 | 0.39 | 1.61 | 0.22 | 0.37 | 0.47 | ME |
| S7_vs_55526 | 0.04 | 0 | 0 | 1.44 | 0.05 | 0.31 | 0.43 | 0.31 | 0.13 | 27.32 | 1.82 | 0.11 | 0.74 | 0.13 | 1.01 | 0.3 | 0.16 | 0.23 | 0.34 | 0.35 | ME |
| S7_vs_56529 | 0 | 0 | 0 | 1.27 | 0.16 | 0.45 | 0.02 | 0.35 | 0.29 | 121.77 | 1033.08 | 0.1 | 0.32 | 0.51 | 0.72 | 0.37 | 1.39 | 0.26 | 7.72 | 1.6 | Shared |
| S7_vs_59465 | 0.01 | 0 | 0 | 3.34 | 0.03 | 0.91 | 0.07 | 0.32 | 0.42 | 37.01 | 2.29 | 0.08 | 0.42 | 0.14 | 0.45 | 1.01 | 0.23 | 0.35 | 0.46 | 0.37 | ME |
| S7_vs_69630 | 0.01 | 0 | 0 | 0.93 | 0.04 | 0.45 | 0.17 | 0.52 | 0.47 | 22.28 | 0.88 | 0.12 | 0.44 | 0.22 | 0.35 | 0.41 | 0.37 | 0.18 | 0.33 | 0.49 | ME |
| S7_vs_70265 | 0 | 0 | 0 | 0.45 | 0.06 | 0.24 | 0.01 | 0.21 | 0.1 | 334.6 | 0.43 | 0.08 | 0.35 | 0.29 | 0.71 | 0.24 | 0.17 | 0.18 | 0.39 | 112.96 | ME |
| S7_vs_73280 | 0.01 | 0 | 0 | 2.54 | 0.02 | 0.31 | 8.65 | 0.24 | 0.12 | 31.55 | 2.3 | 0.07 | 0.44 | 0.09 | 0.27 | 0.89 | 0.17 | 0.19 | 0.25 | 0.41 | ME |
| S7_vs_78507 | 0.02 | 0.01 | 0 | 2.16 | 0.16 | 4.28 | 0.67 | 11.09 | 0.3 | 12.49 | 0.79 | 5.46 | 0.34 | 31.47 | 0.36 | 0.68 | 69.74 | 1.19 | 24.67 | 2.73 | Shared |
| S7_vs_82140 | 0 | 0 | 0 | 0.36 | 0.04 | 0.32 | 0.08 | 0.3 | 0.12 | 28.56 | 0.41 | 0.06 | 6.65 | 0.23 | 0.45 | 1.06 | 0.29 | 2.77 | 0.36 | 0.72 | ME |
| S7_vs_83100 | 0 | 0 | 0 | 1.28 | 0.04 | 0.7 | 1.59 | 0.83 | 0.62 | 28.19 | 8.42 | 0.94 | 0.24 | 0.64 | 0.26 | 0.24 | 9.75 | 0.16 | 1 | 1.51 | ME |
| S7_vs_83474 | 0 | 0 | 0 | 0.74 | 0.43 | 0.35 | 0.11 | 0.29 | 0.05 | 11.18 | 0.89 | 0.09 | 0.62 | 1.19 | 0.48 | 0.27 | 0.53 | 0.19 | 1.91 | 0.25 | ME |
| S7_vs_86377 | 0 | 0 | 0 | 1.15 | 0.12 | 3.17 | 0.08 | 0.71 | 0.17 | 142.16 | 0.98 | 0.71 | 0.48 | 17.88 | 1.16 | 0.58 | 0.54 | 0.49 | 0.78 | 1.2 | ME |
| S7_vs_87279 | 0.01 | 0 | 0.01 | 3.41 | 0.1 | 2.39 | 0.27 | 0.23 | 0.13 | 24.12 | 1.26 | 0.12 | 1.55 | 1.46 | 0.97 | 0.97 | 0.21 | 0.27 | 3.22 | 10.68 | ME |
| S7_vs_87439 | 0 | 0 | 0 | 4.74 | 0.09 | 11.56 | 0.78 | 0.66 | 0.47 | 157.71 | 1.19 | 1.14 | 0.76 | 6.45 | 0.91 | 1.29 | 18.44 | 0.18 | 2.99 | 0.5 | ME |
| S7_vs_90915 | 0 | 0 | 0 | 0.37 | 0.02 | 0.29 | 0.07 | 0.44 | 0.19 | 22.01 | 0.43 | 0.09 | 0.42 | 0.28 | 0.36 | 0.4 | 0.72 | 0.23 | 0.54 | 0.59 | ME |
| S7_vs_97737 | 0.02 | 0 | 0 | 3.99 | 0.13 | 27.8 | 3.77 | 22.65 | 2.18 | 50.57 | 7.2 | 2.21 | 6.14 | 1.55 | 5.05 | 2.76 | 7.16 | 6.59 | 7.9 | 32.16 | ME |
| S7_vs_100640 | 0 | 0 | 0 | 1.08 | 0.01 | 0.37 | 0.12 | 0.2 | 0.17 | 11.7 | 0.49 | 0.09 | 0.71 | 0.51 | 0.31 | 0.24 | 0.86 | 0.26 | 0.78 | 0.97 | ME |
| S7_vs_101751 | 0.01 | 0 | 0 | 1.36 | 0.11 | 0.73 | 0.15 | 418.93 | 1.76 | 23.3 | 1.29 | 0.13 | 1.26 | 5.74 | 1.04 | 1.3 | 84.77 | 0.34 | 0.41 | 1.56 | ME |
| S7_vs_107876 | 0 | 0 | 0 | 0.53 | 103.56 | 0.53 | 0.07 | 3.6 | 5.84 | 149.85 | 0.5 | 0.18 | 0.51 | 167.8 | 0.5 | 0.5 | 0.5 | 0.49 | 51.7 | 0.74 | Shared |

| | | | | | | | | | | | | | | | | | | | | | |
|---|---|---|---|---|---|---|---|---|---|---|---|---|---|---|---|---|---|---|---|---|---|
| S7_vs_111831 | 0.02 | 0 | 0 | 0.89 | 0.03 | 2.58 | 4.54 | 0.42 | 0.09 | 12.88 | 0.52 | 0.22 | 0.29 | 0.58 | 0.49 | 0.34 | 0.46 | 0.73 | 3.29 | 8.57 | ME |
| S7_vs_118295 | 0.01 | 0 | 0 | 0.97 | 0.11 | 0.74 | 0.15 | 0.8 | 0.3 | 10.56 | 2.72 | 0.54 | 0.24 | 2.25 | 1.71 | 0.49 | 5.98 | 0.24 | 1.13 | 5.57 | ME |
| S7_vs_119492 | 0 | 0 | 0 | 0.32 | 0.09 | 0.43 | 0.01 | 29.4 | 140.96 | 93.58 | 0.43 | 0.09 | 0.53 | 9.38 | 0.5 | 0.21 | 0.29 | 0.21 | 0.23 | 0.59 | Shared |
| S7_vs_119679 | 0 | 0 | 0.01 | 7.73 | 0.07 | 12.88 | 0.47 | 38.84 | 9.12 | 16.41 | 11.2 | 1.05 | 1.09 | 4.47 | 4.9 | 1.46 | 5.28 | 15.27 | 7.11 | 5.1 | Shared |
| S7_vs_122151 | 0.01 | 0 | 0 | 1.45 | 0.02 | 2.38 | 0.04 | 0.27 | 0.1 | 16.17 | 0.3 | 0.04 | 0.42 | 0.2 | 0.41 | 0.81 | 1.09 | 0.29 | 2.45 | 0.41 | ME |
| S7_vs_129858 | 0 | 0 | 0 | 0.7 | 0.01 | 0.23 | 0.02 | 0.24 | 0.06 | 11.73 | 0.54 | 0.06 | 0.29 | 1.71 | 0.35 | 0.27 | 0.57 | 0.16 | 0.26 | 0.44 | ME |
| S7_vs_134583 | 0.01 | 0 | 0 | 1.07 | 0.04 | 3.17 | 2.77 | 4.96 | 11.66 | 54.78 | 0.95 | 0.18 | 0.45 | 10.38 | 42.93 | 0.42 | 0.22 | 6.18 | 0.4 | 10.41 | Shared |
| S7_vs_72737 | 0 | 0 | 0 | 7.61 | 1.61 | 12.69 | 1.45 | 33.33 | 3.08 | 59.7 | 3.41 | 1.75 | 5.22 | 24.19 | 3.72 | 4.95 | 24.47 | 0.5 | 9.23 | 2.06 | ME |
| S16_vf_1037 | 0 | 0 | 0 | 3.8 | 0.17 | 2.59 | 0.49 | 0.25 | 0.16 | 0.21 | 0.65 | 0.36 | 0.83 | 0.5 | 3.71 | 1.67 | 0.63 | 0.46 | 1251.41 | 1.05 | SHHC |
| S16_vf_5270 | 0 | 0 | 0 | 0.61 | 0.03 | 0.42 | 0.09 | 0.25 | 0.12 | 0.13 | 0.5 | 0.07 | 0.47 | 0.52 | 0.64 | 0.43 | 0.8 | 0.34 | 37.62 | 0.74 | SHHC |
| S16_vf_5809 | 0 | 0 | 0 | 0.94 | 0.13 | 2.11 | 0.05 | 0.22 | 0.14 | 0.16 | 0.45 | 0.1 | 0.42 | 0.38 | 14.88 | 0.25 | 0.28 | 0.24 | 89.43 | 0.68 | SHHC |
| S16_vf_6102 | 0.01 | 0 | 0 | 59.82 | 0.05 | 0.36 | 0.07 | 0.47 | 0.21 | 0.36 | 19.05 | 0.15 | 42.7 | 0.21 | 0.63 | 18.86 | 0.24 | 0.43 | 130.21 | 0.72 | Shared |
| S16_vf_9270 | 0 | 0 | 0 | 0.44 | 0.02 | 0.51 | 0.04 | 0.18 | 0.13 | 0.09 | 0.58 | 1.87 | 1.36 | 1.04 | 1.28 | 32.52 | 1.15 | 1.01 | 8658.84 | 2.6 | Shared |
| S16_vf_12551 | 0 | 0 | 0 | 5.28 | 0.02 | 0.28 | 0.09 | 0.16 | 0.07 | 0.13 | 1.31 | 0.07 | 0.36 | 0.22 | 0.38 | 0.74 | 0.23 | 1.93 | 183.79 | 0.34 | SHHC |
| S16_vf_16182 | 0 | 0 | 0 | 36.02 | 0.01 | 0.19 | 0.25 | 0.12 | 0.05 | 0.06 | 0.17 | 0.22 | 0.14 | 0.17 | 0.15 | 4.97 | 0.14 | 0.21 | 16.63 | 33.55 | Shared |
| S16_vf_32359 | 0 | 0 | 0 | 1.53 | 0.03 | 0.37 | 0.08 | 0.21 | 0.14 | 0.25 | 22.02 | 0.2 | 0.3 | 0.79 | 0.32 | 0.59 | 0.47 | 0.2 | 158.96 | 0.45 | SHHC |
| S16_vf_33218 | 0.14 | 0 | 0 | 0.8 | 3.18 | 83.19 | 145.5 | 8.86 | 2.27 | 3.4 | 16.34 | 7.17 | 1.36 | 0.98 | 1.7 | 0.47 | 4310.88 | 0.82 | 34.86 | 2015.52 | Shared |
| S16_vf_36146 | 0 | 0 | 0 | 5.8 | 0.1 | 3.57 | 1.52 | 10.21 | 0.13 | 0.85 | 2.46 | 0.2 | 1.33 | 5.19 | 3.53 | 1.73 | 4.05 | 1.09 | 11.95 | 3.88 | Shared |
| S16_vf_52513 | 0 | 0 | 0 | 2.95 | 0.05 | 2.46 | 0.13 | 10.85 | 0.4 | 1.6 | 134.66 | 1.22 | 1.1 | 0.69 | 2.42 | 3.6 | 4.53 | 1.4 | 10.78 | 1.56 | Shared |
| S16_vf_84462 | 0.06 | 0 | 0 | 92.09 | 0.05 | 2.86 | 0.11 | 0.46 | 0.46 | 0.24 | 2.9 | 0.15 | 0.51 | 0.23 | 0.65 | 43.42 | 0.39 | 0.51 | 76.28 | 163.35 | Shared |
| S16_vf_87190 | 0 | 0 | 0 | 0.75 | 0.04 | 0.72 | 0.41 | 0.29 | 0.23 | 0.19 | 0.93 | 0.14 | 0.95 | 0.65 | 0.85 | 0.65 | 1.95 | 0.52 | 62.96 | 1.28 | SHHC |
| S16_vf_90328 | 0 | 0 | 0 | 1.11 | 0.71 | 1.61 | 0.36 | 0.94 | 0.63 | 0.47 | 2.27 | 0.43 | 2.48 | 1.05 | 2.29 | 2.32 | 1.06 | 1.07 | 43.58 | 1.9 | SHHC |
| S16_vf_94866 | 0.03 | 0 | 0.1 | 0.77 | 0.02 | 857.11 | 0.2 | 0.19 | 0.16 | 0.15 | 0.82 | 2.18 | 0.66 | 0.75 | 0.9 | 3.46 | 0.81 | 0.55 | 4513.82 | 1.38 | Shared |
| S16_vf_96483 | 0 | 0 | 0 | 13.8 | 0.03 | 1.56 | 0.45 | 1.36 | 0.15 | 0.15 | 1 | 0.14 | 1.06 | 0.41 | 0.36 | 5.55 | 0.35 | 0.47 | 19.58 | 42.19 | Shared |
| S16_vf_97279 | 0 | 0 | 0 | 0.61 | 0.02 | 0.32 | 0.08 | 0.27 | 0.12 | 10.31 | 0.68 | 0.08 | 0.43 | 0.76 | 69.85 | 6.08 | 0.32 | 0.33 | 68.64 | 3.56 | Shared |
| S16_vf_100235 | 0 | 0 | 0 | 0.48 | 0.04 | 0.25 | 0.08 | 0.15 | 0.09 | 0.1 | 0.24 | 0.56 | 0.58 | 0.87 | 0.42 | 1.89 | 0.49 | 0.38 | 2513.66 | 0.88 | SHHC |
| S16_vf_100378 | 0 | 0 | 0 | 0.31 | 0.02 | 0.14 | 0.75 | 0.16 | 0.08 | 0.06 | 0.31 | 0.05 | 0.21 | 0.03 | 0.12 | 0.1 | 0.07 | 0.13 | 27.27 | 0.21 | SHHC |
| S16_vf_100473 | 0.01 | 0.02 | 0 | 113.58 | 2.74 | 1.29 | 0.02 | 0.07 | 0.08 | 3.7 | 0.33 | 0.33 | 0.19 | 6.12 | 0.16 | 1.12 | 0.1 | 0.21 | 64.53 | 6.98 | Shared |
| S16_vf_108554 | 0 | 0 | 0 | 10.64 | 1.27 | 0.45 | 0.22 | 0.67 | 0.18 | 0.19 | 0.39 | 0.11 | 0.41 | 0.32 | 0.37 | 4.77 | 0.5 | 0.32 | 115.83 | 0.67 | Shared |
| S16_vf_119834 | 0 | 0 | 0 | 0.32 | 0.03 | 0.2 | 0.07 | 0.12 | 0.06 | 0.09 | 0.51 | 0.04 | 0.77 | 0.31 | 0.33 | 0.26 | 0.31 | 0.13 | 26.39 | 0.45 | SHHC |
| S16_vf_147326 | 0 | 0 | 0 | 0.27 | 0.02 | 0.41 | 0.11 | 0.19 | 0.09 | 0.17 | 0.3 | 1.63 | 1.35 | 0.93 | 1.02 | 4.84 | 1.03 | 0.94 | 5924.87 | 1.83 | SHHC |
| S16_vf_151450 | 1.04 | 0.09 | 1.99 | 139.3 | 2.7 | 22255.25 | 0.08 | 2.97 | 3.07 | 4.99 | 564.94 | 36.02 | 4.85 | 4.18 | 2349.41 | 727.54 | 6.22 | 1469 | 30278.91 | 10.22 | Shared |
| S16_vf_164913 | 0 | 0 | 0 | 0.68 | 0.12 | 1.02 | 0.15 | 426.45 | 644.63 | 0.9 | 80.92 | 0.11 | 1.45 | 1.46 | 0.66 | 0.75 | 0.29 | 0.46 | 28.65 | 1.04 | Shared |
| S16_vf_169895 | 0 | 0 | 0 | 0.38 | 45.34 | 0.71 | 0.02 | 0.34 | 0.11 | 31.75 | 1.38 | 0.3 | 0.86 | 0.11 | 0.84 | 0.85 | 0.54 | 0.42 | 420.9 | 2.44 | Shared |
| S16_vf_169985 | 0.01 | 0 | 0 | 0.31 | 0.03 | 19.6 | 0.11 | 1.92 | 0.11 | 0.12 | 0.4 | 1.79 | 2518.18 | 0.86 | 36.24 | 27.3 | 0.69 | 0.61 | 1774.9 | 1.56 | Shared |
| S16_vf_172897 | 0.01 | 0 | 0 | 0.42 | 0.02 | 0.21 | 0.06 | 0.21 | 0.08 | 0.12 | 0.43 | 2.03 | 1.19 | 1.15 | 1.29 | 7.31 | 1.04 | 0.9 | 9540.2 | 2.25 | SHHC |
| S16_vf_173095 | 0 | 0 | 0 | 0.37 | 0.05 | 0.55 | 0.12 | 0.4 | 0.31 | 0.14 | 7.74 | 0.37 | 1.45 | 0.52 | 0.9 | 1.36 | 0.62 | 1.35 | 998.42 | 1.24 | SHHC |
| S16_vf_173099 | 0 | 0 | 0 | 0.67 | 0.04 | 0.46 | 0.36 | 0.28 | 0.13 | 1.01 | 0.71 | 0.14 | 0.46 | 0.24 | 0.28 | 1.73 | 1.64 | 0.22 | 87.37 | 0.65 | SHHC |
| S16_vf_173109 | 0 | 0 | 0 | 0.45 | 0.02 | 0.22 | 0.07 | 0.1 | 0.05 | 0.08 | 0.28 | 0.07 | 0.33 | 0.08 | 0.28 | 0.18 | 0.24 | 0.17 | 80.69 | 0.5 | SHHC |
| S16_vf_173133 | 0.02 | 0 | 0 | 1.21 | 0.01 | 0.27 | 0.2 | 0.2 | 0.07 | 0.15 | 0.64 | 0.06 | 0.3 | 0.36 | 0.21 | 0.12 | 0.32 | 0.12 | 22.88 | 0.44 | SHHC |
| S16_vs_15093 | 0.01 | 0 | 0 | 0.99 | 0.02 | 0.29 | 0.04 | 0.2 | 0.12 | 0.2 | 0.65 | 0.68 | 0.65 | 0.49 | 0.74 | 2.69 | 0.53 | 0.43 | 2969.8 | 1.22 | SHHC |
| S16_vs_20060 | 0 | 0 | 0 | 19.77 | 0.03 | 0.28 | 0.04 | 0.31 | 0.13 | 0.17 | 36.9 | 0.1 | 3.65 | 0.22 | 0.97 | 20.59 | 0.28 | 0.69 | 18.2 | 41 | Shared |
| S16_vs_59697 | 0 | 0 | 0 | 0.64 | 0.02 | 0.46 | 0.11 | 0.22 | 0.17 | 0.13 | 0.61 | 0.11 | 0.44 | 0.55 | 0.37 | 0.47 | 0.28 | 0.3 | 123.62 | 1.24 | SHHC |
| S16_vs_75108 | 0.01 | 0 | 0 | 0.84 | 0.03 | 0.43 | 0.26 | 1.39 | 2.62 | 11.35 | 3.97 | 5.69 | 0.46 | 0.82 | 0.67 | 10.28 | 0.25 | 0.22 | 378.71 | 0.45 | Shared |
| S16_vs_76823 | 0 | 0 | 0 | 3.79 | 0.02 | 0.31 | 0.11 | 0.27 | 0.13 | 0.21 | 30.51 | 0.1 | 0.28 | 0.15 | 0.17 | 0.39 | 0.2 | 0.21 | 58.88 | 0.71 | SHHC |
| S16_vs_78361 | 0.01 | 0 | 0 | 14.56 | 0.01 | 5.66 | 0.01 | 0.12 | 0.05 | 0.06 | 1.45 | 0.06 | 0.29 | 0.12 | 0.24 | 0.83 | 0.15 | 0.17 | 130.38 | 2.15 | Shared |
| S16_vs_78429 | 0 | 0 | 0 | 5.11 | 0.05 | 0.4 | 0.02 | 0.22 | 0.13 | 0.2 | 113.43 | 0.06 | 0.89 | 0.32 | 1.07 | 0.66 | 0.42 | 0.33 | 135.18 | 0.54 | SHHC |
| S16_vs_127905 | 0 | 0 | 0 | 19.59 | 0.16 | 4.42 | 0.4 | 0.61 | 0.54 | 0.45 | 1.03 | 0.37 | 2.45 | 3.03 | 1.61 | 17.88 | 2.17 | 6.09 | 53.99 | 161.61 | Shared |
| S16_vs_172749 | 0.01 | 0 | 0 | 8.32 | 0.03 | 0.19 | 0.08 | 0.25 | 0.09 | 0.19 | 0.35 | 0.07 | 7.17 | 0.1 | 0.28 | 1.73 | 0.12 | 1.18 | 30.75 | 0.29 | SHHC |
| S16_vs_173079 | 0 | 0 | 0 | 9.12 | 0.04 | 0.33 | 0.09 | 0.25 | 0.15 | 0.17 | 0.54 | 0.1 | 2.33 | 5.88 | 2.32 | 2.99 | 5.33 | 6.3 | 33.63 | 0.78 | SHHC |
| S16_vs_173096 | 0 | 0 | 0.01 | 14.74 | 10.39 | 0.65 | 0.01 | 0.26 | 0.12 | 0.87 | 0.65 | 0.59 | 0.46 | 0.73 | 0.56 | 3.32 | 0.51 | 0.57 | 2860.57 | 1.36 | Shared |
| S16_vs_173102 | 0.2 | 0 | 0.18 | 236.24 | 0.63 | 2620.06 | 162.99 | 536.59 | 855.2 | 6.29 | 1.73 | 3.74 | 1.24 | 2.2 | 0.54 | 37.1 | 0.33 | 0.69 | 76.47 | 181.93 | Shared |
| S16_vs_173141 | 0 | 0 | 0 | 0.52 | 0 | 0.28 | 0.06 | 0.13 | 0.16 | 0.04 | 0.4 | 0.06 | 0.11 | 38.91 | 0.48 | 0.14 | 0.07 | 0.11 | 79.75 | 1 | Shared |
| S16_vs_4662 | 0 | 0 | 0 | 0.87 | 0.08 | 0.49 | 0.04 | 0.3 | 0.15 | 0.24 | 0.48 | 0.13 | 0.49 | 0.16 | 0.32 | 0.62 | 0.32 | 0.16 | 61.64 | 0.6 | SHHC |
| S16_vs_10622 | 0 | 0 | 0 | 1.25 | 0.02 | 0.48 | 0.19 | 7.09 | 0.11 | 1.06 | 0.4 | 0.08 | 0.85 | 0.28 | 0.45 | 0.42 | 1.29 | 0.29 | 28.1 | 0.64 | SHHC |
| S16_vs_11649 | 0 | 0 | 0 | 0.26 | 0.04 | 0.43 | 0.11 | 0.58 | 0.92 | 0.13 | 10.71 | 0.13 | 0.54 | 0.34 | 0.34 | 1.32 | 0.34 | 0.3 | 360.76 | 0.79 | SHHC |
| S16_vs_17493 | 0 | 0 | 0 | 0.4 | 0.01 | 0.37 | 0.03 | 30.36 | 0.21 | 17.06 | 11.3 | 0.11 | 1.81 | 4.8 | 0.3 | 0.28 | 2.31 | 0.2 | 15.98 | 4.25 | Shared |
| S16_vs_18293 | 0 | 0 | 0 | 0.77 | 0.05 | 0.39 | 0.05 | 0.42 | 0.09 | 0.43 | 0.76 | 0.11 | 0.5 | 0.29 | 0.79 | 0.71 | 0.41 | 0.26 | 11.19 | 0.62 | SHHC |
| S16_vs_18440 | 0 | 0 | 0 | 1.83 | 0.07 | 5.26 | 1.8 | 7.29 | 0.7 | 0.16 | 1.28 | 0.19 | 0.66 | 0.2 | 0.7 | 0.62 | 1.49 | 0.23 | 57.52 | 4.04 | SHHC |
| S16_vs_19320 | 0 | 0 | 0.01 | 0.51 | 0.01 | 0.2 | 0.01 | 0.26 | 0.09 | 0.06 | 0.47 | 0.06 | 0.14 | 0.26 | 0.24 | 4.91 | 0.17 | 0.78 | 16.54 | 0.37 | SHHC |
| S16_vs_19968 | 0.01 | 0.01 | 0 | 27.63 | 10.53 | 8.85 | 0.25 | 12.99 | 6.35 | 7.42 | 43.66 | 8.45 | 8.12 | 2.59 | 23.32 | 8.42 | 1.89 | 13.36 | 87.81 | 13.22 | Shared |
| S16_vs_26078 | 0 | 0 | 0 | 12.83 | 0.02 | 1 | 0.09 | 1.22 | 0.17 | 1.17 | 0.97 | 0.97 | 0.27 | 9.83 | 0.33 | 2.93 | 5.24 | 0.63 | 334.28 | 23.25 | Shared |
| S16_vs_32764 | 0.01 | 0.01 | 0 | 13.34 | 6.92 | 16.55 | 14.25 | 23.64 | 14.34 | 14.23 | 92.69 | 14.43 | 47.44 | 9.3 | 45 | 9.47 | 41.16 | 36.16 | 91.02 | 13.18 | Shared |
| S16_vs_33281 | 0.02 | 0.01 | 0.04 | 3.15 | 0.85 | 26.38 | 0.16 | 3.76 | 3.8 | 2.33 | 21.9 | 541.44 | 12.57 | 1.67 | 12.22 | 4.69 | 0.54 | 8.82 | 25.85 | 20.86 | Shared |
| S16_vs_34021 | 0.03 | 0 | 0 | 3.92 | 0.57 | 2.18 | 0.12 | 0.44 | 0.24 | 7.13 | 11.57 | 3.17 | 8.7 | 1.89 | 1.23 | 3.99 | 129.23 | 3.08 | 38.41 | 2.31 | Shared |
| S16_vs_34751 | 0 | 0 | 0 | 0.53 | 0.05 | 0.43 | 4.17 | 41.66 | 39.68 | 15.11 | 0.48 | 0.19 | 0.21 | 1.72 | 0.69 | 0.4 | 12.32 | 0.52 | 44.94 | 3.6 | Shared |
| S16_vs_35031 | 0 | 0 | 0 | 1.75 | 0.51 | 1.69 | 0.12 | 0.55 | 0.38 | 1.12 | 3.54 | 0.72 | 0.53 | 1.67 | 0.63 | 0.54 | 2.33 | 0.29 | 18.71 | 0.94 | SHHC |
| S16_vs_38863 | 0 | 0.01 | 0 | 0.46 | 0.07 | 5.81 | 0.13 | 0.35 | 0.18 | 4.11 | 0.68 | 0.14 | 9.29 | 0.25 | 1.19 | 0.78 | 0.45 | 9.53 | 188.36 | 0.97 | SHHC |
| S16_vs_48714 | 0 | 0 | 0 | 1.89 | 0.04 | 3.26 | 0.22 | 0.32 | 0.24 | 3.59 | 6.14 | 2.52 | 3.38 | 5.16 | 1.76 | 2 | 5.59 | 3.01 | 29.85 | 1.9 | SHHC |
| S16_vs_51901 | 0 | 0 | 0 | 0.72 | 0.01 | 0.21 | 0.07 | 0.36 | 0.52 | 0.09 | 0.5 | 0.35 | 0.37 | 0.25 | 0.4 | 1.08 | 0.23 | 0.3 | 1465.95 | 0.88 | SHHC |
| S16_vs_54250 | 0.01 | 0 | 0 | 0.58 | 0.24 | 6.8 | 0.21 | 1.28 | 3.43 | 0.4 | 3.07 | 0.31 | 2.92 | 1.69 | 3.15 | 1.68 | 1.49 | 1.37 | 37.6 | 2.59 | SHHC |
| S16_vs_54616 | 0.02 | 0 | 0 | 0.42 | 0.02 | 0.27 | 0.07 | 0.11 | 0.06 | 0.6 | 1.33 | 0.06 | 0.2 | 0.25 | 1.95 | 0.08 | 0.15 | 0.17 | 21.4 | 0.35 | SHHC |
| S16_vs_64584 | 0 | 0 | 0 | 1.67 | 0.02 | 0.38 | 1.28 | 0.3 | 0.24 | 0.29 | 1.25 | 0.13 | 3.21 | 0.25 | 1.21 | 1.38 | 17.93 | 4.46 | 25.63 | 0.51 | Shared |
| S16_vs_80244 | 0 | 0.01 | 0.01 | 15.36 | 0.16 | 26.72 | 0.43 | 0.67 | 0.38 | 0.59 | 1.31 | 0.5 | 21.69 | 0.65 | 1.06 | 5.31 | 2.06 | 1.28 | 21.57 | 52.13 | Shared |
| S16_vs_81805 | 0.01 | 0 | 0.01 | 1.94 | 0.04 | 1.27 | 0.06 | 1.24 | 0.21 | 1.03 | 0.43 | 1.54 | 0.31 | 0.51 | 0.4 | 0.3 | 6.6 | 0.2 | 24.6 | 0.58 | SHHC |
| S16_vs_82116 | 0 | 0 | 0 | 7.67 | 2.95 | 25.77 | 1.92 | 96.92 | 11.51 | 162.89 | 8.64 | 3.86 | 12.83 | 20.5 | 2.68 | 3.89 | 35.92 | 0.61 | 42.15 | 1.92 | Shared |
| S16_vs_86116 | 0 | 0 | 0 | 1.03 | 0.02 | 0.4 | 0.03 | 0.24 | 0.17 | 2.35 | 0.25 | 0.1 | 0.63 | 26.92 | 1.05 | 0.36 | 2.5 | 0.47 | 35.1 | 0.95 | Shared |
| S16_vs_89511 | 0 | 0 | 0 | 7.23 | 0.17 | 4.18 | 0.08 | 17.71 | 8.07 | 6.65 | 4.38 | 2.06 | 7.68 | 4.26 | 5.28 | 5.3 | 4.01 | 8.43 | 20.45 | 10.7 | Shared |
| S16_vs_92150 | 0.01 | 0 | 0 | 0.68 | 0.06 | 1.1 | 0.1 | 0.66 | 0.09 | 0.51 | 0.49 | 0.06 | 0.54 | 0.29 | 0.21 | 0.27 | 2.32 | 0.23 | 19.98 | 0.37 | SHHC |
| S16_vs_101072 | 0 | 0 | 0 | 1.97 | 0.05 | 7.72 | 0.08 | 0.33 | 0.18 | 0.2 | 0.3 | 2.03 | 0.69 | 0.14 | 0.26 | 0.54 | 0.3 | 0.31 | 12.86 | 1.06 | SHHC |
| S16_vs_101671 | 0.02 | 0 | 0 | 1.85 | 0 | 19.38 | 0 | 0.23 | 0.08 | 0.04 | 0.14 | 0.02 | 0.1 | 0.02 | 0.14 | 0.3 | 0.11 | 0.13 | 20.83 | 0.52 | Shared |
| S16_vs_103726 | 0 | 0 | 0 | 0.72 | 0.06 | 2.29 | 0.18 | 2.76 | 0.44 | 1.36 | 2.15 | 0.81 | 0.68 | 0.4 | 0.38 | 0.67 | 0.98 | 0.62 | 18.65 | 1.44 | SHHC |
| S16_vs_107235 | 0.02 | 0 | 0 | 3.21 | 0.1 | 4.5 | 0.1 | 0.84 | 0.52 | 4.93 | 8.09 | 2.7 | 4.85 | 6.45 | 1.32 | 2.18 | 6.73 | 3.11 | 35.07 | 2.81 | SHHC |
| S16_vs_116956 | 0 | 0 | 0 | 2.04 | 0.28 | 7.25 | 0.11 | 0.37 | 1.4 | 0.46 | 1.02 | 0.22 | 0.91 | 0.46 | 0.95 | 0.96 | 0.38 | 0.7 | 17.12 | 1.83 | SHHC |

| | | | | | | | | | | | | | | | | | | | | | |
|---|---|---|---|---|---|---|---|---|---|---|---|---|---|---|---|---|---|---|---|---|---|
| S16_vs_118029 | 0 | 0 | 0 | 0.6 | 0.08 | 1.34 | 0.14 | 0.32 | 0.25 | 2.55 | 0.56 | 0.74 | 0.77 | 0.32 | 0.63 | 0.51 | 0.47 | 0.42 | 192.8 | 2.49 | SHHC |
| S16_vs_120149 | 0 | 0 | 0 | 0.55 | 0.02 | 0.56 | 0.13 | 0.23 | 0.16 | 0.18 | 0.41 | 0.07 | 1.19 | 0.48 | 0.69 | 0.49 | 0.27 | 0.34 | 12.58 | 0.84 | SHHC |
| S16_vs_120605 | 0 | 0 | 0 | 0.1 | 0 | 0.13 | 0.55 | 0.05 | 0.01 | 0.04 | 0.23 | 0.14 | 0.21 | 0.01 | 0.08 | 0.25 | 0.11 | 0 | 7.07 | 0.09 | SHHC |
| S16_vs_120622 | 0 | 0 | 0 | 0.49 | 0.01 | 0.22 | 0.05 | 4.36 | 1.03 | 0.21 | 0.32 | 0.05 | 0.22 | 0.14 | 3.08 | 0.12 | 3.63 | 0.2 | 68.32 | 0.64 | SHHC |
| S16_vs_124703 | 0 | 0 | 0 | 23.37 | 0.07 | 0.55 | 0.02 | 0.29 | 0.13 | 0.81 | 7.74 | 0.17 | 0.49 | 0.26 | 0.25 | 5.06 | 0.14 | 0.23 | 304.41 | 0.5 | Shared |
| S16_vs_127669 | 1.09 | 0.23 | 1.49 | 28.34 | 2.61 | 18273.7 | 9.93 | 5.95 | 3.88 | 1575.51 | 197.36 | 26.14 | 200.3 | 28.03 | 56.79 | 880.54 | 26.06 | 3867.01 | 81.62 | 18.03 | Shared |
| S16_vs_132979 | 0 | 0 | 0.01 | 1.83 | 0.11 | 1.38 | 2.35 | 4.98 | 7.52 | 1.83 | 3.25 | 1.07 | 1.34 | 0.12 | 2.92 | 0.56 | 4.12 | 0.7 | 11.13 | 17.85 | Shared |
| S16_vs_136935 | 0 | 0 | 0 | 15.93 | 0.2 | 20.73 | 6.56 | 1.92 | 0.33 | 4.63 | 1.58 | 0.61 | 7.62 | 1.94 | 0.97 | 4.87 | 215.52 | 5.37 | 35.26 | 80.05 | Shared |
| S16_vs_137222 | 0 | 0 | 0 | 0.56 | 0.04 | 0.46 | 0.08 | 0.24 | 0.11 | 0.13 | 0.45 | 0.09 | 0.62 | 0.26 | 0.3 | 0.24 | 0.32 | 0.33 | 41.8 | 0.98 | SHHC |
| S16_vs_140468 | 0 | 0 | 0 | 4.71 | 0.57 | 18.17 | 0.07 | 0.58 | 4.5 | 2.11 | 2.31 | 0.76 | 3.62 | 3.46 | 2.58 | 3.12 | 0.34 | 2.63 | 18.01 | 4.85 | Shared |
| S16_vs_140709 | 0 | 0.01 | 0 | 55.25 | 0.16 | 0.42 | 0.08 | 0.25 | 0.12 | 0.19 | 0.88 | 0.21 | 0.52 | 0.64 | 13.96 | 7.98 | 0.34 | 1 | 26.85 | 60.25 | Shared |
| S16_vs_141218 | 0 | 0 | 0 | 1.79 | 0.06 | 2.51 | 0.29 | 0.78 | 0.36 | 0.45 | 24.79 | 1.34 | 3.84 | 12.17 | 3.87 | 17.15 | 2.06 | 2.01 | 4252.9 | 3.13 | Shared |
| S16_vs_142561 | 0.01 | 0 | 0.01 | 1.31 | 0.22 | 3.69 | 126.85 | 21.76 | 3.35 | 1.55 | 12.44 | 1.26 | 1.54 | 5.57 | 7.78 | 0.69 | 3.14 | 1.82 | 26.66 | 1.82 | Shared |
| S16_vs_165662 | 0 | 0 | 0 | 1.06 | 0.55 | 6.36 | 0.65 | 0.6 | 0.42 | 0.47 | 12.41 | 0.19 | 11.57 | 0.69 | 10.84 | 3.74 | 0.87 | 0.64 | 158.61 | 2.22 | SHHC |
| S10_vf_3644 | 0 | 0 | 0.01 | 0.28 | 0.02 | 34.85 | 0.09 | 0.44 | 0.11 | 0.14 | 8.29 | 0.4 | 378.39 | 0.21 | 2.11 | 30.97 | 0.24 | 0.72 | 0.35 | 0.65 | Shared |
| S10_vf_18671 | 0.01 | 0 | 0 | 14.68 | 12.56 | 19.6 | 38.48 | 17.87 | 28.51 | 154.07 | 1.36 | 0.26 | 116.97 | 16.27 | 0.48 | 41.21 | 0.27 | 0.21 | 46.26 | 0.55 | Shared |
| S10_vf_24791 | 0.01 | 0 | 0 | 0.53 | 0.02 | 0.25 | 0.1 | 0.21 | 0.07 | 0.08 | 0.57 | 0.12 | 44.39 | 0.24 | 0.28 | 3.4 | 0.21 | 0.22 | 0.23 | 0.52 | SHHC |
| S10_vf_25241 | 0 | 0 | 0 | 0.4 | 0 | 15.95 | 0.06 | 0.04 | 0.06 | 0.06 | 3.92 | 0.18 | 65.24 | 1.38 | 0.37 | 8.1 | 0.13 | 0.12 | 0.13 | 0.62 | Shared |
| S10_vf_36880 | 0 | 0.02 | 1.61 | 12.2 | 9.72 | 35.39 | 1147.09 | 10.3 | 20 | 45780.42 | 12600.25 | 27872.57 | 1470.28 | 15403.57 | 67.99 | 888.66 | 6.72 | 314.96 | 6.89 | 28.16 | Shared |
| S10_vf_43964 | 0 | 0.02 | 0 | 0.85 | 0.07 | 4.6 | 0.98 | 0.45 | 0.15 | 0.18 | 0.65 | 0.31 | 111.8 | 4.12 | 0.81 | 1.66 | 4.62 | 237.72 | 0.91 | 4.07 | Shared |
| S10_vf_46328 | 0 | 0 | 0 | 0.98 | 0.05 | 2.69 | 0.64 | 0.34 | 0.1 | 0.2 | 0.3 | 0.24 | 27.69 | 0.3 | 0.37 | 3.73 | 0.33 | 0.49 | 0.32 | 0.5 | SHHC |
| S10_vf_47170 | 0 | 0 | 0 | 2 | 0.02 | 12.78 | 0.04 | 0.52 | 0.35 | 0.55 | 6.64 | 0.11 | 57.87 | 20.74 | 0.42 | 7.96 | 0.19 | 0.32 | 0.45 | 1.21 | Shared |
| S10_vf_57705 | 0 | 0 | 0 | 0.49 | 0.02 | 0.47 | 0.04 | 0.43 | 0.12 | 0.11 | 0.85 | 0.41 | 392.42 | 0.35 | 1.92 | 2.9 | 0.24 | 4.82 | 0.41 | 0.73 | SHHC |
| S10_vf_62230 | 0 | 0 | 0 | 1.09 | 0.02 | 1.9 | 0.26 | 0.5 | 0.19 | 0.32 | 1.49 | 0.18 | 26.02 | 0.37 | 1.45 | 3.12 | 0.62 | 0.47 | 0.97 | 1.22 | SHHC |
| S10_vf_64371 | 0 | 0 | 0.35 | 3.03 | 0.62 | 4.13 | 0.09 | 5.45 | 0.1 | 0.12 | 0.35 | 4999.3 | 1898.08 | 1.07 | 1.36 | 37.99 | 1.02 | 0.69 | 1.51 | 2.25 | Shared |
| S10_vf_80712 | 0 | 0 | 0 | 4.57 | 0.04 | 18.35 | 0.1 | 4.98 | 0.35 | 0.3 | 36.29 | 0.37 | 105.84 | 192.77 | 1.36 | 19.18 | 3.18 | 0.65 | 0.77 | 1.41 | Shared |
| S10_vf_113489 | 0 | 0 | 0 | 0.51 | 0.03 | 0.29 | 0.07 | 0.23 | 0.1 | 0.15 | 0.29 | 0.09 | 20.76 | 0.17 | 0.16 | 3.46 | 0.15 | 0.17 | 0.15 | 0.52 | SHHC |
| S10_vf_120738 | 0 | 0.01 | 0.01 | 22.11 | 0.07 | 0.61 | 0.12 | 10.41 | 16.67 | 0.26 | 207.35 | 128.76 | 26.3 | 109.75 | 31.33 | 16.46 | 0.35 | 55.15 | 17.46 | 0.87 | Shared |
| S10_vs_43473 | 0 | 0 | 0 | 1.07 | 0.02 | 2.49 | 0.31 | 0.49 | 1.4 | 0.11 | 0.55 | 0.08 | 98.59 | 0.24 | 1.09 | 0.38 | 0.69 | 0.16 | 0.27 | 1.3 | SHHC |
| S10_vs_120489 | 0 | 0 | 0 | 4.64 | 0.02 | 0.8 | 0.14 | 0.44 | 0.2 | 0.3 | 0.86 | 0.26 | 33.21 | 0.62 | 0.61 | 4.41 | 0.3 | 0.37 | 0.99 | 0.71 | SHHC |
| S10_vs_120541 | 0 | 0 | 0 | 3.37 | 0.01 | 0.47 | 0.04 | 0.26 | 0.1 | 0.19 | 7.09 | 0.06 | 91.24 | 0.28 | 0.7 | 0.78 | 0.23 | 0.27 | 2.19 | 0.59 | SHHC |
| S10_vs_120570 | 0 | 0 | 0 | 1.52 | 0.2 | 0.32 | 0.06 | 0.2 | 0.07 | 0.66 | 9.32 | 0.16 | 41.86 | 0.27 | 0.3 | 0.66 | 22.38 | 1.43 | 2.98 | 0.87 | Shared |
| S10_vs_2538 | 0 | 0 | 0 | 0.56 | 0.03 | 0.39 | 0.11 | 0.44 | 0.22 | 0.25 | 0.68 | 0.55 | 227.09 | 0.16 | 0.51 | 28.74 | 0.27 | 0.38 | 1.27 | 12.95 | Shared |
| S10_vs_14557 | 0.02 | 0.01 | 0.01 | 13.28 | 13.2 | 64.01 | 0.51 | 19.14 | 8.89 | 10.13 | 19.76 | 1 | 162.3 | 9.83 | 14.44 | 25.83 | 58.94 | 50.57 | 44.96 | 56.62 | Shared |
| S10_vs_18901 | 0 | 0 | 0 | 0.9 | 0.04 | 0.39 | 0.03 | 0.26 | 0.15 | 0.16 | 0.42 | 0.08 | 15.97 | 0.63 | 0.48 | 2.33 | 0.21 | 0.2 | 0.51 | 0.63 | SHHC |
| S10_vs_20400 | 0.01 | 0 | 0 | 1.83 | 0 | 0.14 | 0.06 | 0.18 | 0.08 | 0.14 | 25.34 | 0.05 | 16.88 | 0.09 | 0.32 | 1.01 | 0.27 | 0.12 | 0.32 | 0.55 | SHHC |
| S10_vs_26365 | 0 | 0 | 0.01 | 2.41 | 0.02 | 105.56 | 0.06 | 0.42 | 0.37 | 0.12 | 50.64 | 0.39 | 37.61 | 0.71 | 1.28 | 1.16 | 0.25 | 4.39 | 454.7 | 0.87 | Shared |
| S10_vs_35854 | 0 | 0 | 0 | 0.45 | 0.04 | 0.71 | 0.23 | 0.79 | 0.14 | 0.11 | 0.37 | 0.37 | 669.68 | 3.36 | 0.49 | 7.15 | 2.91 | 3.79 | 0.67 | 11.18 | Shared |
| S10_vs_36407 | 0 | 0 | 0 | 0.97 | 0.03 | 0.99 | 0.15 | 1.18 | 0.09 | 5.78 | 6.2 | 1.03 | 16.51 | 1.14 | 0.73 | 4.32 | 1.02 | 0.32 | 0.71 | 1.12 | SHHC |
| S10_vs_36598 | 0.01 | 0 | 0 | 3.09 | 0.45 | 12.79 | 15.92 | 0.24 | 0.15 | 3.46 | 2.51 | 1.52 | 76.32 | 1.45 | 0.45 | 2.72 | 13.7 | 0.39 | 4.37 | 21.58 | Shared |
| S10_vs_37379 | 0 | 0 | 0.01 | 8.61 | 0.08 | 84.82 | 0.08 | 3.25 | 0.37 | 45.21 | 32.23 | 0.24 | 86.49 | 40.31 | 0.26 | 12.39 | 11.49 | 0.13 | 0.21 | 0.37 | Shared |
| S10_vs_37694 | 0.01 | 0.01 | 0 | 19.71 | 8.87 | 1.13 | 0.15 | 1.98 | 1.13 | 7.01 | 92.72 | 0.09 | 17.9 | 12.06 | 60.16 | 4.14 | 0.34 | 18.05 | 32.97 | 1.9 | Shared |
| S10_vs_46819 | 0.01 | 0 | 0.02 | 1.22 | 0.25 | 3.28 | 0.78 | 1.49 | 0.33 | 4.06 | 2.34 | 0.4 | 37.08 | 0.25 | 1.42 | 5.78 | 3.53 | 1.14 | 1.23 | 3.24 | SHHC |
| S10_vs_58014 | 0.01 | 0.01 | 0.01 | 47.32 | 2.83 | 26.93 | 3.46 | 29.56 | 30.43 | 25.11 | 19 | 4.85 | 335.82 | 4.13 | 5.16 | 28.65 | 63.69 | 52.31 | 30.52 | 26.87 | Shared |
| S10_vs_62776 | 0 | 0 | 0 | 1.66 | 0.43 | 0.73 | 2.83 | 0.32 | 0.18 | 0.22 | 0.92 | 0.16 | 33.45 | 1.54 | 0.71 | 2.2 | 29.78 | 1.21 | 2.95 | 3.22 | Shared |
| S10_vs_67471 | 0 | 0 | 0 | 0.51 | 0.01 | 8.55 | 1.99 | 0.18 | 0.09 | 3.5 | 5.03 | 0.1 | 24.35 | 0.15 | 0.23 | 0.39 | 0.24 | 3.98 | 2.57 | 3.71 | SHHC |
| S10_vs_71900 | 0.01 | 0.01 | 0 | 12.17 | 0.52 | 2.06 | 4.38 | 0.69 | 0.21 | 3.37 | 1.62 | 2.3 | 57.91 | 16.14 | 0.62 | 5.27 | 333.16 | 5.41 | 17.06 | 17.96 | Shared |
| S10_vs_77004 | 0 | 0 | 0 | 10.41 | 0.04 | 0.52 | 0.16 | 0.37 | 0.17 | 0.14 | 0.38 | 0.39 | 39.17 | 0.38 | 0.61 | 7.15 | 0.6 | 0.87 | 0.5 | 1.07 | Shared |
| S10_vs_106834 | 0 | 0 | 0 | 0.42 | 0.04 | 0.97 | 0.09 | 0.93 | 0.62 | 0.68 | 1.43 | 0.11 | 27.05 | 0.75 | 1.87 | 0.95 | 0.63 | 2.18 | 0.79 | 1.45 | SHHC |
| S10_vs_119298 | 0 | 0 | 0 | 0.24 | 0.05 | 0.35 | 0.11 | 1 | 2.62 | 0.12 | 19.05 | 0.06 | 18.85 | 0.23 | 0.35 | 1.62 | 0.41 | 0.27 | 3.28 | 0.9 | SHHC |
| S12_vf_55175 | 0.01 | 0 | 0.02 | 3.77 | 0.24 | 98.23 | 1.14 | 0.38 | 0.15 | 0.62 | 0.66 | 0.33 | 0.38 | 4.31 | 263.29 | 0.65 | 0.74 | 0.29 | 0.72 | 1.45 | Shared |
| S12_vf_8173 | 0 | 0 | 0 | 0.37 | 0.01 | 0.45 | 0.13 | 0.18 | 0.14 | 0.5 | 0.74 | 0.18 | 2.3 | 0.28 | 414.49 | 0.32 | 0.2 | 1.24 | 0.45 | 0.67 | SHHC |
| S12_vf_173666 | 0.01 | 0 | 0 | 0.51 | 0.02 | 0.22 | 0.01 | 0.25 | 0.12 | 0.79 | 0.78 | 0.07 | 0.48 | 12.16 | 54.65 | 0.24 | 0.14 | 0.24 | 17.88 | 0.51 | Shared |
| S12_vf_16374 | 0 | 0 | 0 | 0.58 | 0.08 | 1.19 | 555.55 | 0.94 | 0.43 | 7.65 | 3.06 | 0.17 | 20.86 | 40.96 | 38.18 | 3.89 | 2.78 | 30.2 | 1.89 | 12.56 | Shared |
| S12_vf_79666 | 0 | 0 | 0.14 | 4.75 | 0.22 | 1922.78 | 4.14 | 0.4 | 0.34 | 8.02 | 0.75 | 2.72 | 6 | 0.69 | 150.36 | 0.86 | 0.21 | 6.53 | 20.42 | 0.66 | Shared |
| S12_vf_48278 | 0 | 0 | 0 | 0.49 | 0.02 | 0.19 | 0.05 | 0.1 | 0.09 | 0.23 | 0.23 | 0.09 | 0.18 | 0.16 | 191.06 | 0.08 | 0.17 | 0.2 | 0.17 | 0.27 | SHHC |
| S12_vf_32615 | 0.08 | 0.05 | 0.1 | 503.98 | 779.27 | 1001.4 | 0.14 | 0.78 | 0.57 | 1.63 | 59.23 | 1.67 | 1.13 | 0.43 | 1107.68 | 1673.58 | 1.45 | 50.08 | 2.78 | 1.13 | Shared |
| S12_vf_174374 | 0 | 0 | 0 | 4.63 | 0.38 | 79.52 | 0.74 | 17.22 | 0.97 | 1.22 | 25.38 | 0.23 | 1.15 | 1.09 | 411.19 | 0.8 | 1.07 | 0.21 | 0.75 | 1.69 | Shared |
| S12_vf_105542 | 0 | 0 | 0 | 0.54 | 0.06 | 0.91 | 0.23 | 0.34 | 0.14 | 0.43 | 1.75 | 0.21 | 4.44 | 0.42 | 208.32 | 1.39 | 0.56 | 0.49 | 0.96 | 1.74 | SHHC |
| S12_vf_18902 | 0.02 | 0 | 0 | 0.69 | 0.04 | 15.31 | 0.42 | 0.2 | 0.09 | 3.72 | 1.39 | 0.97 | 0.81 | 4.45 | 4481.17 | 0.21 | 0.91 | 10.7 | 20.01 | 1.67 | Shared |
| S12_vf_128794 | 0 | 0 | 0 | 2.47 | 0.11 | 1.84 | 0.08 | 0.18 | 0.12 | 1.62 | 0.28 | 0.34 | 0.48 | 1.93 | 1423.81 | 0.53 | 1.49 | 1.96 | 0.36 | 0.76 | SHHC |
| S12_vf_145044 | 0.03 | 0.02 | 0 | 92.91 | 5.5 | 1.77 | 0.22 | 0.63 | 0.32 | 3.54 | 1.04 | 1.02 | 2.32 | 0.44 | 288.09 | 434.36 | 0.58 | 0.63 | 1.4 | 2.75 | Shared |
| S12_vf_10887 | 0 | 0 | 0 | 0.54 | 0.01 | 0.32 | 0.03 | 0.2 | 0.11 | 0.21 | 0.4 | 0.07 | 0.46 | 0.21 | 30.28 | 0.21 | 0.27 | 0.21 | 0.28 | 0.58 | SHHC |
| S12_vf_176437 | 0 | 0 | 0 | 0.75 | 0.03 | 0.95 | 0.01 | 0.24 | 0.09 | 1.06 | 0.4 | 0.38 | 0.77 | 0.37 | 1383.39 | 0.8 | 0.43 | 0.3 | 0.56 | 1.06 | SHHC |
| S12_vf_169796 | 0 | 0 | 0 | 7.19 | 0.03 | 0.52 | 0.1 | 0.76 | 1.21 | 0.31 | 4.14 | 1.02 | 1.16 | 601.61 | 62.04 | 50.95 | 0.44 | 0.74 | 8.45 | 0.98 | Shared |
| S12_vf_67909 | 0 | 0 | 0 | 0.77 | 0.03 | 1.03 | 0.21 | 0.41 | 0.24 | 3.52 | 0.97 | 0.36 | 1.6 | 0.55 | 429.93 | 0.61 | 0.52 | 0.53 | 0.62 | 2.87 | SHHC |
| S12_vf_60224 | 0 | 0 | 0 | 2.15 | 0.18 | 1.62 | 0.44 | 1.26 | 0.58 | 0.52 | 1.27 | 0.2 | 3.99 | 0.66 | 21.22 | 0.72 | 2.11 | 1.65 | 1.54 | 3.98 | SHHC |
| S12_vf_170614 | 0 | 0 | 0 | 0.37 | 0.01 | 0.2 | 0.01 | 0.12 | 0.03 | 0.6 | 0.34 | 0.04 | 0.34 | 0.07 | 27.05 | 0.16 | 0.12 | 0.2 | 0.14 | 0.34 | SHHC |
| S12_vf_61299 | 0 | 0 | 0 | 3.13 | 0.34 | 0.59 | 0.13 | 0.14 | 0.11 | 0.63 | 0.4 | 0.41 | 0.59 | 0.38 | 900.45 | 0.29 | 0.78 | 0.28 | 0.48 | 1.2 | SHHC |
| S12_vf_69136 | 0 | 0 | 0 | 0.55 | 0.01 | 0.38 | 0.14 | 0.22 | 0.15 | 0.2 | 0.37 | 0.11 | 0.5 | 0.4 | 128.18 | 0.16 | 0.39 | 0.35 | 0.3 | 1.07 | SHHC |
| S12_vf_187941 | 0 | 0 | 0 | 0.48 | 0.03 | 0.61 | 0.59 | 0.31 | 0.15 | 0.24 | 1.2 | 0.13 | 0.81 | 0.79 | 43.19 | 0.44 | 0.39 | 0.41 | 0.47 | 1.22 | SHHC |
| S12_vf_188819 | 0 | 0 | 0 | 0.77 | 0.06 | 0.84 | 0.35 | 0.43 | 0.23 | 0.62 | 1.79 | 0.21 | 1.31 | 0.51 | 433.5 | 0.65 | 0.78 | 0.63 | 0.8 | 1.36 | SHHC |
| S12_vf_188822 | 0 | 0 | 0 | 0.29 | 0.03 | 0.23 | 0.09 | 0.23 | 0.1 | 0.14 | 0.25 | 0.07 | 0.41 | 3.69 | 58.53 | 0.18 | 74.58 | 0.65 | 0.24 | 0.39 | Shared |
| S12_vf_188824 | 0 | 0 | 0 | 0.46 | 0.03 | 9.37 | 0.09 | 0.58 | 0.11 | 2.07 | 0.43 | 0.91 | 605.81 | 0.61 | 2504.75 | 6.52 | 0.69 | 1.21 | 27.06 | 1.35 | SHHC |
| S12_vf_188827 | 0 | 0 | 0 | 0.09 | 0 | 0 | 0 | 0.01 | 0.03 | 0.04 | 0.19 | 0.01 | 0.03 | 0.03 | 27.32 | 0.07 | 0.01 | 0 | 0.09 | 0.18 | SHHC |
| S12_vf_188830 | 0 | 0 | 0 | 0.58 | 0.04 | 0.35 | 0.01 | 0.4 | 0.1 | 0.26 | 0.42 | 0.07 | 0.23 | 0.18 | 58.08 | 0.37 | 0.43 | 0.26 | 0.45 | 0.47 | SHHC |
| S12_vf_188838 | 0 | 0 | 0 | 0.19 | 0.04 | 0.39 | 0.04 | 0.23 | 0.05 | 0.26 | 0.37 | 0.01 | 0.28 | 0.2 | 29.35 | 0.08 | 0.28 | 0.51 | 0.33 | 0.65 | SHHC |
| S12_vf_188846 | 0 | 0 | 0 | 2.18 | 0.01 | 0.2 | 0.12 | 0.25 | 0.12 | 0.1 | 0.35 | 0.05 | 0.13 | 0.12 | 34.97 | 0.37 | 0.15 | 0.13 | 0.49 | 0.82 | SHHC |
| S12_vf_173208 | 0 | 0 | 0 | 0.55 | 0.02 | 155.13 | 0.14 | 0.13 | 0.11 | 0.12 | 5.98 | 0.26 | 7.59 | 0.05 | 33.33 | 0.27 | 0.08 | 0.16 | 1.22 | 0.44 | Shared |
| S12_vf_181024 | 0.01 | 0 | 0 | 0.8 | 0.02 | 0.77 | 0.09 | 0.22 | 0.12 | 0.23 | 0.72 | 0.1 | 4.1 | 0.21 | 79.82 | 0.42 | 0.35 | 0.32 | 1.88 | 3.44 | SHHC |
| S12_vs_86928 | 0 | 0 | 0 | 0.81 | 0.01 | 0.76 | 0.1 | 6.55 | 0.12 | 0.31 | 0.97 | 0.14 | 0.75 | 0.31 | 93.35 | 0.22 | 0.88 | 0.24 | 0.36 | 0.59 | SHHC |
| S12_vs_87088 | 0.02 | 0.01 | 0 | 3.28 | 0.12 | 2.14 | 8.69 | 24.26 | 0.45 | 2.63 | 5.29 | 5.75 | 2.33 | 4.96 | 30.71 | 1.23 | 2.22 | 0.52 | 4.11 | 6.64 | Shared |
| S12_vs_79785 | 0 | 0 | 0 | 1.59 | 0.02 | 0.63 | 0.81 | 0.53 | 0.9 | 0.25 | 0.42 | 0.11 | 0.89 | 0.26 | 17.27 | 0.63 | 0.22 | 0.41 | 99.29 | 0.64 | SHHC |
| S12_vs_167155 | 0 | 0 | 0 | 1 | 0.02 | 0.4 | 0.06 | 0.22 | 0.12 | 0.15 | 0.53 | 0.08 | 0.31 | 0.24 | 27.07 | 0.37 | 0.21 | 0.37 | 0.31 | 0.61 | SHHC |

| | | | | | | | | | | | | | | | | | | | | | |
|---|---|---|---|---|---|---|---|---|---|---|---|---|---|---|---|---|---|---|---|---|---|
| S12_vs_1980 | 0 | 0 | 0.01 | 0.74 | 0.14 | 12.39 | 0.79 | 1.98 | 0.36 | 5.31 | 23.87 | 0.25 | 1.7 | 1.62 | 235.58 | 2.55 | 3.05 | 1.1 | 1.58 | 2.58 | Shared |
| S12_vs_111815 | 0 | 0 | 0 | 0.69 | 0.05 | 0.68 | 0.2 | 0.4 | 0.16 | 6.08 | 0.76 | 1.52 | 1.6 | 3.98 | 7454.59 | 0.5 | 1.57 | 0.99 | 1.29 | 2.8 | SHHC |
| S12_vs_183891 | 0 | 0 | 0 | 0.13 | 0.05 | 0.59 | 0.02 | 0.06 | 0.05 | 0.7 | 1.09 | 9.03 | 0.14 | 0.31 | 276.74 | 0.01 | 0.24 | 0.15 | 0.13 | 0.67 | SHHC |
| S12_vs_137069 | 0 | 0 | 0 | 51.06 | 0.01 | 0.25 | 0.06 | 0.21 | 0.1 | 4.67 | 0.86 | 1.5 | 2.35 | 0.23 | 16.64 | 78.96 | 0.1 | 0.29 | 0.29 | 0.69 | Shared |
| S12_vs_188820 | 0 | 0 | 0 | 0.61 | 0.03 | 0.6 | 0.1 | 0.56 | 0.23 | 0.36 | 2.23 | 0.15 | 2.82 | 0.75 | 105.91 | 0.58 | 1.1 | 0.34 | 0.52 | 0.89 | SHHC |
| S12_vs_188821 | 0 | 0 | 0 | 1.29 | 0.07 | 1.7 | 0.15 | 0.36 | 0.18 | 0.99 | 0.74 | 0.28 | 0.88 | 0.45 | 1058.2 | 0.59 | 0.56 | 0.47 | 1.01 | 1.08 | SHHC |
| S12_vs_188823 | 0 | 0 | 0 | 0.49 | 0.01 | 0.47 | 0.16 | 0.19 | 0.07 | 0.27 | 0.57 | 0.11 | 0.71 | 0.62 | 116.17 | 0.18 | 0.4 | 0.3 | 0.26 | 1.01 | SHHC |
| S12_vs_188825 | 0 | 0 | 0 | 1.37 | 0.04 | 1.14 | 0.03 | 0.22 | 0.1 | 1.67 | 0.45 | 0.08 | 0.33 | 0.25 | 83.12 | 0.35 | 0.19 | 0.22 | 10.78 | 0.54 | SHHC |
| S12_vs_188828 | 0 | 0 | 0 | 2.44 | 0.02 | 0.21 | 0.02 | 0.3 | 0.1 | 0.49 | 0.43 | 0.07 | 0.42 | 0.11 | 101.96 | 0.79 | 0.32 | 0.21 | 0.25 | 0.46 | SHHC |
| S12_vs_188832 | 0 | 0 | 0 | 0.46 | 0.06 | 0.66 | 0.06 | 0.27 | 0.31 | 0.4 | 1.46 | 0.1 | 0.7 | 2.8 | 350.41 | 0.44 | 0.43 | 0.38 | 0.82 | 1.21 | SHHC |
| S12_vs_188833 | 0 | 0 | 0 | 0.79 | 0.02 | 0.2 | 0.07 | 0.19 | 0.08 | 0.91 | 1 | 0.05 | 0.39 | 5.51 | 20.94 | 0.16 | 1.16 | 0.19 | 0.38 | 0.27 | SHHC |
| S12_vs_118154 | 0 | 0 | 0 | 0.34 | 0.02 | 0.26 | 0.21 | 0.21 | 0.16 | 0.19 | 0.57 | 0.1 | 0.42 | 0.1 | 123.61 | 0.2 | 0.19 | 0.27 | 0.47 | 0.5 | SHHC |
| S12_vs_8204 | 0 | 0 | 0 | 0.9 | 0.04 | 2.91 | 0.11 | 0.3 | 0.19 | 14.22 | 17.3 | 0.25 | 0.62 | 0.48 | 39.03 | 0.53 | 0.37 | 1.16 | 0.63 | 5.84 | Shared |
| S12_vs_71525 | 0 | 0 | 0.02 | 1.63 | 0.27 | 49.45 | 0.22 | 0.75 | 0.63 | 0.47 | 15.63 | 0.16 | 2.95 | 1.52 | 25.32 | 0.22 | 0.5 | 1.54 | 3.68 | 5.05 | Shared |
| S12_vs_142625 | 0.01 | 0.01 | 0 | 4.96 | 1.84 | 8.98 | 0.34 | 10.92 | 6.94 | 4.83 | 59.51 | 9.06 | 8.57 | 3.22 | 53.11 | 2.69 | 1.32 | 12.96 | 26.79 | 10.36 | Shared |
| S12_vs_48045 | 0 | 0 | 0 | 0.77 | 0.02 | 0.33 | 0.06 | 0.13 | 0.08 | 0.07 | 0.31 | 0.05 | 0.27 | 0.13 | 33.03 | 0.14 | 0.14 | 0.25 | 0.29 | 0.36 | SHHC |
| S12_vs_111286 | 0 | 0 | 0 | 0.7 | 0.02 | 0.23 | 0.08 | 0.13 | 0.09 | 0.23 | 0.29 | 0.08 | 0.33 | 0.16 | 147.21 | 0.24 | 0.22 | 0.24 | 0.23 | 0.46 | SHHC |
| S12_vs_150812 | 0 | 0 | 0 | 0.55 | 0.02 | 13.33 | 0.1 | 0.3 | 0.15 | 0.68 | 0.64 | 0.17 | 0.4 | 0.27 | 28.13 | 1.59 | 0.23 | 0.17 | 0.32 | 0.57 | Shared |
| S12_vs_151292 | 0.59 | 0.19 | 0.74 | 16.97 | 4.32 | 9419.23 | 4.65 | 20.61 | 7.59 | 620.8 | 79.79 | 16.1 | 111.1 | 21.16 | 112.68 | 463.73 | 53.18 | 2343.87 | 35.87 | 27.01 | Shared |
| S12_vs_183752 | 0 | 0 | 0 | 0.95 | 0.03 | 5.75 | 0.15 | 0.43 | 0.61 | 0.41 | 1.67 | 0.12 | 1.63 | 0.95 | 62.29 | 0.86 | 0.52 | 0.57 | 0.78 | 1.21 | SHHC |
| S12_vs_58148 | 0.02 | 0 | 0 | 2.21 | 0.24 | 1.46 | 0.06 | 0.42 | 0.17 | 0.89 | 0.82 | 0.15 | 0.78 | 0.2 | 367.06 | 0.49 | 0.73 | 0.3 | 0.49 | 0.61 | SHHC |
| S12_vs_26336 | 0.01 | 0 | 0 | 4.33 | 2.21 | 5.26 | 0.21 | 19.27 | 14.89 | 4.18 | 49.42 | 5.23 | 3.41 | 6.33 | 33.55 | 1.91 | 1.36 | 28.1 | 23.07 | 4.35 | Shared |
| S12_vs_81729 | 0 | 0 | 0 | 1.2 | 10.99 | 0.42 | 0.12 | 0.21 | 0.13 | 0.32 | 0.5 | 0.13 | 0.48 | 0.22 | 150.05 | 0.42 | 0.3 | 0.25 | 0.58 | 0.5 | SHHC |
| S12_vs_98175 | 0 | 0 | 0 | 3.94 | 0.13 | 0.33 | 0.04 | 0.21 | 0.11 | 0.26 | 1.28 | 0.13 | 0.93 | 2.19 | 92.57 | 0.69 | 0.39 | 0.29 | 0.86 | 2.99 | SHHC |
| S12_vs_58560 | 0 | 0 | 0 | 0.56 | 0.01 | 0.25 | 0.04 | 0.21 | 0.11 | 0.27 | 0.6 | 0.05 | 0.46 | 1.39 | 18.39 | 0.9 | 0.34 | 0.22 | 0.44 | 0.38 | SHHC |
| S12_vs_26847 | 0 | 0 | 0 | 1.3 | 0.52 | 1.74 | 0.07 | 0.19 | 0.11 | 0.36 | 0.44 | 0.08 | 0.57 | 0.38 | 267.72 | 0.3 | 0.25 | 0.18 | 20.29 | 0.76 | SHHC |
| S12_vs_106329 | 0 | 0 | 0 | 1.21 | 0.04 | 0.52 | 1.89 | 2.2 | 0.8 | 0.93 | 12.38 | 2.09 | 0.76 | 2.02 | 45.96 | 0.4 | 1.36 | 0.51 | 0.56 | 1.61 | SHHC |
| S12_vs_74280 | 0 | 0 | 0 | 0.48 | 0 | 32.76 | 0.17 | 0.2 | 0.09 | 0.32 | 15.83 | 0.04 | 5.05 | 3.47 | 27.28 | 0.09 | 0.11 | 0.66 | 0.2 | 0.28 | Shared |
| S12_vs_99092 | 0 | 0 | 0 | 1.14 | 0.24 | 2.75 | 0.08 | 0.51 | 0.31 | 0.28 | 0.56 | 0.09 | 1.03 | 0.59 | 21.27 | 0.54 | 0.27 | 0.43 | 0.96 | 2.8 | SHHC |
| S12_vs_99219 | 0 | 0 | 0 | 0.74 | 0.08 | 0.74 | 0.29 | 0.28 | 0.17 | 0.25 | 0.96 | 0.19 | 1.09 | 11.06 | 25.72 | 2.4 | 1.81 | 0.48 | 0.9 | 5.73 | Shared |
| S12_vs_138261 | 0 | 0 | 0 | 0.46 | 0.12 | 0.3 | 0.29 | 0.17 | 0.25 | 1.45 | 3.94 | 0.09 | 0.51 | 130.32 | 57.83 | 0.46 | 7.78 | 0.23 | 1.38 | 11.67 | Shared |
| S12_vs_121913 | 0 | 0 | 0 | 2.05 | 0.17 | 2.4 | 0.15 | 3.28 | 1.25 | 0.5 | 4.17 | 0.12 | 2.62 | 4.33 | 25.51 | 0.56 | 1.76 | 0.88 | 4.52 | 4.77 | SHHC |
| S12_vs_153780 | 0 | 0 | 0 | 0.75 | 0.02 | 0.37 | 0.12 | 0.27 | 0.15 | 2.84 | 3.08 | 0.07 | 0.33 | 0.81 | 28.93 | 0.34 | 0.23 | 1.18 | 0.4 | 1.19 | SHHC |
| S12_vs_75098 | 0.01 | 0 | 0 | 6.65 | 0.04 | 1.39 | 0.19 | 1.73 | 0.43 | 0.92 | 2.22 | 0.51 | 1.25 | 1.2 | 46.69 | 3.78 | 1.28 | 0.91 | 3.74 | 1.64 | SHHC |
| S12_vs_28661 | 0.01 | 0 | 0 | 1.25 | 0.01 | 0.21 | 0.02 | 0.23 | 0.17 | 0.8 | 4.16 | 0.09 | 0.19 | 0.18 | 45.48 | 2.16 | 0.2 | 0.22 | 0.26 | 0.54 | SHHC |
| S12_vs_5857 | 0.01 | 0 | 0.01 | 0.62 | 0.01 | 0.19 | 0.02 | 0.21 | 0.08 | 0.23 | 0.69 | 0.08 | 0.37 | 0.15 | 143.52 | 5.34 | 0.22 | 0.74 | 0.67 | 0.45 | SHHC |
| S12_vs_92868 | 0.01 | 0 | 0 | 1.92 | 0.17 | 3.02 | 0.07 | 459.82 | 719.49 | 1.79 | 0.94 | 1.19 | 1.08 | 6.94 | 635.59 | 1.28 | 19.69 | 0.71 | 2.44 | 1.44 | Shared |
| S12_vs_123620 | 0 | 0 | 0 | 1.07 | 0.9 | 2.51 | 0.43 | 0.82 | 0.33 | 1.09 | 86.98 | 0.44 | 185.57 | 0.63 | 793.42 | 5.47 | 2.26 | 0.8 | 18.48 | 2.35 | SHHC |
| S12_vs_93357 | 0 | 0 | 0 | 0.55 | 0.02 | 0.21 | 0.07 | 2.01 | 0.32 | 0.13 | 0.79 | 0.08 | 0.25 | 0.35 | 16.95 | 0.31 | 0.97 | 0.18 | 0.16 | 0.41 | SHHC |
| S12_vs_6836 | 0 | 0 | 0 | 0.64 | 0.02 | 1.31 | 0.23 | 0.42 | 0.28 | 0.39 | 1.67 | 0.26 | 2.26 | 1.04 | 30.01 | 1.3 | 0.77 | 5.33 | 2.82 | 1.76 | SHHC |
| S12_vs_163990 | 0 | 0 | 0 | 0.79 | 0.04 | 1 | 0.21 | 0.25 | 0.19 | 0.88 | 117.21 | 0.31 | 0.44 | 22.06 | 120.28 | 0.32 | 0.26 | 32.49 | 0.97 | 1.1 | Shared |
| S12_vs_164056 | 0.04 | 0 | 0.01 | 5.79 | 0.04 | 1.2 | 0.11 | 4.79 | 0.73 | 1.35 | 3.85 | 0.69 | 1.29 | 0.33 | 16.07 | 18.32 | 3.12 | 0.45 | 0.91 | 2.35 | Shared |
| S12_vs_141337 | 0.01 | 0 | 0.01 | 45.22 | 4.83 | 28.02 | 3.9 | 28.21 | 30.37 | 22.98 | 15.58 | 3.17 | 106.3 | 5.16 | 35.45 | 20.93 | 47.39 | 43.01 | 20.62 | 19.44 | Shared |
| S12_vs_70241 | 0 | 0 | 0 | 0.77 | 0 | 0.77 | 0.3 | 0.1 | 0.3 | 0.4 | 1.31 | 0.19 | 1.77 | 0.35 | 12.63 | 0.19 | 0.18 | 0.93 | 0.54 | 1.09 | SHHC |
| S12_vs_94042 | 0 | 0 | 0.01 | 14.68 | 0.02 | 4.61 | 0.02 | 0.39 | 0.13 | 7.67 | 24.58 | 0.12 | 2.83 | 0.27 | 29.3 | 7.08 | 0.22 | 3.3 | 10.88 | 0.84 | Shared |
| S12_vs_14467 | 0.01 | 0 | 0 | 1.41 | 0.03 | 18.63 | 0.18 | 0.29 | 0.2 | 0.2 | 0.71 | 0.15 | 3.16 | 2.52 | 36.66 | 0.38 | 2.5 | 0.68 | 0.57 | 0.96 | Shared |
| S12_vs_22853 | 0 | 0 | 0 | 0.84 | 0.03 | 0.45 | 0.08 | 0.23 | 0.14 | 0.25 | 0.43 | 0.06 | 0.41 | 0.41 | 30.86 | 0.21 | 0.19 | 0.3 | 0.34 | 0.6 | SHHC |
| S12_vs_133070 | 0.01 | 0 | 0 | 28.73 | 0.17 | 1.29 | 0.24 | 0.83 | 0.26 | 1.13 | 3.35 | 0.74 | 0.95 | 4.46 | 378.54 | 13.24 | 0.85 | 0.56 | 3.02 | 1.57 | Shared |
| S12_vs_172379 | 0.01 | 0 | 0 | 1.04 | 0.12 | 1.82 | 0.24 | 0.64 | 0.31 | 0.64 | 7.59 | 0.26 | 10.09 | 0.59 | 155.57 | 4.24 | 0.94 | 0.78 | 2.23 | 2.07 | SHHC |
| S12_vs_14927 | 0 | 0 | 0 | 27.22 | 0.07 | 2.65 | 0.3 | 0.66 | 0.38 | 2.66 | 1.63 | 0.73 | 3.05 | 2.94 | 1867.34 | 92.11 | 1.23 | 1.27 | 1.47 | 2.08 | Shared |
| S12_vs_23796 | 0.01 | 0 | 0 | 0.64 | 0.15 | 0.74 | 0.2 | 6.66 | 4.16 | 0.32 | 2.88 | 0.31 | 1.16 | 1.6 | 73.56 | 0.38 | 0.58 | 0.71 | 1.75 | 9.34 | SHHC |
| S12_vs_32669 | 0 | 0 | 0 | 0.73 | 0.03 | 0.34 | 0.01 | 0.2 | 0.11 | 0.22 | 0.64 | 0.1 | 0.67 | 0.31 | 105.07 | 0.31 | 0.25 | 0.32 | 0.38 | 0.6 | SHHC |
| S15_vf_2032 | 0 | 0 | 0 | 0.61 | 0.03 | 1.2 | 0.11 | 0.46 | 0.2 | 0.19 | 0.68 | 0.34 | 224.84 | 0.78 | 0.52 | 3.16 | 1.1 | 104.61 | 0.63 | 14.13 | Shared |
| S15_vf_2513 | 0 | 0 | 0 | 1.49 | 0.02 | 7.84 | 0.12 | 0.26 | 0.23 | 0.24 | 0.58 | 0.13 | 0.65 | 0.58 | 0.75 | 0.5 | 0.46 | 93.1 | 0.65 | 1.24 | SHHC |
| S15_vf_20644 | 0 | 0.01 | 0 | 3.13 | 0.02 | 0.48 | 0.07 | 0.26 | 0.11 | 0.15 | 0.55 | 0.12 | 0.42 | 0.25 | 0.47 | 6.8 | 0.93 | 259.66 | 2.52 | 4.95 | SHHC |
| S15_vf_33505 | 0.34 | 0.15 | 0.38 | 931.04 | 2557.69 | 4083.17 | 0.14 | 1.69 | 1.45 | 1.91 | 68.24 | 5.04 | 1.17 | 0.44 | 77.52 | 5433.36 | 3.64 | 757.13 | 7.49 | 1.09 | Shared |
| S15_vf_55639 | 0.01 | 0.02 | 0 | 0.33 | 0.01 | 0.11 | 0.04 | 0.09 | 0.08 | 0.1 | 0.15 | 0.06 | 0.21 | 0.25 | 0.2 | 0.16 | 0.32 | 56.9 | 0.21 | 0.48 | SHHC |
| S15_vf_58467 | 0 | 0.02 | 0 | 0.87 | 0.07 | 1.72 | 0.17 | 0.83 | 0.23 | 0.29 | 1.01 | 0.2 | 2.44 | 0.58 | 1.22 | 0.87 | 1.01 | 140.55 | 0.81 | 1.22 | SHHC |
| S15_vf_71616 | 0 | 0.04 | 0 | 0.87 | 0.01 | 0.61 | 0.07 | 0.23 | 0.12 | 0.16 | 0.5 | 0.16 | 12.36 | 0.21 | 2.86 | 0.54 | 0.25 | 143.88 | 0.31 | 0.73 | SHHC |
| S15_vf_74586 | 0 | 0.04 | 0 | 1 | 0.02 | 0.98 | 0.08 | 0.18 | 0.09 | 0.12 | 0.39 | 0.17 | 28.89 | 0.37 | 2.17 | 0.4 | 0.79 | 561.09 | 0.35 | 0.7 | SHHC |
| S15_vf_75924 | 0 | 0.11 | 0 | 3.55 | 0.16 | 0.75 | 0.2 | 0.79 | 0.38 | 749.04 | 2.16 | 0.83 | 94.05 | 89.92 | 1583.12 | 1.8 | 2.42 | 1696.07 | 43.34 | 102.84 | Shared |
| S15_vs_1407 | 0 | 0 | 0 | 0.38 | 0.02 | 2.64 | 2.14 | 0.17 | 0.04 | 0.08 | 0.23 | 0.1 | 0.8 | 0.5 | 3.65 | 0.36 | 0.31 | 227.97 | 0.18 | 0.49 | SHHC |
| S15_vs_7103 | 0 | 0.01 | 0 | 1.38 | 0.19 | 1.76 | 1.09 | 2.05 | 2.7 | 1.34 | 16.12 | 7.59 | 0.29 | 1.09 | 0.43 | 0.51 | 4.88 | 141.65 | 3.97 | 1.88 | SHHC |
| S15_vs_18274 | 0 | 0.01 | 0 | 0.62 | 0.02 | 0.41 | 0.07 | 0.27 | 0.17 | 0.17 | 0.53 | 0.14 | 0.79 | 0.28 | 0.47 | 0.34 | 0.48 | 50.42 | 0.46 | 1.04 | SHHC |
| S15_vs_76390 | 0.01 | 0.01 | 0 | 1.25 | 0.05 | 0.68 | 0.07 | 0.33 | 0.22 | 0.18 | 0.79 | 0.21 | 0.92 | 0.36 | 0.81 | 0.33 | 0.56 | 212.18 | 0.86 | 0.94 | SHHC |
| S15_vs_3868 | 0 | 0.03 | 0.03 | 62.19 | 1.72 | 423.76 | 0.08 | 1.91 | 1.73 | 10218.34 | 10.17 | 0.99 | 1.04 | 342.38 | 39.1 | 75.85 | 15.07 | 156.23 | 69.49 | 73.42 | Shared |
| S15_vs_27510 | 0 | 0.02 | 0 | 8.34 | 0.18 | 1.51 | 0.3 | 0.31 | 0.13 | 0.41 | 2.03 | 0.64 | 1.6 | 2.46 | 3.24 | 4.41 | 1.56 | 242.89 | 1.27 | 22.3 | Shared |
| S15_vs_35445 | 0.08 | 0.01 | 0 | 29.36 | 0.82 | 30.59 | 0.7 | 53.99 | 53.42 | 6.45 | 80.34 | 2.66 | 41.42 | 5.3 | 12.63 | 5.6 | 42.44 | 85.39 | 23.11 | 35.86 | Shared |
| S15_vs_39804 | 0 | 0 | 0.04 | 0.67 | 0.16 | 603.74 | 0.7 | 2.48 | 0.27 | 0.35 | 5.97 | 0.86 | 2.06 | 0.91 | 1.07 | 1.74 | 0.82 | 56.64 | 0.95 | 1.24 | Shared |
| S15_vs_42043 | 0.01 | 0 | 0 | 11.64 | 2.02 | 22.21 | 0.47 | 20.65 | 30.69 | 2.97 | 10.76 | 0.9 | 25.26 | 4.89 | 6.27 | 3.55 | 13.01 | 92.71 | 10.07 | 22.99 | Shared |
| S15_vs_43913 | 0 | 0 | 0 | 0.66 | 0.05 | 0.48 | 1139.83 | 0.22 | 0.17 | 6.53 | 1.24 | 0.11 | 1.11 | 14.74 | 5.56 | 2.22 | 2.8 | 54.89 | 1.57 | 0.91 | Shared |
| S15_vs_71968 | 0.01 | 0.01 | 0 | 5.11 | 0.28 | 36.97 | 0.93 | 7.01 | 3 | 3.77 | 23.07 | 1 | 1.3 | 8.3 | 8.3 | 1.35 | 0.81 | 35.46 | 5.97 | 3.21 | Shared |
| S2_vf_4130 | 0.29 | 0.04 | 0.01 | 223.63 | 2531.26 | 62.27 | 15.54 | 0.57 | 0.43 | 0.56 | 1.5 | 0.22 | 1.36 | 0.33 | 1.52 | 22.44 | 2.9 | 0.71 | 0.51 | 0.83 | Shared |
| S2_vf_17850 | 0 | 0 | 0 | 10.06 | 42.1 | 0.46 | 0.04 | 0.32 | 0.11 | 0.16 | 0.75 | 0.08 | 0.4 | 0.09 | 0.41 | 0.99 | 2.3 | 0.23 | 0.85 | 18.14 | Shared |
| S2_vf_16829 | 0 | 0 | 0 | 0.5 | 230.53 | 0.45 | 0.07 | 0.12 | 0.11 | 0.07 | 0.27 | 54.21 | 0.36 | 0.04 | 0.13 | 0.25 | 0.46 | 0.16 | 218.56 | 17.5 | Shared |
| S2_vf_3356 | 0 | 0 | 0 | 229.92 | 138.02 | 0.46 | 0.1 | 0.31 | 0.18 | 0.28 | 0.24 | 0.06 | 0.33 | 0.07 | 0.29 | 5.02 | 0.39 | 0.74 | 0.1 | 0.59 | Shared |
| S2_vf_20910 | 0 | 0 | 0 | 0.38 | 38.6 | 0.48 | 0.09 | 0.19 | 0.15 | 4.21 | 0.38 | 0.08 | 0.55 | 5.93 | 0.61 | 0.4 | 0.45 | 0.3 | 0.37 | 2.56 | SHHC |
| S2_vf_4910 | 0 | 0 | 0 | 0.4 | 49.06 | 0.47 | 0.01 | 0.22 | 0.13 | 0.24 | 560.33 | 0.1 | 1.44 | 0.11 | 0.21 | 0.25 | 0.35 | 0.26 | 0.28 | 1.28 | SHHC |
| S2_vf_21511 | 0 | 0 | 0 | 1.19 | 62.4 | 25.44 | 0.13 | 0.28 | 0.25 | 0.26 | 1.33 | 0.1 | 0.74 | 0.38 | 0.48 | 0.45 | 0.41 | 0.34 | 0.63 | 0.84 | Shared |
| S2_vf_21630 | 0 | 0 | 0 | 0.61 | 43.56 | 0.32 | 0.05 | 0.11 | 0.07 | 0.1 | 0.23 | 0.06 | 0.31 | 0.29 | 0.19 | 0.17 | 0.32 | 0.11 | 0.41 | 0.39 | SHHC |
| S2_vs_7306 | 0.2 | 0.09 | 0.01 | 793.18 | 1660.82 | 202.47 | 221.17 | 187.58 | 317.92 | 112.97 | 2.53 | 0.33 | 1.43 | 12.69 | 0.41 | 58.79 | 2.28 | 1.55 | 24.75 | 0.51 | Shared |
| S2_vs_4368 | 0.03 | 0 | 0 | 1.11 | 27.26 | 0.74 | 0.1 | 0.45 | 0.16 | 4.58 | 0.45 | 0.06 | 0.2 | 0.54 | 0.89 | 0.23 | 0.8 | 0.22 | 0.3 | 0.63 | SHHC |
| S2_vs_10289 | 0.01 | 0 | 0 | 13.72 | 37.96 | 15.06 | 0.48 | 6.51 | 3.11 | 13.57 | 3.31 | 1.43 | 27.32 | 3.47 | 13.84 | 8.89 | 31.35 | 16.06 | 74.1 | 14.3 | Shared |
| S4_vf_2524 | 0.01 | 0 | 0 | 0.32 | 0 | 4.97 | 21.65 | 0.22 | 0.1 | 0.1 | 0.23 | 0.01 | 0.51 | 0.69 | 3.18 | 0.27 | 0.2 | 26.26 | 0.2 | 0.36 | SHHC |

| | | | | | | | | | | | | | | | | | | | | | |
|---|---|---|---|---|---|---|---|---|---|---|---|---|---|---|---|---|---|---|---|---|---|
| S4_vf_4838 | 0.01 | 0 | 0 | 3 | 0.2 | 0.8 | 8527.63 | 0.4 | 0.26 | 0.69 | 1380.5 | 0.08 | 0.87 | 1.33 | 0.66 | 14.56 | 0.36 | 0.32 | 0.51 | 2.88 | Shared |
| S4_vf_4891 | 0 | 0 | 0 | 0.96 | 0.02 | 0.46 | 14.81 | 0.27 | 0.17 | 0.16 | 33.26 | 0.22 | 1.06 | 0.34 | 0.39 | 0.25 | 0.33 | 0.35 | 0.38 | 0.84 | SHHC |
| S4_vf_5021 | 0 | 0 | 0 | 0.38 | 0.01 | 0.31 | 212.82 | 0.15 | 0.08 | 0.12 | 0.4 | 0.06 | 1.02 | 0.61 | 0.3 | 0.29 | 0.16 | 0.17 | 0.27 | 1 | SHHC |
| S4_vf_10506 | 0.01 | 0 | 0 | 0.7 | 0.05 | 0.63 | 10658.08 | 0.21 | 0.14 | 0.12 | 57.13 | 0.09 | 0.69 | 0.17 | 0.51 | 1.22 | 0.31 | 0.39 | 0.45 | 0.95 | SHHC |
| S4_vf_11308 | 0 | 0 | 0 | 0.76 | 1.81 | 1.3 | 12.54 | 31.21 | 3.56 | 1.29 | 0.95 | 0.2 | 0.52 | 0.49 | 2.94 | 0.31 | 18.02 | 0.21 | 3.07 | 0.61 | Shared |
| S4_vf_12614 | 0 | 0 | 0 | 0.53 | 0.03 | 0.48 | 76.95 | 0.23 | 1.64 | 0.19 | 0.56 | 0.3 | 0.66 | 0.25 | 1.43 | 0.23 | 0.37 | 0.31 | 1.57 | 6.64 | SHHC |
| S4_vf_12624 | 0.01 | 0 | 0 | 0.4 | 0.05 | 0.09 | 116.26 | 0.17 | 0.07 | 1.48 | 0.36 | 0.08 | 0.22 | 0.21 | 0.23 | 0.17 | 0.35 | 0.13 | 0.17 | 0.25 | SHHC |
| S4_vf_13353 | 0 | 0 | 0 | 1.15 | 0.11 | 1.41 | 4571.4 | 0.23 | 0.16 | 0.22 | 0.86 | 0.39 | 0.55 | 0.25 | 0.7 | 0.46 | 0.39 | 2.87 | 34.31 | 72.32 | Shared |
| S4_vf_13627 | 0 | 0 | 0 | 3.87 | 0.02 | 156.33 | 316.23 | 0.13 | 0.15 | 0.1 | 0.15 | 0.3 | 0.35 | 0.12 | 0.1 | 1.49 | 0.13 | 0.2 | 0.23 | 0.26 | Shared |
| S4_vf_13759 | 0 | 0 | 0 | 0.46 | 0.04 | 2.3 | 9.91 | 0.95 | 0.31 | 0.38 | 2.22 | 0.35 | 2.21 | 0.44 | 1.43 | 0.82 | 0.87 | 0.98 | 2.25 | 1.66 | SHHC |
| S4_vf_13831 | 0 | 0 | 0 | 0.7 | 0.29 | 1.95 | 1822.83 | 0.7 | 0.48 | 0.71 | 1226.1 | 0.29 | 3.14 | 2.03 | 1.52 | 1.83 | 0.93 | 0.61 | 1.03 | 3.56 | SHHC |
| S4_vf_16485 | 0 | 0 | 0 | 0.3 | 0.02 | 0.25 | 50.09 | 0.17 | 0.09 | 0.1 | 0.37 | 0.04 | 0.17 | 0.23 | 0.27 | 0.84 | 0.18 | 0.21 | 0.24 | 0.66 | SHHC |
| S4_vf_17948 | 0.01 | 0 | 0 | 0.29 | 0.02 | 0.39 | 133.3 | 0.1 | 0.1 | 0.08 | 0.92 | 0.05 | 0.29 | 0.11 | 0.39 | 0.19 | 0.21 | 0.12 | 0.17 | 0.51 | SHHC |
| S4_vf_18803 | 0.01 | 0 | 0 | 0.42 | 0.02 | 0.36 | 795.17 | 0.16 | 0.11 | 0.09 | 35.83 | 0.07 | 0.29 | 0.24 | 0.18 | 5.7 | 0.18 | 0.21 | 0.21 | 0.7 | SHHC |
| S4_vf_20440 | 0 | 0 | 0 | 0.57 | 0.08 | 0.4 | 330.61 | 0.23 | 0.18 | 0.22 | 355.21 | 0.19 | 0.68 | 0.59 | 0.27 | 0.25 | 0.22 | 0.3 | 0.36 | 1.14 | SHHC |
| S4_vf_22219 | 0.85 | 0.04 | 0.98 | 1.29 | 1.34 | 11507.36 | 465.16 | 1.6 | 1.67 | 1.65 | 3.42 | 14.57 | 0.83 | 0.67 | 0.48 | 1.53 | 0.37 | 0.41 | 0.46 | 1.02 | Shared |
| S4_vf_26417 | 0 | 0 | 0 | 1.25 | 0.89 | 2.9 | 730.2 | 301.02 | 811.68 | 6.53 | 5532.69 | 34.47 | 3.69 | 5.93 | 6451.22 | 4.81 | 2.7 | 1.73 | 4202.33 | 11.76 | Shared |
| S4_vf_27861 | 0 | 0 | 0 | 0.52 | 0.03 | 13.75 | 239.09 | 0.19 | 0.14 | 0.08 | 0.36 | 0.74 | 47.36 | 39.2 | 0.33 | 0.78 | 0.22 | 0.24 | 0.34 | 0.61 | Shared |
| S4_vf_28601 | 0.02 | 0 | 0 | 0.59 | 0.01 | 0.16 | 164.87 | 0.23 | 0.07 | 0.11 | 0.55 | 0.06 | 0.14 | 0.07 | 0.37 | 0.22 | 0.11 | 0.19 | 0.13 | 0.36 | SHHC |
| S4_vf_30103 | 0.01 | 0 | 0 | 0.45 | 0.03 | 61.17 | 4852.83 | 0.18 | 0.14 | 0.09 | 0.32 | 0.12 | 0.3 | 0.04 | 0.36 | 0.19 | 0.1 | 0.25 | 0.16 | 0.34 | Shared |
| S4_vf_31682 | 0.01 | 0.01 | 0.01 | 81.11 | 0.03 | 0.19 | 24.46 | 0.25 | 0.11 | 0.99 | 0.37 | 0.1 | 5.4 | 0.37 | 0.24 | 16.35 | 0.18 | 8.47 | 0.38 | 0.63 | Shared |
| S4_vf_31825 | 0.39 | 0 | 0.38 | 8.45 | 31.61 | 4417.99 | 1212.55 | 0.66 | 0.62 | 0.7 | 3.55 | 5.5 | 0.43 | 0.39 | 0.76 | 0.91 | 0.22 | 0.18 | 0.31 | 0.51 | Shared |
| S4_vf_31973 | 0.01 | 0 | 0.02 | 0.64 | 0.04 | 189.95 | 70.01 | 2.63 | 3.15 | 0.31 | 0.7 | 134.54 | 0.93 | 0.23 | 22.23 | 0.53 | 0.4 | 0.36 | 0.48 | 0.94 | Shared |
| S4_vf_31978 | 0 | 0 | 0 | 4.05 | 0.07 | 1.39 | 16.78 | 0.27 | 0.14 | 0.19 | 0.66 | 0.12 | 0.8 | 0.42 | 3.62 | 0.82 | 0.45 | 0.39 | 34.2 | 0.91 | SHHC |
| S4_vf_31979 | 0.01 | 0 | 0 | 0.63 | 0.01 | 0.36 | 57.2 | 0.95 | 1.21 | 0.11 | 0.64 | 0.09 | 0.37 | 0.22 | 0.67 | 0.26 | 0.26 | 0.2 | 0.26 | 0.47 | SHHC |
| S4_vf_31981 | 0 | 0 | 0 | 0.44 | 0.05 | 0.85 | 198.44 | 0.38 | 0.22 | 0.18 | 0.96 | 0.1 | 1.34 | 0.31 | 0.77 | 0.53 | 0.44 | 0.42 | 0.62 | 0.99 | SHHC |
| S4_vf_31982 | 0.01 | 0 | 0 | 0.48 | 0.02 | 0.19 | 286.19 | 0.18 | 0.08 | 0.12 | 0.51 | 0.04 | 0.31 | 0.26 | 0.43 | 0.16 | 0.2 | 0.2 | 0.28 | 0.36 | SHHC |
| S4_vs_VIRSorter_ | 0 | 0 | 0 | 3.01 | 0.11 | 1.18 | 24.04 | 0.9 | 0.52 | 0.32 | 0.79 | 0.37 | 1.83 | 31.86 | 0.61 | 0.42 | 1.65 | 1.64 | 1.31 | 4.85 | Shared |
| S4_vs_VIRSorter_ | 0.09 | 0.01 | 0.08 | 44.34 | 32.83 | 1215.41 | 9554.46 | 561.71 | 904.83 | 183.55 | 4.67 | 1.57 | 1.5 | 21.58 | 0.62 | 1.71 | 0.24 | 0.23 | 3.24 | 0.55 | Shared |
| S4_vs_VIRSorter_ | 0 | 0 | 0 | 0.91 | 0.04 | 5.06 | 47.9 | 0.35 | 0.18 | 1.15 | 0.66 | 0.46 | 0.69 | 1.02 | 0.37 | 0.28 | 0.37 | 0.41 | 1.33 | 1.39 | SHHC |
| S4_vs_VIRSorter_ | 0.21 | 0 | 0 | 14.18 | 4.01 | 12.56 | 32.36 | 73.62 | 0.44 | 60.41 | 25.06 | 328.01 | 3.56 | 0.95 | 119.35 | 5.32 | 5291.67 | 21.52 | 196.81 | 15.42 | Shared |
| S4_vs_VIRSorter_ | 1 | 0 | 0.7 | 6.37 | 1.17 | 7413.04 | 758 | 269.92 | 404.13 | 4.81 | 2.62 | 10 | 0.93 | 1.34 | 0.41 | 1.34 | 0.73 | 0.37 | 1.05 | 2.35 | Shared |
| S4_vs_VIRSorter_ | 0 | 0 | 0 | 0.72 | 0.07 | 0.41 | 70.7 | 0.49 | 0.23 | 1.61 | 3 | 1.56 | 0.45 | 1.2 | 0.41 | 1.51 | 0.51 | 0.34 | 0.4 | 1.23 | SHHC |
| S4_vs_VIRSorter_ | 0.01 | 0 | 0 | 1.19 | 0.31 | 5.91 | 115.6 | 2.8 | 2.25 | 3.6 | 6.29 | 2.35 | 0.4 | 4.15 | 1.36 | 0.52 | 142.76 | 0.43 | 1.36 | 2.16 | Shared |
| S4_vs_VIRSorter_ | 0 | 0 | 0 | 5.54 | 0.36 | 3.44 | 33.68 | 5.93 | 0.27 | 3.94 | 3.47 | 0.51 | 1.04 | 8.88 | 2.26 | 1.53 | 20.92 | 1.74 | 30.8 | 6.49 | Shared |
| S4_vs_VIRSorter_ | 0 | 0 | 0 | 1 | 0.01 | 0.7 | 85.36 | 0.53 | 0.13 | 1.23 | 0.89 | 0.09 | 0.6 | 0.09 | 0.52 | 0.32 | 0.29 | 0.2 | 0.31 | 0.89 | SHHC |
| S4_vs_VIRSorter_ | 0.01 | 0 | 0 | 1.02 | 2.86 | 1.1 | 206.57 | 10.99 | 9.24 | 89.57 | 0.51 | 1.87 | 0.42 | 1.07 | 0.34 | 0.41 | 4.01 | 0.65 | 4.92 | 0.68 | Shared |
| S4_vs_VIRSorter_ | 0 | 0 | 0 | 1.37 | 0.11 | 0.43 | 24.12 | 4.43 | 0.09 | 0.18 | 0.46 | 0.1 | 0.32 | 0.42 | 0.2 | 0.41 | 4.03 | 0.22 | 0.25 | 0.53 | SHHC |
| S4_vs_VIRSorter_ | 0 | 0 | 0 | 1.96 | 0.03 | 1.22 | 28.11 | 0.26 | 0.09 | 0.16 | 0.61 | 0.11 | 0.73 | 0.15 | 0.38 | 0.32 | 0.2 | 0.19 | 0.24 | 0.84 | SHHC |
| S4_vs_VIRSorter_ | 0.01 | 0 | 0 | 0.5 | 0.02 | 0.28 | 46.88 | 0.18 | 0.11 | 0.11 | 0.44 | 0.09 | 0.27 | 0.19 | 0.25 | 0.2 | 15.77 | 0.21 | 8.03 | 0.4 | Shared |
| S4_vs_VIRSorter_ | 0 | 0 | 0 | 0.87 | 0.03 | 0.2 | 21.16 | 0.19 | 0.05 | 0.12 | 9.17 | 0.06 | 0.29 | 0.13 | 0.25 | 3.91 | 0.14 | 0.2 | 0.21 | 0.33 | SHHC |
| S4_vs_VIRSorter_ | 0.04 | 0 | 0.04 | 9.31 | 0.07 | 359.34 | 13896.79 | 0.18 | 0.16 | 0.17 | 0.39 | 0.58 | 0.37 | 0.15 | 0.3 | 0.32 | 0.27 | 0.21 | 0.31 | 0.61 | Shared |
| S4_vs_VIRSorter_ | 0 | 0 | 0 | 1.6 | 0.05 | 0.95 | 387.45 | 13.36 | 11.82 | 2.67 | 2.06 | 1.45 | 0.41 | 57.75 | 0.64 | 28.91 | 2.74 | 0.38 | 9.62 | 8.46 | Shared |
| S4_vs_VIRSorter_ | 0.03 | 0 | 0.01 | 10.57 | 0.17 | 87.35 | 35.89 | 0.45 | 0.15 | 0.32 | 1.69 | 2.2 | 1.27 | 0.42 | 1.52 | 3.55 | 41.43 | 1.41 | 1.05 | 1.93 | Shared |
| S4_vs_VIRSorter_ | 0 | 0 | 0 | 0.85 | 0.03 | 0.47 | 60.32 | 1.81 | 1.9 | 0.17 | 8.2 | 0.13 | 2.97 | 0.37 | 1.21 | 0.59 | 1.2 | 0.39 | 0.92 | 0.77 | SHHC |
| S4_vs_VIRSorter_ | 0.02 | 0 | 0 | 1.17 | 15.98 | 51.46 | 21.32 | 1 | 0.18 | 2.59 | 0.97 | 0.19 | 1.46 | 0.15 | 145.84 | 7.03 | 1.16 | 0.2 | 0.65 | 0.87 | Shared |
| S4_vs_VIRSorter_ | 0.01 | 0 | 0 | 1.05 | 0.04 | 0.63 | 10.25 | 1.03 | 0.43 | 6.79 | 6.59 | 0.32 | 0.24 | 0.62 | 0.33 | 0.71 | 18.33 | 0.54 | 1.92 | 3.3 | Shared |
| S4_vs_VIRSorter_ | 0.1 | 0 | 0.02 | 1.11 | 0.03 | 157.63 | 243.84 | 0.43 | 0.19 | 0.15 | 0.67 | 0.29 | 0.26 | 0.1 | 0.29 | 0.39 | 0.21 | 0.27 | 0.42 | 0.72 | Shared |
| S4_vs_VIRSorter_ | 0 | 0 | 0 | 0.61 | 0.02 | 3.84 | 65.84 | 0.29 | 0.18 | 0.25 | 0.46 | 0.15 | 0.5 | 0.19 | 0.44 | 0.3 | 0.39 | 0.3 | 0.39 | 0.89 | SHHC |
| S4_vs_VIRSorter_ | 0.01 | 0 | 0.01 | 0.37 | 0.03 | 2.3 | 166.53 | 0.22 | 0.1 | 2.3 | 1.31 | 0.06 | 0.3 | 0.23 | 0.25 | 0.39 | 0.71 | 0.25 | 0.26 | 0.55 | SHHC |
| S4_vs_VIRSorter_ | 0 | 0 | 0 | 1.41 | 0.04 | 1.48 | 34.5 | 0.27 | 0.21 | 0.31 | 0.57 | 0.09 | 0.45 | 0.71 | 0.32 | 0.4 | 0.41 | 0.34 | 0.63 | 1.47 | SHHC |
| S4_vs_VIRSorter_ | 0.01 | 0 | 0 | 0.78 | 0.14 | 0.39 | 56 | 0.37 | 0.19 | 1.63 | 1.97 | 0.32 | 0.29 | 0.27 | 0.4 | 0.38 | 1.15 | 0.21 | 0.63 | 0.54 | SHHC |
| S4_vs_VIRSorter_ | 0.01 | 0 | 0 | 0.93 | 0.9 | 4.87 | 11.43 | 1.11 | 0.3 | 1.36 | 16.59 | 0.9 | 1.62 | 17.78 | 4.06 | 0.59 | 1.56 | 0.26 | 0.72 | 9.49 | Shared |
| S4_vs_VIRSorter_ | 0 | 0.01 | 0 | 2.82 | 0.06 | 0.64 | 29.13 | 24.46 | 21.02 | 11.02 | 1.14 | 1.44 | 0.63 | 2.02 | 1.31 | 2.11 | 8.39 | 1.29 | 15.07 | 3.71 | Shared |
| S4_vs_VIRSorter_ | 0 | 0 | 0 | 10.23 | 0.1 | 5.27 | 66.64 | 4.04 | 1.03 | 0.15 | 3.1 | 1.2 | 1.04 | 3.38 | 0.86 | 7.36 | 0.71 | 1.31 | 3.56 | 1.11 | Shared |
| S4_vs_VIRSorter_ | 0 | 0.01 | 0 | 1.07 | 0.05 | 1.77 | 18.38 | 0.69 | 0.26 | 8.72 | 1.36 | 0.27 | 2.05 | 0.85 | 1.65 | 0.65 | 0.59 | 0.67 | 1.16 | 1.36 | SHHC |
| S4_vs_VIRSorter_ | 0 | 0 | 0 | 1.68 | 0.99 | 0.28 | 18.11 | 0.24 | 0.1 | 1.43 | 0.75 | 0.08 | 0.61 | 0.42 | 5.94 | 0.64 | 0.98 | 0.18 | 0.3 | 0.33 | SHHC |
| S4_vs_VIRSorter_ | 0 | 0 | 0 | 0.32 | 0 | 35.82 | 661.72 | 0.2 | 0.08 | 2.09 | 0.4 | 0.04 | 0.4 | 0.9 | 0.17 | 0.3 | 0.33 | 0.27 | 0.3 | 0.46 | Shared |
| S4_vs_VIRSorter_ | 0 | 0 | 0 | 0.92 | 0.01 | 1 | 19.91 | 0.2 | 0.1 | 0.08 | 32.43 | 0.06 | 0.45 | 0.15 | 0.22 | 6.79 | 0.16 | 0.18 | 0.16 | 2.47 | SHHC |
| S6_vf_47238 | 0 | 0 | 0 | 0.7 | 0.88 | 2.39 | 0.1 | 1421.63 | 2105.49 | 0.72 | 1.74 | 0.12 | 2.45 | 4.22 | 0.84 | 2.48 | 0.45 | 0.41 | 0.52 | 1 | Shared |
| S6_vf_45228 | 0 | 0 | 0 | 0.67 | 0.11 | 0.51 | 0.02 | 2.98 | 9.28 | 0.18 | 0.51 | 0.08 | 0.61 | 0.57 | 0.4 | 0.29 | 0.37 | 0.33 | 0.33 | 1.06 | SHHC |
| S6_vf_45618 | 0 | 0 | 0 | 3.67 | 0.09 | 0.68 | 6.87 | 15.87 | 23.47 | 1.01 | 16.23 | 4.16 | 0.26 | 2.63 | 2.72 | 0.68 | 6.69 | 15.5 | 6.94 | 6.93 | Shared |
| S6_vf_24553 | 1.07 | 0 | 0.02 | 0.5 | 9.5 | 0.6 | 0 | 52.07 | 81.09 | 2.96 | 0.22 | 710.84 | 1.75 | 1.6 | 2.19 | 27.57 | 16180.59 | 1.26 | 13.67 | 3.56 | Shared |
| S6_vf_3435 | 0 | 0 | 0.03 | 0.54 | 0.12 | 743.78 | 0.07 | 318.56 | 35.84 | 38.87 | 5.47 | 1.32 | 1.21 | 0.55 | 0.9 | 0.46 | 1.28 | 0.25 | 0.32 | 11.25 | Shared |
| S6_vf_6314 | 0 | 0 | 0 | 0.5 | 0.78 | 1.41 | 0.06 | 2223.86 | 3214.06 | 1.08 | 2.34 | 0.01 | 3.4 | 4.8 | 0.4 | 0.78 | 0.15 | 0.18 | 0.22 | 0.55 | Shared |
| S6_vf_25710 | 0 | 0 | 0 | 0.6 | 0.03 | 0.4 | 0.08 | 19.5 | 181.78 | 0.16 | 0.48 | 0.05 | 0.4 | 0.36 | 0.3 | 0.26 | 0.31 | 0.28 | 0.28 | 0.68 | Shared |
| S6_vf_56484 | 0 | 0 | 0 | 4.09 | 0.03 | 1.37 | 0.04 | 9.9 | 25.5 | 0.16 | 2.51 | 0.06 | 0.91 | 0.46 | 40.2 | 2.22 | 0.21 | 0.28 | 0.2 | 0.37 | SHHC |
| S6_vf_56486 | 0 | 0 | 0 | 1.3 | 0.07 | 0.39 | 0.08 | 38.15 | 56.22 | 0.16 | 3.29 | 0.08 | 0.57 | 0.26 | 0.43 | 0.3 | 0.31 | 0.2 | 0.35 | 0.64 | Shared |
| S6_vs_32886 | 0.01 | 0 | 0 | 0.65 | 0.15 | 0.63 | 0.31 | 320.29 | 62.28 | 0.47 | 0.82 | 0.13 | 1.77 | 4.78 | 2.12 | 0.48 | 99.19 | 0.42 | 0.58 | 0.7 | Shared |
| S6_vs_56485 | 0 | 0 | 0 | 0.52 | 0.01 | 0.94 | 0.04 | 10.66 | 14.24 | 2.06 | 0.42 | 0.32 | 0.34 | 0.25 | 0.31 | 0.14 | 0.73 | 0.16 | 0.34 | 1.16 | Shared |
| S6_vs_56490 | 0 | 0 | 0 | 2.23 | 0.21 | 1.95 | 0.09 | 798.6 | 350.27 | 0.7 | 1.14 | 0.85 | 1.91 | 1.89 | 1.05 | 0.77 | 134.41 | 0.59 | 1.27 | 2.17 | Shared |
| S6_vs_56491 | 0 | 0 | 0 | 0.63 | 0.43 | 0.43 | 0.17 | 22.67 | 39.94 | 1.01 | 0.74 | 3.42 | 0.7 | 2 | 0.37 | 0.35 | 0.77 | 0.55 | 0.41 | 0.76 | Shared |
| S6_vs_56492 | 0 | 0 | 0 | 2.08 | 0.14 | 0.52 | 0.14 | 13.35 | 23.19 | 1.07 | 0.51 | 0.54 | 0.5 | 0.54 | 2.6 | 0.43 | 1.44 | 0.24 | 0.61 | 1.21 | Shared |
| S6_vs_56493 | 0 | 0 | 0 | 1.87 | 0.06 | 0.33 | 0.12 | 6.56 | 11.19 | 0.37 | 0.64 | 0.07 | 0.26 | 3.27 | 1.15 | 1.53 | 7.32 | 0.23 | 0.49 | 0.51 | SHHC |
| S6_vs_56496 | 0 | 0 | 0 | 0.88 | 0.02 | 0.18 | 0.05 | 11 | 27.34 | 0.09 | 0.39 | 0.09 | 0.38 | 0.23 | 0.18 | 0.26 | 0.13 | 0.19 | 0.22 | 0.43 | Shared |
| S6_vs_56498 | 0 | 0 | 0 | 0.64 | 0.05 | 0.39 | 0.07 | 30.95 | 55.36 | 1.54 | 0.38 | 0.27 | 0.4 | 0.35 | 0.35 | 2.06 | 0.28 | 0.21 | 0.28 | 0.87 | Shared |
| S6_vs_37812 | 0 | 0 | 0 | 0.99 | 0.07 | 2.41 | 0.07 | 10.51 | 18.42 | 2.95 | 9.03 | 1.2 | 2.05 | 22.18 | 4.02 | 6.27 | 2.26 | 0.43 | 4.2 | 2.59 | Shared |
| S6_vs_38250 | 0 | 0 | 0 | 0.64 | 0.02 | 0.61 | 0.07 | 6.54 | 13.63 | 1.64 | 0.46 | 0.19 | 0.55 | 0.62 | 0.5 | 0.39 | 0.7 | 0.29 | 0.46 | 0.78 | SHHC |
| S6_vs_9784 | 0.01 | 0.01 | 0 | 11.92 | 0.91 | 11.75 | 1.61 | 49.75 | 88.32 | 3.83 | 7.45 | 0.98 | 18.2 | 3.43 | 4.01 | 2.72 | 8.96 | 32 | 6.21 | 19.63 | Shared |
| S6_vs_33711 | 0.01 | 0 | 0 | 0.57 | 0.01 | 0.38 | 0.1 | 5.22 | 16.08 | 0.2 | 0.4 | 0.1 | 0.71 | 1.77 | 0.53 | 0.41 | 0.42 | 0.42 | 5.88 | 0.76 | SHHC |
| S6_vs_54606 | 0.01 | 0 | 0 | 7.69 | 0.22 | 9.15 | 3.48 | 23.83 | 29.5 | 10.23 | 5.75 | 1.32 | 9.34 | 2.38 | 3.3 | 7.48 | 1.66 | 6.59 | 14.64 | 6.08 | Shared |
| S6_vs_29152 | 0 | 0 | 0 | 6.66 | 0.19 | 3.05 | 0.72 | 5.33 | 30.7 | 4.11 | 5.04 | 0.97 | 5.32 | 2.08 | 2.48 | 5.76 | 1.06 | 4.52 | 7.74 | 6.13 | SHHC |
| S6_vs_10314 | 0 | 0 | 0 | 10.37 | 0.12 | 4.7 | 0.56 | 5.59 | 32.36 | 9.29 | 9.21 | 1.84 | 9.43 | 3.81 | 2.1 | 10.47 | 1.08 | 8.23 | 14.83 | 7.37 | Shared |
| S6_vs_1146 | 0.01 | 0.01 | 0.01 | 3.42 | 0.22 | 27.76 | 12.27 | 19.09 | 12.83 | 7.85 | 6.05 | 4.27 | 3.07 | 1.53 | 18.04 | 2.19 | 5.83 | 2 | 14.78 | 25.7 | Shared |

| | | | | | | | | | | | | | | | | | | | | | |
|---|---|---|---|---|---|---|---|---|---|---|---|---|---|---|---|---|---|---|---|---|---|
| S6_vs_46039 | 0.02 | 0 | 0 | 5.82 | 0.78 | 5.84 | 2.02 | 67.12 | 61.8 | 10.42 | 9.8 | 1.2 | 7.71 | 11.12 | 5.39 | 1.84 | 66.65 | 11.23 | 35.08 | 17.95 | Shared |
| S6_vs_12945 | 0.01 | 0 | 0 | 5.71 | 1.15 | 3.67 | 0.22 | 5.05 | 30.17 | 2.17 | 3.67 | 1.12 | 6.17 | 6.86 | 3.34 | 5.5 | 1.6 | 4.09 | 9.62 | 23.37 | Shared |
| S6_vs_34453 | 0 | 0 | 0 | 1.21 | 0.07 | 3.05 | 1.26 | 29.6 | 49.2 | 0.69 | 1.19 | 9.67 | 1.27 | 0.53 | 0.99 | 0.65 | 1.29 | 0.36 | 2.02 | 7.07 | Shared |
| S6_vs_15390 | 0 | 0 | 0 | 0.46 | 0.03 | 0.53 | 0.08 | 35.48 | 86.49 | 0.11 | 1.92 | 0.08 | 0.75 | 0.54 | 0.49 | 1.1 | 0.22 | 0.3 | 1.21 | 0.71 | Shared |
| S6_vs_39025 | 0 | 0 | 0 | 7.4 | 0.25 | 3.88 | 0.11 | 56.3 | 28.34 | 6.78 | 8.7 | 1.39 | 8.95 | 2.87 | 1.44 | 7.42 | 9.27 | 6.54 | 13.4 | 6.71 | Shared |
| S6_vs_46091 | 0 | 0 | 0.02 | 0.54 | 0.04 | 0.66 | 0.06 | 7.77 | 18.53 | 0.16 | 0.35 | 308.61 | 0.39 | 0.39 | 0.36 | 0.21 | 0.75 | 0.23 | 0.36 | 0.62 | Shared |
| S6_vs_15813 | 0.01 | 0 | 0 | 1.83 | 0.19 | 9.71 | 9.3 | 16.39 | 25.37 | 5.57 | 4.21 | 3.51 | 0.42 | 2.93 | 0.85 | 0.68 | 167.46 | 0.32 | 1.04 | 2.62 | Shared |
| S6_vs_22637 | 0 | 0 | 0 | 0.42 | 0.01 | 0.39 | 0.04 | 18.27 | 17.69 | 0.23 | 0.32 | 0.05 | 0.24 | 0.08 | 0.18 | 0.09 | 2.74 | 0.14 | 0.27 | 0.67 | Shared |
| S6_vs_44070 | 0 | 0 | 0 | 2.44 | 0.05 | 0.45 | 0.04 | 10.56 | 17.22 | 0.43 | 1.07 | 0.09 | 0.57 | 1.58 | 0.65 | 0.41 | 0.36 | 0.32 | 0.54 | 0.84 | Shared |
| S6_vs_53966 | 0.01 | 0 | 0 | 1.06 | 0.08 | 1.61 | 0.88 | 6.18 | 8 | 0.33 | 1.44 | 0.39 | 0.55 | 0.12 | 1.27 | 0.3 | 0.66 | 0.52 | 9.49 | 4.92 | SHHC |
| S6_vs_39065 | 0.01 | 0.01 | 0 | 63.26 | 0.1 | 0.54 | 0.03 | 48.09 | 87.79 | 0.3 | 3.59 | 0.14 | 0.8 | 16.49 | 0.86 | 46.57 | 0.31 | 0.42 | 0.65 | 1.53 | Shared |
| S8_vf_43483 | 0.01 | 0 | 0 | 1.1 | 0.05 | 2.93 | 0.01 | 10.55 | 14.89 | 0.15 | 389.95 | 0.13 | 0.28 | 0.13 | 0.41 | 0.17 | 0.1 | 0.18 | 0.37 | 0.96 | Shared |
| S8_vf_122206 | 0 | 0 | 0 | 1.5 | 0.09 | 0.96 | 0.78 | 0.55 | 0.27 | 0.3 | 217.68 | 0.47 | 1.73 | 0.22 | 0.65 | 39.68 | 0.89 | 0.62 | 0.66 | 1.98 | Shared |
| S8_vf_131009 | 0 | 0 | 0 | 1.36 | 0.1 | 2.32 | 0.1 | 0.62 | 0.43 | 0.7 | 337.25 | 0.15 | 1.21 | 0.39 | 1.31 | 1.22 | 2.59 | 0.7 | 7.11 | 1.86 | SHHC |
| S8_vf_87755 | 0.01 | 0 | 0.03 | 2.53 | 1.29 | 238.32 | 4.29 | 2.03 | 1.47 | 1573.69 | 107.74 | 1.04 | 700.65 | 0.57 | 2.92 | 8540.27 | 0.54 | 0.58 | 11.2 | 1.35 | Shared |
| S8_vf_174559 | 0 | 0 | 0 | 1.79 | 0.17 | 2.52 | 0.29 | 0.71 | 0.93 | 0.42 | 677.4 | 0.15 | 1.05 | 0.28 | 0.7 | 26.76 | 0.93 | 0.99 | 0.86 | 2.47 | Shared |
| S8_vf_183045 | 0.03 | 0 | 0 | 0.59 | 0.27 | 0.89 | 182.31 | 0.48 | 0.51 | 0.54 | 2519.1 | 0.06 | 0.47 | 0.27 | 0.27 | 385.66 | 0.26 | 0.3 | 0.74 | 4.35 | Shared |
| S8_vf_166403 | 0 | 0 | 0 | 0.62 | 0.04 | 0.65 | 0.15 | 0.52 | 0.34 | 1.13 | 27.32 | 1.15 | 0.2 | 0.31 | 0.25 | 0.35 | 15.02 | 0.15 | 0.63 | 1.52 | Shared |
| S8_vf_79769 | 0 | 0 | 0 | 0.77 | 0.04 | 0.4 | 3.25 | 0.26 | 0.21 | 0.16 | 418.11 | 0.11 | 0.61 | 0.25 | 0.32 | 20.9 | 0.23 | 0.22 | 0.28 | 1.28 | Shared |
| S8_vf_192724 | 0 | 0 | 0 | 0.36 | 0.1 | 0.33 | 0.14 | 0.23 | 0.11 | 0.19 | 574.78 | 0.07 | 0.54 | 0.45 | 0.42 | 17.03 | 0.29 | 0.23 | 0.33 | 1.48 | Shared |
| S8_vf_131802 | 0 | 0 | 0 | 0.68 | 0.67 | 1.4 | 0.07 | 0.73 | 0.7 | 0.84 | 4703.11 | 0.16 | 0.91 | 0.23 | 0.57 | 0.78 | 0.29 | 0.41 | 0.41 | 6.85 | SHHC |
| S8_vf_166677 | 0.05 | 0 | 0 | 3.2 | 4.07 | 6.29 | 65.39 | 3.78 | 3.39 | 4.55 | 29416.89 | 0.2 | 1.24 | 0.35 | 0.82 | 3.09 | 0.59 | 0.53 | 0.61 | 39.7 | Shared |
| S8_vf_71214 | 0.04 | 0 | 0 | 0.31 | 0.05 | 0.09 | 0.01 | 0.29 | 0.05 | 13.56 | 37.8 | 0.01 | 0.23 | 0.04 | 0.1 | 0.18 | 0.15 | 0.1 | 0.15 | 1.28 | Shared |
| S8_vf_158225 | 0 | 0 | 0 | 1.79 | 2.32 | 4.85 | 0.25 | 837.26 | 1819.65 | 9.73 | 13998.91 | 0.17 | 3.76 | 3.43 | 1.69 | 2.43 | 0.71 | 0.64 | 1.02 | 19.94 | Shared |
| S8_vf_54514 | 0.01 | 0.03 | 0 | 46.65 | 1.69 | 2.99 | 0.96 | 4.69 | 0.98 | 2.02 | 88.67 | 0.83 | 1.28 | 2.74 | 0.81 | 13.71 | 9.8 | 0.46 | 1.93 | 4.09 | Shared |
| S8_vf_201695 | 0 | 0 | 0 | 8.77 | 0.06 | 2.29 | 0.06 | 2.52 | 4.87 | 0.12 | 31.4 | 0.08 | 0.37 | 0.18 | 0.33 | 1.55 | 0.19 | 0.25 | 3.6 | 76.66 | Shared |
| S8_vf_45776 | 0.27 | 0 | 0.02 | 0.61 | 3.37 | 140.59 | 150.67 | 16.11 | 3.93 | 11.43 | 1596.88 | 10.83 | 1.65 | 1.7 | 1.62 | 1.07 | 4891.97 | 0.75 | 9.89 | 532.56 | Shared |
| S8_vf_28956 | 0.01 | 0 | 0 | 14.41 | 1.66 | 22.85 | 1.43 | 43.06 | 7.02 | 7.54 | 38.31 | 3.52 | 121.43 | 3.26 | 18.15 | 15.17 | 120.65 | 31.7 | 48.85 | 120.22 | Shared |
| S8_vf_132697 | 0 | 0 | 0 | 3.25 | 0.64 | 11.82 | 0.16 | 0.58 | 0.26 | 0.29 | 791.25 | 0.11 | 0.58 | 0.44 | 0.93 | 0.76 | 0.36 | 0.27 | 0.7 | 1.87 | Shared |
| S8_vf_106946 | 0 | 0 | 0 | 2.59 | 0.09 | 0.3 | 0.04 | 0.14 | 0.15 | 0.16 | 595.57 | 0.05 | 0.25 | 0.31 | 0.2 | 0.2 | 0.17 | 0.22 | 0.16 | 0.87 | SHHC |
| S8_vf_159173 | 0 | 0 | 0 | 0.53 | 0.01 | 0.39 | 0 | 0.22 | 0.17 | 0.14 | 141.2 | 0.04 | 0.74 | 0.4 | 0.17 | 0.2 | 0.48 | 0.33 | 0.41 | 0.76 | SHHC |
| S8_vf_55476 | 0 | 0 | 0 | 3.85 | 0.3 | 2.65 | 1.13 | 1.73 | 0.26 | 1.29 | 70.76 | 6.9 | 1.13 | 16.33 | 2.4 | 1.16 | 2.47 | 0.91 | 15.67 | 5.9 | Shared |
| S8_vf_3410 | 0 | 0 | 0 | 0.42 | 52.33 | 0.39 | 0.09 | 22.26 | 41.95 | 0.63 | 151.05 | 0.08 | 0.9 | 43.75 | 0.19 | 0.29 | 0.38 | 0.38 | 16.81 | 1.01 | Shared |
| S8_vf_72329 | 0 | 0 | 0 | 10.33 | 3.29 | 1.16 | 0.31 | 0.49 | 0.68 | 384.15 | 246.76 | 0.17 | 0.81 | 39.49 | 71.37 | 3.51 | 3.32 | 0.27 | 1.24 | 1.14 | Shared |
| S8_vf_11794 | 0 | 0 | 0 | 1 | 0.07 | 0.76 | 0.14 | 0.48 | 0.21 | 8.7 | 408.9 | 0.1 | 0.44 | 0.4 | 0.54 | 0.44 | 1.09 | 0.29 | 0.48 | 1.21 | SHHC |
| S8_vf_142700 | 0 | 0 | 0 | 0.29 | 0.03 | 3.71 | 0 | 0.12 | 0.07 | 0.06 | 38 | 0.03 | 0.19 | 0.03 | 0.28 | 0.21 | 0.07 | 0.21 | 0.2 | 0.19 | SHHC |
| S8_vf_72773 | 0.01 | 0.06 | 0 | 827.83 | 0.13 | 20.68 | 0.08 | 0.53 | 0.25 | 0.42 | 131.86 | 0.13 | 1.42 | 0.28 | 38.68 | 338.85 | 0.94 | 1.26 | 0.78 | 0.92 | Shared |
| S8_vf_203044 | 0 | 0 | 0 | 1.13 | 1.65 | 2.55 | 0.04 | 1.63 | 1.63 | 2.08 | 11690.43 | 0.09 | 0.48 | 0.26 | 0.47 | 1.55 | 0.51 | 0.25 | 0.31 | 17.16 | Shared |
| S8_vf_12212 | 0 | 0 | 0 | 1.49 | 0.23 | 1.4 | 0.13 | 0.43 | 0.24 | 0.37 | 202.74 | 0.12 | 1.97 | 0.3 | 165.13 | 9.85 | 0.82 | 0.6 | 0.91 | 1.28 | SHHC |
| S8_vf_195032 | 0 | 0 | 0 | 0.59 | 0.01 | 0.26 | 0.14 | 0.22 | 0.09 | 0.06 | 25.85 | 0.06 | 0.31 | 0.18 | 0.37 | 0.2 | 0.14 | 0.2 | 0.32 | 0.54 | SHHC |
| S8_vf_151384 | 0.01 | 0 | 0.01 | 1.42 | 0.04 | 0.77 | 0.16 | 0.65 | 0.45 | 0.89 | 67.39 | 0.21 | 0.59 | 9.24 | 1.63 | 1.56 | 1.2 | 0.27 | 0.86 | 1.45 | SHHC |
| S8_vf_91033 | 0 | 0 | 0 | 0.22 | 0.03 | 0.21 | 0 | 0.24 | 0.08 | 0.21 | 166.79 | 0.05 | 0.26 | 0.31 | 0.19 | 0.13 | 0.37 | 0.21 | 0.15 | 0.67 | SHHC |
| S8_vf_65522 | 0 | 0.01 | 0 | 13.32 | 0.02 | 0.24 | 0.06 | 0.12 | 0.07 | 8.69 | 28.98 | 0.06 | 0.25 | 0.29 | 1.05 | 5.99 | 0.1 | 0.25 | 0.29 | 0.41 | Shared |
| S8_vf_134943 | 0 | 0 | 0 | 0.35 | 0.03 | 0.31 | 0.14 | 0.21 | 0.2 | 6.35 | 62.14 | 0.1 | 0.71 | 2.94 | 8.25 | 0.36 | 0.2 | 0.2 | 0.27 | 0.8 | SHHC |
| S8_vf_117796 | 0.11 | 0 | 0 | 1.07 | 0.12 | 0.62 | 0.97 | 0.47 | 0.49 | 0.45 | 649.37 | 0.14 | 0.81 | 0.42 | 0.39 | 155.75 | 0.4 | 0.59 | 0.72 | 1.68 | Shared |
| S8_vf_13709 | 0.01 | 0 | 0 | 0.72 | 0.08 | 1.36 | 0.33 | 0.6 | 0.36 | 0.39 | 173.75 | 0.2 | 1.45 | 0.56 | 1.42 | 1.18 | 0.52 | 0.64 | 0.91 | 1.86 | SHHC |
| S8_vf_13758 | 0 | 0 | 0 | 1 | 0.1 | 1.83 | 0.25 | 0.67 | 0.36 | 0.4 | 63.22 | 0.21 | 2.55 | 0.43 | 3.11 | 1.02 | 0.82 | 0.7 | 1.12 | 1.89 | SHHC |
| S8_vf_170430 | 0.03 | 0 | 0 | 0.4 | 0.1 | 0.2 | 0.04 | 0.51 | 0.61 | 0.08 | 72.44 | 0.36 | 0.22 | 0.07 | 0.22 | 2.86 | 1.18 | 1.27 | 0.14 | 0.41 | SHHC |
| S8_vf_196670 | 0 | 0 | 0 | 0.78 | 0.03 | 0.37 | 0 | 0.17 | 0.08 | 10.41 | 380.74 | 0.03 | 0.21 | 0.15 | 0.15 | 0.18 | 0.19 | 0.22 | 0.14 | 0.81 | Shared |
| S8_vf_161703 | 0 | 0 | 0 | 0.88 | 0.01 | 0.68 | 0.09 | 0.33 | 0.13 | 0.28 | 60.5 | 0.15 | 1.18 | 1.7 | 0.77 | 0.41 | 0.91 | 0.41 | 1.15 | 0.98 | SHHC |
| S8_vf_197061 | 0 | 0 | 0 | 0.83 | 0.26 | 0.81 | 0.42 | 0.39 | 0.22 | 0.47 | 2027.94 | 0.11 | 0.18 | 0.11 | 0.32 | 32.25 | 0.33 | 0.21 | 0.26 | 2.62 | Shared |
| S8_vf_170878 | 0.01 | 0 | 0 | 28.58 | 0.04 | 4.35 | 0.23 | 4.33 | 4.32 | 1.5 | 111.55 | 0.46 | 4.52 | 3.13 | 3.23 | 14.8 | 1.64 | 9.36 | 2.05 | 3 | Shared |
| S8_vf_153696 | 0 | 0 | 0.01 | 174.48 | 6.44 | 2.12 | 0.05 | 0.1 | 0.07 | 9.98 | 28.46 | 14.49 | 0.12 | 5.85 | 0.14 | 1.8 | 0.06 | 0.34 | 1.74 | 8.36 | Shared |
| S8_vf_101283 | 0 | 0 | 0 | 1.06 | 0.01 | 2.99 | 1.1 | 0.8 | 0.1 | 0.67 | 39.33 | 0.69 | 1.39 | 0.28 | 2.63 | 0.83 | 9.89 | 0.24 | 2.93 | 1.4 | SHHC |
| S8_vf_23955 | 0 | 0 | 0 | 0.57 | 0.06 | 0.5 | 0.13 | 0.23 | 0.16 | 0.17 | 362.41 | 0.12 | 1.15 | 0.38 | 0.36 | 18.54 | 0.32 | 0.28 | 0.4 | 1.12 | Shared |
| S8_vf_136235 | 0 | 0 | 0 | 0.7 | 0.14 | 0.39 | 8.69 | 0.82 | 0.18 | 324.95 | 427.78 | 0.08 | 0.35 | 0.3 | 0.88 | 0.3 | 0.19 | 0.27 | 0.52 | 0.91 | Shared |
| S8_vf_187742 | 0 | 0 | 0 | 0.6 | 0.02 | 0.31 | 0.1 | 0.23 | 0.13 | 0.26 | 55.14 | 0.05 | 0.45 | 0.26 | 0.34 | 0.24 | 0.25 | 0.22 | 0.29 | 0.65 | SHHC |
| S8_vf_49282 | 0.01 | 0 | 0.02 | 4.81 | 4.16 | 248.25 | 70.57 | 0.23 | 0.13 | 0.17 | 42.12 | 0.48 | 0.76 | 0.28 | 0.54 | 0.46 | 0.28 | 0.29 | 0.42 | 0.69 | Shared |
| S8_vf_41376 | 0 | 0 | 0 | 0.52 | 0.06 | 0.48 | 0.1 | 0.27 | 0.19 | 0.18 | 183.42 | 0.1 | 0.74 | 0.27 | 0.6 | 0.3 | 0.3 | 0.25 | 0.63 | 0.87 | SHHC |
| S8_vf_7243 | 0 | 0 | 0 | 2.15 | 0.06 | 0.12 | 0.04 | 0.12 | 0.04 | 3 | 57.64 | 0.04 | 0.15 | 0.25 | 0.08 | 0.36 | 0.25 | 0.13 | 0.06 | 0.3 | SHHC |
| S8_vf_102508 | 0 | 0 | 0 | 0.31 | 0.02 | 0.14 | 1.03 | 0.14 | 0.08 | 0.26 | 82.09 | 0.03 | 0.17 | 0.2 | 0.26 | 0.15 | 0.11 | 0.26 | 0.16 | 0.32 | SHHC |
| S8_vf_93976 | 0 | 0 | 0 | 0.68 | 0.04 | 0.39 | 0 | 0.28 | 0.19 | 0.16 | 21.78 | 0.1 | 0.36 | 0.14 | 0.42 | 0.25 | 0.29 | 0.23 | 0.31 | 0.79 | SHHC |
| S8_vf_111790 | 0 | 0 | 0 | 3.67 | 0.14 | 1.28 | 0.76 | 1.2 | 1.14 | 0.7 | 182.23 | 12.26 | 0.25 | 1.25 | 0.98 | 1.15 | 3.61 | 13.85 | 4.58 | 4.25 | Shared |
| S8_vf_94361 | 0 | 0 | 0 | 0.52 | 0 | 0.22 | 0.01 | 0.12 | 0.12 | 0.03 | 14.82 | 0.02 | 0.12 | 0.09 | 0.16 | 0.24 | 0.15 | 0.23 | 0.16 | 0.56 | SHHC |
| S8_vf_16172 | 0 | 0 | 0 | 0.51 | 0.02 | 0.33 | 0.09 | 0.11 | 0.09 | 3.33 | 26.22 | 0.03 | 0.32 | 2.22 | 0.53 | 0.18 | 0.1 | 0.19 | 0.18 | 3.32 | SHHC |
| S8_vf_129423 | 0 | 0 | 0 | 0.54 | 0.03 | 0.51 | 0.02 | 0.67 | 0.24 | 0.49 | 36.18 | 0.06 | 0.65 | 0.43 | 0.4 | 0.33 | 2.35 | 0.35 | 0.63 | 0.65 | SHHC |
| S8_vf_163760 | 0 | 0 | 0 | 2.37 | 3.32 | 5.39 | 0.05 | 3.37 | 2.99 | 4.23 | 28581.73 | 0.07 | 3.02 | 0.22 | 0.28 | 2.54 | 0.25 | 0.24 | 0.21 | 38.7 | Shared |
| S8_vf_147128 | 0 | 0 | 0 | 0.71 | 0.09 | 0.63 | 0.11 | 67.37 | 160.79 | 0.28 | 44.28 | 2.19 | 1.1 | 0.43 | 0.56 | 2.47 | 0.37 | 0.43 | 0.49 | 0.94 | Shared |
| S8_vf_173225 | 0 | 0 | 0 | 0.98 | 0.03 | 0.7 | 0.15 | 0.32 | 0.23 | 0.18 | 61.09 | 0.09 | 0.94 | 0.34 | 0.55 | 8.07 | 0.41 | 0.41 | 0.62 | 1.27 | SHHC |
| S8_vf_147338 | 0 | 0 | 0 | 0.69 | 0.02 | 1.66 | 0.13 | 0.19 | 0.13 | 0.12 | 35.79 | 0.09 | 0.6 | 0.28 | 0.29 | 0.51 | 0.28 | 0.24 | 5.74 | 1.14 | SHHC |
| S8_vf_208102 | 0 | 0 | 0 | 4.5 | 0.37 | 0.88 | 0.1 | 0.47 | 0.36 | 0.41 | 2503.78 | 3.77 | 0.33 | 0.18 | 1.49 | 0.93 | 0.25 | 0.24 | 0.25 | 3.8 | SHHC |
| S8_vf_208107 | 0 | 0 | 0 | 0.92 | 0.91 | 1.67 | 0.05 | 0.96 | 1.04 | 1.04 | 7513.08 | 0.05 | 0.7 | 0.16 | 0.26 | 0.82 | 0.19 | 0.18 | 0.3 | 10.73 | Shared |
| S8_vf_52050 | 0 | 0.03 | 0 | 238.46 | 0.24 | 0.71 | 0.06 | 0.85 | 1.03 | 0.25 | 160.41 | 0.06 | 0.59 | 0.3 | 0.74 | 224.42 | 0.54 | 0.7 | 0.69 | 0.83 | Shared |
| S8_vf_208110 | 0 | 0 | 0 | 14.26 | 0.03 | 0.51 | 0.14 | 0.55 | 0.69 | 0.2 | 71.97 | 0.47 | 1.38 | 142.52 | 1.57 | 45.8 | 0.31 | 0.27 | 38.81 | 1 | Shared |
| S8_vf_208111 | 0 | 0.01 | 0 | 113.14 | 0.46 | 1.04 | 0.07 | 849.46 | 2330.71 | 0.62 | 45.62 | 0.03 | 1.22 | 3.59 | 0.18 | 21.69 | 0.21 | 0.35 | 0.2 | 0.49 | Shared |
| S8_vf_208115 | 0 | 0 | 0 | 1.3 | 0.29 | 0.81 | 0.03 | 0.43 | 0.19 | 0.14 | 722.11 | 0.14 | 0.74 | 0.08 | 0.3 | 1.29 | 0.16 | 0.3 | 0.85 | 2.77 | SHHC |
| S8_vf_208119 | 0 | 0 | 0 | 0.4 | 0 | 1.33 | 0.6 | 0.23 | 0.15 | 6.38 | 116.57 | 0.46 | 0.21 | 0.32 | 0.67 | 0.13 | 0.33 | 0.14 | 0.46 | 0.69 | SHHC |
| S8_vf_208120 | 0 | 0 | 0 | 0.55 | 0.08 | 1.75 | 0.27 | 0.2 | 0.26 | 0.22 | 495.83 | 0.09 | 0.44 | 36.88 | 1.47 | 0.26 | 0.2 | 0.26 | 0.63 | 1.61 | Shared |
| S8_vf_208126 | 0.01 | 0 | 0 | 0.72 | 0.07 | 0.3 | 0.1 | 0.29 | 0.1 | 0.22 | 190.78 | 0.1 | 1.51 | 1.1 | 4.24 | 0.4 | 0.27 | 0.25 | 8.76 | 0.74 | SHHC |
| S8_vf_208146 | 0 | 0 | 0 | 0.14 | 0.03 | 0.63 | 0.21 | 0.1 | 0.06 | 0.17 | 137.23 | 0.02 | 0.57 | 0.14 | 0.11 | 0.14 | 0.2 | 0.3 | 0.86 | 0.7 | SHHC |
| S8_vs_182288 | 0.01 | 0 | 0.01 | 16.44 | 0.25 | 94.49 | 11.17 | 2.58 | 0.46 | 23.79 | 1673.04 | 0.17 | 15.06 | 43.11 | 0.21 | 20.4 | 14.72 | 0.2 | 2.32 | 2.49 | Shared |
| S8_vs_9114 | 0 | 0 | 0 | 0.55 | 0.18 | 0.87 | 0.76 | 0.77 | 0.28 | 0.35 | 1462.33 | 0.25 | 0.61 | 0.19 | 1.41 | 0.43 | 0.52 | 0.46 | 0.44 | 2.6 | SHHC |
| S8_vs_35308 | 0.01 | 0 | 0 | 5.99 | 0.64 | 1.03 | 0.09 | 0.79 | 0.61 | 0.76 | 4701.49 | 0.07 | 0.99 | 0.26 | 1.15 | 1.13 | 0.17 | 0.43 | 0.6 | 6.09 | SHHC |
| S8_vs_88013 | 0 | 0 | 0 | 7.85 | 9.76 | 15.16 | 0.07 | 8.61 | 7.87 | 11.7 | 82495.66 | 0.13 | 3.93 | 0.23 | 0.38 | 6.96 | 0.17 | 0.43 | 0.26 | 107.13 | Shared |

| | | | | | | | | | | | | | | | | | | | | | | |
|---|---|---|---|---|---|---|---|---|---|---|---|---|---|---|---|---|---|---|---|---|---|---|
| S8_vs_9987 | 0.01 | 0 | 0.01 | 1.59 | 0.05 | 51.28 | 0.09 | 0.38 | 0.24 | 0.18 | 160.07 | 0.14 | 18.72 | 6.23 | 0.35 | 0.68 | 0.36 | 0.33 | 2.58 | 7.14 | Shared | |
| S8_vs_71212 | 0.01 | 0 | 0 | 1.42 | 0.14 | 0.43 | 0.06 | 0.42 | 0.24 | 0.34 | 941.1 | 0.12 | 0.82 | 0.1 | 0.42 | 0.46 | 0.26 | 0.55 | 0.54 | 1.96 | SHHC | |
| S8_vs_149700 | 0 | 0 | 0 | 6.56 | 0.32 | 16.31 | 0.66 | 30.63 | 3.35 | 9.84 | 2297.71 | 1.8 | 8.5 | 2.08 | 13.89 | 6.21 | 8.52 | 5.12 | 7.21 | 21.23 | Shared | |
| S8_vs_132359 | 0 | 0 | 0 | 0.89 | 0.06 | 0.43 | 0.08 | 0.33 | 0.2 | 0.17 | 42.96 | 0.12 | 0.67 | 0.52 | 0.47 | 0.3 | 0.42 | 0.54 | 0.95 | 0.93 | SHHC | |
| S8_vs_193843 | 0 | 0 | 0 | 0.67 | 0.05 | 0.43 | 0.09 | 0.26 | 0.15 | 0.18 | 532.17 | 0.09 | 0.43 | 0.91 | 0.2 | 0.34 | 0.17 | 0.26 | 0.25 | 1.27 | SHHC | |
| S8_vs_72117 | 0 | 0 | 0 | 1.52 | 1.4 | 2.53 | 0.01 | 1.34 | 1.19 | 1.65 | 9893.41 | 0.07 | 0.53 | 0.14 | 0.31 | 0.88 | 0.26 | 0.27 | 0.29 | 13.59 | Shared | |
| S8_vs_40014 | 0.01 | 0 | 0 | 1.03 | 0.35 | 7.95 | 0.77 | 0.54 | 0.36 | 1.32 | 3008.23 | 31.7 | 1.91 | 0.16 | 0.21 | 0.34 | 0.25 | 0.42 | 0.27 | 5.28 | Shared | |
| S8_vs_162053 | 0 | 0 | 0 | 2.04 | 1.3 | 2.67 | 0.07 | 1.37 | 0.83 | 1.66 | 8943.74 | 0.14 | 0.54 | 0.36 | 0.87 | 1.2 | 0.28 | 0.33 | 0.52 | 11.51 | Shared | |
| S8_vs_59736 | 0 | 0 | 0 | 1.08 | 0.08 | 4.66 | 0.24 | 0.5 | 0.34 | 0.4 | 135.66 | 0.37 | 1.74 | 1.19 | 1.54 | 16.7 | 0.71 | 1.11 | 1.45 | 2.5 | Shared | |
| S8_vs_51368 | 0 | 0 | 0 | 0.84 | 0.05 | 0.23 | 0.02 | 0.15 | 0.1 | 0.11 | 251.81 | 0.03 | 0.21 | 0.21 | 0.34 | 0.13 | 0.11 | 0.32 | 7.22 | 0.73 | SHHC | |
| S8_vs_208103 | 0 | 0 | 0 | 0.48 | 0.09 | 1.23 | 0.08 | 0.23 | 0.11 | 0.15 | 115.42 | 0.07 | 0.64 | 0.24 | 0.45 | 0.29 | 0.32 | 0.26 | 0.31 | 0.83 | SHHC | |
| S8_vs_208113 | 0 | 0 | 0 | 1.19 | 0.11 | 0.46 | 0.03 | 0.29 | 0.16 | 0.26 | 504.67 | 0.07 | 0.28 | 0.1 | 0.51 | 0.52 | 0.24 | 0.19 | 0.62 | 1.17 | SHHC | |
| S8_vs_208114 | 0.01 | 0 | 0 | 0.69 | 0.02 | 0.4 | 0.04 | 0.22 | 0.12 | 0.18 | 119.31 | 0.07 | 0.45 | 0.12 | 0.3 | 0.2 | 0.17 | 0.24 | 0.24 | 0.69 | SHHC | |
| S8_vs_208118 | 0 | 0 | 0 | 2.17 | 3.14 | 4.5 | 0.08 | 0.19 | 0.11 | 0.25 | 40.57 | 0.1 | 0.16 | 0.96 | 1.38 | 0.62 | 0.27 | 0.25 | 0.2 | 0.58 | SHHC | |
| S8_vs_208127 | 0.01 | 0.01 | 0 | 10.51 | 0.04 | 0.26 | 0.01 | 0.28 | 0.1 | 0.19 | 27.61 | 0.06 | 1.32 | 2.05 | 5.31 | 8.08 | 0.27 | 0.26 | 0.27 | 0.53 | Shared | |
| S8_vs_208129 | 0 | 0 | 0 | 0.87 | 0.02 | 0.64 | 0.09 | 0.33 | 0.16 | 0.21 | 33.99 | 0.13 | 0.45 | 0.74 | 0.44 | 0.43 | 0.48 | 0.3 | 0.5 | 1.21 | SHHC | |
| S8_vs_147670 | 0 | 0 | 0 | 0.42 | 0.01 | 0.36 | 0.04 | 0.23 | 0.08 | 0.12 | 19.15 | 0.06 | 0.28 | 0.11 | 0.22 | 2.44 | 0.22 | 0.21 | 0.24 | 0.63 | SHHC | |
| S8_vs_113059 | 0 | 0 | 0 | 0.71 | 0 | 0.16 | 0.42 | 0.11 | 0.05 | 0.04 | 16.32 | 0.03 | 0.1 | 0.02 | 0.15 | 0.44 | 0.06 | 0.2 | 0.34 | 0.36 | SHHC | |
| S8_vs_182331 | 0 | 0.04 | 0 | 80.63 | 0.07 | 0.64 | 0.05 | 20.4 | 29.42 | 0.18 | 26.5 | 0.1 | 0.74 | 0.51 | 0.38 | 23.85 | 0.19 | 0.34 | 0.42 | 0.58 | Shared | |
| S8_vs_147697 | 0.01 | 0 | 0 | 0.88 | 1.55 | 14.17 | 0.81 | 20.57 | 29.77 | 2.61 | 59.46 | 1.57 | 15.22 | 2.61 | 9.22 | 3.13 | 40.43 | 40.23 | 47.64 | 3.49 | Shared | |
| S8_vs_173961 | 0.01 | 0 | 0 | 0.9 | 0.15 | 11.98 | 0.14 | 0.35 | 0.24 | 0.21 | 127.79 | 0.17 | 0.97 | 0.57 | 1.31 | 0.46 | 0.6 | 0.34 | 0.49 | 1.11 | Shared | |
| S8_vs_139391 | 0 | 0 | 0 | 1.71 | 1.1 | 1.16 | 0.17 | 0.33 | 0.16 | 39.44 | 20.42 | 0.2 | 0.45 | 2.91 | 4.21 | 0.37 | 1.45 | 0.31 | 1.06 | 0.92 | Shared | |
| S8_vs_148389 | 0 | 0 | 0 | 0.75 | 0.05 | 0.32 | 0.7 | 0.21 | 0.18 | 0.15 | 110.53 | 0.07 | 0.31 | 0.11 | 0.32 | 0.25 | 0.18 | 0.33 | 0.29 | 0.71 | SHHC | |
| S8_vs_175235 | 0 | 0 | 0 | 0.47 | 0.05 | 0.29 | 0.05 | 0.27 | 0.17 | 0.16 | 380.47 | 0.1 | 0.19 | 0.14 | 0.28 | 0.24 | 0.2 | 0.18 | 0.22 | 1.14 | SHHC | |
| S8_vs_54183 | 0.01 | 0 | 0.01 | 7.16 | 0.06 | 1.22 | 0.2 | 1.27 | 0.2 | 0.8 | 58.27 | 0.3 | 0.71 | 0.47 | 0.98 | 1.75 | 0.8 | 0.66 | 1.39 | 1.05 | SHHC | |
| S8_vs_28311 | 0 | 0 | 0.01 | 0.93 | 0.65 | 25.08 | 0.3 | 0.53 | 0.19 | 1.63 | 20.79 | 0.13 | 57.84 | 1.24 | 2.61 | 2.23 | 21.04 | 0.31 | 2.74 | 8.31 | Shared | |
| S8_vs_132472 | 0 | 0 | 0 | 3.31 | 0.05 | 3.15 | 0.13 | 0.94 | 0.17 | 0.23 | 32.76 | 0.84 | 1.66 | 0.87 | 0.98 | 2.68 | 0.32 | 0.86 | 0.74 | 0.87 | SHHC | |
| S8_vs_63277 | 0 | 0 | 0 | 1.22 | 0.38 | 2.93 | 0.12 | 0.26 | 0.12 | 1.96 | 34.25 | 0.12 | 1.26 | 0.17 | 0.81 | 0.41 | 0.27 | 0.42 | 2.17 | 3.08 | SHHC | |
| S8_vs_176604 | 0 | 0 | 0 | 6.81 | 0.04 | 4.39 | 0.04 | 0.15 | 0.1 | 0.08 | 122.61 | 0.08 | 5.59 | 0.14 | 2.17 | 4.51 | 0.12 | 0.2 | 0.61 | 0.48 | SHHC | |
| S8_vs_176720 | 0 | 0 | 0.01 | 0.99 | 0.04 | 0.24 | 0.14 | 0.41 | 0.14 | 0.13 | 45.23 | 0.13 | 0.31 | 0.31 | 0.37 | 0.61 | 0.37 | 0.27 | 0.35 | 0.57 | SHHC | |
| S8_vs_142097 | 0 | 0 | 0 | 0.48 | 0.19 | 1.16 | 0.15 | 0.54 | 0.29 | 0.35 | 994.31 | 0.17 | 0.49 | 0.54 | 0.81 | 0.29 | 0.36 | 0.5 | 0.37 | 1.91 | SHHC | |
| S8_vs_3224 | 0 | 0 | 0 | 0.51 | 0.04 | 0.31 | 0.1 | 0.28 | 0.13 | 0.18 | 27.37 | 0.11 | 0.81 | 0.24 | 0.46 | 0.37 | 0.25 | 0.32 | 0.38 | 0.82 | SHHC | |
| S8_vs_115958 | 0 | 0 | 0 | 1.47 | 0.27 | 1.63 | 0.19 | 3.44 | 0.37 | 2.6 | 123.17 | 33.93 | 1.3 | 14.11 | 2.02 | 1.11 | 2.74 | 0.44 | 1.32 | 1.75 | Shared | |
| S8_vs_185041 | 0 | 0 | 0 | 5.49 | 0.46 | 0.61 | 0.48 | 0.89 | 0.18 | 10.88 | 69.21 | 0.23 | 23.3 | 0.37 | 5.25 | 5.04 | 1.65 | 1.12 | 0.95 | 1.67 | Shared | |
| S8_vs_99362 | 0.27 | 0.06 | 0 | 242.33 | 0.1 | 83.99 | 16.89 | 0.97 | 0.67 | 12.81 | 49.36 | 0.5 | 2.03 | 131.64 | 24.72 | 60.22 | 1.17 | 319.91 | 173.27 | 112.53 | Shared | |
| S8_vs_125717 | 0.01 | 0 | 0.01 | 6.09 | 0.16 | 3.12 | 0.3 | 9.76 | 1.17 | 2.19 | 371.03 | 1.11 | 6.5 | 1.37 | 16.91 | 15.47 | 8.3 | 2.46 | 1.63 | 2.72 | Shared | |
| S8_vs_4153 | 0.05 | 0 | 0 | 5.32 | 0.49 | 14.02 | 0.21 | 17.18 | 47.49 | 13.14 | 32.64 | 2.39 | 31.23 | 2.07 | 3.9 | 6.03 | 13.98 | 18.46 | 6.66 | 11.67 | Shared | |
| S8_vs_47265 | 0 | 0 | 0.01 | 3.71 | 0.04 | 37.2 | 24.18 | 2.15 | 3.77 | 10.58 | 38.41 | 0.42 | 18.68 | 0.45 | 0.55 | 1.87 | 3.02 | 1.85 | 13.25 | 3.99 | Shared | |
| S8_vs_143608 | 0 | 0 | 0 | 0.9 | 0.01 | 0.27 | 0.25 | 0.21 | 0.11 | 0.12 | 27.15 | 0.07 | 0.26 | 0.78 | 0.64 | 0.35 | 0.11 | 0.26 | 0.5 | 0.39 | SHHC | |
| S8_vs_134805 | 0.01 | 0 | 0 | 3.59 | 0.02 | 1.18 | 0.78 | 1.25 | 0.99 | 2.08 | 39.29 | 4.01 | 3.03 | 6.04 | 13.8 | 2.68 | 0.57 | 4.52 | 5.79 | 16.44 | Shared | |
| S8_vs_204260 | 0 | 0 | 0 | 0.9 | 0.03 | 0.46 | 0.14 | 0.39 | 0.18 | 0.18 | 202.23 | 0.06 | 0.51 | 0.2 | 0.42 | 0.28 | 0.34 | 0.24 | 0.26 | 1.08 | SHHC | |
| S8_vs_4883 | 0 | 0 | 0 | 1.14 | 0.07 | 0.43 | 0.02 | 0.2 | 0.12 | 0.1 | 117.37 | 0.06 | 0.41 | 0.38 | 0.37 | 0.13 | 0.33 | 0.22 | 0.2 | 0.82 | SHHC | |
| S8_vs_152553 | 0 | 0 | 0 | 0.39 | 0.01 | 0.62 | 0.1 | 0.62 | 0.31 | 0.18 | 78.59 | 0.06 | 0.48 | 0.79 | 0.5 | 0.24 | 0.31 | 0.18 | 3.37 | 0.99 | SHHC | |
| S8_vs_126582 | 0 | 0 | 0 | 0.43 | 0.04 | 0.75 | 0.15 | 0.24 | 0.16 | 2.58 | 80.03 | 0.06 | 0.75 | 0.68 | 0.44 | 0.42 | 0.35 | 0.28 | 0.4 | 0.97 | SHHC | |
| S8_vs_196247 | 0.01 | 0 | 0 | 0.86 | 0.04 | 0.38 | 0.03 | 0.2 | 0.13 | 0.15 | 53.78 | 0.11 | 0.45 | 0.11 | 0.81 | 0.4 | 0.13 | 0.3 | 0.28 | 0.76 | SHHC | |
| S8_vs_5211 | 0.17 | 0 | 0 | 2.91 | 0.09 | 2.73 | 0.08 | 8.21 | 9.31 | 0.16 | 40 | 0.16 | 0.45 | 13.34 | 4.24 | 1.23 | 3.23 | 0.64 | 0.44 | 1.17 | Shared | |
| S8_vs_179208 | 0 | 0 | 0.01 | 6.58 | 1.78 | 110.5 | 1.39 | 46.17 | 19.74 | 33.19 | 146.95 | 4.8 | 25.64 | 13.24 | 111.3 | 5.91 | 21.48 | 78.21 | 77.18 | 44.1 | Shared | |
| S8_vs_179529 | 0 | 0 | 0 | 1.09 | 0.03 | 6.36 | 1.1 | 3.5 | 0.17 | 3.52 | 34.56 | 0.64 | 1.87 | 0.25 | 1.03 | 0.9 | 0.76 | 4.09 | 14.77 | 3.65 | Shared | |
| S8_vs_205257 | 0 | 0 | 0 | 7.1 | 0.28 | 6.79 | 0.52 | 14.87 | 1.27 | 5.71 | 43.48 | 2.9 | 4.47 | 163.84 | 12.19 | 15.02 | 4.42 | 2.85 | 5.56 | 5.89 | Shared | |
| S8_vs_145173 | 0 | 0 | 0.01 | 0.53 | 0.14 | 0.27 | 0.9 | 0.47 | 0.12 | 0.47 | 42.04 | 0.18 | 0.16 | 3.91 | 0.43 | 0.48 | 1.44 | 0.28 | 0.65 | 2.49 | SHHC | |
| S8_vs_110692 | 0 | 0 | 0 | 0.49 | 0.04 | 5.39 | 0.19 | 0.12 | 0.05 | 0.09 | 28.54 | 0.13 | 0.12 | 0.22 | 0.84 | 0.18 | 0.15 | 0.3 | 0.22 | 0.48 | SHHC | |
| S8_vs_154231 | 0 | 0 | 0 | 0.58 | 0.02 | 0.29 | 0.04 | 0.3 | 0.12 | 0.26 | 26.04 | 0.14 | 0.85 | 1.47 | 0.65 | 0.35 | 3.08 | 0.2 | 4.5 | 1.12 | SHHC | |
| S8_vs_136922 | 0 | 0 | 0 | 0.7 | 0.05 | 0.41 | 1.26 | 0.34 | 0.18 | 0.19 | 77.29 | 0.13 | 0.67 | 0.19 | 0.55 | 0.3 | 0.54 | 0.36 | 0.51 | 1.19 | SHHC | |
| S8_vs_102608 | 0.01 | 0 | 0 | 0.47 | 0.02 | 0.26 | 0.04 | 0.34 | 0.08 | 0.21 | 214.48 | 0.08 | 0.2 | 0.18 | 0.2 | 0.32 | 0.13 | 0.17 | 5.03 | 0.83 | SHHC | |
| S8_vs_206526 | 0 | 0.01 | 0 | 1.19 | 0.04 | 2.75 | 0.08 | 0.59 | 0.83 | 0.24 | 78.5 | 0.09 | 0.61 | 0.18 | 4.21 | 0.21 | 7.62 | 0.15 | 2.58 | 28.72 | Shared | |
| S8_vs_76852 | 0 | 0 | 0 | 0.9 | 0.05 | 38.97 | 0.17 | 0.23 | 0.16 | 8.64 | 46.95 | 0.1 | 0.51 | 1.49 | 0.57 | 0.12 | 0.18 | 0.59 | 0.16 | 0.5 | Shared | |
| S8_vs_50431 | 0.01 | 0 | 0 | 1.24 | 0.02 | 0.54 | 0.06 | 0.28 | 0.09 | 0.11 | 27.11 | 0.08 | 0.2 | 0.18 | 0.4 | 0.15 | 0.17 | 0.24 | 1.04 | 0.45 | SHHC | |
| #Total | 0.04 | 0.01 | 0.04 | 28.54 | 141.38 | 312.22 | 102.3 | 79.82 | 145.28 | 170.25 | 316.92 | 154.77 | 53.29 | 86.9 | 101.08 | 80.04 | 61.71 | 28.74 | 106.89 | 14.41 | 116.738 | mean |
| | | | | | | | | | | | | | | | | | | | | | 87.0401 | SD |

WMS_avg_cov

| vOTUs | S1_wms_ME | S2_wms_SHHC | S3_wms_ME | S4_wms_SHHC | S5_wms_ME | S6_wms_SHHC | S7_wms_ME | S8_wms_SHHC | S9_wms_ME | S11_wms_ME | S12_wms_SHHC | S17_wms_ME | Status |
|---|---|---|---|---|---|---|---|---|---|---|---|---|---|
| S9-wms-combined_27311 | 0.88 | 0.82 | 0.11 | 0.31 | 1.16 | 0.2 | 0.76 | 0.89 | 15.21 | 0.46 | 1.15 | 0.62 | ME |
| S9-wms-combined_64277 | 2.1 | 1.02 | 0.23 | 0.36 | 2.2 | 0.25 | 2.57 | 0.73 | 130.53 | 0.51 | 1.31 | 0.94 | ME |
| S9-wms-combined_10990 | 17.51 | 57.54 | 1.45 | 8.53 | 12.84 | 5.41 | 30.03 | 6.66 | 151.11 | 6.96 | 24.21 | 17.38 | Shared |
| S9-wms-combined_53991 | 28.18 | 24.59 | 0.09 | 0.72 | 2.81 | 5.09 | 37.75 | 0.74 | 65.91 | 0.56 | 1.16 | 13.93 | Shared |
| S9-wms-combined_97755 | 7.8 | 8.83 | 0.72 | 1.77 | 4.07 | 1.2 | 3.26 | 3.45 | 93.72 | 5.09 | 3.65 | 3.04 | ME |
| S9-wms-combined_33230 | 33.59 | 70.71 | 1.95 | 7.52 | 61.22 | 3.44 | 43.15 | 14.97 | 277.62 | 13.51 | 26.2 | 22.79 | Shared |
| S9-wms-combined_2042 | 4.48 | 6.65 | 0.49 | 1.22 | 3.4 | 1.1 | 2.69 | 3.25 | 98.02 | 3.11 | 3.38 | 2.29 | ME |
| S9-wms-combined_124857 | 35.21 | 50.02 | 3.13 | 4.12 | 39.54 | 27.05 | 12.9 | 31.53 | 61.99 | 41.14 | 25.04 | 139.43 | Shared |
| S9-wms-combined_28805 | 8.88 | 7.24 | 0.19 | 1.73 | 3.67 | 1.33 | 6.54 | 7.46 | 33.68 | 3.5 | 8.91 | 2.75 | ME |
| S9-wms-combined_71647 | 0.82 | 1.8 | 0.06 | 0.79 | 1.22 | 0.16 | 1.64 | 0.44 | 16.7 | 0.36 | 0.72 | 0.48 | ME |
| S9-wms-combined_93232 | 1.06 | 0.99 | 0.06 | 0.42 | 1.37 | 0.19 | 1.06 | 0.63 | 22.09 | 0.82 | 0.81 | 0.58 | ME |
| S9-wms-combined_109322 | 13.34 | 25.83 | 1.02 | 8 | 4.05 | 7.85 | 19.06 | 4.43 | 151.24 | 3.49 | 6.45 | 13.13 | Shared |
| S9-wms-combined_3461 | 1.2 | 1.55 | 0.11 | 0.39 | 1.17 | 0.37 | 1.51 | 0.85 | 15.71 | 0.38 | 1.17 | 0.82 | ME |
| S9-wms-combined_29611 | 33.41 | 39.63 | 3.03 | 3.9 | 31.36 | 21.19 | 13.8 | 24.3 | 50.12 | 34.52 | 22.07 | 113.99 | Shared |
| S9-wms-combined_25007 | 6.76 | 7.85 | 0.4 | 1.21 | 11.39 | 1.41 | 14.72 | 3.18 | 27.45 | 10.82 | 3.53 | 4.29 | ME |
| S9-wms-combined_3878 | 0.31 | 0.7 | 0.03 | 0.27 | 0.34 | 0.05 | 0.53 | 0.32 | 24.59 | 0.18 | 0.72 | 0.34 | ME |
| S9-wms-combined_89249 | 6.69 | 1.71 | 0.26 | 1.1 | 1.04 | 0.37 | 2.95 | 0.75 | 15.4 | 1.5 | 2.87 | 3.17 | ME |
| S9-wms-combined_105462 | 1.65 | 3.79 | 0.16 | 1.46 | 1.29 | 0.62 | 1.9 | 0.67 | 33.54 | 0.53 | 5.87 | 1.44 | ME |
| S9-wms-combined_20485 | 3.85 | 4.93 | 0.13 | 2.35 | 3.77 | 0.24 | 2.26 | 0.76 | 59.95 | 0.51 | 0.82 | 2.94 | ME |
| S9-wms-combined_9931 | 3.17 | 4.91 | 0.2 | 1.25 | 6.02 | 0.94 | 3.64 | 15.44 | 116.02 | 1.07 | 2.26 | 1.19 | Shared |
| S9-wms-combined_36459 | 0.82 | 0.86 | 0.05 | 0.5 | 1.14 | 2.7 | 0.92 | 0.78 | 16.07 | 0.39 | 0.92 | 0.64 | ME |
| S9-wms-combined_105822 | 1.52 | 1.62 | 0.51 | 0.67 | 1.64 | 0.54 | 2.63 | 0.89 | 36.66 | 1.6 | 1.17 | 2.02 | ME |
| S9-wms-combined_26410 | 3.18 | 2.92 | 0.12 | 0.92 | 1.34 | 1.21 | 1.56 | 0.83 | 29.97 | 1.01 | 2.75 | 1.3 | ME |
| S9-wms-combined_31456 | 11.35 | 4.16 | 0.53 | 2.46 | 39.66 | 5.04 | 29.1 | 7.3 | 165.36 | 5.08 | 12.84 | 9.03 | Shared |
| S9-wms-combined_37587 | 16.93 | 40.38 | 0.95 | 6.61 | 29.05 | 2.39 | 17.03 | 5.97 | 289.89 | 5.26 | 18.45 | 10.28 | Shared |
| S9-wms-combined_5526 | 135.59 | 50.51 | 3.16 | 2.28 | 275.21 | 163.01 | 89.43 | 86.71 | 313.82 | 102.89 | 107.44 | 29.54 | Shared |
| S9-wms-combined_33229 | 0.83 | 0.74 | 0.18 | 0.51 | 0.69 | 0.16 | 1.05 | 0.37 | 296.16 | 0.22 | 1.06 | 0.48 | ME |
| S9-wms-combined_76274 | 6.5 | 1.94 | 8.31 | 13.41 | 32.05 | 26 | 8.69 | 2.56 | 98.67 | 1.28 | 2.38 | 4.51 | Shared |
| S9-wms-combined_44402 | 20.19 | 26.47 | 0.24 | 0.6 | 2.05 | 0.16 | 1.05 | 0.45 | 382.37 | 0.71 | 1.59 | 0.74 | Shared |
| S9-wms-combined_56773 | 0.73 | 2.51 | 0.18 | 1.2 | 0.58 | 0.21 | 1.02 | 0.41 | 147.32 | 0.2 | 0.81 | 0.84 | ME |
| S9-wms-combined_122742 | 58.05 | 28.66 | 5.84 | 2.85 | 99.4 | 33.42 | 6.72 | 23.2 | 75.29 | 21.13 | 35.02 | 16.88 | Shared |
| S9-wms_64472 | 9.38 | 10.07 | 0.72 | 1.8 | 4.43 | 3.08 | 6.93 | 1.66 | 34.5 | 3.56 | 5.79 | 11.1 | Shared |
| S9-wms_104169 | 1.13 | 1.55 | 0.09 | 0.42 | 3.55 | 0.52 | 0.29 | 0.54 | 17.23 | 0.62 | 0.26 | 0.22 | ME |
| S9-wms_19025 | 0.33 | 0.79 | 0.04 | 0.48 | 0.39 | 0.08 | 0.62 | 0.19 | 25.74 | 0.14 | 0.44 | 0.22 | ME |
| S9-wms_114971 | 0.45 | 0.6 | 0.04 | 0.45 | 0.41 | 0.05 | 0.53 | 0.18 | 17.95 | 0.14 | 0.33 | 0.26 | ME |
| S9-wms_10480 | 0.55 | 0.35 | 0.06 | 0.08 | 1.31 | 0.71 | 0.24 | 1.68 | 7.31 | 0.46 | 1.47 | 0.68 | ME |
| S9-wms_128232 | 0.52 | 1 | 0.27 | 1.41 | 0.77 | 0.22 | 0.68 | 0.47 | 26.4 | 0.19 | 0.91 | 0.32 | ME |
| S11-wms-combined_41367 | 0.84 | 0.95 | 0.06 | 0.29 | 0.87 | 1.43 | 1.64 | 0.42 | 0.44 | 43.76 | 1.09 | 1.45 | ME |
| S11-wms-combined_193148 | 9.1 | 1.47 | 0.16 | 0.24 | 1.63 | 60.11 | 1.38 | 2.53 | 0.89 | 76.13 | 1.85 | 1.21 | Shared |
| S11-wms-combined_179605 | 11.48 | 11.07 | 0.83 | 2.52 | 10.53 | 3.98 | 6.28 | 6.42 | 10.36 | 52.43 | 6.27 | 24.47 | Shared |
| S11-wms-combined_221310 | 1.44 | 1.06 | 0.16 | 0.34 | 2.44 | 0.65 | 9.83 | 1.74 | 0.6 | 23.55 | 1.53 | 1 | ME |
| S11-wms-combined_14636 | 1.96 | 1.46 | 0.25 | 0.64 | 4.87 | 0.79 | 2.08 | 2.63 | 0.97 | 39.06 | 2.72 | 1.55 | ME |
| S11-wms-combined_83571 | 25.22 | 0.63 | 0.98 | 1.86 | 114.16 | 51.63 | 94.59 | 21.46 | 1.29 | 42.44 | 8.46 | 0.86 | Shared |
| S11-wms-combined_125176 | 206.31 | 34.33 | 8.66 | 17.78 | 388.14 | 79.59 | 232.47 | 115.11 | 22.31 | 136.18 | 80.22 | 23.9 | Shared |
| S11-wms-combined_304239 | 1.54 | 1.32 | 0.19 | 0.4 | 2.38 | 0.7 | 1.61 | 1.41 | 1.01 | 44.5 | 1.54 | 1.74 | ME |
| S11-wms-combined_194205 | 5.36 | 5.68 | 1 | 1.08 | 11.98 | 1.75 | 13.5 | 9.64 | 3.23 | 70.88 | 12.65 | 2.36 | Shared |
| S11-wms-combined_84326 | 2.19 | 1.32 | 0.2 | 7.01 | 2.53 | 0.74 | 1.26 | 2.5 | 0.77 | 53.65 | 2.01 | 1.71 | ME |
| S11-wms-combined_236214 | 76.68 | 10.46 | 7.55 | 10.43 | 7.02 | 9.79 | 8.38 | 35.26 | 1.74 | 43.5 | 25.36 | 79.22 | Shared |
| S11-wms-combined_264050 | 38.92 | 18.29 | 3.59 | 10.15 | 11.36 | 2.98 | 14.54 | 22.33 | 3.45 | 43.84 | 22.9 | 31.05 | Shared |
| S11-wms-combined_112147 | 1.07 | 1.2 | 0.09 | 0.64 | 1.64 | 0.23 | 0.76 | 1.16 | 0.68 | 39.39 | 1.47 | 0.87 | ME |
| S11-wms-combined_140009 | 27.63 | 8.9 | 4.44 | 9.92 | 3.67 | 2.95 | 7.92 | 16.38 | 1.66 | 61.48 | 36.71 | 97.16 | Shared |
| S11-wms-combined_168048 | 4.51 | 1.76 | 0.23 | 1.53 | 1.68 | 0.45 | 8.11 | 4.26 | 3.39 | 42.99 | 12.48 | 0.76 | Shared |
| S11-wms-combined_195425 | 5.24 | 5.39 | 0.65 | 9.55 | 1.38 | 0.39 | 1.24 | 1.11 | 0.84 | 21.05 | 1.52 | 119.63 | ME |
| S11-wms-combined_112733 | 2.16 | 1.55 | 0.23 | 13.99 | 2.97 | 0.91 | 1.55 | 2.15 | 0.86 | 106.45 | 1.99 | 1.9 | Shared |
| S11-wms-combined_58538 | 1.08 | 0.91 | 0.09 | 0.22 | 1.3 | 0.32 | 0.66 | 1 | 0.72 | 20.43 | 0.71 | 0.79 | ME |
| S11-wms-combined_154845 | 0.66 | 0.56 | 0.09 | 0.68 | 1.12 | 0.21 | 0.39 | 0.91 | 0.52 | 31.66 | 0.76 | 0.47 | ME |
| S11-wms-combined_17029 | 0.82 | 0.93 | 0.08 | 0.39 | 0.91 | 0.31 | 0.75 | 0.33 | 0.51 | 103.68 | 2.97 | 1.23 | ME |
| S11-wms-combined_3507 | 117.01 | 25.97 | 10.46 | 4.19 | 328 | 9.16 | 28.85 | 48.94 | 5.45 | 100.16 | 68.16 | 29.42 | Shared |
| S11-wms-combined_265453 | 1.41 | 1.32 | 0.05 | 0.12 | 0.7 | 1.46 | 1.04 | 0.29 | 1.55 | 10.46 | 1.02 | 1.75 | ME |
| S11-wms-combined_320710 | 5.44 | 7.82 | 0.36 | 3.26 | 7.02 | 2.51 | 8.95 | 2.59 | 5.34 | 49.99 | 4.62 | 3.43 | ME |
| S11-wms-combined_101337 | 3.31 | 1.52 | 0.19 | 0.49 | 1.59 | 0.97 | 6.92 | 1.81 | 1.09 | 114.79 | 11.25 | 19.65 | Shared |
| S11-wms-combined_73204 | 190.09 | 107.07 | 4.65 | 1.07 | 430.65 | 12.36 | 100.05 | 18.78 | 8.16 | 177.05 | 122.38 | 36.99 | Shared |
| S11-wms-combined_46816 | 6.46 | 30.95 | 1.27 | 14.73 | 2.15 | 0.5 | 1.33 | 21.2 | 0.65 | 56.18 | 1.72 | 1.16 | Shared |
| S11-wms-combined_226165 | 0.84 | 0.52 | 0.09 | 0.19 | 99.07 | 8.92 | 0.35 | 1.02 | 0.39 | 55.44 | 12.72 | 0.86 | Shared |

| | | | | | | | | | | | | | |
|---|---|---|---|---|---|---|---|---|---|---|---|---|---|
| S11-wms-combined_321938 | 48.96 | 74.68 | 2.03 | 5.24 | 23.04 | 10.54 | 66.69 | 49.16 | 1.67 | 57.85 | 39.56 | 27.22 | Shared |
| S11-wms-combined_198512 | 23.18 | 7.26 | 0.73 | 1.5 | 14.79 | 1.78 | 12.61 | 10.66 | 1.49 | 43.55 | 9.83 | 10.92 | Shared |
| S11-wms-combined_282227 | 2.41 | 4.51 | 0.28 | 1.38 | 6.45 | 0.54 | 13.05 | 1.79 | 6.8 | 20.91 | 2.63 | 2.36 | ME |
| S11-wms-combined_267993 | 1.78 | 1.25 | 0.15 | 0.26 | 4.23 | 0.4 | 2.01 | 2.27 | 0.76 | 27.61 | 2.14 | 1.2 | ME |
| S11-wms-combined_74983 | 2.12 | 1.21 | 0.17 | 0.72 | 2.19 | 18.72 | 14.09 | 1.03 | 0.56 | 18.95 | 96.89 | 143.89 | Shared |
| S11-wms-combined_116589 | 19.19 | 23.94 | 2.28 | 2.41 | 19.06 | 16.54 | 14.65 | 19.17 | 13.79 | 51.53 | 25.6 | 74.52 | Shared |
| S11-wms-combined_48246 | 1.9 | 1.25 | 1.24 | 1.02 | 1.2 | 1.4 | 2.03 | 9.22 | 1.26 | 85.89 | 37.53 | 0.57 | Shared |
| S11-wms-combined_283220 | 4.06 | 4.72 | 0.24 | 0.33 | 3.02 | 2.01 | 1.52 | 2.31 | 1.08 | 71.02 | 2.48 | 1.76 | ME |
| S11-wms-combined_172889 | 1.92 | 1.11 | 24.16 | 0.56 | 1.94 | 0.53 | 9.44 | 2.65 | 1.21 | 137.2 | 1.46 | 1.77 | ME |
| S11-wms-combined_268967 | 2.48 | 1.78 | 0.32 | 3.12 | 3.77 | 0.62 | 1.22 | 2.68 | 1.05 | 62.44 | 2.83 | 1.7 | ME |
| S11-wms-combined_200084 | 9.02 | 0.93 | 0.14 | 0.49 | 0.9 | 0.21 | 1.23 | 0.72 | 0.99 | 27.55 | 2 | 0.68 | ME |
| S11-wms-combined_117756 | 1.4 | 0.94 | 0.27 | 0.59 | 1.72 | 0.48 | 1.07 | 1.22 | 1.72 | 21.57 | 1.31 | 0.85 | ME |
| S11-wms-combined_49632 | 2.31 | 2.78 | 0.43 | 0.17 | 2.03 | 2.87 | 4.21 | 1.64 | 0.9 | 31.06 | 4.11 | 7.39 | ME |
| S11-wms-combined_63818 | 69.86 | 23.88 | 4.89 | 3.74 | 298.95 | 97.54 | 59.98 | 60.58 | 41.81 | 324.38 | 138.49 | 32.26 | Shared |
| S11-wms-combined_215723 | 291.55 | 83.24 | 2.87 | 2.18 | 183.18 | 174.54 | 103.94 | 103.06 | 100.6 | 101.52 | 92.54 | 31.27 | Shared |
| S11-wms-combined_64392 | 2.46 | 1.94 | 0.14 | 0.43 | 8.43 | 2.43 | 7.09 | 14.62 | 0.83 | 77.15 | 11.58 | 1.53 | Shared |
| S11-wms-combined_92114 | 85.13 | 37.47 | 6.97 | 3.77 | 342.54 | 115.61 | 69.82 | 69.07 | 60.4 | 143.84 | 144.77 | 71.81 | Shared |
| S11-wms-combined_119215 | 3.04 | 3.99 | 0.33 | 0.4 | 4.55 | 1.56 | 1.97 | 2.8 | 1.37 | 29.37 | 3.11 | 5.45 | ME |
| S11-wms-combined_120257 | 17.24 | 13.23 | 2.18 | 1.51 | 27.36 | 6.5 | 6.8 | 13.6 | 3.19 | 31.82 | 11.46 | 23.69 | Shared |
| S11-wms-combined_299954 | 2.12 | 6.17 | 0.22 | 0.69 | 3.09 | 1.12 | 1.87 | 1.98 | 0.83 | 38.28 | 2.98 | 6.76 | ME |
| S11-wms-combined_217790 | 87.87 | 24.33 | 2.28 | 0.73 | 184.19 | 183.26 | 65.46 | 50.73 | 64.93 | 46.56 | 41.99 | 16.13 | Shared |
| S11-wms-combined_204522 | 1.01 | 0.95 | 0.06 | 0.3 | 0.88 | 3.4 | 1.02 | 0.62 | 0.69 | 31.2 | 1.03 | 0.49 | ME |
| S11-wms-combined_329493 | 81.49 | 50.92 | 9 | 2.81 | 68.88 | 6.65 | 18.67 | 36.5 | 3.11 | 48.06 | 29.17 | 14.07 | Shared |
| S11-wms_303398 | 3.13 | 49.74 | 0.25 | 0.42 | 2.31 | 8.41 | 3.17 | 3.66 | 0.98 | 27.4 | 3.75 | 8.71 | Shared |
| S11-wms_55610 | 1.5 | 19.96 | 0.22 | 0.67 | 1.79 | 0.5 | 3.75 | 1.74 | 0.45 | 18.05 | 1.53 | 1.58 | Shared |
| S11-wms_14390 | 2.48 | 3.11 | 0.32 | 0.81 | 2.45 | 7.96 | 4.11 | 5.34 | 0.84 | 494.02 | 5.61 | 1.4 | ME |
| S11-wms_3116 | 50.14 | 38.44 | 7.12 | 5.48 | 5.42 | 40.93 | 7.29 | 134.78 | 3.27 | 73.94 | 56.6 | 32.22 | Shared |
| S11-wms_320555 | 0.47 | 0.85 | 0.06 | 0.39 | 0.94 | 0.14 | 0.84 | 0.52 | 0.79 | 24.92 | 0.83 | 0.43 | ME |
| S11-wms_87293 | 0.53 | 0.9 | 0.19 | 0.64 | 0.86 | 0.18 | 0.6 | 0.47 | 0.62 | 362.52 | 1.41 | 0.4 | ME |
| S11-wms_32691 | 0.95 | 9.43 | 0.08 | 0.43 | 1.02 | 0.24 | 0.93 | 0.69 | 0.47 | 49.82 | 1.08 | 0.49 | ME |
| S11-wms_6203 | 0.67 | 1.68 | 0.06 | 0.59 | 1.96 | 0.24 | 1.14 | 1.16 | 2.17 | 38.97 | 0.78 | 0.39 | ME |
| S11-wms_330479 | 0.97 | 1.15 | 2.53 | 0.32 | 1.17 | 0.17 | 0.94 | 0.73 | 0.55 | 211.64 | 0.96 | 0.67 | ME |
| S1-wms-combined_324053 | 137.39 | 0.88 | 0.09 | 0.49 | 0.92 | 8.43 | 0.87 | 0.66 | 0.79 | 0.33 | 0.98 | 0.52 | ME |
| S1-wms-combined_394574 | 67.22 | 850.69 | 0.59 | 4.18 | 0.83 | 2.24 | 1.22 | 0.71 | 0.73 | 0.85 | 1.28 | 0.68 | Shared |
| S1-wms-combined_394628 | 73.85 | 0.52 | 0.09 | 0.39 | 0.47 | 0.23 | 1.54 | 0.77 | 0.32 | 0.31 | 1.84 | 0.36 | ME |
| S1-wms-combined_246923 | 24.55 | 0.61 | 0.11 | 0.52 | 0.89 | 0.65 | 0.94 | 0.89 | 0.46 | 11.71 | 9.47 | 23.95 | ME |
| S1-wms-combined_263562 | 161.54 | 530.16 | 1.47 | 0.59 | 1.07 | 0.25 | 36.7 | 33.56 | 0.69 | 7.35 | 10.13 | 0.66 | Shared |
| S1-wms-combined_164894 | 70.03 | 2.63 | 6.79 | 3.95 | 2.84 | 0.96 | 1.86 | 2.92 | 1.01 | 1.78 | 4.5 | 2.1 | ME |
| S1-wms-combined_231234 | 39.45 | 0.52 | 0.04 | 0.45 | 0.75 | 0.07 | 0.72 | 0.34 | 0.27 | 0.25 | 0.37 | 0.25 | ME |
| S1-wms-combined_231411 | 37.6 | 0.95 | 0.06 | 0.76 | 1.01 | 0.28 | 0.6 | 2.81 | 0.24 | 0.49 | 1.13 | 0.77 | ME |
| S1-wms-combined_132934 | 66.35 | 13.33 | 2.09 | 2.11 | 5.35 | 2.85 | 6.76 | 12.06 | 2.58 | 17.18 | 10.31 | 5.76 | Shared |
| S1-wms-combined_17651 | 32.66 | 10.38 | 0.69 | 1.11 | 12.38 | 0.6 | 1.49 | 4.26 | 0.71 | 2.61 | 17.77 | 3.17 | Shared |
| S1-wms-combined_313851 | 17.95 | 0.74 | 0.04 | 0.42 | 0.39 | 0.13 | 0.53 | 0.33 | 0.64 | 0.16 | 0.77 | 0.47 | ME |
| S1-wms-combined_330329 | 41.33 | 1.21 | 0.23 | 0.64 | 0.69 | 0.16 | 0.66 | 1.23 | 0.58 | 1.54 | 0.72 | 0.64 | ME |
| S1-wms-combined_116938 | 35.08 | 10.59 | 2.93 | 5.56 | 5.79 | 3.25 | 6.09 | 9.86 | 1.49 | 8.34 | 11.87 | 18.62 | Shared |
| S1-wms-combined_248622 | 60.56 | 71.71 | 0.47 | 1.45 | 0.42 | 0.4 | 7.16 | 0.49 | 1.09 | 0.47 | 2.2 | 9.01 | Shared |
| S1-wms-combined_100430 | 32.25 | 0.92 | 3.45 | 8.49 | 1.24 | 0.16 | 1.02 | 0.65 | 1.06 | 0.35 | 0.89 | 15.01 | ME |
| S1-wms-combined_166392 | 49.52 | 6.16 | 2.93 | 0.64 | 2.55 | 0.47 | 4.19 | 3.94 | 0.82 | 1.07 | 1.7 | 22.11 | ME |
| S1-wms-combined_19031 | 38.38 | 0.92 | 0.69 | 0.43 | 1.8 | 1.09 | 2.1 | 2.07 | 0.63 | 2.24 | 4.22 | 1.18 | ME |
| S1-wms-combined_118167 | 61.2 | 1.05 | 0.37 | 0.62 | 1.82 | 11.97 | 1.55 | 1.85 | 0.68 | 0.77 | 2.41 | 1 | Shared |
| S1-wms-combined_331896 | 55.48 | 3.34 | 0.95 | 1.97 | 3.21 | 1.23 | 3.22 | 2.25 | 4.66 | 0.67 | 24.02 | 4.41 | Shared |
| S1-wms-combined_332236 | 179.6 | 10.94 | 2.46 | 2.86 | 38.85 | 3.16 | 44.43 | 8.53 | 2.9 | 49.35 | 17.77 | 5.46 | Shared |
| S1-wms-combined_151708 | 36.81 | 1.66 | 0.6 | 0.82 | 1.74 | 0.27 | 1.4 | 9.53 | 0.92 | 0.46 | 2.27 | 1.44 | ME |
| S1-wms-combined_52643 | 27.33 | 13.95 | 0.08 | 0.48 | 1.34 | 0.24 | 0.86 | 0.91 | 0.8 | 0.52 | 1.07 | 1 | Shared |
| S1-wms-combined_52708 | 210.91 | 129.55 | 16.07 | 28.68 | 11.15 | 18.8 | 16.15 | 97.75 | 3.57 | 67.73 | 64.59 | 203.77 | Shared |
| S1-wms-combined_185336 | 36.86 | 1.61 | 0.34 | 0.27 | 2.63 | 0.13 | 4.81 | 1.73 | 0.32 | 0.32 | 1.26 | 0.26 | ME |
| S1-wms-combined_68998 | 49.4 | 2.75 | 0.98 | 1.85 | 1.66 | 0.83 | 3.21 | 2.1 | 0.9 | 1.01 | 3.65 | 2.44 | ME |
| S1-wms-combined_102801 | 42.57 | 62.59 | 1.59 | 0.84 | 8.43 | 1.77 | 12.26 | 7.15 | 9.23 | 37.13 | 41 | 24.01 | Shared |
| S1-wms-combined_120533 | 53.35 | 4.76 | 1.68 | 2.55 | 2.64 | 2.34 | 5.16 | 50.74 | 3.26 | 5.23 | 15.95 | 8.29 | Shared |
| S1-wms-combined_252439 | 19.05 | 2.6 | 0.04 | 0.12 | 0.84 | 0.12 | 0.56 | 0.85 | 0.47 | 0.47 | 1.27 | 0.64 | ME |
| S1-wms-combined_302166 | 322.93 | 4.13 | 3.07 | 1.23 | 6.98 | 1.38 | 29.18 | 112.39 | 2.48 | 4.29 | 91.36 | 3.67 | Shared |
| S1-wms-combined_285012 | 61.82 | 3.09 | 1.53 | 1.69 | 2.68 | 0.56 | 2.47 | 42.07 | 0.78 | 1.99 | 11.92 | 1.11 | Shared |
| S1-wms-combined_186878 | 68.99 | 1.2 | 0.09 | 1.18 | 1.53 | 0.32 | 1.45 | 1 | 0.88 | 0.56 | 0.91 | 0.61 | ME |
| S1-wms-combined_153670 | 58.04 | 0.46 | 0.07 | 0.14 | 0.88 | 0.32 | 1.53 | 0.56 | 1.27 | 0.48 | 0.87 | 0.59 | ME |
| S1-wms-combined_351889 | 73.16 | 1.06 | 0.28 | 0.6 | 1.86 | 0.49 | 1.56 | 18.21 | 0.64 | 0.77 | 2 | 1.02 | Shared |
| S1-wms-combined_169788 | 222.7 | 1.37 | 0.24 | 0.32 | 1.24 | 3.63 | 1.16 | 1.51 | 1.14 | 0.52 | 60.14 | 1.75 | Shared |

| | | | | | | | | | | | | | |
|---|---|---|---|---|---|---|---|---|---|---|---|---|---|
| S1-wms-combined_220022 | 35.77 | 1.45 | 0.2 | 0.59 | 1.12 | 0.3 | 1.44 | 3.13 | 0.79 | 0.41 | 1.08 | 1.51 | ME |
| S1-wms-combined_169879 | 25.15 | 10.68 | 0.87 | 0.48 | 0.75 | 0.33 | 1.12 | 2.15 | 1.23 | 0.34 | 6.86 | 0.82 | Shared |
| S1-wms-combined_23248 | 228.01 | 111.2 | 26.33 | 28.09 | 21.24 | 46.18 | 26.21 | 88.81 | 5.98 | 71.81 | 71.84 | 265.86 | Shared |
| S1-wms-combined_72167 | 24.04 | 1.11 | 0.22 | 0.27 | 2.74 | 1.05 | 1.15 | 2.66 | 1.04 | 1.32 | 2.11 | 1.8 | ME |
| S1-wms-combined_40115 | 56.99 | 8.11 | 1.08 | 1.96 | 4.3 | 0.74 | 4.97 | 7.49 | 2.61 | 2 | 8.67 | 5.64 | ME |
| S1-wms-combined_304377 | 135 | 44.35 | 9.89 | 15.86 | 6.81 | 12.05 | 14.84 | 42.54 | 2.06 | 27.1 | 35.59 | 94.97 | Shared |
| S1-wms-combined_353757 | 35.85 | 1.37 | 0.29 | 0.36 | 1.98 | 0.55 | 1.08 | 6.03 | 0.87 | 1.08 | 9.29 | 1.08 | ME |
| S1-wms-combined_57370 | 149.94 | 12.95 | 1.65 | 2.01 | 38.58 | 6.93 | 8.01 | 38.63 | 3.24 | 10.69 | 45.61 | 12.67 | Shared |
| S1-wms-combined_354899 | 23.29 | 3.67 | 0.19 | 0.6 | 2.57 | 0.59 | 1.66 | 1.41 | 0.65 | 0.85 | 1.34 | 1.38 | ME |
| S1-wms-combined_41369 | 16.46 | 1.36 | 0.14 | 0.32 | 1.75 | 0.47 | 1.1 | 1.63 | 0.73 | 0.84 | 1.58 | 1.05 | ME |
| S1-wms-combined_156637 | 83.91 | 6.84 | 3.71 | 4.68 | 27.27 | 0.33 | 7.88 | 28.3 | 0.94 | 0.85 | 34.24 | 1.26 | Shared |
| S1-wms-combined_107073 | 14.45 | 9.57 | 0.06 | 0.25 | 0.49 | 0.19 | 0.89 | 0.41 | 0.48 | 0.21 | 0.61 | 0.33 | ME |
| S1-wms-combined_107766 | 281.97 | 81.17 | 3.13 | 1.33 | 218.14 | 266.61 | 95.76 | 72.37 | 94.01 | 71.85 | 64.22 | 29.75 | Shared |
| S1-wms-combined_92523 | 48.16 | 1.59 | 0.2 | 0.73 | 2.13 | 8.12 | 1.5 | 1.99 | 1.21 | 22.93 | 8.11 | 1.95 | ME |
| S1-wms-combined_224578 | 28.88 | 1.54 | 0.07 | 0.42 | 1.04 | 0.21 | 0.95 | 0.61 | 0.64 | 0.73 | 1.02 | 0.59 | ME |
| S1-wms-combined_357373 | 329.29 | 1.24 | 0.34 | 0.3 | 1.59 | 0.59 | 1.46 | 0.54 | 1.38 | 0.24 | 0.59 | 2.17 | ME |
| S1-wms-combined_373308 | 183.22 | 32.77 | 8.36 | 4.21 | 107.7 | 19.81 | 15.33 | 206.55 | 7.74 | 38.47 | 225.64 | 41.1 | Shared |
| S1-wms-combined_143459 | 40.37 | 0.96 | 0.6 | 0.5 | 2.02 | 0.75 | 5.23 | 7.17 | 0.78 | 1.66 | 1.61 | 1.06 | ME |
| S1-wms-combined_308848 | 35.61 | 112.41 | 0.16 | 0.47 | 8.68 | 0.36 | 1.03 | 7.16 | 0.51 | 0.8 | 1.27 | 0.94 | Shared |
| S1-wms-combined_159544 | 32.19 | 1.23 | 0.26 | 0.53 | 2 | 0.55 | 2.52 | 9.04 | 0.63 | 1.94 | 1.45 | 0.77 | ME |
| S1-wms-combined_374219 | 14.66 | 6.33 | 3.06 | 0.49 | 5.47 | 3.8 | 2.8 | 2.76 | 1.36 | 2.73 | 2.02 | 21.49 | ME |
| S1-wms-combined_340558 | 18.09 | 0.84 | 0.25 | 0.3 | 1.47 | 0.46 | 0.83 | 0.99 | 0.75 | 0.29 | 14.41 | 0.51 | Shared |
| S1-wms-combined_227003 | 24.75 | 0.93 | 0.4 | 0.8 | 1.09 | 0.27 | 1.28 | 1.11 | 0.7 | 0.45 | 2.64 | 0.57 | ME |
| S1-wms-combined_227917 | 42.08 | 3.58 | 1.85 | 0.65 | 2.65 | 1.18 | 3.28 | 2.13 | 1.14 | 1.3 | 2.4 | 16.6 | ME |
| S1-wms-combined_360285 | 31.33 | 1.24 | 1.24 | 0.76 | 3.24 | 5.68 | 11.8 | 13.36 | 0.86 | 3.5 | 2.73 | 1.01 | Shared |
| S1-wms-combined_212892 | 45.4 | 1.01 | 0.03 | 0.59 | 0.38 | 0.13 | 0.55 | 0.27 | 0.65 | 0.2 | 0.82 | 0.33 | ME |
| S1-wms-combined_131270 | 20.64 | 1.11 | 0.24 | 0.55 | 1.84 | 0.56 | 1.02 | 1.5 | 1 | 0.71 | 1.34 | 1.23 | ME |
| S1-wms-combined_197401 | 38.77 | 135.28 | 0.43 | 0.79 | 1.52 | 0.5 | 1.19 | 1.3 | 0.83 | 0.56 | 3.23 | 1.15 | Shared |
| S1-wms-combined_279492 | 102.73 | 57.01 | 5.68 | 18.53 | 11.15 | 9.09 | 48.68 | 33.43 | 93.77 | 10.65 | 73.86 | 40.72 | Shared |
| S1-wms-combined_164485 | 63.38 | 7.77 | 4.72 | 6.19 | 12.23 | 1.93 | 4.24 | 47.32 | 1.95 | 4.66 | 22.19 | 11.99 | Shared |
| S1-wms-combined_32863 | 33.44 | 27.87 | 1.85 | 14.37 | 44.01 | 6.5 | 35.09 | 15.61 | 9.04 | 7.48 | 14.86 | 15.13 | Shared |
| S1-wms-combined_331544 | 42.49 | 4.31 | 1.99 | 1.33 | 2.41 | 0.76 | 1.03 | 1.42 | 2.1 | 0.51 | 1.33 | 10.99 | ME |
| S1-wms-combined_85225 | 33.77 | 3.9 | 3.67 | 5.72 | 4.98 | 1.59 | 7.62 | 1.82 | 2.9 | 0.94 | 1.51 | 10.69 | ME |
| S1-wms-combined_136097 | 88.64 | 17.94 | 5.3 | 2.5 | 18.15 | 4.73 | 6.1 | 30.77 | 3.8 | 13.88 | 24.13 | 19.88 | Shared |
| S1-wms-combined_307323 | 38.99 | 3.98 | 3.91 | 1.67 | 3.63 | 1.57 | 2.3 | 1.63 | 3.91 | 0.76 | 1.23 | 6.54 | ME |
| S1-wms-combined_94988 | 200.49 | 16.14 | 4.65 | 3.09 | 75.76 | 10.52 | 11.19 | 90.2 | 5.2 | 19.84 | 98.07 | 15.32 | Shared |
| S1-wms_296850 | 122.94 | 2.12 | 0.9 | 1.01 | 3.35 | 1.03 | 1.63 | 27.87 | 1.21 | 1.74 | 4.52 | 2.36 | Shared |
| S1-wms_313991 | 21.9 | 8.94 | 0.09 | 0.09 | 0.66 | 0.07 | 0.35 | 0.54 | 0.35 | 0.29 | 0.34 | 0.26 | ME |
| S1-wms_100010 | 41.79 | 12.03 | 0.13 | 0.66 | 1.99 | 1.06 | 1.89 | 2.67 | 0.63 | 1.84 | 2.81 | 1.15 | Shared |
| S1-wms_265285 | 27.87 | 2.41 | 0.48 | 0.38 | 1.62 | 0.49 | 1.5 | 5.09 | 0.76 | 4.8 | 7.25 | 1.11 | ME |
| S1-wms_185046 | 15.74 | 1.06 | 0.7 | 0.46 | 2.66 | 0.53 | 1.09 | 1.92 | 0.77 | 2.23 | 38.38 | 1.38 | Shared |
| S1-wms_102342 | 56.58 | 1.18 | 0.17 | 0.69 | 1.19 | 1.46 | 0.84 | 1.1 | 0.84 | 4.74 | 40.6 | 1.65 | Shared |
| S1-wms_4277 | 75.32 | 2.39 | 0.39 | 0.76 | 9.28 | 15.71 | 3.52 | 9.94 | 0.52 | 59.13 | 21.34 | 1.11 | Shared |
| S1-wms_153615 | 283.6 | 0.83 | 0.03 | 0.14 | 1.59 | 0.1 | 0.11 | 0.17 | 0.2 | 0.21 | 0.4 | 1.05 | ME |
| S1-wms_236105 | 79.78 | 0.83 | 0.2 | 0.6 | 1.09 | 0.45 | 0.75 | 46.85 | 0.74 | 0.63 | 1.76 | 0.53 | Shared |
| S1-wms_170115 | 45.28 | 3.45 | 1.91 | 7.75 | 2.45 | 3.54 | 4.26 | 3.8 | 1.05 | 4.42 | 6.35 | 2.87 | ME |
| S1-wms_106115 | 82.7 | 0.83 | 0.04 | 0.44 | 0.39 | 0.16 | 0.61 | 0.23 | 0.76 | 0.16 | 0.6 | 0.48 | ME |
| S1-wms_388292 | 13.07 | 0.77 | 0.2 | 0.35 | 6.19 | 0.41 | 1.06 | 0.69 | 0.48 | 0.47 | 1.14 | 0.51 | ME |
| S1-wms_223620 | 217.43 | 0.85 | 0.4 | 0.53 | 1.2 | 0.43 | 57.97 | 229.9 | 0.69 | 0.58 | 7.47 | 0.76 | Shared |
| S1-wms_208875 | 29.11 | 0.84 | 0.75 | 0.42 | 0.59 | 0.22 | 1.14 | 0.38 | 0.45 | 0.34 | 0.68 | 0.33 | ME |
| S1-wms_225995 | 40.92 | 1.48 | 0.08 | 0.46 | 1.46 | 0.44 | 1.87 | 1.24 | 1.01 | 3.7 | 1.86 | 2.19 | ME |
| S1-wms_29288 | 28.44 | 2.32 | 0.46 | 0.89 | 1.63 | 1.41 | 1.47 | 1.64 | 0.6 | 0.77 | 1.91 | 2.34 | ME |
| S1-wms_244836 | 22.92 | 75.83 | 0.18 | 3.71 | 8.21 | 0.11 | 2.53 | 1.02 | 126.99 | 4.19 | 8.68 | 1.75 | Shared |
| S1-wms_130214 | 27.7 | 1.03 | 0.12 | 0.66 | 1.4 | 0.42 | 1.4 | 1.75 | 0.72 | 0.62 | 1.22 | 0.76 | ME |
| S1-wms_212940 | 75.56 | 1.17 | 1.1 | 6.19 | 0.78 | 1.45 | 1.64 | 1.28 | 0.64 | 2.22 | 51.19 | 0.51 | Shared |
| S1-wms_394571 | 102.54 | 40.3 | 0.11 | 0.35 | 1.31 | 0.32 | 0.74 | 1.8 | 0.48 | 0.5 | 1.04 | 0.63 | Shared |
| S1-wms_32878 | 35.82 | 1.18 | 0.21 | 0.24 | 1.2 | 0.2 | 0.74 | 1.12 | 0.59 | 0.5 | 0.99 | 1.4 | ME |
| S3-wms-combined_135428 | 1.42 | 0.94 | 14.82 | 0.24 | 2.36 | 0.76 | 7.5 | 3.52 | 0.58 | 0.72 | 1.46 | 1.07 | ME |
| S3-wms-combined_73262 | 83.67 | 35.19 | 20.67 | 23.18 | 5.72 | 256.28 | 86.45 | 178.77 | 1.36 | 74.99 | 39.56 | 1.37 | Shared |
| S3-wms-combined_45305 | 181.56 | 138.03 | 46.62 | 8.15 | 12.96 | 3.04 | 95.53 | 62.1 | 1.27 | 8.57 | 11.99 | 56.34 | Shared |
| S3-wms_23958 | 4.91 | 3.22 | 18.02 | 0.56 | 1.28 | 0.81 | 1.28 | 15.41 | 0.63 | 0.63 | 47.59 | 0.83 | Shared |
| S3-wms_94494 | 1.97 | 1.13 | 17.03 | 0.72 | 2.95 | 1.25 | 5.87 | 2.09 | 1.23 | 1.19 | 2.36 | 1.61 | ME |
| S3-wms_22744 | 24.04 | 14.73 | 15.25 | 2.22 | 3.03 | 1.71 | 4.91 | 3.3 | 0.8 | 3.79 | 45.78 | 2.76 | Shared |
| S5-wms-combined_26061 | 10.9 | 7.89 | 2.31 | 66.99 | 35.3 | 58.87 | 53.01 | 15.46 | 1.36 | 15.35 | 54.14 | 19.63 | Shared |
| S5-wms-combined_205046 | 64.47 | 73.67 | 8.09 | 8.93 | 87.5 | 58.6 | 36.88 | 48.85 | 45.58 | 56.26 | 49.54 | 184.36 | Shared |
| S5-wms-combined_18356 | 7.6 | 1.22 | 0.11 | 0.44 | 24.04 | 0.52 | 1.44 | 0.95 | 23.81 | 2.14 | 1.32 | 2.86 | ME |

| | | | | | | | | | | | | | |
|---|---|---|---|---|---|---|---|---|---|---|---|---|---|
| S5-wms-combined_161191 | 22.41 | 16.75 | 3.12 | 2.59 | 41.09 | 16.65 | 14.8 | 18.5 | 6.54 | 22.8 | 14.44 | 65.17 | Shared |
| S5-wms-combined_134381 | 137.54 | 28.79 | 6.89 | 9.04 | 46.04 | 6.17 | 9.28 | 20.52 | 2.42 | 31.16 | 7.17 | 10.37 | Shared |
| S5-wms-combined_90315 | 12.53 | 14.35 | 0.68 | 2.35 | 93.35 | 7.69 | 13.81 | 16.6 | 5.06 | 16.73 | 46.71 | 9.67 | Shared |
| S5-wms-combined_134743 | 1.12 | 2.1 | 0.12 | 0.33 | 42.98 | 0.27 | 1.34 | 1.1 | 0.88 | 0.75 | 1.53 | 1.03 | ME |
| S5-wms-combined_197186 | 1.04 | 1.46 | 0.08 | 0.21 | 28.32 | 0.22 | 1.56 | 0.71 | 0.93 | 0.3 | 0.91 | 0.61 | ME |
| S5-wms-combined_117095 | 11.27 | 5.94 | 0.64 | 5.72 | 38.33 | 8.9 | 33.62 | 27.19 | 9.4 | 8.49 | 5.81 | 4.42 | Shared |
| S5-wms-combined_72410 | 15.99 | 12.78 | 1.83 | 1.96 | 139.03 | 15.68 | 8.58 | 13.85 | 13.34 | 17.67 | 11.06 | 16.75 | Shared |
| S5-wms-combined_206050 | 3.24 | 3.82 | 0.33 | 1.12 | 24.13 | 1.37 | 3.14 | 2.53 | 8.02 | 2.32 | 4.92 | 8.22 | ME |
| S5-wms-combined_73301 | 1.05 | 2.64 | 0.19 | 2.8 | 15.16 | 0.27 | 2.54 | 0.4 | 0.85 | 0.44 | 1.29 | 3.85 | ME |
| S5-wms-combined_108794 | 9.42 | 27.76 | 1.25 | 4.7 | 138.8 | 10.71 | 19.28 | 6.48 | 17.08 | 3.11 | 4.35 | 1.33 | Shared |
| S5-wms-combined_145067 | 4.87 | 3.79 | 0.71 | 1.87 | 23.09 | 26 | 3.43 | 6.91 | 3.04 | 10.48 | 2.33 | 68.83 | Shared |
| S5-wms-combined_3201 | 1.09 | 0.97 | 0.07 | 0.34 | 26.7 | 0.15 | 0.66 | 0.83 | 0.74 | 0.37 | 0.97 | 0.56 | ME |
| S5-wms-combined_207291 | 36.25 | 95.53 | 0.95 | 2.99 | 57.11 | 2.55 | 36.84 | 33.97 | 18.28 | 7.09 | 24.57 | 11.85 | Shared |
| S5-wms-combined_29272 | 14.17 | 3.14 | 0.77 | 1.01 | 30.96 | 0.91 | 6.17 | 8.53 | 0.76 | 4.78 | 8.37 | 1.58 | ME |
| S5-wms-combined_29447 | 14.01 | 5.77 | 0.86 | 1.82 | 114.37 | 16.78 | 11.7 | 40.76 | 6.67 | 29.83 | 34.75 | 5.98 | Shared |
| S5-wms-combined_199135 | 2 | 1.36 | 0.07 | 0.51 | 19.43 | 0.63 | 1.17 | 1.28 | 0.59 | 0.41 | 1.02 | 1.9 | ME |
| S5-wms-combined_83370 | 30.5 | 59.51 | 1.81 | 5.09 | 77.52 | 6.85 | 45.72 | 18.19 | 37.23 | 13.93 | 19.62 | 32.48 | Shared |
| S5-wms-combined_83412 | 0.85 | 0.85 | 0.06 | 0.58 | 17.46 | 0.25 | 0.87 | 0.51 | 0.98 | 0.76 | 0.89 | 0.62 | ME |
| S5-wms-combined_66017 | 0.74 | 1.03 | 0.09 | 0.26 | 28.04 | 0.24 | 0.72 | 0.57 | 0.87 | 0.32 | 0.72 | 0.45 | ME |
| S5-wms-combined_66662 | 18.76 | 11.09 | 2.07 | 2.64 | 159.11 | 13.52 | 8.06 | 13.35 | 12.5 | 14.68 | 11.57 | 20.55 | Shared |
| S5-wms-combined_147993 | 1.19 | 1.18 | 0.28 | 0.4 | 50.13 | 0.19 | 0.85 | 0.5 | 1.35 | 0.21 | 0.7 | 0.48 | ME |
| S5-wms-combined_156329 | 0.94 | 1.1 | 0.09 | 0.21 | 28.92 | 0.26 | 1.46 | 0.62 | 1.25 | 0.21 | 0.83 | 0.46 | ME |
| S5-wms-combined_7292 | 0.69 | 1.2 | 0.07 | 0.36 | 215.01 | 10.36 | 1.23 | 0.4 | 0.29 | 0.2 | 4.49 | 0.38 | Shared |
| S5-wms-combined_59064 | 31.44 | 73.56 | 7.39 | 13.59 | 90.05 | 8.33 | 67.21 | 26.24 | 46.49 | 18.04 | 32.38 | 215.03 | Shared |
| S5-wms-combined_175145 | 0.86 | 1.12 | 0.08 | 0.57 | 19.46 | 0.26 | 0.97 | 0.61 | 0.6 | 0.77 | 0.93 | 6.06 | ME |
| S5-wms-combined_94482 | 1.94 | 2.77 | 0.24 | 1.7 | 60.07 | 2.03 | 16.22 | 1.33 | 1.62 | 1.15 | 3.94 | 1.4 | ME |
| S5-wms-combined_67650 | 195.25 | 13.77 | 10.17 | 11.57 | 365.17 | 135.73 | 198.24 | 136.26 | 19.76 | 171.23 | 81.6 | 15.68 | Shared |
| S5-wms-combined_94786 | 7.4 | 3.92 | 1.93 | 1.37 | 65.83 | 5.72 | 8.02 | 7.92 | 4.5 | 8.09 | 4.95 | 16.94 | ME |
| S5-wms-combined_210959 | 84.51 | 35.3 | 5.07 | 6.9 | 1045.11 | 226.03 | 74.14 | 63.87 | 54.03 | 283.75 | 420.78 | 48.42 | Shared |
| S5-wms-combined_78434 | 2.29 | 2.58 | 0.14 | 0.51 | 28.95 | 0.58 | 1.78 | 1.47 | 0.86 | 1.6 | 2.58 | 4.95 | ME |
| S5-wms-combined_106597 | 0.52 | 0.69 | 0.03 | 0.71 | 42.93 | 0.17 | 0.51 | 0.39 | 0.58 | 0.21 | 0.75 | 0.49 | ME |
| S5-wms-combined_204782 | 1.7 | 1.17 | 0.07 | 0.6 | 29.93 | 1.03 | 2.59 | 0.65 | 0.69 | 1 | 1.38 | 0.64 | ME |
| S5-wms-combined_98202 | 58.72 | 4.18 | 2.68 | 4.94 | 144.73 | 77.21 | 79.68 | 40.05 | 5.05 | 44.21 | 35.64 | 4.41 | Shared |
| S5-wms-combined_26971 | 23.03 | 3.69 | 1.21 | 3.34 | 265.61 | 7.49 | 6.53 | 26.06 | 2.19 | 9.31 | 21.77 | 5.12 | Shared |
| S5-wms-combined_178432 | 13.67 | 6.34 | 1.1 | 1.65 | 947.72 | 151.73 | 8.55 | 18.05 | 2.87 | 101.99 | 210.09 | 3.93 | Shared |
| S5-wms-combined_179241 | 132.71 | 25.84 | 10.84 | 3.83 | 593.91 | 7.1 | 46.01 | 22.14 | 1.75 | 84.76 | 48.65 | 11.81 | Shared |
| S5-wms-combined_117213 | 4.44 | 1.31 | 7.75 | 12.06 | 31.7 | 28.6 | 6.37 | 1.97 | 96.12 | 1.08 | 1.85 | 2.91 | Shared |
| S5-wms-combined_163165 | 1.42 | 8.15 | 0.1 | 0.34 | 27.36 | 0.34 | 1.11 | 3.52 | 0.54 | 13.88 | 2.14 | 0.89 | ME |
| S5-wms-combined_39793 | 0.5 | 0.44 | 0 | 0.13 | 50.98 | 17.1 | 0.72 | 0.77 | 0.53 | 0.6 | 2.5 | 0.57 | Shared |
| S5-wms-combined_183113 | 12.02 | 5.99 | 0.48 | 0.74 | 64.48 | 7.08 | 19.6 | 7.61 | 4.35 | 5.81 | 9.77 | 3.49 | ME |
| S5-wms-combined_52741 | 5.08 | 13.32 | 0.58 | 0.4 | 214.37 | 1.05 | 4.32 | 76.58 | 0.48 | 0.59 | 1.53 | 0.99 | Shared |
| S5-wms_62679 | 1.82 | 0.98 | 0.21 | 0.35 | 41.05 | 0.73 | 1.15 | 1.93 | 0.76 | 1.62 | 1.4 | 1.83 | ME |
| S5-wms_10783 | 40.2 | 41.83 | 2.41 | 11.38 | 68.1 | 5.75 | 34.58 | 27.66 | 22.72 | 19.12 | 34 | 45.71 | Shared |
| S5-wms_164032 | 0.42 | 0.95 | 0.31 | 0.18 | 16.49 | 0.59 | 3.11 | 0.25 | 0.6 | 0.18 | 0.43 | 0.6 | ME |
| S5-wms_49310 | 0.65 | 2.83 | 0.3 | 4.79 | 192.62 | 11.66 | 0.61 | 0.35 | 0.57 | 0.19 | 1.63 | 0.4 | Shared |
| S5-wms_67169 | 2.6 | 1.57 | 0.57 | 1.48 | 75.15 | 1.92 | 2.52 | 2.12 | 1.34 | 1.64 | 1.97 | 6.02 | ME |
| S5-wms_85650 | 7.49 | 23.32 | 1 | 9.31 | 96.43 | 14.35 | 18.88 | 0.81 | 1.21 | 2.55 | 7.93 | 5.82 | Shared |
| S5-wms_210460 | 0.57 | 0.78 | 0.2 | 0.18 | 17.47 | 0.08 | 0.7 | 0.47 | 0.52 | 0.14 | 0.67 | 0.2 | ME |
| S5-wms_94965 | 2.54 | 2 | 0.28 | 0.79 | 22.02 | 1.79 | 1.28 | 2.68 | 1.42 | 1.99 | 2.85 | 1.65 | ME |
| S5-wms_52566 | 127.63 | 98.63 | 11.97 | 12.57 | 134.68 | 45.18 | 52.6 | 103.23 | 49.08 | 100.88 | 110.63 | 196.39 | Shared |
| S5-wms_123499 | 0.69 | 0.36 | 0.05 | 0.11 | 20.53 | 0.05 | 1.8 | 0.18 | 0.77 | 0.2 | 0.67 | 0.32 | ME |
| S5-wms_212596 | 0.95 | 0.77 | 0.09 | 0.64 | 11.31 | 0.1 | 0.48 | 0.32 | 0.55 | 0.11 | 0.23 | 0.11 | ME |
| S7-wms-combined_217226 | 1.07 | 1.46 | 0.07 | 0.38 | 1.53 | 0.32 | 43.87 | 0.83 | 0.63 | 0.5 | 1.28 | 0.66 | ME |
| S7-wms-combined_139683 | 6.55 | 1.75 | 2.02 | 0.69 | 1.76 | 0.62 | 24.09 | 6.75 | 0.52 | 7.4 | 16.69 | 0.53 | Shared |
| S7-wms-combined_71592 | 0.82 | 11.21 | 3.37 | 0.42 | 0.71 | 0.24 | 55.11 | 1.63 | 1.4 | 0.26 | 7.32 | 0.48 | Shared |
| S7-wms-combined_235165 | 2.49 | 3.9 | 1.49 | 1.51 | 2.95 | 0.7 | 86.75 | 2.04 | 1.43 | 0.76 | 3.73 | 0.74 | ME |
| S7-wms-combined_225260 | 1.06 | 1.58 | 0.07 | 0.71 | 0.96 | 0.15 | 19.09 | 0.6 | 0.89 | 0.25 | 1 | 0.69 | ME |
| S7-wms-combined_31068 | 5.03 | 15.06 | 1.4 | 3 | 5.05 | 9.12 | 23.87 | 5.8 | 2.98 | 10.05 | 11.69 | 67.87 | Shared |
| S7-wms-combined_123085 | 16.03 | 25.89 | 1.04 | 11.47 | 4.56 | 4.4 | 50.93 | 5.48 | 87.84 | 1.51 | 5.01 | 10.38 | Shared |
| S7-wms-combined_20921 | 3.06 | 4.31 | 0.48 | 3.76 | 1.24 | 5.98 | 46.44 | 1.61 | 25.98 | 0.52 | 1.65 | 5.68 | ME |
| S7-wms-combined_62119 | 356.97 | 552.92 | 22.18 | 15.03 | 337.24 | 59.16 | 1081.3 | 72.89 | 17.56 | 80.49 | 141.8 | 307.74 | Shared |
| S7-wms-combined_185016 | 24.82 | 59.93 | 1.09 | 6.55 | 42.76 | 5.17 | 50.86 | 16.11 | 1.17 | 7.38 | 8.99 | 18.58 | Shared |
| S7-wms-combined_174935 | 7.97 | 3.01 | 19.53 | 11.01 | 30.36 | 17.21 | 35.73 | 5.87 | 20.09 | 2.42 | 4.41 | 5.17 | Shared |
| S7-wms-combined_134452 | 116.78 | 6.35 | 0.84 | 1.41 | 6.35 | 0.74 | 58.32 | 35.23 | 1.44 | 3.44 | 65.99 | 1.72 | Shared |
| S7-wms-combined_134458 | 21.94 | 13.79 | 2.7 | 3.07 | 13.4 | 20.66 | 39.06 | 26.76 | 2.26 | 5.08 | 14.87 | 12.49 | Shared |
| S7-wms-combined_21693 | 6.93 | 19.59 | 0.43 | 0.75 | 0.85 | 3.8 | 26.75 | 28.82 | 0.81 | 1.2 | 23.51 | 0.92 | Shared |

| | | | | | | | | | | | | | |
|---|---|---|---|---|---|---|---|---|---|---|---|---|---|
| S7-wms-combined_2122 | 3.72 | 5.86 | 0.38 | 3.71 | 1.86 | 2.09 | 58.72 | 1.67 | 22.12 | 0.85 | 2.18 | 3.42 | ME |
| S7-wms-combined_135154 | 4.04 | 7.44 | 0.67 | 7.92 | 47.7 | 8.73 | 51.96 | 4.34 | 1.49 | 2.7 | 6.56 | 13.56 | ME |
| S7-wms-combined_156351 | 0.79 | 1.05 | 0.07 | 0.73 | 0.57 | 0.15 | 15.21 | 1.5 | 0.89 | 0.27 | 0.78 | 0.43 | ME |
| S7-wms-combined_104872 | 56.18 | 160.82 | 0.92 | 0.66 | 4.19 | 0.12 | 27.14 | 25.59 | 0.75 | 4.06 | 12.62 | 0.68 | Shared |
| S7-wms-combined_238231 | 17.11 | 17.39 | 1.32 | 2.19 | 26.72 | 3.39 | 37.33 | 8.86 | 7.69 | 17.48 | 11.35 | 11.38 | Shared |
| S7-wms-combined_43950 | 60.15 | 79.2 | 10.07 | 23.17 | 25.64 | 191.94 | 217.49 | 159.54 | 22.91 | 79.81 | 50.41 | 31.13 | Shared |
| S7-wms-combined_84688 | 23.27 | 21.49 | 2.56 | 5.23 | 37.88 | 22.63 | 40.28 | 10.14 | 19.28 | 11.85 | 8.41 | 53.04 | Shared |
| S7-wms-combined_3669 | 0.22 | 0.7 | 0.03 | 0.19 | 2.93 | 0.05 | 8.19 | 0.4 | 0.54 | 0.37 | 0.74 | 0.35 | ME |
| S7-wms-combined_3697 | 164.6 | 63.77 | 6.39 | 3.92 | 415.49 | 206.31 | 188.41 | 136.75 | 140.07 | 181.77 | 196.97 | 83.41 | Shared |
| S7-wms-combined_157260 | 15.62 | 7.37 | 0.72 | 12.38 | 3.75 | 1.74 | 36.68 | 2.19 | 14.61 | 1.19 | 3.17 | 6.83 | Shared |
| S7-wms-combined_74829 | 23.9 | 18.13 | 0.11 | 0.91 | 1.99 | 4.59 | 54.79 | 0.69 | 12.35 | 0.49 | 1.05 | 10.43 | Shared |
| S7-wms-combined_14267 | 271.07 | 84.02 | 4.53 | 5.15 | 405.75 | 303.54 | 323.17 | 105.8 | 176.48 | 117.28 | 111.5 | 32.68 | Shared |
| S7-wms-combined_157434 | 19.39 | 47.4 | 1.26 | 6.26 | 3.76 | 5.19 | 33.61 | 11.99 | 22.28 | 4.48 | 8.92 | 39.79 | Shared |
| S7-wms-combined_167867 | 1.63 | 1.03 | 8.88 | 0.75 | 1.72 | 0.45 | 27.72 | 1.05 | 0.72 | 0.6 | 1.31 | 1.09 | ME |
| S7-wms-combined_106253 | 6.08 | 8.85 | 0.78 | 13.96 | 5.03 | 1.51 | 40.44 | 2.31 | 15.21 | 1.7 | 3.73 | 8.43 | Shared |
| S7-wms-combined_239784 | 45.21 | 143.6 | 0.73 | 1.27 | 6.61 | 2.67 | 383.44 | 5.92 | 0.85 | 4.37 | 12.18 | 15.23 | Shared |
| S7-wms-combined_14529 | 119.71 | 2.73 | 3.08 | 0.91 | 6.92 | 1.69 | 67.37 | 165.33 | 1.3 | 3.81 | 113.51 | 3.44 | Shared |
| S7-wms-combined_189250 | 2.02 | 1.03 | 0.37 | 0.48 | 3.13 | 0.99 | 26.1 | 16.36 | 0.86 | 1.33 | 1.72 | 1.16 | Shared |
| S7-wms-combined_137858 | 85.09 | 26.45 | 11.87 | 12.47 | 6.3 | 7.96 | 53.25 | 34.34 | 3.26 | 37.65 | 34.51 | 59.74 | Shared |
| S7-wms-combined_169375 | 1.81 | 6.09 | 0.34 | 5.93 | 24.8 | 0.97 | 120.02 | 1.12 | 153.47 | 1.22 | 1.38 | 2.2 | ME |
| S7-wms-combined_5625 | 285.48 | 548.82 | 20.41 | 21.61 | 295.16 | 44.71 | 1137.17 | 54.16 | 22.09 | 71.61 | 118.43 | 270 | Shared |
| S7-wms-combined_190281 | 2.97 | 1.92 | 4.2 | 12.74 | 7.54 | 1.6 | 34.43 | 18.37 | 1.74 | 3.75 | 5.17 | 18.72 | Shared |
| S7-wms-combined_26217 | 163.1 | 10.19 | 9.07 | 16.58 | 229.78 | 98.62 | 294.28 | 122.42 | 15.69 | 137.3 | 68.96 | 23.78 | Shared |
| S7-wms-combined_69024 | 0.75 | 0.92 | 0.08 | 0.8 | 2.68 | 0.7 | 13.79 | 0.56 | 0.84 | 0.24 | 1.13 | 2.02 | ME |
| S7-wms-combined_7205 | 12.43 | 29.98 | 1.42 | 1.07 | 1.97 | 7.59 | 70.59 | 3.94 | 3.94 | 6.52 | 4.57 | 2.26 | Shared |
| S7-wms-combined_121062 | 0.58 | 0.92 | 0.26 | 0.57 | 0.54 | 0.15 | 15.29 | 0.31 | 0.54 | 0.24 | 0.35 | 0.5 | ME |
| S7-wms-combined_90393 | 0.52 | 1.45 | 0.16 | 0.35 | 0.96 | 0.34 | 152.96 | 0.39 | 0.51 | 0.32 | 0.97 | 2.43 | ME |
| S7-wms-combined_111004 | 25.94 | 12.6 | 4.95 | 5.32 | 2.82 | 6.36 | 33.81 | 14.75 | 1.83 | 6.83 | 15.04 | 29.07 | Shared |
| S7-wms-combined_90728 | 0.7 | 1.22 | 0.03 | 0.4 | 0.79 | 0.25 | 84.78 | 0.32 | 0.86 | 0.22 | 0.71 | 3.64 | ME |
| S7-wms-combined_40779 | 14.03 | 7.76 | 0.49 | 8.96 | 2.98 | 7.32 | 50.03 | 2.57 | 1.4 | 4.09 | 3.25 | 274.22 | ME |
| S7-wms-combined_28138 | 1.46 | 3.31 | 0.12 | 0.8 | 16.34 | 1.65 | 33.88 | 0.78 | 1.84 | 0.58 | 1.45 | 0.48 | ME |
| S7-wms-combined_224523 | 3.5 | 62.46 | 0.18 | 0.68 | 0.55 | 0.32 | 44.28 | 7.65 | 0.78 | 0.27 | 7.14 | 0.94 | Shared |
| S7-wms-combined_143218 | 6.78 | 3.17 | 4.32 | 4.12 | 11.72 | 4.39 | 25.37 | 3.7 | 12.6 | 0.87 | 4.15 | 3.47 | ME |
| S7-wms-combined_163980 | 7.13 | 3.42 | 3.23 | 2.18 | 25.6 | 5.61 | 53.84 | 145.74 | 1.47 | 2.56 | 7.2 | 4.79 | ME |
| S7-wms-combined_72070 | 50.23 | 122.56 | 2.6 | 2.35 | 73.8 | 10.15 | 1008.58 | 31.38 | 6.56 | 10.39 | 50.7 | 23.81 | Shared |
| S7-wms-combined_136642 | 6.32 | 1.49 | 3.9 | 6.17 | 6.12 | 2.8 | 37.06 | 2.32 | 7.87 | 1.03 | 1.83 | 3.74 | ME |
| S7-wms-combined_147158 | 2.83 | 1.83 | 6.4 | 8.48 | 6.52 | 3.31 | 30.74 | 1.9 | 10.42 | 1.06 | 1.74 | 1.47 | ME |
| S7-wms-combined_218379 | 12.28 | 7.94 | 4.73 | 2.59 | 7.81 | 1.87 | 105.24 | 15.89 | 5.06 | 4.66 | 9.3 | 7.21 | Shared |
| S7-wms-combined_107679_0 | 147.71 | 6.59 | 7.73 | 9.72 | 247.95 | 120.46 | 218.08 | 108.29 | 13.43 | 144.76 | 58.6 | 11.97 | Shared |
| S7-wms-combined_107679_1 | 61.09 | 9.86 | 1.46 | 2.29 | 191.23 | 35.76 | 267.27 | 73.09 | 6.16 | 56.58 | 28.11 | 11.66 | Shared |
| S7-wms_113137 | 6.05 | 18.23 | 3.73 | 2.35 | 4.09 | 24.15 | 32.98 | 109.66 | 2.17 | 21.77 | 34.47 | 1.13 | Shared |
| S7-wms_123954 | 0.71 | 0.5 | 0.05 | 0.51 | 0.71 | 0.14 | 22.14 | 0.94 | 0.55 | 0.25 | 9.77 | 4.83 | ME |
| S7-wms_53338 | 0.37 | 0.46 | 0.04 | 0.11 | 0.43 | 0.08 | 37.52 | 0.46 | 0.27 | 0.19 | 0.35 | 0.32 | ME |
| S7-wms_157264 | 0.4 | 0.33 | 0.02 | 0.18 | 0.88 | 0.37 | 337.77 | 1.53 | 0.61 | 1.26 | 0.61 | 0.32 | ME |
| S7-wms_239833 | 0.73 | 0.77 | 0.05 | 0.54 | 0.65 | 0.18 | 62.64 | 0.79 | 0.66 | 0.32 | 0.61 | 0.39 | ME |
| S7-wms_127801 | 7.92 | 15.3 | 3.84 | 1.05 | 3.04 | 0.77 | 48.5 | 3.31 | 2.49 | 1.48 | 8.22 | 2.29 | Shared |
| S7-wms_76437 | 0.69 | 0.72 | 0.04 | 0.46 | 0.71 | 0.15 | 34.12 | 0.47 | 0.42 | 0.22 | 0.68 | 0.39 | ME |
| S7-wms_118607 | 2.22 | 1.81 | 0.21 | 0.46 | 1.55 | 11.91 | 53.61 | 1.21 | 0.99 | 27.04 | 12.37 | 0.92 | Shared |
| S7-wms_159432 | 5.7 | 1.03 | 3.48 | 2.38 | 0.8 | 0.28 | 70.38 | 0.67 | 1.36 | 0.26 | 1.91 | 12.24 | ME |
| S7-wms_200936 | 9.63 | 0.77 | 0.17 | 0.58 | 0.74 | 6.77 | 14.21 | 19.13 | 0.26 | 20.98 | 4.42 | 48.81 | Shared |
| S7-wms_151112 | 0.78 | 0.87 | 0.07 | 0.35 | 0.45 | 0.22 | 46.88 | 0.41 | 0.89 | 0.18 | 0.52 | 0.37 | ME |
| S7-wms_80255 | 0.8 | 0.71 | 0.14 | 0.22 | 0.87 | 0.17 | 15.22 | 3.78 | 0.5 | 0.5 | 5.05 | 0.64 | ME |
| S7-wms_244425 | 0.43 | 1.02 | 0.06 | 1.61 | 0.61 | 0.38 | 126.34 | 24.01 | 26.87 | 21.98 | 0.66 | 0.41 | Shared |
| S7-wms_244866 | 2.63 | 1.29 | 0.04 | 0.36 | 0.59 | 0.2 | 36.86 | 0.36 | 0.54 | 0.13 | 0.53 | 0.32 | ME |
| S17-wms-combined_157468 | 1.8 | 2.32 | 0.08 | 0.67 | 9.85 | 9.11 | 2.81 | 4.23 | 1.44 | 1.33 | 1.01 | 28.87 | ME |
| S17-wms-combined_233761 | 73.12 | 108.12 | 5.91 | 1.31 | 71.91 | 9.34 | 140.11 | 5.17 | 1.01 | 12.17 | 20.48 | 127.59 | Shared |
| S17-wms-combined_56006 | 27.09 | 15.78 | 4.15 | 11.61 | 6.63 | 20.75 | 7.87 | 30.21 | 2.23 | 5.58 | 18.91 | 121.26 | Shared |
| S17-wms-combined_56061 | 41.76 | 53.97 | 6.76 | 13.09 | 13.05 | 17.62 | 11.04 | 32.53 | 5.35 | 33.54 | 26.86 | 247.28 | Shared |
| S17-wms-combined_223748 | 0.7 | 3.14 | 0.11 | 0.28 | 1.04 | 1.13 | 0.99 | 0.37 | 0.27 | 0.46 | 0.32 | 20.83 | ME |
| S17-wms-combined_79709 | 1.88 | 3.69 | 0.19 | 0.29 | 2.93 | 1.19 | 1.96 | 1.6 | 0.8 | 2.8 | 2.01 | 35.35 | ME |
| S17-wms-combined_146050 | 24.07 | 50.1 | 4.15 | 6.3 | 23.4 | 8.91 | 75.22 | 4.81 | 7.15 | 55.77 | 49.81 | 139.66 | Shared |
| S17-wms-combined_179363 | 2.02 | 1.43 | 1.16 | 0.15 | 2.94 | 0.27 | 1.16 | 1.65 | 3.98 | 0.6 | 1.8 | 19.18 | ME |
| S17-wms-combined_90378 | 13.79 | 4.07 | 4.43 | 3.2 | 3.29 | 5.03 | 4.45 | 6.51 | 1.13 | 1.51 | 4.77 | 75.72 | ME |
| S17-wms-combined_179771 | 1.43 | 3.48 | 0.27 | 0.77 | 26.92 | 3.13 | 71.86 | 4.67 | 0.78 | 1.14 | 16.93 | 99.72 | Shared |
| S17-wms-combined_2234 | 19.5 | 22.02 | 2.3 | 1.87 | 44.16 | 15.25 | 9.17 | 18.63 | 6.16 | 31.3 | 17.41 | 197.7 | Shared |
| S17-wms-combined_146850 | 3.1 | 1.26 | 0.13 | 0.21 | 2.75 | 0.42 | 1.46 | 1.48 | 0.6 | 1.32 | 2.01 | 27.47 | ME |

| | | | | | | | | | | | | | |
|---|---|---|---|---|---|---|---|---|---|---|---|---|---|
| S17-wms-combined_258087 | 68.39 | 20.36 | 4.06 | 5.94 | 6.14 | 5.35 | 2.63 | 14.35 | 0.71 | 13.34 | 18.31 | 37.73 | Shared |
| S17-wms-combined_225356 | 2.32 | 0.94 | 0.37 | 0.49 | 4.69 | 1.44 | 2.19 | 2.85 | 0.81 | 11.95 | 2.29 | 17.64 | ME |
| S17-wms-combined_136622 | 1.21 | 1.02 | 1.02 | 0.16 | 1.05 | 0.16 | 0.59 | 5.53 | 0.64 | 0.53 | 0.68 | 20.83 | ME |
| S17-wms-combined_203701 | 108.75 | 27.36 | 40.02 | 30.35 | 1077.68 | 114.24 | 721.8 | 354.77 | 19.55 | 256.43 | 825.9 | 86.95 | Shared |
| S17-wms-combined_158354 | 2.83 | 2.75 | 0.29 | 1.13 | 0.81 | 0.35 | 22.41 | 0.45 | 0.68 | 8.29 | 3.17 | 58.7 | ME |
| S17-wms-combined_247494 | 16.87 | 29.66 | 1.89 | 1.5 | 14.2 | 8.15 | 5.52 | 10.3 | 3.19 | 19.58 | 10.58 | 109.69 | Shared |
| S17-wms-combined_70983 | 23.39 | 38.6 | 0.74 | 1.06 | 0.42 | 0.42 | 16.78 | 0.52 | 1.12 | 1.31 | 4.38 | 28.61 | Shared |
| S17-wms-combined_126600 | 16.02 | 23.12 | 0.66 | 1.93 | 21.23 | 6.1 | 13.44 | 10.14 | 4.71 | 27.59 | 31.35 | 70.87 | Shared |
| S17-wms-combined_126774 | 28.51 | 21.33 | 4.04 | 4.53 | 62.62 | 12.92 | 15.43 | 15.8 | 6.95 | 5.68 | 14.53 | 89.36 | Shared |
| S17-wms-combined_171305 | 0.91 | 1.78 | 0.04 | 0.7 | 2.69 | 0.31 | 1.97 | 3.08 | 0.58 | 1.87 | 2.32 | 45.34 | ME |
| S17-wms-combined_248321 | 107.06 | 9.22 | 3.22 | 10.72 | 6.82 | 7.86 | 5.36 | 34.22 | 3.49 | 9.2 | 6.63 | 135.68 | Shared |
| S17-wms-combined_193958 | 89.18 | 75.17 | 10.71 | 12.15 | 7.44 | 16.32 | 11.79 | 55.92 | 1.84 | 40.45 | 33.75 | 347.73 | Shared |
| S17-wms-combined_47860 | 1.11 | 6.85 | 0.15 | 0.64 | 1.73 | 1.25 | 1.3 | 2.17 | 0.58 | 1.45 | 1.91 | 38.86 | ME |
| S17-wms-combined_259811 | 7.17 | 13.57 | 1.23 | 4.85 | 7.78 | 10.19 | 12.41 | 6.07 | 3.14 | 15.14 | 12.58 | 83.27 | Shared |
| S17-wms-combined_127688 | 27.5 | 9.16 | 4.41 | 4.37 | 3.82 | 2.27 | 38.76 | 10.4 | 10.22 | 3.15 | 16.53 | 33.03 | Shared |
| S17-wms-combined_260343 | 5.7 | 2.45 | 1.54 | 0.65 | 2.82 | 0.84 | 14.41 | 1.85 | 0.78 | 3.54 | 2.81 | 86.87 | ME |
| S17-wms-combined_72837 | 10.43 | 4.12 | 0.73 | 3.28 | 2.28 | 0.83 | 6.71 | 5.04 | 0.73 | 3.72 | 3.9 | 26.85 | ME |
| S17-wms-combined_16309 | 0.43 | 0.89 | 0.24 | 0.63 | 0.48 | 0.23 | 0.95 | 0.28 | 0.64 | 0.25 | 1.09 | 41.74 | ME |
| S17-wms-combined_106262 | 4.67 | 4.41 | 0.17 | 0.71 | 2.19 | 1.04 | 4.57 | 1.62 | 0.62 | 3.23 | 2.37 | 69.61 | ME |
| S17-wms-combined_185239 | 40.8 | 48.8 | 4.47 | 4.63 | 46.77 | 22.38 | 18.96 | 32.13 | 17.43 | 93 | 31.92 | 143.66 | Shared |
| S17-wms-combined_39687 | 2.77 | 1.63 | 0.22 | 0.51 | 2 | 8.1 | 2.74 | 6.68 | 0.65 | 3.67 | 1.7 | 18.53 | ME |
| S17-wms-combined_140517 | 0.66 | 0.99 | 0.05 | 0.46 | 0.64 | 0.15 | 1.12 | 0.34 | 1.04 | 0.46 | 1.34 | 18.4 | ME |
| S17-wms-combined_106940 | 102.76 | 293.64 | 4.16 | 10.91 | 60.65 | 8.19 | 117.38 | 13.12 | 4.43 | 17.34 | 36.84 | 151.68 | Shared |
| S17-wms-combined_186314 | 15.51 | 2.29 | 4.31 | 4.45 | 4.79 | 2.78 | 6.56 | 1.58 | 6.12 | 0.68 | 1.23 | 26.1 | ME |
| S17-wms-combined_162786 | 1.5 | 1.79 | 0.11 | 0.43 | 1.27 | 0.26 | 1.55 | 0.87 | 1.48 | 0.51 | 1.81 | 74.24 | ME |
| S17-wms-combined_95360 | 45.23 | 55.02 | 0.36 | 2.27 | 4.75 | 11.09 | 63.36 | 1.86 | 40.05 | 1.39 | 2.84 | 27.14 | Shared |
| S17-wms-combined_130215 | 16.98 | 9.92 | 1.34 | 1.51 | 14.43 | 32.22 | 9.76 | 7.89 | 5.61 | 5.05 | 9.81 | 86.19 | Shared |
| S17-wms-combined_75892 | 6.23 | 3.47 | 0.34 | 0.62 | 2.2 | 0.99 | 2.48 | 1.7 | 0.72 | 1.48 | 1.71 | 26.15 | ME |
| S17-wms-combined_30308 | 0.77 | 1.12 | 0.04 | 0.18 | 1.02 | 0.18 | 0.49 | 0.53 | 0.92 | 0.44 | 0.62 | 13.39 | ME |
| S17-wms-combined_175924 | 6.04 | 6.62 | 0.75 | 2.94 | 4.31 | 5.74 | 3.14 | 4.72 | 4.23 | 4.56 | 5.88 | 112.18 | ME |
| S17-wms-combined_199440 | 64.65 | 9.55 | 3 | 1.49 | 28.53 | 8.04 | 29.93 | 10.41 | 7.3 | 13.7 | 8.17 | 55.36 | Shared |
| S17-wms-combined_199746 | 4.68 | 3.48 | 4.17 | 0.98 | 46.21 | 9.37 | 5.24 | 17.86 | 3.26 | 17.59 | 11.07 | 59.5 | Shared |
| S17-wms-combined_66123 | 7.96 | 2.09 | 0.44 | 2.1 | 1.8 | 1.95 | 5.05 | 9.51 | 1.01 | 2.61 | 1.3 | 39.96 | ME |
| S17-wms-combined_9864 | 3.33 | 3.8 | 1.55 | 0.6 | 21.25 | 6.77 | 16.33 | 13.57 | 1.22 | 8.97 | 6.62 | 22.74 | Shared |
| S17-wms-combined_253499 | 1.02 | 0.89 | 0.09 | 0.96 | 2 | 0.52 | 1.21 | 1.02 | 0.94 | 3.25 | 1.19 | 84.49 | ME |
| S17-wms_124308 | 0.86 | 1.28 | 0.03 | 0.73 | 0.71 | 0.25 | 4.74 | 0.4 | 0.87 | 0.21 | 0.81 | 34.03 | ME |
| S17-wms_235536 | 1.34 | 1.67 | 0.14 | 0.83 | 0.58 | 0.42 | 0.98 | 0.22 | 4.95 | 1.7 | 1.43 | 28.56 | ME |
| S17-wms_116248 | 9.89 | 3.51 | 1.77 | 1.53 | 2.11 | 5.51 | 2.09 | 14.81 | 0.78 | 2.97 | 11.15 | 37.17 | Shared |
| S17-wms_106671 | 2.2 | 6.34 | 0.71 | 2.19 | 2.71 | 0.57 | 1.48 | 2.8 | 3.87 | 8.5 | 5.07 | 35.81 | ME |
| S17-wms_29082 | 8.68 | 4.79 | 2.25 | 4.36 | 7.44 | 2.05 | 4.29 | 5.37 | 3.51 | 10.65 | 5.87 | 43.84 | ME |
| S17-wms_254872 | 0.31 | 0.36 | 0.08 | 0.24 | 0.32 | 0.17 | 0.51 | 0.47 | 0.43 | 0.08 | 0.51 | 23.97 | ME |
| S17-wms_266511 | 0.49 | 0.58 | 0.07 | 0.5 | 0.66 | 0.12 | 0.48 | 0.4 | 0.31 | 0.3 | 0.43 | 239.77 | ME |
| S12-wms-combined_321174 | 5.49 | 0.65 | 0.14 | 0.39 | 2.05 | 1.66 | 6.41 | 5.37 | 0.43 | 14.26 | 64.58 | 0.63 | Shared |
| S12-wms-combined_401379 | 2.05 | 0.69 | 0.05 | 0.24 | 0.84 | 0.26 | 8.74 | 0.66 | 0.79 | 0.34 | 53.45 | 0.84 | SHHC |
| S12-wms-combined_401383 | 0.25 | 0.87 | 0 | 0.19 | 0.36 | 0.08 | 1.04 | 0.3 | 0.34 | 0.06 | 18.31 | 0.13 | SHHC |
| S12-wms-combined_334806 | 27.49 | 12.14 | 2.39 | 1.64 | 34.01 | 8.01 | 7.28 | 20.46 | 4.85 | 16.82 | 42.22 | 25.49 | Shared |
| S12-wms-combined_184338 | 19.14 | 15.04 | 2.2 | 0.9 | 5.71 | 3.07 | 4.4 | 83.9 | 3.52 | 6.26 | 56.22 | 15.92 | Shared |
| S12-wms-combined_368462 | 6.37 | 4.21 | 0.35 | 0.72 | 5.48 | 1.48 | 5.46 | 28.56 | 1.04 | 2.64 | 46.98 | 1.58 | SHHC |
| S12-wms-combined_385270 | 79.58 | 5.82 | 0.55 | 1.27 | 6.76 | 1.14 | 5.37 | 4.33 | 3.4 | 2.87 | 183.56 | 4.6 | Shared |
| S12-wms-combined_201125 | 25.63 | 39 | 1.78 | 14.48 | 27.7 | 6.85 | 54.47 | 7.02 | 10.75 | 12.13 | 160.23 | 30.72 | Shared |
| S12-wms-combined_368562 | 1.64 | 3.07 | 3.09 | 6.27 | 10.41 | 10.8 | 9.81 | 2.07 | 0.72 | 29.85 | 121.9 | 302.89 | Shared |
| S12-wms-combined_351847 | 37.58 | 11.42 | 0.07 | 0.79 | 0.89 | 0.15 | 1.77 | 24.26 | 1.09 | 0.45 | 48.23 | 1.03 | Shared |
| S12-wms-combined_117628 | 112.29 | 42.16 | 3.8 | 6.75 | 28.05 | 7.34 | 16.09 | 35.62 | 11.37 | 18.27 | 55.09 | 27.13 | Shared |
| S12-wms-combined_17617 | 84.11 | 10.04 | 7.27 | 3.66 | 12.07 | 8.8 | 31.73 | 15.97 | 3.77 | 11.57 | 24.76 | 25.79 | Shared |
| S12-wms-combined_51076 | 5.04 | 2.78 | 0.38 | 0.6 | 4.24 | 2.28 | 2.24 | 3.77 | 1.16 | 2.74 | 65.27 | 4.08 | SHHC |
| S12-wms-combined_151843 | 172.69 | 4.46 | 4.74 | 1.39 | 8.06 | 1.6 | 44.47 | 181.19 | 3.7 | 4.19 | 232.91 | 4.48 | Shared |
| S12-wms-combined_302876 | 0.8 | 1.08 | 0.3 | 0.96 | 0.77 | 0.17 | 0.75 | 0.37 | 1 | 0.84 | 16.4 | 0.46 | SHHC |
| S12-wms-combined_18287 | 55.43 | 35.55 | 3.9 | 3.08 | 50.31 | 11.25 | 63.99 | 33.53 | 7.61 | 45.19 | 68.41 | 44.32 | Shared |
| S12-wms-combined_52101 | 1.55 | 1.17 | 0.1 | 0.51 | 1.41 | 0.43 | 1.35 | 1.18 | 1.06 | 0.69 | 13.11 | 0.8 | SHHC |
| S12-wms-combined_320262 | 304.84 | 92.53 | 5.41 | 2.75 | 507.87 | 642.36 | 231.25 | 158.9 | 235.24 | 212.78 | 202.01 | 67.25 | Shared |
| S12-wms-combined_2450 | 5.21 | 3.65 | 0.34 | 1.03 | 78.22 | 8.11 | 5.71 | 26.27 | 2.42 | 13.74 | 22.36 | 2.74 | Shared |
| S12-wms-combined_320727 | 88.64 | 54.27 | 5.03 | 3.3 | 360.96 | 11.27 | 154.8 | 26.46 | 3.5 | 125.87 | 365.62 | 32.42 | Shared |
| S12-wms-combined_371541 | 16.63 | 10.8 | 0.47 | 4.75 | 10.92 | 3.24 | 15.58 | 2.4 | 6.44 | 68.6 | 78.53 | 16.01 | Shared |
| S12-wms-combined_120133 | 11.52 | 29.88 | 0.62 | 3.26 | 5.81 | 2.99 | 8.38 | 1.87 | 22.73 | 1.06 | 25.37 | 11.09 | Shared |
| S12-wms-combined_71235 | 3.14 | 1.64 | 0.7 | 0.54 | 1.38 | 0.54 | 1.04 | 1.46 | 0.85 | 1.12 | 16.55 | 1.18 | SHHC |
| S12-wms-combined_189323 | 50.72 | 21.42 | 3.17 | 6.87 | 591.03 | 156.14 | 39.55 | 39.58 | 31.94 | 222.13 | 496.19 | 42.31 | Shared |

| | | | | | | | | | | | | | |
|---|---|---|---|---|---|---|---|---|---|---|---|---|---|
| S12-wms-combined_21504 | 0.8 | 0.78 | 0.07 | 0.15 | 1.12 | 0.26 | 0.92 | 0.79 | 0.82 | 0.4 | 18.36 | 0.53 | SHHC |
| S12-wms-combined_190114 | 645.11 | 838.25 | 4.22 | 1.83 | 35.52 | 12.12 | 52.89 | 9.45 | 1.56 | 14.15 | 104.29 | 49.87 | Shared |
| S12-wms-combined_256299 | 0.79 | 0.78 | 0.07 | 0.73 | 0.9 | 0.28 | 1.06 | 0.58 | 0.57 | 0.39 | 135.09 | 0.44 | SHHC |
| S12-wms-combined_156234 | 55.16 | 10.1 | 5.4 | 4.8 | 4.12 | 9.55 | 8.82 | 26.44 | 1.29 | 10.65 | 77.91 | 72.26 | Shared |
| S12-wms-combined_223043 | 1.99 | 3.99 | 0.66 | 0.79 | 1.74 | 0.18 | 5.62 | 5.53 | 1.1 | 2.04 | 15.28 | 8.75 | SHHC |
| S12-wms-combined_323943 | 1.9 | 1.27 | 0.23 | 1.69 | 1.8 | 0.63 | 1.13 | 5.37 | 0.94 | 0.74 | 26.94 | 1.16 | SHHC |
| S12-wms-combined_390985 | 0.42 | 0.76 | 0.08 | 0.44 | 0.83 | 0.06 | 0.5 | 0.33 | 0.25 | 0.19 | 32.37 | 0.25 | SHHC |
| S12-wms-combined_191570 | 1.17 | 1.3 | 0.83 | 0.7 | 0.73 | 0.2 | 0.97 | 0.94 | 0.45 | 0.5 | 51.72 | 0.63 | SHHC |
| S12-wms-combined_23517 | 116.25 | 15.61 | 4.61 | 2.22 | 58.24 | 14.43 | 9.2 | 181.15 | 5.08 | 20.08 | 200.92 | 23.55 | Shared |
| S12-wms-combined_375343 | 9.6 | 3.32 | 0.23 | 0.66 | 5.08 | 2.25 | 4.75 | 1.62 | 3.49 | 43.71 | 68.05 | 8.5 | Shared |
| S12-wms-combined_325418 | 4.79 | 1.38 | 0.11 | 0.66 | 1.21 | 0.48 | 2 | 1.54 | 2.54 | 1.66 | 36.36 | 0.86 | SHHC |
| S12-wms-combined_41160 | 4.96 | 2.61 | 0.27 | 0.45 | 1.91 | 0.79 | 1.58 | 42.09 | 0.99 | 1.56 | 34.05 | 1.52 | SHHC |
| S12-wms-combined_7174 | 16.93 | 4.67 | 2.79 | 1.3 | 11.38 | 4.69 | 43.87 | 9.59 | 2.33 | 101.92 | 50.24 | 2.26 | Shared |
| S12-wms-combined_358010 | 5.94 | 5.05 | 2.67 | 1.7 | 3.17 | 0.65 | 2.09 | 7.07 | 1.11 | 1.92 | 41.99 | 1.71 | SHHC |
| S12-wms-combined_376326 | 1.39 | 1.37 | 1.89 | 2.45 | 1.05 | 0.34 | 0.87 | 0.99 | 0.78 | 0.48 | 33.6 | 0.78 | SHHC |
| S12-wms-combined_91231 | 0.74 | 1.03 | 0.05 | 1 | 0.78 | 0.23 | 1.06 | 0.45 | 0.61 | 2.86 | 24.13 | 0.43 | SHHC |
| S12-wms-combined_107942 | 1.53 | 1.2 | 0.12 | 0.31 | 0.64 | 0.26 | 3.22 | 0.53 | 1.18 | 13.81 | 28.91 | 1.21 | Shared |
| S12-wms-combined_293354 | 248.26 | 1.16 | 3.92 | 0.36 | 1.78 | 0.65 | 69.44 | 281.01 | 0.89 | 1.02 | 251.78 | 1.03 | Shared |
| S12-wms-combined_75733 | 0.78 | 0.52 | 0.26 | 1.2 | 1.1 | 0.17 | 0.8 | 0.49 | 0.59 | 0.15 | 25.41 | 0.42 | SHHC |
| S12-wms-combined_393703 | 53.59 | 6.56 | 1.02 | 2.2 | 2.67 | 2.12 | 2.02 | 14.35 | 32.09 | 1.5 | 28.65 | 5.17 | Shared |
| S12-wms-combined_358797 | 107.71 | 9.04 | 8.8 | 7.35 | 173.86 | 63.18 | 96.06 | 65.71 | 17.14 | 101.18 | 99.4 | 14.97 | Shared |
| S12-wms-combined_25459 | 143.21 | 9.6 | 12.28 | 12.68 | 226.39 | 137.86 | 141.15 | 134.69 | 20.29 | 153.48 | 656.64 | 26.72 | Shared |
| S12-wms-combined_359190 | 7.58 | 23.02 | 0.47 | 2.56 | 1.78 | 1.01 | 5.4 | 9.55 | 6.18 | 1.77 | 36.28 | 2.22 | SHHC |
| S12-wms-combined_259783 | 1.34 | 3.74 | 0.38 | 0.44 | 0.84 | 1.39 | 0.76 | 10.53 | 0.43 | 1.2 | 25.72 | 67.32 | Shared |
| S12-wms-combined_260036 | 0.79 | 0.92 | 0.05 | 0.75 | 14.57 | 0.17 | 1.4 | 0.71 | 1.04 | 0.27 | 32.75 | 2.24 | Shared |
| S12-wms-combined_210942 | 70.51 | 15 | 9.66 | 12.92 | 13.76 | 6.13 | 5.51 | 21.1 | 2.62 | 5.34 | 61.75 | 14.96 | Shared |
| S12-wms-combined_328554 | 24.54 | 13.72 | 0.96 | 0.93 | 77.64 | 81.14 | 26.89 | 30.86 | 33.07 | 33.31 | 89.61 | 25.22 | Shared |
| S12-wms-combined_329242 | 14.54 | 17.58 | 0.5 | 2.74 | 2.7 | 42.84 | 2.21 | 6.67 | 2.86 | 1.47 | 24.31 | 5.07 | Shared |
| S12-wms-combined_345948 | 9.04 | 1.97 | 1.73 | 0.95 | 2.73 | 3.55 | 2.63 | 15.26 | 0.75 | 1.96 | 18.59 | 3.17 | SHHC |
| S12-wms-combined_313953 | 1.12 | 1.09 | 0.05 | 0.31 | 1.11 | 0.26 | 8.06 | 1.28 | 0.75 | 0.96 | 65.33 | 0.64 | SHHC |
| S12-wms-combined_245755 | 1.24 | 1.14 | 0.08 | 0.25 | 1.39 | 1.03 | 1.02 | 1.09 | 0.43 | 0.45 | 18.43 | 0.62 | SHHC |
| S12-wms-combined_111524 | 129.73 | 38.88 | 8.21 | 2.69 | 361.77 | 210.32 | 100.25 | 96.18 | 86.68 | 158.42 | 131.63 | 78.06 | Shared |
| S12-wms-combined_263377 | 1.25 | 1.03 | 0.12 | 0.18 | 1.28 | 0.33 | 0.84 | 1.07 | 0.71 | 0.78 | 27.62 | 0.8 | SHHC |
| S12-wms-combined_297545 | 30.79 | 49.52 | 2.21 | 22.63 | 29.17 | 7.72 | 504.61 | 8.19 | 13.35 | 14.62 | 131.73 | 36.81 | Shared |
| S12-wms-combined_129554 | 1.02 | 0.54 | 0.37 | 0.14 | 1.18 | 0.27 | 0.47 | 4.45 | 0.36 | 0.46 | 17.36 | 0.48 | SHHC |
| S12-wms-combined_63056 | 6.21 | 2.97 | 0.24 | 0.51 | 1.19 | 0.36 | 1.11 | 0.74 | 0.64 | 7.91 | 19 | 0.89 | SHHC |
| S12-wms-combined_264077 | 6.62 | 2.99 | 0.88 | 1.41 | 2.06 | 1.32 | 1.71 | 4.76 | 1.1 | 2.03 | 24.77 | 6.23 | SHHC |
| S12-wms-combined_129799 | 85.2 | 26.97 | 7.83 | 5.38 | 65.94 | 4.07 | 9.16 | 57.32 | 2.6 | 21.67 | 103.02 | 12.88 | Shared |
| S12-wms-combined_63649 | 21.19 | 1.04 | 0.3 | 0.18 | 1.58 | 0.49 | 0.93 | 11.48 | 0.5 | 0.71 | 55.86 | 0.84 | Shared |
| S12-wms-combined_282594 | 20.82 | 51.02 | 0.79 | 0.85 | 117.37 | 2.02 | 8.11 | 102.67 | 2.01 | 15.44 | 84.32 | 3.82 | Shared |
| S12-wms-combined_264425 | 5.19 | 2.41 | 2.43 | 5.56 | 0.97 | 0.58 | 2.12 | 0.9 | 0.91 | 0.67 | 65.24 | 0.97 | SHHC |
| S12-wms-combined_147628 | 7.56 | 3.88 | 0.51 | 1.01 | 6.48 | 0.63 | 1.79 | 8.69 | 1.07 | 1.21 | 69.27 | 3.5 | SHHC |
| S12-wms-combined_48648 | 22.48 | 6.78 | 2.96 | 5.71 | 16.22 | 1.7 | 2.79 | 69.38 | 3.29 | 1.78 | 64.75 | 11 | Shared |
| S12-wms-combined_264845 | 43.69 | 12.56 | 5.62 | 10.69 | 16.84 | 4.12 | 5.96 | 23.14 | 4.29 | 13.02 | 51.55 | 26.44 | Shared |
| S12-wms-combined_299293 | 16.35 | 19.05 | 2.9 | 2.64 | 6.74 | 1.49 | 3.88 | 22.55 | 1.91 | 3.49 | 46.99 | 6.32 | Shared |
| S12-wms-combined_98046 | 2.28 | 1.52 | 0.15 | 0.32 | 1.83 | 0.69 | 1.61 | 3.12 | 0.78 | 1.09 | 24.25 | 1.16 | SHHC |
| S12-wms-combined_299872 | 0.58 | 2.88 | 0.07 | 0.21 | 0.52 | 0.23 | 1.12 | 0.35 | 0.58 | 0.29 | 29.8 | 2.67 | SHHC |
| S12-wms-combined_148741 | 30.2 | 1.9 | 0.27 | 0.77 | 3.18 | 0.99 | 1.63 | 7.36 | 1.09 | 1.5 | 68.04 | 2.23 | Shared |
| S12-wms-combined_31761 | 2.85 | 5.36 | 0.2 | 1.1 | 1.47 | 1.17 | 2.82 | 0.94 | 4.54 | 0.67 | 23.69 | 1.7 | SHHC |
| S12-wms-combined_99704 | 2.12 | 1.32 | 0.15 | 0.25 | 1.74 | 0.96 | 2 | 1.56 | 5.05 | 1.2 | 15.46 | 4.91 | SHHC |
| S12-wms-combined_132907 | 12.7 | 1.06 | 0.6 | 0.84 | 3.81 | 2.56 | 2.6 | 5.8 | 0.77 | 3.47 | 36.15 | 0.83 | Shared |
| S12-wms-combined_250551 | 84.11 | 114.72 | 0.63 | 1.55 | 1.2 | 0.98 | 3.83 | 5.79 | 0.73 | 2.95 | 175.57 | 1.05 | Shared |
| S12-wms-combined_368190 | 43.03 | 58.55 | 2.34 | 32.39 | 24.26 | 8.22 | 93.25 | 12.41 | 6.59 | 41.19 | 163.52 | 142.13 | Shared |
| S12-wms-combined_117447 | 0.5 | 0.55 | 0.11 | 0.19 | 0.4 | 0.13 | 0.28 | 0.6 | 0.24 | 0.19 | 136.97 | 0.29 | SHHC |
| S12-wms-combined_168259 | 5.5 | 12.23 | 0.31 | 0.55 | 26.96 | 0.65 | 1.81 | 23.06 | 0.93 | 6.53 | 58.04 | 1.7 | Shared |
| S12-wms-combined_270839 | 28.67 | 4.78 | 1.23 | 0.74 | 23.71 | 2.05 | 3.45 | 24.28 | 1.23 | 5.9 | 66.11 | 3.73 | Shared |
| S12-wms-combined_157167 | 1.95 | 0.44 | 0.05 | 0.04 | 0.56 | 0.28 | 0.31 | 0.43 | 0.52 | 0.17 | 49.54 | 0.25 | SHHC |
| S12-wms-combined_164656 | 1.2 | 1.39 | 0.17 | 0.4 | 1.17 | 0.39 | 1.5 | 0.93 | 0.62 | 0.63 | 232.33 | 0.82 | SHHC |
| S12-wms-combined_133608 | 1.4 | 2.11 | 0.01 | 0.05 | 0.6 | 0.15 | 0.4 | 0.16 | 0.1 | 0.2 | 13.7 | 0.32 | SHHC |
| S12-wms_184276 | 1.3 | 0.82 | 0.12 | 0.18 | 1.43 | 0.91 | 1.38 | 1.52 | 0.47 | 5.32 | 45.8 | 0.81 | SHHC |
| S12-wms_235125 | 2.86 | 1.92 | 4.29 | 1.24 | 2.15 | 0.77 | 1.71 | 2.33 | 0.96 | 2.14 | 258.65 | 2.88 | SHHC |
| S12-wms_17950 | 1.02 | 1.24 | 1.77 | 1.06 | 2.03 | 0.91 | 0.84 | 1.17 | 0.37 | 2.37 | 18.18 | 0.48 | SHHC |
| S12-wms_135291 | 96.22 | 63.95 | 3.74 | 10.41 | 26.27 | 7.24 | 25.99 | 32.26 | 46.78 | 17.22 | 133.24 | 63.82 | Shared |
| S12-wms_352846 | 0.58 | 1 | 0.03 | 0.14 | 0.82 | 0.2 | 0.73 | 0.4 | 0.38 | 0.25 | 49.7 | 0.25 | SHHC |
| S12-wms_386692 | 14.32 | 5.15 | 1.46 | 0.77 | 13.12 | 1.57 | 7.75 | 20.65 | 0.42 | 3.64 | 387.22 | 1.51 | Shared |
| S12-wms_202803 | 62.69 | 1.96 | 3.7 | 0.6 | 2.97 | 1.02 | 2 | 4.33 | 1.06 | 3.21 | 37.5 | 5.81 | Shared |

| | | | | | | | | | | | | | |
|---|---|---|---|---|---|---|---|---|---|---|---|---|---|
| S12-wms_336780 | 17.25 | 1.36 | 0.07 | 0.37 | 0.34 | 0.19 | 0.52 | 0.2 | 0.74 | 0.37 | 32.18 | 0.4 | Shared |
| S12-wms_373775 | 37.66 | 2.75 | 0.7 | 1.56 | 2.43 | 1.03 | 2.48 | 57.33 | 1.1 | 0.98 | 57.82 | 3.04 | Shared |
| S12-wms_223235 | 0.64 | 1.68 | 4.61 | 0.34 | 0.45 | 0.13 | 4.34 | 1.05 | 1.37 | 0.19 | 14.08 | 0.4 | SHHC |
| S12-wms_193565 | 1.82 | 12.37 | 0.09 | 0.26 | 1.09 | 0.4 | 0.61 | 1.93 | 0.59 | 0.99 | 26.5 | 0.82 | SHHC |
| S12-wms_161239 | 59.85 | 17.65 | 5.92 | 3.4 | 114.59 | 6.5 | 15.56 | 34.78 | 2.32 | 31.42 | 50.83 | 22.06 | Shared |
| S12-wms_44817 | 0.86 | 0.74 | 0.06 | 0.42 | 2.33 | 0.43 | 0.94 | 0.94 | 0.56 | 3.14 | 60.19 | 0.45 | SHHC |
| S12-wms_179388 | 11.01 | 2.17 | 0.14 | 6.62 | 1.05 | 0.35 | 1.94 | 0.96 | 2.2 | 10.58 | 18.26 | 1.2 | Shared |
| S12-wms_12658 | 2.54 | 5.21 | 0.38 | 0.81 | 3.02 | 0.88 | 20.7 | 1.02 | 1.76 | 5.33 | 74.96 | 7.98 | Shared |
| S12-wms_383851 | 2.5 | 2.14 | 0.11 | 1.08 | 2.2 | 0.5 | 38.15 | 1.5 | 0.79 | 2.36 | 155.27 | 1.4 | Shared |
| S12-wms_15700 | 0.66 | 1.22 | 0.15 | 0.59 | 0.53 | 0.14 | 1.04 | 0.24 | 0.66 | 4.45 | 26.74 | 0.31 | SHHC |
| S12-wms_401366 | 1.3 | 2 | 0.07 | 0.33 | 0.64 | 0.23 | 0.71 | 0.68 | 0.62 | 2.4 | 43.59 | 0.45 | SHHC |
| S12-wms_401372 | 0.53 | 1.16 | 0.33 | 0.72 | 0.46 | 0.22 | 1.2 | 0.24 | 0.65 | 0.33 | 150.78 | 0.37 | SHHC |
| S2-wms-combined_207832 | 2.73 | 191.02 | 1.99 | 1.18 | 2.28 | 1.07 | 6.62 | 1.53 | 0.71 | 4.16 | 83.41 | 1.56 | SHHC |
| S2-wms-combined_248318 | 3.17 | 26.83 | 0.16 | 0.14 | 2.61 | 0.5 | 1.48 | 10.16 | 1.26 | 0.7 | 3.05 | 9.99 | SHHC |
| S2-wms-combined_248329 | 1.68 | 15.25 | 0.08 | 0.24 | 1.21 | 0.3 | 0.85 | 0.94 | 0.63 | 0.51 | 1.03 | 0.61 | SHHC |
| S2-wms-combined_31084 | 1.61 | 107.67 | 0.14 | 0.68 | 0.98 | 0.82 | 1.79 | 0.51 | 1.72 | 0.34 | 3.93 | 0.77 | SHHC |
| S2-wms-combined_196783 | 16.96 | 32.94 | 1.93 | 2.4 | 23.23 | 9.54 | 8.97 | 11.51 | 9.51 | 20.39 | 12.05 | 50.68 | Shared |
| S2-wms-combined_165833 | 3.61 | 18.15 | 0.12 | 0.38 | 1.86 | 0.31 | 3.37 | 0.54 | 1.17 | 0.32 | 1.06 | 7.22 | SHHC |
| S2-wms-combined_176376 | 1.05 | 15.73 | 0.1 | 0.68 | 0.82 | 0.67 | 0.72 | 1.16 | 0.78 | 0.31 | 1.32 | 4.31 | SHHC |
| S2-wms-combined_62377 | 90.91 | 190.42 | 0.11 | 0.34 | 0.76 | 0.12 | 0.74 | 0.57 | 0.57 | 0.36 | 0.79 | 0.51 | Shared |
| S2-wms-combined_72725 | 42.82 | 88.08 | 1.36 | 3.64 | 50.69 | 13.6 | 16.69 | 43.68 | 16.03 | 20.81 | 24.82 | 60.26 | Shared |
| S2-wms-combined_135014 | 27.51 | 118.6 | 0.27 | 0.65 | 0.99 | 15.83 | 63.37 | 0.88 | 3.75 | 0.73 | 1.05 | 20.94 | Shared |
| S2-wms-combined_62519 | 82.49 | 184.64 | 6.79 | 4.5 | 22.83 | 7.24 | 10.37 | 124.36 | 4.24 | 25.77 | 52.31 | 57.92 | Shared |
| S2-wms-combined_135139 | 36.95 | 109.53 | 0.71 | 0.64 | 154.8 | 1.16 | 8.3 | 133.84 | 1.18 | 14.69 | 37.34 | 1.66 | Shared |
| S2-wms-combined_218002 | 15.51 | 85.74 | 0.97 | 3.24 | 3.04 | 3.97 | 13.54 | 6.83 | 11.65 | 4.59 | 5.16 | 15.79 | Shared |
| S2-wms-combined_135154 | 65.72 | 96.6 | 0.25 | 1.3 | 5.24 | 11.93 | 63.2 | 1.63 | 41.55 | 1.05 | 2.01 | 26.87 | Shared |
| S2-wms-combined_238929 | 16.9 | 38.66 | 0.88 | 0.54 | 2.39 | 0.47 | 1.84 | 6.77 | 0.96 | 1.61 | 7.46 | 1.35 | Shared |
| S2-wms-combined_93845 | 10.87 | 63.06 | 0.68 | 0.95 | 8.07 | 3.3 | 2.25 | 56.36 | 2.24 | 3.25 | 9.56 | 3.84 | Shared |
| S2-wms-combined_73327 | 0.98 | 15.39 | 0.06 | 0.78 | 0.87 | 0.29 | 0.79 | 0.5 | 2.45 | 1.01 | 1.08 | 1.88 | SHHC |
| S2-wms-combined_52568 | 14.82 | 51.74 | 1.51 | 1.27 | 13.85 | 2.91 | 3.79 | 45.49 | 2.11 | 6.1 | 17.17 | 7.75 | Shared |
| S2-wms-combined_166665 | 44.11 | 71.25 | 7.49 | 15.99 | 3.08 | 39.54 | 7.09 | 25.96 | 3.87 | 9.64 | 31.37 | 95.71 | Shared |
| S2-wms-combined_62839 | 23.61 | 57.21 | 0.14 | 0.73 | 0.91 | 0.34 | 1.14 | 0.9 | 0.97 | 0.47 | 1.08 | 0.78 | Shared |
| S2-wms-combined_197577 | 4.99 | 45.19 | 1.98 | 0.41 | 0.65 | 0.5 | 0.89 | 0.27 | 0.86 | 0.19 | 0.58 | 0.37 | SHHC |
| S2-wms-combined_208559 | 149.5 | 56.71 | 11.07 | 5.31 | 447.38 | 129.12 | 100.05 | 134.25 | 30.78 | 109.64 | 136.34 | 29.6 | Shared |
| S2-wms-combined_229529 | 46.37 | 173.2 | 1.69 | 12.15 | 60.36 | 10.49 | 58.92 | 10 | 48 | 13.52 | 21.32 | 28.14 | Shared |
| S2-wms-combined_115352 | 0.86 | 13.12 | 0.02 | 0.16 | 2.34 | 0.81 | 0.99 | 0.88 | 0.9 | 0.52 | 0.77 | 0.54 | SHHC |
| S2-wms-combined_84608 | 102.77 | 58.45 | 2.24 | 1.3 | 1.17 | 0.3 | 5.99 | 2.5 | 0.63 | 2.86 | 1.26 | 0.72 | Shared |
| S2-wms-combined_126134 | 2.53 | 41.42 | 0.79 | 1.65 | 3.06 | 5.13 | 6.84 | 2.57 | 2.9 | 3.94 | 4.29 | 16.22 | Shared |
| S2-wms-combined_74387 | 3.29 | 20.55 | 0.2 | 0.52 | 2.3 | 0.81 | 1.91 | 4.22 | 1.45 | 3.06 | 4.18 | 2.19 | SHHC |
| S2-wms-combined_230282 | 2.95 | 6.46 | 0.67 | 0.31 | 1.6 | 0.86 | 0.52 | 1.42 | 1.47 | 1.05 | 6.88 | 1.31 | SHHC |
| S2-wms-combined_126440 | 1.76 | 24.57 | 0.16 | 0.5 | 0.92 | 1.14 | 1.73 | 0.71 | 0.95 | 0.74 | 1.04 | 0.45 | SHHC |
| S2-wms-combined_147065 | 73.55 | 186.02 | 1.74 | 4.77 | 29.75 | 18.71 | 44.45 | 13.52 | 19.24 | 16.34 | 18.23 | 26.96 | Shared |
| S2-wms-combined_137119 | 8.6 | 53.73 | 0.68 | 0.91 | 9.17 | 0.82 | 4.46 | 5.59 | 1.31 | 10.55 | 12.02 | 9.05 | Shared |
| S2-wms-combined_85381 | 2.55 | 26.34 | 0.26 | 0.82 | 2.08 | 1.5 | 6.96 | 2.39 | 6.53 | 7.81 | 2.92 | 5.99 | SHHC |
| S2-wms-combined_116221 | 21.93 | 20.59 | 0.34 | 4.21 | 4.97 | 5.08 | 2.75 | 20.92 | 1.31 | 5.43 | 22.94 | 3.65 | Shared |
| S2-wms-combined_95987 | 0.96 | 13.03 | 0.13 | 0.53 | 1.66 | 0.29 | 0.43 | 0.61 | 0.77 | 0.49 | 0.84 | 0.54 | SHHC |
| S2-wms-combined_157934 | 7.03 | 66.04 | 7.26 | 25 | 4.18 | 12.37 | 6.67 | 5.01 | 35.94 | 2.38 | 5.45 | 4.53 | Shared |
| S2-wms-combined_64692 | 7.21 | 27.06 | 0.15 | 0.62 | 0.92 | 0.84 | 1.35 | 1.13 | 0.53 | 1.39 | 0.87 | 1.31 | SHHC |
| S2-wms-combined_127080 | 3.42 | 18.12 | 0.21 | 0.94 | 1.45 | 0.48 | 1.22 | 1.63 | 0.82 | 1.11 | 2.48 | 1.08 | SHHC |
| S2-wms-combined_13246 | 146.18 | 134.84 | 19.07 | 24.88 | 12.49 | 24.53 | 20.88 | 76.53 | 3.61 | 47.51 | 59.09 | 201.25 | Shared |
| S2-wms-combined_199384 | 4.37 | 11.65 | 0.66 | 0.26 | 2.65 | 0.55 | 1.96 | 2.33 | 1.42 | 2.56 | 6.88 | 3.46 | SHHC |
| S2-wms-combined_55495 | 3.77 | 77.42 | 0.14 | 0.16 | 1.6 | 2.57 | 0.81 | 0.93 | 0.75 | 2.47 | 3.16 | 0.85 | SHHC |
| S2-wms-combined_158821 | 40.98 | 42.91 | 0.53 | 7.26 | 0.78 | 1.65 | 1.11 | 6.29 | 0.86 | 8.26 | 10.42 | 0.53 | Shared |
| S2-wms-combined_210822 | 675.74 | 819.46 | 47.34 | 37.99 | 1546.97 | 343.03 | 1259.36 | 210.47 | 78.36 | 461.06 | 902.13 | 82.64 | Shared |
| S2-wms-combined_190624 | 0.89 | 19.47 | 0.04 | 0.57 | 0.59 | 0.2 | 0.68 | 0.59 | 0.79 | 0.23 | 0.6 | 0.55 | SHHC |
| S2-wms-combined_24549 | 33.26 | 73.77 | 2.92 | 2.61 | 39.9 | 9.66 | 13.23 | 23.79 | 6.54 | 51.36 | 26.42 | 65.31 | Shared |
| S2-wms-combined_180640 | 19.94 | 49.97 | 0.32 | 0.91 | 0.61 | 1.1 | 1.67 | 0.87 | 1.39 | 0.37 | 1.28 | 1.03 | Shared |
| S2-wms-combined_170845 | 6.22 | 38.05 | 0.44 | 1.67 | 1.4 | 0.52 | 5.23 | 4.3 | 2.99 | 0.79 | 1.77 | 6.36 | SHHC |
| S2-wms-combined_159670 | 16.97 | 19.75 | 1.3 | 0.94 | 30.17 | 4.03 | 3.87 | 7.14 | 1.19 | 7.59 | 9.64 | 6.73 | Shared |
| S2-wms-combined_181391 | 19.46 | 37.87 | 1.58 | 2.28 | 19.92 | 13.15 | 7.75 | 16.44 | 21.79 | 19.55 | 12.73 | 79.82 | Shared |
| S2-wms-combined_200390 | 86.75 | 83.27 | 6.3 | 8.73 | 20.68 | 12.93 | 29.15 | 23.96 | 86.77 | 16.47 | 31.87 | 41.16 | Shared |
| S2-wms-combined_233107 | 18.93 | 23.81 | 0.32 | 1.84 | 43.34 | 2.52 | 13.19 | 10.35 | 12.37 | 9.48 | 6.6 | 28.57 | Shared |
| S2-wms-combined_57293 | 211.1 | 130.77 | 23.34 | 48.29 | 11.22 | 75.16 | 31.08 | 90.99 | 5.25 | 34.28 | 90.33 | 262.42 | Shared |
| S2-wms-combined_191613 | 4.68 | 22.11 | 0.36 | 1.04 | 2.05 | 2.65 | 2.03 | 10.4 | 2.78 | 1.71 | 3.94 | 11.45 | Shared |
| S2-wms-combined_98888 | 77.21 | 95.29 | 6.67 | 25.88 | 48.22 | 10.26 | 39.33 | 30.02 | 62.95 | 22.47 | 45.92 | 48.81 | Shared |
| S2-wms-combined_181849 | 106.7 | 156.85 | 12.55 | 16.35 | 12.41 | 20.77 | 17.67 | 50.45 | 2.6 | 35.66 | 41.89 | 133.28 | Shared |

| | | | | | | | | | | | | | |
|---|---|---|---|---|---|---|---|---|---|---|---|---|---|
| S2-wms-combined_48661 | 4.91 | 16.46 | 0.66 | 3.08 | 3.71 | 1.32 | 4.29 | 2.01 | 6.76 | 2.02 | 5.3 | 3.69 | SHHC |
| S2-wms-combined_201205 | 39.26 | 129.16 | 0.07 | 0.54 | 2.54 | 0.57 | 1.18 | 3.68 | 0.85 | 0.95 | 1.62 | 0.87 | Shared |
| S2-wms-combined_99446 | 8.66 | 64.13 | 0.76 | 3.98 | 1.66 | 1.15 | 9.12 | 2.12 | 9.96 | 0.81 | 2.24 | 5.99 | SHHC |
| S2-wms-combined_25727 | 17.23 | 33.23 | 2.5 | 2.04 | 3.23 | 0.81 | 1.85 | 17.25 | 1 | 1.36 | 29.21 | 1.53 | Shared |
| S2-wms-combined_78670 | 65.28 | 63.69 | 3.6 | 2.65 | 50.54 | 1.97 | 6.77 | 18.95 | 2.79 | 12.57 | 42.37 | 5.87 | Shared |
| S2-wms-combined_58121 | 128.36 | 305.21 | 3.66 | 26 | 73.04 | 39.19 | 312.64 | 14.78 | 7.47 | 24.13 | 163.67 | 155.16 | Shared |
| S2-wms-combined_140996 | 145.41 | 164.68 | 11.51 | 6.47 | 64.81 | 10.48 | 12.37 | 198.85 | 7.57 | 41.91 | 70.52 | 27.8 | Shared |
| S2-wms-combined_89742 | 12.87 | 9.59 | 0.04 | 0.23 | 1.12 | 0.26 | 0.97 | 0.42 | 0.86 | 0.34 | 0.68 | 0.48 | Shared |
| S2-wms-combined_222535 | 1.99 | 54.74 | 0.21 | 2.22 | 0.89 | 0.35 | 1.57 | 0.73 | 3.74 | 0.7 | 0.75 | 0.56 | SHHC |
| S2-wms-combined_58563 | 15.34 | 53.86 | 1.22 | 1.27 | 7.93 | 2.5 | 2.84 | 76.05 | 2.26 | 4.51 | 25.41 | 3.86 | Shared |
| S2-wms-combined_174241 | 21.33 | 28.03 | 0.98 | 1.01 | 9.91 | 0.88 | 9.87 | 3.63 | 3.15 | 13.38 | 4.33 | 2.29 | Shared |
| S2-wms-combined_110215 | 16.49 | 22 | 1.86 | 3.47 | 5.42 | 2.57 | 6.63 | 7.37 | 1.32 | 5.51 | 6.26 | 14.89 | Shared |
| S2-wms-combined_5791 | 36.66 | 53.37 | 3.61 | 4.86 | 7.97 | 2.63 | 10.46 | 7.34 | 25.61 | 7.68 | 11.02 | 10.97 | Shared |
| S2-wms-combined_51446 | 0.31 | 10.91 | 0.05 | 0.22 | 0.55 | 0.12 | 0.4 | 0.78 | 0.43 | 0.28 | 0.87 | 0.31 | SHHC |
| S2-wms-combined_101826 | 4.04 | 44.35 | 2.95 | 0.5 | 1.41 | 0.38 | 22.13 | 7.79 | 1.64 | 0.53 | 9.22 | 0.97 | Shared |
| S2-wms-combined_122457 | 16.39 | 65.87 | 2.11 | 2.26 | 12.19 | 7.3 | 7.86 | 10.02 | 4.18 | 27.62 | 10.15 | 40.34 | Shared |
| S2-wms-combined_244564 | 26.1 | 29.87 | 1.62 | 3.28 | 22.73 | 2.31 | 18.69 | 25.28 | 9.79 | 7.82 | 20.18 | 10.94 | Shared |
| S2-wms-combined_6605 | 13.06 | 21.19 | 0.33 | 2.47 | 2.61 | 40.7 | 2.25 | 6.84 | 3.22 | 1.91 | 22.68 | 6.39 | Shared |
| S2-wms-combined_185494 | 31.63 | 75.2 | 0.12 | 0.66 | 2.28 | 10.86 | 64.53 | 0.77 | 36.7 | 0.52 | 1.09 | 10.94 | Shared |
| S2-wms-combined_132966 | 1.26 | 13.87 | 0.09 | 0.17 | 1.05 | 0.22 | 1.19 | 0.69 | 0.6 | 0.36 | 0.81 | 0.77 | SHHC |
| S2-wms-combined_225908 | 110.2 | 36.86 | 1.52 | 2.72 | 12.11 | 3.09 | 5.13 | 12.21 | 3.38 | 7.45 | 36.48 | 15.06 | Shared |
| S2-wms-combined_246480 | 1.13 | 20.47 | 0.11 | 0.62 | 1.07 | 0.21 | 0.86 | 0.52 | 0.97 | 0.41 | 0.98 | 0.61 | SHHC |
| S2-wms-combined_113619 | 385.13 | 171.37 | 0.29 | 1.41 | 4.47 | 4.78 | 6 | 33.94 | 63.9 | 1.66 | 1.34 | 0.78 | Shared |
| S2-wms-combined_227800 | 9.61 | 37.72 | 1.45 | 1.57 | 22.66 | 15.4 | 9.74 | 12.69 | 14.21 | 9.75 | 12.61 | 28.61 | Shared |
| S2-wms-combined_30750 | 5.95 | 31.7 | 1.4 | 0.46 | 0.88 | 0.31 | 9.16 | 3.76 | 1.08 | 0.39 | 6.93 | 0.64 | SHHC |
| S2-wms-combined_52006 | 36.84 | 36.59 | 3.12 | 1.91 | 467.04 | 1.03 | 47.05 | 28.77 | 0.69 | 6.04 | 18.91 | 5.09 | Shared |
| S2-wms-combined_75144 | 23.43 | 87.15 | 0.25 | 1.91 | 26.93 | 3.47 | 28.47 | 1.3 | 0.65 | 17.15 | 7.25 | 4.74 | Shared |
| S2-wms-combined_181095 | 3.68 | 24.55 | 1.07 | 2.94 | 1.38 | 4.61 | 0.57 | 1.43 | 0.39 | 1.04 | 4.33 | 1.45 | SHHC |
| S2-wms-combined_4685 | 46.28 | 165.47 | 2.25 | 0.66 | 1.88 | 0.56 | 2.04 | 103.73 | 1.12 | 5.16 | 10.23 | 1.48 | Shared |
| S2-wms-combined_143737 | 65.83 | 143.91 | 0.9 | 1.25 | 18.22 | 28.48 | 31.72 | 3.16 | 4.4 | 4.91 | 8.51 | 10.57 | Shared |
| S2-wms-combined_204225 | 0.38 | 12.12 | 0.01 | 0.05 | 0.62 | 0.11 | 0.42 | 0.19 | 1.46 | 0.62 | 0.31 | 0.18 | SHHC |
| S2-wms-combined_7756 | 35.87 | 111.72 | 0.33 | 0.6 | 80.53 | 0.59 | 4.36 | 56.65 | 1.02 | 6.06 | 18.44 | 0.94 | Shared |
| S2-wms-combined_207000 | 2.81 | 41.78 | 1.07 | 3.56 | 3.85 | 1.84 | 2.15 | 1.54 | 6.27 | 1.07 | 1.94 | 1.14 | SHHC |
| S2-wms_124416 | 48.39 | 107.07 | 2.76 | 6.45 | 12.45 | 4.92 | 13.91 | 21.29 | 5.53 | 10.54 | 24.47 | 22.56 | Shared |
| S2-wms_156439 | 26.94 | 106.95 | 0.96 | 3.75 | 58.94 | 3.74 | 30.63 | 7.7 | 215.23 | 7.35 | 30.12 | 11.07 | Shared |
| S2-wms_53881 | 1.55 | 3031.13 | 1.05 | 1.1 | 0.63 | 0.14 | 0.81 | 0.78 | 2.06 | 1.64 | 2.23 | 1.46 | SHHC |
| S2-wms_23562 | 5.99 | 20.88 | 0.07 | 0.31 | 0.65 | 0.12 | 0.7 | 0.3 | 0.86 | 0.19 | 0.8 | 0.33 | SHHC |
| S2-wms_107329 | 6.24 | 29.18 | 0.89 | 0.97 | 8.07 | 3.21 | 7.11 | 5.25 | 1.66 | 17.42 | 5.55 | 19.03 | Shared |
| S2-wms_128831 | 0.53 | 118.94 | 0.17 | 0.54 | 0.51 | 0.16 | 1.56 | 0.31 | 0.7 | 0.3 | 0.5 | 0.51 | SHHC |
| S2-wms_128962 | 17.34 | 3364.14 | 1.16 | 1.93 | 0.49 | 4.1 | 0.57 | 0.29 | 1.53 | 1.26 | 2.51 | 1.42 | Shared |
| S2-wms_140933 | 11.66 | 23.86 | 0.58 | 2.28 | 3.22 | 1.4 | 4.07 | 2.54 | 6.93 | 2.01 | 3.58 | 4.47 | Shared |
| S2-wms_192667 | 2.7 | 29.96 | 0.5 | 1.08 | 0.7 | 3.67 | 3.96 | 2.04 | 1.66 | 7.38 | 5.13 | 0.64 | SHHC |
| S2-wms_79304 | 0.79 | 37.08 | 0.16 | 0.41 | 0.59 | 0.17 | 0.73 | 0.58 | 0.58 | 0.31 | 0.64 | 3.07 | SHHC |
| S2-wms_161986 | 17.09 | 66.05 | 10.37 | 0.76 | 1.59 | 0.78 | 2.54 | 3.13 | 1.63 | 7.05 | 10.78 | 2.09 | Shared |
| S2-wms_79944 | 15.7 | 91.4 | 0.27 | 0.67 | 1.11 | 0.21 | 1.53 | 2.46 | 0.84 | 28.36 | 1.64 | 0.53 | Shared |
| S2-wms_130687 | 8.7 | 52.12 | 1.42 | 6.39 | 132.79 | 8.02 | 16.1 | 4.98 | 6.75 | 3.65 | 4.61 | 1.28 | Shared |
| S2-wms_110761 | 16.7 | 8.63 | 0.03 | 0.16 | 0.45 | 0.13 | 0.27 | 0.3 | 0.62 | 0.19 | 0.47 | 0.08 | Shared |
| S2-wms_6424 | 0.6 | 110.94 | 0.08 | 0.55 | 0.65 | 0.1 | 0.81 | 0.3 | 0.4 | 8.93 | 13.43 | 0.49 | SHHC |
| S2-wms_245232 | 1.48 | 90.61 | 0.22 | 0.5 | 1.16 | 0.31 | 1.43 | 1.02 | 0.98 | 1.07 | 5.88 | 0.91 | SHHC |
| S2-wms_246296 | 12.95 | 51.5 | 0.26 | 0.61 | 1.37 | 0.27 | 1.56 | 2.87 | 0.99 | 1.01 | 22.91 | 0.67 | Shared |
| S2-wms_248315 | 0.79 | 76.59 | 0.09 | 0.55 | 0.77 | 0.24 | 8.8 | 0.33 | 1.14 | 0.31 | 0.71 | 0.41 | SHHC |
| S2-wms_248324 | 0.89 | 52.29 | 0.07 | 0.16 | 0.62 | 0.15 | 0.82 | 0.51 | 0.58 | 0.31 | 0.79 | 0.52 | SHHC |
| S2-wms_248327 | 1.22 | 15.32 | 0.09 | 0.12 | 0.82 | 0.25 | 0.89 | 1.49 | 0.85 | 0.43 | 1.23 | 0.97 | SHHC |
| S4-wms-combined_118531 | 1.48 | 1.63 | 0.94 | 69.21 | 1.47 | 0.33 | 1.37 | 1.05 | 1.38 | 0.85 | 2.01 | 1.06 | SHHC |
| S4-wms-combined_14411 | 2.9 | 24.2 | 0.53 | 29.22 | 0.6 | 3.21 | 0.78 | 0.41 | 0.42 | 0.23 | 0.72 | 0.37 | SHHC |
| S4-wms-combined_112093 | 10.65 | 4.99 | 0.83 | 20.89 | 4.4 | 14.9 | 1.3 | 7.66 | 1.42 | 9.81 | 4.41 | 1.16 | Shared |
| S4-wms-combined_42647 | 94.37 | 70.63 | 9.34 | 25.99 | 32.03 | 37.81 | 25.96 | 36.92 | 39.62 | 28.27 | 62.86 | 58.68 | Shared |
| S4-wms-combined_1246 | 95.52 | 21.33 | 9.38 | 15.15 | 72.49 | 3.87 | 17.88 | 37.49 | 9.37 | 13.47 | 31.55 | 11.4 | Shared |
| S4-wms-combined_22012 | 4.05 | 2.45 | 9.85 | 42.15 | 7.15 | 1.6 | 5.9 | 6 | 1.69 | 2.73 | 3.93 | 8.81 | SHHC |
| S4-wms-combined_57123 | 1.54 | 2.02 | 0.67 | 1125.01 | 8.22 | 0.47 | 3.84 | 6.29 | 0.99 | 2.77 | 3.74 | 1.06 | SHHC |
| S4-wms-combined_64632 | 1.66 | 2.34 | 0.19 | 17.5 | 1.99 | 0.47 | 5.18 | 0.85 | 3.11 | 0.66 | 1.34 | 1.8 | SHHC |
| S4-wms-combined_113721 | 52.2 | 29.2 | 5.59 | 16.48 | 5.16 | 2.12 | 19.17 | 22.45 | 1.63 | 57.5 | 29.79 | 47.69 | Shared |
| S4-wms-combined_57963 | 5.18 | 4.13 | 3.02 | 70.83 | 3.28 | 0.87 | 3.07 | 3.01 | 2.47 | 3.59 | 4.62 | 3.05 | SHHC |
| S4-wms-combined_10758 | 0.45 | 0.73 | 8.88 | 54.01 | 0.75 | 0.23 | 0.77 | 0.3 | 0.7 | 0.21 | 0.52 | 0.38 | SHHC |
| S4-wms-combined_17284 | 3.76 | 1.6 | 7.01 | 18.47 | 15.44 | 9.83 | 5.61 | 2.65 | 39.22 | 1.43 | 2.37 | 2.5 | Shared |
| S4-wms-combined_24659 | 0.48 | 0.86 | 0.36 | 23.02 | 0.62 | 0.18 | 1.11 | 0.73 | 0.62 | 0.66 | 0.75 | 0.4 | SHHC |

| | | | | | | | | | | | | | |
|---|---|---|---|---|---|---|---|---|---|---|---|---|---|
| S4-wms-combined_121978 | 10.95 | 12.58 | 0.65 | 32.94 | 3.96 | 1.61 | 10.1 | 6.16 | 18.81 | 3.67 | 5.04 | 9.71 | Shared |
| S4-wms-combined_25201 | 10.17 | 15.34 | 0.75 | 37.93 | 6.19 | 1.92 | 13.6 | 3.56 | 26.28 | 3.17 | 5.98 | 11.06 | Shared |
| S4-wms-combined_47565 | 0.77 | 0.72 | 0.07 | 49.77 | 0.99 | 0.28 | 0.88 | 0.89 | 0.53 | 0.24 | 0.75 | 0.43 | SHHC |
| S4-wms-combined_83492 | 2.26 | 1.04 | 0.24 | 22.42 | 0.96 | 0.21 | 0.92 | 0.92 | 0.56 | 0.67 | 1.44 | 319.19 | Shared |
| S4-wms-combined_41490 | 131.69 | 9.41 | 8.58 | 20.74 | 300.61 | 132.01 | 130.55 | 98.94 | 17.43 | 166.14 | 68.44 | 13.51 | Shared |
| S4-wms-combined_51079 | 10.89 | 2.68 | 10.24 | 17.17 | 10.68 | 5.57 | 15.02 | 3.53 | 14.54 | 2.6 | 3.12 | 6.31 | Shared |
| S4-wms-combined_143879 | 0.46 | 0.98 | 0.21 | 23.35 | 0.4 | 0.17 | 0.97 | 0.47 | 0.86 | 0.41 | 0.76 | 0.34 | SHHC |
| S4-wms_118690 | 0.46 | 0.86 | 0.05 | 40.73 | 0.53 | 0.15 | 0.76 | 0.23 | 1.21 | 0.14 | 0.65 | 0.44 | SHHC |
| S4-wms_161443 | 0.47 | 0.47 | 0.1 | 21.97 | 0.56 | 0.15 | 0.24 | 0.43 | 0.25 | 0.24 | 1 | 0.22 | SHHC |
| S4-wms_50475 | 4.98 | 6.79 | 0.62 | 589.25 | 5.46 | 1.04 | 7.71 | 22.18 | 2.29 | 2.34 | 3.68 | 5.52 | SHHC |
| S4-wms_141607 | 2.8 | 2.55 | 0.64 | 1303.88 | 7.37 | 0.78 | 3.52 | 3.69 | 1.95 | 2.73 | 4.6 | 1.62 | SHHC |
| S4-wms_107182 | 2.02 | 1.22 | 4.8 | 12.5 | 2.64 | 0.6 | 1.14 | 4.99 | 0.6 | 0.7 | 1.83 | 1.98 | SHHC |
| S4-wms_114120 | 0.45 | 0.69 | 0.03 | 21.49 | 0.9 | 0.16 | 0.81 | 0.38 | 0.64 | 0.26 | 0.78 | 0.51 | SHHC |
| S4-wms_45128 | 0.65 | 1.02 | 0.04 | 36.6 | 0.96 | 0.22 | 1.21 | 0.37 | 0.69 | 0.25 | 0.81 | 0.57 | SHHC |
| S4-wms_73990 | 0.57 | 2.05 | 0.05 | 42.34 | 0.51 | 0.13 | 0.94 | 2.54 | 1.02 | 0.16 | 0.61 | 0.43 | SHHC |
| S4-wms_62635 | 0.88 | 1.39 | 0.15 | 50.56 | 4.42 | 0.38 | 1.13 | 0.41 | 2.64 | 0.31 | 0.96 | 0.58 | SHHC |
| S6-wms-combined_117756 | 1.51 | 2.75 | 0.11 | 0.41 | 1.41 | 144.95 | 2 | 1.15 | 0.73 | 0.57 | 1.23 | 0.9 | SHHC |
| S6-wms-combined_41246 | 44.99 | 39.92 | 5.38 | 3.85 | 130 | 91.67 | 50.33 | 22.84 | 35.19 | 22.05 | 13.99 | 122.8 | Shared |
| S6-wms-combined_135223 | 16.07 | 40.42 | 0.61 | 20.74 | 114.07 | 57.53 | 350.63 | 5.24 | 1.83 | 9.21 | 84.71 | 4.37 | Shared |
| S6-wms-combined_88414 | 37.68 | 146.74 | 5.26 | 6.22 | 4.1 | 66.43 | 26.48 | 15.96 | 15.11 | 22.65 | 18.97 | 9.44 | Shared |
| S6-wms-combined_745 | 7.89 | 0.91 | 0.04 | 0.2 | 1.09 | 30.98 | 1.25 | 0.69 | 0.94 | 0.64 | 0.82 | 0.52 | SHHC |
| S6-wms-combined_82597 | 55.79 | 100.74 | 4.48 | 5.62 | 50.72 | 124.93 | 107.63 | 14.08 | 13.29 | 27.96 | 49.76 | 60.79 | Shared |
| S6-wms-combined_65225 | 0.76 | 0.89 | 0.04 | 0.48 | 0.5 | 18.71 | 0.61 | 0.55 | 0.75 | 0.24 | 0.65 | 0.54 | SHHC |
| S6-wms-combined_118427 | 37.08 | 43.04 | 5.85 | 6.7 | 50.49 | 91.87 | 33.72 | 30.54 | 64.47 | 33.07 | 24.78 | 150.71 | Shared |
| S6-wms-combined_118484 | 5.09 | 1.78 | 0.16 | 1.07 | 1.5 | 43.89 | 7.77 | 1.06 | 0.56 | 1.06 | 1.77 | 28.61 | Shared |
| S6-wms-combined_72228 | 94.37 | 31.26 | 2.09 | 1.62 | 216.35 | 240.44 | 74.68 | 58.57 | 75.05 | 87 | 105.2 | 25.65 | Shared |
| S6-wms-combined_119017 | 9.3 | 0.9 | 0.29 | 0.65 | 1.98 | 40.6 | 2.95 | 16.29 | 0.76 | 3.71 | 1.63 | 0.91 | SHHC |
| S6-wms-combined_119138 | 26.74 | 94.16 | 3.61 | 2.32 | 7.92 | 45.28 | 19.6 | 9.04 | 6.3 | 11.46 | 15.82 | 6.19 | Shared |
| S6-wms-combined_49909 | 6.6 | 4.69 | 1.53 | 2.21 | 1.83 | 68.54 | 1 | 2.59 | 0.87 | 0.92 | 10.38 | 7.54 | SHHC |
| S6-wms-combined_113986 | 15.71 | 5.12 | 0.08 | 0.52 | 1.04 | 29.56 | 1.37 | 0.6 | 0.67 | 0.26 | 11.4 | 0.69 | Shared |
| S6-wms-combined_8508 | 63.84 | 52.31 | 6.43 | 10.47 | 65.26 | 104.53 | 43.36 | 49.5 | 49.48 | 52.31 | 48.69 | 253.49 | Shared |
| S6-wms-combined_32530 | 10.8 | 55.81 | 1.14 | 3.05 | 2 | 29.11 | 5.94 | 2.7 | 5.24 | 10.35 | 8.22 | 5.12 | Shared |
| S6-wms-combined_126236 | 115.95 | 52.47 | 26 | 18.86 | 18.62 | 71.74 | 31.4 | 57.08 | 3.05 | 53.83 | 49.03 | 138.45 | Shared |
| S6-wms-combined_50721 | 234.99 | 17.33 | 11.54 | 14 | 407.73 | 181.98 | 253.18 | 165.94 | 33.61 | 219.07 | 119.19 | 24.82 | Shared |
| S6-wms-combined_138710 | 148.71 | 61.23 | 31 | 13.48 | 52.49 | 64.05 | 22.3 | 65.23 | 3.16 | 43.48 | 48.08 | 67.98 | Shared |
| S6-wms-combined_56464 | 68.38 | 29.64 | 10.82 | 21.13 | 7.02 | 73.72 | 11.03 | 38.27 | 2.05 | 12.47 | 30.28 | 116.35 | Shared |
| S6-wms-combined_110950 | 10.38 | 2.12 | 2.38 | 0.92 | 16.31 | 54.98 | 6.13 | 4.69 | 0.95 | 4.61 | 5.6 | 3.72 | Shared |
| S6-wms-combined_128972 | 2.99 | 3.67 | 0.93 | 0.58 | 13.9 | 85.02 | 9.79 | 2.38 | 1.05 | 2.58 | 2.44 | 19.32 | Shared |
| S6-wms-combined_140731 | 5.83 | 3.06 | 0.31 | 0.6 | 2.76 | 40.12 | 2.83 | 2.22 | 4.51 | 1.1 | 2.78 | 2.5 | SHHC |
| S6-wms-combined_91039 | 84.24 | 35.79 | 5.34 | 4.58 | 940.77 | 159.11 | 67.26 | 25.76 | 16.32 | 259.25 | 385.74 | 29.11 | Shared |
| S6-wms_54578 | 1.09 | 7.67 | 0.93 | 2.97 | 0.59 | 28.42 | 0.77 | 0.39 | 0.29 | 0.26 | 0.81 | 0.39 | SHHC |
| S6-wms_137591 | 1.41 | 1.45 | 0.23 | 0.51 | 2.66 | 28.06 | 1.81 | 1.15 | 1.97 | 0.48 | 1.02 | 0.98 | SHHC |
| S6-wms_132588 | 228.45 | 147.13 | 2.8 | 1.41 | 2.1 | 47.58 | 5.14 | 8.52 | 0.83 | 4.24 | 25.26 | 1.03 | Shared |
| S6-wms_138342 | 0.77 | 49.16 | 0.08 | 0.64 | 0.44 | 353.2 | 0.58 | 0.3 | 0.54 | 0.12 | 1.22 | 0.25 | SHHC |
| S6-wms_140743 | 0.59 | 0.79 | 0.04 | 0.62 | 0.66 | 40.59 | 0.91 | 0.3 | 0.62 | 0.22 | 0.59 | 0.33 | SHHC |
| S8-wms-combined_246389 | 6.51 | 1.37 | 0.15 | 0.55 | 1.51 | 0.55 | 1.11 | 283.01 | 0.77 | 1.03 | 1.75 | 1 | SHHC |
| S8-wms-combined_215102 | 151.81 | 2.73 | 57.84 | 0.66 | 4.16 | 1.17 | 46.93 | 328.33 | 1.35 | 4.3 | 145.1 | 2.79 | Shared |
| S8-wms-combined_276204 | 2.66 | 1.38 | 0.14 | 0.32 | 1.97 | 0.72 | 1.13 | 273.45 | 1.06 | 0.97 | 1.71 | 1.35 | SHHC |
| S8-wms-combined_276211 | 3.98 | 1 | 0.22 | 0.39 | 1.38 | 0.43 | 1.03 | 251.61 | 0.8 | 0.63 | 2.42 | 0.73 | SHHC |
| S8-wms-combined_23583 | 21.88 | 4.84 | 1.56 | 1.19 | 141.65 | 20.79 | 12.2 | 111.01 | 6.92 | 18.47 | 25.26 | 5.73 | Shared |
| S8-wms-combined_81597 | 0.85 | 1.08 | 12.35 | 11.76 | 2.34 | 0.56 | 6.44 | 47.26 | 1.02 | 277.32 | 32.91 | 135.84 | Shared |
| S8-wms-combined_219796 | 1.49 | 0.87 | 0.1 | 0.26 | 1.76 | 0.47 | 0.47 | 188.07 | 0.52 | 0.66 | 15.86 | 0.97 | SHHC |
| S8-wms-combined_185611 | 2.03 | 2.49 | 0.31 | 1 | 77.25 | 0.87 | 10.29 | 47.36 | 1.19 | 16.32 | 10.23 | 1.63 | Shared |
| S8-wms-combined_140059 | 1.58 | 1.13 | 0.11 | 0.29 | 1.44 | 0.56 | 6.25 | 57.05 | 0.51 | 0.5 | 3.34 | 0.66 | SHHC |
| S8-wms-combined_186054 | 23.52 | 10.16 | 3.37 | 2.64 | 40.29 | 5.88 | 19.5 | 537.53 | 15.41 | 24.64 | 48.92 | 8.28 | Shared |
| S8-wms-combined_117281 | 1.8 | 1.55 | 0.26 | 0.53 | 4 | 0.84 | 10.71 | 52.79 | 1.44 | 9.31 | 7.39 | 0.84 | Shared |
| S8-wms-combined_25607 | 2.36 | 1.91 | 0.22 | 0.46 | 2.8 | 1.22 | 2.14 | 18.45 | 0.87 | 2.38 | 2.53 | 9 | SHHC |
| S8-wms-combined_48046 | 7.38 | 3.48 | 0.79 | 0.72 | 8.31 | 1.72 | 3.92 | 68.56 | 1.84 | 3.93 | 10.5 | 3.11 | SHHC |
| S8-wms-combined_26087 | 69.52 | 2.44 | 6.74 | 4.79 | 111.74 | 35.26 | 86.19 | 119.84 | 5.04 | 109.8 | 22.98 | 3.61 | Shared |
| S8-wms-combined_164289 | 3.6 | 1.21 | 0.19 | 0.51 | 1.23 | 0.23 | 1.12 | 30.17 | 0.4 | 0.45 | 5.43 | 0.8 | SHHC |
| S8-wms-combined_72911 | 1.35 | 0.81 | 0.09 | 0.33 | 3.1 | 0.39 | 1.54 | 44.84 | 0.53 | 0.92 | 2.84 | 0.68 | SHHC |
| S8-wms-combined_49369 | 39.47 | 25.1 | 6.57 | 3.72 | 5.53 | 3.4 | 39.33 | 37.92 | 1.94 | 6.26 | 8.36 | 23.75 | Shared |
| S8-wms-combined_234473 | 9.42 | 19.61 | 2.7 | 2.18 | 30.35 | 34.84 | 17.16 | 15.71 | 29.4 | 8.46 | 6.66 | 6.72 | Shared |
| S8-wms-combined_211119 | 0.91 | 0.7 | 0.07 | 0.14 | 1.06 | 0.31 | 0.72 | 125.94 | 0.51 | 0.49 | 2.5 | 0.61 | SHHC |
| S8-wms-combined_74275 | 1.96 | 2.47 | 0.28 | 474.3 | 3.16 | 0.58 | 1.96 | 80.63 | 1.92 | 1.26 | 1.98 | 1.38 | SHHC |
| S8-wms-combined_270362 | 50.84 | 3.22 | 4.48 | 3.73 | 68.94 | 31.21 | 49.39 | 73.83 | 6.91 | 71.84 | 24.05 | 6.66 | Shared |

| | | | | | | | | | | | | | | |
|---|---|---|---|---|---|---|---|---|---|---|---|---|---|---|
| S8-wms-combined_200168 | 123.28 | 46.79 | 2.67 | 5.64 | 37.97 | 11.66 | 24.12 | 131.57 | 21.81 | 36.87 | 33.4 | 75.66 | Shared | |
| S8-wms-combined_97848 | 6.47 | 7.6 | 0.32 | 1.63 | 2.24 | 0.68 | 1.13 | 41.22 | 0.61 | 1.01 | 1.96 | 4.15 | SHHC | |
| S8-wms-combined_108917 | 4.85 | 4.06 | 0.31 | 0.8 | 9.05 | 0.78 | 2.94 | 169.84 | 1.71 | 3.09 | 10.19 | 1.39 | SHHC | |
| S8-wms-combined_40666 | 51.57 | 30.38 | 4.44 | 5.38 | 169.21 | 9.75 | 53.87 | 68.33 | 47.62 | 16.15 | 25.94 | 165.13 | Shared | |
| S8-wms-combined_225114 | 2.1 | 1.43 | 4.13 | 0.36 | 1.24 | 0.55 | 1.14 | 60.04 | 0.61 | 0.68 | 6.27 | 0.92 | SHHC | |
| S8-wms-combined_134098 | 6.67 | 1.13 | 0.5 | 0.31 | 3.19 | 0.62 | 1.14 | 20.78 | 0.88 | 1.3 | 3.47 | 1.28 | SHHC | |
| S8-wms-combined_180957 | 5.09 | 1.66 | 0.42 | 0.84 | 1.49 | 0.57 | 1.33 | 33.98 | 0.78 | 8.6 | 4.36 | 1.27 | SHHC | |
| S8-wms-combined_134284 | 2.1 | 1.18 | 0.18 | 0.3 | 1.89 | 0.87 | 1.29 | 22.46 | 0.84 | 2.01 | 1.9 | 1.75 | SHHC | |
| S8-wms-combined_226076 | 0.6 | 0.76 | 0.04 | 0.17 | 0.99 | 0.22 | 0.89 | 126.89 | 0.44 | 0.32 | 0.61 | 0.38 | SHHC | |
| S8-wms-combined_146562 | 1.38 | 1 | 0.06 | 0.16 | 1.76 | 0.39 | 0.69 | 173.21 | 0.67 | 0.62 | 5.09 | 0.58 | SHHC | |
| S8-wms-combined_169835 | 137.44 | 20.16 | 6.88 | 3.6 | 60.54 | 18.28 | 10.73 | 346.07 | 8.61 | 23.86 | 189.44 | 26.42 | Shared | |
| S8-wms-combined_239791 | 0.72 | 0.79 | 0.05 | 0.13 | 1.35 | 0.22 | 0.81 | 38.65 | 0.72 | 0.3 | 1.08 | 0.42 | SHHC | |
| S8-wms-combined_217866 | 1.15 | 1.12 | 2.8 | 8.36 | 1.5 | 12.71 | 2.86 | 42.65 | 0.87 | 0.34 | 0.88 | 0.8 | SHHC | |
| S8-wms-combined_68780 | 17.1 | 18.98 | 0.92 | 1.21 | 4.55 | 1.69 | 3.21 | 129 | 1.83 | 22.45 | 31.65 | 4.14 | Shared | |
| S8-wms-combined_161181 | 19.13 | 37.97 | 2.77 | 1.95 | 8.22 | 3.54 | 10.1 | 123.95 | 1.78 | 7.9 | 28.35 | 125.74 | Shared | |
| S8-wms-combined_264711 | 1.17 | 0.94 | 0.17 | 0.19 | 1.41 | 0.41 | 0.53 | 15.42 | 0.33 | 0.64 | 1.53 | 0.94 | SHHC | |
| S8-wms-combined_246185 | 12 | 39.24 | 0.12 | 1.28 | 40.88 | 0.35 | 9.35 | 149.72 | 0.38 | 3.12 | 9.95 | 0.7 | Shared | |
| S8-wms_270 | 1.92 | 6.21 | 0.07 | 0.22 | 0.79 | 0.09 | 0.29 | 40.62 | 0.39 | 0.36 | 1.11 | 0.36 | SHHC | |
| S8-wms_127191 | 0.72 | 0.86 | 0.05 | 0.36 | 0.77 | 0.14 | 0.62 | 59.49 | 0.39 | 0.26 | 0.88 | 0.32 | SHHC | |
| S8-wms_58515 | 53.11 | 37.25 | 10.68 | 7.63 | 344.14 | 39.09 | 35.92 | 97.72 | 25.88 | 163.58 | 227.58 | 94.09 | Shared | |
| S8-wms_82880 | 1.37 | 9.2 | 0.14 | 0.48 | 4.05 | 1.56 | 1.09 | 36.57 | 0.79 | 15.08 | 1.39 | 2.04 | Shared | |
| S8-wms_244525 | 0.55 | 0.94 | 0.03 | 0.21 | 0.43 | 0.13 | 0.79 | 28.75 | 0.55 | 0.16 | 0.66 | 0.3 | SHHC | |
| S8-wms_152831 | 9.66 | 1.33 | 3.87 | 7.71 | 1.48 | 0.41 | 1.39 | 96.76 | 0.72 | 0.9 | 17.71 | 2.56 | SHHC | |
| S8-wms_48935 | 42.68 | 8.39 | 2.94 | 2.08 | 11.69 | 6.28 | 3.88 | 290.5 | 2.56 | 7.38 | 26.77 | 9.65 | Shared | |
| S8-wms_190781 | 7.91 | 0.74 | 1.23 | 0.62 | 0.75 | 0.21 | 0.66 | 34.53 | 0.57 | 0.75 | 0.67 | 0.46 | SHHC | |
| S8-wms_30498 | 2.75 | 0.73 | 0.19 | 0.19 | 1.06 | 2.19 | 1.06 | 92.76 | 0.61 | 7.67 | 11.51 | 41.97 | Shared | |
| S8-wms_111056 | 0.84 | 4.77 | 0.06 | 0.48 | 0.56 | 0.19 | 0.79 | 31.96 | 0.4 | 0.14 | 0.45 | 0.28 | SHHC | |
| S8-wms_113693 | 2.75 | 1.2 | 1.67 | 0.38 | 1.71 | 0.51 | 1.25 | 16.71 | 0.34 | 0.62 | 15.97 | 0.66 | SHHC | |
| S8-wms_20528 | 1.81 | 1.56 | 0.24 | 0.75 | 2.76 | 0.56 | 1.38 | 33.9 | 0.75 | 1.1 | 18.87 | 1.27 | SHHC | |
| S8-wms_57121 | 0.7 | 0.75 | 0.05 | 0.25 | 0.98 | 0.2 | 0.93 | 93.54 | 0.64 | 0.33 | 3.49 | 0.55 | SHHC | |
| #Total | 45.75 | 52.2 | 3.29 | 10.86 | 61.41 | 23.04 | 49.76 | 31.39 | 16.74 | 28.29 | 42.1 | 32.49 | 33.11 | mean |
| | | | | | | | | | | | | | 17.7441 | SD |

Eukaryotic_DNA_virus_detection

VLP:

| qseqid | sseqid | matched_id | Virus_name | pident | qcovs | length | mismatch | gapopen | qstart | qend | sstart | send | evalue | bitscore |
|---|---|---|---|---|---|---|---|---|---|---|---|---|---|---|
| S12_vf_188827 | NC_026252.1 | NC_026252.1 | Human smacovirus 1 isolate France/12/2008/3454 | 96.555 | 100 | 2264 | 78 | 0 | 338 | 2601 | 2460 | 197 | 0 | 3749 |
| S13_vf_150407 | NC_026582.1 | NC_026582.1 | Uncultured phage WW-nAnB | 87.358 | 64 | 2025 | 241 | 15 | 1873 | 3890 | 4817 | 2801 | 0 | 2307 |
| S12_vf_188838 | NC_026582.1 | NC_026582.1 | Uncultured phage WW-nAnB | 86.645 | 42 | 1213 | 148 | 10 | 1 | 1204 | 3610 | 4817 | 0 | 1330 |
| S11_vs_49071 | NC_023639.1 | NC_023639.1 | Mimivirus terra2 genome | 89.286 | 0 | 56 | 6 | 0 | 55885 | 55940 | 646433 | 646488 | 1.59E-08 | 71.3 |
| S12_vs_22853 | NC_023639.1 | NC_023639.1 | Mimivirus terra2 genome | 89.286 | 0 | 56 | 4 | 2 | 12040 | 12094 | 5401 | 5455 | 1.97E-08 | 69.4 |
| S8_vf_94361 | NC_040329.1 | NC_040329.1 | Lynx canadensis associated microvirus CLP 9413 | 72.026 | 22 | 765 | 207 | 6 | 2154 | 2913 | 2979 | 3741 | 5.78E-55 | 220 |
| S5_vs_6478 | NC_000896.1 | NC_000896.1 | Lactobacillus prophage phiadh | 92.742 | 60 | 9893 | 652 | 54 | 19996 | 29855 | 40362 | 30503 | 0 | 14231 |
| S14_vf_78139 | NC_019456.1 | NC_019456.1 | Lactobacillus phage JCL1032 | 90.489 | 75 | 11218 | 951 | 67 | 39242 | 50424 | 12062 | 926 | 0 | 14698 |
| S14_vf_106156 | NC_005354.1 | NC_005354.1 | Lactobacillus prophage Lj928 | 84.623 | 16 | 3050 | 453 | 13 | 537 | 3577 | 32577 | 29535 | 0 | 3020 |
| S16_vf_1037 | NC_010821.1 | NC_010821.1 | Pseudomonas phage 201phi2-1 | 97.619 | 0 | 42 | 1 | 0 | 47295 | 47336 | 20456 | 20415 | 2.9E-09 | 73.1 |
| S4_vf_31978 | NC_010821.1 | NC_010821.1 | Pseudomonas phage 201phi2-1 | 97.619 | 0 | 42 | 1 | 0 | 11515 | 11556 | 20415 | 20456 | 2.79E-09 | 73.1 |
| S7_vf_29941 | NC_049348.1 | NC_049348.1 | Lactococcus lactis phage p272 | 92.736 | 89 | 9375 | 649 | 25 | 7233 | 16586 | 16145 | 6782 | 0 | 13511 |
| S3_vs_89485 | NC_005841.1 | NC_005841.1 | Enterobacteria phage ST104 DNA | 88.387 | 1 | 310 | 36 | 0 | 351 | 660 | 9856 | 10165 | 4.4E-100 | 374 |
| S12_vs_183752 | NC_021312.1 | NC_021312.1 | Phaeocystis globosa virus strain 16T | 95.122 | 0 | 41 | 0 | 2 | 6961 | 7000 | 52424 | 52463 | 4.28E-07 | 63.9 |
| S12_vs_81729 | NC_009816.1 | NC_009816.1 | Corynebacterium phage P1201 | 91.071 | 0 | 56 | 4 | 1 | 35099 | 35153 | 51086 | 51031 | 5.15E-10 | 75 |
| S5_vs_60507 | NC_008721.2 | NC_008721.2 | Streptococcus phage SMP | 75.15 | 2 | 668 | 156 | 7 | 13552 | 14214 | 36008 | 35346 | 1.22E-79 | 305 |
| S12_vs_111815 | NC_015287.1 | NC_015287.1 | Synechococcus phage S-SSM7 | 87.931 | 0 | 58 | 5 | 2 | 4212 | 4267 | 216504 | 216561 | 1.23E-07 | 67.6 |
| S8_vf_106946 | NC_011042.1 | NC_011042.1 | Enterobacteria phage EcoDS1 | 77.519 | 0 | 129 | 27 | 2 | 42117 | 42244 | 21953 | 21826 | 1.63E-10 | 76.8 |
| S9_vf_15012 | NC_012756.1 | NC_012756.1 | Streptococcus phage PH10 | 77.617 | 27 | 2971 | 639 | 25 | 26893 | 29851 | 18224 | 15268 | 0 | 1779 |
| S8_vf_163760 | NC_023694.1 | NC_023694.1 | Rhodococcus phage ReqiPoco6 | 70.718 | 2 | 1547 | 417 | 34 | 43456 | 44984 | 15921 | 17449 | 4.72E-82 | 315 |
| S9_vs_90859 | NC_015254.1 | NC_015254.1 | Brochothrix phage BL3 | 82.418 | 0 | 91 | 14 | 2 | 25224 | 25313 | 28922 | 29011 | 4.69E-11 | 78.7 |
| S13_vf_4901 | NC_014636.1 | NC_014636.1 | Aeromonas phage phiAS5 | 87.5 | 0 | 80 | 9 | 1 | 94353 | 94431 | 191395 | 191316 | 1.31E-14 | 91.6 |
| S12_vf_169796 | NC_014636.1 | NC_014636.1 | Aeromonas phage phiAS5 | 87.5 | 0 | 80 | 9 | 1 | 46382 | 46460 | 191316 | 191395 | 1.78E-14 | 91.6 |
| S8_vf_208110 | NC_014636.1 | NC_014636.1 | Aeromonas phage phiAS5 | 84.81 | 0 | 79 | 11 | 1 | 111194 | 111271 | 191395 | 191317 | 1.46E-10 | 78.7 |
| S9_vs_90851 | NC_049345.1 | NC_049345.1 | Roseovarius Plymouth Podovirus 1 partial sequence | 85.714 | 0 | 70 | 7 | 3 | 4559 | 4627 | 71255 | 71322 | 8.16E-09 | 71.3 |
| S11_vf_28515 | NC_028999.1 | NC_028999.1 | Pseudomonas phage PhiPA3 | 85.135 | 0 | 74 | 9 | 2 | 70940 | 71012 | 74690 | 74762 | 1.36E-09 | 75 |
| S5_vs_129970 | NC_019929.1 | NC_019929.1 | Erwinia phage phiEaH2 | 95.122 | 0 | 41 | 2 | 0 | 31535 | 31575 | 167072 | 167032 | 2.69E-07 | 65.8 |
| S10_vs_14557 | NC_019929.1 | NC_019929.1 | Erwinia phage phiEaH2 | 95.122 | 0 | 41 | 2 | 0 | 71918 | 71958 | 167072 | 167032 | 7.75E-07 | 65.8 |
| S13_vs_113593 | NC_016770.1 | NC_016770.1 | Bacteroides phage B124-14 | 75.691 | 0 | 181 | 44 | 0 | 43632 | 43812 | 3904 | 3724 | 7.65E-15 | 91.6 |
| S5_vs_128398 | NC_020201.1 | NC_020201.1 | Pectobacterium phage phiTE | 88.235 | 0 | 51 | 6 | 0 | 15480 | 15530 | 79876 | 79926 | 2.81E-06 | 62.1 |
| S5_vf_74661 | NC_019445.1 | NC_019445.1 | Escherichia phage TL-2011b | 98.221 | 55 | 9838 | 169 | 5 | 10579 | 20413 | 24046 | 14212 | 0 | 17193 |
| S13_vs_45488 | NC_023717.1 | NC_023717.1 | Cronobacter phage CR9 | 91.837 | 0 | 49 | 4 | 0 | 117881 | 117929 | 148189 | 148141 | 7.96E-08 | 69.4 |
| S17_vf_177199 | NC_021775.1 | NC_021775.1 | Salmonella phage FSL SP-031 | 77.761 | 18 | 2001 | 415 | 25 | 11297 | 13282 | 11416 | 13401 | 0 | 1203 |
| S3_vs_98675 | NC_017978.1 | NC_017978.1 | Clostridium phage PhiS63 | 76.077 | 4 | 1555 | 315 | 51 | 20400 | 21923 | 557 | 2085 | 0 | 758 |
| S9_vs_50439 | NC_020082.1 | NC_020082.1 | Edwardsiella phage MSW-3 DNA | 89.007 | 1 | 282 | 31 | 0 | 37198 | 37479 | 40452 | 40733 | 1.15E-92 | 350 |
| S16_vf_32359 | NC_020082.1 | NC_020082.1 | Edwardsiella phage MSW-3 DNA | 89.818 | 1 | 275 | 28 | 0 | 6951 | 7225 | 40726 | 40452 | 4.43E-94 | 353 |
| S3_vf_96222 | NC_049351.1 | NC_049351.1 | Lactococcus phage CaseusJM1 | 95.072 | 79 | 4667 | 192 | 24 | 7566 | 12224 | 17615 | 22251 | 0 | 7310 |
| S10_vs_106834 | NC_021342.2 | NC_021342.2 | Edwardsiella phage PEi21 | 80.851 | 1 | 188 | 36 | 0 | 5 | 192 | 41201 | 41014 | 2.47E-32 | 148 |
| S14_vf_85122 | NC_021792.1 | NC_021792.1 | Cellulophaga phage phi46:3 | 75.602 | 1 | 332 | 75 | 6 | 28594 | 28922 | 5079 | 5407 | 1.22E-35 | 159 |
| S10_vs_2538 | NC_022762.1 | NC_022762.1 | Lactobacillus phage phiLdb | 73.256 | 1 | 172 | 44 | 2 | 1455 | 1625 | 3136 | 3306 | 3.45E-06 | 62.1 |
| S7_vf_60968 | NC_024361.1 | NC_024361.1 | Streptococcus phage DCC1738 | 88.679 | 0 | 53 | 6 | 0 | 33233 | 33285 | 31324 | 31272 | 3.46E-07 | 65.8 |
| S3_vf_35073 | NC_024214.1 | NC_024214.1 | Lactococcus phage P162 | 78.846 | 0 | 104 | 18 | 3 | 1154 | 1255 | 54182 | 54081 | 1.37E-07 | 67.6 |
| S11_vs_83666 | NC_025453.1 | NC_025453.1 | Enterococcus phage EFC-1 | 77.122 | 1 | 271 | 58 | 4 | 20286 | 20554 | 5519 | 5251 | 4.17E-34 | 154 |
| S7_vf_63894 | NC_027358.1 | NC_027358.1 | Leuconostoc phage Ln-9 | 93.258 | 78 | 10026 | 650 | 21 | 14091 | 24098 | 15155 | 5138 | 0 | 14746 |
| S8_vf_129423 | NC_027358.1 | NC_027358.1 | Leuconostoc phage Ln-9 | 91.15 | 87 | 10813 | 918 | 31 | 2032 | 12825 | 2502 | 13294 | 0 | 14630 |
| S7_vs_16344 | NC_027366.1 | NC_027366.1 | Bacillus phage Mater | 91.228 | 0 | 57 | 5 | 0 | 59364 | 59420 | 132697 | 132641 | 1.83E-10 | 78.7 |
| S12_vs_75098 | NC_027366.1 | NC_027366.1 | Bacillus phage Mater | 91.228 | 0 | 57 | 5 | 0 | 36919 | 36975 | 132641 | 132697 | 4.4E-11 | 78.7 |
| S14_vs_56251 | NC_028958.1 | NC_028958.1 | Clostridium phage phiCD146 | 75.192 | 1 | 391 | 87 | 8 | 18405 | 18790 | 1195 | 810 | 1.14E-40 | 176 |
| S9_vs_62803 | NC_028767.1 | NC_028767.1 | Paenibacillus phage Vegas | 70.82 | 3 | 1244 | 282 | 72 | 13277 | 14481 | 36682 | 37883 | 3.58E-51 | 211 |
| S11_vs_13652 | NC_028886.1 | NC_028886.1 | Bacillus phage phi4B1 | 89.583 | 0 | 48 | 5 | 0 | 1689 | 1736 | 100 | 147 | 2.47E-06 | 62.1 |
| S13_vs_27462 | NC_028969.1 | NC_028969.1 | Brevibacillus phage Osiris | 89.474 | 0 | 57 | 6 | 0 | 49018 | 49074 | 6446 | 6502 | 3.22E-09 | 73.1 |
| S14_vf_117157 | NC_031907.1 | NC_031907.1 | Streptococcus phage phiARI0746 | 83.497 | 1 | 509 | 84 | 0 | 43488 | 43996 | 23943 | 24451 | 1.9E-130 | 475 |
| S9_vs_90850 | NC_028783.1 | NC_028783.1 | Lactobacillus phage iLp84 | 92.453 | 0 | 53 | 4 | 0 | 29021 | 29073 | 32392 | 32444 | 1.48E-10 | 76.8 |
| S17_vs_31654 | NC_042140.1 | NC_042140.1 | Bacillus phage Bm15 | 94.737 | 0 | 38 | 2 | 0 | 3136 | 3173 | 62614 | 62577 | 9.6E-06 | 60.2 |
| S16_vf_84462 | NC_047760.1 | NC_047760.1 | Lactobacillus phage SA-C12 | 76.19 | 0 | 231 | 55 | 0 | 33277 | 33507 | 68953 | 69183 | 2.63E-24 | 122 |
| S14_vs_4887 | NC_041974.1 | NC_041974.1 | Erwinia phage vB_EamM_Simmy50 | 97.561 | 0 | 41 | 1 | 0 | 959 | 999 | 254871 | 254911 | 7.21E-09 | 71.3 |
| S14_vs_2870 | NC_030917.1 | NC_030917.1 | Gordonia phage OneUp | 85.294 | 0 | 68 | 9 | 1 | 23157 | 23224 | 35483 | 35549 | 5.65E-08 | 69.4 |
| S11_vf_10907 | NC_048640.1 | NC_048640.1 | Bacillus phage vB_BtS_BMBtp14 | 86.885 | 0 | 61 | 8 | 0 | 32849 | 32909 | 28087 | 28147 | 2.41E-08 | 69.4 |
| S12_vs_133070 | NC_047767.1 | NC_047767.1 | Staphylococcus phage vB_SscM-1 | 84.906 | 0 | 106 | 10 | 6 | 90624 | 90728 | 60235 | 60135 | 8.34E-18 | 102 |
| S6_vs_1146 | NC_031040.1 | NC_031040.1 | Lactococcus phage 50101 | 79.167 | 0 | 120 | 25 | 0 | 11065 | 11184 | 22728 | 22609 | 1.23E-12 | 84.2 |
| S9_vs_58086 | NC_031017.1 | NC_031017.1 | Lactococcus phage 63301 | 81.421 | 1 | 183 | 28 | 6 | 2697 | 2876 | 23218 | 23397 | 2.82E-31 | 145 |
| S5_vf_25423 | NC_049374.1 | NC_049374.1 | Lactococcus phage 10W18 | 94.97 | 72 | 6720 | 334 | 4 | 747 | 7464 | 1209 | 7926 | 0 | 10536 |
| S5_vf_103206 | NC_031029.1 | NC_031029.1 | Gordonia phage Cucurbita | 97.222 | 0 | 36 | 1 | 0 | 18140 | 18175 | 6354 | 6319 | 1.99E-06 | 62.1 |
| S13_vf_151755 | NC_047797.2 | NC_047797.2 | Pectobacterium phage PP81 | 83.871 | 0 | 124 | 16 | 4 | 20350 | 20471 | 16936 | 17057 | 3.51E-22 | 115 |
| S8_vs_208103 | NC_034383.1 | NC_034383.1 | Pacmanvirus A23 genome assembly | 80 | 0 | 120 | 18 | 6 | 90927 | 91043 | 46851 | 46967 | 2.28E-12 | 84.2 |
| S12_vf_18902 | NC_062582.1 | NC_062582.1 | Pseudomonas phage Phabio | 94.737 | 0 | 57 | 3 | 0 | 108637 | 108693 | 296423 | 296479 | 5.75E-14 | 89.8 |
| S1_vs_23682 | NC_042013.1 | NC_042013.1 | Agrobacterium phage Atu_ph07 | 88.462 | 0 | 52 | 4 | 2 | 17883 | 17933 | 337993 | 337943 | 4.76E-06 | 62.1 |
| S7_vf_72493 | NC_042013.1 | NC_042013.1 | Agrobacterium phage Atu_ph07 | 85.714 | 0 | 63 | 8 | 1 | 10753 | 10815 | 222959 | 222898 | 1.9E-07 | 65.8 |
| S12_vs_92868 | NC_042016.1 | NC_042016.1 | Streptomyces phage DrGrey | 74.28 | 1 | 486 | 105 | 17 | 10722 | 11197 | 11945 | 12420 | 1.44E-43 | 187 |
| S4_vs_VIRSorter_30222-cat_2 | NC_042016.1 | NC_042016.1 | Streptomyces phage DrGrey | 75.186 | 0 | 403 | 84 | 12 | 11208 | 11602 | 11945 | 12339 | 3.2E-40 | 176 |
| S6_vf_3435 | NC_054997.1 | NC_054997.1 | Lactococcus phage 62403 | 88.876 | 97 | 9277 | 957 | 69 | 2881 | 12119 | 5264 | 14503 | 0 | 11345 |
| S5_vf_55687 | NC_054998.1 | NC_054998.1 | Lactococcus phage 62606 | 91.844 | 99 | 12910 | 1008 | 36 | 1529 | 14413 | 20214 | 7325 | 0 | 17965 |
| S7_vf_68239 | NC_049424.1 | NC_049424.1 | Lactococcus phage LP1502a | 77.022 | 7 | 1719 | 357 | 36 | 12021 | 13719 | 14122 | 15822 | 0 | 952 |
| S16_vf_33218 | NC_049398.1 | NC_049398.1 | Lactococcus phage 16802 | 93.084 | 75 | 7432 | 487 | 22 | 4627 | 12043 | 12754 | 20173 | 0 | 10852 |
| S8_vf_45776 | NC_049398.1 | NC_049398.1 | Lactococcus phage 16802 | 93.652 | 75 | 8129 | 492 | 9 | 17951 | 26057 | 8127 | 1 | 0 | 12131 |
| S16_vf_16182 | NC_042042.1 | NC_042042.1 | Streptomyces phage Scap1 | 78.049 | 0 | 164 | 34 | 2 | 27065 | 27227 | 7027 | 6865 | 2.09E-18 | 102 |
| S14_vs_117165 | NC_047906.1 | NC_047906.1 | Streptomyces phage Rowa | 95.349 | 0 | 43 | 0 | 2 | 3296 | 3336 | 41905 | 41863 | 9.25E-08 | 67.6 |
| S7_vs_94738 | NC_047906.1 | NC_047906.1 | Streptomyces phage Rowa | 95.349 | 0 | 43 | 0 | 2 | 30381 | 30421 | 41905 | 41863 | 9.16E-08 | 67.6 |
| S7_vs_87439 | NC_047906.1 | NC_047906.1 | Streptomyces phage Rowa | 95.349 | 0 | 43 | 0 | 2 | 9206 | 9246 | 41905 | 41863 | 5.49E-08 | 67.6 |
| S13_vf_54201 | NC_047909.1 | NC_047909.1 | Faecalibacterium phage FP_Brigit | 72.007 | 3 | 1211 | 288 | 48 | 1197 | 2381 | 19642 | 20827 | 3.6E-81 | 311 |
| S16_vs_76823 | NC_047909.1 | NC_047909.1 | Faecalibacterium phage FP_Brigit | 75.676 | 1 | 333 | 81 | 0 | 20139 | 20471 | 26131 | 26463 | 1.2E-37 | 167 |
| S9_vs_27813 | NC_047910.1 | NC_047910.1 | Faecalibacterium phage FP_Epona | 71.342 | 2 | 827 | 207 | 21 | 1515 | 2320 | 21147 | 21964 | 7.35E-44 | 187 |
| S11_vs_23357 | NC_047910.1 | NC_047910.1 | Faecalibacterium phage FP_Epona | 71.826 | 2 | 827 | 203 | 21 | 41071 | 41876 | 21964 | 21147 | 1.57E-50 | 209 |
| S11_vs_10172 | NC_047910.1 | NC_047910.1 | Faecalibacterium phage FP_Epona | 69.836 | 3 | 1220 | 305 | 47 | 1107 | 2290 | 20748 | 21940 | 2.07E-34 | 156 |
| S3_vs_54303 | NC_047910.1 | NC_047910.1 | Faecalibacterium phage FP_Epona | 69.952 | 3 | 1238 | 305 | 51 | 1107 | 2306 | 20748 | 21956 | 1.25E-36 | 163 |
| S5_vs_38949 | NC_047910.1 | NC_047910.1 | Faecalibacterium phage FP_Epona | 71.792 | 2 | 826 | 205 | 22 | 21952 | 22757 | 21147 | 21964 | 1.59E-50 | 209 |
| S7_vf_5439 | NC_047910.1 | NC_047910.1 | Faecalibacterium phage FP_Epona | 74.027 | 7 | 1336 | 303 | 35 | 22329 | 23639 | 20666 | 21982 | 2.1E-139 | 505 |
| S7_vs_4956 | NC_047910.1 | NC_047910.1 | Faecalibacterium phage FP_Epona | 71.235 | 2 | 737 | 179 | 24 | 31016 | 31737 | 21466 | 20748 | 2.12E-34 | 156 |
| S16_vf_97279 | NC_047910.1 | NC_047910.1 | Faecalibacterium phage FP_Epona | 77.843 | 7 | 1530 | 309 | 29 | 42497 | 44011 | 10973 | 12487 | 0 | 920 |
| S4_vs_VIRSorter_7386-circular-cat_2 | NC_047910.1 | NC_047910.1 | Faecalibacterium phage FP_Epona | 72.606 | 2 | 752 | 176 | 22 | 18636 | 19366 | 21222 | 21964 | 7.44E-54 | 220 |
| S8_vf_151384 | NC_047910.1 | NC_047910.1 | Faecalibacterium phage FP_Epona | 75.737 | 2 | 882 | 194 | 18 | 43521 | 44392 | 11477 | 12348 | 1.7E-115 | 425 |
| S8_vs_47265 | NC_047910.1 | NC_047910.1 | Faecalibacterium phage FP_Epona | 71.252 | 1 | 807 | 212 | 20 | 40708 | 41506 | 26460 | 25666 | 1.3E-43 | 187 |
| S9_vf_64844 | NC_047911.1 | NC_047911.1 | Faecalibacterium phage FP_Lagaffe | 75.382 | 2 | 2161 | 480 | 41 | 48568 | 50699 | 323 | 2460 | 0 | 996 |

WMS:

| qseqid | sseqid | matched_id | Virus_name | pident | qcovs | length | mismatch | gapopen | qstart | qend | sstart | send | evalue | bitscore |
|---|---|---|---|---|---|---|---|---|---|---|---|---|---|---|
| S2-wms_130687 | NC_015274.1 | NC_015274.1 | Streptococcus phage Dp-1 | 87.037 | 0 | 54 | 7 | 0 | 4629 | 4682 | 10555 | 10502 | 7.5E-06 | 62.1 |
| S5-wms-combined_18356 | NC_003356.1 | NC_003356.1 | Enterobacteria phage phiP27 | 93.04 | 5 | 704 | 49 | 0 | 13922 | 14625 | 39416 | 40119 | 0 | 1029 |
| S9-wms-combined_109322 | NC_006557.1 | NC_006557.1 | Bacillus phage BCJA1c | 72.919 | 1 | 997 | 235 | 32 | 13129 | 14108 | 13185 | 14163 | 2.77E-81 | 313 |
| S8-wms_127191 | NC_023694.1 | NC_023694.1 | Rhodococcus phage ReqiPoco6 | 70.718 | 3 | 1547 | 417 | 34 | 2024 | 3552 | 15921 | 17449 | 4.46E-82 | 315 |
| S5-wms-combined_207291 | NC_019929.1 | NC_019929.1 | Erwinia phage phiEaH2 | 95.122 | 0 | 41 | 2 | 0 | 13351 | 13391 | 167032 | 167072 | 4.77E-07 | 65.8 |
| S17-wms-combined_95360 | NC_016761.1 | NC_016761.1 | Salmonella phage SPN1S | 92.857 | 0 | 56 | 4 | 0 | 66667 | 66722 | 2882 | 2827 | 1.05E-11 | 82.4 |
| S9-wms-combined_93232 | NC_019720.1 | NC_019720.1 | Enterobacterial phage mEp213 | 76.683 | 13 | 6493 | 1417 | 84 | 28026 | 34478 | 1 | 6436 | 0 | 3513 |
| S17-wms_266511 | NC_021775.1 | NC_021775.1 | Salmonella phage FSL SP-031 | 77.761 | 18 | 2001 | 415 | 25 | 22538 | 24523 | 13401 | 11416 | 0 | 1203 |
| S11-wms-combined_215723 | NC_034248.1 | NC_034248.1 | Rhizobium phage RHEph10 | 76.712 | 0 | 146 | 28 | 5 | 49472 | 49614 | 61871 | 61729 | 2.48E-10 | 76.8 |
| S9-wms-combined_64277 | NC_049941.1 | NC_049941.1 | Stx2-converting phage Stx2a_WGPS2 proviral DNA | 94.052 | 0 | 269 | 16 | 0 | 94870 | 95138 | 52769 | 52501 | 3.2E-110 | 409 |
| S1-wms-combined_85225 | NC_021311.1 | NC_021311.1 | Mycobacterium phage Phaux | 89.474 | 0 | 76 | 8 | 0 | 5770 | 5845 | 42329 | 42404 | 1.98E-16 | 97.1 |
| S17-wms-combined_130215 | NC_023584.1 | NC_023584.1 | Synechococcus phage S-MbCM100 | 93.22 | 0 | 59 | 4 | 0 | 26227 | 26285 | 154576 | 154518 | 5.32E-14 | 87.9 |
| S4-wms-combined_57963 | NC_029046.1 | NC_029046.1 | Sinorhizobium phage phiLM21 | 91.667 | 0 | 48 | 3 | 1 | 60866 | 60913 | 50 | 4 | 1.33E-06 | 65.8 |
| S12-wms-combined_385270 | NC_027366.1 | NC_027366.1 | Bacillus phage Mater | 91.228 | 0 | 57 | 5 | 0 | 4492 | 4548 | 132697 | 132641 | 4.46E-11 | 78.7 |
| S2-wms-combined_159670 | NC_028914.1 | NC_028914.1 | Mycobacterium phage Sheen | 97.297 | 0 | 37 | 1 | 0 | 31720 | 31756 | 4386 | 4422 | 1.09E-06 | 63.9 |
| S9-wms-combined_105822 | NC_028767.1 | NC_028767.1 | Paenibacillus phage Vegas | 69.486 | 3 | 1324 | 361 | 37 | 32732 | 34035 | 37646 | 36346 | 9.48E-38 | 167 |
| S6-wms-combined_49909 | NC_031040.1 | NC_031040.1 | Lactococcus phage 50101 | 79.167 | 0 | 120 | 25 | 0 | 21424 | 21543 | 22609 | 22728 | 6.43E-13 | 84.2 |
| S11-wms-combined_112733 | NC_031059.1 | NC_031059.1 | Rhodovulum phage vB_RhkS_P1 | 74.569 | 2 | 696 | 139 | 30 | 13702 | 14380 | 15331 | 16005 | 5.66E-69 | 270 |
| S5-wms_10783 | NC_031029.1 | NC_031029.1 | Gordonia phage Cucurbita | 97.222 | 0 | 36 | 1 | 0 | 18140 | 18175 | 6354 | 6319 | 2.02E-06 | 62.1 |
| S9-wms-combined_37587 | NC_041997.1 | NC_041997.1 | Acidovorax phage ACP17 | 92.157 | 0 | 51 | 3 | 1 | 46760 | 46810 | 50 | 1 | 1.46E-08 | 71.3 |
| S7-wms-combined_135154 | NC_041997.1 | NC_041997.1 | Acidovorax phage ACP17 | 92.157 | 0 | 51 | 3 | 1 | 28407 | 28457 | 50 | 1 | 8.9E-09 | 71.3 |
| S12-wms-combined_2450 | NC_041997.1 | NC_041997.1 | Acidovorax phage ACP17 | 92.157 | 0 | 51 | 3 | 1 | 20484 | 20534 | 50 | 1 | 1.46E-08 | 71.3 |
| S2-wms-combined_229529 | NC_041997.1 | NC_041997.1 | Acidovorax phage ACP17 | 92.157 | 0 | 51 | 3 | 1 | 87980 | 88030 | 50 | 1 | 2.01E-08 | 71.3 |
| S2-wms-combined_143737 | NC_041997.1 | NC_041997.1 | Acidovorax phage ACP17 | 92.157 | 0 | 51 | 3 | 1 | 777 | 827 | 1 | 50 | 9.38E-09 | 71.3 |
| S1-wms-combined_131270 | NC_042013.1 | NC_042013.1 | Agrobacterium phage Atu_ph07 | 88.235 | 0 | 51 | 6 | 0 | 10231 | 10281 | 337993 | 337943 | 4.17E-06 | 62.1 |
| S2-wms-combined_62519 | NC_047861.1 | NC_047861.1 | Bordetella phage vB_BbrM_PHB04 | 88.462 | 0 | 78 | 9 | 0 | 50353 | 50430 | 45195 | 45118 | 8.99E-16 | 95.3 |
| S12-wms-combined_151843 | NC_048694.1 | NC_048694.1 | Klebsiella phage vB_Kpn_F48 | 96.154 | 0 | 52 | 2 | 0 | 55811 | 55862 | 168343 | 168394 | 8.17E-13 | 86.1 |
| S9-wms-combined_29611 | NC_047909.1 | NC_047909.1 | Faecalibacterium phage FP_Brigit | 78.738 | 0 | 301 | 54 | 8 | 365245 | 365540 | 37415 | 37120 | 1.51E-44 | 193 |
| S11-wms_55610 | NC_047909.1 | NC_047909.1 | Faecalibacterium phage FP_Brigit | 77.073 | 7 | 2316 | 486 | 38 | 14150 | 16435 | 14157 | 16457 | 0 | 1293 |
| S2-wms_107329 | NC_047909.1 | NC_047909.1 | Faecalibacterium phage FP_Brigit | 76.827 | 7 | 2313 | 497 | 32 | 22852 | 25137 | 14157 | 16457 | 0 | 1266 |
| S8-wms_82880 | NC_047909.1 | NC_047909.1 | Faecalibacterium phage FP_Brigit | 75.274 | 1 | 457 | 97 | 15 | 9106 | 9554 | 37568 | 37120 | 8.04E-49 | 204 |
| S11-wms-combined_264050 | NC_047910.1 | NC_047910.1 | Faecalibacterium phage FP_Epona | 75.107 | 1 | 932 | 210 | 20 | 37803 | 38723 | 11477 | 12397 | 1.9E-112 | 416 |
| S7-wms_159432 | NC_047910.1 | NC_047910.1 | Faecalibacterium phage FP_Epona | 74.027 | 6 | 1336 | 303 | 35 | 9504 | 10814 | 21982 | 20666 | 1.4E-139 | 505 |
| S17-wms-combined_127688 | NC_047910.1 | NC_047910.1 | Faecalibacterium phage FP_Epona | 71.138 | 3 | 1573 | 379 | 51 | 37039 | 38578 | 39629 | 41159 | 7.36E-84 | 320 |
| S12-wms-combined_18287 | NC_047910.1 | NC_047910.1 | Faecalibacterium phage FP_Epona | 70.008 | 1 | 1237 | 306 | 47 | 48850 | 50049 | 20748 | 21956 | 6.03E-38 | 169 |
| S12-wms-combined_129799 | NC_047910.1 | NC_047910.1 | Faecalibacterium phage FP_Epona | 77.778 | 5 | 1530 | 310 | 29 | 33925 | 35439 | 10973 | 12487 | 0 | 915 |
| S6-wms-combined_8508 | NC_047910.1 | NC_047910.1 | Faecalibacterium phage FP_Epona | 71.792 | 1 | 826 | 205 | 22 | 14940 | 15745 | 21147 | 21964 | 3.15E-50 | 209 |
| S8-wms-combined_217866 | NC_047910.1 | NC_047910.1 | Faecalibacterium phage FP_Epona | 71.252 | 2 | 807 | 212 | 20 | 22334 | 23132 | 26460 | 25666 | 8.51E-44 | 187 |
| S3-wms_22744 | NC_047911.1 | NC_047911.1 | Faecalibacterium phage FP_Lagaffe | 75.812 | 4 | 2187 | 476 | 43 | 45321 | 47476 | 2481 | 317 | 0 | 1059 |
| S5-wms-combined_134381 | NC_047911.1 | NC_047911.1 | Faecalibacterium phage FP_Lagaffe | 75.497 | 3 | 2163 | 474 | 45 | 49300 | 51431 | 323 | 2460 | 0 | 1007 |
| S7-wms_118607 | NC_047911.1 | NC_047911.1 | Faecalibacterium phage FP_Lagaffe | 97.95 | 99 | 30440 | 614 | 10 | 17901 | 48335 | 1 | 30435 | 0 | 52746 |
| S17-wms-combined_248321 | NC_047911.1 | NC_047911.1 | Faecalibacterium phage FP_Lagaffe | 75.671 | 2 | 2162 | 472 | 43 | 108424 | 110555 | 2460 | 323 | 0 | 1029 |
| S12-wms-combined_259783 | NC_047911.1 | NC_047911.1 | Faecalibacterium phage FP_Lagaffe | 80.473 | 3 | 804 | 153 | 4 | 7175 | 7976 | 42445 | 43246 | 1.1E-171 | 612 |
| S2-wms-combined_207832 | NC_047911.1 | NC_047911.1 | Faecalibacterium phage FP_Lagaffe | 75.903 | 5 | 2187 | 474 | 43 | 3784 | 5939 | 2481 | 317 | 0 | 1070 |
| S11-wms-combined_14636 | NC_047912.1 | NC_047912.1 | Faecalibacterium phage FP_Lugh | 72.508 | 7 | 1495 | 353 | 52 | 1947 | 3412 | 9499 | 10964 | 2.5E-117 | 431 |
| S11-wms-combined_140009 | NC_047912.1 | NC_047912.1 | Faecalibacterium phage FP_Lugh | 93.015 | 48 | 9349 | 560 | 70 | 24305 | 33614 | 31056 | 21762 | 0 | 13561 |
| S1-wms-combined_164894 | NC_047912.1 | NC_047912.1 | Faecalibacterium phage FP_Lugh | 90.173 | 3 | 519 | 51 | 0 | 5867 | 6385 | 5648 | 6166 | 0 | 676 |
| S1-wms-combined_166392 | NC_047912.1 | NC_047912.1 | Faecalibacterium phage FP_Lugh | 73.153 | 4 | 704 | 158 | 23 | 4671 | 5357 | 18063 | 18752 | 2.49E-55 | 224 |
| S1-wms-combined_23248 | NC_047912.1 | NC_047912.1 | Faecalibacterium phage FP_Lugh | 97.254 | 24 | 8557 | 229 | 3 | 34153 | 42706 | 5417 | 13970 | 0 | 14495 |
| S1-wms-combined_227917 | NC_047912.1 | NC_047912.1 | Faecalibacterium phage FP_Lugh | 78.674 | 34 | 3845 | 744 | 66 | 2020 | 5825 | 13238 | 9431 | 0 | 2488 |
| S3-wms-combined_45305 | NC_047912.1 | NC_047912.1 | Faecalibacterium phage FP_Lugh | 78.358 | 37 | 4542 | 829 | 113 | 9190 | 13625 | 13924 | 9431 | 0 | 2800 |
| S17-wms-combined_90378 | NC_047912.1 | NC_047912.1 | Faecalibacterium phage FP_Lugh | 83.794 | 28 | 6047 | 870 | 81 | 22782 | 28790 | 13268 | 7294 | 0 | 5637 |
| S12-wms-combined_71235 | NC_047912.1 | NC_047912.1 | Faecalibacterium phage FP_Lugh | 88.436 | 72 | 9037 | 914 | 112 | 3321 | 12283 | 30741 | 21762 | 0 | 10778 |
| S12-wms-combined_345948 | NC_047912.1 | NC_047912.1 | Faecalibacterium phage FP_Lugh | 81.413 | 19 | 1189 | 209 | 11 | 17602 | 18784 | 17706 | 18888 | 0 | 961 |
| S2-wms-combined_127080 | NC_047912.1 | NC_047912.1 | Faecalibacterium phage FP_Lugh | 78.481 | 2 | 553 | 106 | 12 | 1329 | 1876 | 32466 | 33010 | 5.19E-93 | 350 |
| S2-wms-combined_181849 | NC_047912.1 | NC_047912.1 | Faecalibacterium phage FP_Lugh | 98.216 | 3 | 897 | 16 | 0 | 4155 | 5051 | 33988 | 33092 | 0 | 1568 |
| S4-wms-combined_42647 | NC_047912.1 | NC_047912.1 | Faecalibacterium phage FP_Lugh | 91.235 | 24 | 3799 | 301 | 29 | 23639 | 27420 | 26959 | 30742 | 0 | 5142 |
| S6-wms-combined_56464 | NC_047912.1 | NC_047912.1 | Faecalibacterium phage FP_Lugh | 74.34 | 3 | 1099 | 246 | 31 | 14338 | 15418 | 17810 | 18890 | 1.9E-118 | 435 |
| S11-wms-combined_283220 | NC_047913.1 | NC_047913.1 | Faecalibacterium phage FP_Mushu | 76.159 | 82 | 13221 | 2706 | 358 | 3702 | 16737 | 31198 | 18239 | 0 | 6541 |
| S11-wms_3116 | NC_047913.1 | NC_047913.1 | Faecalibacterium phage FP_Mushu | 98.624 | 82 | 31693 | 392 | 18 | 6855 | 38537 | 36636 | 4978 | 0 | 56070 |
| S1-wms_4277 | NC_047913.1 | NC_047913.1 | Faecalibacterium phage FP_Mushu | 74.264 | 50 | 8937 | 1907 | 299 | 11767 | 20538 | 27745 | 19037 | 0 | 3398 |
| S7-wms_113137 | NC_047913.1 | NC_047913.1 | Faecalibacterium phage FP_Mushu | 96.959 | 96 | 32099 | 899 | 53 | 4154 | 36208 | 4060 | 36125 | 0 | 53797 |
| S7-wms_200936 | NC_047913.1 | NC_047913.1 | Faecalibacterium phage FP_Mushu | 76.395 | 65 | 9909 | 1941 | 293 | 9729 | 19431 | 12372 | 22088 | 0 | 4974 |
| S17-wms-combined_193958 | NC_047913.1 | NC_047913.1 | Faecalibacterium phage FP_Mushu | 78.11 | 26 | 4719 | 929 | 88 | 76711 | 81386 | 12372 | 17029 | 0 | 2896 |
| S17-wms-combined_175924 | NC_047913.1 | NC_047913.1 | Faecalibacterium phage FP_Mushu | 73.235 | 19 | 3258 | 689 | 130 | 19334 | 22490 | 18275 | 21450 | 0 | 1016 |
| S12-wms_193565 | NC_047913.1 | NC_047913.1 | Faecalibacterium phage FP_Mushu | 77.923 | 53 | 18440 | 3591 | 394 | 34422 | 52640 | 18236 | 36416 | 0 | 11057 |
| S8-wms_30498 | NC_047913.1 | NC_047913.1 | Faecalibacterium phage FP_Mushu | 75.218 | 70 | 4588 | 967 | 124 | 13613 | 18128 | 27631 | 23142 | 0 | 2017 |
| S1-wms-combined_68998 | NC_047914.1 | NC_047914.1 | Faecalibacterium phage FP_Taranis | 89.987 | 76 | 11066 | 1062 | 32 | 16349 | 27390 | 41087 | 30044 | 0 | 14255 |
| S1-wms_29288 | NC_047914.1 | NC_047914.1 | Faecalibacterium phage FP_Taranis | 95.193 | 22 | 1581 | 69 | 7 | 32341 | 33917 | 14091 | 12514 | 0 | 2492 |
| S7-wms-combined_111004 | NC_047914.1 | NC_047914.1 | Faecalibacterium phage FP_Taranis | 96.322 | 64 | 17673 | 619 | 25 | 24790 | 42446 | 34902 | 17245 | 0 | 29006 |
| S17-wms-combined_2234 | NC_047914.1 | NC_047914.1 | Faecalibacterium phage FP_Taranis | 82.27 | 3 | 705 | 122 | 3 | 64903 | 65605 | 5055 | 4352 | 7.7E-170 | 606 |
| S12-wms-combined_323943 | NC_047914.1 | NC_047914.1 | Faecalibacterium phage FP_Taranis | 80.919 | 10 | 1698 | 305 | 13 | 29525 | 31214 | 20813 | 19127 | 0 | 1323 |
| S2-wms-combined_140996 | NC_047914.1 | NC_047914.1 | Faecalibacterium phage FP_Taranis | 81.513 | 0 | 714 | 131 | 1 | 95406 | 96118 | 4351 | 5064 | 3.5E-163 | 586 |
| S8-wms-combined_225114 | NC_047914.1 | NC_047914.1 | Faecalibacterium phage FP_Taranis | 89.41 | 16 | 6308 | 590 | 48 | 43271 | 49531 | 43744 | 50020 | 0 | 7877 |
| S1-wms-combined_118167 | NC_047915.1 | NC_047915.1 | Faecalibacterium phage FP_Toutatis | 90.045 | 4 | 1989 | 182 | 11 | 48270 | 50249 | 45532 | 47513 | 0 | 2562 |
| S12-wms-combined_264077 | NC_047915.1 | NC_047915.1 | Faecalibacterium phage FP_Toutatis | 78.82 | 6 | 1525 | 299 | 19 | 1537 | 3052 | 1 | 1510 | 0 | 1005 |
| S2-wms-combined_13246 | NC_047915.1 | NC_047915.1 | Faecalibacterium phage FP_Toutatis | 85.849 | 0 | 106 | 13 | 2 | 172932 | 173036 | 24028 | 23924 | 2.52E-20 | 111 |
| S6-wms-combined_126236 | NC_047915.1 | NC_047915.1 | Faecalibacterium phage FP_Toutatis | 90.16 | 2 | 376 | 34 | 2 | 61928 | 62302 | 23829 | 23456 | 1E-133 | 486 |
| S6-wms-combined_138710 | NC_047915.1 | NC_047915.1 | Faecalibacterium phage FP_Toutatis | 91.954 | 0 | 87 | 4 | 3 | 34619 | 34704 | 53800 | 53884 | 2.48E-23 | 119 |
| S8-wms_152831 | NC_047915.1 | NC_047915.1 | Faecalibacterium phage FP_Toutatis | 99.553 | 89 | 21705 | 95 | 2 | 6916 | 28619 | 46030 | 24327 | 0 | 39543 |
| S9-wms-combined_26410 | NC_047916.1 | NC_047916.1 | Faecalibacterium phage FP_oengus | 77.919 | 1 | 394 | 83 | 4 | 47587 | 47978 | 47512 | 47903 | 2.26E-60 | 243 |
| S11-wms-combined_236214 | NC_047916.1 | NC_047916.1 | Faecalibacterium phage FP_oengus | 77.932 | 10 | 4187 | 837 | 69 | 88862 | 92999 | 31755 | 27607 | 0 | 2532 |
| S1-wms-combined_304377 | NC_047916.1 | NC_047916.1 | Faecalibacterium phage FP_oengus | 87.232 | 26 | 4065 | 500 | 16 | 19096 | 23150 | 12400 | 16455 | 0 | 4614 |
| S2-wms-combined_57293 | NC_047916.1 | NC_047916.1 | Faecalibacterium phage FP_oengus | 87.037 | 0 | 108 | 11 | 2 | 2911 | 3015 | 22379 | 22272 | 2.96E-23 | 119 |
| S2-wms_124416 | NC_047916.1 | NC_047916.1 | Faecalibacterium phage FP_oengus | 100 | 0 | 123 | 0 | 0 | 58769 | 58891 | 22140 | 22262 | 5.87E-56 | 228 |
| S7-wms-combined_137858 | NC_055744.1 | NC_055744.1 | Synechococcus phage S-B64 | 90 | 0 | 60 | 6 | 0 | 38862 | 38921 | 102482 | 102423 | 5.4E-11 | 78.7 |
| S17-wms-combined_56061 | NC_055744.1 | NC_055744.1 | Synechococcus phage S-B64 | 90 | 0 | 60 | 6 | 0 | 106366 | 106425 | 102482 | 102423 | 1.22E-10 | 78.7 |
| S5-wms_164032 | NC_055000.1 | NC_055000.1 | Lactococcus phage vB_Llc_bIBBA3 | 94.585 | 100 | 11283 | 597 | 13 | 1 | 11275 | 7700 | 18976 | 0 | 17439 |
| S2-wms-combined_24549 | NC_049442.1 | NC_049442.1 | Arthrobacter phage KBurrousTX | 97.222 | 0 | 36 | 1 | 0 | 60877 | 60912 | 28935 | 28900 | 7.03E-06 | 62.1 |
| S4-wms-combined_51079 | NC_048101.1 | NC_048101.1 | Microbacterium phage Goodman | 79.574 | 1 | 235 | 46 | 2 | 7804 | 8037 | 14101 | 13868 | 5.44E-38 | 167 |
| S1-wms-combined_279492 | NC_048144.1 | NC_048144.1 | Microbacterium phage Schubert | 84.962 | 0 | 133 | 20 | 0 | 3168 | 3300 | 10342 | 10210 | 1.81E-28 | 135 |
| S7-wms-combined_147158 | NC_048144.1 | NC_048144.1 | Microbacterium phage Schubert | 73.701 | 0 | 308 | 61 | 16 | 8648 | 8948 | 10495 | 10201 | 4.65E-18 | 102 |
| S4-wms_141607 | NC_048176.1 | NC_048176.1 | Gordonia phage GodonK | 83.562 | 0 | 73 | 12 | 0 | 34114 | 34186 | 84174 | 84246 | 2.26E-08 | 69.4 |

| | | | | | | | | | | | | | | |
|---|---|---|---|---|---|---|---|---|---|---|---|---|---|---|
| S13_vs_35816 | NC_047911.1 | NC_047911.1 | Faecalibacterium phage FP_Lagaffe | 75.521 | 4 | 2161 | 477 | 41 | 35109 | 37240 | 2460 | 323 | 0 | 1013 |
| S3_vf_99734 | NC_047911.1 | NC_047911.1 | Faecalibacterium phage FP_Lagaffe | 75.812 | 5 | 2187 | 476 | 43 | 380 | 2535 | 317 | 2481 | 0 | 1059 |
| S16_vs_172749 | NC_047911.1 | NC_047911.1 | Faecalibacterium phage FP_Lagaffe | 78.173 | 1 | 197 | 37 | 6 | 1623 | 1816 | 25584 | 25391 | 6.56E-24 | 121 |
| S16_vs_86116 | NC_047911.1 | NC_047911.1 | Faecalibacterium phage FP_Lagaffe | 78.123 | 56 | 7396 | 1419 | 161 | 16 | 7317 | 96 | 7386 | 0 | 4510 |
| S8_vs_206526 | NC_047911.1 | NC_047911.1 | Faecalibacterium phage FP_Lagaffe | 80.348 | 2 | 804 | 154 | 4 | 41672 | 42473 | 42445 | 43246 | 5E-170 | 606 |
| S11_vs_20225 | NC_047912.1 | NC_047912.1 | Faecalibacterium phage FP_Lugh | 72.508 | 7 | 1495 | 353 | 52 | 1480 | 2945 | 9499 | 10964 | 2.6E-117 | 431 |
| S14_vs_14737 | NC_047912.1 | NC_047912.1 | Faecalibacterium phage FP_Lugh | 73.95 | 2 | 833 | 188 | 24 | 28955 | 29771 | 18892 | 18073 | 1.23E-80 | 309 |
| S14_vs_9162 | NC_047912.1 | NC_047912.1 | Faecalibacterium phage FP_Lugh | 93.029 | 1 | 416 | 26 | 2 | 80807 | 81222 | 2538 | 2950 | 4.1E-169 | 604 |
| S1_vs_133577 | NC_047912.1 | NC_047912.1 | Faecalibacterium phage FP_Lugh | 78.59 | 19 | 3830 | 741 | 69 | 33894 | 37682 | 9431 | 13222 | 0 | 2457 |
| S3_vs_119771 | NC_047912.1 | NC_047912.1 | Faecalibacterium phage FP_Lugh | 78.358 | 24 | 4542 | 829 | 113 | 9454 | 13889 | 13924 | 9431 | 0 | 2800 |
| S16_vs_81805 | NC_047912.1 | NC_047912.1 | Faecalibacterium phage FP_Lugh | 75.436 | 4 | 1433 | 304 | 40 | 25 | 1436 | 9559 | 10964 | 0 | 652 |
| S16_vs_92150 | NC_047912.1 | NC_047912.1 | Faecalibacterium phage FP_Lugh | 75.542 | 4 | 1153 | 251 | 27 | 28510 | 29645 | 11532 | 12670 | 1.2E-149 | 538 |
| S16_vs_132979 | NC_047912.1 | NC_047912.1 | Faecalibacterium phage FP_Lugh | 74.34 | 3 | 1099 | 246 | 31 | 986 | 2066 | 18890 | 17810 | 1.7E-118 | 435 |
| S10_vs_46819 | NC_047912.1 | NC_047912.1 | Faecalibacterium phage FP_Lugh | 79.022 | 3 | 920 | 177 | 12 | 31481 | 32392 | 32201 | 33112 | 6.3E-173 | 616 |
| S12_vs_87088 | NC_047912.1 | NC_047912.1 | Faecalibacterium phage FP_Lugh | 73.628 | 2 | 747 | 164 | 30 | 36865 | 37595 | 26808 | 27537 | 5.07E-65 | 257 |
| S4_vs_VIRSorter_27316-cat_2 | NC_047912.1 | NC_047912.1 | Faecalibacterium phage FP_Lugh | 93.182 | 1 | 308 | 21 | 0 | 60191 | 60498 | 6150 | 5843 | 1.1E-123 | 453 |
| S6_vs_53966 | NC_047912.1 | NC_047912.1 | Faecalibacterium phage FP_Lugh | 74.34 | 4 | 1099 | 246 | 31 | 6367 | 7447 | 18890 | 17810 | 1.6E-118 | 435 |
| S8_vs_63277 | NC_047912.1 | NC_047912.1 | Faecalibacterium phage FP_Lugh | 87.859 | 39 | 1705 | 192 | 15 | 6601 | 8298 | 21909 | 20213 | 0 | 1988 |
| S17_vf_97671 | NC_047913.1 | NC_047913.1 | Faecalibacterium phage FP_Mushu | 75.223 | 84 | 4589 | 965 | 126 | 10362 | 14877 | 27631 | 23142 | 0 | 2017 |
| S11_vs_52577 | NC_047913.1 | NC_047913.1 | Faecalibacterium phage FP_Mushu | 76.159 | 91 | 13221 | 2706 | 358 | 13237 | 26272 | 18239 | 31198 | 0 | 6541 |
| S5_vf_134040 | NC_047913.1 | NC_047913.1 | Faecalibacterium phage FP_Mushu | 98.45 | 68 | 24509 | 343 | 19 | 137 | 24634 | 12154 | 36636 | 0 | 43120 |
| S7_vf_39724 | NC_047913.1 | NC_047913.1 | Faecalibacterium phage FP_Mushu | 96.959 | 95 | 32099 | 899 | 53 | 4152 | 36206 | 4060 | 36125 | 0 | 53797 |
| S8_vs_134805 | NC_047913.1 | NC_047913.1 | Faecalibacterium phage FP_Mushu | 75.218 | 62 | 4588 | 967 | 124 | 25513 | 30028 | 23142 | 27631 | 0 | 2017 |
| S11_vs_11994 | NC_047914.1 | NC_047914.1 | Faecalibacterium phage FP_Taranis | 82.575 | 2 | 637 | 105 | 5 | 37006 | 37638 | 4353 | 4987 | 6.1E-155 | 556 |
| S3_vs_90331 | NC_047914.1 | NC_047914.1 | Faecalibacterium phage FP_Taranis | 96.924 | 28 | 5234 | 149 | 7 | 14 | 5241 | 48971 | 43744 | 0 | 8763 |
| S5_vf_122568 | NC_047914.1 | NC_047914.1 | Faecalibacterium phage FP_Taranis | 82.213 | 2 | 714 | 126 | 1 | 49480 | 50192 | 4351 | 5064 | 3.5E-172 | 614 |
| S5_vs_130958 | NC_047914.1 | NC_047914.1 | Faecalibacterium phage FP_Taranis | 77.215 | 1 | 474 | 103 | 4 | 12284 | 12753 | 20298 | 20770 | 1.65E-69 | 272 |
| S7_vs_135322 | NC_047914.1 | NC_047914.1 | Faecalibacterium phage FP_Taranis | 96.016 | 79 | 16089 | 610 | 25 | 1 | 16073 | 18829 | 34902 | 0 | 26131 |
| S8_vs_179529 | NC_047914.1 | NC_047914.1 | Faecalibacterium phage FP_Taranis | 89.363 | 15 | 6308 | 593 | 48 | 3069 | 9329 | 50020 | 43744 | 0 | 7860 |
| S3_vs_42569 | NC_047915.1 | NC_047915.1 | Faecalibacterium phage FP_Toutatis | 76.708 | 28 | 8196 | 1706 | 166 | 19351 | 27451 | 32606 | 24519 | 0 | 4372 |
| S5_vs_124677 | NC_047915.1 | NC_047915.1 | Faecalibacterium phage FP_Toutatis | 90.16 | 1 | 376 | 34 | 2 | 10705 | 11079 | 23456 | 23829 | 1.6E-133 | 486 |
| S5_vs_130294 | NC_047915.1 | NC_047915.1 | Faecalibacterium phage FP_Toutatis | 91.954 | 0 | 87 | 4 | 3 | 3887 | 3972 | 53884 | 53800 | 2.69E-23 | 119 |
| S16_vs_18440 | NC_047915.1 | NC_047915.1 | Faecalibacterium phage FP_Toutatis | 91.743 | 21 | 8381 | 631 | 46 | 21734 | 30083 | 53591 | 45241 | 0 | 11587 |
| S8_vf_101283 | NC_047915.1 | NC_047915.1 | Faecalibacterium phage FP_Toutatis | 99.383 | 90 | 22538 | 135 | 4 | 1 | 22535 | 23494 | 46030 | 0 | 40847 |
| S11_vs_104064 | NC_047916.1 | NC_047916.1 | Faecalibacterium phage FP_oengus | 76.562 | 0 | 192 | 39 | 6 | 8452 | 8640 | 15848 | 15660 | 1.32E-17 | 100 |
| S11_vs_68151 | NC_047916.1 | NC_047916.1 | Faecalibacterium phage FP_oengus | 77.665 | 1 | 394 | 84 | 4 | 48891 | 49282 | 47903 | 47512 | 1.02E-58 | 237 |
| S3_vs_68031 | NC_047916.1 | NC_047916.1 | Faecalibacterium phage FP_oengus | 76.663 | 15 | 4585 | 975 | 75 | 15802 | 20337 | 16865 | 12327 | 0 | 2451 |
| S5_vs_117319 | NC_047916.1 | NC_047916.1 | Faecalibacterium phage FP_oengus | 76.354 | 14 | 4597 | 984 | 81 | 85786 | 90332 | 16870 | 12327 | 0 | 2372 |
| S7_vs_41098 | NC_047916.1 | NC_047916.1 | Faecalibacterium phage FP_oengus | 78.117 | 1 | 393 | 80 | 6 | 12358 | 12747 | 47527 | 47916 | 5.95E-61 | 244 |
| S7_vs_87279 | NC_047916.1 | NC_047916.1 | Faecalibacterium phage FP_oengus | 87.09 | 37 | 4485 | 554 | 22 | 9912 | 14385 | 16870 | 12400 | 0 | 5051 |
| S6_vs_34453 | NC_047916.1 | NC_047916.1 | Faecalibacterium phage FP_oengus | 76.73 | 18 | 3872 | 830 | 64 | 47192 | 51026 | 16237 | 12400 | 0 | 2093 |
| S3_vf_44405 | NC_042106.1 | NC_042106.1 | Streptomyces phage Bing | 72.328 | 4 | 1684 | 406 | 51 | 34513 | 36172 | 4548 | 2901 | 4.4E-130 | 473 |
| S13_vf_38708 | NC_042119.1 | NC_042119.1 | Microbacterium phage Pikmin | 77.544 | 1 | 285 | 55 | 3 | 21962 | 22237 | 10030 | 10314 | 8.3E-37 | 163 |
| S7_vs_97737 | NC_055744.1 | NC_055744.1 | Synechococcus phage S-B64 | 90 | 0 | 60 | 6 | 0 | 63490 | 63549 | 102482 | 102423 | 7.93E-11 | 78.7 |
| S3_vf_54353 | NC_048021.1 | NC_048021.1 | Gordonia phage Daredevil | 77.558 | 1 | 303 | 57 | 11 | 2320 | 2618 | 1709 | 1414 | 2.73E-39 | 172 |
| S3_vf_5525 | NC_048070.1 | NC_048070.1 | Corynebacterium phage Juicebox | 76.892 | 1 | 251 | 45 | 7 | 31068 | 31313 | 2577 | 2335 | 1.16E-26 | 130 |
| S5_vf_55415 | NC_049977.1 | NC_049977.1 | Bacteroides phage crAss001 | 96.248 | 81 | 10553 | 358 | 23 | 8362 | 18887 | 93048 | 82507 | 0 | 17259 |
| S7_vf_89197 | NC_054999.1 | NC_054999.1 | Lactococcus phage vB_LLc_bIBB14 | 96.588 | 99 | 16941 | 529 | 33 | 1 | 16913 | 249 | 17168 | 0 | 28036 |
| S11_vf_3790 | NC_048077.1 | NC_048077.1 | Dickeya phage vB_DsoM_JA11 | 89.796 | 0 | 49 | 5 | 0 | 51606 | 51654 | 245750 | 245702 | 1.95E-06 | 63.9 |
| S16_vf_173109 | NC_048101.1 | NC_048101.1 | Microbacterium phage Goodman | 74.857 | 1 | 350 | 80 | 8 | 14055 | 14400 | 14101 | 13756 | 1.75E-33 | 152 |
| S4_vf_28601 | NC_048115.1 | NC_048115.1 | Arthrobacter phage Liebe | 75.589 | 1 | 467 | 99 | 13 | 11852 | 12310 | 9484 | 9943 | 8.27E-53 | 217 |
| S7_vs_73280 | NC_048144.1 | NC_048144.1 | Microbacterium phage Schubert | 73.701 | 1 | 308 | 61 | 16 | 28528 | 28828 | 10495 | 10201 | 2.46E-18 | 102 |
| S8_vf_147128 | NC_049445.1 | NC_049445.1 | Acinetobacter phage AbTZA1 | 89.873 | 0 | 79 | 8 | 0 | 38463 | 38541 | 139783 | 139705 | 5.16E-18 | 102 |
| S7_vf_38697 | NC_055027.1 | NC_055027.1 | Lactococcus phage CHPC971 | 83.824 | 0 | 136 | 20 | 2 | 6057 | 6191 | 52420 | 52286 | 6.31E-26 | 128 |
| S4_vf_31682 | NC_048780.1 | NC_048780.1 | Corynebacterium phage StAB | 77.224 | 1 | 281 | 54 | 10 | 22745 | 23020 | 35257 | 34982 | 1.79E-34 | 156 |
| S10_vf_120738 | NC_048804.1 | NC_048804.1 | Stenotrophomonas phage Mendera | 90.741 | 0 | 54 | 5 | 0 | 102755 | 102808 | 82069 | 82016 | 6.87E-09 | 73.1 |
| S8_vf_11794 | NC_048804.1 | NC_048804.1 | Stenotrophomonas phage Mendera | 94.231 | 0 | 52 | 3 | 0 | 47016 | 47067 | 82067 | 82016 | 3.94E-11 | 80.5 |
| S12_vs_163990 | NC_048817.1 | NC_048817.1 | Gordonia phage Tanis | 80.337 | 0 | 178 | 30 | 4 | 1417 | 1593 | 58115 | 57942 | 1.77E-26 | 130 |
| S13_vs_152223 | NC_048822.1 | NC_048822.1 | Gordonia phage Avazak | 78.417 | 0 | 278 | 58 | 2 | 13184 | 13460 | 14167 | 14443 | 1.72E-41 | 180 |
| S10_vs_26365 | NC_048822.1 | NC_048822.1 | Gordonia phage Avazak | 87.273 | 0 | 55 | 5 | 2 | 16510 | 16563 | 45878 | 45825 | 6.24E-06 | 62.1 |
| S3_vf_57422 | NC_049971.1 | NC_049971.1 | Bacillus phage vB_Bpu_PumA1 | 94.872 | 0 | 39 | 2 | 0 | 10740 | 10778 | 2285 | 2323 | 1.71E-06 | 62.1 |
| S3_vf_23042 | NC_049474.1 | NC_049474.1 | Lactococcus phage P1532 | 89.353 | 67 | 5025 | 457 | 50 | 1551 | 6544 | 24984 | 20007 | 0 | 6244 |
| S11_vs_5077 | NC_049395.1 | NC_049395.1 | Escherichia phage ESSI2_ev015 genome assembly | 81.812 | 67 | 5960 | 1006 | 61 | 10057 | 15978 | 20440 | 14521 | 0 | 4929 |
| S5_vf_145579 | NC_055832.1 | NC_055832.1 | Bacteroides phage DAC15 | 71.445 | 0 | 851 | 211 | 29 | 23879 | 24713 | 88907 | 89741 | 5.24E-46 | 196 |
| S13_vf_1770 | NC_048862.1 | NC_048862.1 | Salmonella phage astrithr | 84.27 | 1 | 89 | 14 | 0 | 8738 | 8826 | 7417 | 7329 | 2.22E-14 | 87.9 |
| S3_vf_108959 | NC_055841.1 | NC_055841.1 | Agrobacterium phage OLIVR5 | 90.411 | 0 | 73 | 6 | 1 | 53655 | 53726 | 82011 | 82083 | 6.49E-16 | 95.3 |
| S6_vs_56498 | NC_048848.1 | NC_048848.1 | Butyrivibrio phage Arawn | 81.977 | 1 | 172 | 30 | 1 | 22248 | 22418 | 4967 | 5138 | 3.47E-31 | 145 |
| S6_vs_10314 | NC_048848.1 | NC_048848.1 | Butyrivibrio phage Arawn | 73.244 | 0 | 299 | 74 | 4 | 60131 | 60426 | 3398 | 3693 | 1.47E-18 | 104 |
| S14_vf_117155 | NC_048876.1 | NC_048876.1 | Gordonia phage Secretariat | 77.66 | 1 | 282 | 48 | 11 | 30333 | 30608 | 44247 | 44519 | 5.17E-35 | 158 |
| S5_vs_30490 | NC_055848.1 | NC_055848.1 | Vibrio phage AG74 | 88.571 | 0 | 70 | 6 | 2 | 19400 | 19468 | 75368 | 75436 | 1.29E-12 | 84.2 |
| S5_vs_51416 | NC_049900.1 | NC_049900.1 | Listeria phage LP-HM00113468 | 81.25 | 0 | 96 | 12 | 5 | 32663 | 32752 | 24506 | 24411 | 1.89E-09 | 73.1 |
| S15_vs_27510 | NC_062754.1 | NC_062754.1 | Winogradskyella phage Peternella_1 | 95.122 | 0 | 41 | 2 | 0 | 45444 | 45484 | 13010 | 12970 | 4.75E-07 | 65.8 |
| S5_vf_21869 | NC_054463.1 | NC_054463.1 | Ralstonia phage Bakoly | 89.116 | 1 | 294 | 32 | 0 | 33184 | 33477 | 36736 | 36443 | 8.75E-98 | 366 |
| S9_vs_5676 | NC_054959.1 | NC_054959.1 | Ralstonia phage Gerry | 90.977 | 0 | 133 | 12 | 0 | 34055 | 34187 | 57782 | 57914 | 9.68E-42 | 180 |
| S14_vf_57689 | NC_055859.1 | NC_055859.1 | Gordonia phage Paries | 80.46 | 0 | 87 | 14 | 3 | 5846 | 5932 | 11690 | 11773 | 1.02E-06 | 63.9 |
| S13_vf_143422 | NC_055868.1 | NC_055868.1 | CrAssphage cr50_1 | 95.284 | 87 | 19785 | 901 | 16 | 34611 | 54390 | 92077 | 72320 | 0 | 31338 |
| S4_vf_4838 | NC_055868.1 | NC_055868.1 | CrAssphage cr50_1 | 96.295 | 86 | 23992 | 819 | 39 | 59055 | 83001 | 92077 | 68111 | 0 | 39316 |
| S3_vs_80151 | NC_055869.1 | NC_055869.1 | CrAssphage cr55_1 | 95.699 | 1 | 651 | 28 | 0 | 111921 | 112571 | 85720 | 86370 | 0 | 1048 |
| S10_vs_119298 | NC_055870.1 | NC_055870.1 | CrAssphage cr107_1 | 77.457 | 0 | 173 | 34 | 5 | 40508 | 40677 | 71463 | 71293 | 4.02E-17 | 99 |
| S7_vs_51332 | NC_055872.1 | NC_055872.1 | CrAssphage cr118_1 | 83.478 | 1 | 230 | 32 | 4 | 9535 | 9761 | 92374 | 92600 | 1.72E-50 | 209 |
| S4_vf_31981 | NC_055872.1 | NC_055872.1 | CrAssphage cr118_1 | 80.4 | 20 | 2148 | 385 | 27 | 16351 | 18482 | 76574 | 74447 | 0 | 1602 |
| S17_vf_81232 | NC_055875.1 | NC_055875.1 | CrAssphage cr10_1 | 95.508 | 89 | 22685 | 1006 | 8 | 59720 | 82397 | 95768 | 73090 | 0 | 36234 |
| S16_vs_127905 | NC_055876.1 | NC_055876.1 | CrAssphage cr11_1 | 98.795 | 1 | 332 | 4 | 0 | 16192 | 16523 | 39413 | 39082 | 2E-165 | 592 |
| S9_vf_51656 | NC_055877.1 | NC_055877.1 | CrAssphage cr8_1 | 98.142 | 64 | 18410 | 330 | 8 | 42467 | 60866 | 38979 | 20572 | 0 | 32092 |
| S4_vf_4891 | NC_055877.1 | NC_055877.1 | CrAssphage cr8_1 | 84.615 | 0 | 130 | 19 | 1 | 4265 | 4393 | 40600 | 40729 | 4.49E-26 | 128 |
| S8_vf_166677 | NC_055877.1 | NC_055877.1 | CrAssphage cr8_1 | 85.935 | 42 | 5567 | 714 | 58 | 69538 | 75061 | 62011 | 56471 | 0 | 5879 |
| S3_vf_127406 | NC_055878.1 | NC_055878.1 | CrAssphage cr108_1 | 85.636 | 1 | 550 | 73 | 4 | 16924 | 17470 | 23962 | 23416 | 8.7E-160 | 573 |
| S8_vf_131009 | NC_055881.1 | NC_055881.1 | CrAssphage cr112_1 | 84.146 | 0 | 82 | 8 | 4 | 31627 | 31707 | 64106 | 64029 | 7.88E-10 | 75 |
| S11_vf_104044 | NC_055883.1 | NC_055883.1 | CrAssphage cr85_1 | 99.089 | 95 | 37430 | 329 | 10 | 35690 | 73112 | 64398 | 26974 | 0 | 67221 |
| S3_vf_127447 | NC_055884.1 | NC_055884.1 | CrAssphage cr126_1 | 89.046 | 45 | 5532 | 555 | 36 | 37993 | 43491 | 12089 | 6576 | 0 | 6811 |
| S17_vf_35138 | NC_055885.1 | NC_055885.1 | CrAssphage cr128_1 | 87.018 | 1 | 285 | 30 | 7 | 8407 | 8686 | 36512 | 36794 | 3.87E-82 | 315 |
| S13_vs_151510 | NC_055885.1 | NC_055885.1 | CrAssphage cr128_1 | 83.686 | 2 | 1275 | 187 | 18 | 76253 | 77519 | 76172 | 74911 | 0 | 1182 |
| S3_vs_127445 | NC_055885.1 | NC_055885.1 | CrAssphage cr128_1 | 81.364 | 0 | 220 | 33 | 7 | 28166 | 28381 | 84913 | 84698 | 3.63E-39 | 172 |
| S4_vf_12614 | NC_055885.1 | NC_055885.1 | CrAssphage cr128_1 | 87.723 | 3 | 505 | 53 | 9 | 11623 | 12121 | 36512 | 37013 | 2.9E-162 | 580 |
| S4_vf_26417 | NC_055886.1 | NC_055886.1 | CrAssphage cr127_1 | 97.383 | 99 | 78064 | 1821 | 120 | 3437 | 81347 | 83336 | 5342 | 0 | 132600 |
| S11_vf_104035 | NC_055889.1 | NC_055889.1 | CrAssphage cr53_1 | 91.659 | 71 | 18176 | 1394 | 81 | 6755 | 24886 | 50225 | 32128 | 0 | 25054 |

| | | | | | | | | | | | | | | |
|---|---|---|---|---|---|---|---|---|---|---|---|---|---|---|
| S2-wms_246296 | NC_042094.1 | NC_042094.1 | Vibrio phage Ceto | 90 | 0 | 60 | 5 | 1 | 30103 | 30162 | 77283 | 77341 | 2.07E-10 | 76.8 |
| S1-wms_170115 | NC_048780.1 | NC_048780.1 | Corynebacterium phage StAB | 77.936 | 1 | 281 | 52 | 10 | 9819 | 10094 | 34982 | 35257 | 8.84E-38 | 167 |
| S12-wms-combined_117628 | NC_048804.1 | NC_048804.1 | Stenotrophomonas phage Mendera | 87.805 | 0 | 82 | 7 | 3 | 37091 | 37170 | 67809 | 67889 | 2.23E-15 | 93.5 |
| S2-wms-combined_62377 | NC_048822.1 | NC_048822.1 | Gordonia phage Avazak | 78.417 | 0 | 278 | 58 | 2 | 47444 | 47720 | 14443 | 14167 | 1.72E-41 | 180 |
| S11-wms-combined_154845 | NC_049395.1 | NC_049395.1 | Escherichia phage ESSI2_ev015 genome assembly | 81.812 | 82 | 5960 | 1006 | 61 | 11886 | 17807 | 14521 | 20440 | 0 | 4929 |
| S8-wms-combined_200168 | NC_055832.1 | NC_055832.1 | Bacteroides phage DAC15 | 89.231 | 0 | 65 | 6 | 1 | 28125 | 28189 | 94697 | 94634 | 2.31E-11 | 80.5 |
| S6-wms-combined_88414 | NC_048848.1 | NC_048848.1 | Butyrivibrio phage Arawn | 73.244 | 0 | 299 | 74 | 4 | 60131 | 60426 | 3398 | 3693 | 1.77E-18 | 104 |
| S1-wms-combined_307323 | NC_054924.1 | NC_054924.1 | Escherichia phage vB_EcoM_Lutter | 96.667 | 0 | 60 | 2 | 0 | 33870 | 33929 | 72883 | 72942 | 1.59E-17 | 100 |
| S4-wms_73990 | NC_055868.1 | NC_055868.1 | CrAssphage cr50_1 | 96.295 | 91 | 23992 | 819 | 39 | 32911 | 56857 | 92077 | 68111 | 0 | 39316 |
| S4-wms_45128 | NC_055877.1 | NC_055877.1 | CrAssphage cr8_1 | 84.615 | 0 | 130 | 19 | 1 | 1616 | 1744 | 40600 | 40729 | 4.28E-26 | 128 |
| S7-wms-combined_163980 | NC_055880.1 | NC_055880.1 | CrAssphage cr111_1 | 89.873 | 0 | 79 | 7 | 1 | 1651 | 1729 | 80243 | 80166 | 6.07E-18 | 100 |
| S6-wms-combined_50721 | NC_055880.1 | NC_055880.1 | CrAssphage cr111_1 | 87.342 | 0 | 79 | 6 | 4 | 65402 | 65478 | 80169 | 80245 | 2.33E-13 | 87.9 |
| S12-wms-combined_260036 | NC_055885.1 | NC_055885.1 | CrAssphage cr128_1 | 93.571 | 2 | 560 | 31 | 4 | 8246 | 8805 | 83552 | 84106 | 0 | 830 |
| S2-wms-combined_197577 | NC_055885.1 | NC_055885.1 | CrAssphage cr128_1 | 80.455 | 0 | 220 | 35 | 7 | 21840 | 22055 | 84913 | 84698 | 6.6E-36 | 161 |
| S4-wms_107182 | NC_055910.1 | NC_055910.1 | Providencia phage PSTCR5 | 95.122 | 0 | 41 | 2 | 0 | 1869 | 1909 | 88714 | 88674 | 1.43E-07 | 65.8 |
| S5-wms-combined_210959 | NC_011421.1 | NC_011421.1 | Bacillus phage SPO1 | 87.931 | 0 | 58 | 5 | 2 | 84305 | 84362 | 28214 | 28269 | 4.47E-07 | 67.6 |
| S9-wms_19025 | NC_007065.1 | NC_007065.1 | Staphylococcus phage X2 | 80.743 | 1 | 296 | 51 | 3 | 22910 | 23200 | 25379 | 25085 | 1.98E-55 | 226 |
| S2-wms_128962 | NC_067213.1 | NC_067213.1 | Uncultured phage cr150_1 | 95.885 | 92 | 14800 | 605 | 4 | 67422 | 82219 | 12995 | 27792 | 0 | 23954 |
| S6-wms_138342 | NC_067213.1 | NC_067213.1 | Uncultured phage cr150_1 | 96.281 | 95 | 16754 | 615 | 8 | 82876 | 99625 | 12995 | 29744 | 0 | 27481 |
| S12-wms_401372 | NC_067212.1 | NC_067212.1 | Uncultured phage cr19_1 | 83.951 | 0 | 81 | 11 | 2 | 23440 | 23519 | 63467 | 63388 | 3.36E-10 | 76.8 |
| S1-wms_32878 | NC_001697.1 | NC_001697.1 | Haemophilus phage HP1 strain HP1c1 | 84.615 | 0 | 78 | 12 | 0 | 13642 | 13719 | 433 | 356 | 2E-11 | 78.7 |
| S3-wms_94494 | NC_003278.1 | NC_003278.1 | Pseudomonas phage phiCTX DNA | 93.617 | 0 | 47 | 3 | 0 | 16458 | 16504 | 21404 | 21450 | 3.63E-09 | 71.3 |
| S7-wms-combined_106253 | NC_067270.1 | NC_067270.1 | Bixzunavirus I3 | 83.117 | 0 | 77 | 11 | 2 | 80503 | 80578 | 97823 | 97898 | 6.2E-08 | 69.4 |
| S7-wms_244425 | NC_062780.1 | NC_062780.1 | Uncultured phage cr13_1 | 97.418 | 95 | 27696 | 713 | 2 | 38964 | 66658 | 37871 | 65565 | 0 | 47183 |
| S8-wms_111056 | NC_062769.1 | NC_062769.1 | Uncultured phage cr36_1 | 94.854 | 99 | 29653 | 1408 | 98 | 23884 | 53483 | 1 | 29588 | 0 | 46196 |
| S9-wms-combined_20485 | NC_071054.1 | NC_071054.1 | Streptococcus phage P0093 | 81.305 | 19 | 5488 | 968 | 46 | 57600 | 63069 | 14500 | 9053 | 0 | 4397 |
| S2-wms-combined_7756 | NC_070819.1 | NC_070819.1 | Gordonia phage Sidious | 100 | 0 | 33 | 0 | 0 | 44170 | 44202 | 30553 | 30521 | 6.78E-06 | 62.1 |
| S2-wms-combined_58563 | NC_070882.1 | NC_070882.1 | Pseudomonas phage vB_PaeM_PS119XW | 97.436 | 0 | 39 | 0 | 1 | 26506 | 26543 | 296440 | 296402 | 4.43E-07 | 65.8 |
| S7-wms-combined_167867 | NC_070834.1 | NC_070834.1 | Arthrobacter phage Persistence | 85 | 0 | 80 | 11 | 1 | 37222 | 37301 | 43952 | 44030 | 3.84E-11 | 80.5 |
| S1-wms_212940 | NC_070945.1 | NC_070945.1 | Arthrobacter phage Kardesai | 91.667 | 0 | 48 | 4 | 0 | 35153 | 35200 | 56539 | 56586 | 1.37E-07 | 67.6 |
| S5-wms-combined_72410 | NC_070945.1 | NC_070945.1 | Arthrobacter phage Kardesai | 83.333 | 0 | 120 | 20 | 0 | 113746 | 113865 | 9644 | 9763 | 2.49E-20 | 111 |
| S12-wms-combined_107942 | NC_070946.1 | NC_070946.1 | Arthrobacter phage Dynamite | 97.619 | 0 | 42 | 0 | 1 | 17228 | 17269 | 11668 | 11628 | 5.49E-09 | 71.3 |
| S2-wms_140933 | NC_071044.1 | NC_071044.1 | Bacillus phage vB_BanS_Nate | 94.545 | 0 | 55 | 3 | 0 | 44045 | 44099 | 147119 | 147173 | 2.91E-13 | 86.1 |
| S9-wms-combined_76274 | NC_073367.1 | NC_073367.1 | Microbacterium phage Terij | 75 | 1 | 360 | 74 | 16 | 30899 | 31250 | 40144 | 39793 | 4.25E-33 | 152 |
| S5-wms-combined_117213 | NC_073367.1 | NC_073367.1 | Microbacterium phage Terij | 75 | 0 | 360 | 74 | 16 | 6748 | 7099 | 39793 | 40144 | 4.46E-33 | 152 |

| | | | | | | | | | | | | | | |
|---|---|---|---|---|---|---|---|---|---|---|---|---|---|---|
| S8_vf_131802 | NC_055890.1 | NC_055890.1 | CrAssphage cr56_1 | 98.298 | 92 | 21802 | 353 | 16 | 69322 | 91114 | 48431 | 26639 | 0 | 38189 |
| S10_vf_3644 | NC_055891.1 | NC_055891.1 | CrAssphage cr60_1 | 92.691 | 69 | 24600 | 1761 | 33 | 533 | 25112 | 54332 | 29750 | 0 | 35432 |
| S3_vf_25163 | NC_055892.1 | NC_055892.1 | CrAssphage cr109_1 | 97.436 | 90 | 27072 | 659 | 19 | 76435 | 103478 | 52972 | 80036 | 0 | 46115 |
| S12_vf_32615 | NC_055892.1 | NC_055892.1 | CrAssphage cr109_1 | 73.152 | 1 | 1028 | 237 | 35 | 146468 | 147480 | 87821 | 88824 | 3.59E-87 | 333 |
| S15_vf_2032 | NC_055897.1 | NC_055897.1 | CrAssphage cr114_1 | 90.98 | 0 | 255 | 23 | 0 | 29785 | 30039 | 63364 | 63110 | 6.52E-91 | 344 |
| S10_vf_57705 | NC_055902.1 | NC_055902.1 | CrAssphage cr131_1 | 91.489 | 1 | 47 | 2 | 1 | 13450 | 13494 | 443 | 489 | 4.14E-07 | 63.9 |
| S12_vf_8173 | NC_055903.1 | NC_055903.1 | CrAssphage cr272_1 | 100 | 0 | 33 | 0 | 0 | 784 | 816 | 5549 | 5517 | 1.48E-06 | 62.1 |
| S15_vf_71616 | NC_055903.1 | NC_055903.1 | CrAssphage cr272_1 | 100 | 0 | 32 | 0 | 0 | 706 | 737 | 5548 | 5517 | 5.31E-06 | 60.2 |
| S5_vs_130032 | NC_049955.1 | NC_049955.1 | Escherichia phage Lambda_ev243 genome assembly | 97.378 | 51 | 19911 | 514 | 8 | 1 | 19907 | 188 | 20094 | 0 | 33870 |
| S3_vs_17168 | NC_003309.1 | NC_003309.1 | Burkholderia phage phiE125 | 85.714 | 0 | 98 | 11 | 3 | 70721 | 70816 | 43890 | 43986 | 2.34E-17 | 100 |
| S5_vs_20500 | NC_003309.1 | NC_003309.1 | Burkholderia phage phiE125 | 83.333 | 0 | 102 | 14 | 3 | 54966 | 55065 | 43986 | 43886 | 9.79E-15 | 91.6 |
| S16_vs_142561 | NC_003309.1 | NC_003309.1 | Burkholderia phage phiE125 | 83.333 | 0 | 102 | 14 | 3 | 74254 | 74353 | 43986 | 43886 | 1.13E-14 | 91.6 |
| S8_vs_40014 | NC_004348.1 | NC_004348.1 | Enterobacteria phage ST64T | 96.471 | 0 | 85 | 3 | 0 | 33733 | 33817 | 11350 | 11434 | 4.52E-30 | 141 |
| S3_vf_61860 | NC_007065.1 | NC_007065.1 | Staphylococcus phage X2 | 97.619 | 0 | 42 | 1 | 0 | 56851 | 56892 | 35168 | 35127 | 3.36E-09 | 73.1 |
| S10_vf_64371 | NC_007065.1 | NC_007065.1 | Staphylococcus phage X2 | 80.743 | 0 | 296 | 51 | 3 | 3657 | 3947 | 25085 | 25379 | 2.12E-55 | 226 |
| S17_vf_174033 | NC_007021.1 | NC_007021.1 | Staphylococcus phage Twort | 94 | 0 | 50 | 3 | 0 | 8972 | 9021 | 128621 | 128670 | 2.47E-10 | 76.8 |
| S4_vf_31973 | NC_015251.1 | NC_015251.1 | Aeromonas phage 65 | 84.932 | 0 | 73 | 9 | 2 | 99308 | 99379 | 112484 | 112555 | 5.86E-09 | 73.1 |
| S7_vs_72737 | NC_023724.1 | NC_023724.1 | Mycobacterium phage Larva | 97.297 | 0 | 37 | 1 | 0 | 16059 | 16095 | 594 | 630 | 9.96E-07 | 63.9 |
| S3_vf_17748 | NC_067194.1 | NC_067194.1 | Carjivirus communis | 97.401 | 96 | 26937 | 665 | 17 | 60198 | 87115 | 25491 | 52411 | 0 | 45833 |
| S8_vf_87755 | NC_067194.1 | NC_067194.1 | Carjivirus communis | 95.926 | 94 | 13009 | 505 | 25 | 22448 | 35448 | 88721 | 75730 | 0 | 21064 |
| S3_vs_45147 | NC_067217.1 | NC_067217.1 | Uncultured phage cr35_1 | 93.264 | 0 | 193 | 11 | 2 | 90048 | 90239 | 56083 | 56274 | 1.89E-72 | 283 |
| S10_vf_43964 | NC_067217.1 | NC_067217.1 | Uncultured phage cr35_1 | 90.217 | 0 | 368 | 34 | 2 | 53561 | 53927 | 56449 | 56083 | 2.3E-131 | 479 |
| S4_vf_31825 | NC_067217.1 | NC_067217.1 | Uncultured phage cr35_1 | 99.327 | 93 | 32104 | 210 | 1 | 14184 | 46287 | 65609 | 97706 | 0 | 58083 |
| S8_vf_49282 | NC_067217.1 | NC_067217.1 | Uncultured phage cr35_1 | 97.521 | 86 | 32111 | 779 | 12 | 22164 | 54270 | 97706 | 65609 | 0 | 54872 |
| S9_vs_83108 | NC_067210.1 | NC_067210.1 | Uncultured phage cr12_1 | 92.077 | 1 | 568 | 42 | 3 | 40284 | 40850 | 92387 | 91822 | 0 | 797 |
| S12_vf_145044 | NC_067210.1 | NC_067210.1 | Uncultured phage cr12_1 | 72.212 | 1 | 1058 | 261 | 29 | 107891 | 108930 | 45525 | 46567 | 1.63E-75 | 294 |
| S2_vf_4130 | NC_067210.1 | NC_067210.1 | Uncultured phage cr12_1 | 96.833 | 87 | 23460 | 716 | 17 | 12894 | 36337 | 13231 | 36679 | 0 | 39181 |
| S13_vf_11246 | NC_067213.1 | NC_067213.1 | Uncultured phage cr150_1 | 95.341 | 92 | 9917 | 458 | 4 | 87428 | 97342 | 27792 | 17878 | 0 | 15751 |
| S17_vs_39515 | NC_067211.1 | NC_067211.1 | Uncultured phage cr105_1 | 89.091 | 0 | 55 | 5 | 1 | 17670 | 17724 | 71501 | 71554 | 8.35E-08 | 67.6 |
| S17_vs_160502 | NC_067211.1 | NC_067211.1 | Uncultured phage cr105_1 | 88.189 | 0 | 127 | 9 | 3 | 21236 | 21362 | 71585 | 71465 | 1.06E-31 | 147 |
| S9_vf_76769 | NC_067211.1 | NC_067211.1 | Uncultured phage cr105_1 | 96.556 | 73 | 10771 | 356 | 11 | 7138 | 17901 | 71917 | 82679 | 0 | 17821 |
| S3_vf_73597 | NC_067211.1 | NC_067211.1 | Uncultured phage cr105_1 | 96.999 | 3 | 1766 | 44 | 8 | 17711 | 19473 | 69826 | 71585 | 0 | 2959 |
| S4_vs_VIRSorter_31975-circular-cat_1 | NC_067211.1 | NC_067211.1 | Uncultured phage cr105_1 | 90.625 | 0 | 128 | 9 | 2 | 607 | 734 | 71585 | 71461 | 1.52E-37 | 167 |
| S3_vf_115304 | NC_067218.1 | NC_067218.1 | Uncultured phage cr23_1 | 98.302 | 92 | 40982 | 649 | 32 | 29291 | 70262 | 58425 | 17481 | 0 | 71780 |
| S15_vf_74586 | NC_067218.1 | NC_067218.1 | Uncultured phage cr23_1 | 77.165 | 0 | 127 | 24 | 5 | 16853 | 16977 | 85004 | 84881 | 5.43E-08 | 69.4 |
| S12_vf_188824 | NC_067212.1 | NC_067212.1 | Uncultured phage cr19_1 | 83.951 | 0 | 81 | 11 | 2 | 79322 | 79401 | 63388 | 63467 | 3.36E-10 | 76.8 |
| S15_vf_75924 | NC_067212.1 | NC_067212.1 | Uncultured phage cr19_1 | 84.716 | 0 | 229 | 18 | 11 | 158300 | 158526 | 96100 | 96313 | 5.17E-51 | 213 |
| S16_vf_151450 | NC_062779.1 | NC_062779.1 | Uncultured phage cr2_1 | 95.356 | 87 | 20800 | 898 | 43 | 74509 | 95263 | 54105 | 33329 | 0 | 32997 |
| S10_vf_36880 | NC_062780.1 | NC_062780.1 | Uncultured phage cr13_1 | 97.037 | 95 | 63088 | 1808 | 55 | 28072 | 91123 | 88488 | 25426 | 0 | 106100 |
| S13_vf_3728 | NC_062769.1 | NC_062769.1 | Uncultured phage cr36_1 | 95.725 | 91 | 29430 | 1179 | 47 | 56238 | 85640 | 35362 | 64739 | 0 | 47304 |
| S5_vf_67622 | NC_062769.1 | NC_062769.1 | Uncultured phage cr36_1 | 97.759 | 1 | 357 | 8 | 0 | 135448 | 135804 | 32111 | 32467 | 3.1E-172 | 616 |
| S16_vf_173095 | NC_062774.1 | NC_062774.1 | Uncultured phage cr54_1 | 97.829 | 95 | 29431 | 637 | 2 | 41516 | 70945 | 40042 | 69471 | 0 | 50807 |
| S8_vf_158225 | NC_062776.1 | NC_062776.1 | Uncultured phage cr9_1 | 96.412 | 93 | 23439 | 784 | 40 | 61766 | 85172 | 90388 | 66975 | 0 | 38572 |
| S11_vf_104059 | NC_062766.1 | NC_062766.1 | Uncultured phage cr123_1 | 97.231 | 94 | 16650 | 431 | 22 | 1 | 16640 | 37068 | 20439 | 0 | 28166 |
| S11_vf_43997 | NC_062770.1 | NC_062770.1 | Uncultured phage cr91_1 | 94.486 | 91 | 18772 | 965 | 51 | 29641 | 48378 | 93332 | 74597 | 0 | 28867 |
| S7_vs_557 | NC_062770.1 | NC_062770.1 | Uncultured phage cr91_1 | 93.792 | 2 | 451 | 24 | 2 | 2488 | 2934 | 68732 | 69182 | 0 | 675 |
| S13_vf_56074 | NC_062778.1 | NC_062778.1 | Uncultured phage cr17_1 | 95.506 | 0 | 89 | 4 | 0 | 36748 | 36836 | 54766 | 54854 | 2.89E-30 | 143 |
| S12_vs_14927 | NC_062778.1 | NC_062778.1 | Uncultured phage cr17_1 | 99.067 | 99 | 52318 | 455 | 9 | 26534 | 78836 | 352 | 52651 | 0 | 93879 |
| S15_vf_20644 | NC_062778.1 | NC_062778.1 | Uncultured phage cr17_1 | 95.455 | 0 | 88 | 4 | 0 | 48086 | 48173 | 54853 | 54766 | 1.03E-29 | 141 |
| S14_vs_15228 | NC_062773.1 | NC_062773.1 | Uncultured phage cr44_1 | 92.308 | 0 | 52 | 3 | 1 | 52667 | 52717 | 18334 | 18283 | 7.44E-09 | 73.1 |
| S3_vs_82895 | NC_062773.1 | NC_062773.1 | Uncultured phage cr44_1 | 92.308 | 0 | 52 | 3 | 1 | 71634 | 71684 | 18334 | 18283 | 7.28E-09 | 73.1 |
| S7_vf_28429 | NC_062773.1 | NC_062773.1 | Uncultured phage cr44_1 | 92.308 | 0 | 52 | 3 | 1 | 103972 | 104022 | 18283 | 18334 | 7.75E-09 | 73.1 |
| S15_vf_33505 | NC_062772.1 | NC_062772.1 | Uncultured phage cr77_1 | 76.65 | 0 | 591 | 118 | 17 | 103959 | 104539 | 87145 | 87725 | 6.05E-80 | 309 |
| S4_vs_VIRSorter_13190-cat_2 | NC_062772.1 | NC_062772.1 | Uncultured phage cr77_1 | 95.579 | 1 | 656 | 29 | 0 | 60981 | 61636 | 28060 | 28715 | 0 | 1051 |
| S8_vs_126582 | NC_062767.1 | NC_062767.1 | Uncultured phage cr149_1 | 92.587 | 2 | 634 | 47 | 0 | 40675 | 41308 | 41485 | 40852 | 0 | 911 |
| S13_vf_87873 | NC_070823.1 | NC_070823.1 | Pseudoalteromonas phage vB_PspS-H40/1 | 90.196 | 0 | 51 | 5 | 0 | 26875 | 26925 | 15563 | 15513 | 8.82E-08 | 67.6 |
| S9_vf_4115 | NC_070923.1 | NC_070923.1 | Streptococcus phage IPP54 | 89.942 | 51 | 13482 | 1280 | 56 | 19815 | 33250 | 29716 | 16265 | 0 | 17315 |
| S9_vf_90856 | NC_071054.1 | NC_071054.1 | Streptococcus phage P0093 | 81.305 | 45 | 5488 | 968 | 46 | 11724 | 17193 | 9053 | 14500 | 0 | 4397 |
| S9_vf_90866 | NC_070709.1 | NC_070709.1 | Streptococcus phage P9854 | 96.226 | 1 | 53 | 2 | 0 | 8428 | 8480 | 25657 | 25709 | 1.65E-14 | 87.9 |
| S3_vs_50557 | NC_070832.1 | NC_070832.1 | Arthrobacter phage Auxilium | 75.792 | 2 | 979 | 213 | 23 | 6649 | 7615 | 39987 | 39021 | 4.9E-130 | 473 |
| S3_vf_127423 | NC_070882.1 | NC_070882.1 | Pseudomonas phage vB_PaeM_PS119XW | 93.478 | 0 | 46 | 3 | 0 | 26239 | 26284 | 149962 | 149917 | 3.1E-08 | 69.4 |
| S7_vs_13370 | NC_070882.1 | NC_070882.1 | Pseudomonas phage vB_PaeM_PS119XW | 91.781 | 0 | 73 | 6 | 0 | 10136 | 10208 | 296440 | 296368 | 1.95E-18 | 102 |
| S5_vs_85065 | NC_070945.1 | NC_070945.1 | Arthrobacter phage Kardesai | 83.333 | 0 | 120 | 20 | 0 | 8233 | 8352 | 9644 | 9763 | 6.06E-21 | 111 |
| S12_vs_142625 | NC_070849.1 | NC_070849.1 | Alcaligenes phage vB_AfaP_QDWS595 | 97.368 | 0 | 38 | 1 | 0 | 33489 | 33526 | 55292 | 55255 | 6.68E-07 | 65.8 |
| S10_vf_80712 | NC_071044.1 | NC_071044.1 | Bacillus phage vB_BanS_Nate | 96.491 | 0 | 57 | 2 | 0 | 114841 | 114897 | 147174 | 147118 | 1.23E-15 | 95.3 |
| S5_vs_63924 | NC_073367.1 | NC_073367.1 | Microbacterium phage Terij | 75 | 1 | 360 | 74 | 16 | 15072 | 15423 | 39793 | 40144 | 2.2E-33 | 152 |
| S3_vf_70746 | NC_073428.1 | NC_073428.1 | Microbacterium phage Quenya | 78.421 | 1 | 380 | 72 | 10 | 24012 | 24386 | 10177 | 10551 | 1.42E-59 | 239 |
| S13_vs_149860 | NC_073478.1 | NC_073478.1 | Bifidobacterium phage BD811P2 | 78.539 | 27 | 3299 | 661 | 41 | 12109 | 15383 | 6710 | 3435 | 0 | 2126 |
| S7_vf_69348 | NC_073482.1 | NC_073482.1 | Clostridium phage CI55P1 | 87.906 | 77 | 8938 | 1002 | 57 | 1652 | 10554 | 1326 | 10219 | 0 | 10443 |
| S11_vf_92391 | NC_073481.1 | NC_073481.1 | Clostridium phage CI461P1 | 93.525 | 3 | 278 | 16 | 2 | 14048 | 14324 | 13406 | 13682 | 4.1E-112 | 412 |
| S2_vf_20910 | NC_073481.1 | NC_073481.1 | Clostridium phage CI461P1 | 95.378 | 2 | 238 | 11 | 0 | 14580 | 14817 | 13659 | 13896 | 4.1E-102 | 379 |
